# Predominance of the SARS-CoV-2 lineage P.1 and its sublineage P.1.2 in patients from the metropolitan region of Porto Alegre, Southern Brazil in March 2021: a phylogenomic analysis

**DOI:** 10.1101/2021.05.18.21257420

**Authors:** Vinícius Bonetti Franceschi, Gabriel Dickin Caldana, Christiano Perin, Alexandre Horn, Camila Peter, Gabriela Bettella Cybis, Patrícia Aline Gröhs Ferrareze, Liane Nanci Rotta, Flávio Adsuara Cadegiani, Ricardo Ariel Zimerman, Claudia Elizabeth Thompson

## Abstract

Almost a year after the COVID-19 pandemic had begun, The United Kingdom, South Africa, and Brazil became the epicenter of new lineages, the Variant of Concern (VOCs), B.1.1.7, B.1.351, and P.1, respectively. These VOCs are increasingly associated with enhanced transmissibility, immunity evasion, and mortality. The previous most prevalent lineages in the state of Rio Grande do South (Brazil), B.1.1.28 and B.1.1.33 were rapidly replaced by P.1 and P.2, two B.1.1.28-derived lineages harboring the E484K mutation. To perform a genomic characterization of SARS-CoV-2 samples from COVID-19 patients from the metropolitan region of Porto Alegre (Rio Grande do Sul, Southern Brazil), in this second pandemic wave, we sequenced viral samples from patients of this region to: (i) identify the prevalence of SARS-CoV-2 lineages in the region, the state and bordering countries/states, (ii) characterize the mutation spectra, and (iii) hypothesize possible viral dispersal routes by using phylogenetic and phylogeographic approaches. As results, we not only confirmed that 96.4% of the samples belonged to the P.1 lineage but also that approximately 20% of which could be assigned as the newer P.1.2 (a P.1 derived new sublineage harboring new signature substitutions recently described and present in other Brazilian states and foreign countries). Moreover, P.1 sequences from this study were allocated in several distinct branches (four clades and five clusters) of the P.1 phylogeny, suggesting multiple introductions of P.1 in Rio Grande do Sul still in 2020 and placing this state as a potential core of diffusion and emergence of P.1-derived clades. It is still uncertain if the emergence of P.1.2 and other P.1 clades are related to further virological, clinical, or epidemiological consequences. However, the clear signs of viral molecular diversification from recently introduced P.1 warrant further genomic surveillance.

## Introduction

After its initial emergence in Wuhan, China, in late 2019, the Severe acute respiratory syndrome coronavirus 2 (SARS-CoV-2) spread rapidly around the world leading to the COVID-19 pandemic officially recognized in March 2020 (1). As of May 17, 2021, >163 million cases and >3.3 million deaths were confirmed. In Brazil, the third country most affected by COVID-19, >15.6 million cases and >435.000 deaths were reported. From a virological standpoint, this could be related to the continental magnitude of Brazil, leading to multiple viral introductions (2) and the recent emergence of novel “ Variant of Concern” (VOC) presenting enhanced infectiousness.

Rio Grande do Sul (RS) is the southernmost state in Brazil. It is bordered southerly by Uruguay, westerly by Argentina, and northerly by the state of Santa Catarina, Brazil. With an estimated population of 11.5 million inhabitants and 39.79 inhabitants per square kilometer, RS is the sixth most populous and the 13th most densely populated state in the country (3). Since 2017, the Brazilian Institute of Geography and Statistics (IBGE) has divided RS into eight intermediate geographic regions: Porto Alegre, Pelotas, Uruguaiana, Santa Maria, Santa Cruz do Sul / Lajeado, Ijuí, Passo Fundo, and Caxias do Sul (4). The municipality of Porto Alegre is the state capital, and its metropolitan region comprises 34 municipalities aggregating >4 million inhabitants (∼2% of the country’s population) and is characterized by intense transit of people. COVID-19 was firstly confirmed in RS on March 10, 2020, in a returning traveler from Italy (5). The state implemented on May 2020 the “ controlled distancing system”, which divided the state into 21 regions and 26 areas (R01 to R26) and consisted of a flag system establishing restrictions and flexibilizations of non-essential activities based on the weekly occupation of intensive care unit beds and expected deaths. However, due to economic losses, the more amenable “ shared management system” allowed mayors to appeal in court and adopt less restrictive flag protocols (6).

Important shifts of COVID-19 epicenters have occurred during 2020, starting with Asia and followed by Europe, North America, and South America. After months of relatively slow evolution, novel VOCs (e. g., B.1.1.7, B.1.351, and P.1) harboring a constellation of signature mutations in the Spike protein have emerged. This phenomenon independently arose in the United Kingdom (7), South Africa (8), and Brazil (9,10) and has fueled secondary outbreaks in places where they have emerged, despite previous rates of seroprevalence of up to 75% (11). The city of Manaus (Amazonas, Brazil), the probable place of origin of P.1 lineage, faced a major second wave of COVID-19. An explosive resurgence of cases and deaths became evident in mid-December 2020. Since the P.1 variant carries multiple mutations of potential biological significance (especially E484K, K417T, and N501Y in the Receptor-Binding Domain [RBD] from spike protein), (i) many key substitutions may lead to the immunity evasion, (ii) higher transmissibility when compared with pre-existing lineages have been characterized, (iii) this VOC has been the focus of increased surveillance and has deserved being studied in greater detail (12). After this outbreak, almost all Brazilian states experienced increases in the number of cases, hospitalizations, intensive care unit (ICU) admissions, and deaths, resulting in a reemergence of the public health crisis previously experienced in the first wave of COVID-19 (13).

The diversity of SARS-CoV-2 during the first epidemic wave in Brazil was mainly composed of B.1.1.28 and B.1.1.33 lineages (2,14), although the very low sequencing rate across the country has limited these estimates (14). However, these previous lineages were rapidly replaced by P.1 and P.2, both derived from the common ancestor B.1.1.28 and harbor concerning mutations in the Spike protein (*e*.*g*., E484K and N501Y), from late 2020 and early 2021 (14,15). In the RS state, the most common lineages identified to May 2021 still are B.1.1.33 (n=290) and B.1.1.28 (n=238). Nevertheless, P.1 has emerged as the most prevalent lineage sequenced in more recent samples (16). Recently, newer mutations were detected in addition to the original set presented in P.1, giving rise to the sublineage P.1.2 (17). P.2 probably emerged in the Rio de Janeiro state (Southeast) (18), but was also found in several municipalities of the RS state as of October 2020 (19,20). The first P.1 infection in the state was once thought to be in a patient of Gramado city in February 2021 (21). However, in a more recent study, the actual first P.1 was detected on November 30. This happened in a patient with comorbidities from Campo Bom city, who were reinfected by the P.2 lineage on March 11 (22).

Even though RS was one of the least affected Brazilian states in the first epidemic wave, it suffered a pronounced increase in cases in late 2020 (13). In February 2021, the progressive increases in cases and hospitalizations (3.8-fold) led to the collapse of the local state healthcare system. Since recent findings of the widespread dissemination of the SARS-CoV-2 lineage P.1 in Brazil have been confirmed, we sequenced samples from patients from the metropolitan region of Porto Alegre to (i) identify the prevalence of SARS-CoV-2 lineages in the region, the state and bordering countries/states, (ii) characterize the mutation spectra, and (iii) hypothesize possible viral dispersal routes by using phylogenetic and phylogeographic approaches.

## Methods

### Ethics approval and consent to participate

Ethical approval was obtained from the Brazilian’s National Ethics Committee (Comissão Nacional de Ética em Pesquisa — CONEP) under process number CAAE 41909121.0.0000.5553 and Comitê de Ética em Pesquisa em Seres Humanos da Universidade Federal de Ciências da Saúde de Porto Alegre (CEP - UFCSPA) under process number CAAE 35083220.2.0000.5345. The study was performed in accordance with the Declaration of Helsinki. Patients included in the clinical trial approved by CONEP were informed in detail about the study and gave written informed consent to participate. All samples belonging to the Hospital da Brigada Militar patients that yielded positive RT-qPCR had their laboratory electronic records reviewed to compile metadata such as date of collection, sex, age, symptoms, exposure history, and clinical status, when available. Samples were anonymized before being received by the study investigators, following Brazilian and international ethical standards.

### Sample collection and clinical testing

Samples were obtained from Hospital da Brigada Militar patients, both admitted or visiting the emergency ward, from Porto Alegre, RS, Brazil. Nasopharyngeal swabs were collected and placed in saline solution. Samples were transported to the clinical laboratory (Laboratório Exame) and tested on the same day for SARS-CoV-2 using Real Time Reverse-transcriptase Polymerase Chain Reaction (Charité RT-qPCR assays). The RTq-PCR assay used primers and probes recommended by the World Health Organization targeting the Nucleocapsid (N1 and N2) genes (23). Remnant samples were stored at -20°C.

Between March 9th to March 17th, all the routinely tested samples of the clinical laboratory provenient of the Hospital da Brigada Militar patients and yielded positive RT-qPCR were selected. Subsequently, those positive clinical samples were submitted to a second RT-qPCR performed by BiomeHub (Florianópolis, Santa Catarina, Brazil), using the same protocol (*charite-berlin*). Only samples with quantification cycle (Cq) below 30 for at least one primer were submitted to the SARS-CoV-2 genome sequencing. In total, 56 patients who presented symptoms such as fever, cough, sore throat, dyspnea, anosmia, fatigue, diarrhea, and vomiting (moderate and severe clinical status) (24) were included in the study.

### RNA extraction, library preparation, and sequencing

Total RNAs were prepared as in the reference protocol (25) using SuperScript IV (Invitrogen, Carlsbad, USA) for cDNA synthesis and Platinum Taq High Fidelity (Invitrogen, Carlsbad, USA) for specific viral amplicons. Subsequently, cDNA was used for the library preparation with Nextera Flex (Illumina, San Diego, USA) and quantified with Picogreen and Collibri Library Quantification Kit (Invitrogen, Carlsbad, USA). The sequencing was performed on the Illumina MiSeq (Illumina, San Diego, USA) 150×150 runs with 500xSARS-CoV-2 coverage (50-100 thousand reads/per sample).

### Quality control and consensus calling

Quality control, reference mapping, and consensus calling were performed using an in-house pipeline developed by BiomeHub (Florianópolis, Santa Catarina, Brazil). Briefly, adapters were removed and reads were trimmed by size=150. Reads were mapped to the reference SARS-CoV-2 genome (GenBank accession number NC_045512.2) using Bowtie v2.4.2 (end-to-end and very-sensitive parameters) (26). Mapping coverage and depth were retrieved using samtools v1.11 (27) (minimum base quality per base (Q) ≥ 30). Consensus sequences were generated using bcftools mpileup (Q ≥ 30; depth (d) ≤ 1000) combined with bcftools filter (DP>50) and bcftools consensus v1.11 (28). Coverage values for each genome were plotted using the karyoploteR v1.12.4 R package (29). Finally, we assessed the consensus sequences quality using Nextclade v0.14.2 (https://clades.nextstrain.org/).

### Mutation analysis

Single Nucleotide Polymorphisms (SNPs) and insertions/deletions in each sample were identified using snippy variant calling and core genome alignment pipeline v4.6.0 (https://github.com/tseemann/snippy), which uses FreeBayes v1.3.2 (30) to call variants and snpEff v5.0 (31) to annotate and predict their effects on genes and proteins. Genome map and SNP histogram were generated after running MAFFT v7.475 (32) alignment using the msastats.py script, and plotAlignment and plotSNPHist functions (33). Sequence positions refer to GenBank RefSeq sequence (NC_045512.2), isolated and sequenced from an early case from Wuhan (China) in 2019.

We identified global virus lineages using the dynamic nomenclature implemented in Pangolin v2.3.8 (26; https://github.com/cov-lineages/pangolin) and global clades and mutations using Nextclade v0.14.2 (https://clades.nextstrain.org/). We also used the Pathogenwatch (https://pathogen.watch/) and Microreact (35) to explore mutations and lineages across time and geography initially.

### Maximum likelihood phylogenomic analysis

All available SARS-CoV-2 genomes (1,048,519 sequences) were obtained from GISAID on April 26, 2021 and combined with our 56 sequences to obtain a global representative dataset. These sequences were subjected to analysis inside the NextStrain ncov pipeline (27; https://github.com/nextstrain/ncov). In this workflow, sequences were aligned using nextalign v0.1.6 (https://github.com/neherlab/nextalign). In the initial filtering step, short and low-quality sequences or those with incomplete sampling dates were excluded. Uninformative sites and ends (100 positions in the beginning, 50 in the end) were also masked from the alignment. Genetically closely related genomes to our focal subset were selected, prioritizing sequences geographically closer to Brazil’s state RS. The maximum likelihood (ML) phylogenetic tree was built using IQ-TREE v2.1.2 (37), employing the General time-reversible (GTR) model with unequal rates and base frequencies (38). The tree’s root was placed between lineage A and B (Wuhan/WH01/2019 and Wuhan/Hu-1/2019 representatives), and sequences that deviate more than four interquartile ranges from the root-to-tip regression of genetic distances against sampling dates were excluded from the analysis.

A time-scaled ML tree was generated with TreeTime v0.8.1 (39) under a strict clock under a skyline coalescent prior with a mean rate of 8×10^−4^ substitutions per site per year. Finally, clades and mutations were assigned and geographic movements inferred. The results were exported to JSON format to enable interactive visualization through Auspice. Additionally, as P.1 sequences mostly represent our dataset, we downloaded all complete and high-quality global genomes assigned to P.1 PANGO lineage (4,499 sequences) submitted until April 26th, 2021. These sequences were aligned using MAFFT v7.475, the ends of the alignment (300 in the beginning and 500 in the end) were masked, and the ML tree was built with IQ-TREE v2.0.3 using the GTR+F+R3 nucleotide substitution model as selected by the ModelFinder (40). Branch support was calculated using the Shimodaira-Hasegawa approximate likelihood ratio test (SH-aLRT) (41) with 1,000 replicates.

Local sequences were classified according to the following scheme: monophyletic clades composed by one local genome were classified as “ isolated”, while clades composed by 2 < genomes < 4 were considered “ clusters” and if ≥ 4 local genomes represented, we assigned a “ clade” designation.

ML trees were inspected in TempEst v1.5.3 (42) to investigate the temporal signal through regression of root-to-tip genetic divergence against sampling dates. For the P.1 ML tree, samples with missing days of the collection were filled with the 15th day of the month. ML and time-resolved trees were visualized using FigTree v1.4.4 (http://tree.bio.ed.ac.uk/software/figtree/) and ggtree R package v2.0.4 (43).

### Discrete Bayesian phylogeographic and phylodynamic analysis

Considering the four identified clades composed by ≥ 4 sequences from this study, we extracted the clade members using the caper R package v1.0.1 (44). Clade-specific ML trees and root-to-tip regression assignments were generated as described above. Evolutionary parameter estimates and spatial diffusion were estimated separately for each clade using a Bayesian Markov Chain Monte Carlo (MCMC) approach implemented in BEAST v10.4 (45) using the BEAGLE library (46) to enhance computational time. Time-stamped Bayesian trees were generated using the HKY+Γ nucleotide model (47), a strict molecular clock model with a Continuous Time Markov Chain (CTMC) rate reference prior (48) (mean rate = 8×10^−4^) and a non-parametric skygrid tree prior (49) with grid points defined by the approximate number of weeks spanned by the duration of the phylogeny.

The MCMC chains were run in duplicates for at least 50 million generations, and convergence was checked using Tracer v1.7.1 (50). Log and tree files were combined using LogCombiner v1.10.4 to ensure stationarity and good mixing (43) after removing 10% as burn-in. Maximum clade credibility (MCC) was generated using TreeAnnotator v1.10.4 (45). Viral migrations were reconstructed using a reversible discrete asymmetric phylogeographic model (51) to infer the locations of the internal nodes of the tree. A discretization scheme with a resolution of different Brazilian states and other countries was applied. Location diffusion rates were estimated using the Bayesian stochastic search variable selection (BSSVS) (51) procedure employing Bayes factors to identify well-supported rates.

### Geoplotting

Geographical maps and other plots were generated using R v3.6.1 (52), and the ggplot2 v3.3.2 (53), geobr v.1.4 (54), and sf v0.9.8 (55) packages. For the discrete phylogeographic analysis, the SpreaD3 v0.9.7.1 (56) was used to map spatiotemporal information embedded in MCC trees.

## Results

### Epidemiological information

From the 56 samples of hospitalized patients between March 09-17 2021, 75.0% (n=42) of them were male, and the mean age was 37.2 years (interquartile range (IQR): 13.5 years). The mean cycle threshold (Ct) value for the first RT-qPCR conducted at Laboratório Exame was 19.12 cycles (median: 18.00, IQR: 6.00 cycles). Forty-seven (83.9%) had contact with a confirmed or suspected case. The majority of them were from the RS state capital, Porto Alegre (n=32; 57.1%). In total, 51 (91.1%) were from the intermediate geographic region of Porto Alegre and 5 (8.9%) from the intermediate region of Santa Maria (Table 1, Figure S2C).

**Table 1.** Epidemiological characteristics of the 56 sequenced samples from Rio Grande do Sul, Southern Brazil

| Study ID (HBM-RS) | GISAID ID (EPI_ISL_) | Cycle threshold | Municipality of residence | Gender | Age group | Lineage | Contact with confirmed case |
| --- | --- | --- | --- | --- | --- | --- | --- |
| 39468 | 2139494 | 16 | São Leopoldo | Male | 30-39 | P.1 | Yes |
| 39469 | 2139495 | 19 | Porto Alegre | Female | 20-29 | P.1.2 | Yes |
| 39470 | 2139496 | 19 | Porto Alegre | Male | 60-69 | P.1 | No |
| 39471 | 2139497 | 18 | Porto Alegre | Male | 20-29 | P.1 | Yes |
| 39472 | 2139498 | 17 | Gravataí | Male | 30-39 | P.1 | No |
| 39473 | 2139499 | 26 | Cachoeira do Sul | Female | 20-29 | P.1 | Yes |
| 39474 | 2139500 | 18 | Gravataí | Male | 30-39 | P.1 | Yes |
| 39475 | 2139501 | 18 | Porto Alegre | Female | 20-29 | P.1 | No |
| 39476 | 2139502 | 15 | Porto Alegre | Male | 20-29 | P.1 | Yes |
| 39477 | 2139503 | 21 | Porto Alegre | Male | 30-39 | P.1 | Yes |
| 39478 | 2139504 | 15 | Cachoeira do Sul | Male | 30-39 | P.1 | Yes |
| 39479 | 2139505 | 22 | Porto Alegre | Male | 50-59 | P.1 | Yes |
| 39480 | 2139506 | 17 | Novo Hamburgo | Male | 40-49 | P.1 | Yes |
| 39481 | 2139507 | 14 | Porto Alegre | Female | 70-79 | P.1 | Yes |
| 39482 | 2139508 | 14 | Porto Alegre | Female | 80-89 | P.1.2 | No |
| 39483 | 2139509 | 13 | Gravataí | Male | 30-39 | P.1 | Yes |
| 39484 | 2139510 | 20 | Porto Alegre | Male | 20-29 | P.1 | Yes |
| 39485 | 2139511 | 16 | Porto Alegre | Male | 50-59 | P.1 | Yes |
| 39486 | 2139512 | 27 | Porto Alegre | Male | 30-39 | P.2 | No |
| 39487 | 2139513 | 14 | São Sebastião do Caí | Male | 40-49 | P.1.2 | Yes |
| 39488 | 2139514 | 28 | Santo Antônio da Patrulha | Male | 70-79 | P.1 | Yes |
| 39489 | 2139515 | 27 | Porto Alegre | Female | 20-29 | P.1.2 | Yes |
| 39490 | 2139516 | 18 | Porto Alegre | Male | 20-29 | P.1 | Yes |
| 39491 | 2139517 | 15 | Alvorada | Female | 20-29 | B.1.1.28 | Yes |
| 39492 | 2139518 | 17 | Gravataí | Female | 30-39 | P.1 | Yes |
| 39493 | 2139519 | 22 | Canoas | Male | 30-39 | P.1.2 | Yes |
| 39494 | 2139520 | 17 | Porto Alegre | Female | 30-39 | P.1 | No |
| 39495 | 2139521 | 17 | Porto Alegre | Male | 30-39 | P.1 | Yes |
| 39496 | 2139522 | 17 | Canoas | Female | 30-39 | P.1 | Yes |
| 39497 | 2139523 | 21 | Porto Alegre | Male | 40-49 | P.1.2 | Yes |
| 39498 | 2139524 | 20 | Porto Alegre | Female | 40-49 | P.1 | Yes |
| 39499 | 2139525 | 22 | Portão | Male | 30-39 | P.1 | Yes |
| 39500 | 2139526 | 11 | Porto Alegre | Male | 20-29 | P.1.2 | Yes |
| 39501 | 2139527 | 14 | Santa Maria | Male | 20-29 | P.1.2 | Yes |
| 39502 | 2139528 | 21 | Porto Alegre | Male | 30-39 | P.1 | Yes |
| 39503 | 2139529 | 16 | Porto Alegre | Male | 30-39 | P.1 | No |
| 39504 | 2139530 | 21 | Gravataí | Male | 40-49 | P.1 | Yes |
| 39505 | 2139531 | 13 | Porto Alegre | Male | 30-39 | P.1.2 | Yes |
| 39506 | 2139532 | 23 | Porto Alegre | Female | 40-49 | P.1 | Yes |
| 39507 | 2139533 | 28 | Canoas | Female | 30-39 | P.1 | Yes |
| 39508 | 2139534 | 22 | Porto Alegre | Male | 20-29 | P.1 | Yes |
| 39509 | 2139535 | 23 | Alvorada | Male | 20-29 | P.1 | Yes |
| 39510 | 2139536 | 19 | Canoas | Male | 50-59 | P.1.2 | Yes |
| 39511 | 2139537 | 22 | Porto Alegre | Male | 30-39 | P.1 | No |
| 39512 | 2139538 | 25 | Cachoeira do Sul | Female | 40-49 | P.1 | Yes |
| 39513 | 2139539 | 23 | Santa Maria | Male | 40-49 | P.1 | Yes |
| 39514 | 2139540 | 15 | Porto Alegre | Male | 30-39 | P.1 | No |
| 39515 | 2139541 | 21 | Porto Alegre | Male | 20-29 | P.1 | Yes |
| 39516 | 2139542 | 28 | Porto Alegre | Male | 50-59 | P.1 | Yes |
| 39517 | 2139543 | 17 | Sapiranga | Male | 30-39 | P.1 | Yes |
| 39518 | 2139544 | 17 | Porto Alegre | Male | 30-39 | P.1.2 | Yes |
| 39519 | 2139545 | 23 | Porto Alegre | Male | 20-29 | P.1 | Yes |
| 39520 | 2139546 | 15 | Campo Bom | Male | 20-29 | P.1 | Yes |
| 39521 | 2139547 | 15 | Porto Alegre | Male | 20-29 | P.1 | Yes |
| 39522 | 2139548 | 21 | Porto Alegre | Male | 50-59 | P.1 | Yes |
| 39523 | 2139549 | 18 | Porto Alegre | Male | 20-29 | P.1 | Yes |

All samples were nasopharyngeal swabs collected between March 9—17, 2021, from residents of the RS state. Study ID: Study identifier only known by study investigators. The Ct values are related to the first RT-qPCR conducted at Laboratório Exame.

### SARS-CoV-2 mutations and lineages

Consensus SARS-CoV-2 genomes were obtained with an average coverage depth of 813.2× (median: 820.6×, IQR: 184.7×) (Supplementary File 1). We detected 175 different mutations comprising all samples (Figure 1A). The ORF1ab carried 102 (58.3%) replacements followed by spike (n=24; 13.7%), nucleocapsid (n=18; 10.3%) ORF3a (n=14, 8.0%), ORF7a (n=6; 3.43%), ORF8 (n=5; 2.86%), and membrane (n=3; 1.7%) genes. Remarkably, 50% of the spike substitutions occurred in only one genome, and of these, 9 (75.0%) were missense (Supplementary file 2). Fifty-nine (33.7%) mutations were identified in two or more sequences. From these, 36 (61.0%) are non-synonymous (missense), 21 (35.6%) are synonymous, one (1.7%) intergenic at 5’ Untranslated Region (UTR), and one (1.7%) inframe deletion. Highly frequent (≥ 10 genomes) mutations were found in 34 genomic positions, 24 (70.5%) being missense and 9 (26.5%) synonymous. Fifteen substitutions (10 in the spike protein: L18F, T20N, P26S, D138Y, R190S, K417T, E484K, N501Y, H655Y, T1027I) are P.1 lineage-defining mutations (Figure 1B, Table 2). The only P.1 defining replacement not found at high frequency in our study was the deletion in ORF1ab (del:11288:9), called in only four genomes. This result is due to the stringent coverage depth filter applied (DP>50) for calling the genomic positions in the consensus sequences.

**Table 2.**
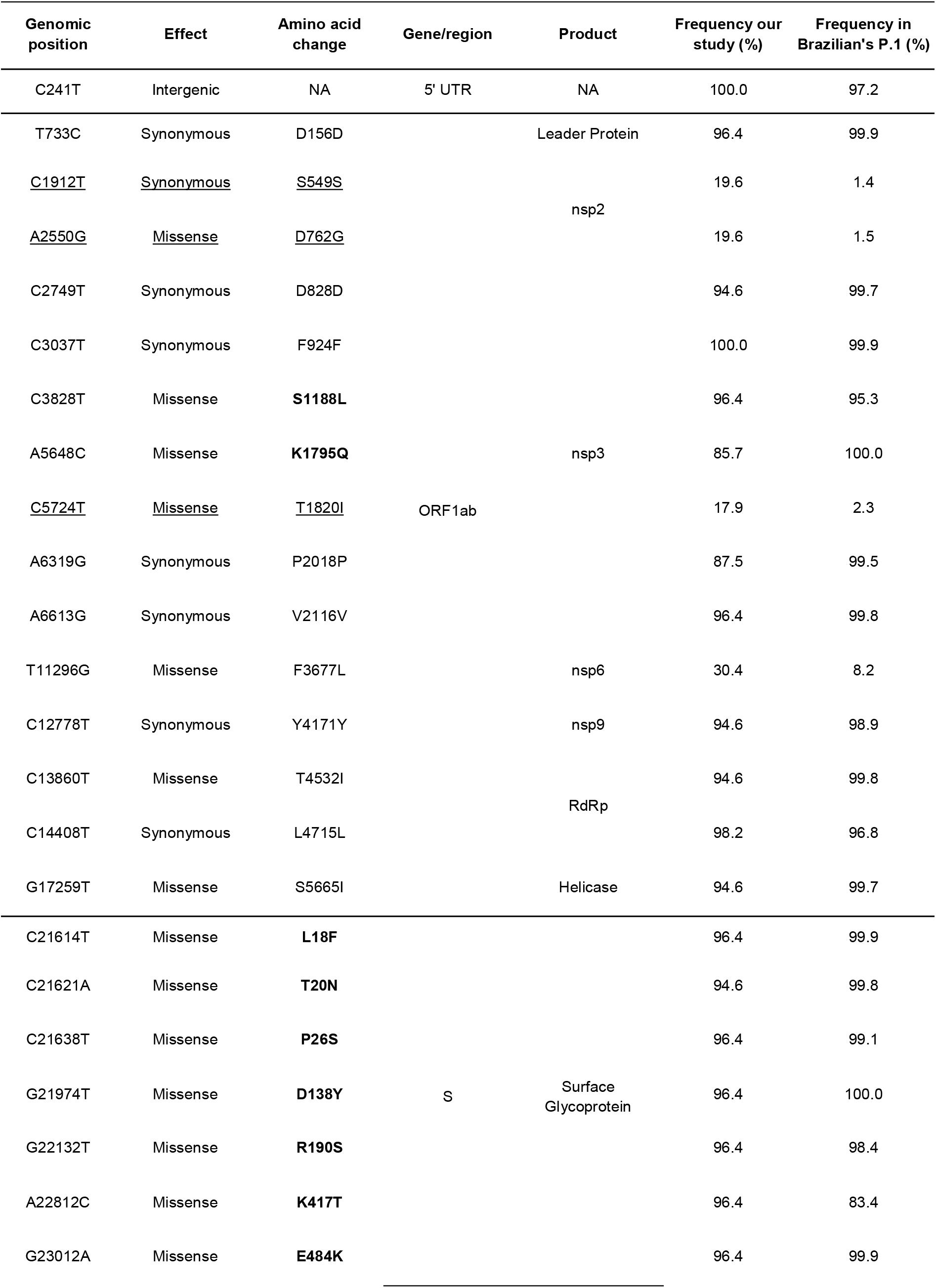
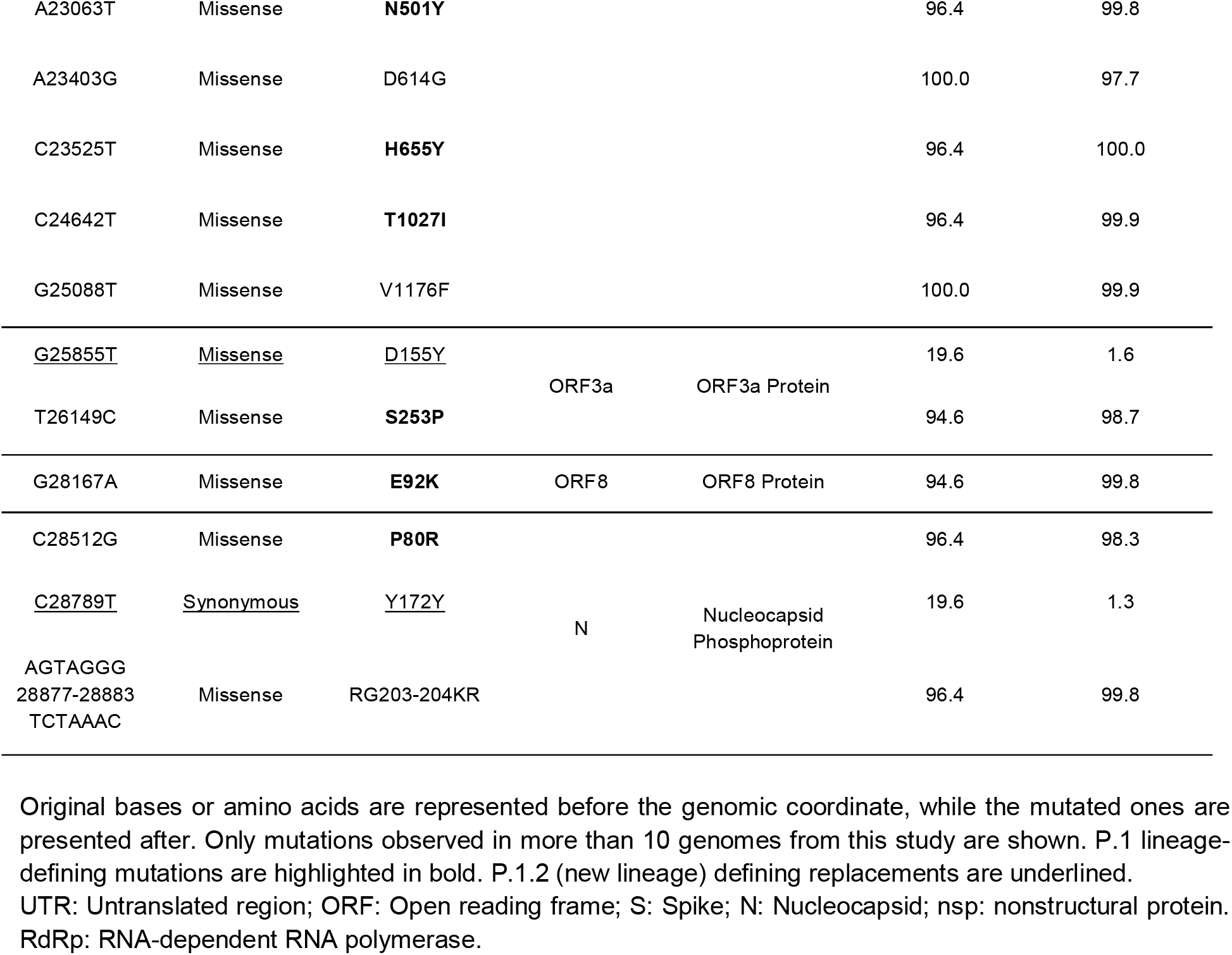
Detailed description and frequency of mutations found in our 56 sequences compared with all Brazilian P.1 sequences until April 26, 2021.

**Figure 1.**
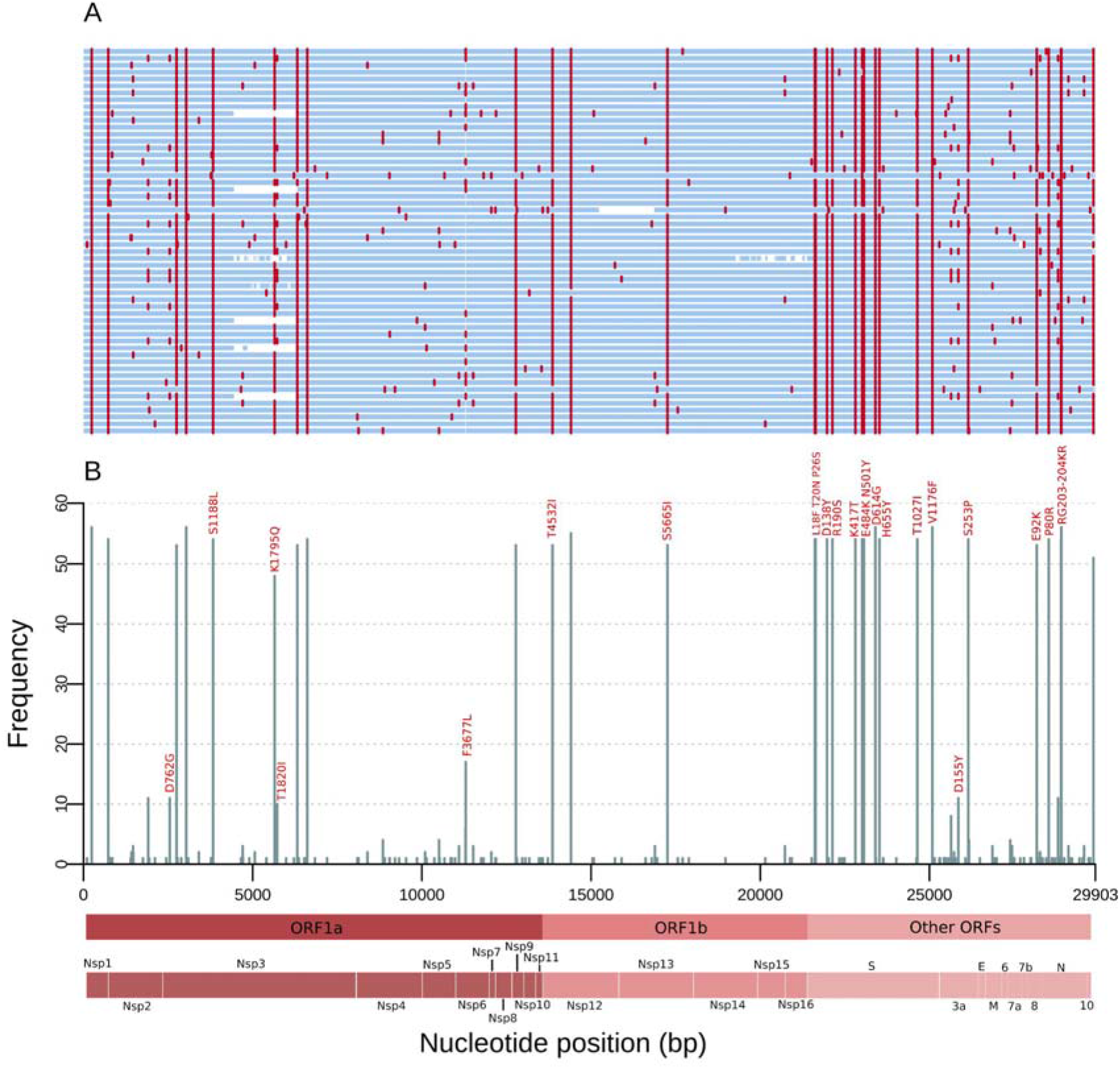
Mutations of the SARS-CoV-2 genomes from the RS state, Southern Brazil sampled in March 2021. (A) Genome map for the 56 genomes sequenced. Nucleotide substitutions are colored in red and blank regions represent low sequencing coverage. (B) Frequency of SNPs per SARS-CoV-2 genome position along the 56 genomes. These mutations are corresponding to the red lines in (A), and only missense substitutions represented by >10 sequences have their respective amino acid changes indicated above the bars. Main Open Reading Frames (ORFs) and SARS-CoV-2 proteins are indicated at the bottom to allow a rapid visualization of the viral proteins affected.

After comparing the frequency of mutations from the recently sequenced samples and the Brazilian P.1 genomes, we observed a combination of mutations that stood out in a significant proportion (n=11; 19.6%) compared with previous P.1 sequences from Brazil. This combination was previously described (17) and gave rise to the P.1.2 lineage, which harbors three ORF1ab replacements (synC1912T, D762G, T1820I), one in ORF3a (D155Y), and one in N protein (synC28789T) (Table 2). Additionally, two of these genomes (18.2%) carry T11296G (ORF1ab nsp6: F3677L) and 8 (72.7%) harbor G25641T (ORF3a: L83F) substitutions. Another cluster, made of four genomes and subsequently named Clade 2, possesses three defining mutations (ORF1ab nsp4: V2862L, synC10507T, ORF3a: M260K) was also detected. This cluster does not fall into a lineage designation at this moment but deserves further monitoring (Table 2, Figure S1).

Considering PANGO lineages, 54 genomes (96.4%) were designated as P.1, one (1.8%) as P.2, and one (1.8%) as B.1.1.28. Even without being classified according to the Pango-designation’s most updated version, the P.1.2 lineage was present in 11/54 (20.4%) of the P.1 sequences (https://github.com/cov-lineages/pango-designation/issues/56) (Figure S1).

### Lineage distribution in neighboring countries and Brazilian regions

The RS state shares borders with Argentina in its west (Figure S2), leading to the transit of people at the frontiers. From March to April 2020, B.1 was the most prevalent lineage in this bordering country. B.1.499 and N.3 were abundant from May to July when the N.5 started to rise and surpassed B.1.499 in November 2020 (Figure 2A). Importantly, N.3 and N.5 are derived from the B.1.1.33 lineage widespread in Brazil (https://cov-lineages.org/lineages.html). The P.2 lineage, which initially emerged in Brazil (18) and derived from another Brazilian disseminated lineage (B.1.1.28), was firstly found in November 2020, and by January and February 2021 it had already outnumbered the other lineages in Argentina (Figure 2A).

**Figure 2.**
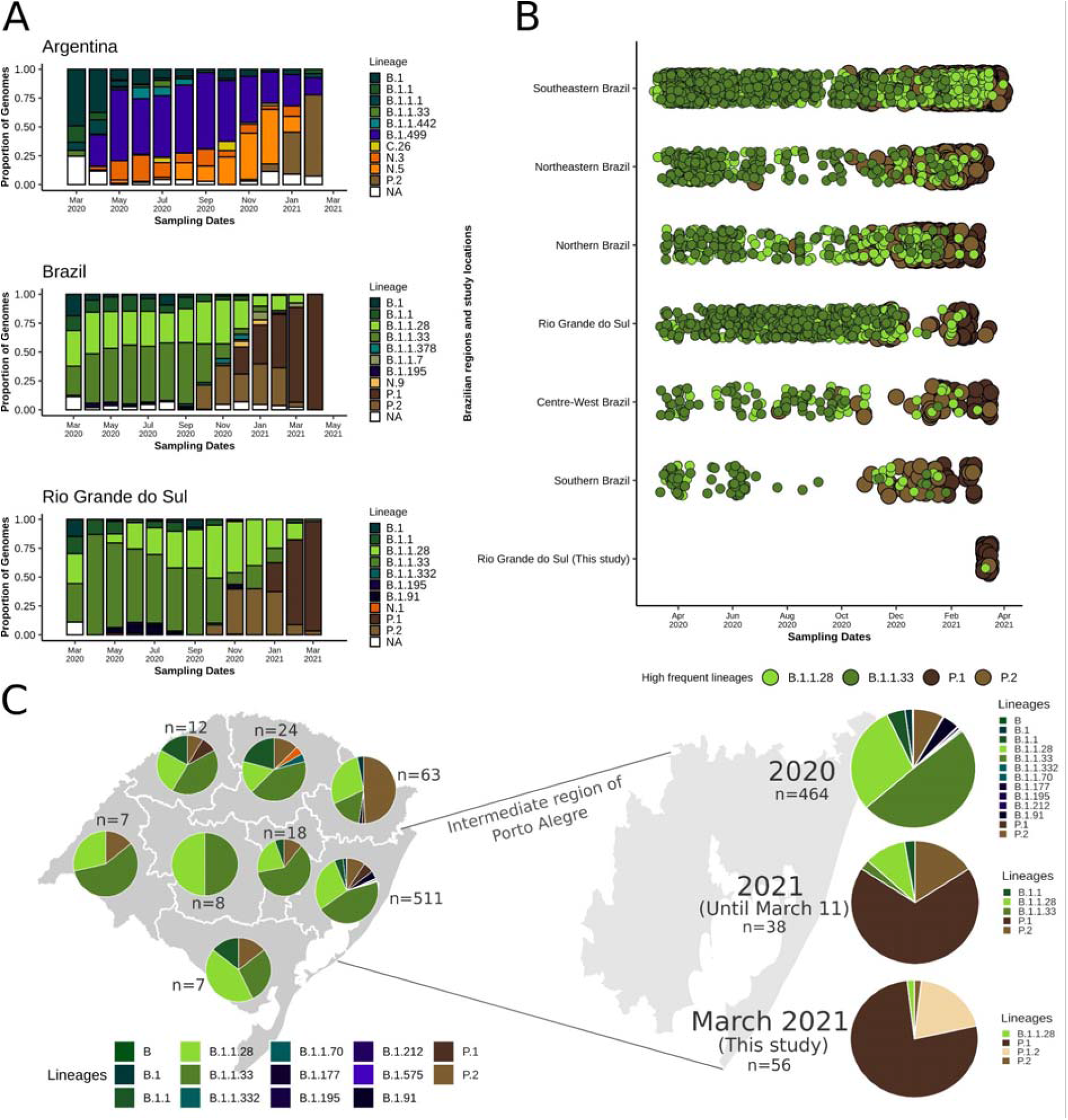
Distribution of SARS-CoV-2 lineages across time in Argentina, other Brazilian regions and in the RS state. (A) Per month lineage distribution in Argentina, entire Brazil and the RS state. Only the 10 most prevalent lineages were considered. (B) Timeline showing the distribution of the most prevalent lineages until the end of 2020 (B.1.1.28 and B.1.1.33) and from the end of 2020 onward (P.1 and P.2) for the fifth Brazilian regions, the RS state and considering only this study (March 2021). (C) Abundance of different lineages in the eight intermediate regions of the RS state defined by IBGE. The region of Porto Alegre was amplified and the frequencies from 2020, 2021 and only considering the present study are presented on the right. NA=Other lineages.

In the entire Brazil, despite early introductions of B.1 and B.1.1, lineages B.1.1.28 and B.1.1.33 were most abundant from March to October 2020. In October, P.2 already represented an important portion of the sequences, and by November it had already surpassed B.1.1.33. In December 2020 and January 2021, with the emergence of P.1, this lineage and P.2 already became the most prevalent, while between February and April P.1 replaced all other lineages (Figure 2A and 2B). Some fluctuations evidently occurred in different Brazilian regions, such as a prevalence of more local lineages (*e. g*. B.1.195 and B.1.1.378 in the Northern region, B.1.1 and N.9 in the Northeast, and B.1.1.7 in the Southeast and Centre-West (Figure S3). In RS, a similar landscape was observed compared to the Brazilian scenario. B.1.1.28 and B.1.1.33 were most prevalent until October 2020, when P.2 emerged and remained until January 2021 along with B.1.1.28 as the most prevalent. After the introduction of P.1 (January 2021), this lineage practically supplanted the others in February and March 2021 (Figure 2A and 2B).

After dividing RS into the intermediate regions proposed by IBGE (Figure S2), it was possible to gain insights into the dynamics of the lineages in the state, despite the low sample size of some regions (Figure 2C). In most regions, the lineages B.1.1.28 and B.1.1.33 were more prevalent, but P.2 was also detected. In fact, in the Caxias do Sul region, more P.2 (n=31; 49.2%) were sequenced in relation to other lineages. Since the Porto Alegre region has a larger sample size, we divided its results by year to check the most recent (2021) evolutionary abundance. In 2020, B.1.1.33 (n=229; 49.3%) and B.1.1.28 (n=137, 29.5%) were the most abundant, followed by P.2 (n=37; 8.0%). In 2021, P.1 (n=26, 68.4%) and P.2 (n=6, 15.8%) have already outperformed the other lineages (Figure 2C). In our study from March 2021, 96.4% of the samples were classified as P.1. We were able to identify a new P.1 sublineage (P.1.2) in 11 (20.4%) genomes from four different municipalities (Porto Alegre, Canoas, São Sebastião do Caí, and Santa Maria), demonstrating the possible diversification of P.1 and its spread within RS (Figure 2C, Figure S1).

### Maximum likelihood phylogenomic analysis

After running the Nextstrain workflow using quality control and subsampling approaches, we obtained a dataset of 8,635 time- and geographical-representative genomes. From these, 861 were from Africa, 1,370 from Asia, 2,219 from Europe, 481 from North America, 218 from Oceania, and 3,486 from South America. Brazil was represented by 2,608 sequences and the RS state by 730 sequences (56 from this study and 674 available in GISAID) (Supplementary file 3).

The time-resolved ML phylogenetic tree confirmed the PANGO lineages assigned since 54 genomes (96.4%) grouped with P.1 representatives, one (1.8%) with B.1.1.28, and one (1.8%) with P.2 sequences. We also observed a strong correlation between genetic distances and sampling dates (R^2^ = 0.71). The P.1 sequences were grouped above the regression line, showing higher evolutionary rates than the other lineages in the SARS-CoV-2 phylogeny as observed in other studies (10,11). We highlighted the most abundant global lineages present in the RS state that passed the quality control criteria (B.1.1 [n=32], P.1 [n=83], P.2 [n=83], B.1.1.28 [n=203], and B.1.1.33 [n=286]). We also noticed the high abundance of B.1.1.28 and B.1.1.33 lineages until October-November 2020, followed by the rise and establishment of P.2 and P.1, respectively (Figure 3A).

**Figure 3.**
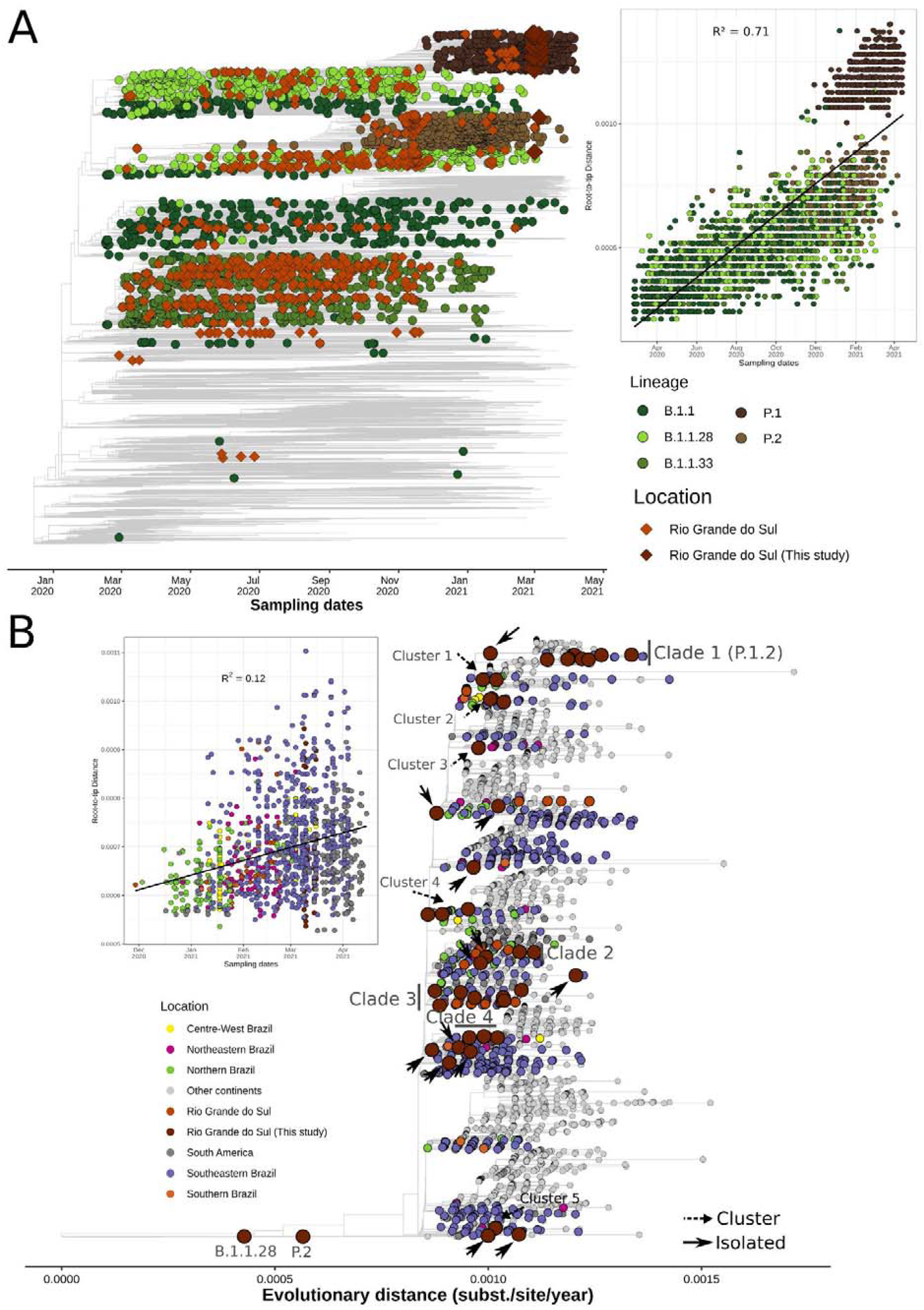
Phylogenetics analysis of genomes sequences in the RS state in global context. (A) Time resolved ML tree of 8,635 global representative SARS-CoV-2 genomes. Circles represent global sequences belonging to the five most abundant lineages in the RS state that passed quality control criteria: B.1.1 (n=32), P.1 (n=83), P.2 (n=83), B.1.1.28 (n=203), B.1.1.33 (n=286), and diamonds represent RS genomes (available in GISAID and sequenced in this study). Root-to-tip regression is represented on the right of the tree. (B) ML tree of 4,499 SARS-CoV-2 genomes belonging to the P.1 lineage. Tips are colored by Brazilian regions, South America or Other continents. Introductions, clusters and clades are annotated in the tree (see Methods). Root-to-tip regression is depicted on the left of the tree and sequences from “ Other continents” were dropped to improve visualization.

To get a more detailed understanding of the P.1 diffusion throughout Rio Grande do Sul, other Brazilian regions, and worldwide countries, we built a ML tree of 4,499 genomes belonging to this lineage (Supplementary file 4). P.1 sequences from this study were allocated in several distinct branches, suggesting multiple introductions and the formation of different P.1-derived clades and clusters.

We identified four clades, five clusters, and 13 isolated sequences (Figure 3B, Supplementary file 5). Most importantly, clade 1 was composed of 11 sequences originated in this study that shared five lineage-defining mutations as previously described (Table 2) and were recently attributed to the P.1.2 sublineage (https://github.com/cov-lineages/pango-designation/issues/56). As of April 26, 2021, this sublineage is already distributed worldwide in 93 sequences (the Netherlands, Spain, England, and the USA) and in other Brazilian states (Rio de Janeiro [RJ] and São Paulo [SP]) (17). Clade 2 sequences harbored two mutations in the ORF1ab:V2862L (nsp4) and synC10507T, one in the ORF3a:M260K, and comprised 81 genomes. Four samples are from this study. The majority is from Amazonas (n=15), São Paulo (n=11), RS (n=8), Bahia (BA) (n=4), and worldwide sequences are mainly represented by French Guiana, USA, Spain, Japan, and Jordan. Clade 3 is represented by three ORF1ab mutations (synC1420T, D1600N [nsp3], and synT8392A) in 3 of 7 local genomes. It is composed of sequences from RS (n=25), SP (n=15), Maranhão (n=10), RJ (n=8), as well as other countries (mainly Spain, French Guiana, and the USA). Clade 4 is characterized by two ORF1ab substitutions (G400S [nsp2] and S6822I [2’-O-ribose methyltransferase]), one N:synT26861C in three genomes and carries other additional mutations (ORF1ab: synG10096A, G3676S [nsp6], F3677L [nsp6]) and M:synT26861C. This clade is mainly found in SP (n=11), RS (n=7), Santa Catarina (n=5), BA (n=4), Goiás (n=4), and other countries (mainly USA, Chile, and England).

Clusters 1 and 3 have respectively one (ORF1ab: G3676S [nsp6]) and two (ORF1ab: synC1471T, A1049V [nsp3]) shared mutations. Among all identified clusters, the most diverse was cluster 5, which contains three samples from this study and has five defining mutations: four in ORF1ab (synT4705C, synC11095T, syn11518, T5541I [helicase]) and one in ORF7a: E16D. Moreover, two sequences share one distinct mutation (ORF1ab: F3677L [nsp6]).

### Bayesian molecular clock and phylogeographic analysis

To date, the time of the most recent common ancestor (TMRCA) and the diffusion of the four P.1 clades identified in our ML analysis, we used coalescent and phylogeographic methods. For clade 1, which is correspondent to the recently labeled P.1.2 lineage, sequences showed a strong correlation of genetic distances and sampling dates (correlation coefficient: 0.59, R^2^ = 0.34) (Figure 4A). We estimated a median evolutionary rate of 7.68×10^−4^ (95% Highest Posterior Density interval [HPD]: 4.18×10^−4^ to 1.14×10^−3^ subst/site/year) and the TMRCA on December 18, 2020 (95% HPD: October 29, 2020 to January 31, 2021). Interestingly, the tree’s root was placed in RS, between a sequence from RS (the oldest sequence from this clade: EPI_ISL_983865) and a subclade from the USA. The divergence of these subclades was dated on January 15, 2021 (95% HPD: January 15 to March 26, 2021). The subclade composed by the RS sequences formed two separate clusters, one with three sequences from this study and one Australian genome and another composed of sequences from RS, SP, UK, Portugal, USA, and transmission clusters from RJ and Netherlands (Figure 4B). The emergence of an important cluster in RJ carrying additional mutations (17) was dated here on March 11, 2021 (95% HPD: March 11 to April 6, 2021). As USA sequences formed a separate subclade, local transmission is probably occurring in the country. The divergence of the USA subclade was dated February 7, 2021 (95% HPD: February 1 to May 11, 2021). Although the BSSVS procedure identified well-supported rates of diffusion from Rio Grande do Sul to other Brazilian states such as São Paulo (Bayes Factor [BF]: 6.82, posterior probability [PP]: 0.52), Rio de Janeiro (BF: 39.18, PP: 0.86), and other countries such as USA (BF: 31.94, PP: 0.84) and Netherlands (BF: 80.16; PP: 0.93), it is possible that this lineage emerged in another Brazilian state but its earlier representatives were not sampled. This is a strong hypothesis since this sequence is associated with community transmission after contact with tourists in a city of RS (Gramado) that receives numerous visitors annually (21).

**Figure 4.**
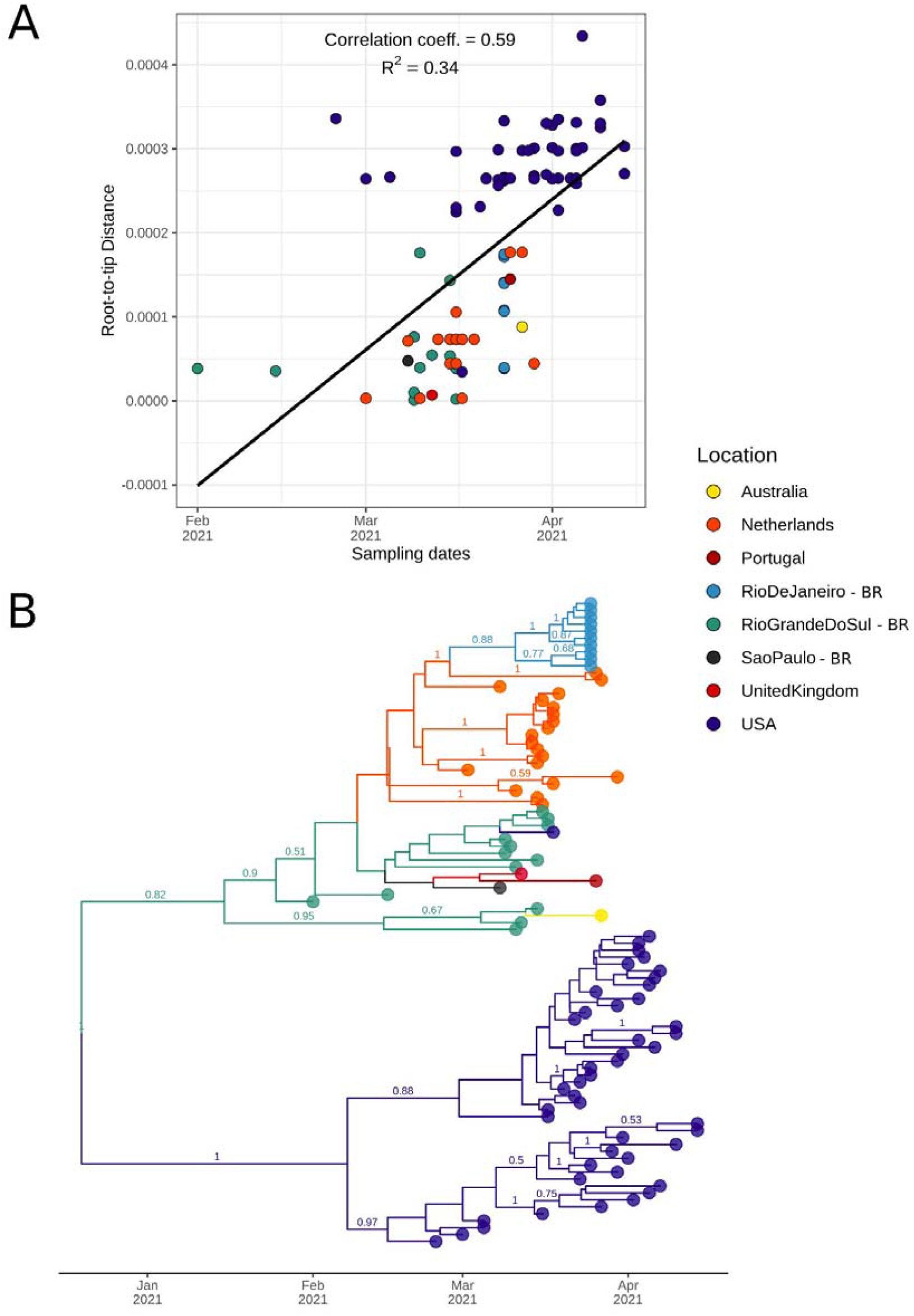
Bayesian discrete asymmetric phylogeographic analysis of the identified Clade 1 (lineage P.1.2). (A) Root-to-tip regression of genetic distances and sampling dates for Clade 1. Correlation coefficient and R squared are depicted above the graph. (B) MCC tree of the 93 sequences included in this analysis up to 26 April, 2021 (82 from GISAID and 11 from this study). Numbers above branches represent the posterior probability of each branch. Only posteriors > 0.5 are shown. Circles indicate countries outside Brazil and Brazilian states (BR suffix).

For clade 2, we estimated a median evolutionary rate of 5.×10^−4^ (95% HPD: 4.18×10^−4^ to 7.71×10^−4^ subst/site/year), and the TMRCA was dated November 30, 2020 (95% HPD: November 2 to December 21, 2020). This clade includes sequences from 11 Brazilian states from all five regions and 9 other countries. We were able to detect at least five introductions from Amazonas, where this clade probably emerged. These introductions ranged from December 28, 2020 (95% HPD: December 28, 2020 to January 5, 2021) to January 28, 2021 (95% HPD: January 28 to March 7, 2021). Importantly, we identified a well-supported subclade (PP = 1) of four genomes from this study (Figure 5A).

**Figure 5.**
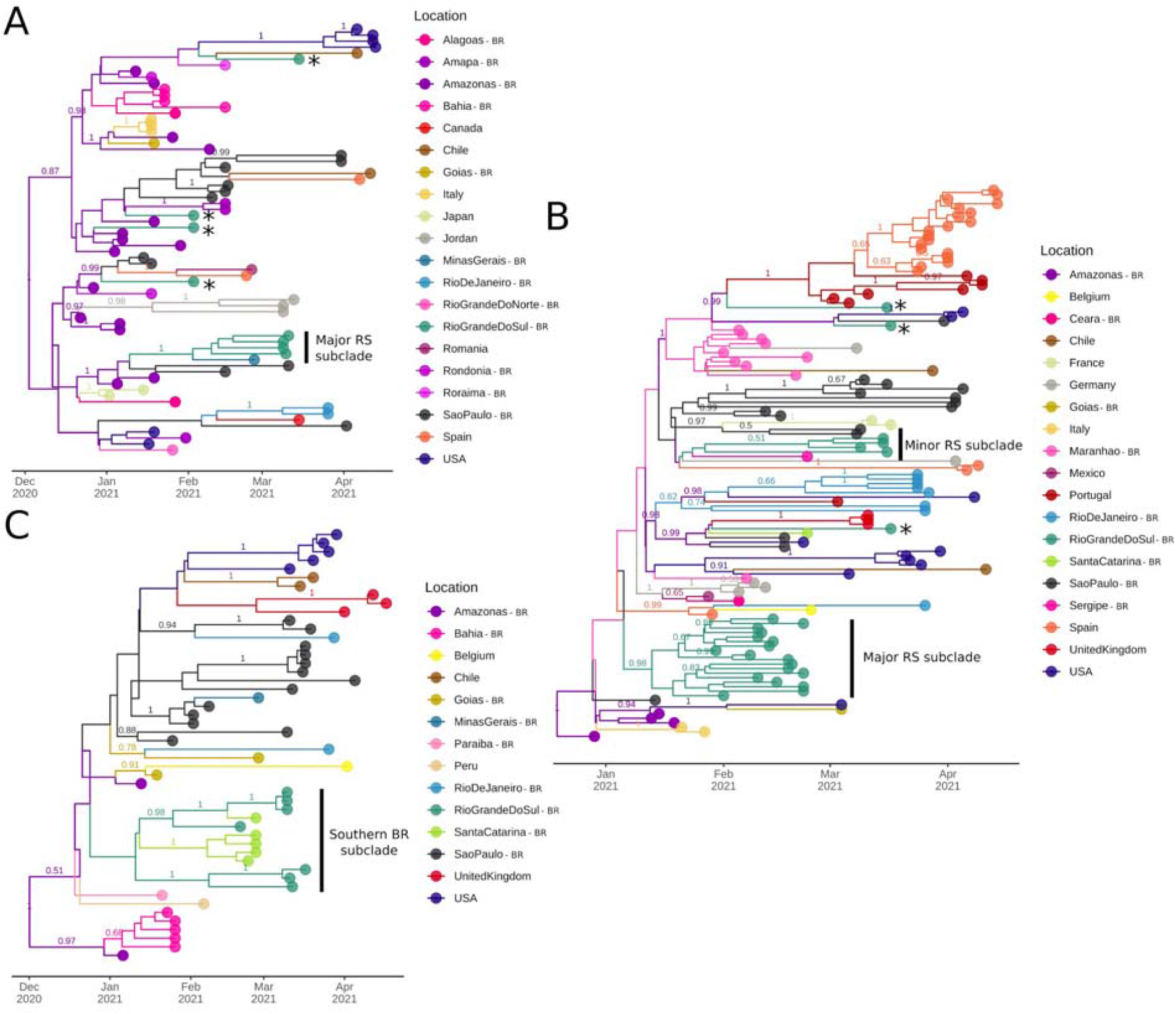
Bayesian discrete asymmetric phylogeographic analysis of the identified clades 2, 3 and 4. (A) MCC tree of the 71 sequences included in this analysis up to 26 April, 2021. (A) MCC tree of the 122 genomes included in this analysis up to 26 April, 2021. (A) MCC tree of the 50 sequences included in this analysis up to 26 April, 2021. For all MCC trees, numbers above branches represent the posterior probability of each branch. Only posteriors > 0.5 are shown. Asterisks represent potential introductions in the RS state and subclades cited in the text are indicated. Circles indicate countries outside Brazil and Brazilian states (BR suffix).

For clade 3, the TMRCA was estimated on December 20, 2020 (95% HPD: November 25 to December 29, 2020) and the median evolutionary rate was 7.85×10^−4^ (95% HPD: 6.06×10^−4^ to 1.02×10^−3^ subst/site/year). This clade harbors sequences from nine Brazilian states and 10 other countries. Amazonas is the most probable source of its emergence. From then onwards, multiple transmission clusters were established in foreign countries (e.g. Spain, Portugal, USA) and Brazilian states (especially Maranhão, SP, and RS). This clade was introduced at least five times in RS, leading to two major subclades represented by 18 and four sequences, respectively. The major subclade (n=18, PP = 0.98) was dated January 11, 2021 (95% HPD: January 11 to February 1, 2021) (Figure 5B).

For clade 4, the TMRCA was dated on December 02, 2020 (95% HPD: October 7, 2020 to January 3, 2021), and the median evolutionary rate was 6.26×10^−4^ (95% HPD: 3.51×10^−4^ to 1.01×10^−3^). This clade comprises nine Brazilian states and five foreign countries. After its initial emergence and spread in Amazonas, it had already formed transmission clusters in SP, BA, United Kingdom, and the USA. Most important, a subclade containing sequences from two neighboring states from Southern Brazil (seven from RS and five from Santa Catarina [SC]) indicates its diffusion from RS to SC, probably leading to two separate introductions. The divergence of this subclade was estimated on December 16, 2020 (95% HPD: December 16, 2020 to January 19, 2021) (Figure 5C).

Phylogenetic and molecular clock approaches suggest the wide circulation of the VOC P.1 both nationally and internationally between late 2020 and early 2021. This lineage has already diversified into some clades that bear characteristic mutations, although they exhibit similar evolutionary rates. We have inferred that P.1 (and its derived clades) was introduced multiple times in the southernmost Brazilian state (RS) still in 2020, probably in December. Remarkably, this date is close to the first P.1 detection in Manaus, which is located ∼4 thousand kilometers away. These early introductions led to the formation of local subclades that could be identified even using a reduced set of sequenced samples.

## Discussion

In this study, the analysis of 56 samples from the state of Rio Grande do Sul (RS), Southern Brazil, confirmed that the P.1 lineage was already highly prevalent. Interestingly, we demonstrated that P.1 is already showing signs of diversification and has originated a new sublineage (P.1.2) by indicating the likely origin and the first clusters of this novel lineage. This sublineage was detected in three Brazilian states, and other countries, and its most recent common ancestor was dated on December 18th, 2020 (95% HPD: October 29th, 2020 to January 31th, 2021). In accordance with the majority of the states from Brazil, this state experienced significant increases in hospitalizations in early 2021. This scenario was related to the emergence and rapid spread of the P.1 variant across the country.

In most RNA viruses, the accuracy of RNA replication is low, leading to the emergence of frequent nucleotide substitutions. However, since its very long RNA genomic strand requires higher fidelity replication in order to keep up genome integrity, coronaviruses behave differently in this regard. In SARS-CoV-2, for example, a proofreading mechanism increases 100 to 1,000 fold the fidelity of RNA synthesis through the activity of an exonuclease present in NSP14. This enzyme corrects nucleotide misincorporation during RNA duplication by the error-prone RNA-dependent RNA polymerase (57–59). As a result, coronaviruses have lower rates of mutation than other RNA viruses that lack proofreading activity (58).

After almost one year of relatively slow SARS-CoV-2 evolution, the emergence of multiple and convergent lineages harboring a constellation of mutations in the spike protein raised concern in the scientific community. This protein is present on the viral surface of SARS-CoV-2 and is composed of two subunits, S1 and S2. S1 contains the receptor□binding domain (RBD), responsible for virus attachment at the cell surface, the N-Terminal Domain (NTD), also critical for binding properties and the more conserved CTS1. S2 is an alpha-helix-rich subunit that contains HR1 and HR2, critical for viral-cell membrane fusion (58). Spike protein, therefore, is responsible for mediating interaction with the human Angiotensin-Converting Enzyme 2 receptor (hACE2) and is a primary target of neutralizing antibodies and vaccine development (60). The variants harboring different mutational signatures, including spike protein substitutions, were classified as VOCs and “ Variant of interest” (VOI), depending on their growing relevance in the current pandemic. The first three VOC emerged in England (B.1.1.7) (7), South Africa (B.1.351) (8), and Brazil (P.1) (9). More recently, B.1.427/429 (California) also was characterized as VOC (61). As far as May 2020, B.1.526 (New York), B.1.617 (India), and P.2 (Brazil) (18) were still categorized as VOIs. B.1.1.7, B.1.351 and P.1, the most studied VOCs, have the D614G and N501Y mutations in common. B.1.351 and P.1 share a mutation in the K417 site (K417N and K417T, respectively) and the E484K replacement, which is also observed in the P.2 lineage. Additionally, B.1.1.7 carries the P681H substitution in the furin-cleavage site and multiple VOIs bear the L452R substitution (62). The presence of common substitutions in different SARS-CoV-2 lineages suggests co-evolutionary and convergent mutational processes (7–9,63).

D614G is a substitution of aspartic acid to glycine at amino acid position 614 at the CTS1 segment of the S1 subunit. It emerged at the end of January 2020, first reported in April 2020, and quickly became prevailing in several B.1-derived lineages, including most VOCs and VOIs described thus far (64–67). This mutation is interesting, since it does not occur near RBD and thus does not directly modify the binding affinity for hACE-2. Instead, it disrupts important hydrogen bonds with neighbor S2, resulting in altered interprotomeric configurations. As a consequence, the active “ one RBD up and two RBD down” is favored. This allows binding to ACE2 more effectively, leading to higher replication rates and viral loads (65). However, G614 mutants are similarly (or even more) susceptible to immune neutralization than the original D614 variant (65,68).

The substitution from asparagine to a tyrosine at position 501 (N501Y) has first appeared in the UK in September 2020. It is one of the six key contact residues within RBD interacting with ACE2 and has been associated with increased binding affinity to human and murine ACE2 due to the formation of an extra hydrogen bond with the ACE2 receptor (69–71). Studies suggest that this mutation could enhance SARS-CoV-2 transmissibility and mortality (72–74).

The substitution of glutamic acid for lysine at the 484 RBD position (E484K) creates a strong ion interaction with the amino acid 75 of ACE2 due to the electrostatic change from a negatively charged to a positively charged amino acid (75). E484K can also lead to immune evasion since it is located at a flexible loop previously irrelevant for receptor binding when the original glutamic acid is in place. This may explain the enhanced dissemination and improved infectivity and dissemination of SARS-CoV-2 (76,77). In fact, as of May 17th 2021, 66,405 sequences with the E484K mutation have been detected (62). Recent reinfection cases of E484K-containing SARS-CoV-2 (78,79) and the recent proof of its fixation in different lineages (80) are suggestive that this mutation must be investigated in more detail.

Despite the residue 452 does not directly contact the ACE2 receptor, L452 with the residues F490 and L492 form a hydrophobic patch on the surface of the spike RBD. This stabilizes the interaction between the spike protein and the ACE2 receptor and promotes an increased virus entry into the cell Moreover, lineages carrying this mutation seem to present a moderate resistance to neutralization by antibodies elicited by prior infection or vaccination (61). The P681H mutation, in turn, has not its significance completely elucidated. The adjacent position to the furin cleavage site (five sites upstream of the arginine residues), which is needed for the cell membrane fusion, can potentially affect the viral infectivity. Furthermore, the independent appearance in different lineages suggests convergent evolution and a possible adaptive advantage since the acquisition of a multibasic S1/S2 cleavage site was essential for SARS-CoV-2 infection in humans (81,82).

In the present study, we noticed that B.1.1.33 and B.1.1.28 lineages, detected at the beginning of the pandemic in Brazil (2), had been similarly prevalent in different regions until September 2020, before the appearance of P.2 (in October) and P.1 (in December 2020). The B.1.1.33 lineage shows variable abundance in different Brazilian states (ranging from 2% in Pernambuco to 80% in Rio de Janeiro), with moderate prevalence in South American countries (5-18%). Surprisingly, this lineage was firstly detected in early March 2020 in other American countries (*e*.*g*. Argentina and the USA). Apparently, an intermediate strain probably emerged in Europe and subsequently spread to Brazil, where its spread gave rise to B.1.1.33 (83) and possibly triggered secondary outbreaks in Argentina and Uruguay (83,84). We had found that N.3 and N.5, both derived from B.1.1.33, represented an important proportion of the sequences from Argentina from May to December 2020, when it was replaced by the P.2 lineage, which probably emerged in Rio de Janeiro (Southeastern Brazil). The B.1.1.28 lineage, despite apparently less abundant than B.1.1.33 in several Brazilian regions, quickly diversified into two variants: VOC P.1 and VOI P.2 (85). Since the end of 2020, these two lineages lead the diversity of SARS-CoV-2 in Brazil (14) and have caused concern in other countries after several introductions.

The emergence of a B.1.1.28 derived lineage carrying the S:E484K mutation (P.2) was dated, in a retrospective study, late February 2020 in the Southeast (São Paulo and Rio de Janeiro), followed by transmission to the South (especially RS). Since then, multiple dispersion routes were observed between Brazilian states, especially in late 2020 and early 2021 (15). However, this lineage went unreported until October 2020, when it was first detected in the state of Rio de Janeiro (18) and in the small municipality of Esteio in RS (19). The increased frequency of B.1.1.28 and derived lineages was corroborated by another study that included samples from several municipalities of RS in November 2020. This study found that 86% of the genomes could be classified as B.1.1.28 and ∼50% of these, in fact, belong to the new lineage P.2 (20). Nonetheless, our current study suggests that P.2 has already been nearly entirely replaced by the lineage or is not particularly well represented among the analyzed patients seeking emergency consultation or requiring hospitalization.

Between June and October 2020, an extremely high seroprevalence (44-76%) was observed in Manaus (Amazonas, Brazil) in a study from blood donors (11). However, despite these numbers, Manaus faced a resurgence of cases and a 6-fold increase in hospitalizations between December 2020 and January 2021. The most plausible hypotheses that would justify this condition are: (i) the previous overestimation of seroprevalence in Manaus, (ii) the immune evasion property of some SARS-CoV-2 mutations found in VOCs, and (iii) higher transmissibility and pathogenicity of SARS-CoV-2 lineages circulating in the second wave compared with pre-existing lineages (12).

A genomic epidemiology study that used 250 SARS-CoV-2 genomes from 25 different municipalities from Amazonas sampled between March 2020, and January 2021 shows that the first exponential phase in the state was driven mainly by the spread of lineage B.1.195, which was gradually replaced by B.1.1.28. The second wave coincided with the emergence of P.1 in November, which rapidly replaced the parental lineage (<2 months) (10) and whose emergence was preceded by a period of rapid molecular evolution (9). Importantly, rapid accumulation of mutations over short timeframes have been reported in chronically infected or immunocompromised hosts (86,87). However, preliminary findings pointed to the existence of P.1 intermediate lineages, suggesting that the constellation of mutations defining P.1 were acquired at sequential steps during multiple rounds of infections instead of within a single long-term infected individual (88). The VOC P.1 carries three deletions, four synonymous substitutions, a four base-pair nucleotide insertion, and at least 17 other lineage-defining replacements, including 10 missense mutations in the Spike protein (L18F, T20N, P26S, D138Y, R190S, K417T, E484K, N501Y, H655Y, T1027I), eight of which are knowingly subjected to positive selection (9).

Regarding infectiousness, transmissibility, and case fatality, the viral load was ∼10-fold higher in P.1 infections than in non-P.1 infections (10). Although another study points to uncertainties regarding viral load and duration of infection after accounting for confounding effects (9). Moreover, it was estimated to be 1.7–2.4-fold more transmissible, raising the probability that reinfections would be caused more frequently in hosts infected by P.1 rather than by older lineages. Remarkably, infections were 1.2–1.9 times more likely to result in death in the period following the emergence of P.1 compared to previous time frames (9). These findings support that successive lineage replacements in Amazonas were driven by a complex combination of factors, including the emergence of the more transmissible VOC P.1 virus (10).

A study conducted in RS described a P.1 lineage infection on November 30th followed by a lineage reinfection on March 11th in a patient with comorbidities. This report was the first detected P.1 in the state (22). Other analyses suggest that the P.1 lineage presumably emerged in Manaus, Brazil, in mid-November 2020 (9,10). Therefore, the P.1 lineage was present in Southern Brazil about just 15 days after the P.1 emergence in Brazil. Our molecular clock analysis supported this scenario. Another study, once thought to be the first P.1 report in RS, documented local transmission of P.1 from a person who had close contact with tourists and was positive to COVID-19 in early February 2021 (21). This happened in the city of Gramado, a town on the mountains that receives around 6.5 million tourists every year and belongs to the Caxias do Sul intermediate region. Interestingly, this sample from Gramado was the earliest representative of a new P.1-derived lineage (P.1.2), described in 11 patients from this study and found in transmission clusters from the RJ state in Southeastern Brazil, the USA, and the Netherlands. Remarkably, these local sequences are more similar to genomes from other countries compared to the RJ cluster, which acquired at least four additional mutations (including S:A262S) (17).

Whether P.1.2 has worse clinical outcomes than its prior variant (P.1) is unknown. However, as described above, the missense mutations characteristic of the new sublineage are located at nsp2 and nsp3 (ORF1ab), ORF3a, and Nucleocapsid. These sites are known for their interaction with human proteome, potentially influencing the immunological and inflammatory response against SARS-CoV-2 infection (89). The ORF3a:D155Y substitution is located near SARS-CoV caveolin-binding Domain IV. The binding interaction of viral ORF3a protein to host caveolin-1 is essential for entry and endomembrane trafficking of SARS-CoV-2. Since this mutation breaks the salt bridge formation between Asp155-Arg134, it can interfere with the binding affinity of ORF3a to host caveolin-1 and change virulence properties. Most importantly, this disrupted interaction may be associated with improved viral fitness, since it can avoid the induction of host cell apoptosis or extend the asymptomatic phase of infection (90). We hypothesize that these new substitutions could, therefore, influence epidemiological and clinical outcomes favouring P.1.2 evolution. This is elusive at best at this time, however, and further sublineage characterization is needed to further exploit its real relevance.

Some limitations should be considered. Firstly, the sample size is low and not necessarily representative of the RS state. Furthermore, publicly available genomes are a result of episodic sequencing efforts, especially in Brazil. This scenario restricts more precise inferences about introductions and diffusion processes in regional and worldwide contexts since samples are not geographical and temporally well distributed. Therefore, more research and surveillance are essential to unravel a more precise genomic characterization of SARS-CoV-2 in Brazil, identifying novel variants promptly to better respond and control its spread.

In summary, our study corroborates the total virtual substitution of previous lineages by P.1 in Southern Brazil in COVID-19 cases sequenced in March 2020. Moreover, we confirmed various cases caused by the novel P.1.2 sublineage and placed its origin in the State of Rio Grande do Sul. The continuous evolution of the VOC P.1 is worrisome, considering its clinical and epidemiological impact, and warrants enhanced genomic surveillance.

## Data Availability

Full tables acknowledging the authors and corresponding labs submitting sequencing data used in this study can be found in Supplementary Files 3 and 4. Consensus genomes generated in this study were deposited in the GISAID database under Accession IDs: EPI_ISL_2139494 to EPI_ISL_2139549. Additional information used and/or analysed during the current study are available from the corresponding author on reasonable request.

https://www.gisaid.org/

## Author’s contributions

**Vinicius B. Franceschi:** Methodology, Software, Validation, Formal analysis, Investigation, Data Curation, Visualization, Writing - Original Draft, Writing - Review & Editing. **Gabriel D. Caldana:** Methodology, Validation, Formal analysis, Investigation, Visualization, Writing - Original Draft, Writing - Review & Editing. **Christiano Perin:** Investigation, Data Curation, Resources, Writing - Review & Editing. **Alexandre Horn:** Investigation, Data Curation, Resources, Writing - Review & Editing. **Camila Peter:** Methodology, Data Curation, Resources. **Gabriela B. Cybis:** Methodology, Validation, Formal analysis, Investigation, Writing - Review & Editing, Supervision. **Patrícia A. G. Ferrareze:** Writing - Review & Editing. **Liane N. Rotta:** Resources, Writing - Review & Editing. **Flávio A. Cadegiani:** Resources, Writing - Review & Editing. **Ricardo A. Zimerman:** Conceptualization, Methodology, Investigation, Resources, Writing - Original Draft, Writing - Review & Editing, Project administration. **Claudia E. Thompson:** Conceptualization, Methodology, Formal analysis, Investigation, Resources, Writing - Original Draft, Writing - Review & Editing, Supervision, Project administration.

All authors have read and approved the manuscript.

## Acknowledgements

We thank the administrators of the GISAID database and research groups across the world for supporting the rapid and transparent sharing of genomic data during the COVID-19 pandemic. We also thank the staff of Hospital da Brigada Militar, Laboratório Exame, Beppler & Puppi Advogados, Smellbox Produtos de Higiene Ltda., Dr. Leonardo Mestre Negri, Florense Brands, and Biome that directly contributed to this study.

## Supplementary files

**Supplementary file 1**. Coverage depth plots for each genome sequenced.

**Supplementary file 2**. Collection of all mutations (n=175) found in the 56 sequenced genomes, including effects on SARS-CoV-2 genes and proteins. The table is ordered by genomic position.

**Supplementary file 3**. GISAID acknowledgment table of the 8,635 global sequences used in the Nextstrain workflow.

**Supplementary file 4**. GISAID acknowledgment table of the 4,499 P.1 sequences analyzed.

**Supplementary File 5**. Expanded ML tree built using 4,499 P.1 sequences deposited in GISAID until 26 April, 2021. Tips represented by sequences sequenced in this study are augmented to enable visualization of the different introductions, clusters and clades reported.

## References

1. World Health Organization. WHO Director-General’s opening remarks at the media briefing on COVID-19 - 11 March 2020 [Internet]. 2020 [cited 2020 Nov 10]. Available from: https://www.who.int/director-general/speeches/detail/who-director-general-s-opening-remarks-at-the-media-briefing-on-covid-19---11-march-2020

2. Candido D, Claro IM, Jesus JG de, Souza WM, Moreira FRR, Dellicour S, et al. Evolution and epidemic spread of SARS-CoV-2 in Brazil. Science. 2020 Sep 4;369(6508):1255–60.

3. IBGE (Brazilian Institute of Geography and Statistics). Rio Grande do Sul - Cidades e Estados [Internet]. 2021 [cited 2021 May 17]. Available from: https://www.ibge.gov.br/cidades-e-estados/rs.html

4. IBGE (Brazilian Institute of Geography and Statistics). Regiões Geográficas [Internet]. 2021 [cited 2021 May 17]. Available from: https://www.ibge.gov.br/apps/regioes_geograficas/

5. Rio Grande do Sul Department of Health - SES-RS. Confirmado o primeiro caso de novo coronavírus no Rio Grande do Sul [Internet]. Secretaria da Saúde. 2020 [cited 2020 Nov 24]. Available from: https://saude.rs.gov.br/confirmado-o-primeiro-caso-de-novo-coronavirus-no-rio-grande-do-sul

6. Secretaria de Planejamento, Governança e Gestão - Governo do Estado do Rio Grande do Sul. Cogestão Regional - Distanciamento Controlado [Internet]. 2021 [cited 2021 May 17]. Available from: https://distanciamentocontrolado.rs.gov.br/

7. Rambaut A, Loman N, Pybus O, Barclay W, Barrett J, Carabelli A, et al. Preliminary genomic characterisation of an emergent SARS-CoV-2 lineage in the UK defined by a novel set of spike mutations [Internet]. Virological. 2020 [cited 2021 Jan 4]. Available from: https://virological.org/t/preliminary-genomic-characterisation-of-an-emergent-sars-cov-2-lineage-in-the-uk-defined-by-a-novel-set-of-spike-mutations/563

8. Tegally H, Wilkinson E, Giovanetti M, Iranzadeh A, Fonseca V, Giandhari J, et al. Detection of a SARS-CoV-2 variant of concern in South Africa. Nature. 2021 Apr;592(7854):438–43.

9. Faria N, Mellan TA, Whittaker C, Claro IM, Candido D da S, Mishra S, et al. Genomics and epidemiology of the P.1 SARS-CoV-2 lineage in Manaus, Brazil. Science [Internet]. 2021 Apr 14 [cited 2021 Apr 20]; Available from: https://science.sciencemag.org/content/early/2021/04/13/science.abh2644

10. Naveca F, Nascimento V, Souza V, Corado A, Nascimento F, Silva G, et al. COVID-19 epidemic in the Brazilian state of Amazonas was driven by long-term persistence of endemic SARS-CoV-2 lineages and the recent emergence of the new Variant of Concern P.1 [Internet]. 2021 [cited 2021 Mar 1]. Available from: https://www.researchsquare.com

11. Buss LF, Prete CA, Abrahim CMM, Mendrone A, Salomon T, Almeida-Neto C de, et al. Three-quarters attack rate of SARS-CoV-2 in the Brazilian Amazon during a largely unmitigated epidemic. Science [Internet]. 2020 Dec 8 [cited 2020 Dec 19]; Available from: https://science.sciencemag.org/content/early/2020/12/07/science.abe9728

12. Sabino EC, Buss LF, Carvalho MPS, Prete CA, Crispim MAE, Fraiji NA, et al. Resurgence of COVID-19 in Manaus, Brazil, despite high seroprevalence. The Lancet. 2021 Feb 6;397(10273):452–5.

13. Brazilian Ministry of Health. Painel Coronavírus Brasil [Internet]. 2021 [cited 2021 May 17]. Available from: https://covid.saude.gov.br/

14. Franceschi VB, Ferrareze PAG, Zimerman RA, Cybis GB, Thompson CE. Mutation hotspots, geographical and temporal distribution of SARS-CoV-2 lineages in Brazil, February 2020 to February 2021: insights and limitations from uneven sequencing efforts. medRxiv. 2021 Mar 12;2021.03.08.21253152.

15. Lamarca AP, Almeida LGP de, Francisco R da S, Lima LFA, Scortecci KC, Perez VP, et al. Genomic surveillance of SARS-CoV-2 tracks early interstate transmission of P.1 lineage and diversification within P.2 clade in Brazil. medRxiv. 2021 Mar 26;2021.03.21.21253418.

16. Rio Grande do Sul Health Surveillance Center. Genomic Bulletin 5 (16/04/2021) [Internet]. 2021 [cited 2021 Apr 20]. Available from: https://coronavirus.rs.gov.br/upload/arquivos/202104/16173629-vigilancia-genomica-rs-boletim05-compactado.pdf

17. De Almeira LG, Lamarca AP, Francisco Junior R da S, Cavalcante L, Gerber AL, Guimarães AP de C, et al. Genomic Surveillance of SARS-CoV-2 in the State of Rio de Janeiro, Brazil: technical briefing - SARS-CoV-2 coronavirus / nCoV-2019 Genomic Epidemiology [Internet]. Virological. 2021 [cited 2021 May 4]. Available from: https://virological.org/t/genomic-surveillance-of-sars-cov-2-in-the-state-of-rio-de-janeiro-brazil-technical-briefing/683

18. Voloch CM, Francisco R da S, Almeida LGP de, Cardoso CC, Brustolini OJ, Gerber AL, et al. Genomic characterization of a novel SARS-CoV-2 lineage from Rio de Janeiro, Brazil. J Virol [Internet]. 2021 Mar 1 [cited 2021 Apr 20]; Available from: https://jvi.asm.org/content/early/2021/02/25/JVI.00119-21

19. Franceschi VB, Caldana GD, Mayer A de M, Cybis GB, Neves CAM, Ferrareze PAG, et al. Genomic Epidemiology of SARS-CoV-2 in Esteio, Rio Grande do Sul, Brazil. medRxiv. 2021 Jan 26;2021.01.21.21249906.

20. Francisco Jr R da S, Benites LF, Lamarca AP, de Almeida LGP, Hansen AW, Gularte JS, et al. Pervasive transmission of E484K and emergence of VUI-NP13L with evidence of SARS-CoV-2 co-infection events by two different lineages in Rio Grande do Sul, Brazil. Virus Res. 2021 Apr 15;296:198345.

21. Salvato RS, Gregianini TS, Campos AAS, Crescente LV, Vallandro MJ, Ranieri TMS, et al. Epidemiological investigation reveals local transmission of SARS-CoV-2 lineage P.1 in Southern Brazil. Rev Epidemiol E Controle Infecção [Internet]. 2021 Apr 6 [cited 2021 Apr 20];1(1). Available from: https://online.unisc.br/seer/index.php/epidemiologia/article/view/16335

22. Soares da Silva M, Demoliner M, Hansen A, Gularte J, Silveira F, Heldt F, et al. Early detection of SARS-CoV-2 P.1 variant in Southern Brazil and reinfection of the same patient by P.2 [Internet]. 2021 [cited 2021 May 14]. Available from: https://www.researchsquare.com

23. Corman VM, Landt O, Kaiser M, Molenkamp R, Meijer A, Chu DK, et al. Detection of 2019 novel coronavirus (2019-nCoV) by real-time RT-PCR. Eurosurveillance. 2020 Jan 23;25(3):2000045.

24. World Health Organization. COVID-19 Clinical management: living guidance [Internet]. 2021 [cited 2021 May 1]. Available from: https://www.who.int/publications-detail-redirect/WHO-2019-nCoV-clinical-2021-1

25. Eden J-S. SARS-CoV-2 Genome Sequencing Using Long Pooled Amplicons on Illumina Platforms. 2020 Apr 4 [cited 2021 May 12]; Available from: https://www.protocols.io/view/sars-cov-2-genome-sequencing-using-long-pooled-amp-befyjbpw

26. Langmead B, Salzberg SL. Fast gapped-read alignment with Bowtie 2. Nat Methods. 2012 Mar 4;9(4):357–9.

27. Li H, Handsaker B, Wysoker A, Fennell T, Ruan J, Homer N, et al. The Sequence Alignment/Map format and SAMtools. Bioinformatics. 2009 Aug 15;25(16):2078–9.

28. Li H. A statistical framework for SNP calling, mutation discovery, association mapping and population genetical parameter estimation from sequencing data. Bioinformatics. 2011 Nov 1;27(21):2987–93.

29. Gel B, Serra E. karyoploteR: an R/Bioconductor package to plot customizable genomes displaying arbitrary data. Bioinforma Oxf Engl. 2017 Oct 1;33(19):3088–90.

30. Garrison E, Marth G. Haplotype-based variant detection from short-read sequencing. ArXiv12073907 Q-Bio [Internet]. 2012 Jul 20 [cited 2020 Nov 14]; Available from: http://arxiv.org/abs/1207.3907

31. Cingolani P, Platts A, Wang LL, Coon M, Nguyen T, Wang L, et al. A program for annotating and predicting the effects of single nucleotide polymorphisms, SnpEff. Fly (Austin). 2012 Apr 1;6(2):80–92.

32. Katoh K, Standley DM. MAFFT Multiple Sequence Alignment Software Version 7: Improvements in Performance and Usability. Mol Biol Evol. 2013 Apr;30(4):772–80.

33. Du Plessis L. laduplessis/SARS-CoV-2_Guangdong_genomic_epidemiology: Initial release [Internet]. Zenodo; 2020 [cited 2021 May 4]. Available from: https://zenodo.org/record/3922606

34. Rambaut A, Holmes EC, O’Toole Á, Hill V, McCrone JT, Ruis C, et al. A dynamic nomenclature proposal for SARS-CoV-2 lineages to assist genomic epidemiology. Nat Microbiol. 2020 Nov;5(11):1403–7.

35. Argimón S, Abudahab K, Goater RJE, Fedosejev A, Bhai J, Glasner C, et al. Microreact: visualizing and sharing data for genomic epidemiology and phylogeography. Microb Genomics [Internet]. 2016 Nov 30 [cited 2020 Jun 26];2(11). Available from: https://www.ncbi.nlm.nih.gov/pmc/articles/PMC5320705/

36. Hadfield J, Megill C, Bell SM, Huddleston J, Potter B, Callender C, et al. Nextstrain: real-time tracking of pathogen evolution. Bioinformatics. 2018 Dec 1;34(23):4121–3.

37. Nguyen L-T, Schmidt HA, von Haeseler A, Minh BQ. IQ-TREE: A Fast and Effective Stochastic Algorithm for Estimating Maximum-Likelihood Phylogenies. Mol Biol Evol. 2015 Jan 1;32(1):268–74.

38. Tavare S. Some probabilistic and statistical problems in the analysis of DNA sequences. Some Math Quest Biol DNA Seq Anal Ed Robert M Miura [Internet]. 1986 [cited 2021 May 1]; Available from: https://agris.fao.org/agris-search/search.do?recordID=US201301755037

39. Sagulenko P, Puller V, Neher RA. TreeTime: Maximum-likelihood phylodynamic analysis. Virus Evol [Internet]. 2018 Jan 1 [cited 2020 Nov 14];4(1). Available from: https://academic.oup.com/ve/article/4/1/vex042/4794731

40. Kalyaanamoorthy S, Minh BQ, Wong TKF, von Haeseler A, Jermiin LS. ModelFinder: fast model selection for accurate phylogenetic estimates. Nat Methods. 2017 Jun;14(6):587–9.

41. Guindon S, Dufayard J-F, Lefort V, Anisimova M, Hordijk W, Gascuel O. New Algorithms and Methods to Estimate Maximum-Likelihood Phylogenies: Assessing the Performance of PhyML 3.0. Syst Biol. 2010 May 1;59(3):307–21.

42. Rambaut A, Lam TT, Max Carvalho L, Pybus OG. Exploring the temporal structure of heterochronous sequences using TempEst (formerly Path-O-Gen). Virus Evol [Internet]. 2016 Apr 9 [cited 2020 Nov 14];2(1). Available from: https://www.ncbi.nlm.nih.gov/pmc/articles/PMC4989882/

43. Yu G, Smith DK, Zhu H, Guan Y, Lam TT-Y. ggtree: an r package for visualization and annotation of phylogenetic trees with their covariates and other associated data. Methods Ecol Evol. 2017;8(1):28–36.

44. Orme D, Freckleton R, Thomas G, Petzoldt T, Fritz S, Isaac N, et al. Caper: Comparative Analyses of Phylogenetics and Evolution in R (R package version 1.0.1) [Internet]. [cited 2021 May 10]. Available from: https://www.scienceopen.com/document?vid=d750bff2-a400-41dd-aa1e-728bb7aaf4d5

45. Suchard MA, Lemey P, Baele G, Ayres DL, Drummond AJ, Rambaut A. Bayesian phylogenetic and phylodynamic data integration using BEAST 1.10. Virus Evol [Internet]. 2018 Jan 1 [cited 2020 Nov 16];4(1). Available from: https://academic.oup.com/ve/article/4/1/vey016/5035211

46. Ayres DL, Darling A, Zwickl DJ, Beerli P, Holder MT, Lewis PO, et al. BEAGLE: An Application Programming Interface and High-Performance Computing Library for Statistical Phylogenetics. Syst Biol. 2012 Jan 1;61(1):170–3.

47. Hasegawa M, Kishino H, Yano T. Dating of the human-ape splitting by a molecular clock of mitochondrial DNA. J Mol Evol. 1985;22(2):160–74.

48. Ferreira MAR, Suchard MA. Bayesian analysis of elapsed times in continuous-time Markov chains. Can J Stat. 2008;36(3):355–68.

49. Gill MS, Lemey P, Faria NR, Rambaut A, Shapiro B, Suchard MA. Improving Bayesian Population Dynamics Inference: A Coalescent-Based Model for Multiple Loci. Mol Biol Evol. 2013 Mar 1;30(3):713–24.

50. Rambaut A, Drummond AJ, Xie D, Baele G, Suchard MA. Posterior Summarization in Bayesian Phylogenetics Using Tracer 1.7. Syst Biol. 2018 Sep 1;67(5):901–4.

51. Lemey P, Rambaut A, Drummond AJ, Suchard MA. Bayesian Phylogeography Finds Its Roots. PLOS Comput Biol. 2009 Sep 25;5(9):e1000520.

52. R Core Team. R: A Language and Environment for Statistical Computing [Internet]. Vienna, Austria: R Foundation for Statistical Computing; 2020. Available from: https://www.R-project.org/

53. Wickham H. ggplot2: Elegant Graphics for Data Analysis [Internet]. New York: Springer-Verlag; 2009 [cited 2021 May 4]. (Use R!). Available from: https://www.springer.com/gp/book/9780387981413

54. Pereira R, Gonçalves C. geobr: Loads Shapefiles of Official Spatial Data Sets of Brazil [Internet]. 2019. Available from: https://github.com/ipeaGIT/geobr

55. Pebesma E. Simple Features for R: Standardized Support for Spatial Vector Data. R J. 2018;10(1):439–46.

56. Bielejec F, Baele G, Vrancken B, Suchard MA, Rambaut A, Lemey P. SpreaD3: Interactive Visualization of Spatiotemporal History and Trait Evolutionary Processes. Mol Biol Evol. 2016 Aug;33(8):2167–9.

57. Wu F, Zhao S, Yu B, Chen Y-M, Wang W, Song Z-G, et al. A new coronavirus associated with human respiratory disease in China. Nature. 2020 Mar;579(7798):265–9.

58. Ma Y, Wu L, Shaw N, Gao Y, Wang J, Sun Y, et al. Structural basis and functional analysis of the SARS coronavirus nsp14–nsp10 complex. Proc Natl Acad Sci. 2015 Jul 28;112(30):9436–41.

59. Ogando NS, Ferron F, Decroly E, Canard B, Posthuma CC, Snijder EJ. The Curious Case of the Nidovirus Exoribonuclease: Its Role in RNA Synthesis and Replication Fidelity. Front Microbiol [Internet]. 2019 [cited 2021 May 15];10. Available from: https://www.frontiersin.org/articles/10.3389/fmicb.2019.01813/full

60. Walls AC, Park Y-J, Tortorici MA, Wall A, McGuire AT, Veesler D. Structure, Function, and Antigenicity of the SARS-CoV-2 Spike Glycoprotein. Cell. 2020 Apr 16;181(2):281–292.e6.

61. Deng X, Garcia-Knight MA, Khalid MM, Servellita V, Wang C, Morris MK, et al. Transmission, infectivity, and neutralization of a spike L452R SARS-CoV-2 variant. Cell [Internet]. 2021 Apr 20 [cited 2021 May 10]; Available from: https://www.sciencedirect.com/science/article/pii/S0092867421005055

62. Mullen JL, Tsueng G, Latif AA, Alkuzweny M, Cano M, Haag E, et al. outbreak.info [Internet]. 2021 [cited 2021 May 17]. Available from: outbreak.info

63. Martin DP, Weaver S, Tegally H, San EJ, Shank SD, Wilkinson E, et al. The emergence and ongoing convergent evolution of the N501Y lineages coincides with a major global shift in the SARS-CoV-2 selective landscape. medRxiv. 2021 Mar 5;2021.02.23.21252268.

64. Guo S, Liu K, Zheng J. The Genetic Variant of SARS-CoV-2: would It Matter for Controlling the Devastating Pandemic? Int J Biol Sci. 2021;17(6):1476–85.

65. Groves DC, Rowland-Jones SL, Angyal A. The D614G mutations in the SARS-CoV-2 spike protein: Implications for viral infectivity, disease severity and vaccine design. Biochem Biophys Res Commun. 2021 Jan 29;538:104–7.

66. Korber B, Fischer WM, Gnanakaran S, Yoon H, Theiler J, Abfalterer W, et al. Tracking Changes in SARS-CoV-2 Spike: Evidence that D614G Increases Infectivity of the COVID-19 Virus. Cell. 2020 Aug 20;182(4):812–827.e19.

67. Laha S, Chakraborty J, Das S, Manna SK, Biswas S, Chatterjee R. Characterizations of SARS-CoV-2 mutational profile, spike protein stability and viral transmission. Infect Genet Evol. 2020 Nov 1;85:104445.

68. Jackson CB, Zhang L, Farzan M, Choe H. Functional importance of the D614G mutation in the SARS-CoV-2 spike protein. Biochem Biophys Res Commun. 2021 Jan 29;538:108–15.

69. Khan A, Zia T, Suleman M, Khan T, Ali SS, Abbasi AA, et al. Higher infectivity of the SARS-CoV-2 new variants is associated with K417N/T, E484K, and N501Y mutants: An insight from structural data. J Cell Physiol [Internet]. 2021 [cited 2021 May 13];n/a(n/a). Available from: https://onlinelibrary.wiley.com/doi/abs/10.1002/jcp.30367

70. Starr TN, Greaney AJ, Hilton SK, Ellis D, Crawford KHD, Dingens AS, et al. Deep Mutational Scanning of SARS-CoV-2 Receptor Binding Domain Reveals Constraints on Folding and ACE2 Binding. Cell. 2020 Sep 3;182(5):1295–1310.e20.

71. Leung K, Shum MH, Leung GM, Lam TT, Wu JT. Early transmissibility assessment of the N501Y mutant strains of SARS-CoV-2 in the United Kingdom, October to November 2020. Eurosurveillance. 2021 Jan 7;26(1):2002106.

72. Tang JW, Tambyah PA, Hui DS. Emergence of a new SARS-CoV-2 variant in the UK. J Infect. 2021 Apr 1;82(4):e27–8.

73. Gu H, Chen Q, Yang G, He L, Fan H, Deng Y-Q, et al. Adaptation of SARS-CoV-2 in BALB/c mice for testing vaccine efficacy. Science. 2020 Sep 25;369(6511):1603–7.

74. Ali F, Kasry A, Amin M. The new SARS-CoV-2 strain shows a stronger binding affinity to ACE2 due to N501Y mutant. Med Drug Discov. 2021 Jun 1;10:100086.

75. Nelson G, Buzko O, Spilman P, Niazi K, Rabizadeh S, Soon-Shiong P. Molecular dynamic simulation reveals E484K mutation enhances spike RBD-ACE2 affinity and the combination of E484K, K417N and N501Y mutations (501Y.V2 variant) induces conformational change greater than N501Y mutant alone, potentially resulting in an escape mutant. bioRxiv. 2021 Jan 13;2021.01.13.426558.

76. Baum A, Fulton BO, Wloga E, Copin R, Pascal KE, Russo V, et al. Antibody cocktail to SARS-CoV-2 spike protein prevents rapid mutational escape seen with individual antibodies. Science. 2020 Aug 21;369(6506):1014–8.

77. Greaney AJ, Starr TN, Gilchuk P, Zost SJ, Binshtein E, Loes AN, et al. Complete mapping of mutations to the SARS-CoV-2 spike receptor-binding domain that escape antibody recognition. Cell Host Microbe [Internet]. 2020 Nov 19 [cited 2020 Nov 24]; Available from: http://www.sciencedirect.com/science/article/pii/S1931312820306247

78. Nonaka CKV, Franco MM, Gräf T, Barcia CA de L, Mendonça RN de Á, Sousa KAF de, et al. Genomic Evidence of SARS-CoV-2 Reinfection Involving E484K Spike Mutation, Brazil. Emerg Infect Dis [Internet]. 2021 May [cited 2021 Feb 24];27(5). Available from: https://www.nc.cdc.gov/eid/article/27/5/21-0191_article

79. Resende PC, Bezerra JF, Vasconcelos RHT, Arantes I, Appolinario L, Mendonça AC, et al. Severe Acute Respiratory Syndrome Coronavirus 2 P.2 Lineage Associated with Reinfection Case, Brazil, June–October 2020. Emerg Infect Dis. 27:July 2021.

80. Ferrareze PAG, Franceschi VB, Mayer A de M, Caldana GD, Zimerman RA, Thompson CE. E484K as an innovative phylogenetic event for viral evolution: Genomic analysis of the E484K spike mutation in SARS-CoV-2 lineages from Brazil. bioRxiv. 2021 Jan 27;2021.01.27.426895.

81. Lasek-Nesselquist E, Pata J, Schneider E, George KS. A tale of three SARS-CoV-2 variants with independently acquired P681H mutations in New York State. medRxiv. 2021 Mar 12;2021.03.10.21253285.

82. Hoffmann M, Kleine-Weber H, Pöhlmann S. A Multibasic Cleavage Site in the Spike Protein of SARS-CoV-2 Is Essential for Infection of Human Lung Cells. Mol Cell. 2020 May 21;78(4):779–784.e5.

83. Resende PC, Delatorre E, Gräf T, Mir D, Motta FC, Appolinario LR, et al. Evolutionary Dynamics and Dissemination Pattern of the SARS-CoV-2 Lineage B.1.1.33 During the Early Pandemic Phase in Brazil. Front Microbiol [Internet]. 2021 [cited 2021 Feb 24];11. Available from: https://www.frontiersin.org/articles/10.3389/fmicb.2020.615280/full

84. Mir D, Rego N, Resende PC, López-Tort F, Fernandez-Calero T, Noya V, et al. Recurrent dissemination of SARS-CoV-2 through the Uruguayan-Brazilian border. medRxiv. 2021 Jan 8;2021.01.06.20249026.

85. Naveca F, Nascimento V, Souza V, Corado A, Nascimento F, Silva G, et al. Phylogenetic relationship of SARS-CoV-2 sequences from Amazonas with emerging Brazilian variants harboring mutations E484K and N501Y in the Spike protein [Internet]. Virological. 2021 [cited 2021 Feb 24]. Available from: https://virological.org/t/phylogenetic-relationship-of-sars-cov-2-sequences-from-amazonas-with-emerging-brazilian-variants-harboring-mutations-e484k-and-n501y-in-the-spike-protein/585

86. Choi B, Choudhary MC, Regan J, Sparks JA, Padera RF, Qiu X, et al. Persistence and Evolution of SARS-CoV-2 in an Immunocompromised Host. N Engl J Med. 2020 Dec 3;383(23):2291–3.

87. Kemp SA, Collier DA, Datir RP, Ferreira IATM, Gayed S, Jahun A, et al. SARS-CoV-2 evolution during treatment of chronic infection. Nature. 2021 Feb 5;1–10.

88. Gräf T, Bello G, Venas TMM, Pereira EC, Paixão ACD, Appolinario LR, et al. Identification of SARS-CoV-2 P.1-related lineages in Brazil provides new insights about the mechanisms of emergence of Variants of Concern - SARS-CoV-2 coronavirus / nCoV-2019 Genomic Epidemiology [Internet]. Virological. 2021 [cited 2021 May 17]. Available from: https://virological.org/t/identification-of-sars-cov-2-p-1-related-lineages-in-brazil-provides-new-insights-about-the-mechanisms-of-emergence-of-variants-of-concern/694/1

89. Plante JA, Mitchell BM, Plante KS, Debbink K, Weaver SC, Menachery VD. The variant gambit: COVID-19’s next move. Cell Host Microbe. 2021 Apr 14;29(4):508–15.

90. Gupta S, Mallick D, Banerjee K, Sarkar S, Lee STM, Basuchowdhuri P, et al. D155Y Substitution of SARS-CoV-2 ORF3a Weakens Binding with Caveolin-1: An in silico Study. bioRxiv. 2021 Mar 26;2021.03.26.437194.

