## Supplementary Material for "Predominance of the SARS-CoV-2 lineage P.1 and its sublineage P.1.2 in patients from the metropolitan region of Porto Alegre, Southern Brazil in March 2021: a phylogenomic analysis"

### **Table of Contents**

**Figure S1.** Spatiotemporal distribution of the 56 sequenced samples from RS.	2

**Figure S2.** Neighbouring countries, Brazilian divisions and RS intermediate regions.	3

**Figure S3.** Proportion of the 10 most frequent lineages of SARS-CoV-2 across time in four Brazilian regions.	4

#### **Figure S1**. Spatiotemporal distribution of the 56 sequenced samples from RS.

(A) Geographic distribution of the 13 municipalities included.

(B) ML tree of the local genomes.

(C) Timeline of sampling dates from March 2021.

Colors in the tree panels are correspondent and depict each of the 13 municipalities.

Interactive visualization is available at: <https://microreact.org/project/bTjQ665cumQQtbv4B8fmXy>

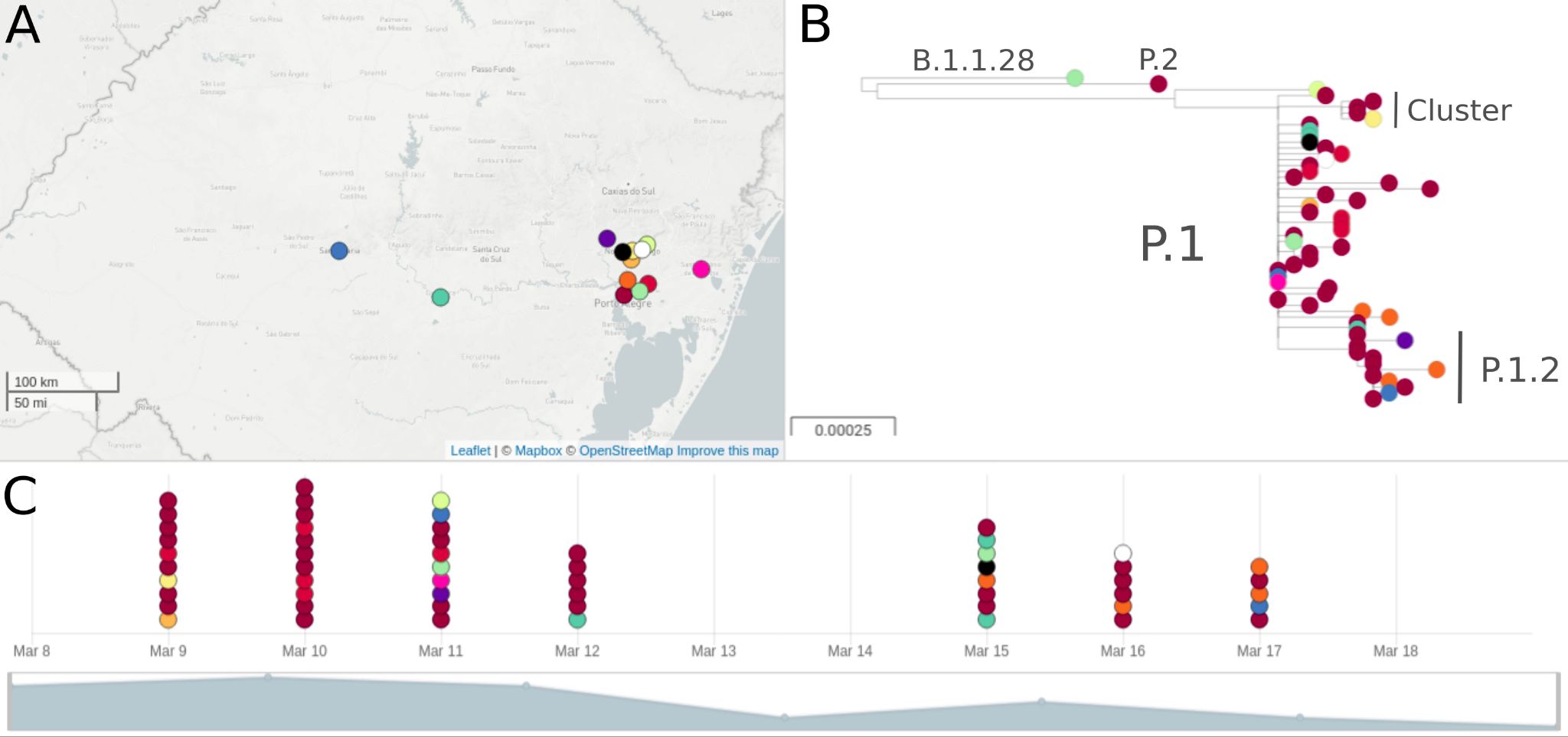

#### **Figure S2**. Neighbouring countries, Brazilian divisions and RS intermediate regions.

(A) Map from South America showing countries that share borders with Brazil (Argentina and Uruguay).

(B) Brazilian map displaying the RS state (dark blue) and its bordering state Santa Catarina (grey), as well as the divisions of the 26 states plus the Federal District.

(C) RS state map presenting the 8 intermediate regions defined by IBGE. The metropolitan region of Porto Alegre and its component municipalities are represented in light grey, while the state capital (Porto Alegre) is shown in dark grey.

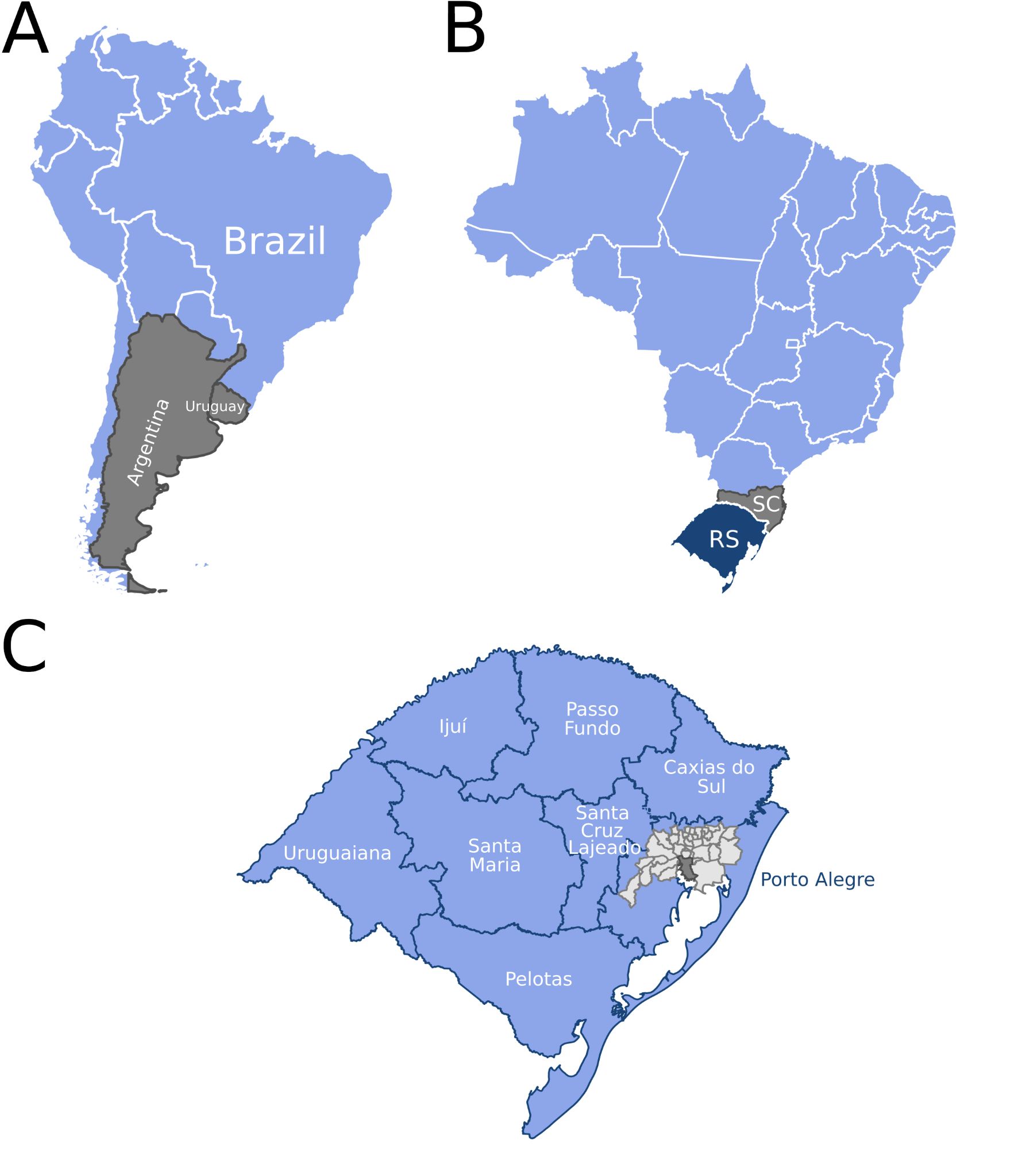

#### **Figure S3**. Proportion of the 10 most frequent lineages of SARS-CoV-2 across time in four Brazilian regions.

NA=Other lineages.

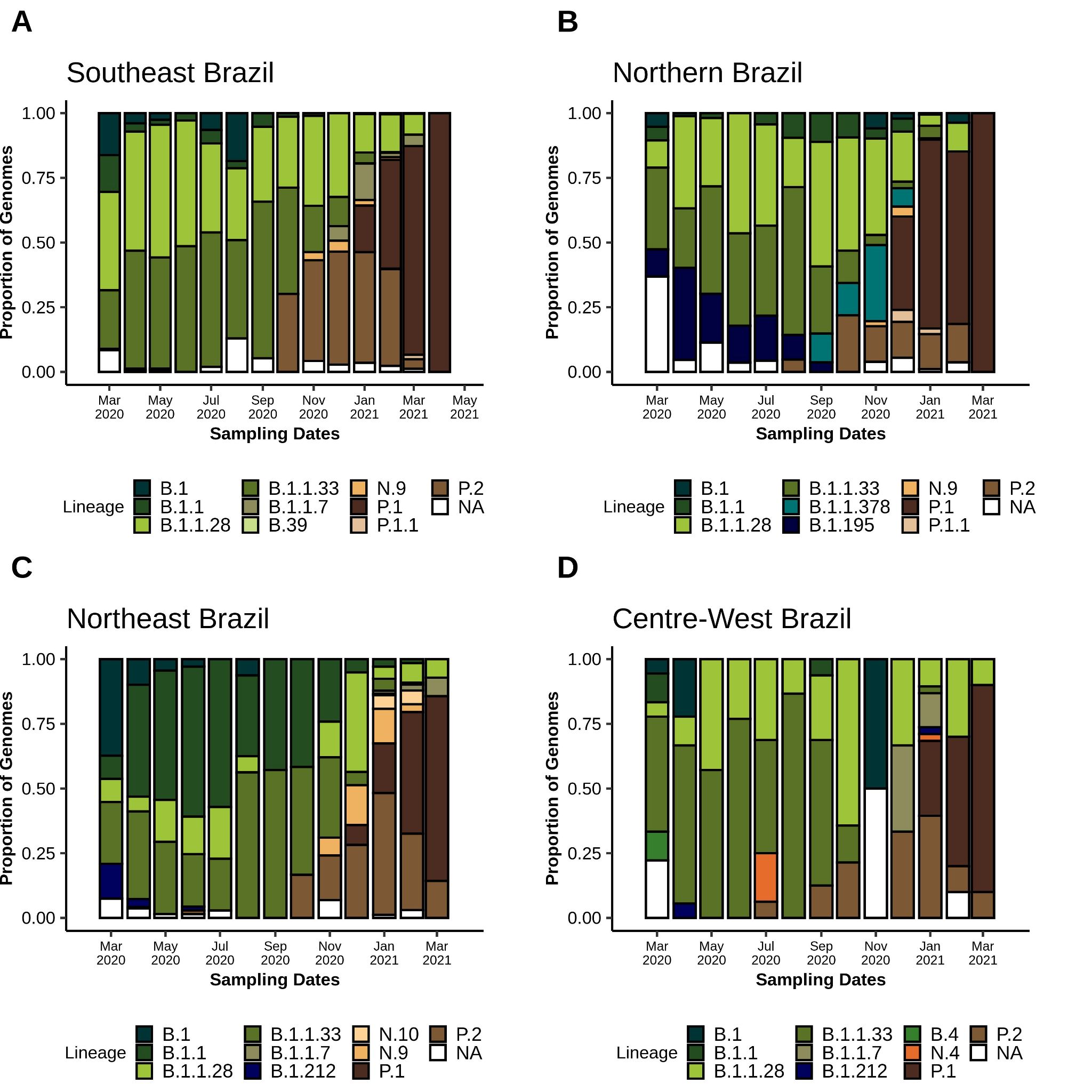
