## Supplementary file 1 for "Predominance of the SARS-CoV-2 lineage P.1 and its sublineage P.1.2 in patients from the metropolitan region of Porto Alegre, Southern Brazil in March 2021: a phylogenomic analysis"

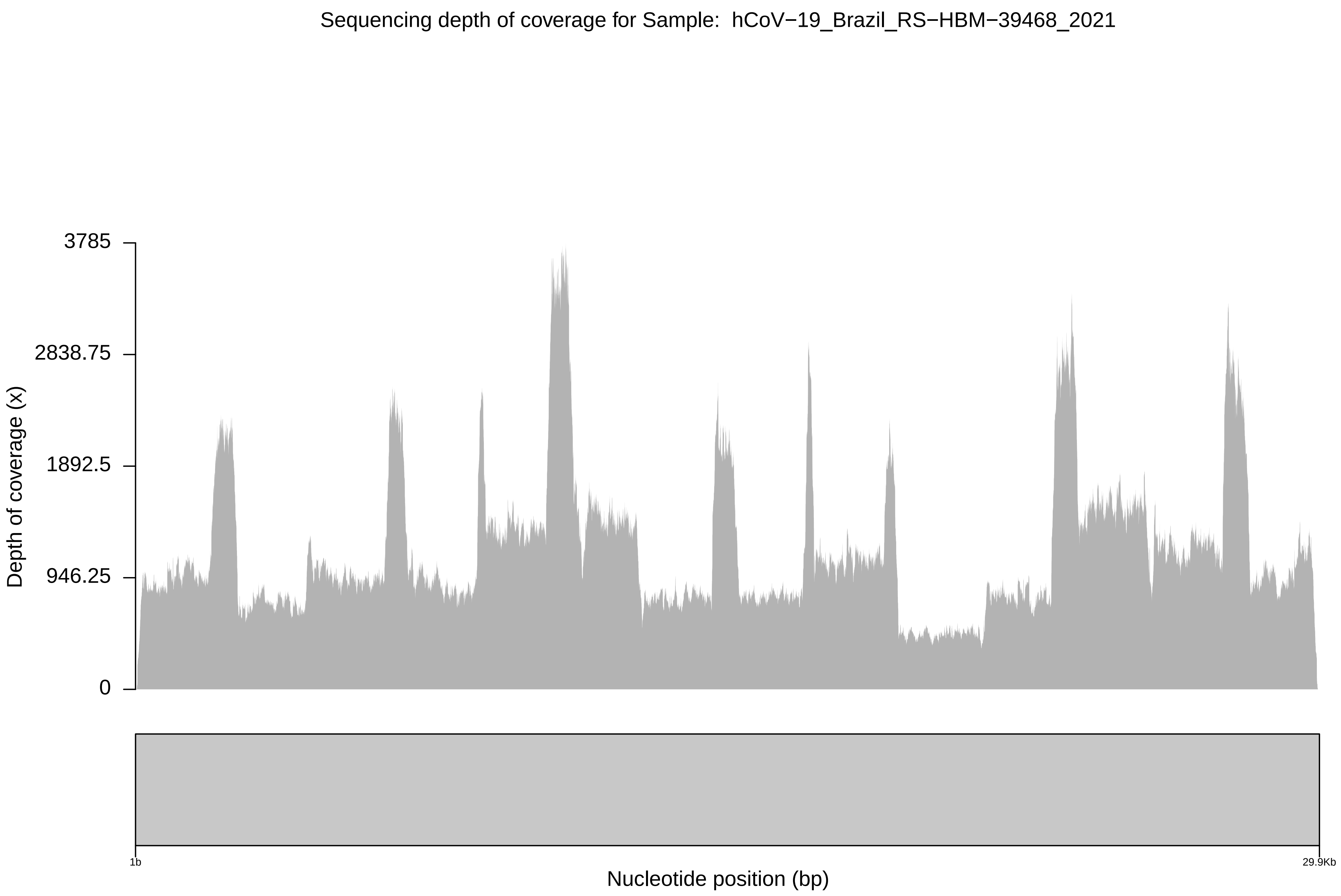
Sequencing depth of coverage for Sample: hCoV−19_Brazil_RS−HBM−39468_2021
3785
2838.75
1892.5
946.25
0
Depth of coverage (x)
1b
29.9Kb
Nucleotide position (bp)

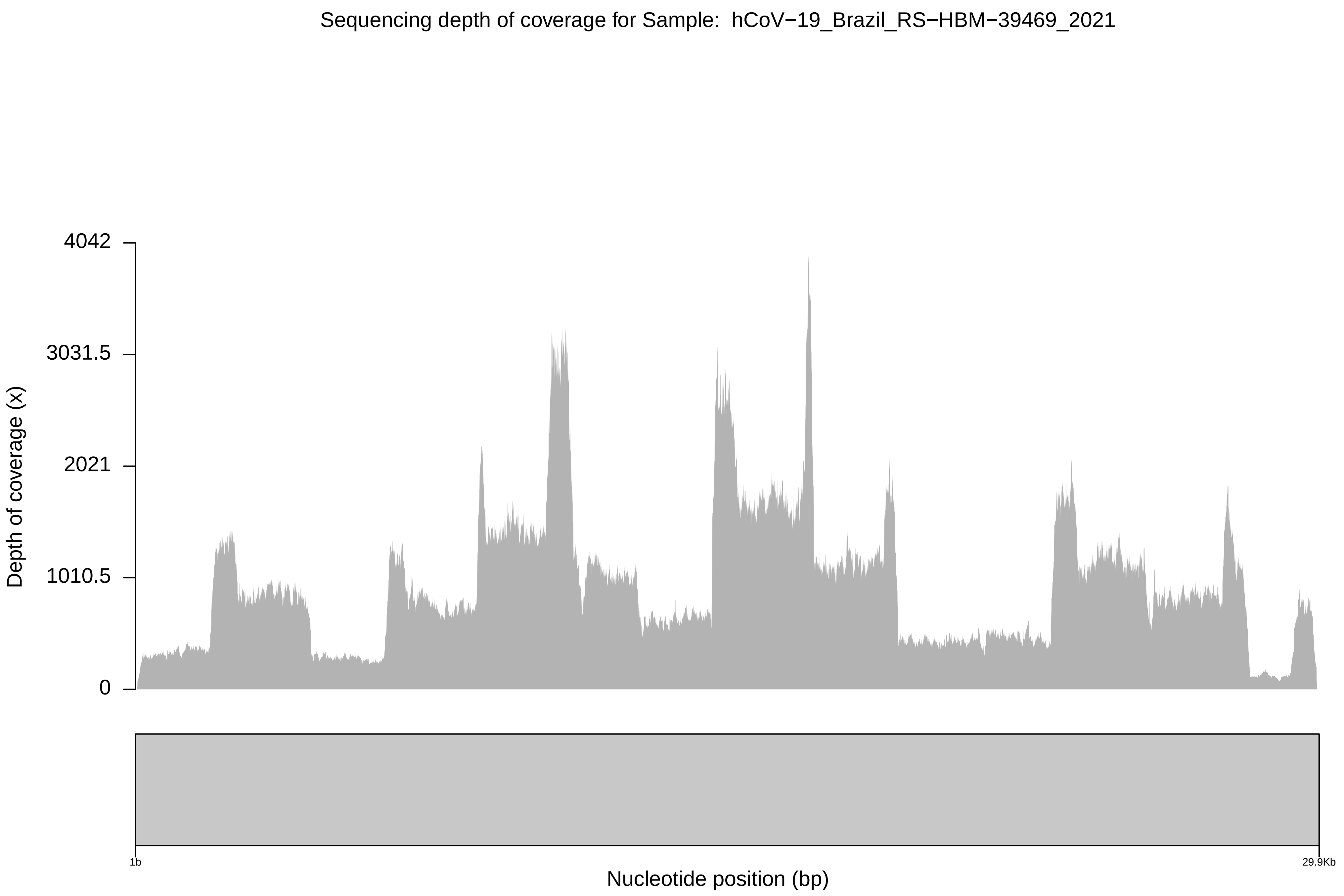
Sequencing depth of coverage for Sample: hCoV−19_Brazil_RS−HBM−39469_2021
Depth of coverage (x)
4042
3031.5
2021
1010.5
0
1b
29.9Kb
Nucleotide position (bp)

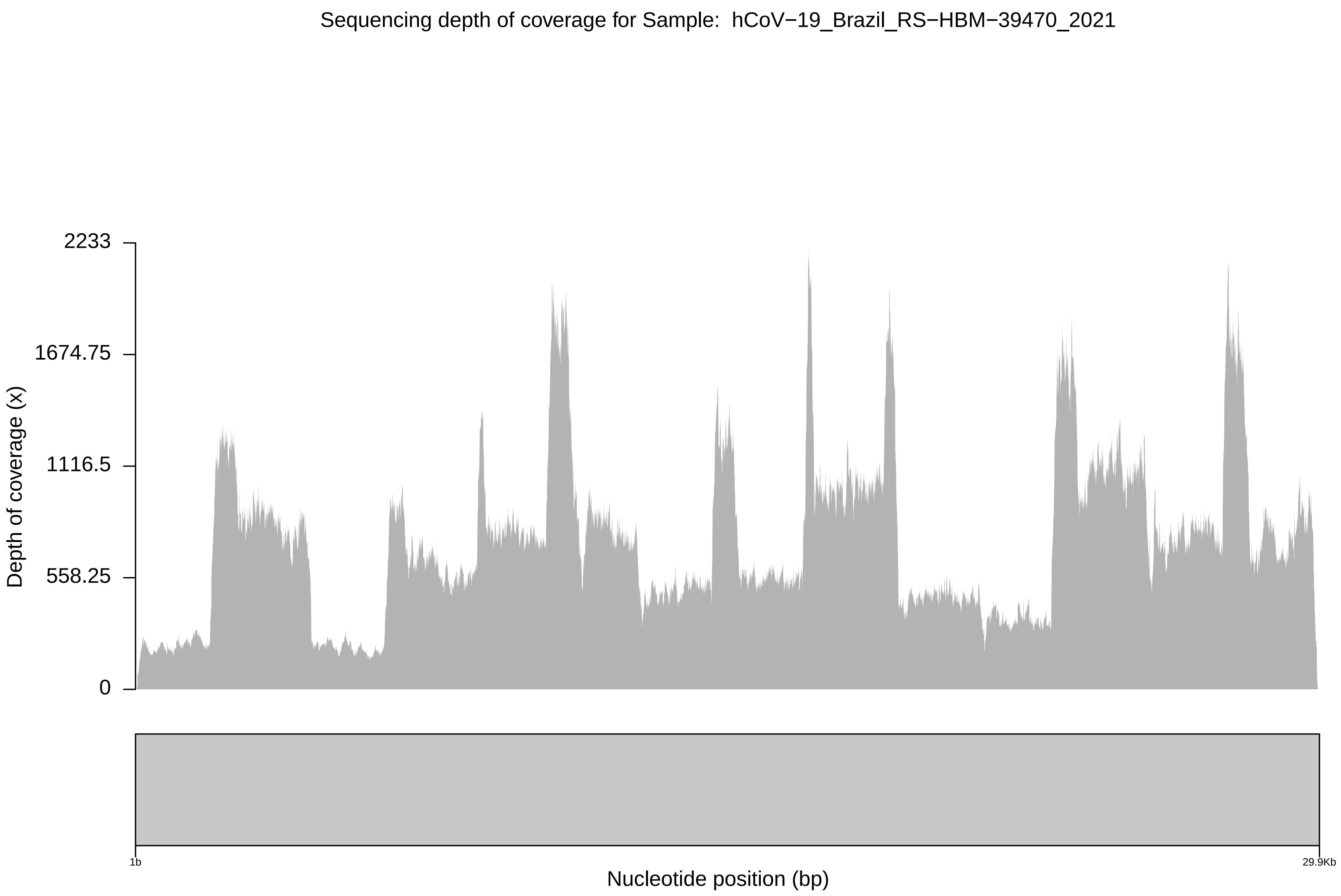
Sequencing depth of coverage for Sample: hCoV−19_Brazil_RS−HBM−39470_2021
Depth of coverage (x)
2233
1674.75
1116.5
558.25
0
1b
29.9Kb
Nucleotide position (bp)

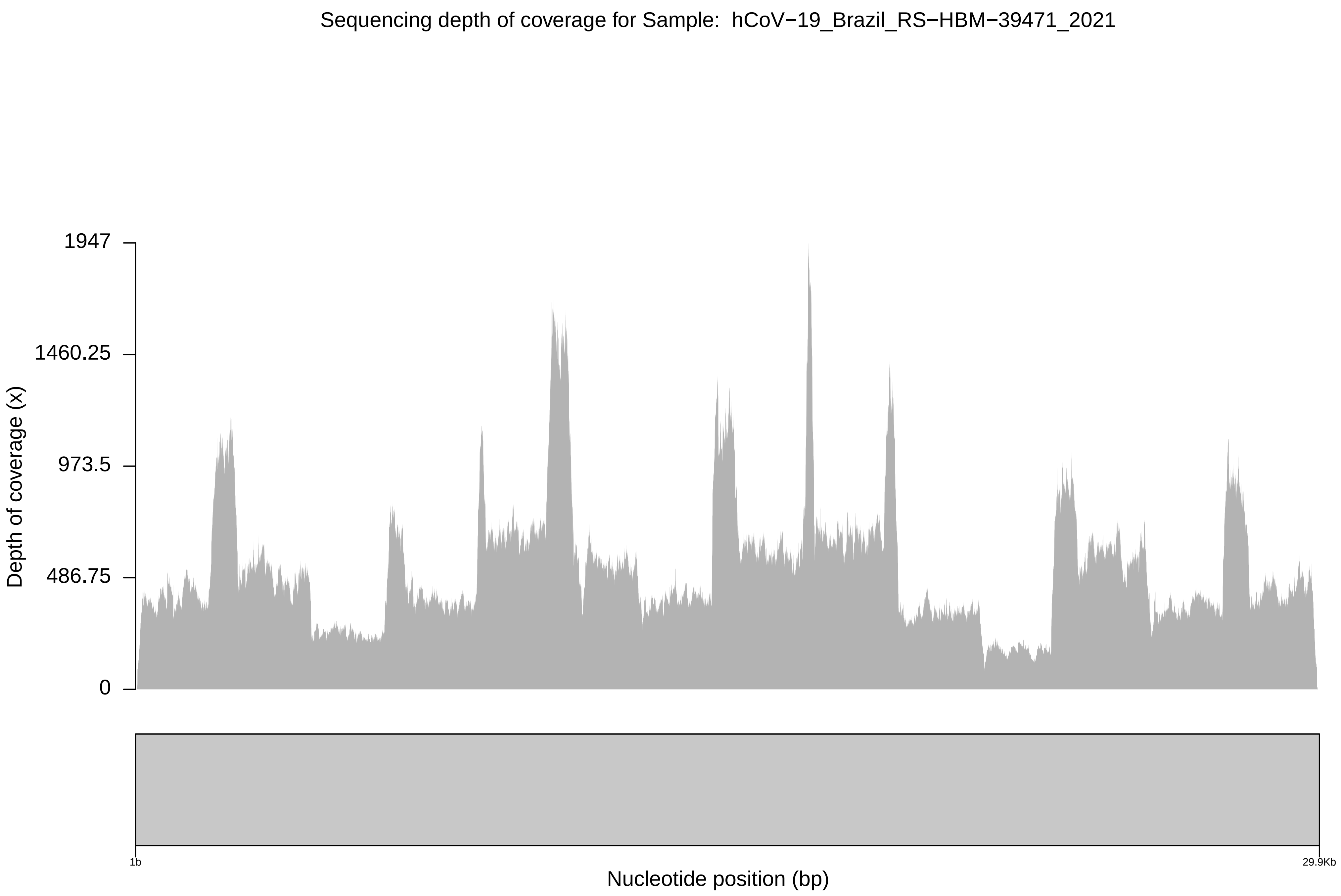
Sequencing depth of coverage for Sample: hCoV−19_Brazil_RS−HBM−39471_2021
Depth of coverage (x)
1947
1460.25
973.5
486.75
0
1b
29.9Kb
Nucleotide position (bp)

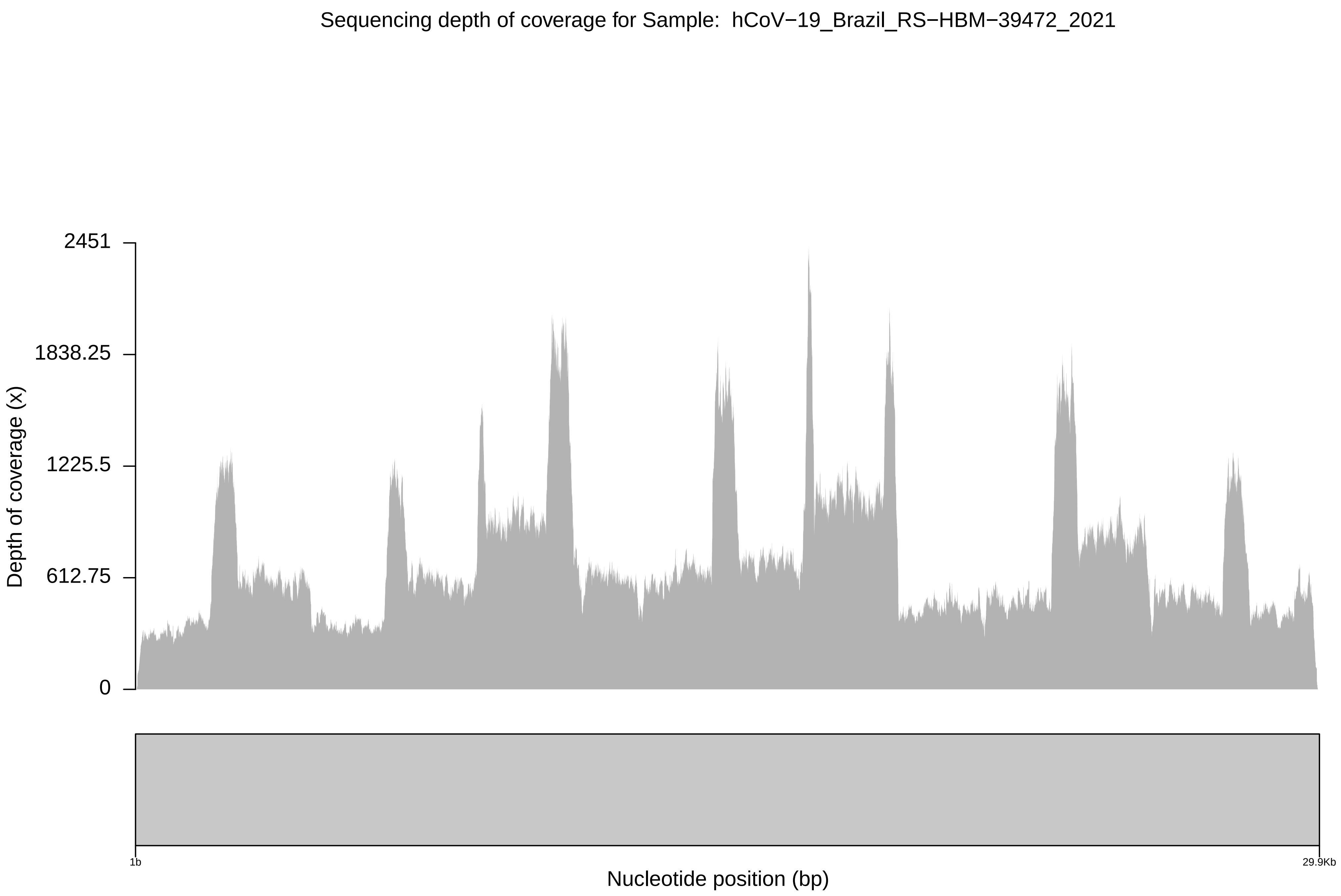

Sequencing depth of coverage for Sample: hCoV−19_Brazil_RS−HBM−39472_2021
2451
1838.25
1225.5
612.75
0
Depth of coverage (x)
1b
29.9Kb
Nucleotide position (bp)

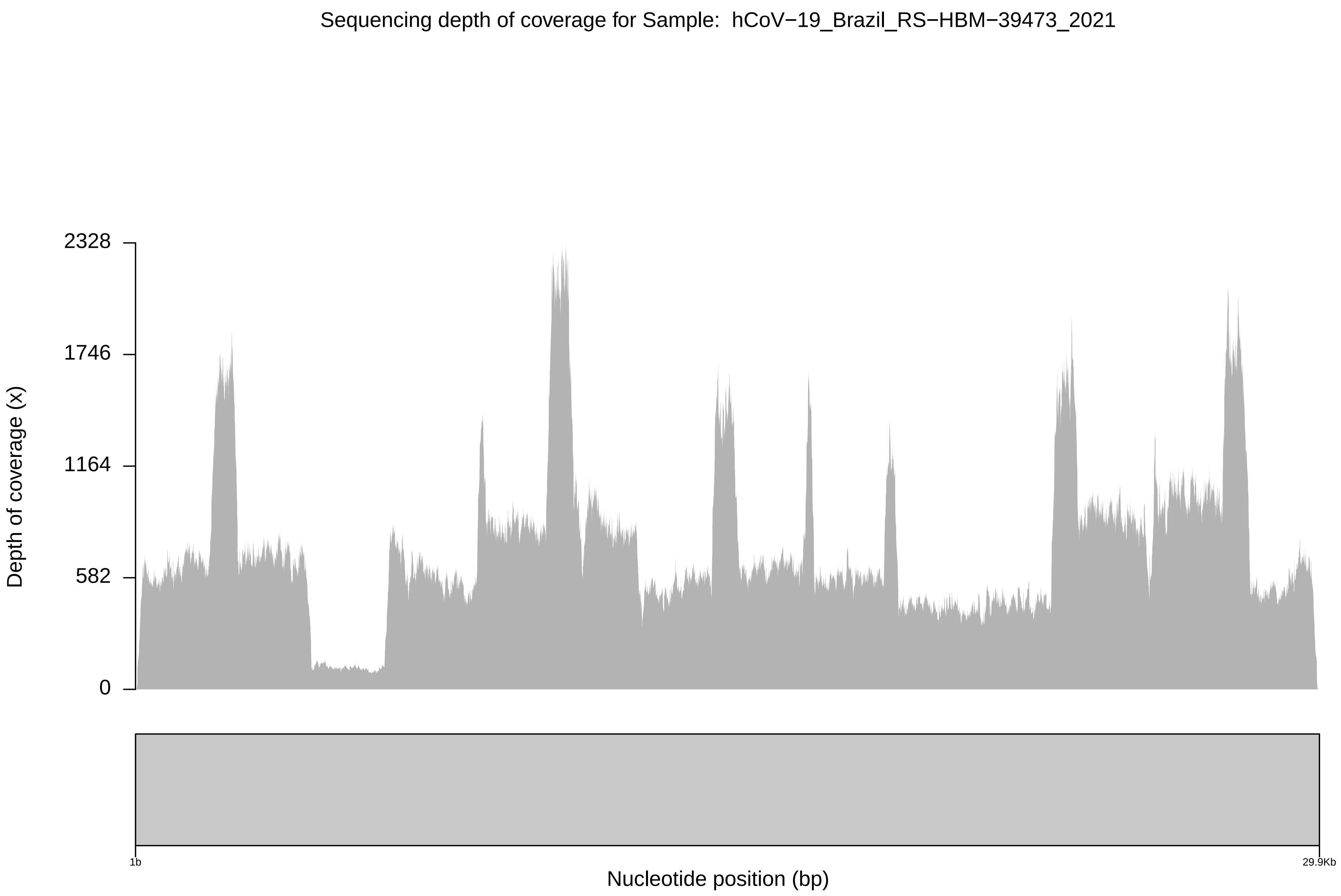

Sequencing depth of coverage for Sample: hCoV−19_Brazil_RS−HBM−39473_2021
Depth of coverage (x)
2328
1746
1164
582
0
1b
29.9Kb
Nucleotide position (bp)

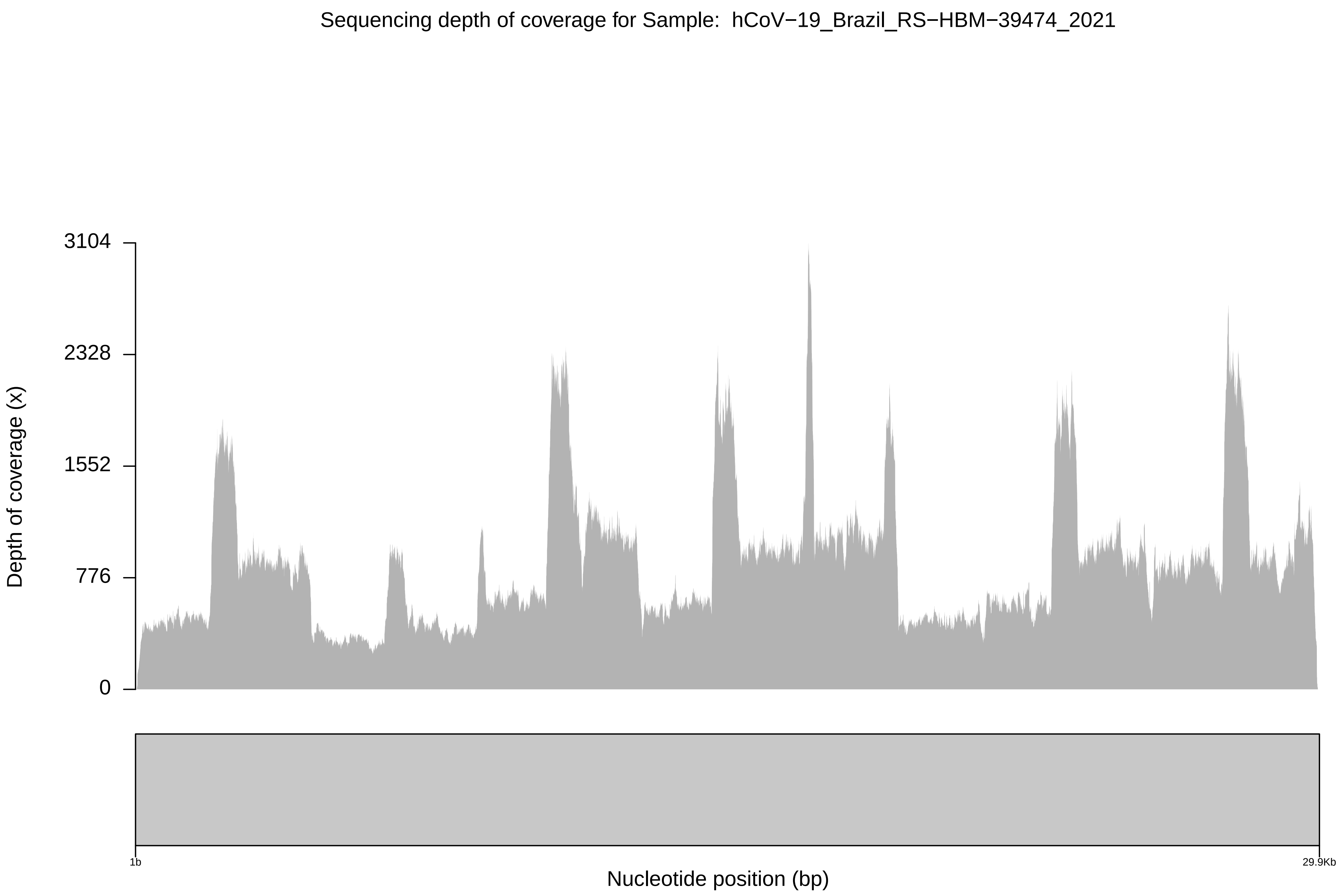

Sequencing depth of coverage for Sample: hCoV−19_Brazil_RS−HBM−39474_2021
3104
2328
1552
776
0
Depth of coverage (x)
1b
29.9Kb
Nucleotide position (bp)

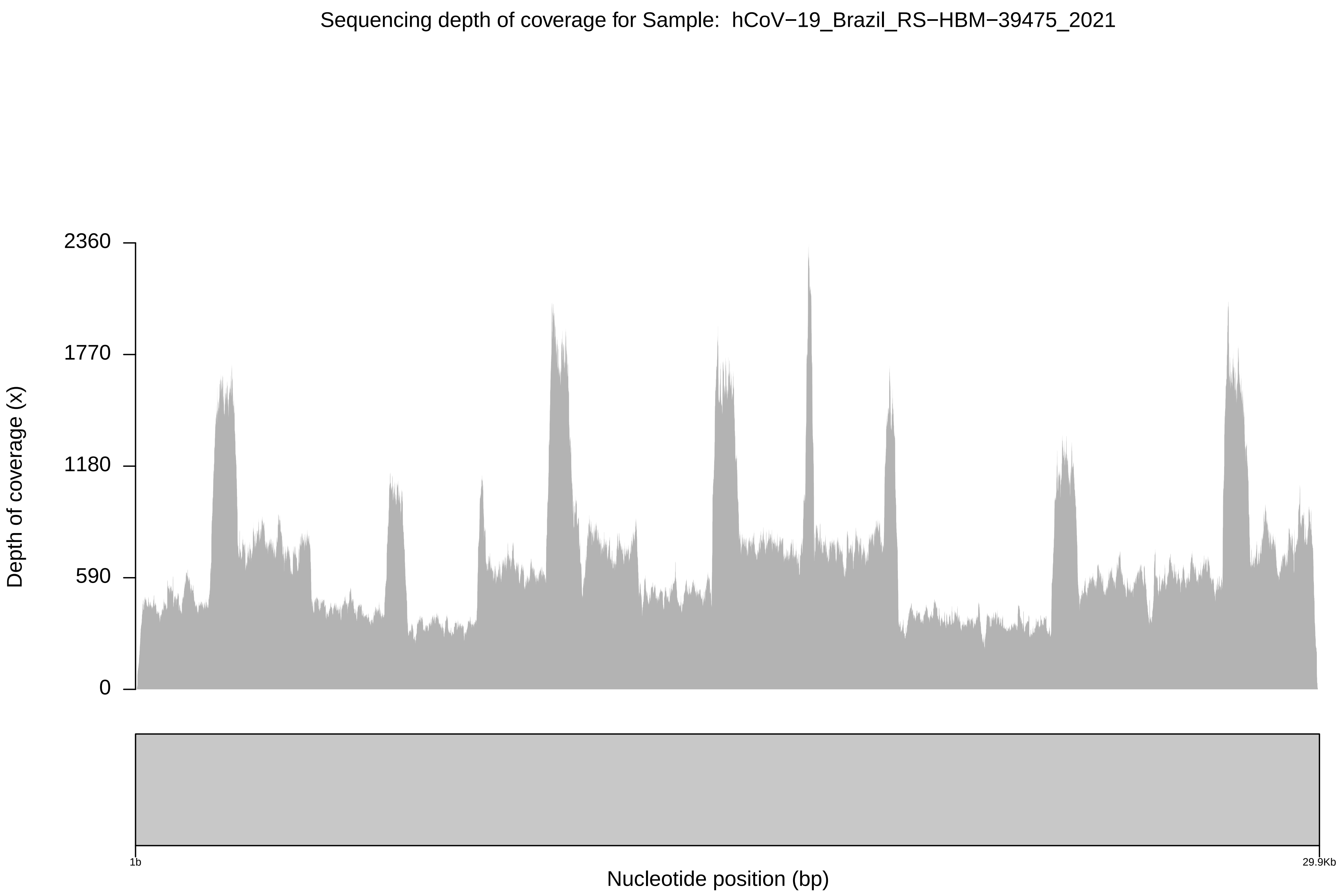

Sequencing depth of coverage for Sample: hCoV−19_Brazil_RS−HBM−39475_2021
Depth of coverage (x)
2360
1770
1180
590
0
1b
29.9Kb
Nucleotide position (bp)

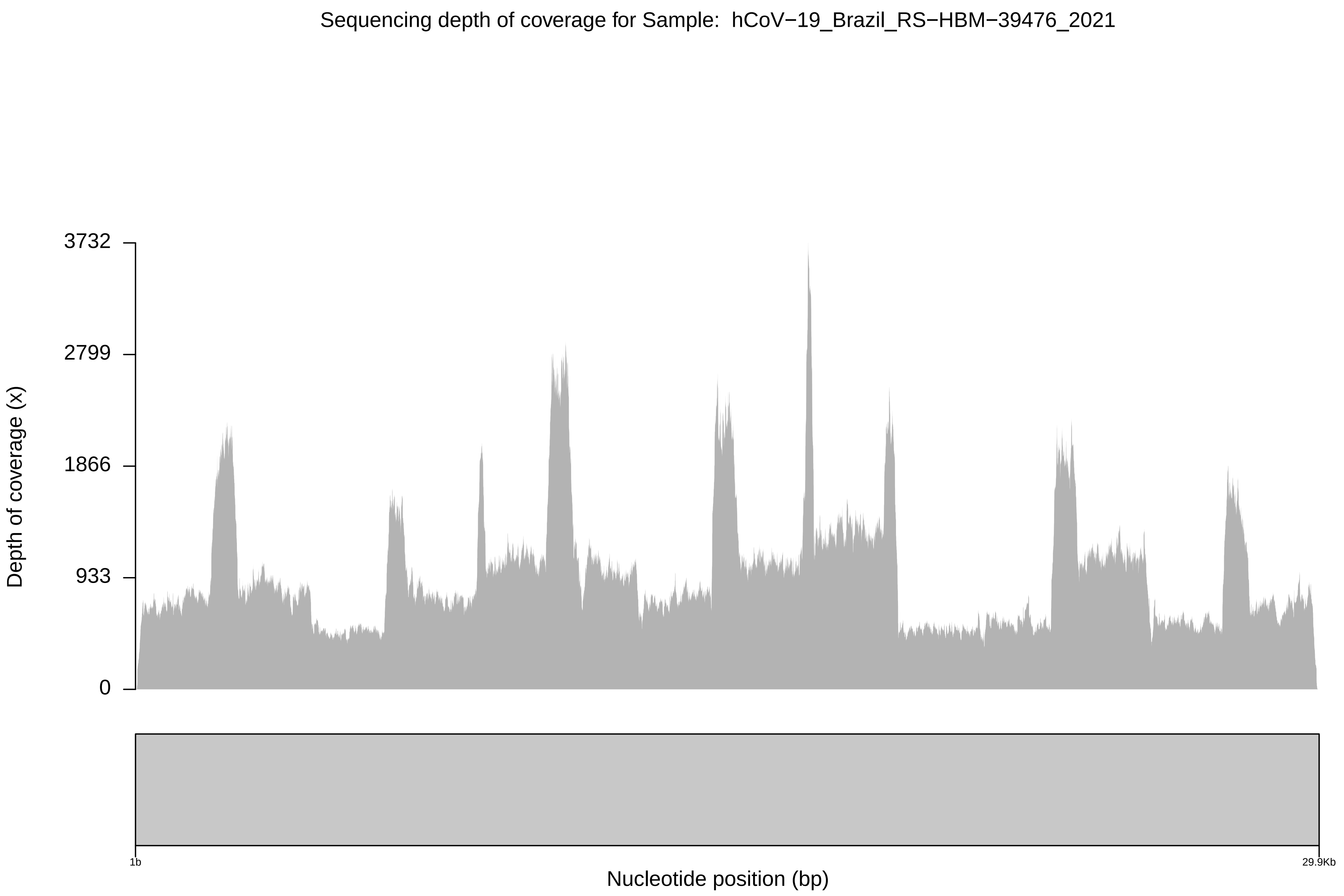
Sequencing depth of coverage for Sample: hCoV−19_Brazil_RS−HBM−39476_2021
3732
2799
1866
933
0
Depth of coverage (x)
1b
29.9Kb
Nucleotide position (bp)

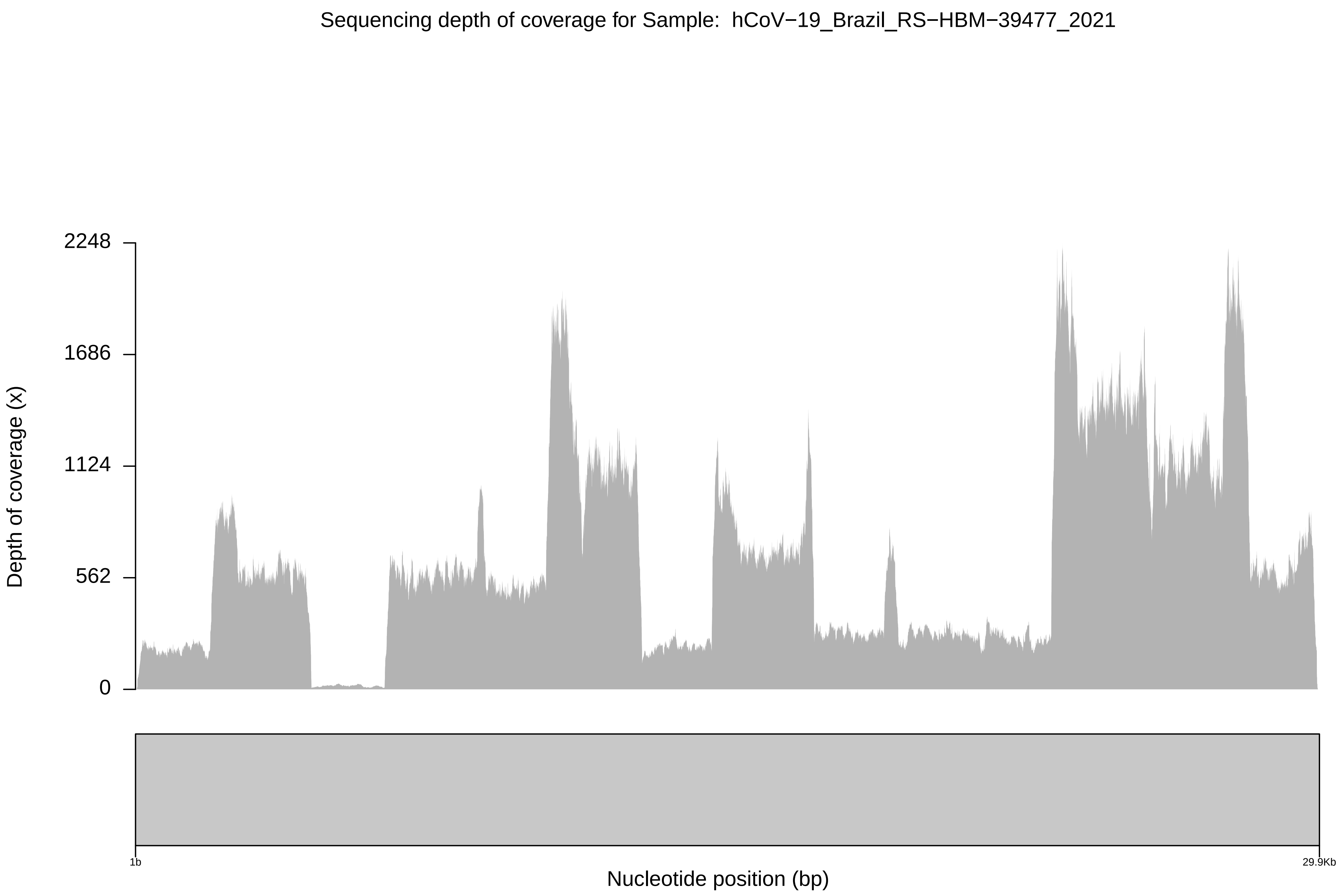

Sequencing depth of coverage for Sample: hCoV−19_Brazil_RS−HBM−39477_2021
Depth of coverage (x)
2248
1686
1124
562
0
1b
29.9Kb
Nucleotide position (bp)

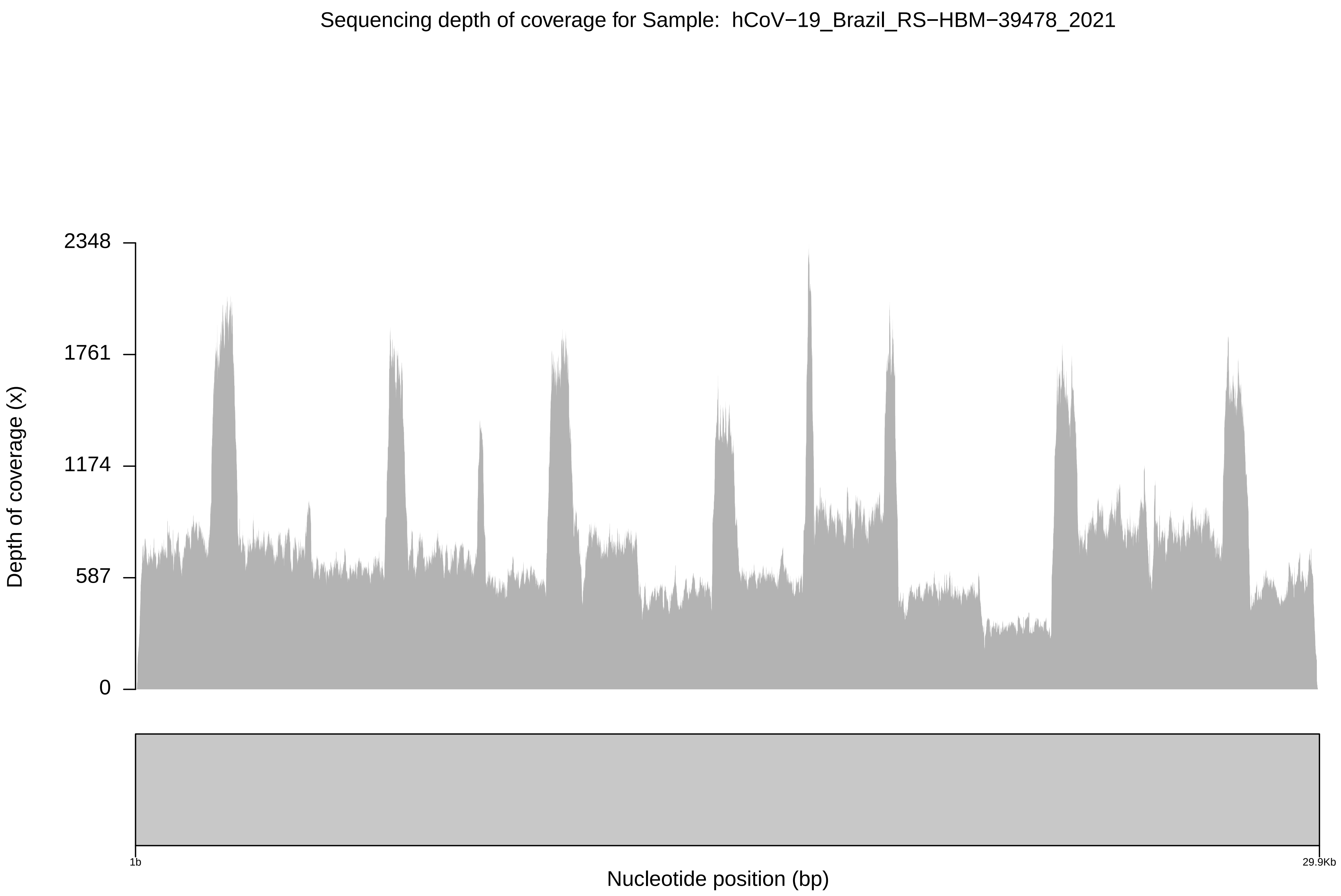

Sequencing depth of coverage for Sample: hCoV−19_Brazil_RS−HBM−39478_2021
2348
1761
1174
587
0
Depth of coverage (x)
1b
29.9Kb
Nucleotide position (bp)

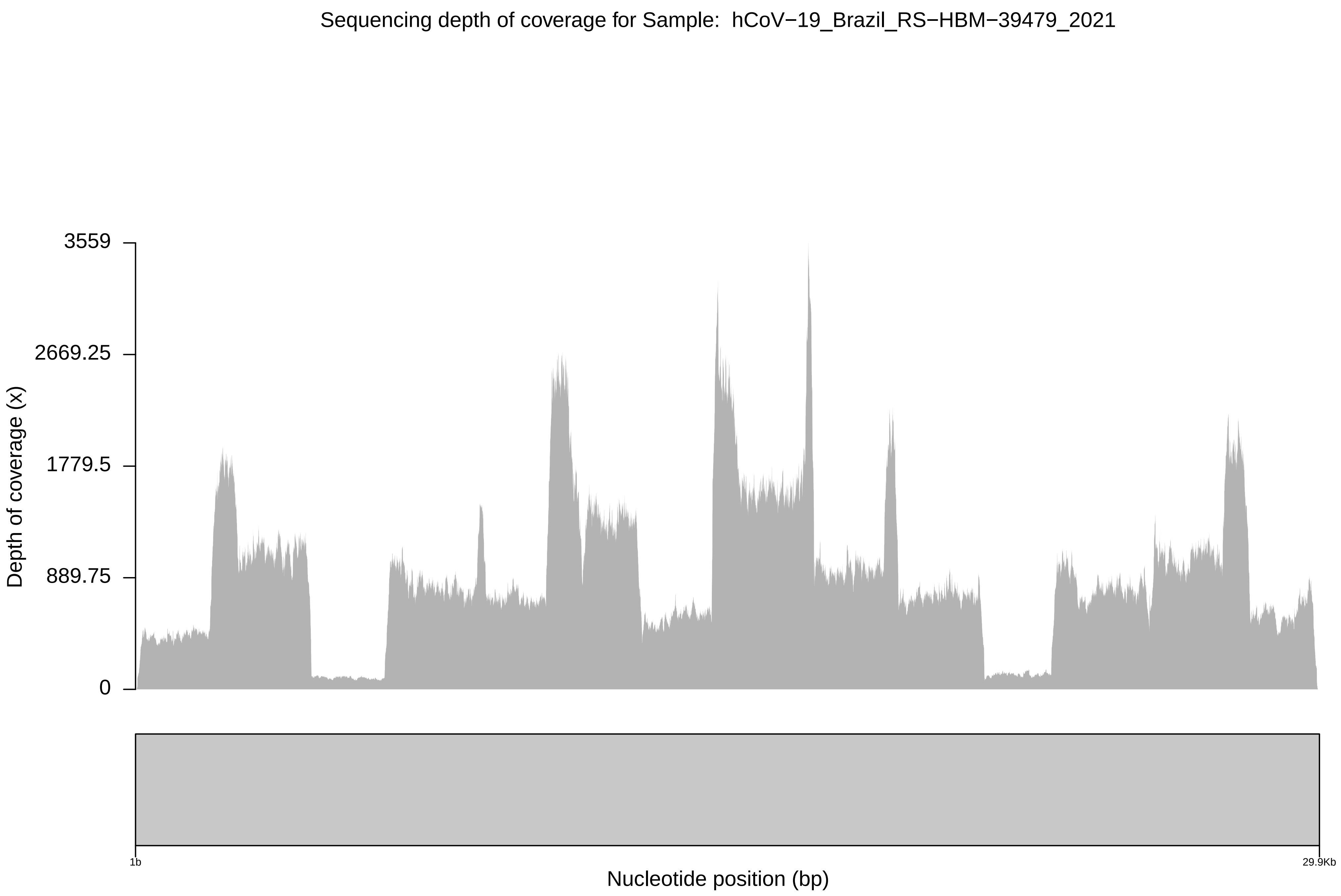

Sequencing depth of coverage for Sample: hCoV−19_Brazil_RS−HBM−39479_2021
3559
2669.25
1779.5
889.75
0
Depth of coverage (x)
1b
29.9Kb
Nucleotide position (bp)

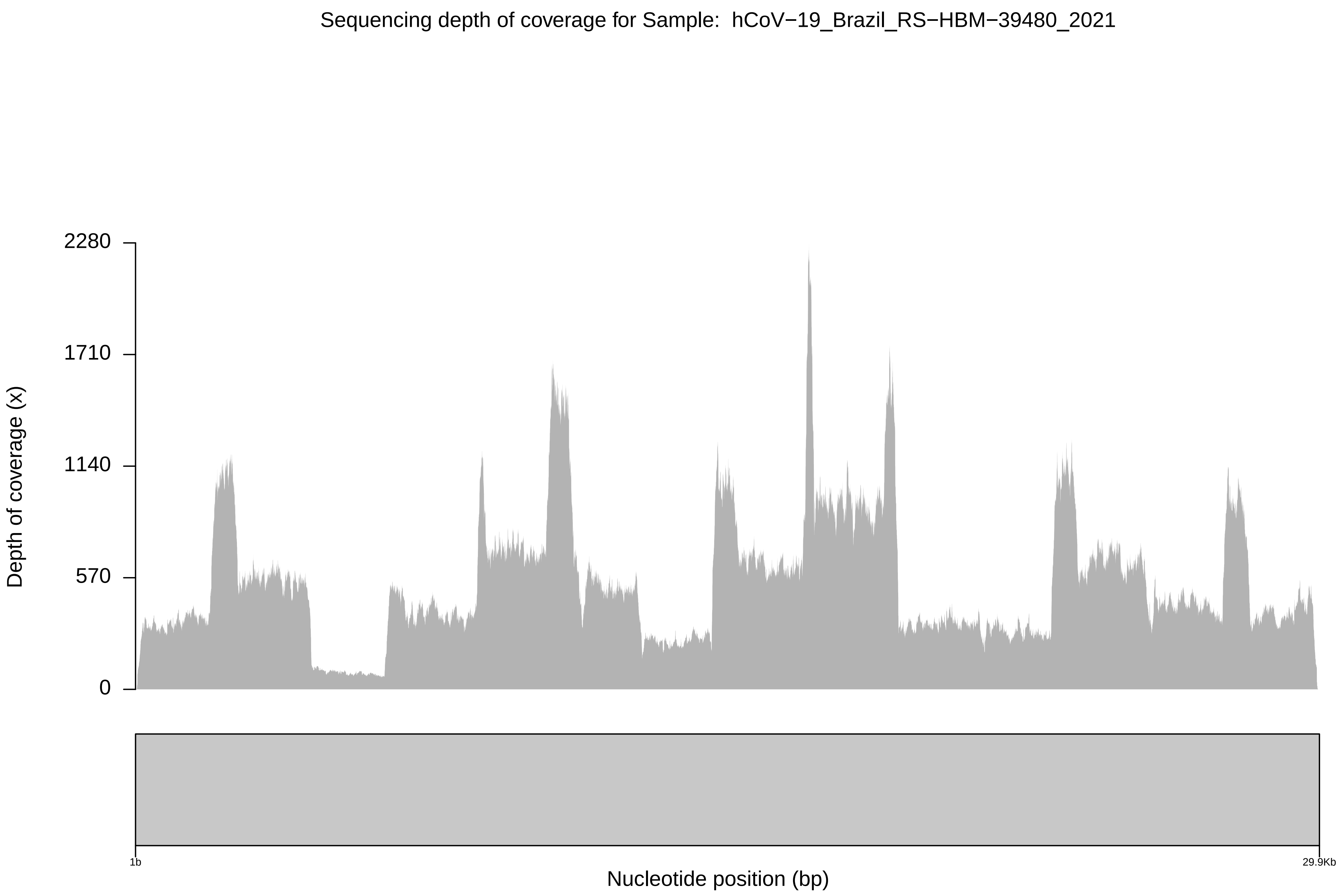

Sequencing depth of coverage for Sample: hCoV−19_Brazil_RS−HBM−39480_2021
2280
1710
1140
570
0
Depth of coverage (x)
1b
29.9Kb
Nucleotide position (bp)

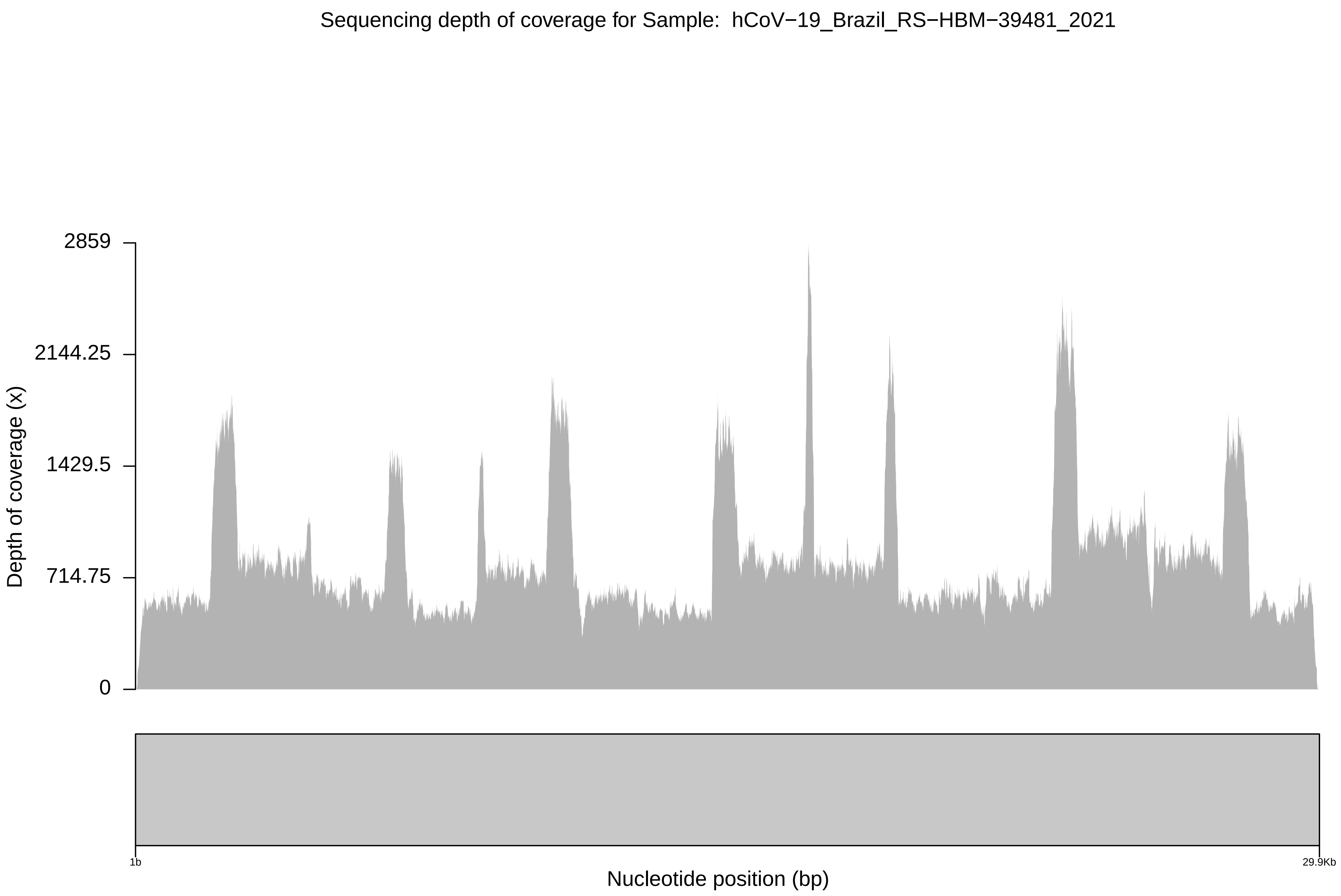

Sequencing depth of coverage for Sample: hCoV−19_Brazil_RS−HBM−39481_2021
2859
2144.25
1429.5
714.75
0
Depth of coverage (x)
1b
29.9Kb
Nucleotide position (bp)

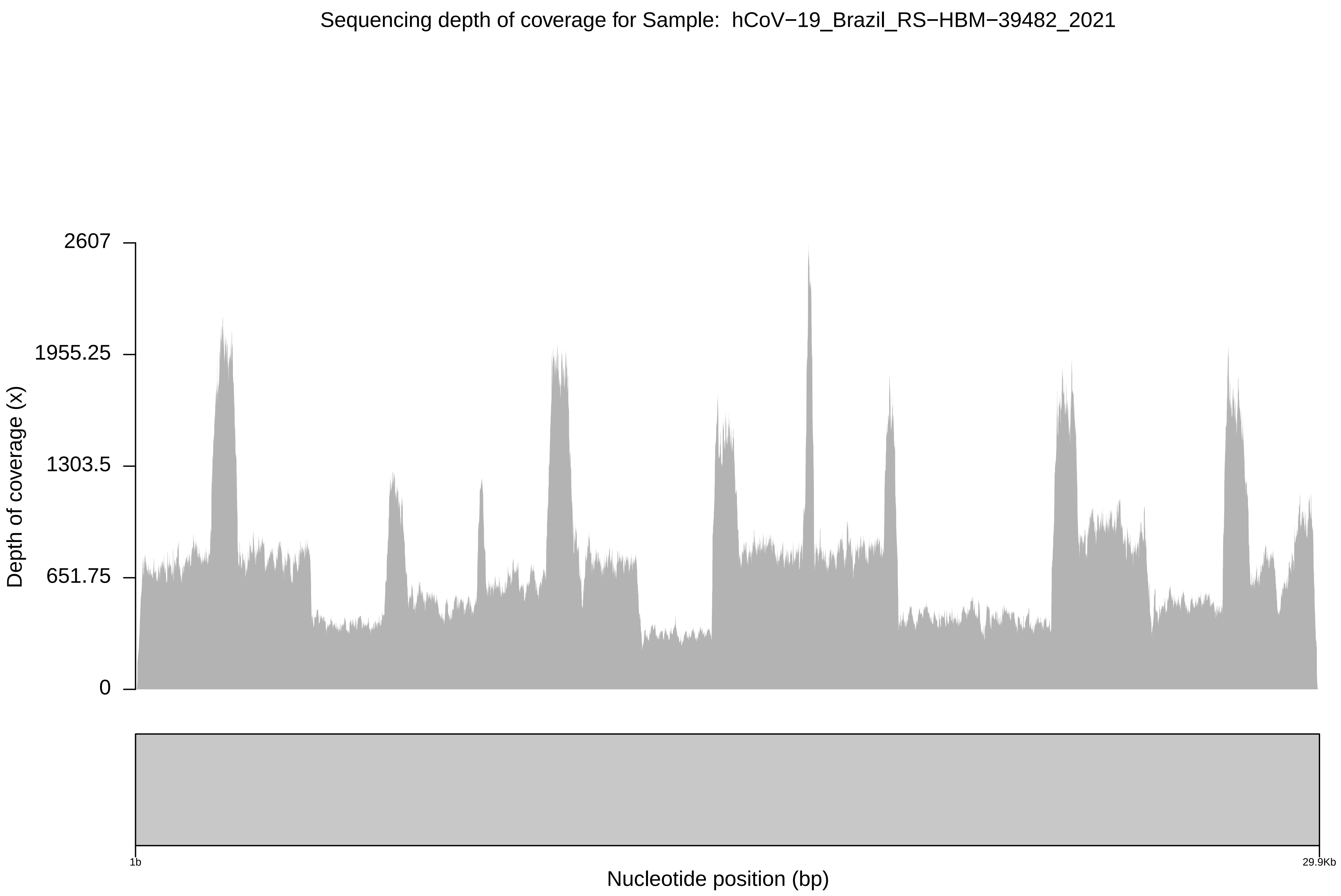

Sequencing depth of coverage for Sample: hCoV−19_Brazil_RS−HBM−39482_2021
2607
1955.25
1303.5
651.75
0
Depth of coverage (x)
1b
29.9Kb
Nucleotide position (bp)

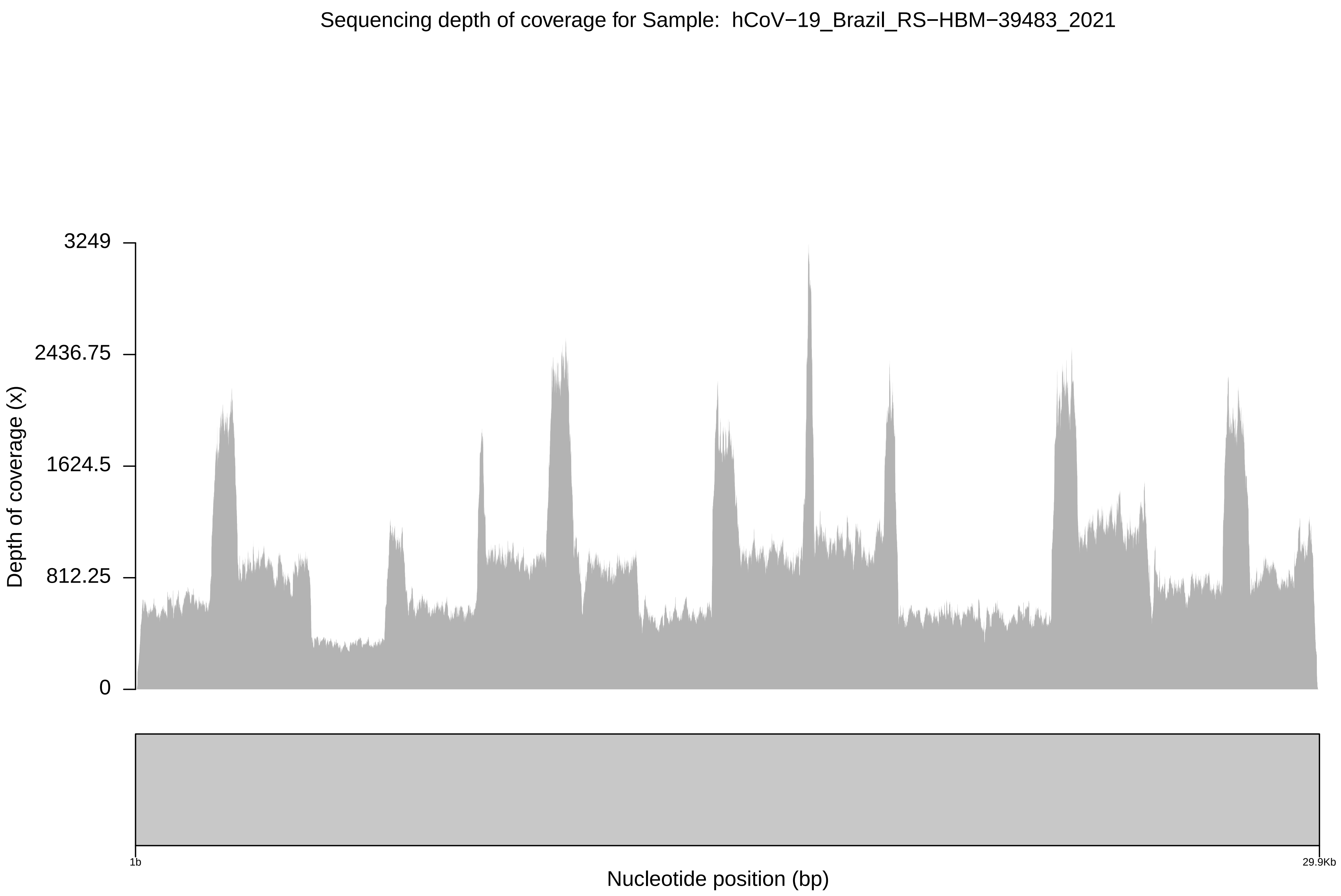
Sequencing depth of coverage for Sample: hCoV−19_Brazil_RS−HBM−39483_2021
3249
2436.75
1624.5
812.25
0
Depth of coverage (x)
1b
29.9Kb
Nucleotide position (bp)

Sequencing depth of coverage for Sample: hCoV−19_Brazil_RS−HBM−39484_2021

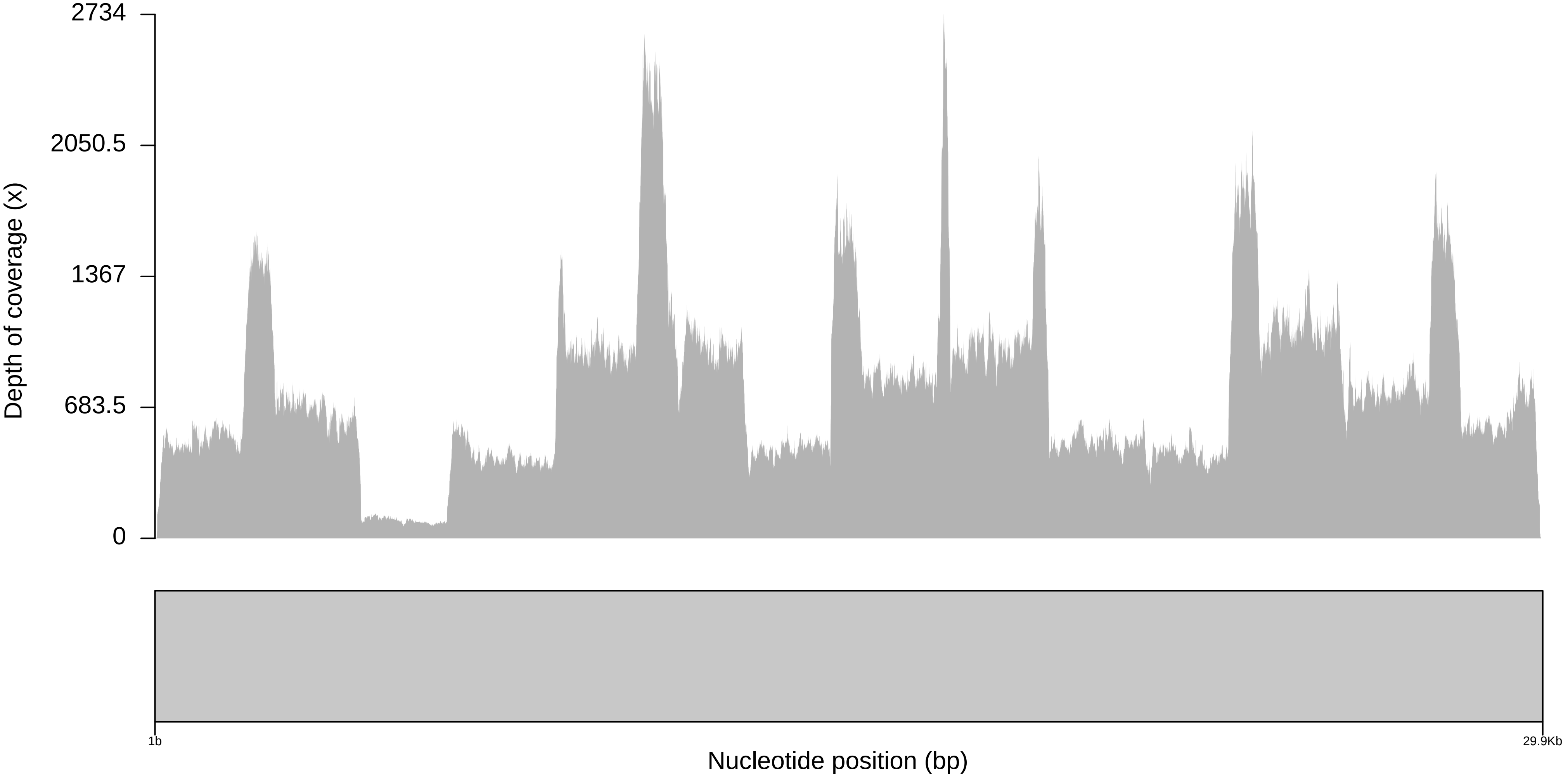

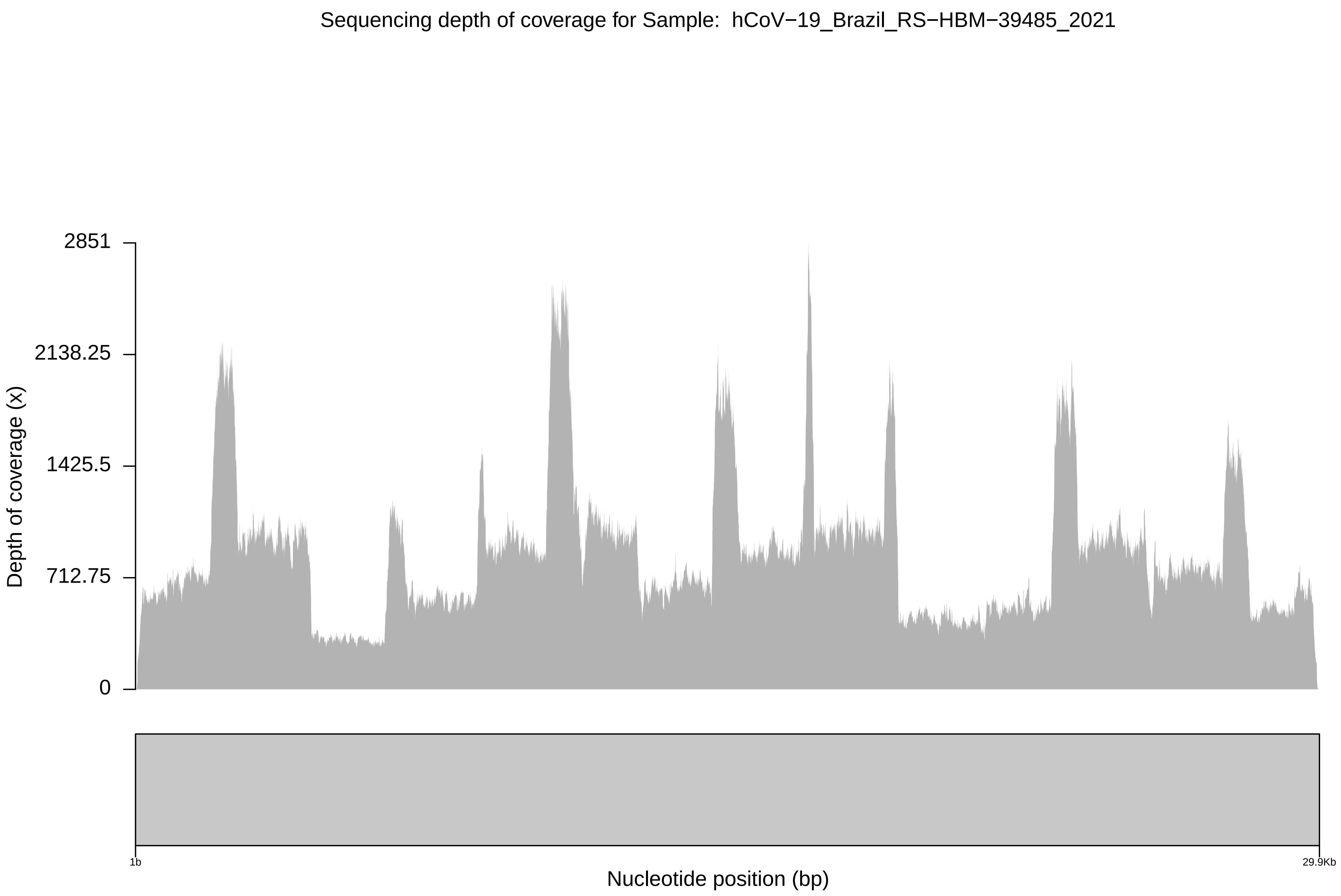

Sequencing depth of coverage for Sample: hCoV−19_Brazil_RS−HBM−39485_2021
2851
2138.25
1425.5
712.75
0
Depth of coverage (x)
1b
29.9Kb
Nucleotide position (bp)

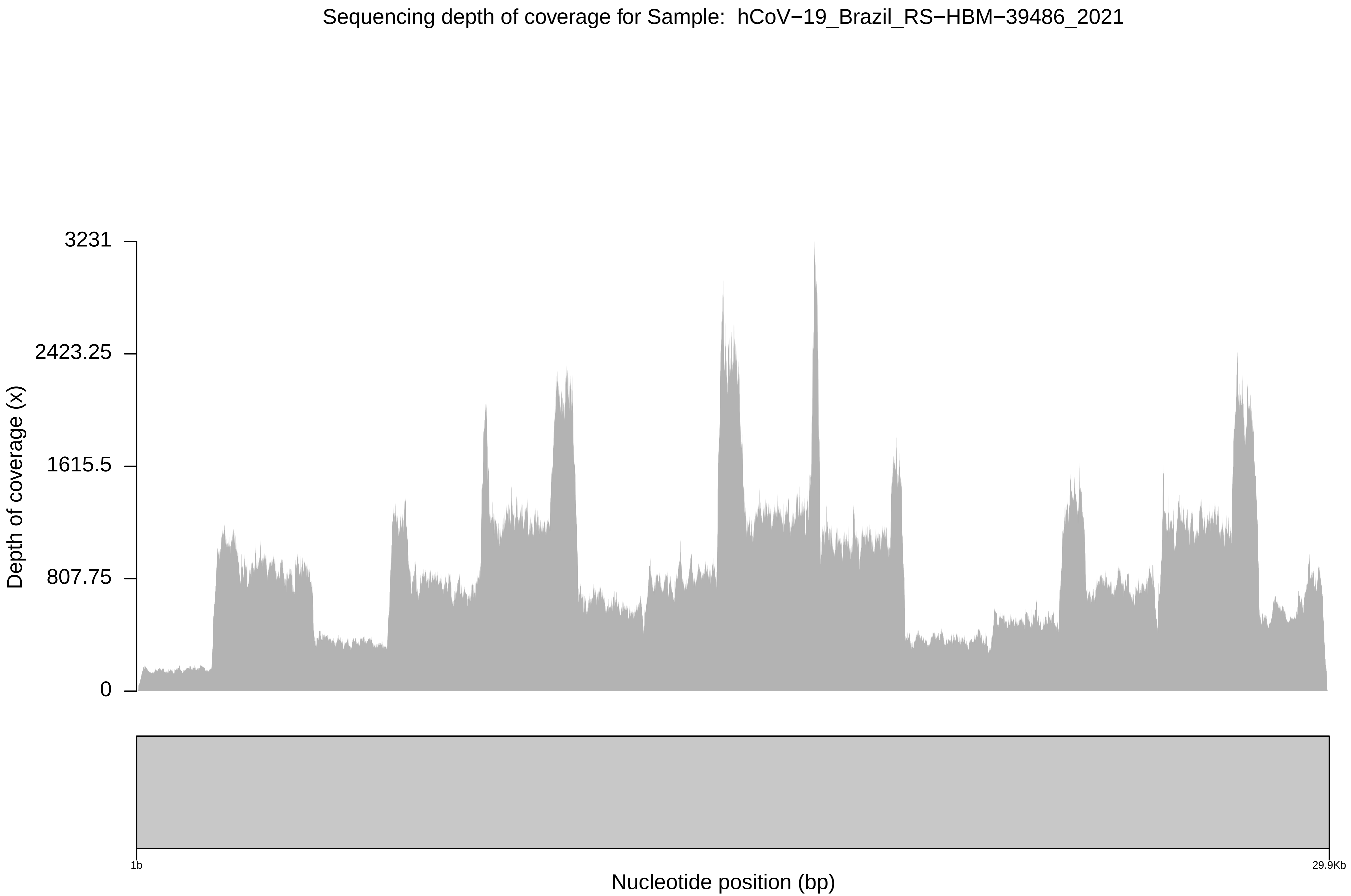

Sequencing depth of coverage for Sample: hCoV−19_Brazil_RS−HBM−39486_2021
3231
2423.25
1615.5
807.75
0
Depth of coverage (x)
1b
29.9Kb
Nucleotide position (bp)

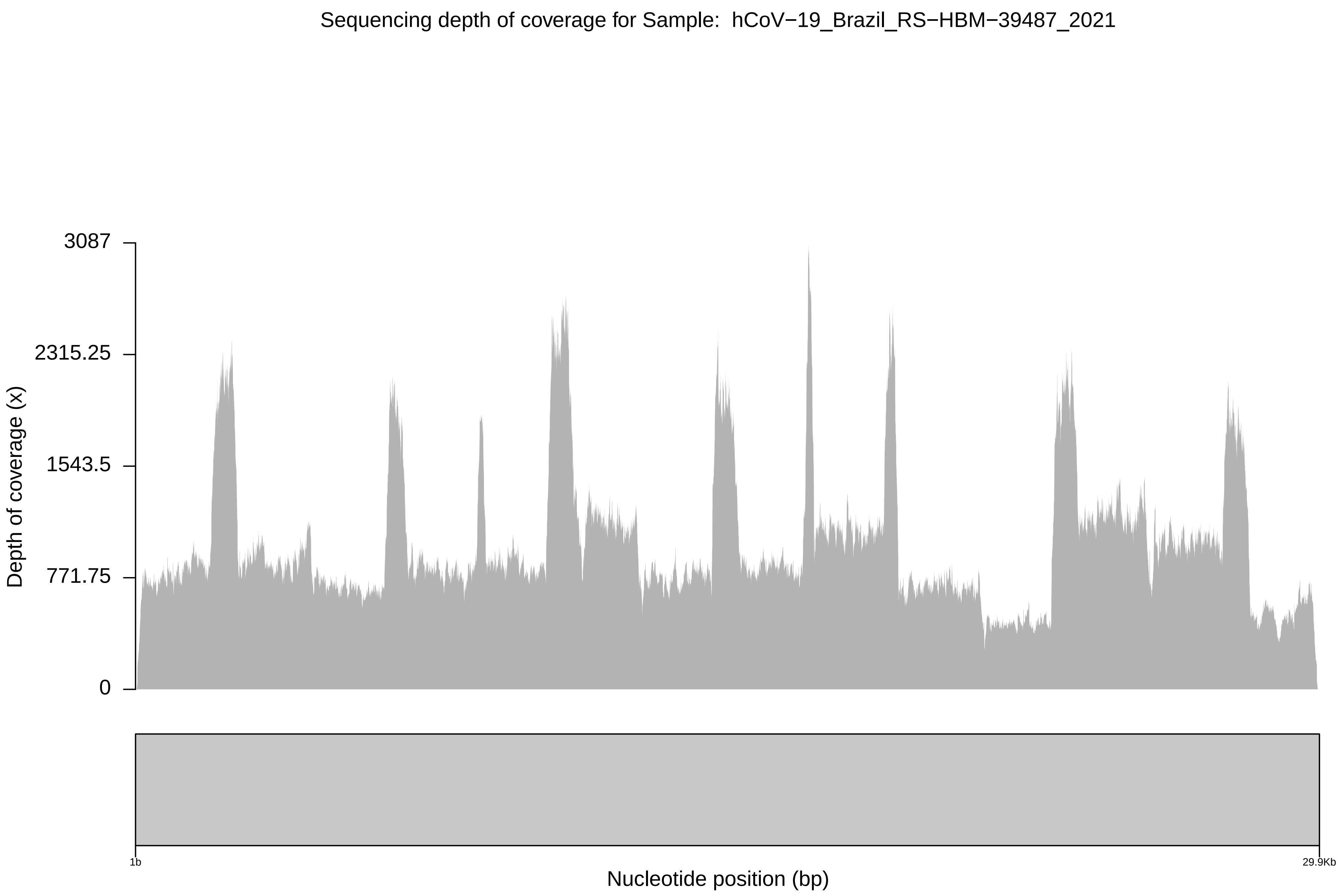

Sequencing depth of coverage for Sample: hCoV−19_Brazil_RS−HBM−39487_2021
3087
2315.25
1543.5
771.75
0
Depth of coverage (x)
1b
29.9Kb
Nucleotide position (bp)

Sequencing depth of coverage for Sample:  hCoV−19_Brazil_RS−HBM−39488_2021

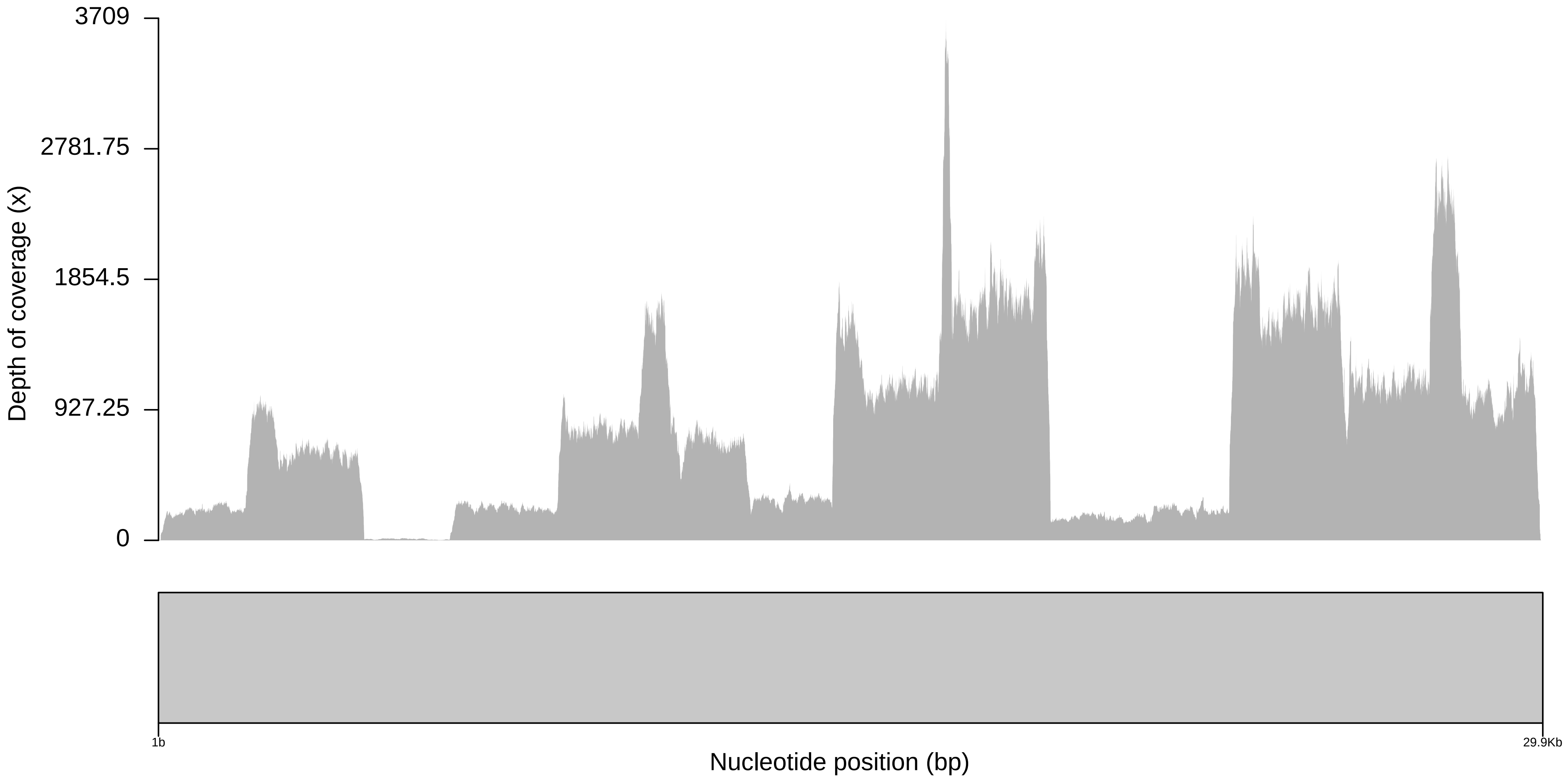

Sequencing depth of coverage for Sample:  hCoV−19_Brazil_RS−HBM−39489_2021

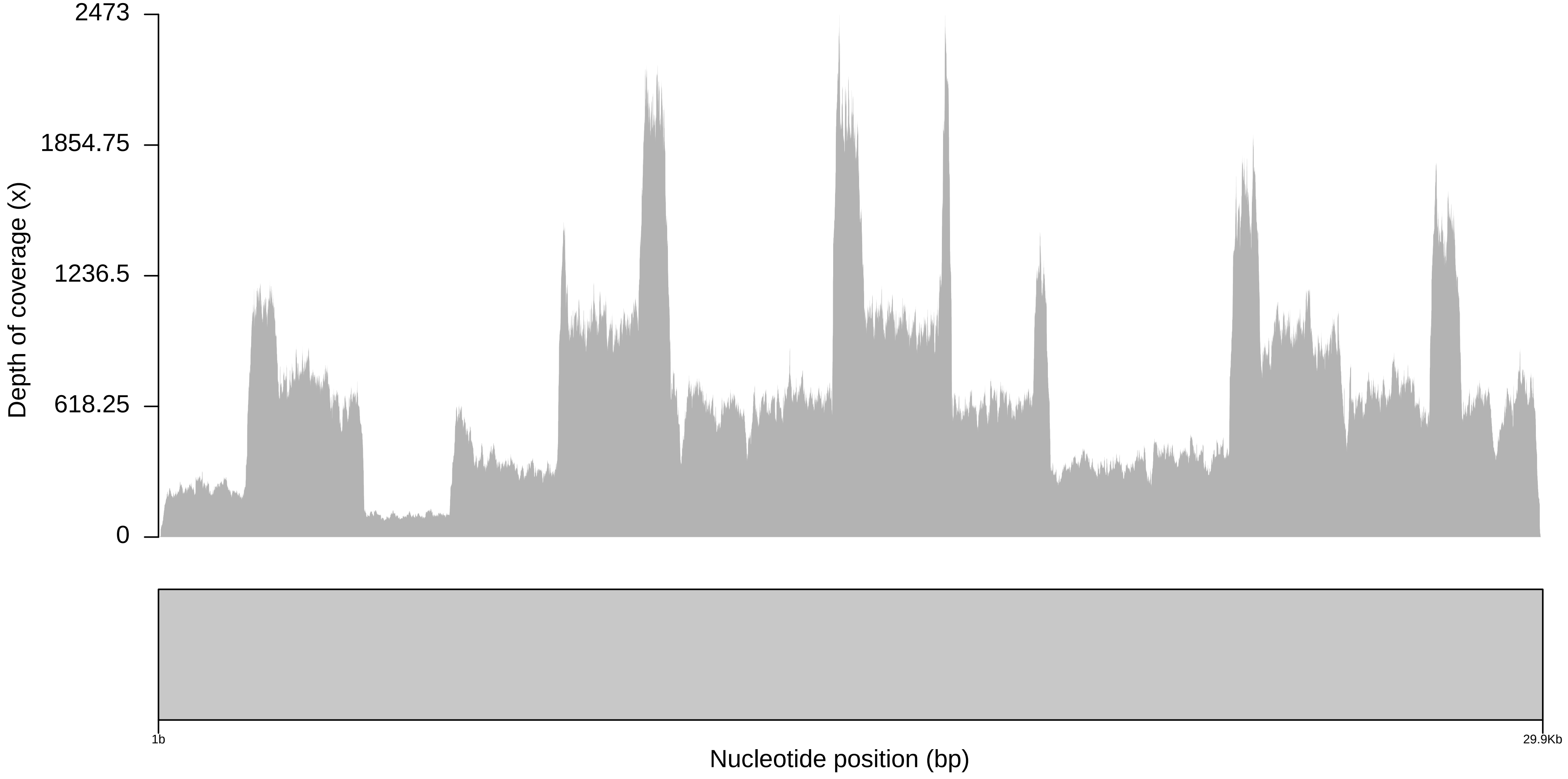

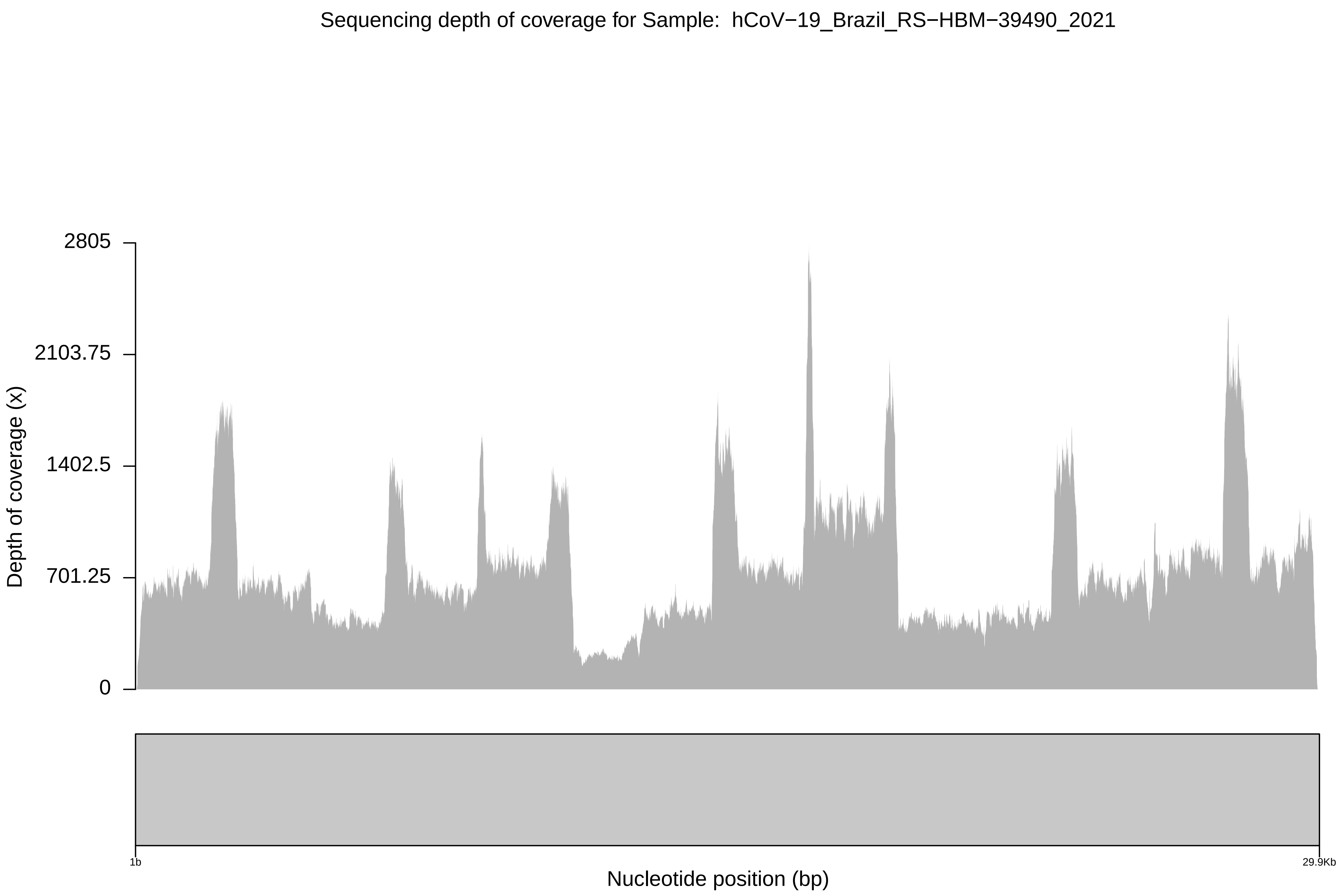
Sequencing depth of coverage for Sample: hCoV−19_Brazil_RS−HBM−39490_2021
Depth of coverage (x)
2805
2103.75
1402.5
701.25
0
1b
29.9Kb
Nucleotide position (bp)

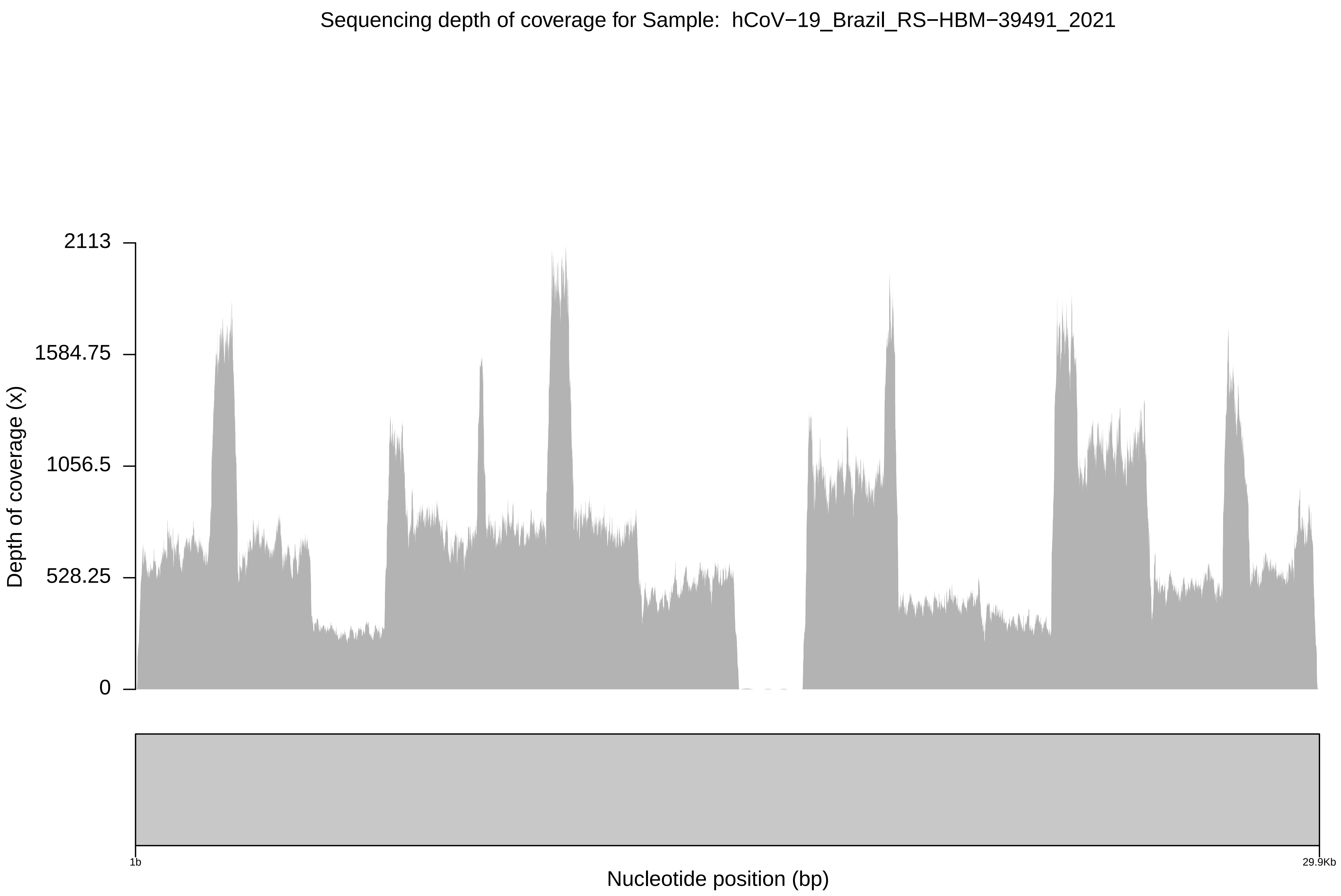
Sequencing depth of coverage for Sample: hCoV−19_Brazil_RS−HBM−39491_2021
2113
1584.75
1056.5
528.25
0
Depth of coverage (x)
1b
29.9Kb
Nucleotide position (bp)

Sequencing depth of coverage for Sample: hCoV−19_Brazil_RS−HBM−39492_2021

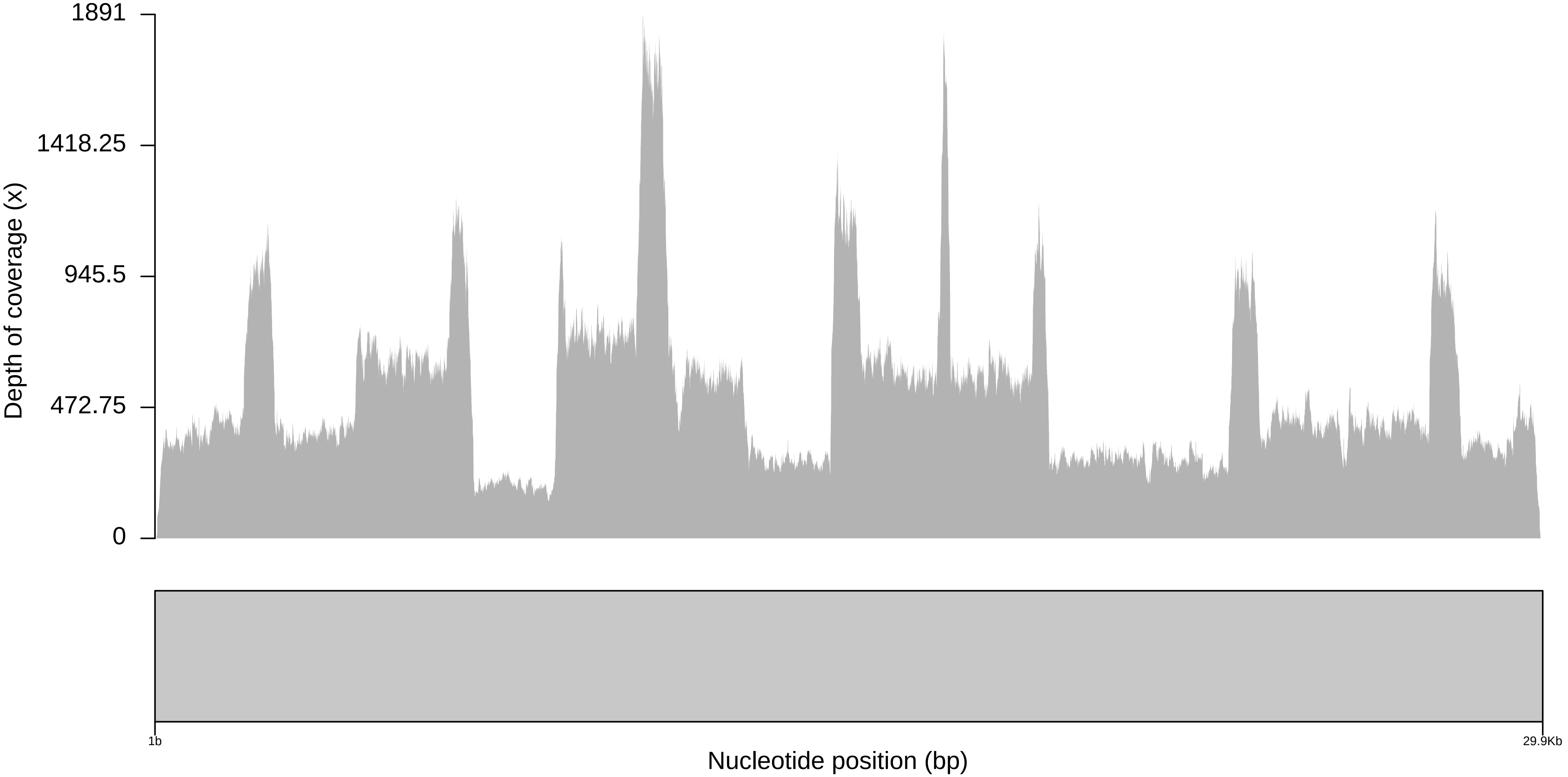

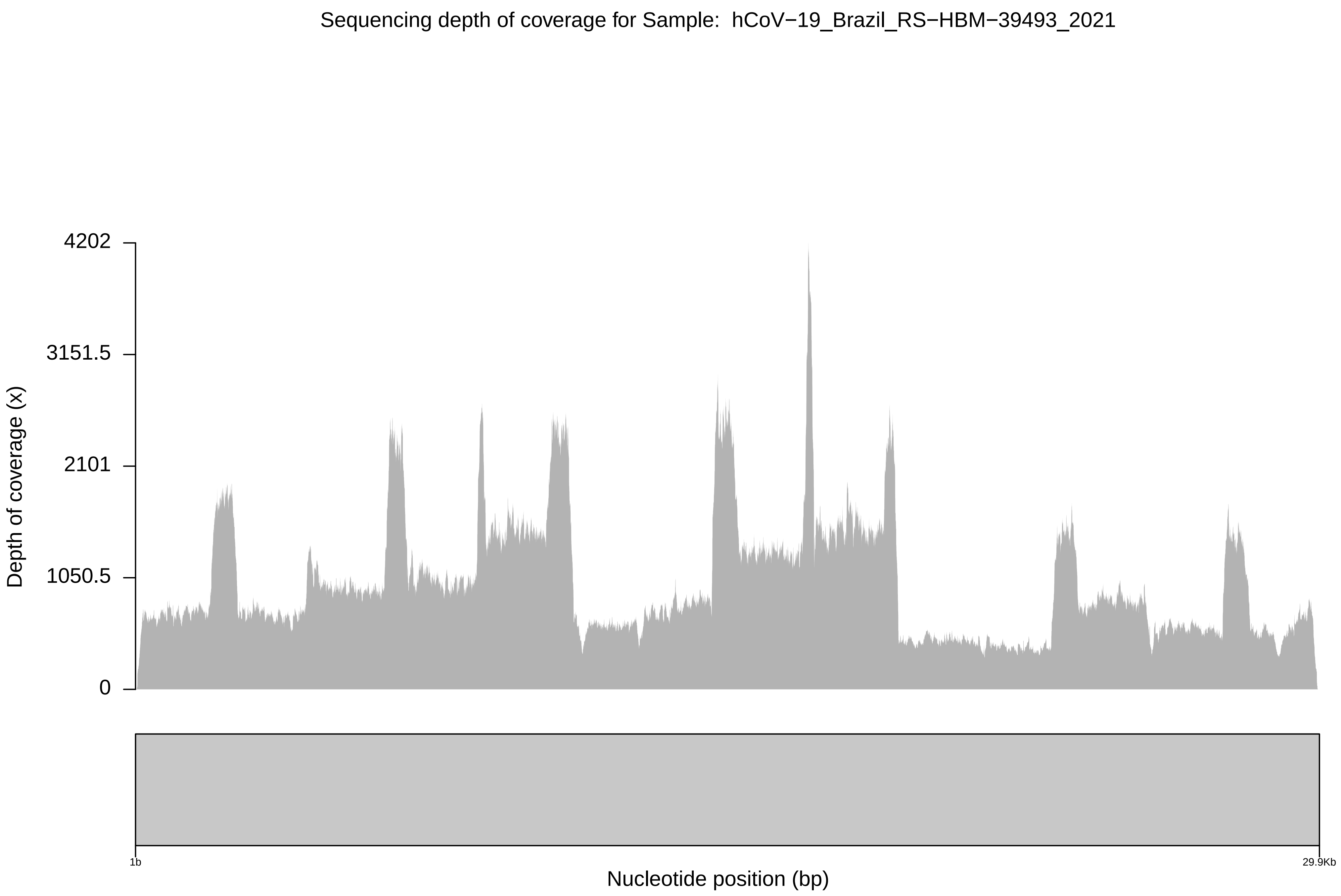

Sequencing depth of coverage for Sample: hCoV−19_Brazil_RS−HBM−39493_2021
4202
3151.5
2101
1050.5
0
Depth of coverage (x)
1b
29.9Kb
Nucleotide position (bp)

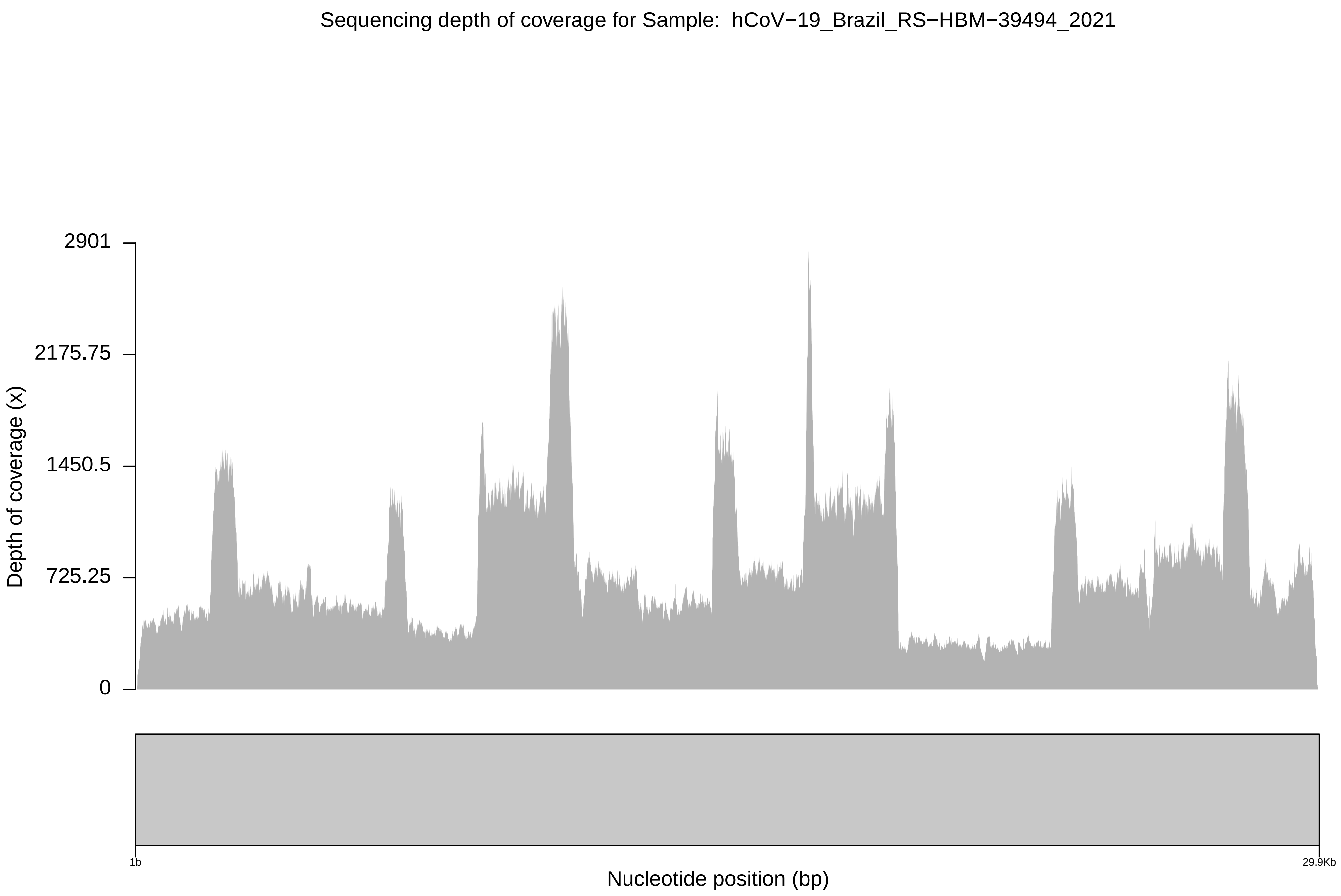

Sequencing depth of coverage for Sample: hCoV−19_Brazil_RS−HBM−39494_2021
2901
2175.75
1450.5
725.25
0
Depth of coverage (x)
1b
29.9Kb
Nucleotide position (bp)

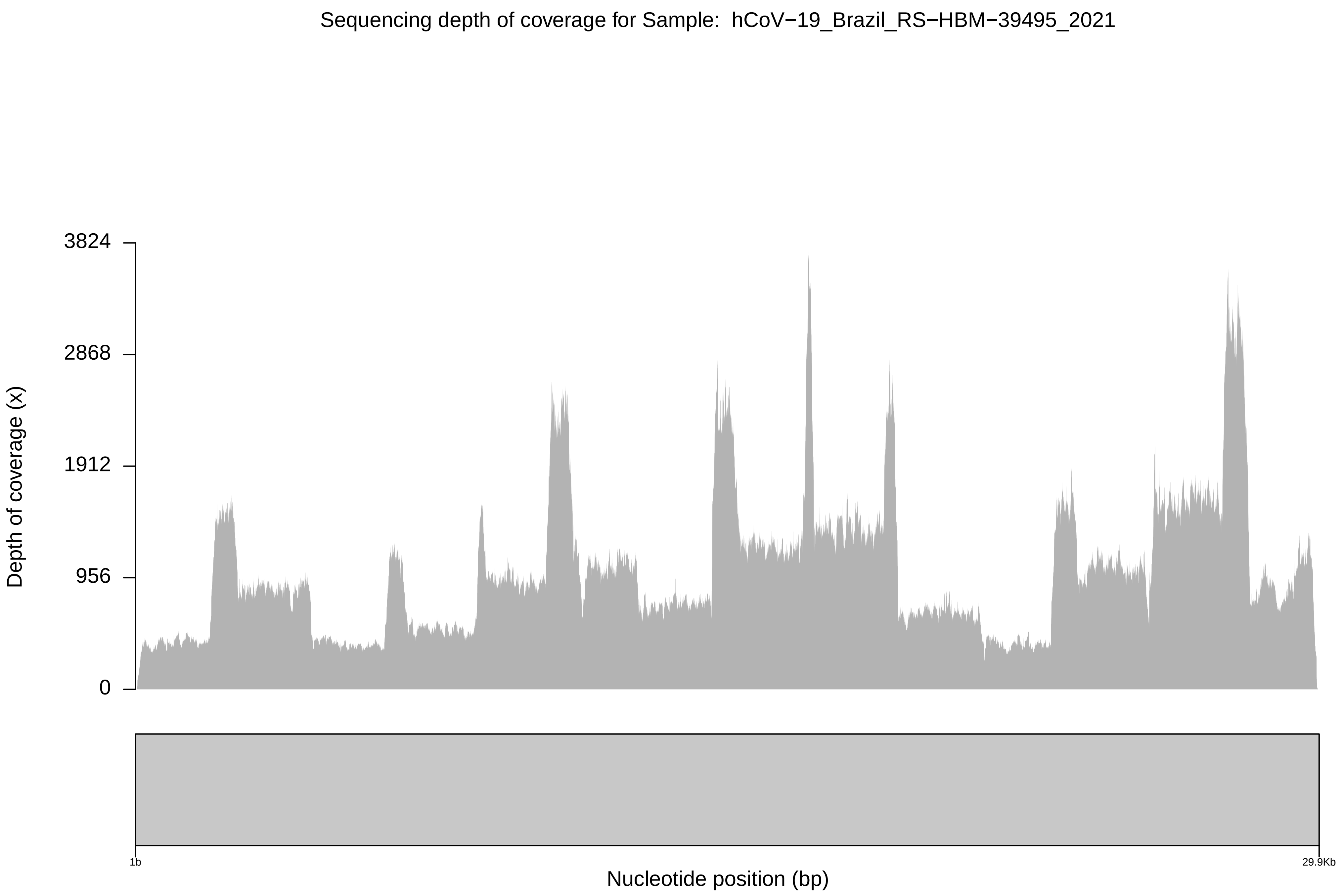
Sequencing depth of coverage for Sample: hCoV−19_Brazil_RS−HBM−39495_2021
Depth of coverage (x)
3824
2868
1912
956
0
1b
29.9Kb
Nucleotide position (bp)

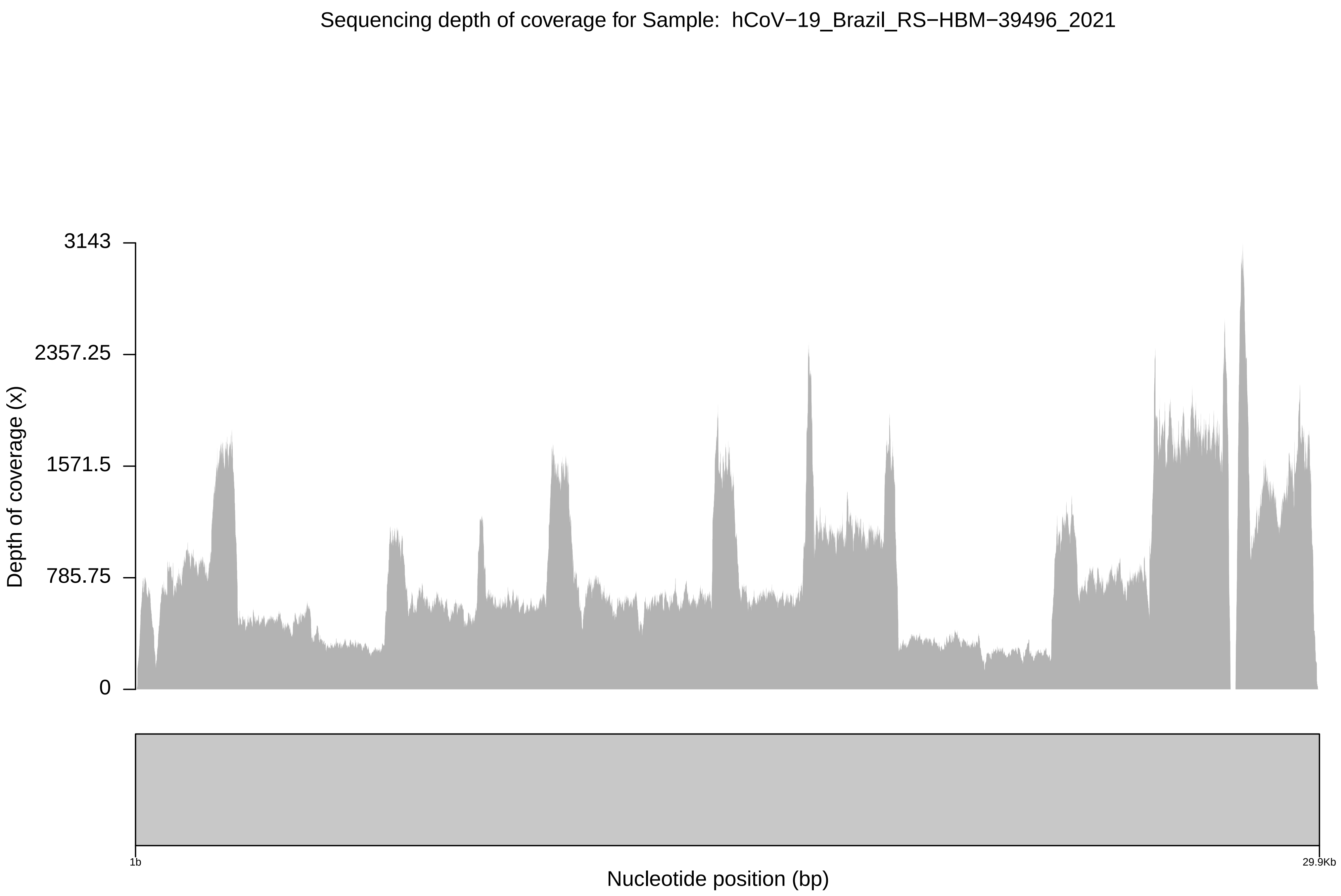

Sequencing depth of coverage for Sample: hCoV−19_Brazil_RS−HBM−39496_2021
3143
2357.25
1571.5
785.75
0
Depth of coverage (x)
1b
29.9Kb
Nucleotide position (bp)

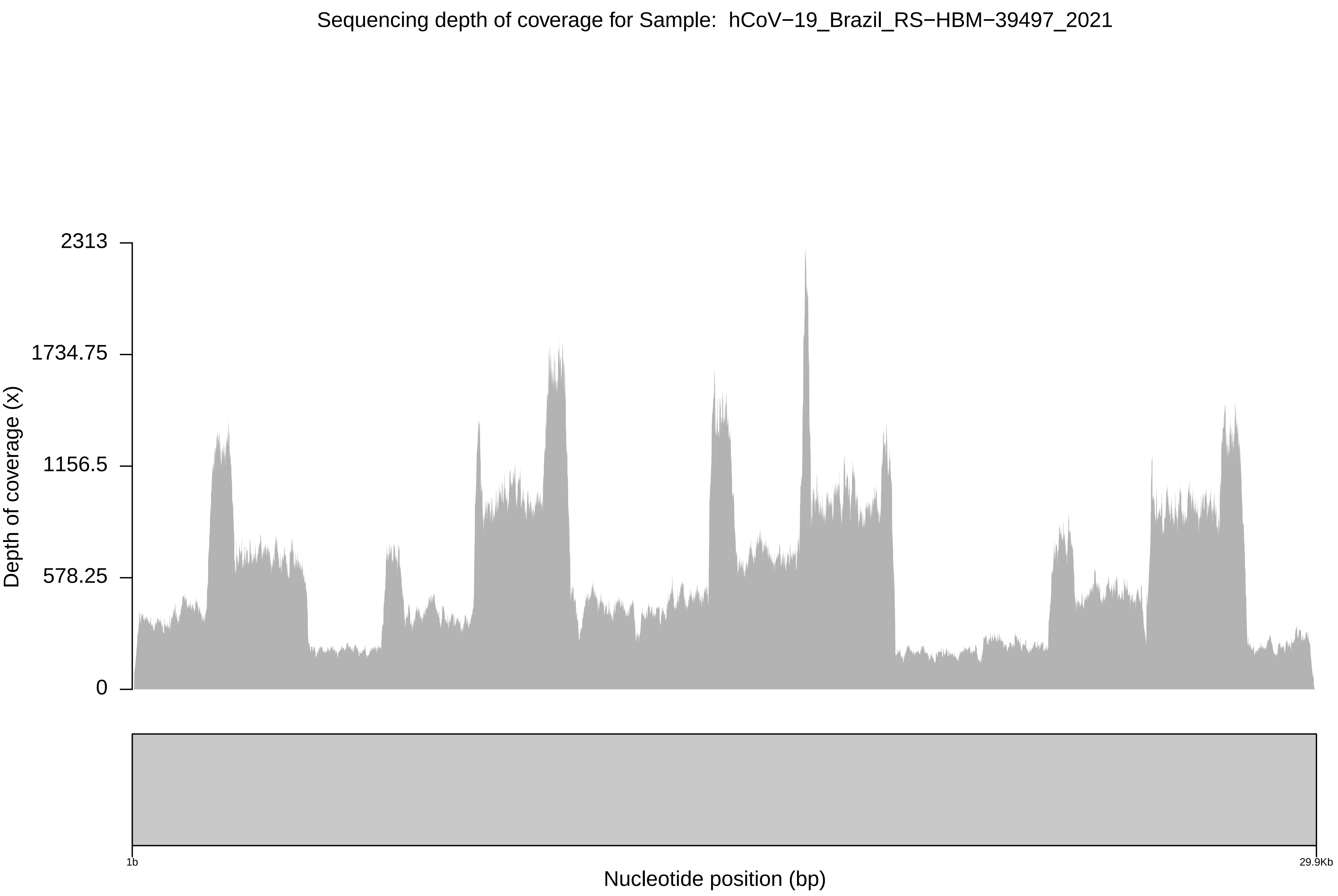

Sequencing depth of coverage for Sample: hCoV−19_Brazil_RS−HBM−39497_2021
2313
1734.75
1156.5
578.25
0
Depth of coverage (x)
1b
29.9Kb
Nucleotide position (bp)

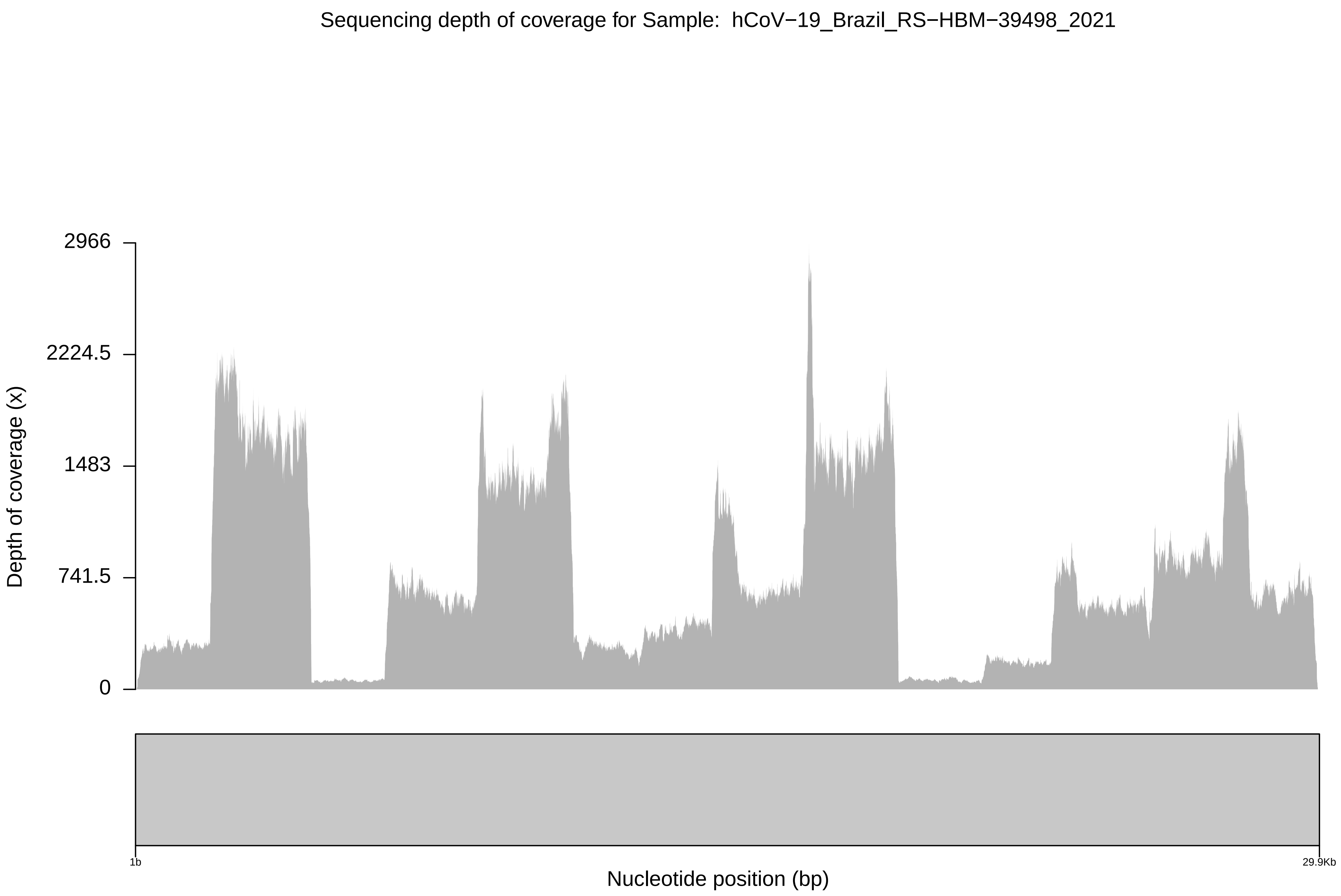
Sequencing depth of coverage for Sample:  hCoV−19_Brazil_RS−HBM−39498_2021
2966
2224.5
1483
741.5
0
Depth of coverage (x)
1b
29.9Kb
Nucleotide position (bp)

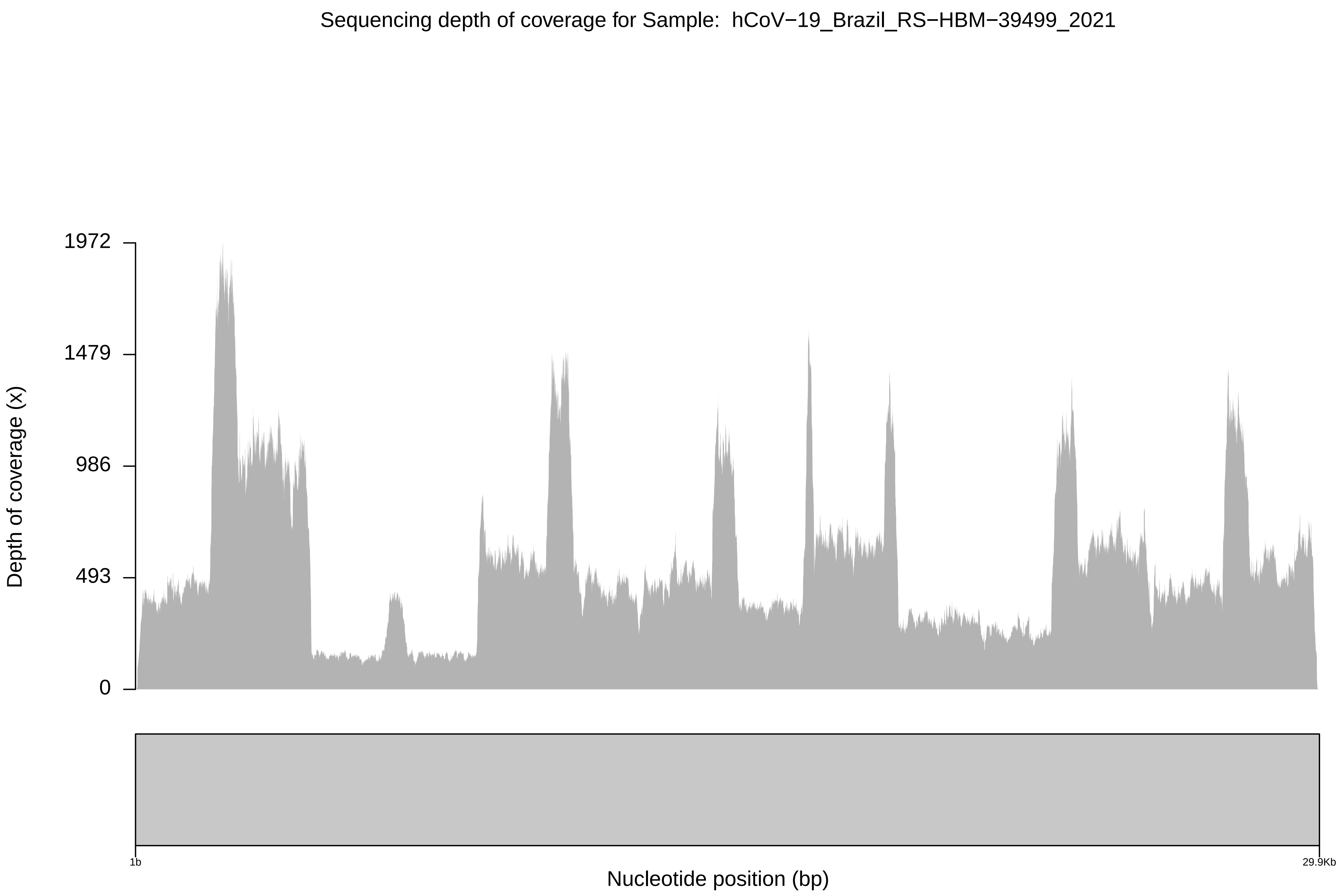
Sequencing depth of coverage for Sample: hCoV−19_Brazil_RS−HBM−39499_2021
Depth of coverage (x)
1972
1479
986
493
0
1b
29.9Kb
Nucleotide position (bp)

Sequencing depth of coverage for Sample: hCoV−19_Brazil_RS−HBM−39500_2021

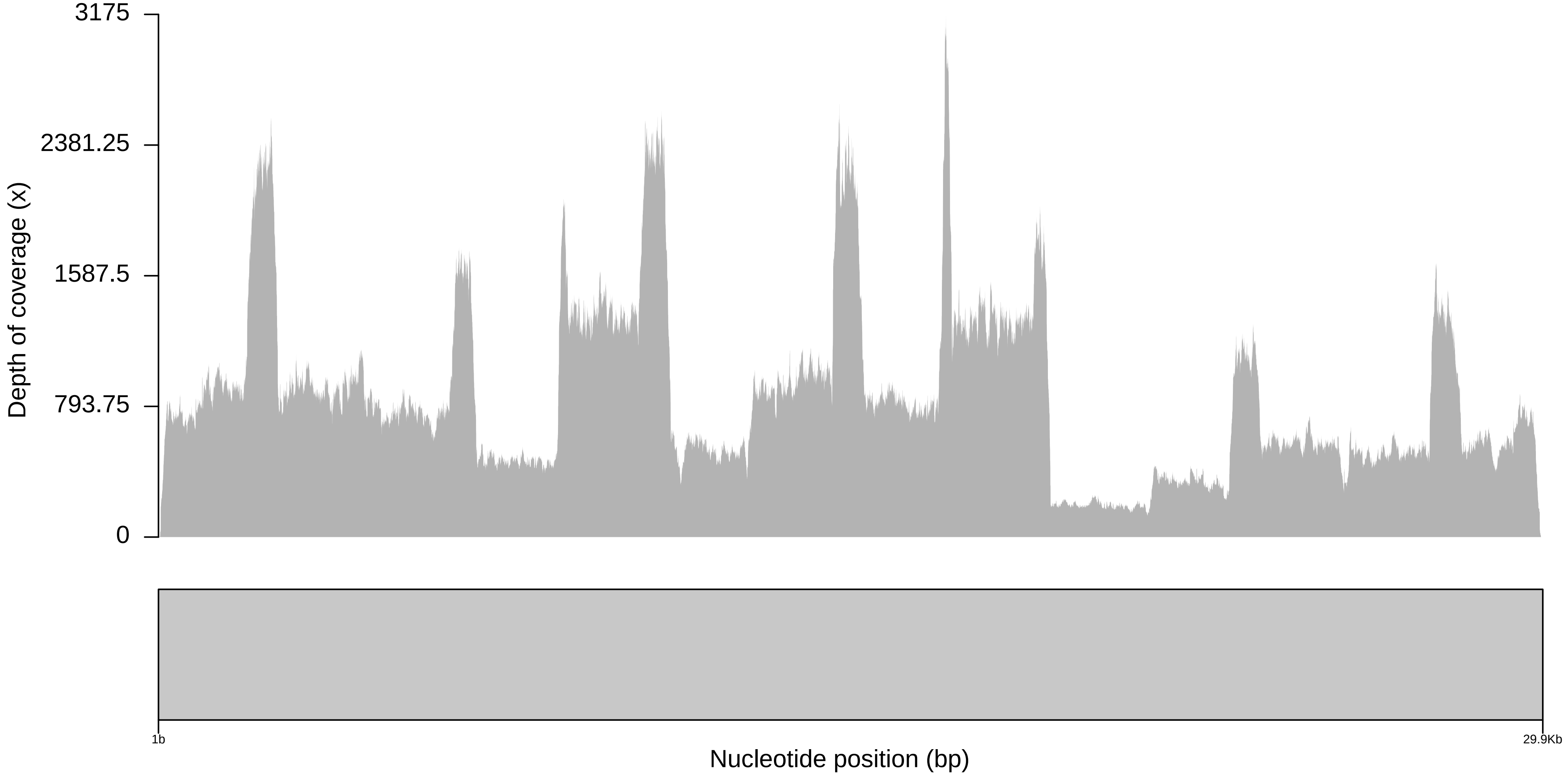

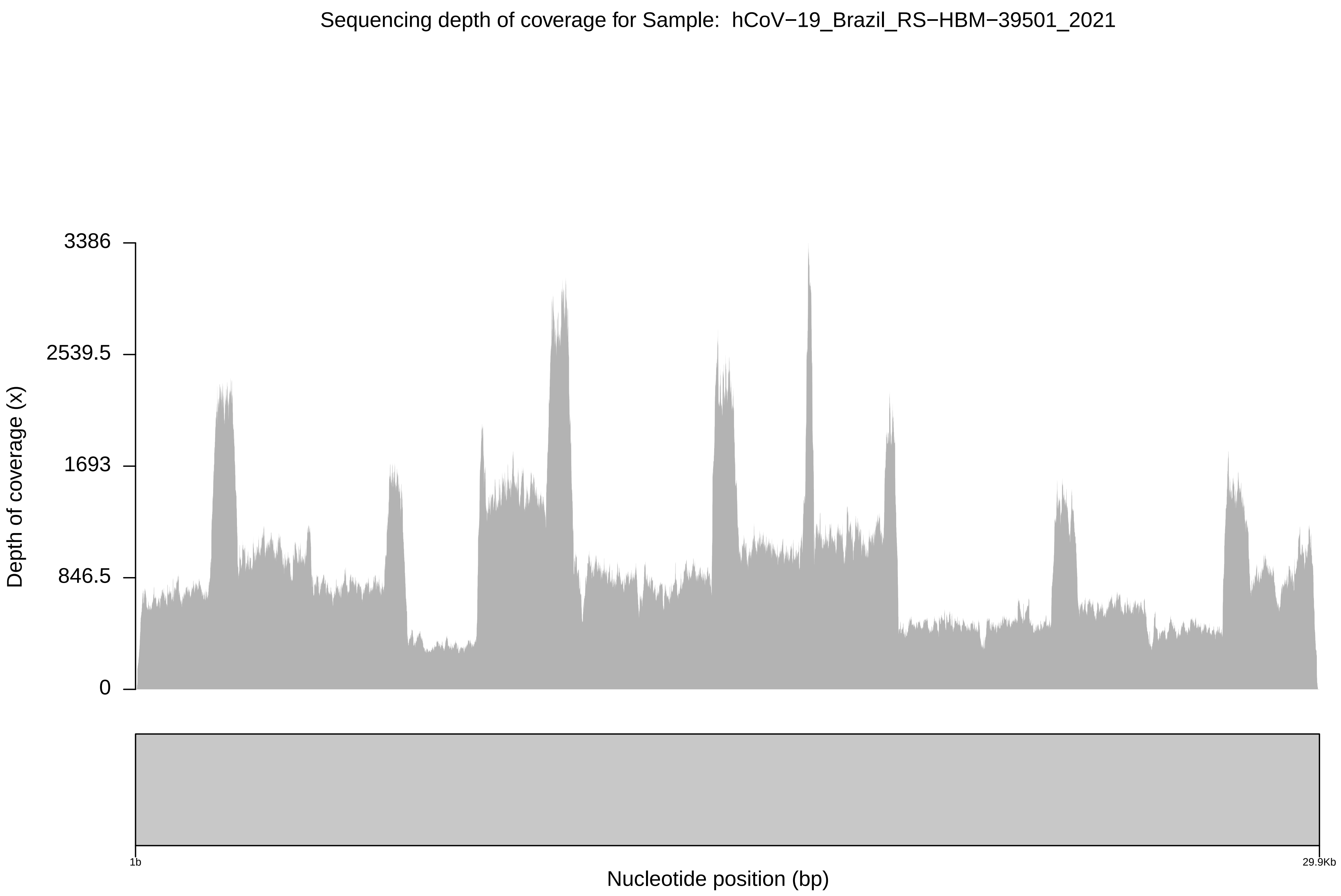
Sequencing depth of coverage for Sample: hCoV−19_Brazil_RS−HBM−39501_2021
3386
2539.5
1693
846.5
0
Depth of coverage (x)
1b
29.9Kb
Nucleotide position (bp)

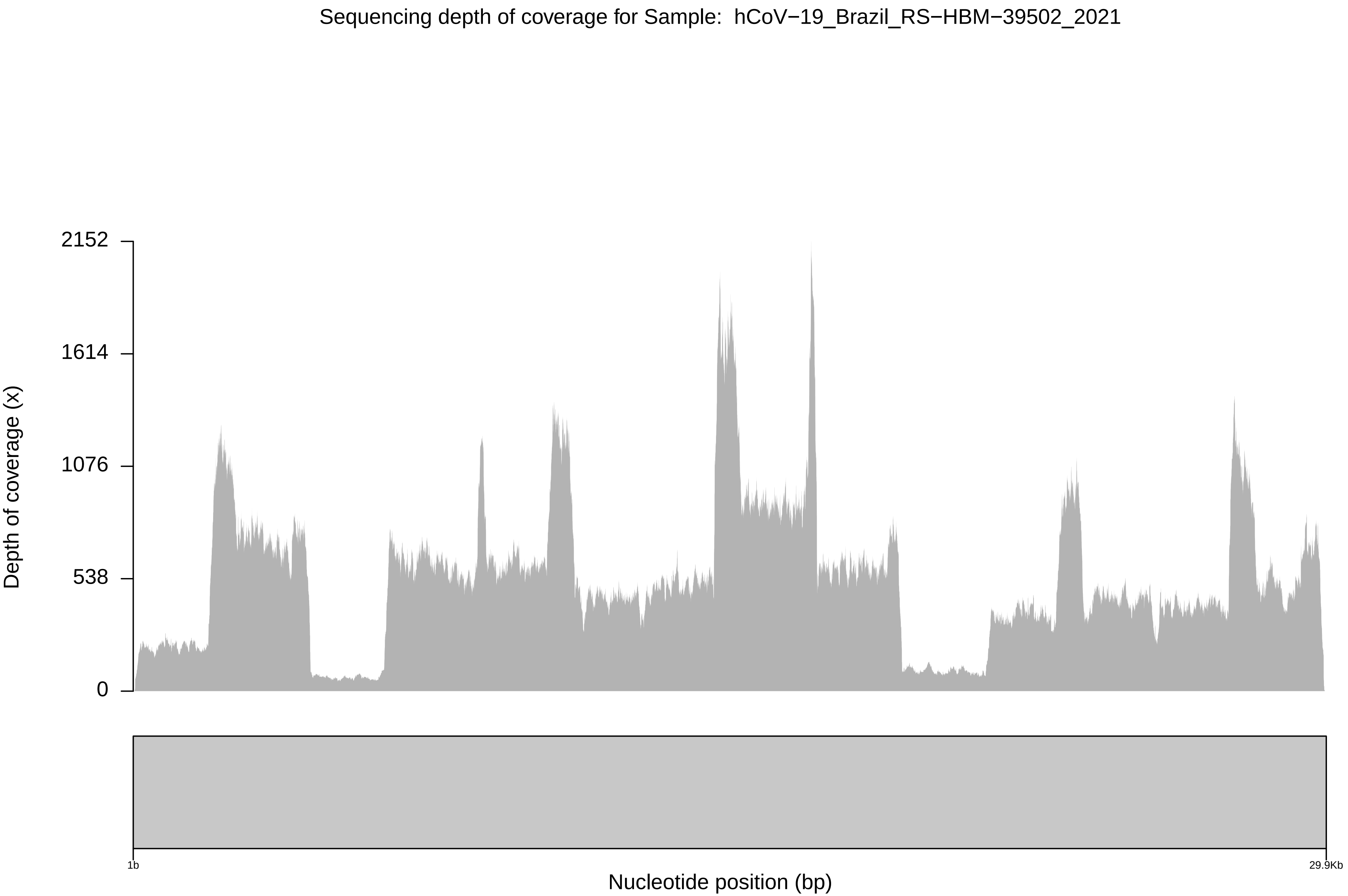

Sequencing depth of coverage for Sample: hCoV−19_Brazil_RS−HBM−39502_2021
2152
1614
1076
538
0
Depth of coverage (x)
1b
29.9Kb
Nucleotide position (bp)

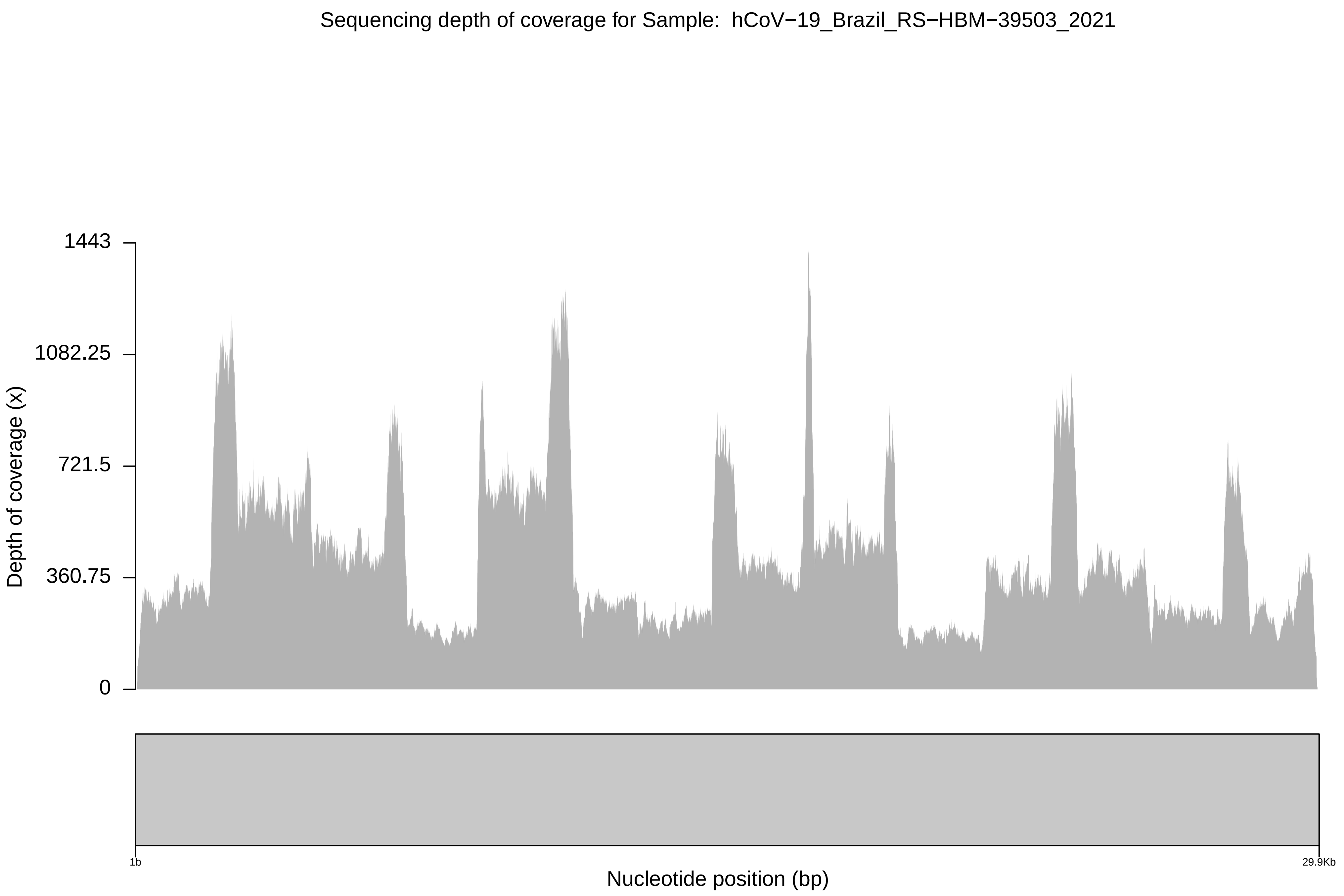

Sequencing depth of coverage for Sample: hCoV−19_Brazil_RS−HBM−39503_2021
1443
1082.25
721.5
360.75
0
Depth of coverage (x)
1b
29.9Kb
Nucleotide position (bp)

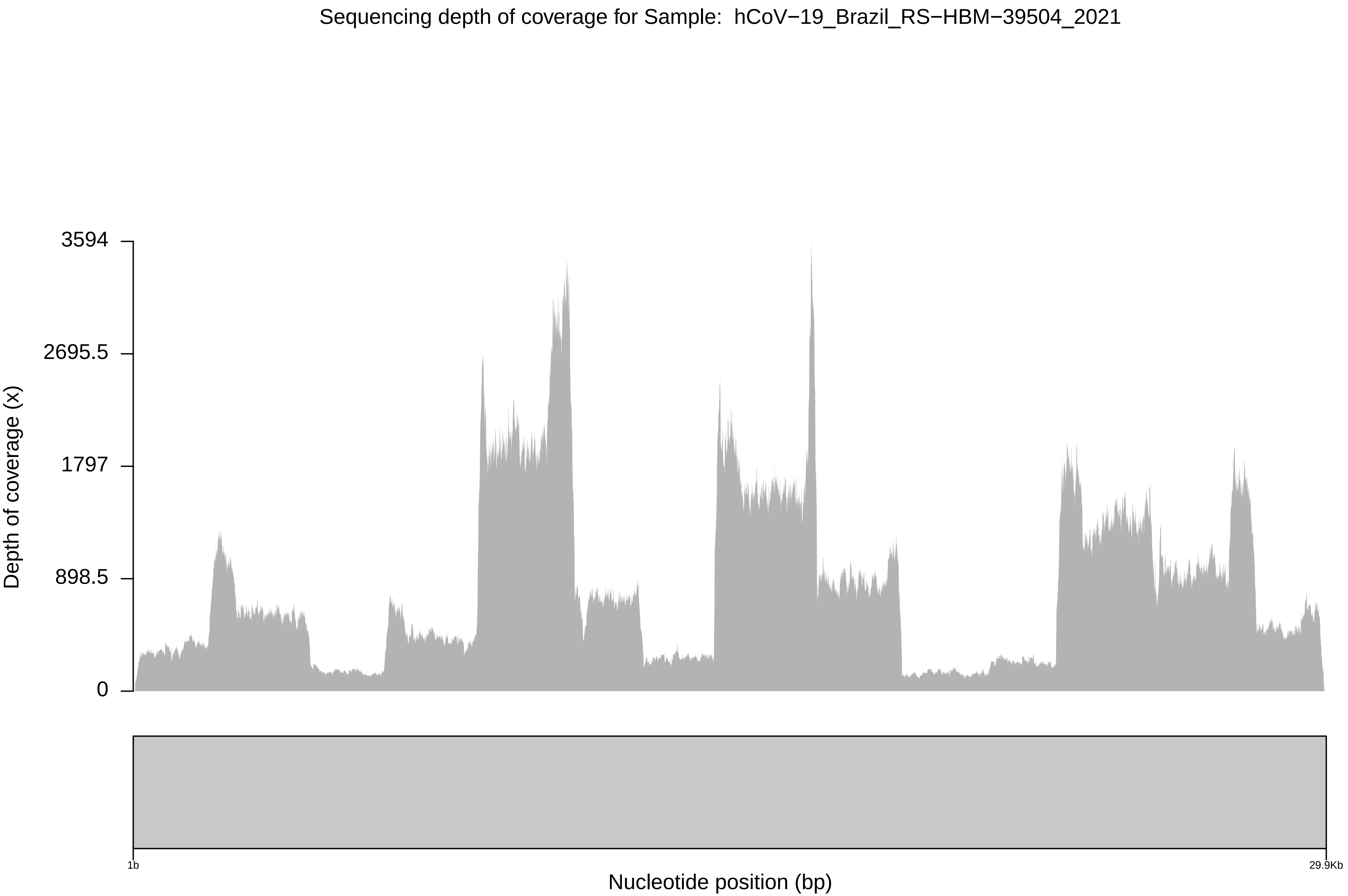
Sequencing depth of coverage for Sample: hCoV−19_Brazil_RS−HBM−39504_2021
3594
2695.5
1797
898.5
0
Depth of coverage (x)
1b
29.9Kb
Nucleotide position (bp)

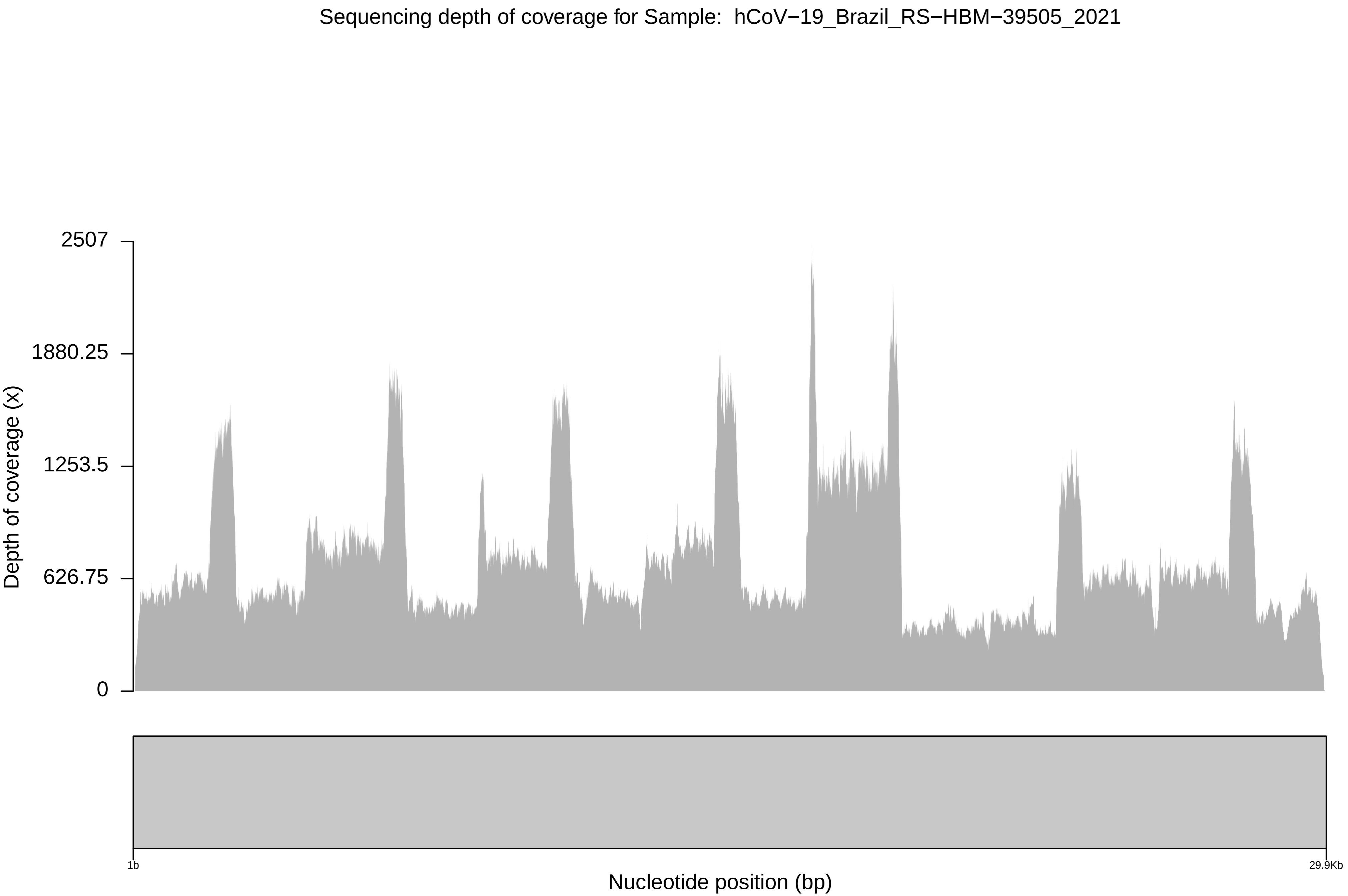

Sequencing depth of coverage for Sample: hCoV−19_Brazil_RS−HBM−39505_2021
2507
1880.25
1253.5
626.75
0
Depth of coverage (x)
1b
29.9Kb
Nucleotide position (bp)

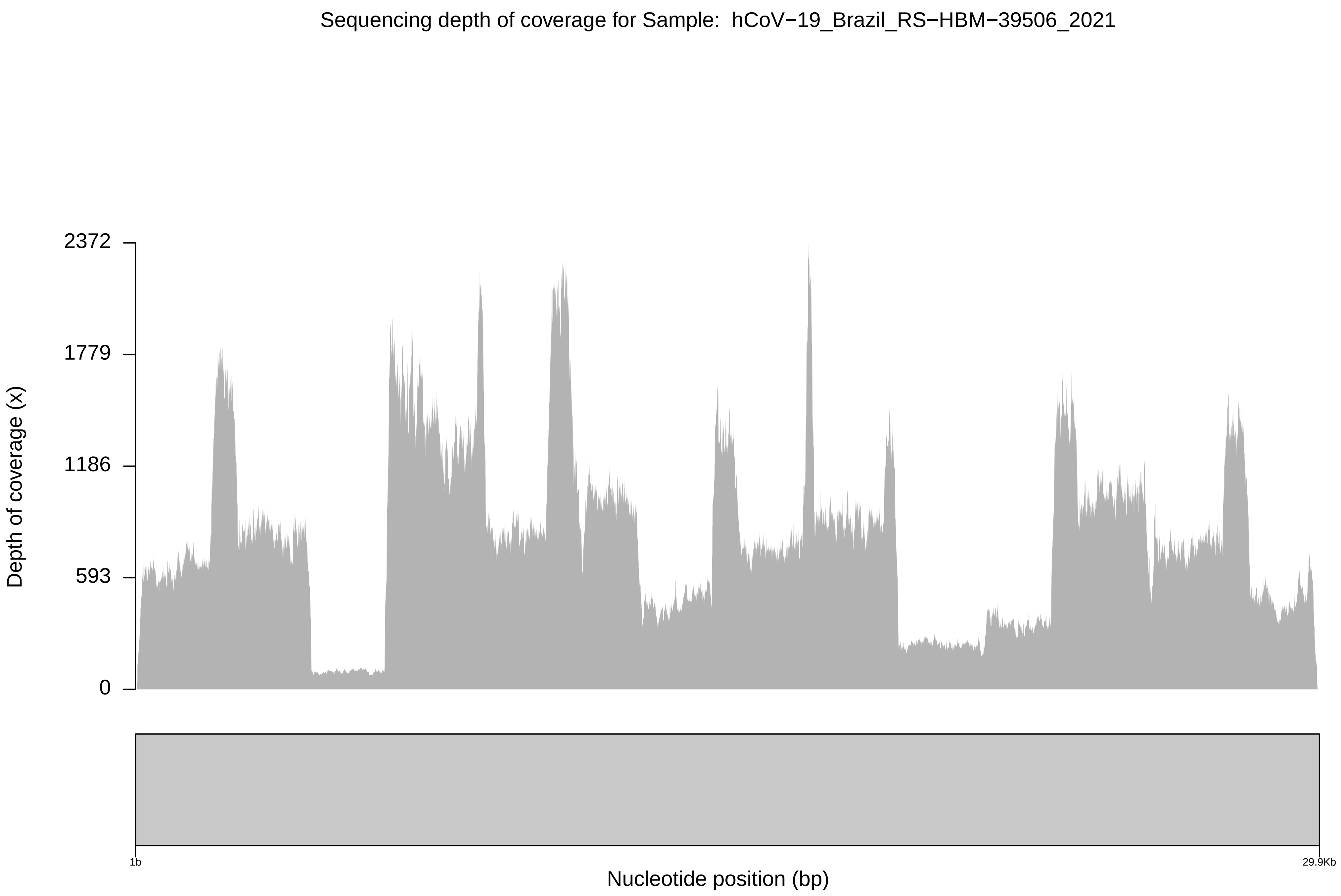

Sequencing depth of coverage for Sample: hCoV−19_Brazil_RS−HBM−39506_2021
2372
1779
1186
593
0
Depth of coverage (x)
1b
29.9Kb
Nucleotide position (bp)

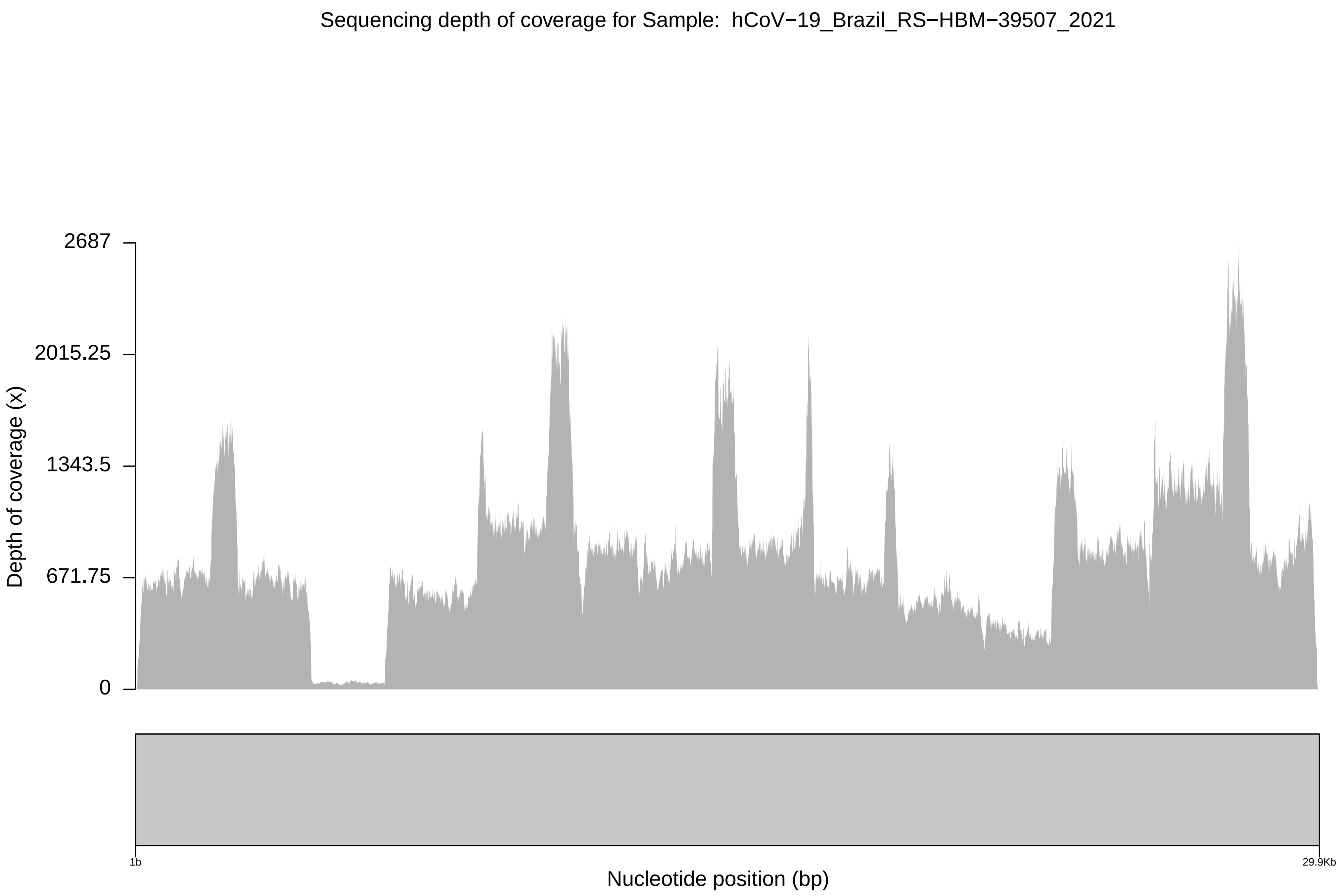
Sequencing depth of coverage for Sample: hCoV−19_Brazil_RS−HBM−39507_2021
Depth of coverage (x)
2687
2015.25
1343.5
671.75
0
1b
29.9Kb
Nucleotide position (bp)

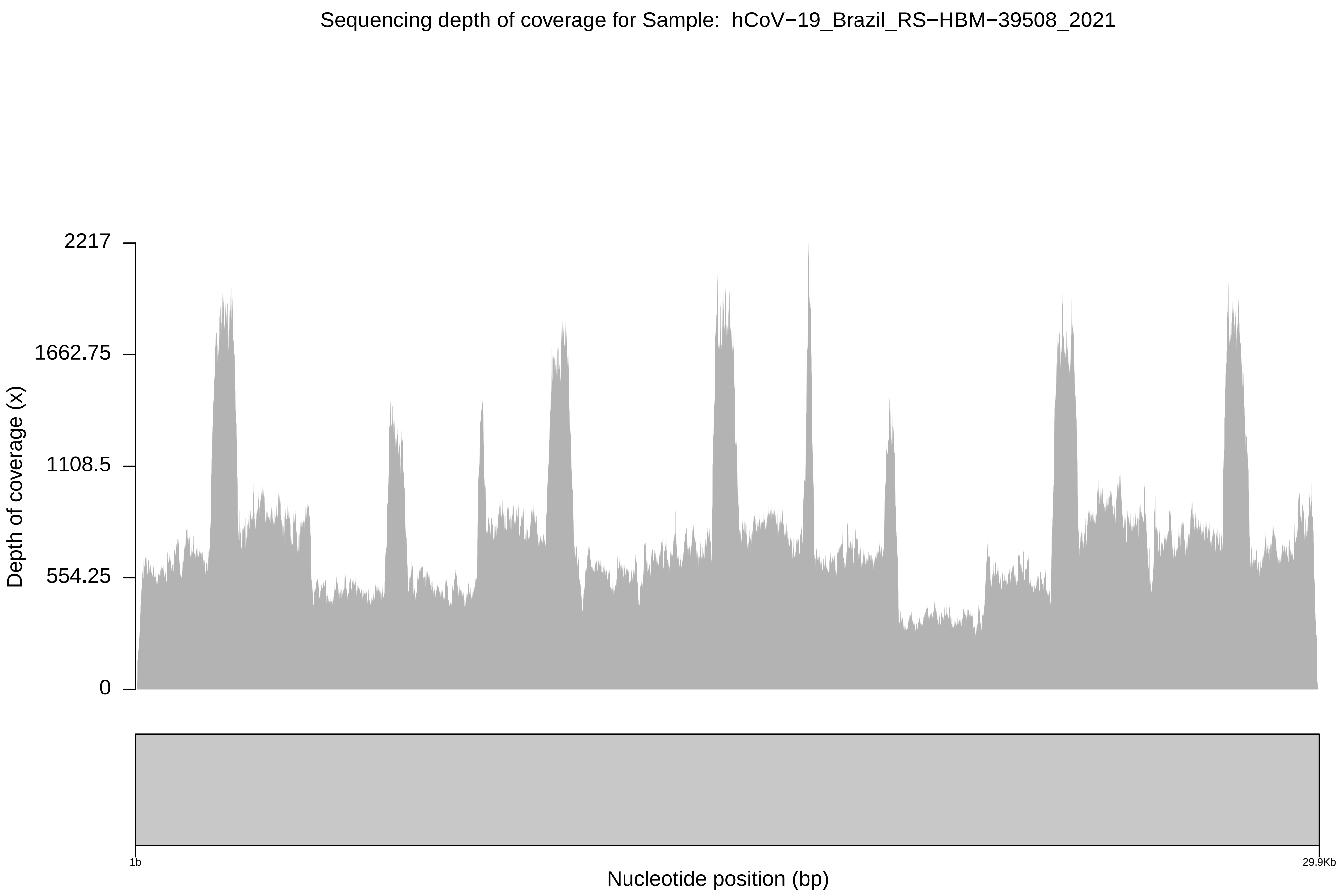

Sequencing depth of coverage for Sample: hCoV−19_Brazil_RS−HBM−39508_2021
Depth of coverage (x)
2217
1662.75
1108.5
554.25
0
1b
29.9Kb
Nucleotide position (bp)

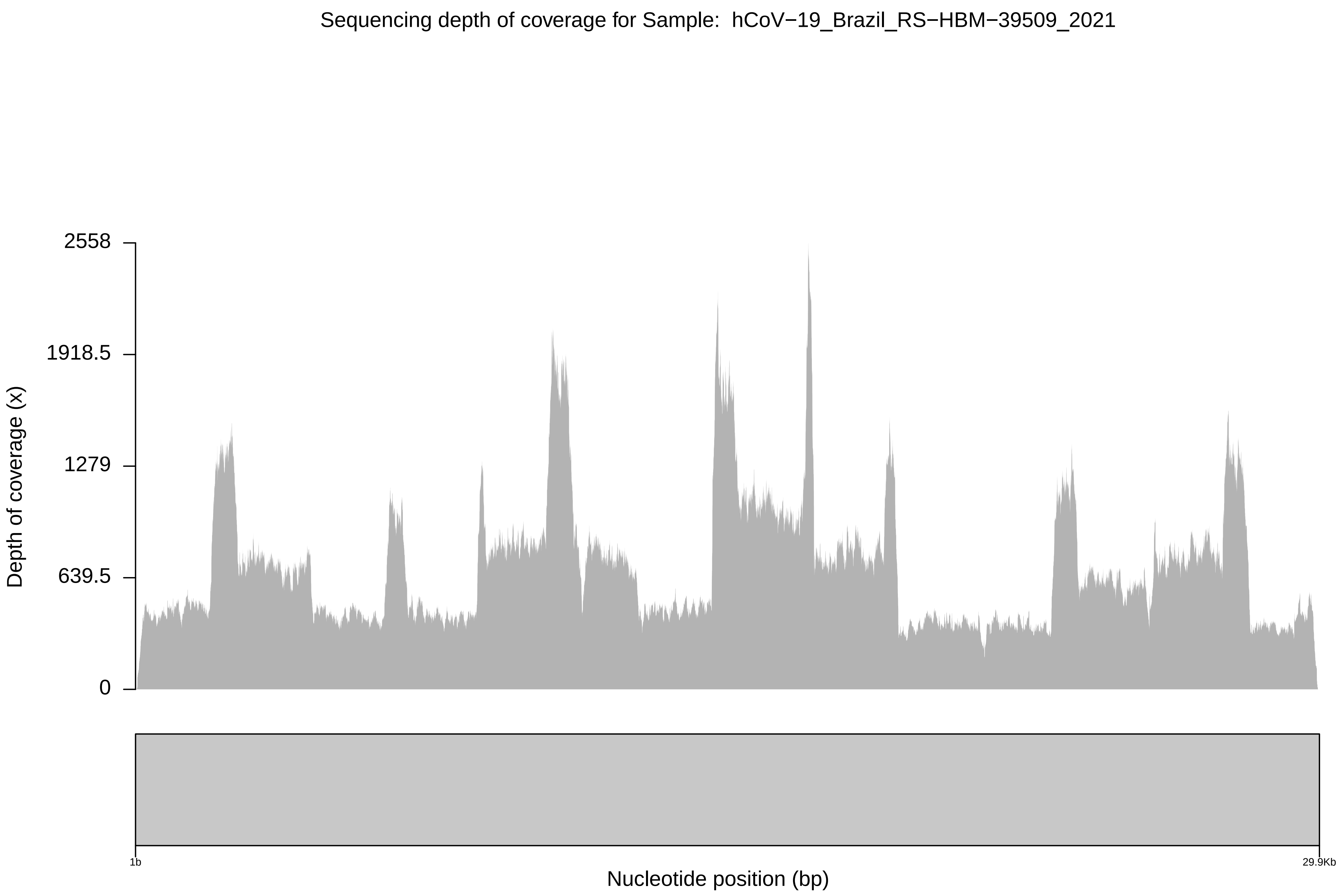
Sequencing depth of coverage for Sample: hCoV−19_Brazil_RS−HBM−39509_2021
2558
1918.5
1279
639.5
0
Depth of coverage (x)
1b
29.9Kb
Nucleotide position (bp)

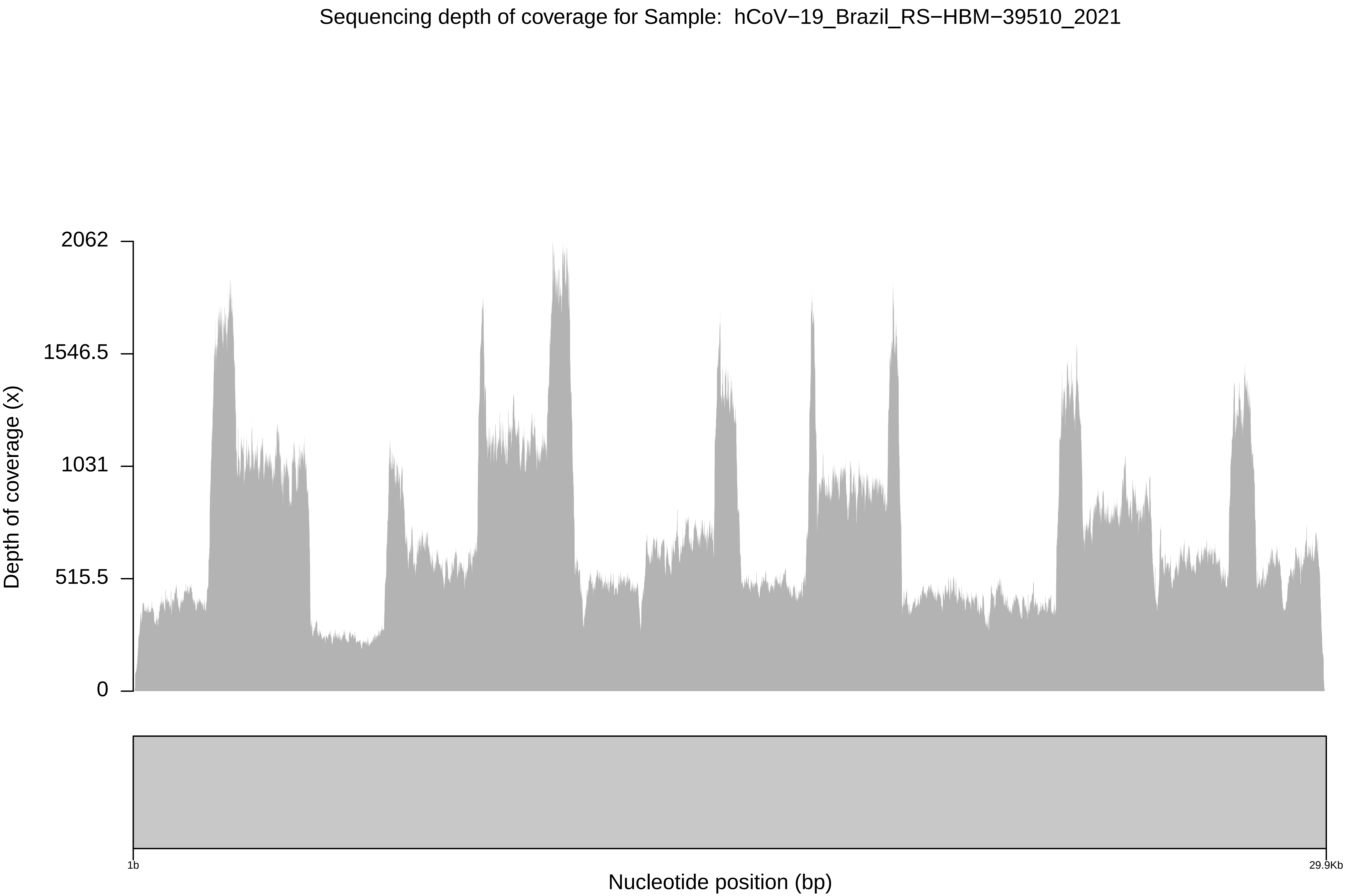

Sequencing depth of coverage for Sample: hCoV−19_Brazil_RS−HBM−39510_2021
Depth of coverage (x)
2062
1546.5
1031
515.5
0
1b
29.9Kb
Nucleotide position (bp)

Sequencing depth of coverage for Sample: hCoV−19_Brazil_RS−HBM−39511_2021

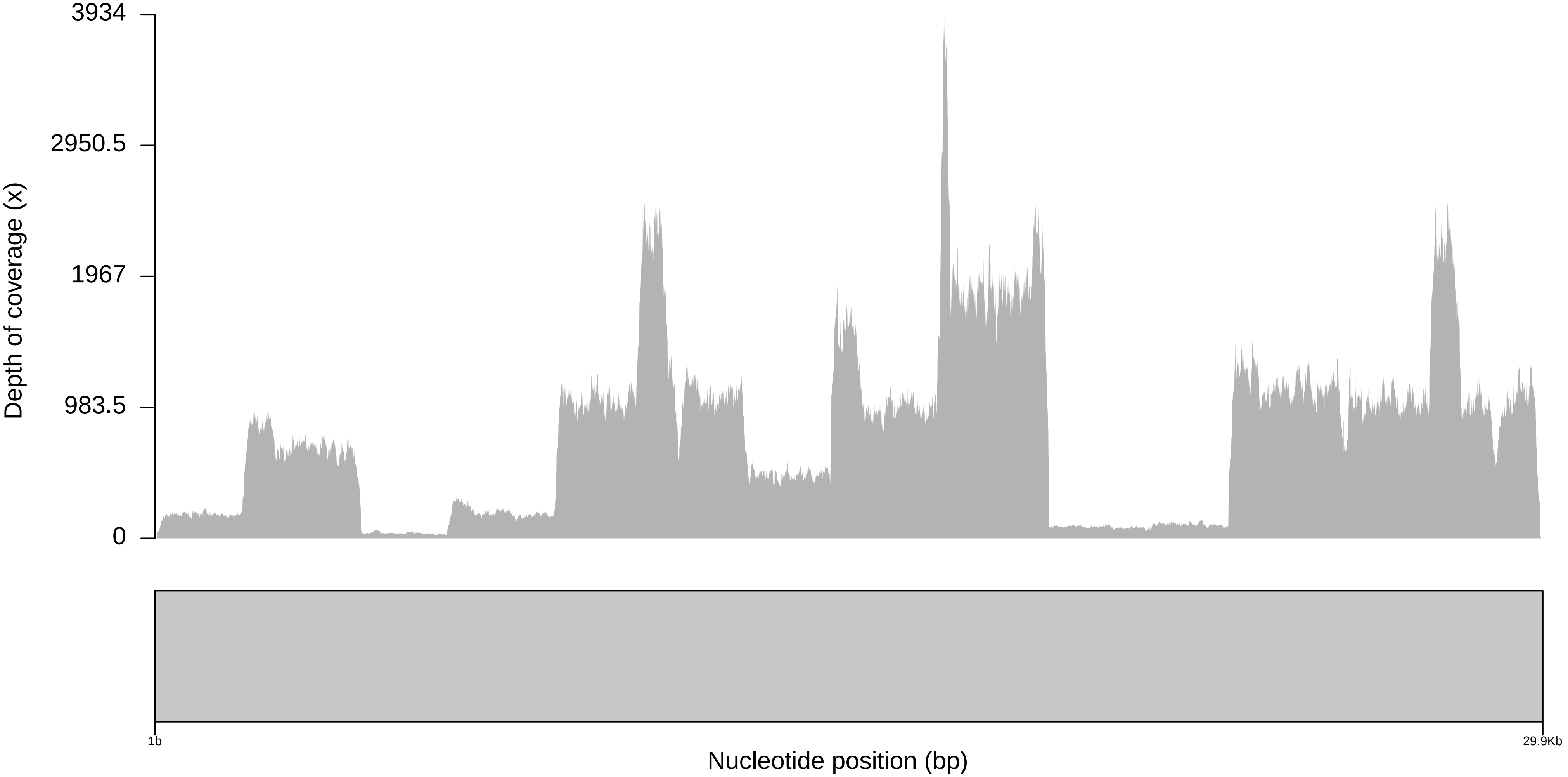

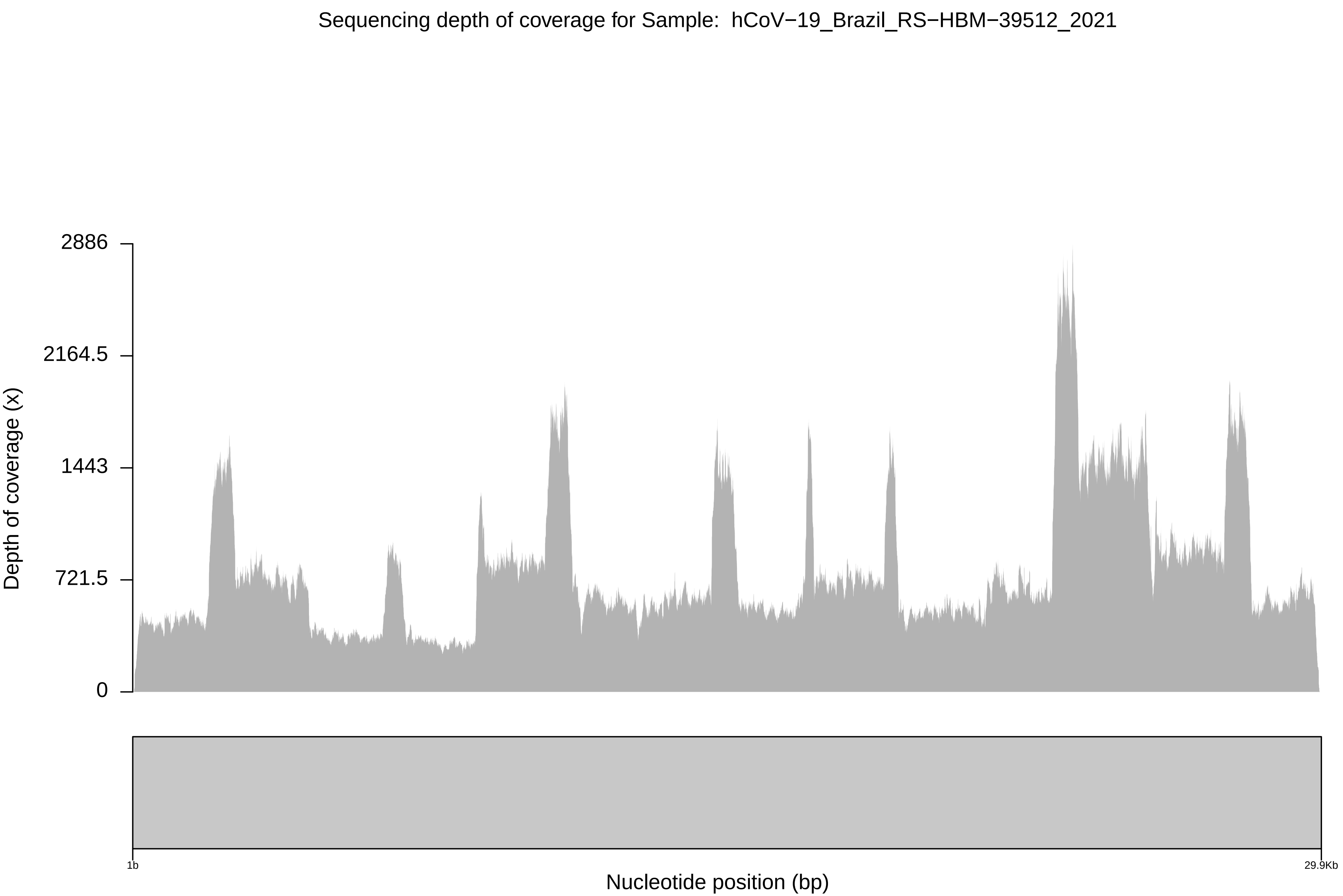

Sequencing depth of coverage for Sample: hCoV−19_Brazil_RS−HBM−39512_2021
2886
2164.5
1443
721.5
0
Depth of coverage (x)
1b
29.9Kb
Nucleotide position (bp)

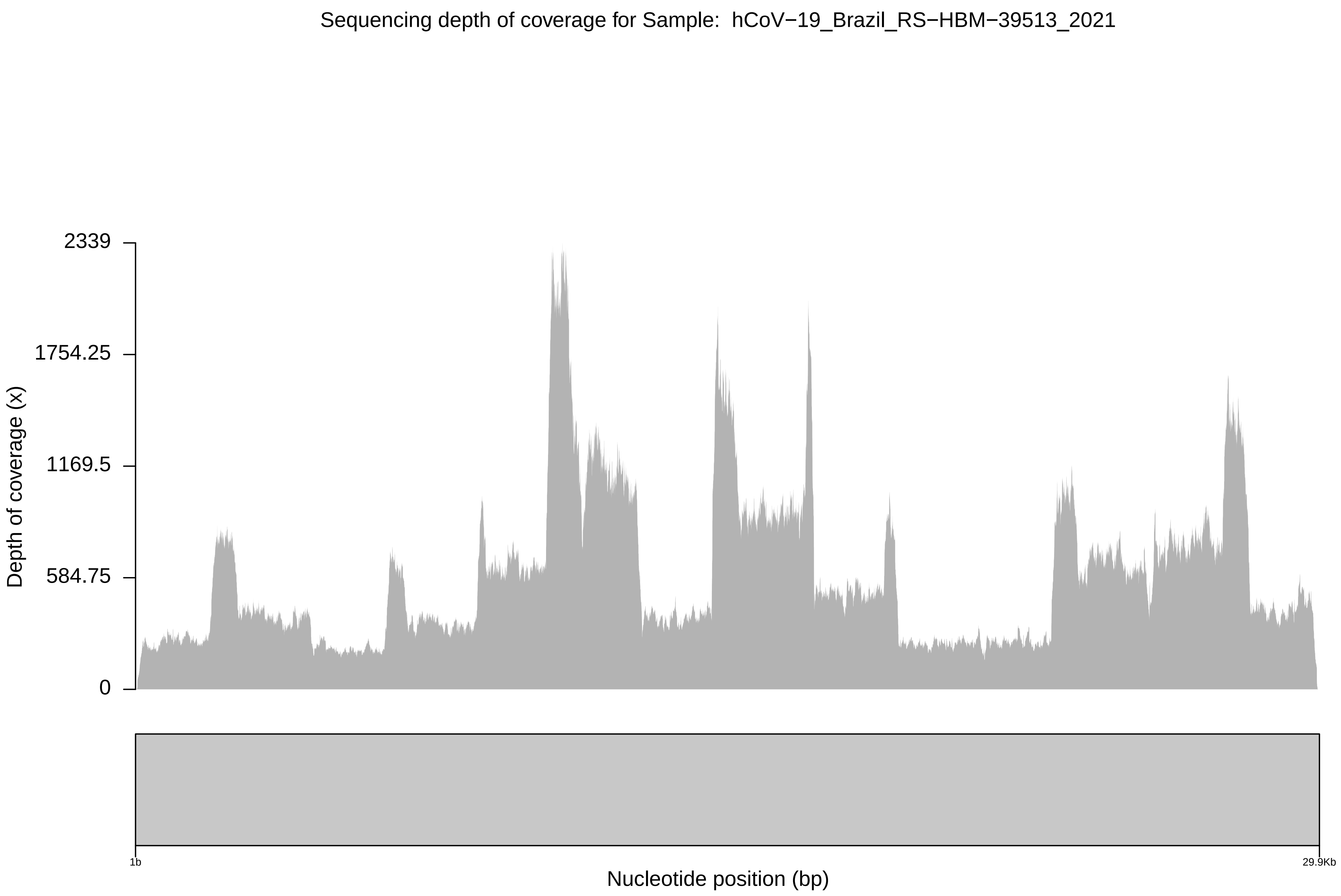
Sequencing depth of coverage for Sample: hCoV−19_Brazil_RS−HBM−39513_2021
Depth of coverage (x)
2339
1754.25
1169.5
584.75
0
1b
29.9Kb
Nucleotide position (bp)

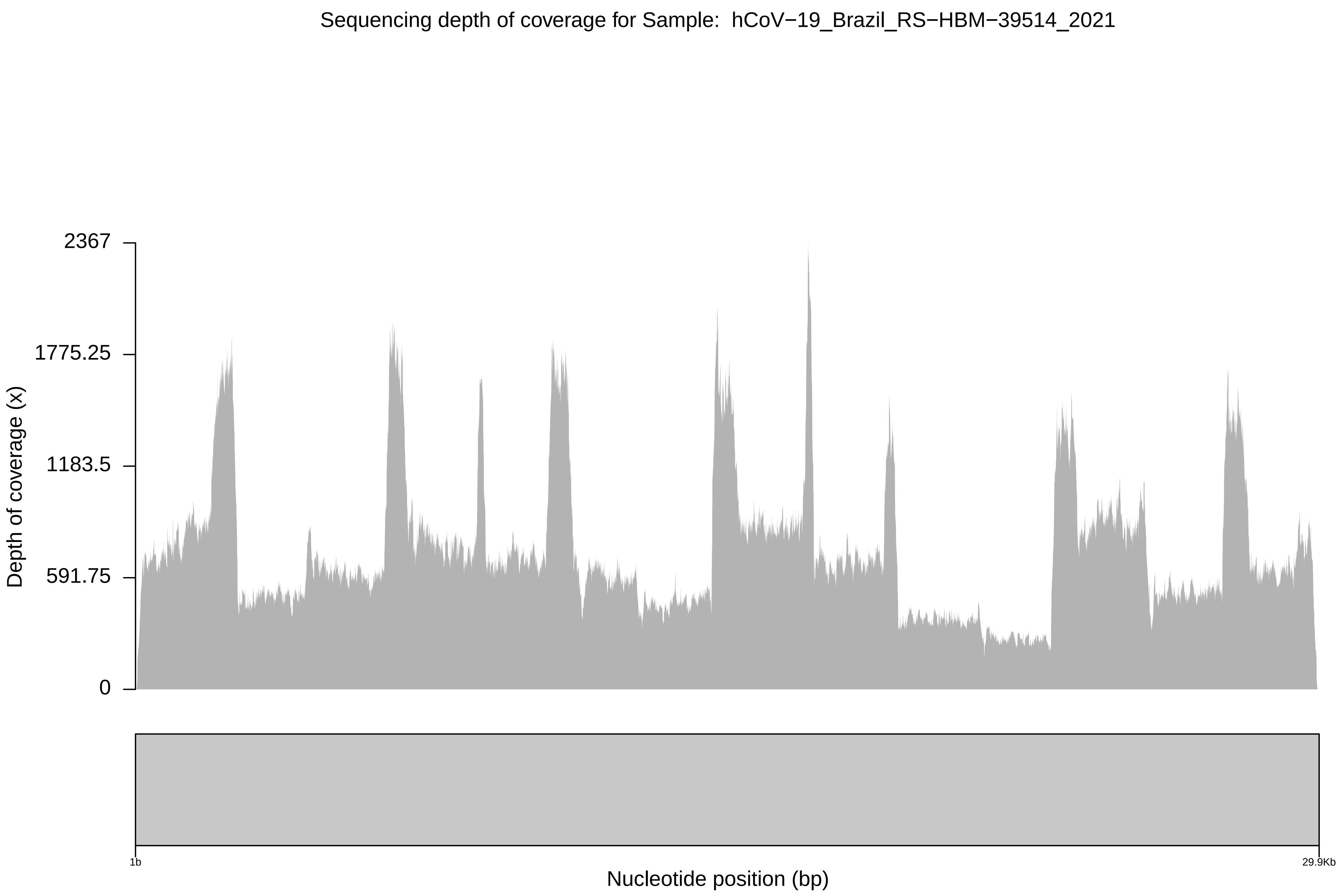

Sequencing depth of coverage for Sample:  hCoV−19_Brazil_RS−HBM−39514_2021
2367
1775.25
1183.5
591.75
0
Depth of coverage (x)
1b
29.9Kb
Nucleotide position (bp)

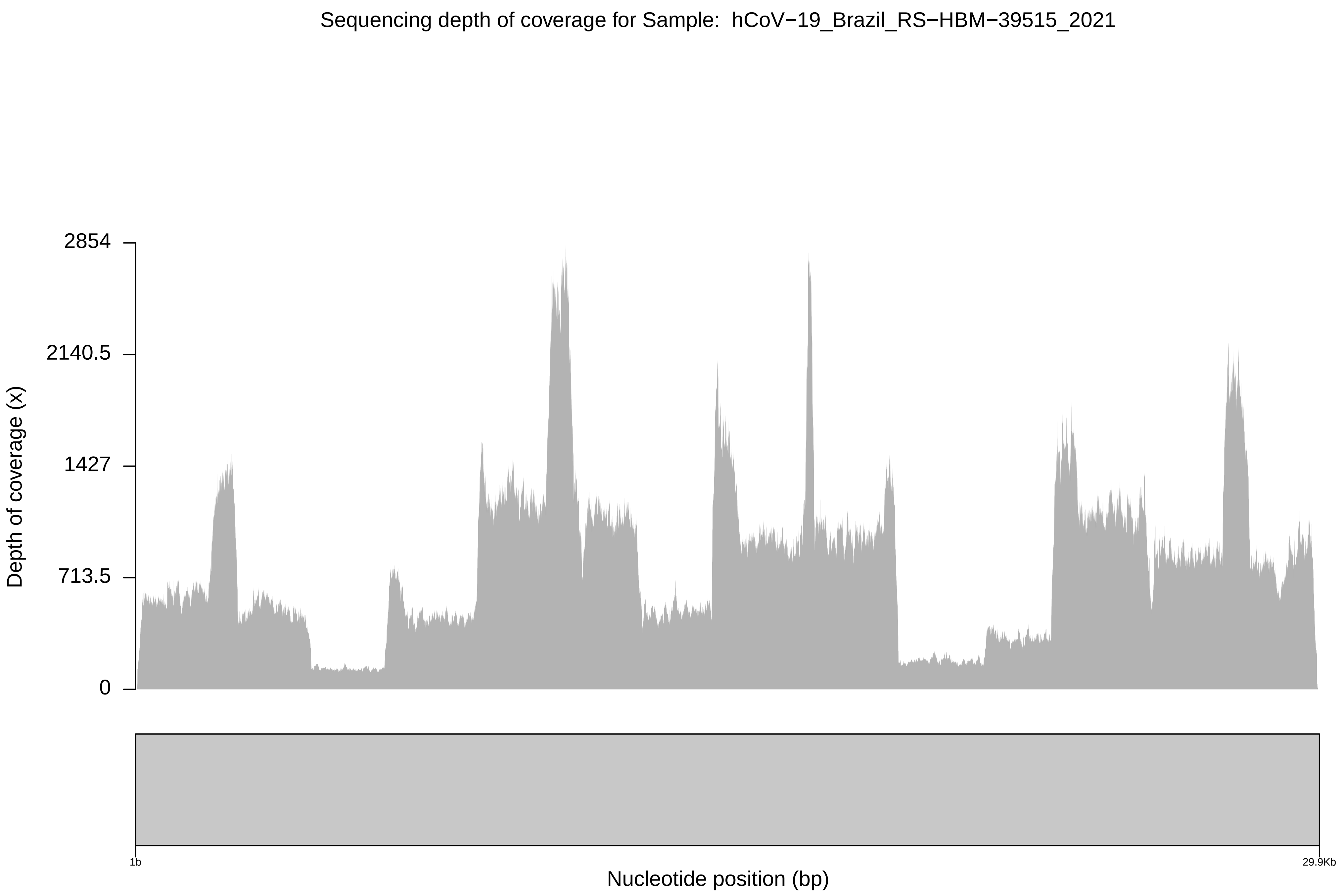

Sequencing depth of coverage for Sample: hCoV−19_Brazil_RS−HBM−39515_2021
Depth of coverage (x)
2854
2140.5
1427
713.5
0
1b
29.9Kb
Nucleotide position (bp)

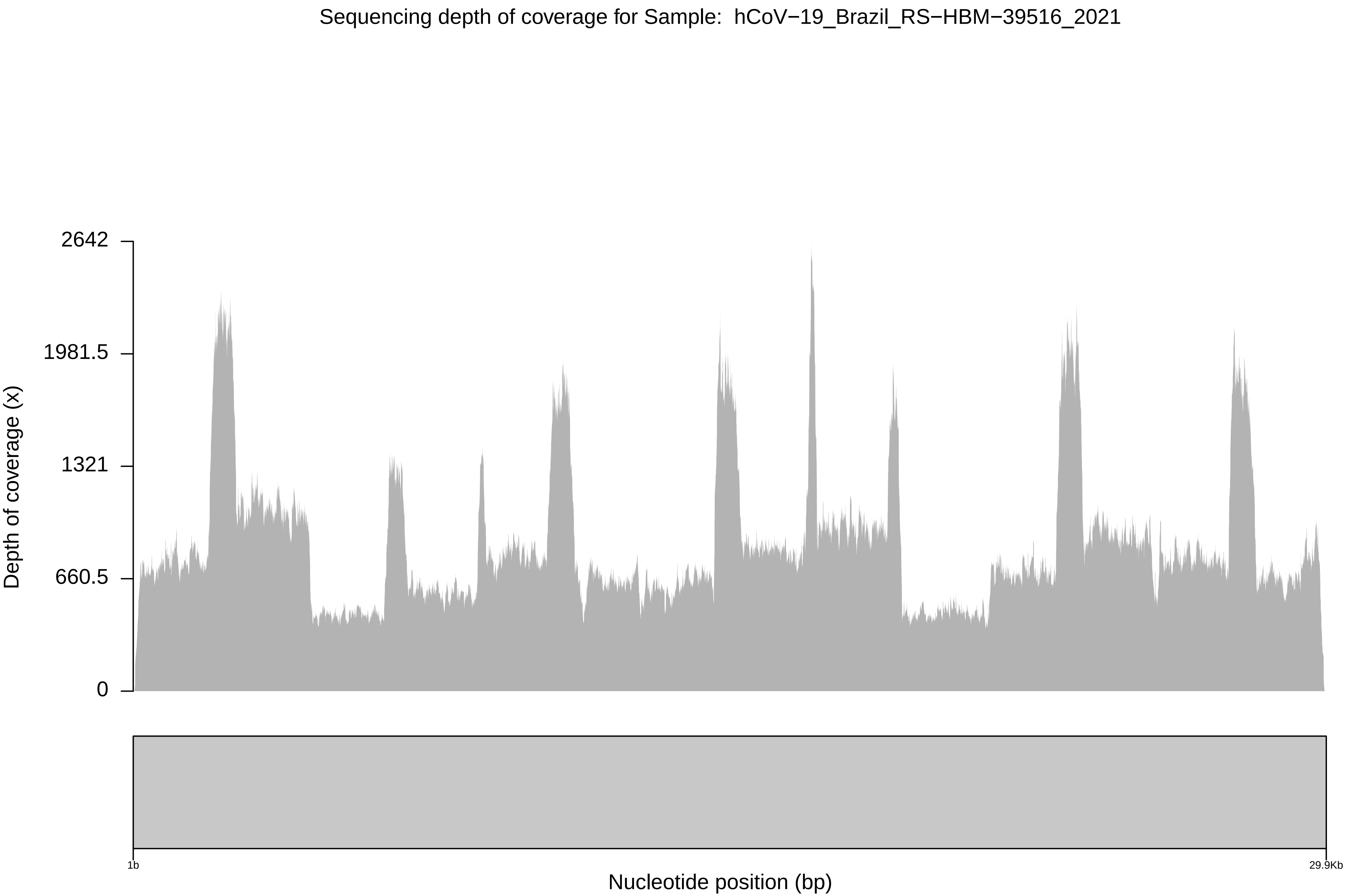

Sequencing depth of coverage for Sample: hCoV−19_Brazil_RS−HBM−39516_2021
2642
1981.5
1321
660.5
0
Depth of coverage (x)
1b
29.9Kb
Nucleotide position (bp)

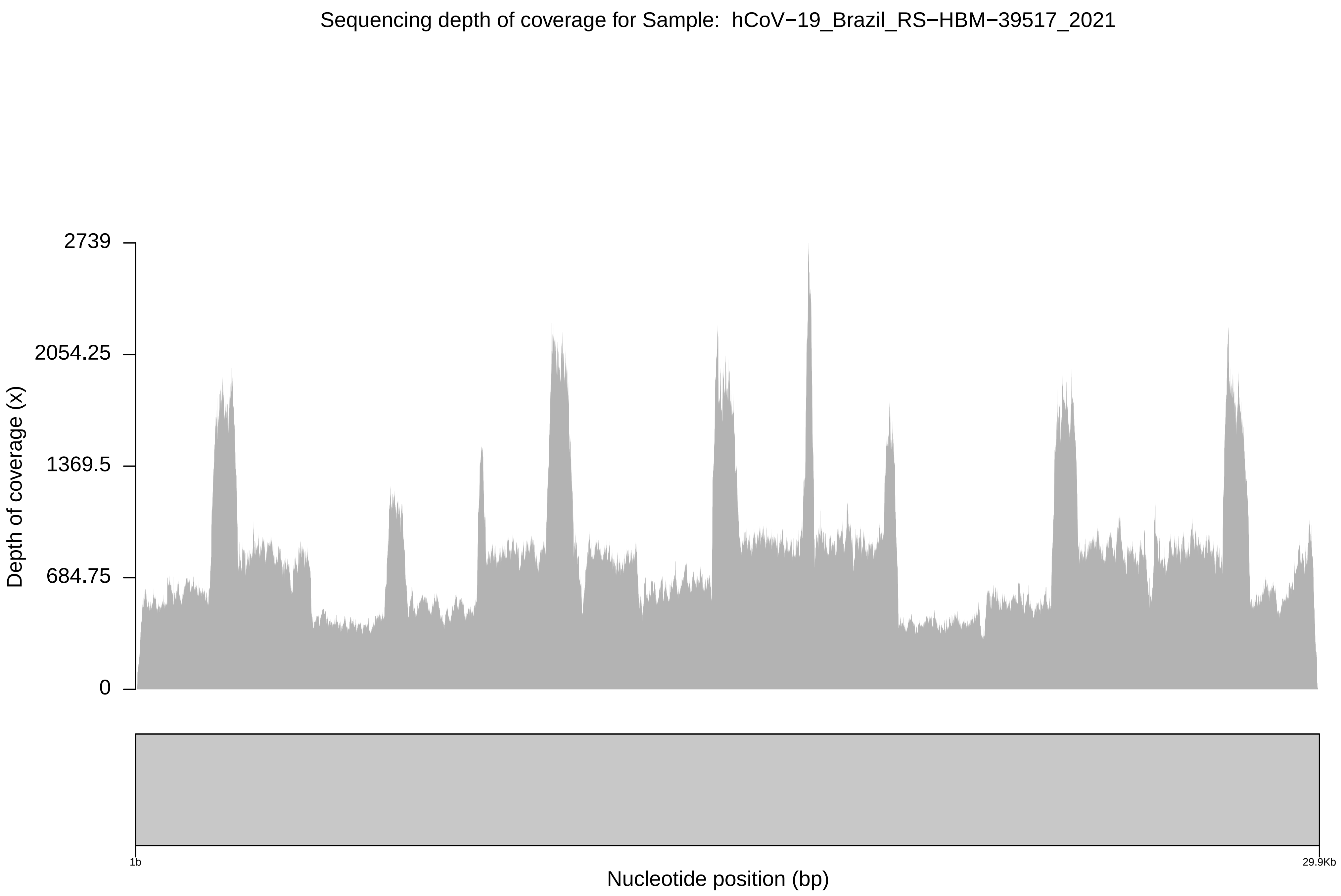
Sequencing depth of coverage for Sample: hCoV−19_Brazil_RS−HBM−39517_2021
2739
2054.25
1369.5
684.75
0
Depth of coverage (x)
1b
29.9Kb
Nucleotide position (bp)

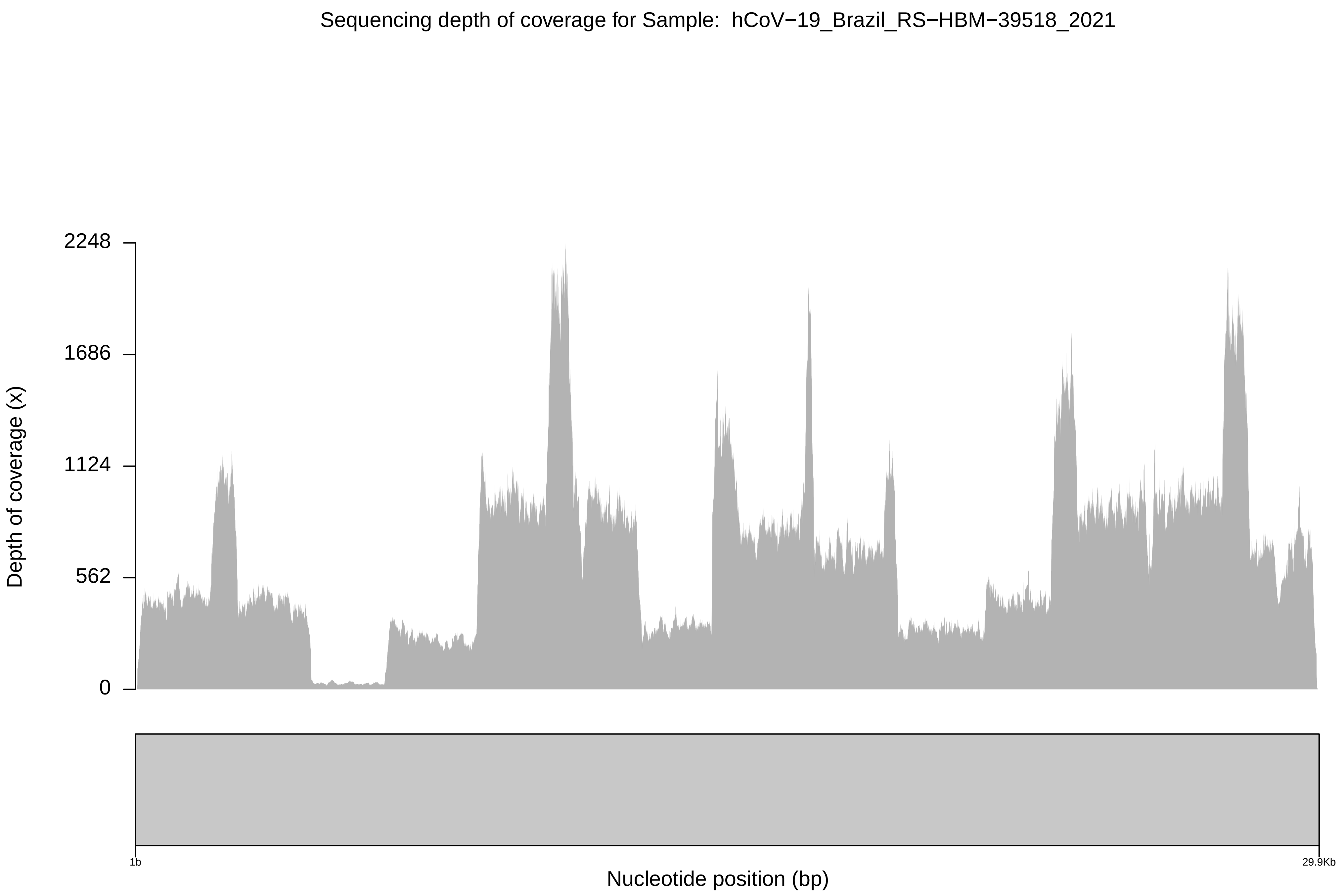
Sequencing depth of coverage for Sample: hCoV−19_Brazil_RS−HBM−39518_2021
Depth of coverage (x)
2248
1686
1124
562
0
1b
29.9Kb
Nucleotide position (bp)

Sequencing depth of coverage for Sample: hCoV−19_Brazil_RS−HBM−39519_2021

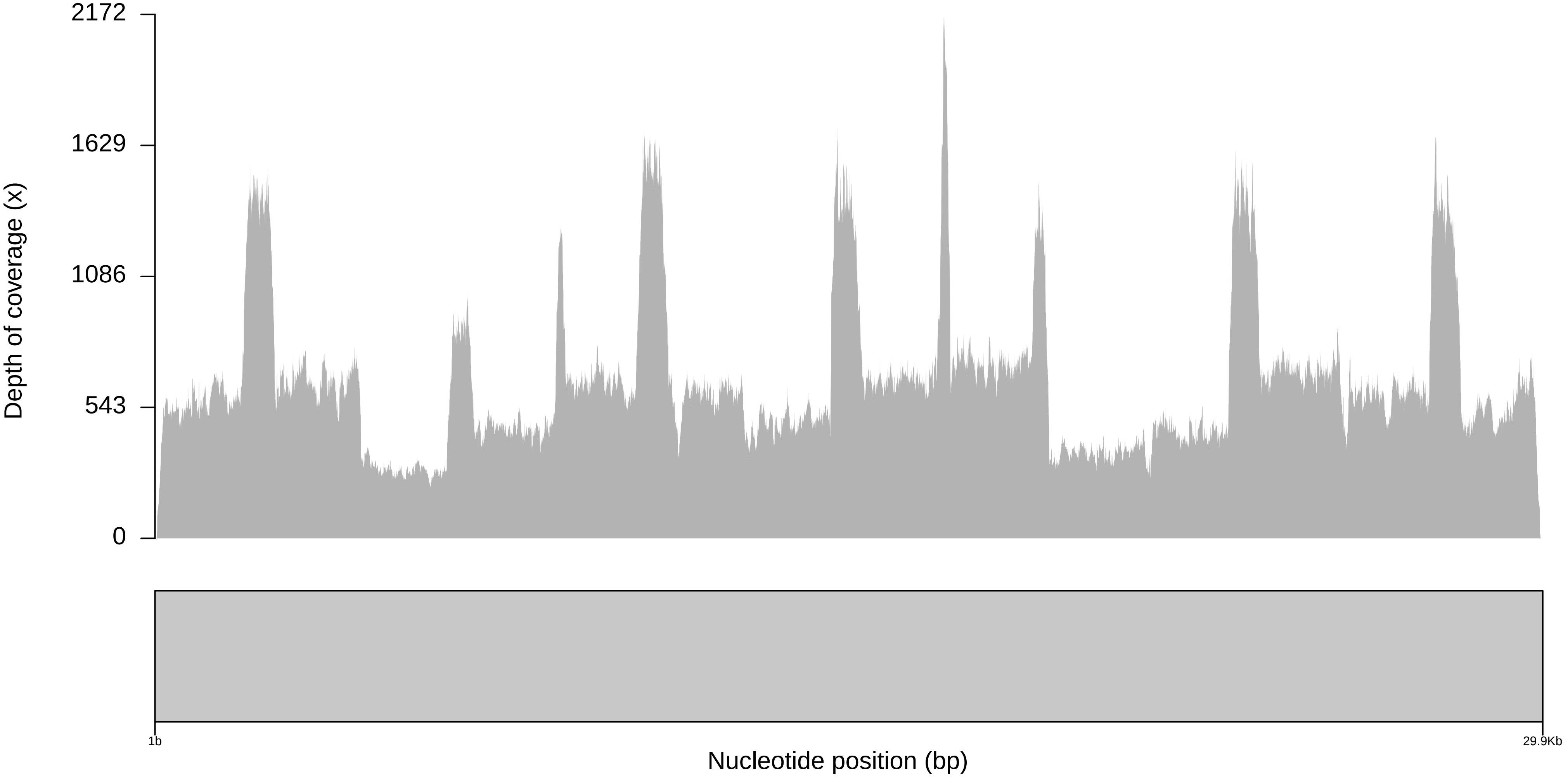

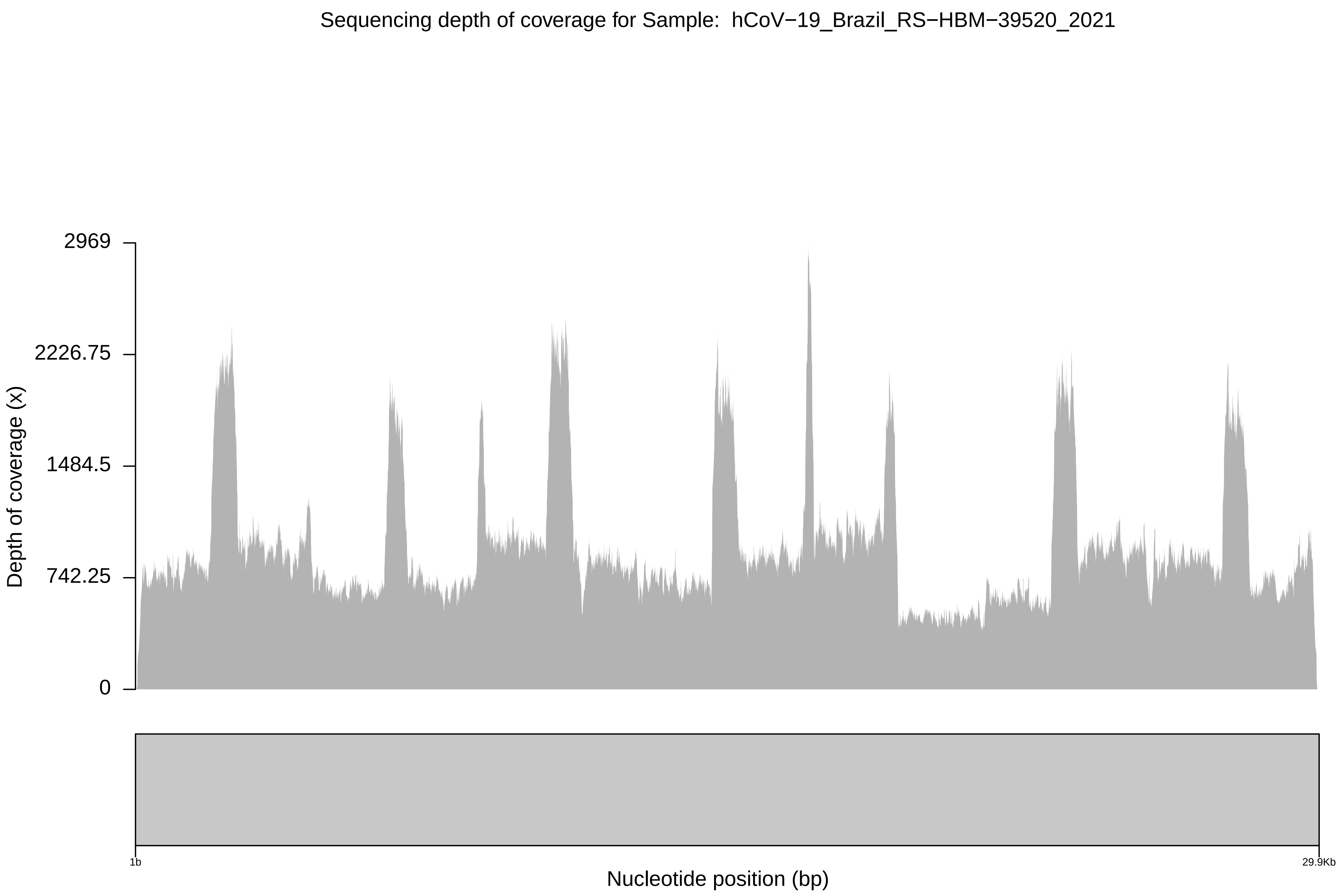

Sequencing depth of coverage for Sample: hCoV−19_Brazil_RS−HBM−39520_2021
2969
2226.75
1484.5
742.25
0
Depth of coverage (x)
1b
29.9Kb
Nucleotide position (bp)

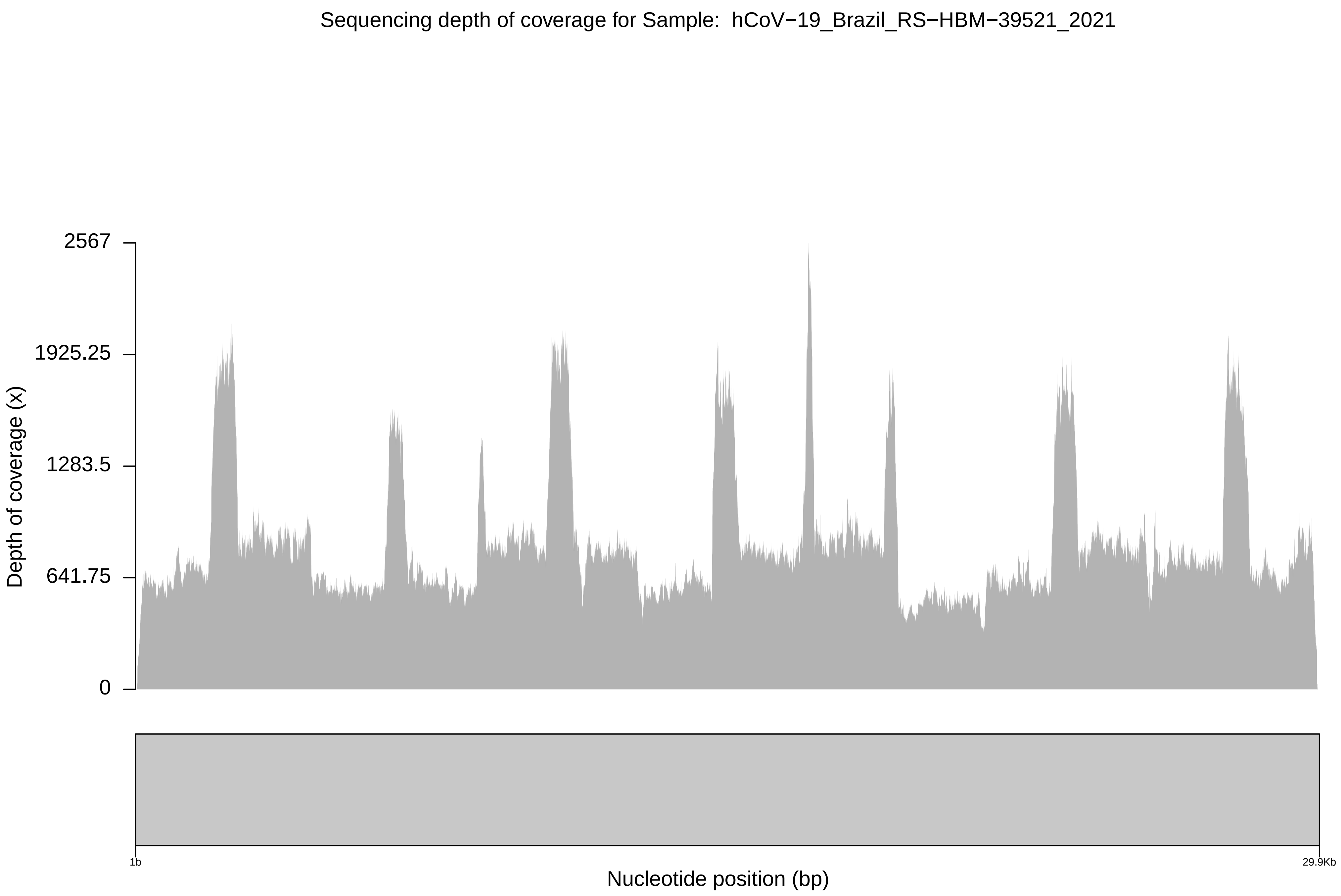

Sequencing depth of coverage for Sample: hCoV−19_Brazil_RS−HBM−39521_2021
Depth of coverage (x)
2567
1925.25
1283.5
641.75
0
1b
29.9Kb
Nucleotide position (bp)

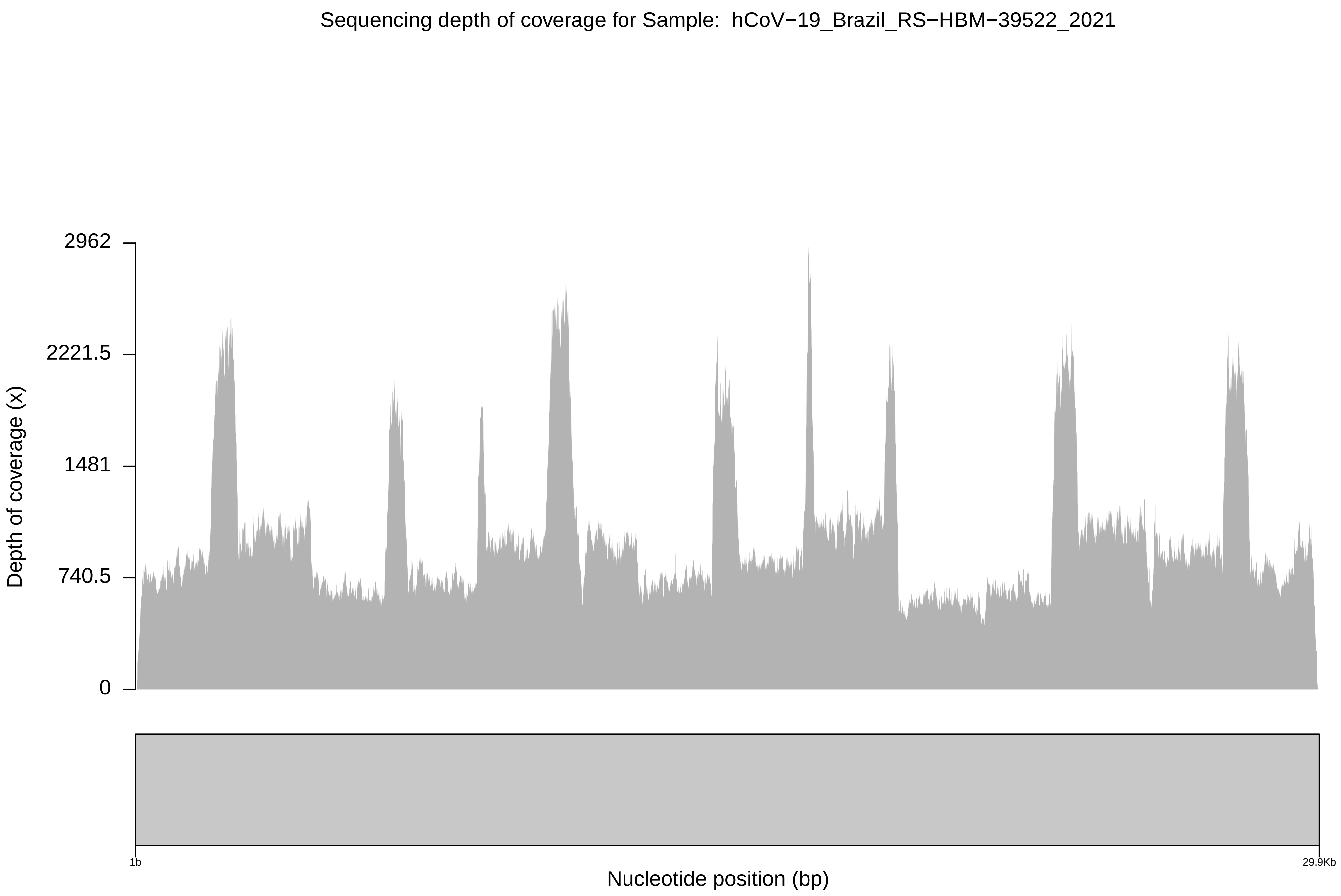
Sequencing depth of coverage for Sample: hCoV−19_Brazil_RS−HBM−39522_2021
2962
2221.5
1481
740.5
0
Depth of coverage (x)
1b
29.9Kb
Nucleotide position (bp)

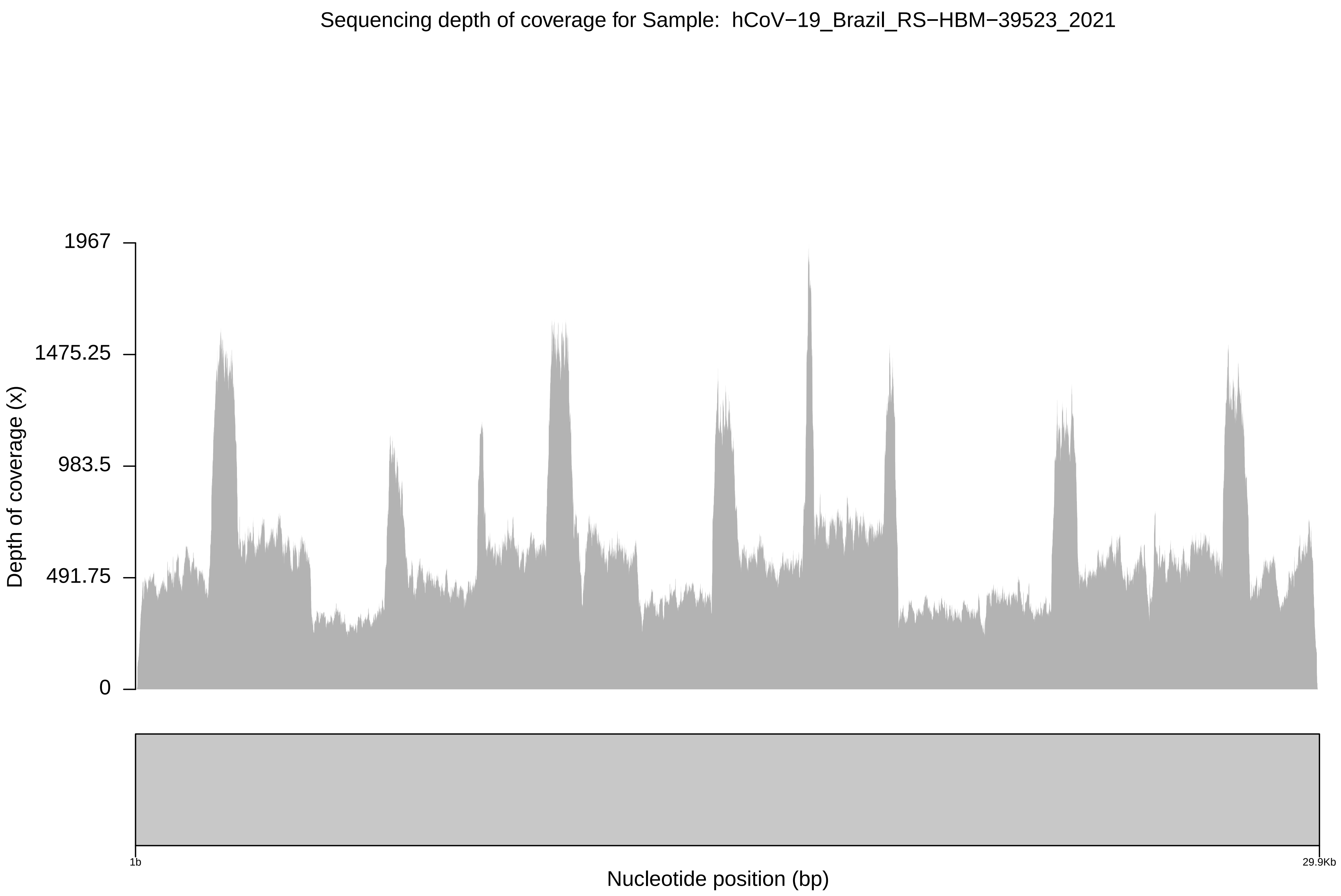

Sequencing depth of coverage for Sample: hCoV−19_Brazil_RS−HBM−39523_2021
1967
1475.25
983.5
491.75
0
Depth of coverage (x)
1b
29.9Kb
Nucleotide position (bp)
