## Supplementary file 2 for "Predominance of the SARS-CoV-2 lineage P.1 and its sublineage P.1.2 in patients from the metropolitan region of Porto Alegre, Southern Brazil in March 2021: a phylogenomic analysis"

| Genomic position | Reference nt | Changed nt | Gene / region | Effect, position inside gene and amino acid replacement | Mutation count | Frequency (%) |
|---|---|---|---|---|---|---|
| 241 | C | T | 5' UTR | intergenic_region n.241C>T | 56 | 100.0 |
| 507 | ATGGTCATGTT ATGGT | A | ORF1ab | disruptive_inframe_deletion c.245_259delGTCATGTTATGGTTG p.Gly82_Val86del | 1 | 1.8 |
| 733 | T | C | | synonymous_variant c.468T>C p.Asp156Asp | 54 | 96.4 |
| 775 | C | T | | synonymous_variant c.510C>T p.Thr170Thr | 1 | 1.8 |
| 792 | A | G | | missense_variant c.527A>G p.Glu176Gly | 1 | 1.8 |
| 843 | C | T | | missense_variant c.578C>T p.Pro193Leu | 1 | 1.8 |
| 851 | T | C | | missense_variant c.586T>C p.Tyr196His | 1 | 1.8 |
| 1395 | A | G | | missense_variant c.1130A>G p.Glu377Gly | 1 | 1.8 |
| 1420 | C | T | | synonymous_variant c.1155C>T p.Ala385Ala | 2 | 3.6 |
| 1463 | G | A | | missense_variant c.1198G>A p.Gly400Ser | 3 | 5.4 |
| 1471 | C | T | | synonymous_variant c.1206C>T p.Arg402Arg | 2 | 3.6 |
| 1757 | G | T | | missense_variant c.1492G>T p.Ala498Ser | 1 | 1.8 |
| 1912 | C | T | | synonymous_variant c.1647C>T p.Ser549Ser | 11 | 19.6 |
| 1942 | G | T | | synonymous_variant c.1677G>T p.Val559Val | 1 | 1.8 |
| 2113 | C | T | | synonymous_variant c.1848C>T p.Ile616Ile | 1 | 1.8 |
| 2445 | C | T | | missense_variant c.2180C>T p.Thr727Ile | 1 | 1.8 |
| 2550 | A | G | | missense_variant c.2285A>G p.Asp762Gly | 11 | 19.6 |
| 2749 | C | T | | synonymous_variant c.2484C>T p.Asp828Asp | 53 | 94.6 |
| 2783 | A | G | | missense_variant c.2518A>G p.Asn840Asp | 1 | 1.8 |
| 2890 | G | T | | synonymous_variant c.2625G>T p.Val875Val | 1 | 1.8 |
| 3037 | C | T | | synonymous_variant c.2772C>T p.Phe924Phe | 56 | 100.0 |
| 3096 | C | T | | missense_variant c.2831C>T p.Ser944Leu | 1 | 1.8 |
| 3411 | C | T | | missense_variant c.3146C>T p.Ala1049Val | 2 | 3.6 |

| | | | | | |
|---|---|---|---|---|---|
| 3766 | T | C | synonymous_variant c.3501T>C p.Asp1167Asp | 1 | 1.8 |
| 3783 | T | C | missense_variant c.3518T>C p.Val1173Ala | 1 | 1.8 |
| 3787 | C | T | synonymous_variant c.3522C>T p.Tyr1174Tyr | 1 | 1.8 |
| 3828 | C | T | missense_variant c.3563C>T p.Ser1188Leu | 54 | 96.4 |
| 4655 | C | T | missense_variant c.4390C>T p.Arg1464Trp | 1 | 1.8 |
| 4705 | T | C | synonymous_variant c.4440T>C p.Pro1480Pro | 3 | 5.4 |
| 4708 | T | C | synonymous_variant c.4443T>C p.Asp1481Asp | 1 | 1.8 |
| 4901 | C | T | synonymous_variant c.4636C>T p.Leu1546Leu | 1 | 1.8 |
| 5063 | G | A | missense_variant c.4798G>A p.Asp1600Asn | 2 | 3.6 |
| 5407 | C | T | synonymous_variant c.5142C>T p.Ile1714Ile | 1 | 1.8 |
| 5648 | A | C | missense_variant c.5383A>C p.Lys1795Gln | 48 | 85.7 |
| 5724 | C | T | missense_variant c.5459C>T p.Thr1820Ile | 10 | 17.9 |
| 5986 | C | T | synonymous_variant c.5721C>T p.Phe1907Phe | 1 | 1.8 |
| 6218 | T | C | synonymous_variant c.5953T>C p.Leu1985Leu | 1 | 1.8 |
| 6319 | A | G | synonymous_variant c.6054A>G p.Pro2018Pro | 49 | 87.5 |
| 6363 | C | T | missense_variant c.6098C>T p.Ala2033Val | 1 | 1.8 |
| 6525 | C | T | missense_variant c.6260C>T p.Thr2087Ile | 1 | 1.8 |
| 6576 | G | T | missense_variant c.6311G>T p.Ser2104Ile | 1 | 1.8 |
| 6613 | A | G | synonymous_variant c.6348A>G p.Val2116Val | 54 | 96.4 |
| 6840 | C | T | missense_variant c.6575C>T p.Ala2192Val | 1 | 1.8 |
| 7201 | A | G | synonymous_variant c.6936A>G p.Ser2312Ser | 1 | 1.8 |
| 8092 | C | T | synonymous_variant c.7827C>T p.Leu2609Leu | 1 | 1.8 |
| 8128 | A | T | synonymous_variant c.7863A>T p.Ala2621Ala | 1 | 1.8 |
| 8392 | T | A | synonymous_variant c.8127T>A p.Ile2709Ile | 2 | 3.6 |
| 8849 | G | T | missense_variant c.8584G>T p.Val2862Leu | 4 | 7.1 |
| 8905 | C | T | synonymous_variant c.8640C>T p.Asp2880Asp | 1 | 1.8 |
| 9042 | C | T | missense_variant c.8777C>T p.Ser2926Phe | 1 | 1.8 |

| | | | | | |
|---|---|---|---|---|---|
| 9052 | A | G | synonymous_variant c.8787A>G p.Pro2929Pro | 1 | 1.8 |
| 9203 | G | C | missense_variant c.8938G>C p.Asp2980His | 1 | 1.8 |
| 9329 | G | A | missense_variant c.9064G>A p.Asp3022Asn | 1 | 1.8 |
| 9532 | C | T | synonymous_variant c.9267C>T p.Phe3089Phe | 1 | 1.8 |
| 9856 | G | T | synonymous_variant c.9591G>T p.Val3197Val | 1 | 1.8 |
| 10096 | G | A | synonymous_variant c.9831G>A p.Glu3277Glu | 2 | 3.6 |
| 10138 | C | T | synonymous_variant c.9873C>T p.Asn3291Asn | 1 | 1.8 |
| 10369 | C | T | synonymous_variant c.10104C>T p.Arg3368Arg | 1 | 1.8 |
| 10507 | C | T | synonymous_variant c.10242C>T p.Asn3414Asn | 4 | 7.1 |
| 10525 | C | T | synonymous_variant c.10260C>T p.Val3420Val | 1 | 1.8 |
| 10667 | T | G | missense_variant c.10402T>G p.Leu3468Val | 1 | 1.8 |
| 10851 | C | T | missense_variant c.10586C>T p.Ala3529Val | 1 | 1.8 |
| 10885 | T | C | synonymous_variant c.10620T>C p.Asn3540Asn | 1 | 1.8 |
| 10981 | G | T | synonymous_variant c.10716G>T p.Val3572Val | 1 | 1.8 |
| 11095 | C | T | synonymous_variant c.10830C>T p.Ala3610Ala | 3 | 5.4 |
| 11287 | GTCTGGTTTT | G | conservative_inframe_deletion c.11023_11031delTCTGGTTTT p.Ser3675_Phe3677del | 4 | 7.1 |
| 11291 | G | A | missense_variant c.11026G>A p.Gly3676Ser | 6 | 10.7 |
| 11296 | T | G | missense_variant c.11031T>G p.Phe3677Leu | 17 | 30.4 |
| 11518 | C | T | synonymous_variant c.11253C>T p.Val3751Val | 3 | 5.4 |
| 11750 | C | T | missense_variant c.11485C>T p.Leu3829Phe | 1 | 1.8 |
| 11824 | C | T | synonymous_variant c.11559C>T p.Ile3853Ile | 1 | 1.8 |
| 12053 | C | T | missense_variant c.11788C>T p.Leu3930Phe | 2 | 3.6 |
| 12174 | A | G | missense_variant c.11909A>G p.Asn3970Ser | 1 | 1.8 |
| 12191 | G | T | missense_variant c.11926G>T p.Val3976Phe | 1 | 1.8 |
| 12778 | C | T | synonymous_variant c.12513C>T p.Tyr4171Tyr | 53 | 94.6 |
| 12809 | C | T | missense_variant c.12544C>T p.Leu4182Phe | 1 | 1.8 |

| | | | | | |
|---|---|---|---|---|---|
| 12964 | A | G | synonymous_variant c.12699A>G p.Gly4233Gly | 1 | 1.8 |
| 13059 | C | T | missense_variant c.12794C>T p.Thr4265Ile | 1 | 1.8 |
| 13176 | C | T | missense_variant c.12911C>T p.Thr4304Ile | 1 | 1.8 |
| 13459 | G | T | synonymous_variant c.13194G>T p.Ser4398Ser | 1 | 1.8 |
| 13536 | C | T | missense_variant c.13271C>T p.Thr4424Ile | 1 | 1.8 |
| 13572 | T | C | missense_variant c.13307T>C p.Val4436Ala | 1 | 1.8 |
| 13724 | C | T | synonymous_variant c.13459C>T p.Leu4487Leu | 1 | 1.8 |
| 13860 | C | T | missense_variant c.13595C>T p.Thr4532Ile | 53 | 94.6 |
| 14408 | C | T | synonymous_variant c.14143C>T p.Leu4715Leu | 55 | 98.2 |
| 15043 | G | A | missense_variant c.14778G>A p.Met4926Ile | 1 | 1.8 |
| 15082 | A | G | synonymous_variant c.14817A>G p.Pro4939Pro | 1 | 1.8 |
| 15708 | G | T | stop_lost c.15443G>T p.Ter5148Leuext*? | 1 | 1.8 |
| 15903 | A | G | missense_variant c.15638A>G p.Asn5213Ser | 1 | 1.8 |
| 16611 | C | T | missense_variant c.16346C>T p.Pro5449Leu | 1 | 1.8 |
| 16794 | T | C | missense_variant c.16529T>C p.Val5510Ala | 1 | 1.8 |
| 16887 | C | T | missense_variant c.16622C>T p.Thr5541Ile | 3 | 5.4 |
| 16954 | C | T | synonymous_variant c.16689C>T p.His5563His | 1 | 1.8 |
| 17259 | G | T | missense_variant c.16994G>T p.Ser5665Ile | 53 | 94.6 |
| 17562 | G | T | missense_variant c.17297G>T p.Gly5766Val | 1 | 1.8 |
| 17706 | G | A | missense_variant c.17441G>A p.Gly5814Asp | 1 | 1.8 |
| 17889 | A | G | missense_variant c.17624A>G p.Lys5875Arg | 1 | 1.8 |
| 18973 | G | A | synonymous_variant c.18708G>A p.Arg6236Arg | 1 | 1.8 |
| 20149 | A | C | synonymous_variant c.19884A>C p.Ser6628Ser | 1 | 1.8 |
| 20730 | G | T | missense_variant c.20465G>T p.Ser6822Ile | 3 | 5.4 |
| 20877 | T | C | missense_variant c.20612T>C p.Val6871Ala | 1 | 1.8 |
| 20931 | G | A | missense_variant c.20666G>A p.Arg6889Gln | 1 | 1.8 |
| 21519 | A | C | missense_variant c.21254A>C p.Glu7085Ala | 1 | 1.8 |

| | | | | | | |
|---|---|---|---|---|---|---|
| 21614 | C | T | | missense_variant c.52C>T p.Leu18Phe | 54 | 96.4 |
| 21621 | C | A | | missense_variant c.59C>A p.Thr20Asn | 53 | 94.6 |
| 21638 | C | T | | missense_variant c.76C>T p.Pro26Ser | 54 | 96.4 |
| 21974 | G | T | | missense_variant c.412G>T p.Asp138Tyr | 54 | 96.4 |
| 22020 | T | C | | missense_variant c.458T>C p.Met153Thr | 1 | 1.8 |
| 22132 | G | T | | missense_variant c.570G>T p.Arg190Ser | 54 | 96.4 |
| 22336 | G | T | | missense_variant c.774G>T p.Trp258Cys | 1 | 1.8 |
| 22413 | C | T | | missense_variant c.851C>T p.Thr284Ile | 1 | 1.8 |
| 22487 | G | A | | missense_variant c.925G>A p.Glu309Lys | 1 | 1.8 |
| 22812 | A | C | | missense_variant c.1250A>C p.Lys417Thr | 54 | 96.4 |
| 23012 | G | A | | missense_variant c.1450G>A p.Glu484Lys | 54 | 96.4 |
| 23063 | A | T | Spike | missense_variant c.1501A>T p.Asn501Tyr | 54 | 96.4 |
| 23403 | A | G | | missense_variant c.1841A>G p.Asp614Gly | 56 | 100.0 |
| 23417 | G | C | | missense_variant c.1855G>C p.Glu619Gln | 1 | 1.8 |
| 23525 | C | T | | missense_variant c.1963C>T p.His655Tyr | 54 | 96.4 |
| 23628 | G | T | | missense_variant c.2066G>T p.Ser689Ile | 1 | 1.8 |
| 23639 | A | T | | missense_variant c.2077A>T p.Ile693Phe | 1 | 1.8 |
| 24023 | C | T | | synonymous_variant c.2461C>T p.Leu821Leu | 1 | 1.8 |
| 24616 | C | T | | synonymous_variant c.3054C>T p.Ile1018Ile | 1 | 1.8 |
| 24642 | C | T | | missense_variant c.3080C>T p.Thr1027Ile | 54 | 96.4 |
| 25088 | G | T | | missense_variant c.3526G>T p.Val1176Phe | 56 | 100.0 |
| 25158 | A | G | | missense_variant c.3596A>G p.Asp1199Gly | 1 | 1.8 |
| 25294 | C | T | | synonymous_variant c.3732C>T p.Leu1244Leu | 1 | 1.8 |
| 25314 | G | T | | missense_variant c.3752G>T p.Gly1251Val | 1 | 1.8 |
| 25423 | G | C | | missense_variant c.31G>C p.Gly11Arg | 1 | 1.8 |
| 25471 | G | T | ORF3a | missense_variant c.79G>T p.Asp27Tyr | 1 | 1.8 |
| 25483 | G | A | | missense_variant c.91G>A p.Ala31Thr | 1 | 1.8 |

| | | | | | | |
|---|---|---|---|---|---|---|
| 25563 | G | T | | missense_variant c.171G>T p.Gln57His | 1 | 1.8 |
| 25641 | G | T | | missense_variant c.249G>T p.Leu83Phe | 8 | 14.3 |
| 25658 | C | T | | missense_variant c.266C>T p.Thr89Ile | 1 | 1.8 |
| 25725 | A | G | | synonymous_variant c.333A>G p.Leu111Leu | 1 | 1.8 |
| 25728 | C | T | | synonymous_variant c.336C>T p.Val112Val | 2 | 3.6 |
| 25771 | C | T | | missense_variant c.379C>T p.Leu127Phe | 1 | 1.8 |
| 25855 | G | T | | missense_variant c.463G>T p.Asp155Tyr | 11 | 19.6 |
| 26063 | G | T | | missense_variant c.671G>T p.Gly224Val | 1 | 1.8 |
| 26149 | T | C | | missense_variant c.757T>C p.Ser253Pro | 53 | 94.6 |
| 26149 | TCCG | CCCT | | stop_gained c.757_760delTCCGinsCCCT p.SerGly253* | 1 | 1.8 |
| 26171 | T | A | | missense_variant c.779T>A p.Met260Lys | 4 | 7.1 |
| 26861 | T | C | Membrane | synonymous_variant c.339T>C p.Asn113Asn | 3 | 5.4 |
| 26936 | C | T | | synonymous_variant c.414C>T p.Leu138Leu | 1 | 1.8 |
| 26988 | C | T | | synonymous_variant c.466C>T p.Leu156Leu | 1 | 1.8 |
| 27441 | G | T | ORF7a | missense_variant c.48G>T p.Glu16Asp | 3 | 5.4 |
| 27467 | G | T | | missense_variant c.74G>T p.Arg25Ile | 1 | 1.8 |
| 27496 | T | A | | missense_variant c.103T>A p.Cys35Ser | 1 | 1.8 |
| 27506 | G | T | | missense_variant c.113G>T p.Gly38Val | 1 | 1.8 |
| 27513 | C | T | | synonymous_variant c.120C>T p.Tyr40Tyr | 1 | 1.8 |
| 27684 | C | T | | synonymous_variant c.291C>T p.Tyr97Tyr | 1 | 1.8 |
| 27982 | C | T | ORF8 | missense_variant c.89C>T p.Pro30Leu | 1 | 1.8 |
| 28003 | G | T | | missense_variant c.110G>T p.Cys37Phe | 1 | 1.8 |
| 28167 | G | A | | missense_variant c.274G>A p.Glu92Lys | 53 | 94.6 |
| 28199 | T | C | | synonymous_variant c.306T>C p.Cys102Cys | 1 | 1.8 |
| 28253 | C | T | | synonymous_variant c.360C>T p.Phe120Phe | 2 | 3.6 |
| 28333 | C | T | Nucleocapsid | synonymous_variant c.60C>T p.Pro20Pro | 1 | 1.8 |
| 28436 | G | T | | missense_variant c.163G>T p.Ala55Ser | 1 | 1.8 |

| | | | | | | |
|---|---|---|---|---|---|---|
| 28512 | C | G | | missense_variant c.239C>G p.Pro80Arg | 54 | 96.4 |
| 28603 | C | T | | synonymous_variant c.330C>T p.Phe110Phe | 1 | 1.8 |
| 28628 | G | T | | missense_variant c.355G>T p.Ala119Ser | 1 | 1.8 |
| 28705 | T | C | | synonymous_variant c.432T>C p.Asp144Asp | 1 | 1.8 |
| 28789 | C | T | | synonymous_variant c.516C>T p.Tyr172Tyr | 11 | 19.6 |
| 28806 | G | T | | missense_variant c.533G>T p.Gly178Val | 1 | 1.8 |
| 28817 | G | T | | missense_variant c.544G>T p.Ala182Ser | 1 | 1.8 |
| 28877 | AGTAGGG | TCTAAAC | | missense_variant c.604_610delAGTAGGGinsTCTAAAC p.ArgGly203LysArg | 54 | 96.4 |
| 28881 | GGG | AAC | | missense_variant c.608_610delGGGinsAAC p.ArgGly203LysArg | 2 | 3.6 |
| 28893 | C | T | | missense_variant c.620C>T p.Pro207Leu | 1 | 1.8 |
| 28975 | G | T | | missense_variant c.702G>T p.Met234Ile | 1 | 1.8 |
| 29095 | C | T | | synonymous_variant c.822C>T p.Phe274Phe | 3 | 5.4 |
| 29160 | C | T | | missense_variant c.887C>T p.Thr296Ile | 1 | 1.8 |
| 29197 | C | T | | synonymous_variant c.924C>T p.Ala308Ala | 1 | 1.8 |
| 29421 | C | T | | missense_variant c.1148C>T p.Pro383Leu | 1 | 1.8 |
| 29511 | G | T | | missense_variant c.1238G>T p.Ser413Ile | 1 | 1.8 |
| 29688 | G | T | 3' UTR | intergenic_region n.29688G>T | 1 | 1.8 |
| 29736 | G | T | | intergenic_region n.29736G>T | 1 | 1.8 |
