## Supplementary file 3 for "Predominance of the SARS-CoV-2 lineage P.1 and its sublineage P.1.2 in patients from the metropolitan region of Porto Alegre, Southern Brazil in March 2021: a phylogenomic analysis"

**We gratefully acknowledge the following Authors from the Originating laboratories responsible for obtaining the specimens, as well as the Submitting laboratories where the genome data were generated and shared via GISAID, on which this research is based.**

**All Submitters of data may be contacted directly via www.gisaid.org**

**Authors are sorted alphabetically.**

| Accession ID | Originating Laboratory | Submitting Laboratory | Authors |
|---|---|---|---|
| EPI_ISL_1349621 | Landesgesundheitsamt Baden-Württemberg | Robert Koch Institute | ? |
| EPI_ISL_1432849 | Eurofins LifeCodexx GmbH | Robert Koch Institute | ? |
| EPI_ISL_1349034 | Bayerisches Landesamt für Gesundheit und Lebensmittelsicherheit (LGL) | Robert Koch Institute | ? |
| EPI_ISL_1349038 | Bayerisches Landesamt für Gesundheit und Lebensmittelsicherheit (LGL) | Robert Koch Institute | ? |
| EPI_ISL_1349120 | Bayerisches Landesamt für Gesundheit und Lebensmittelsicherheit (LGL) | Robert Koch Institute | ? |
| EPI_ISL_1357204 | Bayerisches Landesamt für Gesundheit und Lebensmittelsicherheit (LGL) | Robert Koch Institute | ? |
| EPI_ISL_1570339 | Bayerisches Landesamt fÃ¼r Gesundheit und Lebensmittelsicherheit (LGL) | Robert Koch Institute | ? |
| EPI_ISL_1152591 | MVZ Medizinisches Labor Hannover GmbH | Robert Koch Institute | ? |
| EPI_ISL_1144958 | MVZ Labor Krone GbR | Robert Koch Institute | ? |
| EPI_ISL_1644947 | MVZ Labor Krone GbR | Robert Koch Institute | ? |
| EPI_ISL_1573258 | Labor Prof. Dr. G. Enders MVZ GbR | Robert Koch Institute | ? |
| EPI_ISL_1573260 | Labor Prof. Dr. G. Enders MVZ GbR | Robert Koch Institute | ? |
| EPI_ISL_1014679 | National Influenza Center, Virology Department | National Influenza Center | A Nejati et al |
| EPI_ISL_862078 | National Influenza Center, Virology Department | National Influenza Center | A Nejati et al |
| EPI_ISL_1014684 | National Influenza Center, Virology Department | National Influenza Center | A Nejati et al |
| EPI_ISL_791502 | The Hosptial for Sick Children | The Hospital for Sick Children | Aaron Campigotto et al |
| EPI_ISL_445088 | Human Genetic Research Center, Kawsar Biotech Company | Human Genetic Research Center, Kawsar Biotech Company | Abbasalipour Bashash et al |
| EPI_ISL_1341149 | Laboratory of Virology, National center of expertise | RSE National Center of Expertise and RSE National center for Biotechnology | Abdaliyev Askar et al |
| EPI_ISL_1341150 | Laboratory of Virology, National center of expertise | RSE National Center of Expertise and RSE National center for Biotechnology | Abdaliyev Askar et al |
| EPI_ISL_454585 | Laboratory of virology, National Center of Expertise | Laboratory of molecular-genetic research, National Center for Expertise, Kazakhstan National Center for Biotechnology, Kazakhstan | Abdaliyev Askar et al |
| EPI_ISL_1341503 | Laboratory of Virology, National center of expertise | RSE National Center of Expertise and RSE National center for Biotechnology | Abdaliyev Askar et al |
| EPI_ISL_454575 | Laboratory of virology, National Center of Expertise | Laboratory of molecular-genetic research, National Center for Expertise, Kazakhstan National Center for Biotechnology, Kazakhstan | Abdaliyev Askar et al |
| EPI_ISL_1365697 | Laboratory of Virology, National center of expertise | RSE National Center of Expertise and RSE National center for Biotechnology | Abdaliyev Askar et al |

| | | | |
|---|---|---|---|
| EPI_ISL_454571 | National Center of Expertise | National Center for Expertise, National Center for Biotechnology, Kazakhstan | Abdaliyev Askar et al |
| EPI_ISL_454572 | National Center of Expertise | National Center for Expertise, Kazakhstan National Center for Biotechnology, Kazakhstan | Abdaliyev Askar et al |
| EPI_ISL_1341643 | Laboratory of Virology, National center of expertise | RSE National Center of Expertise and RSE National center for Biotechnology | Abdaliyev Askar et al |
| EPI_ISL_1341504 | Laboratory of Virology, National center of expertise | RSE National Center of Expertise and RSE National center for Biotechnology | Abdaliyev Askar et al |
| EPI_ISL_1341641 | Laboratory of Virology, National center of expertise | RSE National Center of Expertise and RSE National center for Biotechnology | Abdaliyev Askar et al |
| EPI_ISL_660446 | Laboratoire de Microbiologie CHU Souro Sanou | Centre Muraz | Abdoul-Salam Ouedraogo et al |
| EPI_ISL_660478 | Laboratoire de Microbiologie CHU Souro Sanou | Centre Muraz | Abdoul-Salam Ouedraogo et al |
| EPI_ISL_660450 | Laboratoire de Microbiologie CHU Souro Sanou | Centre Muraz | Abdoul-Salam Ouedraogo et al |
| EPI_ISL_660451 | Laboratoire de Microbiologie CHU Souro Sanou | Centre Muraz | Abdoul-Salam Ouedraogo et al |
| EPI_ISL_660479 | Laboratoire de Microbiologie CHU Souro Sanou | Centre Muraz | Abdoul-Salam Ouedraogo et al |
| EPI_ISL_660452 | Laboratoire de Microbiologie CHU Souro Sanou | Centre Muraz | Abdoul-Salam Ouedraogo et al |
| EPI_ISL_660496 | Laboratoire de Microbiologie CHU Souro Sanou | Centre Muraz | Abdoul-Salam Ouedraogo et al |
| EPI_ISL_660510 | Laboratoire de Microbiologie CHU Souro Sanou | Centre Muraz | Abdoul-Salam Ouedraogo et al |
| EPI_ISL_660460 | Laboratoire de Microbiologie CHU Souro Sanou | Centre Muraz | Abdoul-Salam Ouedraogo et al |
| EPI_ISL_660520 | Laboratoire de Microbiologie CHU Souro Sanou | Centre Muraz | Abdoul-Salam Ouedraogo et al |
| EPI_ISL_660464 | Laboratoire de Microbiologie CHU Souro Sanou | Centre Muraz | Abdoul-Salam Ouedraogo et al |
| EPI_ISL_660518 | Laboratoire de Microbiologie CHU Souro Sanou | Centre Muraz | Abdoul-Salam Ouedraogo et al |
| EPI_ISL_660506 | Laboratoire de Microbiologie CHU Souro Sanou | Centre Muraz | Abdoul-Salam Ouedraogo et al |
| EPI_ISL_660466 | Laboratoire de Microbiologie CHU Souro Sanou | Centre Muraz | Abdoul-Salam Ouedraogo et al |
| EPI_ISL_660521 | Laboratoire de Microbiologie CHU Souro Sanou | Centre Muraz | Abdoul-Salam Ouedraogo et al |
| EPI_ISL_660507 | Laboratoire de Microbiologie CHU Souro Sanou | Centre Muraz | Abdoul-Salam Ouedraogo et al |
| EPI_ISL_660511 | Laboratoire de Microbiologie CHU Souro Sanou | Centre Muraz | Abdoul-Salam Ouedraogo et al |
| EPI_ISL_660497 | Laboratoire de Microbiologie CHU Souro Sanou | Centre Muraz | Abdoul-Salam Ouedraogo et al |
| EPI_ISL_660473 | Laboratoire de Microbiologie CHU Souro Sanou | Centre Muraz | Abdoul-Salam Ouedraogo et al |
| EPI_ISL_660519 | Laboratoire de Microbiologie CHU Souro Sanou | Centre Muraz | Abdoul-Salam Ouedraogo et al |
| EPI_ISL_660476 | Laboratoire de Microbiologie CHU Souro Sanou | Centre Muraz | Abdoul-Salam Ouedraogo et al |
| EPI_ISL_660522 | Laboratoire de Microbiologie CHU Souro Sanou | Centre Muraz | Abdoul-Salam Ouedraogo et al |
| EPI_ISL_915192 | MRCG at LSHTM Genomics lab | MRCG at LSHTM Genomics lab | Abdul Karim sesay et al |

| | | | |
|---|---|---|---|
| EPI_ISL_1620184 | MRCG at LSHTM Genomics lab | MRCG at LSHTM Genomics lab | Abdul Karim sesay et al |
| EPI_ISL_1620185 | MRCG at LSHTM Genomics lab | MRCG at LSHTM Genomics lab | Abdul Karim sesay et al |
| EPI_ISL_1620177 | MRCG at LSHTM Genomics lab | MRCG at LSHTM Genomics lab | Abdul Karim sesay et al |
| EPI_ISL_1234531 | MRCG at LSHTM Genomics lab | MRCG at LSHTM Genomics lab | Abdul Karim sesay et al |
| EPI_ISL_1234527 | MRCG at LSHTM Genomics lab | MRCG at LSHTM Genomics lab | Abdul Karim sesay et al |
| EPI_ISL_1234534 | MRCG at LSHTM Genomics lab | MRCG at LSHTM Genomics lab | Abdul Karim sesay et al |
| EPI_ISL_1234535 | MRCG at LSHTM Genomics lab | MRCG at LSHTM Genomics lab | Abdul Karim sesay et al |
| EPI_ISL_561024 | MRCG at LSHTM Genomics lab | MRCG at LSHTM Genomics lab | Abdul Karim sesay et al |
| EPI_ISL_561026 | MRCG at LSHTM Genomics lab | MRCG at LSHTM Genomics lab | Abdul Karim sesay et al |
| EPI_ISL_561029 | MRCG at LSHTM Genomics lab | MRCG at LSHTM Genomics lab | Abdul Karim sesay et al |
| EPI_ISL_561033 | MRCG at LSHTM Genomics lab | MRCG at LSHTM Genomics lab | Abdul Karim sesay et al |
| EPI_ISL_561136 | MRCG at LSHTM Genomics lab | MRCG at LSHTM Genomics lab | Abdul Karim sesay et al |
| EPI_ISL_561088 | MRCG at LSHTM Genomics lab | MRCG at LSHTM Genomics lab | Abdul Karim sesay et al |
| EPI_ISL_561207 | MRCG at LSHTM Genomics lab | MRCG at LSHTM Genomics lab | Abdul Karim sesay et al |
| EPI_ISL_561212 | MRCG at LSHTM Genomics lab | MRCG at LSHTM Genomics lab | Abdul Karim sesay et al |
| EPI_ISL_561215 | MRCG at LSHTM Genomics lab | MRCG at LSHTM Genomics lab | Abdul Karim sesay et al |
| EPI_ISL_561216 | MRCG at LSHTM Genomics lab | MRCG at LSHTM Genomics lab | Abdul Karim sesay et al |
| EPI_ISL_561264 | MRCG at LSHTM Genomics lab | MRCG at LSHTM Genomics lab | Abdul Karim sesay et al |
| EPI_ISL_561302 | MRCG at LSHTM Genomics lab | MRCG at LSHTM Genomics lab | Abdul Karim sesay et al |
| EPI_ISL_561312 | MRCG at LSHTM Genomics lab | MRCG at LSHTM Genomics lab | Abdul Karim sesay et al |
| EPI_ISL_1216072 | MRCG at LSHTM Genomics lab | MRCG at LSHTM Genomics lab | Abdul Karim sesay et al |
| EPI_ISL_1216076 | MRCG at LSHTM Genomics lab | MRCG at LSHTM Genomics lab | Abdul Karim sesay et al |
| EPI_ISL_1216077 | MRCG at LSHTM Genomics lab | MRCG at LSHTM Genomics lab | Abdul Karim sesay et al |
| EPI_ISL_1216085 | MRCG at LSHTM Genomics lab | MRCG at LSHTM Genomics lab | Abdul Karim sesay et al |
| EPI_ISL_1216087 | MRCG at LSHTM Genomics lab | MRCG at LSHTM Genomics lab | Abdul Karim sesay et al |
| EPI_ISL_1216119 | MRCG at LSHTM Genomics lab | MRCG at LSHTM Genomics lab | Abdul Karim sesay et al |
| EPI_ISL_1216123 | MRCG at LSHTM Genomics lab | MRCG at LSHTM Genomics lab | Abdul Karim sesay et al |
| EPI_ISL_1216128 | MRCG at LSHTM Genomics lab | MRCG at LSHTM Genomics lab | Abdul Karim sesay et al |
| EPI_ISL_1216130 | MRCG at LSHTM Genomics lab | MRCG at LSHTM Genomics lab | Abdul Karim sesay et al |
| EPI_ISL_1216133 | MRCG at LSHTM Genomics lab | MRCG at LSHTM Genomics lab | Abdul Karim sesay et al |

| | | | |
|---|---|---|---|
| EPI_ISL_810975 | MRCG at LSHTM Genomics lab | MRCG at LSHTM Genomics lab | Abdul Karim sesay et al |
| EPI_ISL_810974 | MRCG at LSHTM Genomics lab | MRCG at LSHTM Genomics lab | Abdul Karim sesay et al |
| EPI_ISL_811015 | MRCG at LSHTM Genomics lab | MRCG at LSHTM Genomics lab | Abdul Karim sesay et al |
| EPI_ISL_810976 | MRCG at LSHTM Genomics lab | MRCG at LSHTM Genomics lab | Abdul Karim sesay et al |
| EPI_ISL_810973 | MRCG at LSHTM Genomics lab | MRCG at LSHTM Genomics lab | Abdul Karim sesay et al |
| EPI_ISL_810984 | MRCG at LSHTM Genomics lab | MRCG at LSHTM Genomics lab | Abdul Karim sesay et al |
| EPI_ISL_811037 | MRCG at LSHTM Genomics lab | MRCG at LSHTM Genomics lab | Abdul Karim sesay et al |
| EPI_ISL_810982 | MRCG at LSHTM Genomics lab | MRCG at LSHTM Genomics lab | Abdul Karim sesay et al |
| EPI_ISL_810986 | MRCG at LSHTM Genomics lab | MRCG at LSHTM Genomics lab | Abdul Karim sesay et al |
| EPI_ISL_1532295 | Oman-National Influenza Center | Biotechnology & OMICs Laboratory | Abdul Latif Khan et al |
| EPI_ISL_1532297 | Oman-National Influenza Center | Biotechnology & OMICs Laboratory | Abdul Latif Khan et al |
| EPI_ISL_1532299 | Oman-National Influenza Center | Biotechnology & OMICs Laboratory | Abdul Latif Khan et al |
| EPI_ISL_1532300 | Oman-National Influenza Center | Biotechnology & OMICs Laboratory | Abdul Latif Khan et al |
| EPI_ISL_1532305 | Oman-National Influenza Center | Biotechnology & OMICs Laboratory | Abdul Latif Khan et al |
| EPI_ISL_518848 | Oman-National Influenza Center | Biotechnology & OMICs Laboratory, Natural & Medical Sciences Research Center, University of Nizwa | Abdul Latif Khan et al |
| EPI_ISL_498554 | Lebanese American University | Lebanese American University | Abi Habib et al |
| EPI_ISL_498556 | Lebanese American University | Lebanese American University | Abi Habib et al |
| EPI_ISL_498551 | Lebanese American University | Lebanese American University | Abi Habib et al |
| EPI_ISL_1533610 | Shiraz University | Shiraz University | Abozar Ghorbani et al |
| EPI_ISL_1588437 | Armed Forces Institute of Pathology (AFIP), Dhaka Cantonment | Genomic Research Lab, BCSIR | Abu Sayeed Mohammad Mahmud et al |
| EPI_ISL_1653926 | DNA Solution Ltd. | Genomic Research Lab, BCSIR | Abu Sayeed Mohammad Mahmud et al |
| EPI_ISL_498818 | National Institute of Laboratory Medicine and Referral Center | Genomic Research Lab, BCSIR | Abu Sayeed Mohammad Mahmud et al |
| EPI_ISL_498830 | National Institute of Laboratory Medicine and Referral Center | Genomic Research Lab, BCSIR | Abu Sayeed Mohammad Mahmud et al |
| EPI_ISL_803867 | National Institute of Laboratory Medicine and Referral Center | Genomic Research Lab, BCSIR | Abu Sayeed Mohammad Mahmud et al |
| EPI_ISL_956332 | Department of Biology, University of Basrah | Department of Biology, University of Basrah | Abu-Ali et al |
| EPI_ISL_907075 | Department of Biology, University of Basrah | Department of Biology, University of Basrah | Abu-Ali et al |
| EPI_ISL_547445 | Dutch COVID-19 response team | National Institute for Public Health and the Environment (RIVM) | Adam Meijer et al |
| EPI_ISL_547446 | Dutch COVID-19 response team | National Institute for Public Health and the Environment (RIVM) | Adam Meijer et al |
| EPI_ISL_547447 | Dutch COVID-19 response team | National Institute for Public Health and the Environment (RIVM) | Adam Meijer et al |
| EPI_ISL_547448 | Dutch COVID-19 response team | National Institute for Public Health and the Environment (RIVM) | Adam Meijer et al |

| | | | |
|---|---|---|---|
| EPI_ISL_547449 | Dutch COVID-19 response team | National Institute for Public Health and the Environment (RIVM) | Adam Meijer et al |
| EPI_ISL_636513 | Dutch COVID-19 response team | National Institute for Public Health and the Environment (RIVM) | Adam Meijer et al |
| EPI_ISL_636514 | Dutch COVID-19 response team | National Institute for Public Health and the Environment (RIVM) | Adam Meijer et al |
| EPI_ISL_636515 | Dutch COVID-19 response team | National Institute for Public Health and the Environment (RIVM) | Adam Meijer et al |
| EPI_ISL_636516 | Dutch COVID-19 response team | National Institute for Public Health and the Environment (RIVM) | Adam Meijer et al |
| EPI_ISL_636517 | Dutch COVID-19 response team | National Institute for Public Health and the Environment (RIVM) | Adam Meijer et al |
| EPI_ISL_636518 | Dutch COVID-19 response team | National Institute for Public Health and the Environment (RIVM) | Adam Meijer et al |
| EPI_ISL_1059910 | Dutch COVID-19 response team | National Institute for Public Health and the Environment (RIVM) | Adam Meijer et al |
| EPI_ISL_1014240 | Dutch COVID-19 response team | National Institute for Public Health and the Environment (RIVM) | Adam Meijer et al |
| EPI_ISL_1014646 | Dutch COVID-19 response team | National Institute for Public Health and the Environment (RIVM) | Adam Meijer et al |
| EPI_ISL_1014645 | Dutch COVID-19 response team | National Institute for Public Health and the Environment (RIVM) | Adam Meijer et al |
| EPI_ISL_1089592 | Dutch COVID-19 response team | National Institute for Public Health and the Environment (RIVM) | Adam Meijer et al |
| EPI_ISL_1090206 | Dutch COVID-19 response team | National Institute for Public Health and the Environment (RIVM) | Adam Meijer et al |
| EPI_ISL_1165384 | Dutch COVID-19 response team | National Institute for Public Health and the Environment (RIVM) | Adam Meijer et al |
| EPI_ISL_1165608 | Dutch COVID-19 response team | National Institute for Public Health and the Environment (RIVM) | Adam Meijer et al |
| EPI_ISL_1165565 | Dutch COVID-19 response team | National Institute for Public Health and the Environment (RIVM) | Adam Meijer et al |
| EPI_ISL_1165616 | Dutch COVID-19 response team | National Institute for Public Health and the Environment (RIVM) | Adam Meijer et al |
| EPI_ISL_1232545 | Dutch COVID-19 response team | National Institute for Public Health and the Environment (RIVM) | Adam Meijer et al |
| EPI_ISL_1232593 | Dutch COVID-19 response team | National Institute for Public Health and the Environment (RIVM) | Adam Meijer et al |
| EPI_ISL_1232714 | Dutch COVID-19 response team | National Institute for Public Health and the Environment (RIVM) | Adam Meijer et al |
| EPI_ISL_1289134 | Dutch COVID-19 response team | National Institute for Public Health and the Environment (RIVM) | Adam Meijer et al |
| EPI_ISL_1289392 | Dutch COVID-19 response team | National Institute for Public Health and the Environment (RIVM) | Adam Meijer et al |
| EPI_ISL_1289394 | Dutch COVID-19 response team | National Institute for Public Health and the Environment (RIVM) | Adam Meijer et al |
| EPI_ISL_1457612 | Dutch COVID-19 response team | National Institute for Public Health and the Environment (RIVM) | Adam Meijer et al |
| EPI_ISL_1370577 | Dutch COVID-19 response team | National Institute for Public Health and the Environment (RIVM) | Adam Meijer et al |
| EPI_ISL_1521157 | Dutch COVID-19 response team | National Institute for Public Health and the Environment (RIVM) | Adam Meijer et al |
| EPI_ISL_1521159 | Dutch COVID-19 response team | National Institute for Public Health and the Environment (RIVM) | Adam Meijer et al |
| EPI_ISL_1521168 | Dutch COVID-19 response team | National Institute for Public Health and the Environment (RIVM) | Adam Meijer et al |
| EPI_ISL_1521731 | Dutch COVID-19 response team | National Institute for Public Health and the Environment (RIVM) | Adam Meijer et al |
| EPI_ISL_1596060 | Dutch COVID-19 response team | National Institute for Public Health and the Environment (RIVM) | Adam Meijer et al |

| | | | |
|---|---|---|---|
| EPI_ISL_1596208 | Dutch COVID-19 response team | National Institute for Public Health and the Environment (RIVM) | Adam Meijer et al |
| EPI_ISL_1596268 | Dutch COVID-19 response team | National Institute for Public Health and the Environment (RIVM) | Adam Meijer et al |
| EPI_ISL_1232249 | Dutch COVID-19 response team | National Institute for Public Health and the Environment (RIVM) | Adam Meijer et al |
| EPI_ISL_1232329 | Dutch COVID-19 response team | National Institute for Public Health and the Environment (RIVM) | Adam Meijer et al |
| EPI_ISL_1232330 | Dutch COVID-19 response team | National Institute for Public Health and the Environment (RIVM) | Adam Meijer et al |
| EPI_ISL_1232632 | Dutch COVID-19 response team | National Institute for Public Health and the Environment (RIVM) | Adam Meijer et al |
| EPI_ISL_1232640 | Dutch COVID-19 response team | National Institute for Public Health and the Environment (RIVM) | Adam Meijer et al |
| EPI_ISL_1232862 | Dutch COVID-19 response team | National Institute for Public Health and the Environment (RIVM) | Adam Meijer et al |
| EPI_ISL_1232869 | Dutch COVID-19 response team | National Institute for Public Health and the Environment (RIVM) | Adam Meijer et al |
| EPI_ISL_1288120 | Dutch COVID-19 response team | National Institute for Public Health and the Environment (RIVM) | Adam Meijer et al |
| EPI_ISL_1288129 | Dutch COVID-19 response team | National Institute for Public Health and the Environment (RIVM) | Adam Meijer et al |
| EPI_ISL_1289398 | Dutch COVID-19 response team | National Institute for Public Health and the Environment (RIVM) | Adam Meijer et al |
| EPI_ISL_1288131 | Dutch COVID-19 response team | National Institute for Public Health and the Environment (RIVM) | Adam Meijer et al |
| EPI_ISL_1289399 | Dutch COVID-19 response team | National Institute for Public Health and the Environment (RIVM) | Adam Meijer et al |
| EPI_ISL_1371226 | Dutch COVID-19 response team | National Institute for Public Health and the Environment (RIVM) | Adam Meijer et al |
| EPI_ISL_1371251 | Dutch COVID-19 response team | National Institute for Public Health and the Environment (RIVM) | Adam Meijer et al |
| EPI_ISL_1371258 | Dutch COVID-19 response team | National Institute for Public Health and the Environment (RIVM) | Adam Meijer et al |
| EPI_ISL_1457749 | Dutch COVID-19 response team | National Institute for Public Health and the Environment (RIVM) | Adam Meijer et al |
| EPI_ISL_1457751 | Dutch COVID-19 response team | National Institute for Public Health and the Environment (RIVM) | Adam Meijer et al |
| EPI_ISL_1521848 | Dutch COVID-19 response team | National Institute for Public Health and the Environment (RIVM) | Adam Meijer et al |
| EPI_ISL_1521860 | Dutch COVID-19 response team | National Institute for Public Health and the Environment (RIVM) | Adam Meijer et al |
| EPI_ISL_1596423 | Dutch COVID-19 response team | National Institute for Public Health and the Environment (RIVM) | Adam Meijer et al |
| EPI_ISL_1596437 | Dutch COVID-19 response team | National Institute for Public Health and the Environment (RIVM) | Adam Meijer et al |
| EPI_ISL_547450 | Dutch COVID-19 response team | National Institute for Public Health and the Environment (RIVM) | Adam Meijer et al |
| EPI_ISL_547451 | Dutch COVID-19 response team | National Institute for Public Health and the Environment (RIVM) | Adam Meijer et al |
| EPI_ISL_547452 | Dutch COVID-19 response team | National Institute for Public Health and the Environment (RIVM) | Adam Meijer et al |
| EPI_ISL_547453 | Dutch COVID-19 response team | National Institute for Public Health and the Environment (RIVM) | Adam Meijer et al |
| EPI_ISL_636492 | Dutch COVID-19 response team | National Institute for Public Health and the Environment (RIVM) | Adam Meijer et al |
| EPI_ISL_636493 | Dutch COVID-19 response team | National Institute for Public Health and the Environment (RIVM) | Adam Meijer et al |
| EPI_ISL_636519 | Dutch COVID-19 response team | National Institute for Public Health and the Environment (RIVM) | Adam Meijer et al |

| | | | |
|---|---|---|---|
| EPI_ISL_636520 | Dutch COVID-19 response team | National Institute for Public Health and the Environment (RIVM) | Adam Meijer et al |
| EPI_ISL_636521 | Dutch COVID-19 response team | National Institute for Public Health and the Environment (RIVM) | Adam Meijer et al |
| EPI_ISL_1014596 | Dutch COVID-19 response team | National Institute for Public Health and the Environment (RIVM) | Adam Meijer et al |
| EPI_ISL_1014595 | Dutch COVID-19 response team | National Institute for Public Health and the Environment (RIVM) | Adam Meijer et al |
| EPI_ISL_1014591 | Dutch COVID-19 response team | National Institute for Public Health and the Environment (RIVM) | Adam Meijer et al |
| EPI_ISL_1014590 | Dutch COVID-19 response team | National Institute for Public Health and the Environment (RIVM) | Adam Meijer et al |
| EPI_ISL_1014589 | Dutch COVID-19 response team | National Institute for Public Health and the Environment (RIVM) | Adam Meijer et al |
| EPI_ISL_1014587 | Dutch COVID-19 response team | National Institute for Public Health and the Environment (RIVM) | Adam Meijer et al |
| EPI_ISL_1014586 | Dutch COVID-19 response team | National Institute for Public Health and the Environment (RIVM) | Adam Meijer et al |
| EPI_ISL_1014583 | Dutch COVID-19 response team | National Institute for Public Health and the Environment (RIVM) | Adam Meijer et al |
| EPI_ISL_1035771 | Dutch COVID-19 response team | National Institute for Public Health and the Environment (RIVM) | Adam Meijer et al |
| EPI_ISL_1035770 | Dutch COVID-19 response team | National Institute for Public Health and the Environment (RIVM) | Adam Meijer et al |
| EPI_ISL_1035767 | Dutch COVID-19 response team | National Institute for Public Health and the Environment (RIVM) | Adam Meijer et al |
| EPI_ISL_1035766 | Dutch COVID-19 response team | National Institute for Public Health and the Environment (RIVM) | Adam Meijer et al |
| EPI_ISL_1232248 | Dutch COVID-19 response team | National Institute for Public Health and the Environment (RIVM) | Adam Meijer et al |
| EPI_ISL_1232269 | Dutch COVID-19 response team | National Institute for Public Health and the Environment (RIVM) | Adam Meijer et al |
| EPI_ISL_1232282 | Dutch COVID-19 response team | National Institute for Public Health and the Environment (RIVM) | Adam Meijer et al |
| EPI_ISL_1232290 | Dutch COVID-19 response team | National Institute for Public Health and the Environment (RIVM) | Adam Meijer et al |
| EPI_ISL_1232292 | Dutch COVID-19 response team | National Institute for Public Health and the Environment (RIVM) | Adam Meijer et al |
| EPI_ISL_1232326 | Dutch COVID-19 response team | National Institute for Public Health and the Environment (RIVM) | Adam Meijer et al |
| EPI_ISL_1232327 | Dutch COVID-19 response team | National Institute for Public Health and the Environment (RIVM) | Adam Meijer et al |
| EPI_ISL_1232328 | Dutch COVID-19 response team | National Institute for Public Health and the Environment (RIVM) | Adam Meijer et al |
| EPI_ISL_1233007 | Dutch COVID-19 response team | National Institute for Public Health and the Environment (RIVM) | Adam Meijer et al |
| EPI_ISL_1288141 | Dutch COVID-19 response team | National Institute for Public Health and the Environment (RIVM) | Adam Meijer et al |
| EPI_ISL_1288143 | Dutch COVID-19 response team | National Institute for Public Health and the Environment (RIVM) | Adam Meijer et al |
| EPI_ISL_1288144 | Dutch COVID-19 response team | National Institute for Public Health and the Environment (RIVM) | Adam Meijer et al |
| EPI_ISL_1288149 | Dutch COVID-19 response team | National Institute for Public Health and the Environment (RIVM) | Adam Meijer et al |
| EPI_ISL_1370641 | Dutch COVID-19 response team | National Institute for Public Health and the Environment (RIVM) | Adam Meijer et al |
| EPI_ISL_1369778 | Dutch COVID-19 response team | National Institute for Public Health and the Environment (RIVM) | Adam Meijer et al |
| EPI_ISL_1370667 | Dutch COVID-19 response team | National Institute for Public Health and the Environment (RIVM) | Adam Meijer et al |

| | | | |
|---|---|---|---|
| EPI_ISL_1369786 | Dutch COVID-19 response team | National Institute for Public Health and the Environment (RIVM) | Adam Meijer et al |
| EPI_ISL_1457573 | Dutch COVID-19 response team | National Institute for Public Health and the Environment (RIVM) | Adam Meijer et al |
| EPI_ISL_1457641 | Dutch COVID-19 response team | National Institute for Public Health and the Environment (RIVM) | Adam Meijer et al |
| EPI_ISL_1521490 | Dutch COVID-19 response team | National Institute for Public Health and the Environment (RIVM) | Adam Meijer et al |
| EPI_ISL_1014579 | Dutch COVID-19 response team | National Institute for Public Health and the Environment (RIVM) | Adam Meijer et al |
| EPI_ISL_1014578 | Dutch COVID-19 response team | National Institute for Public Health and the Environment (RIVM) | Adam Meijer et al |
| EPI_ISL_1014577 | Dutch COVID-19 response team | National Institute for Public Health and the Environment (RIVM) | Adam Meijer et al |
| EPI_ISL_1014576 | Dutch COVID-19 response team | National Institute for Public Health and the Environment (RIVM) | Adam Meijer et al |
| EPI_ISL_1014575 | Dutch COVID-19 response team | National Institute for Public Health and the Environment (RIVM) | Adam Meijer et al |
| EPI_ISL_1014574 | Dutch COVID-19 response team | National Institute for Public Health and the Environment (RIVM) | Adam Meijer et al |
| EPI_ISL_1014573 | Dutch COVID-19 response team | National Institute for Public Health and the Environment (RIVM) | Adam Meijer et al |
| EPI_ISL_1014572 | Dutch COVID-19 response team | National Institute for Public Health and the Environment (RIVM) | Adam Meijer et al |
| EPI_ISL_1014570 | Dutch COVID-19 response team | National Institute for Public Health and the Environment (RIVM) | Adam Meijer et al |
| EPI_ISL_1014569 | Dutch COVID-19 response team | National Institute for Public Health and the Environment (RIVM) | Adam Meijer et al |
| EPI_ISL_1014568 | Dutch COVID-19 response team | National Institute for Public Health and the Environment (RIVM) | Adam Meijer et al |
| EPI_ISL_1014567 | Dutch COVID-19 response team | National Institute for Public Health and the Environment (RIVM) | Adam Meijer et al |
| EPI_ISL_1014566 | Dutch COVID-19 response team | National Institute for Public Health and the Environment (RIVM) | Adam Meijer et al |
| EPI_ISL_1014564 | Dutch COVID-19 response team | National Institute for Public Health and the Environment (RIVM) | Adam Meijer et al |
| EPI_ISL_1014563 | Dutch COVID-19 response team | National Institute for Public Health and the Environment (RIVM) | Adam Meijer et al |
| EPI_ISL_1014562 | Dutch COVID-19 response team | National Institute for Public Health and the Environment (RIVM) | Adam Meijer et al |
| EPI_ISL_1014560 | Dutch COVID-19 response team | National Institute for Public Health and the Environment (RIVM) | Adam Meijer et al |
| EPI_ISL_1014559 | Dutch COVID-19 response team | National Institute for Public Health and the Environment (RIVM) | Adam Meijer et al |
| EPI_ISL_1014557 | Dutch COVID-19 response team | National Institute for Public Health and the Environment (RIVM) | Adam Meijer et al |
| EPI_ISL_1014556 | Dutch COVID-19 response team | National Institute for Public Health and the Environment (RIVM) | Adam Meijer et al |
| EPI_ISL_1014555 | Dutch COVID-19 response team | National Institute for Public Health and the Environment (RIVM) | Adam Meijer et al |
| EPI_ISL_1014554 | Dutch COVID-19 response team | National Institute for Public Health and the Environment (RIVM) | Adam Meijer et al |
| EPI_ISL_1014553 | Dutch COVID-19 response team | National Institute for Public Health and the Environment (RIVM) | Adam Meijer et al |
| EPI_ISL_1014552 | Dutch COVID-19 response team | National Institute for Public Health and the Environment (RIVM) | Adam Meijer et al |
| EPI_ISL_1014551 | Dutch COVID-19 response team | National Institute for Public Health and the Environment (RIVM) | Adam Meijer et al |
| EPI_ISL_1014549 | Dutch COVID-19 response team | National Institute for Public Health and the Environment (RIVM) | Adam Meijer et al |

| | | | |
|---|---|---|---|
| EPI_ISL_1596269 | Dutch COVID-19 response team | National Institute for Public Health and the Environment (RIVM) | Adam Meijer et al |
| EPI_ISL_1596270 | Dutch COVID-19 response team | National Institute for Public Health and the Environment (RIVM) | Adam Meijer et al |
| EPI_ISL_1289218 | Dutch COVID-19 response team | National Institute for Public Health and the Environment (RIVM) | Adam Meijer et al |
| EPI_ISL_1456646 | Dutch COVID-19 response team | National Institute for Public Health and the Environment (RIVM) | Adam Meijer et al |
| EPI_ISL_1456652 | Dutch COVID-19 response team | National Institute for Public Health and the Environment (RIVM) | Adam Meijer et al |
| EPI_ISL_1521313 | Dutch COVID-19 response team | National Institute for Public Health and the Environment (RIVM) | Adam Meijer et al |
| EPI_ISL_1596573 | Dutch COVID-19 response team | National Institute for Public Health and the Environment (RIVM) | Adam Meijer et al |
| EPI_ISL_547478 | Dutch COVID-19 response team | National Institute for Public Health and the Environment (RIVM) | Adam Meijer et al |
| EPI_ISL_1035663 | Dutch COVID-19 response team | National Institute for Public Health and the Environment (RIVM) | Adam Meijer et al |
| EPI_ISL_1597267 | Dutch COVID-19 response team | National Institute for Public Health and the Environment (RIVM) | Adam Meijer et al |
| EPI_ISL_1090134 | Dutch COVID-19 response team | National Institute for Public Health and the Environment (RIVM) | Adam Meijer et al |
| EPI_ISL_905383 | Dutch COVID-19 response team | National Institute for Public Health and the Environment (RIVM) | Adam Meijer et al |
| EPI_ISL_1596936 | Dutch COVID-19 response team | National Institute for Public Health and the Environment (RIVM) | Adam Meijer et al |
| EPI_ISL_547499 | Dutch COVID-19 response team | National Institute for Public Health and the Environment (RIVM) | Adam Meijer et al |
| EPI_ISL_1014436 | Dutch COVID-19 response team | National Institute for Public Health and the Environment (RIVM) | Adam Meijer et al |
| EPI_ISL_1014435 | Dutch COVID-19 response team | National Institute for Public Health and the Environment (RIVM) | Adam Meijer et al |
| EPI_ISL_454750 | Dutch COVID-19 response team | National Institute for Public Health and the Environment (RIVM) | Adam Meijer et al |
| EPI_ISL_454751 | Dutch COVID-19 response team | National Institute for Public Health and the Environment (RIVM) | Adam Meijer et al |
| EPI_ISL_1014104 | Dutch COVID-19 response team | National Institute for Public Health and the Environment (RIVM) | Adam Meijer et al |
| EPI_ISL_1035443 | Dutch COVID-19 response team | National Institute for Public Health and the Environment (RIVM) | Adam Meijer et al |
| EPI_ISL_1035246 | Dutch COVID-19 response team | National Institute for Public Health and the Environment (RIVM) | Adam Meijer et al |
| EPI_ISL_1035244 | Dutch COVID-19 response team | National Institute for Public Health and the Environment (RIVM) | Adam Meijer et al |
| EPI_ISL_1035243 | Dutch COVID-19 response team | National Institute for Public Health and the Environment (RIVM) | Adam Meijer et al |
| EPI_ISL_1035242 | Dutch COVID-19 response team | National Institute for Public Health and the Environment (RIVM) | Adam Meijer et al |
| EPI_ISL_1035240 | Dutch COVID-19 response team | National Institute for Public Health and the Environment (RIVM) | Adam Meijer et al |
| EPI_ISL_1035239 | Dutch COVID-19 response team | National Institute for Public Health and the Environment (RIVM) | Adam Meijer et al |
| EPI_ISL_1089591 | Dutch COVID-19 response team | National Institute for Public Health and the Environment (RIVM) | Adam Meijer et al |
| EPI_ISL_1595963 | Dutch COVID-19 response team | National Institute for Public Health and the Environment (RIVM) | Adam Meijer et al |
| EPI_ISL_1595971 | Dutch COVID-19 response team | National Institute for Public Health and the Environment (RIVM) | Adam Meijer et al |
| EPI_ISL_1595972 | Dutch COVID-19 response team | National Institute for Public Health and the Environment (RIVM) | Adam Meijer et al |

| | | | |
|---|---|---|---|
| EPI_ISL_1595982 | Dutch COVID-19 response team | National Institute for Public Health and the Environment (RIVM) | Adam Meijer et al |
| EPI_ISL_1595983 | Dutch COVID-19 response team | National Institute for Public Health and the Environment (RIVM) | Adam Meijer et al |
| EPI_ISL_1596056 | Dutch COVID-19 response team | National Institute for Public Health and the Environment (RIVM) | Adam Meijer et al |
| EPI_ISL_1596057 | Dutch COVID-19 response team | National Institute for Public Health and the Environment (RIVM) | Adam Meijer et al |
| EPI_ISL_1596078 | Dutch COVID-19 response team | National Institute for Public Health and the Environment (RIVM) | Adam Meijer et al |
| EPI_ISL_1596090 | Dutch COVID-19 response team | National Institute for Public Health and the Environment (RIVM) | Adam Meijer et al |
| EPI_ISL_1522177 | Dutch COVID-19 response team | National Institute for Public Health and the Environment (RIVM) | Adam Meijer et al |
| EPI_ISL_1522184 | Dutch COVID-19 response team | National Institute for Public Health and the Environment (RIVM) | Adam Meijer et al |
| EPI_ISL_1522185 | Dutch COVID-19 response team | National Institute for Public Health and the Environment (RIVM) | Adam Meijer et al |
| EPI_ISL_1522186 | Dutch COVID-19 response team | National Institute for Public Health and the Environment (RIVM) | Adam Meijer et al |
| EPI_ISL_1522189 | Dutch COVID-19 response team | National Institute for Public Health and the Environment (RIVM) | Adam Meijer et al |
| EPI_ISL_1521324 | Dutch COVID-19 response team | National Institute for Public Health and the Environment (RIVM) | Adam Meijer et al |
| EPI_ISL_1522195 | Dutch COVID-19 response team | National Institute for Public Health and the Environment (RIVM) | Adam Meijer et al |
| EPI_ISL_1522200 | Dutch COVID-19 response team | National Institute for Public Health and the Environment (RIVM) | Adam Meijer et al |
| EPI_ISL_1522201 | Dutch COVID-19 response team | National Institute for Public Health and the Environment (RIVM) | Adam Meijer et al |
| EPI_ISL_1522203 | Dutch COVID-19 response team | National Institute for Public Health and the Environment (RIVM) | Adam Meijer et al |
| EPI_ISL_1522205 | Dutch COVID-19 response team | National Institute for Public Health and the Environment (RIVM) | Adam Meijer et al |
| EPI_ISL_1522208 | Dutch COVID-19 response team | National Institute for Public Health and the Environment (RIVM) | Adam Meijer et al |
| EPI_ISL_1522211 | Dutch COVID-19 response team | National Institute for Public Health and the Environment (RIVM) | Adam Meijer et al |
| EPI_ISL_1522213 | Dutch COVID-19 response team | National Institute for Public Health and the Environment (RIVM) | Adam Meijer et al |
| EPI_ISL_833494 | RS PMI Bogor, Indonesia | Biosafety Level-3 Laboratory, Indonesian Institute of Sciences (LIPI) | Ade Andriani et al |
| EPI_ISL_977171 | Microbiologia e Virologia | Istituto Zooprofilattico Sperimentale delle Venezie | Adelaide Milani et al |
| EPI_ISL_977172 | Microbiologia e Virologia | Istituto Zooprofilattico Sperimentale delle Venezie | Adelaide Milani et al |
| EPI_ISL_875556 | ULSS 2 Treviso | Istituto Zooprofilattico Sperimentale delle Venezie | Adelaide Milani et al |
| EPI_ISL_875553 | ULSS 2 Treviso | Istituto Zooprofilattico Sperimentale delle Venezie | Adelaide Milani et al |
| EPI_ISL_875554 | ULSS 2 Treviso | Istituto Zooprofilattico Sperimentale delle Venezie | Adelaide Milani et al |
| EPI_ISL_875555 | ULSS 2 Treviso | Istituto Zooprofilattico Sperimentale delle Venezie | Adelaide Milani et al |
| EPI_ISL_1260963 | ULSS 04 Veneto Orientale | Istituto Zooprofilattico Sperimentale delle Venezie | Adelaide Milani et al |
| EPI_ISL_424673 | Instituto de Diagnostico y Referencia Epidemiologicos | Instituto de Diagnostico y Referencia Epidemiologicos | Adnan Araiza Rodríguez et al |
| EPI_ISL_1018088 | Immunology, Noguchi Memorial Institute for Medical Research | Immunology, Noguchi Memorial Institute for Medical Research | Adu et al |

| | | | |
|---|---|---|---|
| EPI_ISL_1018089 | Immunology, Noguchi Memorial Institute for Medical Research | Immunology, Noguchi Memorial Institute for Medical Research | Adu et al |
| EPI_ISL_1018091 | Immunology, Noguchi Memorial Institute for Medical Research | Immunology, Noguchi Memorial Institute for Medical Research | Adu et al |
| EPI_ISL_1018087 | Immunology, Noguchi Memorial Institute for Medical Research | Immunology, Noguchi Memorial Institute for Medical Research | Adu et al |
| EPI_ISL_747304 | Division of Emerging Infectious Diseases, Bureau of Infectious Diseases Diagnosis Control, Korea Disease Control and Prevention Agency | Division of Emerging Infectious Diseases, Bureau of Infectious Diseases Diagnosis Control, Korea Disease Control and Prevention Agency | Ae Kyung Park et al |
| EPI_ISL_747307 | Division of Emerging Infectious Diseases, Bureau of Infectious Diseases Diagnosis Control, Korea Disease Control and Prevention Agency | Division of Emerging Infectious Diseases, Bureau of Infectious Diseases Diagnosis Control, Korea Disease Control and Prevention Agency | Ae Kyung Park et al |
| EPI_ISL_747318 | Division of Emerging Infectious Diseases, Bureau of Infectious Diseases Diagnosis Control, Korea Disease Control and Prevention Agency | Division of Emerging Infectious Diseases, Bureau of Infectious Diseases Diagnosis Control, Korea Disease Control and Prevention Agency | Ae Kyung Park et al |
| EPI_ISL_747333 | Division of Emerging Infectious Diseases, Bureau of Infectious Diseases Diagnosis Control, Korea Disease Control and Prevention Agency | Division of Emerging Infectious Diseases, Bureau of Infectious Diseases Diagnosis Control, Korea Disease Control and Prevention Agency | Ae Kyung Park et al |
| EPI_ISL_747337 | Division of Emerging Infectious Diseases, Bureau of Infectious Diseases Diagnosis Control, Korea Disease Control and Prevention Agency | Division of Emerging Infectious Diseases, Bureau of Infectious Diseases Diagnosis Control, Korea Disease Control and Prevention Agency | Ae Kyung Park et al |
| EPI_ISL_747342 | Division of Emerging Infectious Diseases, Bureau of Infectious Diseases Diagnosis Control, Korea Disease Control and Prevention Agency | Division of Emerging Infectious Diseases, Bureau of Infectious Diseases Diagnosis Control, Korea Disease Control and Prevention Agency | Ae Kyung Park et al |
| EPI_ISL_747362 | Division of Emerging Infectious Diseases, Bureau of Infectious Diseases Diagnosis Control, Korea Disease Control and Prevention Agency | Division of Emerging Infectious Diseases, Bureau of Infectious Diseases Diagnosis Control, Korea Disease Control and Prevention Agency | Ae Kyung Park et al |
| EPI_ISL_747449 | Division of Emerging Infectious Diseases, Bureau of Infectious Diseases Diagnosis Control, Korea Disease Control and Prevention Agency | Division of Emerging Infectious Diseases, Bureau of Infectious Diseases Diagnosis Control, Korea Disease Control and Prevention Agency | Ae Kyung Park et al |
| EPI_ISL_760102 | Division of Emerging Infectious Diseases, Bureau of Infectious Diseases Diagnosis Control, Korea Disease Control and Prevention Agency | Division of Emerging Infectious Diseases, Bureau of Infectious Diseases Diagnosis Control, Korea Disease Control and Prevention Agency | Ae Kyung Park et al |
| EPI_ISL_760103 | Division of Emerging Infectious Diseases, Bureau of Infectious Diseases Diagnosis Control, Korea Disease Control and Prevention Agency | Division of Emerging Infectious Diseases, Bureau of Infectious Diseases Diagnosis Control, Korea Disease Control and Prevention Agency | Ae Kyung Park et al |
| EPI_ISL_833249 | Division of Emerging Infectious Diseases, Bureau of Infectious Diseases Diagnosis Control, Korea Disease Control and Prevention Agency | Division of Emerging Infectious Diseases, Bureau of Infectious Diseases Diagnosis Control, Korea Disease Control and Prevention Agency | Ae Kyung Park et al |
| EPI_ISL_850198 | Division of Emerging Infectious Diseases, Bureau of Infectious Diseases Diagnosis Control, Korea Disease Control and Prevention Agency | Division of Emerging Infectious Diseases, Bureau of Infectious Diseases Diagnosis Control, Korea Disease Control and Prevention Agency | Ae Kyung Park et al |
| EPI_ISL_850200 | Division of Emerging Infectious Diseases, Bureau of Infectious Diseases Diagnosis Control, Korea Disease Control and Prevention Agency | Division of Emerging Infectious Diseases, Bureau of Infectious Diseases Diagnosis Control, Korea Disease Control and Prevention Agency | Ae Kyung Park et al |
| EPI_ISL_850211 | Division of Emerging Infectious Diseases, Bureau of Infectious Diseases Diagnosis Control, Korea Disease Control and Prevention Agency | Division of Emerging Infectious Diseases, Bureau of Infectious Diseases Diagnosis Control, Korea Disease Control and Prevention Agency | Ae Kyung Park et al |
| EPI_ISL_850215 | Division of Emerging Infectious Diseases, Bureau of Infectious Diseases Diagnosis Control, Korea Disease Control and Prevention Agency | Division of Emerging Infectious Diseases, Bureau of Infectious Diseases Diagnosis Control, Korea Disease Control and Prevention Agency | Ae Kyung Park et al |
| EPI_ISL_850391 | Division of Emerging Infectious Diseases, Bureau of Infectious Diseases Diagnosis Control, Korea Disease Control and Prevention Agency | Division of Emerging Infectious Diseases, Bureau of Infectious Diseases Diagnosis Control, Korea Disease Control and Prevention Agency | Ae Kyung Park et al |
| EPI_ISL_995230 | Division of Emerging Infectious Diseases, Bureau of Infectious Diseases Diagnosis Control, Korea Disease Control and Prevention Agency | Division of Emerging Infectious Diseases, Bureau of Infectious Diseases Diagnosis Control, Korea Disease Control and Prevention Agency | Ae Kyung Park et al |
| EPI_ISL_1063647 | Division of Emerging Infectious Diseases, Bureau of Infectious Diseases Diagnosis Control, Korea Disease Control and Prevention Agency | Division of Emerging Infectious Diseases, Bureau of Infectious Diseases Diagnosis Control, Korea Disease Control and Prevention Agency | Ae Kyung Park et al |
| EPI_ISL_1063656 | Division of Emerging Infectious Diseases, Bureau of Infectious Diseases Diagnosis Control, Korea Disease Control and Prevention Agency | Division of Emerging Infectious Diseases, Bureau of Infectious Diseases Diagnosis Control, Korea Disease Control and Prevention Agency | Ae Kyung Park et al |
| EPI_ISL_1063614 | Division of Emerging Infectious Diseases, Bureau of Infectious Diseases Diagnosis Control, Korea Disease Control and Prevention Agency | Division of Emerging Infectious Diseases, Bureau of Infectious Diseases Diagnosis Control, Korea Disease Control and Prevention Agency | Ae Kyung Park et al |

| | | | |
|---|---|---|---|
| EPI_ISL_1063615 | Division of Emerging Infectious Diseases, Bureau of Infectious Diseases Diagnosis Control, Korea Disease Control and Prevention Agency | Division of Emerging Infectious Diseases, Bureau of Infectious Diseases Diagnosis Control, Korea Disease Control and Prevention Agency | Ae Kyung Park et al |
| EPI_ISL_1063563 | Division of Emerging Infectious Diseases, Bureau of Infectious Diseases Diagnosis Control, Korea Disease Control and Prevention Agency | Division of Emerging Infectious Diseases, Bureau of Infectious Diseases Diagnosis Control, Korea Disease Control and Prevention Agency | Ae Kyung Park et al |
| EPI_ISL_1063589 | Division of Emerging Infectious Diseases, Bureau of Infectious Diseases Diagnosis Control, Korea Disease Control and Prevention Agency | Division of Emerging Infectious Diseases, Bureau of Infectious Diseases Diagnosis Control, Korea Disease Control and Prevention Agency | Ae Kyung Park et al |
| EPI_ISL_1063559 | Division of Emerging Infectious Diseases, Bureau of Infectious Diseases Diagnosis Control, Korea Disease Control and Prevention Agency | Division of Emerging Infectious Diseases, Bureau of Infectious Diseases Diagnosis Control, Korea Disease Control and Prevention Agency | Ae Kyung Park et al |
| EPI_ISL_1063554 | Division of Emerging Infectious Diseases, Bureau of Infectious Diseases Diagnosis Control, Korea Disease Control and Prevention Agency | Division of Emerging Infectious Diseases, Bureau of Infectious Diseases Diagnosis Control, Korea Disease Control and Prevention Agency | Ae Kyung Park et al |
| EPI_ISL_1063558 | Division of Emerging Infectious Diseases, Bureau of Infectious Diseases Diagnosis Control, Korea Disease Control and Prevention Agency | Division of Emerging Infectious Diseases, Bureau of Infectious Diseases Diagnosis Control, Korea Disease Control and Prevention Agency | Ae Kyung Park et al |
| EPI_ISL_1063556 | Division of Emerging Infectious Diseases, Bureau of Infectious Diseases Diagnosis Control, Korea Disease Control and Prevention Agency | Division of Emerging Infectious Diseases, Bureau of Infectious Diseases Diagnosis Control, Korea Disease Control and Prevention Agency | Ae Kyung Park et al |
| EPI_ISL_1063616 | Division of Emerging Infectious Diseases, Bureau of Infectious Diseases Diagnosis Control, Korea Disease Control and Prevention Agency | Division of Emerging Infectious Diseases, Bureau of Infectious Diseases Diagnosis Control, Korea Disease Control and Prevention Agency | Ae Kyung Park et al |
| EPI_ISL_1063678 | Division of Emerging Infectious Diseases, Bureau of Infectious Diseases Diagnosis Control, Korea Disease Control and Prevention Agency | Division of Emerging Infectious Diseases, Bureau of Infectious Diseases Diagnosis Control, Korea Disease Control and Prevention Agency | Ae Kyung Park et al |
| EPI_ISL_1063600 | Division of Emerging Infectious Diseases, Bureau of Infectious Diseases Diagnosis Control, Korea Disease Control and Prevention Agency | Division of Emerging Infectious Diseases, Bureau of Infectious Diseases Diagnosis Control, Korea Disease Control and Prevention Agency | Ae Kyung Park et al |
| EPI_ISL_1063702 | Division of Emerging Infectious Diseases, Bureau of Infectious Diseases Diagnosis Control, Korea Disease Control and Prevention Agency | Division of Emerging Infectious Diseases, Bureau of Infectious Diseases Diagnosis Control, Korea Disease Control and Prevention Agency | Ae Kyung Park et al |
| EPI_ISL_1063599 | Division of Emerging Infectious Diseases, Bureau of Infectious Diseases Diagnosis Control, Korea Disease Control and Prevention Agency | Division of Emerging Infectious Diseases, Bureau of Infectious Diseases Diagnosis Control, Korea Disease Control and Prevention Agency | Ae Kyung Park et al |
| EPI_ISL_1063601 | Division of Emerging Infectious Diseases, Bureau of Infectious Diseases Diagnosis Control, Korea Disease Control and Prevention Agency | Division of Emerging Infectious Diseases, Bureau of Infectious Diseases Diagnosis Control, Korea Disease Control and Prevention Agency | Ae Kyung Park et al |
| EPI_ISL_1063570 | Division of Emerging Infectious Diseases, Bureau of Infectious Diseases Diagnosis Control, Korea Disease Control and Prevention Agency | Division of Emerging Infectious Diseases, Bureau of Infectious Diseases Diagnosis Control, Korea Disease Control and Prevention Agency | Ae Kyung Park et al |
| EPI_ISL_1165001 | Division of Emerging Infectious Diseases, Bureau of Infectious Diseases Diagnosis Control, Korea Disease Control and Prevention Agency | Division of Emerging Infectious Diseases, Bureau of Infectious Diseases Diagnosis Control, Korea Disease Control and Prevention Agency | Ae Kyung Park et al |
| EPI_ISL_1165058 | Division of Emerging Infectious Diseases, Bureau of Infectious Diseases Diagnosis Control, Korea Disease Control and Prevention Agency | Division of Emerging Infectious Diseases, Bureau of Infectious Diseases Diagnosis Control, Korea Disease Control and Prevention Agency | Ae Kyung Park et al |
| EPI_ISL_1209415 | Division of Emerging Infectious Diseases, Bureau of Infectious Diseases Diagnosis Control, Korea Disease Control and Prevention Agency | Division of Emerging Infectious Diseases, Bureau of Infectious Diseases Diagnosis Control, Korea Disease Control and Prevention Agency | Ae Kyung Park et al |
| EPI_ISL_1490007 | Division of Emerging Infectious Diseases, Bureau of Infectious Diseases Diagnosis Control, Korea Disease Control and Prevention Agency | Division of Emerging Infectious Diseases, Bureau of Infectious Diseases Diagnosis Control, Korea Disease Control and Prevention Agency | Ae Kyung Park et al |
| EPI_ISL_1587522 | Division of Emerging Infectious Diseases, Bureau of Infectious Diseases Diagnosis Control, Korea Disease Control and Prevention Agency | Division of Emerging Infectious Diseases, Bureau of Infectious Diseases Diagnosis Control, Korea Disease Control and Prevention Agency | Ae Kyung Park et al |
| EPI_ISL_1587525 | Division of Emerging Infectious Diseases, Bureau of Infectious Diseases Diagnosis Control, Korea Disease Control and Prevention Agency | Division of Emerging Infectious Diseases, Bureau of Infectious Diseases Diagnosis Control, Korea Disease Control and Prevention Agency | Ae Kyung Park et al |
| EPI_ISL_1587547 | Division of Emerging Infectious Diseases, Bureau of Infectious Diseases Diagnosis Control, Korea Disease Control and Prevention Agency | Division of Emerging Infectious Diseases, Bureau of Infectious Diseases Diagnosis Control, Korea Disease Control and Prevention Agency | Ae Kyung Park et al |
| EPI_ISL_1587550 | Division of Emerging Infectious Diseases, Bureau of Infectious Diseases Diagnosis Control, Korea Disease Control and Prevention Agency | Division of Emerging Infectious Diseases, Bureau of Infectious Diseases Diagnosis Control, Korea Disease Control and Prevention Agency | Ae Kyung Park et al |

| | | | |
|---|---|---|---|
| EPI_ISL_1622483 | Division of Emerging Infectious Diseases, Bureau of Infectious Diseases Diagnosis Control, Korea Disease Control and Prevention Agency | Division of Emerging Infectious Diseases, Bureau of Infectious Diseases Diagnosis Control, Korea Disease Control and Prevention Agency | Ae Kyung Park et al |
| EPI_ISL_1622487 | Division of Emerging Infectious Diseases, Bureau of Infectious Diseases Diagnosis Control, Korea Disease Control and Prevention Agency | Division of Emerging Infectious Diseases, Bureau of Infectious Diseases Diagnosis Control, Korea Disease Control and Prevention Agency | Ae Kyung Park et al |
| EPI_ISL_1622497 | Division of Emerging Infectious Diseases, Bureau of Infectious Diseases Diagnosis Control, Korea Disease Control and Prevention Agency | Division of Emerging Infectious Diseases, Bureau of Infectious Diseases Diagnosis Control, Korea Disease Control and Prevention Agency | Ae Kyung Park et al |
| EPI_ISL_1622498 | Division of Emerging Infectious Diseases, Bureau of Infectious Diseases Diagnosis Control, Korea Disease Control and Prevention Agency | Division of Emerging Infectious Diseases, Bureau of Infectious Diseases Diagnosis Control, Korea Disease Control and Prevention Agency | Ae Kyung Park et al |
| EPI_ISL_1622499 | Division of Emerging Infectious Diseases, Bureau of Infectious Diseases Diagnosis Control, Korea Disease Control and Prevention Agency | Division of Emerging Infectious Diseases, Bureau of Infectious Diseases Diagnosis Control, Korea Disease Control and Prevention Agency | Ae Kyung Park et al |
| EPI_ISL_1647347 | Division of Emerging Infectious Diseases, Bureau of Infectious Diseases Diagnosis Control, Korea Disease Control and Prevention Agency | Division of Emerging Infectious Diseases, Bureau of Infectious Diseases Diagnosis Control, Korea Disease Control and Prevention Agency | Ae Kyung Park et al |
| EPI_ISL_1647350 | Division of Emerging Infectious Diseases, Bureau of Infectious Diseases Diagnosis Control, Korea Disease Control and Prevention Agency | Division of Emerging Infectious Diseases, Bureau of Infectious Diseases Diagnosis Control, Korea Disease Control and Prevention Agency | Ae Kyung Park et al |
| EPI_ISL_513008 | Pathogen Genomics Lab King Abdullah University of Science and Technology (KAUST) | Pathogen Genomics Lab King Abdullah University of Science and Technology(KAUST) | Afrah Alsomali et al |
| EPI_ISL_513021 | Pathogen Genomics Lab King Abdullah University of Science and Technology (KAUST) | Pathogen Genomics Lab King Abdullah University of Science and Technology(KAUST) | Afrah Alsomali et al |
| EPI_ISL_513192 | Pathogen Genomics Lab King Abdullah University of Science and Technology (KAUST) | Pathogen Genomics Lab King Abdullah University of Science and Technology(KAUST) | Afrah Alsomali et al |
| EPI_ISL_882637 | Azerbaijan National Hematology Center Division of Medical Genetics | Azerbaijan National Hematology Center Division of Medical Genetics | Aghayev Agha Rza et al |
| EPI_ISL_882639 | Azerbaijan National Hematology Center Division of Medical Genetics | Azerbaijan National Hematology Center Division of Medical Genetics | Aghayev Agha Rza et al |
| EPI_ISL_882641 | Azerbaijan National Hematology Center Division of Medical Genetics | Azerbaijan National Hematology Center Division of Medical Genetics | Aghayev Agha Rza et al |
| EPI_ISL_833130 | National Institute of Health Research and Development | National Institute of Health Research and Development | Agustiningsih et al |
| EPI_ISL_1517192 | Oman-National Influenza Center | Biotechnology & OMICs Laboratory | Ahmed Al Harrasi et al |
| EPI_ISL_1532284 | Oman-National Influenza Center | Biotechnology & OMICs Laboratory | Ahmed Al Harrasi et al |
| EPI_ISL_1532285 | Oman-National Influenza Center | Biotechnology & OMICs Laboratory | Ahmed Al Harrasi et al |
| EPI_ISL_1532301 | Oman-National Influenza Center | Biotechnology & OMICs Laboratory | Ahmed Al Harrasi et al |
| EPI_ISL_1009123 | School of Pharmacy | School of Pharmacy | Ahmed Kandeil et al |
| EPI_ISL_1009128 | School of Pharmacy | School of Pharmacy | Ahmed Kandeil et al |
| EPI_ISL_1009160 | School of Pharmacy | School of Pharmacy | Ahmed Kandeil et al |
| EPI_ISL_1532290 | Oman-National Influenza Center | Biotechnology & OMICs Laboratory | Aisha Al-Amri et al |
| EPI_ISL_1532291 | Oman-National Influenza Center | Biotechnology & OMICs Laboratory | Aisha Al-Amri et al |
| EPI_ISL_1532303 | Oman-National Influenza Center | Biotechnology & OMICs Laboratory | Aisha Al-Amri et al |
| EPI_ISL_510528 | Communicable Disease Laboratory, Public Health Directorate | Communicable Disease Laboratory, Public Health Directorate | Al Wasti et al |
| EPI_ISL_710483 | Department of Medical Laboratory Sciences, Arab American University | Department of Medical Laboratory Sciences, Arab American University | Al-Jawabreh et al |
| EPI_ISL_710484 | Department of Medical Laboratory Sciences, Arab American University | Department of Medical Laboratory Sciences, Arab American University | Al-Jawabreh et al |

| | | | |
|---|---|---|---|
| EPI_ISL_632900 | Communicable Disease Laboratory, Public Health Directorate | Communicable Disease Laboratory, Public Health Directorate | AlAbbas et al |
| EPI_ISL_733579 | Temporary Specimen Collection Centre | Hong Kong Department of Health | Alan K.L. Tsang et al |
| EPI_ISL_759892 | Temporary Specimen Collection Centre | Hong Kong Department of Health | Alan K.L. Tsang et al |
| EPI_ISL_955905 | Pamela Youde Nethersole Eastern Hospital | Hong Kong Department of Health | Alan K.L. Tsang et al |
| EPI_ISL_1630158 | Laboratório de Microbiologia Molecular - Universidade FEEVALE | Laboratório de Microbiologia Molecular - Universidade FEEVALE | Alana Witt Hansen et al |
| EPI_ISL_1629809 | Laboratório de Microbiologia Molecular - Universidade FEEVALE | Molecular Microbiology Laboratory | Alana Witt Hansen et al |
| EPI_ISL_458079 | Mitra Keluarga Hospital Kenjeran | Institute of Tropical Disease, Universitas Airlangga | Aldise M Nastri et al |
| EPI_ISL_529964 | RSUD Bangil Pasuruan | Institute of Tropical Disease, Universitas Airlangga | Aldise M Nastri et al |
| EPI_ISL_1335890 | Elisabeth Hospital Medan | Faculty of Medicine, Universitas Sumatera Utara; Institute of Tropical Disease, Universitas Airlangga | Aldise M Nastri et al |
| EPI_ISL_956404 | General Hospital - Strumica | Research Center for Genetic Engineering and Biotechnology Georgi D. Efremov" , Macedonian Academy of Sciences and Arts | Aleksandar J. Dimovski et al |
| EPI_ISL_943995 | General Hospital - Veles | Research Center for Genetic Engineering and Biotechnology Georgi D. Efremov" , Macedonian Academy of Sciences and Arts | Aleksandar J. Dimovski et al |
| EPI_ISL_944106 | General Hospital - Strumica | Research Center for Genetic Engineering and Biotechnology Georgi D. Efremov" , Macedonian Academy of Sciences and Arts | Aleksandar J. Dimovski et al |
| EPI_ISL_947678 | General Hospital - Tetovo | Research Center for Genetic Engineering and Biotechnology Georgi D. Efremov" , Macedonian Academy of Sciences and Arts | Aleksandar J. Dimovski et al |
| EPI_ISL_954611 | General Hospital - Struga | Research Center for Genetic Engineering and Biotechnology Georgi D. Efremov" , Macedonian Academy of Sciences and Arts | Aleksandar J. Dimovski et al |
| EPI_ISL_954745 | General Hospital - Ohrid | Research Center for Genetic Engineering and Biotechnology Georgi D. Efremov" , Macedonian Academy of Sciences and Arts | Aleksandar J. Dimovski et al |
| EPI_ISL_956383 | General Hospital - Prilep | Research Center for Genetic Engineering and Biotechnology Georgi D. Efremov" , Macedonian Academy of Sciences and Arts | Aleksandar J. Dimovski et al |
| EPI_ISL_1013583 | General Hospital - Prilep | Research Center for Genetic Engineering and Biotechnology Georgi D. Efremov" , Macedonian Academy of Sciences and Arts | Aleksandar J. Dimovski et al |
| EPI_ISL_1138412 | General Hospital - Shtip | Research Center for Genetic Engineering and Biotechnology Georgi D. Efremov" , Macedonian Academy of Sciences and Arts | Aleksandar J. Dimovski et al |
| EPI_ISL_1014939 | General Hospital - Struga | Research Center for Genetic Engineering and Biotechnology Georgi D. Efremov" , Macedonian Academy of Sciences and Arts | Aleksandar J. Dimovski et al |
| EPI_ISL_1138424 | General Hospital - Struga | Research Center for Genetic Engineering and Biotechnology Georgi D. Efremov" , Macedonian Academy of Sciences and Arts | Aleksandar J. Dimovski et al |
| EPI_ISL_1018109 | General Hospital - Stuga | Research Center for Genetic Engineering and Biotechnology Georgi D. Efremov" , Macedonian Academy of Sciences and Arts | Aleksandar J. Dimovski et al |
| EPI_ISL_1143746 | General Hospital - Kumanovo | Research Center for Genetic Engineering and Biotechnology Georgi D. Efremov" , Macedonian Academy of Sciences and Arts | Aleksandar J. Dimovski et al |
| EPI_ISL_942961 | General Hospital - Bitola | Research Center for Genetic Engineering and Biotechnology Georgi D. Efremov" , Macedonian Academy of Sciences and Arts | Aleksandar J.Dimovski et al |
| EPI_ISL_1499088 | Institute of Microbiology and Immunology, Faculty of Medicine, University of Ljubljana | Institute of Microbiology and Immunology, Faculty of Medicine, University of Ljubljana | Alen Suljič et al |

| | | | |
|---|---|---|---|
| EPI_ISL_1499099 | Institute of Microbiology and Immunology, Faculty of Medicine, University of Ljubljana | Institute of Microbiology and Immunology, Faculty of Medicine, University of Ljubljana | Alen Suljič et al |
| EPI_ISL_1499104 | Institute of Microbiology and Immunology, Faculty of Medicine, University of Ljubljana | Institute of Microbiology and Immunology, Faculty of Medicine, University of Ljubljana | Alen Suljič et al |
| EPI_ISL_1098776 | Institute of Microbiology and Immunology, Faculty of Medicine, University of Ljubljana | Institute of Microbiology and Immunology, Faculty of Medicine, University of Ljubljana | Alen Suljič et al |
| EPI_ISL_1098778 | Institute of Microbiology and Immunology, Faculty of Medicine, University of Ljubljana | Institute of Microbiology and Immunology, Faculty of Medicine, University of Ljubljana | Alen Suljič et al |
| EPI_ISL_1668553 | Institute of Microbiology and Immunology, Faculty of Medicine, University of Ljubljana | Institute of Microbiology and Immunology, Faculty of Medicine, University of Ljubljana | Alen Suljič et al |
| EPI_ISL_1098789 | Institute of Microbiology and Immunology, Faculty of Medicine, University of Ljubljana | Institute of Microbiology and Immunology, Faculty of Medicine, University of Ljubljana | Alen Suljič et al |
| EPI_ISL_1098795 | Institute of Microbiology and Immunology, Faculty of Medicine, University of Ljubljana | Institute of Microbiology and Immunology, Faculty of Medicine, University of Ljubljana | Alen Suljič et al |
| EPI_ISL_1668820 | Institute of Microbiology and Immunology, Faculty of Medicine, University of Ljubljana | Institute of Microbiology and Immunology, Faculty of Medicine, University of Ljubljana | Alen Suljič et al |
| EPI_ISL_1098796 | Institute of Microbiology and Immunology, Faculty of Medicine, University of Ljubljana | Institute of Microbiology and Immunology, Faculty of Medicine, University of Ljubljana | Alen Suljič et al |
| EPI_ISL_1181825 | Institute of Microbiology and Immunology, Faculty of Medicine, University of Ljubljana | Institute of Microbiology and Immunology, Faculty of Medicine, University of Ljubljana | Alen Suljič et al |
| EPI_ISL_1181830 | Institute of Microbiology and Immunology, Faculty of Medicine, University of Ljubljana | Institute of Microbiology and Immunology, Faculty of Medicine, University of Ljubljana | Alen Suljič et al |
| EPI_ISL_1510994 | Institute of Microbiology and Immunology, Faculty of Medicine, University of Ljubljana | Institute of Microbiology and Immunology, Faculty of Medicine, University of Ljubljana | Alen Suljič et al |
| EPI_ISL_1510968 | Institute of Microbiology and Immunology, Faculty of Medicine, University of Ljubljana | Institute of Microbiology and Immunology, Faculty of Medicine, University of Ljubljana | Alen Suljič et al |
| EPI_ISL_1510958 | Institute of Microbiology and Immunology, Faculty of Medicine, University of Ljubljana | Institute of Microbiology and Immunology, Faculty of Medicine, University of Ljubljana | Alen Suljič et al |
| EPI_ISL_1511108 | Institute of Microbiology and Immunology, Faculty of Medicine, University of Ljubljana | Institute of Microbiology and Immunology, Faculty of Medicine, University of Ljubljana | Alen Suljič et al |
| EPI_ISL_1511077 | Institute of Microbiology and Immunology, Faculty of Medicine, University of Ljubljana | Institute of Microbiology and Immunology, Faculty of Medicine, University of Ljubljana | Alen Suljič et al |
| EPI_ISL_1511003 | Institute of Microbiology and Immunology, Faculty of Medicine, University of Ljubljana | Institute of Microbiology and Immunology, Faculty of Medicine, University of Ljubljana | Alen Suljič et al |
| EPI_ISL_1511004 | Institute of Microbiology and Immunology, Faculty of Medicine, University of Ljubljana | Institute of Microbiology and Immunology, Faculty of Medicine, University of Ljubljana | Alen Suljič et al |
| EPI_ISL_1511097 | Institute of Microbiology and Immunology, Faculty of Medicine, University of Ljubljana | Institute of Microbiology and Immunology, Faculty of Medicine, University of Ljubljana | Alen Suljič et al |
| EPI_ISL_1511040 | Institute of Microbiology and Immunology, Faculty of Medicine, University of Ljubljana | Institute of Microbiology and Immunology, Faculty of Medicine, University of Ljubljana | Alen Suljič et al |
| EPI_ISL_1511093 | Institute of Microbiology and Immunology, Faculty of Medicine, University of Ljubljana | Institute of Microbiology and Immunology, Faculty of Medicine, University of Ljubljana | Alen Suljič et al |
| EPI_ISL_1511102 | Institute of Microbiology and Immunology, Faculty of Medicine, University of Ljubljana | Institute of Microbiology and Immunology, Faculty of Medicine, University of Ljubljana | Alen Suljič et al |

| | | | |
|---|---|---|---|
| EPI_ISL_1511013 | Institute of Microbiology and Immunology, Faculty of Medicine, University of Ljubljana | Institute of Microbiology and Immunology, Faculty of Medicine, University of Ljubljana | Alen Suljič et al |
| EPI_ISL_1498931 | Institute of Microbiology and Immunology, Faculty of Medicine, University of Ljubljana | Institute of Microbiology and Immunology, Faculty of Medicine, University of Ljubljana | Alen Suljič et al |
| EPI_ISL_1498951 | Institute of Microbiology and Immunology, Faculty of Medicine, University of Ljubljana | Institute of Microbiology and Immunology, Faculty of Medicine, University of Ljubljana | Alen Suljič et al |
| EPI_ISL_1498956 | Institute of Microbiology and Immunology, Faculty of Medicine, University of Ljubljana | Institute of Microbiology and Immunology, Faculty of Medicine, University of Ljubljana | Alen Suljič et al |
| EPI_ISL_1181860 | Institute of Microbiology and Immunology, Faculty of Medicine, University of Ljubljana | Institute of Microbiology and Immunology, Faculty of Medicine, University of Ljubljana | Alen Suljič et al |
| EPI_ISL_1668569 | Institute of Microbiology and Immunology, Faculty of Medicine, University of Ljubljana | Institute of Microbiology and Immunology, Faculty of Medicine, University of Ljubljana | Alen Suljič et al |
| EPI_ISL_1213399 | LAFEM/UESC | Bioinformatics Laboratory / LNCC | Alessandra P Lamarca et al |
| EPI_ISL_1213226 | LAFEM/UESC | Bioinformatics Laboratory / LNCC | Alessandra P Lamarca et al |
| EPI_ISL_1213294 | LAFEM/UESC | Bioinformatics Laboratory / LNCC | Alessandra P Lamarca et al |
| EPI_ISL_1213401 | LAFEM/UESC | Bioinformatics Laboratory / LNCC | Alessandra P Lamarca et al |
| EPI_ISL_1213384 | LAFEM/UESC | Bioinformatics Laboratory / LNCC | Alessandra P Lamarca et al |
| EPI_ISL_1213342 | LAFEM/UESC | Bioinformatics Laboratory / LNCC | Alessandra P Lamarca et al |
| EPI_ISL_1213232 | LAFEM/UESC | Bioinformatics Laboratory / LNCC | Alessandra P Lamarca et al |
| EPI_ISL_1213298 | LAFEM/UESC | Bioinformatics Laboratory / LNCC | Alessandra P Lamarca et al |
| EPI_ISL_1213390 | LAFEM/UESC | Bioinformatics Laboratory / LNCC | Alessandra P Lamarca et al |
| EPI_ISL_1213170 | LAFEM/UESC | Bioinformatics Laboratory / LNCC | Alessandra P Lamarca et al |
| EPI_ISL_1213302 | LAFEM/UESC | Bioinformatics Laboratory / LNCC | Alessandra P Lamarca et al |
| EPI_ISL_1213303 | LAFEM/UESC | Bioinformatics Laboratory / LNCC | Alessandra P Lamarca et al |
| EPI_ISL_1213220 | LAFEM/UESC | Bioinformatics Laboratory / LNCC | Alessandra P Lamarca et al |
| EPI_ISL_1213237 | LAFEM/UESC | Bioinformatics Laboratory / LNCC | Alessandra P Lamarca et al |
| EPI_ISL_1213413 | LAFEM/UESC | Bioinformatics Laboratory / LNCC | Alessandra P Lamarca et al |
| EPI_ISL_1213269 | LAFEM/UESC | Bioinformatics Laboratory / LNCC | Alessandra P Lamarca et al |
| EPI_ISL_1213241 | LAFEM/UESC | Bioinformatics Laboratory / LNCC | Alessandra P Lamarca et al |
| EPI_ISL_1213402 | LAFEM/UESC | Bioinformatics Laboratory / LNCC | Alessandra P Lamarca et al |
| EPI_ISL_1213328 | LAFEM/UESC | Bioinformatics Laboratory / LNCC | Alessandra P Lamarca et al |
| EPI_ISL_1213441 | LBM/UFPB | Bioinformatics Laboratory / LNCC | Alessandra P Lamarca et al |
| EPI_ISL_1213433 | LBM/UFPB | Bioinformatics Laboratory / LNCC | Alessandra P Lamarca et al |
| EPI_ISL_1213427 | LBM/UFPB | Bioinformatics Laboratory / LNCC | Alessandra P Lamarca et al |

| | | | |
|---|---|---|---|
| EPI_ISL_1213404 | LBM/UFPB | Bioinformatics Laboratory / LNCC | Alessandra P Lamarca et al |
| EPI_ISL_1213367 | LBM/UFPB | Bioinformatics Laboratory / LNCC | Alessandra P Lamarca et al |
| EPI_ISL_1213392 | LBM/UFPB | Bioinformatics Laboratory / LNCC | Alessandra P Lamarca et al |
| EPI_ISL_1213153 | LBM/UFPB | Bioinformatics Laboratory / LNCC | Alessandra P Lamarca et al |
| EPI_ISL_1213213 | LBM/UFPB | Bioinformatics Laboratory / LNCC | Alessandra P Lamarca et al |
| EPI_ISL_1213161 | LBM/UFPB | Bioinformatics Laboratory / LNCC | Alessandra P Lamarca et al |
| EPI_ISL_1213438 | LBM/UFPB | Bioinformatics Laboratory / LNCC | Alessandra P Lamarca et al |
| EPI_ISL_1213454 | LBM/UFPB | Bioinformatics Laboratory / LNCC | Alessandra P Lamarca et al |
| EPI_ISL_1213461 | LBM/UFPB | Bioinformatics Laboratory / LNCC | Alessandra P Lamarca et al |
| EPI_ISL_1213443 | LBM/UFPB | Bioinformatics Laboratory / LNCC | Alessandra P Lamarca et al |
| EPI_ISL_1213201 | LBM/UFPB | Bioinformatics Laboratory / LNCC | Alessandra P Lamarca et al |
| EPI_ISL_1213429 | LBM/UFPB | Bioinformatics Laboratory / LNCC | Alessandra P Lamarca et al |
| EPI_ISL_1213453 | LBM/UFPB | Bioinformatics Laboratory / LNCC | Alessandra P Lamarca et al |
| EPI_ISL_1213439 | LBM/UFPB | Bioinformatics Laboratory / LNCC | Alessandra P Lamarca et al |
| EPI_ISL_1213431 | LBM/UFPB | Bioinformatics Laboratory / LNCC | Alessandra P Lamarca et al |
| EPI_ISL_1213456 | LBM/UFPB | Bioinformatics Laboratory / LNCC | Alessandra P Lamarca et al |
| EPI_ISL_1213369 | LBM/UFPB | Bioinformatics Laboratory / LNCC | Alessandra P Lamarca et al |
| EPI_ISL_1213458 | LBM/UFPB | Bioinformatics Laboratory / LNCC | Alessandra P Lamarca et al |
| EPI_ISL_1213459 | LBM/UFPB | Bioinformatics Laboratory / LNCC | Alessandra P Lamarca et al |
| EPI_ISL_1213444 | LBM/UFPB | Bioinformatics Laboratory / LNCC | Alessandra P Lamarca et al |
| EPI_ISL_1213344 | LBM/UFPB | Bioinformatics Laboratory / LNCC | Alessandra P Lamarca et al |
| EPI_ISL_1213436 | LBM/UFPB | Bioinformatics Laboratory / LNCC | Alessandra P Lamarca et al |
| EPI_ISL_1213336 | LBM/UFPB | Bioinformatics Laboratory / LNCC | Alessandra P Lamarca et al |
| EPI_ISL_1213156 | LBM/UFPB | Bioinformatics Laboratory / LNCC | Alessandra P Lamarca et al |
| EPI_ISL_1213449 | LBM/UFPB | Bioinformatics Laboratory / LNCC | Alessandra P Lamarca et al |
| EPI_ISL_1213406 | LBM/UFPB | Bioinformatics Laboratory / LNCC | Alessandra P Lamarca et al |
| EPI_ISL_1213370 | LBM/UFPB | Bioinformatics Laboratory / LNCC | Alessandra P Lamarca et al |
| EPI_ISL_1213451 | LBM/UFPB | Bioinformatics Laboratory / LNCC | Alessandra P Lamarca et al |
| EPI_ISL_1213372 | Laboratório HLA/UERJ | Bioinformatics Laboratory / LNCC | Alessandra P Lamarca et al |
| EPI_ISL_1213339 | Laboratório HLA/UERJ | Bioinformatics Laboratory / LNCC | Alessandra P Lamarca et al |

| | | | |
|---|---|---|---|
| EPI_ISL_1213248 | Laboratório HLA/UERJ | Bioinformatics Laboratory / LNCC | Alessandra P Lamarca et al |
| EPI_ISL_1213422 | Laboratório HLA/UERJ | Bioinformatics Laboratory / LNCC | Alessandra P Lamarca et al |
| EPI_ISL_1213424 | Laboratório HLA/UERJ | Bioinformatics Laboratory / LNCC | Alessandra P Lamarca et al |
| EPI_ISL_1213309 | Laboratório HLA/UERJ | Bioinformatics Laboratory / LNCC | Alessandra P Lamarca et al |
| EPI_ISL_1213425 | Laboratório HLA/UERJ | Bioinformatics Laboratory / LNCC | Alessandra P Lamarca et al |
| EPI_ISL_1213171 | Laboratório HLA/UERJ | Bioinformatics Laboratory / LNCC | Alessandra P Lamarca et al |
| EPI_ISL_1213378 | Laboratório HLA/UERJ | Bioinformatics Laboratory / LNCC | Alessandra P Lamarca et al |
| EPI_ISL_1213270 | Laboratório HLA/UERJ | Bioinformatics Laboratory / LNCC | Alessandra P Lamarca et al |
| EPI_ISL_1213151 | Laboratório HLA/UERJ | Bioinformatics Laboratory / LNCC | Alessandra P Lamarca et al |
| EPI_ISL_1213408 | Laboratório HLA/UERJ | Bioinformatics Laboratory / LNCC | Alessandra P Lamarca et al |
| EPI_ISL_1213381 | Laboratório HLA/UERJ | Bioinformatics Laboratory / LNCC | Alessandra P Lamarca et al |
| EPI_ISL_1213393 | Laboratório HLA/UERJ | Bioinformatics Laboratory / LNCC | Alessandra P Lamarca et al |
| EPI_ISL_1213382 | Laboratório HLA/UERJ | Bioinformatics Laboratory / LNCC | Alessandra P Lamarca et al |
| EPI_ISL_1213272 | Laboratório HLA/UERJ | Bioinformatics Laboratory / LNCC | Alessandra P Lamarca et al |
| EPI_ISL_1213249 | Laboratório HLA/UERJ | Bioinformatics Laboratory / LNCC | Alessandra P Lamarca et al |
| EPI_ISL_1213149 | Laboratório HLA/UERJ | Bioinformatics Laboratory / LNCC | Alessandra P Lamarca et al |
| EPI_ISL_1213148 | Laboratório HLA/UERJ | Bioinformatics Laboratory / LNCC | Alessandra P Lamarca et al |
| EPI_ISL_1213155 | Laboratório HLA/UERJ | Bioinformatics Laboratory / LNCC | Alessandra P Lamarca et al |
| EPI_ISL_1213158 | Laboratório HLA/UERJ | Bioinformatics Laboratory / LNCC | Alessandra P Lamarca et al |
| EPI_ISL_1213315 | IMT-UFRN/RN | Bioinformatics Laboratory / LNCC | Alessandra P Lamarca et al |
| EPI_ISL_1213350 | IMT-UFRN/RN | Bioinformatics Laboratory / LNCC | Alessandra P Lamarca et al |
| EPI_ISL_1213275 | IMT-UFRN/RN | Bioinformatics Laboratory / LNCC | Alessandra P Lamarca et al |
| EPI_ISL_1213277 | IMT-UFRN/RN | Bioinformatics Laboratory / LNCC | Alessandra P Lamarca et al |
| EPI_ISL_1213173 | IMT-UFRN/RN | Bioinformatics Laboratory / LNCC | Alessandra P Lamarca et al |
| EPI_ISL_1213175 | IMT-UFRN/RN | Bioinformatics Laboratory / LNCC | Alessandra P Lamarca et al |
| EPI_ISL_1213177 | IMT-UFRN/RN | Bioinformatics Laboratory / LNCC | Alessandra P Lamarca et al |
| EPI_ISL_1213317 | IMT-UFRN/RN | Bioinformatics Laboratory / LNCC | Alessandra P Lamarca et al |
| EPI_ISL_1213178 | IMT-UFRN/RN | Bioinformatics Laboratory / LNCC | Alessandra P Lamarca et al |
| EPI_ISL_1213185 | IMT-UFRN/RN | Bioinformatics Laboratory / LNCC | Alessandra P Lamarca et al |
| EPI_ISL_1213357 | IMT-UFRN/RN | Bioinformatics Laboratory / LNCC | Alessandra P Lamarca et al |

| | | | |
|---|---|---|---|
| EPI_ISL_1213180 | IMT-UFRN/RN | Bioinformatics Laboratory / LNCC | Alessandra P Lamarca et al |
| EPI_ISL_1213329 | IMT-UFRN/RN | Bioinformatics Laboratory / LNCC | Alessandra P Lamarca et al |
| EPI_ISL_1213279 | IMT-UFRN/RN | Bioinformatics Laboratory / LNCC | Alessandra P Lamarca et al |
| EPI_ISL_1213281 | IMT-UFRN/RN | Bioinformatics Laboratory / LNCC | Alessandra P Lamarca et al |
| EPI_ISL_1213282 | IMT-UFRN/RN | Bioinformatics Laboratory / LNCC | Alessandra P Lamarca et al |
| EPI_ISL_1213284 | IMT-UFRN/RN | Bioinformatics Laboratory / LNCC | Alessandra P Lamarca et al |
| EPI_ISL_1213286 | IMT-UFRN/RN | Bioinformatics Laboratory / LNCC | Alessandra P Lamarca et al |
| EPI_ISL_1213322 | IMT-UFRN/RN | Bioinformatics Laboratory / LNCC | Alessandra P Lamarca et al |
| EPI_ISL_1213288 | IMT-UFRN/RN | Bioinformatics Laboratory / LNCC | Alessandra P Lamarca et al |
| EPI_ISL_1213182 | IMT-UFRN/RN | Bioinformatics Laboratory / LNCC | Alessandra P Lamarca et al |
| EPI_ISL_1213183 | IMT-UFRN/RN | Bioinformatics Laboratory / LNCC | Alessandra P Lamarca et al |
| EPI_ISL_1213289 | IMT-UFRN/RN | Bioinformatics Laboratory / LNCC | Alessandra P Lamarca et al |
| EPI_ISL_1213364 | IMT-UFRN/RN | Bioinformatics Laboratory / LNCC | Alessandra P Lamarca et al |
| EPI_ISL_1213291 | IMT-UFRN/RN | Bioinformatics Laboratory / LNCC | Alessandra P Lamarca et al |
| EPI_ISL_1213293 | IMT-UFRN/RN | Bioinformatics Laboratory / LNCC | Alessandra P Lamarca et al |
| EPI_ISL_1213199 | IMT-UFRN/RN | Bioinformatics Laboratory / LNCC | Alessandra P Lamarca et al |
| EPI_ISL_1620638 | Hospital Municipal Dr. Ignacio Proença de Gouvea | Instituto Butantan / Mendelics | Alexander Roberto Precioso et al |
| EPI_ISL_454497 | RSE National Center for Biotechnology | RSE National Center for Biotechnology | Alexandr Shevtsov et al |
| EPI_ISL_454507 | RSE National Center for Biotechnology | RSE National Center for Biotechnology | Alexandr Shevtsov et al |
| EPI_ISL_454508 | RSE National Center for Biotechnology | RSE National Center for Biotechnology | Alexandr Shevtsov et al |
| EPI_ISL_454511 | RSE National Center for Biotechnology | RSE National Center for Biotechnology | Alexandr Shevtsov et al |
| EPI_ISL_454512 | RSE National Center for Biotechnology | RSE National Center for Biotechnology | Alexandr Shevtsov et al |
| EPI_ISL_454520 | RSE National Center for Biotechnology | RSE National Center for Biotechnology | Alexandr Shevtsov et al |
| EPI_ISL_435045 | Laboratory of Applied Genetics | RSE National Center for Biotechnology | Alexandr Shevtsov et al |
| EPI_ISL_435046 | Laboratory of Applied Genetics | RSE National Center for Biotechnology | Alexandr Shevtsov et al |
| EPI_ISL_435047 | Laboratory of Applied Genetics | RSE National Center for Biotechnology | Alexandr Shevtsov et al |
| EPI_ISL_470568 | Hermes Pardini | Bioinformatics Laboratory / LNCC | Alexandra Gerber et al |
| EPI_ISL_470570 | Hermes Pardini | Bioinformatics Laboratory / LNCC | Alexandra Gerber et al |
| EPI_ISL_470571 | Hermes Pardini | Bioinformatics Laboratory / LNCC | Alexandra Gerber et al |
| EPI_ISL_470572 | Hermes Pardini | Bioinformatics Laboratory / LNCC | Alexandra Gerber et al |

| | | | |
|---|---|---|---|
| EPI_ISL_470573 | Hermes Pardini | Bioinformatics Laboratory / LNCC | Alexandra Gerber et al |
| EPI_ISL_470574 | Hermes Pardini | Bioinformatics Laboratory / LNCC | Alexandra Gerber et al |
| EPI_ISL_470575 | Hermes Pardini | Bioinformatics Laboratory / LNCC | Alexandra Gerber et al |
| EPI_ISL_470576 | Hermes Pardini | Bioinformatics Laboratory / LNCC | Alexandra Gerber et al |
| EPI_ISL_470577 | Hermes Pardini | Bioinformatics Laboratory / LNCC | Alexandra Gerber et al |
| EPI_ISL_470580 | Hermes Pardini | Bioinformatics Laboratory / LNCC | Alexandra Gerber et al |
| EPI_ISL_470582 | Hermes Pardini | Bioinformatics Laboratory / LNCC | Alexandra Gerber et al |
| EPI_ISL_470583 | Hermes Pardini | Bioinformatics Laboratory / LNCC | Alexandra Gerber et al |
| EPI_ISL_470584 | Hermes Pardini | Bioinformatics Laboratory / LNCC | Alexandra Gerber et al |
| EPI_ISL_470585 | Hermes Pardini | Bioinformatics Laboratory / LNCC | Alexandra Gerber et al |
| EPI_ISL_470586 | Hermes Pardini | Bioinformatics Laboratory / LNCC | Alexandra Gerber et al |
| EPI_ISL_470587 | Hermes Pardini | Bioinformatics Laboratory / LNCC | Alexandra Gerber et al |
| EPI_ISL_470588 | Hermes Pardini | Bioinformatics Laboratory / LNCC | Alexandra Gerber et al |
| EPI_ISL_470589 | Simile | Bioinformatics Laboratory / LNCC | Alexandra Gerber et al |
| EPI_ISL_470590 | Simile | Bioinformatics Laboratory / LNCC | Alexandra Gerber et al |
| EPI_ISL_470591 | Simile | Bioinformatics Laboratory / LNCC | Alexandra Gerber et al |
| EPI_ISL_470592 | Simile | Bioinformatics Laboratory / LNCC | Alexandra Gerber et al |
| EPI_ISL_470593 | Simile | Bioinformatics Laboratory / LNCC | Alexandra Gerber et al |
| EPI_ISL_470594 | Simile | Bioinformatics Laboratory / LNCC | Alexandra Gerber et al |
| EPI_ISL_470595 | Simile | Bioinformatics Laboratory / LNCC | Alexandra Gerber et al |
| EPI_ISL_470596 | Simile | Bioinformatics Laboratory / LNCC | Alexandra Gerber et al |
| EPI_ISL_470599 | Hermes Pardini | Bioinformatics Laboratory / LNCC | Alexandra Gerber et al |
| EPI_ISL_470600 | Hermes Pardini | Bioinformatics Laboratory / LNCC | Alexandra Gerber et al |
| EPI_ISL_470602 | Hermes Pardini | Bioinformatics Laboratory / LNCC | Alexandra Gerber et al |
| EPI_ISL_470604 | Hermes Pardini | Bioinformatics Laboratory / LNCC | Alexandra Gerber et al |
| EPI_ISL_470605 | Hermes Pardini | Bioinformatics Laboratory / LNCC | Alexandra Gerber et al |
| EPI_ISL_470612 | Hermes Pardini | Bioinformatics Laboratory / LNCC | Alexandra Gerber et al |
| EPI_ISL_470613 | Hermes Pardini | Bioinformatics Laboratory / LNCC | Alexandra Gerber et al |
| EPI_ISL_470614 | Hermes Pardini | Bioinformatics Laboratory / LNCC | Alexandra Gerber et al |
| EPI_ISL_470615 | Laboratorio de Virologia Molecular / UFRJ | Bioinformatics Laboratory / LNCC | Alexandra Gerber et al |

| | | | |
|---|---|---|---|
| EPI_ISL_470617 | Laboratorio de Virologia Molecular / UFRJ | Bioinformatics Laboratory / LNCC | Alexandra Gerber et al |
| EPI_ISL_470618 | Laboratorio de Virologia Molecular / UFRJ | Bioinformatics Laboratory / LNCC | Alexandra Gerber et al |
| EPI_ISL_470620 | Laboratorio de Virologia Molecular / UFRJ | Bioinformatics Laboratory / LNCC | Alexandra Gerber et al |
| EPI_ISL_470621 | Laboratorio de Virologia Molecular / UFRJ | Bioinformatics Laboratory / LNCC | Alexandra Gerber et al |
| EPI_ISL_470623 | Laboratorio de Virologia Molecular / UFRJ | Bioinformatics Laboratory / LNCC | Alexandra Gerber et al |
| EPI_ISL_470638 | Laboratorio de Virologia Molecular / UFRJ | Bioinformatics Laboratory / LNCC | Alexandra Gerber et al |
| EPI_ISL_470651 | Hermes Pardini | Bioinformatics Laboratory / LNCC | Alexandra Gerber et al |
| EPI_ISL_470652 | Hermes Pardini | Bioinformatics Laboratory / LNCC | Alexandra Gerber et al |
| EPI_ISL_470653 | Hermes Pardini | Bioinformatics Laboratory / LNCC | Alexandra Gerber et al |
| EPI_ISL_470654 | Hermes Pardini | Bioinformatics Laboratory / LNCC | Alexandra Gerber et al |
| EPI_ISL_470655 | Hermes Pardini | Bioinformatics Laboratory / LNCC | Alexandra Gerber et al |
| EPI_ISL_419655 | Center for Virology, Medical University of Vienna | Bergthaler laboratory, CeMM Research Center for Molecular Medicine of the Austrian Academy of Sciences | Alexandra Popa et al |
| EPI_ISL_419657 | Center for Virology, Medical University of Vienna | Bergthaler laboratory, CeMM Research Center for Molecular Medicine of the Austrian Academy of Sciences | Alexandra Popa et al |
| EPI_ISL_583563 | Center for Virology, Medical University of Vienna | Bergthaler laboratory, CeMM Research Center for Molecular Medicine of the Austrian Academy of Sciences | Alexandra Popa et al |
| EPI_ISL_437993 | Center for Virology, Medical University of Vienna | Bergthaler laboratory, CeMM Research Center for Molecular Medicine of the Austrian Academy of Sciences | Alexandra Popa et al |
| EPI_ISL_437995 | Center for Virology, Medical University of Vienna | Bergthaler laboratory, CeMM Research Center for Molecular Medicine of the Austrian Academy of Sciences | Alexandra Popa et al |
| EPI_ISL_475789 | Center for Virology, Medical University of Vienna | Bergthaler laboratory, CeMM Research Center for Molecular Medicine of the Austrian Academy of Sciences | Alexandra Popa et al |
| EPI_ISL_475790 | Center for Virology, Medical University of Vienna | Bergthaler laboratory, CeMM Research Center for Molecular Medicine of the Austrian Academy of Sciences | Alexandra Popa et al |
| EPI_ISL_475796 | Center for Virology, Medical University of Vienna | Bergthaler laboratory, CeMM Research Center for Molecular Medicine of the Austrian Academy of Sciences | Alexandra Popa et al |
| EPI_ISL_475805 | Center for Virology, Medical University of Vienna | Bergthaler laboratory, CeMM Research Center for Molecular Medicine of the Austrian Academy of Sciences | Alexandra Popa et al |
| EPI_ISL_475832 | Austrian Agency for Health and Food Safety (AGES) | Bergthaler laboratory, CeMM Research Center for Molecular Medicine of the Austrian Academy of Sciences | Alexandra Popa et al |
| EPI_ISL_583573 | Austrian Agency for Health and Food Safety (AGES) | Bergthaler laboratory, CeMM Research Center for Molecular Medicine of the Austrian Academy of Sciences | Alexandra Popa et al |
| EPI_ISL_475836 | Austrian Agency for Health and Food Safety (AGES) | Bergthaler laboratory, CeMM Research Center for Molecular Medicine of the Austrian Academy of Sciences | Alexandra Popa et al |
| EPI_ISL_583575 | Austrian Agency for Health and Food Safety (AGES) | Bergthaler laboratory, CeMM Research Center for Molecular Medicine of the Austrian Academy of Sciences | Alexandra Popa et al |
| EPI_ISL_475896 | Zentralinstitut für medizinische und chemische Labordiagnostik, Universitätskliniken Insbruck | Bergthaler laboratory, CeMM Research Center for Molecular Medicine of the Austrian Academy of Sciences | Alexandra Popa et al |

| | | | |
|---|---|---|---|
| EPI_ISL_583604 | Institut für Virologie am Department für Hygiene, Mikrobiologie und Public Health | Bergthaler laboratory, CeMM Research Center for Molecular Medicine of the Austrian Academy of Sciences | Alexandra Popa et al |
| EPI_ISL_583632 | Austrian Agency for Health and Food Safety (AGES) | Bergthaler laboratory, CeMM Research Center for Molecular Medicine of the Austrian Academy of Sciences | Alexandra Popa et al |
| EPI_ISL_583638 | Austrian Agency for Health and Food Safety (AGES) | Bergthaler laboratory, CeMM Research Center for Molecular Medicine of the Austrian Academy of Sciences | Alexandra Popa et al |
| EPI_ISL_583666 | Austrian Agency for Health and Food Safety (AGES) | Bergthaler laboratory, CeMM Research Center for Molecular Medicine of the Austrian Academy of Sciences | Alexandra Popa et al |
| EPI_ISL_583670 | Austrian Agency for Health and Food Safety (AGES) | Bergthaler laboratory, CeMM Research Center for Molecular Medicine of the Austrian Academy of Sciences | Alexandra Popa et al |
| EPI_ISL_583677 | Austrian Agency for Health and Food Safety (AGES) | Bergthaler laboratory, CeMM Research Center for Molecular Medicine of the Austrian Academy of Sciences | Alexandra Popa et al |
| EPI_ISL_583717 | Center for Virology, Medical University of Vienna | Bergthaler laboratory, CeMM Research Center for Molecular Medicine of the Austrian Academy of Sciences | Alexandra Popa et al |
| EPI_ISL_583734 | Dr. Gernot Walder GmbH | Bergthaler laboratory, CeMM Research Center for Molecular Medicine of the Austrian Academy of Sciences | Alexandra Popa et al |
| EPI_ISL_583762 | Dr. Gernot Walder GmbH | Bergthaler laboratory, CeMM Research Center for Molecular Medicine of the Austrian Academy of Sciences | Alexandra Popa et al |
| EPI_ISL_583854 | Institute for Laboratory Diagnostics and Microbiology, Klinikum Klagenfurt am Worthersee | Bergthaler laboratory, CeMM Research Center for Molecular Medicine of the Austrian Academy of Sciences | Alexandra Popa et al |
| EPI_ISL_583883 | Austrian Agency for Health and Food Safety (AGES) | Bergthaler laboratory, CeMM Research Center for Molecular Medicine of the Austrian Academy of Sciences | Alexandra Popa et al |
| EPI_ISL_583884 | Austrian Agency for Health and Food Safety (AGES) | Bergthaler laboratory, CeMM Research Center for Molecular Medicine of the Austrian Academy of Sciences | Alexandra Popa et al |
| EPI_ISL_583885 | Austrian Agency for Health and Food Safety (AGES) | Bergthaler laboratory, CeMM Research Center for Molecular Medicine of the Austrian Academy of Sciences | Alexandra Popa et al |
| EPI_ISL_583890 | Austrian Agency for Health and Food Safety (AGES) | Bergthaler laboratory, CeMM Research Center for Molecular Medicine of the Austrian Academy of Sciences | Alexandra Popa et al |
| EPI_ISL_1396166 | Laboratorio de Virología del Hospital de Niños Dr. Ricardo Gutierrez | Área de Secuenciación del Laboratorio de Virología del Hospital de Niños Dr. Ricardo Gutierrez on behalf of 'Proyecto Argentino Interinstitucional de genomica de SARS-CoV-2' (PAIS Consortium) | Alexay et al |
| EPI_ISL_1396170 | Laboratorio de Virología del Hospital de Niños Dr. Ricardo Gutierrez | Área de Secuenciación del Laboratorio de Virología del Hospital de Niños Dr. Ricardo Gutierrez on behalf of 'Proyecto Argentino Interinstitucional de genomica de SARS-CoV-2' (PAIS Consortium) | Alexay et al |
| EPI_ISL_1396173 | Laboratorio de Virología del Hospital de Niños Dr. Ricardo Gutierrez | Área de Secuenciación del Laboratorio de Virología del Hospital de Niños Dr. Ricardo Gutierrez on behalf of 'Proyecto Argentino Interinstitucional de genomica de SARS-CoV-2' (PAIS Consortium) | Alexay et al |
| EPI_ISL_1396175 | Laboratorio de Virología del Hospital de Niños Dr. Ricardo Gutierrez | Área de Secuenciación del Laboratorio de Virología del Hospital de Niños Dr. Ricardo Gutierrez on behalf of 'Proyecto Argentino Interinstitucional de genomica de SARS-CoV-2' (PAIS Consortium) | Alexay et al |
| EPI_ISL_1396185 | Laboratorio de Virología del Hospital de Niños Dr. Ricardo Gutierrez | Área de Secuenciación del Laboratorio de Virología del Hospital de Niños Dr. Ricardo Gutierrez on behalf of 'Proyecto Argentino Interinstitucional de genomica de SARS-CoV-2' (PAIS Consortium) | Alexay et al |

| | | | |
|---|---|---|---|
| EPI_ISL_1396191 | Laboratorio de Virología del Hospital de Niños Dr. Ricardo Gutierrez | Área de Secuenciación del Laboratorio de Virología del Hospital de Niños Dr. Ricardo Gutierrez on behalf of 'Proyecto Argentino Interinstitucional de genomica de SARS-CoV-2' (PAIS Consortium) | Alexay et al |
| EPI_ISL_1396192 | Laboratorio de Virología del Hospital de Niños Dr. Ricardo Gutierrez | Área de Secuenciación del Laboratorio de Virología del Hospital de Niños Dr. Ricardo Gutierrez on behalf of 'Proyecto Argentino Interinstitucional de genomica de SARS-CoV-2' (PAIS Consortium) | Alexay et al |
| EPI_ISL_1396193 | Laboratorio de Virología del Hospital de Niños Dr. Ricardo Gutierrez | Área de Secuenciación del Laboratorio de Virología del Hospital de Niños Dr. Ricardo Gutierrez on behalf of 'Proyecto Argentino Interinstitucional de genomica de SARS-CoV-2' (PAIS Consortium) | Alexay et al |
| EPI_ISL_1396194 | Laboratorio de Virología del Hospital de Niños Dr. Ricardo Gutierrez | Área de Secuenciación del Laboratorio de Virología del Hospital de Niños Dr. Ricardo Gutierrez on behalf of 'Proyecto Argentino Interinstitucional de genomica de SARS-CoV-2' (PAIS Consortium) | Alexay et al |
| EPI_ISL_1396197 | Laboratorio de Virología del Hospital de Niños Dr. Ricardo Gutierrez | Área de Secuenciación del Laboratorio de Virología del Hospital de Niños Dr. Ricardo Gutierrez on behalf of 'Proyecto Argentino Interinstitucional de genomica de SARS-CoV-2' (PAIS Consortium) | Alexay et al |
| EPI_ISL_1396201 | Laboratorio de Virología del Hospital de Niños Dr. Ricardo Gutierrez | Área de Secuenciación del Laboratorio de Virología del Hospital de Niños Dr. Ricardo Gutierrez on behalf of 'Proyecto Argentino Interinstitucional de genomica de SARS-CoV-2' (PAIS Consortium) | Alexay et al |
| EPI_ISL_1396204 | Laboratorio de Virología del Hospital de Niños Dr. Ricardo Gutierrez | Área de Secuenciación del Laboratorio de Virología del Hospital de Niños Dr. Ricardo Gutierrez on behalf of 'Proyecto Argentino Interinstitucional de genomica de SARS-CoV-2' (PAIS Consortium) | Alexay et al |
| EPI_ISL_1396206 | Laboratorio de Virología del Hospital de Niños Dr. Ricardo Gutierrez | Área de Secuenciación del Laboratorio de Virología del Hospital de Niños Dr. Ricardo Gutierrez on behalf of 'Proyecto Argentino Interinstitucional de genomica de SARS-CoV-2' (PAIS Consortium) | Alexay et al |
| EPI_ISL_1396215 | Laboratorio de Virología del Hospital de Niños Dr. Ricardo Gutierrez | Área de Secuenciación del Laboratorio de Virología del Hospital de Niños Dr. Ricardo Gutierrez on behalf of 'Proyecto Argentino Interinstitucional de genomica de SARS-CoV-2' (PAIS Consortium) | Alexay et al |
| EPI_ISL_1396224 | Laboratorio de Virología del Hospital de Niños Dr. Ricardo Gutierrez | Área de Secuenciación del Laboratorio de Virología del Hospital de Niños Dr. Ricardo Gutierrez on behalf of 'Proyecto Argentino Interinstitucional de genomica de SARS-CoV-2' (PAIS Consortium) | Alexay et al |
| EPI_ISL_1396225 | Laboratorio de Virología del Hospital de Niños Dr. Ricardo Gutierrez | Área de Secuenciación del Laboratorio de Virología del Hospital de Niños Dr. Ricardo Gutierrez on behalf of 'Proyecto Argentino Interinstitucional de genomica de SARS-CoV-2' (PAIS Consortium) | Alexay et al |
| EPI_ISL_1396244 | Laboratorio de Virología del Hospital de Niños Dr. Ricardo Gutierrez | Área de Secuenciación del Laboratorio de Virología del Hospital de Niños Dr. Ricardo Gutierrez on behalf of 'Proyecto Argentino Interinstitucional de genomica de SARS-CoV-2' (PAIS Consortium) | Alexay et al |
| EPI_ISL_1395778 | Laboratorio de Virología del Hospital de Niños Dr. Ricardo Gutierrez | Área de Secuenciación del Laboratorio de Virología del Hospital de Niños Dr. Ricardo Gutierrez on behalf of 'Proyecto Argentino Interinstitucional de genomica de SARS-CoV-2' (PAIS Consortium) | Alexay et al |
| EPI_ISL_1395779 | Laboratorio de Virología del Hospital de Niños Dr. Ricardo Gutierrez | Área de Secuenciación del Laboratorio de Virología del Hospital de Niños Dr. Ricardo Gutierrez on behalf of 'Proyecto Argentino Interinstitucional de genomica de SARS-CoV-2' (PAIS Consortium) | Alexay et al |
| EPI_ISL_1395780 | Laboratorio de Virología del Hospital de Niños Dr. Ricardo Gutierrez | Área de Secuenciación del Laboratorio de Virología del Hospital de Niños Dr. Ricardo Gutierrez on behalf of 'Proyecto Argentino Interinstitucional de genomica de SARS-CoV-2' (PAIS Consortium) | Alexay et al |

| | | | |
|---|---|---|---|
| EPI_ISL_1395792 | Laboratorio de Virología del Hospital de Niños Dr. Ricardo Gutierrez | Área de Secuenciación del Laboratorio de Virología del Hospital de Niños Dr. Ricardo Gutierrez on behalf of 'Proyecto Argentino Interinstitucional de genomica de SARS-CoV-2' (PAIS Consortium) | Alexay et al |
| EPI_ISL_1395793 | Laboratorio de Virología del Hospital de Niños Dr. Ricardo Gutierrez | Área de Secuenciación del Laboratorio de Virología del Hospital de Niños Dr. Ricardo Gutierrez on behalf of 'Proyecto Argentino Interinstitucional de genomica de SARS-CoV-2' (PAIS Consortium) | Alexay et al |
| EPI_ISL_1396246 | Laboratorio de Virología del Hospital de Niños Dr. Ricardo Gutierrez | Biocódices SA. on behalf of 'Proyecto Argentino Interinstitucional de genomica de SARS-CoV-2' (PAIS Consortium) | Alexay et al |
| EPI_ISL_1396304 | Laboratorio de Virología del Hospital de Niños Dr. Ricardo Gutierrez | Biocódices SA. on behalf of 'Proyecto Argentino Interinstitucional de genomica de SARS-CoV-2' (PAIS Consortium) | Alexay et al |
| EPI_ISL_1396318 | Laboratorio de Virología del Hospital de Niños Dr. Ricardo Gutierrez | Biocódices SA. on behalf of 'Proyecto Argentino Interinstitucional de genomica de SARS-CoV-2' (PAIS Consortium) | Alexay et al |
| EPI_ISL_1396327 | Laboratorio de Virología del Hospital de Niños Dr. Ricardo Gutierrez | Biocódices SA. on behalf of 'Proyecto Argentino Interinstitucional de genomica de SARS-CoV-2' (PAIS Consortium) | Alexay et al |
| EPI_ISL_872637 | Pathogenic Microorganisms Variability Laboratory | Pathogenic Microorganisms Variability Laboratory | Alexey Shchetinin et al |
| EPI_ISL_872638 | Pathogenic Microorganisms Variability Laboratory | Pathogenic Microorganisms Variability Laboratory | Alexey Shchetinin et al |
| EPI_ISL_1233441 | National Center for Infectious and Parasitic Diseases (NCIPD) | National Center for Infectious and Parasitic Diseases (NCIPD) | Alexiev et al |
| EPI_ISL_1233454 | National Center for Infectious and Parasitic Diseases (NCIPD) | National Center for Infectious and Parasitic Diseases (NCIPD) | Alexiev et al |
| EPI_ISL_1233495 | National Center for Infectious and Parasitic Diseases (NCIPD) | National Center for Infectious and Parasitic Diseases (NCIPD) | Alexiev et al |
| EPI_ISL_1233531 | National Center for Infectious and Parasitic Diseases (NCIPD) | National Center for Infectious and Parasitic Diseases (NCIPD) | Alexiev et al |
| EPI_ISL_1233561 | National Center for Infectious and Parasitic Diseases (NCIPD) | National Center for Infectious and Parasitic Diseases (NCIPD) | Alexiev et al |
| EPI_ISL_1233611 | National Center for Infectious and Parasitic Diseases (NCIPD) | National Center for Infectious and Parasitic Diseases (NCIPD) | Alexiev et al |
| EPI_ISL_1233617 | National Center for Infectious and Parasitic Diseases (NCIPD) | National Center for Infectious and Parasitic Diseases (NCIPD) | Alexiev et al |
| EPI_ISL_1233623 | National Center for Infectious and Parasitic Diseases (NCIPD) | National Center for Infectious and Parasitic Diseases (NCIPD) | Alexiev et al |
| EPI_ISL_1231635 | National Center for Infectious and Parasitic Diseases (NCIPD) | National Center for Infectious and Parasitic Diseases (NCIPD) | Alexiev et al |
| EPI_ISL_1401123 | National Center of Infectious and Parasitic Diseases | National Center of Infectious and Parasitic Diseases | Alexiev et al |
| EPI_ISL_1401141 | National Center of Infectious and Parasitic Diseases | National Center of Infectious and Parasitic Diseases | Alexiev et al |
| EPI_ISL_1401437 | National Center of Infectious and Parasitic Diseases | National Center of Infectious and Parasitic Diseases | Alexiev et al |
| EPI_ISL_1608340 | National Center of Infectious and Parasitic Diseases | National Center of Infectious and Parasitic Diseases | Alexiev et al |
| EPI_ISL_1608347 | National Center of Infectious and Parasitic Diseases | National Center of Infectious and Parasitic Diseases | Alexiev et al |
| EPI_ISL_1608354 | National Center of Infectious and Parasitic Diseases | National Center of Infectious and Parasitic Diseases | Alexiev et al |
| EPI_ISL_1608362 | National Center of Infectious and Parasitic Diseases | National Center of Infectious and Parasitic Diseases | Alexiev et al |
| EPI_ISL_1608370 | National Center of Infectious and Parasitic Diseases | National Center of Infectious and Parasitic Diseases | Alexiev et al |
| EPI_ISL_1608436 | National Center of Infectious and Parasitic Diseases | National Center of Infectious and Parasitic Diseases | Alexiev et al |
| EPI_ISL_1608511 | National Center of Infectious and Parasitic Diseases | National Center of Infectious and Parasitic Diseases | Alexiev et al |

| | | | |
|---|---|---|---|
| EPI_ISL_1302126 | National Center of Infectious and Parasitic Diseases | National Center of Infectious and Parasitic Diseases | Alexiev et al |
| EPI_ISL_1302130 | National Center of Infectious and Parasitic Diseases | National Center of Infectious and Parasitic Diseases | Alexiev et al |
| EPI_ISL_1302046 | National Center of Infectious and Parasitic Diseases | National Center of Infectious and Parasitic Diseases | Alexiev et al |
| EPI_ISL_1509778 | National Center of Infectious and Parasitic Diseases | National Center of Infectious and Parasitic Diseases | Alexiev et al |
| EPI_ISL_1509800 | National Center of Infectious and Parasitic Diseases | National Center of Infectious and Parasitic Diseases | Alexiev et al |
| EPI_ISL_1509887 | National Center of Infectious and Parasitic Diseases | National Center of Infectious and Parasitic Diseases | Alexiev et al |
| EPI_ISL_959923 | University Medical Center Hamburg Eppendorf | Heinrich Pette Institute, Leibniz Institute for Experimental Virology | Alexis Robitaille et al |
| EPI_ISL_960056 | University Medical Center Hamburg Eppendorf | Heinrich Pette Institute, Leibniz Institute for Experimental Virology | Alexis Robitaille et al |
| EPI_ISL_775788 | University Medical Center Hamburg Eppendorf | Heinrich Pette Institute, Leibniz Institute for Experimental Virology | Alexis Robitaille et al |
| EPI_ISL_775789 | University Medical Center Hamburg Eppendorf | Heinrich Pette Institute, Leibniz Institute for Experimental Virology | Alexis Robitaille et al |
| EPI_ISL_776012 | University Medical Center Hamburg Eppendorf | Heinrich Pette Institute, Leibniz Institute for Experimental Virology | Alexis Robitaille et al |
| EPI_ISL_776539 | University Medical Center Hamburg Eppendorf | Heinrich Pette Institute, Leibniz Institute for Experimental Virology | Alexis Robitaille et al |
| EPI_ISL_491945 | Instituto Nacional de Investigación en Salud Pública - INSPI | INSPI - Charité | Alfredo Bruno Caicedo et al |
| EPI_ISL_491944 | Instituto Nacional de Investigación en Salud Pública - INSPI | INSPI - Charité | Alfredo Bruno Caicedo et al |
| EPI_ISL_491948 | Instituto Nacional de Investigación en Salud Pública - INSPI | INSPI - Charité | Alfredo Bruno Caicedo et al |
| EPI_ISL_491953 | Instituto Nacional de Investigación en Salud Pública - INSPI | INSPI - Charité | Alfredo Bruno Caicedo et al |
| EPI_ISL_491950 | Instituto Nacional de Investigación en Salud Pública - INSPI | INSPI - Charité | Alfredo Bruno Caicedo et al |
| EPI_ISL_491949 | Instituto Nacional de Investigación en Salud Pública - INSPI | INSPI - Charité | Alfredo Bruno Caicedo et al |
| EPI_ISL_491952 | Instituto Nacional de Investigación en Salud Pública - INSPI | INSPI - Charité | Alfredo Bruno Caicedo et al |
| EPI_ISL_491954 | Instituto Nacional de Investigación en Salud Pública - INSPI | INSPI - Charité | Alfredo Bruno Caicedo et al |
| EPI_ISL_632905 | Communicable Disease Laboratory, Public Health Directorate | Communicable Disease Laboratory, Public Health Directorate | AlHujairi et al |
| EPI_ISL_632908 | Genomic Sciences, Rehman Medical Institute | Genomic Sciences, Rehman Medical Institute | Ali et al |
| EPI_ISL_943595 | Central Laboratory of Public Health of Rio Grande do Sul (Lacen-RS) | State Center for Health Surveillance of the Health Department of the State of Rio Grande do Sul (CEVS/SES-RS) | Aline Campos et al |
| EPI_ISL_943596 | Central Laboratory of Public Health of Rio Grande do Sul (Lacen-RS) | State Center for Health Surveillance of the Health Department of the State of Rio Grande do Sul (CEVS/SES-RS) | Aline Campos et al |
| EPI_ISL_943589 | Central Laboratory of Public Health of Rio Grande do Sul (Lacen-RS) | State Center for Health Surveillance of the Health Department of the State of Rio Grande do Sul (CEVS/SES-RS) | Aline Campos et al |
| EPI_ISL_943597 | Central Laboratory of Public Health of Rio Grande do Sul (Lacen-RS) | State Center for Health Surveillance of the Health Department of the State of Rio Grande do Sul (CEVS/SES-RS) | Aline Campos et al |
| EPI_ISL_943598 | Central Laboratory of Public Health of Rio Grande do Sul (Lacen-RS) | State Center for Health Surveillance of the Health Department of the State of Rio Grande do Sul (CEVS/SES-RS) | Aline Campos et al |
| EPI_ISL_943590 | Central Laboratory of Public Health of Rio Grande do Sul (Lacen-RS) | State Center for Health Surveillance of the Health Department of the State of Rio Grande do Sul (CEVS/SES-RS) | Aline Campos et al |

| | | | |
|---|---|---|---|
| EPI_ISL_943591 | Central Laboratory of Public Health of Rio Grande do Sul (Lacen-RS) | State Center for Health Surveillance of the Health Department of the State of Rio Grande do Sul (CEVS/SES-RS) | Aline Campos et al |
| EPI_ISL_943593 | Central Laboratory of Public Health of Rio Grande do Sul (Lacen-RS) | State Center for Health Surveillance of the Health Department of the State of Rio Grande do Sul (CEVS/SES-RS) | Aline Campos et al |
| EPI_ISL_943594 | Central Laboratory of Public Health of Rio Grande do Sul (Lacen-RS) | State Center for Health Surveillance of the Health Department of the State of Rio Grande do Sul (CEVS/SES-RS) | Aline Campos et al |
| EPI_ISL_943600 | Central Laboratory of Public Health of Rio Grande do Sul (Lacen-RS) | State Center for Health Surveillance of the Health Department of the State of Rio Grande do Sul (CEVS/SES-RS) | Aline Campos et al |
| EPI_ISL_943579 | Central Laboratory of Public Health of Rio Grande do Sul (Lacen-RS) | State Center for Health Surveillance of the Health Department of the State of Rio Grande do Sul (CEVS/SES-RS) | Aline Campos et al |
| EPI_ISL_943580 | Central Laboratory of Public Health of Rio Grande do Sul (Lacen-RS) | State Center for Health Surveillance of the Health Department of the State of Rio Grande do Sul (CEVS/SES-RS) | Aline Campos et al |
| EPI_ISL_943577 | Central Laboratory of Public Health of Rio Grande do Sul (Lacen-RS) | State Center for Health Surveillance of the Health Department of the State of Rio Grande do Sul (CEVS/SES-RS) | Aline Campos et al |
| EPI_ISL_943578 | Central Laboratory of Public Health of Rio Grande do Sul (Lacen-RS) | State Center for Health Surveillance of the Health Department of the State of Rio Grande do Sul (CEVS/SES-RS) | Aline Campos et al |
| EPI_ISL_943576 | Central Laboratory of Public Health of Rio Grande do Sul (Lacen-RS) | State Center for Health Surveillance of the Health Department of the State of Rio Grande do Sul (CEVS/SES-RS) | Aline Campos et al |
| EPI_ISL_943581 | Central Laboratory of Public Health of Rio Grande do Sul (Lacen-RS) | State Center for Health Surveillance of the Health Department of the State of Rio Grande do Sul (CEVS/SES-RS) | Aline Campos et al |
| EPI_ISL_943582 | Central Laboratory of Public Health of Rio Grande do Sul (Lacen-RS) | State Center for Health Surveillance of the Health Department of the State of Rio Grande do Sul (CEVS/SES-RS) | Aline Campos et al |
| EPI_ISL_943583 | Central Laboratory of Public Health of Rio Grande do Sul (Lacen-RS) | State Center for Health Surveillance of the Health Department of the State of Rio Grande do Sul (CEVS/SES-RS) | Aline Campos et al |
| EPI_ISL_943584 | Central Laboratory of Public Health of Rio Grande do Sul (Lacen-RS) | State Center for Health Surveillance of the Health Department of the State of Rio Grande do Sul (CEVS/SES-RS) | Aline Campos et al |
| EPI_ISL_943585 | Central Laboratory of Public Health of Rio Grande do Sul (Lacen-RS) | State Center for Health Surveillance of the Health Department of the State of Rio Grande do Sul (CEVS/SES-RS) | Aline Campos et al |
| EPI_ISL_943588 | Central Laboratory of Public Health of Rio Grande do Sul (Lacen-RS) | State Center for Health Surveillance of the Health Department of the State of Rio Grande do Sul (CEVS/SES-RS) | Aline Campos et al |
| EPI_ISL_943586 | Central Laboratory of Public Health of Rio Grande do Sul (Lacen-RS) | State Center for Health Surveillance of the Health Department of the State of Rio Grande do Sul (CEVS/SES-RS) | Aline Campos et al |
| EPI_ISL_943587 | Central Laboratory of Public Health of Rio Grande do Sul (Lacen-RS) | State Center for Health Surveillance of the Health Department of the State of Rio Grande do Sul (CEVS/SES-RS) | Aline Campos et al |
| EPI_ISL_943574 | Central Laboratory of Public Health of Rio Grande do Sul (Lacen-RS) | State Center for Health Surveillance of the Health Department of the State of Rio Grande do Sul (CEVS/SES-RS) | Aline Campos et al |
| EPI_ISL_943575 | Central Laboratory of Public Health of Rio Grande do Sul (Lacen-RS) | State Center for Health Surveillance of the Health Department of the State of Rio Grande do Sul (CEVS/SES-RS) | Aline Campos et al |
| EPI_ISL_943592 | Central Laboratory of Public Health of Rio Grande do Sul (Lacen-RS) | State Center for Health Surveillance of the Health Department of the State of Rio Grande do Sul (CEVS/SES-RS) | Aline Campos et al |
| EPI_ISL_943605 | Central Laboratory of Public Health of Rio Grande do Sul (Lacen-RS) | State Center for Health Surveillance of the Health Department of the State of Rio Grande do Sul (CEVS/SES-RS) | Aline Campos et al |
| EPI_ISL_943606 | Central Laboratory of Public Health of Rio Grande do Sul (Lacen-RS) | State Center for Health Surveillance of the Health Department of the State of Rio Grande do Sul (CEVS/SES-RS) | Aline Campos et al |

| | | | |
|---|---|---|---|
| EPI_ISL_943607 | Central Laboratory of Public Health of Rio Grande do Sul (Lacen-RS) | State Center for Health Surveillance of the Health Department of the State of Rio Grande do Sul (CEVS/SES-RS) | Aline Campos et al |
| EPI_ISL_943608 | Central Laboratory of Public Health of Rio Grande do Sul (Lacen-RS) | State Center for Health Surveillance of the Health Department of the State of Rio Grande do Sul (CEVS/SES-RS) | Aline Campos et al |
| EPI_ISL_943602 | Central Laboratory of Public Health of Rio Grande do Sul (Lacen-RS) | State Center for Health Surveillance of the Health Department of the State of Rio Grande do Sul (CEVS/SES-RS) | Aline Campos et al |
| EPI_ISL_943603 | Central Laboratory of Public Health of Rio Grande do Sul (Lacen-RS) | State Center for Health Surveillance of the Health Department of the State of Rio Grande do Sul (CEVS/SES-RS) | Aline Campos et al |
| EPI_ISL_943609 | Central Laboratory of Public Health of Rio Grande do Sul (Lacen-RS) | State Center for Health Surveillance of the Health Department of the State of Rio Grande do Sul (CEVS/SES-RS) | Aline Campos et al |
| EPI_ISL_943611 | Central Laboratory of Public Health of Rio Grande do Sul (Lacen-RS) | State Center for Health Surveillance of the Health Department of the State of Rio Grande do Sul (CEVS/SES-RS) | Aline Campos et al |
| EPI_ISL_943610 | Central Laboratory of Public Health of Rio Grande do Sul (Lacen-RS) | State Center for Health Surveillance of the Health Department of the State of Rio Grande do Sul (CEVS/SES-RS) | Aline Campos et al |
| EPI_ISL_943612 | Central Laboratory of Public Health of Rio Grande do Sul (Lacen-RS) | State Center for Health Surveillance of the Health Department of the State of Rio Grande do Sul (CEVS/SES-RS) | Aline Campos et al |
| EPI_ISL_943613 | Central Laboratory of Public Health of Rio Grande do Sul (Lacen-RS) | State Center for Health Surveillance of the Health Department of the State of Rio Grande do Sul (CEVS/SES-RS) | Aline Campos et al |
| EPI_ISL_943604 | Central Laboratory of Public Health of Rio Grande do Sul (Lacen-RS) | State Center for Health Surveillance of the Health Department of the State of Rio Grande do Sul (CEVS/SES-RS) | Aline Campos et al |
| EPI_ISL_983863 | Central Laboratory of Public Health of Rio Grande do Sul (Lacen-RS) | State Center for Health Surveillance of the Health Department of the State of Rio Grande do Sul (CEVS/SES-RS) | Aline Campos et al |
| EPI_ISL_983864 | Central Laboratory of Public Health of Rio Grande do Sul (Lacen-RS) | State Center for Health Surveillance of the Health Department of the State of Rio Grande do Sul (CEVS/SES-RS) | Aline Campos et al |
| EPI_ISL_983867 | Central Laboratory of Public Health of Rio Grande do Sul (Lacen-RS) | State Center for Health Surveillance of the Health Department of the State of Rio Grande do Sul (CEVS/SES-RS) | Aline Campos et al |
| EPI_ISL_983869 | Central Laboratory of Public Health of Rio Grande do Sul (Lacen-RS) | State Center for Health Surveillance of the Health Department of the State of Rio Grande do Sul (CEVS/SES-RS) | Aline Campos et al |
| EPI_ISL_983865 | Central Laboratory of Public Health of Rio Grande do Sul (Lacen-RS) | State Center for Health Surveillance of the Health Department of the State of Rio Grande do Sul (CEVS/SES-RS) | Aline Campos et al |
| EPI_ISL_983868 | Central Laboratory of Public Health of Rio Grande do Sul (Lacen-RS) | State Center for Health Surveillance of the Health Department of the State of Rio Grande do Sul (CEVS/SES-RS) | Aline Campos et al |
| EPI_ISL_983866 | Central Laboratory of Public Health of Rio Grande do Sul (Lacen-RS) | State Center for Health Surveillance of the Health Department of the State of Rio Grande do Sul (CEVS/SES-RS) | Aline Campos et al |
| EPI_ISL_984620 | Central Laboratory of Public Health of Rio Grande do Sul (Lacen-RS) | State Center for Health Surveillance of the Health Department of the State of Rio Grande do Sul (CEVS/SES-RS) | Aline Campos et al |
| EPI_ISL_984621 | Central Laboratory of Public Health of Rio Grande do Sul (Lacen-RS) | State Center for Health Surveillance of the Health Department of the State of Rio Grande do Sul (CEVS/SES-RS) | Aline Campos et al |
| EPI_ISL_984619 | Central Laboratory of Public Health of Rio Grande do Sul (Lacen-RS) | State Center for Health Surveillance of the Health Department of the State of Rio Grande do Sul (CEVS/SES-RS) | Aline Campos et al |
| EPI_ISL_943601 | Central Laboratory of Public Health of Rio Grande do Sul (Lacen-RS) | State Center for Health Surveillance of the Health Department of the State of Rio Grande do Sul (CEVS/SES-RS) | Aline Campos et al |
| EPI_ISL_515155 | National Institute for Communicable Diseases of the National Health Laboratory Service | National Institute for Communicable Diseases of the National Health Laboratory Service | Allam M et al |

| | | | |
|---|---|---|---|
| EPI_ISL_514386 | National Institute for Communicable Diseases of the National Health Laboratory Service | National Institute for Communicable Diseases of the National Health Laboratory Service | Allam M et al |
| EPI_ISL_514408 | National Institute for Communicable Diseases of the National Health Laboratory Service | National Institute for Communicable Diseases of the National Health Laboratory Service | Allam M et al |
| EPI_ISL_514417 | National Institute for Communicable Diseases of the National Health Laboratory Service | National Institute for Communicable Diseases of the National Health Laboratory Service | Allam M et al |
| EPI_ISL_622982 | National Health Laboratory Service | National Institute for Communicable Diseases of the National Health Laboratory Service | Allam M et al |
| EPI_ISL_622976 | National Health Laboratory Service | National Institute for Communicable Diseases of the National Health Laboratory Service | Allam M et al |
| EPI_ISL_622977 | National Health Laboratory Service | National Institute for Communicable Diseases of the National Health Laboratory Service | Allam M et al |
| EPI_ISL_450297 | National Institute for Communicable Diseases of the National Health Laboratory Service | National Institute for Communicable Diseases of the National Health Laboratory Service | Allam M et al |
| EPI_ISL_470878 | National Institute for Communicable Diseases of the National Health Laboratory Service | National Institute for Communicable Diseases of the National Health Laboratory Service | Allam M et al |
| EPI_ISL_470879 | National Institute for Communicable Diseases of the National Health Laboratory Service | National Institute for Communicable Diseases of the National Health Laboratory Service | Allam M et al |
| EPI_ISL_853127 | Toronto Invasive Bacterial Diseases Network | Ontario Institute for Cancer Research | Allison McGeer et al |
| EPI_ISL_853120 | Toronto Invasive Bacterial Diseases Network | Ontario Institute for Cancer Research | Allison McGeer et al |
| EPI_ISL_755696 | Toronto Invasive Bacterial Diseases Network | McMaster University | Allison McGeer et al |
| EPI_ISL_962186 | Toronto Invasive Bacterial Diseases Network | McMaster University | Allison McGeer et al |
| EPI_ISL_962196 | Toronto Invasive Bacterial Diseases Network | McMaster University | Allison McGeer et al |
| EPI_ISL_591267 | Toronto Invasive Bacterial Diseases Network | McMaster University | Allison McGeer et al |
| EPI_ISL_489996 | King Fahad Medical City | King Fahad Medical City | Alosaimi et al |
| EPI_ISL_489997 | King Fahad Medical City | King Fahad Medical City | Alosaimi et al |
| EPI_ISL_489998 | King Fahad Medical City | King Fahad Medical City | Alosaimi et al |
| EPI_ISL_490000 | King Fahad Medical City | King Fahad Medical City | Alosaimi et al |
| EPI_ISL_490002 | King Fahad Medical City | King Fahad Medical City | Alosaimi et al |
| EPI_ISL_490003 | King Fahad Medical City | King Fahad Medical City | Alosaimi et al |
| EPI_ISL_490004 | King Fahad Medical City | King Fahad Medical City | Alosaimi et al |
| EPI_ISL_788933 | Centre de Recherche et de Formation en Infectiologie Guinée | TransVIHMI, IRD/INSERM/Monpellier University | Alpha Kabinet KEITA et al |
| EPI_ISL_788934 | Centre de Recherche et de Formation en Infectiologie Guinée | TransVIHMI, IRD/INSERM/Monpellier University | Alpha Kabinet KEITA et al |
| EPI_ISL_788935 | Centre de Recherche et de Formation en Infectiologie Guinée | TransVIHMI, IRD/INSERM/Monpellier University | Alpha Kabinet KEITA et al |
| EPI_ISL_788936 | Centre de Recherche et de Formation en Infectiologie Guinée | TransVIHMI, IRD/INSERM/Monpellier University | Alpha Kabinet KEITA et al |
| EPI_ISL_788937 | Centre de Recherche et de Formation en Infectiologie Guinée | TransVIHMI, IRD/INSERM/Monpellier University | Alpha Kabinet KEITA et al |

| | | | |
|---|---|---|---|
| EPI_ISL_788938 | Centre de Recherche et de Formation en Infectiologie Guinée | TransVIHMI, IRD/INSERM/Monpellier University | Alpha Kabinet KEITA et al |
| EPI_ISL_632283 | Communicable Disease Laboratory, Public Health Directorate | Communicable Disease Laboratory, Public Health Directorate | AlWasti et al |
| EPI_ISL_636973 | Public Health Lab | Public Health Lab | Alwasti et al |
| EPI_ISL_632285 | Communicable Disease Laboratory, Public Health Directorate | Communicable Disease Laboratory, Public Health Directorate | AlWasti et al |
| EPI_ISL_632262 | Communicable Disease Laboratory, Public Health Directorate | Communicable Disease Laboratory, Public Health Directorate | AlWasti et al |
| EPI_ISL_632263 | Communicable Disease Laboratory, Public Health Directorate | Communicable Disease Laboratory, Public Health Directorate | AlWasti et al |
| EPI_ISL_681309 | Communicable Disease Laboratory, Public Health Directorate | Communicable Disease Laboratory, Public Health Directorate | Alwasti et al |
| EPI_ISL_682300 | Communicable Disease Laboratory, Public Health Directorate | Communicable Disease Laboratory, Public Health Directorate | Alwasti et al |
| EPI_ISL_632269 | Communicable Disease Laboratory, Public Health Directorate | Communicable Disease Laboratory, Public Health Directorate | AlWasti et al |
| EPI_ISL_632273 | Communicable Disease Laboratory, Public Health Directorate | Communicable Disease Laboratory, Public Health Directorate | AlWasti et al |
| EPI_ISL_632274 | Communicable Disease Laboratory, Public Health Directorate | Communicable Disease Laboratory, Public Health Directorate | AlWasti et al |
| EPI_ISL_632279 | Communicable Disease Laboratory, Public Health Directorate | Communicable Disease Laboratory, Public Health Directorate | AlWasti et al |
| EPI_ISL_632252 | Communicable Disease Laboratory, Public Health Directorate | Communicable Disease Laboratory, Public Health Directorate | AlWasti et al |
| EPI_ISL_632268 | Communicable Disease Laboratory, Public Health Directorate | Communicable Disease Laboratory, Public Health Directorate | AlWasti et al |
| EPI_ISL_681315 | Communicable Disease Laboratory, Public Health Directorate | Communicable Disease Laboratory, Public Health Directorate | Alwasti et al |
| EPI_ISL_684033 | Communicable Disease Laboratory, Public Health Directorate | Communicable Disease Laboratory, Public Health Directorate | Alwasti et al |
| EPI_ISL_684032 | Communicable Disease Laboratory, Public Health Directorate | Communicable Disease Laboratory, Public Health Directorate | Alwasti et al |
| EPI_ISL_682320 | Communicable Disease Laboratory, Public Health Directorate | Communicable Disease Laboratory, Public Health Directorate | Alwasti et al |
| EPI_ISL_678270 | Communicable Disease Laboratory, Public Health Directorate | Communicable Disease Laboratory, Public Health Directorate | Alwasti et al |
| EPI_ISL_1660462 | Communicable Disease Laboratory, Public Health Directorate | Communicable Disease Laboratory, Public Health Directorate | Alwasti et al |
| EPI_ISL_1660461 | Communicable Disease Laboratory, Public Health Directorate | Communicable Disease Laboratory, Public Health Directorate | Alwasti et al |
| EPI_ISL_1660463 | Communicable Disease Laboratory, Public Health Directorate | Communicable Disease Laboratory, Public Health Directorate | Alwasti et al |
| EPI_ISL_1660464 | Communicable Disease Laboratory, Public Health Directorate | Communicable Disease Laboratory, Public Health Directorate | Alwasti et al |
| EPI_ISL_1660465 | Communicable Disease Laboratory, Public Health Directorate | Communicable Disease Laboratory, Public Health Directorate | Alwasti et al |
| EPI_ISL_1660466 | Communicable Disease Laboratory, Public Health Directorate | Communicable Disease Laboratory, Public Health Directorate | Alwasti et al |
| EPI_ISL_1660467 | Communicable Disease Laboratory, Public Health Directorate | Communicable Disease Laboratory, Public Health Directorate | Alwasti et al |
| EPI_ISL_678261 | Communicable Disease Laboratory, Public Health Directorate | Communicable Disease Laboratory, Public Health Directorate | Alwasti et al |
| EPI_ISL_1660437 | Communicable Disease Laboratory, Public Health Directorate | Communicable Disease Laboratory, Public Health Directorate | Alwasti et al |
| EPI_ISL_1660426 | Communicable Disease Laboratory, Public Health Directorate | Communicable Disease Laboratory, Public Health Directorate | Alwasti et al |
| EPI_ISL_1660428 | Communicable Disease Laboratory, Public Health Directorate | Communicable Disease Laboratory, Public Health Directorate | Alwasti et al |

| | | | |
|---|---|---|---|
| EPI_ISL_1660434 | Communicable Disease Laboratory, Public Health Directorate | Communicable Disease Laboratory, Public Health Directorate | Alwasti et al |
| EPI_ISL_1660435 | Communicable Disease Laboratory, Public Health Directorate | Communicable Disease Laboratory, Public Health Directorate | Alwasti et al |
| EPI_ISL_632266 | Communicable Disease Laboratory, Public Health Directorate | Communicable Disease Laboratory, Public Health Directorate | AlWasti et al |
| EPI_ISL_632267 | Communicable Disease Laboratory, Public Health Directorate | Communicable Disease Laboratory, Public Health Directorate | AlWasti et al |
| EPI_ISL_681301 | Communicable Disease Laboratory, Public Health Directorate | Communicable Disease Laboratory, Public Health Directorate | Alwasti et al |
| EPI_ISL_682303 | Communicable Disease Laboratory, Public Health Directorate | Communicable Disease Laboratory, Public Health Directorate | Alwasti et al |
| EPI_ISL_1660443 | Communicable Disease Laboratory, Public Health Directorate | Communicable Disease Laboratory, Public Health Directorate | Alwasti et al |
| EPI_ISL_1660448 | Communicable Disease Laboratory, Public Health Directorate | Communicable Disease Laboratory, Public Health Directorate | Alwasti et al |
| EPI_ISL_1660453 | Communicable Disease Laboratory, Public Health Directorate | Communicable Disease Laboratory, Public Health Directorate | Alwasti et al |
| EPI_ISL_792525 | Laboratorio del Hospital Interzonal General de Agudos Evita | Grupo de Genómica y Bioinformática del Instituto de Investigación de la Cadena Láctea CONICET-INTA on behalf of 'Proyecto Argentino Interinstitucional de genomica de SARS-CoV-2' (PAIS Consortium) | Amadio et al |
| EPI_ISL_1398364 | Emam Ali Hospital | Razi Vaccine and Serum Research Institute | Amir Kaffashi et al |
| EPI_ISL_1398925 | Emam Ali Hospital | Razi Vaccine and Serum Research Institute | Amir Kaffashi et al |
| EPI_ISL_1398937 | Emam Ali Hospital | Razi Vaccine and Serum Research Institute | Amir Kaffashi et al |
| EPI_ISL_1239225 | Kimberley Hospital, National Health Laboratory Services, Northern Cape, South Africa | National Institute for Communicable Diseases of the National Health Laboratory Service | Amoako DG et al |
| EPI_ISL_1239860 | National Institute for Communicable Diseases,National Health Laboratory Services, Gauteng, South Africa | National Institute for Communicable Diseases of the National Health Laboratory Service | Amoako DG et al |
| EPI_ISL_425177 | Public Health Ontario | Public Health Agency of Canada - National Microbiology Laboratory | Amrit S. Boese et al |
| EPI_ISL_534612 | NHSGGC West of Scotland Specialist Virology Centre / MRC-University of Glasgow Centre for Virus Research | Wellcome Sanger Institute for the COVID-19 Genomics UK (COG-UK) consortium | Ana da Silva Filipe et al |
| EPI_ISL_534543 | NHSGGC West of Scotland Specialist Virology Centre / MRC-University of Glasgow Centre for Virus Research | Wellcome Sanger Institute for the COVID-19 Genomics UK (COG-UK) consortium | Ana da Silva Filipe et al |
| EPI_ISL_802105 | MSHS Clinical Microbiology Laboratories | MSHS Pathogen Surveillance Program | Ana S. Gonzalez-Reiche et al |
| EPI_ISL_802106 | MSHS Clinical Microbiology Laboratories | MSHS Pathogen Surveillance Program | Ana S. Gonzalez-Reiche et al |
| EPI_ISL_1300937 | MSHS Clinical Microbiology Laboratories | MSHS Pathogen Surveillance Program | Ana S. Gonzalez-Reiche et al |
| EPI_ISL_1301233 | MSHS Clinical Microbiology Laboratories | MSHS Pathogen Surveillance Program | Ana S. Gonzalez-Reiche et al |
| EPI_ISL_1301238 | MSHS Clinical Microbiology Laboratories | MSHS Pathogen Surveillance Program | Ana S. Gonzalez-Reiche et al |
| EPI_ISL_1395788 | Laboratorio del Hospital Teresa Germani | Área de Secuenciación del Laboratorio de Virología del Hospital de Niños Dr. Ricardo Gutierrez on behalf of 'Proyecto Argentino Interinstitucional de genomica de SARS-CoV-2' (PAIS Consortium) | Anabel Luzio et al |
| EPI_ISL_1395807 | Laboratorio del Hospital Teresa Germani | Área de Secuenciación del Laboratorio de Virología del Hospital de Niños Dr. Ricardo Gutierrez on behalf of 'Proyecto Argentino Interinstitucional de genomica de SARS-CoV-2' (PAIS Consortium) | Anabel Luzio et al |
| EPI_ISL_1081961 | Stefan S. NicolauInstitute of Virology | Stefan S. NicolauInstitute of Virology | Anca Botezatu et al |

| | | | |
|---|---|---|---|
| EPI_ISL_1251275 | Stefan S. NicolauInstitute of Virology | Stefan S. NicolauInstitute of Virology | Anca Botezatu et al |
| EPI_ISL_1508848 | Medical Genetics Laboratory, Regional Centre of Medical Genetics, Emergency County Hospital Craiova | Medical Genetics Laboratory, Regional Centre of Medical Genetics, Emergency County Hospital Craiova | Anca-Lelia (Riza) Costache et al |
| EPI_ISL_1533052 | Diagnostic and Research Center of Infectious Diseases, Medical Faculty, Andalas University | Diagnostic and Research Center of Infectious Diseases, Medical Faculty, Andalas University | Andani Eka Putra et al |
| EPI_ISL_1533065 | Diagnostic and Research Center of Infectious Diseases, Medical Faculty, Andalas University | Diagnostic and Research Center of Infectious Diseases, Medical Faculty, Andalas University | Andani Eka Putra et al |
| EPI_ISL_1533066 | Diagnostic and Research Center of Infectious Diseases, Medical Faculty, Andalas University | Diagnostic and Research Center of Infectious Diseases, Medical Faculty, Andalas University | Andani Eka Putra et al |
| EPI_ISL_1447336 | Diagnostic and Research Center of Infectious Diseases, Medical Faculty, Andalas University | Diagnostic and Research Center of Infectious Diseases, Medical Faculty, Andalas University | Andani Eka Putra et al |
| EPI_ISL_539787 | Institute of Microbiology, Universidad San Francisco de Quito | Institute of Microbiology, Universidad San Francisco de Quito | Andrea Macias et al |
| EPI_ISL_539788 | Institute of Microbiology, Universidad San Francisco de Quito | Institute of Microbiology, Universidad San Francisco de Quito | Andrea Macias et al |
| EPI_ISL_697794 | Institute of Microbiology, Universidad San Francisco de Quito | Institute of Microbiology, Universidad San Francisco de Quito | Andrea Macias et al |
| EPI_ISL_697795 | Institute of Microbiology, Universidad San Francisco de Quito | Institute of Microbiology, Universidad San Francisco de Quito | Andrea Macias et al |
| EPI_ISL_949202 | Departamento de Microbiología, CDB, Hospital Clínic, Barcelona | SeqCOVID-SPAIN consortium/IBV(CSIC) | Andrea Vergara et al |
| EPI_ISL_1469196 | Klinisk mikrobiologi, Region Västerbotten | CBRN Defence and Security, Swedish Defence Research Agency | Andreas Sjödin et al |
| EPI_ISL_1469197 | Klinisk mikrobiologi, Region Västerbotten | CBRN Defence and Security, Swedish Defence Research Agency | Andreas Sjödin et al |
| EPI_ISL_445259 | CLINICA LAS CONDES S.A. | Instituto de Salud Publica de Chile | Andrés E Castillo et al |
| EPI_ISL_445260 | CLINICA ALEMANA DE SANTIAGO S.A. | Instituto de Salud Publica de Chile | Andrés E Castillo et al |
| EPI_ISL_445318 | C.DE SALUD FAMILIAR PABLO NERUDA | Instituto de Salud Publica de Chile | Andrés E Castillo et al |
| EPI_ISL_445247 | UNIVERSIDAD DE LOS ANDES | Instituto de Salud Publica de Chile | Andrés E Castillo et al |
| EPI_ISL_445367 | ASISTENCIA PUBLICA DR.ALEJANDRO DEL RIO | Instituto de Salud Publica de Chile | Andrés E Castillo et al |
| EPI_ISL_1091784 | LDSP | Universidad Nacional de Colombia - Laboratorio Genómico One Health | Andres F. Cardona-Rios et al |
| EPI_ISL_1091785 | LDSP | Universidad Nacional de Colombia - Laboratorio Genómico One Health | Andres F. Cardona-Rios et al |
| EPI_ISL_1091786 | LDSP | Universidad Nacional de Colombia - Laboratorio Genómico One Health | Andres F. Cardona-Rios et al |
| EPI_ISL_1629760 | LABMÉDICO | Universidad Nacional de Colombia - Laboratorio Genómico One Health | Andres F. Cardona-Rios et al |
| EPI_ISL_480321 | Laboratorio Clínico San José | Charité Virology-University of Costa Rica | Andres Moreira-Soto et al |
| EPI_ISL_480326 | Hospital Nacional de Niños | Charité Virology-University of Costa Rica | Andres Moreira-Soto et al |
| EPI_ISL_480327 | Hospital Nacional de Niños | Charité Virology-University of Costa Rica | Andres Moreira-Soto et al |
| EPI_ISL_480328 | Laboratorio LABIN | Charité Virology-University of Costa Rica | Andres Moreira-Soto et al |
| EPI_ISL_733077 | HELIX LLC | WHO National Influenza Centre Russian Federation | Andrey Komissarov et al |
| EPI_ISL_733078 | HELIX LLC | WHO National Influenza Centre Russian Federation | Andrey Komissarov et al |

| | | | |
|---|---|---|---|
| EPI_ISL_733151 | HELIX LLC | WHO National Influenza Centre Russian Federation | Andrey Komissarov et al |
| EPI_ISL_733433 | HELIX LLC | WHO National Influenza Centre Russian Federation | Andrey Komissarov et al |
| EPI_ISL_1400527 | HELIX LLC | WHO National Influenza Centre Russian Federation | Andrey Komissarov et al |
| EPI_ISL_733452 | HELIX LLC | WHO National Influenza Centre Russian Federation | Andrey Komissarov et al |
| EPI_ISL_733462 | WHO National Influenza Centre Russian Federation | WHO National Influenza Centre Russian Federation | Andrey Komissarov et al |
| EPI_ISL_1372409 | Pathogenic Microorganisms Variability Laboratory | WHO National Influenza Centre Russian Federation | Andrey Komissarov et al |
| EPI_ISL_1491590 | Pathogenic Microorganisms Variability Laboratory | WHO National Influenza Centre Russian Federation | Andrey Komissarov et al |
| EPI_ISL_733166 | Pathogenic Microorganisms Variability Laboratory | WHO National Influenza Centre Russian Federation | Andrey Komissarov et al |
| EPI_ISL_602423 | HELIX LLC | WHO National Influenza Centre Russian Federation | Andrey Komissarov et al |
| EPI_ISL_733098 | HELIX LLC | WHO National Influenza Centre Russian Federation | Andrey Komissarov et al |
| EPI_ISL_733444 | HELIX LLC | WHO National Influenza Centre Russian Federation | Andrey Komissarov et al |
| EPI_ISL_733300 | WHO National Influenza Centre Russian Federation | WHO National Influenza Centre Russian Federation | Andrey Komissarov et al |
| EPI_ISL_596324 | Pathogenic Microorganisms Variability Laboratory | WHO National Influenza Centre Russian Federation | Andrey Komissarov et al |
| EPI_ISL_596337 | Pathogenic Microorganisms Variability Laboratory | WHO National Influenza Centre Russian Federation | Andrey Komissarov et al |
| EPI_ISL_596338 | Pathogenic Microorganisms Variability Laboratory | WHO National Influenza Centre Russian Federation | Andrey Komissarov et al |
| EPI_ISL_596347 | Pathogenic Microorganisms Variability Laboratory | WHO National Influenza Centre Russian Federation | Andrey Komissarov et al |
| EPI_ISL_596348 | Pathogenic Microorganisms Variability Laboratory | WHO National Influenza Centre Russian Federation | Andrey Komissarov et al |
| EPI_ISL_596352 | Pathogenic Microorganisms Variability Laboratory | WHO National Influenza Centre Russian Federation | Andrey Komissarov et al |
| EPI_ISL_770670 | Pathogenic Microorganisms Variability Laboratory | WHO National Influenza Centre Russian Federation | Andrey Komissarov et al |
| EPI_ISL_602408 | HELIX LLC | WHO National Influenza Centre Russian Federation | Andrey Komissarov et al |
| EPI_ISL_1372355 | HELIX LLC | WHO National Influenza Centre Russian Federation | Andrey Komissarov et al |
| EPI_ISL_596266 | WHO National Influenza Centre Russian Federation | WHO National Influenza Centre Russian Federation | Andrey Komissarov et al |
| EPI_ISL_733008 | WHO National Influenza Centre Russian Federation | WHO National Influenza Centre Russian Federation | Andrey Komissarov et al |
| EPI_ISL_733015 | WHO National Influenza Centre Russian Federation | WHO National Influenza Centre Russian Federation | Andrey Komissarov et al |
| EPI_ISL_733016 | WHO National Influenza Centre Russian Federation | WHO National Influenza Centre Russian Federation | Andrey Komissarov et al |
| EPI_ISL_733018 | WHO National Influenza Centre Russian Federation | WHO National Influenza Centre Russian Federation | Andrey Komissarov et al |
| EPI_ISL_733021 | WHO National Influenza Centre Russian Federation | WHO National Influenza Centre Russian Federation | Andrey Komissarov et al |
| EPI_ISL_733314 | HELIX LLC | WHO National Influenza Centre Russian Federation | Andrey Komissarov et al |
| EPI_ISL_524015 | WHO National Influenza Centre Russian Federation | WHO National Influenza Centre Russian Federation | Andrey Komissarov et al |
| EPI_ISL_1372283 | WHO National Influenza Centre Russian Federation | WHO National Influenza Centre Russian Federation | Andrey Komissarov et al |

| | | | |
|---|---|---|---|
| EPI_ISL_1372287 | WHO National Influenza Centre Russian Federation | WHO National Influenza Centre Russian Federation | Andrey Komissarov et al |
| EPI_ISL_1039159 | WHO National Influenza Centre Russian Federation | WHO National Influenza Centre Russian Federation | Andrey Komissarov et al |
| EPI_ISL_732973 | WHO National Influenza Centre Russian Federation | WHO National Influenza Centre Russian Federation | Andrey Komissarov et al |
| EPI_ISL_732972 | WHO National Influenza Centre Russian Federation | WHO National Influenza Centre Russian Federation | Andrey Komissarov et al |
| EPI_ISL_1372345 | HELIX LLC | WHO National Influenza Centre Russian Federation | Andrey Komissarov et al |
| EPI_ISL_1372347 | HELIX LLC | WHO National Influenza Centre Russian Federation | Andrey Komissarov et al |
| EPI_ISL_1372379 | HELIX LLC | WHO National Influenza Centre Russian Federation | Andrey Komissarov et al |
| EPI_ISL_1372392 | HELIX LLC | WHO National Influenza Centre Russian Federation | Andrey Komissarov et al |
| EPI_ISL_1491700 | HELIX LLC | WHO National Influenza Centre Russian Federation | Andrey Komissarov et al |
| EPI_ISL_1664642 | HELIX LLC | WHO National Influenza Centre Russian Federation | Andrey Komissarov et al |
| EPI_ISL_1664718 | HELIX LLC | WHO National Influenza Centre Russian Federation | Andrey Komissarov et al |
| EPI_ISL_1664720 | HELIX LLC | WHO National Influenza Centre Russian Federation | Andrey Komissarov et al |
| EPI_ISL_1664726 | HELIX LLC | WHO National Influenza Centre Russian Federation | Andrey Komissarov et al |
| EPI_ISL_1652545 | HELIX LLC | WHO National Influenza Centre Russian Federation | Andrey Komissarov et al |
| EPI_ISL_1652551 | HELIX LLC | WHO National Influenza Centre Russian Federation | Andrey Komissarov et al |
| EPI_ISL_1652560 | HELIX LLC | WHO National Influenza Centre Russian Federation | Andrey Komissarov et al |
| EPI_ISL_1652563 | HELIX LLC | WHO National Influenza Centre Russian Federation | Andrey Komissarov et al |
| EPI_ISL_1652577 | HELIX LLC | WHO National Influenza Centre Russian Federation | Andrey Komissarov et al |
| EPI_ISL_1652584 | HELIX LLC | WHO National Influenza Centre Russian Federation | Andrey Komissarov et al |
| EPI_ISL_1652601 | HELIX LLC | WHO National Influenza Centre Russian Federation | Andrey Komissarov et al |
| EPI_ISL_1652611 | HELIX LLC | WHO National Influenza Centre Russian Federation | Andrey Komissarov et al |
| EPI_ISL_1652643 | HELIX LLC | WHO National Influenza Centre Russian Federation | Andrey Komissarov et al |
| EPI_ISL_596301 | HELIX LCC | WHO National Influenza Centre Russian Federation | Andrey Komissarov et al |
| EPI_ISL_596279 | HELIX LCC | WHO National Influenza Centre Russian Federation | Andrey Komissarov et al |
| EPI_ISL_1664772 | UMMC-Health | WHO National Influenza Centre Russian Federation | Andrey Komissarov et al |
| EPI_ISL_732996 | UMMC-Health | WHO National Influenza Centre Russian Federation | Andrey Komissarov et al |
| EPI_ISL_430091 | WHO National Influenza Centre Russian Federation | WHO National Influenza Centre Russian Federation | Andrey Komissarov et al |
| EPI_ISL_450277 | WHO National Influenza Centre Russian Federation | WHO National Influenza Centre Russian Federation | Andrey Komissarov et al |
| EPI_ISL_733121 | HELIX LLC | WHO National Influenza Centre Russian Federation | Andrey Komissarov et al |
| EPI_ISL_1300661 | RSUD Pasarminggu | Stem Cell Lab, Universitas Pembangunan Nasional Veteran Jakarta (UPNVJ) | Andri Pramesyanti Pramono et al |

| | | | |
|---|---|---|---|
| EPI_ISL_1300540 | RSUD Pasarminggu | Stem Cell Lab, Universitas Pembangunan Nasional Veteran Jakarta (UPNVJ) | Andri Pramesyanti Pramono et al |
| EPI_ISL_833391 | Klinik Apotek Dein, Jakarta, Indonesia | Biosafety Level-3 Laboratory, Indonesian Institute of Sciences (LIPI) | Andri Wardiana et al |
| EPI_ISL_833508 | Klinik Apotek Dein, Jakarta, Indonesia | Biosafety Level-3 Laboratory, Indonesian Institute of Sciences (LIPI) | Andri Wardiana et al |
| EPI_ISL_1250844 | Tanjungpura University Hospital | Tanjungpura University Hospital | Andriani et al |
| EPI_ISL_912354 | Fondation Congolaise pour la recherche medicale (FCRM), Francine Ntoumi | NGS Competence Center Tuebingen, Institut für Medizinische Mikrobiologie und Hygiene, Universitaetsklinikum Tübingen | Angel Angelov et al |
| EPI_ISL_912361 | Fondation Congolaise pour la recherche medicale (FCRM), Francine Ntoumi | NGS Competence Center Tuebingen, Institut für Medizinische Mikrobiologie und Hygiene, Universitaetsklinikum Tübingen | Angel Angelov et al |
| EPI_ISL_912363 | Fondation Congolaise pour la recherche medicale (FCRM), Francine Ntoumi | NGS Competence Center Tuebingen, Institut für Medizinische Mikrobiologie und Hygiene, Universitaetsklinikum Tübingen | Angel Angelov et al |
| EPI_ISL_912367 | Fondation Congolaise pour la recherche medicale (FCRM), Francine Ntoumi | NGS Competence Center Tuebingen, Institut für Medizinische Mikrobiologie und Hygiene, Universitaetsklinikum Tübingen | Angel Angelov et al |
| EPI_ISL_912368 | Fondation Congolaise pour la recherche medicale (FCRM), Francine Ntoumi | NGS Competence Center Tuebingen, Institut für Medizinische Mikrobiologie und Hygiene, Universitaetsklinikum Tübingen | Angel Angelov et al |
| EPI_ISL_912375 | Fondation Congolaise pour la recherche medicale (FCRM), Francine Ntoumi | NGS Competence Center Tuebingen, Institut für Medizinische Mikrobiologie und Hygiene, Universitaetsklinikum Tübingen | Angel Angelov et al |
| EPI_ISL_912376 | Fondation Congolaise pour la recherche medicale (FCRM), Francine Ntoumi | NGS Competence Center Tuebingen, Institut für Medizinische Mikrobiologie und Hygiene, Universitaetsklinikum Tübingen | Angel Angelov et al |
| EPI_ISL_912377 | Fondation Congolaise pour la recherche medicale (FCRM), Francine Ntoumi | NGS Competence Center Tuebingen, Institut für Medizinische Mikrobiologie und Hygiene, Universitaetsklinikum Tübingen | Angel Angelov et al |
| EPI_ISL_912380 | Fondation Congolaise pour la recherche medicale (FCRM), Francine Ntoumi | NGS Competence Center Tuebingen, Institut für Medizinische Mikrobiologie und Hygiene, Universitaetsklinikum Tübingen | Angel Angelov et al |
| EPI_ISL_912387 | Fondation Congolaise pour la recherche medicale (FCRM), Francine Ntoumi | NGS Competence Center Tuebingen, Institut für Medizinische Mikrobiologie und Hygiene, Universitaetsklinikum Tübingen | Angel Angelov et al |
| EPI_ISL_912388 | Fondation Congolaise pour la recherche medicale (FCRM), Francine Ntoumi | NGS Competence Center Tuebingen, Institut für Medizinische Mikrobiologie und Hygiene, Universitaetsklinikum Tübingen | Angel Angelov et al |
| EPI_ISL_912389 | Fondation Congolaise pour la recherche medicale (FCRM), Francine Ntoumi | NGS Competence Center Tuebingen, Institut für Medizinische Mikrobiologie und Hygiene, Universitaetsklinikum Tübingen | Angel Angelov et al |
| EPI_ISL_912394 | Fondation Congolaise pour la recherche medicale (FCRM), Francine Ntoumi | NGS Competence Center Tuebingen, Institut für Medizinische Mikrobiologie und Hygiene, Universitaetsklinikum Tübingen | Angel Angelov et al |
| EPI_ISL_581487 | Fondation Congolaise pour la recherche medicale (FCRM) | NGS Competence Center Tübingen, Institut für Medizinische Mikrobiologie und Hygiene, Universitätsklinikum Tübingen | Angel Angelov et al |
| EPI_ISL_581489 | Fondation Congolaise pour la recherche medicale (FCRM) | NGS Competence Center Tübingen, Institut für Medizinische Mikrobiologie und Hygiene, Universitätsklinikum Tübingen | Angel Angelov et al |
| EPI_ISL_581455 | Fondation Congolaise pour la recherche medicale (FCRM) | NGS Competence Center Tübingen, Institut für Medizinische Mikrobiologie und Hygiene, Universitätsklinikum Tübingen | Angel Angelov et al |
| EPI_ISL_581492 | Fondation Congolaise pour la recherche medicale (FCRM) | NGS Competence Center Tübingen, Institut für Medizinische Mikrobiologie und Hygiene, Universitätsklinikum Tübingen | Angel Angelov et al |
| EPI_ISL_581493 | Fondation Congolaise pour la recherche medicale (FCRM) | NGS Competence Center Tübingen, Institut für Medizinische Mikrobiologie und Hygiene, Universitätsklinikum Tübingen | Angel Angelov et al |
| EPI_ISL_485810 | Institut für Virologie und Epidemiologie der Viruskrankheiten, Universitätsklinikum Tübingen | NGS Competence Center Tübingen, Institut für Medizinische Mikrobiologie und Hygiene, Universitätsklinikum Tübingen | Angelov et al. |

| | | | |
|---|---|---|---|
| EPI_ISL_1020195 | Melania Hospital, Bogor , West Java | Biosafety Level-3 Laboratory, Indonesian Institute of Sciences (LIPI) | Anggia Prasetyoputri et al |
| EPI_ISL_1020192 | Melania Hospital, Bagor , west Java | Biosafety Level-3 Laboratory, Indonesian Institute of Sciences (LIPI) | Anik Budhi Dharmayanthi et al |
| EPI_ISL_1020197 | Dinkes, Bogor, West Java | Biosafety Level-3 Laboratory, Indonesian Institute of Sciences (LIPI) | Anik Budhi Dharmayanthi et al |
| EPI_ISL_419562 | Laboratoire National de Santé, Microbiology, Virology | Laboratoire National de Santé, Microbiology, Epidemiology and Microbial Genomics | Anke Wienecke-Baldacchino et al |
| EPI_ISL_1383410 | Laboratoire national de sante, Microbiology, Virology | Laboratoire national de sante, Microbiology, Microbial Genomics Platform | Anke Wienecke-Baldacchino et al |
| EPI_ISL_739793 | Laboratoire national de santé, Microbiology, Virology | Laboratoire national de santé, Microbiology, Microbial Genomics Platform | Anke Wienecke-Baldacchino et al |
| EPI_ISL_419565 | Laboratoire National de Santé, Microbiology, Virology | Laboratoire National de Santé, Microbiology, Epidemiology and Microbial Genomics | Anke Wienecke-Baldacchino et al |
| EPI_ISL_1383771 | Laboratoire national de sante, Microbiology, Virology | Laboratoire national de sante, Microbiology, Microbial Genomics Platform | Anke Wienecke-Baldacchino et al |
| EPI_ISL_739835 | Laboratoire national de santé, Microbiology, Virology | Laboratoire national de santé, Microbiology, Microbial Genomics Platform | Anke Wienecke-Baldacchino et al |
| EPI_ISL_911195 | Laboratoire national de sante, Microbiology, Virology | Laboratoire national de sante, Microbiology, Microbial Genomics Platform | Anke Wienecke-Baldacchino et al |
| EPI_ISL_739880 | Laboratoire national de santé, Microbiology, Virology | Laboratoire national de santé, Microbiology, Epidemiology and Microbial Genomics | Anke Wienecke-Baldacchino et al |
| EPI_ISL_1384017 | Hospital Center Emile Mayrisch | Laboratoire national de sante, Microbiology, Microbial Genomics Platform | Anke Wienecke-Baldacchino et al |
| EPI_ISL_1383653 | Laboratoire national de sante, Microbiology, Virology | Laboratoire national de sante, Microbiology, Microbial Genomics Platform | Anke Wienecke-Baldacchino et al |
| EPI_ISL_739934 | Laboratoire national de santé, Microbiology, Virology | Laboratoire national de santé, Microbiology, Microbial Genomics Platform | Anke Wienecke-Baldacchino et al |
| EPI_ISL_1384349 | Laboratoires d'analyses medicales - Ketterthill | Laboratoire national de sante, Microbiology, Microbial Genomics Platform | Anke Wienecke-Baldacchino et al |
| EPI_ISL_739947 | Laboratoire national de santé, Microbiology, Virology | Laboratoire national de santé, Microbiology, Microbial Genomics Platform | Anke Wienecke-Baldacchino et al |
| EPI_ISL_739956 | Laboratoire national de santé, Microbiology, Virology | Laboratoire national de santé, Microbiology, Microbial Genomics Platform | Anke Wienecke-Baldacchino et al |
| EPI_ISL_739977 | Laboratoire national de santé, Microbiology, Virology | Laboratoire national de santé, Microbiology, Microbial Genomics Platform | Anke Wienecke-Baldacchino et al |
| EPI_ISL_739978 | Laboratoire national de santé, Microbiology, Virology | Laboratoire national de santé, Microbiology, Microbial Genomics Platform | Anke Wienecke-Baldacchino et al |
| EPI_ISL_739987 | Laboratoire national de santé, Microbiology, Virology | Laboratoire national de santé, Microbiology, Microbial Genomics Platform | Anke Wienecke-Baldacchino et al |
| EPI_ISL_740013 | Laboratoire national de santé, Microbiology, Virology | Laboratoire national de santé, Microbiology, Microbial Genomics Platform | Anke Wienecke-Baldacchino et al |
| EPI_ISL_1498404 | BioneXt Lab | Laboratoire national de sante, Microbiology, Microbial Genomics Platform | Anke Wienecke-Baldacchino et al |
| EPI_ISL_740098 | Laboratoire national de santé, Microbiology, Virology | Laboratoire national de santé, Microbiology, Microbial Genomics Platform | Anke Wienecke-Baldacchino et al |
| EPI_ISL_740129 | Laboratoire national de santé, Microbiology, Virology | Laboratoire national de santé, Microbiology, Epidemiology and Microbial Genomics | Anke Wienecke-Baldacchino et al |
| EPI_ISL_740202 | Laboratoire national de santé, Microbiology, Virology | Laboratoire national de santé, Microbiology, Microbial Genomics Platform | Anke Wienecke-Baldacchino et al |
| EPI_ISL_1383541 | Laboratoire national de sante, Microbiology, Virology | Laboratoire national de sante, Microbiology, Microbial Genomics Platform | Anke Wienecke-Baldacchino et al |
| EPI_ISL_740259 | Laboratoire national de santé, Microbiology, Virology | Laboratoire national de santé, Microbiology, Microbial Genomics Platform | Anke Wienecke-Baldacchino et al |
| EPI_ISL_421735 | Laboratoire National de Sante, Microbiology, Virology | Laboratoire National de Sante, Microbiology, Epidemiology and Microbial Genomics | Anke Wienecke-Baldacchino et al |

| | | | |
|---|---|---|---|
| EPI_ISL_740289 | Laboratoire national de santé, Microbiology, Virology | Laboratoire national de santé, Microbiology, Microbial Genomics Platform | Anke Wienecke-Baldacchino et al |
| EPI_ISL_459896 | Laboratoire National de Sante, Microbiology, Virology | Laboratoire National de Sante, Microbiology, Epidemiology and Microbial Genomics | Anke Wienecke-Baldacchino et al |
| EPI_ISL_459898 | Laboratoire National de Sante, Microbiology, Virology | Laboratoire National de Sante, Microbiology, Epidemiology and Microbial Genomics | Anke Wienecke-Baldacchino et al |
| EPI_ISL_740408 | Laboratoire national de santé, Microbiology, Virology | Laboratoire national de santé, Microbiology, Microbial Genomics Platform | Anke Wienecke-Baldacchino et al |
| EPI_ISL_740413 | Laboratoire national de santé, Microbiology, Virology | Laboratoire national de santé, Microbiology, Microbial Genomics Platform | Anke Wienecke-Baldacchino et al |
| EPI_ISL_740486 | Laboratoire national de santé, Microbiology, Virology | Laboratoire national de santé, Microbiology, Microbial Genomics Platform | Anke Wienecke-Baldacchino et al |
| EPI_ISL_434509 | Laboratoire National de Sante, Microbiology, Virology | Laboratoire National de Sante, Microbiology, Epidemiology and Microbial Genomics | Anke Wienecke-Baldacchino et al |
| EPI_ISL_744135 | Laboratoire national de santé, Microbiology, Virology | Laboratoire national de santé, Microbiology, Microbial Genomics Platform | Anke Wienecke-Baldacchino et al |
| EPI_ISL_744139 | Laboratoire national de santé, Microbiology, Virology | Laboratoire national de santé, Microbiology, Microbial Genomics Platform | Anke Wienecke-Baldacchino et al |
| EPI_ISL_1522973 | Laboratoire national de sante, Microbiology, Virology | Laboratoire national de sante, Microbiology, Microbial Genomics Platform | Anke Wienecke-Baldacchino et al |
| EPI_ISL_429738 | Laboratoire National de Sante, Microbiology, Virology | Laboratoire National de Sante, Microbiology, Epidemiology and Microbial Genomics | Anke Wienecke-Baldacchino et al |
| EPI_ISL_744245 | Laboratoire national de santé, Microbiology, Virology | Laboratoire national de santé, Microbiology, Microbial Genomics Platform | Anke Wienecke-Baldacchino et al |
| EPI_ISL_429774 | Laboratoire National de Sante, Microbiology, Virology | Laboratoire National de Sante, Microbiology, Epidemiology and Microbial Genomics | Anke Wienecke-Baldacchino et al |
| EPI_ISL_744291 | Laboratoire national de santé, Microbiology, Virology | Laboratoire national de santé, Microbiology, Microbial Genomics Platform | Anke Wienecke-Baldacchino et al |
| EPI_ISL_1383729 | Laboratoire national de sante, Microbiology, Virology | Laboratoire national de sante, Microbiology, Microbial Genomics Platform | Anke Wienecke-Baldacchino et al |
| EPI_ISL_429731 | Laboratoire National de Sante, Microbiology, Virology | Laboratoire National de Sante, Microbiology, Epidemiology and Microbial Genomics | Anke Wienecke-Baldacchino et al |
| EPI_ISL_429743 | Laboratoire National de Sante, Microbiology, Virology | Laboratoire National de Sante, Microbiology, Epidemiology and Microbial Genomics | Anke Wienecke-Baldacchino et al |
| EPI_ISL_1384478 | Laboratoires d'analyses medicales - Ketterthill | Laboratoire national de sante, Microbiology, Microbial Genomics Platform | Anke Wienecke-Baldacchino et al |
| EPI_ISL_459901 | Laboratoire National de Sante, Microbiology, Virology | Laboratoire National de Sante, Microbiology, Epidemiology and Microbial Genomics | Anke Wienecke-Baldacchino et al |
| EPI_ISL_1384382 | Laboratoires d'analyses medicales - Ketterthill | Laboratoire national de sante, Microbiology, Microbial Genomics Platform | Anke Wienecke-Baldacchino et al |
| EPI_ISL_1384184 | Laboratoires d'analyses medicales - Ketterthill | Laboratoire national de sante, Microbiology, Microbial Genomics Platform | Anke Wienecke-Baldacchino et al |
| EPI_ISL_744480 | Laboratoire national de santé, Microbiology, Virology | Laboratoire national de santé, Microbiology, Microbial Genomics Platform | Anke Wienecke-Baldacchino et al |
| EPI_ISL_1384156 | Laboratoire national de sante, Microbiology, Virology | Laboratoire national de sante, Microbiology, Microbial Genomics Platform | Anke Wienecke-Baldacchino et al |
| EPI_ISL_744561 | Laboratoire national de santé, Microbiology, Virology | Laboratoire national de santé, Microbiology, Microbial Genomics Platform | Anke Wienecke-Baldacchino et al |
| EPI_ISL_744574 | Laboratoire national de santé, Microbiology, Virology | Laboratoire national de santé, Microbiology, Microbial Genomics Platform | Anke Wienecke-Baldacchino et al |
| EPI_ISL_744577 | Laboratoire national de santé, Microbiology, Virology | Laboratoire national de santé, Microbiology, Microbial Genomics Platform | Anke Wienecke-Baldacchino et al |
| EPI_ISL_445062 | Laboratoire National de Sante, Microbiology, Virology | Laboratoire National de Sante, Microbiology, Epidemiology and Microbial Genomics | Anke Wienecke-Baldacchino et al |

| | | | |
|---|---|---|---|
| EPI_ISL_459906 | Laboratoire National de Sante, Microbiology, Virology | Laboratoire National de Sante, Microbiology, Epidemiology and Microbial Genomics | Anke Wienecke-Baldacchino et al |
| EPI_ISL_428940 | Laboratoire National de Sante, Microbiology, Virology | Laboratoire National de Sante, Microbiology, Epidemiology and Microbial Genomics | Anke Wienecke-Baldacchino et al |
| EPI_ISL_1498386 | BioneXt Lab | Laboratoire national de sante, Microbiology, Microbial Genomics Platform | Anke Wienecke-Baldacchino et al |
| EPI_ISL_1522970 | Laboratoire national de sante, Microbiology, Virology | Laboratoire national de sante, Microbiology, Microbial Genomics Platform | Anke Wienecke-Baldacchino et al |
| EPI_ISL_1384419 | Laboratoires d'analyses medicales - Ketterthill | Laboratoire national de sante, Microbiology, Microbial Genomics Platform | Anke Wienecke-Baldacchino et al |
| EPI_ISL_744674 | Laboratoire national de santé, Microbiology, Virology | Laboratoire national de santé, Microbiology, Microbial Genomics Platform | Anke Wienecke-Baldacchino et al |
| EPI_ISL_744766 | Laboratoire national de santé, Microbiology, Virology | Laboratoire national de santé, Microbiology, Microbial Genomics Platform | Anke Wienecke-Baldacchino et al |
| EPI_ISL_744773 | Laboratoire national de santé, Microbiology, Virology | Laboratoire national de santé, Microbiology, Microbial Genomics Platform | Anke Wienecke-Baldacchino et al |
| EPI_ISL_1384207 | Laboratoires d'analyses medicales - Ketterthill | Laboratoire national de sante, Microbiology, Microbial Genomics Platform | Anke Wienecke-Baldacchino et al |
| EPI_ISL_1384442 | Laboratoires d'analyses medicales - Ketterthill | Laboratoire national de sante, Microbiology, Microbial Genomics Platform | Anke Wienecke-Baldacchino et al |
| EPI_ISL_744958 | Laboratoire national de santé, Microbiology, Virology | Laboratoire national de santé, Microbiology, Microbial Genomics Platform | Anke Wienecke-Baldacchino et al |
| EPI_ISL_910820 | Laboratoire national de sante, Microbiology, Virology | Laboratoire national de sante, Microbiology, Microbial Genomics Platform | Anke Wienecke-Baldacchino et al |
| EPI_ISL_745023 | Laboratoire national de santé, Microbiology, Virology | Laboratoire national de santé, Microbiology, Microbial Genomics Platform | Anke Wienecke-Baldacchino et al |
| EPI_ISL_416480 | R. G. Lugar Center for Public Health Research, National Center for Disease Control and Public Health (NCDC) of Georgia. | R. G. Lugar Center for Public Health Research, National Center for Disease Control and Public Health (NCDC) of Georgia. | Ann Machablishvili et al |
| EPI_ISL_935788 | Cadham Provincial laboratory | National Microbiology Laboratory (NML) | Anna Majer et al |
| EPI_ISL_935789 | Cadham Provincial laboratory | National Microbiology Laboratory (NML) | Anna Majer et al |
| EPI_ISL_935828 | Cadham Provincial laboratory | National Microbiology Laboratory (NML) | Anna Majer et al |
| EPI_ISL_935834 | Cadham Provincial laboratory | National Microbiology Laboratory (NML) | Anna Majer et al |
| EPI_ISL_935843 | Cadham Provincial laboratory | National Microbiology Laboratory (NML) | Anna Majer et al |
| EPI_ISL_935847 | Cadham Provincial laboratory | National Microbiology Laboratory (NML) | Anna Majer et al |
| EPI_ISL_935848 | Cadham Provincial laboratory | National Microbiology Laboratory (NML) | Anna Majer et al |
| EPI_ISL_935872 | Cadham Provincial laboratory | National Microbiology Laboratory (NML) | Anna Majer et al |
| EPI_ISL_935873 | Cadham Provincial laboratory | National Microbiology Laboratory (NML) | Anna Majer et al |
| EPI_ISL_935875 | Cadham Provincial laboratory | National Microbiology Laboratory (NML) | Anna Majer et al |
| EPI_ISL_1594113 | MB-Cadham Provincial laboratory | National Microbiology Laboratory (NML) | Anna Majer et al |
| EPI_ISL_1594051 | MB-Cadham Provincial laboratory | National Microbiology Laboratory (NML) | Anna Majer et al |
| EPI_ISL_1258081 | MB-Cadham Provincial laboratory | National Microbiology Laboratory (NML) | Anna Majer et al |
| EPI_ISL_1258090 | MB-Cadham Provincial laboratory | National Microbiology Laboratory (NML) | Anna Majer et al |
| EPI_ISL_1594071 | MB-Cadham Provincial laboratory | National Microbiology Laboratory (NML) | Anna Majer et al |

| | | | |
|---|---|---|---|
| EPI_ISL_582342 | Cadham Provincial Laboratory | National Microbiology Laboratory (NML) | Anna Majer et al |
| EPI_ISL_1164956 | Hôpital Georges L. Dumont | National Microbiology Laboratory (NML) | Anna Majer et al |
| EPI_ISL_1272270 | NB-Hôpital Georges L. Dumont | National Microbiology Laboratory (NML) | Anna Majer et al |
| EPI_ISL_1272256 | NB-Hôpital Georges L. Dumont | National Microbiology Laboratory (NML) | Anna Majer et al |
| EPI_ISL_1588150 | NB-Hôpital Georges L. Dumont | National Microbiology Laboratory (NML) | Anna Majer et al |
| EPI_ISL_1588200 | NB-Hôpital Georges L. Dumont | National Microbiology Laboratory (NML) | Anna Majer et al |
| EPI_ISL_1588269 | NB-Hôpital Georges L. Dumont | National Microbiology Laboratory (NML) | Anna Majer et al |
| EPI_ISL_481756 | Dr. Georges-L.-Dumont University Hospital Centre | National Microbiology Laboratory | Anna Majer et al |
| EPI_ISL_1278151 | NL-Dr. Leonard A. Miller Centre for Health Services | National Microbiology Laboratory (NML) | Anna Majer et al |
| EPI_ISL_1055752 | QEII Health Sciences Centre | National Microbiology Laboratory (NML) | Anna Majer et al |
| EPI_ISL_1666730 | NS-QEII Health Sciences Centre | National Microbiology Laboratory (NML) | Anna Majer et al |
| EPI_ISL_1055479 | QEII Health Sciences Centre | National Microbiology Laboratory (NML) | Anna Majer et al |
| EPI_ISL_856714 | Infectious Diseases Unit, Department of Internal Medicine, Azienda Ospedaliera-Universitaria di Padova | Laboratory of Infectious Diseases, Department of Biomedical and Clinical Sciences L. Sacco, University of Milan | Anna Maria Cattelan et al |
| EPI_ISL_856715 | Infectious Diseases Unit, Department of Internal Medicine, Azienda Ospedaliera-Universitaria di Padova | Laboratory of Infectious Diseases, Department of Biomedical and Clinical Sciences L. Sacco, University of Milan | Anna Maria Cattelan et al |
| EPI_ISL_856721 | Infectious Diseases Unit, Department of Internal Medicine, Azienda Ospedaliera-Universitaria di Padova | Laboratory of Infectious Diseases, Department of Biomedical and Clinical Sciences L. Sacco, University of Milan | Anna Maria Cattelan et al |
| EPI_ISL_406034 | California Department of Public Health | Pathogen Discovery, Respiratory Viruses Branch, Division of Viral Diseases, Centers for Disease Control and Prevention | Anna Uehara et al |
| EPI_ISL_406036 | California Department of Public Health | Pathogen Discovery, Respiratory Viruses Branch, Division of Viral Diseases, Centers for Disease Control and Prevention | Anna Uehara et al |
| EPI_ISL_419560 | FL Bureau of Public Health Laboratories-Tampa | Pathogen Discovery, Respiratory Viruses Branch, Division of Viral Diseases, Centers for Disease Control and Prevention | Anna Uehara et al |
| EPI_ISL_454481 | Karolinska Universitetslaboratoriet | The Public Health Agency of Sweden | Anna-Malin Linde et al |
| EPI_ISL_455902 | Halmstad klinisk mikrobiologi | The Public Health Agency of Sweden | Anna-Malin Linde et al |
| EPI_ISL_534224 | Universitetssjukhuset i Linkoping | The Public Health Agency of Sweden | Anna-Malin Linde et al |
| EPI_ISL_582789 | Orebro klinisk mikrobiologi | The Public Health Agency of Sweden | Anna-Malin Linde et al |
| EPI_ISL_582791 | Orebro klinisk mikrobiologi | The Public Health Agency of Sweden | Anna-Malin Linde et al |
| EPI_ISL_623085 | Klinisk mikrobiologi, Skanes universitetssjukhus, Lund | The Public Health Agency of Sweden | Anna-Malin Linde et al |
| EPI_ISL_648189 | The Public Health Agency of Sweden | The Public Health Agency of Sweden | Anna-Malin Linde et al |
| EPI_ISL_560973 | Klinisk mikrobiologi NAL Trollhattan | The Public Health Agency of Sweden | Anna-Malin Linde et al |
| EPI_ISL_623087 | Klinisk mikrobiologi NAL Trollhattan | The Public Health Agency of Sweden | Anna-Malin Linde et al |
| EPI_ISL_582797 | Klinisk mikrobiologi Vasternorrland | The Public Health Agency of Sweden | Anna-Malin Linde et al |

| | | | |
|---|---|---|---|
| EPI_ISL_582798 | Klinisk mikrobiologi Vasternorrland | The Public Health Agency of Sweden | Anna-Malin Linde et al |
| EPI_ISL_582800 | Klinisk mikrobiologi Vasternorrland | The Public Health Agency of Sweden | Anna-Malin Linde et al |
| EPI_ISL_534248 | Laboratoriemedicin Vasternorrland | The Public Health Agency of Sweden | Anna-Malin Linde et al |
| EPI_ISL_582807 | Klinisk mikrobiologi SAS Boras | The Public Health Agency of Sweden | Anna-Malin Linde et al |
| EPI_ISL_648163 | Gavle klinisk mikrobiologi | The Public Health Agency of Sweden | Anna-Malin Linde et al |
| EPI_ISL_648167 | Eskilstuna | The Public Health Agency of Sweden | Anna-Malin Linde et al |
| EPI_ISL_648166 | Eskilstuna | The Public Health Agency of Sweden | Anna-Malin Linde et al |
| EPI_ISL_913478 | Klinisk mikrobiologi | The Public Health Agency of Sweden | Anna-Malin Linde et al |
| EPI_ISL_1008415 | Klinisk mikrobiologi | The Public Health Agency of Sweden | Anna-Malin Linde et al |
| EPI_ISL_913475 | Klinisk mikrobiologi | The Public Health Agency of Sweden | Anna-Malin Linde et al |
| EPI_ISL_913363 | Klinisk mikrobiologi | The Public Health Agency of Sweden | Anna-Malin Linde et al |
| EPI_ISL_1008444 | Klinisk mikrobiologi | The Public Health Agency of Sweden | Anna-Malin Linde et al |
| EPI_ISL_1008548 | Klinisk mikrobiologi | The Public Health Agency of Sweden | Anna-Malin Linde et al |
| EPI_ISL_430846 | General Intensive Care Unit, Raymond Poincaré Hospital (AP-HP), Lab Inflammation & Infection, U1173 University Paris Saclay-UVSQ/INSERM, Garches, France. | Institut Pasteur, Laboratory for Urgent Response to biological Threats | Annane Djillali et al |
| EPI_ISL_1593977 | CHC Andrée Rosemon | Institut Pasteur de la Guyane | Anne Lavergne et al |
| EPI_ISL_1593978 | CHC Andrée Rosemon | Institut Pasteur de la Guyane | Anne Lavergne et al |
| EPI_ISL_1627097 | CHC Andrée Rosemon | Institut Pasteur de la Guyane | Anne Lavergne et al |
| EPI_ISL_1628333 | Laboratoire Carage | Institut Pasteur de la Guyane | Anne Lavergne et al |
| EPI_ISL_1628334 | CHC Andrée Rosemon | Institut Pasteur de la Guyane | Anne Lavergne et al |
| EPI_ISL_1628336 | CNR Institut Pasteur de la Guyane | Institut Pasteur de la Guyane | Anne Lavergne et al |
| EPI_ISL_1628338 | CNR Institut Pasteur de la Guyane | Institut Pasteur de la Guyane | Anne Lavergne et al |
| EPI_ISL_812267 | Landstuhl Regional Medical Center | United States Air Force School of Aerospace Medicine | Anthony Fries et al |
| EPI_ISL_812271 | Landstuhl Regional Medical Center | United States Air Force School of Aerospace Medicine | Anthony Fries et al |
| EPI_ISL_812275 | Landstuhl Regional Medical Center | United States Air Force School of Aerospace Medicine | Anthony Fries et al |
| EPI_ISL_812280 | Landstuhl Regional Medical Center | United States Air Force School of Aerospace Medicine | Anthony Fries et al |
| EPI_ISL_738323 | Landstuhl Regional Medical Center | United States Air Force School of Aerospace Medicine | Anthony Fries et al |
| EPI_ISL_738321 | Landstuhl Regional Medical Center | United States Air Force School of Aerospace Medicine | Anthony Fries et al |
| EPI_ISL_1038791 | US Air Force School of Aerospace Medicine | US Air Force School of Aerospace Medicine | Anthony Fries et al |
| EPI_ISL_568946 | MEPHI, Aix Marseille University | MEPHI, Aix Marseille University | Anthony LEVASSEUR et al |

| | | | |
|---|---|---|---|
| EPI_ISL_568968 | MEPHI, Aix Marseille University | MEPHI, Aix Marseille University | Anthony LEVASSEUR et al |
| EPI_ISL_568978 | MEPHI, Aix Marseille University | MEPHI, Aix Marseille University | Anthony LEVASSEUR et al |
| EPI_ISL_568983 | MEPHI, Aix Marseille University | MEPHI, Aix Marseille University | Anthony LEVASSEUR et al |
| EPI_ISL_569007 | MEPHI, Aix Marseille University | MEPHI, Aix Marseille University | Anthony LEVASSEUR et al |
| EPI_ISL_569055 | MEPHI, Aix Marseille University | MEPHI, Aix Marseille University | Anthony LEVASSEUR et al |
| EPI_ISL_569095 | MEPHI, Aix Marseille University | MEPHI, Aix Marseille University | Anthony LEVASSEUR et al |
| EPI_ISL_569171 | MEPHI, Aix Marseille University | MEPHI, Aix Marseille University | Anthony LEVASSEUR et al |
| EPI_ISL_569218 | MEPHI, Aix Marseille University | MEPHI, Aix Marseille University | Anthony LEVASSEUR et al |
| EPI_ISL_569241 | MEPHI, Aix Marseille University | MEPHI, Aix Marseille University | Anthony LEVASSEUR et al |
| EPI_ISL_982230 | MEPHI, Aix Marseille University | MEPHI, Aix Marseille University | Anthony LEVASSEUR et al |
| EPI_ISL_900144 | MEPHI, Aix Marseille University | MEPHI, Aix Marseille University | Anthony LEVASSEUR et al |
| EPI_ISL_900163 | MEPHI, Aix Marseille University | MEPHI, Aix Marseille University | Anthony LEVASSEUR et al |
| EPI_ISL_900165 | MEPHI, Aix Marseille University | MEPHI, Aix Marseille University | Anthony LEVASSEUR et al |
| EPI_ISL_900080 | MEPHI, Aix Marseille University | MEPHI, Aix Marseille University | Anthony LEVASSEUR et al |
| EPI_ISL_900192 | MEPHI, Aix Marseille University | MEPHI, Aix Marseille University | Anthony LEVASSEUR et al |
| EPI_ISL_896130 | MEPHI, Aix Marseille University | MEPHI, Aix Marseille University | Anthony LEVASSEUR et al |
| EPI_ISL_900082 | MEPHI, Aix Marseille University | MEPHI, Aix Marseille University | Anthony LEVASSEUR et al |
| EPI_ISL_900168 | MEPHI, Aix Marseille University | MEPHI, Aix Marseille University | Anthony LEVASSEUR et al |
| EPI_ISL_900176 | MEPHI, Aix Marseille University | MEPHI, Aix Marseille University | Anthony LEVASSEUR et al |
| EPI_ISL_900196 | MEPHI, Aix Marseille University | MEPHI, Aix Marseille University | Anthony LEVASSEUR et al |
| EPI_ISL_900169 | MEPHI, Aix Marseille University | MEPHI, Aix Marseille University | Anthony LEVASSEUR et al |
| EPI_ISL_896134 | MEPHI, Aix Marseille University | MEPHI, Aix Marseille University | Anthony LEVASSEUR et al |
| EPI_ISL_900181 | MEPHI, Aix Marseille University | MEPHI, Aix Marseille University | Anthony LEVASSEUR et al |
| EPI_ISL_900125 | MEPHI, Aix Marseille University | MEPHI, Aix Marseille University | Anthony LEVASSEUR et al |
| EPI_ISL_804478 | MEPHI, Aix Marseille University | MEPHI, Aix Marseille University | Anthony LEVASSEUR et al |
| EPI_ISL_644375 | MEPHI, Aix Marseille University | MEPHI, Aix Marseille University | Anthony LEVASSEUR et al |
| EPI_ISL_644475 | MEPHI, Aix Marseille University | MEPHI, Aix Marseille University | Anthony LEVASSEUR et al |
| EPI_ISL_940274 | Hôpital Bichat Claude Bernard, Laboratoire de Virologie | IAME UMR1137 Inserm, Université de Paris, Hôpital Bichat | Antoine Bridier-Nahmias et al |
| EPI_ISL_940552 | Hôpital Bichat Claude Bernard, Laboratoire de Virologie | IAME UMR1137 Inserm, Université de Paris, Hôpital Bichat | Antoine Bridier-Nahmias et al |
| EPI_ISL_940530 | Hôpital Bichat Claude Bernard, Laboratoire de Virologie | IAME UMR1137 Inserm, Université de Paris, Hôpital Bichat | Antoine Bridier-Nahmias et al |

| | | | |
|---|---|---|---|
| EPI_ISL_940372 | Hôpital Bichat Claude Bernard, Laboratoire de Virologie | IAME UMR1137 Inserm, Université de Paris, Hôpital Bichat | Antoine Bridier-Nahmias et al |
| EPI_ISL_940325 | Hôpital Bichat Claude Bernard, Laboratoire de Virologie | IAME UMR1137 Inserm, Université de Paris, Hôpital Bichat | Antoine Bridier-Nahmias et al |
| EPI_ISL_940186 | Hôpital Bichat Claude Bernard, Laboratoire de Virologie | IAME UMR1137 Inserm, Université de Paris, Hôpital Bichat | Antoine Bridier-Nahmias et al |
| EPI_ISL_940175 | Hôpital Bichat Claude Bernard, Laboratoire de Virologie | IAME UMR1137 Inserm, Université de Paris, Hôpital Bichat | Antoine Bridier-Nahmias et al |
| EPI_ISL_940177 | Hôpital Bichat Claude Bernard, Laboratoire de Virologie | IAME UMR1137 Inserm, Université de Paris, Hôpital Bichat | Antoine Bridier-Nahmias et al |
| EPI_ISL_940165 | Hôpital Bichat Claude Bernard, Laboratoire de Virologie | IAME UMR1137 Inserm, Université de Paris, Hôpital Bichat | Antoine Bridier-Nahmias et al |
| EPI_ISL_940340 | Hôpital Bichat Claude Bernard, Laboratoire de Virologie | IAME UMR1137 Inserm, Université de Paris, Hôpital Bichat | Antoine Bridier-Nahmias et al |
| EPI_ISL_940427 | Hôpital Bichat Claude Bernard, Laboratoire de Virologie | IAME UMR1137 Inserm, Université de Paris, Hôpital Bichat | Antoine Bridier-Nahmias et al |
| EPI_ISL_683835 | CICM | Malaria Research and Training Center (MRTC-Parasito) | Antoine Dara et al |
| EPI_ISL_1135058 | ORIAPOLE - BACTERIOLOGIE | CNR Virus des Infections Respiratoires - France SUD | Antonin Bal et al |
| EPI_ISL_660360 | CHU Clermont-Ferrand | CNR Virus des Infections Respiratoires - France SUD | Antonin Bal et al |
| EPI_ISL_1371872 | CERBALIANCE PROVENCE | CNR Virus des Infections Respiratoires - France SUD | Antonin Bal et al |
| EPI_ISL_645213 | Unité des Virus Émergents | CNR Virus des Infections Respiratoires - France SUD | Antonin Bal et al |
| EPI_ISL_913666 | Cerballiance | CNR Virus des Infections Respiratoires - France SUD | Antonin Bal et al |
| EPI_ISL_693387 | CHU de Nice - Hôpital Archet 2 | CNR Virus des Infections Respiratoires - France SUD | Antonin Bal et al |
| EPI_ISL_1336649 | HOPITAL PRINCESSE GRACE | CNR Virus des Infections Respiratoires - France SUD | Antonin Bal et al |
| EPI_ISL_1135056 | C.H. PRINCESSE GRACE | CNR Virus des Infections Respiratoires - France SUD | Antonin Bal et al |
| EPI_ISL_1371898 | C H DE LA POLYNESIE FRANCAISE | CNR Virus des Infections Respiratoires - France SUD | Antonin Bal et al |
| EPI_ISL_1371903 | C H DE LA POLYNESIE FRANCAISE | CNR Virus des Infections Respiratoires - France SUD | Antonin Bal et al |
| EPI_ISL_1371896 | C H DE LA POLYNESIE FRANCAISE | CNR Virus des Infections Respiratoires - France SUD | Antonin Bal et al |
| EPI_ISL_1371902 | C H DE LA POLYNESIE FRANCAISE | CNR Virus des Infections Respiratoires - France SUD | Antonin Bal et al |
| EPI_ISL_1371900 | C H DE LA POLYNESIE FRANCAISE | CNR Virus des Infections Respiratoires - France SUD | Antonin Bal et al |
| EPI_ISL_1371904 | C H DE LA POLYNESIE FRANCAISE | CNR Virus des Infections Respiratoires - France SUD | Antonin Bal et al |
| EPI_ISL_1371901 | C H DE LA POLYNESIE FRANCAISE | CNR Virus des Infections Respiratoires - France SUD | Antonin Bal et al |
| EPI_ISL_1371905 | C H DE LA POLYNESIE FRANCAISE | CNR Virus des Infections Respiratoires - France SUD | Antonin Bal et al |
| EPI_ISL_837279 | Istituto Zooprofilattico Sperimentale del Mezzogiorno | TIGEM | Antonio Grimaldi et al |
| EPI_ISL_1086087 | IZSM | TIGEM | Antonio Grimaldi et al |
| EPI_ISL_1086093 | IZSM | TIGEM | Antonio Grimaldi et al |
| EPI_ISL_1086096 | IZSM | TIGEM | Antonio Grimaldi et al |
| EPI_ISL_776981 | Istituto Zooprofilattico Sperimentale del Mezzogiorno | TIGEM | Antonio Grimaldi et al |

| | | | |
|---|---|---|---|
| EPI_ISL_776973 | Istituto Zooprofilattico Sperimentale del Mezzogiorno | TIGEM | Antonio Grimaldi et al |
| EPI_ISL_1086086 | IZSM | TIGEM | Antonio Grimaldi et al |
| EPI_ISL_837300 | Istituto Zooprofilattico Sperimentale del Mezzogiorno | TIGEM | Antonio Grimaldi et al |
| EPI_ISL_1656405 | Istituto Zooprofilattico Sperimentale del Mezzogiorno | TIGEM | Antonio Grimaldi Patrizia Annunziata Franc |
| EPI_ISL_1196991 | IZSM | TIGEM | Antonio Grimaldi Patrizia Annunziata Franc |
| EPI_ISL_1197001 | IZSM | TIGEM | Antonio Grimaldi Patrizia Annunziata Franc |
| EPI_ISL_1197002 | IZSM | TIGEM | Antonio Grimaldi Patrizia Annunziata Franc |
| EPI_ISL_1197005 | IZSM | TIGEM | Antonio Grimaldi Patrizia Annunziata Franc |
| EPI_ISL_751339 | IRCCS Sacro Cuore Don Calabria Hospital, Department of Infectious, Tropical Diseases & Microbiology | University of Verona, Department of Biotechnology | Antonio Mori et al |
| EPI_ISL_751342 | IRCCS Sacro Cuore Don Calabria Hospital, Department of Infectious, Tropical Diseases & Microbiology | University of Verona, Department of Biotechnology | Antonio Mori et al |
| EPI_ISL_541031 | Servicio de Microbiología, Hospital Miguel Servet, Zaragoza | SeqCOVID-SPAIN consortium/Institute of Biomedicine of Valencia, IBV-CSIC | Antonio Rezusta López et al |
| EPI_ISL_541034 | Servicio de Microbiología, Hospital Miguel Servet, Zaragoza | SeqCOVID-SPAIN consortium/Institute of Biomedicine of Valencia, IBV-CSIC | Antonio Rezusta López et al |
| EPI_ISL_700595 | Heideveld Emergency Centre | NHLS/UCT | Arash Iranzadeh et al |
| EPI_ISL_960172 | Dr Abdurahman CDC wc DAC | National Health Laboratory Service/UCT | Arash Iranzadeh et al |
| EPI_ISL_640054 | Groote Schuur Hospital wc GSH | NHLS/UCT | Arash Iranzadeh et al |
| EPI_ISL_640041 | Green Point CDC wc GPH | NHLS/UCT | Arash Iranzadeh et al |
| EPI_ISL_640059 | Du Noon CDC wc DNC | NHLS/UCT | Arash Iranzadeh et al |
| EPI_ISL_960142 | Groote Schuur Hospital wc GSH | National Health Laboratory Service/UCT | Arash Iranzadeh et al |
| EPI_ISL_1040795 | Groote Schuur Hospital wc GSH | NHLS/UCT | Arash Iranzadeh et al |
| EPI_ISL_1040796 | Groote Schuur Hospital wc GSH | NHLS/UCT | Arash Iranzadeh et al |
| EPI_ISL_1040787 | Groote Schuur Hospital wc GSH | NHLS/UCT | Arash Iranzadeh et al |
| EPI_ISL_1040793 | Guguletu CHC wc GDH | NHLS/UCT | Arash Iranzadeh et al |
| EPI_ISL_1040783 | Groote Schuur Hospital wc GSH | NHLS/UCT | Arash Iranzadeh et al |
| EPI_ISL_1040770 | 2 Military Hospital wc MAA | NHLS/UCT | Arash Iranzadeh et al |
| EPI_ISL_1040771 | 2 Military Hospital wc MAA | NHLS/UCT | Arash Iranzadeh et al |
| EPI_ISL_640129 | Groote Schuur Hospital wc GSH | NHLS/UCT | Arash Iranzadeh et al |
| EPI_ISL_640130 | Groote Schuur Hospital wc GSH | NHLS/UCT | Arash Iranzadeh et al |
| EPI_ISL_1040816 | Groote Schuur Hospital wc GSH | NHLS/UCT | Arash Iranzadeh et al |
| EPI_ISL_700411 | Hanover Park CHC wc HPH | NHLS/UCT | Arash Iranzadeh et al |

esco Panariello Biancamaria Pierri Claudia T
esco Panariello Biancamaria Pierri Valentina
esco Panariello Biancamaria Pierri Valentina
esco Panariello Biancamaria Pierri Valentina
esco Panariello Biancamaria Pierri Valentina

| | | | |
|---|---|---|---|
| EPI_ISL_700455 | Guguletu CHC wc GDH | NHLS/UCT | Arash Iranzadeh et al |
| EPI_ISL_700481 | Guguletu CHC wc GDH | NHLS/UCT | Arash Iranzadeh et al |
| EPI_ISL_1040815 | Groote Schuur Hospital wc GSH | NHLS/UCT | Arash Iranzadeh et al |
| EPI_ISL_640138 | Groote Schuur Hospital wc GSH | NHLS/UCT | Arash Iranzadeh et al |
| EPI_ISL_640091 | Du Noon CDC wc DNC | NHLS/UCT | Arash Iranzadeh et al |
| EPI_ISL_960162 | Heideveld Emergency Centre | National Health Laboratory Service/UCT | Arash Iranzadeh et al |
| EPI_ISL_1040667 | Guguletu CHC wc GDH | NHLS/UCT | Arash Iranzadeh et al |
| EPI_ISL_1040761 | Guguletu CHC | NHLS/UCT | Arash Iranzadeh et al |
| EPI_ISL_1040762 | Bredasdorp clinic | NHLS/UCT | Arash Iranzadeh et al |
| EPI_ISL_1040760 | Kensington CDC | NHLS/UCT | Arash Iranzadeh et al |
| EPI_ISL_1040647 | Swellendam PHC | NHLS/UCT | Arash Iranzadeh et al |
| EPI_ISL_1040649 | Groote Schuur Hospital wc GSH | NHLS/UCT | Arash Iranzadeh et al |
| EPI_ISL_1040767 | Groote Schuur Hospital wc GSH | NHLS/UCT | Arash Iranzadeh et al |
| EPI_ISL_1040765 | Groote Schuur Hospital wc GSH | NHLS/UCT | Arash Iranzadeh et al |
| EPI_ISL_1534309 | Groote Schuur Hospital wc GSH | NHLS/UCT | Arash Iranzadeh et al |
| EPI_ISL_508337 | Indian Institute of Science | National Institute of Biomedical Genomics | Arindam Maitra et al |
| EPI_ISL_508214 | Government Medical College | National Institute of Biomedical Genomics | Arindam Maitra et al |
| EPI_ISL_508266 | Government Medical College | National Institute of Biomedical Genomics | Arindam Maitra et al |
| EPI_ISL_1589741 | INSACOG-WB | National Institute of Biomedical Genomics – INSACOG | Arindam Maitra et al |
| EPI_ISL_1589853 | INSACOG-WB | National Institute of Biomedical Genomics – INSACOG | Arindam Maitra et al |
| EPI_ISL_1419217 | INSACOG-WB | National Institute of Biomedical Genomics – INSACOG | Arindam Maitra et al |
| EPI_ISL_1589869 | INSACOG-WB | National Institute of Biomedical Genomics – INSACOG | Arindam Maitra et al |
| EPI_ISL_1419562 | INSACOG-WB | National Institute of Biomedical Genomics – INSACOG | Arindam Maitra et al |
| EPI_ISL_1419609 | INSACOG-WB | National Institute of Biomedical Genomics – INSACOG | Arindam Maitra et al |
| EPI_ISL_1419620 | INSACOG-WB | National Institute of Biomedical Genomics – INSACOG | Arindam Maitra et al |
| EPI_ISL_1419643 | INSACOG-WB | National Institute of Biomedical Genomics – INSACOG | Arindam Maitra et al |
| EPI_ISL_1419631 | INSACOG-WB | National Institute of Biomedical Genomics – INSACOG | Arindam Maitra et al |
| EPI_ISL_1419888 | INSACOG-WB | National Institute of Biomedical Genomics – INSACOG | Arindam Maitra et al |
| EPI_ISL_1589966 | INSACOG-WB | National Institute of Biomedical Genomics – INSACOG | Arindam Maitra et al |
| EPI_ISL_1419660 | INSACOG-WB | National Institute of Biomedical Genomics – INSACOG | Arindam Maitra et al |

| | | | |
|---|---|---|---|
| EPI_ISL_1419659 | INSACOG-WB | National Institute of Biomedical Genomics – INSACOG | Arindam Maitra et al |
| EPI_ISL_430852 | Klinisk mikrobiologi och vardhygien Halmstad | The Public Health Agency of Sweden | Arne Kotz et al |
| EPI_ISL_1139157 | National Center of Disease Control and Prevention of the Republic of Armenia | Institute of Molecular Biology NAS RA, Republic of Armenia, Department of Bioengineering, BioinformaticsInstitute and Molecular Biology IBMPh RAU, Republic of Armenia | Arsen Arakelyan et al |
| EPI_ISL_1139149 | National Center of Disease Control and Prevention of the Republic of Armenia | Institute of Molecular Biology NAS RA, Republic of Armenia, Department of Bioengineering, BioinformaticsInstitute and Molecular Biology IBMPh RAU, Republic of Armenia | Arsen Arakelyan et al |
| EPI_ISL_1139150 | National Center of Disease Control and Prevention of the Republic of Armenia | Institute of Molecular Biology NAS RA, Republic of Armenia, Department of Bioengineering, BioinformaticsInstitute and Molecular Biology IBMPh RAU, Republic of Armenia | Arsen Arakelyan et al |
| EPI_ISL_1139151 | National Center of Disease Control and Prevention of the Republic of Armenia | Institute of Molecular Biology NAS RA, Republic of Armenia, Department of Bioengineering, BioinformaticsInstitute and Molecular Biology IBMPh RAU, Republic of Armenia | Arsen Arakelyan et al |
| EPI_ISL_1139153 | National Center of Disease Control and Prevention of the Republic of Armenia | Institute of Molecular Biology NAS RA, Republic of Armenia, Department of Bioengineering, BioinformaticsInstitute and Molecular Biology IBMPh RAU, Republic of Armenia | Arsen Arakelyan et al |
| EPI_ISL_1591100 | CMA Dano | Centre Muraz | Arsène Zongo et al |
| EPI_ISL_569776 | Omsk Research Institute of Natural Focal Infections | WHO National Influenza Centre Russian Federation | Artem Fadeev et al |
| EPI_ISL_639919 | Omsk Research Institute of Natural Focal Infections | WHO National Influenza Centre Russian Federation | Artem Fadeev et al |
| EPI_ISL_639920 | Omsk Research Institute of Natural Focal Infections | WHO National Influenza Centre Russian Federation | Artem Fadeev et al |
| EPI_ISL_569821 | Omsk Research Institute of Natural Focal Infections | WHO National Influenza Centre Russian Federation | Artem Fadeev et al |
| EPI_ISL_639932 | Omsk Research Institute of Natural Focal Infections | WHO National Influenza Centre Russian Federation | Artem Fadeev et al |
| EPI_ISL_569803 | Omsk Research Institute of Natural Focal Infections | WHO National Influenza Centre Russian Federation | Artem Fadeev et al |
| EPI_ISL_639933 | Omsk Research Institute of Natural Focal Infections | WHO National Influenza Centre Russian Federation | Artem Fadeev et al |
| EPI_ISL_639935 | Omsk Research Institute of Natural Focal Infections | WHO National Influenza Centre Russian Federation | Artem Fadeev et al |
| EPI_ISL_639943 | Omsk Research Institute of Natural Focal Infections | WHO National Influenza Centre Russian Federation | Artem Fadeev et al |
| EPI_ISL_1299227 | OLVZ Aalst | OLVZ Aalst | Astrid Holderbeke et al |
| EPI_ISL_1302291 | Instituto Nacional de Enfermedades Respiratorias (INER) | Instituto de Biotecnología de la UNAM | Authors from IBT et al |
| EPI_ISL_1302152 | Instituto Nacional de Enfermedades Respiratorias (INER) | Instituto de Biotecnología de la UNAM | Authors from IBT et al |
| EPI_ISL_1301499 | Instituto Nacional de Enfermedades Respiratorias (INER) | Instituto de Biotecnología de la UNAM | Authors from IBT et al |
| EPI_ISL_1301507 | Instituto Nacional de Enfermedades Respiratorias (INER) | Instituto de Biotecnología de la UNAM | Authors from IBT et al |
| EPI_ISL_1301474 | Instituto de Diagnostico y Referencia Epidemiologicos InDRE_RNLSP | Instituto de Biotecnología de la UNAM | Authors from IBT et al |
| EPI_ISL_1302210 | Instituto de Diagnostico y Referencia Epidemiologicos InDRE_RNLSP | Instituto de Biotecnología de la UNAM | Authors from IBT et al |
| EPI_ISL_1301519 | Instituto de Diagnostico y Referencia Epidemiologicos InDRE_RNLSP | Instituto de Biotecnología de la UNAM | Authors from IBT et al |

| | | | |
|---|---|---|---|
| EPI_ISL_1340864 | West Bandung Public Health | West Java Health Laboratory; School of Life Sciences and Technology, Institut Teknologi Bandung | Azzania Fibriani et al |
| EPI_ISL_1634465 | Bandung Public Health | West Java Health Laboratory; School of Life Sciences and Technology, Institut Teknologi Bandung | Azzania Fibriani et al |
| EPI_ISL_1615589 | Sumedang Public Health | West Java Health Laboratory; School of Life Sciences and Technology, Institut Teknologi Bandung | Azzania Fibriani et al |
| EPI_ISL_1615590 | Subang Public Health | West Java Health Laboratory; School of Life Sciences and Technology, Institut Teknologi Bandung | Azzania Fibriani et al |
| EPI_ISL_1634442 | Mitra Kasih Hospital | West Java Health Laboratory; School of Life Sciences and Technology, Institut Teknologi Bandung | Azzania Fibriani et al |
| EPI_ISL_1660419 | Santosa Kopo Hospital | West Java Health Laboratory; School of Life Sciences and Technology, Institut Teknologi Bandung | Azzania Fibriani et al |
| EPI_ISL_747240 | Al Islam Hospital | West Java Health Laboratory; School of Life Sciences and Technology, Institut Teknologi Bandung | Azzania Fibriani et al |
| EPI_ISL_1056959 | Institute of Biocides and Medical Ecology, Belgarde, Serbia | Virology Department Institute of Microbiology and Immunology Faculty of Medicine University of Belgrade | Banko Ana et al |
| EPI_ISL_891272 | Institute of Biocides and Medical Ecology, Belgarde, Serbia | Virology department Institute of microbiology and immunology Faculty of Medicine University of Belgrade | Banko Ana et al |
| EPI_ISL_1056968 | Institute of Biocides and Medical Ecology, Belgarde, Serbia | Virology Department Institute of Microbiology and Immunology Faculty of Medicine University of Belgrade | Banko Ana et al |
| EPI_ISL_1057105 | Institute of Biocides and Medical Ecology, Belgarde, Serbia | Virology Department Institute of Microbiology and Immunology Faculty of Medicine University of Belgrade | Banko Ana et al |
| EPI_ISL_1061431 | Institute of Biocides and Medical Ecology, Belgarde, Serbia | Virology Department Institute of Microbiology and Immunology Faculty of Medicine University of Belgrade | Banko Ana et al |
| EPI_ISL_909737 | Institute of Biocides and Medical Ecology, Belgrade, Serbia | Virology Department, Institute of microbiology and immunology, Faculty of Medicine University of Blegrade | Banko Ana et al |
| EPI_ISL_891271 | Institute of Biocides and Medical Ecology, Belgarde, Serbia | Virology department Institute of microbiology and immunology Faculty of Medicine University of Belgrade | Banko Ana et al |
| EPI_ISL_1057040 | Institute of Biocides and Medical Ecology, Belgarde, Serbia | Virology Department Institute of Microbiology and Immunology Faculty of Medicine University of Belgrade | Banko Ana et al |
| EPI_ISL_930854 | Central Laboratory of Public Health of Rio Grande do Sul (Lacen-RS) | State Center for Health Surveillance of the Health Department of the State of Rio Grande do Sul (CEVS/SES-RS) | Barcellos R et al |
| EPI_ISL_930855 | Central Laboratory of Public Health of Rio Grande do Sul (Lacen-RS) | State Center for Health Surveillance of the Health Department of the State of Rio Grande do Sul (CEVS/SES-RS) | Barcellos R et al |
| EPI_ISL_942407 | Central Laboratory of Public Health of Rio Grande do Sul (Lacen-RS) | State Center for Health Surveillance of the Health Department of the State of Rio Grande do Sul (CEVS/SES-RS) | Barcellos R et al |
| EPI_ISL_930856 | Central Laboratory of Public Health of Rio Grande do Sul (Lacen-RS) | State Center for Health Surveillance of the Health Department of the State of Rio Grande do Sul (CEVS/SES-RS) | Barcellos R et al |
| EPI_ISL_930857 | Central Laboratory of Public Health of Rio Grande do Sul (Lacen-RS) | State Center for Health Surveillance of the Health Department of the State of Rio Grande do Sul (CEVS/SES-RS) | Barcellos R et al |
| EPI_ISL_930858 | Central Laboratory of Public Health of Rio Grande do Sul (Lacen-RS) | State Center for Health Surveillance of the Health Department of the State of Rio Grande do Sul (CEVS/SES-RS) | Barcellos R et al |
| EPI_ISL_942374 | Central Laboratory of Public Health of Rio Grande do Sul (Lacen-RS) | State Center for Health Surveillance of the Health Department of the State of Rio Grande do Sul (CEVS/SES-RS) | Barcellos R et al |

| | | | |
|---|---|---|---|
| EPI_ISL_942375 | Central Laboratory of Public Health of Rio Grande do Sul (Lacen-RS) | State Center for Health Surveillance of the Health Department of the State of Rio Grande do Sul (CEVS/SES-RS) | Barcellos R et al |
| EPI_ISL_942896 | Central Laboratory of Public Health of Rio Grande do Sul (Lacen-RS) | State Center for Health Surveillance of the Health Department of the State of Rio Grande do Sul (CEVS/SES-RS) | Barcellos R et al |
| EPI_ISL_942897 | Central Laboratory of Public Health of Rio Grande do Sul (Lacen-RS) | State Center for Health Surveillance of the Health Department of the State of Rio Grande do Sul (CEVS/SES-RS) | Barcellos R et al |
| EPI_ISL_942898 | Central Laboratory of Public Health of Rio Grande do Sul (Lacen-RS) | State Center for Health Surveillance of the Health Department of the State of Rio Grande do Sul (CEVS/SES-RS) | Barcellos R et al |
| EPI_ISL_942899 | Central Laboratory of Public Health of Rio Grande do Sul (Lacen-RS) | State Center for Health Surveillance of the Health Department of the State of Rio Grande do Sul (CEVS/SES-RS) | Barcellos R et al |
| EPI_ISL_942930 | Central Laboratory of Public Health of Rio Grande do Sul (Lacen-RS) | State Center for Health Surveillance of the Health Department of the State of Rio Grande do Sul (CEVS/SES-RS) | Barcellos R et al |
| EPI_ISL_942931 | Central Laboratory of Public Health of Rio Grande do Sul (Lacen-RS) | State Center for Health Surveillance of the Health Department of the State of Rio Grande do Sul (CEVS/SES-RS) | Barcellos R et al |
| EPI_ISL_982236 | National Institute of Laboratory Medicine and Referral Center | Genomic Research Lab, BCSIR | Barna Goswami et al |
| EPI_ISL_1040481 | National Institute of Laboratory Medicine and Referral Center | Genomic Research Lab, BCSIR | Barna Goswami et al |
| EPI_ISL_483551 | Kingdom of Bahrein Ministry of Health | Erasmus Medical Center | Bas Oude Munnink et al |
| EPI_ISL_483552 | Kingdom of Bahrein Ministry of Health | Erasmus Medical Center | Bas Oude Munnink et al |
| EPI_ISL_483555 | Kingdom of Bahrein Ministry of Health | Erasmus Medical Center | Bas Oude Munnink et al |
| EPI_ISL_483558 | Kingdom of Bahrein Ministry of Health | Erasmus Medical Center | Bas Oude Munnink et al |
| EPI_ISL_483560 | Kingdom of Bahrein Ministry of Health | Erasmus Medical Center | Bas Oude Munnink et al |
| EPI_ISL_483561 | Kingdom of Bahrein Ministry of Health | Erasmus Medical Center | Bas Oude Munnink et al |
| EPI_ISL_483565 | Kingdom of Bahrein Ministry of Health | Erasmus Medical Center | Bas Oude Munnink et al |
| EPI_ISL_523137 | Dutch COVID-19 response team | Erasmus Medical Center | Bas Oude Munnink et al |
| EPI_ISL_523165 | Dutch COVID-19 response team | Erasmus Medical Center | Bas Oude Munnink et al |
| EPI_ISL_801406 | Dutch COVID-19 response team | Erasmus Medical Center | Bas Oude Munnink et al |
| EPI_ISL_801429 | Dutch COVID-19 response team | Erasmus Medical Center | Bas Oude Munnink et al |
| EPI_ISL_422757 | Dutch COVID-19 response team | Erasmus Medical Center | Bas Oude Munnink et al |
| EPI_ISL_422588 | Dutch COVID-19 response team | Erasmus Medical Center | Bas Oude Munnink et al |
| EPI_ISL_455136 | Dutch COVID-19 response team | Erasmus Medical Center | Bas Oude Munnink et al |
| EPI_ISL_455138 | Dutch COVID-19 response team | Erasmus Medical Center | Bas Oude Munnink et al |
| EPI_ISL_455158 | Dutch COVID-19 response team | Erasmus Medical Center | Bas Oude Munnink et al |
| EPI_ISL_455169 | Dutch COVID-19 response team | Erasmus Medical Center | Bas Oude Munnink et al |
| EPI_ISL_460715 | Dutch COVID-19 response team | Erasmus Medical Center | Bas Oude Munnink et al |
| EPI_ISL_461061 | Dutch COVID-19 response team | Erasmus Medical Center | Bas Oude Munnink et al |

| | | | |
|---|---|---|---|
| EPI_ISL_461124 | Dutch COVID-19 response team | Erasmus Medical Center | Bas Oude Munnink et al |
| EPI_ISL_422850 | Dutch COVID-19 response team | Erasmus Medical Center | Bas Oude Munnink et al |
| EPI_ISL_523308 | Dutch COVID-19 response team | Erasmus Medical Center | Bas Oude Munnink et al |
| EPI_ISL_523334 | Dutch COVID-19 response team | Erasmus Medical Center | Bas Oude Munnink et al |
| EPI_ISL_523337 | Dutch COVID-19 response team | Erasmus Medical Center | Bas Oude Munnink et al |
| EPI_ISL_523384 | Dutch COVID-19 response team | Erasmus Medical Center | Bas Oude Munnink et al |
| EPI_ISL_632359 | Dutch COVID-19 response team | Erasmus Medical Center | Bas Oude Munnink et al |
| EPI_ISL_722378 | Dutch COVID-19 response team | Erasmus Medical Center | Bas Oude Munnink et al |
| EPI_ISL_763139 | Dutch COVID-19 response team | Erasmus Medical Center | Bas Oude Munnink et al |
| EPI_ISL_1311245 | Dutch COVID-19 response team | Erasmus Medical Center | Bas Oude Munnink et al |
| EPI_ISL_1311685 | Dutch COVID-19 response team | Erasmus Medical Center | Bas Oude Munnink et al |
| EPI_ISL_1311275 | Dutch COVID-19 response team | Erasmus Medical Center | Bas Oude Munnink et al |
| EPI_ISL_577849 | Dutch COVID-19 response team | Erasmus Medical Center | Bas Oude Munnink et al |
| EPI_ISL_461158 | Dutch COVID-19 response team | Erasmus Medical Center | Bas Oude Munnink et al |
| EPI_ISL_722292 | Dutch COVID-19 response team | Erasmus Medical Center | Bas Oude Munnink et al |
| EPI_ISL_632327 | Dutch COVID-19 response team | Erasmus Medical Center | Bas Oude Munnink et al |
| EPI_ISL_632417 | Dutch COVID-19 response team | Erasmus Medical Center | Bas Oude Munnink et al |
| EPI_ISL_523547 | Dutch COVID-19 response team | Erasmus Medical Center | Bas Oude Munnink et al |
| EPI_ISL_722396 | Dutch COVID-19 response team | Erasmus Medical Center | Bas Oude Munnink et al |
| EPI_ISL_801441 | Dutch COVID-19 response team | Erasmus Medical Center | Bas Oude Munnink et al |
| EPI_ISL_801442 | Dutch COVID-19 response team | Erasmus Medical Center | Bas Oude Munnink et al |
| EPI_ISL_1310851 | Dutch COVID-19 response team | Erasmus Medical Center | Bas Oude Munnink et al |
| EPI_ISL_1311252 | Dutch COVID-19 response team | Erasmus Medical Center | Bas Oude Munnink et al |
| EPI_ISL_523645 | Dutch COVID-19 response team | Erasmus Medical Center | Bas Oude Munnink et al |
| EPI_ISL_574789 | Dutch COVID-19 response team | Erasmus Medical Center | Bas Oude Munnink et al |
| EPI_ISL_577932 | Dutch COVID-19 response team | Erasmus Medical Center | Bas Oude Munnink et al |
| EPI_ISL_632400 | Dutch COVID-19 response team | Erasmus Medical Center | Bas Oude Munnink et al |
| EPI_ISL_632769 | Dutch COVID-19 response team | Erasmus Medical Center | Bas Oude Munnink et al |
| EPI_ISL_461315 | Dutch COVID-19 response team | Erasmus Medical Center | Bas Oude Munnink et al |
| EPI_ISL_422933 | Dutch COVID-19 response team | Erasmus Medical Center | Bas Oude Munnink et al |

| | | | |
|---|---|---|---|
| EPI_ISL_523510 | Dutch COVID-19 response team | Erasmus Medical Center | Bas Oude Munnink et al |
| EPI_ISL_517616 | Academic Hospital Paramaribo | Erasmus Medical Center | Bas Oude Munnink et al |
| EPI_ISL_517628 | Academic Hospital Paramaribo | Erasmus Medical Center | Bas Oude Munnink et al |
| EPI_ISL_517629 | Academic Hospital Paramaribo | Erasmus Medical Center | Bas Oude Munnink et al |
| EPI_ISL_517631 | Academic Hospital Paramaribo | Erasmus Medical Center | Bas Oude Munnink et al |
| EPI_ISL_517632 | Academic Hospital Paramaribo | Erasmus Medical Center | Bas Oude Munnink et al |
| EPI_ISL_517634 | Academic Hospital Paramaribo | Erasmus Medical Center | Bas Oude Munnink et al |
| EPI_ISL_517635 | Academic Hospital Paramaribo | Erasmus Medical Center | Bas Oude Munnink et al |
| EPI_ISL_517642 | Academic Hospital Paramaribo | Erasmus Medical Center | Bas Oude Munnink et al |
| EPI_ISL_517643 | Academic Hospital Paramaribo | Erasmus Medical Center | Bas Oude Munnink et al |
| EPI_ISL_517645 | Academic Hospital Paramaribo | Erasmus Medical Center | Bas Oude Munnink et al |
| EPI_ISL_517657 | Academic Hospital Paramaribo | Erasmus Medical Center | Bas Oude Munnink et al |
| EPI_ISL_518799 | Academic Hospital Paramaribo | Erasmus Medical Center | Bas Oude Munnink et al |
| EPI_ISL_518800 | Academic Hospital Paramaribo | Erasmus Medical Center | Bas Oude Munnink et al |
| EPI_ISL_518805 | Academic Hospital Paramaribo | Erasmus Medical Center | Bas Oude Munnink et al |
| EPI_ISL_518807 | Academic Hospital Paramaribo | Erasmus Medical Center | Bas Oude Munnink et al |
| EPI_ISL_518811 | Academic Hospital Paramaribo | Erasmus Medical Center | Bas Oude Munnink et al |
| EPI_ISL_518812 | Academic Hospital Paramaribo | Erasmus Medical Center | Bas Oude Munnink et al |
| EPI_ISL_517661 | Academic Hospital Paramaribo | Erasmus Medical Center | Bas Oude Munnink et al |
| EPI_ISL_517662 | Academic Hospital Paramaribo | Erasmus Medical Center | Bas Oude Munnink et al |
| EPI_ISL_518818 | Academic Hospital Paramaribo | Erasmus Medical Center | Bas Oude Munnink et al |
| EPI_ISL_1259261 | LHUB-ULB | UAntwerp, Laboratory of Medical Microbiology | Basil Britto Xavier et al |
| EPI_ISL_1259276 | LHUB-ULB | UAntwerp, Laboratory of Medical Microbiology | Basil Britto Xavier et al |
| EPI_ISL_629023 | Centro de Biotecnología Vegetal, Universidad Andrés Bello, Center for Genome Regulation | Center for Mathematical Modeling and Center for Genome Regulation. Santiago, Chile | Bastias M et al |
| EPI_ISL_629015 | Centro de Biotecnología Vegetal, Universidad Andrés Bello, Center for Genome Regulation | Center for Mathematical Modeling and Center for Genome Regulation. Santiago, Chile | Bastias M et al |
| EPI_ISL_629016 | Centro de Biotecnología Vegetal, Universidad Andrés Bello, Center for Genome Regulation | Center for Mathematical Modeling and Center for Genome Regulation. Santiago, Chile | Bastias M et al |
| EPI_ISL_849151 | Servicio Virosis Respiratorias-Departamento Virología-INEI | Instituto Nacional Enfermedades Infecciosas C.G.Malbran | Baumeister E. et al |
| EPI_ISL_849154 | Servicio Virosis Respiratorias-Departamento Virología-INEI | Instituto Nacional Enfermedades Infecciosas C.G.Malbran | Baumeister E. et al |
| EPI_ISL_849318 | Servicio Virosis Respiratorias-Departamento Virología-INEI | Instituto Nacional Enfermedades Infecciosas C.G.Malbran | Baumeister E. et al |

| | | | |
|---|---|---|---|
| EPI_ISL_849301 | Servicio Virosis Respiratorias-Departamento Virología-INEI | Instituto Nacional Enfermedades Infecciosas C.G.Malbran | Baumeister E. et al |
| EPI_ISL_849295 | Servicio Virosis Respiratorias-Departamento Virología-INEI | Instituto Nacional Enfermedades Infecciosas C.G.Malbran | Baumeister E. et al |
| EPI_ISL_849298 | Servicio Virosis Respiratorias-Departamento Virología-INEI | Instituto Nacional Enfermedades Infecciosas C.G.Malbran | Baumeister E. et al |
| EPI_ISL_849280 | Servicio Virosis Respiratorias-Departamento Virología-INEI | Instituto Nacional Enfermedades Infecciosas C.G.Malbran | Baumeister E. et al |
| EPI_ISL_849285 | Servicio Virosis Respiratorias-Departamento Virología-INEI | Instituto Nacional Enfermedades Infecciosas C.G.Malbran | Baumeister E. et al |
| EPI_ISL_849306 | Servicio Virosis Respiratorias-Departamento Virología-INEI | Instituto Nacional Enfermedades Infecciosas C.G.Malbran | Baumeister E. et al |
| EPI_ISL_849281 | Servicio Virosis Respiratorias-Departamento Virología-INEI | Instituto Nacional Enfermedades Infecciosas C.G.Malbran | Baumeister E. et al |
| EPI_ISL_849289 | Servicio Virosis Respiratorias-Departamento Virología-INEI | Instituto Nacional Enfermedades Infecciosas C.G.Malbran | Baumeister E. et al |
| EPI_ISL_849284 | Servicio Virosis Respiratorias-Departamento Virología-INEI | Instituto Nacional Enfermedades Infecciosas C.G.Malbran | Baumeister E. et al |
| EPI_ISL_849291 | Servicio Virosis Respiratorias-Departamento Virología-INEI | Instituto Nacional Enfermedades Infecciosas C.G.Malbran | Baumeister E. et al |
| EPI_ISL_849373 | Servicio Virosis Respiratorias-Departamento Virología-INEI | Instituto Nacional Enfermedades Infecciosas C.G.Malbran | Baumeister E. et al |
| EPI_ISL_849353 | Servicio Virosis Respiratorias-Departamento Virología-INEI | Instituto Nacional Enfermedades Infecciosas C.G.Malbran | Baumeister E. et al |
| EPI_ISL_849361 | Servicio Virosis Respiratorias-Departamento Virología-INEI | Instituto Nacional Enfermedades Infecciosas C.G.Malbran | Baumeister E. et al |
| EPI_ISL_849362 | Servicio Virosis Respiratorias-Departamento Virología-INEI | Instituto Nacional Enfermedades Infecciosas C.G.Malbran | Baumeister E. et al |
| EPI_ISL_849369 | Servicio Virosis Respiratorias-Departamento Virología-INEI | Instituto Nacional Enfermedades Infecciosas C.G.Malbran | Baumeister E. et al |
| EPI_ISL_856792 | Servicio Virosis Respiratorias-Departamento Virología-INEI | Instituto Nacional Enfermedades Infecciosas C.G.Malbran | Baumeister E. et al |
| EPI_ISL_856766 | Servicio Virosis Respiratorias-Departamento Virología-INEI | Instituto Nacional Enfermedades Infecciosas C.G.Malbran | Baumeister E. et al |
| EPI_ISL_856771 | Servicio Virosis Respiratorias-Departamento Virología-INEI | Instituto Nacional Enfermedades Infecciosas C.G.Malbran | Baumeister E. et al |
| EPI_ISL_422563 | Institute of Microbiology Universidad San Francisco de Quito | Institute of Microbiology Universidad San Francisco de Quito | Belen Prado-Vivar et al |
| EPI_ISL_477014 | Institute of Microbiology, Universidad San Francisco de Quito | Institute of Microbiology, Universidad San Francisco de Quito | Belen Prado-Vivar et al |
| EPI_ISL_481244 | Hospital IESS Babahoyo | Institute of Microbiology, Universidad San Francisco de Quito | Belén Prado-Vivar et al |
| EPI_ISL_481245 | Hospital IESS Babahoyo | Institute of Microbiology, Universidad San Francisco de Quito | Belén Prado-Vivar et al |
| EPI_ISL_471268 | Hospital IESS Babahoyo | Institute of Microbiology, Universidad San Francisco de Quito | Belén Prado-Vivar et al |
| EPI_ISL_481246 | Hospital IESS Babahoyo | Institute of Microbiology, Universidad San Francisco de Quito | Belén Prado-Vivar et al |
| EPI_ISL_481248 | Hospital IESS Babahoyo | Institute of Microbiology, Universidad San Francisco de Quito | Belén Prado-Vivar et al |
| EPI_ISL_491939 | Institute of Microbiology, Universidad San Francisco de Quito | Institute of Microbiology, Universidad San Francisco de Quito | Belén Prado-Vivar et al |
| EPI_ISL_491940 | Institute of Microbiology, Universidad San Francisco de Quito | Institute of Microbiology, Universidad San Francisco de Quito | Belén Prado-Vivar et al |
| EPI_ISL_491936 | Institute of Microbiology, Universidad San Francisco de Quito | Institute of Microbiology, Universidad San Francisco de Quito | Belén Prado-Vivar et al |
| EPI_ISL_491938 | Institute of Microbiology, Universidad San Francisco de Quito | Institute of Microbiology, Universidad San Francisco de Quito | Belén Prado-Vivar et al |
| EPI_ISL_697783 | Institute of Microbiology, Universidad San Francisco de Quito | Institute of Microbiology, Universidad San Francisco de Quito | Belén Prado-Vivar et al |

| | | | |
|---|---|---|---|
| EPI_ISL_697784 | Institute of Microbiology, Universidad San Francisco de Quito | Institute of Microbiology, Universidad San Francisco de Quito | Belén Prado-Vivar et al |
| EPI_ISL_697786 | Institute of Microbiology, Universidad San Francisco de Quito | Institute of Microbiology, Universidad San Francisco de Quito | Belén Prado-Vivar et al |
| EPI_ISL_527812 | Institute of Microbiology, Universidad San Francisco de Quito | Institute of Microbiology, Universidad San Francisco de Quito | Belén Prado-Vivar et al |
| EPI_ISL_527814 | Institute of Microbiology, Universidad San Francisco de Quito | Institute of Microbiology, Universidad San Francisco de Quito | Belén Prado-Vivar et al |
| EPI_ISL_527817 | Institute of Microbiology, Universidad San Francisco de Quito | Institute of Microbiology, Universidad San Francisco de Quito | Belén Prado-Vivar et al |
| EPI_ISL_539791 | Institute of Microbiology, Universidad San Francisco de Quito | Institute of Microbiology, Universidad San Francisco de Quito | Belén Prado-Vivar et al |
| EPI_ISL_539792 | Institute of Microbiology, Universidad San Francisco de Quito | Institute of Microbiology, Universidad San Francisco de Quito | Belén Prado-Vivar et al |
| EPI_ISL_697789 | Institute of Microbiology, Universidad San Francisco de Quito | Institute of Microbiology, Universidad San Francisco de Quito | Belén Prado-Vivar et al |
| EPI_ISL_697790 | Institute of Microbiology, Universidad San Francisco de Quito | Institute of Microbiology, Universidad San Francisco de Quito | Belén Prado-Vivar et al |
| EPI_ISL_697791 | Institute of Microbiology, Universidad San Francisco de Quito | Institute of Microbiology, Universidad San Francisco de Quito | Belén Prado-Vivar et al |
| EPI_ISL_697792 | Institute of Microbiology, Universidad San Francisco de Quito | Institute of Microbiology, Universidad San Francisco de Quito | Belén Prado-Vivar et al |
| EPI_ISL_697793 | Institute of Microbiology, Universidad San Francisco de Quito | Institute of Microbiology, Universidad San Francisco de Quito | Belén Prado-Vivar et al |
| EPI_ISL_697798 | Institute of Microbiology, Universidad San Francisco de Quito | Institute of Microbiology, Universidad San Francisco de Quito | Belén Prado-Vivar et al |
| EPI_ISL_697799 | Institute of Microbiology, Universidad San Francisco de Quito | Institute of Microbiology, Universidad San Francisco de Quito | Belén Prado-Vivar et al |
| EPI_ISL_697800 | Institute of Microbiology, Universidad San Francisco de Quito | Institute of Microbiology, Universidad San Francisco de Quito | Belén Prado-Vivar et al |
| EPI_ISL_824290 | Institute of Microbiology, Universidad San Francisco de Quito | Institute of Microbiology, Universidad San Francisco de Quito | Belén Prado-Vivar et al |
| EPI_ISL_824286 | Institute of Microbiology, Universidad San Francisco de Quito | Institute of Microbiology, Universidad San Francisco de Quito | Belén Prado-Vivar et al |
| EPI_ISL_824288 | Institute of Microbiology, Universidad San Francisco de Quito | Institute of Microbiology, Universidad San Francisco de Quito | Belén Prado-Vivar et al |
| EPI_ISL_824289 | Institute of Microbiology, Universidad San Francisco de Quito | Institute of Microbiology, Universidad San Francisco de Quito | Belén Prado-Vivar et al |
| EPI_ISL_824285 | Institute of Microbiology, Universidad San Francisco de Quito | Institute of Microbiology, Universidad San Francisco de Quito | Belén Prado-Vivar et al |
| EPI_ISL_824293 | Institute of Microbiology, Universidad San Francisco de Quito | Institute of Microbiology, Universidad San Francisco de Quito | Belén Prado-Vivar et al |
| EPI_ISL_824294 | Institute of Microbiology, Universidad San Francisco de Quito | Institute of Microbiology, Universidad San Francisco de Quito | Belén Prado-Vivar et al |
| EPI_ISL_877561 | Institute of Microbiology, Universidad San Francisco de Quito | Institute of Microbiology, Universidad San Francisco de Quito | Belén Prado-Vivar et al |
| EPI_ISL_877562 | Institute of Microbiology, Universidad San Francisco de Quito | Institute of Microbiology, Universidad San Francisco de Quito | Belén Prado-Vivar et al |
| EPI_ISL_1656147 | Institute of Microbiology, Universidad San Francisco de Quito | Institute of Microbiology, Universidad San Francisco de Quito | Belén Prado-Vivar et al |
| EPI_ISL_1540707 | Universidad Tecnica Particular de Loja | Institute of Microbiology, Universidad San Francisco de Quito | Belén Prado-Vivar et al |
| EPI_ISL_1656146 | Universidad Tecnica Particular de Loja | Institute of Microbiology, Universidad San Francisco de Quito | Belén Prado-Vivar et al |
| EPI_ISL_407896 | Pathology Queensland | Public Health Virology Laboratory | Ben Huang et al |
| EPI_ISL_411951 | Unit for Laboratory Development and Technology Transfer, Public Health Agency of Sweden | Unit for Laboratory Development and Technology Transfer, Public Health Agency of Sweden | Bengner et al |
| EPI_ISL_476778 | Stanford clinical virology lab | Chan-Zuckerberg Biohub | Benjamin Pinksy et al |

| | | | |
|---|---|---|---|
| EPI_ISL_476782 | Stanford clinical virology lab | Chan-Zuckerberg Biohub | Benjamin Pinksy et al |
| EPI_ISL_476790 | Stanford clinical virology lab | Chan-Zuckerberg Biohub | Benjamin Pinksy et al |
| EPI_ISL_1540836 | The Jackson Laboratory | The Jackson Laboratory | Bergeron D et al |
| EPI_ISL_1039831 | National Public Health Center, COVID Laboratory | National Public Health Center, National Biosafety Laboratory | Bernadett Pályi et al |
| EPI_ISL_1172027 | National Public Health Center, COVID Laboratory | National Public Health Center, National Biosafety Laboratory | Bernadett Pályi et al |
| EPI_ISL_1039986 | National Public Health Center, COVID Laboratory | National Public Health Center, National Biosafety Laboratory | Bernadett Pályi et al |
| EPI_ISL_1039983 | National Public Health Center, COVID Laboratory | National Public Health Center, National Biosafety Laboratory | Bernadett Pályi et al |
| EPI_ISL_1039981 | National Public Health Center, COVID Laboratory | National Public Health Center, National Biosafety Laboratory | Bernadett Pályi et al |
| EPI_ISL_1039980 | National Public Health Center, COVID Laboratory | National Public Health Center, National Biosafety Laboratory | Bernadett Pályi et al |
| EPI_ISL_1039961 | National Public Health Center, COVID Laboratory | National Public Health Center, National Biosafety Laboratory | Bernadett Pályi et al |
| EPI_ISL_1039964 | National Public Health Center, COVID Laboratory | National Public Health Center, National Biosafety Laboratory | Bernadett Pályi et al |
| EPI_ISL_1039837 | National Public Health Center, COVID Laboratory | National Public Health Center, National Biosafety Laboratory | Bernadett Pályi et al |
| EPI_ISL_1039959 | National Public Health Center, COVID Laboratory | National Public Health Center, National Biosafety Laboratory | Bernadett Pályi et al |
| EPI_ISL_1172022 | National Public Health Center, COVID Laboratory | National Public Health Center, National Biosafety Laboratory | Bernadett Pályi et al |
| EPI_ISL_1095612 | National Public Health Center, COVID Laboratory | National Public Health Center, National Biosafety Laboratory | Bernadett Pályi et al |
| EPI_ISL_1039993 | National Public Health Center, COVID Laboratory | National Public Health Center, National Biosafety Laboratory | Bernadett Pályi et al |
| EPI_ISL_1195207 | National Public Health Center, COVID Laboratory | National Public Health Center, National Biosafety Laboratory | Bernadett Pályi et al |
| EPI_ISL_1447296 | National Public Health Center, COVID Laboratory | National Public Health Center, National Biosafety Laboratory | Bernadett Pályi et al |
| EPI_ISL_1195192 | National Public Health Center, COVID Laboratory | National Public Health Center, National Biosafety Laboratory | Bernadett Pályi et al |
| EPI_ISL_1095621 | National Public Health Center, COVID Laboratory | National Public Health Center, National Biosafety Laboratory | Bernadett Pályi et al |
| EPI_ISL_1095620 | National Public Health Center, COVID Laboratory | National Public Health Center, National Biosafety Laboratory | Bernadett Pályi et al |
| EPI_ISL_415159 | KU Leuven, Clinical and Epidemiological Virology | KU Leuven, Clinical and Epidemiological Virology | Bert Vanmechelen et al |
| EPI_ISL_407976 | KU Leuven, Clinical and Epidemiological Virology | KU Leuven, Clinical and Epidemiological Virology | Bert Vanmechelen et al |
| EPI_ISL_763064 | RS Kasih Ibu, Surakarta | Universitas Sebelas Maret (UNS); Rumah Sakit UNS (RS-UNS); National Institute of Health Research and Development, Indonesian Ministry of Health. | Betty Suryawati et al |
| EPI_ISL_1402340 | CMBG FN Brno | CMBG FN Brno | Bezdicek M. et al |
| EPI_ISL_1402342 | CMBG FN Brno | CMBG FN Brno | Bezdicek M. et al |
| EPI_ISL_1402345 | CMBG FN Brno | CMBG FN Brno | Bezdicek M. et al |
| EPI_ISL_1402349 | CMBG FN Brno | CMBG FN Brno | Bezdicek M. et al |
| EPI_ISL_1402369 | Vyskov hospital | CMBG FN Brno | Bezdicek M. et al |

| | | | |
|---|---|---|---|
| EPI_ISL_492035 | Instituto de Biologia do Exército | Laboratório Metabolismo Macromolecular FirminoTorres de Castro, Instituto de Biofísica Carlos Chagas Filho, Universidade Federal do Rio de Janeiro | Bianca Catarina Azevedo Cabral et al |
| EPI_ISL_492043 | Instituto de Biologia do Exército | Laboratório Metabolismo Macromolecular FirminoTorres de Castro, Instituto de Biofísica Carlos Chagas Filho, Universidade Federal do Rio de Janeiro | Bianca Catarina Azevedo Cabral et al |
| EPI_ISL_492044 | Instituto de Biologia do Exército | Laboratório Metabolismo Macromolecular FirminoTorres de Castro, Instituto de Biofísica Carlos Chagas Filho, Universidade Federal do Rio de Janeiro | Bianca Catarina Azevedo Cabral et al |
| EPI_ISL_492036 | Instituto de Biologia do Exército | Laboratório Metabolismo Macromolecular FirminoTorres de Castro, Instituto de Biofísica Carlos Chagas Filho, Universidade Federal do Rio de Janeiro | Bianca Catarina Azevedo Cabral et al |
| EPI_ISL_576119 | Laboratory, The Bio Arte Limited | Laboratory, The Bio Arte Limited | Biazzo et al |
| EPI_ISL_576121 | Laboratory, The Bio Arte Limited | Laboratory, The Bio Arte Limited | Biazzo et al |
| EPI_ISL_576122 | Laboratory, The Bio Arte Limited | Laboratory, The Bio Arte Limited | Biazzo et al |
| EPI_ISL_576123 | Laboratory, The Bio Arte Limited | Laboratory, The Bio Arte Limited | Biazzo et al |
| EPI_ISL_576124 | Laboratory, The Bio Arte Limited | Laboratory, The Bio Arte Limited | Biazzo et al |
| EPI_ISL_1629593 | Clinical Microbiology, Infection Prevention and Control | Section for Molecular Diagnostics | Björn Hallström et al |
| EPI_ISL_1629603 | Clinical Microbiology, Infection Prevention and Control | Section for Molecular Diagnostics | Björn Hallström et al |
| EPI_ISL_1629615 | Clinical Microbiology, Infection Prevention and Control | Section for Molecular Diagnostics | Björn Hallström et al |
| EPI_ISL_1629655 | Clinical Microbiology, Infection Prevention and Control | Section for Molecular Diagnostics | Björn Hallström et al |
| EPI_ISL_450209 | Department of Virology | Department of Virology | Boehmer et al |
| EPI_ISL_450199 | Department of Virology | Department of Virology | Boehmer et al |
| EPI_ISL_450200 | Department of Virology | Department of Virology | Boehmer et al |
| EPI_ISL_450198 | Department of Virology | Department of Virology | Boehmer et al |
| EPI_ISL_454057 | unknown | Instituto Nacional de Saude (INSA) | Borges et al |
| EPI_ISL_454110 | unknown | Instituto Nacional de Saude (INSA) | Borges et al |
| EPI_ISL_454172 | unknown | Instituto Nacional de Saude (INSA) | Borges et al |
| EPI_ISL_454259 | unknown | Instituto Nacional de Saude (INSA) | Borges et al |
| EPI_ISL_511103 | Instituto Nacional de Saude (INSA) | Instituto Nacional de Saude (INSA) | Borges et al |
| EPI_ISL_511105 | Instituto Nacional de Saude (INSA) | Instituto Nacional de Saude (INSA) | Borges et al |
| EPI_ISL_511142 | Instituto Nacional de Saude (INSA) | Instituto Nacional de Saude (INSA) | Borges et al |
| EPI_ISL_511579 | Instituto Nacional de Saude (INSA) | Instituto Nacional de Saude (INSA) | Borges et al |
| EPI_ISL_511190 | Instituto Nacional de Saude (INSA) and Instituto Gulbenkian de Ciencia (IGC) | Instituto Nacional de Saude (INSA) and Instituto Gulbenkian de Ciencia (IGC) | Borges et al |
| EPI_ISL_693550 | Instituto Nacional de Saude (INSA) | Instituto Nacional de Saude (INSA) | Borges et al |
| EPI_ISL_693596 | Instituto Nacional de Saude (INSA) | Instituto Nacional de Saude (INSA) | Borges et al |

| | | | |
|---|---|---|---|
| EPI_ISL_693531 | Instituto Nacional de Saude (INSA) | Instituto Nacional de Saude (INSA) | Borges et al |
| EPI_ISL_693618 | Instituto Nacional de Saude (INSA) | Instituto Nacional de Saude (INSA) | Borges et al |
| EPI_ISL_693619 | Instituto Nacional de Saude (INSA) | Instituto Nacional de Saude (INSA) | Borges et al |
| EPI_ISL_693532 | Instituto Nacional de Saude (INSA) | Instituto Nacional de Saude (INSA) | Borges et al |
| EPI_ISL_693537 | Hospital Vila Franca de Xira | Instituto Nacional de Saude (INSA) | Borges et al |
| EPI_ISL_693536 | Hospital Vila Franca de Xira | Instituto Nacional de Saude (INSA) | Borges et al |
| EPI_ISL_693535 | Hospital Vila Franca de Xira | Instituto Nacional de Saude (INSA) | Borges et al |
| EPI_ISL_732116 | Instituto Nacional de Saude (INSA) | Instituto Nacional de Saude (INSA) | Borges et al |
| EPI_ISL_732179 | Instituto Nacional de Saude (INSA) and Instituto Gulbenkian de Ciencia (IGC) | Instituto Nacional de Saude (INSA) and Instituto Gulbenkian de Ciencia (IGC) | Borges et al |
| EPI_ISL_732180 | Instituto Nacional de Saude (INSA) and Instituto Gulbenkian de Ciencia (IGC) | Instituto Nacional de Saude (INSA) and Instituto Gulbenkian de Ciencia (IGC) | Borges et al |
| EPI_ISL_732183 | Instituto Nacional de Saude (INSA) and Instituto Gulbenkian de Ciencia (IGC) | Instituto Nacional de Saude (INSA) and Instituto Gulbenkian de Ciencia (IGC) | Borges et al |
| EPI_ISL_732184 | Instituto Nacional de Saude (INSA) and Instituto Gulbenkian de Ciencia (IGC) | Instituto Nacional de Saude (INSA) and Instituto Gulbenkian de Ciencia (IGC) | Borges et al |
| EPI_ISL_861535 | Instituto Nacional de Saude (INSA) | Instituto Nacional de Saude (INSA) | Borges et al |
| EPI_ISL_941550 | Instituto Nacional de Saude (INSA) | Instituto Nacional de Saude (INSA) | Borges et al |
| EPI_ISL_941552 | Instituto Nacional de Saude (INSA) | Instituto Nacional de Saude (INSA) | Borges et al |
| EPI_ISL_941583 | Instituto Nacional de Saude (INSA) | Instituto Nacional de Saude (INSA) | Borges et al |
| EPI_ISL_941896 | Instituto Nacional de Saude (INSA) and Instituto Gulbenkian de Ciencia (IGC) | Instituto Nacional de Saude (INSA) and Instituto Gulbenkian de Ciencia (IGC) | Borges et al |
| EPI_ISL_1023463 | Instituto Nacional de Saude (INSA) | Instituto Nacional de Saude (INSA) | Borges et al |
| EPI_ISL_1260860 | Instituto Nacional de Saude (INSA) | Instituto Nacional de Saude (INSA) | Borges et al |
| EPI_ISL_1399630 | Instituto Nacional de Saude (INSA) and BioSystems & Integrative Sciences Institute (BioISI) Genomics Unit, FCUL | Instituto Nacional de Saude (INSA) and BioSystems & Integrative Sciences Institute (BioISI) Genomics Unit, FCUL | Borges et al |
| EPI_ISL_1399631 | Instituto Nacional de Saude (INSA) and Instituto Gulbenkian de Ciencia (IGC) | Instituto Nacional de Saude (INSA) and Instituto Gulbenkian de Ciencia (IGC) | Borges et al |
| EPI_ISL_1399632 | Instituto Nacional de Saude (INSA) and Instituto Gulbenkian de Ciencia (IGC) | Instituto Nacional de Saude (INSA) and Instituto Gulbenkian de Ciencia (IGC) | Borges et al |
| EPI_ISL_428451 | Guangdong Provincial Center for Diseases Control and Prevention;Guangdong Provincial Institute of Public Health | School of Public Health, The University of Hong Kong | Bosheng Li et al |
| EPI_ISL_1060884 | DB Diagnosticos do Brasil | Instituto de Medicina Tropical de Sao Paulo | Brazil-UK Centre for Arbovirus Discovery Di |
| EPI_ISL_1060918 | DB Diagnosticos do Brasil | Instituto de Medicina Tropical de Sao Paulo | Brazil-UK Centre for Arbovirus Discovery Di |
| EPI_ISL_1060943 | DB Diagnosticos do Brasil | Instituto de Medicina Tropical de Sao Paulo | Brazil-UK Centre for Arbovirus Discovery Di |
| EPI_ISL_1060954 | DB Diagnosticos do Brasil | Instituto de Medicina Tropical de Sao Paulo | Brazil-UK Centre for Arbovirus Discovery Di |

agnosis Genomics et al
agnosis Genomics et al
agnosis Genomics et al
agnosis Genomics et al

| | | | |
|---|---|---|---|
| EPI_ISL_1060926 | DB Diagnosticos do Brasil | Instituto de Medicina Tropical de Sao Paulo | Brazil-UK Centre for Arbovirus Discovery Di |
| EPI_ISL_1060947 | DB Diagnosticos do Brasil | Instituto de Medicina Tropical de Sao Paulo | Brazil-UK Centre for Arbovirus Discovery Di |
| EPI_ISL_1060889 | CDL Laboratorio Santos e Vidal LTDA. | Instituto de Medicina Tropical de Sao Paulo | Brazil-UK Centre for Arbovirus Discovery Di |
| EPI_ISL_1060897 | CDL Laboratorio Santos e Vidal LTDA. | Instituto de Medicina Tropical de Sao Paulo | Brazil-UK Centre for Arbovirus Discovery Di |
| EPI_ISL_1060880 | CDL Laboratorio Santos e Vidal LTDA. | Instituto de Medicina Tropical de Sao Paulo | Brazil-UK Centre for Arbovirus Discovery Di |
| EPI_ISL_1064736 | CDL Laboratorio Santos e Vidal LTDA. | Instituto de Medicina Tropical de Sao Paulo | Brazil-UK Centre for Arbovirus Discovery Di |
| EPI_ISL_1060892 | CDL Laboratorio Santos e Vidal LTDA. | Instituto de Medicina Tropical de Sao Paulo | Brazil-UK Centre for Arbovirus Discovery Di |
| EPI_ISL_1060896 | CDL Laboratorio Santos e Vidal LTDA. | Instituto de Medicina Tropical de Sao Paulo | Brazil-UK Centre for Arbovirus Discovery Di |
| EPI_ISL_1060931 | CDL Laboratorio Santos e Vidal LTDA. | Instituto de Medicina Tropical de Sao Paulo | Brazil-UK Centre for Arbovirus Discovery Di |
| EPI_ISL_1060955 | CDL Laboratorio Santos e Vidal LTDA. | Instituto de Medicina Tropical de Sao Paulo | Brazil-UK Centre for Arbovirus Discovery Di |
| EPI_ISL_1060956 | CDL Laboratorio Santos e Vidal LTDA. | Instituto de Medicina Tropical de Sao Paulo | Brazil-UK Centre for Arbovirus Discovery Di |
| EPI_ISL_1060959 | CDL Laboratorio Santos e Vidal LTDA. | Instituto de Medicina Tropical de Sao Paulo | Brazil-UK Centre for Arbovirus Discovery Di |
| EPI_ISL_1060898 | CDL Laboratorio Santos e Vidal LTDA. | Instituto de Medicina Tropical de Sao Paulo | Brazil-UK Centre for Arbovirus Discovery Di |
| EPI_ISL_1060948 | CDL Laboratorio Santos e Vidal LTDA. | Instituto de Medicina Tropical de Sao Paulo | Brazil-UK Centre for Arbovirus Discovery Di |
| EPI_ISL_1060911 | CDL Laboratorio Santos e Vidal LTDA. | Instituto de Medicina Tropical de Sao Paulo | Brazil-UK Centre for Arbovirus Discovery Di |
| EPI_ISL_1060899 | CDL Laboratorio Santos e Vidal LTDA. | Instituto de Medicina Tropical de Sao Paulo | Brazil-UK Centre for Arbovirus Discovery Di |
| EPI_ISL_1060915 | CDL Laboratorio Santos e Vidal LTDA. | Instituto de Medicina Tropical de Sao Paulo | Brazil-UK Centre for Arbovirus Discovery Di |
| EPI_ISL_1060890 | CDL Laboratorio Santos e Vidal LTDA. | Instituto de Medicina Tropical de Sao Paulo | Brazil-UK Centre for Arbovirus Discovery Di |
| EPI_ISL_1064745 | CDL Laboratorio Santos e Vidal LTDA. | Instituto de Medicina Tropical de Sao Paulo | Brazil-UK Centre for Arbovirus Discovery Di |
| EPI_ISL_1060894 | CDL Laboratorio Santos e Vidal LTDA. | Instituto de Medicina Tropical de Sao Paulo | Brazil-UK Centre for Arbovirus Discovery Di |
| EPI_ISL_1060944 | CDL Laboratorio Santos e Vidal LTDA. | Instituto de Medicina Tropical de Sao Paulo | Brazil-UK Centre for Arbovirus Discovery Di |
| EPI_ISL_1060879 | CDL Laboratorio Santos e Vidal LTDA. | Instituto de Medicina Tropical de Sao Paulo | Brazil-UK Centre for Arbovirus Discovery Di |
| EPI_ISL_1060908 | CDL Laboratorio Santos e Vidal LTDA. | Instituto de Medicina Tropical de Sao Paulo | Brazil-UK Centre for Arbovirus Discovery Di |
| EPI_ISL_1060877 | CDL Laboratorio Santos e Vidal LTDA. | Instituto de Medicina Tropical de Sao Paulo | Brazil-UK Centre for Arbovirus Discovery Di |
| EPI_ISL_1060921 | CDL Laboratorio Santos e Vidal LTDA. | Instituto de Medicina Tropical de Sao Paulo | Brazil-UK Centre for Arbovirus Discovery Di |
| EPI_ISL_1060881 | CDL Laboratorio Santos e Vidal LTDA. | Instituto de Medicina Tropical de Sao Paulo | Brazil-UK Centre for Arbovirus Discovery Di |
| EPI_ISL_1060907 | CDL Laboratorio Santos e Vidal LTDA. | Instituto de Medicina Tropical de Sao Paulo | Brazil-UK Centre for Arbovirus Discovery Di |
| EPI_ISL_722139 | DB Diagnosticos do Brasil | Laboratório de Parasitologia Médica - Instituto de Medicina Tropical - Universidade de São Paulo | Brazil-UK Centre for Arbovirus Discovery Di |
| EPI_ISL_722136 | DB Diagnosticos do Brasil | Laboratório de Parasitologia Médica - Instituto de Medicina Tropical - Universidade de São Paulo | Brazil-UK Centre for Arbovirus Discovery Di |

agnosis Genomics et al
agnosis Genomics et al
agnosis Genomics et al
agnosis Genomics et al
agnosis Genomics et al
agnosis Genomics et al
agnosis Genomics et al
agnosis Genomics et al
agnosis Genomics et al
agnosis Genomics et al
agnosis Genomics et al
agnosis Genomics et al
agnosis Genomics et al
agnosis Genomics et al
agnosis Genomics et al
agnosis Genomics et al
agnosis Genomics et al
agnosis Genomics et al
agnosis Genomics et al
agnosis Genomics et al
agnosis Genomics et al
agnosis Genomics et al
agnosis Genomics et al
agnosis Genomics et al
agnosis Genomics et al
agnosis Genomics et al
agnosis Genomics et al
agnosis Genomics et al
agnosis Genomics et al

| | | | |
|---|---|---|---|
| EPI_ISL_1084726 | DB Diagnosticos do Brasil | Laboratório de Parasitologia Médica - Instituto de Medicina Tropical - Universidade de São Paulo | Brazil-UK Centre for Arbovirus Discovery Di |
| EPI_ISL_1084730 | DB Diagnosticos do Brasil | Laboratório de Parasitologia Médica - Instituto de Medicina Tropical - Universidade de São Paulo | Brazil-UK Centre for Arbovirus Discovery Di |
| EPI_ISL_672666 | DB Diagnosticos do Brasil | Laboratório de Parasitologia Médica - Instituto de Medicina Tropical - Universidade de São Paulo | Brazil-UK Centre for Arbovirus Discovery Di |
| EPI_ISL_672667 | DB Diagnosticos do Brasil | Laboratório de Parasitologia Médica - Instituto de Medicina Tropical - Universidade de São Paulo | Brazil-UK Centre for Arbovirus Discovery Di |
| EPI_ISL_722146 | DB Diagnosticos do Brasil | Laboratório de Parasitologia Médica - Instituto de Medicina Tropical - Universidade de São Paulo | Brazil-UK Centre for Arbovirus Discovery Di |
| EPI_ISL_722140 | DB Diagnosticos do Brasil | Laboratório de Parasitologia Médica - Instituto de Medicina Tropical - Universidade de São Paulo | Brazil-UK Centre for Arbovirus Discovery Di |
| EPI_ISL_904018 | DB Diagnosticos do Brasil | Laboratório de Parasitologia Médica - Instituto de Medicina Tropical - Universidade de São Paulo | Brazil-UK Centre for Arbovirus Discovery Di |
| EPI_ISL_672672 | DB Diagnosticos do Brasil | Laboratório de Parasitologia Médica - Instituto de Medicina Tropical - Universidade de São Paulo | Brazil-UK Centre for Arbovirus Discovery Di |
| EPI_ISL_904024 | DB Diagnosticos do Brasil | Laboratório de Parasitologia Médica - Instituto de Medicina Tropical - Universidade de São Paulo | Brazil-UK Centre for Arbovirus Discovery Di |
| EPI_ISL_904025 | DB Diagnosticos do Brasil | Laboratório de Parasitologia Médica - Instituto de Medicina Tropical - Universidade de São Paulo | Brazil-UK Centre for Arbovirus Discovery Di |
| EPI_ISL_904026 | DB Diagnosticos do Brasil | Laboratório de Parasitologia Médica - Instituto de Medicina Tropical - Universidade de São Paulo | Brazil-UK Centre for Arbovirus Discovery Di |
| EPI_ISL_904028 | DB Diagnosticos do Brasil | Laboratório de Parasitologia Médica - Instituto de Medicina Tropical - Universidade de São Paulo | Brazil-UK Centre for Arbovirus Discovery Di |
| EPI_ISL_904029 | DB Diagnosticos do Brasil | Laboratório de Parasitologia Médica - Instituto de Medicina Tropical - Universidade de São Paulo | Brazil-UK Centre for Arbovirus Discovery Di |
| EPI_ISL_672670 | DB Diagnosticos do Brasil | Laboratório de Parasitologia Médica - Instituto de Medicina Tropical - Universidade de São Paulo | Brazil-UK Centre for Arbovirus Discovery Di |
| EPI_ISL_672675 | DB Diagnosticos do Brasil | Laboratório de Parasitologia Médica - Instituto de Medicina Tropical - Universidade de São Paulo | Brazil-UK Centre for Arbovirus Discovery Di |
| EPI_ISL_672679 | DB Diagnosticos do Brasil | Laboratório de Parasitologia Médica - Instituto de Medicina Tropical - Universidade de São Paulo | Brazil-UK Centre for Arbovirus Discovery Di |
| EPI_ISL_672681 | DB Diagnosticos do Brasil | Laboratório de Parasitologia Médica - Instituto de Medicina Tropical - Universidade de São Paulo | Brazil-UK Centre for Arbovirus Discovery Di |
| EPI_ISL_722148 | DB Diagnosticos do Brasil | Laboratório de Parasitologia Médica - Instituto de Medicina Tropical - Universidade de São Paulo | Brazil-UK Centre for Arbovirus Discovery Di |
| EPI_ISL_722134 | DB Diagnosticos do Brasil | Laboratório de Parasitologia Médica - Instituto de Medicina Tropical - Universidade de São Paulo | Brazil-UK Centre for Arbovirus Discovery Di |
| EPI_ISL_722135 | DB Diagnosticos do Brasil | Laboratório de Parasitologia Médica - Instituto de Medicina Tropical - Universidade de São Paulo | Brazil-UK Centre for Arbovirus Discovery Di |
| EPI_ISL_722149 | DB Diagnosticos do Brasil | Laboratório de Parasitologia Médica - Instituto de Medicina Tropical - Universidade de São Paulo | Brazil-UK Centre for Arbovirus Discovery Di |
| EPI_ISL_1009675 | Diagnósticos da América - DASA | Instituto de Medicina Tropical de Sao Paulo | Brazil-UK Centre for Arbovirus Discovery Di |

agnosis Genomics et al
agnosis Genomics et al
agnosis Genomics et al
agnosis Genomics et al
agnosis Genomics et al
agnosis Genomics et al
agnosis Genomics et al
agnosis Genomics et al
agnosis Genomics et al
agnosis Genomics et al
agnosis Genomics et al
agnosis Genomics et al
agnosis Genomics et al
agnosis Genomics et al
agnosis Genomics et al
agnosis Genomics et al
agnosis Genomics et al
agnosis Genomics et al
agnosis Genomics et al
agnosis Genomics et al
agnosis Genomics et al
agnosis Genomics et al

| | | | |
|---|---|---|---|
| EPI_ISL_722147 | DB Diagnosticos do Brasil | Laboratório de Parasitologia Médica - Instituto de Medicina Tropical - Universidade de São Paulo | Brazil-UK Centre for Arbovirus Discovery Di |
| EPI_ISL_672684 | DB Diagnosticos do Brasil | Laboratório de Parasitologia Médica - Instituto de Medicina Tropical - Universidade de São Paulo | Brazil-UK Centre for Arbovirus Discovery Di |
| EPI_ISL_722144 | DB Diagnosticos do Brasil | Laboratório de Parasitologia Médica - Instituto de Medicina Tropical - Universidade de São Paulo | Brazil-UK Centre for Arbovirus Discovery Di |
| EPI_ISL_672705 | Institute of Tropical Medicine at the University of São Paulo (IMT-USP) | Laboratório de Parasitologia Médica - Instituto de Medicina Tropical - Universidade de São Paulo | Brazil-UK Centre for Arbovirus Discovery Di |
| EPI_ISL_672712 | Institute of Tropical Medicine at the University of São Paulo (IMT-USP) | Laboratório de Parasitologia Médica - Instituto de Medicina Tropical - Universidade de São Paulo | Brazil-UK Centre for Arbovirus Discovery Di |
| EPI_ISL_672720 | Institute of Tropical Medicine at the University of São Paulo (IMT-USP) | Laboratório de Parasitologia Médica - Instituto de Medicina Tropical - Universidade de São Paulo | Brazil-UK Centre for Arbovirus Discovery Di |
| EPI_ISL_722004 | Hospital das Clínicas Universidade de São Paulo Medical School | Laboratório de Parasitologia Médica - Instituto de Medicina Tropical - Universidade de São Paulo | Brazil-UK Centre for Arbovirus Discovery Di |
| EPI_ISL_722002 | Hospital das Clínicas Universidade de São Paulo Medical School | Laboratório de Parasitologia Médica - Instituto de Medicina Tropical - Universidade de São Paulo | Brazil-UK Centre for Arbovirus Discovery Di |
| EPI_ISL_722040 | Hospital das Clínicas Universidade de São Paulo Medical School | Laboratório de Parasitologia Médica - Instituto de Medicina Tropical - Universidade de São Paulo | Brazil-UK Centre for Arbovirus Discovery Di |
| EPI_ISL_722022 | Hospital das Clínicas Universidade de São Paulo Medical School | Laboratório de Parasitologia Médica - Instituto de Medicina Tropical - Universidade de São Paulo | Brazil-UK Centre for Arbovirus Discovery Di |
| EPI_ISL_721995 | Hospital das Clínicas Universidade de São Paulo Medical School | Laboratório de Parasitologia Médica - Instituto de Medicina Tropical - Universidade de São Paulo | Brazil-UK Centre for Arbovirus Discovery Di |
| EPI_ISL_722010 | Hospital das Clínicas Universidade de São Paulo Medical School | Laboratório de Parasitologia Médica - Instituto de Medicina Tropical - Universidade de São Paulo | Brazil-UK Centre for Arbovirus Discovery Di |
| EPI_ISL_722017 | Hospital das Clínicas Universidade de São Paulo Medical School | Laboratório de Parasitologia Médica - Instituto de Medicina Tropical - Universidade de São Paulo | Brazil-UK Centre for Arbovirus Discovery Di |
| EPI_ISL_1171871 | IMT_USP | Laboratório de Parasitologia Médica - Instituto de Medicina Tropical - Universidade de São Paulo | Brazil-UK Centre for Arbovirus Discovery Di |
| EPI_ISL_1084729 | IMT_USP | Laboratório de Parasitologia Médica - Instituto de Medicina Tropical - Universidade de São Paulo | Brazil-UK Centre for Arbovirus Discovery Di |
| EPI_ISL_525996 | OHSU Lab Services Molecular Microbiology Lab | Oregon SARS-CoV-2 Genome Sequencing Center | Brendan L. O'Connell et al |
| EPI_ISL_1167936 | PA Department of Health, Bureau of Laboratories | Genomics and Discovery, Respiratory Viruses Branch, Division of Viral Diseases, Centers for Disease Control and Prevention | Brian Lynch et al |
| EPI_ISL_1492545 | Institute of Virology, Biomedical Research Center of the Slovak Academy of Sciences, Bratislava | Faculty of Natural Sciences, Comenius University, Bratislava | Brona Brejova et al |
| EPI_ISL_903986 | Institute of Virology, Biomedical Research Center of the Slovak Academy of Sciences, Bratislava | Faculty of Natural Sciences, Comenius University, Bratislava | Broňa Brejová et al |
| EPI_ISL_959632 | Institute of Virology, Biomedical Research Center of the Slovak Academy of Sciences, Bratislava | Faculty of Natural Sciences, Comenius University, Bratislava | Broňa Brejová et al |
| EPI_ISL_1239415 | Institute of Virology, Biomedical Research Center of the Slovak Academy of Sciences, Bratislava | Faculty of Natural Sciences, Comenius University, Bratislava | Broňa Brejová et al |
| EPI_ISL_577740 | Institute of Virology, Biomedical Research Center of the Slovak Academy of Sciences, Bratislava | Faculty of Natural Sciences, Comenius University, Bratislava | Broňa Brejová et al |

agnosis Genomics et al
agnosis Genomics et al
agnosis Genomics et al
agnosis Genomics et al
agnosis Genomics et al
agnosis Genomics et al
agnosis Genomics et al
agnosis Genomics et al
agnosis Genomics et al
agnosis Genomics et al
agnosis Genomics et al
agnosis Genomics et al
agnosis Genomics et al
agnosis Genomics et al
agnosis Genomics et al

| | | | |
|---|---|---|---|
| EPI_ISL_577741 | Institute of Virology, Biomedical Research Center of the Slovak Academy of Sciences, Bratislava | Faculty of Natural Sciences, Comenius University, Bratislava | Broňa Brejová et al |
| EPI_ISL_583487 | Institute of Virology, Biomedical Research Center of the Slovak Academy of Sciences, Bratislava | Faculty of Natural Sciences, Comenius University, Bratislava | Broňa Brejová et al |
| EPI_ISL_1405498 | Johns Hopkins Hospital Department of Pathology | Johns Hopkins Hospital Department of Pathology | C. Paul Morris et al |
| EPI_ISL_671343 | National Virus Reference Laboratory | Irish Coronavirus Sequencing Consortium - Teagasc Moorepark | Calm Walsh et al |
| EPI_ISL_767724 | National Virus Reference Laboratory | Irish Coronavirus Sequencing Consortium - Teagasc Moorepark | Calum Walsh et al |
| EPI_ISL_671381 | National Virus Reference Laboratory | Irish Coronavirus Sequencing Consortium - Teagasc Moorepark | Calum Walsh et al |
| EPI_ISL_671400 | National Virus Reference Laboratory | Irish Coronavirus Sequencing Consortium - Teagasc Moorepark | Calum Walsh et al |
| EPI_ISL_562239 | Victorian Infectious Diseases Reference Laboratory (VIDRL) | VIDRL and MDU-PHL | Caly et al |
| EPI_ISL_480778 | Victorian Infectious Diseases Reference Laboratory (VIDRL) | VIDRL and MDU-PHL | Caly L. et al |
| EPI_ISL_480779 | Victorian Infectious Diseases Reference Laboratory (VIDRL) | VIDRL and MDU-PHL | Caly L. et al |
| EPI_ISL_480781 | Victorian Infectious Diseases Reference Laboratory (VIDRL) | VIDRL and MDU-PHL | Caly L. et al |
| EPI_ISL_430637 | Victorian Infectious Diseases Reference Laboratory (VIDRL) | Microbiological Diagnostic Unit Public Health Laboratory and Victorian Infectious Diseases Reference Laboratory, The Peter Doherty Institute for Infection and Immunity | Caly L. et al |
| EPI_ISL_419733 | Victorian Infectious Diseases Reference Laboratory (VIDRL) | Victorian Infectious Diseases Reference Laboratory and Microbiological Diagnostic Unit Public Health Laboratory, Doherty Institute | Caly L. et al |
| EPI_ISL_419735 | Victorian Infectious Diseases Reference Laboratory (VIDRL) | Victorian Infectious Diseases Reference Laboratory and Microbiological Diagnostic Unit Public Health Laboratory, Doherty Institute | Caly L. et al |
| EPI_ISL_456535 | Victorian Infectious Diseases Reference Laboratory (VIDRL) | Microbiological Diagnostic Unit Public Health Laboratory and Victorian Infectious Diseases Reference Laboratory, Doherty Institute | Caly L. et al |
| EPI_ISL_456552 | Victorian Infectious Diseases Reference Laboratory (VIDRL) | Microbiological Diagnostic Unit Public Health Laboratory and Victorian Infectious Diseases Reference Laboratory, Doherty Institute | Caly L. et al |
| EPI_ISL_456571 | Victorian Infectious Diseases Reference Laboratory (VIDRL) | Microbiological Diagnostic Unit Public Health Laboratory and Victorian Infectious Diseases Reference Laboratory, Doherty Institute | Caly L. et al |
| EPI_ISL_456633 | Victorian Infectious Diseases Reference Laboratory (VIDRL) | Microbiological Diagnostic Unit Public Health Laboratory and Victorian Infectious Diseases Reference Laboratory, Doherty Institute | Caly L. et al |
| EPI_ISL_456635 | Victorian Infectious Diseases Reference Laboratory (VIDRL) | Microbiological Diagnostic Unit Public Health Laboratory and Victorian Infectious Diseases Reference Laboratory, Doherty Institute | Caly L. et al |
| EPI_ISL_779617 | Victorian Infectious Diseases Reference Laboratory (VIDRL) | VIDRL and MDU-PHL | Caly L. et al |
| EPI_ISL_521976 | Victorian Infectious Diseases Reference Laboratory (VIDRL) | VIDRL and MDU-PHL | Caly L. et al |
| EPI_ISL_520762 | Victorian Infectious Diseases Reference Laboratory (VIDRL) | VIDRL and MDU-PHL | Caly L. et al |
| EPI_ISL_1396344 | Laboratorio del Hospital Regional Ushuaia Gdor. Ernesto Campos | Nodo de Secuenciación Tierra del Fuego - Hospital Regional Ushuaia - Centro Austral De Investigaciones Científicas - Universidad Nacional De Tierra Del Fuego on behalf of 'Proyecto Argentino Interinstitucional de genomica de SARS-CoV-2' (PAIS Consortium) | Carina Andrea De Roccis et al |

| | | | |
|---|---|---|---|
| EPI_ISL_1396351 | Laboratorio del Hospital Regional Ushuaia Gdor. Ernesto Campos | Nodo de Secuenciación Tierra del Fuego - Hospital Regional Ushuaia - Centro Austral De Investigaciones Científicas - Universidad Nacional De Tierra Del Fuego on behalf of 'Proyecto Argentino Interinstitucional de genomica de SARS-CoV-2' (PAIS Consortium) | Carina Andrea De Roccis et al |
| EPI_ISL_1396353 | Laboratorio del Hospital Regional Ushuaia Gdor. Ernesto Campos | Nodo de Secuenciación Tierra del Fuego - Hospital Regional Ushuaia - Centro Austral De Investigaciones Científicas - Universidad Nacional De Tierra Del Fuego on behalf of 'Proyecto Argentino Interinstitucional de genomica de SARS-CoV-2' (PAIS Consortium) | Carina Andrea De Roccis et al |
| EPI_ISL_1396356 | Laboratorio del Hospital Regional Ushuaia Gdor. Ernesto Campos | Nodo de Secuenciación Tierra del Fuego - Hospital Regional Ushuaia - Centro Austral De Investigaciones Científicas - Universidad Nacional De Tierra Del Fuego on behalf of 'Proyecto Argentino Interinstitucional de genomica de SARS-CoV-2' (PAIS Consortium) | Carina Andrea De Roccis et al |
| EPI_ISL_691625 | Servicio de Microbiología, Hospital Universitario Son Espases | SeqCOVID-SPAIN consortium/IBV(CSIC) | Carla López-Causapé et al |
| EPI_ISL_431833 | National Institutes of Health, University of the Philippines Manila | Philippine Genome Center, University of the Philippines System | Carlo M. Lapid et al |
| EPI_ISL_434554 | National Institutes of Health, University of the Philippines Manila | Philippine Genome Center | Carlo M. Lapid et al |
| EPI_ISL_434555 | National Institutes of Health, University of the Philippines Manila | Philippine Genome Center | Carlo M. Lapid et al |
| EPI_ISL_434556 | National Institutes of Health, University of the Philippines Manila | Philippine Genome Center | Carlo M. Lapid et al |
| EPI_ISL_434557 | National Institutes of Health, University of the Philippines Manila | Philippine Genome Center | Carlo M. Lapid et al |
| EPI_ISL_649155 | Laboratorio de Investigaciones de Baney | University Hospital Basel, Clinical Bacteriology | Carlos Cortes et al |
| EPI_ISL_648305 | Laboratorio de Investigaciones de Baney | University Hospital Basel, Clinical Bacteriology | Carlos Cortes et al |
| EPI_ISL_953402 | Laboratorio de Investigaciones de Baney | Swiss Tropical and Public Health Institute | Carlos Cortes et al |
| EPI_ISL_648311 | Laboratorio de Investigaciones de Baney | University Hospital Basel, Clinical Bacteriology | Carlos Cortes et al |
| EPI_ISL_649160 | Laboratorio de Investigaciones de Baney | University Hospital Basel, Clinical Bacteriology | Carlos Cortes et al |
| EPI_ISL_648314 | Laboratorio de Investigaciones de Baney | University Hospital Basel, Clinical Bacteriology | Carlos Cortes et al |
| EPI_ISL_648364 | Laboratorio de Investigaciones de Baney | University Hospital Basel, Clinical Bacteriology | Carlos Cortes et al |
| EPI_ISL_953405 | Laboratorio de Investigaciones de Baney | Swiss Tropical and Public Health Institute | Carlos Cortes et al |
| EPI_ISL_648368 | Laboratorio de Investigaciones de Baney | University Hospital Basel, Clinical Bacteriology | Carlos Cortes et al |
| EPI_ISL_648316 | Laboratorio de Investigaciones de Baney | University Hospital Basel, Clinical Bacteriology | Carlos Cortes et al |
| EPI_ISL_648318 | Laboratorio de Investigaciones de Baney | University Hospital Basel, Clinical Bacteriology | Carlos Cortes et al |
| EPI_ISL_648319 | Laboratorio de Investigaciones de Baney | University Hospital Basel, Clinical Bacteriology | Carlos Cortes et al |
| EPI_ISL_953412 | Laboratorio de Investigaciones de Baney | Swiss Tropical and Public Health Institute | Carlos Cortes et al |
| EPI_ISL_648323 | Laboratorio de Investigaciones de Baney | University Hospital Basel, Clinical Bacteriology | Carlos Cortes et al |
| EPI_ISL_953408 | Laboratorio de Investigaciones de Baney | Swiss Tropical and Public Health Institute | Carlos Cortes et al |
| EPI_ISL_648329 | Laboratorio de Investigaciones de Baney | University Hospital Basel, Clinical Bacteriology | Carlos Cortes et al |
| EPI_ISL_648332 | Laboratorio de Investigaciones de Baney | University Hospital Basel, Clinical Bacteriology | Carlos Cortes et al |

| | | | |
|---|---|---|---|
| EPI_ISL_648378 | Laboratorio de Investigaciones de Baney | University Hospital Basel, Clinical Bacteriology | Carlos Cortes et al |
| EPI_ISL_648337 | Laboratorio de Investigaciones de Baney | University Hospital Basel, Clinical Bacteriology | Carlos Cortes et al |
| EPI_ISL_953411 | Laboratorio de Investigaciones de Baney | Swiss Tropical and Public Health Institute | Carlos Cortes et al |
| EPI_ISL_648338 | Laboratorio de Investigaciones de Baney | University Hospital Basel, Clinical Bacteriology | Carlos Cortes et al |
| EPI_ISL_648340 | Laboratorio de Investigaciones de Baney | University Hospital Basel, Clinical Bacteriology | Carlos Cortes et al |
| EPI_ISL_953403 | Laboratorio de Investigaciones de Baney | Swiss Tropical and Public Health Institute | Carlos Cortes et al |
| EPI_ISL_648349 | Laboratorio de Investigaciones de Baney | University Hospital Basel, Clinical Bacteriology | Carlos Cortes et al |
| EPI_ISL_648355 | Laboratorio de Investigaciones de Baney | University Hospital Basel, Clinical Bacteriology | Carlos Cortes et al |
| EPI_ISL_1093178 | Laboratorio de Referencia Nacional de Virus Respiratorio. Instituto Nacional de Salud Perú | Laboratorio de Referencia Nacional de Biotecnología y Biología Molecular. Instituto Nacional de Salud Perú | Carlos Padilla Rojas et al |
| EPI_ISL_1534641 | Laboratorio de Referencia Nacional de Virus Respiratorio. Instituto Nacional de Salud Perú | Laboratorio de Referencia Nacional de Biotecnología y Biología Molecular. Instituto Nacional de Salud Perú | Carlos Padilla Rojas et al |
| EPI_ISL_1532151 | Laboratorio de Referencia Nacional de Virus Respiratorio. Instituto Nacional de Salud Perú | Laboratorio de Referencia Nacional de Biotecnología y Biología Molecular. Instituto Nacional de Salud Perú | Carlos Padilla Rojas et al |
| EPI_ISL_1532158 | Laboratorio de Referencia Nacional de Virus Respiratorio. Instituto Nacional de Salud Perú | Laboratorio de Referencia Nacional de Biotecnología y Biología Molecular. Instituto Nacional de Salud Perú | Carlos Padilla Rojas et al |
| EPI_ISL_1534653 | Laboratorio de Referencia Nacional de Virus Respiratorio. Instituto Nacional de Salud Perú | Laboratorio de Referencia Nacional de Biotecnología y Biología Molecular. Instituto Nacional de Salud Perú | Carlos Padilla Rojas et al |
| EPI_ISL_1534655 | Laboratorio de Referencia Nacional de Virus Respiratorio. Instituto Nacional de Salud Perú | Laboratorio de Referencia Nacional de Biotecnología y Biología Molecular. Instituto Nacional de Salud Perú | Carlos Padilla Rojas et al |
| EPI_ISL_1093162 | Laboratorio de Referencia Nacional de Virus Respiratorio. Instituto Nacional de Salud Perú | Laboratorio de Referencia Nacional de Biotecnología y Biología Molecular. Instituto Nacional de Salud Perú | Carlos Padilla Rojas et al |
| EPI_ISL_1532197 | Laboratorio de Referencia Nacional de Virus Respiratorio. Instituto Nacional de Salud Perú | Laboratorio de Referencia Nacional de Biotecnología y Biología Molecular. Instituto Nacional de Salud Perú | Carlos Padilla Rojas et al |
| EPI_ISL_517531 | Laboratorio de Referencia Nacional de Virus Respiratorio. Centro Nacional de Salud Publica. Instituto Nacional de Salud Peru. | Laboratorio de Referencia Nacional de Biotecnología y Biología Molecular. Centro Nacional de Salud Publica. Instituto Nacional de Salud Peru. | Carlos Padilla Rojas et al |
| EPI_ISL_516647 | Laboratorio de Referencia Nacional de Virus Respiratorio. Centro Nacional de Salud Publica. Instituto Nacional de Salud Peru. | Laboratorio de Referencia Nacional de Biotecnología y Biología Molecular. Centro Nacional de Salud Publica. Instituto Nacional de Salud Peru. | Carlos Padilla Rojas et al |
| EPI_ISL_1532163 | Laboratorio de Referencia Nacional de Virus Respiratorio. Instituto Nacional de Salud Perú | Laboratorio de Referencia Nacional de Biotecnología y Biología Molecular. Instituto Nacional de Salud Perú | Carlos Padilla Rojas et al |
| EPI_ISL_1532164 | Laboratorio de Referencia Nacional de Virus Respiratorio. Instituto Nacional de Salud Perú | Laboratorio de Referencia Nacional de Biotecnología y Biología Molecular. Instituto Nacional de Salud Perú | Carlos Padilla Rojas et al |
| EPI_ISL_1093172 | Laboratorio de Referencia Nacional de Virus Respiratorio. Instituto Nacional de Salud Perú | Laboratorio de Referencia Nacional de Biotecnología y Biología Molecular. Instituto Nacional de Salud Perú | Carlos Padilla Rojas et al |
| EPI_ISL_1532199 | Laboratorio de Referencia Nacional de Virus Respiratorio. Instituto Nacional de Salud Perú | Laboratorio de Referencia Nacional de Biotecnología y Biología Molecular. Instituto Nacional de Salud Perú | Carlos Padilla Rojas et al |
| EPI_ISL_489988 | Laboratorio de Referencia Nacional de Virus Respiratorio. Instituto Nacional de Salud Perú | Laboratorio de Referencia Nacional de Biotecnología y Biología Molecular. Instituto Nacional de Salud Perú | Carlos Padilla Rojas et al |
| EPI_ISL_491427 | Laboratorio de Referencia Nacional de Virus Respiratorio. Instituto Nacional de Salud Perú | Laboratorio de Referencia Nacional de Biotecnología y Biología Molecular. Instituto Nacional de Salud Perú | Carlos Padilla Rojas et al |

| | | | |
|---|---|---|---|
| EPI_ISL_491429 | Laboratorio de Referencia Nacional de Virus Respiratorio. Instituto Nacional de Salud Perú | Laboratorio de Referencia Nacional de Biotecnología y Biología Molecular. Instituto Nacional de Salud Perú | Carlos Padilla Rojas et al |
| EPI_ISL_491430 | Laboratorio de Referencia Nacional de Virus Respiratorio. Instituto Nacional de Salud Perú | Laboratorio de Referencia Nacional de Biotecnología y Biología Molecular. Instituto Nacional de Salud Perú | Carlos Padilla Rojas et al |
| EPI_ISL_491432 | Laboratorio de Referencia Nacional de Virus Respiratorio. Instituto Nacional de Salud.Perú | Laboratorio de Referencia Nacional de Biotecnología y Biología Molecular. Instituto Nacional de Salud.Perú | Carlos Padilla Rojas et al |
| EPI_ISL_517959 | Laboratorio de Referencia Nacional de Virus Respiratorio. Centro Nacional de Salud Publica. Instituto Nacional de Salud Peru | Laboratorio de Referencia Nacional de Biotecnología y Biología Molecular. Centro Nacional de Salud Publica. Instituto Nacional de Salud Peru. | Carlos Padilla Rojas et al |
| EPI_ISL_516722 | Laboratorio de Referencia Nacional de Virus Respiratorio. Centro Nacional de Salud Publica. Instituto Nacional de Salud Peru. | Laboratorio de Referencia Nacional de Biotecnología y Biología Molecular. Centro Nacional de Salud Publica. Instituto Nacional de Salud Peru. | Carlos Padilla Rojas et al |
| EPI_ISL_1092344 | Laboratorio de Referencia Nacional de Virus Respiratorio. Instituto Nacional de Salud Perú | Laboratorio de Referencia Nacional de Biotecnología y Biología Molecular. Instituto Nacional de Salud Perú | Carlos Padilla Rojas et al |
| EPI_ISL_1092336 | Laboratorio de Referencia Nacional de Virus Respiratorio. Instituto Nacional de Salud Perú | Laboratorio de Referencia Nacional de Biotecnología y Biología Molecular. Instituto Nacional de Salud Perú | Carlos Padilla Rojas et al |
| EPI_ISL_1092353 | Laboratorio de Referencia Nacional de Virus Respiratorio. Instituto Nacional de Salud Perú | Laboratorio de Referencia Nacional de Biotecnología y Biología Molecular. Instituto Nacional de Salud Perú | Carlos Padilla Rojas et al |
| EPI_ISL_1093157 | Laboratorio de Referencia Nacional de Virus Respiratorio. Instituto Nacional de Salud Perú | Laboratorio de Referencia Nacional de Biotecnología y Biología Molecular. Instituto Nacional de Salud Perú | Carlos Padilla Rojas et al |
| EPI_ISL_1137488 | Laboratorio de Referencia Nacional de Virus Respiratorio. Instituto Nacional de Salud Perú | Laboratorio de Referencia Nacional de Biotecnología y Biología Molecular. Instituto Nacional de Salud Perú | Carlos Padilla Rojas et al |
| EPI_ISL_1137489 | Laboratorio de Referencia Nacional de Virus Respiratorio. Instituto Nacional de Salud Perú | Laboratorio de Referencia Nacional de Biotecnología y Biología Molecular. Instituto Nacional de Salud Perú | Carlos Padilla Rojas et al |
| EPI_ISL_1664258 | Laboratorio de Referencia Nacional de Virus Respiratorio. Instituto Nacional de Salud Perú | Laboratorio de Referencia Nacional de Biotecnología y Biología Molecular. Instituto Nacional de Salud Perú | Carlos Padilla Rojas et al |
| EPI_ISL_1532206 | Laboratorio de Referencia Nacional de Virus Respiratorio. Instituto Nacional de Salud Perú | Laboratorio de Referencia Nacional de Biotecnología y Biología Molecular. Instituto Nacional de Salud Perú | Carlos Padilla Rojas et al |
| EPI_ISL_1532225 | Laboratorio de Referencia Nacional de Virus Respiratorio. Instituto Nacional de Salud Perú | Laboratorio de Referencia Nacional de Biotecnología y Biología Molecular. Instituto Nacional de Salud Perú | Carlos Padilla Rojas et al |
| EPI_ISL_1532226 | Laboratorio de Referencia Nacional de Virus Respiratorio. Instituto Nacional de Salud Perú | Laboratorio de Referencia Nacional de Biotecnología y Biología Molecular. Instituto Nacional de Salud Perú | Carlos Padilla Rojas et al |
| EPI_ISL_1534627 | Laboratorio de Referencia Nacional de Virus Respiratorio. Instituto Nacional de Salud Perú | Laboratorio de Referencia Nacional de Biotecnología y Biología Molecular. Instituto Nacional de Salud Perú | Carlos Padilla Rojas et al |
| EPI_ISL_1534629 | Laboratorio de Referencia Nacional de Virus Respiratorio. Instituto Nacional de Salud Perú | Laboratorio de Referencia Nacional de Biotecnología y Biología Molecular. Instituto Nacional de Salud Perú | Carlos Padilla Rojas et al |
| EPI_ISL_1534630 | Laboratorio de Referencia Nacional de Virus Respiratorio. Instituto Nacional de Salud Perú | Laboratorio de Referencia Nacional de Biotecnología y Biología Molecular. Instituto Nacional de Salud Perú | Carlos Padilla Rojas et al |
| EPI_ISL_756293 | Center for Biotechnology and Cell Therapy, São Rafael Hospital, Salvador, Brazil | Center for Biotechnology and Cell Therapy, São Rafael Hospital, Salvador, Brazil | Carolina Kymie Vasques Nonaka et al |
| EPI_ISL_756294 | Center for Biotechnology and Cell Therapy, São Rafael Hospital, Salvador, Brazil | Center for Biotechnology and Cell Therapy, São Rafael Hospital, Salvador, Brazil | Carolina Kymie Vasques Nonaka et al |
| EPI_ISL_1608170 | Hospital São Rafael - IDOR | Hospital São Rafael - IDOR | Carolina Kymie Vasques Nonaka et al |
| EPI_ISL_623104 | Simile Medicina Diagnóstica | Bioinformatics Laboratory / LNCC | Carolina M Voloch et al |
| EPI_ISL_623105 | Simile Medicina Diagnóstica | Bioinformatics Laboratory / LNCC | Carolina M Voloch et al |

| | | | |
|---|---|---|---|
| EPI_ISL_623107 | Laboratorio de Virologia Molecular / UFRJ | Bioinformatics Laboratory / LNCC | Carolina M Voloch et al |
| EPI_ISL_623113 | Laboratorio de Virologia Molecular / UFRJ | Bioinformatics Laboratory / LNCC | Carolina M Voloch et al |
| EPI_ISL_623121 | Laboratorio de Virologia Molecular / UFRJ | Bioinformatics Laboratory / LNCC | Carolina M Voloch et al |
| EPI_ISL_623125 | Laboratorio de Virologia Molecular / UFRJ | Bioinformatics Laboratory / LNCC | Carolina M Voloch et al |
| EPI_ISL_623130 | Laboratorio de Virologia Molecular / UFRJ | Bioinformatics Laboratory / LNCC | Carolina M Voloch et al |
| EPI_ISL_623131 | Laboratorio de Virologia Molecular / UFRJ | Bioinformatics Laboratory / LNCC | Carolina M Voloch et al |
| EPI_ISL_623151 | Laboratorio de Virologia Molecular / UFRJ | Bioinformatics Laboratory / LNCC | Carolina M Voloch et al |
| EPI_ISL_623155 | Laboratorio de Virologia Molecular / UFRJ | Bioinformatics Laboratory / LNCC | Carolina M Voloch et al |
| EPI_ISL_623159 | Laboratorio de Virologia Molecular / UFRJ | Bioinformatics Laboratory / LNCC | Carolina M Voloch et al |
| EPI_ISL_623164 | Laboratorio de Virologia Molecular / UFRJ | Bioinformatics Laboratory / LNCC | Carolina M Voloch et al |
| EPI_ISL_717807 | Laboratorio de Virologia Molecular / UFRJ | Bioinformatics Laboratory / LNCC | Carolina M Voloch et al |
| EPI_ISL_717808 | Laboratorio de Virologia Molecular / UFRJ | Bioinformatics Laboratory / LNCC | Carolina M Voloch et al |
| EPI_ISL_717810 | Laboratorio de Virologia Molecular / UFRJ | Bioinformatics Laboratory / LNCC | Carolina M Voloch et al |
| EPI_ISL_717811 | Laboratorio de Virologia Molecular / UFRJ | Bioinformatics Laboratory / LNCC | Carolina M Voloch et al |
| EPI_ISL_717812 | Laboratorio de Virologia Molecular / UFRJ | Bioinformatics Laboratory / LNCC | Carolina M Voloch et al |
| EPI_ISL_717813 | Laboratorio de Virologia Molecular / UFRJ | Bioinformatics Laboratory / LNCC | Carolina M Voloch et al |
| EPI_ISL_717814 | Laboratorio de Virologia Molecular / UFRJ | Bioinformatics Laboratory / LNCC | Carolina M Voloch et al |
| EPI_ISL_717815 | Laboratorio de Virologia Molecular / UFRJ | Bioinformatics Laboratory / LNCC | Carolina M Voloch et al |
| EPI_ISL_717816 | Laboratorio de Virologia Molecular / UFRJ | Bioinformatics Laboratory / LNCC | Carolina M Voloch et al |
| EPI_ISL_717817 | Laboratorio de Virologia Molecular / UFRJ | Bioinformatics Laboratory / LNCC | Carolina M Voloch et al |
| EPI_ISL_717819 | Laboratorio de Virologia Molecular / UFRJ | Bioinformatics Laboratory / LNCC | Carolina M Voloch et al |
| EPI_ISL_717821 | Laboratorio de Virologia Molecular / UFRJ | Bioinformatics Laboratory / LNCC | Carolina M Voloch et al |
| EPI_ISL_717823 | Laboratorio de Virologia Molecular / UFRJ | Bioinformatics Laboratory / LNCC | Carolina M Voloch et al |
| EPI_ISL_717825 | Laboratorio de Virologia Molecular / UFRJ | Bioinformatics Laboratory / LNCC | Carolina M Voloch et al |
| EPI_ISL_717826 | Laboratorio de Virologia Molecular / UFRJ | Bioinformatics Laboratory / LNCC | Carolina M Voloch et al |
| EPI_ISL_717827 | Laboratorio de Virologia Molecular / UFRJ | Bioinformatics Laboratory / LNCC | Carolina M Voloch et al |
| EPI_ISL_717828 | Laboratorio de Virologia Molecular / UFRJ | Bioinformatics Laboratory / LNCC | Carolina M Voloch et al |
| EPI_ISL_717830 | Laboratorio de Virologia Molecular / UFRJ | Bioinformatics Laboratory / LNCC | Carolina M Voloch et al |
| EPI_ISL_717787 | LACEN RJ - Noel Nutels | Bioinformatics Laboratory / LNCC | Carolina M Voloch et al |
| EPI_ISL_717794 | LACEN RJ - Noel Nutels | Bioinformatics Laboratory / LNCC | Carolina M Voloch et al |

| | | | |
|---|---|---|---|
| EPI_ISL_717832 | LACEN Dr. Francisco Rimolo Neto | Bioinformatics Laboratory / LNCC | Carolina M Voloch et al |
| EPI_ISL_717835 | LACEN Dr. Francisco Rimolo Neto | Bioinformatics Laboratory / LNCC | Carolina M Voloch et al |
| EPI_ISL_717851 | Laboratorio de Virologia Molecular / UFRJ | Bioinformatics Laboratory / LNCC | Carolina M Voloch et al |
| EPI_ISL_717857 | Laboratorio de Virologia Molecular / UFRJ | Bioinformatics Laboratory / LNCC | Carolina M Voloch et al |
| EPI_ISL_717863 | Laboratorio de Virologia Molecular / UFRJ | Bioinformatics Laboratory / LNCC | Carolina M Voloch et al |
| EPI_ISL_717869 | Laboratorio de Virologia Molecular / UFRJ | Bioinformatics Laboratory / LNCC | Carolina M Voloch et al |
| EPI_ISL_717870 | Laboratorio de Virologia Molecular / UFRJ | Bioinformatics Laboratory / LNCC | Carolina M Voloch et al |
| EPI_ISL_717871 | Laboratorio de Virologia Molecular / UFRJ | Bioinformatics Laboratory / LNCC | Carolina M Voloch et al |
| EPI_ISL_717872 | Laboratorio de Virologia Molecular / UFRJ | Bioinformatics Laboratory / LNCC | Carolina M Voloch et al |
| EPI_ISL_717873 | Laboratorio de Virologia Molecular / UFRJ | Bioinformatics Laboratory / LNCC | Carolina M Voloch et al |
| EPI_ISL_717793 | Laboratorio de Virologia Molecular / UFRJ | Bioinformatics Laboratory / LNCC | Carolina M Voloch et al |
| EPI_ISL_717877 | Laboratorio de Virologia Molecular / UFRJ | Bioinformatics Laboratory / LNCC | Carolina M Voloch et al |
| EPI_ISL_717878 | Laboratorio de Virologia Molecular / UFRJ | Bioinformatics Laboratory / LNCC | Carolina M Voloch et al |
| EPI_ISL_717887 | Laboratorio de Virologia Molecular / UFRJ | Bioinformatics Laboratory / LNCC | Carolina M Voloch et al |
| EPI_ISL_717889 | Laboratorio de Virologia Molecular / UFRJ | Bioinformatics Laboratory / LNCC | Carolina M Voloch et al |
| EPI_ISL_717890 | Laboratorio de Virologia Molecular / UFRJ | Bioinformatics Laboratory / LNCC | Carolina M Voloch et al |
| EPI_ISL_717891 | Laboratorio de Virologia Molecular / UFRJ | Bioinformatics Laboratory / LNCC | Carolina M Voloch et al |
| EPI_ISL_717893 | Laboratorio de Virologia Molecular / UFRJ | Bioinformatics Laboratory / LNCC | Carolina M Voloch et al |
| EPI_ISL_717894 | Laboratorio de Virologia Molecular / UFRJ | Bioinformatics Laboratory / LNCC | Carolina M Voloch et al |
| EPI_ISL_717895 | Laboratorio de Virologia Molecular / UFRJ | Bioinformatics Laboratory / LNCC | Carolina M Voloch et al |
| EPI_ISL_717896 | Laboratorio de Virologia Molecular / UFRJ | Bioinformatics Laboratory / LNCC | Carolina M Voloch et al |
| EPI_ISL_717900 | LACEN RJ - Noel Nutels | Bioinformatics Laboratory / LNCC | Carolina M Voloch et al |
| EPI_ISL_717901 | LACEN RJ - Noel Nutels | Bioinformatics Laboratory / LNCC | Carolina M Voloch et al |
| EPI_ISL_717797 | LACEN RJ - Noel Nutels | Bioinformatics Laboratory / LNCC | Carolina M Voloch et al |
| EPI_ISL_717788 | LACEN RJ - Noel Nutels | Bioinformatics Laboratory / LNCC | Carolina M Voloch et al |
| EPI_ISL_717903 | LACEN RJ - Noel Nutels | Bioinformatics Laboratory / LNCC | Carolina M Voloch et al |
| EPI_ISL_717904 | LACEN RJ - Noel Nutels | Bioinformatics Laboratory / LNCC | Carolina M Voloch et al |
| EPI_ISL_717907 | LACEN RJ - Noel Nutels | Bioinformatics Laboratory / LNCC | Carolina M Voloch et al |
| EPI_ISL_717908 | LACEN RJ - Noel Nutels | Bioinformatics Laboratory / LNCC | Carolina M Voloch et al |
| EPI_ISL_717789 | LACEN RJ - Noel Nutels | Bioinformatics Laboratory / LNCC | Carolina M Voloch et al |

| | | | |
|---|---|---|---|
| EPI_ISL_717916 | LACEN Dr. Francisco Rimolo Neto | Bioinformatics Laboratory / LNCC | Carolina M Voloch et al |
| EPI_ISL_717919 | LACEN Dr. Francisco Rimolo Neto | Bioinformatics Laboratory / LNCC | Carolina M Voloch et al |
| EPI_ISL_717785 | LACEN RJ - Noel Nutels | Bioinformatics Laboratory / LNCC | Carolina M Voloch et al |
| EPI_ISL_717921 | Laboratorio de Virologia Molecular / UFRJ | Bioinformatics Laboratory / LNCC | Carolina M Voloch et al |
| EPI_ISL_717922 | Laboratorio de Virologia Molecular / UFRJ | Bioinformatics Laboratory / LNCC | Carolina M Voloch et al |
| EPI_ISL_717924 | Laboratorio de Virologia Molecular / UFRJ | Bioinformatics Laboratory / LNCC | Carolina M Voloch et al |
| EPI_ISL_717925 | Laboratorio de Virologia Molecular / UFRJ | Bioinformatics Laboratory / LNCC | Carolina M Voloch et al |
| EPI_ISL_717926 | Laboratorio de Virologia Molecular / UFRJ | Bioinformatics Laboratory / LNCC | Carolina M Voloch et al |
| EPI_ISL_717929 | Laboratorio de Virologia Molecular / UFRJ | Bioinformatics Laboratory / LNCC | Carolina M Voloch et al |
| EPI_ISL_717930 | Laboratorio de Virologia Molecular / UFRJ | Bioinformatics Laboratory / LNCC | Carolina M Voloch et al |
| EPI_ISL_717931 | Laboratorio de Virologia Molecular / UFRJ | Bioinformatics Laboratory / LNCC | Carolina M Voloch et al |
| EPI_ISL_717934 | Laboratorio de Virologia Molecular / UFRJ | Bioinformatics Laboratory / LNCC | Carolina M Voloch et al |
| EPI_ISL_717935 | Laboratorio de Virologia Molecular / UFRJ | Bioinformatics Laboratory / LNCC | Carolina M Voloch et al |
| EPI_ISL_717941 | Laboratorio de Virologia Molecular / UFRJ | Bioinformatics Laboratory / LNCC | Carolina M Voloch et al |
| EPI_ISL_717944 | Laboratorio de Virologia Molecular / UFRJ | Bioinformatics Laboratory / LNCC | Carolina M Voloch et al |
| EPI_ISL_717945 | Laboratorio de Virologia Molecular / UFRJ | Bioinformatics Laboratory / LNCC | Carolina M Voloch et al |
| EPI_ISL_717946 | Laboratorio de Virologia Molecular / UFRJ | Bioinformatics Laboratory / LNCC | Carolina M Voloch et al |
| EPI_ISL_717955 | Laboratorio de Virologia Molecular / UFRJ | Bioinformatics Laboratory / LNCC | Carolina M Voloch et al |
| EPI_ISL_717957 | Laboratorio de Virologia Molecular / UFRJ | Bioinformatics Laboratory / LNCC | Carolina M Voloch et al |
| EPI_ISL_717961 | Laboratorio de Virologia Molecular / UFRJ | Bioinformatics Laboratory / LNCC | Carolina M Voloch et al |
| EPI_ISL_717963 | LACEN Dr. Francisco Rimolo Neto | Bioinformatics Laboratory / LNCC | Carolina M Voloch et al |
| EPI_ISL_1632717 | Klinisk mikrobiologi, Region Västerbotten | CBRN Defence and Security, Swedish Defence Research Agency | Caroline Öhrman et al |
| EPI_ISL_960703 | Germano de sousa | Instituto Gulbenkian de Ciencia | Cathy Paulino et al |
| EPI_ISL_960728 | Germano de sousa | Instituto Gulbenkian de Ciencia | Cathy Paulino et al |
| EPI_ISL_515911 | California Department of Public Health | California Department of Public Health | CDPH IDLB COVIDNet et al |
| EPI_ISL_792467 | Laboratorio del Hospital Regional Ushuaia Gdor. Ernesto Campos | Hospital Regional Ushuaia - Centro Austral De Investigaciones Científicas - Universidad Nacional De Tierra Del Fuego on behalf of 'Proyecto Argentino Interinstitucional de genomica de SARS-CoV-2' (PAIS Consortium) | Ceballos et al |
| EPI_ISL_1190749 | CREMER(Centre de Rechercherches sur les Maladies Emergentes et Ré-émergentes) | TransVIHMI(Recherches Translationnelles sur le VIH et les Maladies Infectieuses) | Celestin Godwe et al |
| EPI_ISL_1190755 | CREMER(Centre de Rechercherches sur les Maladies Emergentes et Ré-émergentes) | TransVIHMI(Recherches Translationnelles sur le VIH et les Maladies Infectieuses) | Celestin Godwe et al |

| | | | |
|---|---|---|---|
| EPI_ISL_1190756 | CREMER(Centre de Rechercherches sur les Maladies Emergentes et Ré-émergentes) | TransVIHMI(Recherches Translationnelles sur le VIH et les Maladies Infectieuses) | Celestin Godwe et al |
| EPI_ISL_1190757 | CREMER(Centre de Rechercherches sur les Maladies Emergentes et Ré-émergentes) | TransVIHMI(Recherches Translationnelles sur le VIH et les Maladies Infectieuses) | Celestin Godwe et al |
| EPI_ISL_1190758 | CREMER(Centre de Rechercherches sur les Maladies Emergentes et Ré-émergentes) | TransVIHMI(Recherches Translationnelles sur le VIH et les Maladies Infectieuses) | Celestin Godwe et al |
| EPI_ISL_1190759 | CREMER(Centre de Rechercherches sur les Maladies Emergentes et Ré-émergentes) | TransVIHMI(Recherches Translationnelles sur le VIH et les Maladies Infectieuses) | Celestin Godwe et al |
| EPI_ISL_1190760 | CREMER(Centre de Rechercherches sur les Maladies Emergentes et Ré-émergentes) | TransVIHMI(Recherches Translationnelles sur le VIH et les Maladies Infectieuses) | Celestin Godwe et al |
| EPI_ISL_1190761 | CREMER(Centre de Rechercherches sur les Maladies Emergentes et Ré-émergentes) | TransVIHMI(Recherches Translationnelles sur le VIH et les Maladies Infectieuses) | Celestin Godwe et al |
| EPI_ISL_1190765 | CREMER(Centre de Rechercherches sur les Maladies Emergentes et Ré-émergentes) | TransVIHMI(Recherches Translationnelles sur le VIH et les Maladies Infectieuses) | Celestin Godwe et al |
| EPI_ISL_1190768 | CREMER(Centre de Rechercherches sur les Maladies Emergentes et Ré-émergentes) | TransVIHMI(Recherches Translationnelles sur le VIH et les Maladies Infectieuses) | Celestin Godwe et al |
| EPI_ISL_1190752 | CREMER(Centre de Rechercherches sur les Maladies Emergentes et Ré-émergentes) | TransVIHMI(Recherches Translationnelles sur le VIH et les Maladies Infectieuses) | Celestin Godwe et al |
| EPI_ISL_837621 | Instituto Nacional de Enfermedades Respiratorias (INER) | Instituto Nacional de Enfermedades Respiratorias (INER) | Celia Boukadida et al |
| EPI_ISL_837694 | Instituto Nacional de Enfermedades Respiratorias (INER) | Instituto Nacional de Enfermedades Respiratorias (INER) | Celia Boukadida et al |
| EPI_ISL_837736 | Instituto Nacional de Enfermedades Respiratorias (INER) | Instituto Nacional de Enfermedades Respiratorias (INER) | Celia Boukadida et al |
| EPI_ISL_1620797 | Plateforme de testing Namuroise | Plateforme de testing Namuroise | Céline Maschietto et al |
| EPI_ISL_1620773 | Plateforme de testing Namuroise | Plateforme de testing Namuroise | Céline Maschietto et al |
| EPI_ISL_1670621 | Plateforme de testing Namuroise | Plateforme de testing Namuroise | Céline Maschietto et al |
| EPI_ISL_451302 | Laboratory of Virology, INMI Lazzaro Spallanzani IRCCS | Laboratory of Virology, INMI Lazzaro Spallanzani IRCCS | Cesare E.M. Gruber et al |
| EPI_ISL_1318216 | Ministry of Health Hospitals | Institute of Health and Community Medicine | Chan Chia Jui et al |
| EPI_ISL_1318217 | Ministry of Health Hospitals | Institute of Health and Community Medicine | Chan Chia Jui et al |
| EPI_ISL_434571 | Microbiology, The University of Hong Kong | Microbiology, The University of Hong Kong | Chan et al |
| EPI_ISL_1184582 | Genelabs Medical (Pvt) Ltd | Genelabs Medical (Pvt) Ltd | Chandanamali Punchihewa et al |
| EPI_ISL_862717 | Centre for Dengue Research and AICBU, Department of Immunology and Molecular Medicine | Centre for Dengue Research and AICBU, Department of Immunology and Molecular Medicine | Chandima Jeewandara et al |
| EPI_ISL_862719 | Centre for Dengue Research and AICBU, Department of Immunology and Molecular Medicine | Centre for Dengue Research and AICBU, Department of Immunology and Molecular Medicine | Chandima Jeewandara et al |
| EPI_ISL_668447 | Centre for Dengue Research, Department of Immunology and Molecular Medicine | Centre for Dengue Research, Department of Immunology and Molecular Medicine | Chandima Jeewandara et al |
| EPI_ISL_668448 | Centre for Dengue Research, Department of Immunology and Molecular Medicine | Centre for Dengue Research, Department of Immunology and Molecular Medicine | Chandima Jeewandara et al |
| EPI_ISL_668453 | Centre for Dengue Research, Department of Immunology and Molecular Medicine | Centre for Dengue Research, Department of Immunology and Molecular Medicine | Chandima Jeewandara et al |

| | | | |
|---|---|---|---|
| EPI_ISL_668449 | Centre for Dengue Research, Department of Immunology and Molecular Medicine | Centre for Dengue Research, Department of Immunology and Molecular Medicine | Chandima Jeewandara et al |
| EPI_ISL_668450 | Centre for Dengue Research, Department of Immunology and Molecular Medicine | Centre for Dengue Research, Department of Immunology and Molecular Medicine | Chandima Jeewandara et al |
| EPI_ISL_668452 | Centre for Dengue Research, Department of Immunology and Molecular Medicine | Centre for Dengue Research, Department of Immunology and Molecular Medicine | Chandima Jeewandara et al |
| EPI_ISL_978919 | Centre for Dengue Research and AICBU, Department of Immunology and Molecular Medicine | Centre for Dengue Research and AICBU, Department of Immunology and Molecular Medicine | Chandima Jeewandara et al |
| EPI_ISL_525488 | Centre for Dengue Research | Centre for Dengue Research | Chandima Jeewandara et al |
| EPI_ISL_525489 | Centre for Dengue Research | Centre for Dengue Research | Chandima Jeewandara et al |
| EPI_ISL_525486 | Centre for Dengue Research | Centre for Dengue Research | Chandima Jeewandara et al |
| EPI_ISL_525474 | Centre for Dengue Research | Centre for Dengue Research, USJ, SL | Chandima Jeewandara et al |
| EPI_ISL_525476 | Centre for Dengue Research | Centre for Dengue Research | Chandima Jeewandara et al |
| EPI_ISL_525478 | Centre for Dengue Research | Centre for Dengue Research | Chandima Jeewandara et al |
| EPI_ISL_525479 | Centre for Dengue Research | Centre for Dengue Research | Chandima Jeewandara et al |
| EPI_ISL_1533806 | Centre for Dengue Research and AICBU, Department of Immunology and Molecular Medicine | Centre for Dengue Research and AICBU, Department of Immunology and Molecular Medicine | Chandima Jeewandara et al |
| EPI_ISL_1233112 | Centre for Dengue Research and AICBU, Department of Immunology and Molecular Medicine | Centre for Dengue Research and AICBU, Department of Immunology and Molecular Medicine | Chandima Jeewandara et al |
| EPI_ISL_1533821 | Centre for Dengue Research and AICBU, Department of Immunology and Molecular Medicine | Centre for Dengue Research and AICBU, Department of Immunology and Molecular Medicine | Chandima Jeewandara et al |
| EPI_ISL_602565 | Centre for Dengue Research, Department of Immunology and Molecular Medicine | Centre for Dengue Research, Department of Immunology and Molecular Medicine, | Chandima Jeewandara et al |
| EPI_ISL_792548 | Centre for Dengue Research and AICBU, Department of Immunology and Molecular Medicine | Centre for Dengue Research and AICBU, Department of Immunology and Molecular Medicine | Chandima Jeewandara et al |
| EPI_ISL_792549 | Centre for Dengue Research and AICBU, Department of Immunology and Molecular Medicine | Centre for Dengue Research and AICBU, Department of Immunology and Molecular Medicine | Chandima Jeewandara et al |
| EPI_ISL_978920 | Centre for Dengue Research and AICBU, Department of Immunology and Molecular Medicine | Centre for Dengue Research and AICBU, Department of Immunology and Molecular Medicine | Chandima Jeewandara et al |
| EPI_ISL_792550 | Centre for Dengue Research and AICBU, Department of Immunology and Molecular Medicine | Centre for Dengue Research and AICBU, Department of Immunology and Molecular Medicine | Chandima Jeewandara et al |
| EPI_ISL_978921 | Centre for Dengue Research and AICBU, Department of Immunology and Molecular Medicine | Centre for Dengue Research and AICBU, Department of Immunology and Molecular Medicine | Chandima Jeewandara et al |
| EPI_ISL_978923 | Centre for Dengue Research and AICBU, Department of Immunology and Molecular Medicine | Centre for Dengue Research and AICBU, Department of Immunology and Molecular Medicine | Chandima Jeewandara et al |
| EPI_ISL_978986 | Centre for Dengue Research and AICBU, Department of Immunology and Molecular Medicine | Centre for Dengue Research and AICBU, Department of Immunology and Molecular Medicine | Chandima Jeewandara et al |
| EPI_ISL_1233096 | Centre for Dengue Research and AICBU, Department of Immunology and Molecular Medicine | Centre for Dengue Research and AICBU, Department of Immunology and Molecular Medicine | Chandima Jeewandara et al |
| EPI_ISL_1533823 | Centre for Dengue Research and AICBU, Department of Immunology and Molecular Medicine | Centre for Dengue Research and AICBU, Department of Immunology and Molecular Medicine | Chandima Jeewandara et al |

| | | | |
|---|---|---|---|
| EPI_ISL_1533831 | Centre for Dengue Research and AICBU, Department of Immunology and Molecular Medicine | Centre for Dengue Research and AICBU, Department of Immunology and Molecular Medicine | Chandima Jeewandara et al |
| EPI_ISL_1533836 | Centre for Dengue Research and AICBU, Department of Immunology and Molecular Medicine | Centre for Dengue Research and AICBU, Department of Immunology and Molecular Medicine | Chandima Jeewandara et al |
| EPI_ISL_428670 | Centre for Dengue Research | Centre for Dengue Research | Chandima Jeewandara et al |
| EPI_ISL_428673 | Centre for Dengue Research | Centre for Dengue Research | Chandima Jeewandara et al |
| EPI_ISL_428672 | Centre for Dengue Research | Centre for Dengue Research | Chandima Jeewandara et al |
| EPI_ISL_1533838 | Centre for Dengue Research and AICBU, Department of Immunology and Molecular Medicine | Centre for Dengue Research and AICBU, Department of Immunology and Molecular Medicine | Chandima Jeewandara et al |
| EPI_ISL_996060 | Centre for Dengue Research and AICBU, Department of Immunology and Molecular Medicine | Centre for Dengue Research and AICBU, Department of Immunology and Molecular Medicine | Chandima Jeewandara et al |
| EPI_ISL_996036 | Centre for Dengue Research and AICBU, Department of Immunology and Molecular Medicine | Centre for Dengue Research and AICBU, Department of Immunology and Molecular Medicine | Chandima Jeewandara et al |
| EPI_ISL_1233114 | Centre for Dengue Research and AICBU, Department of Immunology and Molecular Medicine | Centre for Dengue Research and AICBU, Department of Immunology and Molecular Medicine | Chandima Jeewandara et al |
| EPI_ISL_792552 | Centre for Dengue Research and AICBU, Department of Immunology and Molecular Medicine | Centre for Dengue Research and AICBU, Department of Immunology and Molecular Medicine | Chandima Jeewandara et al |
| EPI_ISL_1233061 | Centre for Dengue Research and AICBU, Department of Immunology and Molecular Medicine | Centre for Dengue Research and AICBU, Department of Immunology and Molecular Medicine | Chandima Jeewandara et al |
| EPI_ISL_1533848 | Centre for Dengue Research and AICBU, Department of Immunology and Molecular Medicine | Centre for Dengue Research and AICBU, Department of Immunology and Molecular Medicine | Chandima Jeewandara et al |
| EPI_ISL_1582415 | Centre for Dengue Research and AICBU, Department of Immunology and Molecular Medicine | Centre for Dengue Research and AICBU, Department of Immunology and Molecular Medicine | Chandima Jeewandara et al |
| EPI_ISL_792556 | Centre for Dengue Research and AICBU, Department of Immunology and Molecular Medicine | Centre for Dengue Research and AICBU, Department of Immunology and Molecular Medicine | Chandima Jeewandara et al |
| EPI_ISL_602566 | Centre for Dengue Research, Department of Immunology and Molecular Medicine | Centre for Dengue Research, Department of Immunology and Molecular Medicine | Chandima Jeewandara et al |
| EPI_ISL_668454 | Centre for Dengue Research, Department of Immunology and Molecular Medicine | Centre for Dengue Research, Department of Immunology and Molecular Medicine | Chandima Jeewandara et al |
| EPI_ISL_1662584 | Molecular diagnostic unit for viral haemorrhagic fevers and emerging viruses, Bouaké CHU Laboratory | Project group Epidemology of Highly Pathogenic Microorganisms, Robert Koch-Institute | Chantal Akoua-Koffi et al |
| EPI_ISL_1662591 | Molecular diagnostic unit for viral haemorrhagic fevers and emerging viruses, Bouaké CHU Laboratory | Project group Epidemology of Highly Pathogenic Microorganisms, Robert Koch-Institute | Chantal Akoua-Koffi et al |
| EPI_ISL_1663657 | Molecular diagnostic unit for viral haemorrhagic fevers and emerging viruses, Bouaké CHU Laboratory | Molecular diagnostic unit for viral haemorrhagic fevers and emerging viruses, Bouaké CHU Laboratory | Chantal Akoua-Koffi et al |
| EPI_ISL_1663664 | Molecular diagnostic unit for viral haemorrhagic fevers and emerging viruses, Bouaké CHU Laboratory | Molecular diagnostic unit for viral haemorrhagic fevers and emerging viruses, Bouaké CHU Laboratory | Chantal Akoua-Koffi et al |
| EPI_ISL_1663669 | Molecular diagnostic unit for viral haemorrhagic fevers and emerging viruses, Bouaké CHU Laboratory | Molecular diagnostic unit for viral haemorrhagic fevers and emerging viruses, Bouaké CHU Laboratory | Chantal Akoua-Koffi et al |
| EPI_ISL_1663661 | Molecular diagnostic unit for viral haemorrhagic fevers and emerging viruses, Bouaké CHU Laboratory | Molecular diagnostic unit for viral haemorrhagic fevers and emerging viruses, Bouaké CHU Laboratory | Chantal Akoua-Koffi et al |
| EPI_ISL_1663662 | Molecular diagnostic unit for viral haemorrhagic fevers and emerging viruses, Bouaké CHU Laboratory | Molecular diagnostic unit for viral haemorrhagic fevers and emerging viruses, Bouaké CHU Laboratory | Chantal Akoua-Koffi et al |

| | | | |
|---|---|---|---|
| EPI_ISL_1663670 | Molecular diagnostic unit for viral haemorrhagic fevers and emerging viruses, Bouaké CHU Laboratory | Molecular diagnostic unit for viral haemorrhagic fevers and emerging viruses, Bouaké CHU Laboratory | Chantal Akoua-Koffi et al |
| EPI_ISL_1663673 | Molecular diagnostic unit for viral haemorrhagic fevers and emerging viruses, Bouaké CHU Laboratory | Molecular diagnostic unit for viral haemorrhagic fevers and emerging viruses, Bouaké CHU Laboratory | Chantal Akoua-Koffi et al |
| EPI_ISL_1663659 | Molecular diagnostic unit for viral haemorrhagic fevers and emerging viruses, Bouaké CHU Laboratory | Molecular diagnostic unit for viral haemorrhagic fevers and emerging viruses, Bouaké CHU Laboratory | Chantal Akoua-Koffi et al |
| EPI_ISL_1663656 | Molecular diagnostic unit for viral haemorrhagic fevers and emerging viruses, Bouaké CHU Laboratory | Molecular diagnostic unit for viral haemorrhagic fevers and emerging viruses, Bouaké CHU Laboratory | Chantal Akoua-Koffi et al |
| EPI_ISL_1663674 | Molecular diagnostic unit for viral haemorrhagic fevers and emerging viruses, Bouaké CHU Laboratory | Molecular diagnostic unit for viral haemorrhagic fevers and emerging viruses, Bouaké CHU Laboratory | Chantal Akoua-Koffi et al |
| EPI_ISL_1663677 | Molecular diagnostic unit for viral haemorrhagic fevers and emerging viruses, Bouaké CHU Laboratory | Molecular diagnostic unit for viral haemorrhagic fevers and emerging viruses, Bouaké CHU Laboratory | Chantal Akoua-Koffi et al |
| EPI_ISL_1663680 | Molecular diagnostic unit for viral haemorrhagic fevers and emerging viruses, Bouaké CHU Laboratory | Molecular diagnostic unit for viral haemorrhagic fevers and emerging viruses, Bouaké CHU Laboratory | Chantal Akoua-Koffi et al |
| EPI_ISL_1663678 | Molecular diagnostic unit for viral haemorrhagic fevers and emerging viruses, Bouaké CHU Laboratory | Molecular diagnostic unit for viral haemorrhagic fevers and emerging viruses, Bouaké CHU Laboratory | Chantal Akoua-Koffi et al |
| EPI_ISL_1663679 | Molecular diagnostic unit for viral haemorrhagic fevers and emerging viruses, Bouaké CHU Laboratory | Molecular diagnostic unit for viral haemorrhagic fevers and emerging viruses, Bouaké CHU Laboratory | Chantal Akoua-Koffi et al |
| EPI_ISL_1365032 | Molecular diagnostic unit for viral haemorrhagic fevers and emerging viruses, Bouaké CHU Laboratory | Molecular diagnostic unit for viral haemorrhagic fevers and emerging viruses, Bouaké CHU Laboratory | Chantal Akoua-Koffi et al |
| EPI_ISL_1365027 | Molecular diagnostic unit for viral haemorrhagic fevers and emerging viruses, Bouaké CHU Laboratory | Molecular diagnostic unit for viral haemorrhagic fevers and emerging viruses, Bouaké CHU Laboratory | Chantal Akoua-Koffi et al |
| EPI_ISL_614348 | Molecular diagnostic unit for viral haemorrhagic fevers and emerging viruses, Bouaké CHU Laboratory | Project group Epidemology of Highly Pathogenic Microorganisms, Robert Koch-Institute | Chantal Akoua-Koffi et al |
| EPI_ISL_614349 | Molecular diagnostic unit for viral haemorrhagic fevers and emerging viruses, Bouaké CHU Laboratory | Project group Epidemology of Highly Pathogenic Microorganisms, Robert Koch-Institute | Chantal Akoua-Koffi et al |
| EPI_ISL_614350 | Molecular diagnostic unit for viral haemorrhagic fevers and emerging viruses, Bouaké CHU Laboratory | Project group Epidemology of Highly Pathogenic Microorganisms, Robert Koch-Institute | Chantal Akoua-Koffi et al |
| EPI_ISL_614363 | Molecular diagnostic unit for viral haemorrhagic fevers and emerging viruses, Bouaké CHU Laboratory | Project group Epidemology of Highly Pathogenic Microorganisms, Robert Koch-Institute | Chantal Akoua-Koffi et al |
| EPI_ISL_614366 | Molecular diagnostic unit for viral haemorrhagic fevers and emerging viruses, Bouaké CHU Laboratory | Project group Epidemology of Highly Pathogenic Microorganisms, Robert Koch-Institute | Chantal Akoua-Koffi et al |
| EPI_ISL_614370 | Molecular diagnostic unit for viral haemorrhagic fevers and emerging viruses, Bouaké CHU Laboratory | Project group Epidemology of Highly Pathogenic Microorganisms, Robert Koch-Institute | Chantal Akoua-Koffi et al |
| EPI_ISL_614371 | Molecular diagnostic unit for viral haemorrhagic fevers and emerging viruses, Bouaké CHU Laboratory | Project group Epidemology of Highly Pathogenic Microorganisms, Robert Koch-Institute | Chantal Akoua-Koffi et al |
| EPI_ISL_614373 | Molecular diagnostic unit for viral haemorrhagic fevers and emerging viruses, Bouaké CHU Laboratory | Project group Epidemology of Highly Pathogenic Microorganisms, Robert Koch-Institute | Chantal Akoua-Koffi et al |
| EPI_ISL_614374 | Molecular diagnostic unit for viral haemorrhagic fevers and emerging viruses, Bouaké CHU Laboratory | Project group Epidemology of Highly Pathogenic Microorganisms, Robert Koch-Institute | Chantal Akoua-Koffi et al |
| EPI_ISL_682059 | Molecular diagnostic unit for viral haemorrhagic fevers and emerging viruses, Bouaké CHU Laboratory | Project group Epidemology of Highly Pathogenic Microorganisms, Robert Koch-Institute | Chantal Akoua-Koffi et al |
| EPI_ISL_681837 | Molecular diagnostic unit for viral haemorrhagic fevers and emerging viruses, Bouaké CHU Laboratory | Project group Epidemology of Highly Pathogenic Microorganisms, Robert Koch-Institute | Chantal Akoua-Koffi et al |

| | | | |
|---|---|---|---|
| EPI_ISL_681839 | Molecular diagnostic unit for viral haemorrhagic fevers and emerging viruses, Bouaké CHU Laboratory | Project group Epidemology of Highly Pathogenic Microorganisms, Robert Koch-Institute | Chantal Akoua-Koffi et al |
| EPI_ISL_614387 | Molecular diagnostic unit for viral haemorrhagic fevers and emerging viruses, Bouaké CHU Laboratory | Project group Epidemology of Highly Pathogenic Microorganisms, Robert Koch-Institute | Chantal Akoua-Koffi et al |
| EPI_ISL_614390 | Molecular diagnostic unit for viral haemorrhagic fevers and emerging viruses, Bouaké CHU Laboratory | Project group Epidemology of Highly Pathogenic Microorganisms, Robert Koch-Institute | Chantal Akoua-Koffi et al |
| EPI_ISL_614391 | Molecular diagnostic unit for viral haemorrhagic fevers and emerging viruses, Bouaké CHU Laboratory | Project group Epidemology of Highly Pathogenic Microorganisms, Robert Koch-Institute | Chantal Akoua-Koffi et al |
| EPI_ISL_614392 | Molecular diagnostic unit for viral haemorrhagic fevers and emerging viruses, Bouaké CHU Laboratory | Project group Epidemology of Highly Pathogenic Microorganisms, Robert Koch-Institute | Chantal Akoua-Koffi et al |
| EPI_ISL_1367697 | Molecular diagnostic unit for viral haemorrhagic fevers and emerging viruses, Bouaké CHU Laboratory | Molecular diagnostic unit for viral haemorrhagic fevers and emerging viruses, Bouaké CHU Laboratory | Chantal Akoua-Koffi et al |
| EPI_ISL_1367689 | Molecular diagnostic unit for viral haemorrhagic fevers and emerging viruses, Bouaké CHU Laboratory | Molecular diagnostic unit for viral haemorrhagic fevers and emerging viruses, Bouaké CHU Laboratory | Chantal Akoua-Koffi et al |
| EPI_ISL_1367698 | Molecular diagnostic unit for viral haemorrhagic fevers and emerging viruses, Bouaké CHU Laboratory | Molecular diagnostic unit for viral haemorrhagic fevers and emerging viruses, Bouaké CHU Laboratory | Chantal Akoua-Koffi et al |
| EPI_ISL_1367695 | Molecular diagnostic unit for viral haemorrhagic fevers and emerging viruses, Bouaké CHU Laboratory | Molecular diagnostic unit for viral haemorrhagic fevers and emerging viruses, Bouaké CHU Laboratory | Chantal Akoua-Koffi et al |
| EPI_ISL_682058 | Molecular diagnostic unit for viral haemorrhagic fevers and emerging viruses, Bouaké CHU Laboratory | Project group Epidemology of Highly Pathogenic Microorganisms, Robert Koch-Institute | Chantal Akoua-Koffi et al |
| EPI_ISL_729091 | Viollier AG | Department of Biosystems Science and Engineering, ETH Zürich | Chaoran Chen et al |
| EPI_ISL_1408304 | Viollier AG | Department of Biosystems Science and Engineering, ETH Zürich | Chaoran Chen et al |
| EPI_ISL_1408276 | Viollier AG | Department of Biosystems Science and Engineering, ETH Zürich | Chaoran Chen et al |
| EPI_ISL_1408279 | Viollier AG | Department of Biosystems Science and Engineering, ETH Zürich | Chaoran Chen et al |
| EPI_ISL_1407582 | Viollier AG | Department of Biosystems Science and Engineering, ETH Zürich | Chaoran Chen et al |
| EPI_ISL_1408265 | Viollier AG | Department of Biosystems Science and Engineering, ETH Zürich | Chaoran Chen et al |
| EPI_ISL_1407598 | Viollier AG | Department of Biosystems Science and Engineering, ETH Zürich | Chaoran Chen et al |
| EPI_ISL_1407551 | Viollier AG | Department of Biosystems Science and Engineering, ETH Zürich | Chaoran Chen et al |
| EPI_ISL_1408290 | Viollier AG | Department of Biosystems Science and Engineering, ETH Zürich | Chaoran Chen et al |
| EPI_ISL_1407619 | Viollier AG | Department of Biosystems Science and Engineering, ETH Zürich | Chaoran Chen et al |
| EPI_ISL_1408278 | Viollier AG | Department of Biosystems Science and Engineering, ETH Zürich | Chaoran Chen et al |
| EPI_ISL_1659173 | Viollier AG | Department of Biosystems Science and Engineering, ETH Zürich | Chaoran Chen et al |
| EPI_ISL_648803 | Department of Laboratory Medicine, Tan Tock Seng Hospital | Department of Laboratory Medicine, Tan Tock Seng Hospital | Chen YYC et al |
| EPI_ISL_648805 | Department of Laboratory Medicine, Tan Tock Seng Hospital | Department of Laboratory Medicine, Tan Tock Seng Hospital | Chen YYC et al |
| EPI_ISL_648806 | Department of Laboratory Medicine, Tan Tock Seng Hospital | Department of Laboratory Medicine, Tan Tock Seng Hospital | Chen YYC et al |
| EPI_ISL_648818 | Department of Laboratory Medicine, Tan Tock Seng Hospital | Department of Laboratory Medicine, Tan Tock Seng Hospital | Chen YYC et al |
| EPI_ISL_648706 | Department of Laboratory Medicine, Tan Tock Seng Hospital | Department of Laboratory Medicine, Tan Tock Seng Hospital | Chen YYC et al |

| | | | |
|---|---|---|---|
| EPI_ISL_648715 | Department of Laboratory Medicine, Tan Tock Seng Hospital | Department of Laboratory Medicine, Tan Tock Seng Hospital | Chen YYC et al |
| EPI_ISL_648718 | Department of Laboratory Medicine, Tan Tock Seng Hospital | Department of Laboratory Medicine, Tan Tock Seng Hospital | Chen YYC et al |
| EPI_ISL_648736 | Department of Laboratory Medicine, Tan Tock Seng Hospital | Department of Laboratory Medicine, Tan Tock Seng Hospital | Chen YYC et al |
| EPI_ISL_507006 | Department of Laboratory Medicine, Tan Tock Seng Hospital | Department of Laboratory Medicine, Tan Tock Seng Hospital | Chen YYC et al |
| EPI_ISL_538436 | Department of Laboratory Medicine, Tan Tock Seng Hospital | Department of Laboratory Medicine, Tan Tock Seng Hospital | Chen YYC et al |
| EPI_ISL_538456 | Department of Laboratory Medicine, Tan Tock Seng Hospital | Department of Laboratory Medicine, Tan Tock Seng Hospital | Chen YYC et al |
| EPI_ISL_538458 | Department of Laboratory Medicine, Tan Tock Seng Hospital | Department of Laboratory Medicine, Tan Tock Seng Hospital | Chen YYC et al |
| EPI_ISL_426629 | TSGH-CP molecular lab | TSGH-CP molecular lab | Cherng-Lih Perng et al |
| EPI_ISL_437336 | TSGH-CP molecular lab, Division of Clinical Pathology, Department of Pathology | TSGH-CP molecular lab, Division of Clinical Pathology, Department of Pathology | Cherng-Lih Perng et al |
| EPI_ISL_447252 | TSGH-CP molecular lab | TSGH-CP molecular lab | Cherng-Lih Perng et al |
| EPI_ISL_447256 | TSGH-CP molecular lab | TSGH-CP molecular lab | Cherng-Lih Perng et al |
| EPI_ISL_447593 | TSGH-CP molecular lab | TSGH-CP molecular lab | Cherng-Lih Perng et al |
| EPI_ISL_803911 | BBMP Urban PHC | Department of Neurovirology, National Institute of Mental Health and Neurosciences (NIMHANS) | Chitra Pattabiraman et al |
| EPI_ISL_803903 | Shimoga Institute of Medical Sciences | Department of Neurovirology, National Institute of Mental Health and Neurosciences (NIMHANS) | Chitra Pattabiraman et al |
| EPI_ISL_995747 | Railway Hospital | Department of Neurovirology, National Institute of Mental Health and Neurosciences (NIMHANS) | Chitra Pattabiraman et al |
| EPI_ISL_995753 | RAILWAY HOSPITAL | Department of Neurovirology, National Institute of Mental Health and Neurosciences (NIMHANS) | Chitra Pattabiraman et al |
| EPI_ISL_484073 | Centre for Clinical Infection and Diagnostics Research and Genomics Innovation Unit, Guy's and St. Thomas' NHS Trust | COVID-19 Genomics UK (COG-UK) Consortium | Chloe Fisher et al |
| EPI_ISL_1360415 | Child Health Research Foundation | Child Health Research Foundation | CHRF Bangladesh Genomics Team et al |
| EPI_ISL_1360430 | Child Health Research Foundation | Child Health Research Foundation | CHRF Bangladesh Genomics Team et al |
| EPI_ISL_1360437 | Shimantik Pathology and Diagnostic Center | Child Health Research Foundation | CHRF Bangladesh Genomics Team et al |
| EPI_ISL_1360439 | Shimantik Pathology and Diagnostic Center | Child Health Research Foundation | CHRF Bangladesh Genomics Team et al |
| EPI_ISL_1360445 | Shimantik Pathology and Diagnostic Center | Child Health Research Foundation | CHRF Bangladesh Genomics Team et al |
| EPI_ISL_1360451 | Child Health Research Foundation | Child Health Research Foundation | CHRF Bangladesh Genomics Team et al |
| EPI_ISL_1469973 | Child Health Research Foundation | Child Health Research Foundation | CHRF Bangladesh Genomics Team et al |
| EPI_ISL_1469978 | Child Health Research Foundation | Child Health Research Foundation | CHRF Bangladesh Genomics Team et al |
| EPI_ISL_1469982 | Child Health Research Foundation | Child Health Research Foundation | CHRF Bangladesh Genomics Team et al |
| EPI_ISL_1634449 | Child Health Research Foundation | Child Health Research Foundation | CHRF Bangladesh Genomics Team et al |
| EPI_ISL_486462 | Viollier AG | Department of Biosystems Science and Engineering, ETH Zürich | Christian Beisel et al |

| | | | |
|---|---|---|---|
| EPI_ISL_1129407 | Viollier AG | Department of Biosystems Science and Engineering, ETH Zürich | Christian Beisel et al |
| EPI_ISL_468285 | Viollier AG | Department of Biosystems Science and Engineering, ETH Zürich | Christian Beisel et al |
| EPI_ISL_523870 | Viollier AG | Department of Biosystems Science and Engineering, ETH Zürich | Christian Beisel et al |
| EPI_ISL_1658258 | Viollier AG | Department of Biosystems Science and Engineering, ETH Zurich | Christian Beisel et al |
| EPI_ISL_574887 | Viollier AG | Department of Biosystems Science and Engineering, ETH Zürich | Christian Beisel et al |
| EPI_ISL_468290 | Viollier AG | Department of Biosystems Science and Engineering, ETH Zürich | Christian Beisel et al |
| EPI_ISL_476094 | Viollier AG | Department of Biosystems Science and Engineering, ETH Zürich | Christian Beisel et al |
| EPI_ISL_486521 | Viollier AG | Department of Biosystems Science and Engineering, ETH Zürich | Christian Beisel et al |
| EPI_ISL_486528 | Viollier AG | Department of Biosystems Science and Engineering, ETH Zürich | Christian Beisel et al |
| EPI_ISL_1406815 | Viollier AG | Department of Biosystems Science and Engineering, ETH Zürich | Christian Beisel et al |
| EPI_ISL_560458 | Viollier AG | Department of Biosystems Science and Engineering, ETH Zürich | Christian Beisel et al |
| EPI_ISL_1496225 | Viollier AG | Department of Biosystems Science and Engineering, ETH Zürich | Christian Beisel et al |
| EPI_ISL_451774 | Viollier AG | Department of Biosystems Science and Engineering, ETH Zürich | Christian Beisel et al |
| EPI_ISL_467001 | Viollier AG | Department of Biosystems Science and Engineering, ETH Zürich | Christian Beisel et al |
| EPI_ISL_1130121 | Viollier AG | Department of Biosystems Science and Engineering, ETH Zürich | Christian Beisel et al |
| EPI_ISL_468269 | Viollier AG | Department of Biosystems Science and Engineering, ETH Zürich | Christian Beisel et al |
| EPI_ISL_1130149 | Viollier AG | Department of Biosystems Science and Engineering, ETH Zürich | Christian Beisel et al |
| EPI_ISL_486506 | Viollier AG | Department of Biosystems Science and Engineering, ETH Zürich | Christian Beisel et al |
| EPI_ISL_468295 | Viollier AG | Department of Biosystems Science and Engineering, ETH Zürich | Christian Beisel et al |
| EPI_ISL_1130695 | Viollier AG | Department of Biosystems Science and Engineering, ETH Zürich | Christian Beisel et al |
| EPI_ISL_1130704 | Viollier AG | Department of Biosystems Science and Engineering, ETH Zürich | Christian Beisel et al |
| EPI_ISL_539418 | Viollier AG | Department of Biosystems Science and Engineering, ETH Zürich | Christian Beisel et al |
| EPI_ISL_693858 | Viollier AG | Department of Biosystems Science and Engineering, ETH Zürich | Christian Beisel et al |
| EPI_ISL_468281 | Viollier AG | Department of Biosystems Science and Engineering, ETH Zürich | Christian Beisel et al |
| EPI_ISL_523920 | Viollier AG | Department of Biosystems Science and Engineering, ETH Zürich | Christian Beisel et al |
| EPI_ISL_541509 | Viollier AG | Department of Biosystems Science and Engineering, ETH Zürich | Christian Beisel et al |
| EPI_ISL_451726 | Viollier AG | Department of Biosystems Science and Engineering, ETH Zürich | Christian Beisel et al |
| EPI_ISL_483659 | Viollier AG | Department of Biosystems Science and Engineering, ETH Zürich | Christian Beisel et al |
| EPI_ISL_523913 | Viollier AG | Department of Biosystems Science and Engineering, ETH Zürich | Christian Beisel et al |
| EPI_ISL_541512 | Viollier AG | Department of Biosystems Science and Engineering, ETH Zürich | Christian Beisel et al |

| | | | |
|---|---|---|---|
| EPI_ISL_1260530 | Viollier AG | Department of Biosystems Science and Engineering, ETH Zürich | Christian Beisel et al |
| EPI_ISL_1260508 | Viollier AG | Department of Biosystems Science and Engineering, ETH Zürich | Christian Beisel et al |
| EPI_ISL_1361446 | Viollier AG | Department of Biosystems Science and Engineering, ETH Zürich | Christian Beisel et al |
| EPI_ISL_508862 | Virology Unit, Institut Pasteur de Madagascar | Virology Unit, Institut Pasteur de Madagascar | Christian Ranaivoson et al |
| EPI_ISL_508863 | Virology Unit, Institut Pasteur de Madagascar | Virology Unit, Institut Pasteur de Madagascar | Christian Ranaivoson et al |
| EPI_ISL_1660239 | Virology Unit, Institut Pasteur de Madagascar | Virology Unit, Institut Pasteur de Madagascar | Christian Ranaivoson et al |
| EPI_ISL_1660240 | Virology Unit, Institut Pasteur de Madagascar | Virology Unit, Institut Pasteur de Madagascar | Christian Ranaivoson et al |
| EPI_ISL_1660263 | Virology Unit, Institut Pasteur de Madagascar | Virology Unit, Institut Pasteur de Madagascar | Christian Ranaivoson et al |
| EPI_ISL_1660272 | Virology Unit, Institut Pasteur de Madagascar | Virology Unit, Institut Pasteur de Madagascar | Christian Ranaivoson et al |
| EPI_ISL_1660273 | Virology Unit, Institut Pasteur de Madagascar | Virology Unit, Institut Pasteur de Madagascar | Christian Ranaivoson et al |
| EPI_ISL_1660274 | Virology Unit, Institut Pasteur de Madagascar | Virology Unit, Institut Pasteur de Madagascar | Christian Ranaivoson et al |
| EPI_ISL_1660279 | Virology Unit, Institut Pasteur de Madagascar | Virology Unit, Institut Pasteur de Madagascar | Christian Ranaivoson et al |
| EPI_ISL_1660286 | Virology Unit, Institut Pasteur de Madagascar | Virology Unit, Institut Pasteur de Madagascar | Christian Ranaivoson et al |
| EPI_ISL_1660288 | Virology Unit, Institut Pasteur de Madagascar | Virology Unit, Institut Pasteur de Madagascar | Christian Ranaivoson et al |
| EPI_ISL_1660289 | Virology Unit, Institut Pasteur de Madagascar | Virology Unit, Institut Pasteur de Madagascar | Christian Ranaivoson et al |
| EPI_ISL_1660290 | Virology Unit, Institut Pasteur de Madagascar | Virology Unit, Institut Pasteur de Madagascar | Christian Ranaivoson et al |
| EPI_ISL_1660291 | Virology Unit, Institut Pasteur de Madagascar | Virology Unit, Institut Pasteur de Madagascar | Christian Ranaivoson et al |
| EPI_ISL_1660225 | Virology Unit, Institut Pasteur de Madagascar | Virology Unit, Institut Pasteur de Madagascar | Christian Ranaivoson et al |
| EPI_ISL_1660226 | Virology Unit, Institut Pasteur de Madagascar | Virology Unit, Institut Pasteur de Madagascar | Christian Ranaivoson et al |
| EPI_ISL_1660227 | Virology Unit, Institut Pasteur de Madagascar | Virology Unit, Institut Pasteur de Madagascar | Christian Ranaivoson et al |
| EPI_ISL_1660234 | Virology Unit, Institut Pasteur de Madagascar | Virology Unit, Institut Pasteur de Madagascar | Christian Ranaivoson et al |
| EPI_ISL_677635 | Virology Unit, Institut Pasteur de Madagascar | Virology Unit, Institut Pasteur de Madagascar | Christian Ranaivoson et al |
| EPI_ISL_677636 | Virology Unit, Institut Pasteur de Madagascar | Virology Unit, Institut Pasteur de Madagascar | Christian Ranaivoson et al |
| EPI_ISL_677634 | Virology Unit, Institut Pasteur de Madagascar | Virology Unit, Institut Pasteur de Madagascar | Christian Ranaivoson et al |
| EPI_ISL_625456 | Virology Unit, Institut Pasteur de Madagascar | Virology Unit, Institut Pasteur de Madagascar | Christian Ranaivoson et al |
| EPI_ISL_1404498 | Hôpital Henri Mondor | Department of Virology, Henri Mondor University Hospital, Assistance Publique Hôpitaux de Paris, Université Paris-Est Créteil, INSERM U955 | Christophe Rodriguez et al |
| EPI_ISL_1404538 | Hôpital Henri Mondor | Department of Virology, Henri Mondor University Hospital, Assistance Publique Hôpitaux de Paris, Université Paris-Est Créteil, INSERM U955 | Christophe Rodriguez et al |
| EPI_ISL_451957 | Molecular Pathology Division, Department of Pathology, Hong Kong Sanatorium & Hospital | Molecular Pathology Division, Department of Pathology, Hong Kong Sanatorium & Hospital | Chun Hang AU et al |
| EPI_ISL_578701 | Molecular Pathology Division, Department of Pathology, Hong Kong Sanatorium & Hospital | Molecular Pathology Division, Department of Pathology, Hong Kong Sanatorium & Hospital | Chun Hang AU et al |

| | | | |
|---|---|---|---|
| EPI_ISL_578702 | Molecular Pathology Division, Department of Pathology, Hong Kong Sanatorium & Hospital | Molecular Pathology Division, Department of Pathology, Hong Kong Sanatorium & Hospital | Chun Hang AU et al |
| EPI_ISL_593698 | South Eastern Area Laboratory Services (SEALS) | NSW Health Pathology - Institute of Clinical Pathology and Medical Research; Westmead Hospital; University of Sydney | CIDM-PH et al. |
| EPI_ISL_593737 | South Eastern Area Laboratory Services (SEALS) | NSW Health Pathology - Institute of Clinical Pathology and Medical Research; Westmead Hospital; University of Sydney | CIDM-PH et al. |
| EPI_ISL_593739 | South Eastern Area Laboratory Services (SEALS) | NSW Health Pathology - Institute of Clinical Pathology and Medical Research; Westmead Hospital; University of Sydney | CIDM-PH et al. |
| EPI_ISL_629004 | South Eastern Area Laboratory Services (SEALS) | NSW Health Pathology - Institute of Clinical Pathology and Medical Research; Westmead Hospital; University of Sydney | CIDM-PH et al. |
| EPI_ISL_667800 | South Eastern Area Laboratory Services (SEALS) | NSW Health Pathology - Institute of Clinical Pathology and Medical Research; Westmead Hospital; University of Sydney | CIDM-PH et al. |
| EPI_ISL_767864 | Sydney South West Pathology Service (SSWPS) - Royal Prince Alfred Hospital - NSW Health Pathology | NSW Health Pathology - Institute of Clinical Pathology and Medical Research; Westmead Hospital; University of Sydney | CIDM-PH et al. |
| EPI_ISL_767865 | Sydney South West Pathology Service (SSWPS) - Royal Prince Alfred Hospital - NSW Health Pathology | NSW Health Pathology - Institute of Clinical Pathology and Medical Research; Westmead Hospital; University of Sydney | CIDM-PH et al. |
| EPI_ISL_1098652 | South Eastern Area Laboratory Services (SEALS) | NSW Health Pathology - Institute of Clinical Pathology and Medical Research; Westmead Hospital; University of Sydney | CIDM-PH et al. |
| EPI_ISL_451558 | Medlab Pathology | NSW Health Pathology - Institute of Clinical Pathology and Medical Research; Westmead Hospital; University of Sydney | CIDM-PH et al. |
| EPI_ISL_451594 | Childrens Hospital Westmead | NSW Health Pathology - Institute of Clinical Pathology and Medical Research; Westmead Hospital; University of Sydney | CIDM-PH et al. |
| EPI_ISL_478718 | Sydney South West Pathology Service (SSWPS) - Concord Repatriation General Hospital - NSW Health Pathology | NSW Health Pathology - Institute of Clinical Pathology and Medical Research; Westmead Hospital; University of Sydney | CIDM-PH et al. |
| EPI_ISL_513316 | South Eastern Area Laboratory Services (SEALS) | NSW Health Pathology - Institute of Clinical Pathology and Medical Research; Westmead Hospital; University of Sydney | CIDM-PH et al. |
| EPI_ISL_542246 | ASST GOM Niguarda | Dep. Of Oncology and Hemato-Oncology University of Milan | Claudia Alteri et al |
| EPI_ISL_1424051 | LESP Ciudad de Mexico | Instituto de Diagnostico y Referencia Epidemiologicos (INDRE) | Claudia Wong-Arambula et al |
| EPI_ISL_1626820 | LESP Guanajuato | Instituto de Diagnostico y Referencia Epidemiologicos (INDRE) | Claudia Wong-Arambula et al |
| EPI_ISL_1093146 | Laboratorio de Diagnostico de Enfermedades Emergentes y Reemergentes | Instituto de Diagnostico y Referencia Epidemiologicos (INDRE) | Claudia Wong-Arambula et al |
| EPI_ISL_1093147 | Laboratorio de Diagnostico de Enfermedades Emergentes y Reemergentes | Instituto de Diagnostico y Referencia Epidemiologicos (INDRE) | Claudia Wong-Arambula et al |
| EPI_ISL_1516762 | LESP Jalisco/UdeG | Instituto de Diagnostico y Referencia Epidemiologicos (INDRE) | Claudia Wong-Arambula et al |
| EPI_ISL_913968 | Instituto de Diagnostico y Referencia Epidemiologicos INDRE_RNLSP | Instituto de Diagnostico y Referencia Epidemiologicos (INDRE) | Claudia Wong-Arambula et al |
| EPI_ISL_1168466 | Instituto de Diagnostico y Referencia Epidemiologicos INDRE_RNLSP | Instituto de Diagnostico y Referencia Epidemiologicos (INDRE) | Claudia Wong-Arambula et al |
| EPI_ISL_1324767 | LESP Quintana Roo | Instituto de Diagnostico y Referencia Epidemiologicos (INDRE) | Claudia Wong-Arambula et al |
| EPI_ISL_1626821 | LESP Quintana Roo | Instituto de Diagnostico y Referencia Epidemiologicos (INDRE) | Claudia Wong-Arambula et al |
| EPI_ISL_913952 | Instituto de Diagnostico y Referencia Epidemiologicos INDRE_RNLSP | Instituto de Diagnostico y Referencia Epidemiologicos (INDRE) | Claudia Wong-Arambula et al |
| EPI_ISL_913966 | Instituto de Diagnostico y Referencia Epidemiologicos INDRE_RNLSP | Instituto de Diagnostico y Referencia Epidemiologicos (INDRE) | Claudia Wong-Arambula et al |

| | | | |
|---|---|---|---|
| EPI_ISL_913944 | Instituto de Diagnostico y Referencia Epidemiologicos INDRE_RNLSP | Instituto de Diagnostico y Referencia Epidemiologicos (INDRE) | Claudia Wong-Arambula et al |
| EPI_ISL_940617 | LACEN-PI DR. Costa Alvarenga | Instituto Adolfo Lutz, Interdiciplinary Procedures Center, Strategic Laboratory | Claudio Tavares Sacchi et al |
| EPI_ISL_882663 | LACEN do Distrito Federal | Instituto Adolfo Lutz, Interdiciplinary Procedures Center, Strategic Laboratory | Claudio Tavares Sacchi et al |
| EPI_ISL_882664 | LACEN do Distrito Federal | Instituto Adolfo Lutz, Interdiciplinary Procedures Center, Strategic Laboratory | Claudio Tavares Sacchi et al |
| EPI_ISL_882657 | LACEN do Estado do Piaui, Dr. Costa Alvarenga | Instituto Adolfo Lutz, Interdiciplinary Procedures Center, Strategic Laboratory | Claudio Tavares Sacchi et al |
| EPI_ISL_1293051 | LACEN do Distrito Federal | Instituto Adolfo Lutz, Interdiciplinary Procedures Center, Strategic Laboratory | Claudio Tavares Sacchi et al |
| EPI_ISL_1196285 | LACEN do Distrito Federal | Instituto Adolfo Lutz, Interdiciplinary Procedures Center, Strategic Laboratory | Claudio Tavares Sacchi et al |
| EPI_ISL_1196286 | LACEN do Distrito Federal | Instituto Adolfo Lutz, Interdiciplinary Procedures Center, Strategic Laboratory | Claudio Tavares Sacchi et al |
| EPI_ISL_1196287 | LACEN do Distrito Federal | Instituto Adolfo Lutz, Interdiciplinary Procedures Center, Strategic Laboratory | Claudio Tavares Sacchi et al |
| EPI_ISL_1196288 | LACEN do Distrito Federal | Instituto Adolfo Lutz, Interdiciplinary Procedures Center, Strategic Laboratory | Claudio Tavares Sacchi et al |
| EPI_ISL_1196289 | LACEN do Distrito Federal | Instituto Adolfo Lutz, Interdiciplinary Procedures Center, Strategic Laboratory | Claudio Tavares Sacchi et al |
| EPI_ISL_1196290 | LACEN do Distrito Federal | Instituto Adolfo Lutz, Interdiciplinary Procedures Center, Strategic Laboratory | Claudio Tavares Sacchi et al |
| EPI_ISL_1196291 | LACEN do Distrito Federal | Instituto Adolfo Lutz, Interdiciplinary Procedures Center, Strategic Laboratory | Claudio Tavares Sacchi et al |
| EPI_ISL_1196292 | LACEN do Distrito Federal | Instituto Adolfo Lutz, Interdiciplinary Procedures Center, Strategic Laboratory | Claudio Tavares Sacchi et al |
| EPI_ISL_1196293 | LACEN do Distrito Federal | Instituto Adolfo Lutz, Interdiciplinary Procedures Center, Strategic Laboratory | Claudio Tavares Sacchi et al |
| EPI_ISL_1196294 | LACEN do Distrito Federal | Instituto Adolfo Lutz, Interdiciplinary Procedures Center, Strategic Laboratory | Claudio Tavares Sacchi et al |
| EPI_ISL_1303506 | LACEN do Distrito Federal | Instituto Adolfo Lutz, Interdiciplinary Procedures Center, Strategic Laboratory | Claudio Tavares Sacchi et al |
| EPI_ISL_1303507 | LACEN do Distrito Federal | Instituto Adolfo Lutz, Interdiciplinary Procedures Center, Strategic Laboratory | Claudio Tavares Sacchi et al |
| EPI_ISL_1303508 | LACEN do Distrito Federal | Instituto Adolfo Lutz, Interdiciplinary Procedures Center, Strategic Laboratory | Claudio Tavares Sacchi et al |
| EPI_ISL_882661 | Hospital de Santa Barbara de Goias | Instituto Adolfo Lutz, Interdiciplinary Procedures Center, Strategic Laboratory | Claudio Tavares Sacchi et al |
| EPI_ISL_882662 | Hospital de Santa Barbara de Goias | Instituto Adolfo Lutz, Interdiciplinary Procedures Center, Strategic Laboratory | Claudio Tavares Sacchi et al |
| EPI_ISL_940612 | Hospital de Campanha para Enfrentamento do Coronavirus - Goiania | Instituto Adolfo Lutz, Interdiciplinary Procedures Center, Strategic Laboratory | Claudio Tavares Sacchi et al |

| | | | |
|---|---|---|---|
| EPI_ISL_943988 | LACEN do Estado de Goias | Instituto Adolfo Lutz, Interdiciplinary Procedures Center, Strategic Laboratory | Claudio Tavares Sacchi et al |
| EPI_ISL_943989 | LACEN do Estado de Goias | Instituto Adolfo Lutz, Interdiciplinary Procedures Center, Strategic Laboratory | Claudio Tavares Sacchi et al |
| EPI_ISL_1039691 | LACEN do Estado de Goias | Instituto Adolfo Lutz, Interdiciplinary Procedures Center, Strategic Laboratory | Claudio Tavares Sacchi et al |
| EPI_ISL_1039692 | LACEN do Estado de Goias | Instituto Adolfo Lutz, Interdiciplinary Procedures Center, Strategic Laboratory | Claudio Tavares Sacchi et al |
| EPI_ISL_1039693 | LACEN do Estado de Goias | Instituto Adolfo Lutz, Interdiciplinary Procedures Center, Strategic Laboratory | Claudio Tavares Sacchi et al |
| EPI_ISL_1039694 | LACEN do Estado de Goias | Instituto Adolfo Lutz, Interdiciplinary Procedures Center, Strategic Laboratory | Claudio Tavares Sacchi et al |
| EPI_ISL_1039695 | LACEN do Estado de Goias | Instituto Adolfo Lutz, Interdiciplinary Procedures Center, Strategic Laboratory | Claudio Tavares Sacchi et al |
| EPI_ISL_1041509 | LACEN do Estado de Goias | Instituto Adolfo Lutz, Interdiciplinary Procedures Center, Strategic Laboratory | Claudio Tavares Sacchi et al |
| EPI_ISL_1303510 | LACEN do Estado de Goias | Instituto Adolfo Lutz, Interdiciplinary Procedures Center, Strategic Laboratory | Claudio Tavares Sacchi et al |
| EPI_ISL_1303511 | LACEN do Estado de Goias | Instituto Adolfo Lutz, Interdiciplinary Procedures Center, Strategic Laboratory | Claudio Tavares Sacchi et al |
| EPI_ISL_1303512 | LACEN do Estado de Goias | Instituto Adolfo Lutz, Interdiciplinary Procedures Center, Strategic Laboratory | Claudio Tavares Sacchi et al |
| EPI_ISL_1303513 | LACEN do Estado de Goias | Instituto Adolfo Lutz, Interdiciplinary Procedures Center, Strategic Laboratory | Claudio Tavares Sacchi et al |
| EPI_ISL_1303514 | LACEN do Estado de Goias | Instituto Adolfo Lutz, Interdiciplinary Procedures Center, Strategic Laboratory | Claudio Tavares Sacchi et al |
| EPI_ISL_1303515 | LACEN do Estado de Goias | Instituto Adolfo Lutz, Interdiciplinary Procedures Center, Strategic Laboratory | Claudio Tavares Sacchi et al |
| EPI_ISL_1303516 | LACEN do Estado de Goias | Instituto Adolfo Lutz, Interdiciplinary Procedures Center, Strategic Laboratory | Claudio Tavares Sacchi et al |
| EPI_ISL_1303517 | LACEN do Estado de Goias | Instituto Adolfo Lutz, Interdiciplinary Procedures Center, Strategic Laboratory | Claudio Tavares Sacchi et al |
| EPI_ISL_1201896 | Diagnosticos da America - DASA | Instituto Adolfo Lutz, Interdiciplinary Procedures Center, Strategic Laboratory | Claudio Tavares Sacchi et al |
| EPI_ISL_1468431 | LACEN do Estado de Goias | Instituto Adolfo Lutz, Interdiciplinary Procedures Center, Strategic Laboratory | Claudio Tavares Sacchi et al |
| EPI_ISL_1468413 | LACEN do Estado de Goias | Instituto Adolfo Lutz, Interdiciplinary Procedures Center, Strategic Laboratory | Claudio Tavares Sacchi et al |
| EPI_ISL_1468414 | LACEN do Estado de Goias | Instituto Adolfo Lutz, Interdiciplinary Procedures Center, Strategic Laboratory | Claudio Tavares Sacchi et al |
| EPI_ISL_1468415 | LACEN do Estado de Goias | Instituto Adolfo Lutz, Interdiciplinary Procedures Center, Strategic Laboratory | Claudio Tavares Sacchi et al |
| EPI_ISL_1628363 | LACEN do Estado de Goias | Instituto Adolfo Lutz, Interdiciplinary Procedures Center, Strategic Laboratory | Claudio Tavares Sacchi et al |

| | | | |
|---|---|---|---|
| EPI_ISL_1628364 | LACEN do Estado de Goias | Instituto Adolfo Lutz, Interdiciplinary Procedures Center, Strategic Laboratory | Claudio Tavares Sacchi et al |
| EPI_ISL_1628365 | LACEN do Estado de Goias | Instituto Adolfo Lutz, Interdiciplinary Procedures Center, Strategic Laboratory | Claudio Tavares Sacchi et al |
| EPI_ISL_1493577 | LACEN do Estado de Goias | Instituto Adolfo Lutz, Interdiciplinary Procedures Center, Strategic Laboratory | Claudio Tavares Sacchi et al |
| EPI_ISL_1493576 | LACEN do Estado de Goias | Instituto Adolfo Lutz, Interdiciplinary Procedures Center, Strategic Laboratory | Claudio Tavares Sacchi et al |
| EPI_ISL_1493575 | LACEN do Estado de Goias | Instituto Adolfo Lutz, Interdiciplinary Procedures Center, Strategic Laboratory | Claudio Tavares Sacchi et al |
| EPI_ISL_1493574 | LACEN do Estado de Goias | Instituto Adolfo Lutz, Interdiciplinary Procedures Center, Strategic Laboratory | Claudio Tavares Sacchi et al |
| EPI_ISL_1493573 | LACEN do Estado de Goias | Instituto Adolfo Lutz, Interdiciplinary Procedures Center, Strategic Laboratory | Claudio Tavares Sacchi et al |
| EPI_ISL_1040825 | LACEN do Mato Grosso do Sul | Instituto Adolfo Lutz, Interdiciplinary Procedures Center, Strategic Laboratory | Claudio Tavares Sacchi et al |
| EPI_ISL_1040826 | LACEN do Mato Grosso do Sul | Instituto Adolfo Lutz, Interdiciplinary Procedures Center, Strategic Laboratory | Claudio Tavares Sacchi et al |
| EPI_ISL_1171621 | LACEN do Mato Grosso do Sul | Instituto Adolfo Lutz, Interdiciplinary Procedures Center, Strategic Laboratory | Claudio Tavares Sacchi et al |
| EPI_ISL_1040832 | LACEN do Mato Grosso do Sul | Instituto Adolfo Lutz, Interdiciplinary Procedures Center, Strategic Laboratory | Claudio Tavares Sacchi et al |
| EPI_ISL_1040830 | LACEN do Mato Grosso do Sul | Instituto Adolfo Lutz, Interdiciplinary Procedures Center, Strategic Laboratory | Claudio Tavares Sacchi et al |
| EPI_ISL_1040831 | LACEN do Mato Grosso do Sul | Instituto Adolfo Lutz, Interdiciplinary Procedures Center, Strategic Laboratory | Claudio Tavares Sacchi et al |
| EPI_ISL_1040827 | LACEN do Mato Grosso do Sul | Instituto Adolfo Lutz, Interdiciplinary Procedures Center, Strategic Laboratory | Claudio Tavares Sacchi et al |
| EPI_ISL_1040828 | LACEN do Mato Grosso do Sul | Instituto Adolfo Lutz, Interdiciplinary Procedures Center, Strategic Laboratory | Claudio Tavares Sacchi et al |
| EPI_ISL_1040829 | LACEN do Mato Grosso do Sul | Instituto Adolfo Lutz, Interdiciplinary Procedures Center, Strategic Laboratory | Claudio Tavares Sacchi et al |
| EPI_ISL_1040824 | LACEN do Mato Grosso do Sul | Instituto Adolfo Lutz, Interdiciplinary Procedures Center, Strategic Laboratory | Claudio Tavares Sacchi et al |
| EPI_ISL_1040833 | LACEN do Mato Grosso do Sul | Instituto Adolfo Lutz, Interdiciplinary Procedures Center, Strategic Laboratory | Claudio Tavares Sacchi et al |
| EPI_ISL_1040834 | LACEN do Mato Grosso do Sul | Instituto Adolfo Lutz, Interdiciplinary Procedures Center, Strategic Laboratory | Claudio Tavares Sacchi et al |
| EPI_ISL_1040835 | LACEN do Mato Grosso do Sul | Instituto Adolfo Lutz, Interdiciplinary Procedures Center, Strategic Laboratory | Claudio Tavares Sacchi et al |
| EPI_ISL_1040836 | LACEN do Mato Grosso do Sul | Instituto Adolfo Lutz, Interdiciplinary Procedures Center, Strategic Laboratory | Claudio Tavares Sacchi et al |
| EPI_ISL_1040837 | LACEN do Mato Grosso do Sul | Instituto Adolfo Lutz, Interdiciplinary Procedures Center, Strategic Laboratory | Claudio Tavares Sacchi et al |

| | | | |
|---|---|---|---|
| EPI_ISL_1040838 | LACEN do Mato Grosso do Sul | Instituto Adolfo Lutz, Interdiciplinary Procedures Center, Strategic Laboratory | Claudio Tavares Sacchi et al |
| EPI_ISL_1040839 | LACEN do Mato Grosso do Sul | Instituto Adolfo Lutz, Interdiciplinary Procedures Center, Strategic Laboratory | Claudio Tavares Sacchi et al |
| EPI_ISL_1040840 | LACEN do Mato Grosso do Sul | Instituto Adolfo Lutz, Interdiciplinary Procedures Center, Strategic Laboratory | Claudio Tavares Sacchi et al |
| EPI_ISL_1040842 | LACEN do Mato Grosso do Sul | Instituto Adolfo Lutz, Interdiciplinary Procedures Center, Strategic Laboratory | Claudio Tavares Sacchi et al |
| EPI_ISL_1040841 | LACEN do Mato Grosso do Sul | Instituto Adolfo Lutz, Interdiciplinary Procedures Center, Strategic Laboratory | Claudio Tavares Sacchi et al |
| EPI_ISL_1040843 | LACEN do Mato Grosso do Sul | Instituto Adolfo Lutz, Interdiciplinary Procedures Center, Strategic Laboratory | Claudio Tavares Sacchi et al |
| EPI_ISL_1121327 | LACEN do Mato Grosso do Sul | Instituto Adolfo Lutz, Interdiciplinary Procedures Center, Strategic Laboratory | Claudio Tavares Sacchi et al |
| EPI_ISL_1040847 | LACEN do Mato Grosso do Sul | Instituto Adolfo Lutz, Interdiciplinary Procedures Center, Strategic Laboratory | Claudio Tavares Sacchi et al |
| EPI_ISL_1040848 | LACEN do Mato Grosso do Sul | Instituto Adolfo Lutz, Interdiciplinary Procedures Center, Strategic Laboratory | Claudio Tavares Sacchi et al |
| EPI_ISL_1040846 | LACEN do Mato Grosso do Sul | Instituto Adolfo Lutz, Interdiciplinary Procedures Center, Strategic Laboratory | Claudio Tavares Sacchi et al |
| EPI_ISL_1040844 | LACEN do Mato Grosso do Sul | Instituto Adolfo Lutz, Interdiciplinary Procedures Center, Strategic Laboratory | Claudio Tavares Sacchi et al |
| EPI_ISL_1040845 | LACEN do Mato Grosso do Sul | Instituto Adolfo Lutz, Interdiciplinary Procedures Center, Strategic Laboratory | Claudio Tavares Sacchi et al |
| EPI_ISL_1040850 | LACEN do Mato Grosso do Sul | Instituto Adolfo Lutz, Interdiciplinary Procedures Center, Strategic Laboratory | Claudio Tavares Sacchi et al |
| EPI_ISL_1040851 | LACEN do Mato Grosso do Sul | Instituto Adolfo Lutz, Interdiciplinary Procedures Center, Strategic Laboratory | Claudio Tavares Sacchi et al |
| EPI_ISL_1040849 | LACEN do Mato Grosso do Sul | Instituto Adolfo Lutz, Interdiciplinary Procedures Center, Strategic Laboratory | Claudio Tavares Sacchi et al |
| EPI_ISL_1121328 | LACEN do Mato Grosso do Sul | Instituto Adolfo Lutz, Interdiciplinary Procedures Center, Strategic Laboratory | Claudio Tavares Sacchi et al |
| EPI_ISL_1121329 | LACEN do Mato Grosso do Sul | Instituto Adolfo Lutz, Interdiciplinary Procedures Center, Strategic Laboratory | Claudio Tavares Sacchi et al |
| EPI_ISL_1121330 | LACEN do Mato Grosso do Sul | Instituto Adolfo Lutz, Interdiciplinary Procedures Center, Strategic Laboratory | Claudio Tavares Sacchi et al |
| EPI_ISL_1196282 | LACEN do Mato Grosso do Sul | Instituto Adolfo Lutz, Interdiciplinary Procedures Center, Strategic Laboratory | Claudio Tavares Sacchi et al |
| EPI_ISL_1196283 | LACEN do Mato Grosso do Sul | Instituto Adolfo Lutz, Interdiciplinary Procedures Center, Strategic Laboratory | Claudio Tavares Sacchi et al |
| EPI_ISL_1196284 | LACEN do Mato Grosso do Sul | Instituto Adolfo Lutz, Interdiciplinary Procedures Center, Strategic Laboratory | Claudio Tavares Sacchi et al |
| EPI_ISL_1139052 | LACEN do Mato Grosso do Sul | Instituto Adolfo Lutz, Interdiciplinary Procedures Center, Strategic Laboratory | Claudio Tavares Sacchi et al |

| | | | |
|---|---|---|---|
| EPI_ISL_1139053 | LACEN do Mato Grosso do Sul | Instituto Adolfo Lutz, Interdiciplinary Procedures Center, Strategic Laboratory | Claudio Tavares Sacchi et al |
| EPI_ISL_1139054 | LACEN do Mato Grosso do Sul | Instituto Adolfo Lutz, Interdiciplinary Procedures Center, Strategic Laboratory | Claudio Tavares Sacchi et al |
| EPI_ISL_1139055 | LACEN do Mato Grosso do Sul | Instituto Adolfo Lutz, Interdiciplinary Procedures Center, Strategic Laboratory | Claudio Tavares Sacchi et al |
| EPI_ISL_1139056 | LACEN do Mato Grosso do Sul | Instituto Adolfo Lutz, Interdiciplinary Procedures Center, Strategic Laboratory | Claudio Tavares Sacchi et al |
| EPI_ISL_1139057 | LACEN do Mato Grosso do Sul | Instituto Adolfo Lutz, Interdiciplinary Procedures Center, Strategic Laboratory | Claudio Tavares Sacchi et al |
| EPI_ISL_1139058 | LACEN do Mato Grosso do Sul | Instituto Adolfo Lutz, Interdiciplinary Procedures Center, Strategic Laboratory | Claudio Tavares Sacchi et al |
| EPI_ISL_1139059 | LACEN do Mato Grosso do Sul | Instituto Adolfo Lutz, Interdiciplinary Procedures Center, Strategic Laboratory | Claudio Tavares Sacchi et al |
| EPI_ISL_1139060 | LACEN do Mato Grosso do Sul | Instituto Adolfo Lutz, Interdiciplinary Procedures Center, Strategic Laboratory | Claudio Tavares Sacchi et al |
| EPI_ISL_1139061 | LACEN do Mato Grosso do Sul | Instituto Adolfo Lutz, Interdiciplinary Procedures Center, Strategic Laboratory | Claudio Tavares Sacchi et al |
| EPI_ISL_1139062 | LACEN do Mato Grosso do Sul | Instituto Adolfo Lutz, Interdiciplinary Procedures Center, Strategic Laboratory | Claudio Tavares Sacchi et al |
| EPI_ISL_1139063 | LACEN do Mato Grosso do Sul | Instituto Adolfo Lutz, Interdiciplinary Procedures Center, Strategic Laboratory | Claudio Tavares Sacchi et al |
| EPI_ISL_1139064 | LACEN do Mato Grosso do Sul | Instituto Adolfo Lutz, Interdiciplinary Procedures Center, Strategic Laboratory | Claudio Tavares Sacchi et al |
| EPI_ISL_1139065 | LACEN do Mato Grosso do Sul | Instituto Adolfo Lutz, Interdiciplinary Procedures Center, Strategic Laboratory | Claudio Tavares Sacchi et al |
| EPI_ISL_1139066 | LACEN do Mato Grosso do Sul | Instituto Adolfo Lutz, Interdiciplinary Procedures Center, Strategic Laboratory | Claudio Tavares Sacchi et al |
| EPI_ISL_1139067 | LACEN do Mato Grosso do Sul | Instituto Adolfo Lutz, Interdiciplinary Procedures Center, Strategic Laboratory | Claudio Tavares Sacchi et al |
| EPI_ISL_1139068 | LACEN do Mato Grosso do Sul | Instituto Adolfo Lutz, Interdiciplinary Procedures Center, Strategic Laboratory | Claudio Tavares Sacchi et al |
| EPI_ISL_1201883 | LACEN do Mato Grosso do Sul | Instituto Adolfo Lutz, Interdiciplinary Procedures Center, Strategic Laboratory | Claudio Tavares Sacchi et al |
| EPI_ISL_1358307 | LACEN do Mato Grosso do Sul | Instituto Adolfo Lutz, Interdiciplinary Procedures Center, Strategic Laboratory | Claudio Tavares Sacchi et al |
| EPI_ISL_1358305 | LACEN do Mato Grosso do Sul | Instituto Adolfo Lutz, Interdiciplinary Procedures Center, Strategic Laboratory | Claudio Tavares Sacchi et al |
| EPI_ISL_1358309 | LACEN do Mato Grosso do Sul | Instituto Adolfo Lutz, Interdiciplinary Procedures Center, Strategic Laboratory | Claudio Tavares Sacchi et al |
| EPI_ISL_1358304 | LACEN do Mato Grosso do Sul | Instituto Adolfo Lutz, Interdiciplinary Procedures Center, Strategic Laboratory | Claudio Tavares Sacchi et al |
| EPI_ISL_1358308 | LACEN do Mato Grosso do Sul | Instituto Adolfo Lutz, Interdiciplinary Procedures Center, Strategic Laboratory | Claudio Tavares Sacchi et al |

| | | | |
|---|---|---|---|
| EPI_ISL_1358306 | LACEN do Mato Grosso do Sul | Instituto Adolfo Lutz, Interdiciplinary Procedures Center, Strategic Laboratory | Claudio Tavares Sacchi et al |
| EPI_ISL_1358316 | LACEN do Mato Grosso do Sul | Instituto Adolfo Lutz, Interdiciplinary Procedures Center, Strategic Laboratory | Claudio Tavares Sacchi et al |
| EPI_ISL_1358317 | LACEN do Mato Grosso do Sul | Instituto Adolfo Lutz, Interdiciplinary Procedures Center, Strategic Laboratory | Claudio Tavares Sacchi et al |
| EPI_ISL_1358311 | LACEN do Mato Grosso do Sul | Instituto Adolfo Lutz, Interdiciplinary Procedures Center, Strategic Laboratory | Claudio Tavares Sacchi et al |
| EPI_ISL_1358312 | LACEN do Mato Grosso do Sul | Instituto Adolfo Lutz, Interdiciplinary Procedures Center, Strategic Laboratory | Claudio Tavares Sacchi et al |
| EPI_ISL_1358313 | LACEN do Mato Grosso do Sul | Instituto Adolfo Lutz, Interdiciplinary Procedures Center, Strategic Laboratory | Claudio Tavares Sacchi et al |
| EPI_ISL_1358310 | LACEN do Mato Grosso do Sul | Instituto Adolfo Lutz, Interdiciplinary Procedures Center, Strategic Laboratory | Claudio Tavares Sacchi et al |
| EPI_ISL_1358314 | LACEN do Mato Grosso do Sul | Instituto Adolfo Lutz, Interdiciplinary Procedures Center, Strategic Laboratory | Claudio Tavares Sacchi et al |
| EPI_ISL_1358315 | LACEN do Mato Grosso do Sul | Instituto Adolfo Lutz, Interdiciplinary Procedures Center, Strategic Laboratory | Claudio Tavares Sacchi et al |
| EPI_ISL_1468432 | LACEN do Mato Grosso do Sul | Instituto Adolfo Lutz, Interdiciplinary Procedures Center, Strategic Laboratory | Claudio Tavares Sacchi et al |
| EPI_ISL_1468433 | LACEN do Mato Grosso do Sul | Instituto Adolfo Lutz, Interdiciplinary Procedures Center, Strategic Laboratory | Claudio Tavares Sacchi et al |
| EPI_ISL_1468434 | LACEN do Mato Grosso do Sul | Instituto Adolfo Lutz, Interdiciplinary Procedures Center, Strategic Laboratory | Claudio Tavares Sacchi et al |
| EPI_ISL_1468435 | LACEN do Mato Grosso do Sul | Instituto Adolfo Lutz, Interdiciplinary Procedures Center, Strategic Laboratory | Claudio Tavares Sacchi et al |
| EPI_ISL_1468436 | LACEN do Mato Grosso do Sul | Instituto Adolfo Lutz, Interdiciplinary Procedures Center, Strategic Laboratory | Claudio Tavares Sacchi et al |
| EPI_ISL_1468437 | LACEN do Mato Grosso do Sul | Instituto Adolfo Lutz, Interdiciplinary Procedures Center, Strategic Laboratory | Claudio Tavares Sacchi et al |
| EPI_ISL_1468438 | LACEN do Mato Grosso do Sul | Instituto Adolfo Lutz, Interdiciplinary Procedures Center, Strategic Laboratory | Claudio Tavares Sacchi et al |
| EPI_ISL_1468439 | LACEN do Mato Grosso do Sul | Instituto Adolfo Lutz, Interdiciplinary Procedures Center, Strategic Laboratory | Claudio Tavares Sacchi et al |
| EPI_ISL_1468440 | LACEN do Mato Grosso do Sul | Instituto Adolfo Lutz, Interdiciplinary Procedures Center, Strategic Laboratory | Claudio Tavares Sacchi et al |
| EPI_ISL_1468441 | LACEN do Mato Grosso do Sul | Instituto Adolfo Lutz, Interdiciplinary Procedures Center, Strategic Laboratory | Claudio Tavares Sacchi et al |
| EPI_ISL_1381066 | LACEN do Mato Grosso do Sul | Instituto Adolfo Lutz, Interdiciplinary Procedures Center, Strategic Laboratory | Claudio Tavares Sacchi et al |
| EPI_ISL_1381067 | LACEN do Mato Grosso do Sul | Instituto Adolfo Lutz, Interdiciplinary Procedures Center, Strategic Laboratory | Claudio Tavares Sacchi et al |
| EPI_ISL_940613 | LACEN-PI DR. Costa Alvarenga | Instituto Adolfo Lutz, Interdiciplinary Procedures Center, Strategic Laboratory | Claudio Tavares Sacchi et al |

| | | | |
|---|---|---|---|
| EPI_ISL_1293052 | LACEN de Rondonia | Instituto Adolfo Lutz, Interdiciplinary Procedures Center, Strategic Laboratory | Claudio Tavares Sacchi et al |
| EPI_ISL_1293053 | LACEN de Rondonia | Instituto Adolfo Lutz, Interdiciplinary Procedures Center, Strategic Laboratory | Claudio Tavares Sacchi et al |
| EPI_ISL_1293054 | LACEN de Rondonia | Instituto Adolfo Lutz, Interdiciplinary Procedures Center, Strategic Laboratory | Claudio Tavares Sacchi et al |
| EPI_ISL_1303499 | LACEN de Rondonia | Instituto Adolfo Lutz, Interdiciplinary Procedures Center, Strategic Laboratory | Claudio Tavares Sacchi et al |
| EPI_ISL_1303500 | LACEN de Rondonia | Instituto Adolfo Lutz, Interdiciplinary Procedures Center, Strategic Laboratory | Claudio Tavares Sacchi et al |
| EPI_ISL_1303501 | LACEN de Rondonia | Instituto Adolfo Lutz, Interdiciplinary Procedures Center, Strategic Laboratory | Claudio Tavares Sacchi et al |
| EPI_ISL_1303502 | LACEN de Rondonia | Instituto Adolfo Lutz, Interdiciplinary Procedures Center, Strategic Laboratory | Claudio Tavares Sacchi et al |
| EPI_ISL_1303504 | LACEN de Rondonia | Instituto Adolfo Lutz, Interdiciplinary Procedures Center, Strategic Laboratory | Claudio Tavares Sacchi et al |
| EPI_ISL_1303505 | LACEN de Rondonia | Instituto Adolfo Lutz, Interdiciplinary Procedures Center, Strategic Laboratory | Claudio Tavares Sacchi et al |
| EPI_ISL_1520107 | LACEN do Estado de Rondonia | Instituto Adolfo Lutz, Interdiciplinary Procedures Center, Strategic Laboratory | Claudio Tavares Sacchi et al |
| EPI_ISL_1520108 | LACEN do Estado de Rondonia | Instituto Adolfo Lutz, Interdiciplinary Procedures Center, Strategic Laboratory | Claudio Tavares Sacchi et al |
| EPI_ISL_1520109 | LACEN do Estado de Rondonia | Instituto Adolfo Lutz, Interdiciplinary Procedures Center, Strategic Laboratory | Claudio Tavares Sacchi et al |
| EPI_ISL_1493600 | LACEN do Estado de Rondonia | Instituto Adolfo Lutz, Interdiciplinary Procedures Center, Strategic Laboratory | Claudio Tavares Sacchi et al |
| EPI_ISL_1493599 | LACEN do Estado de Rondonia | Instituto Adolfo Lutz, Interdiciplinary Procedures Center, Strategic Laboratory | Claudio Tavares Sacchi et al |
| EPI_ISL_1493597 | LACEN do Estado de Rondonia | Instituto Adolfo Lutz, Interdiciplinary Procedures Center, Strategic Laboratory | Claudio Tavares Sacchi et al |
| EPI_ISL_1493598 | LACEN do Estado de Rondonia | Instituto Adolfo Lutz, Interdiciplinary Procedures Center, Strategic Laboratory | Claudio Tavares Sacchi et al |
| EPI_ISL_1493595 | LACEN do Estado de Rondonia | Instituto Adolfo Lutz, Interdiciplinary Procedures Center, Strategic Laboratory | Claudio Tavares Sacchi et al |
| EPI_ISL_1493596 | LACEN do Estado de Rondonia | Instituto Adolfo Lutz, Interdiciplinary Procedures Center, Strategic Laboratory | Claudio Tavares Sacchi et al |
| EPI_ISL_1494924 | LACEN do Estado de Rondonia | Instituto Adolfo Lutz, Interdiciplinary Procedures Center, Strategic Laboratory | Claudio Tavares Sacchi et al |
| EPI_ISL_1493584 | LACEN do Estado de Rondonia | Instituto Adolfo Lutz, Interdiciplinary Procedures Center, Strategic Laboratory | Claudio Tavares Sacchi et al |
| EPI_ISL_1493579 | LACEN do Estado de Rondonia | Instituto Adolfo Lutz, Interdiciplinary Procedures Center, Strategic Laboratory | Claudio Tavares Sacchi et al |
| EPI_ISL_1493578 | LACEN do Estado de Rondonia | Instituto Adolfo Lutz, Interdiciplinary Procedures Center, Strategic Laboratory | Claudio Tavares Sacchi et al |

| | | | |
|---|---|---|---|
| EPI_ISL_940630 | Hospital Geral de Sao Paulo | Instituto Adolfo Lutz, Interdiciplinary Procedures Center, Strategic Laboratory | Claudio Tavares Sacchi et al |
| EPI_ISL_943967 | Hospital Geral de Sao Paulo | Instituto Adolfo Lutz, Interdiciplinary Procedures Center, Strategic Laboratory | Claudio Tavares Sacchi et al |
| EPI_ISL_943968 | Hospital Geral de Sao Paulo | Instituto Adolfo Lutz, Interdiciplinary Procedures Center, Strategic Laboratory | Claudio Tavares Sacchi et al |
| EPI_ISL_943969 | Hospital Geral de Sao Paulo | Instituto Adolfo Lutz, Interdiciplinary Procedures Center, Strategic Laboratory | Claudio Tavares Sacchi et al |
| EPI_ISL_943970 | Hospital Geral de Sao Paulo | Instituto Adolfo Lutz, Interdiciplinary Procedures Center, Strategic Laboratory | Claudio Tavares Sacchi et al |
| EPI_ISL_943971 | Hospital Geral de Sao Paulo | Instituto Adolfo Lutz, Interdiciplinary Procedures Center, Strategic Laboratory | Claudio Tavares Sacchi et al |
| EPI_ISL_1121316 | LACEN do Rio Grande do Sul | Instituto Adolfo Lutz, Interdiciplinary Procedures Center, Strategic Laboratory | Claudio Tavares Sacchi et al |
| EPI_ISL_985318 | LACEN de Santa Catarina | Instituto Adolfo Lutz, Interdiciplinary Procedures Center, Strategic Laboratory | Claudio Tavares Sacchi et al |
| EPI_ISL_985319 | LACEN de Santa Catarina | Instituto Adolfo Lutz, Interdiciplinary Procedures Center, Strategic Laboratory | Claudio Tavares Sacchi et al |
| EPI_ISL_414016 | Hospital São Joaquim Beneficencia Portuguesa | Instituto Adolfo Lutz, Interdiciplinary Procedures Center, Strategic Laboratory | Claudio Tavares Sacchi et al |
| EPI_ISL_414015 | Hospital São Joaquim Beneficencia Portuguesa | Instituto Adolfo Lutz, Interdiciplinary Procedures Center, Strategic Laboratory | Claudio Tavares Sacchi et al |
| EPI_ISL_416032 | National Influenza Center - Instituto Adolfo Lutz | Instituto Adolfo Lutz, Interdiciplinary Procedures Center, Strategic Laboratory | Claudio Tavares Sacchi et al |
| EPI_ISL_861680 | Hospital e Pronto Socorro Portinari | Instituto Adolfo Lutz, Interdiciplinary Procedures Center, Strategic Laboratory | Claudio Tavares Sacchi et al |
| EPI_ISL_861682 | UPA Vila Santa Catarina | Instituto Adolfo Lutz, Interdiciplinary Procedures Center, Strategic Laboratory | Claudio Tavares Sacchi et al |
| EPI_ISL_882665 | Unidade de Pronto Atendimento Dra Zilda Arns | Instituto Adolfo Lutz, Interdiciplinary Procedures Center, Strategic Laboratory | Claudio Tavares Sacchi et al |
| EPI_ISL_882671 | Hospital Municipal Dr. Guido Guida | Instituto Adolfo Lutz, Interdiciplinary Procedures Center, Strategic Laboratory | Claudio Tavares Sacchi et al |
| EPI_ISL_882672 | Hospital Municipal Dr. Guido Guida | Instituto Adolfo Lutz, Interdiciplinary Procedures Center, Strategic Laboratory | Claudio Tavares Sacchi et al |
| EPI_ISL_977471 | Instituto Adolfo Lutz - Regional de Presidente Prudente | Instituto Adolfo Lutz, Interdiciplinary Procedures Center, Strategic Laboratory | Claudio Tavares Sacchi et al |
| EPI_ISL_977472 | Instituto Adolfo Lutz Central | Instituto Adolfo Lutz, Interdiciplinary Procedures Center, Strategic Laboratory | Claudio Tavares Sacchi et al |
| EPI_ISL_977474 | Instituto Adolfo Lutz Central | Instituto Adolfo Lutz, Interdiciplinary Procedures Center, Strategic Laboratory | Claudio Tavares Sacchi et al |
| EPI_ISL_985172 | Instituto Adolfo Lutz - Regional de Taubate | Instituto Adolfo Lutz, Interdiciplinary Procedures Center, Strategic Laboratory | Claudio Tavares Sacchi et al |
| EPI_ISL_985173 | Instituto Adolfo Lutz - Regional de Taubate | Instituto Adolfo Lutz, Interdiciplinary Procedures Center, Strategic Laboratory | Claudio Tavares Sacchi et al |

| | | | |
|---|---|---|---|
| EPI_ISL_985175 | Instituto Adolfo Lutz Central | Instituto Adolfo Lutz, Interdiciplinary Procedures Center, Strategic Laboratory | Claudio Tavares Sacchi et al |
| EPI_ISL_985171 | Instituto Adolfo Lutz - Regional de Taubate | Instituto Adolfo Lutz, Interdiciplinary Procedures Center, Strategic Laboratory | Claudio Tavares Sacchi et al |
| EPI_ISL_1171620 | Instituto Adolfo Lutz Central | Instituto Adolfo Lutz, Interdiciplinary Procedures Center, Strategic Laboratory | Claudio Tavares Sacchi et al |
| EPI_ISL_977476 | Instituto Adolfo Lutz Central | Instituto Adolfo Lutz, Interdiciplinary Procedures Center, Strategic Laboratory | Claudio Tavares Sacchi et al |
| EPI_ISL_977477 | Instituto Adolfo Lutz Central | Instituto Adolfo Lutz, Interdiciplinary Procedures Center, Strategic Laboratory | Claudio Tavares Sacchi et al |
| EPI_ISL_984243 | Instituto Adolfo Lutz - Regional de Marilia | Instituto Adolfo Lutz, Interdiciplinary Procedures Center, Strategic Laboratory | Claudio Tavares Sacchi et al |
| EPI_ISL_984244 | Instituto Adolfo Lutz - Regional de Marilia | Instituto Adolfo Lutz, Interdiciplinary Procedures Center, Strategic Laboratory | Claudio Tavares Sacchi et al |
| EPI_ISL_984246 | Instituto Adolfo Lutz Central | Instituto Adolfo Lutz, Interdiciplinary Procedures Center, Strategic Laboratory | Claudio Tavares Sacchi et al |
| EPI_ISL_1039697 | Instituto Adolfo Lutz Central | Instituto Adolfo Lutz, Interdiciplinary Procedures Center, Strategic Laboratory | Claudio Tavares Sacchi et al |
| EPI_ISL_1039698 | Lab Loc - Itapecerica da Serra | Instituto Adolfo Lutz, Interdiciplinary Procedures Center, Strategic Laboratory | Claudio Tavares Sacchi et al |
| EPI_ISL_1039699 | Instituto Adolfo Lutz - Regional de Taubate | Instituto Adolfo Lutz, Interdiciplinary Procedures Center, Strategic Laboratory | Claudio Tavares Sacchi et al |
| EPI_ISL_1039700 | Instituto Adolfo Lutz Central | Instituto Adolfo Lutz, Interdiciplinary Procedures Center, Strategic Laboratory | Claudio Tavares Sacchi et al |
| EPI_ISL_1039701 | Instituto Adolfo Lutz Central | Instituto Adolfo Lutz, Interdiciplinary Procedures Center, Strategic Laboratory | Claudio Tavares Sacchi et al |
| EPI_ISL_1039703 | Instituto Adolfo Lutz - Regional de Taubate | Instituto Adolfo Lutz, Interdiciplinary Procedures Center, Strategic Laboratory | Claudio Tavares Sacchi et al |
| EPI_ISL_1039704 | Lab Loc - Itapecerica da Serra | Instituto Adolfo Lutz, Interdiciplinary Procedures Center, Strategic Laboratory | Claudio Tavares Sacchi et al |
| EPI_ISL_1039705 | Instituto Adolfo Lutz Central | Instituto Adolfo Lutz, Interdiciplinary Procedures Center, Strategic Laboratory | Claudio Tavares Sacchi et al |
| EPI_ISL_1039706 | Instituto Adolfo Lutz Central | Instituto Adolfo Lutz, Interdiciplinary Procedures Center, Strategic Laboratory | Claudio Tavares Sacchi et al |
| EPI_ISL_1039707 | Instituto Adolfo Lutz Central | Instituto Adolfo Lutz, Interdiciplinary Procedures Center, Strategic Laboratory | Claudio Tavares Sacchi et al |
| EPI_ISL_1039708 | Instituto Adolfo Lutz Central | Instituto Adolfo Lutz, Interdiciplinary Procedures Center, Strategic Laboratory | Claudio Tavares Sacchi et al |
| EPI_ISL_1039709 | Instituto Adolfo Lutz Central | Instituto Adolfo Lutz, Interdiciplinary Procedures Center, Strategic Laboratory | Claudio Tavares Sacchi et al |
| EPI_ISL_1039710 | Instituto Adolfo Lutz Central | Instituto Adolfo Lutz, Interdiciplinary Procedures Center, Strategic Laboratory | Claudio Tavares Sacchi et al |
| EPI_ISL_977482 | Instituto Adolfo Lutz - Regional de Aracatuba | Instituto Adolfo Lutz, Interdiciplinary Procedures Center, Strategic Laboratory | Claudio Tavares Sacchi et al |

| | | | |
|---|---|---|---|
| EPI_ISL_977483 | Instituto Adolfo Lutz Central | Instituto Adolfo Lutz, Interdiciplinary Procedures Center, Strategic Laboratory | Claudio Tavares Sacchi et al |
| EPI_ISL_977487 | Instituto Adolfo Lutz Central | Instituto Adolfo Lutz, Interdiciplinary Procedures Center, Strategic Laboratory | Claudio Tavares Sacchi et al |
| EPI_ISL_977488 | Instituto Adolfo Lutz - Regional de Presidente Prudente | Instituto Adolfo Lutz, Interdiciplinary Procedures Center, Strategic Laboratory | Claudio Tavares Sacchi et al |
| EPI_ISL_984252 | IAL Regional de Marilia | Instituto Adolfo Lutz, Interdiciplinary Procedures Center, Strategic Laboratory | Claudio Tavares Sacchi et al |
| EPI_ISL_984253 | IAL Regional de Marilia | Instituto Adolfo Lutz, Interdiciplinary Procedures Center, Strategic Laboratory | Claudio Tavares Sacchi et al |
| EPI_ISL_984254 | IAL Regional de Marilia | Instituto Adolfo Lutz, Interdiciplinary Procedures Center, Strategic Laboratory | Claudio Tavares Sacchi et al |
| EPI_ISL_984263 | IAL Regional de Bauru | Instituto Adolfo Lutz, Interdiciplinary Procedures Center, Strategic Laboratory | Claudio Tavares Sacchi et al |
| EPI_ISL_1121326 | IAL Regional de Bauru | Instituto Adolfo Lutz, Interdiciplinary Procedures Center, Strategic Laboratory | Claudio Tavares Sacchi et al |
| EPI_ISL_468311 | Hospital Municipal Dr Ignacio Proenca de Gouvea | Instituto Adolfo Lutz, Interdiciplinary Procedures Center, Strategic Laboratory | Claudio Tavares Sacchi et al |
| EPI_ISL_1219031 | Aeroporto Internacional de Guarulhos | Instituto Adolfo Lutz, Interdiciplinary Procedures Center, Strategic Laboratory | Claudio Tavares Sacchi et al |
| EPI_ISL_1171625 | IAL Regional de Santos | Instituto Adolfo Lutz, Interdiciplinary Procedures Center, Strategic Laboratory | Claudio Tavares Sacchi et al |
| EPI_ISL_1171627 | IAL Regional de Santos | Instituto Adolfo Lutz, Interdiciplinary Procedures Center, Strategic Laboratory | Claudio Tavares Sacchi et al |
| EPI_ISL_1171631 | IAL Regional de Santos | Instituto Adolfo Lutz, Interdiciplinary Procedures Center, Strategic Laboratory | Claudio Tavares Sacchi et al |
| EPI_ISL_1171633 | IAL Regional de Santos | Instituto Adolfo Lutz, Interdiciplinary Procedures Center, Strategic Laboratory | Claudio Tavares Sacchi et al |
| EPI_ISL_1171635 | IAL Regional de Santos | Instituto Adolfo Lutz, Interdiciplinary Procedures Center, Strategic Laboratory | Claudio Tavares Sacchi et al |
| EPI_ISL_1196297 | IAL Regional de Marilia | Instituto Adolfo Lutz, Interdiciplinary Procedures Center, Strategic Laboratory | Claudio Tavares Sacchi et al |
| EPI_ISL_1171645 | IAL Regional de Marilia | Instituto Adolfo Lutz, Interdiciplinary Procedures Center, Strategic Laboratory | Claudio Tavares Sacchi et al |
| EPI_ISL_1201890 | IAL Regional de Marilia | Instituto Adolfo Lutz, Interdiciplinary Procedures Center, Strategic Laboratory | Claudio Tavares Sacchi et al |
| EPI_ISL_1201891 | IAL Regional de Marilia | Instituto Adolfo Lutz, Interdiciplinary Procedures Center, Strategic Laboratory | Claudio Tavares Sacchi et al |
| EPI_ISL_1201892 | IAL Regional de Marilia | Instituto Adolfo Lutz, Interdiciplinary Procedures Center, Strategic Laboratory | Claudio Tavares Sacchi et al |
| EPI_ISL_523969 | Hospital Sao Paulo de Ensino da Unifesp | Instituto Adolfo Lutz, Interdiciplinary Procedures Center, Strategic Laboratory | Claudio Tavares Sacchi et al |
| EPI_ISL_1079008 | IAL Regional de Bauru | Instituto Adolfo Lutz, Interdiciplinary Procedures Center, Strategic Laboratory | Claudio Tavares Sacchi et al |

| | | | |
|---|---|---|---|
| EPI_ISL_1086051 | IAL Regional de Bauru | Instituto Adolfo Lutz, Interdiciplinary Procedures Center, Strategic Laboratory | Claudio Tavares Sacchi et al |
| EPI_ISL_1078999 | IAL Regional de Bauru | Instituto Adolfo Lutz, Interdiciplinary Procedures Center, Strategic Laboratory | Claudio Tavares Sacchi et al |
| EPI_ISL_1171654 | IAL Regional de Presidente Prudente | Instituto Adolfo Lutz, Interdiciplinary Procedures Center, Strategic Laboratory | Claudio Tavares Sacchi et al |
| EPI_ISL_1171667 | IAL Regional de Presidente Prudente | Instituto Adolfo Lutz, Interdiciplinary Procedures Center, Strategic Laboratory | Claudio Tavares Sacchi et al |
| EPI_ISL_1171668 | IAL Regional de Presidente Prudente | Instituto Adolfo Lutz, Interdiciplinary Procedures Center, Strategic Laboratory | Claudio Tavares Sacchi et al |
| EPI_ISL_1171669 | IAL Regional de Presidente Prudente | Instituto Adolfo Lutz, Interdiciplinary Procedures Center, Strategic Laboratory | Claudio Tavares Sacchi et al |
| EPI_ISL_1219027 | IAL Regional de Presidente Prudente | Instituto Adolfo Lutz, Interdiciplinary Procedures Center, Strategic Laboratory | Claudio Tavares Sacchi et al |
| EPI_ISL_1293057 | IAL Regional de Sorocaba | Instituto Adolfo Lutz, Interdiciplinary Procedures Center, Strategic Laboratory | Claudio Tavares Sacchi et al |
| EPI_ISL_1293069 | IAL Regional de Sorocaba | Instituto Adolfo Lutz, Interdiciplinary Procedures Center, Strategic Laboratory | Claudio Tavares Sacchi et al |
| EPI_ISL_1293071 | IAL Regional de Sorocaba | Instituto Adolfo Lutz, Interdiciplinary Procedures Center, Strategic Laboratory | Claudio Tavares Sacchi et al |
| EPI_ISL_1303527 | IAL Regional de São Jose do Rio Preto | Instituto Adolfo Lutz, Interdiciplinary Procedures Center, Strategic Laboratory | Claudio Tavares Sacchi et al |
| EPI_ISL_1358295 | IAL Regional de Santo Andre | Instituto Adolfo Lutz, Interdiciplinary Procedures Center, Strategic Laboratory | Claudio Tavares Sacchi et al |
| EPI_ISL_1381051 | IAL Regional de Santo Andre | Instituto Adolfo Lutz, Interdiciplinary Procedures Center, Strategic Laboratory | Claudio Tavares Sacchi et al |
| EPI_ISL_1468449 | Santa Casa de Aracatuba Hospital Sagrado Coracao de Jesus | Instituto Adolfo Lutz, Interdiciplinary Procedures Center, Strategic Laboratory | Claudio Tavares Sacchi et al |
| EPI_ISL_1468453 | Santa Casa de Misericordia de Pereira Barreto | Instituto Adolfo Lutz, Interdiciplinary Procedures Center, Strategic Laboratory | Claudio Tavares Sacchi et al |
| EPI_ISL_1468454 | Santa Casa de Andradina | Instituto Adolfo Lutz, Interdiciplinary Procedures Center, Strategic Laboratory | Claudio Tavares Sacchi et al |
| EPI_ISL_1468459 | Secretaria Municipal de Saude de Valparaiso SP | Instituto Adolfo Lutz, Interdiciplinary Procedures Center, Strategic Laboratory | Claudio Tavares Sacchi et al |
| EPI_ISL_1468469 | Secretaria Municipal de Saude Porto Ferreira | Instituto Adolfo Lutz, Interdiciplinary Procedures Center, Strategic Laboratory | Claudio Tavares Sacchi et al |
| EPI_ISL_1520113 | Hospital Santo Antonio de Juquia Juquia | Instituto Adolfo Lutz, Interdiciplinary Procedures Center, Strategic Laboratory | Claudio Tavares Sacchi et al |
| EPI_ISL_1533697 | Hospital Estadual de Vila Alpina | Instituto Adolfo Lutz, Interdiciplinary Procedures Center, Strategic Laboratory | Claudio Tavares Sacchi et al |
| EPI_ISL_1533700 | Hospital Estadual de Sapopemba Sao Paulo | Instituto Adolfo Lutz, Interdiciplinary Procedures Center, Strategic Laboratory | Claudio Tavares Sacchi et al |
| EPI_ISL_527868 | Hospital e Maternidade do Braz | Instituto Adolfo Lutz, Interdiciplinary Procedures Center, Strategic Laboratory | Claudio Tavares Sacchi et al |

| | | | |
|---|---|---|---|
| EPI_ISL_1628346 | Instituto Adolfo Lutz - Regional de Rio Claro | Instituto Adolfo Lutz, Interdiciplinary Procedures Center, Strategic Laboratory | Claudio Tavares Sacchi et al |
| EPI_ISL_524468 | Hospital Municipal Vereador Jose Storopolli | Instituto Adolfo Lutz, Interdiciplinary Procedures Center, Strategic Laboratory | Claudio Tavares Sacchi et al |
| EPI_ISL_693196 | Hospital Santa Clara | Instituto Adolfo Lutz, Interdiciplinary Procedures Center, Strategic Laboratory | Claudio Tavares Sacchi et al |
| EPI_ISL_693198 | Santa Casa de Misericordia de Sao Paulo - Hospital Central | Instituto Adolfo Lutz, Interdiciplinary Procedures Center, Strategic Laboratory | Claudio Tavares Sacchi et al |
| EPI_ISL_693199 | Hospital do Servidor Publico Estadual Francisco Morato de Oliveira | Instituto Adolfo Lutz, Interdiciplinary Procedures Center, Strategic Laboratory | Claudio Tavares Sacchi et al |
| EPI_ISL_534319 | Hospital do Serv Pub ESTAFCO Morato de Oliveira | Instituto Adolfo Lutz, Interdiciplinary Procedures Center, Strategic Laboratory | Claudio Tavares Sacchi et al |
| EPI_ISL_534320 | Hospital do Serv Pub ESTAFCO Morato de Oliveira | Instituto Adolfo Lutz, Interdiciplinary Procedures Center, Strategic Laboratory | Claudio Tavares Sacchi et al |
| EPI_ISL_524464 | Santa Casa de Santa Isabel | Instituto Adolfo Lutz, Interdiciplinary Procedures Center, Strategic Laboratory | Claudio Tavares Sacchi et al |
| EPI_ISL_527858 | Pronto Atendimento Sancta Maggiore Jardim Paulista | Instituto Adolfo Lutz, Interdiciplinary Procedures Center, Strategic Laboratory | Claudio Tavares Sacchi et al |
| EPI_ISL_527856 | Hospital Municipal Prof. Waldomiro de Paula | Instituto Adolfo Lutz, Interdiciplinary Procedures Center, Strategic Laboratory | Claudio Tavares Sacchi et al |
| EPI_ISL_534314 | Hospital Universitario da USP de SP | Instituto Adolfo Lutz, Interdiciplinary Procedures Center, Strategic Laboratory | Claudio Tavares Sacchi et al |
| EPI_ISL_534318 | Hospital Municipal Antonio Giglio | Instituto Adolfo Lutz, Interdiciplinary Procedures Center, Strategic Laboratory | Claudio Tavares Sacchi et al |
| EPI_ISL_547573 | Vigilância em Saúde de Cajamar | Instituto Adolfo Lutz, Interdiciplinary Procedures Center, Strategic Laboratory | Claudio Tavares Sacchi et al |
| EPI_ISL_583495 | Serviço de Verificação de Óbitos SVO Guaruulhos | Instituto Adolfo Lutz, Interdiciplinary Procedures Center, Strategic Laboratory | Claudio Tavares Sacchi et al |
| EPI_ISL_547575 | SVO Jundiaí | Instituto Adolfo Lutz, Interdiciplinary Procedures Center, Strategic Laboratory | Claudio Tavares Sacchi et al |
| EPI_ISL_693207 | Cs II Doutor Antonio Vicoso Moreira de Rezende | Instituto Adolfo Lutz, Interdiciplinary Procedures Center, Strategic Laboratory | Claudio Tavares Sacchi et al |
| EPI_ISL_583499 | Distrito Sanitario Sul Campinas | Instituto Adolfo Lutz, Interdiciplinary Procedures Center, Strategic Laboratory | Claudio Tavares Sacchi et al |
| EPI_ISL_574597 | Secretaria Municipal de Saude de Jarinu | Instituto Adolfo Lutz, Interdiciplinary Procedures Center, Strategic Laboratory | Claudio Tavares Sacchi et al |
| EPI_ISL_574598 | Servico de Verificacao de Obito SVO | Instituto Adolfo Lutz, Interdiciplinary Procedures Center, Strategic Laboratory | Claudio Tavares Sacchi et al |
| EPI_ISL_583502 | Serv de Vig Sanitaria Epidemio e CTRL de Zoonoses Guaruja | Instituto Adolfo Lutz, Interdiciplinary Procedures Center, Strategic Laboratory | Claudio Tavares Sacchi et al |
| EPI_ISL_583505 | Casa de Saude Stella Maris | Instituto Adolfo Lutz, Interdiciplinary Procedures Center, Strategic Laboratory | Claudio Tavares Sacchi et al |
| EPI_ISL_693208 | Hospital Municipal Antonio Giglio | Instituto Adolfo Lutz, Interdiciplinary Procedures Center, Strategic Laboratory | Claudio Tavares Sacchi et al |

| | | | |
|---|---|---|---|
| EPI_ISL_547577 | Hospital e Maternidade Nossa Senhora das Graças | Instituto Adolfo Lutz, Interdiciplinary Procedures Center, Strategic Laboratory | Claudio Tavares Sacchi et al |
| EPI_ISL_547579 | Santa Casa de Misericórdia de Araçatuba | Instituto Adolfo Lutz, Interdiciplinary Procedures Center, Strategic Laboratory | Claudio Tavares Sacchi et al |
| EPI_ISL_574594 | Hospital Escola da Universidade de Taubate | Instituto Adolfo Lutz, Interdiciplinary Procedures Center, Strategic Laboratory | Claudio Tavares Sacchi et al |
| EPI_ISL_603037 | Hospital Geral de Pedreira | Instituto Adolfo Lutz, Interdiciplinary Procedures Center, Strategic Laboratory | Claudio Tavares Sacchi et al |
| EPI_ISL_471552 | Hospital Sancta Maggiore | Instituto Adolfo Lutz, Interdiciplinary Procedures Center, Strategic Laboratory | Claudio Tavares Sacchi et al |
| EPI_ISL_735396 | Hospital de Camplanha COVID 19 SER | Instituto Adolfo Lutz, Interdiciplinary Procedures Center, Strategic Laboratory | Claudio Tavares Sacchi et al |
| EPI_ISL_693215 | Secretaria Municipal de Saúde de Iracemapolis | Instituto Adolfo Lutz, Interdiciplinary Procedures Center, Strategic Laboratory | Claudio Tavares Sacchi et al |
| EPI_ISL_471545 | Hospital Sao Paulo de Ensino da Unifesp | Instituto Adolfo Lutz, Interdiciplinary Procedures Center, Strategic Laboratory | Claudio Tavares Sacchi et al |
| EPI_ISL_515566 | PS Municipal Dr Lauro Ribas Braga | Instituto Adolfo Lutz, Interdiciplinary Procedures Center, Strategic Laboratory | Claudio Tavares Sacchi et al |
| EPI_ISL_515554 | Pronto Socorro Municipal de Perus | Instituto Adolfo Lutz, Interdiciplinary Procedures Center, Strategic Laboratory | Claudio Tavares Sacchi et al |
| EPI_ISL_735402 | Instituto Adolfo Lutz - Regional de Rio Claro | Instituto Adolfo Lutz, Interdiciplinary Procedures Center, Strategic Laboratory | Claudio Tavares Sacchi et al |
| EPI_ISL_735403 | Instituto Adolfo Lutz - Regional de Rio Claro | Instituto Adolfo Lutz, Interdiciplinary Procedures Center, Strategic Laboratory | Claudio Tavares Sacchi et al |
| EPI_ISL_735404 | Instituto Adolfo Lutz - Regional de Rio Claro | Instituto Adolfo Lutz, Interdiciplinary Procedures Center, Strategic Laboratory | Claudio Tavares Sacchi et al |
| EPI_ISL_735406 | Unidade de Pronto Atendimento UPA I Sta Isabel | Instituto Adolfo Lutz, Interdiciplinary Procedures Center, Strategic Laboratory | Claudio Tavares Sacchi et al |
| EPI_ISL_735412 | Hospital e Pronto Socorro Portinari | Instituto Adolfo Lutz, Interdiciplinary Procedures Center, Strategic Laboratory | Claudio Tavares Sacchi et al |
| EPI_ISL_735413 | Militello Centro de Diagnosticos e Biopesquisa Clinica | Instituto Adolfo Lutz, Interdiciplinary Procedures Center, Strategic Laboratory | Claudio Tavares Sacchi et al |
| EPI_ISL_735418 | Hospital Regional do Vale do Paraiba | Instituto Adolfo Lutz, Interdiciplinary Procedures Center, Strategic Laboratory | Claudio Tavares Sacchi et al |
| EPI_ISL_735422 | UBS Dematchi | Instituto Adolfo Lutz, Interdiciplinary Procedures Center, Strategic Laboratory | Claudio Tavares Sacchi et al |
| EPI_ISL_776764 | Instituto Adolfo Lutz - Regional de Santo Andre | Instituto Adolfo Lutz, Interdiciplinary Procedures Center, Strategic Laboratory | Claudio Tavares Sacchi et al |
| EPI_ISL_776765 | Instituto Adolfo Lutz - Regional de Santo Andre | Instituto Adolfo Lutz, Interdiciplinary Procedures Center, Strategic Laboratory | Claudio Tavares Sacchi et al |
| EPI_ISL_792116 | Instituto Adolfo Lutz - Regional de Taubate | Instituto Adolfo Lutz, Interdiciplinary Procedures Center, Strategic Laboratory | Claudio Tavares Sacchi et al |
| EPI_ISL_735433 | Posto de Atendimento Saude Cidade Pasc Cajati | Instituto Adolfo Lutz, Interdiciplinary Procedures Center, Strategic Laboratory | Claudio Tavares Sacchi et al |

| | | | |
|---|---|---|---|
| EPI_ISL_940608 | Laboratório Sao Lucas | Instituto Adolfo Lutz, Interdiciplinary Procedures Center, Strategic Laboratory | Claudio Tavares Sacchi et al |
| EPI_ISL_833156 | Instituto Adolfo Lutz - Regional de Sorocaba | Instituto Adolfo Lutz, Interdiciplinary Procedures Center, Strategic Laboratory | Claudio Tavares Sacchi et al |
| EPI_ISL_861627 | Instituto Adolfo Lutz - Central | Instituto Adolfo Lutz, Interdiciplinary Procedures Center, Strategic Laboratory | Claudio Tavares Sacchi et al |
| EPI_ISL_776752 | Instituto Adolfo Lutz - Central | Instituto Adolfo Lutz, Interdiciplinary Procedures Center, Strategic Laboratory | Claudio Tavares Sacchi et al |
| EPI_ISL_792108 | Instituto Adolfo Lutz - Central | Instituto Adolfo Lutz, Interdiciplinary Procedures Center, Strategic Laboratory | Claudio Tavares Sacchi et al |
| EPI_ISL_792107 | Instituto Adolfo Lutz - Central | Instituto Adolfo Lutz, Interdiciplinary Procedures Center, Strategic Laboratory | Claudio Tavares Sacchi et al |
| EPI_ISL_708530 | Secretaria Municipal de Saude de Fernandópolis | Instituto Adolfo Lutz, Interdiciplinary Procedures Center, Strategic Laboratory | Claudio Tavares Sacchi et al |
| EPI_ISL_861636 | Hospital Geral de Sao Mateus São Paulo | Instituto Adolfo Lutz, Interdiciplinary Procedures Center, Strategic Laboratory | Claudio Tavares Sacchi et al |
| EPI_ISL_861639 | Hospital Sao Paulo de Ensino da Unifesp | Instituto Adolfo Lutz, Interdiciplinary Procedures Center, Strategic Laboratory | Claudio Tavares Sacchi et al |
| EPI_ISL_776766 | Instituto Adolfo Lutz - Regional de Santo Andre | Instituto Adolfo Lutz, Interdiciplinary Procedures Center, Strategic Laboratory | Claudio Tavares Sacchi et al |
| EPI_ISL_776767 | Instituto Adolfo Lutz - Regional de Marilia | Instituto Adolfo Lutz, Interdiciplinary Procedures Center, Strategic Laboratory | Claudio Tavares Sacchi et al |
| EPI_ISL_776768 | Instituto Adolfo Lutz - Regional de Aracatuba | Instituto Adolfo Lutz, Interdiciplinary Procedures Center, Strategic Laboratory | Claudio Tavares Sacchi et al |
| EPI_ISL_755641 | Instituto Adolfo Lutz - Regional de Santo Andre | Instituto Adolfo Lutz, Interdiciplinary Procedures Center, Strategic Laboratory | Claudio Tavares Sacchi et al |
| EPI_ISL_755642 | Instituto Adolfo Lutz - Central | Instituto Adolfo Lutz, Interdiciplinary Procedures Center, Strategic Laboratory | Claudio Tavares Sacchi et al |
| EPI_ISL_755647 | Instituto Adolfo Lutz - Regional de Santo Andre | Instituto Adolfo Lutz, Interdiciplinary Procedures Center, Strategic Laboratory | Claudio Tavares Sacchi et al |
| EPI_ISL_755648 | Instituto Adolfo Lutz - Regional de Taubate | Instituto Adolfo Lutz, Interdiciplinary Procedures Center, Strategic Laboratory | Claudio Tavares Sacchi et al |
| EPI_ISL_755653 | Instituto Adolfo Lutz - Central | Instituto Adolfo Lutz, Interdiciplinary Procedures Center, Strategic Laboratory | Claudio Tavares Sacchi et al |
| EPI_ISL_755654 | Instituto Adolfo Lutz - Central | Instituto Adolfo Lutz, Interdiciplinary Procedures Center, Strategic Laboratory | Claudio Tavares Sacchi et al |
| EPI_ISL_861668 | Centro de Triagem Covid19 | Instituto Adolfo Lutz, Interdiciplinary Procedures Center, Strategic Laboratory | Claudio Tavares Sacchi et al |
| EPI_ISL_833157 | Instituto Adolfo Lutz - Regional de Santo Andre | Instituto Adolfo Lutz, Interdiciplinary Procedures Center, Strategic Laboratory | Claudio Tavares Sacchi et al |
| EPI_ISL_833158 | Instituto Adolfo Lutz - Regional de Santo Andre | Instituto Adolfo Lutz, Interdiciplinary Procedures Center, Strategic Laboratory | Claudio Tavares Sacchi et al |
| EPI_ISL_861669 | Lab LOC - Itapecerica da Serra | Instituto Adolfo Lutz, Interdiciplinary Procedures Center, Strategic Laboratory | Claudio Tavares Sacchi et al |

| | | | |
|---|---|---|---|
| EPI_ISL_837053 | UBS Darcy Alves e Robalinho | Instituto Adolfo Lutz, Interdiciplinary Procedures Center, Strategic Laboratory | Claudio Tavares Sacchi et al |
| EPI_ISL_837054 | UBS Jose Sabino Ferreira | Instituto Adolfo Lutz, Interdiciplinary Procedures Center, Strategic Laboratory | Claudio Tavares Sacchi et al |
| EPI_ISL_833163 | Instituto Adolfo Lutz - Regional de Marilia | Instituto Adolfo Lutz, Interdiciplinary Procedures Center, Strategic Laboratory | Claudio Tavares Sacchi et al |
| EPI_ISL_833164 | Secretaria Municipal de Saude de Santa Barbara d'oeste | Instituto Adolfo Lutz, Interdiciplinary Procedures Center, Strategic Laboratory | Claudio Tavares Sacchi et al |
| EPI_ISL_882658 | Secretaria Municipal de Saude | Instituto Adolfo Lutz, Interdiciplinary Procedures Center, Strategic Laboratory | Claudio Tavares Sacchi et al |
| EPI_ISL_882659 | Centro de Triagem Covid19 | Instituto Adolfo Lutz, Interdiciplinary Procedures Center, Strategic Laboratory | Claudio Tavares Sacchi et al |
| EPI_ISL_882660 | Hospital Municipal Prefeito Waldemar Costa Filho | Instituto Adolfo Lutz, Interdiciplinary Procedures Center, Strategic Laboratory | Claudio Tavares Sacchi et al |
| EPI_ISL_861671 | Hospital Municipal Prefeito Waldemar Costa Filho | Instituto Adolfo Lutz, Interdiciplinary Procedures Center, Strategic Laboratory | Claudio Tavares Sacchi et al |
| EPI_ISL_861672 | Day Hospital de Ermelino Matarazzo | Instituto Adolfo Lutz, Interdiciplinary Procedures Center, Strategic Laboratory | Claudio Tavares Sacchi et al |
| EPI_ISL_861673 | PA Novo Osasco | Instituto Adolfo Lutz, Interdiciplinary Procedures Center, Strategic Laboratory | Claudio Tavares Sacchi et al |
| EPI_ISL_861645 | Hospital e Pronto Socorro Comunitario Vila Iolanda | Instituto Adolfo Lutz, Interdiciplinary Procedures Center, Strategic Laboratory | Claudio Tavares Sacchi et al |
| EPI_ISL_861646 | Hospital Santa Marcelina Sao Paulo | Instituto Adolfo Lutz, Interdiciplinary Procedures Center, Strategic Laboratory | Claudio Tavares Sacchi et al |
| EPI_ISL_861652 | AMA Wamberto Dias da Costa | Instituto Adolfo Lutz, Interdiciplinary Procedures Center, Strategic Laboratory | Claudio Tavares Sacchi et al |
| EPI_ISL_861658 | Hospital Municipal Antônio Giglio | Instituto Adolfo Lutz, Interdiciplinary Procedures Center, Strategic Laboratory | Claudio Tavares Sacchi et al |
| EPI_ISL_861659 | PS e Maternidade Nair Fonseca Leitao Arantes | Instituto Adolfo Lutz, Interdiciplinary Procedures Center, Strategic Laboratory | Claudio Tavares Sacchi et al |
| EPI_ISL_861660 | PS e Maternidade Nair Fonseca Leitao Arantes | Instituto Adolfo Lutz, Interdiciplinary Procedures Center, Strategic Laboratory | Claudio Tavares Sacchi et al |
| EPI_ISL_861676 | UPA Vila Santa Catarina | Instituto Adolfo Lutz, Interdiciplinary Procedures Center, Strategic Laboratory | Claudio Tavares Sacchi et al |
| EPI_ISL_861679 | Instituto Adolfo Lutz - Regional de Taubate | Instituto Adolfo Lutz, Interdiciplinary Procedures Center, Strategic Laboratory | Claudio Tavares Sacchi et al |
| EPI_ISL_1493591 | Centro de Saude II Dr Alcides Facundo Arroyo | Instituto Adolfo Lutz, Interdiciplinary Procedures Center, Strategic Laboratory | Claudio Tavares Sacchi et al |
| EPI_ISL_1201886 | Aeroporto Internacional de Guarulhos | Instituto Adolfo Lutz, Interdiciplinary Procedures Center, Strategic Laboratory | Claudio Tavares Sacchi et al |
| EPI_ISL_943973 | LACEN do Estado de Tocantins | Instituto Adolfo Lutz, Interdiciplinary Procedures Center, Strategic Laboratory | Claudio Tavares Sacchi et al |
| EPI_ISL_943974 | LACEN do Estado de Tocantins | Instituto Adolfo Lutz, Interdiciplinary Procedures Center, Strategic Laboratory | Claudio Tavares Sacchi et al |

| | | | |
|---|---|---|---|
| EPI_ISL_943975 | LACEN do Estado de Tocantins | Instituto Adolfo Lutz, Interdiciplinary Procedures Center, Strategic Laboratory | Claudio Tavares Sacchi et al |
| EPI_ISL_943976 | LACEN do Estado de Tocantins | Instituto Adolfo Lutz, Interdiciplinary Procedures Center, Strategic Laboratory | Claudio Tavares Sacchi et al |
| EPI_ISL_943977 | LACEN do Estado de Tocantins | Instituto Adolfo Lutz, Interdiciplinary Procedures Center, Strategic Laboratory | Claudio Tavares Sacchi et al |
| EPI_ISL_943978 | LACEN do Estado de Tocantins | Instituto Adolfo Lutz, Interdiciplinary Procedures Center, Strategic Laboratory | Claudio Tavares Sacchi et al |
| EPI_ISL_943979 | LACEN do Estado de Tocantins | Instituto Adolfo Lutz, Interdiciplinary Procedures Center, Strategic Laboratory | Claudio Tavares Sacchi et al |
| EPI_ISL_943980 | LACEN do Estado de Tocantins | Instituto Adolfo Lutz, Interdiciplinary Procedures Center, Strategic Laboratory | Claudio Tavares Sacchi et al |
| EPI_ISL_943981 | LACEN do Estado de Tocantins | Instituto Adolfo Lutz, Interdiciplinary Procedures Center, Strategic Laboratory | Claudio Tavares Sacchi et al |
| EPI_ISL_943982 | LACEN do Estado de Tocantins | Instituto Adolfo Lutz, Interdiciplinary Procedures Center, Strategic Laboratory | Claudio Tavares Sacchi et al |
| EPI_ISL_943983 | LACEN do Estado de Tocantins | Instituto Adolfo Lutz, Interdiciplinary Procedures Center, Strategic Laboratory | Claudio Tavares Sacchi et al |
| EPI_ISL_943991 | LACEN do Estado de Tocantins | Instituto Adolfo Lutz, Interdiciplinary Procedures Center, Strategic Laboratory | Claudio Tavares Sacchi et al |
| EPI_ISL_943985 | LACEN do Estado de Tocantins | Instituto Adolfo Lutz, Interdiciplinary Procedures Center, Strategic Laboratory | Claudio Tavares Sacchi et al |
| EPI_ISL_943986 | LACEN do Estado de Tocantins | Instituto Adolfo Lutz, Interdiciplinary Procedures Center, Strategic Laboratory | Claudio Tavares Sacchi et al |
| EPI_ISL_943987 | LACEN do Estado de Tocantins | Instituto Adolfo Lutz, Interdiciplinary Procedures Center, Strategic Laboratory | Claudio Tavares Sacchi et al |
| EPI_ISL_1303509 | LACEN do Estado de Tocantins | Instituto Adolfo Lutz, Interdiciplinary Procedures Center, Strategic Laboratory | Claudio Tavares Sacchi et al |
| EPI_ISL_1358300 | Lacen de Tocantins | Instituto Adolfo Lutz, Interdiciplinary Procedures Center, Strategic Laboratory | Claudio Tavares Sacchi et al |
| EPI_ISL_1358301 | Lacen de Tocantins | Instituto Adolfo Lutz, Interdiciplinary Procedures Center, Strategic Laboratory | Claudio Tavares Sacchi et al |
| EPI_ISL_1358302 | Lacen de Tocantins | Instituto Adolfo Lutz, Interdiciplinary Procedures Center, Strategic Laboratory | Claudio Tavares Sacchi et al |
| EPI_ISL_1358303 | Lacen de Tocantins | Instituto Adolfo Lutz, Interdiciplinary Procedures Center, Strategic Laboratory | Claudio Tavares Sacchi et al |
| EPI_ISL_409067 | Massachusetts Department of Public Health | Pathogen Discovery, Respiratory Viruses Branch, Division of Viral Diseases, Centers for Disease Control and Prevention | Clinton R. Paden et al |
| EPI_ISL_1255105 | West African Centre for Cell Biology of Infectious Pathogens (WACCBIP), University of Ghana, Volta Road, Legon-Accra, Ghana | West African Centre for Cell Biology of Infectious Pathogens (WACCBIP), University of Ghana, Volta Road, Legon-Accra, Ghana | Collins M. Morang'a et al |
| EPI_ISL_1255109 | West African Centre for Cell Biology of Infectious Pathogens (WACCBIP), University of Ghana, Accra, Ghana | West African Centre for Cell Biology of Infectious Pathogens (WACCBIP), University of Ghana, Volta Road, Legon-Accra, Ghana | Collins M. Morang'a et al |
| EPI_ISL_1255112 | West African Centre for Cell Biology of Infectious Pathogens (WACCBIP), University of Ghana, Accra, Ghana | West African Centre for Cell Biology of Infectious Pathogens (WACCBIP), University of Ghana, Volta Road, Legon-Accra, Ghana | Collins M. Morang'a et al |

| | | | |
|---|---|---|---|
| EPI_ISL_1255113 | West African Centre for Cell Biology of Infectious Pathogens (WACCBIP), University of Ghana, Accra, Ghana | West African Centre for Cell Biology of Infectious Pathogens (WACCBIP), University of Ghana, Volta Road, Legon-Accra, Ghana | Collins M. Morang'a et al |
| EPI_ISL_1255142 | West African Centre for Cell Biology of Infectious Pathogens (WACCBIP), University of Ghana, Accra, Ghana | West African Centre for Cell Biology of Infectious Pathogens (WACCBIP), University of Ghana, Volta Road, Legon-Accra, Ghana | Collins M. Morang'a et al |
| EPI_ISL_1255145 | West African Centre for Cell Biology of Infectious Pathogens (WACCBIP), University of Ghana, Accra, Ghana | West African Centre for Cell Biology of Infectious Pathogens (WACCBIP), University of Ghana, Volta Road, Legon-Accra, Ghana | Collins M. Morang'a et al |
| EPI_ISL_1255153 | West African Centre for Cell Biology of Infectious Pathogens (WACCBIP), University of Ghana, Accra, Ghana | West African Centre for Cell Biology of Infectious Pathogens (WACCBIP), University of Ghana, Volta Road, Legon-Accra, Ghana | Collins M. Morang'a et al |
| EPI_ISL_1255164 | West African Centre for Cell Biology of Infectious Pathogens (WACCBIP), University of Ghana, Accra, Ghana | West African Centre for Cell Biology of Infectious Pathogens (WACCBIP), University of Ghana, Volta Road, Legon-Accra, Ghana | Collins M. Morang'a et al |
| EPI_ISL_1255175 | West African Centre for Cell Biology of Infectious Pathogens (WACCBIP), University of Ghana, Accra, Ghana | West African Centre for Cell Biology of Infectious Pathogens (WACCBIP), University of Ghana, Volta Road, Legon-Accra, Ghana | Collins M. Morang'a et al |
| EPI_ISL_1255178 | West African Centre for Cell Biology of Infectious Pathogens (WACCBIP), University of Ghana, Accra, Ghana | West African Centre for Cell Biology of Infectious Pathogens (WACCBIP), University of Ghana, Volta Road, Legon-Accra, Ghana | Collins M. Morang'a et al |
| EPI_ISL_1255186 | West African Centre for Cell Biology of Infectious Pathogens (WACCBIP), University of Ghana, Accra, Ghana | West African Centre for Cell Biology of Infectious Pathogens (WACCBIP), University of Ghana, Volta Road, Legon-Accra, Ghana | Collins M. Morang'a et al |
| EPI_ISL_1255196 | West African Centre for Cell Biology of Infectious Pathogens (WACCBIP), University of Ghana, Accra, Ghana | West African Centre for Cell Biology of Infectious Pathogens (WACCBIP), University of Ghana, Volta Road, Legon-Accra, Ghana | Collins M. Morang'a et al |
| EPI_ISL_1255197 | West African Centre for Cell Biology of Infectious Pathogens (WACCBIP), University of Ghana, Accra, Ghana | West African Centre for Cell Biology of Infectious Pathogens (WACCBIP), University of Ghana, Volta Road, Legon-Accra, Ghana | Collins M. Morang'a et al |
| EPI_ISL_1255198 | West African Centre for Cell Biology of Infectious Pathogens (WACCBIP), University of Ghana, Accra, Ghana | West African Centre for Cell Biology of Infectious Pathogens (WACCBIP), University of Ghana, Volta Road, Legon-Accra, Ghana | Collins M. Morang'a et al |
| EPI_ISL_1255199 | West African Centre for Cell Biology of Infectious Pathogens (WACCBIP), University of Ghana, Accra, Ghana | West African Centre for Cell Biology of Infectious Pathogens (WACCBIP), University of Ghana, Volta Road, Legon-Accra, Ghana | Collins M. Morang'a et al |
| EPI_ISL_1255211 | West African Centre for Cell Biology of Infectious Pathogens (WACCBIP), University of Ghana, Accra, Ghana | West African Centre for Cell Biology of Infectious Pathogens (WACCBIP), University of Ghana, Volta Road, Legon-Accra, Ghana | Collins M. Morang'a et al |
| EPI_ISL_1255231 | West African Centre for Cell Biology of Infectious Pathogens (WACCBIP), University of Ghana, Accra, Ghana | West African Centre for Cell Biology of Infectious Pathogens (WACCBIP), University of Ghana, Volta Road, Legon-Accra, Ghana | Collins M. Morang'a et al |
| EPI_ISL_1255233 | West African Centre for Cell Biology of Infectious Pathogens (WACCBIP), University of Ghana, Accra, Ghana | West African Centre for Cell Biology of Infectious Pathogens (WACCBIP), University of Ghana, Volta Road, Legon-Accra, Ghana | Collins M. Morang'a et al |
| EPI_ISL_1255234 | West African Centre for Cell Biology of Infectious Pathogens (WACCBIP), University of Ghana, Accra, Ghana | West African Centre for Cell Biology of Infectious Pathogens (WACCBIP), University of Ghana, Volta Road, Legon-Accra, Ghana | Collins M. Morang'a et al |
| EPI_ISL_1255240 | West African Centre for Cell Biology of Infectious Pathogens (WACCBIP), University of Ghana, Accra, Ghana | West African Centre for Cell Biology of Infectious Pathogens (WACCBIP), University of Ghana, Volta Road, Legon-Accra, Ghana | Collins M. Morang'a et al |
| EPI_ISL_1255241 | West African Centre for Cell Biology of Infectious Pathogens (WACCBIP), University of Ghana, Accra, Ghana | West African Centre for Cell Biology of Infectious Pathogens (WACCBIP), University of Ghana, Volta Road, Legon-Accra, Ghana | Collins M. Morang'a et al |
| EPI_ISL_1255243 | West African Centre for Cell Biology of Infectious Pathogens (WACCBIP), University of Ghana, Accra, Ghana | West African Centre for Cell Biology of Infectious Pathogens (WACCBIP), University of Ghana, Volta Road, Legon-Accra, Ghana | Collins M. Morang'a et al |
| EPI_ISL_1255250 | West African Centre for Cell Biology of Infectious Pathogens (WACCBIP), University of Ghana, Accra, Ghana | West African Centre for Cell Biology of Infectious Pathogens (WACCBIP), University of Ghana, Volta Road, Legon-Accra, Ghana | Collins M. Morang'a et al |
| EPI_ISL_1255256 | West African Centre for Cell Biology of Infectious Pathogens (WACCBIP), University of Ghana, Accra, Ghana | West African Centre for Cell Biology of Infectious Pathogens (WACCBIP), University of Ghana, Volta Road, Legon-Accra, Ghana | Collins M. Morang'a et al |
| EPI_ISL_1255257 | West African Centre for Cell Biology of Infectious Pathogens (WACCBIP), University of Ghana, Accra, Ghana | West African Centre for Cell Biology of Infectious Pathogens (WACCBIP), University of Ghana, Volta Road, Legon-Accra, Ghana | Collins M. Morang'a et al |

| | | | |
|---|---|---|---|
| EPI_ISL_1255258 | West African Centre for Cell Biology of Infectious Pathogens (WACCBIP), University of Ghana, Accra, Ghana | West African Centre for Cell Biology of Infectious Pathogens (WACCBIP), University of Ghana, Volta Road, Legon-Accra, Ghana | Collins M. Morang'a et al |
| EPI_ISL_1255264 | West African Centre for Cell Biology of Infectious Pathogens (WACCBIP), University of Ghana, Accra, Ghana | West African Centre for Cell Biology of Infectious Pathogens (WACCBIP), University of Ghana, Volta Road, Legon-Accra, Ghana | Collins M. Morang'a et al |
| EPI_ISL_1255267 | West African Centre for Cell Biology of Infectious Pathogens (WACCBIP), University of Ghana, Accra, Ghana | West African Centre for Cell Biology of Infectious Pathogens (WACCBIP), University of Ghana, Volta Road, Legon-Accra, Ghana | Collins M. Morang'a et al |
| EPI_ISL_1255270 | West African Centre for Cell Biology of Infectious Pathogens (WACCBIP), University of Ghana, Accra, Ghana | West African Centre for Cell Biology of Infectious Pathogens (WACCBIP), University of Ghana, Volta Road, Legon-Accra, Ghana | Collins M. Morang'a et al |
| EPI_ISL_448951 | Regional Virus Laboratory, Belfast Health and Social Care Trust | COVID-19 Genomics UK (COG-UK) Consortium | Conall McCaughey et al |
| EPI_ISL_1662047 | Centro de Investigacion Biomedica del Noreste (CIBIN) | Unidad de Genomica Avanzada | Consorcio Mexicano de Vigilancia Genomic |
| EPI_ISL_1662101 | Centro de Investigacion Biomedica de Occidente (CIBO) | Unidad de Genomica Avanzada | Consorcio Mexicano de Vigilancia Genomic |
| EPI_ISL_1279276 | Centro de Investigación Biomédica del Noreste (CIBIN) | Instituto Nacional de Enfermedades Respiratorias (INER): Centro de Investigación en Enfermedades Infecciosas (CIENI) | Consorcio Mexicano de Vigilancia Genómic |
| EPI_ISL_1416455 | Laboratorio Central de Epidemiología (LCE) | Instituto de Biotecnología de la UNAM | Consorcio Mexicano de Vigilancia Genómic |
| EPI_ISL_522686 | Royal Hobart Hospital Microbiology Department | MDU-PHL | Cooley L. et al |
| EPI_ISL_1663229 | National Institute of Infectious Diseases-Prof. Dr. Matei Bals Molecular Diagnostics Laboratory | National Institute of Infectious Diseases-Prof. Dr. Matei Bals Molecular Diagnostics Laboratory | Corina Casangiu et al |
| EPI_ISL_1657093 | National Institute of Infectious Diseases-Prof. Dr. Matei Bals Molecular Diagnostics Laboratory | National Institute of Infectious Diseases-Prof. Dr. Matei Bals Molecular Diagnostics Laboratory | Corina Casangiu et al |
| EPI_ISL_1657104 | Department of Public Health Bucharest | National Institute of Infectious Diseases-Prof. Dr. Matei Bals Molecular Diagnostics Laboratory | Corina Casangiu et al |
| EPI_ISL_1657105 | Department of Public Health Bucharest | National Institute of Infectious Diseases-Prof. Dr. Matei Bals Molecular Diagnostics Laboratory | Corina Casangiu et al |
| EPI_ISL_1657100 | National Institute of Infectious Diseases-Prof. Dr. Matei Bals Molecular Diagnostics Laboratory | National Institute of Infectious Diseases-Prof. Dr. Matei Bals Molecular Diagnostics Laboratory | Corina Casangiu et al |
| EPI_ISL_1517387 | Inciensa, Instituto Costarricense de Investigación y Enseñanza en Nutrición y Salud | Inciensa, Instituto Costarricense de Investigación y Enseñanza en Nutrición y Salud | Cristian Peréz-Corrales et al |
| EPI_ISL_1517392 | Inciensa, Instituto Costarricense de Investigación y Enseñanza en Nutrición y Salud | Inciensa, Instituto Costarricense de Investigación y Enseñanza en Nutrición y Salud | Cristian Peréz-Corrales et al |
| EPI_ISL_1517395 | Inciensa, Instituto Costarricense de Investigación y Enseñanza en Nutrición y Salud | Inciensa, Instituto Costarricense de Investigación y Enseñanza en Nutrición y Salud | Cristian Peréz-Corrales et al |
| EPI_ISL_1517400 | Inciensa, Instituto Costarricense de Investigación y Enseñanza en Nutrición y Salud | Inciensa, Instituto Costarricense de Investigación y Enseñanza en Nutrición y Salud | Cristian Peréz-Corrales et al |
| EPI_ISL_1517403 | Inciensa, Instituto Costarricense de Investigación y Enseñanza en Nutrición y Salud | Inciensa, Instituto Costarricense de Investigación y Enseñanza en Nutrición y Salud | Cristian Peréz-Corrales et al |
| EPI_ISL_1517411 | Inciensa, Instituto Costarricense de Investigación y Enseñanza en Nutrición y Salud | Inciensa, Instituto Costarricense de Investigación y Enseñanza en Nutrición y Salud | Cristian Peréz-Corrales et al |
| EPI_ISL_1517432 | Inciensa, Instituto Costarricense de Investigación y Enseñanza en Nutrición y Salud | Inciensa, Instituto Costarricense de Investigación y Enseñanza en Nutrición y Salud | Cristian Peréz-Corrales et al |
| EPI_ISL_1517433 | Inciensa, Instituto Costarricense de Investigación y Enseñanza en Nutrición y Salud | Inciensa, Instituto Costarricense de Investigación y Enseñanza en Nutrición y Salud | Cristian Peréz-Corrales et al |
| EPI_ISL_824483 | Hospital Universitari Vall d'Hebron - Vall d'Hebron Institut de Recerca | Hospital Universitari Vall d'Hebron - Vall d'Hebron Institut de Recerca | Cristina Andrés et al |

a (CoViGen-Mex). Authors (in alphabetical c

a (CoViGen-Mex). Authors (in alphabetical c

a (CoViGen-Mex). Authors (in alphabetical c

a (CoViGen-Mex). Authors (in alphabetical c

| | | | |
|---|---|---|---|
| EPI_ISL_1116556 | Hospital Universitari Vall d'Hebron - Vall d'Hebron Institut de Recerca | Hospital Universitari Vall d'Hebron - Vall d'Hebron Institut de Recerca | Cristina Andrés et al |
| EPI_ISL_824425 | Hospital Universitari Vall d'Hebron - Vall d'Hebron Institut de Recerca | Hospital Universitari Vall d'Hebron - Vall d'Hebron Institut de Recerca | Cristina Andrés et al |
| EPI_ISL_824490 | Hospital Universitari Vall d'Hebron - Vall d'Hebron Institut de Recerca | Hospital Universitari Vall d'Hebron - Vall d'Hebron Institut de Recerca | Cristina Andrés et al |
| EPI_ISL_1058178 | Hospital Universitari Arnau de Vilanova | Hospital Universitari Vall d'Hebron - Vall d'Hebron Institut de Recerca | Cristina Andrés et al |
| EPI_ISL_824481 | Hospital Universitari Vall d'Hebron - Vall d'Hebron Institut de Recerca | Hospital Universitari Vall d'Hebron - Vall d'Hebron Institut de Recerca | Cristina Andrés et al |
| EPI_ISL_445171 | UCSF Clinical Microbiology Laboratory | Chan-Zuckerberg Biohub | CZB Cliahub Consortium et al |
| EPI_ISL_445177 | UCSF Clinical Microbiology Laboratory | Chan-Zuckerberg Biohub | CZB Cliahub Consortium et al |
| EPI_ISL_445179 | UCSF Clinical Microbiology Laboratory | Chan-Zuckerberg Biohub | CZB Cliahub Consortium et al |
| EPI_ISL_739361 | Alameda County Public Health Lab | Chan-Zuckerberg Biohub | CZB Cliahub Consortium et al |
| EPI_ISL_1093065 | Santa Clara County Public Health Laboratory | Chan-Zuckerberg Biohub | CZB Cliahub Consortium et al |
| EPI_ISL_582914 | County of Santa Clara Public Health Department | Chan-Zuckerberg Biohub | CZB Cliahub Consortium et al |
| EPI_ISL_1648355 | Quest Diagnostics Incorporated | Centers for Disease Control and Prevention Division of Viral Diseases, Pathogen Discovery | Dakota Howard et al |
| EPI_ISL_1649154 | Aegis Sciences Corporation | Centers for Disease Control and Prevention Division of Viral Diseases, Pathogen Discovery | Dakota Howard et al |
| EPI_ISL_476496 | Hospital Garrahan | Héritas | Dalmacio Pereyra et al |
| EPI_ISL_1357603 | Centre For Biotechnology Research And Development | THE AFRICA GENOMICS CENTRE AND CONSULTANCY | Damaris Matoke-Muhia et al |
| EPI_ISL_760034 | Hong Kong Department of Health | School of Public Health, The University of Hong Kong | Daniel Chu et al |
| EPI_ISL_1586072 | The National University Hospital of Iceland | deCODE genetics | Daniel F Gudbjartsson et al |
| EPI_ISL_417765 | The National University Hospital of Iceland | deCODE genetics | Daniel F Gudbjartsson et al |
| EPI_ISL_1586507 | deCODE genetics | deCODE genetics | Daniel F Gudbjartsson et al |
| EPI_ISL_1586817 | deCODE genetics | deCODE genetics | Daniel F Gudbjartsson et al |
| EPI_ISL_1586864 | The National University Hospital of Iceland | deCODE genetics | Daniel F Gudbjartsson et al |
| EPI_ISL_1586118 | The National University Hospital of Iceland | deCODE genetics | Daniel F Gudbjartsson et al |
| EPI_ISL_1586099 | deCODE genetics | deCODE genetics | Daniel F Gudbjartsson et al |
| EPI_ISL_417554 | The National University Hospital of Iceland | deCODE genetics | Daniel F Gudbjartsson et al |
| EPI_ISL_826728 | deCODE genetics | deCODE genetics | Daniel F Gudbjartsson et al |
| EPI_ISL_826731 | deCODE genetics | deCODE genetics | Daniel F Gudbjartsson et al |
| EPI_ISL_826896 | deCODE genetics | deCODE genetics | Daniel F Gudbjartsson et al |
| EPI_ISL_826918 | deCODE genetics | deCODE genetics | Daniel F Gudbjartsson et al |
| EPI_ISL_827051 | The National University Hospital of Iceland | deCODE genetics | Daniel F Gudbjartsson et al |
| EPI_ISL_827128 | deCODE genetics | deCODE genetics | Daniel F Gudbjartsson et al |

| | | | |
|---|---|---|---|
| EPI_ISL_827202 | The National University Hospital of Iceland | deCODE genetics | Daniel F Gudbjartsson et al |
| EPI_ISL_827280 | deCODE genetics | deCODE genetics | Daniel F Gudbjartsson et al |
| EPI_ISL_827382 | deCODE genetics | deCODE genetics | Daniel F Gudbjartsson et al |
| EPI_ISL_827862 | deCODE genetics | deCODE genetics | Daniel F Gudbjartsson et al |
| EPI_ISL_827924 | deCODE genetics | deCODE genetics | Daniel F Gudbjartsson et al |
| EPI_ISL_828009 | The National University Hospital of Iceland | deCODE genetics | Daniel F Gudbjartsson et al |
| EPI_ISL_828071 | deCODE genetics | deCODE genetics | Daniel F Gudbjartsson et al |
| EPI_ISL_828215 | The National University Hospital of Iceland | deCODE genetics | Daniel F Gudbjartsson et al |
| EPI_ISL_828262 | deCODE genetics | deCODE genetics | Daniel F Gudbjartsson et al |
| EPI_ISL_828618 | The National University Hospital of Iceland | deCODE genetics | Daniel F Gudbjartsson et al |
| EPI_ISL_828826 | The National University Hospital of Iceland | deCODE genetics | Daniel F Gudbjartsson et al |
| EPI_ISL_424544 | deCODE genetics | deCODE genetics | Daniel F Gudbjartsson et al |
| EPI_ISL_829138 | deCODE genetics | deCODE genetics | Daniel F Gudbjartsson et al |
| EPI_ISL_829198 | deCODE genetics | deCODE genetics | Daniel F Gudbjartsson et al |
| EPI_ISL_829202 | deCODE genetics | deCODE genetics | Daniel F Gudbjartsson et al |
| EPI_ISL_829731 | deCODE genetics | deCODE genetics | Daniel F Gudbjartsson et al |
| EPI_ISL_829740 | deCODE genetics | deCODE genetics | Daniel F Gudbjartsson et al |
| EPI_ISL_829777 | deCODE genetics | deCODE genetics | Daniel F Gudbjartsson et al |
| EPI_ISL_829891 | The National University Hospital of Iceland | deCODE genetics | Daniel F Gudbjartsson et al |
| EPI_ISL_830189 | deCODE genetics | deCODE genetics | Daniel F Gudbjartsson et al |
| EPI_ISL_830209 | The National University Hospital of Iceland | deCODE genetics | Daniel F Gudbjartsson et al |
| EPI_ISL_830398 | deCODE genetics | deCODE genetics | Daniel F Gudbjartsson et al |
| EPI_ISL_830529 | The National University Hospital of Iceland | deCODE genetics | Daniel F Gudbjartsson et al |
| EPI_ISL_1586696 | The National University Hospital of Iceland | deCODE genetics | Daniel F Gudbjartsson et al |
| EPI_ISL_414378 | National Centre for Infectious Diseases | Programme in Emerging Infectious Diseases, Duke-NUS Medical School | Danielle E Anderson et al |
| EPI_ISL_410535 | National Centre for Infectious Diseases | Programme in Emerging Infectious Diseases, Duke-NUS Medical School | Danielle E Anderson et al |
| EPI_ISL_496605 | Gorgas Memorial Laboratory of Health Studies | Gorgas Memorial Laboratory of Health Studies | Danilo Franco et al |
| EPI_ISL_496606 | Gorgas Memorial Laboratory of Health Studies | Gorgas Memorial Laboratory of Health Studies | Danilo Franco et al |
| EPI_ISL_496607 | Gorgas Memorial Laboratory of Health Studies | Gorgas Memorial Laboratory of Health Studies | Danilo Franco et al |
| EPI_ISL_496609 | Gorgas Memorial Laboratory of Health Studies | Gorgas Memorial Laboratory of Health Studies | Danilo Franco et al |

| | | | |
|---|---|---|---|
| EPI_ISL_496615 | Gorgas Memorial Laboratory of Health Studies | Gorgas Memorial Laboratory of Health Studies | Danilo Franco et al |
| EPI_ISL_496649 | Gorgas Memorial Laboratory of Health Studies | Gorgas Memorial Laboratory of Health Studies | Danilo Franco et al |
| EPI_ISL_496685 | Gorgas Memorial Laboratory of Health Studies | Gorgas Memorial Laboratory of Health Studies | Danilo Franco et al |
| EPI_ISL_496736 | Gorgas Memorial Laboratory of Health Studies | Gorgas Memorial Laboratory of Health Studies | Danilo Franco et al |
| EPI_ISL_496771 | Gorgas Memorial Laboratory of Health Studies | Gorgas Memorial Laboratory of Health Studies | Danilo Franco et al |
| EPI_ISL_496776 | Gorgas Memorial Laboratory of Health Studies | Gorgas Memorial Laboratory of Health Studies | Danilo Franco et al |
| EPI_ISL_496785 | Gorgas Memorial Laboratory of Health Studies | Gorgas Memorial Laboratory of Health Studies | Danilo Franco et al |
| EPI_ISL_496799 | Gorgas Memorial Laboratory of Health Studies | Gorgas Memorial Laboratory of Health Studies | Danilo Franco et al |
| EPI_ISL_496804 | Gorgas Memorial Laboratory of Health Studies | Gorgas Memorial Laboratory of Health Studies | Danilo Franco et al |
| EPI_ISL_496825 | Gorgas Memorial Laboratory of Health Studies | Gorgas Memorial Laboratory of Health Studies | Danilo Franco et al |
| EPI_ISL_496832 | Gorgas Memorial Laboratory of Health Studies | Gorgas Memorial Laboratory of Health Studies | Danilo Franco et al |
| EPI_ISL_496836 | Gorgas Memorial Laboratory of Health Studies | Gorgas Memorial Laboratory of Health Studies | Danilo Franco et al |
| EPI_ISL_613544 | Department of Virus and Microbiological Special Diagnostics, Statens Serum Institut, Denmark | Albertsen lab, Department of Chemistry and Bioscience, Aalborg University, Denmark | Danish Corona Genome Consortia et al |
| EPI_ISL_617943 | Department of Virus and Microbiological Special Diagnostics, Statens Serum Institut, Denmark | Albertsen lab, Department of Chemistry and Bioscience, Aalborg University, Denmark | Danish Covid-19 Genome Consortia et al |
| EPI_ISL_622420 | Department of Virus and Microbiological Special Diagnostics, Statens Serum Institut, Denmark | Albertsen lab, Department of Chemistry and Bioscience, Aalborg University, Denmark | Danish Covid-19 Genome Consortia et al |
| EPI_ISL_618102 | Department of Virus and Microbiological Special Diagnostics, Statens Serum Institut, Denmark | Albertsen lab, Department of Chemistry and Bioscience, Aalborg University, Denmark | Danish Covid-19 Genome Consortia et al |
| EPI_ISL_617982 | Department of Virus and Microbiological Special Diagnostics, Statens Serum Institut, Denmark | Albertsen lab, Department of Chemistry and Bioscience, Aalborg University, Denmark | Danish Covid-19 Genome Consortia et al |
| EPI_ISL_618127 | Department of Virus and Microbiological Special Diagnostics, Statens Serum Institut, Denmark | Albertsen lab, Department of Chemistry and Bioscience, Aalborg University, Denmark | Danish Covid-19 Genome Consortia et al |
| EPI_ISL_618137 | Department of Virus and Microbiological Special Diagnostics, Statens Serum Institut, Denmark | Albertsen lab, Department of Chemistry and Bioscience, Aalborg University, Denmark | Danish Covid-19 Genome Consortia et al |
| EPI_ISL_618138 | Department of Virus and Microbiological Special Diagnostics, Statens Serum Institut, Denmark | Albertsen lab, Department of Chemistry and Bioscience, Aalborg University, Denmark | Danish Covid-19 Genome Consortia et al |
| EPI_ISL_619318 | Department of Virus and Microbiological Special Diagnostics, Statens Serum Institut, Denmark | Albertsen lab, Department of Chemistry and Bioscience, Aalborg University, Denmark | Danish Covid-19 Genome Consortia et al |
| EPI_ISL_617946 | Department of Virus and Microbiological Special Diagnostics, Statens Serum Institut, Denmark | Albertsen lab, Department of Chemistry and Bioscience, Aalborg University, Denmark | Danish Covid-19 Genome Consortia et al |
| EPI_ISL_618212 | Department of Virus and Microbiological Special Diagnostics, Statens Serum Institut, Denmark | Albertsen lab, Department of Chemistry and Bioscience, Aalborg University, Denmark | Danish Covid-19 Genome Consortia et al |
| EPI_ISL_622515 | Department of Virus and Microbiological Special Diagnostics, Statens Serum Institut, Denmark | Albertsen lab, Department of Chemistry and Bioscience, Aalborg University, Denmark | Danish Covid-19 Genome Consortia et al |
| EPI_ISL_622531 | Department of Virus and Microbiological Special Diagnostics, Statens Serum Institut, Denmark | Albertsen lab, Department of Chemistry and Bioscience, Aalborg University, Denmark | Danish Covid-19 Genome Consortia et al |

| | | | |
|---|---|---|---|
| EPI_ISL_618328 | Department of Virus and Microbiological Special Diagnostics, Statens Serum Institut, Denmark | Albertsen lab, Department of Chemistry and Bioscience, Aalborg University, Denmark | Danish Covid-19 Genome Consortia et al |
| EPI_ISL_618323 | Department of Virus and Microbiological Special Diagnostics, Statens Serum Institut, Denmark | Albertsen lab, Department of Chemistry and Bioscience, Aalborg University, Denmark | Danish Covid-19 Genome Consortia et al |
| EPI_ISL_618223 | Department of Virus and Microbiological Special Diagnostics, Statens Serum Institut, Denmark | Albertsen lab, Department of Chemistry and Bioscience, Aalborg University, Denmark | Danish Covid-19 Genome Consortia et al |
| EPI_ISL_619321 | Department of Virus and Microbiological Special Diagnostics, Statens Serum Institut, Denmark | Albertsen lab, Department of Chemistry and Bioscience, Aalborg University, Denmark | Danish Covid-19 Genome Consortia et al |
| EPI_ISL_617658 | Department of Virus and Microbiological Special Diagnostics, Statens Serum Institut, Denmark | Albertsen lab, Department of Chemistry and Bioscience, Aalborg University, Denmark | Danish Covid-19 Genome Consortia et al |
| EPI_ISL_617424 | Department of Virus and Microbiological Special Diagnostics, Statens Serum Institut, Denmark | Albertsen lab, Department of Chemistry and Bioscience, Aalborg University, Denmark | Danish Covid-19 Genome Consortia et al |
| EPI_ISL_616408 | Department of Virus and Microbiological Special Diagnostics, Statens Serum Institut, Denmark | Albertsen lab, Department of Chemistry and Bioscience, Aalborg University, Denmark | Danish Covid-19 Genome Consortia et al |
| EPI_ISL_622643 | Department of Virus and Microbiological Special Diagnostics, Statens Serum Institut, Denmark | Albertsen lab, Department of Chemistry and Bioscience, Aalborg University, Denmark | Danish Covid-19 Genome Consortia et al |
| EPI_ISL_622733 | Department of Virus and Microbiological Special Diagnostics, Statens Serum Institut, Denmark | Albertsen lab, Department of Chemistry and Bioscience, Aalborg University, Denmark | Danish Covid-19 Genome Consortia et al |
| EPI_ISL_622356 | Department of Virus and Microbiological Special Diagnostics, Statens Serum Institut, Denmark | Albertsen lab, Department of Chemistry and Bioscience, Aalborg University, Denmark | Danish Covid-19 Genome Consortia et al |
| EPI_ISL_619715 | Department of Virus and Microbiological Special Diagnostics, Statens Serum Institut, Denmark | Albertsen lab, Department of Chemistry and Bioscience, Aalborg University, Denmark | Danish Covid-19 Genome Consortia et al |
| EPI_ISL_619846 | Department of Virus and Microbiological Special Diagnostics, Statens Serum Institut, Denmark | Albertsen lab, Department of Chemistry and Bioscience, Aalborg University, Denmark | Danish Covid-19 Genome Consortia et al |
| EPI_ISL_619301 | Department of Virus and Microbiological Special Diagnostics, Statens Serum Institut, Denmark | Albertsen lab, Department of Chemistry and Bioscience, Aalborg University, Denmark | Danish Covid-19 Genome Consortia et al |
| EPI_ISL_619291 | Department of Virus and Microbiological Special Diagnostics, Statens Serum Institut, Denmark | Albertsen lab, Department of Chemistry and Bioscience, Aalborg University, Denmark | Danish Covid-19 Genome Consortia et al |
| EPI_ISL_619393 | Department of Virus and Microbiological Special Diagnostics, Statens Serum Institut, Denmark | Albertsen lab, Department of Chemistry and Bioscience, Aalborg University, Denmark | Danish Covid-19 Genome Consortia et al |
| EPI_ISL_614421 | Department of Virus and Microbiological Special Diagnostics, Statens Serum Institut, Denmark | Albertsen lab, Department of Chemistry and Bioscience, Aalborg University, Denmark | Danish Covid-19 Genome Consortia et al |
| EPI_ISL_614754 | Department of Virus and Microbiological Special Diagnostics, Statens Serum Institut, Denmark | Albertsen lab, Department of Chemistry and Bioscience, Aalborg University, Denmark | Danish Covid-19 Genome Consortia et al |
| EPI_ISL_614815 | Department of Virus and Microbiological Special Diagnostics, Statens Serum Institut, Denmark | Albertsen lab, Department of Chemistry and Bioscience, Aalborg University, Denmark | Danish Covid-19 Genome Consortia et al |
| EPI_ISL_614799 | Department of Virus and Microbiological Special Diagnostics, Statens Serum Institut, Denmark | Albertsen lab, Department of Chemistry and Bioscience, Aalborg University, Denmark | Danish Covid-19 Genome Consortia et al |
| EPI_ISL_614426 | Department of Virus and Microbiological Special Diagnostics, Statens Serum Institut, Denmark | Albertsen lab, Department of Chemistry and Bioscience, Aalborg University, Denmark | Danish Covid-19 Genome Consortia et al |
| EPI_ISL_614424 | Department of Virus and Microbiological Special Diagnostics, Statens Serum Institut, Denmark | Albertsen lab, Department of Chemistry and Bioscience, Aalborg University, Denmark | Danish Covid-19 Genome Consortia et al |
| EPI_ISL_614753 | Department of Virus and Microbiological Special Diagnostics, Statens Serum Institut, Denmark | Albertsen lab, Department of Chemistry and Bioscience, Aalborg University, Denmark | Danish Covid-19 Genome Consortia et al |

| | | | |
|---|---|---|---|
| EPI_ISL_614411 | Department of Virus and Microbiological Special Diagnostics, Statens Serum Institut, Denmark | Albertsen lab, Department of Chemistry and Bioscience, Aalborg University, Denmark | Danish Covid-19 Genome Consortia et al |
| EPI_ISL_614802 | Department of Virus and Microbiological Special Diagnostics, Statens Serum Institut, Denmark | Albertsen lab, Department of Chemistry and Bioscience, Aalborg University, Denmark | Danish Covid-19 Genome Consortia et al |
| EPI_ISL_670281 | Department of Virus and Microbiological Special Diagnostics, Statens Serum Institut, Copenhagen, Denmark | Albertsen Lab, Department of Chemistry and Bioscience, Aalborg University, Denmark | Danish Covid-19 Genome Consortium et al |
| EPI_ISL_670299 | Department of Virus and Microbiological Special Diagnostics, Statens Serum Institut, Copenhagen, Denmark | Albertsen Lab, Department of Chemistry and Bioscience, Aalborg University, Denmark | Danish Covid-19 Genome Consortium et al |
| EPI_ISL_682605 | Department of Virus and Microbiological Special Diagnostics, Statens Serum Institut, Copenhagen, Denmark | Albertsen Lab, Department of Chemistry and Bioscience, Aalborg University, Denmark | Danish Covid-19 Genome Consortium et al |
| EPI_ISL_845385 | Department of Virus and Microbiological Special Diagnostics, Statens Serum Institut, Copenhagen, Denmark | Albertsen Lab, Department of Chemistry and Bioscience, Aalborg University, Denmark | Danish Covid-19 Genome Consortium et al |
| EPI_ISL_847449 | Department of Virus and Microbiological Special Diagnostics, Statens Serum Institut, Copenhagen, Denmark | Albertsen Lab, Department of Chemistry and Bioscience, Aalborg University, Denmark | Danish Covid-19 Genome Consortium et al |
| EPI_ISL_926776 | Department of Virus and Microbiological Special Diagnostics, Statens Serum Institut, Copenhagen, Denmark | Aalborg University | Danish Covid-19 Genome Consortium et al |
| EPI_ISL_927497 | Department of Virus and Microbiological Special Diagnostics, Statens Serum Institut, Copenhagen, Denmark | Aalborg University | Danish Covid-19 Genome Consortium et al |
| EPI_ISL_971627 | Department of Virus and Microbiological Special Diagnostics, Statens Serum Institut, Copenhagen, Denmark | Aalborg University | Danish Covid-19 Genome Consortium et al |
| EPI_ISL_973071 | Department of Virus and Microbiological Special Diagnostics, Statens Serum Institut, Copenhagen, Denmark | Aalborg University | Danish Covid-19 Genome Consortium et al |
| EPI_ISL_973216 | Department of Virus and Microbiological Special Diagnostics, Statens Serum Institut, Copenhagen, Denmark | Aalborg University | Danish Covid-19 Genome Consortium et al |
| EPI_ISL_1023986 | Department of Virus and Microbiological Special Diagnostics, Statens Serum Institut, Copenhagen, Denmark | Aalborg University | Danish Covid-19 Genome Consortium et al |
| EPI_ISL_1024655 | Department of Virus and Microbiological Special Diagnostics, Statens Serum Institut, Copenhagen, Denmark | Aalborg University | Danish Covid-19 Genome Consortium et al |
| EPI_ISL_1065177 | Department of Virus and Microbiological Special Diagnostics, Statens Serum Institut, Copenhagen, Denmark | Aalborg University | Danish Covid-19 Genome Consortium et al |
| EPI_ISL_1066314 | Department of Virus and Microbiological Special Diagnostics, Statens Serum Institut, Copenhagen, Denmark | Aalborg University | Danish Covid-19 Genome Consortium et al |
| EPI_ISL_654194 | Hospital General Universitario Gregorio Marañón | SeqCOVID-SPAIN consortium/IBV(CSIC) | Darío García de Viedma et al |
| EPI_ISL_654170 | Hospital General Universitario Gregorio Marañón | SeqCOVID-SPAIN consortium/IBV(CSIC) | Darío García de Viedma et al |
| EPI_ISL_813997 | Hospital General Universitario Gregorio Marañón | SeqCOVID-SPAIN consortium/IBV(CSIC) | Darío García de Viedma et al |
| EPI_ISL_852976 | Hospital General Universitario Gregorio Marañón | SeqCOVID-SPAIN consortium/IBV(CSIC) | Darío García de Viedma et al |
| EPI_ISL_610275 | New Mexico Department of Health Scientific Laboratory Division | Center for Global Health, University of New Mexico Health Sciences Center | Daryl Domman et al |
| EPI_ISL_610278 | New Mexico Department of Health Scientific Laboratory Division | Center for Global Health, University of New Mexico Health Sciences Center | Daryl Domman et al |
| EPI_ISL_1181310 | MVZ Dr. Eberhard & Partner Dortmund | Bielefeld University | David Brandt et al |
| EPI_ISL_654496 | Servicio de Microbiología. Hospital Clínico Universitario de Valencia | SeqCOVID-SPAIN consortium/IBV(CSIC) | David Navarro Ortega et al |

| | | | |
|---|---|---|---|
| EPI_ISL_413593 | Laboratoire National de Santé | Erasmus Medical Center | David Nieuwenhuijse et al |
| EPI_ISL_413570 | RIVM | Erasmus Medical Center | David Nieuwenhuijse et al |
| EPI_ISL_413574 | MHC West-Brabant | Erasmus Medical Center | David Nieuwenhuijse et al |
| EPI_ISL_414546 | Dutch COVID-19 response team | Erasmus Medical Center | David Nieuwenhuijse et al |
| EPI_ISL_413586 | Foundation Elisabeth-Tweesteden Ziekenhuis | Erasmus Medical Center | David Nieuwenhuijse et al |
| EPI_ISL_718146 | Ministry of Health Hospitals | Institute of Health and Community Medicine | David Perera et al |
| EPI_ISL_718143 | Ministry of Health Hospitals | Institute of Health and Community Medicine | David Perera et al |
| EPI_ISL_718214 | Ministry of Health Hospitals | Institute of Health and Community Medicine | David Perera et al |
| EPI_ISL_718213 | Ministry of Health Hospitals | Institute of Health and Community Medicine | David Perera et al |
| EPI_ISL_718168 | Ministry of Health Hospitals | Institute of Health and Community Medicine | David Perera et al |
| EPI_ISL_718169 | Ministry of Health Hospitals | Institute of Health and Community Medicine | David Perera et al |
| EPI_ISL_718170 | Ministry of Health Hospitals | Institute of Health and Community Medicine | David Perera et al |
| EPI_ISL_804016 | Facultad de Medicina, Universidad de Atacama | Facultad de Ciencias de la Vida, UNAB | Dayán Sanhueza et al |
| EPI_ISL_676519 | Klinsisk mikrobiologi Linkoping | The Public Health Agency of Sweden | Department of Microbiology et al |
| EPI_ISL_676520 | Klinisk mikrobiologi | The Public Health Agency of Sweden | Department of Microbiology et al |
| EPI_ISL_676528 | Uppsala klinisk mikrobiologi | The Public Health Agency of Sweden | Department of Microbiology et al |
| EPI_ISL_710577 | Vardcentralen Brinken | The Public Health Agency of Sweden | Department of Microbiology et al |
| EPI_ISL_766625 | Klinisk mikrobiologi | The Public Health Agency of Sweden | Department of Microbiology et al |
| EPI_ISL_766628 | Klinisk mikrobiologi | The Public Health Agency of Sweden | Department of Microbiology et al |
| EPI_ISL_491933 | Centro de Investigaciones, Universidad de Especialidades Espíritu Santo | Institute of Microbiology, Universidad San Francisco de Quito | Derly Andrade et al |
| EPI_ISL_491935 | Centro de Investigaciones, Universidad de Especialidades Espíritu Santo | Institute of Microbiology, Universidad San Francisco de Quito | Derly Andrade et al |
| EPI_ISL_491941 | Centro de Investigaciones, Universidad de Especialidades Espíritu Santo | Institute of Microbiology, Universidad San Francisco de Quito | Derly Andrade et al |
| EPI_ISL_527819 | Centro de Investigaciones, Universidad de Especialidades Espíritu Santo | Institute of Microbiology, Universidad San Francisco de Quito | Derly Andrade et al |
| EPI_ISL_1381304 | Omics Sciences Laboratory | Omics Sciences Laboratory | Derly Andrade Molina et al |
| EPI_ISL_1381303 | Omics Sciences Laboratory | Omics Sciences Laboratory | Derly Andrade Molina et al |
| EPI_ISL_1443672 | Omics Sciences Laboratory | Omics Sciences Laboratory | Derly Andrade Molina et al |
| EPI_ISL_1443676 | Omics Sciences Laboratory | Omics Sciences Laboratory | Derly Andrade Molina et al |
| EPI_ISL_1381300 | Omics Sciences Laboratory | Omics Sciences Laboratory | Derly Andrade Molina et al |
| EPI_ISL_1381301 | Omics Sciences Laboratory | Omics Sciences Laboratory | Derly Andrade Molina et al |
| EPI_ISL_1443680 | Omics Sciences Laboratory | Omics Sciences Laboratory | Derly Andrade Molina et al |

| | | | |
|---|---|---|---|
| EPI_ISL_1381305 | Omics Sciences Laboratory | Omics Sciences Laboratory | Derly Andrade Molina et al |
| EPI_ISL_1381302 | Omics Sciences Laboratory | Omics Sciences Laboratory | Derly Andrade Molina et al |
| EPI_ISL_1443681 | Omics Sciences Laboratory | Omics Sciences Laboratory | Derly Andrade Molina et al |
| EPI_ISL_1443682 | Omics Sciences Laboratory | Omics Sciences Laboratory | Derly Andrade Molina et al |
| EPI_ISL_1443641 | Institute of Microbiology, Universidad San Francisco de Quito | Omics Sciences Laboratory | Derly Andrade Molina et al |
| EPI_ISL_1443642 | Institute of Microbiology, Universidad San Francisco de Quito | Omics Sciences Laboratory | Derly Andrade Molina et al |
| EPI_ISL_1443643 | Institute of Microbiology, Universidad San Francisco de Quito | Omics Sciences Laboratory | Derly Andrade Molina et al |
| EPI_ISL_1443650 | Institute of Microbiology, Universidad San Francisco de Quito | Omics Sciences Laboratory | Derly Andrade Molina et al |
| EPI_ISL_1443651 | Institute of Microbiology, Universidad San Francisco de Quito | Omics Sciences Laboratory | Derly Andrade Molina et al |
| EPI_ISL_1443652 | Institute of Microbiology, Universidad San Francisco de Quito | Omics Sciences Laboratory | Derly Andrade Molina et al |
| EPI_ISL_1443656 | Institute of Microbiology, Universidad San Francisco de Quito | Omics Sciences Laboratory | Derly Andrade Molina et al |
| EPI_ISL_1443658 | Institute of Microbiology, Universidad San Francisco de Quito | Omics Sciences Laboratory | Derly Andrade Molina et al |
| EPI_ISL_1443664 | Institute of Microbiology, Universidad San Francisco de Quito | Omics Sciences Laboratory | Derly Andrade Molina et al |
| EPI_ISL_1443665 | Institute of Microbiology, Universidad San Francisco de Quito | Omics Sciences Laboratory | Derly Andrade Molina et al |
| EPI_ISL_861914 | Genomika Einstein | LATE - Laboratório de Técnicas Especiais - Hospital Israelita Albert Einstein | Deyvid Amgarten et al |
| EPI_ISL_861868 | LATE - Laboratório de Técnicas Especiais - Hospital Israelita Albert Einstein | LATE - Laboratório de Técnicas Especiais - Hospital Israelita Albert Einstein | Deyvid Amgarten et al |
| EPI_ISL_861870 | LATE - Laboratório de Técnicas Especiais - Hospital Israelita Albert Einstein | LATE - Laboratório de Técnicas Especiais - Hospital Israelita Albert Einstein | Deyvid Amgarten et al |
| EPI_ISL_861871 | LATE - Laboratório de Técnicas Especiais - Hospital Israelita Albert Einstein | LATE - Laboratório de Técnicas Especiais - Hospital Israelita Albert Einstein | Deyvid Amgarten et al |
| EPI_ISL_861873 | LATE - Laboratório de Técnicas Especiais - Hospital Israelita Albert Einstein | LATE - Laboratório de Técnicas Especiais - Hospital Israelita Albert Einstein | Deyvid Amgarten et al |
| EPI_ISL_861875 | LATE - Laboratório de Técnicas Especiais - Hospital Israelita Albert Einstein | LATE - Laboratório de Técnicas Especiais - Hospital Israelita Albert Einstein | Deyvid Amgarten et al |
| EPI_ISL_861876 | LATE - Laboratório de Técnicas Especiais - Hospital Israelita Albert Einstein | LATE - Laboratório de Técnicas Especiais - Hospital Israelita Albert Einstein | Deyvid Amgarten et al |
| EPI_ISL_861878 | LATE - Laboratório de Técnicas Especiais - Hospital Israelita Albert Einstein | LATE - Laboratório de Técnicas Especiais - Hospital Israelita Albert Einstein | Deyvid Amgarten et al |
| EPI_ISL_861881 | LATE - Laboratório de Técnicas Especiais - Hospital Israelita Albert Einstein | LATE - Laboratório de Técnicas Especiais - Hospital Israelita Albert Einstein | Deyvid Amgarten et al |
| EPI_ISL_861884 | LATE - Laboratório de Técnicas Especiais - Hospital Israelita Albert Einstein | LATE - Laboratório de Técnicas Especiais - Hospital Israelita Albert Einstein | Deyvid Amgarten et al |
| EPI_ISL_861885 | LATE - Laboratório de Técnicas Especiais - Hospital Israelita Albert Einstein | LATE - Laboratório de Técnicas Especiais - Hospital Israelita Albert Einstein | Deyvid Amgarten et al |
| EPI_ISL_861886 | LATE - Laboratório de Técnicas Especiais - Hospital Israelita Albert Einstein | LATE - Laboratório de Técnicas Especiais - Hospital Israelita Albert Einstein | Deyvid Amgarten et al |

| | | | |
|---|---|---|---|
| EPI_ISL_861888 | LATE - Laboratório de Técnicas Especiais - Hospital Israelita Albert Einstein | LATE - Laboratório de Técnicas Especiais - Hospital Israelita Albert Einstein | Deyvid Amgarten et al |
| EPI_ISL_861891 | LATE - Laboratório de Técnicas Especiais - Hospital Israelita Albert Einstein | LATE - Laboratório de Técnicas Especiais - Hospital Israelita Albert Einstein | Deyvid Amgarten et al |
| EPI_ISL_861892 | LATE - Laboratório de Técnicas Especiais - Hospital Israelita Albert Einstein | LATE - Laboratório de Técnicas Especiais - Hospital Israelita Albert Einstein | Deyvid Amgarten et al |
| EPI_ISL_884250 | LATE - Laboratório de Técnicas Especiais - Hospital Israelita Albert Einstein | LATE - Laboratório de Técnicas Especiais - Hospital Israelita Albert Einstein | Deyvid Amgarten et al |
| EPI_ISL_861895 | LATE - Laboratório de Técnicas Especiais - Hospital Israelita Albert Einstein | LATE - Laboratório de Técnicas Especiais - Hospital Israelita Albert Einstein | Deyvid Amgarten et al |
| EPI_ISL_861896 | LATE - Laboratório de Técnicas Especiais - Hospital Israelita Albert Einstein | LATE - Laboratório de Técnicas Especiais - Hospital Israelita Albert Einstein | Deyvid Amgarten et al |
| EPI_ISL_861897 | LATE - Laboratório de Técnicas Especiais - Hospital Israelita Albert Einstein | LATE - Laboratório de Técnicas Especiais - Hospital Israelita Albert Einstein | Deyvid Amgarten et al |
| EPI_ISL_861901 | LATE - Laboratório de Técnicas Especiais - Hospital Israelita Albert Einstein | LATE - Laboratório de Técnicas Especiais - Hospital Israelita Albert Einstein | Deyvid Amgarten et al |
| EPI_ISL_861902 | LATE - Laboratório de Técnicas Especiais - Hospital Israelita Albert Einstein | LATE - Laboratório de Técnicas Especiais - Hospital Israelita Albert Einstein | Deyvid Amgarten et al |
| EPI_ISL_861903 | LATE - Laboratório de Técnicas Especiais - Hospital Israelita Albert Einstein | LATE - Laboratório de Técnicas Especiais - Hospital Israelita Albert Einstein | Deyvid Amgarten et al |
| EPI_ISL_861905 | LATE - Laboratório de Técnicas Especiais - Hospital Israelita Albert Einstein | LATE - Laboratório de Técnicas Especiais - Hospital Israelita Albert Einstein | Deyvid Amgarten et al |
| EPI_ISL_861908 | LATE - Laboratório de Técnicas Especiais - Hospital Israelita Albert Einstein | LATE - Laboratório de Técnicas Especiais - Hospital Israelita Albert Einstein | Deyvid Amgarten et al |
| EPI_ISL_861909 | LATE - Laboratório de Técnicas Especiais - Hospital Israelita Albert Einstein | LATE - Laboratório de Técnicas Especiais - Hospital Israelita Albert Einstein | Deyvid Amgarten et al |
| EPI_ISL_1544070 | National Centre For Cell Science | National Centre For Cell Science – INSACOG | Dhiraj Paul et al |
| EPI_ISL_1544073 | National Centre For Cell Science | National Centre For Cell Science – INSACOG | Dhiraj Paul et al |
| EPI_ISL_1415196 | National Centre For Cell Science | National Centre For Cell Science – INSACOG | Dhiraj Paul et al |
| EPI_ISL_812967 | University Clinical Research Center, University of Sciences | University Clinical Research Center, University of Sciences | Diarra et al |
| EPI_ISL_812966 | University Clinical Research Center, University of Sciences | University Clinical Research Center, University of Sciences | Diarra et al |
| EPI_ISL_525438 | Institute of Microbiology, Universidad San Francisco de Quito | Institute of Microbiology, Universidad San Francisco de Quito | Diego Egas et al |
| EPI_ISL_1445093 | SECRETARIA MUNICIPAL DE SAUDE DE PIRACAIA | Instituto Butantan / Mendelics | Dimas Tadeu Covas et al |
| EPI_ISL_1445095 | SECRETARIA MUNICIPAL DE SAUDE DE PIRACAIA | Instituto Butantan / Mendelics | Dimas Tadeu Covas et al |
| EPI_ISL_1445102 | PRONTO ATENDIMENTO VILA PADRE ANCHIETA | Instituto Butantan / Mendelics | Dimas Tadeu Covas et al |
| EPI_ISL_1445104 | PRONTO ATENDIMENTO VILA PADRE ANCHIETA | Instituto Butantan / Mendelics | Dimas Tadeu Covas et al |
| EPI_ISL_1445129 | UNIDADE RESPIRATORIA NOVA HORTOLANDIA | Instituto Butantan / Mendelics | Dimas Tadeu Covas et al |
| EPI_ISL_1445136 | SMS IPERO | Instituto Butantan / Mendelics | Dimas Tadeu Covas et al |

| | | | |
|---|---|---|---|
| EPI_ISL_1445138 | SECRETARIA MUNICIPAL DE SAUDE SOROCABA | Instituto Butantan / Mendelics | Dimas Tadeu Covas et al |
| EPI_ISL_1445142 | CS II EGIDIO BRUNHARA MORRO AGUDO | Instituto Butantan / Mendelics | Dimas Tadeu Covas et al |
| EPI_ISL_1445157 | SECAO CENTRO DE DIAGNOSTICO SECEDI | Instituto Butantan / Mendelics | Dimas Tadeu Covas et al |
| EPI_ISL_1445159 | SECAO CENTRO DE DIAGNOSTICO SECEDI | Instituto Butantan / Mendelics | Dimas Tadeu Covas et al |
| EPI_ISL_1445212 | UBS IRMA AGUEDA MARIA JAIME | Instituto Butantan / Mendelics | Dimas Tadeu Covas et al |
| EPI_ISL_1445213 | UBS OTACILIO FIRMINO LOPES | Instituto Butantan / Mendelics | Dimas Tadeu Covas et al |
| EPI_ISL_1445214 | UBS JOSE HILARIO DOS SANTOS | Instituto Butantan / Mendelics | Dimas Tadeu Covas et al |
| EPI_ISL_1445217 | UBS EMILIA COSME CERQUEIRA | Instituto Butantan / Mendelics | Dimas Tadeu Covas et al |
| EPI_ISL_1445231 | VIGILANCIA EPIDEMIOLOGICA | Instituto Butantan / Mendelics | Dimas Tadeu Covas et al |
| EPI_ISL_1445232 | VIGILANCIA EPIDEMIOLOGICA | Instituto Butantan / Mendelics | Dimas Tadeu Covas et al |
| EPI_ISL_1445250 | VIGILANCIA EPIDEMIOLOGICA | Instituto Butantan / Mendelics | Dimas Tadeu Covas et al |
| EPI_ISL_1445255 | VIGILANCIA EPIDEMIOLOGICA | Instituto Butantan / Mendelics | Dimas Tadeu Covas et al |
| EPI_ISL_1445260 | VIGILANCIA EPIDEMIOLOGICA | Instituto Butantan / Mendelics | Dimas Tadeu Covas et al |
| EPI_ISL_1445261 | VIGILANCIA EPIDEMIOLOGICA | Instituto Butantan / Mendelics | Dimas Tadeu Covas et al |
| EPI_ISL_1445269 | SERV DE VIG SANITARIA EPIDEMIO E CTRL DE ZOONOZES GUARUJA | Instituto Butantan / Mendelics | Dimas Tadeu Covas et al |
| EPI_ISL_1445271 | VIGILANCIA EPIDEMIOLOGICA | Instituto Butantan / Mendelics | Dimas Tadeu Covas et al |
| EPI_ISL_885143 | Central public health laboratory | Molecular Diagnostics Department, Central public health laboratory | Dler et al |
| EPI_ISL_412028 | Hong Kong Department of Health | School of Public Health, The University of Hon g Kong | Dominic N.C. Tsang et al |
| EPI_ISL_572329 | Public Health Authority of the Slovak Republic, Bratislava | Faculty of Natural Sciences, Comenius University in Bratislava | Dominika Fričová et al |
| EPI_ISL_737200 | National Reference Laboratory, Nigeria Centre for Disease Control. | National Reference Laboratory, Nigeria Centre for Disease Control, Gaduwa,Abuja, Nigeria | Dr Ndodo Nnaemeka et al |
| EPI_ISL_825807 | Nigeria Centre For Disease Control | National reference Laboratory, NCDC, Gaduwa, Abuja | Dr Ndodo Nnaemeka et al |
| EPI_ISL_1073640 | Nigeria Centre For Disease Control | National reference Laboratory, NCDC, Gaduwa, Abuja | Dr Ndodo Nnaemeka et al |
| EPI_ISL_1627088 | National reference Laboratory, NCDC, Gaduwa, Abuja, Nigeria | National reference Laboratory, NCDC, Gaduwa, Abuja, Nigeria | Dr Ndodo Nnaemeka et al |
| EPI_ISL_1197066 | Nigeria Centre For Disease Control | National reference Laboratory, NCDC, Gaduwa, Abuja | Dr Ndodo Nnaemeka et al |
| EPI_ISL_848196 | Microbiology Lab | NBCC Sequencing Facility | Dr. Jeff Wrana et al |
| EPI_ISL_873164 | Medical Laboratory Sciences, Arab American University | Medical Laboratory Sciences, Arab American University | Dumaidi et al |
| EPI_ISL_873165 | Medical Laboratory Sciences, Arab American University | Medical Laboratory Sciences, Arab American University | Dumaidi et al |
| EPI_ISL_792516 | Laboratorio Central de la Ciudad de Santa Fe | Grupo de Genómica y Bioinformática del Instituto de Investigación de la Cadena Láctea CONICET-INTA on behalf of 'Proyecto Argentino Interinstitucional de genomica de SARS-CoV-2' (PAIS Consortium) | Eberhardt et al |

| | | | |
|---|---|---|---|
| EPI_ISL_792517 | Laboratorio Central de la Ciudad de Santa Fe | Grupo de Genómica y Bioinformática del Instituto de Investigación de la Cadena Láctea CONICET-INTA on behalf of 'Proyecto Argentino Interinstitucional de genomica de SARS-CoV-2' (PAIS Consortium) | Eberhardt et al |
| EPI_ISL_1395880 | Laboratorio Central de la Ciudad de Santa Fe | Grupo de Genómica y Bioinformática del Instituto de Investigación de la Cadena Láctea CONICET-INTA on behalf of 'Proyecto Argentino Interinstitucional de genomica de SARS-CoV-2' (PAIS Consortium) | Eberhardt et al |
| EPI_ISL_1395886 | Laboratorio Central de la Ciudad de Santa Fe | Grupo de Genómica y Bioinformática del Instituto de Investigación de la Cadena Láctea CONICET-INTA on behalf of 'Proyecto Argentino Interinstitucional de genomica de SARS-CoV-2' (PAIS Consortium) | Eberhardt et al |
| EPI_ISL_626550 | Laboratorio de Biología Molecular, Facultad de Medicina, Universidad de Atacama, Copiapo, Chile/ FONDAP CRG, Universidad Andrés Bello, Santiago, Chile | Center for Mathematical Modeling and Center for Genome Regulation. Santiago, Chile | Echeverría C et al |
| EPI_ISL_626552 | Laboratorio de Biología Molecular, Facultad de Medicina, Universidad de Atacama, Copiapo, Chile/ FONDAP CRG, Universidad Andrés Bello, Santiago, Chile | Center for Mathematical Modeling and Center for Genome Regulation. Santiago, Chile | Echeverría C et al |
| EPI_ISL_417030 | Centre for Infectious Diseases and Microbiology Laboratory Services | NSW Health Pathology - Institute of Clinical Pathology and Medical Research; Westmead Hospital; University of Sydney | Eden J-S et al |
| EPI_ISL_413213 | Centre for Infectious Diseases and Microbiology Laboratory Services | NSW Health Pathology - Institute of Clinical Pathology and Medical Research; Westmead Hospital; University of Sydney | Eden J-S et al |
| EPI_ISL_889008 | RS Sentra Medika Cibinong | Eijkman Institute for Molecular Biology, Ministry of Research and Technology/National Agency for Research and Innovation | Edison Johar et al |
| EPI_ISL_947249 | RS Bina Husada Cibinong | Eijkman Institute for Molecular Biology, Ministry of Research and Technology/National Agency for Research and Innovation | Edison Johar et al |
| EPI_ISL_947252 | RS Bina Husada Cibinong | Eijkman Institute for Molecular Biology, Ministry of Research and Technology/National Agency for Research and Innovation | Edison Johar et al |
| EPI_ISL_536479 | Instituto Nacional de Salud | Laboratorio de Infecciones Respiratorias Agudas | Eduardo Juscamayta Lopez et al |
| EPI_ISL_536506 | Instituto Nacional de Salud | Laboratorio de Infecciones Respiratorias Agudas | Eduardo Juscamayta Lopez et al |
| EPI_ISL_536526 | Instituto Nacional de Salud | Laboratorio de Infecciones Respiratorias Agudas | Eduardo Juscamayta Lopez et al |
| EPI_ISL_536564 | Instituto Nacional de Salud | Laboratorio de Infecciones Respiratorias Agudas | Eduardo Juscamayta Lopez et al |
| EPI_ISL_536485 | Instituto Nacional de Salud | Laboratorio de Infecciones Respiratorias Agudas | Eduardo Juscamayta Lopez et al |
| EPI_ISL_536489 | Instituto Nacional de Salud | Laboratorio de Infecciones Respiratorias Agudas | Eduardo Juscamayta Lopez et al |
| EPI_ISL_536490 | Instituto Nacional de Salud | Laboratorio de Infecciones Respiratorias Agudas | Eduardo Juscamayta Lopez et al |
| EPI_ISL_536497 | Instituto Nacional de Salud | Laboratorio de Infecciones Respiratorias Agudas | Eduardo Juscamayta Lopez et al |
| EPI_ISL_536500 | Instituto Nacional de Salud | Laboratorio de Infecciones Respiratorias Agudas | Eduardo Juscamayta Lopez et al |
| EPI_ISL_536510 | Instituto Nacional de Salud | Laboratorio de Infecciones Respiratorias Agudas | Eduardo Juscamayta Lopez et al |
| EPI_ISL_536511 | Instituto Nacional de Salud | Laboratorio de Infecciones Respiratorias Agudas | Eduardo Juscamayta Lopez et al |
| EPI_ISL_536512 | Instituto Nacional de Salud | Laboratorio de Infecciones Respiratorias Agudas | Eduardo Juscamayta Lopez et al |
| EPI_ISL_536521 | Instituto Nacional de Salud | Laboratorio de Infecciones Respiratorias Agudas | Eduardo Juscamayta Lopez et al |
| EPI_ISL_536535 | Instituto Nacional de Salud | Laboratorio de Infecciones Respiratorias Agudas | Eduardo Juscamayta Lopez et al |

| | | | |
|---|---|---|---|
| EPI_ISL_536539 | Instituto Nacional de Salud | Laboratorio de Infecciones Respiratorias Agudas | Eduardo Juscamayta Lopez et al |
| EPI_ISL_536540 | Instituto Nacional de Salud | Laboratorio de Infecciones Respiratorias Agudas | Eduardo Juscamayta Lopez et al |
| EPI_ISL_536541 | Instituto Nacional de Salud | Laboratorio de Infecciones Respiratorias Agudas | Eduardo Juscamayta Lopez et al |
| EPI_ISL_536561 | Instituto Nacional de Salud | Laboratorio de Infecciones Respiratorias Agudas | Eduardo Juscamayta Lopez et al |
| EPI_ISL_794606 | Central Public Health Laboratories, Egyptian Ministry of Health and Population | Central Public Health Laboratories, Egyptian Ministry of Health and Population | El-Shesheny et al |
| EPI_ISL_1138899 | Laboratory for HIV and opportunistic infections diagnosis The Republican Research and Practical Center for Epidemiology and Microbiology (RRPCEM) | Laboratory for HIV and opportunistic infections diagnosis The Republican Research and Practical Center for Epidemiology and Microbiology (RRPCEM) | Elena Gasich et al |
| EPI_ISL_933533 | Laboratory for HIV and opportunistic infections diagnosis The Republican Research and Practical Center for Epidemiology and Microbiology (RRPCEM) | Laboratory for HIV and opportunistic infections diagnosis The Republican Research and Practical Center for Epidemiology and Microbiology (RRPCEM) | Elena Gasich et al |
| EPI_ISL_754229 | The Republican Research and Practical Center for Epidemiology and Microbiology (RRPCEM) | WHO National Influenza Centre Russian Federation | Elena Gasich et al |
| EPI_ISL_754230 | The Republican Research and Practical Center for Epidemiology and Microbiology (RRPCEM) | WHO National Influenza Centre Russian Federation | Elena Gasich et al |
| EPI_ISL_754231 | The Republican Research and Practical Center for Epidemiology and Microbiology (RRPCEM) | WHO National Influenza Centre Russian Federation | Elena Gasich et al |
| EPI_ISL_1209407 | Laboratory for HIV and opportunistic infections diagnosis The Republican Research and Practical Center for Epidemiology and Microbiology (RRPCEM) | Laboratory for HIV and opportunistic infections diagnosis The Republican Research and Practical Center for Epidemiology and Microbiology (RRPCEM) | Elena Gasich et al |
| EPI_ISL_1138983 | Laboratory for HIV and opportunistic infections diagnosis The Republican Research and Practical Center for Epidemiology and Microbiology (RRPCEM) | Laboratory for HIV and opportunistic infections diagnosis The Republican Research and Practical Center for Epidemiology and Microbiology (RRPCEM) | Elena Gasich et al |
| EPI_ISL_1508895 | Laboratory for HIV and opportunistic infections diagnosis The Republican Research and Practical Center for Epidemiology and Microbiology (RRPCEM) | Laboratory for HIV and opportunistic infections diagnosis The Republican Research and Practical Center for Epidemiology and Microbiology (RRPCEM) | Elena Gasich et al |
| EPI_ISL_754234 | The Republican Research and Practical Center for Epidemiology and Microbiology (RRPCEM) | WHO National Influenza Centre Russian Federation | Elena Gasich et al |
| EPI_ISL_1181779 | Microbiology and Virology Unit,Azienda Ospedale Padova,Padova,Italy | Department of Molecular Medicine,Computational Medicine Group, Univeresity of Padova,Padova,Italy | Elisa Franchin et al |
| EPI_ISL_1181797 | Microbiology and Virology Unit,Azienda Ospedale Padova,Padova,Italy | Department of Molecular Medicine,Computational Medicine Group, Univeresity of Padova,Padova,Italy | Elisa Franchin et al |
| EPI_ISL_1181767 | Microbiology and Virology Unit,Azienda Ospedale Padova,Padova,Italy | Department of Molecular Medicine,Computational Medicine Group, Univeresity of Padova,Padova,Italy | Elisa Franchin et al |
| EPI_ISL_1181776 | Microbiology and Virology Unit,Azienda Ospedale Padova,Padova,Italy | Department of Molecular Medicine,Computational Medicine Group, Univeresity of Padova,Padova,Italy | Elisa Franchin et al |
| EPI_ISL_1181804 | Microbiology and Virology Unit,Azienda Ospedale Padova,Padova,Italy | Department of Molecular Medicine,Computational Medicine Group, Univeresity of Padova,Padova,Italy | Elisa Franchin et al |
| EPI_ISL_1064084 | Microbiology and Virology Unit, Azienda Ospedale Padova, Padova, Italy | Department of Molecular Medicine, Computational Medicine Group, Univeresity of Padova, Padova, Italy | Elisa Franchin et al |
| EPI_ISL_1064098 | Microbiology and Virology Unit, Azienda Ospedale Padova, Padova, Italy | Department of Molecular Medicine, Computational Medicine Group, Univeresity of Padova, Padova, Italy | Elisa Franchin et al |

| | | | |
|---|---|---|---|
| EPI_ISL_1064028 | Microbiology and Virology Unit, Azienda Ospedale Padova, Padova, Italy | Department of Molecular Medicine, Computational Medicine Group, Univeresity of Padova, Padova, Italy | Elisa Franchin et al |
| EPI_ISL_429168 | Ramathibodi Hospital | COVID-19 Network Investigations (CONI) Alliance | Elizabeth Batty et al |
| EPI_ISL_447004 | Ramathibodi Hospital | COVID-19 Network Investigations (CONI) Alliance | Elizabeth Batty et al |
| EPI_ISL_445380 | Ramathibodi Hospital | COVID-19 Network Investigations (CONI) Alliance | Elizabeth Batty et al |
| EPI_ISL_447029 | Ramathibodi Hospital | COVID-19 Network Investigations (CONI) Alliance | Elizabeth Batty et al |
| EPI_ISL_458028 | Hospital for Tropical Diseases | COVID-19 Network Investigations (CONI) Alliance | Elizabeth Batty et al |
| EPI_ISL_455916 | Ramathibodi Hospital | COVID-19 Network Investigations (CONI) Alliance | Elizabeth Batty et al |
| EPI_ISL_1138829 | Institute for Urban Disease Control and Prevention | COVID-19 Network Investigations (CONI) Alliance | Elizabeth Batty et al |
| EPI_ISL_1073954 | Ramathibodi Hospital | COVID-19 Network Investigations (CONI) Alliance | Elizabeth Batty et al |
| EPI_ISL_1491570 | Institute for Urban Disease Control and Prevention | COVID-19 Network Investigations (CONI) Alliance | Elizabeth Batty et al |
| EPI_ISL_1511124 | Institute for Urban Disease Control and Prevention | COVID-19 Network Investigations (CONI) Alliance | Elizabeth Batty et al |
| EPI_ISL_1511128 | Institute for Urban Disease Control and Prevention | COVID-19 Network Investigations (CONI) Alliance | Elizabeth Batty et al |
| EPI_ISL_1538423 | Institute for Urban Disease Control and Prevention | COVID-19 Network Investigations (CONI) Alliance | Elizabeth Batty et al |
| EPI_ISL_1538419 | Institute for Urban Disease Control and Prevention | COVID-19 Network Investigations (CONI) Alliance | Elizabeth Batty et al |
| EPI_ISL_1538418 | Institute for Urban Disease Control and Prevention | COVID-19 Network Investigations (CONI) Alliance | Elizabeth Batty et al |
| EPI_ISL_1538431 | Faculty of Medicine Vajira Hospital | COVID-19 Network Investigations (CONI) Alliance | Elizabeth Batty et al |
| EPI_ISL_1491572 | Division of Epidemiology | COVID-19 Network Investigations (CONI) Alliance | Elizabeth Batty et al |
| EPI_ISL_1491563 | Division of Epidemiology | COVID-19 Network Investigations (CONI) Alliance | Elizabeth Batty et al |
| EPI_ISL_1491564 | Division of Epidemiology | COVID-19 Network Investigations (CONI) Alliance | Elizabeth Batty et al |
| EPI_ISL_1491566 | Division of Epidemiology | COVID-19 Network Investigations (CONI) Alliance | Elizabeth Batty et al |
| EPI_ISL_1073965 | Office of Diseases Prevention and Control Region 4 Saraburi | COVID-19 Network Investigations (CONI) Alliance | Elizabeth Batty et al |
| EPI_ISL_1491576 | Faculty of Medicine Vajira Hospital | COVID-19 Network Investigations (CONI) Alliance | Elizabeth Batty et al |
| EPI_ISL_1491579 | Faculty of Medicine Vajira Hospital | COVID-19 Network Investigations (CONI) Alliance | Elizabeth Batty et al |
| EPI_ISL_1491581 | Faculty of Medicine Vajira Hospital | COVID-19 Network Investigations (CONI) Alliance | Elizabeth Batty et al |
| EPI_ISL_1538420 | Faculty of Medicine Vajira Hospital | COVID-19 Network Investigations (CONI) Alliance | Elizabeth Batty et al |
| EPI_ISL_882780 | Ramathibodi Chakri Naruebodindra Hospital | COVID-19 Network Investigations (CONI) Alliance | Elizabeth Batty et al |
| EPI_ISL_1073958 | Ramathibodi Hospital | COVID-19 Network Investigations (CONI) Alliance | Elizabeth Batty et al |
| EPI_ISL_978544 | Centre de Virologie des Maladies infectueuses Tropicales | Functional Genomic Platform UATRS-biology, CNRST | Elmostafa EL FAHIME et al |
| EPI_ISL_978549 | Centre de Virologie des Maladies infectueuses Tropicales | Functional Genomic Platform UATRS-biology, CNRST | Elmostafa EL FAHIME et al |
| EPI_ISL_802856 | Vilnius University Hospital Santaros Klinikos, Vilnius University | Institute of Biotechnology, Life Sciences Center, Vilnius University | Emilija Vasiliunaite et al |

| | | | |
|---|---|---|---|
| EPI_ISL_802860 | Vilnius University Hospital Santaros Klinikos, Vilnius University | Institute of Biotechnology, Life Sciences Center, Vilnius University | Emilija Vasiliunaite et al |
| EPI_ISL_802861 | Vilnius University Hospital Santaros Klinikos, Vilnius University | Institute of Biotechnology, Life Sciences Center, Vilnius University | Emilija Vasiliunaite et al |
| EPI_ISL_904939 | Vilnius University Hospital Santaros Klinikos, Vilnius University | Institute of Biotechnology, Life Sciences Center, Vilnius University | Emilija Vasiliunaite et al |
| EPI_ISL_904945 | Vilnius University Hospital Santaros Klinikos, Vilnius University | Institute of Biotechnology, Life Sciences Center, Vilnius University | Emilija Vasiliunaite et al |
| EPI_ISL_1420640 | Biomedical Research Foundation of the Academy of Athens (BRFAA) | Greek Genome Center, Biomedical Research Foundation of the Academy of Athens (BRFAA) | Emmanouil Athanasiadis et al |
| EPI_ISL_1420691 | Biomedical Research Foundation of the Academy of Athens (BRFAA) | Greek Genome Center, Biomedical Research Foundation of the Academy of Athens (BRFAA) | Emmanouil Athanasiadis et al |
| EPI_ISL_1420707 | Biomedical Research Foundation of the Academy of Athens (BRFAA) | Greek Genome Center, Biomedical Research Foundation of the Academy of Athens (BRFAA) | Emmanouil Athanasiadis et al |
| EPI_ISL_1420709 | Biomedical Research Foundation of the Academy of Athens (BRFAA) | Greek Genome Center, Biomedical Research Foundation of the Academy of Athens (BRFAA) | Emmanouil Athanasiadis et al |
| EPI_ISL_1063900 | Rwanda National Reference Laboratory | Rwanda National Reference Laboratory | Enatha Mukantwari et al |
| EPI_ISL_1064147 | Rwanda National Reference Laboratory | Rwanda National Reference Laboratory | ENatha Mukantwari et al |
| EPI_ISL_1064148 | Rwanda National Reference Laboratory | Rwanda National Reference Laboratory | Enatha Mukantwari et al |
| EPI_ISL_1064168 | Rwanda National Reference Laboratory | Rwanda National Reference Laboratory | Enatha Mukantwari et al |
| EPI_ISL_1498123 | National Reference Laboratory | National Reference Laboratory | Enatha Mukantwari et al |
| EPI_ISL_1064170 | Rwanda National Reference Laboratory | Rwanda National Reference Laboratory | Enatha Mukantwari et al |
| EPI_ISL_1041199 | Hungarian Defence Forces Military Medical Centre | National Laboratory of Virology, Szentágothai Research Centre | Endre Gábor Tóth et al |
| EPI_ISL_1041202 | Hungarian Defence Forces Military Medical Centre | National Laboratory of Virology, Szentágothai Research Centre | Endre Gábor Tóth et al |
| EPI_ISL_1041201 | Hungarian Defence Forces Military Medical Centre | National Laboratory of Virology, Szentágothai Research Centre | Endre Gábor Tóth et al |
| EPI_ISL_1041200 | Hungarian Defence Forces Military Medical Centre | National Laboratory of Virology, Szentágothai Research Centre | Endre Gábor Tóth et al |
| EPI_ISL_1041207 | Hungarian Defence Forces Military Medical Centre | National Laboratory of Virology, Szentágothai Research Centre | Endre Gábor Tóth et al |
| EPI_ISL_1041208 | Hungarian Defence Forces Military Medical Centre | National Laboratory of Virology, Szentágothai Research Centre | Endre Gábor Tóth et al |
| EPI_ISL_1041211 | Hungarian Defence Forces Military Medical Centre | National Laboratory of Virology, Szentágothai Research Centre | Endre Gábor Tóth et al |
| EPI_ISL_1041212 | Hungarian Defence Forces Military Medical Centre | National Laboratory of Virology, Szentágothai Research Centre | Endre Gábor Tóth et al |
| EPI_ISL_526225 | Hungarian Defence Forces Military Medical Centre | National Laboratory of Virology, Szentágothai Research Centre | Endre Gábor Tóth et al |
| EPI_ISL_526236 | Hungarian Defence Forces Military Medical Centre | National Laboratory of Virology, Szentágothai Research Centre | Endre Gábor Tóth et al |
| EPI_ISL_526220 | Hungarian Defence Forces Military Medical Centre | National Laboratory of Virology, Szentágothai Research Centre | Endre Gábor Tóth et al |
| EPI_ISL_526221 | Hungarian Defence Forces Military Medical Centre | National Laboratory of Virology, Szentágothai Research Centre | Endre Gábor Tóth et al |
| EPI_ISL_526223 | Hungarian Defence Forces Military Medical Centre | National Laboratory of Virology, Szentágothai Research Centre | Endre Gábor Tóth et al |
| EPI_ISL_671425 | University of Debrecen, Department of Medical Microbiology | National Laboratory of Virology, Szentágothai Research Centre | Endre Gábor Tóth et al |
| EPI_ISL_671426 | University of Debrecen, Department of Medical Microbiology | National Laboratory of Virology, Szentágothai Research Centre | Endre Gábor Tóth et al |

| | | | |
|---|---|---|---|
| EPI_ISL_671429 | University of Debrecen, Department of Medical Microbiology | National Laboratory of Virology, Szentágothai Research Centre | Endre Gábor Tóth et al |
| EPI_ISL_671437 | University of Debrecen, Department of Medical Microbiology | National Laboratory of Virology, Szentágothai Research Centre | Endre Gábor Tóth et al |
| EPI_ISL_671439 | University of Debrecen, Department of Medical Microbiology | National Laboratory of Virology, Szentágothai Research Centre | Endre Gábor Tóth et al |
| EPI_ISL_671441 | University of Debrecen, Department of Medical Microbiology | National Laboratory of Virology, Szentágothai Research Centre | Endre Gábor Tóth et al |
| EPI_ISL_671442 | University of Debrecen, Department of Medical Microbiology | National Laboratory of Virology, Szentágothai Research Centre | Endre Gábor Tóth et al |
| EPI_ISL_671451 | University of Debrecen, Department of Medical Microbiology | National Laboratory of Virology, Szentágothai Research Centre | Endre Gábor Tóth et al |
| EPI_ISL_678396 | University of Debrecen, Department of Medical Microbiology | National Laboratory of Virology, Szentágothai Research Centre | Endre Gábor Tóth et al |
| EPI_ISL_477620 | University of Szeged, Institute of Clinical Microbiology | National Laboratory of Virology, Szentágothai Research Centre | Endre Gábor Tóth et al |
| EPI_ISL_677729 | University of Szeged, Institute of Clinical Microbiology | National Laboratory of Virology, Szentágothai Research Centre | Endre Gábor Tóth et al |
| EPI_ISL_477622 | University of Szeged, Institute of Clinical Microbiology | National Laboratory of Virology, Szentágothai Research Centre | Endre Gábor Tóth et al |
| EPI_ISL_677770 | University of Szeged, Institute of Clinical Microbiology | National Laboratory of Virology, Szentágothai Research Centre | Endre Gábor Tóth et al |
| EPI_ISL_677798 | University of Szeged, Institute of Clinical Microbiology | National Laboratory of Virology, Szentágothai Research Centre | Endre Gábor Tóth et al |
| EPI_ISL_984749 | Gazi University Faculty of Medicine, Medical Virology Laboratory | Gazi University Faculty of Medicine, Medical Virology Laboratory | Erdem Şahin et al |
| EPI_ISL_984750 | Gazi University Faculty of Medicine, Medical Virology Laboratory | Gazi University Faculty of Medicine, Medical Virology Laboratory | Erdem Şahin et al |
| EPI_ISL_984990 | Gazi University Faculty of Medicine, Medical Virology Laboratory | Gazi University Faculty of Medicine, Medical Virology Laboratory | Erdem Şahin et al |
| EPI_ISL_730575 | Gazi University Faculty of Medicine, Medical Virology Laboratory | Gazi University Faculty of Medicine, Medical Virology Laboratory | Erdem Şahin et al |
| EPI_ISL_730572 | Gazi University Faculty of Medicine, Medical Virology Laboratory | Gazi University Faculty of Medicine, Medical Virology Laboratory | Erdem Şahin et al |
| EPI_ISL_730571 | Gazi University Faculty of Medicine, Medical Virology Laboratory | Gazi University Faculty of Medicine, Medical Virology Laboratory | Erdem Şahin et al |
| EPI_ISL_730568 | Gazi University Faculty of Medicine, Medical Virology Laboratory | Gazi University Faculty of Medicine, Medical Virology Laboratory | Erdem Şahin et al |
| EPI_ISL_730569 | Gazi University Faculty of Medicine, Medical Virology Laboratory | Gazi University Faculty of Medicine, Medical Virology Laboratory | Erdem Şahin et al |
| EPI_ISL_730570 | Gazi University Faculty of Medicine, Medical Virology Laboratory | Gazi University Faculty of Medicine, Medical Virology Laboratory | Erdem Şahin et al |
| EPI_ISL_730620 | Gazi University Faculty of Medicine, Medical Virology Laboratory | Gazi University Faculty of Medicine, Medical Virology Laboratory | Erdem Şahin et al |
| EPI_ISL_730577 | Gazi University Faculty of Medicine, Medical Virology Laboratory | Gazi University Faculty of Medicine, Medical Virology Laboratory | Erdem Şahin et al |
| EPI_ISL_1273090 | Biochemistry and Molecular Biology Department-Faculty of Medicine, Al-Quds University | Biochemistry and Molecular Biology Department-Faculty of Medicine, Al-Quds University | Ereqat et al |
| EPI_ISL_1273091 | Biochemistry and Molecular Biology Department-Faculty of Medicine, Al-Quds University | Biochemistry and Molecular Biology Department-Faculty of Medicine, Al-Quds University | Ereqat et al |
| EPI_ISL_1273092 | Biochemistry and Molecular Biology Department-Faculty of Medicine, Al-Quds University | Biochemistry and Molecular Biology Department-Faculty of Medicine, Al-Quds University | Ereqat et al |
| EPI_ISL_1273093 | Biochemistry and Molecular Biology Department-Faculty of Medicine, Al-Quds University | Biochemistry and Molecular Biology Department-Faculty of Medicine, Al-Quds University | Ereqat et al |
| EPI_ISL_1273099 | Faculty of Medicine, Al-Quds University | Faculty of Medicine, Al-Quds University | Ereqat et al |
| EPI_ISL_1273102 | Faculty of Medicine, Al-Quds University | Faculty of Medicine, Al-Quds University | Ereqat et al |

| | | | |
|---|---|---|---|
| EPI_ISL_661272 | Al-Quds Nutrition and Health Research Institute, Al-Quds University | Al-Quds Nutrition and Health Research Institute, Al-Quds University | Ereqat et al |
| EPI_ISL_411902 | Virology Unit, Institut Pasteur du Cambodge. | Virology Unit, Institut Pasteur du Cambodge (Sequencing done by: Jessica E Manning/Jennifer A Bohl at Malaria and Vector Research Research Laboratory, National Institute of Allergy and Infectious Diseases and Vida Ahyong from Chan-Zuckerberg Biohub) | Erik A Karlsson et al |
| EPI_ISL_516613 | Instituto de Diagnostico y Referencia Epidemiologicos (INDRE) | Instituto de Diagnostico y Referencia Epidemiologicos (INDRE) | Ernesto Ramirez-Gonzalez et al |
| EPI_ISL_516614 | Instituto de Diagnostico y Referencia Epidemiologicos (INDRE) | Instituto de Diagnostico y Referencia Epidemiologicos (INDRE) | Ernesto Ramirez-Gonzalez et al |
| EPI_ISL_516615 | Instituto de Diagnostico y Referencia Epidemiologicos (INDRE) | Instituto de Diagnostico y Referencia Epidemiologicos (INDRE) | Ernesto Ramirez-Gonzalez et al |
| EPI_ISL_516616 | Instituto de Diagnostico y Referencia Epidemiologicos (INDRE) | Instituto de Diagnostico y Referencia Epidemiologicos (INDRE) | Ernesto Ramirez-Gonzalez et al |
| EPI_ISL_1195537 | Hospital Universitario 12 de Octubre | SeqCOVID-SPAIN consortium/IBV(CSIC) | Esther Viedma et al |
| EPI_ISL_1195547 | Hospital Universitario 12 de Octubre | SeqCOVID-SPAIN consortium/IBV(CSIC) | Esther Viedma et al |
| EPI_ISL_1195548 | Hospital Universitario 12 de Octubre | SeqCOVID-SPAIN consortium/IBV(CSIC) | Esther Viedma et al |
| EPI_ISL_1195557 | Hospital Universitario 12 de Octubre | SeqCOVID-SPAIN consortium/IBV(CSIC) | Esther Viedma et al |
| EPI_ISL_1272574 | Hospital Universitario 12 de Octubre | SeqCOVID-SPAIN consortium/IBV(CSIC) | Esther Viedma et al |
| EPI_ISL_1159375 | Bahman Hospital | Laboratory of Molecular Biology and Cancer Immunology,Lebanese University Public Health England | Fadi Abdel Sater et al |
| EPI_ISL_1196793 | Clinical Microbiology Laboratory, Faculty of Medicine, Universitas Indonesia | Faculty of Medicine, Universitas Indonesia | Fadilah et al |
| EPI_ISL_512922 | Pathogen Genomics Lab King Abdullah University of Science and Technology (KAUST) | Pathogen Genomics Lab King Abdullah University of Science and Technology(KAUST) | Fadwa Alofi et al |
| EPI_ISL_512923 | Pathogen Genomics Lab King Abdullah University of Science and Technology (KAUST) | Pathogen Genomics Lab King Abdullah University of Science and Technology(KAUST) | Fadwa Alofi et al |
| EPI_ISL_458125 | Oman National Influenza Centre | Department of Microbiology and Immunology-SQUH | Fahad Zadjali et al |
| EPI_ISL_457984 | Oman-NIC | Department of Microbiology and Immunology-SQUH | Fahad Zadjali et al |
| EPI_ISL_491972 | Oman-NIC | Department of Microbiology and Immunology-SQUH | Fahad Zadjali et al |
| EPI_ISL_491985 | Oman-NIC | Department of Microbiology and Immunology-SQUH | Fahad Zadjali et al |
| EPI_ISL_491986 | Oman-NIC | Department of Microbiology and Immunology-SQUH | Fahad Zadjali et al |
| EPI_ISL_491989 | Oman-NIC | Department of Microbiology and Immunology-SQUH | Fahad Zadjali et al |
| EPI_ISL_492015 | Oman-NIC | Department of Microbiology and Immunology-SQUH | Fahad Zadjali et al |
| EPI_ISL_492016 | Oman-NIC | Department of Microbiology and Immunology-SQUH | Fahad Zadjali et al |
| EPI_ISL_492018 | Oman-NIC | Department of Microbiology and Immunology-SQUH | Fahad Zadjali et al |
| EPI_ISL_492020 | Oman-NIC | Department of Microbiology and Immunology-SQUH | Fahad Zadjali et al |
| EPI_ISL_492025 | Oman-NIC | Department of Microbiology and Immunology-SQUH | Fahad Zadjali et al |
| EPI_ISL_422424 | Jaber Al Ahmad Al Sabah Hospital | Dasman diabetes Institute | Fahd Al-Mulla et al |
| EPI_ISL_416541 | Dasman Diabetes Institute and Virology Laboratory Ministry of Health | Dasman Diabetes Institute | Fahd Al-Mulla et al |

| | | | |
|---|---|---|---|
| EPI_ISL_416543 | Dasman Diabetes Institute | Dasman Diabetes Institute | Fahd Al-Mulla et al |
| EPI_ISL_463001 | unknown | Clinical virology | Fares et al |
| EPI_ISL_463002 | unknown | Clinical virology | Fares et al |
| EPI_ISL_477227 | Institute for Stem Cell Science and Regenerative Medicine | National Centre for Biological Sciences | Farhan Ali et al |
| EPI_ISL_978536 | Centre de Virologie des Maladies infectueuses Tropicales | Functional Genomic Platform UATRS-biology, CNRST | Farida HILALI et al |
| EPI_ISL_978546 | Centre de Virologie des Maladies infectueuses Tropicales | Functional Genomic Platform UATRS-biology, CNRST | Farida HILALI et al |
| EPI_ISL_437334 | Ministry of Health Turkey | Ministry of Health Turkey | Fatma Bayrakdar et al |
| EPI_ISL_812774 | Ministry of Health Turkey | Ministry of Health Turkey | Fatma Bayrakdar et al |
| EPI_ISL_812765 | Ministry of Health Turkey | Ministry of Health Turkey | Fatma Bayrakdar et al |
| EPI_ISL_812777 | Ministry of Health Turkey | Ministry of Health Turkey | Fatma Bayrakdar et al |
| EPI_ISL_1379950 | Ministry of Health Turkey | Ministry of Health Turkey | Fatma Bayrakdar et al |
| EPI_ISL_437319 | Ministry of Health Turkey | Ministry of Health Turkey | Fatma Bayrakdar et al |
| EPI_ISL_437325 | Ministry of Health Turkey | Ministry of Health Turkey | Fatma Bayrakdar et al |
| EPI_ISL_437321 | Ministry of Health Turkey | Ministry of Health Turkey | Fatma Bayrakdar et al |
| EPI_ISL_1534486 | Ministry of Health Turkey | Ministry of Health Turkey | Fatma Bayrakdar et al |
| EPI_ISL_1008621 | Ministry of Health Turkey | Ministry of Health Turkey | Fatma Bayrakdar et al |
| EPI_ISL_812876 | Ministry of Health Turkey | Ministry of Health Turkey | Fatma Bayrakdar et al |
| EPI_ISL_812877 | Ministry of Health Turkey | Ministry of Health Turkey | Fatma Bayrakdar et al |
| EPI_ISL_814079 | Ministry of Health Turkey | Ministry of Health Turkey | Fatma Bayrakdar et al |
| EPI_ISL_814091 | Ministry of Health Turkey | Ministry of Health Turkey | Fatma Bayrakdar et al |
| EPI_ISL_814067 | Ministry of Health Turkey | Ministry of Health Turkey | Fatma Bayrakdar et al |
| EPI_ISL_814065 | Ministry of Health Turkey | Ministry of Health Turkey | Fatma Bayrakdar et al |
| EPI_ISL_814066 | Ministry of Health Turkey | Ministry of Health Turkey | Fatma Bayrakdar et al |
| EPI_ISL_1063969 | Ministry of Health Turkey | Ministry of Health Turkey | Fatma Bayrakdar et al |
| EPI_ISL_437307 | Ministry of Health Turkey | Ministry of Health Turkey | Fatma Bayrakdar et al |
| EPI_ISL_1170075 | Ministry of Health Turkey | Ministry of Health Turkey | Fatma Bayrakdar et al |
| EPI_ISL_1183638 | Ministry of Health Turkey | Ministry of Health Turkey | Fatma Bayrakdar et al |
| EPI_ISL_1211725 | Ministry of Health Turkey | Ministry of Health Turkey | Fatma Bayrakdar et al |
| EPI_ISL_1211766 | Ministry of Health Turkey | Ministry of Health Turkey | Fatma Bayrakdar et al |
| EPI_ISL_1263298 | Ministry of Health Turkey | Ministry of Health Turkey | Fatma Bayrakdar et al |

| | | | |
|---|---|---|---|
| EPI_ISL_812886 | Ministry of Health Turkey | Ministry of Health Turkey | Fatma Bayrakdar et al |
| EPI_ISL_812897 | Ministry of Health Turkey | Ministry of Health Turkey | Fatma Bayrakdar et al |
| EPI_ISL_812902 | Ministry of Health Turkey | Ministry of Health Turkey | Fatma Bayrakdar et al |
| EPI_ISL_1403611 | Molecular Virology Unit, Microbiology and Virology Depertment, Fondazione IRCCS Policlinico San Matteo, Pavia | Molecular Virology Unit, Microbiology and Virology Depertment, Fondazione IRCCS Policlinico San Matteo, Pavia | Federica Giardina et al |
| EPI_ISL_779155 | Laboratório de Microbiologia Molecular - Universidade FEEVALE | Bioinformatics Laboratory / LNCC | Felipe Benites et al |
| EPI_ISL_779156 | Laboratório de Microbiologia Molecular - Universidade FEEVALE | Bioinformatics Laboratory / LNCC | Felipe Benites et al |
| EPI_ISL_770555 | Laboratório de Microbiologia Molecular - Universidade FEEVALE | Bioinformatics Laboratory / LNCC | Felipe Benites et al |
| EPI_ISL_770556 | Laboratório de Microbiologia Molecular - Universidade FEEVALE | Bioinformatics Laboratory / LNCC | Felipe Benites et al |
| EPI_ISL_770557 | Laboratório de Microbiologia Molecular - Universidade FEEVALE | Bioinformatics Laboratory / LNCC | Felipe Benites et al |
| EPI_ISL_770558 | Laboratório de Microbiologia Molecular - Universidade FEEVALE | Bioinformatics Laboratory / LNCC | Felipe Benites et al |
| EPI_ISL_770559 | Laboratório de Microbiologia Molecular - Universidade FEEVALE | Bioinformatics Laboratory / LNCC | Felipe Benites et al |
| EPI_ISL_770560 | Laboratório de Microbiologia Molecular - Universidade FEEVALE | Bioinformatics Laboratory / LNCC | Felipe Benites et al |
| EPI_ISL_770561 | Laboratório de Microbiologia Molecular - Universidade FEEVALE | Bioinformatics Laboratory / LNCC | Felipe Benites et al |
| EPI_ISL_770562 | Laboratório de Microbiologia Molecular - Universidade FEEVALE | Bioinformatics Laboratory / LNCC | Felipe Benites et al |
| EPI_ISL_770563 | Laboratório de Microbiologia Molecular - Universidade FEEVALE | Bioinformatics Laboratory / LNCC | Felipe Benites et al |
| EPI_ISL_770564 | Laboratório de Microbiologia Molecular - Universidade FEEVALE | Bioinformatics Laboratory / LNCC | Felipe Benites et al |
| EPI_ISL_770565 | Laboratório de Microbiologia Molecular - Universidade FEEVALE | Bioinformatics Laboratory / LNCC | Felipe Benites et al |
| EPI_ISL_770566 | Laboratório de Microbiologia Molecular - Universidade FEEVALE | Bioinformatics Laboratory / LNCC | Felipe Benites et al |
| EPI_ISL_770567 | Laboratório de Microbiologia Molecular - Universidade FEEVALE | Bioinformatics Laboratory / LNCC | Felipe Benites et al |
| EPI_ISL_770568 | Laboratório de Microbiologia Molecular - Universidade FEEVALE | Bioinformatics Laboratory / LNCC | Felipe Benites et al |
| EPI_ISL_770569 | Laboratório de Microbiologia Molecular - Universidade FEEVALE | Bioinformatics Laboratory / LNCC | Felipe Benites et al |
| EPI_ISL_770570 | Laboratório de Microbiologia Molecular - Universidade FEEVALE | Bioinformatics Laboratory / LNCC | Felipe Benites et al |
| EPI_ISL_770571 | Laboratório de Microbiologia Molecular - Universidade FEEVALE | Bioinformatics Laboratory / LNCC | Felipe Benites et al |
| EPI_ISL_770572 | Laboratório de Microbiologia Molecular - Universidade FEEVALE | Bioinformatics Laboratory / LNCC | Felipe Benites et al |
| EPI_ISL_779157 | Laboratório de Microbiologia Molecular - Universidade FEEVALE | Bioinformatics Laboratory / LNCC | Felipe Benites et al |
| EPI_ISL_770573 | Laboratório de Microbiologia Molecular - Universidade FEEVALE | Bioinformatics Laboratory / LNCC | Felipe Benites et al |
| EPI_ISL_770574 | Laboratório de Microbiologia Molecular - Universidade FEEVALE | Bioinformatics Laboratory / LNCC | Felipe Benites et al |
| EPI_ISL_779158 | Laboratório de Microbiologia Molecular - Universidade FEEVALE | Bioinformatics Laboratory / LNCC | Felipe Benites et al |
| EPI_ISL_770575 | Laboratório de Microbiologia Molecular - Universidade FEEVALE | Bioinformatics Laboratory / LNCC | Felipe Benites et al |
| EPI_ISL_770576 | Laboratório de Microbiologia Molecular - Universidade FEEVALE | Bioinformatics Laboratory / LNCC | Felipe Benites et al |

| | | | |
|---|---|---|---|
| EPI_ISL_770577 | Laboratório de Microbiologia Molecular - Universidade FEEVALE | Bioinformatics Laboratory / LNCC | Felipe Benites et al |
| EPI_ISL_770553 | Laboratório de Microbiologia Molecular - Universidade FEEVALE | Bioinformatics Laboratory / LNCC | Felipe Benites et al |
| EPI_ISL_770578 | Laboratório de Microbiologia Molecular - Universidade FEEVALE | Bioinformatics Laboratory / LNCC | Felipe Benites et al |
| EPI_ISL_770579 | Laboratório de Microbiologia Molecular - Universidade FEEVALE | Bioinformatics Laboratory / LNCC | Felipe Benites et al |
| EPI_ISL_779159 | Laboratório de Microbiologia Molecular - Universidade FEEVALE | Bioinformatics Laboratory / LNCC | Felipe Benites et al |
| EPI_ISL_770580 | Laboratório de Microbiologia Molecular - Universidade FEEVALE | Bioinformatics Laboratory / LNCC | Felipe Benites et al |
| EPI_ISL_770581 | Laboratório de Microbiologia Molecular - Universidade FEEVALE | Bioinformatics Laboratory / LNCC | Felipe Benites et al |
| EPI_ISL_770582 | Laboratório de Microbiologia Molecular - Universidade FEEVALE | Bioinformatics Laboratory / LNCC | Felipe Benites et al |
| EPI_ISL_770583 | Laboratório de Microbiologia Molecular - Universidade FEEVALE | Bioinformatics Laboratory / LNCC | Felipe Benites et al |
| EPI_ISL_770584 | Laboratório de Microbiologia Molecular - Universidade FEEVALE | Bioinformatics Laboratory / LNCC | Felipe Benites et al |
| EPI_ISL_770585 | Laboratório de Microbiologia Molecular - Universidade FEEVALE | Bioinformatics Laboratory / LNCC | Felipe Benites et al |
| EPI_ISL_770586 | Laboratório de Microbiologia Molecular - Universidade FEEVALE | Bioinformatics Laboratory / LNCC | Felipe Benites et al |
| EPI_ISL_770587 | Laboratório de Microbiologia Molecular - Universidade FEEVALE | Bioinformatics Laboratory / LNCC | Felipe Benites et al |
| EPI_ISL_770588 | Laboratório de Microbiologia Molecular - Universidade FEEVALE | Bioinformatics Laboratory / LNCC | Felipe Benites et al |
| EPI_ISL_779160 | Laboratório de Microbiologia Molecular - Universidade FEEVALE | Bioinformatics Laboratory / LNCC | Felipe Benites et al |
| EPI_ISL_770589 | Laboratório de Microbiologia Molecular - Universidade FEEVALE | Bioinformatics Laboratory / LNCC | Felipe Benites et al |
| EPI_ISL_770590 | Laboratório de Microbiologia Molecular - Universidade FEEVALE | Bioinformatics Laboratory / LNCC | Felipe Benites et al |
| EPI_ISL_779161 | Laboratório de Microbiologia Molecular - Universidade FEEVALE | Bioinformatics Laboratory / LNCC | Felipe Benites et al |
| EPI_ISL_770591 | Laboratório de Microbiologia Molecular - Universidade FEEVALE | Bioinformatics Laboratory / LNCC | Felipe Benites et al |
| EPI_ISL_770592 | Laboratório de Microbiologia Molecular - Universidade FEEVALE | Bioinformatics Laboratory / LNCC | Felipe Benites et al |
| EPI_ISL_770593 | Laboratório de Microbiologia Molecular - Universidade FEEVALE | Bioinformatics Laboratory / LNCC | Felipe Benites et al |
| EPI_ISL_770594 | Laboratório de Microbiologia Molecular - Universidade FEEVALE | Bioinformatics Laboratory / LNCC | Felipe Benites et al |
| EPI_ISL_770595 | Laboratório de Microbiologia Molecular - Universidade FEEVALE | Bioinformatics Laboratory / LNCC | Felipe Benites et al |
| EPI_ISL_770596 | Laboratório de Microbiologia Molecular - Universidade FEEVALE | Bioinformatics Laboratory / LNCC | Felipe Benites et al |
| EPI_ISL_770551 | Laboratório de Microbiologia Molecular - Universidade FEEVALE | Bioinformatics Laboratory / LNCC | Felipe Benites et al |
| EPI_ISL_770597 | Laboratório de Microbiologia Molecular - Universidade FEEVALE | Bioinformatics Laboratory / LNCC | Felipe Benites et al |
| EPI_ISL_770598 | Laboratório de Microbiologia Molecular - Universidade FEEVALE | Bioinformatics Laboratory / LNCC | Felipe Benites et al |
| EPI_ISL_770599 | Laboratório de Microbiologia Molecular - Universidade FEEVALE | Bioinformatics Laboratory / LNCC | Felipe Benites et al |
| EPI_ISL_770552 | Laboratório de Microbiologia Molecular - Universidade FEEVALE | Bioinformatics Laboratory / LNCC | Felipe Benites et al |
| EPI_ISL_770600 | Laboratório de Microbiologia Molecular - Universidade FEEVALE | Bioinformatics Laboratory / LNCC | Felipe Benites et al |

| | | | |
|---|---|---|---|
| EPI_ISL_770601 | Laboratório de Microbiologia Molecular - Universidade FEEVALE | Bioinformatics Laboratory / LNCC | Felipe Benites et al |
| EPI_ISL_779162 | Laboratório de Microbiologia Molecular - Universidade FEEVALE | Bioinformatics Laboratory / LNCC | Felipe Benites et al |
| EPI_ISL_770602 | Laboratório de Microbiologia Molecular - Universidade FEEVALE | Bioinformatics Laboratory / LNCC | Felipe Benites et al |
| EPI_ISL_770603 | Laboratório de Microbiologia Molecular - Universidade FEEVALE | Bioinformatics Laboratory / LNCC | Felipe Benites et al |
| EPI_ISL_770604 | Laboratório de Microbiologia Molecular - Universidade FEEVALE | Bioinformatics Laboratory / LNCC | Felipe Benites et al |
| EPI_ISL_770605 | Laboratório de Microbiologia Molecular - Universidade FEEVALE | Bioinformatics Laboratory / LNCC | Felipe Benites et al |
| EPI_ISL_770606 | Laboratório de Microbiologia Molecular - Universidade FEEVALE | Bioinformatics Laboratory / LNCC | Felipe Benites et al |
| EPI_ISL_779163 | Laboratório de Microbiologia Molecular - Universidade FEEVALE | Bioinformatics Laboratory / LNCC | Felipe Benites et al |
| EPI_ISL_770607 | Laboratório de Microbiologia Molecular - Universidade FEEVALE | Bioinformatics Laboratory / LNCC | Felipe Benites et al |
| EPI_ISL_770608 | Laboratório de Microbiologia Molecular - Universidade FEEVALE | Bioinformatics Laboratory / LNCC | Felipe Benites et al |
| EPI_ISL_770609 | Laboratório de Microbiologia Molecular - Universidade FEEVALE | Bioinformatics Laboratory / LNCC | Felipe Benites et al |
| EPI_ISL_770610 | Laboratório de Microbiologia Molecular - Universidade FEEVALE | Bioinformatics Laboratory / LNCC | Felipe Benites et al |
| EPI_ISL_770611 | Laboratório de Microbiologia Molecular - Universidade FEEVALE | Bioinformatics Laboratory / LNCC | Felipe Benites et al |
| EPI_ISL_770612 | Laboratório de Microbiologia Molecular - Universidade FEEVALE | Bioinformatics Laboratory / LNCC | Felipe Benites et al |
| EPI_ISL_779164 | Laboratório de Microbiologia Molecular - Universidade FEEVALE | Bioinformatics Laboratory / LNCC | Felipe Benites et al |
| EPI_ISL_770613 | Laboratório de Microbiologia Molecular - Universidade FEEVALE | Bioinformatics Laboratory / LNCC | Felipe Benites et al |
| EPI_ISL_770614 | Laboratório de Microbiologia Molecular - Universidade FEEVALE | Bioinformatics Laboratory / LNCC | Felipe Benites et al |
| EPI_ISL_770615 | Laboratório de Microbiologia Molecular - Universidade FEEVALE | Bioinformatics Laboratory / LNCC | Felipe Benites et al |
| EPI_ISL_770616 | Laboratório de Microbiologia Molecular - Universidade FEEVALE | Bioinformatics Laboratory / LNCC | Felipe Benites et al |
| EPI_ISL_770617 | Laboratório de Microbiologia Molecular - Universidade FEEVALE | Bioinformatics Laboratory / LNCC | Felipe Benites et al |
| EPI_ISL_779165 | Laboratório de Microbiologia Molecular - Universidade FEEVALE | Bioinformatics Laboratory / LNCC | Felipe Benites et al |
| EPI_ISL_779166 | Laboratório de Microbiologia Molecular - Universidade FEEVALE | Bioinformatics Laboratory / LNCC | Felipe Benites et al |
| EPI_ISL_770618 | Laboratório de Microbiologia Molecular - Universidade FEEVALE | Bioinformatics Laboratory / LNCC | Felipe Benites et al |
| EPI_ISL_770619 | Laboratório de Microbiologia Molecular - Universidade FEEVALE | Bioinformatics Laboratory / LNCC | Felipe Benites et al |
| EPI_ISL_770620 | Laboratório de Microbiologia Molecular - Universidade FEEVALE | Bioinformatics Laboratory / LNCC | Felipe Benites et al |
| EPI_ISL_770554 | Laboratório de Microbiologia Molecular - Universidade FEEVALE | Bioinformatics Laboratory / LNCC | Felipe Benites et al |
| EPI_ISL_770621 | Laboratório de Microbiologia Molecular - Universidade FEEVALE | Bioinformatics Laboratory / LNCC | Felipe Benites et al |
| EPI_ISL_770622 | Laboratório de Microbiologia Molecular - Universidade FEEVALE | Bioinformatics Laboratory / LNCC | Felipe Benites et al |
| EPI_ISL_770623 | Laboratório de Microbiologia Molecular - Universidade FEEVALE | Bioinformatics Laboratory / LNCC | Felipe Benites et al |
| EPI_ISL_770624 | Laboratório de Microbiologia Molecular - Universidade FEEVALE | Bioinformatics Laboratory / LNCC | Felipe Benites et al |

| | | | |
|---|---|---|---|
| EPI_ISL_779167 | Laboratório de Microbiologia Molecular - Universidade FEEVALE | Bioinformatics Laboratory / LNCC | Felipe Benites et al |
| EPI_ISL_779168 | Laboratório de Microbiologia Molecular - Universidade FEEVALE | Bioinformatics Laboratory / LNCC | Felipe Benites et al |
| EPI_ISL_770625 | Laboratório de Microbiologia Molecular - Universidade FEEVALE | Bioinformatics Laboratory / LNCC | Felipe Benites et al |
| EPI_ISL_770626 | Laboratório de Microbiologia Molecular - Universidade FEEVALE | Bioinformatics Laboratory / LNCC | Felipe Benites et al |
| EPI_ISL_770627 | Laboratório de Microbiologia Molecular - Universidade FEEVALE | Bioinformatics Laboratory / LNCC | Felipe Benites et al |
| EPI_ISL_770628 | Laboratório de Microbiologia Molecular - Universidade FEEVALE | Bioinformatics Laboratory / LNCC | Felipe Benites et al |
| EPI_ISL_770629 | Laboratório de Microbiologia Molecular - Universidade FEEVALE | Bioinformatics Laboratory / LNCC | Felipe Benites et al |
| EPI_ISL_770630 | Laboratório de Microbiologia Molecular - Universidade FEEVALE | Bioinformatics Laboratory / LNCC | Felipe Benites et al |
| EPI_ISL_792528 | Laboratorio Central, Ministerio de Salud Córdoba | Instituto de Patologia Vegetal (CIAP-INTA) on behalf of 'Proyecto Argentino Interinstitucional de genomica de SARS-CoV-2' (PAIS Consortium) | Fernández et al |
| EPI_ISL_792531 | Laboratorio Central, Ministerio de Salud Córdoba | Instituto de Patologia Vegetal (CIAP-INTA) on behalf of 'Proyecto Argentino Interinstitucional de genomica de SARS-CoV-2' (PAIS Consortium) | Fernández et al |
| EPI_ISL_792533 | Laboratorio Central, Ministerio de Salud Córdoba | Instituto de Patologia Vegetal (CIAP-INTA) on behalf of 'Proyecto Argentino Interinstitucional de genomica de SARS-CoV-2' (PAIS Consortium) | Fernández et al |
| EPI_ISL_792537 | Laboratorio Central, Ministerio de Salud Córdoba | Instituto de Patologia Vegetal (CIAP-INTA) on behalf of 'Proyecto Argentino Interinstitucional de genomica de SARS-CoV-2' (PAIS Consortium) | Fernández et al |
| EPI_ISL_792540 | Laboratorio Central, Ministerio de Salud Córdoba | Instituto de Patologia Vegetal (CIAP-INTA) on behalf of 'Proyecto Argentino Interinstitucional de genomica de SARS-CoV-2' (PAIS Consortium) | Fernández et al |
| EPI_ISL_1396016 | Laboratorio Central, Ministerio de Salud Cordoba | Instituto de Patologia Vegetal (CIAP-INTA) on behalf of 'Proyecto Argentino Interinstitucional de genomica de SARS-CoV-2' (PAIS Consortium) | Fernández et al |
| EPI_ISL_1395973 | Laboratorio Central, Ministerio de Salud Cordoba | Instituto de Patologia Vegetal (CIAP-INTA) on behalf of 'Proyecto Argentino Interinstitucional de genomica de SARS-CoV-2' (PAIS Consortium) | Fernández et al |
| EPI_ISL_1396019 | Laboratorio Central, Ministerio de Salud Cordoba | Instituto de Patologia Vegetal (CIAP-INTA) on behalf of 'Proyecto Argentino Interinstitucional de genomica de SARS-CoV-2' (PAIS Consortium) | Fernández et al |
| EPI_ISL_1395975 | Laboratorio Central, Ministerio de Salud Cordoba | Instituto de Patologia Vegetal (CIAP-INTA) on behalf of 'Proyecto Argentino Interinstitucional de genomica de SARS-CoV-2' (PAIS Consortium) | Fernández et al |
| EPI_ISL_1395979 | Laboratorio Central, Ministerio de Salud Cordoba | Instituto de Patologia Vegetal (CIAP-INTA) on behalf of 'Proyecto Argentino Interinstitucional de genomica de SARS-CoV-2' (PAIS Consortium) | Fernández et al |
| EPI_ISL_1395980 | Laboratorio Central, Ministerio de Salud Cordoba | Instituto de Patologia Vegetal (CIAP-INTA) on behalf of 'Proyecto Argentino Interinstitucional de genomica de SARS-CoV-2' (PAIS Consortium) | Fernández et al |
| EPI_ISL_1395982 | Laboratorio Central, Ministerio de Salud Cordoba | Instituto de Patologia Vegetal (CIAP-INTA) on behalf of 'Proyecto Argentino Interinstitucional de genomica de SARS-CoV-2' (PAIS Consortium) | Fernández et al |
| EPI_ISL_1395971 | Laboratorio Central, Ministerio de Salud Cordoba | Instituto de Patologia Vegetal (CIAP-INTA) on behalf of 'Proyecto Argentino Interinstitucional de genomica de SARS-CoV-2' (PAIS Consortium) | Fernández et al |
| EPI_ISL_1395988 | Laboratorio Central, Ministerio de Salud Cordoba | Instituto de Patologia Vegetal (CIAP-INTA) on behalf of 'Proyecto Argentino Interinstitucional de genomica de SARS-CoV-2' (PAIS Consortium) | Fernández et al |
| EPI_ISL_1396040 | Laboratorio Central, Ministerio de Salud Cordoba | Instituto de Patologia Vegetal (CIAP-INTA) on behalf of 'Proyecto Argentino Interinstitucional de genomica de SARS-CoV-2' (PAIS Consortium) | Fernández et al |
| EPI_ISL_1395991 | Laboratorio Central, Ministerio de Salud Cordoba | Instituto de Patologia Vegetal (CIAP-INTA) on behalf of 'Proyecto Argentino Interinstitucional de genomica de SARS-CoV-2' (PAIS Consortium) | Fernández et al |

| | | | |
|---|---|---|---|
| EPI_ISL_1396047 | Laboratorio Central, Ministerio de Salud Cordoba | Instituto de Patologia Vegetal (CIAP-INTA) on behalf of 'Proyecto Argentino Interinstitucional de genomica de SARS-CoV-2' (PAIS Consortium) | Fernández et al |
| EPI_ISL_1396000 | Laboratorio Central, Ministerio de Salud Cordoba | Instituto de Patologia Vegetal (CIAP-INTA) on behalf of 'Proyecto Argentino Interinstitucional de genomica de SARS-CoV-2' (PAIS Consortium) | Fernández et al |
| EPI_ISL_1396041 | Laboratorio Central, Ministerio de Salud Cordoba | Instituto de Patologia Vegetal (CIAP-INTA) on behalf of 'Proyecto Argentino Interinstitucional de genomica de SARS-CoV-2' (PAIS Consortium) | Fernández et al |
| EPI_ISL_1396002 | Laboratorio Central, Ministerio de Salud Cordoba | Instituto de Patologia Vegetal (CIAP-INTA) on behalf of 'Proyecto Argentino Interinstitucional de genomica de SARS-CoV-2' (PAIS Consortium) | Fernández et al |
| EPI_ISL_1396003 | Laboratorio Central, Ministerio de Salud Cordoba | Instituto de Patologia Vegetal (CIAP-INTA) on behalf of 'Proyecto Argentino Interinstitucional de genomica de SARS-CoV-2' (PAIS Consortium) | Fernández et al |
| EPI_ISL_1396060 | Laboratorio Central, Ministerio de Salud Cordoba | Instituto de Patologia Vegetal (CIAP-INTA) on behalf of 'Proyecto Argentino Interinstitucional de genomica de SARS-CoV-2' (PAIS Consortium) | Fernández et al |
| EPI_ISL_1396004 | Laboratorio Central, Ministerio de Salud Cordoba | Instituto de Patologia Vegetal (CIAP-INTA) on behalf of 'Proyecto Argentino Interinstitucional de genomica de SARS-CoV-2' (PAIS Consortium) | Fernández et al |
| EPI_ISL_1396023 | Laboratorio Central, Ministerio de Salud Cordoba | Instituto de Patologia Vegetal (CIAP-INTA) on behalf of 'Proyecto Argentino Interinstitucional de genomica de SARS-CoV-2' (PAIS Consortium) | Fernández et al |
| EPI_ISL_1395968 | Laboratorio Central, Ministerio de Salud Cordoba | Instituto de Patologia Vegetal (CIAP-INTA) on behalf of 'Proyecto Argentino Interinstitucional de genomica de SARS-CoV-2' (PAIS Consortium) | Fernández et al |
| EPI_ISL_1396076 | Laboratorio de Salud Pública | Instituto de Patologia Vegetal (CIAP-INTA) on behalf of 'Proyecto Argentino Interinstitucional de genomica de SARS-CoV-2' (PAIS Consortium) | Fernández et al |
| EPI_ISL_1469606 | Fundação Hospitalar de Sapucaia do Sul | Epiclin | Fernando Hayashi Sant'Anna et al |
| EPI_ISL_1469805 | Fundação Hospitalar de Sapucaia do Sul | Epiclin | Fernando Hayashi Sant'Anna et al |
| EPI_ISL_1469703 | Unidade de Pronto Atendimento de Sapucaia do Sul | Epiclin | Fernando Hayashi Sant'Anna et al |
| EPI_ISL_1469787 | Fundação Hospitalar de Sapucaia do Sul | Epiclin | Fernando Hayashi Sant'Anna et al |
| EPI_ISL_1469695 | Unidade de Pronto Atendimento de Sapucaia do Sul | Epiclin | Fernando Hayashi Sant'Anna et al |
| EPI_ISL_1469599 | Hospital Montenegro | Epiclin | Fernando Hayashi Sant'Anna et al |
| EPI_ISL_1469799 | Hospital Regina | Epiclin | Fernando Hayashi Sant'Anna et al |
| EPI_ISL_1469817 | Fundação Hospitalar de Sapucaia do Sul | Epiclin | Fernando Hayashi Sant'Anna et al |
| EPI_ISL_1469551 | Secretaria Municipal de Saúde de São Leopoldo | Epiclin | Fernando Hayashi Sant'Anna et al |
| EPI_ISL_1469727 | Secretaria Municipal de Saúde de São Leopoldo | Epiclin | Fernando Hayashi Sant'Anna et al |
| EPI_ISL_1469600 | Hospital de Portão | Epiclin | Fernando Hayashi Sant'Anna et al |
| EPI_ISL_1469668 | Hospital Unimed Vale do Caí | Epiclin | Fernando Hayashi Sant'Anna et al |
| EPI_ISL_1469654 | Fundação Hospitalar de Sapucaia do Sul | Epiclin | Fernando Hayashi Sant'Anna et al |
| EPI_ISL_1469773 | Hospital Universitário | Epiclin | Fernando Hayashi Sant'Anna et al |
| EPI_ISL_1469729 | Centro de Especialidades Triunfo | Epiclin | Fernando Hayashi Sant'Anna et al |
| EPI_ISL_1469831 | Fundação Hospitalar de Sapucaia do Sul | Epiclin | Fernando Hayashi Sant'Anna et al |
| EPI_ISL_1469849 | Hospital Universitário de Canoas | Epiclin | Fernando Hayashi Sant'Anna et al |

| | | | |
|---|---|---|---|
| EPI_ISL_1469827 | Hospital de Portão | Epiclin | Fernando Hayashi Sant'Anna et al |
| EPI_ISL_1469752 | Fundação Hospitalar de Sapucaia do Sul | Epiclin | Fernando Hayashi Sant'Anna et al |
| EPI_ISL_1469844 | Hospital Universitário de Canoas | Epiclin | Fernando Hayashi Sant'Anna et al |
| EPI_ISL_1469671 | Vigilância em Saúde de Sapucaia do Sul | Epiclin | Fernando Hayashi Sant'Anna et al |
| EPI_ISL_1469724 | Fundação Hospitalar de Sapucaia do Sul | Epiclin | Fernando Hayashi Sant'Anna et al |
| EPI_ISL_1469621 | Pronto Atendimento Campo Bom | Epiclin | Fernando Hayashi Sant'Anna et al |
| EPI_ISL_1469824 | Hospital Regina | Epiclin | Fernando Hayashi Sant'Anna et al |
| EPI_ISL_1469807 | Hospital São Francisco de Assis | Epiclin | Fernando Hayashi Sant'Anna et al |
| EPI_ISL_1469585 | Fundação Hospitalar de Sapucaia do Sul | Epiclin | Fernando Hayashi Sant'Anna et al |
| EPI_ISL_1469842 | Hospital São Francisco de Assis | Epiclin | Fernando Hayashi Sant'Anna et al |
| EPI_ISL_1469837 | Fundação de Saúde Pública São Camilo de Esteio | Epiclin | Fernando Hayashi Sant'Anna et al |
| EPI_ISL_1469561 | Fundação de Saúde Pública São Camilo de Esteio | Epiclin | Fernando Hayashi Sant'Anna et al |
| EPI_ISL_1469581 | Hospital Dia e Pronto Atendimento | Epiclin | Fernando Hayashi Sant'Anna et al |
| EPI_ISL_1469847 | Secretaria Municipal de Saúde de Três Coroas | Epiclin | Fernando Hayashi Sant'Anna et al |
| EPI_ISL_1469838 | Secretaria Municipal de Saúde de Três Coroas | Epiclin | Fernando Hayashi Sant'Anna et al |
| EPI_ISL_1469666 | Secretaria Municipal de Saúde de Montenegro | Epiclin | Fernando Hayashi Sant'Anna et al |
| EPI_ISL_1469825 | Pronto Atendimento Campo Bom | Epiclin | Fernando Hayashi Sant'Anna et al |
| EPI_ISL_1469615 | Pronto Atendimento Campo Bom | Epiclin | Fernando Hayashi Sant'Anna et al |
| EPI_ISL_1469670 | Hospital São Francisco de Assis | Epiclin | Fernando Hayashi Sant'Anna et al |
| EPI_ISL_1469567 | Unidade Sanitária de Igrejinha | Epiclin | Fernando Hayashi Sant'Anna et al |
| EPI_ISL_1469830 | Hospital São Francisco de Assis | Epiclin | Fernando Hayashi Sant'Anna et al |
| EPI_ISL_1469552 | Unidade Sanitária de Igrejinha | Epiclin | Fernando Hayashi Sant'Anna et al |
| EPI_ISL_1469569 | Fundação de Saúde Pública São Camilo de Esteio | Epiclin | Fernando Hayashi Sant'Anna et al |
| EPI_ISL_1469771 | Hospital de Portão | Epiclin | Fernando Hayashi Sant'Anna et al |
| EPI_ISL_1469667 | Centro de Referência em Síndromes Gripais | Epiclin | Fernando Hayashi Sant'Anna et al |
| EPI_ISL_1469794 | Hospital Dia e Pronto Atendimento | Epiclin | Fernando Hayashi Sant'Anna et al |
| EPI_ISL_1469835 | Diretoria de Vigilância em Saúde | Epiclin | Fernando Hayashi Sant'Anna et al |
| EPI_ISL_1469609 | Diretoria de Vigilância em Saúde | Epiclin | Fernando Hayashi Sant'Anna et al |
| EPI_ISL_1469769 | Diretoria de Vigilância em Saúde | Epiclin | Fernando Hayashi Sant'Anna et al |
| EPI_ISL_1469560 | Diretoria de Vigilância em Saúde | Epiclin | Fernando Hayashi Sant'Anna et al |

| | | | |
|---|---|---|---|
| EPI_ISL_1469557 | Centro de Serviços Especializados Santa Rita | Epiclin | Fernando Hayashi Sant’Anna et al |
| EPI_ISL_1469790 | Unidade de Pronto Atendimento de Sapucaia do Sul | Epiclin | Fernando Hayashi Sant’Anna et al |
| EPI_ISL_1469602 | Pronto Atendimento Campo Bom | Epiclin | Fernando Hayashi Sant’Anna et al |
| EPI_ISL_1469614 | Unidade Sanitária de Igrejinha | Epiclin | Fernando Hayashi Sant’Anna et al |
| EPI_ISL_1469562 | Hospital Dia e Pronto Atendimento | Epiclin | Fernando Hayashi Sant’Anna et al |
| EPI_ISL_1469616 | Diretoria de Vigilância em Saúde | Epiclin | Fernando Hayashi Sant’Anna et al |
| EPI_ISL_1469850 | Fundação de Saúde Pública São Camilo de Esteio | Epiclin | Fernando Hayashi Sant’Anna et al |
| EPI_ISL_1469809 | Fundação Hospitalar de Sapucaia do Sul | Epiclin | Fernando Hayashi Sant’Anna et al |
| EPI_ISL_1469845 | Fundação Hospitalar de Sapucaia do Sul | Epiclin | Fernando Hayashi Sant’Anna et al |
| EPI_ISL_1469582 | Unidade de Pronto Atendimento de Sapucaia do Sul | Epiclin | Fernando Hayashi Sant’Anna et al |
| EPI_ISL_1469810 | Unidade de Pronto Atendimento de Sapucaia do Sul | Epiclin | Fernando Hayashi Sant’Anna et al |
| EPI_ISL_1469647 | Secretaria Municipal de Saúde de Taquara | Epiclin | Fernando Hayashi Sant’Anna et al |
| EPI_ISL_1469576 | Unidade Sanitária de Igrejinha | Epiclin | Fernando Hayashi Sant’Anna et al |
| EPI_ISL_1479120 | Diretoria de Vigilância em Saúde | Epiclin | Fernando Hayashi Sant’Anna et al |
| EPI_ISL_1469669 | Diretoria de Vigilância em Saúde | Epiclin | Fernando Hayashi Sant’Anna et al |
| EPI_ISL_1469623 | Diretoria de Vigilância em Saúde | Epiclin | Fernando Hayashi Sant’Anna et al |
| EPI_ISL_1469619 | Fundação Hospitalar de Sapucaia do Sul | Epiclin | Fernando Hayashi Sant’Anna et al |
| EPI_ISL_1469707 | Hospital Bom Jesus | Epiclin | Fernando Hayashi Sant’Anna et al |
| EPI_ISL_1469811 | Hospital Municipal Getúlio Vargas | Epiclin | Fernando Hayashi Sant’Anna et al |
| EPI_ISL_1469556 | Secretaria Municipal de Saúde de São Leopoldo | Epiclin | Fernando Hayashi Sant’Anna et al |
| EPI_ISL_1469617 | Fundação Hospitalar de Sapucaia do Sul | Epiclin | Fernando Hayashi Sant’Anna et al |
| EPI_ISL_1469618 | Diretoria de Vigilância em Saúde | Epiclin | Fernando Hayashi Sant’Anna et al |
| EPI_ISL_1469820 | Hospital Sapiranga | Epiclin | Fernando Hayashi Sant’Anna et al |
| EPI_ISL_1479127 | Hospital Regina | Epiclin | Fernando Hayashi Sant’Anna et al |
| EPI_ISL_1469795 | Hospital Dia e Pronto Atendimento | Epiclin | Fernando Hayashi Sant’Anna et al |
| EPI_ISL_1469818 | Hospital São Francisco de Assis | Epiclin | Fernando Hayashi Sant’Anna et al |
| EPI_ISL_1469595 | Fundação de Saúde Pública de Novo Hamburgo | Epiclin | Fernando Hayashi Sant’Anna et al |
| EPI_ISL_1469741 | Centro de Referência em Síndromes Gripais | Epiclin | Fernando Hayashi Sant’Anna et al |
| EPI_ISL_1469800 | Fundação de Saúde Pública de Novo Hamburgo | Epiclin | Fernando Hayashi Sant’Anna et al |
| EPI_ISL_1469563 | Unidade Básica de Saúde de Riozinho | Epiclin | Fernando Hayashi Sant’Anna et al |

| | | | |
|---|---|---|---|
| EPI_ISL_1469797 | Diretoria de Vigilância em Saúde | Epiclin | Fernando Hayashi Sant'Anna et al |
| EPI_ISL_1469793 | Hospital Universitário de Canoas | Epiclin | Fernando Hayashi Sant'Anna et al |
| EPI_ISL_1469650 | Diretoria de Vigilância em Saúde | Epiclin | Fernando Hayashi Sant'Anna et al |
| EPI_ISL_1469653 | Fundação de Saúde Pública de Novo Hamburgo | Epiclin | Fernando Hayashi Sant'Anna et al |
| EPI_ISL_1469696 | Vigilância em Saúde de Sapucaia do Sul | Epiclin | Fernando Hayashi Sant'Anna et al |
| EPI_ISL_1469605 | Diretoria de Vigilância em Saúde | Epiclin | Fernando Hayashi Sant'Anna et al |
| EPI_ISL_1469713 | Fundação Hospitalar de Sapucaia do Sul | Epiclin | Fernando Hayashi Sant'Anna et al |
| EPI_ISL_1469607 | Secretaria Municipal de Saúde de Três Coroas | Epiclin | Fernando Hayashi Sant'Anna et al |
| EPI_ISL_1469789 | Hospital São Francisco de Assis | Epiclin | Fernando Hayashi Sant'Anna et al |
| EPI_ISL_1469783 | Diretoria de Vigilância em Saúde | Epiclin | Fernando Hayashi Sant'Anna et al |
| EPI_ISL_1469756 | Diretoria de Vigilância em Saúde | Epiclin | Fernando Hayashi Sant'Anna et al |
| EPI_ISL_1469593 | Hospital Sapiranga | Epiclin | Fernando Hayashi Sant'Anna et al |
| EPI_ISL_1469749 | Centro de Referência em Síndromes Gripais | Epiclin | Fernando Hayashi Sant'Anna et al |
| EPI_ISL_1469770 | Fundação de Saúde Pública de Novo Hamburgo | Epiclin | Fernando Hayashi Sant'Anna et al |
| EPI_ISL_1469711 | Hospital Dia e Pronto Atendimento | Epiclin | Fernando Hayashi Sant'Anna et al |
| EPI_ISL_1469803 | Secretaria Municipal de Saúde de Três Coroas | Epiclin | Fernando Hayashi Sant'Anna et al |
| EPI_ISL_1469786 | Diretoria de Vigilância em Saúde | Epiclin | Fernando Hayashi Sant'Anna et al |
| EPI_ISL_1479129 | Pronto Atendimento Campo Bom | Epiclin | Fernando Hayashi Sant'Anna et al |
| EPI_ISL_1469550 | Vigilância em Saúde de Sapucaia do Sul | Epiclin | Fernando Hayashi Sant'Anna et al |
| EPI_ISL_1469777 | Centro de Referência em Síndromes Gripais | Epiclin | Fernando Hayashi Sant'Anna et al |
| EPI_ISL_1469714 | Unidade de Atendimento DST AIDS TB e Han | Epiclin | Fernando Hayashi Sant'Anna et al |
| EPI_ISL_1469554 | Fundação de Saúde Pública São Camilo de Esteio | Epiclin | Fernando Hayashi Sant'Anna et al |
| EPI_ISL_1469778 | Hospital Municipal Getúlio Vargas | Epiclin | Fernando Hayashi Sant'Anna et al |
| EPI_ISL_1469846 | Hospital Nossa Senhora das Graças | Epiclin | Fernando Hayashi Sant'Anna et al |
| EPI_ISL_1469821 | Centro de Saúde Bruno Cassel | Epiclin | Fernando Hayashi Sant'Anna et al |
| EPI_ISL_1469736 | SECRETARIA MUNICIPAL DE SAUDE DE SAO LEOPOLDO | Epiclin | Fernando Hayashi Sant'Anna et al |
| EPI_ISL_1469575 | HOSPITAL SAO FRANCISCO DE ASSIS | Epiclin | Fernando Hayashi Sant'Anna et al |
| EPI_ISL_1469663 | DIRETORIA DE VIGILANCIA EM SAUDE | Epiclin | Fernando Hayashi Sant'Anna et al |
| EPI_ISL_1469590 | DIRETORIA DE VIGILANCIA EM SAUDE | Epiclin | Fernando Hayashi Sant'Anna et al |
| EPI_ISL_1469681 | DIRETORIA DE VIGILANCIA EM SAUDE | Epiclin | Fernando Hayashi Sant'Anna et al |

| | | | |
|---|---|---|---|
| EPI_ISL_1469751 | UNIDADE BASICA DE SAUDE DE RIOZINHO | Epiclin | Fernando Hayashi Sant'Anna et al |
| EPI_ISL_1469826 | Fundação de Saúde Pública de Novo Hamburgo | Epiclin | Fernando Hayashi Sant'Anna et al |
| EPI_ISL_1469659 | DIRETORIA DE VIGILANCIA EM SAUDE | Epiclin | Fernando Hayashi Sant'Anna et al |
| EPI_ISL_1469767 | Fundação de Saúde Pública de Novo Hamburgo | Epiclin | Fernando Hayashi Sant'Anna et al |
| EPI_ISL_1469558 | DIRETORIA DE VIGILANCIA EM SAUDE | Epiclin | Fernando Hayashi Sant'Anna et al |
| EPI_ISL_1469804 | UNIDADE DE PRONTO ATENDIMENTO DE SAPUCAIA DO SUL UPA | Epiclin | Fernando Hayashi Sant'Anna et al |
| EPI_ISL_1469583 | UNIDADE SANITARIA DE IGREJINHA | Epiclin | Fernando Hayashi Sant'Anna et al |
| EPI_ISL_1469792 | Diretoria de Vigilância em Saúde | Epiclin | Fernando Hayashi Sant'Anna et al |
| EPI_ISL_1469708 | Diretoria de Vigilância em Saúde | Epiclin | Fernando Hayashi Sant'Anna et al |
| EPI_ISL_1469731 | DIRETORIA DE VIGILANCIA EM SAUDE | Epiclin | Fernando Hayashi Sant'Anna et al |
| EPI_ISL_1469784 | DIRETORIA DE VIGILANCIA EM SAUDE | Epiclin | Fernando Hayashi Sant'Anna et al |
| EPI_ISL_1469813 | HOSPITAL BOM JESUS | Epiclin | Fernando Hayashi Sant'Anna et al |
| EPI_ISL_1469664 | Unidade de Atendimento DST AIDS TB e Han | Epiclin | Fernando Hayashi Sant'Anna et al |
| EPI_ISL_1469662 | CENTRO DE SAUDE DR BRUNO CASSEL | Epiclin | Fernando Hayashi Sant'Anna et al |
| EPI_ISL_1469594 | DIRETORIA DE VIGILANCIA EM SAUDE | Epiclin | Fernando Hayashi Sant'Anna et al |
| EPI_ISL_1469684 | DIRETORIA DE VIGILANCIA EM SAUDE | Epiclin | Fernando Hayashi Sant'Anna et al |
| EPI_ISL_1469620 | FUNDACAO DE SAUDE PUBLICA DE NOVO HAMBURGO FSNH | Epiclin | Fernando Hayashi Sant'Anna et al |
| EPI_ISL_1469571 | Unidade de Pronto Atendimento de Sapucaia do Sul | Epiclin | Fernando Hayashi Sant'Anna et al |
| EPI_ISL_1469698 | CENTRO DE SERVICOS ESPECIALIZADOS SANTA RITA DE CASSIA | Epiclin | Fernando Hayashi Sant'Anna et al |
| EPI_ISL_1469596 | CENTRO DE REFERENCIA EM SINDROMES GRIPAIS | Epiclin | Fernando Hayashi Sant'Anna et al |
| EPI_ISL_1469655 | Centro Municipal de Saúde de Rolante | Epiclin | Fernando Hayashi Sant'Anna et al |
| EPI_ISL_1469734 | Hospital Dia e Pronto Atendimento | Epiclin | Fernando Hayashi Sant'Anna et al |
| EPI_ISL_1469676 | HOSPITAL SAO FRANCISCO DE ASSIS | Epiclin | Fernando Hayashi Sant'Anna et al |
| EPI_ISL_1469692 | DIRETORIA DE VIGILANCIA EM SAUDE | Epiclin | Fernando Hayashi Sant'Anna et al |
| EPI_ISL_1469679 | DIRETORIA DE VIGILANCIA EM SAUDE | Epiclin | Fernando Hayashi Sant'Anna et al |
| EPI_ISL_1469629 | CENTRO MUNICIPAL DE SAUDE DE ROLANTE | Epiclin | Fernando Hayashi Sant'Anna et al |
| EPI_ISL_1469685 | HOSPITAL SAO FRANCISCO DE ASSIS | Epiclin | Fernando Hayashi Sant'Anna et al |
| EPI_ISL_1469686 | DIRETORIA DE VIGILANCIA EM SAUDE | Epiclin | Fernando Hayashi Sant'Anna et al |
| EPI_ISL_1469710 | DIRETORIA DE VIGILANCIA EM SAUDE | Epiclin | Fernando Hayashi Sant'Anna et al |
| EPI_ISL_1469579 | DIRETORIA DE VIGILANCIA EM SAUDE | Epiclin | Fernando Hayashi Sant'Anna et al |

| | | | |
|---|---|---|---|
| EPI_ISL_1469680 | UNIDADE DE ATENDIMENTO DST AIDS TB E HAN | Epiclin | Fernando Hayashi Sant'Anna et al |
| EPI_ISL_1479126 | UNIDADE DE ATENDIMENTO DST AIDS TB E HAN | Epiclin | Fernando Hayashi Sant'Anna et al |
| EPI_ISL_1469572 | FUNDACAO DE SAUDE PUBLICA DE NOVO HAMBURGO FSNH | Epiclin | Fernando Hayashi Sant'Anna et al |
| EPI_ISL_1469754 | Hospital Municipal Getúlio Vargas | Epiclin | Fernando Hayashi Sant'Anna et al |
| EPI_ISL_1469768 | UNIDADE DE PRONTO ATENDIMENTO DE SAPUCAIA DO SUL UPA | Epiclin | Fernando Hayashi Sant'Anna et al |
| EPI_ISL_1469651 | FUNDACAO DE SAUDE PUBLICA DE NOVO HAMBURGO FSNH | Epiclin | Fernando Hayashi Sant'Anna et al |
| EPI_ISL_1469566 | DIRETORIA DE VIGILANCIA EM SAUDE | Epiclin | Fernando Hayashi Sant'Anna et al |
| EPI_ISL_1469775 | Diretoria de Vigilância em Saúde | Epiclin | Fernando Hayashi Sant'Anna et al |
| EPI_ISL_1469791 | DIRETORIA DE VIGILANCIA EM SAUDE | Epiclin | Fernando Hayashi Sant'Anna et al |
| EPI_ISL_1469603 | Centro de Referência em Síndromes Gripais | Epiclin | Fernando Hayashi Sant'Anna et al |
| EPI_ISL_1469753 | Diretoria de Vigilância em Saúde | Epiclin | Fernando Hayashi Sant'Anna et al |
| EPI_ISL_1469592 | Diretoria de Vigilância em Saúde | Epiclin | Fernando Hayashi Sant'Anna et al |
| EPI_ISL_1469633 | DIRETORIA DE VIGILANCIA EM SAUDE | Epiclin | Fernando Hayashi Sant'Anna et al |
| EPI_ISL_1469584 | CENTRO MUNICIPAL DE SAUDE DE ROLANTE | Epiclin | Fernando Hayashi Sant'Anna et al |
| EPI_ISL_1469549 | FUNDACAO HOSPITALAR DE SAPUCAIA DO SUL | Epiclin | Fernando Hayashi Sant'Anna et al |
| EPI_ISL_1469673 | Fundação de Saúde Pública de Novo Hamburgo | Epiclin | Fernando Hayashi Sant'Anna et al |
| EPI_ISL_1469674 | DIRETORIA DE VIGILANCIA EM SAUDE | Epiclin | Fernando Hayashi Sant'Anna et al |
| EPI_ISL_1479123 | Fundação de Saúde Pública de Novo Hamburgo | Epiclin | Fernando Hayashi Sant'Anna et al |
| EPI_ISL_1469630 | FUNDACAO DE SAUDE PUBLICA DE NOVO HAMBURGO FSNH | Epiclin | Fernando Hayashi Sant'Anna et al |
| EPI_ISL_1469578 | HOSPITAL SAO FRANCISCO DE ASSIS | Epiclin | Fernando Hayashi Sant'Anna et al |
| EPI_ISL_1469742 | CENTRO DE REFERENCIA EM SINDROMES GRIPAIS | Epiclin | Fernando Hayashi Sant'Anna et al |
| EPI_ISL_1469688 | DIRETORIA DE VIGILANCIA EM SAUDE | Epiclin | Fernando Hayashi Sant'Anna et al |
| EPI_ISL_1469725 | DIRETORIA DE VIGILANCIA EM SAUDE | Epiclin | Fernando Hayashi Sant'Anna et al |
| EPI_ISL_1469702 | Diretoria de Vigilância em Saúde | Epiclin | Fernando Hayashi Sant'Anna et al |
| EPI_ISL_1469761 | Diretoria de Vigilância em Saúde | Epiclin | Fernando Hayashi Sant'Anna et al |
| EPI_ISL_1469694 | UNIDADE BASICA DE SAUDE PARECI NOVO | Epiclin | Fernando Hayashi Sant'Anna et al |
| EPI_ISL_1469836 | Diretoria de Vigilância em Saúde | Epiclin | Fernando Hayashi Sant'Anna et al |
| EPI_ISL_1469705 | HOSPITAL DE CAMPO BOM DR LAURO REUS | Epiclin | Fernando Hayashi Sant'Anna et al |
| EPI_ISL_1479133 | HOSPITAL SAO FRANCISCO DE ASSIS | Epiclin | Fernando Hayashi Sant'Anna et al |
| EPI_ISL_1469631 | HOSPITAL MUNICIPAL GETULIO VARGAS | Epiclin | Fernando Hayashi Sant'Anna et al |

| | | | |
|---|---|---|---|
| EPI_ISL_1469738 | HOSPITAL SAO FRANCISCO DE ASSIS | Epiclin | Fernando Hayashi Sant'Anna et al |
| EPI_ISL_1469604 | COORDENADORIA GERAL DE VIGILANCIA EM SAUDE | Epiclin | Fernando Hayashi Sant'Anna et al |
| EPI_ISL_1469745 | UNIDADE DE PRONTO ATENDIMENTO DE SAPUCAIA DO SUL UPA | Epiclin | Fernando Hayashi Sant'Anna et al |
| EPI_ISL_1469687 | SECRETARIA MUNICIPAL DE SAUDE DE TAQUARA | Epiclin | Fernando Hayashi Sant'Anna et al |
| EPI_ISL_1469781 | DIRETORIA DE VIGILANCIA EM SAUDE | Epiclin | Fernando Hayashi Sant'Anna et al |
| EPI_ISL_1469661 | FUNDACAO DE SAUDE PUBLICA DE NOVO HAMBURGO FSNH | Epiclin | Fernando Hayashi Sant'Anna et al |
| EPI_ISL_1469726 | HOSPITAL SAPIRANGA | Epiclin | Fernando Hayashi Sant'Anna et al |
| EPI_ISL_1479125 | CENTRO DE REFERENCIA EM SINDROMES GRIPAIS | Epiclin | Fernando Hayashi Sant'Anna et al |
| EPI_ISL_1469802 | Diretoria de Vigilância em Saúde | Epiclin | Fernando Hayashi Sant'Anna et al |
| EPI_ISL_1469785 | FUNDACAO DE SAUDE PUBLICA DE NOVO HAMBURGO FSNH | Epiclin | Fernando Hayashi Sant'Anna et al |
| EPI_ISL_1195293 | SECRETARIA MUNICIPAL DE SAUDE DE TAQUARA | Epiclin | Fernando Hayashi Sant'Anna et al |
| EPI_ISL_1469709 | SECRETARIA MUNICIPAL DE SAUDE DE TAQUARA | Epiclin | Fernando Hayashi Sant'Anna et al |
| EPI_ISL_1469646 | UNIDADE DE PRONTO ATENDIMENTO DE SAPUCAIA DO SUL UPA | Epiclin | Fernando Hayashi Sant'Anna et al |
| EPI_ISL_1479128 | HOSPITAL SAO FRANCISCO DE ASSIS | Epiclin | Fernando Hayashi Sant'Anna et al |
| EPI_ISL_1469580 | Unidade de Pronto Atendimento de Sapucaia do Sul | Epiclin | Fernando Hayashi Sant'Anna et al |
| EPI_ISL_1469766 | Coordenadoria Geral de Vigilância em Saúde · Vigilância em Saúde | Epiclin | Fernando Hayashi Sant'Anna et al |
| EPI_ISL_1469635 | COORDENADORIA GERAL DE VIGILANCIA EM SAUDE | Epiclin | Fernando Hayashi Sant'Anna et al |
| EPI_ISL_1469677 | DIRETORIA DE VIGILANCIA EM SAUDE | Epiclin | Fernando Hayashi Sant'Anna et al |
| EPI_ISL_1469589 | UNIDADE DE PRONTO ATENDIMENTO DE SAPUCAIA DO SUL UPA | Epiclin | Fernando Hayashi Sant'Anna et al |
| EPI_ISL_1469665 | FUNDACAO DE SAUDE PUBLICA SAO CAMILO DE ESTEIO | Epiclin | Fernando Hayashi Sant'Anna et al |
| EPI_ISL_1469814 | SECRETARIA MUNICIPAL DE SAUDE DE SAO LEOPOLDO | Epiclin | Fernando Hayashi Sant'Anna et al |
| EPI_ISL_1469690 | SECRETARIA MUNICIPAL DE SAUDE DE SAO LEOPOLDO | Epiclin | Fernando Hayashi Sant'Anna et al |
| EPI_ISL_1469624 | HOSPITAL MUNICIPAL GETULIO VARGAS | Epiclin | Fernando Hayashi Sant'Anna et al |
| EPI_ISL_1469683 | SECRETARIA MUNICIPAL DE SAUDE DE TAQUARA | Epiclin | Fernando Hayashi Sant'Anna et al |
| EPI_ISL_1469675 | DIRETORIA DE VIGILANCIA EM SAUDE | Epiclin | Fernando Hayashi Sant'Anna et al |
| EPI_ISL_1469645 | SECRETARIA MUNICIPAL DE SAUDE DE TRES COROAS | Epiclin | Fernando Hayashi Sant'Anna et al |
| EPI_ISL_1469637 | DIRETORIA DE VIGILANCIA EM SAUDE | Epiclin | Fernando Hayashi Sant'Anna et al |
| EPI_ISL_1469755 | Hospital São Francisco de Assis | Epiclin | Fernando Hayashi Sant'Anna et al |
| EPI_ISL_1469648 | HOSPITAL SAO FRANCISCO DE ASSIS | Epiclin | Fernando Hayashi Sant'Anna et al |
| EPI_ISL_1469642 | CENTRO DE REFERENCIA EM SINDROMES GRIPAIS | Epiclin | Fernando Hayashi Sant'Anna et al |

| | | | |
|---|---|---|---|
| EPI_ISL_1469823 | Diretoria de Vigilância em Saúde | Epiclin | Fernando Hayashi Sant'Anna et al |
| EPI_ISL_1479130 | SECRETARIA MUNICIPAL DE SAUDE DE TAQUARA | Epiclin | Fernando Hayashi Sant'Anna et al |
| EPI_ISL_1469628 | CENTRO DE SERVICOS ESPECIALIZADOS SANTA RITA DE CASSIA | Epiclin | Fernando Hayashi Sant'Anna et al |
| EPI_ISL_1469573 | VIGILANCIA EM SAUDE NH | Epiclin | Fernando Hayashi Sant'Anna et al |
| EPI_ISL_1469733 | UNIDADE DE PRONTO ATENDIMENTO DE SAPUCAIA DO SUL UPA | Epiclin | Fernando Hayashi Sant'Anna et al |
| EPI_ISL_1469735 | COORDENADORIA GERAL DE VIGILANCIA EM SAUDE | Epiclin | Fernando Hayashi Sant'Anna et al |
| EPI_ISL_1469715 | COORDENADORIA GERAL DE VIGILANCIA EM SAUDE | Epiclin | Fernando Hayashi Sant'Anna et al |
| EPI_ISL_1469782 | COORDENADORIA GERAL DE VIGILANCIA EM SAUDE | Epiclin | Fernando Hayashi Sant'Anna et al |
| EPI_ISL_1469601 | SECRETARIA MUNICIPAL DE SAUDE DE TAQUARA | Epiclin | Fernando Hayashi Sant'Anna et al |
| EPI_ISL_1469730 | CENTRO DE REFERENCIA EM SINDROMES GRIPAIS | Epiclin | Fernando Hayashi Sant'Anna et al |
| EPI_ISL_1469701 | Hospital Sapiranga | Epiclin | Fernando Hayashi Sant'Anna et al |
| EPI_ISL_1469611 | DIRETORIA DE VIGILANCIA EM SAUDE | Epiclin | Fernando Hayashi Sant'Anna et al |
| EPI_ISL_1469765 | UNIDADE SANITARIA DE IGREJINHA | Epiclin | Fernando Hayashi Sant'Anna et al |
| EPI_ISL_1469722 | UNIDADE SANITARIA DE IGREJINHA | Epiclin | Fernando Hayashi Sant'Anna et al |
| EPI_ISL_1469759 | HOSPITAL MUNICIPAL GETULIO VARGAS | Epiclin | Fernando Hayashi Sant'Anna et al |
| EPI_ISL_1469691 | Fundação de Saúde Pública São Camilo de Esteio | Epiclin | Fernando Hayashi Sant'Anna et al |
| EPI_ISL_1479121 | DIRETORIA DE VIGILANCIA EM SAUDE | Epiclin | Fernando Hayashi Sant'Anna et al |
| EPI_ISL_1469632 | Diretoria de Vigilância em Saúde | Epiclin | Fernando Hayashi Sant'Anna et al |
| EPI_ISL_1469586 | FUNDACAO HOSPITALAR SAO JOSE | Epiclin | Fernando Hayashi Sant'Anna et al |
| EPI_ISL_1469643 | HOSPITAL SAO FRANCISCO DE ASSIS | Epiclin | Fernando Hayashi Sant'Anna et al |
| EPI_ISL_1195273 | FUNDACAO DE SAUDE PUBLICA SAO CAMILO DE ESTEIO | Epiclin | Fernando Hayashi Sant'Anna et al |
| EPI_ISL_1479131 | Fundação de Saúde Pública de Novo Hamburgo | Epiclin | Fernando Hayashi Sant'Anna et al |
| EPI_ISL_1469721 | Pronto Atendimento Cruzeiro do Sul | Epiclin | Fernando Hayashi Sant'Anna et al |
| EPI_ISL_1479132 | Fundação de Saúde Pública de Novo Hamburgo | Epiclin | Fernando Hayashi Sant'Anna et al |
| EPI_ISL_1479119 | DIRETORIA DE VIGILANCIA EM SAUDE | Epiclin | Fernando Hayashi Sant'Anna et al |
| EPI_ISL_1195279 | CENTRO DE REFERENCIA EM SINDROMES GRIPAIS | Epiclin | Fernando Hayashi Sant'Anna et al |
| EPI_ISL_1195280 | SECRETARIA MUNICIPAL DE SAUDE DE ARARICA | Epiclin | Fernando Hayashi Sant'Anna et al |
| EPI_ISL_1479124 | UNIDADE DE SAUDE NOVA HARTZ | Epiclin | Fernando Hayashi Sant'Anna et al |
| EPI_ISL_1195292 | SECRETARIA MUNICIPAL DE SAUDE DE TRES COROAS | Epiclin | Fernando Hayashi Sant'Anna et al |
| EPI_ISL_1469720 | UNIDADE SANITARIA DE IGREJINHA | Epiclin | Fernando Hayashi Sant'Anna et al |

| | | | |
|---|---|---|---|
| EPI_ISL_1469574 | CENTRO DE SERVICOS ESPECIALIZADOS SANTA RITA DE CASSIA | Epiclin | Fernando Hayashi Sant'Anna et al |
| EPI_ISL_1469652 | DIRETORIA DE VIGILANCIA EM SAUDE | Epiclin | Fernando Hayashi Sant'Anna et al |
| EPI_ISL_1469612 | FUNDACAO DE SAUDE PUBLICA DE NOVO HAMBURGO FSNH | Epiclin | Fernando Hayashi Sant'Anna et al |
| EPI_ISL_1195291 | DIRETORIA DE VIGILANCIA EM SAUDE | Epiclin | Fernando Hayashi Sant'Anna et al |
| EPI_ISL_1469723 | FUNDACAO DE SAUDE PUBLICA SAO CAMILO DE ESTEIO | Epiclin | Fernando Hayashi Sant'Anna et al |
| EPI_ISL_1469772 | DIRETORIA DE VIGILANCIA EM SAUDE | Epiclin | Fernando Hayashi Sant'Anna et al |
| EPI_ISL_1469848 | Diretoria de Vigilância em Saúde | Epiclin | Fernando Hayashi Sant'Anna et al |
| EPI_ISL_1195290 | SECRETARIA MUNICIPAL DE SAUDE DE TRES COROAS | Epiclin | Fernando Hayashi Sant'Anna et al |
| EPI_ISL_1469798 | DIRETORIA DE VIGILANCIA EM SAUDE | Epiclin | Fernando Hayashi Sant'Anna et al |
| EPI_ISL_1469764 | DIRETORIA DE VIGILANCIA EM SAUDE | Epiclin | Fernando Hayashi Sant'Anna et al |
| EPI_ISL_1469760 | DIRETORIA DE VIGILANCIA EM SAUDE | Epiclin | Fernando Hayashi Sant'Anna et al |
| EPI_ISL_1469699 | HOSPITAL MUNICIPAL GETULIO VARGAS | Epiclin | Fernando Hayashi Sant'Anna et al |
| EPI_ISL_1469834 | HOSPITAL SAO FRANCISCO DE ASSIS | Epiclin | Fernando Hayashi Sant'Anna et al |
| EPI_ISL_1195278 | DIRETORIA DE VIGILANCIA EM SAUDE | Epiclin | Fernando Hayashi Sant'Anna et al |
| EPI_ISL_1469740 | SECRETARIA MUNICIPAL DE SAUDE DE TRES COROAS | Epiclin | Fernando Hayashi Sant'Anna et al |
| EPI_ISL_1469568 | SECRETARIA MUNICIPAL DE SAUDE DE SAO LEOPOLDO | Epiclin | Fernando Hayashi Sant'Anna et al |
| EPI_ISL_1469555 | FUNDACAO DE SAUDE PUBLICA DE NOVO HAMBURGO FSNH | Epiclin | Fernando Hayashi Sant'Anna et al |
| EPI_ISL_1195289 | SECRETARIA MUNICIPAL DE SAUDE DE SAO LEOPOLDO | Epiclin | Fernando Hayashi Sant'Anna et al |
| EPI_ISL_1469719 | UNIDADE DE PRONTO ATENDIMENTO DE SAPUCAIA DO SUL UPA | Epiclin | Fernando Hayashi Sant'Anna et al |
| EPI_ISL_1469822 | Hospital São Francisco de Assis | Epiclin | Fernando Hayashi Sant'Anna et al |
| EPI_ISL_1469839 | HOSPITAL DE CAMPO BOM DR LAURO REUS | Epiclin | Fernando Hayashi Sant'Anna et al |
| EPI_ISL_1469704 | FUNDACAO DE SAUDE PUBLICA SAO CAMILO DE ESTEIO | Epiclin | Fernando Hayashi Sant'Anna et al |
| EPI_ISL_1469840 | Hospital Sapiranga | Epiclin | Fernando Hayashi Sant'Anna et al |
| EPI_ISL_1195288 | DIRETORIA DE VIGILANCIA EM SAUDE | Epiclin | Fernando Hayashi Sant'Anna et al |
| EPI_ISL_1469638 | DIRETORIA DE VIGILANCIA EM SAUDE | Epiclin | Fernando Hayashi Sant'Anna et al |
| EPI_ISL_1469743 | DIRETORIA DE VIGILANCIA EM SAUDE | Epiclin | Fernando Hayashi Sant'Anna et al |
| EPI_ISL_1469832 | Diretoria de Vigilância em Saúde | Epiclin | Fernando Hayashi Sant'Anna et al |
| EPI_ISL_1195277 | Unidade de Atendimento DST AIDS TB e Han | Epiclin | Fernando Hayashi Sant'Anna et al |
| EPI_ISL_1469774 | Hospital Universitário de Canoas | Epiclin | Fernando Hayashi Sant'Anna et al |
| EPI_ISL_1469553 | Diretoria de Vigilância em Saúde | Epiclin | Fernando Hayashi Sant'Anna et al |

| | | | |
|---|---|---|---|
| EPI_ISL_1469808 | Diretoria de Vigilância em Saúde | Epiclin | Fernando Hayashi Sant'Anna et al |
| EPI_ISL_1469689 | FUNDACAO DE SAUDE PUBLICA SAO CAMILO DE ESTEIO | Epiclin | Fernando Hayashi Sant'Anna et al |
| EPI_ISL_1195287 | SECRETARIA MUNICIPAL DE SAUDE DE SAO LEOPOLDO | Epiclin | Fernando Hayashi Sant'Anna et al |
| EPI_ISL_1195274 | SECRETARIA MUNICIPAL DE SAUDE DE TRES COROAS | Epiclin | Fernando Hayashi Sant'Anna et al |
| EPI_ISL_1469658 | CENTRO DE ESPECIALIDADES TRIUNFO | Epiclin | Fernando Hayashi Sant'Anna et al |
| EPI_ISL_1469788 | SECRETARIA MUNICIPAL DE SAUDE DE SAO LEOPOLDO | Epiclin | Fernando Hayashi Sant'Anna et al |
| EPI_ISL_1469660 | UNIDADE BASICA DE SAUDE DE RIOZINHO | Epiclin | Fernando Hayashi Sant'Anna et al |
| EPI_ISL_1469815 | Hospital Universitário | Epiclin | Fernando Hayashi Sant'Anna et al |
| EPI_ISL_1469776 | Centro Municipal de Saúde de Rolante | Epiclin | Fernando Hayashi Sant'Anna et al |
| EPI_ISL_1469634 | Diretoria de Vigilância em Saúde | Epiclin | Fernando Hayashi Sant'Anna et al |
| EPI_ISL_1469570 | Hospital Universitário | Epiclin | Fernando Hayashi Sant'Anna et al |
| EPI_ISL_1195284 | Centro de Especialidades Triunfo | Epiclin | Fernando Hayashi Sant'Anna et al |
| EPI_ISL_1469706 | CENTRO DE SAUDE DR BRUNO CASSEL | Epiclin | Fernando Hayashi Sant'Anna et al |
| EPI_ISL_1469757 | Secretaria Municipal de Saúde de São Leopoldo | Epiclin | Fernando Hayashi Sant'Anna et al |
| EPI_ISL_1469639 | SECRETARIA MUNICIPAL DE SAUDE DE SAO LEOPOLDO | Epiclin | Fernando Hayashi Sant'Anna et al |
| EPI_ISL_1195285 | SECRETARIA MUNICIPAL DE SAUDE DE TRES COROAS | Epiclin | Fernando Hayashi Sant'Anna et al |
| EPI_ISL_1195286 | SECRETARIA MUNICIPAL DE SAUDE DE TRES COROAS | Epiclin | Fernando Hayashi Sant'Anna et al |
| EPI_ISL_1469641 | COORDENADORIA GERAL DE VIGILANCIA EM SAUDE | Epiclin | Fernando Hayashi Sant'Anna et al |
| EPI_ISL_1469636 | COORDENADORIA GERAL DE VIGILANCIA EM SAUDE | Epiclin | Fernando Hayashi Sant'Anna et al |
| EPI_ISL_1469608 | COORDENADORIA GERAL DE VIGILANCIA EM SAUDE | Epiclin | Fernando Hayashi Sant'Anna et al |
| EPI_ISL_1195282 | UNIDADE SANITARIA DE IGREJINHA | Epiclin | Fernando Hayashi Sant'Anna et al |
| EPI_ISL_1195272 | FUNDACAO HOSPITALAR DE SAPUCAIA DO SUL | Epiclin | Fernando Hayashi Sant'Anna et al |
| EPI_ISL_1469693 | FUNDACAO DE SAUDE PUBLICA SAO CAMILO DE ESTEIO | Epiclin | Fernando Hayashi Sant'Anna et al |
| EPI_ISL_1195281 | FUNDACAO DE SAUDE PUBLICA SAO CAMILO DE ESTEIO | Epiclin | Fernando Hayashi Sant'Anna et al |
| EPI_ISL_1469746 | HOSPITAL MONTENEGRO | Epiclin | Fernando Hayashi Sant'Anna et al |
| EPI_ISL_1195275 | CENTRO DE REFERENCIA EM SINDROMES GRIPAIS | Epiclin | Fernando Hayashi Sant'Anna et al |
| EPI_ISL_1195276 | CENTRO DE REFERENCIA EM SINDROMES GRIPAIS | Epiclin | Fernando Hayashi Sant'Anna et al |
| EPI_ISL_1469737 | CENTRO DE REFERENCIA EM SINDROMES GRIPAIS | Epiclin | Fernando Hayashi Sant'Anna et al |
| EPI_ISL_1469732 | CENTRO DE REFERENCIA EM SINDROMES GRIPAIS | Epiclin | Fernando Hayashi Sant'Anna et al |
| EPI_ISL_1469682 | CENTRO DE SERVICOS ESPECIALIZADOS SANTA RITA DE CASSIA | Epiclin | Fernando Hayashi Sant'Anna et al |

| | | | |
|---|---|---|---|
| EPI_ISL_1469744 | HOSPITAL SAO FRANCISCO DE ASSIS | Epiclin | Fernando Hayashi Sant'Anna et al |
| EPI_ISL_1469678 | SECRETARIA MUNICIPAL DE SAUDE DE TRES COROAS | Epiclin | Fernando Hayashi Sant'Anna et al |
| EPI_ISL_1195283 | SECRETARIA MUNICIPAL DE SAUDE DE TRES COROAS | Epiclin | Fernando Hayashi Sant'Anna et al |
| EPI_ISL_1469672 | Diretoria de Vigilância em Saúde | Epiclin | Fernando Hayashi Sant'Anna et al |
| EPI_ISL_1469819 | Diretoria de Vigilância em Saúde | Epiclin | Fernando Hayashi Sant'Anna et al |
| EPI_ISL_1469627 | Diretoria de Vigilância em Saúde | Epiclin | Fernando Hayashi Sant'Anna et al |
| EPI_ISL_1469828 | Fundação de Saúde Pública de Novo Hamburgo | Epiclin | Fernando Hayashi Sant'Anna et al |
| EPI_ISL_1469758 | Secretaria Municipal de Saúde de Três Coroas | Epiclin | Fernando Hayashi Sant'Anna et al |
| EPI_ISL_1469812 | Posto de Saúde Cambará do Sul | Epiclin | Fernando Hayashi Sant'Anna et al |
| EPI_ISL_1469843 | Hospital Sapiranga | Epiclin | Fernando Hayashi Sant'Anna et al |
| EPI_ISL_1469779 | Centro de Especialidades Triunfo | Epiclin | Fernando Hayashi Sant'Anna et al |
| EPI_ISL_1469841 | Diretoria de Vigilância em Saúde | Epiclin | Fernando Hayashi Sant'Anna et al |
| EPI_ISL_1469763 | Unidade Básica de Saúde de Riozinho | Epiclin | Fernando Hayashi Sant'Anna et al |
| EPI_ISL_1469598 | Diretoria de Vigilância em Saúde | Epiclin | Fernando Hayashi Sant'Anna et al |
| EPI_ISL_1469588 | Secretaria Municipal de Saúde de Taquara | Epiclin | Fernando Hayashi Sant'Anna et al |
| EPI_ISL_1469806 | Centro de Referência em Síndromes Gripais | Epiclin | Fernando Hayashi Sant'Anna et al |
| EPI_ISL_1469829 | Diretoria de Vigilância em Saúde | Epiclin | Fernando Hayashi Sant'Anna et al |
| EPI_ISL_1469801 | Hospital Sapiranga | Epiclin | Fernando Hayashi Sant'Anna et al |
| EPI_ISL_1469610 | Diretoria de Vigilância em Saúde | Epiclin | Fernando Hayashi Sant'Anna et al |
| EPI_ISL_1469748 | Diretoria de Vigilância em Saúde | Epiclin | Fernando Hayashi Sant'Anna et al |
| EPI_ISL_1469697 | Hospital Regina | Epiclin | Fernando Hayashi Sant'Anna et al |
| EPI_ISL_1479122 | Fundação Hospitalar São José | Epiclin | Fernando Hayashi Sant'Anna et al |
| EPI_ISL_1469739 | Hospital São Francisco de Assis | Epiclin | Fernando Hayashi Sant'Anna et al |
| EPI_ISL_1469762 | Hospital Nossa Senhora das Graças | Epiclin | Fernando Hayashi Sant'Anna et al |
| EPI_ISL_1469656 | Diretoria de Vigilância em Saúde | Epiclin | Fernando Hayashi Sant'Anna et al |
| EPI_ISL_1469796 | Diretoria de Vigilância em Saúde | Epiclin | Fernando Hayashi Sant'Anna et al |
| EPI_ISL_1469587 | Hospital Sapiranga | Epiclin | Fernando Hayashi Sant'Anna et al |
| EPI_ISL_1469644 | Diretoria de Vigilância em Saúde | Epiclin | Fernando Hayashi Sant'Anna et al |
| EPI_ISL_1469613 | Fundação de Saúde Pública de Novo Hamburgo | Epiclin | Fernando Hayashi Sant'Anna et al |
| EPI_ISL_1469816 | Diretoria de Vigilância em Saúde | Epiclin | Fernando Hayashi Sant'Anna et al |

| | | | |
|---|---|---|---|
| EPI_ISL_1469780 | Secretaria Municipal de Saúde de Taquara | Epiclin | Fernando Hayashi Sant'Anna et al |
| EPI_ISL_1469649 | Unidade de Atendimento DST AIDS TB e Han | Epiclin | Fernando Hayashi Sant'Anna et al |
| EPI_ISL_1469640 | Fundação de Saúde Pública São Camilo de Esteio | Epiclin | Fernando Hayashi Sant'Anna et al |
| EPI_ISL_1469591 | Hospital Nossa Senhora das Graças | Epiclin | Fernando Hayashi Sant'Anna et al |
| EPI_ISL_1469626 | Fundação de Saúde Pública de Novo Hamburgo | Epiclin | Fernando Hayashi Sant'Anna et al |
| EPI_ISL_1469622 | Fundação de Saúde Pública de Novo Hamburgo | Epiclin | Fernando Hayashi Sant'Anna et al |
| EPI_ISL_1469716 | Unidade Sanitária de Igrejinha | Epiclin | Fernando Hayashi Sant'Anna et al |
| EPI_ISL_1469851 | Hospital Nossa Senhora das Graças | Epiclin | Fernando Hayashi Sant'Anna et al |
| EPI_ISL_1469717 | Hospital São Francisco de Assis | Epiclin | Fernando Hayashi Sant'Anna et al |
| EPI_ISL_1469700 | Diretoria de Vigilância em Saúde | Epiclin | Fernando Hayashi Sant'Anna et al |
| EPI_ISL_1469747 | Unidade de Pronto Atendimento de Sapucaia do Sul | Epiclin | Fernando Hayashi Sant'Anna et al |
| EPI_ISL_1469577 | Unidade de Pronto Atendimento de Sapucaia do Sul | Epiclin | Fernando Hayashi Sant'Anna et al |
| EPI_ISL_1469657 | Pronto Atendimento Campo Bom | Epiclin | Fernando Hayashi Sant'Anna et al |
| EPI_ISL_1469750 | Hospital São Francisco de Assis | Epiclin | Fernando Hayashi Sant'Anna et al |
| EPI_ISL_1469712 | Pronto Atendimento Campo Bom | Epiclin | Fernando Hayashi Sant'Anna et al |
| EPI_ISL_1469833 | Hospital Nossa Senhora das Graças | Epiclin | Fernando Hayashi Sant'Anna et al |
| EPI_ISL_1469625 | Diretoria de Vigilância em Saúde | Epiclin | Fernando Hayashi Sant'Anna et al |
| EPI_ISL_1469728 | Diretoria de Vigilância em Saúde | Epiclin | Fernando Hayashi Sant'Anna et al |
| EPI_ISL_1469718 | Unidade de Pronto Atendimento de Sapucaia do Sul | Epiclin | Fernando Hayashi Sant'Anna et al |
| EPI_ISL_1469564 | Pronto Atendimento Campo Bom | Epiclin | Fernando Hayashi Sant'Anna et al |
| EPI_ISL_1469559 | Secretaria Municipal de Saúde de Três Coroas | Epiclin | Fernando Hayashi Sant'Anna et al |
| EPI_ISL_1469565 | Unidade de Pronto Atendimento de Sapucaia do Sul | Epiclin | Fernando Hayashi Sant'Anna et al |
| EPI_ISL_1469597 | Fundação de Saúde Pública de Novo Hamburgo | Epiclin | Fernando Hayashi Sant'Anna et al |
| EPI_ISL_426580 | Instituto Sabin | Laboratory of Virology | Fernando L Melo et al |
| EPI_ISL_1323013 | Hospital Universitario La Paz (Madrid) | SeqCOVID-SPAIN consortium/IBV(CSIC) | Fernando Lázaro-Perona et al |
| EPI_ISL_1323010 | Hospital Universitario La Paz (Madrid) | SeqCOVID-SPAIN consortium/IBV(CSIC) | Fernando Lázaro-Perona et al |
| EPI_ISL_455566 | Institute for Public Health | Laboratory for advanced genomics | Filip Rokić et al |
| EPI_ISL_454605 | Institute for Public Health | Laboratory for advanced genomics | Filip Rokić et al |
| EPI_ISL_468656 | Institute for Public Health | Laboratory for advanced genomics | Filip Rokić et al |
| EPI_ISL_468591 | Institute for Public Health | Laboratory for advanced genomics | Filip Rokić et al |

| | | | |
|---|---|---|---|
| EPI_ISL_454574 | nstitute for Public Health | Laboratory for advanced genomics | Filip Rokić et al |
| EPI_ISL_417949 | Laboratório Hermes Pardini | Bioinformatics Laboratory - LNCC | Filipe Romero et al |
| EPI_ISL_1592227 | Laboratório Hermes Pardini | Laboratório de Biologia Integrativa, Instituto de Ciências Biológicas, Universidade Federal de Minas Gerais | Filipe Romero Rebello Moreira et al |
| EPI_ISL_1592229 | Laboratório Hermes Pardini | Laboratório de Biologia Integrativa, Instituto de Ciências Biológicas, Universidade Federal de Minas Gerais | Filipe Romero Rebello Moreira et al |
| EPI_ISL_1133267 | Laboratório Hermes Pardini | Laboratório de Biologia Integrativa, Instituto de Ciências Biológicas, Universidade Federal de Minas Gerais | Filipe Romero Rebello Moreira et al |
| EPI_ISL_1494967 | Laboratório de Biologia Integrativa | Laboratório de Biologia Integrativa | Filipe Romero Rebello Moreira et al |
| EPI_ISL_1494968 | Laboratório de Biologia Integrativa | Laboratório de Biologia Integrativa | Filipe Romero Rebello Moreira et al |
| EPI_ISL_1494969 | Laboratório de Biologia Integrativa | Laboratório de Biologia Integrativa | Filipe Romero Rebello Moreira et al |
| EPI_ISL_1494971 | Laboratório de Biologia Integrativa | Laboratório de Biologia Integrativa | Filipe Romero Rebello Moreira et al |
| EPI_ISL_1494972 | Laboratório de Biologia Integrativa | Laboratório de Biologia Integrativa | Filipe Romero Rebello Moreira et al |
| EPI_ISL_1494973 | Laboratório de Biologia Integrativa | Laboratório de Biologia Integrativa | Filipe Romero Rebello Moreira et al |
| EPI_ISL_1494974 | Laboratório de Biologia Integrativa | Laboratório de Biologia Integrativa | Filipe Romero Rebello Moreira et al |
| EPI_ISL_1494975 | Laboratório de Biologia Integrativa | Laboratório de Biologia Integrativa | Filipe Romero Rebello Moreira et al |
| EPI_ISL_1494977 | Laboratório de Biologia Integrativa | Laboratório de Biologia Integrativa | Filipe Romero Rebello Moreira et al |
| EPI_ISL_1494978 | Laboratório de Biologia Integrativa | Laboratório de Biologia Integrativa | Filipe Romero Rebello Moreira et al |
| EPI_ISL_1494982 | Laboratório de Biologia Integrativa | Laboratório de Biologia Integrativa | Filipe Romero Rebello Moreira et al |
| EPI_ISL_1494986 | Laboratório de Biologia Integrativa | Laboratório de Biologia Integrativa | Filipe Romero Rebello Moreira et al |
| EPI_ISL_1494987 | Laboratório de Biologia Integrativa | Laboratório de Biologia Integrativa | Filipe Romero Rebello Moreira et al |
| EPI_ISL_1494989 | Laboratório de Biologia Integrativa | Laboratório de Biologia Integrativa | Filipe Romero Rebello Moreira et al |
| EPI_ISL_1494994 | Laboratório de Biologia Integrativa | Laboratório de Biologia Integrativa | Filipe Romero Rebello Moreira et al |
| EPI_ISL_1494995 | Laboratório de Biologia Integrativa | Laboratório de Biologia Integrativa | Filipe Romero Rebello Moreira et al |
| EPI_ISL_1494997 | Laboratório de Biologia Integrativa | Laboratório de Biologia Integrativa | Filipe Romero Rebello Moreira et al |
| EPI_ISL_1494998 | Laboratório de Biologia Integrativa | Laboratório de Biologia Integrativa | Filipe Romero Rebello Moreira et al |
| EPI_ISL_1494999 | Laboratório de Biologia Integrativa | Laboratório de Biologia Integrativa | Filipe Romero Rebello Moreira et al |
| EPI_ISL_1495004 | Laboratório de Biologia Integrativa | Laboratório de Biologia Integrativa | Filipe Romero Rebello Moreira et al |
| EPI_ISL_1495005 | Laboratório de Biologia Integrativa | Laboratório de Biologia Integrativa | Filipe Romero Rebello Moreira et al |
| EPI_ISL_1495006 | Laboratório de Biologia Integrativa | Laboratório de Biologia Integrativa | Filipe Romero Rebello Moreira et al |
| EPI_ISL_1495007 | Laboratório de Biologia Integrativa | Laboratório de Biologia Integrativa | Filipe Romero Rebello Moreira et al |
| EPI_ISL_1495009 | Laboratório de Biologia Integrativa | Laboratório de Biologia Integrativa | Filipe Romero Rebello Moreira et al |

| | | | |
|---|---|---|---|
| EPI_ISL_1495010 | Laboratório de Biologia Integrativa | Laboratório de Biologia Integrativa | Filipe Romero Rebello Moreira et al |
| EPI_ISL_1495011 | Laboratório de Biologia Integrativa | Laboratório de Biologia Integrativa | Filipe Romero Rebello Moreira et al |
| EPI_ISL_1495012 | Laboratório de Biologia Integrativa | Laboratório de Biologia Integrativa | Filipe Romero Rebello Moreira et al |
| EPI_ISL_1495013 | Laboratório de Biologia Integrativa | Laboratório de Biologia Integrativa | Filipe Romero Rebello Moreira et al |
| EPI_ISL_1495014 | Laboratório de Biologia Integrativa | Laboratório de Biologia Integrativa | Filipe Romero Rebello Moreira et al |
| EPI_ISL_1495015 | Laboratório de Biologia Integrativa | Laboratório de Biologia Integrativa | Filipe Romero Rebello Moreira et al |
| EPI_ISL_1495016 | Laboratório de Biologia Integrativa | Laboratório de Biologia Integrativa | Filipe Romero Rebello Moreira et al |
| EPI_ISL_1495019 | Laboratório de Biologia Integrativa | Laboratório de Biologia Integrativa | Filipe Romero Rebello Moreira et al |
| EPI_ISL_1495020 | Laboratório de Biologia Integrativa | Laboratório de Biologia Integrativa | Filipe Romero Rebello Moreira et al |
| EPI_ISL_1495021 | Laboratório de Biologia Integrativa | Laboratório de Biologia Integrativa | Filipe Romero Rebello Moreira et al |
| EPI_ISL_1495022 | Laboratório de Biologia Integrativa | Laboratório de Biologia Integrativa | Filipe Romero Rebello Moreira et al |
| EPI_ISL_1495023 | Laboratório de Biologia Integrativa | Laboratório de Biologia Integrativa | Filipe Romero Rebello Moreira et al |
| EPI_ISL_1495024 | Laboratório de Biologia Integrativa | Laboratório de Biologia Integrativa | Filipe Romero Rebello Moreira et al |
| EPI_ISL_1495025 | Laboratório de Biologia Integrativa | Laboratório de Biologia Integrativa | Filipe Romero Rebello Moreira et al |
| EPI_ISL_1495026 | Laboratório de Biologia Integrativa | Laboratório de Biologia Integrativa | Filipe Romero Rebello Moreira et al |
| EPI_ISL_1495027 | Laboratório de Biologia Integrativa | Laboratório de Biologia Integrativa | Filipe Romero Rebello Moreira et al |
| EPI_ISL_1495030 | Laboratório de Biologia Integrativa | Laboratório de Biologia Integrativa | Filipe Romero Rebello Moreira et al |
| EPI_ISL_1495031 | Laboratório de Biologia Integrativa | Laboratório de Biologia Integrativa | Filipe Romero Rebello Moreira et al |
| EPI_ISL_1495032 | Laboratório de Biologia Integrativa | Laboratório de Biologia Integrativa | Filipe Romero Rebello Moreira et al |
| EPI_ISL_1495035 | Laboratório de Biologia Integrativa | Laboratório de Biologia Integrativa | Filipe Romero Rebello Moreira et al |
| EPI_ISL_1495036 | Laboratório de Biologia Integrativa | Laboratório de Biologia Integrativa | Filipe Romero Rebello Moreira et al |
| EPI_ISL_1495037 | Laboratório de Biologia Integrativa | Laboratório de Biologia Integrativa | Filipe Romero Rebello Moreira et al |
| EPI_ISL_1495038 | Laboratório de Biologia Integrativa | Laboratório de Biologia Integrativa | Filipe Romero Rebello Moreira et al |
| EPI_ISL_1495042 | Laboratório de Biologia Integrativa | Laboratório de Biologia Integrativa | Filipe Romero Rebello Moreira et al |
| EPI_ISL_1494960 | Laboratório de Biologia Integrativa | Laboratório de Biologia Integrativa | Filipe Romero Rebello Moreira et al |
| EPI_ISL_1494961 | Laboratório de Biologia Integrativa | Laboratório de Biologia Integrativa | Filipe Romero Rebello Moreira et al |
| EPI_ISL_1494962 | Laboratório de Biologia Integrativa | Laboratório de Biologia Integrativa | Filipe Romero Rebello Moreira et al |
| EPI_ISL_1494963 | Laboratório de Biologia Integrativa | Laboratório de Biologia Integrativa | Filipe Romero Rebello Moreira et al |
| EPI_ISL_1494964 | Laboratório de Biologia Integrativa | Laboratório de Biologia Integrativa | Filipe Romero Rebello Moreira et al |
| EPI_ISL_1494965 | Laboratório de Biologia Integrativa | Laboratório de Biologia Integrativa | Filipe Romero Rebello Moreira et al |

| | | | |
|---|---|---|---|
| EPI_ISL_1494966 | Laboratório de Biologia Integrativa | Laboratório de Biologia Integrativa | Filipe Romero Rebello Moreira et al |
| EPI_ISL_1133274 | Laboratório Hermes Pardini | Laboratório de Biologia Integrativa, Instituto de Ciências Biológicas, Universidade Federal de Minas Gerais | Filipe Romero Rebello Moreira et al |
| EPI_ISL_1133256 | Laboratório Hermes Pardini | Laboratório de Biologia Integrativa, Instituto de Ciências Biológicas, Universidade Federal de Minas Gerais | Filipe Romero Rebello Moreira et al |
| EPI_ISL_1592230 | Laboratório Hermes Pardini | Laboratório de Biologia Integrativa, Instituto de Ciências Biológicas, Universidade Federal de Minas Gerais | Filipe Romero Rebello Moreira et al |
| EPI_ISL_1133261 | Laboratório Hermes Pardini | Laboratório de Biologia Integrativa, Instituto de Ciências Biológicas, Universidade Federal de Minas Gerais | Filipe Romero Rebello Moreira et al |
| EPI_ISL_1357775 | National Virus Reference Laboratory | National Virus Reference Laboratory | Fiona Crispie et al |
| EPI_ISL_1357776 | National Virus Reference Laboratory | National Virus Reference Laboratory | Fiona Crispie et al |
| EPI_ISL_1010714 | New South Wales Health Pathology Royal Prince Alfred Hospital | Microbiology RPAH | Foster et al |
| EPI_ISL_1061033 | New South Wales Health Pathology Royal Prince Alfred Hospital | Microbiology RPAH | Foster et al |
| EPI_ISL_1061037 | New South Wales Health Pathology Royal Prince Alfred Hospital | Microbiology RPAH | Foster et al |
| EPI_ISL_1121987 | New South Wales Health Pathology Royal Prince Alfred Hospital | Microbiology RPAH | Foster et al |
| EPI_ISL_1315067 | New South Wales Health Pathology Royal Prince Alfred Hospital | Microbiology RPAH | Foster et al |
| EPI_ISL_1660408 | New South Wales Health Pathology Royal Prince Alfred Hospital | Microbiology RPAH | Foster et al |
| EPI_ISL_678320 | Area of Virology, Serology and Virology Division (SAViD), New South Wales Health Pathology Randwick | Virology Research Laboratory; Area of Virology, Serology and Virology Division (SAViD), New South Wales Health Pathology Randwick | Foster et al |
| EPI_ISL_678344 | Area of Virology, Serology and Virology Division (SAViD), New South Wales Health Pathology Randwick | Virology Research Laboratory; Area of Virology, Serology and Virology Division (SAViD), New South Wales Health Pathology Randwick | Foster et al |
| EPI_ISL_1121976 | Area of Virology, Serology and Virology Division (SAViD), New South Wales Health Pathology Randwick | Virology Research Laboratory; Area of Virology, Serology and Virology Division (SAViD), New South Wales Health Pathology Randwick | Foster et al |
| EPI_ISL_1406435 | Area of Virology, Serology and Virology Division (SAViD), New South Wales Health Pathology Randwick | Virology Research Laboratory; Area of Virology, Serology and Virology Division (SAViD), New South Wales Health Pathology Randwick | Foster et al |
| EPI_ISL_1516747 | Area of Virology, Serology and Virology Division (SAViD), New South Wales Health Pathology Randwick | Virology Research Laboratory; Area of Virology, Serology and Virology Division (SAViD), New South Wales Health Pathology Randwick | Foster et al |
| EPI_ISL_1081793 | Philippine Red Cross | Philippine Genome Center | Francis A. Tablizo et al |
| EPI_ISL_1081795 | SOUTH SUPER HI-WAY | Philippine Genome Center | Francis A. Tablizo et al |
| EPI_ISL_1122458 | The Lord's Grace Medical and Industrial Clinic | Philippine Genome Center | Francis A. Tablizo et al |
| EPI_ISL_1213568 | Lung Center of the Philippines (LCP) | Philippine Genome Center | Francis A. Tablizo et al |
| EPI_ISL_1122457 | Hi-Precision Diagnostics | Philippine Genome Center | Francis A. Tablizo et al |
| EPI_ISL_1122455 | Southern Philippines Medical Center | Philippine Genome Center | Francis A. Tablizo et al |
| EPI_ISL_1122456 | Manila Doctors Hospital | Philippine Genome Center | Francis A. Tablizo et al |
| EPI_ISL_1213571 | Negros Oriental Provincial Hospital | Philippine Genome Center | Francis A. Tablizo et al |
| EPI_ISL_1213573 | Negros Oriental Provincial Hospital | Philippine Genome Center | Francis A. Tablizo et al |

| | | | |
|---|---|---|---|
| EPI_ISL_1213554 | Cebu TB Reference Laboratory | Philippine Genome Center | Francis A. Tablizo et al |
| EPI_ISL_1213518 | Bohol Containerized PCR Laboratory | Philippine Genome Center | Francis A. Tablizo et al |
| EPI_ISL_1213509 | Bohol Containerized PCR Laboratory | Philippine Genome Center | Francis A. Tablizo et al |
| EPI_ISL_1213530 | Bohol Containerized PCR Laboratory | Philippine Genome Center | Francis A. Tablizo et al |
| EPI_ISL_434536 | Hospital San Vicente de Paul | Inciensa, Instituto Costarricense de Investigación y Enseñanza en Nutrición y Salud | Francisco Duarte et al |
| EPI_ISL_491451 | Hospital México | Inciensa, Instituto Costarricense de Investigación y Enseñanza en Nutrición y Salud | Francisco Duarte et al |
| EPI_ISL_512655 | Hospital De Niños Dr. Carlos Saenz Herrera [San Jose/San Jose] | Inciensa, Instituto Costarricense de Investigación y Enseñanza en Nutrición y Salud | Francisco Duarte et al |
| EPI_ISL_512668 | Area De Salud Corredores | Inciensa, Instituto Costarricense de Investigación y Enseñanza en Nutrición y Salud | Francisco Duarte et al |
| EPI_ISL_512669 | Area De Salud Alajuela Norte - Clinica Dr. Marcial Rodriguez | Inciensa, Instituto Costarricense de Investigación y Enseñanza en Nutrición y Salud | Francisco Duarte et al |
| EPI_ISL_527740 | Area De Salud Alajuela Norte - Clinica Dr. Marcial Rodriguez | Inciensa, Instituto Costarricense de Investigación y Enseñanza en Nutrición y Salud | Francisco Duarte et al |
| EPI_ISL_527756 | Area De Salud Aserri | Inciensa, Instituto Costarricense de Investigación y Enseñanza en Nutrición y Salud | Francisco Duarte et al |
| EPI_ISL_682253 | HOSPITAL SAN JUAN DE DIOS | Inciensa, Instituto Costarricense de Investigación y Enseñanza en Nutrición y Salud | Francisco Duarte et al |
| EPI_ISL_682271 | HOSPITAL SAN JUAN DE DIOS | Inciensa, Instituto Costarricense de Investigación y Enseñanza en Nutrición y Salud | Francisco Duarte et al |
| EPI_ISL_682276 | LABORATORIOS LABIN | Inciensa, Instituto Costarricense de Investigación y Enseñanza en Nutrición y Salud | Francisco Duarte et al |
| EPI_ISL_770006 | Hle - Asociacion Hogar De Ancianos De Palmar Sur De Osa | Inciensa, Instituto Costarricense de Investigación y Enseñanza en Nutrición y Salud | Francisco Duarte et al |
| EPI_ISL_770028 | Hospital Dr. Raul Blanco Cervantes | Inciensa, Instituto Costarricense de Investigación y Enseñanza en Nutrición y Salud | Francisco Duarte et al |
| EPI_ISL_770007 | Area De Salud San Rafael | Inciensa, Instituto Costarricense de Investigación y Enseñanza en Nutrición y Salud | Francisco Duarte et al |
| EPI_ISL_770000 | Area De Salud La Cruz | Inciensa, Instituto Costarricense de Investigación y Enseñanza en Nutrición y Salud | Francisco Duarte et al |
| EPI_ISL_770001 | Area De Salud La Cruz | Inciensa, Instituto Costarricense de Investigación y Enseñanza en Nutrición y Salud | Francisco Duarte et al |
| EPI_ISL_770002 | Area De Salud La Cruz | Inciensa, Instituto Costarricense de Investigación y Enseñanza en Nutrición y Salud | Francisco Duarte et al |
| EPI_ISL_770025 | Area De Salud San Juan-San Diego-Concepcion 2 | Inciensa, Instituto Costarricense de Investigación y Enseñanza en Nutrición y Salud | Francisco Duarte et al |
| EPI_ISL_770017 | Area De Salud Perez Zeledon | Inciensa, Instituto Costarricense de Investigación y Enseñanza en Nutrición y Salud | Francisco Duarte et al |
| EPI_ISL_770029 | Area De Salud Corredores | Inciensa, Instituto Costarricense de Investigación y Enseñanza en Nutrición y Salud | Francisco Duarte et al |

| | | | |
|---|---|---|---|
| EPI_ISL_770030 | Area De Salud Buenos Aires | Inciensa, Instituto Costarricense de Investigación y Enseñanza en Nutrición y Salud | Francisco Duarte et al |
| EPI_ISL_769989 | Area De Salud San Francisco-San Antonio (Coopesana) | Inciensa, Instituto Costarricense de Investigación y Enseñanza en Nutrición y Salud | Francisco Duarte et al |
| EPI_ISL_769986 | Area De Salud Catedral Noreste | Inciensa, Instituto Costarricense de Investigación y Enseñanza en Nutrición y Salud | Francisco Duarte et al |
| EPI_ISL_770022 | Area De Salud Perez Zeledon | Inciensa, Instituto Costarricense de Investigación y Enseñanza en Nutrición y Salud | Francisco Duarte et al |
| EPI_ISL_770023 | Area De Salud San Francisco-San Antonio (Coopesana) | Inciensa, Instituto Costarricense de Investigación y Enseñanza en Nutrición y Salud | Francisco Duarte et al |
| EPI_ISL_914800 | AREA DE SALUD SAN FRANCISCO-SAN ANTONIO (COOPESANA) | Inciensa, Instituto Costarricense de Investigación y Enseñanza en Nutrición y Salud | Francisco Duarte et al |
| EPI_ISL_914807 | HLE - ASOCIACION HOGAR DE ANCIANOS SANTIAGO CRESPO CALVO | Inciensa, Instituto Costarricense de Investigación y Enseñanza en Nutrición y Salud | Francisco Duarte et al |
| EPI_ISL_914811 | AREA DE SALUD GOICOECHEA 2 - CLINICA DR. JIMENEZ NUÑEZ | Inciensa, Instituto Costarricense de Investigación y Enseñanza en Nutrición y Salud | Francisco Duarte et al |
| EPI_ISL_914824 | TAMIZAJE COMUNITARIO - PASO CANOAS | Inciensa, Instituto Costarricense de Investigación y Enseñanza en Nutrición y Salud | Francisco Duarte et al |
| EPI_ISL_914834 | HOSPITAL METROPOLITANO | Inciensa, Instituto Costarricense de Investigación y Enseñanza en Nutrición y Salud | Francisco Duarte et al |
| EPI_ISL_1067600 | TAMIZAJE COMUNITARIO - PASO CANOAS | Inciensa, Instituto Costarricense de Investigación y Enseñanza en Nutrición y Salud | Francisco Duarte et al |
| EPI_ISL_1067593 | LABORATORIO CLINICO LABIN | Inciensa, Instituto Costarricense de Investigación y Enseñanza en Nutrición y Salud | Francisco Duarte et al |
| EPI_ISL_1196438 | AREA DE SALUD CORRALILLO | Inciensa, Instituto Costarricense de Investigación y Enseñanza en Nutrición y Salud | Francisco Duarte et al |
| EPI_ISL_1379415 | AREA DE SALUD PAVAS (COOPESALUD) | Inciensa, Instituto Costarricense de Investigación y Enseñanza en Nutrición y Salud | Francisco Duarte et al |
| EPI_ISL_1379422 | HOSPITAL SAN JUAN DE DIOS | Inciensa, Instituto Costarricense de Investigación y Enseñanza en Nutrición y Salud | Francisco Duarte et al |
| EPI_ISL_1576835 | Universidad Industrial de Santander (Laboratorio Central de Investigaciones - Clínica Chicamocha). | Universidad Industrial de Santander. | Francisco Martinez-Perez et al |
| EPI_ISL_1577026 | Universidad Industrial de Santander (Laboratorio Central de Investigaciones - Clínica Chicamocha). | Universidad Industrial de Santander. | Francisco Martinez-Perez et al |
| EPI_ISL_1629710 | Universidad Industrial de Santander (Laboratorio Central de Investigaciones - Clínica Chicamocha). | Universidad Industrial de Santander. | Francisco Martinez-Perez et al |
| EPI_ISL_1553838 | CAP SANT JULIÀ DE VILATORTA | Banc de Sang i Teixits | Francisco Vidal et al |
| EPI_ISL_1665222 | National Platform bis UMONS/Jolimont | National Platform bis UMONS/Jolimont | François Dufrasne et al |
| EPI_ISL_1623659 | National Platform bis UMONS/Jolimont | National Platform bis UMONS/Jolimont | François Dufrasne et al |
| EPI_ISL_1665233 | National Platform bis UMONS/Jolimont | National Platform bis UMONS/Jolimont | François Dufrasne et al |
| EPI_ISL_513312 | Public Health, United States Air Force School of Aerospace Medicine | Public Health, United States Air Force School of Aerospace Medicine | Fries et al |
| EPI_ISL_513313 | Public Health, United States Air Force School of Aerospace Medicine | Public Health, United States Air Force School of Aerospace Medicine | Fries et al |

| | | | |
|---|---|---|---|
| EPI_ISL_766034 | RS Mitra Keluarga Bintaro | Eijkman Institute for Molecular Biology, Ministry of Research and Technology/National Agency for Research and Innovation | Frilasita A Yudhaputri et al |
| EPI_ISL_574611 | RS Premier Bintaro | Eijkman Institute for Molecular Biology, Ministry of Research and Technology/National Agency for Research and Innovation | Frilasita A Yudhaputri et al |
| EPI_ISL_947270 | RS Mitra Keluarga Gading Serpong | Eijkman Institute for Molecular Biology, Ministry of Research and Technology/National Agency for Research and Innovation | Frilasita A Yudhaputri et al |
| EPI_ISL_574606 | RS Siloam Purwakarta | Eijkman Institute for Molecular Biology, Ministry of Research and Technology/National Agency for Research and Innovation | Frilasita A Yudhaputri et al |
| EPI_ISL_574612 | RS Hermina Mekarsari | Eijkman Institute for Molecular Biology, Ministry of Research and Technology/National Agency for Research and Innovation | Frilasita A Yudhaputri et al |
| EPI_ISL_568690 | RSUP Fatmawati | Eijkman Institute for Molecular Biology, Ministry of Research and Technology/National Agency for Research and Innovation | Frilasita A Yudhaputri et al |
| EPI_ISL_568691 | RSUP Fatmawati | Eijkman Institute for Molecular Biology, Ministry of Research and Technology/National Agency for Research and Innovation | Frilasita A Yudhaputri et al |
| EPI_ISL_574608 | RS Mitra Keluarga Kelapa Gading | Eijkman Institute for Molecular Biology, Ministry of Research and Technology/National Agency for Research and Innovation | Frilasita A Yudhaputri et al |
| EPI_ISL_766043 | RS Pelni | Eijkman Institute for Molecular Biology, Ministry of Research and Technology/National Agency for Research and Innovation | Frilasita A Yudhaputri et al |
| EPI_ISL_766046 | RSUP Fatmawati | Eijkman Institute for Molecular Biology, Ministry of Research and Technology/National Agency for Research and Innovation | Frilasita A Yudhaputri et al |
| EPI_ISL_574614 | RS Kramat 128 | Eijkman Institute for Molecular Biology, Ministry of Research and Technology/National Agency for Research and Innovation | Frilasita A Yudhaputri et al |
| EPI_ISL_766033 | RSUD Prof. DR. W. Z. Johannes Kupang | Eijkman Institute for Molecular Biology, Ministry of Research and Technology/National Agency for Research and Innovation | Frilasita A Yudhaputri et al |
| EPI_ISL_766039 | RS Bhayangkara Tk. III | Eijkman Institute for Molecular Biology, Ministry of Research and Technology/National Agency for Research and Innovation | Frilasita A Yudhaputri et al |
| EPI_ISL_574604 | RSUP Prof. Dr. R. Kandou Manado | Eijkman Institute for Molecular Biology, Ministry of Research and Technology/National Agency for Research and Innovation | Frilasita A Yudhaputri et al |
| EPI_ISL_574605 | RSUP dr. Sardjito | Eijkman Institute for Molecular Biology, Ministry of Research and Technology/National Agency for Research and Innovation | Frilasita A Yudhaputri et al |
| EPI_ISL_902739 | Hospital Universitari Germans Trias i Pujol (HUGTiP) / Fundació Lluita contra la SIDA (FLSida) | IrsiCaixa - Can Ruti CovidSeq | Fundació irsiCaixa. Hospital Universitari Ge |
| EPI_ISL_902747 | Hospital Universitari Germans Trias i Pujol (HUGTiP) / Fundació Lluita contra la SIDA (FLSida) | IrsiCaixa - Can Ruti CovidSeq | Fundació irsiCaixa. Hospital Universitari Ge |
| EPI_ISL_660069 | Zurita & Zurita Laboratorios | Zurita & Zurita Laboratorios | Gabriela Sevillano Camilo Zurita-Salinas Ka |
| EPI_ISL_660070 | Zurita & Zurita Laboratorios | Zurita & Zurita Laboratorios | Gabriela Sevillano Camilo Zurita-Salinas Ka |
| EPI_ISL_681703 | Zurita & Zurita Laboratorios | Zurita & Zurita Laboratorios | Gabriela Sevillano et al |
| EPI_ISL_468752 | Center for Genome Regulation (CRG) | Center for Mathematical Modeling and Center for Genome Regulation. Santiago, Chile | Gaete A et al |
| EPI_ISL_459858 | Center for Genome Regulation (CRG) | Center for Mathematical Modeling and Center for Genome Regulation. Santiago, Chile | Gaete A et al |
| EPI_ISL_753417 | Clinical virology Laboratory, Children's Hospital Los Angeles | Center for Personalized Medicine, Children's Hospital Los Angeles | Gai et al |

rmans Trias i Pujol(HUGTiP) et al

rmans Trias i Pujol(HUGTiP) et al

aren Loaiza David Ortega-Paredes Jeannete

aren Loaiza David Ortega-Paredes Jeannete

| | | | |
|---|---|---|---|
| EPI_ISL_602623 | AHRI-Sigal | KRISP, KZN Research Innovation and Sequencing Platform | Gazy I et al |
| EPI_ISL_1615655 | Centre de Recherches Médicales de Lambaréné (CERMEL) | Centre de Recherches Médicales de Lambaréné (CERMEL) | Gédéon Prince Manouana et al |
| EPI_ISL_637110 | Rafik Hariri University Hospital | Microbial Pathogenomics Lab | Georgi Merhi et al |
| EPI_ISL_637112 | Rafik Hariri University Hospital | Microbial Pathogenomics Lab | Georgi Merhi et al |
| EPI_ISL_637113 | Rafik Hariri University Hospital | Microbial Pathogenomics Lab | Georgi Merhi et al |
| EPI_ISL_1116466 | Laboratoire central de Virologie | Laboratoire de Biotechnologie | Ghizlane EL Amin et al |
| EPI_ISL_1116555 | Laboratoire central de Virologie | Laboratoire de Biotechnologie | Ghizlane EL Amin et al |
| EPI_ISL_467431 | Molecular Diagnostics Services (MDS) | KRISP, KZN Research Innovation and Sequencing Platform | Giandhari J et al |
| EPI_ISL_482706 | Molecular Diagnostics Services (MDS) | KRISP, KZN Research Innovation and Sequencing Platform | Giandhari J et al |
| EPI_ISL_482854 | Molecular Diagnostics Services (MDS) | KRISP, KZN Research Innovation and Sequencing Platform | Giandhari J et al |
| EPI_ISL_482871 | Molecular Diagnostics Services (MDS) | KRISP, KZN Research Innovation and Sequencing Platform | Giandhari J et al |
| EPI_ISL_482872 | Molecular Diagnostics Services (MDS) | KRISP, KZN Research Innovation and Sequencing Platform | Giandhari J et al |
| EPI_ISL_495526 | NHLS-IALCH | KRISP, KZN Research Innovation and Sequencing Platform | Giandhari J et al |
| EPI_ISL_495554 | NHLS-IALCH | KRISP, KZN Research Innovation and Sequencing Platform | Giandhari J et al |
| EPI_ISL_498076 | NHLS-IALCH | KRISP, KZN Research Innovation and Sequencing Platform | Giandhari J et al |
| EPI_ISL_455634 | KRISP, KZN Research Innovation and Sequencing Platform | KRISP, KZN Research Innovation and Sequencing Platform | Giandhari J et al |
| EPI_ISL_455638 | KRISP, KZN Research Innovation and Sequencing Platform | KRISP, KZN Research Innovation and Sequencing Platform | Giandhari J et al |
| EPI_ISL_535402 | NHLS-IALCH | KRISP, KZN Research Innovation and Sequencing Platform | Giandhari J et al |
| EPI_ISL_535474 | NHLS-IALCH | KRISP, KZN Research Innovation and Sequencing Platform | Giandhari J et al |
| EPI_ISL_535521 | NHLS-IALCH | KRISP, KZN Research Innovation and Sequencing Platform | Giandhari J et al |
| EPI_ISL_529792 | NHLS-IALCH | KRISP, KZN Research Innovation and Sequencing Platform | Giandhari J et al |
| EPI_ISL_529793 | NHLS-IALCH | KRISP, KZN Research Innovation and Sequencing Platform | Giandhari J et al |
| EPI_ISL_602640 | NHLS-IALCH | KRISP, KZN Research Innovation and Sequencing Platform | Giandhari J et al |
| EPI_ISL_602751 | NHLS-IALCH | KRISP, KZN Research Innovation and Sequencing Platform | Giandhari J et al |
| EPI_ISL_602755 | NHLS-IALCH | KRISP, KZN Research Innovation and Sequencing Platform | Giandhari J et al |
| EPI_ISL_1085997 | Netcare/AMPATH | KRISP, KZN Research Innovation and Sequencing Platform | Giandhari J et al |
| EPI_ISL_1085999 | Netcare/AMPATH | KRISP, KZN Research Innovation and Sequencing Platform | Giandhari J et al |
| EPI_ISL_1371935 | National Health Laboratory Service, South Africa | KRISP, KZn Research Innovation and Sequencing Platform | Giandhari J et al |
| EPI_ISL_1550934 | National Health Laboratory Service, South Africa | KRISP, KZN Research Innovation and Sequencing Platform | Giandhari J et al |
| EPI_ISL_1550952 | National Health Laboratory Service, South Africa | KRISP, KZN Research Innovation and Sequencing Platform | Giandhari J et al |

| | | | |
|---|---|---|---|
| EPI_ISL_1289446 | Department of Health Technology and Informatics, The Hong Kong Polytechnic University | Department of Health Technology and Informatics, The Hong Kong Polytechnic University | Gilman Kit-Hang Siu et al |
| EPI_ISL_1018118 | Department of Health Technology and Informatics, The Hong Kong Polytechnic University | Department of Health Technology and Informatics, The Hong Kong Polytechnic University | Gilman Kit-Hang Siu et al |
| EPI_ISL_1018119 | Department of Health Technology and Informatics, The Hong Kong Polytechnic University | Department of Health Technology and Informatics, The Hong Kong Polytechnic University | Gilman Kit-Hang Siu et al |
| EPI_ISL_1018123 | Department of Health Technology and Informatics, The Hong Kong Polytechnic University | Department of Health Technology and Informatics, The Hong Kong Polytechnic University | Gilman Kit-Hang Siu et al |
| EPI_ISL_1018312 | Department of Health Technology and Informatics, The Hong Kong Polytechnic University | Department of Health Technology and Informatics, The Hong Kong Polytechnic University | Gilman Kit-Hang Siu et al |
| EPI_ISL_1019642 | Department of Health Technology and Informatics, The Hong Kong Polytechnic University | Department of Health Technology and Informatics, The Hong Kong Polytechnic University | Gilman Kit-Hang Siu et al |
| EPI_ISL_1019674 | Department of Health Technology and Informatics, The Hong Kong Polytechnic University | Department of Health Technology and Informatics, The Hong Kong Polytechnic University | Gilman Kit-Hang Siu et al |
| EPI_ISL_1019678 | Department of Health Technology and Informatics, The Hong Kong Polytechnic University | Department of Health Technology and Informatics, The Hong Kong Polytechnic University | Gilman Kit-Hang Siu et al |
| EPI_ISL_1019734 | Department of Health Technology and Informatics, The Hong Kong Polytechnic University | Department of Health Technology and Informatics, The Hong Kong Polytechnic University | Gilman Kit-Hang Siu et al |
| EPI_ISL_1019764 | Department of Health Technology and Informatics, The Hong Kong Polytechnic University | Department of Health Technology and Informatics, The Hong Kong Polytechnic University | Gilman Kit-Hang Siu et al |
| EPI_ISL_1020215 | Department of Health Technology and Informatics, The Hong Kong Polytechnic University | Department of Health Technology and Informatics, The Hong Kong Polytechnic University | Gilman Kit-Hang Siu et al |
| EPI_ISL_763062 | Department for Virology, Molecular Biology and Genome Research, R. G. Lugar Center for Public Health Research, National Center for Disease Control and Public Health (NCDC) of Georgia. | Department for Virology, Molecular Biology and Genome Research, R. G. Lugar Center for Public Health Research, National Center for Disease Control and Public Health (NCDC) of Georgia. | Giorgi Tomashvili et al |
| EPI_ISL_1048371 | Megalab, Molecular and Cytogenetics Diagnostics | Department for Virology, Molecular Biology and Genome Research, R. G. Lugar Center for Public Health Research, National Center for Disease Control and Public Health (NCDC) of Georgia. | Giorgi Tomashvili et al |
| EPI_ISL_876976 | 1.AO Universitaria 'S. Giovanni di Dio e Ruggi D'Aragona, Scuola Medica Salernitana' Hospital / 2.UOC di Virologia e Microbiologia, Università della Campania 'L. Vanvitelli' / 3.AO Universitaria 'Federico II' Napoli Hospital / 4. AORN 'San Giuseppe Moscati' Avellino Hospital / 5.AO 'San Pio - presidio G. Rummo' Benevento Hospital / 6.AO 'Sant'Anna e San Sebastiano' Caserta Hospital / 7.PO 'Maria Santissima Addolorata' Eboli Hospital / 8.Biogem Istituto di Ricerche Genetiche | 1. Genome Research Center for Health (CRGS) / 2. Laboratory of Molecular Medicine and Genomics(LMMGe) / 3. Center for Research in Pure and Applied Mathematics (CRMPA) | Giorgio Giurato (Corresponding Author) et a |
| EPI_ISL_1366893 | 1.AO Universitaria 'S. Giovanni di Dio e Ruggi D'Aragona, Scuola Medica Salernitana' Hospital / 2.UOC di Virologia e Microbiologia, Università della Campania 'L. Vanvitelli' / 3.AO Universitaria 'Federico II' Napoli Hospital / 4. AORN 'San Giuseppe Moscati' Avellino Hospital / 5.AO 'San Pio - presidio G. Rummo' Benevento Hospital / 6.AO 'Sant'Anna e San Sebastiano' Caserta Hospital / 7.PO 'Maria Santissima Addolorata' Eboli Hospital / 8.Biogem Istituto di Ricerche Genetiche | 1. Genome Research Center for Health (CRGS) / 2. Laboratory of Molecular Medicine and Genomics(LMMGe) / 3. Center for Research in Pure and Applied Mathematics (CRMPA) | Giorgio Giurato (Corresponding Author) et a |

| | | | |
|---|---|---|---|
| EPI_ISL_1361596 | 1.AO Universitaria 'S. Giovanni di Dio e Ruggi D'Aragona, Scuola Medica Salernitana' Hospital / 2.UOC di Virologia e Microbiologia, Università della Campania 'L. Vanvitelli' / 3.AO Universitaria 'Federico II' Napoli Hospital / 4. AORN 'San Giuseppe Moscati' Avellino Hospital / 5.AO 'San Pio - presidio G. Rummo' Benevento Hospital / 6.AO 'Sant'Anna e San Sebastiano' Caserta Hospital / 7.PO 'Maria Santissima Addolorata' Eboli Hospital / 8.Biogem Istituto di Ricerche Genetiche / 9. U.O.C. di Genetica Medica e di Laboratorio A.O.R. N., Azienda Ospedaliera di Rilievo Nazionale Antonio Cardarelli, Napoli / 10. Centro di riferimento Oncologico della Basilicata (IRCCS-CROB), Rionero in Vulture (PZ) / 11. Presidio Ospedaliero di Agropoli, Agropoli (SA). | 1. Genome Research Center for Health (CRGS) / 2. Laboratory of Molecular Medicine and Genomics(LMMGe) / 3. Center for Research in Pure and Applied Mathematics (CRMPA) | Giorgio Giurato (Corresponding Author) et a |
| EPI_ISL_1298510 | ASL Napoli 1 Centro | AMES Centro Polidiagnostico Strumentale S.r.l. | Giovanni Savarese et al |
| EPI_ISL_1298614 | ASL Napoli 1 Centro | AMES Centro Polidiagnostico Strumentale S.r.l. | Giovanni Savarese et al |
| EPI_ISL_516620 | Instituto de Diagnostico y Referencia Epidemiologicos (INDRE) | Instituto de Diagnostico y Referencia Epidemiologicos (INDRE) | Gisela Barrera-Badillo et al |
| EPI_ISL_806623 | KEMRI-Wellcome Trust Research Programme/KEMRI-CGMR-C Kilifi | KEMRI-Wellcome Trust Research Programme/KEMRI-CGMR-C Kilifi | Githinji et al |
| EPI_ISL_806624 | KEMRI-Wellcome Trust Research Programme/KEMRI-CGMR-C Kilifi | KEMRI-Wellcome Trust Research Programme/KEMRI-CGMR-C Kilifi | Githinji et al |
| EPI_ISL_806681 | KEMRI-Wellcome Trust Research Programme/KEMRI-CGMR-C Kilifi | KEMRI-Wellcome Trust Research Programme/KEMRI-CGMR-C Kilifi | Githinji et al |
| EPI_ISL_806630 | KEMRI-Wellcome Trust Research Programme/KEMRI-CGMR-C Kilifi | KEMRI-Wellcome Trust Research Programme/KEMRI-CGMR-C Kilifi | Githinji et al |
| EPI_ISL_806632 | KEMRI-Wellcome Trust Research Programme/KEMRI-CGMR-C Kilifi | KEMRI-Wellcome Trust Research Programme/KEMRI-CGMR-C Kilifi | Githinji et al |
| EPI_ISL_806554 | KEMRI-Wellcome Trust Research Programme/KEMRI-CGMR-C Kilifi | KEMRI-Wellcome Trust Research Programme/KEMRI-CGMR-C Kilifi | Githinji et al |
| EPI_ISL_806558 | KEMRI-Wellcome Trust Research Programme/KEMRI-CGMR-C Kilifi | KEMRI-Wellcome Trust Research Programme/KEMRI-CGMR-C Kilifi | Githinji et al |
| EPI_ISL_806566 | KEMRI-Wellcome Trust Research Programme/KEMRI-CGMR-C Kilifi | KEMRI-Wellcome Trust Research Programme/KEMRI-CGMR-C Kilifi | Githinji et al |
| EPI_ISL_806570 | KEMRI-Wellcome Trust Research Programme/KEMRI-CGMR-C Kilifi | KEMRI-Wellcome Trust Research Programme/KEMRI-CGMR-C Kilifi | Githinji et al |
| EPI_ISL_806709 | KEMRI-Wellcome Trust Research Programme/KEMRI-CGMR-C Kilifi | KEMRI-Wellcome Trust Research Programme/KEMRI-CGMR-C Kilifi | Githinji et al |
| EPI_ISL_806710 | KEMRI-Wellcome Trust Research Programme/KEMRI-CGMR-C Kilifi | KEMRI-Wellcome Trust Research Programme/KEMRI-CGMR-C Kilifi | Githinji et al |
| EPI_ISL_806712 | KEMRI-Wellcome Trust Research Programme/KEMRI-CGMR-C Kilifi | KEMRI-Wellcome Trust Research Programme/KEMRI-CGMR-C Kilifi | Githinji et al |
| EPI_ISL_806715 | KEMRI-Wellcome Trust Research Programme/KEMRI-CGMR-C Kilifi | KEMRI-Wellcome Trust Research Programme/KEMRI-CGMR-C Kilifi | Githinji et al |
| EPI_ISL_806716 | KEMRI-Wellcome Trust Research Programme/KEMRI-CGMR-C Kilifi | KEMRI-Wellcome Trust Research Programme/KEMRI-CGMR-C Kilifi | Githinji et al |
| EPI_ISL_806611 | KEMRI-Wellcome Trust Research Programme/KEMRI-CGMR-C Kilifi | KEMRI-Wellcome Trust Research Programme/KEMRI-CGMR-C Kilifi | Githinji et al |
| EPI_ISL_806617 | KEMRI-Wellcome Trust Research Programme/KEMRI-CGMR-C Kilifi | KEMRI-Wellcome Trust Research Programme/KEMRI-CGMR-C Kilifi | Githinji et al |
| EPI_ISL_968881 | KEMRI-Wellcome Trust Research Programme/KEMRI-CGMR-C Kilifi | KEMRI-Wellcome Trust Research Programme/KEMRI-CGMR-C Kilifi | Githinji et al |
| EPI_ISL_968892 | KEMRI-Wellcome Trust Research Programme/KEMRI-CGMR-C Kilifi | KEMRI-Wellcome Trust Research Programme/KEMRI-CGMR-C Kilifi | Githinji et al |
| EPI_ISL_855536 | KEMRI-Wellcome Trust Research Programme/KEMRI-CGMR-C Kilifi | KEMRI-Wellcome Trust Research Programme/KEMRI-CGMR-C Kilifi | Githinji et al |
| EPI_ISL_968901 | KEMRI-Wellcome Trust Research Programme/KEMRI-CGMR-C Kilifi | KEMRI-Wellcome Trust Research Programme/KEMRI-CGMR-C Kilifi | Githinji et al |
| EPI_ISL_855533 | KEMRI-Wellcome Trust Research Programme/KEMRI-CGMR-C Kilifi | KEMRI-Wellcome Trust Research Programme/KEMRI-CGMR-C Kilifi | Githinji et al |

| | | | |
|---|---|---|---|
| EPI_ISL_968902 | KEMRI-Wellcome Trust Research Programme/KEMRI-CGMR-C Kilifi | KEMRI-Wellcome Trust Research Programme/KEMRI-CGMR-C Kilifi | Githinji et al |
| EPI_ISL_968903 | KEMRI-Wellcome Trust Research Programme/KEMRI-CGMR-C Kilifi | KEMRI-Wellcome Trust Research Programme/KEMRI-CGMR-C Kilifi | Githinji et al |
| EPI_ISL_855538 | KEMRI-Wellcome Trust Research Programme/KEMRI-CGMR-C Kilifi | KEMRI-Wellcome Trust Research Programme/KEMRI-CGMR-C Kilifi | Githinji et al |
| EPI_ISL_968911 | KEMRI-Wellcome Trust Research Programme/KEMRI-CGMR-C Kilifi | KEMRI-Wellcome Trust Research Programme/KEMRI-CGMR-C Kilifi | Githinji et al |
| EPI_ISL_968930 | KEMRI-Wellcome Trust Research Programme/KEMRI-CGMR-C Kilifi | KEMRI-Wellcome Trust Research Programme/KEMRI-CGMR-C Kilifi | Githinji et al |
| EPI_ISL_1039225 | KEMRI-Wellcome Trust Research Programme/KEMRI-CGMR-C Kilifi | KEMRI-Wellcome Trust Research Programme/KEMRI-CGMR-C Kilifi | Githinji et al |
| EPI_ISL_968997 | KEMRI-Wellcome Trust Research Programme/KEMRI-CGMR-C Kilifi | KEMRI-Wellcome Trust Research Programme/KEMRI-CGMR-C Kilifi | Githinji et al |
| EPI_ISL_855535 | KEMRI-Wellcome Trust Research Programme/KEMRI-CGMR-C Kilifi | KEMRI-Wellcome Trust Research Programme/KEMRI-CGMR-C Kilifi | Githinji et al |
| EPI_ISL_969003 | KEMRI-Wellcome Trust Research Programme/KEMRI-CGMR-C Kilifi | KEMRI-Wellcome Trust Research Programme/KEMRI-CGMR-C Kilifi | Githinji et al |
| EPI_ISL_969017 | KEMRI-Wellcome Trust Research Programme/KEMRI-CGMR-C Kilifi | KEMRI-Wellcome Trust Research Programme/KEMRI-CGMR-C Kilifi | Githinji et al |
| EPI_ISL_568708 | KEMRI-Wellcome Trust Research Programme/KEMRI-CGMR-C Kilifi | KEMRI-Wellcome Trust Research Programme/KEMRI-CGMR-C Kilifi | Githinji et al 2020 et al |
| EPI_ISL_568790 | KEMRI-Wellcome Trust Research Programme/KEMRI-CGMR-C Kilifi | KEMRI-Wellcome Trust Research Programme/KEMRI-CGMR-C Kilifi | Githinji et al 2020 et al |
| EPI_ISL_568804 | KEMRI-Wellcome Trust Research Programme/KEMRI-CGMR-C Kilifi | KEMRI-Wellcome Trust Research Programme/KEMRI-CGMR-C Kilifi | Githinji et al 2020 et al |
| EPI_ISL_568815 | KEMRI-Wellcome Trust Research Programme/KEMRI-CGMR-C Kilifi | KEMRI-Wellcome Trust Research Programme/KEMRI-CGMR-C Kilifi | Githinji et al 2020 et al |
| EPI_ISL_568863 | KEMRI-Wellcome Trust Research Programme/KEMRI-CGMR-C Kilifi | KEMRI-Wellcome Trust Research Programme/KEMRI-CGMR-C Kilifi | Githinji et al 2020 et al |
| EPI_ISL_1440078 | KEMRI-Wellcome Trust Research Programme,Kilifi | KEMRI-Wellcome Trust Research Programme,Kilifi | Githinji G. et al |
| EPI_ISL_1440084 | KEMRI-Wellcome Trust Research Programme,Kilifi | KEMRI-Wellcome Trust Research Programme,Kilifi | Githinji G. et al |
| EPI_ISL_1440102 | KEMRI-Wellcome Trust Research Programme,Kilifi | KEMRI-Wellcome Trust Research Programme,Kilifi | Githinji G. et al |
| EPI_ISL_1440106 | KEMRI-Wellcome Trust Research Programme,Kilifi | KEMRI-Wellcome Trust Research Programme,Kilifi | Githinji G. et al |
| EPI_ISL_1440096 | KEMRI-Wellcome Trust Research Programme,Kilifi | KEMRI-Wellcome Trust Research Programme,Kilifi | Githinji G. et al |
| EPI_ISL_1440115 | KEMRI-Wellcome Trust Research Programme,Kilifi | KEMRI-Wellcome Trust Research Programme,Kilifi | Githinji G. et al |
| EPI_ISL_1440118 | KEMRI-Wellcome Trust Research Programme,Kilifi | KEMRI-Wellcome Trust Research Programme,Kilifi | Githinji G. et al |
| EPI_ISL_1440119 | KEMRI-Wellcome Trust Research Programme,Kilifi | KEMRI-Wellcome Trust Research Programme,Kilifi | Githinji G. et al |
| EPI_ISL_1440136 | KEMRI-Wellcome Trust Research Programme,Kilifi | KEMRI-Wellcome Trust Research Programme,Kilifi | Githinji G. et al |
| EPI_ISL_1440181 | KEMRI-Wellcome Trust Research Programme,Kilifi | KEMRI-Wellcome Trust Research Programme,Kilifi | Githinji G. et al |
| EPI_ISL_1440292 | KEMRI-Wellcome Trust Research Programme,Kilifi | KEMRI-Wellcome Trust Research Programme,Kilifi | Githinji G. et al |
| EPI_ISL_1610624 | KEMRI Center for Biotechnology Research and Development | KEMRI-Wellcome Trust Research Programme,Kilifi | Githinji G. et al |
| EPI_ISL_1610629 | KEMRI Center for Biotechnology Research and Development | KEMRI-Wellcome Trust Research Programme,Kilifi | Githinji G. et al |
| EPI_ISL_457874 | KEMRI-CGMR-C | KEMRI-Wellcome Trust Research Programme/KEMRI-CGMR-C Kilifi | Githinji G. et al 2020 et al |
| EPI_ISL_457876 | KEMRI-CGMR-C | KEMRI-Wellcome Trust Research Programme/KEMRI-CGMR-C Kilifi | Githinji G. et al 2020 et al |

| | | | |
|---|---|---|---|
| EPI_ISL_457890 | KEMRI-CGMR-C | KEMRI-Wellcome Trust Research Programme/KEMRI-CGMR-C Kilifi | Githinji G. et al 2020 et al |
| EPI_ISL_457916 | KEMRI-CGMR-C | KEMRI-Wellcome Trust Research Programme/KEMRI-CGMR-C Kilifi | Githinji G. et al 2020 et al |
| EPI_ISL_457836 | National Public Health Laboratory | KEMRI-Wellcome Trust Research Programme/KEMRI-CGMR-C Kilifi | Githinji G. et al 2020 et al |
| EPI_ISL_457932 | KEMRI-Centre for Virus Research | KEMRI-Wellcome Trust Research Programme/KEMRI-CGMR-C Kilifi | Githinji G. et al 2020 et al |
| EPI_ISL_457933 | KEMRI-Centre for Virus Research | KEMRI-Wellcome Trust Research Programme/KEMRI-CGMR-C Kilifi | Githinji G. et al 2020 et al |
| EPI_ISL_419687 | Servicio de Microbiología. Consorcio Hospital General Universitario de Valencia | Sequencing and Bioinformatics Service and Molecular Epidemiology Research Group. FISABIO-Public Health | Giuseppe D'Auria et al |
| EPI_ISL_512811 | Kenema Government Hospital, Ministry of Health and Sanitation | Kenema Government Hospital, Ministry of Health and Sanitation | Goba et al |
| EPI_ISL_512812 | Kenema Government Hospital, Ministry of Health and Sanitation | Kenema Government Hospital, Ministry of Health and Sanitation | Goba et al |
| EPI_ISL_512821 | Kenema Government Hospital, Ministry of Health and Sanitation | Kenema Government Hospital, Ministry of Health and Sanitation | Goba et al |
| EPI_ISL_462753 | University Clinical Hospital of Mostar | University of Sarajevo Veterinary Faculty | Goletic et al |
| EPI_ISL_1016885 | University of Sarajevo, Veterinary Faculty, Laboratory for Molecular Diagnostic and Research Laboratory | University of Sarajevo, Veterinary Faculty, Laboratory for Molecular Diagnostic and Research Laboratory | Goletic S. et al |
| EPI_ISL_1016970 | University of Sarajevo, Veterinary Faculty, Laboratory for Molecular Diagnostic and Research Laboratory | University of Sarajevo, Veterinary Faculty, Laboratory for Molecular Diagnostic and Research Laboratory | Goletic S. et al |
| EPI_ISL_955169 | University of Sarajevo, Veterinary Faculty, Laboratory for Molecular Diagnostic and Research Laboratory | University of Sarajevo, Veterinary Faculty, Laboratory for Molecular Diagnostic and Research Laboratory | Goletić Š. et al |
| EPI_ISL_955171 | University of Sarajevo, Veterinary Faculty, Laboratory for Molecular Diagnostic and Research Laboratory | University of Sarajevo, Veterinary Faculty, Laboratory for Molecular Diagnostic and Research Laboratory | Goletić Š. et al |
| EPI_ISL_955173 | University of Sarajevo, Veterinary Faculty, Laboratory for Molecular Diagnostic and Research Laboratory | University of Sarajevo, Veterinary Faculty, Laboratory for Molecular Diagnostic and Research Laboratory | Goletić Š. et al |
| EPI_ISL_955211 | University of Sarajevo, Veterinary Faculty, Laboratory for Molecular Diagnostic and Research Laboratory | University of Sarajevo, Veterinary Faculty, Laboratory for Molecular Diagnostic and Research Laboratory | Goletić Š. et al |
| EPI_ISL_1016690 | University of Sarajevo, Veterinary Faculty, Laboratory for Molecular Diagnostic and Research Laboratory | University of Sarajevo, Veterinary Faculty, Laboratory for Molecular Diagnostic and Research Laboratory | Goletic T. et al |
| EPI_ISL_1016884 | University of Sarajevo, Veterinary Faculty, Laboratory for Molecular Diagnostic and Research Laboratory | University of Sarajevo, Veterinary Faculty, Laboratory for Molecular Diagnostic and Research Laboratory | Goletic T. et al |
| EPI_ISL_955142 | University of Sarajevo, Veterinary Faculty, Laboratory for Molecular Diagnostic and Research Laboratory | University of Sarajevo, Veterinary Faculty, Laboratory for Molecular Diagnostic and Research Laboratory | Goletić T. et al |
| EPI_ISL_955144 | University of Sarajevo, Veterinary Faculty, Laboratory for Molecular Diagnostic and Research Laboratory | University of Sarajevo, Veterinary Faculty, Laboratory for Molecular Diagnostic and Research Laboratory | Goletić T. et al |
| EPI_ISL_955146 | University of Sarajevo, Veterinary Faculty, Laboratory for Molecular Diagnostic and Research Laboratory | University of Sarajevo, Veterinary Faculty, Laboratory for Molecular Diagnostic and Research Laboratory | Goletić T. et al |
| EPI_ISL_955148 | University of Sarajevo, Veterinary Faculty, Laboratory for Molecular Diagnostic and Research Laboratory | University of Sarajevo, Veterinary Faculty, Laboratory for Molecular Diagnostic and Research Laboratory | Goletić T. et al |
| EPI_ISL_955150 | University of Sarajevo, Veterinary Faculty, Laboratory for Molecular Diagnostic and Research Laboratory | University of Sarajevo, Veterinary Faculty, Laboratory for Molecular Diagnostic and Research Laboratory | Goletić T. et al |
| EPI_ISL_955151 | University of Sarajevo, Veterinary Faculty, Laboratory for Molecular Diagnostic and Research Laboratory | University of Sarajevo, Veterinary Faculty, Laboratory for Molecular Diagnostic and Research Laboratory | Goletić T. et al |

| | | | |
|---|---|---|---|
| EPI_ISL_955155 | University of Sarajevo, Veterinary Faculty, Laboratory for Molecular Diagnostic and Research Laboratory | University of Sarajevo, Veterinary Faculty, Laboratory for Molecular Diagnostic and Research Laboratory | Goletić T. et al |
| EPI_ISL_955156 | University of Sarajevo, Veterinary Faculty, Laboratory for Molecular Diagnostic and Research Laboratory | University of Sarajevo, Veterinary Faculty, Laboratory for Molecular Diagnostic and Research Laboratory | Goletić T. et al |
| EPI_ISL_955170 | University of Sarajevo, Veterinary Faculty, Laboratory for Molecular Diagnostic and Research Laboratory | University of Sarajevo, Veterinary Faculty, Laboratory for Molecular Diagnostic and Research Laboratory | Goletić T. et al |
| EPI_ISL_1502889 | Gorgas Memorial Laboratory of Health Studies | Gorgas Memorial Laboratory of Health Studies | Gonzalez Claudia et al |
| EPI_ISL_1502895 | Gorgas Memorial Laboratory of Health Studies | Gorgas Memorial Laboratory of Health Studies | Gonzalez Claudia et al |
| EPI_ISL_1502903 | Gorgas Memorial Laboratory of Health Studies | Gorgas Memorial Laboratory of Health Studies | Gonzalez Claudia et al |
| EPI_ISL_1502908 | Gorgas Memorial Laboratory of Health Studies | Gorgas Memorial Laboratory of Health Studies | Gonzalez Claudia et al |
| EPI_ISL_1502945 | Gorgas Memorial Laboratory of Health Studies | Gorgas Memorial Laboratory of Health Studies | Gonzalez Claudia et al |
| EPI_ISL_1502980 | Gorgas Memorial Laboratory of Health Studies | Gorgas Memorial Laboratory of Health Studies | Gonzalez Claudia et al |
| EPI_ISL_1502982 | Gorgas Memorial Laboratory of Health Studies | Gorgas Memorial Laboratory of Health Studies | Gonzalez Claudia et al |
| EPI_ISL_1503122 | Gorgas Memorial Laboratory of Health Studies | Gorgas Memorial Laboratory of Health Studies | Gonzalez Claudia et al |
| EPI_ISL_1503124 | Hospital Regional Anita Moreno Los Santos | Gorgas Memorial Laboratory of Health Studies | Gonzalez Claudia et al |
| EPI_ISL_1503134 | Gorgas Memorial Laboratory of Health Studies | Gorgas Memorial Laboratory of Health Studies | Gonzalez Claudia et al |
| EPI_ISL_1503139 | Gorgas Memorial Laboratory of Health Studies | Gorgas Memorial Laboratory of Health Studies | Gonzalez Claudia et al |
| EPI_ISL_1503017 | Gorgas Memorial Laboratory of Health Studies | Gorgas Memorial Laboratory of Health Studies | Gonzalez Claudia et al |
| EPI_ISL_1503021 | Gorgas Memorial Laboratory of Health Studies | Gorgas Memorial Laboratory of Health Studies | Gonzalez Claudia et al |
| EPI_ISL_1503024 | Gorgas Memorial Laboratory of Health Studies | Gorgas Memorial Laboratory of Health Studies | Gonzalez Claudia et al |
| EPI_ISL_1503051 | Gorgas Memorial Laboratory of Health Studies | Gorgas Memorial Laboratory of Health Studies | Gonzalez Claudia et al |
| EPI_ISL_1503052 | Gorgas Memorial Laboratory of Health Studies | Gorgas Memorial Laboratory of Health Studies | Gonzalez Claudia et al |
| EPI_ISL_1503054 | Gorgas Memorial Laboratory of Health Studies | Gorgas Memorial Laboratory of Health Studies | Gonzalez Claudia et al |
| EPI_ISL_1503065 | Gorgas Memorial Laboratory of Health Studies | Gorgas Memorial Laboratory of Health Studies | Gonzalez Claudia et al |
| EPI_ISL_1503084 | Gorgas Memorial Laboratory of Health Studies | Gorgas Memorial Laboratory of Health Studies | Gonzalez Claudia et al |
| EPI_ISL_1503086 | Gorgas Memorial Laboratory of Health Studies | Gorgas Memorial Laboratory of Health Studies | Gonzalez Claudia et al |
| EPI_ISL_1502867 | Hospital Regional Anita Moreno Los Santos | Gorgas Memorial Laboratory of Health Studies | Gonzalez Claudia et al |
| EPI_ISL_1502879 | Gorgas Memorial Laboratory of Health Studies | Gorgas Memorial Laboratory of Health Studies | Gonzalez Claudia et al |
| EPI_ISL_1502882 | Gorgas Memorial Laboratory of Health Studies | Gorgas Memorial Laboratory of Health Studies | Gonzalez Claudia et al |
| EPI_ISL_1502883 | Gorgas Memorial Laboratory of Health Studies | Gorgas Memorial Laboratory of Health Studies | Gonzalez Claudia et al |
| EPI_ISL_1502884 | Gorgas Memorial Laboratory of Health Studies | Gorgas Memorial Laboratory of Health Studies | Gonzalez Claudia et al |
| EPI_ISL_414476 | MSHS Clinical Microbiology Laboratories | MSHS Pathogen Surveillance Program | Gopi Patel et al |

| | | | |
|---|---|---|---|
| EPI_ISL_1273391 | National Reference Laboratory - Ministry of Health Maseru Lesotho | National Institute for Communicable Diseases of the National Health Laboratory Service | Gorova V et al |
| EPI_ISL_1273392 | National Reference Laboratory - Ministry of Health Maseru Lesotho | National Institute for Communicable Diseases of the National Health Laboratory Service | Gorova V et al |
| EPI_ISL_1273393 | National Reference Laboratory - Ministry of Health Maseru Lesotho | National Institute for Communicable Diseases of the National Health Laboratory Service | Gorova V et al |
| EPI_ISL_1273394 | National Reference Laboratory - Ministry of Health Maseru Lesotho | National Institute for Communicable Diseases of the National Health Laboratory Service | Gorova V et al |
| EPI_ISL_1273395 | National Reference Laboratory - Ministry of Health Maseru Lesotho | National Institute for Communicable Diseases of the National Health Laboratory Service | Gorova V et al |
| EPI_ISL_1273399 | National Reference Laboratory - Ministry of Health Maseru Lesotho | National Institute for Communicable Diseases of the National Health Laboratory Service | Gorova V et al |
| EPI_ISL_1273401 | National Reference Laboratory - Ministry of Health Maseru Lesotho | National Institute for Communicable Diseases of the National Health Laboratory Service | Gorova V et al |
| EPI_ISL_1273406 | National Reference Laboratory - Ministry of Health Maseru Lesotho | National Institute for Communicable Diseases of the National Health Laboratory Service | Gorova V et al |
| EPI_ISL_1273407 | National Reference Laboratory - Ministry of Health Maseru Lesotho | National Institute for Communicable Diseases of the National Health Laboratory Service | Gorova V et al |
| EPI_ISL_1273408 | National Reference Laboratory - Ministry of Health Maseru Lesotho | National Institute for Communicable Diseases of the National Health Laboratory Service | Gorova V et al |
| EPI_ISL_427731 | Centre for Infectious Diseases and Microbiology Public Health | NSW Health Pathology - Institute of Clinical Pathology and Medical Research; Westmead Hospital; University of Sydney | Gray K et al |
| EPI_ISL_999032 | Institut Pasteur de Guinée | Institut Pasteur de Dakar | Grayo Solene et al |
| EPI_ISL_416485 | Servicio de Microbiologia. Consorcio Hospital General Universitario de Valencia | Sequencing and Bioinformatics Service and Molecular Epidemiology Research Group. FISABIO-Public Health | Griselda De Marco et al |
| EPI_ISL_1055164 | National Virus Reference Laboratory | National Virus Reference Laboratory | Guerrino Macori et al |
| EPI_ISL_426361 | Instituto Nacional de Ciencias Medicas y Nutricion Salvador Zubiran | Instituto Nacional de Ciencias Medicas y Nutricion Salvador Zubiran | Guillermo M. Ruiz-Palacios et al |
| EPI_ISL_538142 | Servicio de Microbiología y Parasitología clínica. UCEIMP. Hospital Universitario Virgen del Rocío/IBIS/CSIC/US | SeqCOVID-SPAIN consortium/IBV(CSIC) | Guillermo Martín Gutiérrez et al |
| EPI_ISL_610162 | RSUD Dr. Tjitrowardojo | Genetics Working Group (Pokja Genetik) Faculty of Medicine, Public Health and Nursing Universitas Gadjah Mada (FK-KMK UGM); Disease Investigation Center Wates Ministry of Agriculture Indonesia; Department of Microbiology FK-KMK UGM; Laboratorium Diagnostik Yayasan Tahija World Mosquito Program (WMP) Yogyakarta Center for Tropical Medicine FK-KMK UGM; Integrated Research Center FK-KMK UGM; Department of Computer Science and Electronics FMIPA UGM; Balai Besar Teknik Kesehatan Lingkungan dan Pengendalian Penyakit (BBTKLPP) Yogyakarta | Gunadi et al |
| EPI_ISL_516806 | Rumah Sakit PKU Gamping | Genetics Working Group (Pokja Genetik) Faculty of Medicine, Public Health and Nursing Universitas Gadjah Mada (FK-KMK UGM); Disease Investigation Center Wates Ministry of Agriculture Indonesia; Department of Microbiology FK-KMK UGM; Laboratorium Diagnostik Yayasan Tahija World Mosquito Program (WMP) Yogyakarta Center for Tropical Medicine FK-KMK UGM; Integrated Research center FK-KMK UGM; Department of Computer Science and Electronics FMIPA UGM | Gunadi et al |

| | | | |
|---|---|---|---|
| EPI_ISL_537558 | UCLA Pathology Clinical Microbiology Lab | Kruglyak Lab | Guo et al. |
| EPI_ISL_509615 | Servicio de Microbiología. Hospital Universitario Donostia. OSI Donostialdea. Área de Enfermedades Infecciosas, Grupo de Infección Respiratoria y Resistencia Antimicrobiana. Instituto de Investigación Sanitaria Biodonostia | SeqCOVID-SPAIN consortium/IBV(CSIC) | Gustavo Cilla et al |
| EPI_ISL_509610 | Servicio de Microbiología. Hospital Universitario Donostia. OSI Donostialdea. Área de Enfermedades Infecciosas, Grupo de Infección Respiratoria y Resistencia Antimicrobiana. Instituto de Investigación Sanitaria Biodonostia | SeqCOVID-SPAIN consortium/IBV(CSIC) | Gustavo Cilla et al |
| EPI_ISL_565921 | Servicio de Microbiología. Hospital Universitario Donostia. OSI Donostialdea. Área de Enfermedades Infecciosas, Grupo de Infección Respiratoria y Resistencia Antimicrobiana. Instituto de Investigación Sanitaria Biodonostia | SeqCOVID-SPAIN consortium/IBV(CSIC) | Gustavo Cilla et al |
| EPI_ISL_528637 | LVM/UFRJ | Bioinformatics Laboratory / LNCC | Gustavo D. P. Silva et al |
| EPI_ISL_470877 | Department for Virology, Molecular Biology and Genome Research, R. G. Lugar Center for Public Health Research, National Center for Disease Control and Public Health (NCDC) of Georgia. | Department for Virology, Molecular Biology and Genome Research, R. G. Lugar Center for Public Health Research, National Center for Disease Control and Public Health (NCDC) of Georgia. | Gvantsa Brachveli et al |
| EPI_ISL_1048367 | Department for Virology, Molecular Biology and Genome Research, R. G. Lugar Center for Public Health Research, National Center for Disease Control and Public Health (NCDC) of Georgia. | Department for Virology, Molecular Biology and Genome Research, R. G. Lugar Center for Public Health Research, National Center for Disease Control and Public Health (NCDC) of Georgia. | Gvantsa Brachveli et al |
| EPI_ISL_1005231 | Vilnius university hospital Santaros Klinikos, Center of Laboratory Medicine | Vilnius university hospital Santaros Klinikos, Center of Laboratory Medicine | Gytis Dudas et al |
| EPI_ISL_1005225 | Vilnius university hospital Santaros Klinikos, Center of Laboratory Medicine | Vilnius university hospital Santaros Klinikos, Center of Laboratory Medicine | Gytis Dudas et al |
| EPI_ISL_1005227 | Vilnius university hospital Santaros Klinikos, Center of Laboratory Medicine | Vilnius university hospital Santaros Klinikos, Center of Laboratory Medicine | Gytis Dudas et al |
| EPI_ISL_1005228 | Vilnius university hospital Santaros Klinikos, Center of Laboratory Medicine | Vilnius university hospital Santaros Klinikos, Center of Laboratory Medicine | Gytis Dudas et al |
| EPI_ISL_1005230 | Vilnius university hospital Santaros Klinikos, Center of Laboratory Medicine | Vilnius university hospital Santaros Klinikos, Center of Laboratory Medicine | Gytis Dudas et al |
| EPI_ISL_1661599 | Life Sciences Center, Vilnius University | Vilnius University Hospital Santaros Klinikos, Center of Laboratory Medicine | Gytis Dudas et al |
| EPI_ISL_1661660 | Vilnius University Hospital Santaros Klinikos | Vilnius University Hospital Santaros Klinikos, Center of Laboratory Medicine | Gytis Dudas et al |
| EPI_ISL_1661664 | UAB Medicina practica laboratorija | Vilnius University Hospital Santaros Klinikos, Center of Laboratory Medicine | Gytis Dudas et al |
| EPI_ISL_1661745 | Vilnius University Hospital Santaros Klinikos | Vilnius University Hospital Santaros Klinikos, Center of Laboratory Medicine | Gytis Dudas et al |
| EPI_ISL_1661761 | Anteja laboratorija (UAB Diagnostikos laboratorija) | Vilnius University Hospital Santaros Klinikos, Center of Laboratory Medicine | Gytis Dudas et al |
| EPI_ISL_1133254 | University of Chittagong | Central Biological Research Laboratory and Department of Biochemistry and Molecular Biology | H. M. Abdullah Al Masud et al |
| EPI_ISL_735500 | University of Chittagong | Central Biological Research Laboratory and Department of Biochemistry and Molecular Biology | H. M. Abdullah Al Masud et al |
| EPI_ISL_476559 | unknown | Laboratoire Sciences et Technologies de la Santé (STS) Institut Supérieur des Sciences de la Santé Université Hassan 1er, Settat, Morocco | Hajar Lemriss et al |
| EPI_ISL_1107189 | Laboratoire central de Virologie | Laboratoire de Biotechnologie | Hakima Kabbaj et al |
| EPI_ISL_1116470 | Laboratoire central de Virologie | Laboratoire de Biotechnologie | Hakima Kabbaj et al |
| EPI_ISL_1117289 | Laboratoire central de Virologie | Laboratoire de Biotechnologie | Hakima Kabbaj et al |
| EPI_ISL_754179 | Specialized Lab for COVID-19 Detection, Department of Genetic Engineering and Biotechnology | Specialized Lab for COVID-19 Detection, Department of Genetic Engineering and Biotechnology | Hammadul Hoque et al |

| | | | |
|---|---|---|---|
| EPI_ISL_1257859 | KKP Soekarno Hatta | National Institute of Health Research and Development | Hana Apsari Pawestri et al |
| EPI_ISL_458286 | unknown | Bundeswehr Institute of Microbiology | Handrick et al |
| EPI_ISL_833336 | Bureau of Quarantine | Research Institute for Tropical Medicine | Hannah Leah Morito et al |
| EPI_ISL_833338 | Siniloan Rural Health Unit | Research Institute for Tropical Medicine | Hannah Leah Morito et al |
| EPI_ISL_833334 | Batangas City Health Office | Research Institute for Tropical Medicine | Hannah Leah Morito et al |
| EPI_ISL_833333 | Batangas City Health Office | Research Institute for Tropical Medicine | Hannah Leah Morito et al |
| EPI_ISL_1069188 | Lumban Rural Health Unit | Research Institute for Tropical Medicine | Hannah Leah Morito et al |
| EPI_ISL_1332747 | Lighthouse Lab in Glasgow | Wellcome Sanger Institute for the COVID-19 Genomics UK (COG-UK) Consortium | Harper VanSteenhouse et al |
| EPI_ISL_532970 | Lighthouse Lab in Glasgow | Wellcome Sanger Institute for the COVID-19 Genomics UK (COG-UK) consortium | Harper VanSteenhouse et al |
| EPI_ISL_549568 | Lighthouse Lab in Glasgow | Wellcome Sanger Institute for the COVID-19 Genomics UK (COG-UK) consortium | Harper VanSteenhouse et al |
| EPI_ISL_567404 | Lighthouse Lab in Glasgow | Wellcome Sanger Institute for the COVID-19 Genomics UK (COG-UK) consortium | Harper VanSteenhouse et al |
| EPI_ISL_567449 | Lighthouse Lab in Glasgow | Wellcome Sanger Institute for the COVID-19 Genomics UK (COG-UK) consortium | Harper VanSteenhouse et al |
| EPI_ISL_567526 | Lighthouse Lab in Glasgow | Wellcome Sanger Institute for the COVID-19 Genomics UK (COG-UK) consortium | Harper VanSteenhouse et al |
| EPI_ISL_599760 | Lighthouse Lab in Glasgow | Wellcome Sanger Institute for the COVID-19 Genomics UK (COG-UK) consortium | Harper VanSteenhouse et al |
| EPI_ISL_587521 | Lighthouse Lab in Glasgow | Wellcome Sanger Institute for the COVID-19 Genomics UK (COG-UK) consortium | Harper VanSteenhouse et al |
| EPI_ISL_641980 | Lighthouse Lab in Glasgow | Wellcome Sanger Institute for the COVID-19 Genomics UK (COG-UK) Consortium | Harper VanSteenhouse et al |
| EPI_ISL_760740 | Lighthouse Lab in Glasgow | Wellcome Sanger Institute for the COVID-19 Genomics UK (COG-UK) Consortium | Harper VanSteenhouse et al |
| EPI_ISL_1507943 | Lighthouse Lab in Glasgow | Wellcome Sanger Institute for the COVID-19 Genomics UK (COG-UK) Consortium | Harper VanSteenhouse et al |
| EPI_ISL_1634819 | Lighthouse Lab in Glasgow | Wellcome Sanger Institute for the COVID-19 Genomics UK (COG-UK) Consortium | Harper VanSteenhouse et al |
| EPI_ISL_1634918 | Lighthouse Lab in Glasgow | Wellcome Sanger Institute for the COVID-19 Genomics UK (COG-UK) Consortium | Harper VanSteenhouse et al |
| EPI_ISL_415578 | BCCDC Public Health Laboratory | BCCDC Public Health Laboratory | Harrigan et al |
| EPI_ISL_415577 | BCCDC Public Health Laboratory | BCCDC Public Health Laboratory | Harrigan et al |
| EPI_ISL_1469256 | RS Khusus Paru Karawang | National Institute of Health Research and Development | Hartanti Dian Ikawati et al |
| EPI_ISL_1257851 | BBTKLPP Banjarbaru | National Institute of Health Research and Development | Hartanti Dian Ikawati et al |
| EPI_ISL_539573 | Centre de Recherches Medicales de Lambarene (CERMEL) | Department of Emerging Infectious Diseases, Institute of Tropical Medicine, Nagasaki University | Haruka Abe et al |

| | | | |
|---|---|---|---|
| EPI_ISL_539574 | Centre de Recherches Medicales de Lambarene (CERMEL) | Department of Emerging Infectious Diseases, Institute of Tropical Medicine, Nagasaki University | Haruka Abe et al |
| EPI_ISL_539575 | Centre de Recherches Medicales de Lambarene (CERMEL) | Department of Emerging Infectious Diseases, Institute of Tropical Medicine, Nagasaki University | Haruka Abe et al |
| EPI_ISL_539576 | Centre de Recherches Medicales de Lambarene (CERMEL) | Department of Emerging Infectious Diseases, Institute of Tropical Medicine, Nagasaki University | Haruka Abe et al |
| EPI_ISL_1498152 | Institute of Epidemiology, Disease Control and Research (IEDCR) | Institute for Developing Science and Health Initiatives (ideSHi) | Hassan Afrad et al |
| EPI_ISL_1498142 | Institute of Epidemiology, Disease Control and Research (IEDCR) | Institute for Developing Science and Health Initiatives (ideSHi) | Hassan Afrad et al |
| EPI_ISL_1498146 | Institute of Epidemiology, Disease Control and Research (IEDCR) | Institute for Developing Science and Health Initiatives (ideSHi) | Hassan Afrad et al |
| EPI_ISL_1582394 | Institute of Epidemiology, Disease Control and Research (IEDCR) | Institute for Developing Science and Health Initiatives (ideSHi) | Hassan Afrad et al |
| EPI_ISL_1582397 | Institute of Epidemiology, Disease Control and Research (IEDCR) | Institute for Developing Science and Health Initiatives (ideSHi) | Hassan Afrad et al |
| EPI_ISL_1582393 | Institute of Epidemiology, Disease Control and Research (IEDCR) | Institute for Developing Science and Health Initiatives (ideSHi) | Hassan Afrad et al |
| EPI_ISL_1587441 | Institute for Developing Science and Health Initiatives (ideSHi) | Institute for Developing Science and Health Initiatives (ideSHi) | Hassan Afrad et al |
| EPI_ISL_1587432 | Institute for Developing Science and Health Initiatives (ideSHi) | Institute for Developing Science and Health Initiatives (ideSHi) | Hassan Afrad et al |
| EPI_ISL_746318 | Genome Center | Genome Center | Hassan M. Al-Emran et al |
| EPI_ISL_812793 | Genomics Program, Children Cancer Hospital | Genomics Program, Children Cancer Hospital | Hatem et al |
| EPI_ISL_812800 | Genomics Program, Children Cancer Hospital | Genomics Program, Children Cancer Hospital | Hatem et al |
| EPI_ISL_812812 | Genomics Program, Children Cancer Hospital | Genomics Program, Children Cancer Hospital | Hatem et al |
| EPI_ISL_812815 | Genomics Program, Children Cancer Hospital | Genomics Program, Children Cancer Hospital | Hatem et al |
| EPI_ISL_812820 | Genomics Program, Children Cancer Hospital | Genomics Program, Children Cancer Hospital | Hatem et al |
| EPI_ISL_812821 | Genomics Program, Children Cancer Hospital | Genomics Program, Children Cancer Hospital | Hatem et al |
| EPI_ISL_812824 | Genomics Program, Children Cancer Hospital | Genomics Program, Children Cancer Hospital | Hatem et al |
| EPI_ISL_812829 | Genomics Program, Children Cancer Hospital | Genomics Program, Children Cancer Hospital | Hatem et al |
| EPI_ISL_812837 | Genomics Program, Children Cancer Hospital | Genomics Program, Children Cancer Hospital | Hatem et al |
| EPI_ISL_812842 | Genomics Program, Children Cancer Hospital | Genomics Program, Children Cancer Hospital | Hatem et al |
| EPI_ISL_812846 | Genomics Program, Children Cancer Hospital | Genomics Program, Children Cancer Hospital | Hatem et al |
| EPI_ISL_812864 | Genomics Program, Children Cancer Hospital | Genomics Program, Children Cancer Hospital | Hatem et al |
| EPI_ISL_812872 | Genomics Program, Children Cancer Hospital | Genomics Program, Children Cancer Hospital | Hatem et al |
| EPI_ISL_1647215 | Dept. of Microbiology and Infection Control, Akershus University Hospital HF | Dept. of Microbiology and Infection Control, Akershus University Hospital HF | Hege Vangstein Aamot et al |
| EPI_ISL_1647217 | Dept. of Microbiology and Infection Control, Akershus University Hospital HF | Dept. of Microbiology and Infection Control, Akershus University Hospital HF | Hege Vangstein Aamot et al |
| EPI_ISL_1647256 | Dept. of Microbiology and Infection Control, Akershus University Hospital HF | Dept. of Microbiology and Infection Control, Akershus University Hospital HF | Hege Vangstein Aamot et al |

| | | | |
|---|---|---|---|
| EPI_ISL_1647260 | Dept. of Microbiology and Infection Control, Akershus University Hospital HF | Dept. of Microbiology and Infection Control, Akershus University Hospital HF | Hege Vangstein Aamot et al |
| EPI_ISL_665258 | Dept. of Microbiology and Infection Control, Akershus University Hospital HF | Dept. of Microbiology and Infection Control, Akershus University Hospital HF | Hege Vangstein Aamot et al |
| EPI_ISL_665259 | Dept. of Microbiology and Infection Control, Akershus University Hospital HF | Dept. of Microbiology and Infection Control, Akershus University Hospital HF | Hege Vangstein Aamot et al |
| EPI_ISL_665260 | Dept. of Microbiology and Infection Control, Akershus University Hospital HF | Dept. of Microbiology and Infection Control, Akershus University Hospital HF | Hege Vangstein Aamot et al |
| EPI_ISL_666596 | Dept. of Microbiology and Infection Control, Akershus University Hospital HF | Dept. of Microbiology and Infection Control, Akershus University Hospital HF | Hege Vangstein Aamot et al |
| EPI_ISL_666599 | Dept. of Microbiology and Infection Control, Akershus University Hospital HF | Dept. of Microbiology and Infection Control, Akershus University Hospital HF | Hege Vangstein Aamot et al |
| EPI_ISL_666601 | Dept. of Microbiology and Infection Control, Akershus University Hospital HF | Dept. of Microbiology and Infection Control, Akershus University Hospital HF | Hege Vangstein Aamot et al |
| EPI_ISL_666603 | Dept. of Microbiology and Infection Control, Akershus University Hospital HF | Dept. of Microbiology and Infection Control, Akershus University Hospital HF | Hege Vangstein Aamot et al |
| EPI_ISL_666604 | Dept. of Microbiology and Infection Control, Akershus University Hospital HF | Dept. of Microbiology and Infection Control, Akershus University Hospital HF | Hege Vangstein Aamot et al |
| EPI_ISL_445240 | Ulltuna Vardcentral | The Public Health Agency of Sweden | Heidi Lindback et al |
| EPI_ISL_450824 | Ulltuna Vardcentral | The Public Health Agency of Sweden | Heidi Lindback et al |
| EPI_ISL_1492070 | National Institute of Public Health | National Institute of Public Health | Helena Jirincova et al |
| EPI_ISL_1492117 | National Institute of Public Health | National Institute of Public Health | Helena Jirincova et al |
| EPI_ISL_1588586 | National Institute of Public Health | National Institute of Public Health | Helena Jirincova et al |
| EPI_ISL_1588589 | National Institute of Public Health | National Institute of Public Health | Helena Jirincova et al |
| EPI_ISL_895763 | Molecular biology division, Institute of Clinical Biochemistry and Diagnostics, Charles University, Faculty of Medicine in Hradec Králové and University Hospital Hradec Králové | Molecular biology division, Institute of Clinical Biochemistry and Diagnostics, Charles University, Faculty of Medicine in Hradec Králové and University Hospital Hradec Králové | Helena Kovaříková et al |
| EPI_ISL_895751 | Molecular biology division, Institute of Clinical Biochemistry and Diagnostics, Charles University, Faculty of Medicine in Hradec Králové and University Hospital Hradec Králové | Molecular biology division, Institute of Clinical Biochemistry and Diagnostics, Charles University, Faculty of Medicine in Hradec Králové and University Hospital Hradec Králové | Helena Kovaříková et al |
| EPI_ISL_895781 | Molecular biology division, Institute of Clinical Biochemistry and Diagnostics, Charles University, Faculty of Medicine in Hradec Králové and University Hospital Hradec Králové | Molecular biology division, Institute of Clinical Biochemistry and Diagnostics, Charles University, Faculty of Medicine in Hradec Králové and University Hospital Hradec Králové | Helena Kovaříková et al |
| EPI_ISL_895784 | Molecular biology division, Institute of Clinical Biochemistry and Diagnostics, Charles University, Faculty of Medicine in Hradec Králové and University Hospital Hradec Králové | Molecular biology division, Institute of Clinical Biochemistry and Diagnostics, Charles University, Faculty of Medicine in Hradec Králové and University Hospital Hradec Králové | Helena Kovaříková et al |
| EPI_ISL_895827 | Molecular biology division, Institute of Clinical Biochemistry and Diagnostics, Charles University, Faculty of Medicine in Hradec Králové and University Hospital Hradec Králové | Molecular biology division, Institute of Clinical Biochemistry and Diagnostics, Charles University, Faculty of Medicine in Hradec Králové and University Hospital Hradec Králové | Helena Kovaříková et al |
| EPI_ISL_895737 | Molecular biology division, Institute of Clinical Biochemistry and Diagnostics, Charles University, Faculty of Medicine in Hradec Králové and University Hospital Hradec Králové | Molecular biology division, Institute of Clinical Biochemistry and Diagnostics, Charles University, Faculty of Medicine in Hradec Králové and University Hospital Hradec Králové | Helena Kovaříková et al |

| | | | |
|---|---|---|---|
| EPI_ISL_895740 | Molecular biology division, Institute of Clinical Biochemistry and Diagnostics, Charles University, Faculty of Medicine in Hradec Králové and University Hospital Hradec Králové | Molecular biology division, Institute of Clinical Biochemistry and Diagnostics, Charles University, Faculty of Medicine in Hradec Králové and University Hospital Hradec Králové | Helena Kovaříková et al |
| EPI_ISL_895787 | Molecular biology division, Institute of Clinical Biochemistry and Diagnostics, Charles University, Faculty of Medicine in Hradec Králové and University Hospital Hradec Králové | Molecular biology division, Institute of Clinical Biochemistry and Diagnostics, Charles University, Faculty of Medicine in Hradec Králové and University Hospital Hradec Králové | Helena Kovaříková et al |
| EPI_ISL_895789 | Molecular biology division, Institute of Clinical Biochemistry and Diagnostics, Charles University, Faculty of Medicine in Hradec Králové and University Hospital Hradec Králové | Molecular biology division, Institute of Clinical Biochemistry and Diagnostics, Charles University, Faculty of Medicine in Hradec Králové and University Hospital Hradec Králové | Helena Kovaříková et al |
| EPI_ISL_895819 | Molecular biology division, Institute of Clinical Biochemistry and Diagnostics, Charles University, Faculty of Medicine in Hradec Králové and University Hospital Hradec Králové | Molecular biology division, Institute of Clinical Biochemistry and Diagnostics, Charles University, Faculty of Medicine in Hradec Králové and University Hospital Hradec Králové | Helena Kovaříková et al |
| EPI_ISL_895729 | Molecular biology division, Institute of Clinical Biochemistry and Diagnostics, Charles University, Faculty of Medicine in Hradec Králové and University Hospital Hradec Králové | Molecular biology division, Institute of Clinical Biochemistry and Diagnostics, Charles University, Faculty of Medicine in Hradec Králové and University Hospital Hradec Králové | Helena Kovaříková et al |
| EPI_ISL_895730 | Molecular biology division, Institute of Clinical Biochemistry and Diagnostics, Charles University, Faculty of Medicine in Hradec Králové and University Hospital Hradec Králové | Molecular biology division, Institute of Clinical Biochemistry and Diagnostics, Charles University, Faculty of Medicine in Hradec Králové and University Hospital Hradec Králové | Helena Kovaříková et al |
| EPI_ISL_895813 | Molecular biology division, Institute of Clinical Biochemistry and Diagnostics, Charles University, Faculty of Medicine in Hradec Králové and University Hospital Hradec Králové | Molecular biology division, Institute of Clinical Biochemistry and Diagnostics, Charles University, Faculty of Medicine in Hradec Králové and University Hospital Hradec Králové | Helena Kovaříková et al |
| EPI_ISL_895814 | Molecular biology division, Institute of Clinical Biochemistry and Diagnostics, Charles University, Faculty of Medicine in Hradec Králové and University Hospital Hradec Králové | Molecular biology division, Institute of Clinical Biochemistry and Diagnostics, Charles University, Faculty of Medicine in Hradec Králové and University Hospital Hradec Králové | Helena Kovaříková et al |
| EPI_ISL_895792 | Molecular biology division, Institute of Clinical Biochemistry and Diagnostics, Charles University, Faculty of Medicine in Hradec Králové and University Hospital Hradec Králové | Molecular biology division, Institute of Clinical Biochemistry and Diagnostics, Charles University, Faculty of Medicine in Hradec Králové and University Hospital Hradec Králové | Helena Kovaříková et al |
| EPI_ISL_895793 | Molecular biology division, Institute of Clinical Biochemistry and Diagnostics, Charles University, Faculty of Medicine in Hradec Králové and University Hospital Hradec Králové | Molecular biology division, Institute of Clinical Biochemistry and Diagnostics, Charles University, Faculty of Medicine in Hradec Králové and University Hospital Hradec Králové | Helena Kovaříková et al |
| EPI_ISL_895834 | Molecular biology division, Institute of Clinical Biochemistry and Diagnostics, Charles University, Faculty of Medicine in Hradec Králové and University Hospital Hradec Králové | Molecular biology division, Institute of Clinical Biochemistry and Diagnostics, Charles University, Faculty of Medicine in Hradec Králové and University Hospital Hradec Králové | Helena Kovaříková et al |
| EPI_ISL_895824 | Molecular biology division, Institute of Clinical Biochemistry and Diagnostics, Charles University, Faculty of Medicine in Hradec Králové and University Hospital Hradec Králové | Molecular biology division, Institute of Clinical Biochemistry and Diagnostics, Charles University, Faculty of Medicine in Hradec Králové and University Hospital Hradec Králové | Helena Kovaříková et al |
| EPI_ISL_895815 | Molecular biology division, Institute of Clinical Biochemistry and Diagnostics, Charles University, Faculty of Medicine in Hradec Králové and University Hospital Hradec Králové | Molecular biology division, Institute of Clinical Biochemistry and Diagnostics, Charles University, Faculty of Medicine in Hradec Králové and University Hospital Hradec Králové | Helena Kovaříková et al |
| EPI_ISL_895761 | Molecular biology division, Institute of Clinical Biochemistry and Diagnostics, Charles University, Faculty of Medicine in Hradec Králové and University Hospital Hradec Králové | Molecular biology division, Institute of Clinical Biochemistry and Diagnostics, Charles University, Faculty of Medicine in Hradec Králové and University Hospital Hradec Králové | Helena Kovaříková et al |
| EPI_ISL_700167 | Hematopathology Laboratory, ACTREC, TMC | Hematopathology Laboratory, ACTREC, TMC | Hematopathology Laboratory et al |
| EPI_ISL_978550 | Centre de Virologie des Maladies infectueuses Tropicales | Functional Genomic Platform UATRS-biology, CNRST | Hemlali Mouhssine et al |
| EPI_ISL_1542694 | Pandemic Response Lab - NYC | Pandemic Response Lab, R&D | Henry Lee et al |

| | | | |
|---|---|---|---|
| EPI_ISL_1543367 | Pandemic Response Lab - NYC | Pandemic Response Lab, R&D | Henry Lee et al |
| EPI_ISL_1633869 | Pandemic Response Lab - NYC | Pandemic Response Lab, R&D | Henry Lee et al |
| EPI_ISL_1633841 | Pandemic Response Lab - NYC | Pandemic Response Lab, R&D | Henry Lee et al |
| EPI_ISL_1633723 | Pandemic Response Lab - NYC | Pandemic Response Lab, R&D | Henry Lee et al |
| EPI_ISL_483035 | Medical Ain Shams Research Institute (MASRI), Ain Shams University | Medical Ain Shams Research Institute (MASRI), Ain Shams University | Hesham Elghazaly et al |
| EPI_ISL_482765 | Medical Ain Shams Research Institute (MASRI), Ain Shams University | Medical Ain Shams Research Institute (MASRI), Ain Shams University | Hesham Elghazaly et al |
| EPI_ISL_483036 | Medical Ain Shams Research Institute (MASRI), Ain Shams University | Medical Ain Shams Research Institute (MASRI), Ain Shams University | Hesham Elghazaly et al |
| EPI_ISL_1109485 | Medical Ain Shams Research Institute (MASRI), Ain Shams University | Medical Ain Shams Research Institute (MASRI), Ain Shams University | Hesham Elghazaly et al |
| EPI_ISL_471456 | Centre de Virologie des Maladies Tropicales | Functional Genomic Platform/Service Analyses Biologique/UATRS/ Centre National Pour la Recherche Scientifique Et Technique (CNRST) | Hicham EL ANNAZ et al |
| EPI_ISL_471458 | Centre de Virologie des Maladies Tropicales | Functional Genomic Platform/Service Analyses Biologique/UATRS/ Centre National Pour la Recherche Scientifique Et Technique (CNRST) | Hicham EL ANNAZ et al |
| EPI_ISL_471460 | Centre de Virologie des Maladies Tropicales | Functional Genomic Platform/Service Analyses Biologique/UATRS/ Centre National Pour la Recherche Scientifique Et Technique (CNRST) | Hicham EL ANNAZ et al |
| EPI_ISL_1591658 | Instituto Nacional de Medicina Genomica | Instituto Nacional de Medicina Genomica | Hidalgo-Miranda A et al |
| EPI_ISL_1585919 | Instituto Nacional de Medicina Genomica | Instituto Nacional de Medicina Genomica | Hidalgo-Miranda A et al |
| EPI_ISL_1585916 | Instituto Nacional de Medicina Genomica | Instituto Nacional de Medicina Genomica | Hidalgo-Miranda A et al |
| EPI_ISL_1585918 | Instituto Nacional de Medicina Genomica | Instituto Nacional de Medicina Genomica | Hidalgo-Miranda A et al |
| EPI_ISL_933525 | Instituto Nacional de Medicina Genomica | Instituto Nacional de Medicina Genomica | Hidalgo-Miranda A et al |
| EPI_ISL_955192 | Instituto Nacional de Medicina Genomica | Instituto Nacional de Medicina Genomica | Hidalgo-Miranda A et al |
| EPI_ISL_955207 | Instituto Nacional de Medicina Genomica | Instituto Nacional de Medicina Genomica | Hidalgo-Miranda A et al |
| EPI_ISL_1005563 | Instituto Nacional de Medicina Genomica | Instituto Nacional de Medicina Genomica | Hidalgo-Miranda A et al |
| EPI_ISL_522942 | Instituto Nacional de Medicina Genómica | Instituto Nacional de Medicina Genómica | Hidalgo-Miranda A et al |
| EPI_ISL_1628603 | InDRE | Instituto Nacional de Medicina Genomica | Hidalgo-Miranda A et al |
| EPI_ISL_574623 | BTKLPP Kelas I Manado | Eijkman Institute for Molecular Biology, Ministry of Research and Technology/National Agency for Research and Innovation | Hidayat Trimarsanto et al |
| EPI_ISL_418275 | University Hospital Basel, Clinical Virology | University Hospital Basel, Clinical Bacteriology | Hirsch et al |
| EPI_ISL_418273 | University Hospital Basel, Clinical Virology | University Hospital Basel, Labormedizin | Hirsch et al |
| EPI_ISL_1390768 | Ohio Department of Health Laboratory | Ohio Department of Health Laboratory | Holmes et al |
| EPI_ISL_824389 | Ohio Department of Health Laboratory | Ohio Department of Health Laboratory | Holmes et al |
| EPI_ISL_909772 | National Institute of Health Research and Development | National Institute of Health Research and Development | Holy Arif Wibowo et al |

| | | | |
|---|---|---|---|
| EPI_ISL_1096141 | Departamento de Cuidados Intensivos-Unidad de Investigación Médica en Inmunoquímica, UMAE Hospital de Especialidades “Bernardo Sepúlveda Gutiérrez”, Centro Médico Nacional Siglo XXI, Instituto Mexicano del Seguro Social (IMSS), México. | Laboratorio de Secuenciación-Centro de Instrumentos, IMSS México /Unidad de Investigación Médica en Inmunoquímica, UMAE Hospital de Especialidades “Bernardo Sepúlveda Gutiérrez”, Centro Médico Nacional Siglo XXI, Instituto Mexicano del Seguro Social (IMSS), México. | Horacio Zamudio-Meza et al |
| EPI_ISL_591270 | National Institute for Viral Disease Control and Prevention, China CDC | National Institute for Viral Disease Control and Prevention, China CDC | Huilai Ma et al |
| EPI_ISL_591271 | National Institute for Viral Disease Control and Prevention, China CDC | National Institute for Viral Disease Control and Prevention, China CDC | Huilai Ma et al |
| EPI_ISL_539496 | Hospital Nostra Senyora de Meritxell | Instituto de Salud Carlos III | Iglesias-Caballero et al |
| EPI_ISL_455314 | Hospital Virgen del Rocio | Instituto de Salud Carlos III | Iglesias-Caballero et al |
| EPI_ISL_802730 | Hospital Clínic de Barcelona | Instituto de Salud Carlos III | Iglesias-Caballero et al |
| EPI_ISL_539558 | Hospital San Pedro de Alcántara | Instituto de Salud Carlos III | Iglesias-Caballero et al |
| EPI_ISL_804972 | Hospital Comarcal de Melilla | Instituto de Salud Carlos III | Iglesias-Caballero et al |
| EPI_ISL_455336 | Hospital San Pedro | Instituto de Salud Carlos III | Iglesias-Caballero et al |
| EPI_ISL_794738 | 1-Laboratory of Microbiology, National Reference Lab, Charles Nicolle Hospital; 2-University of Tunis ElManar, Faculty of Medicine of Tunis, LR99ES09, Tunis, Tunisia | 1-Clinical and Experimental Pharmacology Lab, LR16SP02, National Center of Pharmacovigilance, University of Tunis El Manar, Tunis, Tunisia. 2-Neurodegenerative diseases and psychiatric troubles, LR18SP03, Razi Hospital, University of Tunis El Manar, Tunis, Tunisia. 3- Ministry of Health, National Observatory of New and Emerging Diseases, 1006, Tunis, Tunisia | Ilhem Boutiba-Ben Boubaker et al |
| EPI_ISL_794736 | 1-Laboratory of Microbiology, National Reference Lab, Charles Nicolle Hospital; 2-University of Tunis ElManar, Faculty of Medicine of Tunis, LR99ES09, Tunis, Tunisia | 1-Clinical and Experimental Pharmacology Lab, LR16SP02, National Center of Pharmacovigilance, University of Tunis El Manar, Tunis, Tunisia. 2-Neurodegenerative diseases and psychiatric troubles, LR18SP03, Razi Hospital, University of Tunis El Manar, Tunis, Tunisia. 3- Ministry of Health, National Observatory of New and Emerging Diseases, 1006, Tunis, Tunisia | Ilhem Boutiba-Ben Boubaker et al |
| EPI_ISL_733500 | 1-Laboratory of Microbiology, National Reference Lab, Charles Nicolle Hospital; 2-University of Tunis ElManar, Faculty of Medicine of Tunis, LR99ES09, Tunis, Tunisia | 1-Clinical and Experimental Pharmacology Lab, LR16SP02, National Center of Pharmacovigilance, University of Tunis El Manar, Tunis, Tunisia. 2-Neurodegenerative diseases and psychiatric troubles, LR18SP03, Razi Hospital, University of Tunis El Manar, Tunis, Tunisia. 3- Ministry of Health, National Observatory of New and Emerging Diseases, 1006, Tunis, Tunisia | Ilhem Boutiba-Ben Boubaker et al |
| EPI_ISL_654020 | Laboratory of Microbiology, National Reference Lab, Charles Nicolle Hospital; 2-University of Tunis ElManar, Faculty of Medicine of Tunis, LR99ES09, Tunis, Tunisia | 1-Clinical and Experimental Pharmacology Lab, LR16SP02, National Center of Pharmacovigilance, University of Tunis El Manar, Tunis, Tunisia. 2-Neurodegenerative diseases and psychiatric troubles, LR18SP03, Razi Hospital, University of Tunis El Manar, Tunis, Tunisia. 3- Ministry of Health, National Observatory of New and Emerging Diseases, 1006, Tunis, Tunisia | Ilhem Boutiba-Ben Boubaker et al |
| EPI_ISL_632310 | 1-Laboratory of Microbiology, National Reference Lab, Charles Nicolle Hospital; 2-University of Tunis ElManar, Faculty of Medicine of Tunis, LR99ES09, Tunis, Tunisia | 1-Clinical and Experimental Pharmacology Lab, LR16SP02, National Center of Pharmacovigilance, University of Tunis El Manar, Tunis, Tunisia. 2-Neurodegenerative diseases and psychiatric troubles, LR18SP03, Razi Hospital, University of Tunis El Manar, Tunis, Tunisia. 3- Ministry of Health, National Observatory of New and Emerging Diseases, 1006, Tunis, Tunisia | Ilhem Boutiba-Ben Boubaker et al |
| EPI_ISL_634977 | 1-Laboratory of Microbiology, National Reference Lab, Charles Nicolle Hospital; 2-University of Tunis ElManar, Faculty of Medicine of Tunis, LR99ES09, Tunis, Tunisia | 1-Clinical and Experimental Pharmacology Lab, LR16SP02, National Center of Pharmacovigilance, University of Tunis El Manar, Tunis, Tunisia. 2-Neurodegenerative diseases and psychiatric troubles, LR18SP03, Razi Hospital, University of Tunis El Manar, Tunis, Tunisia. 3- Ministry of Health, National Observatory of New and Emerging Diseases, 1006, Tunis, Tunisia | Ilhem Boutiba-Ben Boubaker et al |

| | | | |
|---|---|---|---|
| EPI_ISL_635062 | 1-Laboratory of Microbiology, National Reference Lab, Charles Nicolle Hospital; 2-University of Tunis ElManar, Faculty of Medicine of Tunis, LR99ES09, Tunis, Tunisia | 1-Clinical and Experimental Pharmacology Lab, LR16SP02, National Center of Pharmacovigilance, University of Tunis El Manar, Tunis, Tunisia. 2-Neurodegenerative diseases and psychiatric troubles, LR18SP03, Razi Hospital, University of Tunis El Manar, Tunis, Tunisia. 3- Ministry of Health, National Observatory of New and Emerging Diseases, 1006, Tunis, Tunisia | Ilhem Boutiba-Ben Boubaker et al |
| EPI_ISL_707700 | 1-Laboratory of Microbiology, National Reference Lab, Charles Nicolle Hospital; 2-University of Tunis ElManar, Faculty of Medicine of Tunis, LR99ES09, Tunis, Tunisia | 1-Clinical and Experimental Pharmacology Lab, LR16SP02, National Center of Pharmacovigilance, University of Tunis El Manar, Tunis, Tunisia. 2-Neurodegenerative diseases and psychiatric troubles, LR18SP03, Razi Hospital, University of Tunis El Manar, Tunis, Tunisia. 3- Ministry of Health, National Observatory of New and Emerging Diseases, 1006, Tunis, Tunisia | Ilhem Boutiba-Ben Boubaker et al |
| EPI_ISL_733499 | 1-Laboratory of Microbiology, National Reference Lab, Charles Nicolle Hospital; 2-University of Tunis ElManar, Faculty of Medicine of Tunis, LR99ES09, Tunis, Tunisia | 1-Clinical and Experimental Pharmacology Lab, LR16SP02, National Center of Pharmacovigilance, University of Tunis El Manar, Tunis, Tunisia. 2-Neurodegenerative diseases and psychiatric troubles, LR18SP03, Razi Hospital, University of Tunis El Manar, Tunis, Tunisia. 3- Ministry of Health, National Observatory of New and Emerging Diseases, 1006, Tunis, Tunisia | Ilhem Boutiba-Ben Boubaker et al |
| EPI_ISL_707792 | 1-Laboratory of Microbiology, National Reference Lab, Charles Nicolle Hospital; 2-University of Tunis ElManar, Faculty of Medicine of Tunis, LR99ES09, Tunis, Tunisia | 1-Clinical and Experimental Pharmacology Lab, LR16SP02, National Center of Pharmacovigilance, University of Tunis El Manar, Tunis, Tunisia. 2-Neurodegenerative diseases and psychiatric troubles, LR18SP03, Razi Hospital, University of Tunis El Manar, Tunis, Tunisia. 3- Ministry of Health, National Observatory of New and Emerging Diseases, 1006, Tunis, Tunisia | Ilhem Boutiba-Ben Boubaker et al |
| EPI_ISL_763065 | 1-Laboratory of Microbiology, National Reference Lab, Charles Nicolle Hospital; 2-University of Tunis ElManar, Faculty of Medicine of Tunis, LR99ES09, Tunis, Tunisia | 1-Clinical and Experimental Pharmacology Lab, LR16SP02, National Center of Pharmacovigilance, University of Tunis El Manar, Tunis, Tunisia. 2-Neurodegenerative diseases and psychiatric troubles, LR18SP03, Razi Hospital, University of Tunis El Manar, Tunis, Tunisia. 3- Ministry of Health, National Observatory of New and Emerging Diseases, 1006, Tunis, Tunisia | Ilhem Boutiba-Ben Boubaker et al |
| EPI_ISL_796782 | Laboratory of Microbiology, National Reference Lab, Charles Nicolle Hospital; 2-University of Tunis ElManar, Faculty of Medicine of Tunis, LR99ES09, Tunis, Tunisia | Clinical and Experimental Pharmacology Lab, LR16SP02, National Center of Pharmacovigilance, University of Tunis El Manar, Tunis, Tunisia. 2-Neurodegenerative diseases and psychiatric troubles, LR18SP03, Razi Hospital, University of Tunis El Manar, Tunis, Tunisia. 3- Ministry of Health, National Observatory of New and Emerging Diseases, 1006, Tunis, Tunisia | Ilhem Boutiba-Ben Boubaker et al |
| EPI_ISL_763067 | 1-Laboratory of Microbiology, National Reference Lab, Charles Nicolle Hospital; 2-University of Tunis ElManar, Faculty of Medicine of Tunis, LR99ES09, Tunis, Tunisia | 1-Clinical and Experimental Pharmacology Lab, LR16SP02, National Center of Pharmacovigilance, University of Tunis El Manar, Tunis, Tunisia. 2-Neurodegenerative diseases and psychiatric troubles, LR18SP03, Razi Hospital, University of Tunis El Manar, Tunis, Tunisia. 3- Ministry of Health, National Observatory of New and Emerging Diseases, 1006, Tunis, Tunisia | Ilhem Boutiba-Ben Boubaker et al |
| EPI_ISL_480246 | Genomic Laboratory (GLAB) (Conjoint lab of Health Directorate of Istanbul and Istanbul Technical University) | Genomic Laboratory (GLAB), Istanbul Technical University | Ilker Karacan et al |
| EPI_ISL_735272 | Genomic Laboratory (GLAB) (Conjoint lab of Health Directorate of Istanbul and Istanbul Technical University) | Genomic Laboratory (GLAB), Istanbul Technical University | Ilker Karacan et al |
| EPI_ISL_735275 | Genomic Laboratory (GLAB) (Conjoint lab of Health Directorate of Istanbul and Istanbul Technical University) | Genomic Laboratory (GLAB), Istanbul Technical University | Ilker Karacan et al |
| EPI_ISL_735277 | Genomic Laboratory (GLAB) (Conjoint lab of Health Directorate of Istanbul and Istanbul Technical University) | Genomic Laboratory (GLAB), Istanbul Technical University | Ilker Karacan et al |
| EPI_ISL_480256 | Genomic Laboratory (GLAB) (Conjoint lab of Health Directorate of Istanbul and Istanbul Technical University) | Genomic Laboratory (GLAB), Istanbul Technical University | Ilker Karacan et al |
| EPI_ISL_480271 | Genomic Laboratory (GLAB) (Conjoint lab of Health Directorate of Istanbul and Istanbul Technical University) | Genomic Laboratory (GLAB), Istanbul Technical University | Ilker Karacan et al |

| | | | |
|---|---|---|---|
| EPI_ISL_480276 | Genomic Laboratory (GLAB) (Conjoint lab of Health Directorate of Istanbul and Istanbul Technical University) | Genomic Laboratory (GLAB), Istanbul Technical University | Ilker Karacan et al |
| EPI_ISL_735328 | Genomic Laboratory (GLAB) (Conjoint lab of Health Directorate of Istanbul and Istanbul Technical University) | Genomic Laboratory (GLAB), Istanbul Technical University | Ilker Karacan et al |
| EPI_ISL_735344 | Genomic Laboratory (GLAB) (Conjoint lab of Health Directorate of Istanbul and Istanbul Technical University) | Genomic Laboratory (GLAB), Istanbul Technical University | Ilker Karacan et al |
| EPI_ISL_735352 | Genomic Laboratory (GLAB) (Conjoint lab of Health Directorate of Istanbul and Istanbul Technical University) | Genomic Laboratory (GLAB), Istanbul Technical University | Ilker Karacan et al |
| EPI_ISL_735360 | Genomic Laboratory (GLAB) (Conjoint lab of Health Directorate of Istanbul and Istanbul Technical University) | Genomic Laboratory (GLAB), Istanbul Technical University | Ilker Karacan et al |
| EPI_ISL_955154 | Genomic Laboratory (GLAB) (Conjoint lab of Health Directorate of Istanbul and Istanbul Technical University) | Genomic Laboratory (GLAB), Istanbul Technical University | Ilker Karacan et al |
| EPI_ISL_419211 | Central Virology Laboratory | Israel Institute for Biological Research | Inbar Cohen-Gihon et al |
| EPI_ISL_933929 | Vilnius university hospital Santaros Klinikos, Center of Laboratory Medicine | Vilnius university hospital Santaros Klinikos, Center of Laboratory Medicine | Ingrida Olendraite et al |
| EPI_ISL_933939 | Vilnius university hospital Santaros Klinikos, Center of Laboratory Medicine | Vilnius university hospital Santaros Klinikos, Center of Laboratory Medicine | Ingrida Olendraite et al |
| EPI_ISL_933949 | Vilnius university hospital Santaros Klinikos, Center of Laboratory Medicine | Vilnius university hospital Santaros Klinikos, Center of Laboratory Medicine | Ingrida Olendraite et al |
| EPI_ISL_933959 | Vilnius university hospital Santaros Klinikos, Center of Laboratory Medicine | Vilnius university hospital Santaros Klinikos, Center of Laboratory Medicine | Ingrida Olendraite et al |
| EPI_ISL_934211 | Vilnius university hospital Santaros Klinikos, Center of Laboratory Medicine | Vilnius university hospital Santaros Klinikos, Center of Laboratory Medicine | Ingrida Olendraite et al |
| EPI_ISL_934272 | Vilnius university hospital Santaros Klinikos, Center of Laboratory Medicine | Vilnius university hospital Santaros Klinikos, Center of Laboratory Medicine | Ingrida Olendraite et al |
| EPI_ISL_914881 | Vilnius university hospital Santaros Klinikos, Center of Laboratory Medicine | Vilnius University Hospital Santaros Klinikos | Ingrida Olendraite et al |
| EPI_ISL_914882 | Vilnius university hospital Santaros Klinikos, Center of Laboratory Medicine | Vilnius University Hospital Santaros Klinikos | Ingrida Olendraite et al |
| EPI_ISL_914884 | Vilnius university hospital Santaros Klinikos, Center of Laboratory Medicine | Vilnius University Hospital Santaros Klinikos | Ingrida Olendraite et al |
| EPI_ISL_914886 | Vilnius university hospital Santaros Klinikos, Center of Laboratory Medicine | Vilnius University Hospital Santaros Klinikos | Ingrida Olendraite et al |
| EPI_ISL_1591041 | Servicio de Microbiología. HRU de Málaga. Servicio Andaluz de Salud | SeqCOVID-SPAIN consortium/IBV(CSIC) | Inmaculada de Toro Peinado et al |
| EPI_ISL_1591046 | Servicio de Microbiología. HRU de Málaga. Servicio Andaluz de Salud | SeqCOVID-SPAIN consortium/IBV(CSIC) | Inmaculada de Toro Peinado et al |
| EPI_ISL_1591050 | Servicio de Microbiología. HRU de Málaga. Servicio Andaluz de Salud | SeqCOVID-SPAIN consortium/IBV(CSIC) | Inmaculada de Toro Peinado et al |
| EPI_ISL_1591051 | Servicio de Microbiología. HRU de Málaga. Servicio Andaluz de Salud | SeqCOVID-SPAIN consortium/IBV(CSIC) | Inmaculada de Toro Peinado et al |
| EPI_ISL_510063 | Servicio de Microbiología. HRU de Málaga. Servicio Andaluz de Salud | SeqCOVID-SPAIN consortium/IBV(CSIC) | Inmaculada de Toro Peinado. MªConcepció |
| EPI_ISL_1532286 | Oman-National Influenza Center | Biotechnology & OMICs Laboratory | Intisar Al-Shukri et al |
| EPI_ISL_1532287 | Oman-National Influenza Center | Biotechnology & OMICs Laboratory | Intisar Al-Shukri et al |
| EPI_ISL_1532288 | Oman-National Influenza Center | Biotechnology & OMICs Laboratory | Intisar Al-Shukri et al |
| EPI_ISL_1532289 | Oman-National Influenza Center | Biotechnology & OMICs Laboratory | Intisar Al-Shukri et al |
| EPI_ISL_1499505 | Institute of Public Health of Međimurje county | Croatian Institute of Public Health | Irena Tabain et al |
| EPI_ISL_1499515 | Institute of Public Health of Karlovac County | Croatian Institute of Public Health | Irena Tabain et al |

n Mediavilla Gradolph. Begoña Palop Borrás

| | | | |
|---|---|---|---|
| EPI_ISL_1499519 | Institute of Public Health of Dubrovnik-Neretva County | Croatian Institute of Public Health | Irena Tabain et al |
| EPI_ISL_1578483 | Uversity hospital Dubrava | Croatian Institute of Public Health | Irena Tabain et al |
| EPI_ISL_1578501 | University Hospital Centre Zagreb | Croatian Institute of Public Health | Irena Tabain et al |
| EPI_ISL_1591279 | Institute for Public Health of Šibenik-Knin County | Croatian Institute of Public Health | Irena Tabain et al |
| EPI_ISL_1591311 | Teaching Institute for Public Health | Croatian Institute of Public Health | Irena Tabain et al |
| EPI_ISL_454602 | Croatian Institute of Public Health | University of Zagreb, Centre for research and knowledge transfer in biotechnology | Irena Tabain et al |
| EPI_ISL_1272020 | University Hospital Centre Zagreb | Croatian Institute of Public Health | Irena Tabain et al |
| EPI_ISL_1287400 | Croatian Institute of Public Health | Croatian Institute of Public Health | Irena Tabain et al |
| EPI_ISL_1435902 | Institute of Public Health of Varazdin county | Croatian Institute of Public Health | Irena Tabain et al |
| EPI_ISL_1195672 | Croatian Institute of Public Health | Croatian Institute of Public Health | Irena Tabain et al |
| EPI_ISL_1195728 | Public Health Institute of Šibenik-Knin County | Croatian Institute of Public Health | Irena Tabain et al |
| EPI_ISL_1091196 | Croatian Institute of Public Health | Croatian Institute of Public Health | Irena Tabain et al |
| EPI_ISL_1300659 | Clinical Center, University of Sarajevo; Unit for Clinical Microbiology | Clinical Center, University of Sarajevo; Unit for Clinical Microbiology | Irma Salimović-Bešić et al |
| EPI_ISL_1300645 | Clinical Center, University of Sarajevo; Unit for Clinical Microbiology | Clinical Center, University of Sarajevo; Unit for Clinical Microbiology | Irma Salimović-Bešić et al |
| EPI_ISL_1300658 | Clinical Center, University of Sarajevo; Unit for Clinical Microbiology | Clinical Center, University of Sarajevo; Unit for Clinical Microbiology | Irma Salimović-Bešić et al |
| EPI_ISL_1300651 | Clinical Center, University of Sarajevo; Unit for Clinical Microbiology | Clinical Center, University of Sarajevo; Unit for Clinical Microbiology | Irma Salimović-Bešić et al |
| EPI_ISL_1020189 | RS Trimitra, Bogor, West java | Biosafety Level-3 Laboratory, Indonesian Institute of Sciences (LIPI) | Isa Nuryana et al |
| EPI_ISL_1395798 | Laboratorio del Hospital Interzonal General de Agudos “Evita” | Área de Secuenciación del Laboratorio de Virología del Hospital de Niños Dr. Ricardo Gutierrez on behalf of 'Proyecto Argentino Interinstitucional de genomica de SARS-CoV-2' (PAIS Consortium) | Isabel Desimone et al |
| EPI_ISL_1395800 | Laboratorio del Hospital Interzonal General de Agudos “Evita” | Área de Secuenciación del Laboratorio de Virología del Hospital de Niños Dr. Ricardo Gutierrez on behalf of 'Proyecto Argentino Interinstitucional de genomica de SARS-CoV-2' (PAIS Consortium) | Isabel Desimone et al |
| EPI_ISL_1443196 | Hospital Aliança | Hospital São Rafael - IDOR | Isadora Cristina de Siqueira et al |
| EPI_ISL_1443197 | Hospital Aliança | Hospital São Rafael - IDOR | Isadora Cristina de Siqueira et al |
| EPI_ISL_1443198 | Hospital Aliança | Hospital São Rafael - IDOR | Isadora Cristina de Siqueira et al |
| EPI_ISL_636979 | HP Pemba | KRISP, KZN Research Innovation and Sequencing Platform | Ismael N et al |
| EPI_ISL_636975 | HP Pemba | KRISP, KZN Research Innovation and Sequencing Platform | Ismael N et al |
| EPI_ISL_636981 | City of Chimoio | KRISP, KZN Research Innovation and Sequencing Platform | Ismael N et al |
| EPI_ISL_636977 | HP Pemba | KRISP, KZN Research Innovation and Sequencing Platform | Ismael N et al |
| EPI_ISL_636976 | HP Pemba | KRISP, KZN Research Innovation and Sequencing Platform | Ismael N et al |
| EPI_ISL_635778 | Biolab Diagnostic Laboratories | Andersen lab at Scripps Research | Issa Abu-Dayyeh et al |

| | | | |
|---|---|---|---|
| EPI_ISL_635782 | Biolab Diagnostic Laboratories | Andersen lab at Scripps Research | Issa Abu-Dayyeh et al |
| EPI_ISL_730240 | Biolab Diagnostic Laboratories | Andersen lab at Scripps Research | Issa Abu-Dayyeh et al |
| EPI_ISL_730266 | Biolab Diagnostic Laboratories | Andersen lab at Scripps Research | Issa Abu-Dayyeh et al |
| EPI_ISL_730378 | Biolab Diagnostic Laboratories | Andersen lab at Scripps Research | Issa Abu-Dayyeh et al |
| EPI_ISL_730385 | Biolab Diagnostic Laboratories | Andersen lab at Scripps Research | Issa Abu-Dayyeh et al |
| EPI_ISL_730398 | Biolab Diagnostic Laboratories | Andersen lab at Scripps Research | Issa Abu-Dayyeh et al |
| EPI_ISL_730520 | Biolab Diagnostic Laboratories | Andersen lab at Scripps Research | Issa Abu-Dayyeh et al |
| EPI_ISL_730521 | Biolab Diagnostic Laboratories | Andersen lab at Scripps Research | Issa Abu-Dayyeh et al |
| EPI_ISL_730524 | Biolab Diagnostic Laboratories | Andersen lab at Scripps Research | Issa Abu-Dayyeh et al |
| EPI_ISL_730544 | Biolab Diagnostic Laboratories | Andersen lab at Scripps Research | Issa Abu-Dayyeh et al |
| EPI_ISL_730560 | Biolab Diagnostic Laboratories | Andersen lab at Scripps Research | Issa Abu-Dayyeh et al |
| EPI_ISL_878498 | Biolab Diagnostic Laboratories | Andersen lab at Scripps Research | Issa Abu-Dayyeh et al |
| EPI_ISL_878571 | Biolab Diagnostic Laboratories | Andersen lab at Scripps Research | Issa Abu-Dayyeh et al |
| EPI_ISL_878574 | Biolab Diagnostic Laboratories | Andersen lab at Scripps Research | Issa Abu-Dayyeh et al |
| EPI_ISL_1406151 | Biolab Diagnostic Laboratories | Biolab Diagnostic Laboratories | Issa Abu-Dayyeh et al |
| EPI_ISL_1406179 | Biolab Diagnostic Laboratories | Biolab Diagnostic Laboratories | Issa Abu-Dayyeh et al |
| EPI_ISL_1406183 | Biolab Diagnostic Laboratories | Biolab Diagnostic Laboratories | Issa Abu-Dayyeh et al |
| EPI_ISL_1406189 | Biolab Diagnostic Laboratories | Biolab Diagnostic Laboratories | Issa Abu-Dayyeh et al |
| EPI_ISL_1406195 | Biolab Diagnostic Laboratories | Biolab Diagnostic Laboratories | Issa Abu-Dayyeh et al |
| EPI_ISL_1406239 | Biolab Diagnostic Laboratories | Biolab Diagnostic Laboratories | Issa Abu-Dayyeh et al |
| EPI_ISL_977602 | Biolab Diagnostic Laboratories | Biolab Diagnostic Laboratories | Issa Abu-Dayyeh et al |
| EPI_ISL_635777 | Biolab Diagnostic Laboratories | Andersen lab at Scripps Research | Issa Abu-Dayyeh et al |
| EPI_ISL_755236 | Biolab Diagnostic Laboratories | Andersen lab at Scripps Research | Issa Abu-Dayyeh et al |
| EPI_ISL_755237 | Biolab Diagnostic Laboratories | Andersen lab at Scripps Research | Issa Abu-Dayyeh et al |
| EPI_ISL_755238 | Biolab Diagnostic Laboratories | Andersen lab at Scripps Research | Issa Abu-Dayyeh et al |
| EPI_ISL_755128 | Biolab Diagnostic Laboratories | Andersen lab at Scripps Research | Issa Abu-Dayyeh et al |
| EPI_ISL_755124 | Biolab Diagnostic Laboratories | Andersen lab at Scripps Research | Issa Abu-Dayyeh et al |
| EPI_ISL_755253 | Biolab Diagnostic Laboratories | Andersen lab at Scripps Research | Issa Abu-Dayyeh et al |
| EPI_ISL_755257 | Biolab Diagnostic Laboratories | Andersen lab at Scripps Research | Issa Abu-Dayyeh et al |
| EPI_ISL_755145 | Biolab Diagnostic Laboratories | Andersen lab at Scripps Research | Issa Abu-Dayyeh et al |

| | | | |
|---|---|---|---|
| EPI_ISL_755263 | Biolab Diagnostic Laboratories | Andersen lab at Scripps Research | Issa Abu-Dayyeh et al |
| EPI_ISL_450188 | Biolab Diagnostic Laboratories | Andersen lab at Scripps Research | Issa Abu-Dayyeh et al |
| EPI_ISL_450189 | Biolab Diagnostic Laboratories | Andersen lab at Scripps Research | Issa Abu-Dayyeh et al |
| EPI_ISL_429995 | Biolab Diagnostic Laboratories | Andersen lab at Scripps Research | Issa Abu-Dayyeh et al |
| EPI_ISL_429998 | Biolab Diagnostic Laboratories | Andersen lab at Scripps Research | Issa Abu-Dayyeh et al |
| EPI_ISL_430000 | Biolab Diagnostic Laboratories | Andersen lab at Scripps Research | Issa Abu-Dayyeh et al |
| EPI_ISL_430008 | Biolab Diagnostic Laboratories | Andersen lab at Scripps Research | Issa Abu-Dayyeh et al |
| EPI_ISL_430011 | Biolab Diagnostic Laboratories | Andersen lab at Scripps Research | Issa Abu-Dayyeh et al |
| EPI_ISL_430015 | Biolab Diagnostic Laboratories | Andersen lab at Scripps Research | Issa Abu-Dayyeh et al |
| EPI_ISL_480297 | National Reference Laboratory Influenza and acute respiratory diseases | NRL-HIV | Ivan Ivanov et al |
| EPI_ISL_480298 | National Reference Laboratory Influenza and acute respiratory diseases | NRL-HIV | Ivan Ivanov et al |
| EPI_ISL_480300 | National Reference Laboratory Influenza and acute respiratory diseases | NRL-HIV | Ivan Ivanov et al |
| EPI_ISL_480302 | National Reference Laboratory Influenza and acute respiratory diseases | NRL-HIV | Ivan Ivanov et al |
| EPI_ISL_480303 | National Reference Laboratory Influenza and acute respiratory diseases | NRL-HIV | Ivan Ivanov et al |
| EPI_ISL_480305 | National Reference Laboratory Influenza and acute respiratory diseases | NRL-HIV | Ivan Ivanov et al |
| EPI_ISL_480307 | National Reference Laboratory Influenza and acute respiratory diseases | NRL-HIV | Ivan Ivanov et al |
| EPI_ISL_480308 | National Reference Laboratory Influenza and acute respiratory diseases | NRL-HIV | Ivan Ivanov et al |
| EPI_ISL_480310 | National Reference Laboratory Influenza and acute respiratory diseases | NRL-HIV | Ivan Ivanov et al |
| EPI_ISL_454581 | University Hospital for Infectious Diseases Dr. Fran Mihaljević, Research Unit | University of Zagreb, Centre for research and knowledge transfer in biotechnology | Ivan-Christian Kurolt et al |
| EPI_ISL_454588 | University Hospital for Infectious Diseases Dr. Fran Mihaljević, Research Unit | University of Zagreb, Centre for research and knowledge transfer in biotechnology | Ivan-Christian Kurolt et al |
| EPI_ISL_454592 | University Hospital for Infectious Diseases Dr. Fran Mihaljević, Research Unit | University of Zagreb, Centre for research and knowledge transfer in biotechnology | Ivan-Christian Kurolt et al |
| EPI_ISL_450518 | E. Gulbja Laboratorija | Latvian Biomedical Research and Study Centre | Ivars Silamiķelis et al |
| EPI_ISL_450519 | E. Gulbja Laboratorija | Latvian Biomedical Research and Study Centre | Ivars Silamiķelis et al |
| EPI_ISL_421654 | E. Gulbja Laboratorija | Latvian Biomedical Research and Study Centre | Ivars Silamiķelis et al |
| EPI_ISL_450522 | Centrālā Laboratorija | Latvian Biomedical Research and Study Centre | Ivars Silamiķelis et al |
| EPI_ISL_450523 | Centrālā Laboratorija | Latvian Biomedical Research and Study Centre | Ivars Silamiķelis et al |
| EPI_ISL_450524 | Centrālā Laboratorija | Latvian Biomedical Research and Study Centre | Ivars Silamiķelis et al |
| EPI_ISL_486390 | Centrālā laboratorija | Latvian Biomedical Research and Study Centre | Ivars Silamiķelis et al |
| EPI_ISL_486410 | Centrala laboratorija | Latvian Biomedical Research and Study Centre | Ivars Silamiķelis et al |

| | | | |
|---|---|---|---|
| EPI_ISL_486412 | Centrālā laboratorija | Latvian Biomedical Research and Study Centre | Ivars Silamiķelis et al |
| EPI_ISL_421655 | E. Gulbja Laboratorija | Latvian Biomedical Research and Study Centre | Ivars Silamiķelis et al |
| EPI_ISL_486414 | Centrālā laboratorija | Latvian Biomedical Research and Study Centre | Ivars Silamiķelis et al |
| EPI_ISL_486415 | Centrālā laboratorija | Latvian Biomedical Research and Study Centre | Ivars Silamiķelis et al |
| EPI_ISL_486416 | Centrālā laboratorija | Latvian Biomedical Research and Study Centre | Ivars Silamiķelis et al |
| EPI_ISL_421656 | E. Gulbja Laboratorija | Latvian Biomedical Research and Study Centre | Ivars Silamiķelis et al |
| EPI_ISL_486423 | Latvijas Infektoloģijas centrs | Latvian Biomedical Research and Study Centre | Ivars Silamiķelis et al |
| EPI_ISL_492988 | Centrālā laboratorija | Latvian Biomedical Research and Study Centre | Ivars Silamiķelis et al |
| EPI_ISL_486426 | Latvijas Infektoloģijas centrs | Latvian Biomedical Research and Study Centre | Ivars Silamiķelis et al |
| EPI_ISL_486430 | Latvijas Infektoloģijas centrs | Latvian Biomedical Research and Study Centre | Ivars Silamiķelis et al |
| EPI_ISL_426285 | E. Gulbja Laboratorija | Latvian Biomedical Research and Study Centre | Ivars Silamiķelis et al |
| EPI_ISL_501286 | Centrālā laboratorija | Latvian Biomedical Research and Study Centre | Ivars Silamiķelis et al |
| EPI_ISL_501829 | E. Gulbja Laboratorija | Latvian Biomedical Research and Study Centre | Ivars Silamiķelis et al |
| EPI_ISL_501849 | E. Gulbja Laboratorija | Latvian Biomedical Research and Study Centre | Ivars Silamiķelis et al |
| EPI_ISL_501915 | Centrālā laboratorija | Latvian Biomedical Research and Study Centre | Ivars Silamiķelis et al |
| EPI_ISL_501922 | Centrālā laboratorija | Latvian Biomedical Research and Study Centre | Ivars Silamiķelis et al |
| EPI_ISL_512313 | E. Gulbja Laboratorija | Latvian Biomedical Research and Study Centre | Ivars Silamiķelis et al |
| EPI_ISL_426289 | E. Gulbja Laboratorija | Latvian Biomedical Research and Study Centre | Ivars Silamiķelis et al |
| EPI_ISL_512647 | Latvijas Infektoloģijas centrs | Latvian Biomedical Research and Study Centre | Ivars Silamiķelis et al |
| EPI_ISL_512649 | Latvijas Infektoloģijas centrs | Latvian Biomedical Research and Study Centre | Ivars Silamiķelis et al |
| EPI_ISL_515185 | Latvijas Infektoloģijas centrs | Latvian Biomedical Research and Study Centre | Ivars Silamiķelis et al |
| EPI_ISL_515186 | Latvijas Infektoloģijas centrs | Latvian Biomedical Research and Study Centre | Ivars Silamiķelis et al |
| EPI_ISL_639637 | E. Gulbja Laboratorija | Latvian Biomedical Research and Study Centre | Ivars Silamiķelis et al |
| EPI_ISL_639658 | E. Gulbja Laboratorija | Latvian Biomedical Research and Study Centre | Ivars Silamiķelis et al |
| EPI_ISL_856707 | MFD Veselības centrs "Dziedniecība" | Latvian Biomedical Research and Study Centre | Ivars Silamiķelis et al |
| EPI_ISL_856709 | MFD Veselības centrs "Dziedniecība" | Latvian Biomedical Research and Study Centre | Ivars Silamiķelis et al |
| EPI_ISL_1014678 | National Influenza Center, Virology Department | National Influenza Center | J Yavarian et al |
| EPI_ISL_959282 | National Influenza Center, Virology Department | National Influenza Center | J Yavarian et al |
| EPI_ISL_862080 | National Influenza Center, Virology Department | National Influenza Center | J Yavarian et al |
| EPI_ISL_1014687 | National Influenza Center, Virology Department | National Influenza Center | J Yavarian et al |

| | | | |
|---|---|---|---|
| EPI_ISL_862077 | National Influenza Center, Virology Department | National Influenza Center | J Yavarian et al |
| EPI_ISL_1014683 | National Influenza Center, Virology Department | National Influenza Center | J Yavarian et al |
| EPI_ISL_886066 | Lighthouse Lab in Alderley Park | Wellcome Sanger Institute for the COVID-19 Genomics UK (COG-UK) Consortium | Jacquelyn Wynn et al |
| EPI_ISL_886148 | Lighthouse Lab in Alderley Park | Wellcome Sanger Institute for the COVID-19 Genomics UK (COG-UK) Consortium | Jacquelyn Wynn et al |
| EPI_ISL_1584044 | Lighthouse Lab in Alderley Park | Wellcome Sanger Institute for the COVID-19 Genomics UK (COG-UK) Consortium | Jacquelyn Wynn et al |
| EPI_ISL_567767 | Lighthouse Lab in Alderley Park | Wellcome Sanger Institute for the COVID-19 Genomics UK (COG-UK) consortium | Jacquelyn Wynn et al |
| EPI_ISL_674292 | Lighthouse Lab in Alderley Park | Wellcome Sanger Institute for the COVID-19 Genomics UK (COG-UK) Consortium | Jacquelyn Wynn et al |
| EPI_ISL_703236 | Lighthouse Lab in Alderley Park | Wellcome Sanger Institute for the COVID-19 Genomics UK (COG-UK) Consortium | Jacquelyn Wynn et al |
| EPI_ISL_833971 | Lighthouse Lab in Alderley Park | Wellcome Sanger Institute for the COVID-19 Genomics UK (COG-UK) Consortium | Jacquelyn Wynn et al |
| EPI_ISL_463741 | Department of Molecular Virology, Cyprus Institute of Neurology and Genetics | Department of Molecular Virology, Cyprus Institute of Neurology and Genetics | Jan Richter et al |
| EPI_ISL_463742 | Department of Molecular Virology, Cyprus Institute of Neurology and Genetics | Department of Molecular Virology, Cyprus Institute of Neurology and Genetics | Jan Richter et al |
| EPI_ISL_463743 | Department of Molecular Virology, Cyprus Institute of Neurology and Genetics | Department of Molecular Virology, Cyprus Institute of Neurology and Genetics | Jan Richter et al |
| EPI_ISL_463744 | Department of Molecular Virology, Cyprus Institute of Neurology and Genetics | Department of Molecular Virology, Cyprus Institute of Neurology and Genetics | Jan Richter et al |
| EPI_ISL_463745 | Department of Molecular Virology, Cyprus Institute of Neurology and Genetics | Department of Molecular Virology, Cyprus Institute of Neurology and Genetics | Jan Richter et al |
| EPI_ISL_463747 | Department of Molecular Virology, Cyprus Institute of Neurology and Genetics | Department of Molecular Virology, Cyprus Institute of Neurology and Genetics | Jan Richter et al |
| EPI_ISL_463748 | Department of Molecular Virology, Cyprus Institute of Neurology and Genetics | Department of Molecular Virology, Cyprus Institute of Neurology and Genetics | Jan Richter et al |
| EPI_ISL_1164626 | Department of Molecular Virology, Cyprus Institute of Neurology and Genetics | Department of Molecular Virology, Cyprus Institute of Neurology and Genetics | Jan Richter et al |
| EPI_ISL_1164627 | Department of Molecular Virology, Cyprus Institute of Neurology and Genetics | Department of Molecular Virology, Cyprus Institute of Neurology and Genetics | Jan Richter et al |
| EPI_ISL_1164628 | Department of Molecular Virology, Cyprus Institute of Neurology and Genetics | Department of Molecular Virology, Cyprus Institute of Neurology and Genetics | Jan Richter et al |
| EPI_ISL_1164629 | Department of Molecular Virology, Cyprus Institute of Neurology and Genetics | Department of Molecular Virology, Cyprus Institute of Neurology and Genetics | Jan Richter et al |
| EPI_ISL_1164630 | Department of Molecular Virology, Cyprus Institute of Neurology and Genetics | Department of Molecular Virology, Cyprus Institute of Neurology and Genetics | Jan Richter et al |
| EPI_ISL_1164631 | Department of Molecular Virology, Cyprus Institute of Neurology and Genetics | Department of Molecular Virology, Cyprus Institute of Neurology and Genetics | Jan Richter et al |

| | | | |
|---|---|---|---|
| EPI_ISL_1164632 | Department of Molecular Virology, Cyprus Institute of Neurology and Genetics | Department of Molecular Virology, Cyprus Institute of Neurology and Genetics | Jan Richter et al |
| EPI_ISL_1164633 | Department of Molecular Virology, Cyprus Institute of Neurology and Genetics | Department of Molecular Virology, Cyprus Institute of Neurology and Genetics | Jan Richter et al |
| EPI_ISL_1164634 | Department of Molecular Virology, Cyprus Institute of Neurology and Genetics | Department of Molecular Virology, Cyprus Institute of Neurology and Genetics | Jan Richter et al |
| EPI_ISL_1164635 | Department of Molecular Virology, Cyprus Institute of Neurology and Genetics | Department of Molecular Virology, Cyprus Institute of Neurology and Genetics | Jan Richter et al |
| EPI_ISL_1164636 | Department of Molecular Virology, Cyprus Institute of Neurology and Genetics | Department of Molecular Virology, Cyprus Institute of Neurology and Genetics | Jan Richter et al |
| EPI_ISL_1164637 | Department of Molecular Virology, Cyprus Institute of Neurology and Genetics | Department of Molecular Virology, Cyprus Institute of Neurology and Genetics | Jan Richter et al |
| EPI_ISL_1164641 | Department of Molecular Virology, Cyprus Institute of Neurology and Genetics | Department of Molecular Virology, Cyprus Institute of Neurology and Genetics | Jan Richter et al |
| EPI_ISL_1164642 | Department of Molecular Virology, Cyprus Institute of Neurology and Genetics | Department of Molecular Virology, Cyprus Institute of Neurology and Genetics | Jan Richter et al |
| EPI_ISL_1164643 | Department of Molecular Virology, Cyprus Institute of Neurology and Genetics | Department of Molecular Virology, Cyprus Institute of Neurology and Genetics | Jan Richter et al |
| EPI_ISL_1164646 | Department of Molecular Virology, Cyprus Institute of Neurology and Genetics | Department of Molecular Virology, Cyprus Institute of Neurology and Genetics | Jan Richter et al |
| EPI_ISL_1164647 | Department of Molecular Virology, Cyprus Institute of Neurology and Genetics | Department of Molecular Virology, Cyprus Institute of Neurology and Genetics | Jan Richter et al |
| EPI_ISL_1164648 | Department of Molecular Virology, Cyprus Institute of Neurology and Genetics | Department of Molecular Virology, Cyprus Institute of Neurology and Genetics | Jan Richter et al |
| EPI_ISL_1164651 | Department of Molecular Virology, Cyprus Institute of Neurology and Genetics | Department of Molecular Virology, Cyprus Institute of Neurology and Genetics | Jan Richter et al |
| EPI_ISL_1164652 | Department of Molecular Virology, Cyprus Institute of Neurology and Genetics | Department of Molecular Virology, Cyprus Institute of Neurology and Genetics | Jan Richter et al |
| EPI_ISL_1164654 | Department of Molecular Virology, Cyprus Institute of Neurology and Genetics | Department of Molecular Virology, Cyprus Institute of Neurology and Genetics | Jan Richter et al |
| EPI_ISL_1164658 | Department of Molecular Virology, Cyprus Institute of Neurology and Genetics | Department of Molecular Virology, Cyprus Institute of Neurology and Genetics | Jan Richter et al |
| EPI_ISL_1164675 | Department of Molecular Virology, Cyprus Institute of Neurology and Genetics | Department of Molecular Virology, Cyprus Institute of Neurology and Genetics | Jan Richter et al |
| EPI_ISL_1164678 | Department of Molecular Virology, Cyprus Institute of Neurology and Genetics | Department of Molecular Virology, Cyprus Institute of Neurology and Genetics | Jan Richter et al |
| EPI_ISL_1164685 | Department of Molecular Virology, Cyprus Institute of Neurology and Genetics | Department of Molecular Virology, Cyprus Institute of Neurology and Genetics | Jan Richter et al |
| EPI_ISL_1164687 | Department of Molecular Virology, Cyprus Institute of Neurology and Genetics | Department of Molecular Virology, Cyprus Institute of Neurology and Genetics | Jan Richter et al |
| EPI_ISL_1164689 | Department of Molecular Virology, Cyprus Institute of Neurology and Genetics | Department of Molecular Virology, Cyprus Institute of Neurology and Genetics | Jan Richter et al |
| EPI_ISL_1164690 | Department of Molecular Virology, Cyprus Institute of Neurology and Genetics | Department of Molecular Virology, Cyprus Institute of Neurology and Genetics | Jan Richter et al |

| | | | |
|---|---|---|---|
| EPI_ISL_1164691 | Department of Molecular Virology, Cyprus Institute of Neurology and Genetics | Department of Molecular Virology, Cyprus Institute of Neurology and Genetics | Jan Richter et al |
| EPI_ISL_1164694 | Department of Molecular Virology, Cyprus Institute of Neurology and Genetics | Department of Molecular Virology, Cyprus Institute of Neurology and Genetics | Jan Richter et al |
| EPI_ISL_1164695 | Department of Molecular Virology, Cyprus Institute of Neurology and Genetics | Department of Molecular Virology, Cyprus Institute of Neurology and Genetics | Jan Richter et al |
| EPI_ISL_1164696 | Department of Molecular Virology, Cyprus Institute of Neurology and Genetics | Department of Molecular Virology, Cyprus Institute of Neurology and Genetics | Jan Richter et al |
| EPI_ISL_1164698 | Department of Molecular Virology, Cyprus Institute of Neurology and Genetics | Department of Molecular Virology, Cyprus Institute of Neurology and Genetics | Jan Richter et al |
| EPI_ISL_1164708 | Department of Molecular Virology, Cyprus Institute of Neurology and Genetics | Department of Molecular Virology, Cyprus Institute of Neurology and Genetics | Jan Richter et al |
| EPI_ISL_1164711 | Department of Molecular Virology, Cyprus Institute of Neurology and Genetics | Department of Molecular Virology, Cyprus Institute of Neurology and Genetics | Jan Richter et al |
| EPI_ISL_1164712 | Department of Molecular Virology, Cyprus Institute of Neurology and Genetics | Department of Molecular Virology, Cyprus Institute of Neurology and Genetics | Jan Richter et al |
| EPI_ISL_1164715 | Department of Molecular Virology, Cyprus Institute of Neurology and Genetics | Department of Molecular Virology, Cyprus Institute of Neurology and Genetics | Jan Richter et al |
| EPI_ISL_1164731 | Department of Molecular Virology, Cyprus Institute of Neurology and Genetics | Department of Molecular Virology, Cyprus Institute of Neurology and Genetics | Jan Richter et al |
| EPI_ISL_1164744 | Department of Molecular Virology, Cyprus Institute of Neurology and Genetics | Department of Molecular Virology, Cyprus Institute of Neurology and Genetics | Jan Richter et al |
| EPI_ISL_1164746 | Department of Molecular Virology, Cyprus Institute of Neurology and Genetics | Department of Molecular Virology, Cyprus Institute of Neurology and Genetics | Jan Richter et al |
| EPI_ISL_483150 | Robert Koch Institute, ZBS1 Highly Pathogenic Viruses, Berlin, Germany | Robert Koch Institute, Bioinformatics MF1, Berlin, Germany | Janine Michel et al |
| EPI_ISL_1312555 | Centrala laboratorija | Latvian Biomedical Research and Study Centre | Janis Pjalkovskis et al |
| EPI_ISL_1312639 | E. Gulbja laboratorija | Latvian Biomedical Research and Study Centre | Janis Pjalkovskis et al |
| EPI_ISL_1312655 | E. Gulbja laboratorija | Latvian Biomedical Research and Study Centre | Janis Pjalkovskis et al |
| EPI_ISL_1312683 | E. Gulbja laboratorija | Latvian Biomedical Research and Study Centre | Janis Pjalkovskis et al |
| EPI_ISL_1312689 | E. Gulbja laboratorija | Latvian Biomedical Research and Study Centre | Janis Pjalkovskis et al |
| EPI_ISL_1312690 | E. Gulbja laboratorija | Latvian Biomedical Research and Study Centre | Janis Pjalkovskis et al |
| EPI_ISL_1312724 | E. Gulbja laboratorija | Latvian Biomedical Research and Study Centre | Janis Pjalkovskis et al |
| EPI_ISL_1321930 | E. Gulbja laboratorija | Latvian Biomedical Research and Study Centre | Janis Pjalkovskis et al |
| EPI_ISL_1613124 | LIC | Latvian Biomedical Research and Study Centre | Janis Pjalkovskis et al |
| EPI_ISL_1613125 | LIC | Latvian Biomedical Research and Study Centre | Janis Pjalkovskis et al |
| EPI_ISL_1613127 | LIC | Latvian Biomedical Research and Study Centre | Janis Pjalkovskis et al |
| EPI_ISL_1613776 | E. Gulbja laboratorija | Latvian Biomedical Research and Study Centre | Janis Pjalkovskis et al |
| EPI_ISL_961801 | E. Gulbja laboratorija | Latvian Biomedical Research and Study Centre | Janis Pjalkovskis et al |

| | | | |
|---|---|---|---|
| EPI_ISL_1219217 | E. Gulbja laboratorija | Latvian Biomedical Research and Study Centre | Janis Pjalkovskis et al |
| EPI_ISL_1219423 | BIOR | Latvian Biomedical Research and Study Centre | Janis Pjalkovskis et al |
| EPI_ISL_1219558 | E. Gulbja laboratorija | Latvian Biomedical Research and Study Centre | Janis Pjalkovskis et al |
| EPI_ISL_1219603 | E. Gulbja laboratorija | Latvian Biomedical Research and Study Centre | Janis Pjalkovskis et al |
| EPI_ISL_1219663 | E. Gulbja laboratorija | Latvian Biomedical Research and Study Centre | Janis Pjalkovskis et al |
| EPI_ISL_1312403 | E. Gulbja laboratorija | Latvian Biomedical Research and Study Centre | Janis Pjalkovskis et al |
| EPI_ISL_1312417 | E. Gulbja laboratorija | Latvian Biomedical Research and Study Centre | Janis Pjalkovskis et al |
| EPI_ISL_1312455 | E. Gulbja laboratorija | Latvian Biomedical Research and Study Centre | Janis Pjalkovskis et al |
| EPI_ISL_1312476 | E. Gulbja laboratorija | Latvian Biomedical Research and Study Centre | Janis Pjalkovskis et al |
| EPI_ISL_1312496 | E. Gulbja laboratorija | Latvian Biomedical Research and Study Centre | Janis Pjalkovskis et al |
| EPI_ISL_1312499 | LIC | Latvian Biomedical Research and Study Centre | Janis Pjalkovskis et al |
| EPI_ISL_1544184 | Government Medical College (GMC), Surat | Gujarat Biotechnology Research Centre | Janvi Raval et al |
| EPI_ISL_412964 | Hospital Israelita Albert Einstein | Instituto Adolfo Lutz Interdisciplinary Procedures Center Strategic Laboratory | Jaqueline Goes de Jesus et al |
| EPI_ISL_413016 | Hospital Israelita Albert Einstein | Instituto Adolfo Lutz, Interdisciplinary Procedures Center, Strategic Laboratory | Jaqueline Goes de Jesus et al |
| EPI_ISL_994658 | Dr. Andrija Stampar“ Teaching Institute of Public Health, Department of Clinical Microbiology | Institute of Applied Genomics | Jasmina Vranes et al |
| EPI_ISL_994659 | Dr. Andrija Stampar“ Teaching Institute of Public Health, Department of Clinical Microbiology | Institute of Applied Genomics | Jasmina Vranes et al |
| EPI_ISL_994667 | Dr. Andrija Stampar“ Teaching Institute of Public Health, Department of Clinical Microbiology | Institute of Applied Genomics | Jasmina Vranes et al |
| EPI_ISL_994661 | Dr. Andrija Stampar“ Teaching Institute of Public Health, Department of Clinical Microbiology | Institute of Applied Genomics | Jasmina Vranes et al |
| EPI_ISL_994740 | Dr. Andrija Stampar“ Teaching Institute of Public Health, Department of Clinical Microbiology | Institute of Applied Genomics | Jasmina Vranes et al |
| EPI_ISL_996004 | Dr. Andrija Stampar“ Teaching Institute of Public Health, Department of Clinical Microbiology | Institute of Applied Genomics | Jasmina Vranes et al |
| EPI_ISL_994739 | Dr. Andrija Stampar“ Teaching Institute of Public Health, Department of Clinical Microbiology | Institute of Applied Genomics | Jasmina Vranes et al |
| EPI_ISL_1167040 | Dr. Andrija Stampar Teaching Institute of Public Health, Department of Clinical Microbiology | Istituto di Genomica Applicata | Jasmina Vranes et al |
| EPI_ISL_1167044 | Dr. Andrija Stampar Teaching Institute of Public Health, Department of Clinical Microbiology | Istituto di Genomica Applicata | Jasmina Vranes et al |
| EPI_ISL_1167045 | Dr. Andrija Stampar Teaching Institute of Public Health, Department of Clinical Microbiology | Istituto di Genomica Applicata | Jasmina Vranes et al |
| EPI_ISL_1167046 | Dr. Andrija Stampar Teaching Institute of Public Health, Department of Clinical Microbiology | Istituto di Genomica Applicata | Jasmina Vranes et al |

| | | | |
|---|---|---|---|
| EPI_ISL_1167049 | Dr. Andrija Stampar Teaching Institute of Public Health, Department of Clinical Microbiology | Istituto di Genomica Applicata | Jasmina Vranes et al |
| EPI_ISL_1167053 | Dr. Andrija Stampar Teaching Institute of Public Health, Department of Clinical Microbiology | Istituto di Genomica Applicata | Jasmina Vranes et al |
| EPI_ISL_1167054 | Dr. Andrija Stampar Teaching Institute of Public Health, Department of Clinical Microbiology | Istituto di Genomica Applicata | Jasmina Vranes et al |
| EPI_ISL_1167057 | Dr. Andrija Stampar Teaching Institute of Public Health, Department of Clinical Microbiology | Istituto di Genomica Applicata | Jasmina Vranes et al |
| EPI_ISL_1167067 | Dr. Andrija Stampar Teaching Institute of Public Health, Department of Clinical Microbiology | Istituto di Genomica Applicata | Jasmina Vranes et al |
| EPI_ISL_1167082 | Dr. Andrija Stampar Teaching Institute of Public Health, Department of Clinical Microbiology | Istituto di Genomica Applicata | Jasmina Vranes et al |
| EPI_ISL_1167083 | Dr. Andrija Stampar Teaching Institute of Public Health, Department of Clinical Microbiology | Istituto di Genomica Applicata | Jasmina Vranes et al |
| EPI_ISL_1167084 | Dr. Andrija Stampar Teaching Institute of Public Health, Department of Clinical Microbiology | Istituto di Genomica Applicata | Jasmina Vranes et al |
| EPI_ISL_1167085 | Dr. Andrija Stampar Teaching Institute of Public Health, Department of Clinical Microbiology | Istituto di Genomica Applicata | Jasmina Vranes et al |
| EPI_ISL_1167028 | Dr. Andrija Stampar Teaching Institute of Public Health, Department of Clinical Microbiology | Istituto di Genomica Applicata | Jasmina Vranes et al |
| EPI_ISL_1661222 | Platform BIS UZA/UAntwerpen | Labo Klinische Biologie, UZA | Jasmine Coppens et al |
| EPI_ISL_1661211 | Platform BIS UZA/UAntwerpen | Labo Klinische Biologie, UZA | Jasmine Coppens et al |
| EPI_ISL_417444 | Department of Healthcare Biotechnology, National University of Sciences and Technology (NUST) | Department of Healthcare Biotechnology, National University of Sciences and Technology (NUST) | Javed et al |
| EPI_ISL_1300508 | Genetica Molecular and Subdepartamento de Virologia ISP Chile | Instituto de Salud Publica de Chile | Javier Tognarelli et al |
| EPI_ISL_1167794 | Genetica Molecular and Subdepartamento de Virologia ISP Chile | Instituto de Salud Publica de Chile | Javier Tognarelli et al |
| EPI_ISL_1541058 | Genetica Molecular and Subdepartamento de Virologia ISP Chile | Instituto de Salud Publica de Chile | Javier Tognarelli et al |
| EPI_ISL_746499 | Genetica Molecular and Subdepartamento de Virologia ISP Chile | Instituto de Salud Publica de Chile | Javier Tognarelli et al |
| EPI_ISL_746503 | Genetica Molecular and Subdepartamento de Virologia ISP Chile | Instituto de Salud Publica de Chile | Javier Tognarelli et al |
| EPI_ISL_1167844 | Genetica Molecular and Subdepartamento de Virologia ISP Chile | Instituto de Salud Publica de Chile | Javier Tognarelli et al |
| EPI_ISL_1167835 | Genetica Molecular and Subdepartamento de Virologia ISP Chile | Instituto de Salud Publica de Chile | Javier Tognarelli et al |
| EPI_ISL_1300511 | Genetica Molecular and Subdepartamento de Virologia ISP Chile | Instituto de Salud Publica de Chile | Javier Tognarelli et al |
| EPI_ISL_1321495 | Genetica Molecular and Subdepartamento de Virologia ISP Chile | Instituto de Salud Publica de Chile | Javier Tognarelli et al |
| EPI_ISL_1321498 | Genetica Molecular and Subdepartamento de Virologia ISP Chile | Instituto de Salud Publica de Chile | Javier Tognarelli et al |
| EPI_ISL_1167875 | Genetica Molecular and Subdepartamento de Virologia ISP Chile | Instituto de Salud Publica de Chile | Javier Tognarelli et al |
| EPI_ISL_1167871 | Genetica Molecular and Subdepartamento de Virologia ISP Chile | Instituto de Salud Publica de Chile | Javier Tognarelli et al |
| EPI_ISL_1633549 | Genetica Molecular and Subdepartamento de Virologia ISP Chile | Instituto de Salud Publica de Chile | Javier Tognarelli et al |

| | | | |
|---|---|---|---|
| EPI_ISL_1167804 | Genetica Molecular and Subdepartamento de Virologia ISP Chile | Instituto de Salud Publica de Chile | Javier Tognarelli et al |
| EPI_ISL_746509 | Genetica Molecular and Subdepartamento de Virologia ISP Chile | Instituto de Salud Publica de Chile | Javier Tognarelli et al |
| EPI_ISL_746518 | Genetica Molecular and Subdepartamento de Virologia ISP Chile | Instituto de Salud Publica de Chile | Javier Tognarelli et al |
| EPI_ISL_1167810 | Genetica Molecular and Subdepartamento de Virologia ISP Chile | Instituto de Salud Publica de Chile | Javier Tognarelli et al |
| EPI_ISL_1300502 | Genetica Molecular and Subdepartamento de Virologia ISP Chile | Instituto de Salud Publica de Chile | Javier Tognarelli et al |
| EPI_ISL_1470478 | Genetica Molecular and Subdepartamento de Virologia ISP Chile | Instituto de Salud Publica de Chile | Javier Tognarelli et al |
| EPI_ISL_1167907 | Genetica Molecular and Subdepartamento de Virologia ISP Chile | Instituto de Salud Publica de Chile | Javier Tognarelli et al |
| EPI_ISL_1321578 | Genetica Molecular and Subdepartamento de Virologia ISP Chile | Instituto de Salud Publica de Chile | Javier Tognarelli et al |
| EPI_ISL_1321468 | Genetica Molecular and Subdepartamento de Virologia ISP Chile | Instituto de Salud Publica de Chile | Javier Tognarelli et al |
| EPI_ISL_1167796 | Genetica Molecular and Subdepartamento de Virologia ISP Chile | Instituto de Salud Publica de Chile | Javier Tognarelli et al |
| EPI_ISL_1167861 | Genetica Molecular and Subdepartamento de Virologia ISP Chile | Instituto de Salud Publica de Chile | Javier Tognarelli et al |
| EPI_ISL_1321469 | Genetica Molecular and Subdepartamento de Virologia ISP Chile | Instituto de Salud Publica de Chile | Javier Tognarelli et al |
| EPI_ISL_1321505 | Genetica Molecular and Subdepartamento de Virologia ISP Chile | Instituto de Salud Publica de Chile | Javier Tognarelli et al |
| EPI_ISL_1321470 | Genetica Molecular and Subdepartamento de Virologia ISP Chile | Instituto de Salud Publica de Chile | Javier Tognarelli et al |
| EPI_ISL_1470531 | Genetica Molecular and Subdepartamento de Virologia ISP Chile | Instituto de Salud Publica de Chile | Javier Tognarelli et al |
| EPI_ISL_1541032 | Genetica Molecular and Subdepartamento de Virologia ISP Chile | Instituto de Salud Publica de Chile | Javier Tognarelli et al |
| EPI_ISL_1541033 | Genetica Molecular and Subdepartamento de Virologia ISP Chile | Instituto de Salud Publica de Chile | Javier Tognarelli et al |
| EPI_ISL_1167918 | Genetica Molecular and Subdepartamento de Virologia ISP Chile | Instituto de Salud Publica de Chile | Javier Tognarelli et al |
| EPI_ISL_1167668 | Genetica Molecular and Subdepartamento de Virologia ISP Chile | Instituto de Salud Publica de Chile | Javier Tognarelli et al |
| EPI_ISL_746545 | Genetica Molecular and Subdepartamento de Virologia ISP Chile | Instituto de Salud Publica de Chile | Javier Tognarelli et al |
| EPI_ISL_746559 | Genetica Molecular and Subdepartamento de Virologia ISP Chile | Instituto de Salud Publica de Chile | Javier Tognarelli et al |
| EPI_ISL_1167842 | Genetica Molecular and Subdepartamento de Virologia ISP Chile | Instituto de Salud Publica de Chile | Javier Tognarelli et al |
| EPI_ISL_746570 | Genetica Molecular and Subdepartamento de Virologia ISP Chile | Instituto de Salud Publica de Chile | Javier Tognarelli et al |
| EPI_ISL_746589 | Genetica Molecular and Subdepartamento de Virologia ISP Chile | Instituto de Salud Publica de Chile | Javier Tognarelli et al |
| EPI_ISL_746590 | Genetica Molecular and Subdepartamento de Virologia ISP Chile | Instituto de Salud Publica de Chile | Javier Tognarelli et al |
| EPI_ISL_746622 | Genetica Molecular and Subdepartamento de Virologia ISP Chile | Instituto de Salud Publica de Chile | Javier Tognarelli et al |
| EPI_ISL_746624 | Genetica Molecular and Subdepartamento de Virologia ISP Chile | Instituto de Salud Publica de Chile | Javier Tognarelli et al |
| EPI_ISL_746628 | Genetica Molecular and Subdepartamento de Virologia ISP Chile | Instituto de Salud Publica de Chile | Javier Tognarelli et al |
| EPI_ISL_746635 | Genetica Molecular and Subdepartamento de Virologia ISP Chile | Instituto de Salud Publica de Chile | Javier Tognarelli et al |
| EPI_ISL_746640 | Genetica Molecular and Subdepartamento de Virologia ISP Chile | Instituto de Salud Publica de Chile | Javier Tognarelli et al |

| | | | |
|---|---|---|---|
| EPI_ISL_746644 | Genetica Molecular and Subdepartamento de Virologia ISP Chile | Instituto de Salud Publica de Chile | Javier Tognarelli et al |
| EPI_ISL_746660 | Genetica Molecular and Subdepartamento de Virologia ISP Chile | Instituto de Salud Publica de Chile | Javier Tognarelli et al |
| EPI_ISL_746666 | Genetica Molecular and Subdepartamento de Virologia ISP Chile | Instituto de Salud Publica de Chile | Javier Tognarelli et al |
| EPI_ISL_746673 | Genetica Molecular and Subdepartamento de Virologia ISP Chile | Instituto de Salud Publica de Chile | Javier Tognarelli et al |
| EPI_ISL_746674 | Genetica Molecular and Subdepartamento de Virologia ISP Chile | Instituto de Salud Publica de Chile | Javier Tognarelli et al |
| EPI_ISL_746675 | Genetica Molecular and Subdepartamento de Virologia ISP Chile | Instituto de Salud Publica de Chile | Javier Tognarelli et al |
| EPI_ISL_746682 | Genetica Molecular and Subdepartamento de Virologia ISP Chile | Instituto de Salud Publica de Chile | Javier Tognarelli et al |
| EPI_ISL_746685 | Genetica Molecular and Subdepartamento de Virologia ISP Chile | Instituto de Salud Publica de Chile | Javier Tognarelli et al |
| EPI_ISL_746686 | Genetica Molecular and Subdepartamento de Virologia ISP Chile | Instituto de Salud Publica de Chile | Javier Tognarelli et al |
| EPI_ISL_746687 | Genetica Molecular and Subdepartamento de Virologia ISP Chile | Instituto de Salud Publica de Chile | Javier Tognarelli et al |
| EPI_ISL_746706 | Genetica Molecular and Subdepartamento de Virologia ISP Chile | Instituto de Salud Publica de Chile | Javier Tognarelli et al |
| EPI_ISL_746723 | Genetica Molecular and Subdepartamento de Virologia ISP Chile | Instituto de Salud Publica de Chile | Javier Tognarelli et al |
| EPI_ISL_746736 | Genetica Molecular and Subdepartamento de Virologia ISP Chile | Instituto de Salud Publica de Chile | Javier Tognarelli et al |
| EPI_ISL_746743 | Genetica Molecular and Subdepartamento de Virologia ISP Chile | Instituto de Salud Publica de Chile | Javier Tognarelli et al |
| EPI_ISL_746750 | Genetica Molecular and Subdepartamento de Virologia ISP Chile | Instituto de Salud Publica de Chile | Javier Tognarelli et al |
| EPI_ISL_746754 | Genetica Molecular and Subdepartamento de Virologia ISP Chile | Instituto de Salud Publica de Chile | Javier Tognarelli et al |
| EPI_ISL_1300459 | Genetica Molecular and Subdepartamento de Virologia ISP Chile | Instituto de Salud Publica de Chile | Javier Tognarelli et al |
| EPI_ISL_746759 | Genetica Molecular and Subdepartamento de Virologia ISP Chile | Instituto de Salud Publica de Chile | Javier Tognarelli et al |
| EPI_ISL_746761 | Genetica Molecular and Subdepartamento de Virologia ISP Chile | Instituto de Salud Publica de Chile | Javier Tognarelli et al |
| EPI_ISL_746767 | Genetica Molecular and Subdepartamento de Virologia ISP Chile | Instituto de Salud Publica de Chile | Javier Tognarelli et al |
| EPI_ISL_1167696 | Genetica Molecular and Subdepartamento de Virologia ISP Chile | Instituto de Salud Publica de Chile | Javier Tognarelli et al |
| EPI_ISL_1167705 | Genetica Molecular and Subdepartamento de Virologia ISP Chile | Instituto de Salud Publica de Chile | Javier Tognarelli et al |
| EPI_ISL_1167707 | Genetica Molecular and Subdepartamento de Virologia ISP Chile | Instituto de Salud Publica de Chile | Javier Tognarelli et al |
| EPI_ISL_1167710 | Genetica Molecular and Subdepartamento de Virologia ISP Chile | Instituto de Salud Publica de Chile | Javier Tognarelli et al |
| EPI_ISL_1167711 | Genetica Molecular and Subdepartamento de Virologia ISP Chile | Instituto de Salud Publica de Chile | Javier Tognarelli et al |
| EPI_ISL_1167716 | Genetica Molecular and Subdepartamento de Virologia ISP Chile | Instituto de Salud Publica de Chile | Javier Tognarelli et al |
| EPI_ISL_1167722 | Genetica Molecular and Subdepartamento de Virologia ISP Chile | Instituto de Salud Publica de Chile | Javier Tognarelli et al |
| EPI_ISL_1167724 | Genetica Molecular and Subdepartamento de Virologia ISP Chile | Instituto de Salud Publica de Chile | Javier Tognarelli et al |
| EPI_ISL_1167736 | Genetica Molecular and Subdepartamento de Virologia ISP Chile | Instituto de Salud Publica de Chile | Javier Tognarelli et al |
| EPI_ISL_1167847 | Genetica Molecular and Subdepartamento de Virologia ISP Chile | Instituto de Salud Publica de Chile | Javier Tognarelli et al |

| | | | |
|---|---|---|---|
| EPI_ISL_1167877 | Genetica Molecular and Subdepartamento de Virologia ISP Chile | Instituto de Salud Publica de Chile | Javier Tognarelli et al |
| EPI_ISL_1167883 | Genetica Molecular and Subdepartamento de Virologia ISP Chile | Instituto de Salud Publica de Chile | Javier Tognarelli et al |
| EPI_ISL_1321466 | Genetica Molecular and Subdepartamento de Virologia ISP Chile | Instituto de Salud Publica de Chile | Javier Tognarelli et al |
| EPI_ISL_1321560 | Genetica Molecular and Subdepartamento de Virologia ISP Chile | Instituto de Salud Publica de Chile | Javier Tognarelli et al |
| EPI_ISL_1321512 | Genetica Molecular and Subdepartamento de Virologia ISP Chile | Instituto de Salud Publica de Chile | Javier Tognarelli et al |
| EPI_ISL_1321515 | Genetica Molecular and Subdepartamento de Virologia ISP Chile | Instituto de Salud Publica de Chile | Javier Tognarelli et al |
| EPI_ISL_1167815 | Genetica Molecular and Subdepartamento de Virologia ISP Chile | Instituto de Salud Publica de Chile | Javier Tognarelli et al |
| EPI_ISL_1470501 | Genetica Molecular and Subdepartamento de Virologia ISP Chile | Instituto de Salud Publica de Chile | Javier Tognarelli et al |
| EPI_ISL_1470440 | Genetica Molecular and Subdepartamento de Virologia ISP Chile | Instituto de Salud Publica de Chile | Javier Tognarelli et al |
| EPI_ISL_1470457 | Genetica Molecular and Subdepartamento de Virologia ISP Chile | Instituto de Salud Publica de Chile | Javier Tognarelli et al |
| EPI_ISL_1534590 | Genetica Molecular and Subdepartamento de Virologia ISP Chile | Instituto de Salud Publica de Chile | Javier Tognarelli et al |
| EPI_ISL_1633493 | Genetica Molecular and Subdepartamento de Virologia ISP Chile | Instituto de Salud Publica de Chile | Javier Tognarelli et al |
| EPI_ISL_1541038 | Genetica Molecular and Subdepartamento de Virologia ISP Chile | Instituto de Salud Publica de Chile | Javier Tognarelli et al |
| EPI_ISL_1167771 | Genetica Molecular and Subdepartamento de Virologia ISP Chile | Instituto de Salud Publica de Chile | Javier Tognarelli et al |
| EPI_ISL_1541064 | Genetica Molecular and Subdepartamento de Virologia ISP Chile | Instituto de Salud Publica de Chile | Javier Tognarelli et al |
| EPI_ISL_1633360 | Genetica Molecular and Subdepartamento de Virologia ISP Chile | Instituto de Salud Publica de Chile | Javier Tognarelli et al |
| EPI_ISL_1633362 | Genetica Molecular and Subdepartamento de Virologia ISP Chile | Instituto de Salud Publica de Chile | Javier Tognarelli et al |
| EPI_ISL_1167772 | Genetica Molecular and Subdepartamento de Virologia ISP Chile | Instituto de Salud Publica de Chile | Javier Tognarelli et al |
| EPI_ISL_746807 | Genetica Molecular and Subdepartamento de Virologia ISP Chile | Instituto de Salud Publica de Chile | Javier Tognarelli et al |
| EPI_ISL_746484 | Genetica Molecular and Subdepartamento de Virologia ISP Chile | Instituto de Salud Publica de Chile | Javier Tognarelli et al |
| EPI_ISL_1300499 | Genetica Molecular and Subdepartamento de Virologia ISP Chile | Instituto de Salud Publica de Chile | Javier Tognarelli et al |
| EPI_ISL_1321471 | Genetica Molecular and Subdepartamento de Virologia ISP Chile | Instituto de Salud Publica de Chile | Javier Tognarelli et al |
| EPI_ISL_1534607 | Genetica Molecular and Subdepartamento de Virologia ISP Chile | Instituto de Salud Publica de Chile | Javier Tognarelli et al |
| EPI_ISL_826830 | INSPI-CRN DE INFLUENZA Y OTROS VIRUS RESPIRATORIOS | Instituto de Salud Publica de Chile | Javier Tognarelli et al |
| EPI_ISL_826832 | INSPI-CRN DE INFLUENZA Y OTROS VIRUS RESPIRATORIOS | Instituto de Salud Publica de Chile | Javier Tognarelli et al |
| EPI_ISL_826834 | INSPI-CRN DE INFLUENZA Y OTROS VIRUS RESPIRATORIOS | Instituto de Salud Publica de Chile | Javier Tognarelli et al |
| EPI_ISL_826800 | INSPI-CRN DE INFLUENZA Y OTROS VIRUS RESPIRATORIOS | Instituto de Salud Publica de Chile | Javier Tognarelli et al |
| EPI_ISL_826818 | INSPI-CRN DE INFLUENZA Y OTROS VIRUS RESPIRATORIOS | Instituto de Salud Publica de Chile | Javier Tognarelli et al |
| EPI_ISL_826836 | INSPI-CRN DE INFLUENZA Y OTROS VIRUS RESPIRATORIOS | Instituto de Salud Publica de Chile | Javier Tognarelli et al |
| EPI_ISL_831020 | INSPI-CRN DE INFLUENZA Y OTROS VIRUS RESPIRATORIOS | Instituto de Salud Publica de Chile | Javier Tognarelli et al |

| | | | |
|---|---|---|---|
| EPI_ISL_826820 | INSPI-CRN DE INFLUENZA Y OTROS VIRUS RESPIRATORIOS | Instituto de Salud Publica de Chile | Javier Tognarelli et al |
| EPI_ISL_826822 | INSPI-CRN DE INFLUENZA Y OTROS VIRUS RESPIRATORIOS | Instituto de Salud Publica de Chile | Javier Tognarelli et al |
| EPI_ISL_826825 | INSPI-CRN DE INFLUENZA Y OTROS VIRUS RESPIRATORIOS | Instituto de Salud Publica de Chile | Javier Tognarelli et al |
| EPI_ISL_636987 | Virology Lab, National Institute for Biomedical Research (INRB) | Project group Epidemology of Highly Pathogenic Microorganisms, Robert Koch-Institute | Jean-Jacques Muyembe Tamfum et al |
| EPI_ISL_1014102 | Zurita & Zurita Laboratorios | Zurita & Zurita Laboratorios | Jeannete Zurita et al |
| EPI_ISL_1040028 | Original detection - Virology Unit, Institut Pasteur du Cambodge; Sequencing - US National Institute of Allergy and Infectious Diseases Cambodia | Virology Unit, Institut Pasteur du Cambodge | Jennifer Bohl et al |
| EPI_ISL_412869 | Division of Viral Diseases, Center for Laboratory Control of Infectious Diseases, Korea Centers for Diseases Control and Prevention | Division of Viral Diseases, Center for Laboratory Control of Infectious Diseases, Korea Centers for Diseases Control and Prevention | Jeong-Min Kim et al |
| EPI_ISL_412871 | Division of Viral Diseases, Center for Laboratory Control of Infectious Diseases, Korea Centers for Diseases Control and Prevention | Division of Viral Diseases, Center for Laboratory Control of Infectious Diseases, Korea Centers for Diseases Control and Prevention | Jeong-Min Kim et al |
| EPI_ISL_425118 | Division of Viral Diseases, Center for Laboratory Control of Infectious Diseases, Korea Centers for Diseases Control and Prevention | Division of Viral Diseases, Center for Laboratory Control of Infectious Diseases, Korea Centers for Diseases Control and Prevention | Jeong-Min Kim et al |
| EPI_ISL_426163 | Division of Viral Diseases, Center for Laboratory Control of Infectious Diseases, Korea Centers for Diseases Control and Prevention | Division of Viral Diseases, Center for Laboratory Control of Infectious Diseases, Korea Centers for Diseases Control and Prevention | Jeong-Min Kim et al |
| EPI_ISL_426187 | Division of Viral Diseases, Center for Laboratory Control of Infectious Diseases, Korea Centers for Diseases Control and Prevention | Division of Viral Diseases, Center for Laboratory Control of Infectious Diseases, Korea Centers for Diseases Control and Prevention | Jeong-Min Kim et al |
| EPI_ISL_497962 | Division of Viral Diseases, Center for Laboratory Control of Infectious Diseases, Korea Centers for Diseases Control and Prevention | Division of Viral Diseases, Center for Laboratory Control of Infectious Diseases, Korea Centers for Diseases Control and Prevention | Jeong-Min Kim et al |
| EPI_ISL_497966 | Division of Viral Diseases, Center for Laboratory Control of Infectious Diseases, Korea Centers for Diseases Control and Prevention | Division of Viral Diseases, Center for Laboratory Control of Infectious Diseases, Korea Centers for Diseases Control and Prevention | Jeong-Min Kim et al |
| EPI_ISL_497970 | Division of Viral Diseases, Center for Laboratory Control of Infectious Diseases, Korea Centers for Diseases Control and Prevention | Division of Viral Diseases, Center for Laboratory Control of Infectious Diseases, Korea Centers for Diseases Control and Prevention | Jeong-Min Kim et al |
| EPI_ISL_497974 | Division of Viral Diseases, Center for Laboratory Control of Infectious Diseases, Korea Centers for Diseases Control and Prevention | Division of Viral Diseases, Center for Laboratory Control of Infectious Diseases, Korea Centers for Diseases Control and Prevention | Jeong-Min Kim et al |
| EPI_ISL_498037 | Division of Viral Diseases, Center for Laboratory Control of Infectious Diseases, Korea Centers for Diseases Control and Prevention | Division of Viral Diseases, Center for Laboratory Control of Infectious Diseases, Korea Centers for Diseases Control and Prevention | Jeong-Min Kim et al |
| EPI_ISL_498049 | Division of Viral Diseases, Center for Laboratory Control of Infectious Diseases, Korea Centers for Diseases Control and Prevention | Division of Viral Diseases, Center for Laboratory Control of Infectious Diseases, Korea Centers for Diseases Control and Prevention | Jeong-Min Kim et al |
| EPI_ISL_506982 | Division of Viral Diseases, Center for Laboratory Control of Infectious Diseases, Korea Centers for Diseases Control and Prevention | Division of Viral Diseases, Center for Laboratory Control of Infectious Diseases, Korea Centers for Diseases Control and Prevention | Jeong-Min Kim et al |
| EPI_ISL_506987 | Division of Viral Diseases, Center for Laboratory Control of Infectious Diseases, Korea Centers for Diseases Control and Prevention | Division of Viral Diseases, Center for Laboratory Control of Infectious Diseases, Korea Centers for Diseases Control and Prevention | Jeong-Min Kim et al |
| EPI_ISL_506991 | Division of Viral Diseases, Center for Laboratory Control of Infectious Diseases, Korea Centers for Diseases Control and Prevention | Division of Viral Diseases, Center for Laboratory Control of Infectious Diseases, Korea Centers for Diseases Control and Prevention | Jeong-Min Kim et al |
| EPI_ISL_510627 | Division of Viral Diseases, Center for Laboratory Control of Infectious Diseases, Korea Centers for Diseases Control and Prevention | Division of Viral Diseases, Center for Laboratory Control of Infectious Diseases, Korea Centers for Diseases Control and Prevention | Jeong-Min Kim et al |
| EPI_ISL_510636 | Division of Viral Diseases, Center for Laboratory Control of Infectious Diseases, Korea Centers for Diseases Control and Prevention | Division of Viral Diseases, Center for Laboratory Control of Infectious Diseases, Korea Centers for Diseases Control and Prevention | Jeong-Min Kim et al |
| EPI_ISL_514830 | Division of Viral Diseases, Center for Laboratory Control of Infectious Diseases, Korea Centers for Diseases Control and Prevention | Division of Viral Diseases, Center for Laboratory Control of Infectious Diseases, Korea Centers for Diseases Control and Prevention | Jeong-Min Kim et al |

| | | | |
|---|---|---|---|
| EPI_ISL_514884 | Division of Viral Diseases, Center for Laboratory Control of Infectious Diseases, Korea Centers for Diseases Control and Prevention | Division of Viral Diseases, Center for Laboratory Control of Infectious Diseases, Korea Centers for Diseases Control and Prevention | Jeong-Min Kim et al |
| EPI_ISL_522490 | Division of Viral Diseases, Center for Laboratory Control of Infectious Diseases, Korea Centers for Diseases Control and Prevention | Division of Viral Diseases, Center for Laboratory Control of Infectious Diseases, Korea Centers for Diseases Control and Prevention | Jeong-Min Kim et al |
| EPI_ISL_1039541 | LSUHS Emerging Viral Threat Laboratory | Microbial Genome Sequencing Center | Jeremy P. Kamil et al |
| EPI_ISL_913073 | Center for Virology | Center for Virology | Jeremy V. Camp et al |
| EPI_ISL_913064 | Center for Virology | Center for Virology | Jeremy V. Camp et al |
| EPI_ISL_913091 | Center for Virology | Center for Virology | Jeremy V. Camp et al |
| EPI_ISL_913063 | Labor Mustafa Wien | Center for Virology | Jeremy V. Camp et al |
| EPI_ISL_913079 | Center for Virology | Center for Virology | Jeremy V. Camp et al |
| EPI_ISL_1056465 | National Laboratory for Health, Environment and Food | CISLD (Clinical Institute of Special Laboratory Diagnostics), University Children's Hospital, University Medical Center Ljubljana | Jernej Kovač et al |
| EPI_ISL_1056460 | National Laboratory for Health, Environment and Food | CISLD (Clinical Institute of Special Laboratory Diagnostics), University Children's Hospital, University Medical Center Ljubljana | Jernej Kovač et al |
| EPI_ISL_1240606 | National Laboratory for Health, Environment and Food, OMM, Maribor | CISLD (Clinical Institute of Special Laboratory Diagnostics), University Children's Hospital, University Medical Center Ljubljana | Jernej Kovač et al |
| EPI_ISL_1267014 | National Laboratory for Health, Environment and Food, OMM, Maribor | CISLD (Clinical Institute of Special Laboratory Diagnostics), University Children's Hospital, University Medical Center Ljubljana | Jernej Kovač et al |
| EPI_ISL_1266463 | National Laboratory for Health, Environment and Food, OMM, Kranj | CISLD (Clinical Institute of Special Laboratory Diagnostics), University Children's Hospital, University Medical Center Ljubljana | Jernej Kovač et al |
| EPI_ISL_1335466 | National Laboratory for Health, Environment and Food, OMM, Celje | CISLD (Clinical Institute of Special Laboratory Diagnostics), University Children's Hospital, University Medical Center Ljubljana | Jernej Kovač et al |
| EPI_ISL_1335712 | National Laboratory for Health, Environment and Food, OMM, Celje | CISLD (Clinical Institute of Special Laboratory Diagnostics), University Children's Hospital, University Medical Center Ljubljana | Jernej Kovač et al |
| EPI_ISL_1056256 | National Laboratory for Health, Environment and Food | CISLD (Clinical Institute of Special Laboratory Diagnostics), University Children's Hospital, University Medical Center Ljubljana | Jernej Kovač et al |
| EPI_ISL_1056454 | National Laboratory for Health, Environment and Food | CISLD (Clinical Institute of Special Laboratory Diagnostics), University Children's Hospital, University Medical Center Ljubljana | Jernej Kovač et al |
| EPI_ISL_1055853 | National Laboratory for Health, Environment and Food | CISLD (Clinical Institute of Special Laboratory Diagnostics), University Children's Hospital, University Medical Center Ljubljana | Jernej Kovač et al |
| EPI_ISL_458081 | RSUD Bangil Pasuruan | Institute of Tropical Disease, Universitas Airlangga | Jezzy R Dewantari et al |
| EPI_ISL_759961 | Universitas Airlangga Hospital | Institute of Tropical Disease, Universitas Airlangga | Jezzy R Dewantari et al |
| EPI_ISL_406031 | Centers for Disease Control, R.O.C. (Taiwan) | Centers for Disease Control, R.O.C. (Taiwan) | Ji-Rong Yang et al |
| EPI_ISL_411926 | Taiwan Centers for Disease Control | Taiwan Centers for Disease Control | Ji-Rong Yang et al |
| EPI_ISL_428489 | Centers for Disease Control, R.O.C. (Taiwan) | Centers for Disease Control, R.O.C. (Taiwan) | Ji-Rong Yang et al |
| EPI_ISL_428488 | Centers for Disease Control, R.O.C. (Taiwan) | Centers for Disease Control, R.O.C. (Taiwan) | Ji-Rong Yang et al |
| EPI_ISL_413868 | Guangdong Provincial Institution of Public Health, Guangdong Provinical Center for Disease Control and Prevention | Guangdong Provincial Institution of Public Health | Jing Lu et al |

| | | | |
|---|---|---|---|
| EPI_ISL_413873 | Guangdong Provincial Institution of Public Health, Guangdong Provinical Center for Disease Control and Prevention | Guangdong Provincial Institution of Public Health | Jing Lu et al |
| EPI_ISL_509711 | Belize Ministry of Health | Pathogen Discovery, Respiratory Viruses Branch, Division of Viral Diseases, Centers for Disease Control and Prevention | Jing Zhang et al |
| EPI_ISL_509712 | Belize Ministry of Health | Pathogen Discovery, Respiratory Viruses Branch, Division of Viral Diseases, Centers for Disease Control and Prevention | Jing Zhang et al |
| EPI_ISL_509713 | Belize Ministry of Health | Pathogen Discovery, Respiratory Viruses Branch, Division of Viral Diseases, Centers for Disease Control and Prevention | Jing Zhang et al |
| EPI_ISL_509714 | Belize Ministry of Health | Pathogen Discovery, Respiratory Viruses Branch, Division of Viral Diseases, Centers for Disease Control and Prevention | Jing Zhang et al |
| EPI_ISL_463902 | Shaoxing Center for Disease Control and Prevention | Department of Pathology and Laboratory Medicine, University of California Los Angeles | Jinkun Chen et al |
| EPI_ISL_1346626 | Center of Excellence in Clinical Virology | Research Unit of Systems Microbiology | Jiratchaya Puenpa et al |
| EPI_ISL_420412 | KU Leuven, Clinical and Epidemiological Virology | KU Leuven, Clinical and Epidemiological Virology | Joan Marti-Carreras et al |
| EPI_ISL_960776 | Germano de sousa | Instituto Gulbenkian de Ciencia | João Costa et al |
| EPI_ISL_491234 | Instituto Gulbenkian de Ciência | Instituto Gulbenkian de Ciência | Joao Sobral et al |
| EPI_ISL_1396423 | Centro de Tecnología en Salud Pública de la Universidad Nacional de Rosario | Laboratorio Mixto de Biotecnología Acuática (LMBA) on behalf of 'Proyecto Argentino Interinstitucional de genomica de SARS-CoV-2' (PAIS Consortium) | Joaquín Ezpeleta et al |
| EPI_ISL_1396459 | Centro de Tecnología en Salud Pública de la Universidad Nacional de Rosario | Laboratorio Mixto de Biotecnología Acuática (LMBA) on behalf of 'Proyecto Argentino Interinstitucional de genomica de SARS-CoV-2' (PAIS Consortium) | Joaquín Ezpeleta et al |
| EPI_ISL_1396460 | Centro de Tecnología en Salud Pública de la Universidad Nacional de Rosario | Laboratorio Mixto de Biotecnología Acuática (LMBA) on behalf of 'Proyecto Argentino Interinstitucional de genomica de SARS-CoV-2' (PAIS Consortium) | Joaquín Ezpeleta et al |
| EPI_ISL_648209 | INBIRS-UBA | Laboratorio Mixto de Biotecnología Acuática (LMBA) | Joaquín Ezpeleta et al |
| EPI_ISL_648214 | INBIRS-UBA | Laboratorio Mixto de Biotecnología Acuática (LMBA) | Joaquín Ezpeleta et al |
| EPI_ISL_424627 | Instituto Nacional de Enfermedades Respiratorias | Instituto Nacional de Enfermedades Respiratorias | Joel Armando Vázquez Pérez et al |
| EPI_ISL_695611 | AZ SPHL, Arizona Department of Health Services | TGen North | Jolene Bowers et al |
| EPI_ISL_1289905 | Department of Public Health Microbiology Ljubljana, National Laboratory for Health, Environment and Food | Department of Public Health Microbiology Ljubljana, National Laboratory for Health, Environment and Food | José Gonçalves et al |
| EPI_ISL_1289911 | Department of Public Health Microbiology Ljubljana, National Laboratory for Health, Environment and Food | Department of Public Health Microbiology Ljubljana, National Laboratory for Health, Environment and Food | José Gonçalves et al |
| EPI_ISL_1289912 | Department of Public Health Microbiology Ljubljana, National Laboratory for Health, Environment and Food | Department of Public Health Microbiology Ljubljana, National Laboratory for Health, Environment and Food | José Gonçalves et al |
| EPI_ISL_1289913 | Department of Public Health Microbiology Ljubljana, National Laboratory for Health, Environment and Food | Department of Public Health Microbiology Ljubljana, National Laboratory for Health, Environment and Food | José Gonçalves et al |
| EPI_ISL_1289915 | Department of Public Health Microbiology Ljubljana, National Laboratory for Health, Environment and Food | Department of Public Health Microbiology Ljubljana, National Laboratory for Health, Environment and Food | José Gonçalves et al |
| EPI_ISL_1289916 | Department of Public Health Microbiology Ljubljana, National Laboratory for Health, Environment and Food | Department of Public Health Microbiology Ljubljana, National Laboratory for Health, Environment and Food | José Gonçalves et al |

| | | | |
|---|---|---|---|
| EPI_ISL_1289917 | Department of Public Health Microbiology Ljubljana, National Laboratory for Health, Environment and Food | Department of Public Health Microbiology Ljubljana, National Laboratory for Health, Environment and Food | José Gonçalves et al |
| EPI_ISL_1289919 | Department of Public Health Microbiology Ljubljana, National Laboratory for Health, Environment and Food | Department of Public Health Microbiology Ljubljana, National Laboratory for Health, Environment and Food | José Gonçalves et al |
| EPI_ISL_1289925 | Department of Public Health Microbiology Ljubljana, National Laboratory for Health, Environment and Food | Department of Public Health Microbiology Ljubljana, National Laboratory for Health, Environment and Food | José Gonçalves et al |
| EPI_ISL_1289927 | Department of Public Health Microbiology Ljubljana, National Laboratory for Health, Environment and Food | Department of Public Health Microbiology Ljubljana, National Laboratory for Health, Environment and Food | José Gonçalves et al |
| EPI_ISL_1289875 | Department of Public Health Microbiology Ljubljana, National Laboratory for Health, Environment and Food | Department of Public Health Microbiology Ljubljana, National Laboratory for Health, Environment and Food | José Gonçalves et al |
| EPI_ISL_1289876 | Department of Public Health Microbiology Ljubljana, National Laboratory for Health, Environment and Food | Department of Public Health Microbiology Ljubljana, National Laboratory for Health, Environment and Food | José Gonçalves et al |
| EPI_ISL_1289892 | Department of Public Health Microbiology Ljubljana, National Laboratory for Health, Environment and Food | Department of Public Health Microbiology Ljubljana, National Laboratory for Health, Environment and Food | José Gonçalves et al |
| EPI_ISL_541038 | Hospital Clínico Universitario de Santiago de Compostela | SeqCOVID-SPAIN consortium/Institute of Biomedicine of Valencia, IBV-CSIC | José Javier Costa Alcalde et al |
| EPI_ISL_541066 | Hospital Clínico Universitario de Santiago de Compostela | SeqCOVID-SPAIN consortium/Institute of Biomedicine of Valencia, IBV-CSIC | José Javier Costa Alcalde et al |
| EPI_ISL_476161 | Laboratório de Patologia Clínica - UNICAMP | Laboratório de Estudos de Vírus Emergentes - UNICAMP | José Luiz Proença-Modena et al |
| EPI_ISL_1620750 | National Virus Reference Laboratory | Irish Coronavirus Sequencing Consortium - Teagasc Moorepark | Jose Sanchez-Morgado et al |
| EPI_ISL_1620748 | National Virus Reference Laboratory | Irish Coronavirus Sequencing Consortium - Teagasc Moorepark | Jose Sanchez-Morgado et al |
| EPI_ISL_1620704 | National Virus Reference Laboratory | Irish Coronavirus Sequencing Consortium - Teagasc Moorepark | Jose Sanchez-Morgado et al |
| EPI_ISL_1620740 | National Virus Reference Laboratory | Irish Coronavirus Sequencing Consortium - Teagasc Moorepark | Jose Sanchez-Morgado et al |
| EPI_ISL_1620712 | National Virus Reference Laboratory | Irish Coronavirus Sequencing Consortium - Teagasc Moorepark | Jose Sanchez-Morgado et al |
| EPI_ISL_1378833 | Instituto de Medicina Tropical & Salud Global Universidad Iberoamericana | Grubaugh Lab - Yale School of Public Health | Joseph Fauver et al |
| EPI_ISL_1378834 | Instituto de Medicina Tropical & Salud Global Universidad Iberoamericana | Grubaugh Lab - Yale School of Public Health | Joseph Fauver et al |
| EPI_ISL_1378837 | Instituto de Medicina Tropical & Salud Global Universidad Iberoamericana | Grubaugh Lab - Yale School of Public Health | Joseph Fauver et al |
| EPI_ISL_1378839 | Instituto de Medicina Tropical & Salud Global Universidad Iberoamericana | Grubaugh Lab - Yale School of Public Health | Joseph Fauver et al |
| EPI_ISL_1378842 | Instituto de Medicina Tropical & Salud Global Universidad Iberoamericana | Grubaugh Lab - Yale School of Public Health | Joseph Fauver et al |
| EPI_ISL_1378844 | Instituto de Medicina Tropical & Salud Global Universidad Iberoamericana | Grubaugh Lab - Yale School of Public Health | Joseph Fauver et al |
| EPI_ISL_729754 | Connecticut Department of Health | Grubaugh Lab - Yale School of Public Health | Joseph Fauver et al |
| EPI_ISL_729744 | Connecticut Department of Health | Grubaugh Lab - Yale School of Public Health | Joseph Fauver et al |
| EPI_ISL_729748 | Connecticut Department of Health | Grubaugh Lab - Yale School of Public Health | Joseph Fauver et al |
| EPI_ISL_422397 | NMIMR, Department of Virology | WACCBIP, University of Ghana | Joyce M. Ngoi et al |
| EPI_ISL_422402 | NMIMR, Department of Virology | WACCBIP, University of Ghana | Joyce M. Ngoi et al |
| EPI_ISL_422403 | NMIMR, Department of Virology | WACCBIP, University of Ghana | Joyce M. Ngoi et al |

| | | | |
|---|---|---|---|
| EPI_ISL_1120498 | Dutch COVID-19 response team | Medical Microbiology, Maastricht University Medical Centre | Jozef Dingemans* et al |
| EPI_ISL_965656 | Dutch COVID-19 response team | Medical Microbiology, Maastricht University Medical Centre | Jozef Dingemans* et al |
| EPI_ISL_1120515 | Dutch COVID-19 response team | Medical Microbiology, Maastricht University Medical Centre | Jozef Dingemans* et al |
| EPI_ISL_965567 | Dutch COVID-19 response team | Medical Microbiology, Maastricht University Medical Centre | Jozef Dingemans* et al |
| EPI_ISL_447767 | Instituto Nacional de Salud, Bogotá, Colombia | Grupo de Investigaciones Microbiológicas-UR (GIMUR), Departamento de Biología, Facultad de Ciencias Naturales, Universidad del Rosario, Bogotá, Colombia Instituto Nacional de Salud, Bogotá, Colombia Icahn School of Medicine at Mount Sinai, New York, USA | Juan David Ramírez et al |
| EPI_ISL_447734 | Grupo de Investigaciones Microbiológicas-UR (GIMUR), Departamento de Biología, Facultad de Ciencias Naturales, Universidad del Rosario, Bogotá, Colombia | Grupo de Investigaciones Microbiológicas-UR (GIMUR), Departamento de Biología, Facultad de Ciencias Naturales, Universidad del Rosario, Bogotá, Colombia Instituto Nacional de Salud, Bogotá, Colombia Icahn School of Medicine at Mount Sinai, New York, USA | Juan David Ramírez et al |
| EPI_ISL_447751 | Grupo de Investigaciones Microbiológicas-UR (GIMUR), Departamento de Biología, Facultad de Ciencias Naturales, Universidad del Rosario, Bogotá, Colombia | Grupo de Investigaciones Microbiológicas-UR (GIMUR), Departamento de Biología, Facultad de Ciencias Naturales, Universidad del Rosario, Bogotá, Colombia Instituto Nacional de Salud, Bogotá, Colombia Icahn School of Medicine at Mount Sinai, New York, USA | Juan David Ramírez et al |
| EPI_ISL_1040924 | Grupo de Investigación en Enfermedades Tropicales del Ejército (GINETEJ), Laboratorio de Referencia e Investigación, Dirección de Sanidad Ejército, Bogotá, Colombia | Centro de Investigaciones en Microbiología y Biotecnología-UR (CIMBIUR), Facultad de Ciencias Naturales, Universidad del Rosario, Bogotá, Colombia | Juan David Ramírez et al |
| EPI_ISL_447757 | Instituto Nacional de Salud, Bogotá, Colombia | Grupo de Investigaciones Microbiológicas-UR (GIMUR), Departamento de Biología, Facultad de Ciencias Naturales, Universidad del Rosario, Bogotá, Colombia Instituto Nacional de Salud, Bogotá, Colombia Icahn School of Medicine at Mount Sinai, New York, USA | Juan David Ramírez et al |
| EPI_ISL_447784 | Instituto Nacional de Salud, Bogotá, Colombia | Grupo de Investigaciones Microbiológicas-UR (GIMUR), Departamento de Biología, Facultad de Ciencias Naturales, Universidad del Rosario, Bogotá, Colombia Instituto Nacional de Salud, Bogotá, Colombia Icahn School of Medicine at Mount Sinai, New York, USA | Juan David Ramírez et al |
| EPI_ISL_477016 | Institute of Microbiology, Universidad San Francisco de Quito | Institute of Microbiology, Universidad San Francisco de Quito | Juan José Guadalupe et al |
| EPI_ISL_516648 | Institute of Microbiology, Universidad San Francisco de Quito | Institute of Microbiology, Universidad San Francisco de Quito | Juan José Guadalupe et al |
| EPI_ISL_516649 | Institute of Microbiology, Universidad San Francisco de Quito | Institute of Microbiology, Universidad San Francisco de Quito | Juan José Guadalupe et al |
| EPI_ISL_525430 | Institute of Microbiology, Universidad San Francisco de Quito | Institute of Microbiology, Universidad San Francisco de Quito | Juan José Guadalupe et al |
| EPI_ISL_525432 | Institute of Microbiology, Universidad San Francisco de Quito | Institute of Microbiology, Universidad San Francisco de Quito | Juan José Guadalupe et al |
| EPI_ISL_525433 | Institute of Microbiology, Universidad San Francisco de Quito | Institute of Microbiology, Universidad San Francisco de Quito | Juan José Guadalupe et al |
| EPI_ISL_525436 | Institute of Microbiology, Universidad San Francisco de Quito | Institute of Microbiology, Universidad San Francisco de Quito | Juan José Guadalupe et al |
| EPI_ISL_513514 | Programa de Oncovirologia, Instituto Nacional de Câncer | Programa de Oncovirologia, Instituto Nacional de Câncer | Juliana D. Siqueira et al |
| EPI_ISL_513519 | Programa de Oncovirologia, Instituto Nacional de Câncer | Programa de Oncovirologia, Instituto Nacional de Câncer | Juliana D. Siqueira et al |
| EPI_ISL_513532 | Programa de Oncovirologia, Instituto Nacional de Câncer | Programa de Oncovirologia, Instituto Nacional de Câncer | Juliana D. Siqueira et al |
| EPI_ISL_513557 | Programa de Oncovirologia, Instituto Nacional de Câncer | Programa de Oncovirologia, Instituto Nacional de Câncer | Juliana D. Siqueira et al |
| EPI_ISL_513566 | Programa de Oncovirologia, Instituto Nacional de Câncer | Programa de Oncovirologia, Instituto Nacional de Câncer | Juliana D. Siqueira et al |

| | | | |
|---|---|---|---|
| EPI_ISL_513573 | Programa de Oncovirologia, Instituto Nacional de Câncer | Programa de Oncovirologia, Instituto Nacional de Câncer | Juliana D. Siqueira et al |
| EPI_ISL_513577 | Programa de Oncovirologia, Instituto Nacional de Câncer | Programa de Oncovirologia, Instituto Nacional de Câncer | Juliana D. Siqueira et al |
| EPI_ISL_513578 | Programa de Oncovirologia, Instituto Nacional de Câncer | Programa de Oncovirologia, Instituto Nacional de Câncer | Juliana D. Siqueira et al |
| EPI_ISL_516783 | Center for Laboratory Control of Infectious Diseases, Korea Centers for Diseases Control and Prevention | Center for Laboratory Control of Infectious Diseases, Korea Centers for Diseases Control and Prevention | Junyoung Kim et al |
| EPI_ISL_522491 | Center for Laboratory Control of Infectious Diseases, Korea Centers for Diseases Control and Prevention | Center for Laboratory Control of Infectious Diseases, Korea Centers for Diseases Control and Prevention | Junyoung Kim et al |
| EPI_ISL_522502 | Center for Laboratory Control of Infectious Diseases, Korea Centers for Diseases Control and Prevention | Center for Laboratory Control of Infectious Diseases, Korea Centers for Diseases Control and Prevention | Junyoung Kim et al |
| EPI_ISL_560391 | Vilnius University Hospital Santaros Klinikos, Vilnius University | Institute of Biotechnology, Life Sciences Center, Vilnius University and Thermo Fisher Scientific | Justinas Slikas et al |
| EPI_ISL_560393 | Vilnius University Hospital Santaros Klinikos, Vilnius University | Institute of Biotechnology, Life Sciences Center, Vilnius University and Thermo Fisher Scientific | Justinas Slikas et al |
| EPI_ISL_560395 | Vilnius University Hospital Santaros Klinikos, Vilnius University | Institute of Biotechnology, Life Sciences Center, Vilnius University and Thermo Fisher Scientific | Justinas Slikas et al |
| EPI_ISL_560396 | Vilnius University Hospital Santaros Klinikos, Vilnius University | Institute of Biotechnology, Life Sciences Center, Vilnius University and Thermo Fisher Scientific | Justinas Slikas et al |
| EPI_ISL_560398 | Vilnius University Hospital Santaros Klinikos, Vilnius University | Institute of Biotechnology, Life Sciences Center, Vilnius University and Thermo Fisher Scientific | Justinas Slikas et al |
| EPI_ISL_560399 | Vilnius University Hospital Santaros Klinikos, Vilnius University | Institute of Biotechnology, Life Sciences Center, Vilnius University and Thermo Fisher Scientific | Justinas Slikas et al |
| EPI_ISL_560405 | Vilnius University Hospital Santaros Klinikos, Vilnius University | Institute of Biotechnology, Life Sciences Center, Vilnius University and Thermo Fisher Scientific | Justinas Slikas et al |
| EPI_ISL_1014680 | National Influenza Center, Virology Department | National Influenza Center | K Sadeghi et al |
| EPI_ISL_1014685 | National Influenza Center, Virology Department | National Influenza Center | K Sadeghi et al |
| EPI_ISL_862079 | National Influenza Center, Virology Department | National Influenza Center | K Sadeghi et al |
| EPI_ISL_959279 | National Influenza Center, Virology Department | National Influenza Center | K Sadeghi et al |
| EPI_ISL_779169 | Laboratorio de Infectología, Servicio de Infectología, Hospital Universitario Dr. José Eleuterio González - Universidad Autónoma de Nuevo León | Laboratorio de Infectología Molecular, Departamento de Bioquímica y Medicina Molecular,Facultad de Medicina - Universidad Autónoma de Nuevo León | Kame A. Galán-Huerta et al |
| EPI_ISL_779176 | Laboratorio de Infectología, Servicio de Infectología, Hospital Universitario Dr. José Eleuterio González - Universidad Autónoma de Nuevo León | Laboratorio de Infectología Molecular, Departamento de Bioquímica y Medicina Molecular,Facultad de Medicina - Universidad Autónoma de Nuevo León | Kame A. Galán-Huerta et al |
| EPI_ISL_779188 | Laboratorio Estatal de Salud Pública de Nuevo León | Laboratorio de Infectología Molecular, Departamento de Bioquímica y Medicina Molecular,Facultad de Medicina - Universidad Autónoma de Nuevo León | Kame A. Galán-Huerta et al |
| EPI_ISL_979331 | Laboratorio Estatal de Salud Pública de Nuevo León | Laboratorio de Infectología Molecular, Departamento de Bioquímica y Medicina Molecular,Facultad de Medicina - Universidad Autónoma de Nuevo León | Kame A. Galán-Huerta et al |
| EPI_ISL_961794 | Laboratorio de Infectología, Servicio de Infectología, Hospital Universitario Dr. José Eleuterio González - Universidad Autónoma de Nuevo León | Laboratorio de Infectología Molecular, Departamento de Bioquímica y Medicina Molecular,Facultad de Medicina - Universidad Autónoma de Nuevo León | Kame A. Galán-Huerta et al |

| | | | |
|---|---|---|---|
| EPI_ISL_961791 | Laboratorio de Infectología, Servicio de Infectología, Hospital Universitario Dr. José Eleuterio González - Universidad Autónoma de Nuevo León | Laboratorio de Infectología Molecular, Departamento de Bioquímica y Medicina Molecular,Facultad de Medicina - Universidad Autónoma de Nuevo León | Kame A. Galán-Huerta et al |
| EPI_ISL_961793 | Laboratorio de Infectología, Servicio de Infectología, Hospital Universitario Dr. José Eleuterio González - Universidad Autónoma de Nuevo León | Laboratorio de Infectología Molecular, Departamento de Bioquímica y Medicina Molecular,Facultad de Medicina - Universidad Autónoma de Nuevo León | Kame A. Galán-Huerta et al |
| EPI_ISL_961775 | Laboratorio de Infectología, Servicio de Infectología, Hospital Universitario Dr. José Eleuterio González - Universidad Autónoma de Nuevo León | Laboratorio de Infectología Molecular, Departamento de Bioquímica y Medicina Molecular,Facultad de Medicina - Universidad Autónoma de Nuevo León | Kame A. Galán-Huerta et al |
| EPI_ISL_961780 | Laboratorio de Infectología, Servicio de Infectología, Hospital Universitario Dr. José Eleuterio González - Universidad Autónoma de Nuevo León | Laboratorio de Infectología Molecular, Departamento de Bioquímica y Medicina Molecular,Facultad de Medicina - Universidad Autónoma de Nuevo León | Kame A. Galán-Huerta et al |
| EPI_ISL_961773 | Laboratorio de Infectología, Servicio de Infectología, Hospital Universitario Dr. José Eleuterio González - Universidad Autónoma de Nuevo León | Laboratorio de Infectología Molecular, Departamento de Bioquímica y Medicina Molecular,Facultad de Medicina - Universidad Autónoma de Nuevo León | Kame A. Galán-Huerta et al |
| EPI_ISL_961766 | Laboratorio de Infectología, Servicio de Infectología, Hospital Universitario Dr. José Eleuterio González - Universidad Autónoma de Nuevo León | Laboratorio de Infectología Molecular, Departamento de Bioquímica y Medicina Molecular,Facultad de Medicina - Universidad Autónoma de Nuevo León | Kame A. Galán-Huerta et al |
| EPI_ISL_1091266 | Laboratorio PGM | Laboratorio de Infectología Molecular, Departamento de Bioquímica y Medicina Molecular,Facultad de Medicina - Universidad Autónoma de Nuevo León | Kame A. Galán-Huerta et al |
| EPI_ISL_1091259 | Laboratorio PGM | Laboratorio de Infectología Molecular, Departamento de Bioquímica y Medicina Molecular,Facultad de Medicina - Universidad Autónoma de Nuevo León | Kame A. Galán-Huerta et al |
| EPI_ISL_728561 | Institute for Urban Disease Control and Prevention | COVID-19 Network Investigations (CONI) Alliance | Kamolthip Atsawawaranunt et al |
| EPI_ISL_882773 | Institute for Urban Disease Control and Prevention | COVID-19 Network Investigations (CONI) Alliance | Kamolthip Atsawawaranunt et al |
| EPI_ISL_733552 | Institute for Urban Disease Control and Prevention | COVID-19 Network Investigations (CONI) Alliance | Kamolthip Atsawawaranunt et al |
| EPI_ISL_812941 | Institute for Urban Disease Control and Prevention | COVID-19 Network Investigations (CONI) Alliance | Kamolthip Atsawawaranunt et al |
| EPI_ISL_561338 | Civil Hospital, Panchkula | CSIR-Institute of Microbial Technology | Kanika Bansal et al |
| EPI_ISL_493137 | Center for Research and Innovation, Faculty of Medical Technology, Mahidol University | Center for Research and Innovation, Faculty of Medical Technology, Mahidol University | Kantima Sangsiriwut et al |
| EPI_ISL_437300 | Diagnostic- and Research Institute of Pathology, Medical University of Graz | Diagnostic- and Research Institute of Pathology, Medical University of Graz | Karl Kashofer et al |
| EPI_ISL_1169051 | National Institute of Health Research and Development | National Institute of Health Research and Development | Kartika Dewi Puspa et al |
| EPI_ISL_1257844 | RSUP Fatmawati | National Institute of Health Research and Development | Kartika Dewi Puspa et al |
| EPI_ISL_437895 | Laboratory of Microbiology, Medical School, National and Kapodistrian University of Athens | Laboratory of Biology, Department of Medicine, Democritus University of Thrace | Kassela K. et al |
| EPI_ISL_437908 | Laboratory of Microbiology, Medical School, National and Kapodistrian University of Athens | Laboratory of Biology, Department of Medicine, Democritus University of Thrace | Kassela K. et al |
| EPI_ISL_418809 | University of Wisconsin - Madison: Influenza Research Institute | University of Wisconsin Madison, AIDS Vaccine Research Laboratories | Katarina Braun et al |
| EPI_ISL_960308 | NLZOH, Laboratory for Virology | NLZOH, Laboratory for Virology | Katarina Prosenc (Laboratory for Virology) e |
| EPI_ISL_960306 | NLZOH, Laboratory for Virology | NLZOH, Laboratory for Virology | Katarina Prosenc (Laboratory for Virology) e |

et al
et al

| | | | |
|---|---|---|---|
| EPI_ISL_960304 | NLZOH, Laboratory for Virology | NLZOH, Laboratory for Virology | Katarina Prosenc (Laboratory for Virology) e |
| EPI_ISL_455441 | 1. ViroGenetics - BSL3 Laboratory of Virology, Małopolska Centre of Biotechnology, Jagiellonian University; 2. II Department of Internal Medicine, Faculty of Medicine, Jagiellonian University Medical College; 3. Narodowy Instytut Zdrowia Publicznego – Państwowy Zakład Higieny (NIZP-PZH) | 1. ViroGenetics - BSL3 Laboratory of Virology, Małopolska Centre of Biotechnology, Jagiellonian University; 2. II Department of Internal Medicine, Faculty of Medicine, Jagiellonian University Medical College; 3. Narodowy Instytut Zdrowia Publicznego – Państwowy Zakład Higieny (NIZP-PZH). | Katarzyna Pancer et al |
| EPI_ISL_455443 | 1. ViroGenetics - BSL3 Laboratory of Virology, Małopolska Centre of Biotechnology, Jagiellonian University; 2. II Department of Internal Medicine, Faculty of Medicine, Jagiellonian University Medical College; 3. Narodowy Instytut Zdrowia Publicznego – Państwowy Zakład Higieny (NIZP-PZH) | 1. ViroGenetics - BSL3 Laboratory of Virology, Małopolska Centre of Biotechnology, Jagiellonian University; 2. II Department of Internal Medicine, Faculty of Medicine, Jagiellonian University Medical College; 3. Narodowy Instytut Zdrowia Publicznego – Państwowy Zakład Higieny (NIZP-PZH). | Katarzyna Pancer et al |
| EPI_ISL_455448 | 1. ViroGenetics - BSL3 Laboratory of Virology, Małopolska Centre of Biotechnology, Jagiellonian University; 2. II Department of Internal Medicine, Faculty of Medicine, Jagiellonian University Medical College; 3. Narodowy Instytut Zdrowia Publicznego – Państwowy Zakład Higieny (NIZP-PZH) | 1. ViroGenetics - BSL3 Laboratory of Virology, Małopolska Centre of Biotechnology, Jagiellonian University; 2. II Department of Internal Medicine, Faculty of Medicine, Jagiellonian University Medical College; 3. Narodowy Instytut Zdrowia Publicznego – Państwowy Zakład Higieny (NIZP-PZH). | Katarzyna Pancer et al |
| EPI_ISL_848146 | National Virus Reference Laboratory | Irish Coronavirus Sequencing Consortium - National University of Ireland Galway | Kate Reddington et al |
| EPI_ISL_848084 | National Virus Reference Laboratory | Irish Coronavirus Sequencing Consortium - National University of Ireland Galway | Kate Reddington et al |
| EPI_ISL_848101 | National Virus Reference Laboratory | Irish Coronavirus Sequencing Consortium - National University of Ireland Galway | Kate Reddington et al |
| EPI_ISL_848113 | National Virus Reference Laboratory | Irish Coronavirus Sequencing Consortium - National University of Ireland Galway | Kate Reddington et al |
| EPI_ISL_1137618 | Dirección de Sanidad Policia Nacional | Instituto Nacional de Salud- Dirección de Investigación en Salud Pública | Katherine Laiton-Donato et al |
| EPI_ISL_1137619 | Laboratorio de Salud Publica de Cauca | Instituto Nacional de Salud- Dirección de Investigación en Salud Pública | Katherine Laiton-Donato et al |
| EPI_ISL_1137615 | Laboratorio de Salud Publica de Cauca | Instituto Nacional de Salud- Dirección de Investigación en Salud Pública | Katherine Laiton-Donato et al |
| EPI_ISL_1303374 | CLINICA COLSANITAS CENTRAL DE REFERENCIA | Instituto Nacional de Salud- Dirección de Investigación en Salud Pública | Katherine Laiton-Donato et al |
| EPI_ISL_1303373 | Dirección de Sanidad Policia Nacional | Instituto Nacional de Salud- Dirección de Investigación en Salud Pública | Katherine Laiton-Donato et al |
| EPI_ISL_1303372 | Dirección de Sanidad Policia Nacional | Instituto Nacional de Salud- Dirección de Investigación en Salud Pública | Katherine Laiton-Donato et al |
| EPI_ISL_1303371 | Dirección de Sanidad Policia Nacional | Instituto Nacional de Salud- Dirección de Investigación en Salud Pública | Katherine Laiton-Donato et al |
| EPI_ISL_1303366 | Instituto Nacional de Salud- Dirección de Redes de Laboratorios de Salud Pública | Instituto Nacional de Salud- Dirección de Investigación en Salud Pública | Katherine Laiton-Donato et al |
| EPI_ISL_1303367 | Instituto Nacional de Salud- Dirección de Redes de Laboratorios de Salud Pública | Instituto Nacional de Salud- Dirección de Investigación en Salud Pública | Katherine Laiton-Donato et al |
| EPI_ISL_1303377 | Instituto Nacional de Salud- Dirección de Redes de Laboratorios de Salud Pública | Instituto Nacional de Salud- Dirección de Investigación en Salud Pública | Katherine Laiton-Donato et al |
| EPI_ISL_1632511 | LDSP Amazonas | Instituto Nacional de Salud- Dirección de Investigación en Salud Pública | Katherine Laiton-Donato et al |
| EPI_ISL_1652041 | LDSP BOGOTA | Instituto Nacional de Salud- Dirección de Investigación en Salud Pública | Katherine Laiton-Donato et al |
| EPI_ISL_956297 | Instituto Nacional de Salud- Dirección de Redes de Laboratorios de Salud Pública | Instituto Nacional de Salud- Dirección de Investigación en Salud Pública | Katherine Laiton-Donato et al |

et al

| | | | |
|---|---|---|---|
| EPI_ISL_956288 | Instituto Nacional de Salud- Dirección de Redes de Laboratorios de Salud Pública | Instituto Nacional de Salud- Dirección de Investigación en Salud Pública | Katherine Laiton-Donato et al |
| EPI_ISL_956289 | Instituto Nacional de Salud- Dirección de Redes de Laboratorios de Salud Pública | Instituto Nacional de Salud- Dirección de Investigación en Salud Pública | Katherine Laiton-Donato et al |
| EPI_ISL_956291 | Instituto Nacional de Salud- Dirección de Redes de Laboratorios de Salud Pública | Instituto Nacional de Salud- Dirección de Investigación en Salud Pública | Katherine Laiton-Donato et al |
| EPI_ISL_956292 | Instituto Nacional de Salud- Dirección de Redes de Laboratorios de Salud Pública | Instituto Nacional de Salud- Dirección de Investigación en Salud Pública | Katherine Laiton-Donato et al |
| EPI_ISL_956293 | Instituto Nacional de Salud- Dirección de Redes de Laboratorios de Salud Pública | Instituto Nacional de Salud- Dirección de Investigación en Salud Pública | Katherine Laiton-Donato et al |
| EPI_ISL_956295 | Instituto Nacional de Salud- Dirección de Redes de Laboratorios de Salud Pública | Instituto Nacional de Salud- Dirección de Investigación en Salud Pública | Katherine Laiton-Donato et al |
| EPI_ISL_653756 | Instituto Nacional de Salud, Bogotá, Colombia | Instituto Nacional de Salud, Bogotá, Colombia | Katherine Laiton-Donato et al |
| EPI_ISL_739680 | Instituto Nacional de Salud, Bogotá, Colombia | Instituto Nacional de Salud, Bogotá, Colombia | Katherine Laiton-Donato et al |
| EPI_ISL_739675 | Instituto Nacional de Salud, Bogotá, Colombia | Instituto Nacional de Salud, Bogotá, Colombia | Katherine Laiton-Donato et al |
| EPI_ISL_739674 | Instituto Nacional de Salud, Bogotá, Colombia | Instituto Nacional de Salud, Bogotá, Colombia | Katherine Laiton-Donato et al |
| EPI_ISL_739677 | Instituto Nacional de Salud, Bogotá, Colombia | Instituto Nacional de Salud, Bogotá, Colombia | Katherine Laiton-Donato et al |
| EPI_ISL_739676 | Instituto Nacional de Salud, Bogotá, Colombia | Instituto Nacional de Salud, Bogotá, Colombia | Katherine Laiton-Donato et al |
| EPI_ISL_653758 | Instituto Nacional de Salud, Bogotá, Colombia | Instituto Nacional de Salud, Bogotá, Colombia | Katherine Laiton-Donato et al |
| EPI_ISL_1424059 | Instituto Nacional de Salud- Dirección de Investigación en Salud Pública | Instituto Nacional de Salud- Dirección de Investigación en Salud Pública | Katherine Laiton-Donato et al |
| EPI_ISL_1424060 | Instituto Nacional de Salud- Dirección de Investigación en Salud Pública | Instituto Nacional de Salud- Dirección de Investigación en Salud Pública | Katherine Laiton-Donato et al |
| EPI_ISL_1424061 | Instituto Nacional de Salud- Dirección de Investigación en Salud Pública | Instituto Nacional de Salud- Dirección de Investigación en Salud Pública | Katherine Laiton-Donato et al |
| EPI_ISL_956300 | Laboratorio de salud publica Arauca | Instituto Nacional de Salud- Dirección de Investigación en Salud Pública | Katherine Laiton-Donato et al |
| EPI_ISL_653761 | Instituto Nacional de Salud, Bogotá, Colombia | Instituto Nacional de Salud, Bogotá, Colombia | Katherine Laiton-Donato et al |
| EPI_ISL_653760 | Instituto Nacional de Salud, Bogotá, Colombia | Instituto Nacional de Salud, Bogotá, Colombia | Katherine Laiton-Donato et al |
| EPI_ISL_653762 | Instituto Nacional de Salud, Bogotá, Colombia | Instituto Nacional de Salud, Bogotá, Colombia | Katherine Laiton-Donato et al |
| EPI_ISL_1220050 | Laboratorio Clinico Clinica General del Norte | Instituto Nacional de Salud- Dirección de Investigación en Salud Pública | Katherine Laiton-Donato et al |
| EPI_ISL_845620 | compensar calle 63 | Instituto Nacional de Salud - Dirección de Investigación en Salud Pública | Katherine Laiton-Donato et al |
| EPI_ISL_845621 | Dirección de Sanidad Ejército | Instituto Nacional de Salud - Dirección de Investigación en Salud Pública | Katherine Laiton-Donato et al |
| EPI_ISL_1632531 | UNIVERSIDAD DEL SINU | Instituto Nacional de Salud- Dirección de Investigación en Salud Pública | Katherine Laiton-Donato et al |
| EPI_ISL_498158 | Instituto Nacional de Salud, Bogotá, Colombia | Instituto Nacional de Salud, Bogotá, Colombia | Katherine Laiton-Donato et al |
| EPI_ISL_456123 | Instituto Nacional de Salud - Unidad de Secuenciación y Análisis Genómico | Instituto Nacional de Salud, Universidad Cooperativa de Colombia, Instituto Alexander von Humboldt, Imperial College-London, London School of Hygiene & Tropical Medicine | Katherine Laiton-Donato et al |
| EPI_ISL_498168 | Instituto Nacional de Salud, Bogotá, Colombia | Instituto Nacional de Salud, Bogotá, Colombia | Katherine Laiton-Donato et al |

| | | | |
|---|---|---|---|
| EPI_ISL_498170 | Instituto Nacional de Salud, Bogotá, Colombia | Instituto Nacional de Salud, Bogotá, Colombia | Katherine Laiton-Donato et al |
| EPI_ISL_526969 | Instituto Nacional de Salud, Bogotá, Colombia | Instituto Nacional de Salud, Bogotá, Colombia | Katherine Laiton-Donato et al |
| EPI_ISL_526974 | Instituto Nacional de Salud, Bogotá, Colombia | Instituto Nacional de Salud, Bogotá, Colombia | Katherine Laiton-Donato et al |
| EPI_ISL_526971 | Instituto Nacional de Salud, Bogotá, Colombia | Instituto Nacional de Salud, Bogotá, Colombia | Katherine Laiton-Donato et al |
| EPI_ISL_653745 | Instituto Nacional de Salud, Bogotá, Colombia | Instituto Nacional de Salud, Bogotá, Colombia | Katherine Laiton-Donato et al |
| EPI_ISL_653746 | Instituto Nacional de Salud, Bogotá, Colombia | Instituto Nacional de Salud, Bogotá, Colombia | Katherine Laiton-Donato et al |
| EPI_ISL_653747 | Instituto Nacional de Salud, Bogotá, Colombia | Instituto Nacional de Salud, Bogotá, Colombia | Katherine Laiton-Donato et al |
| EPI_ISL_653754 | Instituto Nacional de Salud, Bogotá, Colombia | Instituto Nacional de Salud, Bogotá, Colombia | Katherine Laiton-Donato et al |
| EPI_ISL_739673 | Instituto Nacional de Salud, Bogotá, Colombia | Instituto Nacional de Salud, Bogotá, Colombia | Katherine Laiton-Donato et al |
| EPI_ISL_791086 | Instituto Nacional de Salud - Unidad de Secuenciación y Análisis Genómico | Instituto Nacional de Salud - Dirección de Investigación en Salud Pública | Katherine Laiton-Donato et al |
| EPI_ISL_791084 | Instituto Nacional de Salud - Unidad de Secuenciación y Análisis Genómico | Instituto Nacional de Salud - Dirección de Investigación en Salud Pública | Katherine Laiton-Donato et al |
| EPI_ISL_1092008 | Laboratorio de Salud Publica de Bogota | Instituto Nacional de Salud- Dirección de Investigación en Salud Pública | Katherine Laiton-Donato et al |
| EPI_ISL_1494944 | HOSPITAL DEPARTAMENTAL SAN VICENTE DE PAUL | Instituto Nacional de Salud- Dirección de Investigación en Salud Pública | Katherine Laiton-Donato et al |
| EPI_ISL_526959 | Instituto Nacional de Salud, Bogotá, Colombia | Instituto Nacional de Salud, Bogotá, Colombia | Katherine Laiton-Donato et al |
| EPI_ISL_526958 | Instituto Nacional de Salud, Bogotá, Colombia | Instituto Nacional de Salud, Bogotá, Colombia | Katherine Laiton-Donato et al |
| EPI_ISL_1582978 | E.S.E. HOSPITAL SAN JOSE DE MAICAO | Instituto Nacional de Salud- Dirección de Investigación en Salud Pública | Katherine Laiton-Donato et al |
| EPI_ISL_1582979 | E.S.E. HOSPITAL SAN JOSE DE MAICAO | Instituto Nacional de Salud- Dirección de Investigación en Salud Pública | Katherine Laiton-Donato et al |
| EPI_ISL_1582980 | E.S.E. HOSPITAL SAN JOSE DE MAICAO | Instituto Nacional de Salud- Dirección de Investigación en Salud Pública | Katherine Laiton-Donato et al |
| EPI_ISL_1582989 | Laboratorio de Biología Molecular – Universidad del Magdalena | Instituto Nacional de Salud- Dirección de Investigación en Salud Pública | Katherine Laiton-Donato et al |
| EPI_ISL_1582993 | Universidad del Atlántico Laboratorio de Investigación en Biología Molecular | Instituto Nacional de Salud- Dirección de Investigación en Salud Pública | Katherine Laiton-Donato et al |
| EPI_ISL_1582983 | HOSPITAL DEPARTAMENTAL DE VILLAVICENCIO | Instituto Nacional de Salud- Dirección de Investigación en Salud Pública | Katherine Laiton-Donato et al |
| EPI_ISL_498162 | Instituto Nacional de Salud, Bogotá, Colombia | Instituto Nacional de Salud, Bogotá, Colombia | Katherine Laiton-Donato et al |
| EPI_ISL_956306 | Laboratorio Gencore- Universidad de los Andes | Instituto Nacional de Salud- Dirección de Investigación en Salud Pública, Universidad de los Andes- Gencore | Katherine Laiton-Donato et al |
| EPI_ISL_1017707 | Hospital Universitario Hernando Moncaleano Perdomo | Instituto Nacional de Salud- Dirección de Investigación en Salud Pública | Katherine Laiton-Donato et al |
| EPI_ISL_456128 | Instituto Nacional de Salud - Unidad de Secuenciación y Análisis Genómico | Instituto Nacional de Salud, Universidad Cooperativa de Colombia, Instituto Alexander von Humboldt, Imperial College-London, London School of Hygiene & Tropical Medicine | Katherine Laiton-Donato et al |
| EPI_ISL_739663 | Instituto Nacional de Salud, Bogotá, Colombia | Instituto Nacional de Salud, Bogotá, Colombia | Katherine Laiton-Donato et al |
| EPI_ISL_1494950 | Laboratorio de Salud Publica de Santander | Instituto Nacional de Salud- Dirección de Investigación en Salud Pública | Katherine Laiton-Donato et al |
| EPI_ISL_1220065 | Investigaciones Biomédicas - Universidad de Sucre | Instituto Nacional de Salud- Dirección de Investigación en Salud Pública | Katherine Laiton-Donato et al |
| EPI_ISL_1494940 | Investigaciones Biomédicas - Universidad de Sucre | Instituto Nacional de Salud- Dirección de Investigación en Salud Pública | Katherine Laiton-Donato et al |

| | | | |
|---|---|---|---|
| EPI_ISL_653749 | Instituto Nacional de Salud, Bogotá, Colombia | Instituto Nacional de Salud, Bogotá, Colombia | Katherine Laiton-Donato et al |
| EPI_ISL_456152 | Instituto Nacional de Salud - Unidad de Secuenciación y Análisis Genómico | Instituto Nacional de Salud, Universidad Cooperativa de Colombia, Instituto Alexander von Humboldt, Imperial College-London, London School of Hygiene & Tropical Medicine | Katherine Laiton-Donato et al |
| EPI_ISL_707947 | Virology, Universitätsklinikum des Saarlandes | Epigenetics, Saarland University | Kathrin Kattler et al |
| EPI_ISL_707948 | Virology, Universitätsklinikum des Saarlandes | Epigenetics, Saarland University | Kathrin Kattler et al |
| EPI_ISL_708025 | Virology, Universitätsklinikum des Saarlandes | Epigenetics, Saarland University | Kathrin Kattler et al |
| EPI_ISL_860285 | Department of Medical Microbiology, St. Olavs hospital | Norwegian Institute of Public Health, Department of Virology | Kathrine Stene-Johansen et al |
| EPI_ISL_860287 | Department of Medical Microbiology, St. Olavs hospital | Norwegian Institute of Public Health, Department of Virology | Kathrine Stene-Johansen et al |
| EPI_ISL_1034356 | Department of Medical Microbiology, St. Olavs hospital | Norwegian Institute of Public Health, Department of Virology | Kathrine Stene-Johansen et al |
| EPI_ISL_1034357 | Department of Medical Microbiology, St. Olavs hospital | Norwegian Institute of Public Health, Department of Virology | Kathrine Stene-Johansen et al |
| EPI_ISL_1034361 | Department of Medical Microbiology, St. Olavs hospital | Norwegian Institute of Public Health, Department of Virology | Kathrine Stene-Johansen et al |
| EPI_ISL_417483 | Oslo University Hospital, Department of Medical Microbiology | Norwegian Institute of Public Health | Kathrine Stene-Johansen et al |
| EPI_ISL_417484 | Oslo University Hospital, Department of Medical Microbiology | Norwegian Institute of Public Health, Department of Virology | Kathrine Stene-Johansen et al |
| EPI_ISL_417490 | Oslo University Hospital, Department of Medical Microbiology | Norwegian Institute of Public Health, Department of Virology | Kathrine Stene-Johansen et al |
| EPI_ISL_420135 | Oslo University Hospital, Department of Medical Microbiology | Norwegian Institute of Public Health, Department of Virology | Kathrine Stene-Johansen et al |
| EPI_ISL_417488 | Oslo University Hospital, Department of Medical Microbiology | Norwegian Institute of Public Health, Department of Virology | Kathrine Stene-Johansen et al |
| EPI_ISL_417489 | Oslo University Hospital, Department of Medical Microbiology | Norwegian Institute of Public Health, Department of Virology | Kathrine Stene-Johansen et al |
| EPI_ISL_420134 | Akershus University Hospital, Department for Microbiology and Infectious Disease Control | Norwegian Institute of Public Health, Department of Virology | Kathrine Stene-Johansen et al |
| EPI_ISL_417486 | Hospital of Southern Norway - Kristiansand, Department of Medical Microbiology | Norwegian Institute of Public Health, Department of Virology | Kathrine Stene-Johansen et al |
| EPI_ISL_1118497 | Department of Medical Microbiology, St. Olavs hospital | Norwegian Institute of Public Health, Department of Virology | Kathrine Stene-Johansen et al |
| EPI_ISL_420136 | Akershus University Hospital, Department for Microbiology and Infectious Disease Control | Norwegian Institute of Public Health, Department of Virology | Kathrine Stene-Johansen et al |
| EPI_ISL_590915 | Hospital of Southern Norway - Kristiansand, Department of Medical Microbiology | Norwegian Institute of Public Health, Department of Virology | Kathrine Stene-Johansen et al |
| EPI_ISL_420148 | Ostfold Hospital Trust -Kalnes Centre for Laboratory Medicine Section for gene technology and infection serology | Norwegian Institute of Public Health, Department of Virology | Kathrine Stene-Johansen et al |
| EPI_ISL_420151 | Nordland Hospital - Bodo, Laboratory Department, Molecular Biology Unit | Norwegian Institute of Public Health, Department of Virology | Kathrine Stene-Johansen et al |
| EPI_ISL_449788 | Furst Medical Laboratory | Norwegian Institute of Public Health, Department of Virology | Kathrine Stene-Johansen et al |
| EPI_ISL_1317842 | Furst Medical Laboratory | Norwegian Institute of Public Health, Department of Virology | Kathrine Stene-Johansen et al |
| EPI_ISL_481210 | Ostfold Hospital Trust - Kalnes, Centre for Laboratory Medicine, Section for gene technology and infection serology | Norwegian Institute of Public Health, Department of Virology | Kathrine Stene-Johansen et al |
| EPI_ISL_481211 | Ostfold Hospital Trust - Kalnes, Centre for Laboratory Medicine, Section for gene technology and infection serology | Norwegian Institute of Public Health, Department of Virology | Kathrine Stene-Johansen et al |

| | | | |
|---|---|---|---|
| EPI_ISL_481212 | Ostfold Hospital Trust - Kalnes, Centre for Laboratory Medicine, Section for gene technology and infection serology | Norwegian Institute of Public Health, Department of Virology | Kathrine Stene-Johansen et al |
| EPI_ISL_481215 | Oslo University Hospital, Department of Medical Microbiology | Norwegian Institute of Public Health, Department of Virology | Kathrine Stene-Johansen et al |
| EPI_ISL_1251957 | Department of Medical Microbiology, St. Olavs hospital | Norwegian Institute of Public Health, Department of Virology | Kathrine Stene-Johansen et al |
| EPI_ISL_493355 | Oslo University Hospital, Department of Medical Microbiology | Norwegian Institute of Public Health, Department of Virology | Kathrine Stene-Johansen et al |
| EPI_ISL_493358 | Furst Medical Laboratory | Norwegian Institute of Public Health, Department of Virology | Kathrine Stene-Johansen et al |
| EPI_ISL_493369 | Furst Medical Laboratory | Norwegian Institute of Public Health, Department of Virology | Kathrine Stene-Johansen et al |
| EPI_ISL_493380 | Furst Medical Laboratory | Norwegian Institute of Public Health, Department of Virology | Kathrine Stene-Johansen et al |
| EPI_ISL_493382 | Medical Microbiology Unit, Department for Laboratory Medicine, Drammen Hospital, Vestre Viken Health Trust, | Norwegian Institute of Public Health, Department of Virology | Kathrine Stene-Johansen et al |
| EPI_ISL_549038 | Furst Medical Laboratory | Norwegian Institute of Public Health, Department of Virology | Kathrine Stene-Johansen et al |
| EPI_ISL_549068 | Furst Medical Laboratory | Norwegian Institute of Public Health, Department of Virology | Kathrine Stene-Johansen et al |
| EPI_ISL_549056 | Furst Medical Laboratory | Norwegian Institute of Public Health, Department of Virology | Kathrine Stene-Johansen et al |
| EPI_ISL_549057 | Furst Medical Laboratory | Norwegian Institute of Public Health, Department of Virology | Kathrine Stene-Johansen et al |
| EPI_ISL_549084 | Akershus University Hospital, Department for Microbiology and Infectious Disease Control | Norwegian Institute of Public Health, Department of Virology | Kathrine Stene-Johansen et al |
| EPI_ISL_1317969 | Furst Medical Laboratory | Norwegian Institute of Public Health, Department of Virology | Kathrine Stene-Johansen et al |
| EPI_ISL_1192300 | Department of Medical Microbiology, St. Olavs hospital | Norwegian Institute of Public Health, Department of Virology | Kathrine Stene-Johansen et al |
| EPI_ISL_1318016 | Furst Medical Laboratory | Norwegian Institute of Public Health, Department of Virology | Kathrine Stene-Johansen et al |
| EPI_ISL_1490489 | Furst Medical Laboratory | Norwegian Institute of Public Health, Department of Virology | Kathrine Stene-Johansen et al |
| EPI_ISL_635161 | Hospital of Southern Norway - Kristiansand, Department of Medical Microbiology | Norwegian Institute of Public Health, Department of Virology | Kathrine Stene-Johansen et al |
| EPI_ISL_668394 | Hospital of Southern Norway - Kristiansand, Department of Medical Microbiology | Norwegian Institute of Public Health, Department of Virology | Kathrine Stene-Johansen et al |
| EPI_ISL_1252548 | Furst Medical Laboratory | Norwegian Institute of Public Health, Department of Virology | Kathrine Stene-Johansen et al |
| EPI_ISL_708059 | Furst Medical Laboratory | Norwegian Institute of Public Health, Department of Virology | Kathrine Stene-Johansen et al |
| EPI_ISL_1547525 | Furst Medical Laboratory | Norwegian Institute of Public Health, Department of Virology | Kathrine Stene-Johansen et al |
| EPI_ISL_1579172 | Furst Medical Laboratory | Norwegian Institute of Public Health, Department of Virology | Kathrine Stene-Johansen et al |
| EPI_ISL_1547553 | Furst Medical Laboratory | Norwegian Institute of Public Health, Department of Virology | Kathrine Stene-Johansen et al |
| EPI_ISL_775398 | Nordland Hospital - Bodo, Laboratory Department, Molecular Biology Unit | Norwegian Institute of Public Health, Department of Virology | Kathrine Stene-Johansen et al |
| EPI_ISL_1547634 | Ostfold Hospital Trust - Kalnes, Centre for Laboratory Medicine, Section for gene technology and infection serology | Norwegian Institute of Public Health, Department of Virology | Kathrine Stene-Johansen et al |
| EPI_ISL_775528 | Akershus University Hospital, Department for Microbiology and Infectious Disease Control | Norwegian Institute of Public Health, Department of Virology | Kathrine Stene-Johansen et al |

| | | | |
|---|---|---|---|
| EPI_ISL_775529 | Akershus University Hospital, Department for Microbiology and Infectious Disease Control | Norwegian Institute of Public Health, Department of Virology | Kathrine Stene-Johansen et al |
| EPI_ISL_775530 | Akershus University Hospital, Department for Microbiology and Infectious Disease Control | Norwegian Institute of Public Health, Department of Virology | Kathrine Stene-Johansen et al |
| EPI_ISL_1251304 | Akershus University Hospital, Department for Microbiology and Infectious Disease Control | Norwegian Institute of Public Health, Department of Virology | Kathrine Stene-Johansen et al |
| EPI_ISL_796703 | Oslo University Hospital, Department of Medical Microbiology | Norwegian Institute of Public Health, Department of Virology | Kathrine Stene-Johansen et al |
| EPI_ISL_1251378 | University Hospital of Northern Norway, Department for Microbiology and Infectious Disease Control | Norwegian Institute of Public Health, Department of Virology | Kathrine Stene-Johansen et al |
| EPI_ISL_860233 | Norwegian Institute of Public Health, Department of Virology | Norwegian Institute of Public Health, Department of Virology | Kathrine Stene-Johansen et al |
| EPI_ISL_1599419 | Akershus University Hospital, Department for Microbiology and Infectious Disease Control | Norwegian Institute of Public Health, Department of Virology | Kathrine Stene-Johansen et al |
| EPI_ISL_794602 | Central Laboratories, Egyptian Ministry of Health and Population | Central Laboratories, Egyptian Ministry of Health and Population | Kayed et al |
| EPI_ISL_794600 | Central Laboratories, Egyptian Ministry of Health and Population | Central Laboratories, Egyptian Ministry of Health and Population | Kayed et al |
| EPI_ISL_1335899 | Department of Microbiology, Faculty of Medicine, Universitas Sumatera Utara | Faculty of Medicine, Universitas Sumatera Utara; Institute of Tropical Disease, Universitas Airlangga | Kazufumi Shimizu et al |
| EPI_ISL_770472 | National Health laboratory | Botswana Institute for Technology Research and Innovation | Kefentse Arnold Tumedi et al |
| EPI_ISL_770475 | National health Laboratory | Botswana Institute for Technology Research and Innovation | Kefentse Arnold Tumedi et al |
| EPI_ISL_770470 | National Health laboratory | Botswana Institute for Technology Research and Innovation | Kefentse Arnold Tumedi et al |
| EPI_ISL_560386 | National Health Laboratory | Botswana Institute for Technology Research and innovation | Kefentse Arnold Tumedi et al |
| EPI_ISL_560390 | National Health Laboratory | Botswana Institute for Technology Research and Innovation | Kefentse Arnold Tumedi et al |
| EPI_ISL_560385 | National Health Laboratory | Botswana Institute for Technology Research and Innovation | Kefentse Arnold Tumedi et al |
| EPI_ISL_560387 | National Health Laboratory | Botswana Institute for Technology Research and Innovation | Kefentse Arnold Tumedi et al |
| EPI_ISL_560389 | National Health Laboratory | Botswana Institute for Technology Research and Innovation | Kefentse Arnold Tumedi et al |
| EPI_ISL_560388 | National Health Laboratory | Botswana Institute for Technology Research and Innovation | Kefentse Arnold Tumedi et al |
| EPI_ISL_770474 | National Health laboratory | Botswana Institute for Technology Research and Innovation | Kefentse Arnold Tumedi et al |
| EPI_ISL_427389 | Department of Clinical Microbiology | GIGA Medical Genomics | Keith Durkin et al |
| EPI_ISL_455975 | Department of Clinical Microbiology | GIGA Medical Genomics | Keith Durkin et al |
| EPI_ISL_484708 | Department of Clinical Microbiology | GIGA Medical Genomics | Keith Durkin et al |
| EPI_ISL_540451 | University of Liège COVID-19 testing center | GIGA Medical Genomics | Keith Durkin et al |
| EPI_ISL_540466 | University of Liège COVID-19 testing center | GIGA Medical Genomics | Keith Durkin et al |
| EPI_ISL_581596 | Department of Clinical Microbiology | GIGA Medical Genomics | Keith Durkin et al |
| EPI_ISL_581622 | Department of Clinical Microbiology | GIGA Medical Genomics | Keith Durkin et al |
| EPI_ISL_626279 | Department of Clinical Microbiology | GIGA Medical Genomics | Keith Durkin et al |

| | | | |
|---|---|---|---|
| EPI_ISL_626236 | Department of Clinical Microbiology | GIGA Medical Genomics | Keith Durkin et al |
| EPI_ISL_626232 | Department of Clinical Microbiology | GIGA Medical Genomics | Keith Durkin et al |
| EPI_ISL_641597 | Department of Clinical Microbiology | GIGA Medical Genomics | Keith Durkin et al |
| EPI_ISL_661243 | Department of Clinical Microbiology | GIGA Medical Genomics | Keith Durkin et al |
| EPI_ISL_678432 | Department of Clinical Microbiology | GIGA Medical Genomics | Keith Durkin et al |
| EPI_ISL_678452 | Department of Clinical Microbiology | GIGA Medical Genomics | Keith Durkin et al |
| EPI_ISL_707735 | Department of Clinical Microbiology | GIGA Medical Genomics | Keith Durkin et al |
| EPI_ISL_707722 | Department of Clinical Microbiology | GIGA Medical Genomics | Keith Durkin et al |
| EPI_ISL_708455 | Department of Clinical Microbiology | GIGA Medical Genomics | Keith Durkin et al |
| EPI_ISL_708456 | Department of Clinical Microbiology | GIGA Medical Genomics | Keith Durkin et al |
| EPI_ISL_737330 | Department of Clinical Microbiology | GIGA Medical Genomics | Keith Durkin et al |
| EPI_ISL_737372 | Department of Clinical Microbiology | GIGA Medical Genomics | Keith Durkin et al |
| EPI_ISL_737383 | Department of Clinical Microbiology | GIGA Medical Genomics | Keith Durkin et al |
| EPI_ISL_925421 | Department of Clinical Microbiology | GIGA Medical Genomics | Keith Durkin et al |
| EPI_ISL_925470 | Department of Clinical Microbiology | GIGA Medical Genomics | Keith Durkin et al |
| EPI_ISL_925472 | Department of Clinical Microbiology | GIGA Medical Genomics | Keith Durkin et al |
| EPI_ISL_1660775 | University of Liège COVID-19 testing center | GIGA Medical Genomics | Keith Durkin et al |
| EPI_ISL_1660782 | University of Liège COVID-19 testing center | GIGA Medical Genomics | Keith Durkin et al |
| EPI_ISL_848344 | Illinois Department of Public Health | Gagnon Lab, Southern Illinois University | Keith Gagnon et al |
| EPI_ISL_497768 | Department of Microbiology, The University of Hong Kong | Department of Microbiology, The University of Hong Kong | Kelvin K.W. To et al |
| EPI_ISL_497770 | Department of Microbiology, The University of Hong Kong | Department of Microbiology, The University of Hong Kong | Kelvin K.W. To et al |
| EPI_ISL_497789 | Department of Microbiology, The University of Hong Kong | Department of Microbiology, The University of Hong Kong | Kelvin K.W. To et al |
| EPI_ISL_497818 | Department of Microbiology, The University of Hong Kong | Department of Microbiology, The University of Hong Kong | Kelvin K.W. To et al |
| EPI_ISL_497823 | Department of Microbiology, The University of Hong Kong | Department of Microbiology, The University of Hong Kong | Kelvin K.W. To et al |
| EPI_ISL_497851 | Department of Microbiology, The University of Hong Kong | Department of Microbiology, The University of Hong Kong | Kelvin K.W. To et al |
| EPI_ISL_497865 | Department of Microbiology, The University of Hong Kong | Department of Microbiology, The University of Hong Kong | Kelvin K.W. To et al |
| EPI_ISL_498270 | Department of Microbiology, The University of Hong Kong | Department of Microbiology, The University of Hong Kong | Kelvin K.W. To et al |
| EPI_ISL_498271 | Department of Microbiology, The University of Hong Kong | Department of Microbiology, The University of Hong Kong | Kelvin K.W. To et al |
| EPI_ISL_510540 | Department of Microbiology, The University of Hong Kong | Department of Microbiology, The University of Hong Kong | Kelvin K.W. To et al |
| EPI_ISL_1034641 | Department of Microbiology, The University of Hong Kong | Department of Microbiology, The University of Hong Kong | Kelvin K.W. To et al |

| | | | |
|---|---|---|---|
| EPI_ISL_1034642 | Department of Microbiology, The University of Hong Kong | Department of Microbiology, The University of Hong Kong | Kelvin K.W. To et al |
| EPI_ISL_1034656 | Department of Microbiology, The University of Hong Kong | Department of Microbiology, The University of Hong Kong | Kelvin K.W. To et al |
| EPI_ISL_1034382 | Department of Microbiology, The University of Hong Kong | Department of Microbiology, The University of Hong Kong | Kelvin K.W. To et al |
| EPI_ISL_1034592 | Department of Microbiology, The University of Hong Kong | Department of Microbiology, The University of Hong Kong | Kelvin K.W. To et al |
| EPI_ISL_1034373 | Department of Microbiology, The University of Hong Kong | Department of Microbiology, The University of Hong Kong | Kelvin K.W. To et al |
| EPI_ISL_1034600 | Department of Microbiology, The University of Hong Kong | Department of Microbiology, The University of Hong Kong | Kelvin K.W. To et al |
| EPI_ISL_1034601 | Department of Microbiology, The University of Hong Kong | Department of Microbiology, The University of Hong Kong | Kelvin K.W. To et al |
| EPI_ISL_1034603 | Department of Microbiology, The University of Hong Kong | Department of Microbiology, The University of Hong Kong | Kelvin K.W. To et al |
| EPI_ISL_1034368 | Department of Microbiology, The University of Hong Kong | Department of Microbiology, The University of Hong Kong | Kelvin K.W. To et al |
| EPI_ISL_1034668 | Department of Microbiology, The University of Hong Kong | Department of Microbiology, The University of Hong Kong | Kelvin K.W. To et al |
| EPI_ISL_1197081 | Department of Microbiology, The University of Hong Kong | Department of Microbiology, The University of Hong Kong | Kelvin K.W. To et al |
| EPI_ISL_1197114 | Department of Microbiology, The University of Hong Kong | Department of Microbiology, The University of Hong Kong | Kelvin K.W. To et al |
| EPI_ISL_1205467 | Department of Microbiology, The University of Hong Kong | Department of Microbiology, The University of Hong Kong | Kelvin K.W. To et al |
| EPI_ISL_1197083 | Department of Microbiology, The University of Hong Kong | Department of Microbiology, The University of Hong Kong | Kelvin K.W. To et al |
| EPI_ISL_1197084 | Department of Microbiology, The University of Hong Kong | Department of Microbiology, The University of Hong Kong | Kelvin K.W. To et al |
| EPI_ISL_1197109 | Department of Microbiology, The University of Hong Kong | Department of Microbiology, The University of Hong Kong | Kelvin K.W. To et al |
| EPI_ISL_1660475 | Department of Microbiology, The University of Hong Kong | The University of Hong Kong Department of Microbiology | Kelvin K.W. To et al |
| EPI_ISL_469291 | Keio University Hospital | Keio University Hospital | Kenjiro Kosaki et al |
| EPI_ISL_469292 | Keio University Hospital | Keio University Hospital | Kenjiro Kosaki et al |
| EPI_ISL_469295 | Keio University Hospital | Keio University Hospital | Kenjiro Kosaki et al |
| EPI_ISL_417179 | Department of Pathology, Princess Margaret Hospital | Department of Health Technology and Informatics, Faculty of Health and Social Science, The Hong Kong Polytechnic University | Kenneth Siu-Sing LEUNG et al |
| EPI_ISL_417183 | Department of Clinical Pathology, Pamela Youde Nethersole Eastern Hospital | Department of Health Technology and Informatics, Faculty of Health and Social Science, The Hong Kong Polytechnic University | Kenneth Siu-Sing LEUNG et al |
| EPI_ISL_417177 | Department of Pathology, Princess Margaret Hospital | Department of Health Technology and Informatics, Faculty of Health and Social Science, The Hong Kong Polytechnic University | Kenneth Siu-Sing LEUNG et al |
| EPI_ISL_419244 | Department of Clinical Pathology, Pamela Youde Nethersole Eastern Hospital | Department of Health Technology and Informatics, Faculty of Health and Social Science, The Hong Kong Polytechnic University | Kenneth Siu-Sing LEUNG et al |
| EPI_ISL_419245 | Department of Clinical Pathology, Pamela Youde Nethersole Eastern Hospital | Department of Health Technology and Informatics, Faculty of Health and Social Science, The Hong Kong Polytechnic University | Kenneth Siu-Sing LEUNG et al |
| EPI_ISL_419246 | Department of Clinical Pathology, Pamela Youde Nethersole Eastern Hospital | Department of Health Technology and Informatics, Faculty of Health and Social Science, The Hong Kong Polytechnic University | Kenneth Siu-Sing LEUNG et al |
| EPI_ISL_419248 | Department of Clinical Pathology, Pamela Youde Nethersole Eastern Hospital | Department of Health Technology and Informatics, Faculty of Health and Social Science, The Hong Kong Polytechnic University | Kenneth Siu-Sing LEUNG et al |

| | | | |
|---|---|---|---|
| EPI_ISL_419249 | Department of Clinical Pathology, Pamela Youde Nethersole Eastern Hospital | Department of Health Technology and Informatics, Faculty of Health and Social Science, The Hong Kong Polytechnic University | Kenneth Siu-Sing LEUNG et al |
| EPI_ISL_419251 | Department of Clinical Pathology, Pamela Youde Nethersole Eastern Hospital | Department of Health Technology and Informatics, Faculty of Health and Social Science, The Hong Kong Polytechnic University | Kenneth Siu-Sing LEUNG et al |
| EPI_ISL_419252 | Department of Clinical Pathology, Pamela Youde Nethersole Eastern Hospital | Department of Health Technology and Informatics, Faculty of Health and Social Science, The Hong Kong Polytechnic University | Kenneth Siu-Sing LEUNG et al |
| EPI_ISL_419253 | Department of Pathology, United Christian Hospital | Department of Health Technology and Informatics, Faculty of Health and Social Science, The Hong Kong Polytechnic University | Kenneth Siu-Sing LEUNG et al |
| EPI_ISL_605907 | NGS Lab, DNA SOLUTION LTD. | NGS Lab, DNA SOLUTION LTD. | Khan et al |
| EPI_ISL_605908 | NGS Lab, DNA SOLUTION LTD. | NGS Lab, DNA SOLUTION LTD. | Khan et al |
| EPI_ISL_513925 | Microbiology & Bioinformatics and Biostatistics, Kohat University of Science and Technology (Pakistan) & Shanghai Jiao Tong University (China) | Microbiology & Bioinformatics and Biostatistics, Kohat University of Science and Technology (Pakistan) & Shanghai Jiao Tong University (China) | Khan et al |
| EPI_ISL_896284 | SUNY UPSTATE MEDICAL UNIVERSITY | Wadsworth Center, New York State Department of Health | Kirsten St. George et al |
| EPI_ISL_1228374 | WESTCHESTER MEDICAL CENTER | Wadsworth Center, New York State Department of Health | Kirsten St. George et al |
| EPI_ISL_754411 | Wadsworth Center, New York State Department.of Health | Wadsworth Center, New York State Department.of Health | Kirsten St. George et al |
| EPI_ISL_1265406 | 1. Wojewódzka Stacja Sanitarno - Epidemiologiczna w Katowicach (WSSE Katowice); 2. Wojewódzka Stacja Sanitarno - Epidemiologiczna w Kielcach (WSSE Kielce); 3. Wojewódzka Stacja Sanitarno - Epidemiologiczna w Szczecinie (WSSE Szczecin); 4. Wojewódzka Stacja Sanitarno - Epidemiologiczna w Lublinie (WSSE Lublin); 5. Wojewódzka Stacja Sanitarno - Epidemiologiczna w Gorzowie Wielkopolskim (WSSE Gorzów Wielkopolski); 6. Wojewódzka Stacja Sanitarno-Epidemiologiczna w Gdańsku (WSSE Gdańsk); 7. Wojewódzka Stacja Sanitarno-Epidemiologiczna w Poznaniu (WSSE Poznań); 8. Wojewódzka Stacja Sanitarno-Epidemiologiczna w Opolu (WSSE Opole); 9. Wojewódzka Stacja Sanitarno-Epidemiologiczna we Wrocławiu (WSSE Wrocław); 10. Wojewódzka Stacja Sanitarno-Epidemiologiczna w Krakowie (WSSE Kraków); 11. Wojewódzka Stacja Sanitarno-Epidemiologiczna w Olsztynie (WSSE Olsztyn); 12. Wojewódzka Stacja Sanitarno-Epidemiologiczna w Bydgoszczy (WSSE Bydgoszcz); 13. Wojewódzka Stacja Sanitarno-Epidemiologiczna w Rzeszowie (WSSE Rzeszów); 14. Wojewódzka Stacja Sanitarno-Epidemiologiczna w Białymstoku (WSSE Białystok); | 1. ViroGenetics - BSL3 Laboratory of Virology, Małopolska Centre of Biotechnology, Jagiellonian University; 2. Human Genome Variation Research Group, Malopolska Centre of Biotechnology, Jagiellonian University; | Klajmon et al |
| EPI_ISL_1265440 | 1. Wojewódzka Stacja Sanitarno - Epidemiologiczna w Katowicach (WSSE Katowice); 2. Wojewódzka Stacja Sanitarno - Epidemiologiczna w Kielcach (WSSE Kielce); 3. Wojewódzka Stacja Sanitarno - Epidemiologiczna w Szczecinie (WSSE Szczecin); 4. Wojewódzka Stacja Sanitarno - Epidemiologiczna w Lublinie (WSSE Lublin); 5. Wojewódzka Stacja Sanitarno - Epidemiologiczna w Gorzowie Wielkopolskim (WSSE Gorzów Wielkopolski); 6. Wojewódzka Stacja Sanitarno-Epidemiologiczna w Gdańsku (WSSE Gdańsk); 7. Wojewódzka Stacja Sanitarno-Epidemiologiczna w Poznaniu (WSSE Poznań); 8. Wojewódzka Stacja Sanitarno-Epidemiologiczna w Opolu (WSSE Opole); 9. Wojewódzka Stacja Sanitarno-Epidemiologiczna we Wrocławiu (WSSE Wrocław); 10. Wojewódzka Stacja Sanitarno-Epidemiologiczna w Krakowie (WSSE Kraków); 11. Wojewódzka Stacja Sanitarno-Epidemiologiczna w Olsztynie (WSSE Olsztyn); 12. Wojewódzka Stacja Sanitarno-Epidemiologiczna w Bydgoszczy (WSSE Bydgoszcz); 13. Wojewódzka Stacja Sanitarno-Epidemiologiczna w Rzeszowie (WSSE Rzeszów); 14. Wojewódzka Stacja Sanitarno-Epidemiologiczna w Białymstoku (WSSE Białystok); | 1. ViroGenetics - BSL3 Laboratory of Virology, Małopolska Centre of Biotechnology, Jagiellonian University; 2. Human Genome Variation Research Group, Malopolska Centre of Biotechnology, Jagiellonian University; | Klajmon et al |

| | | | |
|---|---|---|---|
| EPI_ISL_1654841 | Laboratory Batajnica | Institute of microbiology and Immunology, Faculty of Medicine, University of Belgrade | Knezevic et al |
| EPI_ISL_1017680 | Institute of Virology, Vaccines and Sera "Torlak" | Institute of microbiology and Immunology, Faculty of Medicine, University of Belgrade | Knezevic et al |
| EPI_ISL_1654811 | Institute of Virology, Vaccines and Sera Torlak | Institute of microbiology and Immunology, Faculty of Medicine, University of Belgrade | Knezevic et al |
| EPI_ISL_1654815 | Institute of Virology, Vaccines and Sera Torlak | Institute of microbiology and Immunology, Faculty of Medicine, University of Belgrade | Knezevic et al |
| EPI_ISL_1017691 | Institute of Virology, Vaccines and Sera "Torlak" | Institute of microbiology and Immunology, Faculty of Medicine, University of Belgrade | Knezevic et al |
| EPI_ISL_1017679 | Institute of Virology, Vaccines and Sera "Torlak" | Institute of microbiology and Immunology, Faculty of Medicine, University of Belgrade | Knezevic et al |
| EPI_ISL_1017688 | Institute of Virology, Vaccines and Sera "Torlak" | Institute of microbiology and Immunology, Faculty of Medicine, University of Belgrade | Knezevic et al |
| EPI_ISL_1017689 | Institute of Virology, Vaccines and Sera "Torlak" | Institute of microbiology and Immunology, Faculty of Medicine, University of Belgrade | Knezevic et al |
| EPI_ISL_1017681 | Institute of Virology, Vaccines and Sera "Torlak" | Institute of microbiology and Immunology, Faculty of Medicine, University of Belgrade | Knezevic et al |
| EPI_ISL_1017699 | Institute of Virology, Vaccines and Sera "Torlak" | Institute of microbiology and Immunology, Faculty of Medicine, University of Belgrade | Knezevic et al |
| EPI_ISL_1017700 | Institute of Virology, Vaccines and Sera "Torlak" | Institute of microbiology and Immunology, Faculty of Medicine, University of Belgrade | Knezevic et al |
| EPI_ISL_1654817 | Institute of Virology, Vaccines and Sera Torlak | Institute of microbiology and Immunology, Faculty of Medicine, University of Belgrade | Knezevic et al |
| EPI_ISL_1654818 | Institute of Virology, Vaccines and Sera Torlak | Institute of microbiology and Immunology, Faculty of Medicine, University of Belgrade | Knezevic et al |
| EPI_ISL_1654809 | Institute of Virology, Vaccines and Sera Torlak | Institute of microbiology and Immunology, Faculty of Medicine, University of Belgrade | Knezevic et al |
| EPI_ISL_1654813 | Institute of Virology, Vaccines and Sera Torlak | Institute of microbiology and Immunology, Faculty of Medicine, University of Belgrade | Knezevic et al |
| EPI_ISL_1654837 | Institute of Virology, Vaccines and Sera Torlak | Institute of microbiology and Immunology, Faculty of Medicine, University of Belgrade | Knezevic et al |
| EPI_ISL_1654835 | Institute of Virology, Vaccines and Sera Torlak | Institute of microbiology and Immunology, Faculty of Medicine, University of Belgrade | Knezevic et al |
| EPI_ISL_955160 | Institute for Biocides and Medical Ecology | Institute of microbiology and Immunology, Faculty of Medicine, University of Belgrade | Knezevic et al |
| EPI_ISL_955166 | Institute for Biocides and Medical Ecology | Institute of microbiology and Immunology, Faculty of Medicine, University of Belgrade | Knezevic et al |
| EPI_ISL_955167 | Institute for Biocides and Medical Ecology | Institute of microbiology and Immunology, Faculty of Medicine, University of Belgrade | Knezevic et al |
| EPI_ISL_1654842 | Laboratory for COVID19 diagnostics, Clinical Centre of Serbia | Institute of microbiology and Immunology, Faculty of Medicine, University of Belgrade | Knezevic et al |
| EPI_ISL_586542 | Government General Hospital, Jam Khambhaliya, Devbhoomi Dwarka | Gujarat Biotechnology Research Centre | Komal Patel et al |

| | | | |
|---|---|---|---|
| EPI_ISL_487446 | CICM-Mali | Bundeswehr Institut of Microbiology | Kouriba et al |
| EPI_ISL_487449 | CICM-Mali | Bundeswehr Institut of Microbiology | Kouriba et al |
| EPI_ISL_487450 | CICM-Mali | Bundeswehr Institut of Microbiology | Kouriba et al |
| EPI_ISL_487452 | CICM-Mali | Bundeswehr Institut of Microbiology | Kouriba et al |
| EPI_ISL_487454 | CICM-Mali | Bundeswehr Institut of Microbiology | Kouriba et al |
| EPI_ISL_487455 | CICM-Mali | Bundeswehr Institut of Microbiology | Kouriba et al |
| EPI_ISL_487461 | CICM-Mali | Bundeswehr Institut of Microbiology | Kouriba et al |
| EPI_ISL_487463 | CICM-Mali | Bundeswehr Institut of Microbiology | Kouriba et al |
| EPI_ISL_645011 | Human Genome Variation Research Group, Malopolska Centre of Biotechnology | Human Genome Variation Research Group, Malopolska Centre of Biotechnology | Kowalski et al |
| EPI_ISL_645030 | Human Genome Variation Research Group, Malopolska Centre of Biotechnology | Human Genome Variation Research Group, Malopolska Centre of Biotechnology | Kowalski et al |
| EPI_ISL_645049 | Human Genome Variation Research Group, Malopolska Centre of Biotechnology | Human Genome Variation Research Group, Malopolska Centre of Biotechnology | Kowalski et al |
| EPI_ISL_645056 | Human Genome Variation Research Group, Malopolska Centre of Biotechnology | Human Genome Variation Research Group, Malopolska Centre of Biotechnology | Kowalski et al |
| EPI_ISL_645057 | Human Genome Variation Research Group, Malopolska Centre of Biotechnology | Human Genome Variation Research Group, Malopolska Centre of Biotechnology | Kowalski et al |
| EPI_ISL_645060 | Human Genome Variation Research Group, Malopolska Centre of Biotechnology | Human Genome Variation Research Group, Malopolska Centre of Biotechnology | Kowalski et al |
| EPI_ISL_645061 | Human Genome Variation Research Group, Malopolska Centre of Biotechnology | Human Genome Variation Research Group, Malopolska Centre of Biotechnology | Kowalski et al |
| EPI_ISL_645062 | Human Genome Variation Research Group, Malopolska Centre of Biotechnology | Human Genome Variation Research Group, Malopolska Centre of Biotechnology | Kowalski et al |
| EPI_ISL_645063 | Human Genome Variation Research Group, Malopolska Centre of Biotechnology | Human Genome Variation Research Group, Malopolska Centre of Biotechnology | Kowalski et al |
| EPI_ISL_645064 | Human Genome Variation Research Group, Malopolska Centre of Biotechnology | Human Genome Variation Research Group, Malopolska Centre of Biotechnology | Kowalski et al |
| EPI_ISL_645065 | Human Genome Variation Research Group, Malopolska Centre of Biotechnology | Human Genome Variation Research Group, Malopolska Centre of Biotechnology | Kowalski et al |
| EPI_ISL_645068 | Human Genome Variation Research Group, Malopolska Centre of Biotechnology | Human Genome Variation Research Group, Malopolska Centre of Biotechnology | Kowalski et al |
| EPI_ISL_645084 | Human Genome Variation Research Group, Malopolska Centre of Biotechnology | Human Genome Variation Research Group, Malopolska Centre of Biotechnology | Kowalski et al |
| EPI_ISL_645085 | Human Genome Variation Research Group, Malopolska Centre of Biotechnology | Human Genome Variation Research Group, Malopolska Centre of Biotechnology | Kowalski et al |
| EPI_ISL_678279 | Mikrobiologie, RARI | Mikrobiologie, RARI | Krasnov et al |
| EPI_ISL_678288 | Mikrobiologie, RARI | Mikrobiologie, RARI | Krasnov et al |
| EPI_ISL_678273 | Mikrobiologie, RARI | Mikrobiologie, RARI | Krasnov et al |

| | | | |
|---|---|---|---|
| EPI_ISL_877227 | Thai Red Cross Emerging Infectious Diseases Health Science Centre, Chulalongkorn Hospital, Faculty of Medicine, Chulalongkorn University | Thai Red Cross Emerging Infectious Diseases Center and Faculty of Medicine, Chulalongkorn University | Krirk Asavametha et al |
| EPI_ISL_759959 | Premier Hospital Surabaya | Institute of Tropical Disease, Universitas Airlangga | Krisnoadi Rahardjo et al |
| EPI_ISL_1072986 | Department of Microbiology, Faculty of Medicine, Universitas Sumatera Utara | Institute of Tropical Disease, Universitas Airlangga; Faculty of Medicine, Universitas Sumatera Utara | Krisnoadi Rahardjo et al |
| EPI_ISL_1273049 | Guam Public Health Laboratory | Centers for Disease Control and Prevention Division of Viral Diseases, Pathogen Discovery | Krista Queen et al |
| EPI_ISL_1273073 | Guam Public Health Laboratory | Centers for Disease Control and Prevention Division of Viral Diseases, Pathogen Discovery | Krista Queen et al |
| EPI_ISL_1273057 | Guam Public Health Laboratory | Centers for Disease Control and Prevention Division of Viral Diseases, Pathogen Discovery | Krista Queen et al |
| EPI_ISL_1273069 | Guam Public Health Laboratory | Centers for Disease Control and Prevention Division of Viral Diseases, Pathogen Discovery | Krista Queen et al |
| EPI_ISL_1273052 | Guam Public Health Laboratory | Centers for Disease Control and Prevention Division of Viral Diseases, Pathogen Discovery | Krista Queen et al |
| EPI_ISL_1273072 | Guam Public Health Laboratory | Centers for Disease Control and Prevention Division of Viral Diseases, Pathogen Discovery | Krista Queen et al |
| EPI_ISL_1273050 | Guam Public Health Laboratory | Centers for Disease Control and Prevention Division of Viral Diseases, Pathogen Discovery | Krista Queen et al |
| EPI_ISL_1273062 | Guam Public Health Laboratory | Centers for Disease Control and Prevention Division of Viral Diseases, Pathogen Discovery | Krista Queen et al |
| EPI_ISL_1273059 | Guam Public Health Laboratory | Centers for Disease Control and Prevention Division of Viral Diseases, Pathogen Discovery | Krista Queen et al |
| EPI_ISL_1273067 | Guam Public Health Laboratory | Centers for Disease Control and Prevention Division of Viral Diseases, Pathogen Discovery | Krista Queen et al |
| EPI_ISL_1273068 | Guam Public Health Laboratory | Centers for Disease Control and Prevention Division of Viral Diseases, Pathogen Discovery | Krista Queen et al |
| EPI_ISL_1273053 | Guam Public Health Laboratory | Centers for Disease Control and Prevention Division of Viral Diseases, Pathogen Discovery | Krista Queen et al |
| EPI_ISL_1273048 | Guam Public Health Laboratory | Centers for Disease Control and Prevention Division of Viral Diseases, Pathogen Discovery | Krista Queen et al |
| EPI_ISL_1273064 | Guam Public Health Laboratory | Centers for Disease Control and Prevention Division of Viral Diseases, Pathogen Discovery | Krista Queen et al |
| EPI_ISL_1273054 | Guam Public Health Laboratory | Centers for Disease Control and Prevention Division of Viral Diseases, Pathogen Discovery | Krista Queen et al |
| EPI_ISL_1273071 | Guam Public Health Laboratory | Centers for Disease Control and Prevention Division of Viral Diseases, Pathogen Discovery | Krista Queen et al |
| EPI_ISL_1273063 | Guam Public Health Laboratory | Centers for Disease Control and Prevention Division of Viral Diseases, Pathogen Discovery | Krista Queen et al |
| EPI_ISL_1273056 | Guam Public Health Laboratory | Centers for Disease Control and Prevention Division of Viral Diseases, Pathogen Discovery | Krista Queen et al |
| EPI_ISL_1273061 | Guam Public Health Laboratory | Centers for Disease Control and Prevention Division of Viral Diseases, Pathogen Discovery | Krista Queen et al |

| | | | |
|---|---|---|---|
| EPI_ISL_450792 | Jamaica Ministry of Health and Wellness | Pathogen Discovery, Respiratory Viruses Branch, Division of Viral Diseases, Centers for Disease Control and Prevention | Krista Queen et al |
| EPI_ISL_450798 | Jamaica Ministry of Health and Wellness | Pathogen Discovery, Respiratory Viruses Branch, Division of Viral Diseases, Centers for Disease Control and Prevention | Krista Queen et al |
| EPI_ISL_408008 | California Department of Health | Pathogen Discovery, Respiratory Viruses Branch, Division of Viral Diseases, Centers for Disease Control and Prevention | Krista Queen et al |
| EPI_ISL_408009 | California Department of Health | Pathogen Discovery, Respiratory Viruses Branch, Division of Viral Diseases, Centers for Disease Control and Prevention | Krista Queen et al |
| EPI_ISL_411954 | California Department of Public Health | Pathogen Discovery, Respiratory Viruses Branch, Division of Viral Diseases, Centers for Disease Control and Prevention | Krista Queen et al |
| EPI_ISL_959618 | Institute of Virology, Biomedical Research Center of the Slovak Academy of Sciences, Bratislava | Faculty of Natural Sciences, Comenius University, Bratislava | Kristína Boršová et al |
| EPI_ISL_1239407 | Institute of Virology, Biomedical Research Center of the Slovak Academy of Sciences, Bratislava | Faculty of Natural Sciences, Comenius University, Bratislava | Kristína Boršová et al |
| EPI_ISL_577737 | Institute of Virology, Biomedical Research Center of the Slovak Academy of Sciences, Bratislava | Faculty of Natural Sciences, Comenius University, Bratislava | Kristína Boršová et al |
| EPI_ISL_577739 | Institute of Virology, Biomedical Research Center of the Slovak Academy of Sciences, Bratislava | Faculty of Natural Sciences, Comenius University, Bratislava | Kristína Boršová et al |
| EPI_ISL_718250 | Institute of Virology, Biomedical Research Center of the Slovak Academy of Sciences, Bratislava | Faculty of Natural Sciences, Comenius University, Bratislava | Kristína Boršová et al |
| EPI_ISL_485389 | University of Ulsan College of Medicine and Asan Medical Center | University of Ulsan College of Medicine and Asan Medical Center | Kuenyoul Park et al |
| EPI_ISL_485390 | University of Ulsan College of Medicine and Asan Medical Center | University of Ulsan College of Medicine and Asan Medical Center | Kuenyoul Park et al |
| EPI_ISL_485391 | University of Ulsan College of Medicine and Asan Medical Center | University of Ulsan College of Medicine and Asan Medical Center | Kuenyoul Park et al |
| EPI_ISL_485393 | University of Ulsan College of Medicine and Asan Medical Center | University of Ulsan College of Medicine and Asan Medical Center | Kuenyoul Park et al |
| EPI_ISL_485395 | University of Ulsan College of Medicine and Asan Medical Center | University of Ulsan College of Medicine and Asan Medical Center | Kuenyoul Park et al |
| EPI_ISL_485396 | University of Ulsan College of Medicine and Asan Medical Center | University of Ulsan College of Medicine and Asan Medical Center | Kuenyoul Park et al |
| EPI_ISL_930843 | Virology, ICAR-National Research Centre on Equines | Virology, ICAR-National Research Centre on Equines | Kumar et al |
| EPI_ISL_760059 | Laboratory of Genetics and Personalized Medicine, Zan Mitrev Clinic | Laboratory of Genetics and Personalized Medicine, Zan Mitrev Clinic | Kungulovski et al. |
| EPI_ISL_760246 | Laboratory of Genetics and Personalized Medicine, Zan Mitrev Clinic | Laboratory of Genetics and Personalized Medicine, Zan Mitrev Clinic | Kungulovski et al. |
| EPI_ISL_1201428 | Laboratory of Genetics and Personalized Medicine, Zan Mitrev Clinic | Laboratory of Genetics and Personalized Medicine, Zan Mitrev Clinic | Kungulovski G. et al. |
| EPI_ISL_1201429 | Laboratory of Genetics and Personalized Medicine, Zan Mitrev Clinic | Laboratory of Genetics and Personalized Medicine, Zan Mitrev Clinic | Kungulovski G. et al. |
| EPI_ISL_1201433 | Laboratory of Genetics and Personalized Medicine, Zan Mitrev Clinic | Laboratory of Genetics and Personalized Medicine, Zan Mitrev Clinic | Kungulovski G. et al. |
| EPI_ISL_1096341 | Laboratory of Genetics and Personalized Medicine, Zan Mitrev Clinic | Laboratory of Genetics and Personalized Medicine, Zan Mitrev Clinic | Kungulovski G. et al. |
| EPI_ISL_415741 | Laboratory Medicine | Department of Laboratory Medicine, Lin-Kou Chang Gung Memorial Hospital, Taoyuan, Taiwan | Kuo-Chien Tsao et al |
| EPI_ISL_415742 | Laboratory Medicine | Department of Laboratory Medicine, Lin-Kou Chang Gung Memorial Hospital, Taoyuan, Taiwan | Kuo-Chien Tsao et al |

| | | | |
|---|---|---|---|
| EPI_ISL_415743 | Laboratory Medicine | Department of Laboratory Medicine, Lin-Kou Chang Gung Memorial Hospital, Taoyuan, Taiwan | Kuo-Chien Tsao et al |
| EPI_ISL_444274 | Laboratory Medicine | Department of Laboratory Medicine, Lin-Kou Chang Gung Memorial Hospital, Taoyuan, Taiwan | Kuo-Chien Tsao et al |
| EPI_ISL_452178 | Laboratory Medicine | Department of Laboratory Medicine, Lin-Kou Chang Gung Memorial Hospital, Taoyuan, Taiwan | Kuo-Chien Tsao et al |
| EPI_ISL_452179 | Laboratory Medicine | Department of Laboratory Medicine, Lin-Kou Chang Gung Memorial Hospital, Taoyuan, Taiwan | Kuo-Chien Tsao et al |
| EPI_ISL_464093 | Laboratory Medicine | Department of Laboratory Medicine, Lin-Kou Chang Gung Memorial Hospital, Taoyuan, Taiwan | Kuo-Chien Tsao et al |
| EPI_ISL_660543 | Laboratory Medicine | Department of Laboratory Medicine, Lin-Kou Chang Gung Memorial Hospital, Taoyuan, Taiwan | Kuo-Chien Tsao et al |
| EPI_ISL_660544 | Laboratory Medicine | Department of Laboratory Medicine, Lin-Kou Chang Gung Memorial Hospital, Taoyuan, Taiwan | Kuo-Chien Tsao et al |
| EPI_ISL_660545 | Laboratory Medicine | Department of Laboratory Medicine, Lin-Kou Chang Gung Memorial Hospital, Taoyuan, Taiwan | Kuo-Chien Tsao et al |
| EPI_ISL_667809 | Laboratory Medicine | Department of Laboratory Medicine, Lin-Kou Chang Gung Memorial Hospital, Taoyuan, Taiwan | Kuo-Chien Tsao et al |
| EPI_ISL_956317 | Laboratory Medicine | Department of Laboratory Medicine, Lin-Kou Chang Gung Memorial Hospital, Taoyuan, Taiwan | Kuo-Chien Tsao et al |
| EPI_ISL_956318 | Laboratory Medicine | Department of Laboratory Medicine, Lin-Kou Chang Gung Memorial Hospital, Taoyuan, Taiwan | Kuo-Chien Tsao et al |
| EPI_ISL_956320 | Laboratory Medicine | Department of Laboratory Medicine, Lin-Kou Chang Gung Memorial Hospital, Taoyuan, Taiwan | Kuo-Chien Tsao et al |
| EPI_ISL_956322 | Laboratory Medicine | Department of Laboratory Medicine, Lin-Kou Chang Gung Memorial Hospital, Taoyuan, Taiwan | Kuo-Chien Tsao et al |
| EPI_ISL_956323 | Laboratory Medicine | Department of Laboratory Medicine, Lin-Kou Chang Gung Memorial Hospital, Taoyuan, Taiwan | Kuo-Chien Tsao et al |
| EPI_ISL_956324 | Laboratory Medicine | Department of Laboratory Medicine, Lin-Kou Chang Gung Memorial Hospital, Taoyuan, Taiwan | Kuo-Chien Tsao et al |
| EPI_ISL_956325 | Laboratory Medicine | Department of Laboratory Medicine, Lin-Kou Chang Gung Memorial Hospital, Taoyuan, Taiwan | Kuo-Chien Tsao et al |
| EPI_ISL_956326 | Laboratory Medicine | Department of Laboratory Medicine, Lin-Kou Chang Gung Memorial Hospital, Taoyuan, Taiwan | Kuo-Chien Tsao et al |
| EPI_ISL_956331 | Laboratory Medicine | Department of Laboratory Medicine, Lin-Kou Chang Gung Memorial Hospital, Taoyuan, Taiwan | Kuo-Chien Tsao et al |
| EPI_ISL_1669972 | Laboratory of virology and molecular diagnostics, Institute of Public Health | Laboratory of virology and molecular diagnostics, Institute of Public Health | Kuzmanovska M et al |
| EPI_ISL_1669992 | Laboratory of virology and molecular diagnostics, Institute of Public Health | Laboratory of virology and molecular diagnostics, Institute of Public Health | Kuzmanovska M et al |
| EPI_ISL_1669993 | Laboratory of virology and molecular diagnostics, Institute of Public Health | Laboratory of virology and molecular diagnostics, Institute of Public Health | Kuzmanovska M et al |
| EPI_ISL_1191759 | Laboratory of virology and molecular diagnostics | Laboratory of virology and molecular diagnostics | Kuzmanovska Maja et al |
| EPI_ISL_1180157 | Laboratory of virology and molecular diagnostics | Laboratory of virology and molecular diagnostics | Kuzmanovska Maja et al |
| EPI_ISL_1180159 | Laboratory of virology and molecular diagnostics | Laboratory of virology and molecular diagnostics | Kuzmanovska Maja et al |

| | | | |
|---|---|---|---|
| EPI_ISL_1180153 | Laboratory of virology and molecular diagnostics | Laboratory of virology and molecular diagnostics | Kuzmanovska Maja et al |
| EPI_ISL_648124 | UHAS COVID-19 Lab | UHAS COVID-19 Lab | Kwabena O. Duedu et al |
| EPI_ISL_648125 | UHAS COVID-19 Lab | UHAS COVID-19 Lab | Kwabena O. Duedu et al |
| EPI_ISL_628761 | UHAS COVID-19 Lab | UHAS COVID-19 Lab | Kwabena O. Duedu et al |
| EPI_ISL_1503159 | National Public Health Organization | National Public Health Organization | Kyriaki Tryfinopoulou et al |
| EPI_ISL_1503160 | National Public Health Organization | National Public Health Organization | Kyriaki Tryfinopoulou et al |
| EPI_ISL_1630201 | Central Public Health Lab, National Public Health Organization | Central Public Health Lab, National Public Health Organization | Kyriaki Tryfinopoulou et al |
| EPI_ISL_1497476 | National Public Health Organization | National Public Health Organization | Kyriaki Tryfinopoulou et al |
| EPI_ISL_1497478 | National Public Health Organization | National Public Health Organization | Kyriaki Tryfinopoulou et al |
| EPI_ISL_1497327 | National Public Health Organization | National Public Health Organization | Kyriaki Tryfinopoulou et al |
| EPI_ISL_1497491 | National Public Health Organization | National Public Health Organization | Kyriaki Tryfinopoulou et al |
| EPI_ISL_1497492 | National Public Health Organization | National Public Health Organization | Kyriaki Tryfinopoulou et al |
| EPI_ISL_1497493 | National Public Health Organization | National Public Health Organization | Kyriaki Tryfinopoulou et al |
| EPI_ISL_1497494 | National Public Health Organization | National Public Health Organization | Kyriaki Tryfinopoulou et al |
| EPI_ISL_1497495 | National Public Health Organization | National Public Health Organization | Kyriaki Tryfinopoulou et al |
| EPI_ISL_1497496 | National Public Health Organization | National Public Health Organization | Kyriaki Tryfinopoulou et al |
| EPI_ISL_1497497 | National Public Health Organization | National Public Health Organization | Kyriaki Tryfinopoulou et al |
| EPI_ISL_1497498 | National Public Health Organization | National Public Health Organization | Kyriaki Tryfinopoulou et al |
| EPI_ISL_1497477 | National Public Health Organization | National Public Health Organization | Kyriaki Tryfinopoulou et al |
| EPI_ISL_1497290 | National Public Health Organization | National Public Health Organization | Kyriaki Tryfinopoulou et al |
| EPI_ISL_1048453 | National Health Laboratory Service, South Africa | KRISP, KZn Research Innovation and Sequencing Platform | Laguda-Akingba O et al |
| EPI_ISL_1366838 | National Health Laboratory Service, South Africa | KRISP, KZN Research Innovation and Sequencing Platform | Laguda-Akingba O et al |
| EPI_ISL_528833 | CSIR-Centre for Cellular and Molecular Biology | CSIR-Centre for Cellular and Molecular Biology | Lamuk Zaveri et al |
| EPI_ISL_751486 | CHU Purpan - Laboratoire de Virologie - Institut Fédératif de Biologie | CHU Purpan - Laboratoire de Virologie - Institut Fédératif de Biologie | Latour J. et al |
| EPI_ISL_415457 | University Hospitals of Geneva Laboratory of Virology | University Hospitals of Geneva Laboratory of Virology | Laubscher F. et al |
| EPI_ISL_415700 | University Hospitals of Geneva Laboratory of Virology | University Hospitals of Geneva Laboratory of Virology | Laubscher F. et al |
| EPI_ISL_415454 | University Hospitals of Geneva Laboratory of Virology | University Hospitals of Geneva Laboratory of Virology | Laubscher F. et al |
| EPI_ISL_429203 | University Hospitals of Geneva Laboratory of Virology | University Hospitals of Geneva Laboratory of Virology | Laubscher F. et al |
| EPI_ISL_413999 | Laboratoire de Virologie, HUG | Swiss National Reference Centre for Influenza | LAUBSCHER Florian et al. |
| EPI_ISL_414019 | Laboratoire de Virologie, HUG | Swiss National Reference Centre for Influenza | LAUBSCHER Florian et al. |

| | | | |
|---|---|---|---|
| EPI_ISL_414020 | Laboratoire de Virologie, HUG | Swiss National Reference Centre for Influenza | LAUBSCHER Florian et al. |
| EPI_ISL_413996 | Laboratoire de Virologie, HUG | Swiss National Reference Centre for Influenza | LAUBSCHER Florian et al. |
| EPI_ISL_510116 | Hospital General Universitario Gregorio Marañón | SeqCOVID-SPAIN consortium/IBV(CSIC) | Laura Pérez-Lago et al |
| EPI_ISL_600492 | Institute of Epidemiology Disease Control And Research | Institute for Developing Science and Health Initiatives | Lauren Cowley et al |
| EPI_ISL_600498 | Institute of Epidemiology Disease Control And Research | Institute for Developing Science and Health Initiatives | Lauren Cowley et al |
| EPI_ISL_600512 | Institute of Epidemiology Disease Control And Research | Institute for Developing Science and Health Initiatives | Lauren Cowley et al |
| EPI_ISL_600562 | Institute of Epidemiology Disease Control And Research | Institute for Developing Science and Health Initiatives | Lauren Cowley et al |
| EPI_ISL_451172 | Lab voor klinische biologie | Onderzoeksgroep Virologie | Laurens Lambrechts et al |
| EPI_ISL_940821 | INSPI-CRN de Influenza y otros virus respiratorios | INSPI-Centro de Investigación Multidisciplinaria de la DTIDI | Leandro Patiño et al |
| EPI_ISL_940775 | INSPI-CRN de Influenza y otros virus respiratorios | INSPI-Centro de Investigación Multidisciplinaria de la DTIDI | Leandro Patiño et al |
| EPI_ISL_940769 | INSPI-CRN de Influenza y otros virus respiratorios | INSPI-Centro de Investigación Multidisciplinaria de la DTIDI | Leandro Patiño et al |
| EPI_ISL_806544 | INSPI Instituto Nacional de Investigación en Salud Pública | Av. Julián Coronel 905 entre Esmeraldas y José Mascote Av. Juan Tanca Marengo No. 100 y Av. de las Américas | Leandro Patiño et al |
| EPI_ISL_574292 | Instituto Nacional de Investigación en Salud Pública | Instituto Nacional de Investigación en Salud Pública | Leandro Patino Patino et al |
| EPI_ISL_574294 | Instituto Nacional de Investigacion en Salud Pública | Instituto Nacional de Investigación en Salud Pública | Leandro Patino Patino et al |
| EPI_ISL_574293 | Instituto Nacional de Investigación en Salud Pública | Instituto Nacional de Investigación en Salud Publica | Leandro Patino Patino et al |
| EPI_ISL_1663230 | Fimlab Laboratories | Fimlab Laboratories | Leena Huhti et al |
| EPI_ISL_1663231 | Fimlab Laboratories | Fimlab Laboratories | Leena Huhti et al |
| EPI_ISL_1663233 | Fimlab Laboratories | Fimlab Laboratories | Leena Huhti et al |
| EPI_ISL_875545 | Instituto de Biotecnologia - UNESP-Botucatu-SP | Instituto de Biotecnologia - UNESP-Botucatu-SP | Leila Sabrina Ullmann et al |
| EPI_ISL_875542 | Instituto de Biotecnologia - UNESP-Botucatu-SP | Instituto de Biotecnologia - UNESP-Botucatu-SP | Leila Sabrina Ullmann et al |
| EPI_ISL_875547 | Instituto de Biotecnologia - UNESP-Botucatu-SP | Instituto de Biotecnologia - UNESP-Botucatu-SP | Leila Sabrina Ullmann et al |
| EPI_ISL_861242 | Instituto de Biotecnologia - UNESP-Botucatu-SP | Instituto de Biotecnologia - UNESP-Botucatu-SP | Leila Sabrina Ullmann et al |
| EPI_ISL_765890 | Massachusetts General Hospital | Infectious Disease Program, Broad Institute of Harvard and MIT | Lemieux et al |
| EPI_ISL_1629785 | Laboratorio de Referencia Nacional de Virus Respiratorios, Instituto Nacional de Salud Peru | Laboratorio de Genómica Microbiana, Universidad Peruana Cayetano Heredia | Lenin Maturrano et al |
| EPI_ISL_1629787 | Laboratorio de Referencia Nacional de Virus Respiratorios, Instituto Nacional de Salud Peru | Laboratorio de Genómica Microbiana, Universidad Peruana Cayetano Heredia | Lenin Maturrano et al |
| EPI_ISL_1629792 | Instituto de Medicina Tropical Alexander Von Humboldt, Universidad Peruana Cayetano Heredia | Laboratorio de Genómica Microbiana, Universidad Peruana Cayetano Heredia | Lenin Maturrano et al |
| EPI_ISL_1629793 | Instituto de Medicina Tropical Alexander Von Humboldt, Universidad Peruana Cayetano Heredia | Laboratorio de Genómica Microbiana, Universidad Peruana Cayetano Heredia | Lenin Maturrano et al |
| EPI_ISL_1629794 | Instituto de Medicina Tropical Alexander Von Humboldt, Universidad Peruana Cayetano Heredia | Laboratorio de Genómica Microbiana, Universidad Peruana Cayetano Heredia | Lenin Maturrano et al |

| | | | |
|---|---|---|---|
| EPI_ISL_1629795 | Instituto de Medicina Tropical Alexander Von Humboldt, Universidad Peruana Cayetano Heredia | Laboratorio de Genómica Microbiana, Universidad Peruana Cayetano Heredia | Lenin Maturrano et al |
| EPI_ISL_1629796 | Instituto de Medicina Tropical Alexander Von Humboldt, Universidad Peruana Cayetano Heredia | Laboratorio de Genómica Microbiana, Universidad Peruana Cayetano Heredia | Lenin Maturrano et al |
| EPI_ISL_1629798 | Instituto de Medicina Tropical Alexander Von Humboldt, Universidad Peruana Cayetano Heredia | Laboratorio de Genómica Microbiana, Universidad Peruana Cayetano Heredia | Lenin Maturrano et al |
| EPI_ISL_1629799 | Instituto de Medicina Tropical Alexander Von Humboldt, Universidad Peruana Cayetano Heredia | Laboratorio de Genómica Microbiana, Universidad Peruana Cayetano Heredia | Lenin Maturrano et al |
| EPI_ISL_1629800 | Instituto de Medicina Tropical Alexander Von Humboldt, Universidad Peruana Cayetano Heredia | Laboratorio de Genómica Microbiana, Universidad Peruana Cayetano Heredia | Lenin Maturrano et al |
| EPI_ISL_1629801 | Instituto de Medicina Tropical Alexander Von Humboldt, Universidad Peruana Cayetano Heredia | Laboratorio de Genómica Microbiana, Universidad Peruana Cayetano Heredia | Lenin Maturrano et al |
| EPI_ISL_1629802 | Instituto de Medicina Tropical Alexander Von Humboldt, Universidad Peruana Cayetano Heredia | Laboratorio de Genómica Microbiana, Universidad Peruana Cayetano Heredia | Lenin Maturrano et al |
| EPI_ISL_1629803 | Instituto de Medicina Tropical Alexander Von Humboldt, Universidad Peruana Cayetano Heredia | Laboratorio de Genómica Microbiana, Universidad Peruana Cayetano Heredia | Lenin Maturrano et al |
| EPI_ISL_1629804 | Instituto de Medicina Tropical Alexander Von Humboldt, Universidad Peruana Cayetano Heredia | Laboratorio de Genómica Microbiana, Universidad Peruana Cayetano Heredia | Lenin Maturrano et al |
| EPI_ISL_1629805 | Instituto de Medicina Tropical Alexander Von Humboldt, Universidad Peruana Cayetano Heredia | Laboratorio de Genómica Microbiana, Universidad Peruana Cayetano Heredia | Lenin Maturrano et al |
| EPI_ISL_1629806 | Instituto de Medicina Tropical Alexander Von Humboldt, Universidad Peruana Cayetano Heredia | Laboratorio de Genómica Microbiana, Universidad Peruana Cayetano Heredia | Lenin Maturrano et al |
| EPI_ISL_1629807 | Instituto de Medicina Tropical Alexander Von Humboldt, Universidad Peruana Cayetano Heredia | Laboratorio de Genómica Microbiana, Universidad Peruana Cayetano Heredia | Lenin Maturrano et al |
| EPI_ISL_801559 | Laboratory of Molecular Virology, Pontificia Universidad Católica de Chile | MSHS Pathogen Surveillance Program | Leonardo I. Almonacid et al |
| EPI_ISL_801566 | Laboratory of Molecular Virology, Pontificia Universidad Católica de Chile | MSHS Pathogen Surveillance Program | Leonardo I. Almonacid et al |
| EPI_ISL_801579 | Laboratory of Molecular Virology, Pontificia Universidad Católica de Chile | MSHS Pathogen Surveillance Program | Leonardo I. Almonacid et al |
| EPI_ISL_801549 | Laboratory of Molecular Virology, Pontificia Universidad Católica de Chile | MSHS Pathogen Surveillance Program | Leonardo I. Almonacid et al |
| EPI_ISL_801606 | Laboratory of Molecular Virology, Pontificia Universidad Católica de Chile | MSHS Pathogen Surveillance Program | Leonardo I. Almonacid et al |
| EPI_ISL_801607 | Laboratory of Molecular Virology, Pontificia Universidad Católica de Chile | MSHS Pathogen Surveillance Program | Leonardo I. Almonacid et al |
| EPI_ISL_801612 | Laboratory of Molecular Virology, Pontificia Universidad Católica de Chile | MSHS Pathogen Surveillance Program | Leonardo I. Almonacid et al |
| EPI_ISL_801613 | Laboratory of Molecular Virology, Pontificia Universidad Católica de Chile | MSHS Pathogen Surveillance Program | Leonardo I. Almonacid et al |
| EPI_ISL_801647 | Laboratory of Molecular Virology, Pontificia Universidad Católica de Chile | MSHS Pathogen Surveillance Program | Leonardo I. Almonacid et al |
| EPI_ISL_801663 | Laboratory of Molecular Virology, Pontificia Universidad Católica de Chile | MSHS Pathogen Surveillance Program | Leonardo I. Almonacid et al |
| EPI_ISL_801731 | Laboratory of Molecular Virology, Pontificia Universidad Católica de Chile | MSHS Pathogen Surveillance Program | Leonardo I. Almonacid et al |
| EPI_ISL_801735 | Laboratory of Molecular Virology, Pontificia Universidad Católica de Chile | MSHS Pathogen Surveillance Program | Leonardo I. Almonacid et al |
| EPI_ISL_801816 | Laboratory of Molecular Virology, Pontificia Universidad Católica de Chile | MSHS Pathogen Surveillance Program | Leonardo I. Almonacid et al |
| EPI_ISL_801848 | Laboratory of Molecular Virology, Pontificia Universidad Católica de Chile | MSHS Pathogen Surveillance Program | Leonardo I. Almonacid et al |

| | | | |
|---|---|---|---|
| EPI_ISL_801853 | Laboratory of Molecular Virology, Pontificia Universidad Católica de Chile | MSHS Pathogen Surveillance Program | Leonardo I. Almonacid et al |
| EPI_ISL_801593 | Laboratory of Molecular Virology, Pontificia Universidad Católica de Chile | MSHS Pathogen Surveillance Program | Leonardo I. Almonacid et al |
| EPI_ISL_985395 | National Institute of Infectious Diseases-Prof. Dr. Matei Bals Molecular Diagnostics Laboratory | National Institute of Infectious Diseases-Prof. Dr. Matei Bals Molecular Diagnostics Laboratory | Leontina Banica et al |
| EPI_ISL_979544 | National Institute of Infectious Diseases-Prof. Dr. Matei Bals Molecular Diagnostics Laboratory | National Institute of Infectious Diseases-Prof. Dr. Matei Bals Molecular Diagnostics Laboratory | Leontina Banica et al |
| EPI_ISL_885154 | National Institute of Infectious Diseases-Prof. Dr. Matei Bals Molecular Diagnostics Laboratory | National Institute of Infectious Diseases-Prof. Dr. Matei Bals Molecular Diagnostics Laboratory | Leontina Banica et al |
| EPI_ISL_979959 | National Institute of Infectious Diseases-Prof. Dr. Matei Bals Molecular Diagnostics Laboratory | National Institute of Infectious Diseases-Prof. Dr. Matei Bals Molecular Diagnostics Laboratory | Leontina Banica et al |
| EPI_ISL_1116445 | National Institute of Infectious Diseases-Prof. Dr. Matei Bals Molecular Diagnostics Laboratory | National Institute of Infectious Diseases-Prof. Dr. Matei Bals Molecular Diagnostics Laboratory | Leontina Banica et al |
| EPI_ISL_1116446 | National Institute of Infectious Diseases-Prof. Dr. Matei Bals Molecular Diagnostics Laboratory | National Institute of Infectious Diseases-Prof. Dr. Matei Bals Molecular Diagnostics Laboratory | Leontina Banica et al |
| EPI_ISL_1184825 | National Institute of Infectious Diseases-Prof. Dr. Matei Bals Molecular Diagnostics Laboratory | National Institute of Infectious Diseases-Prof. Dr. Matei Bals Molecular Diagnostics Laboratory | Leontina Banica et al |
| EPI_ISL_1184826 | National Institute of Infectious Diseases-Prof. Dr. Matei Bals Molecular Diagnostics Laboratory | National Institute of Infectious Diseases-Prof. Dr. Matei Bals Molecular Diagnostics Laboratory | Leontina Banica et al |
| EPI_ISL_1279947 | National Institute of Infectious Diseases-Prof. Dr. Matei Bals Molecular Diagnostics Laboratory | National Institute of Infectious Diseases-Prof. Dr. Matei Bals Molecular Diagnostics Laboratory | Leontina Banica et al |
| EPI_ISL_1279949 | National Institute of Infectious Diseases-Prof. Dr. Matei Bals Molecular Diagnostics Laboratory | National Institute of Infectious Diseases-Prof. Dr. Matei Bals Molecular Diagnostics Laboratory | Leontina Banica et al |
| EPI_ISL_1279953 | National Institute of Infectious Diseases-Prof. Dr. Matei Bals Molecular Diagnostics Laboratory | National Institute of Infectious Diseases-Prof. Dr. Matei Bals Molecular Diagnostics Laboratory | Leontina Banica et al |
| EPI_ISL_1279956 | National Institute of Infectious Diseases-Prof. Dr. Matei Bals Molecular Diagnostics Laboratory | National Institute of Infectious Diseases-Prof. Dr. Matei Bals Molecular Diagnostics Laboratory | Leontina Banica et al |
| EPI_ISL_1279959 | National Institute of Infectious Diseases-Prof. Dr. Matei Bals Molecular Diagnostics Laboratory | National Institute of Infectious Diseases-Prof. Dr. Matei Bals Molecular Diagnostics Laboratory | Leontina Banica et al |
| EPI_ISL_1279965 | National Institute of Infectious Diseases-Prof. Dr. Matei Bals Molecular Diagnostics Laboratory | National Institute of Infectious Diseases-Prof. Dr. Matei Bals Molecular Diagnostics Laboratory | Leontina Banica et al |
| EPI_ISL_1279966 | National Institute of Infectious Diseases-Prof. Dr. Matei Bals Molecular Diagnostics Laboratory | National Institute of Infectious Diseases-Prof. Dr. Matei Bals Molecular Diagnostics Laboratory | Leontina Banica et al |
| EPI_ISL_468136 | [Romania, Bucharest] National Institute for Infectious Diseases “Prof. Dr. Matei Balș” | [Romania, Bucharest] National Institute for Infectious Diseases “Prof. Dr. Matei Balș” | Leontina Banica et al |
| EPI_ISL_1279960 | National Institute of Infectious Diseases-Prof. Dr. Matei Bals Molecular Diagnostics Laboratory | National Institute of Infectious Diseases-Prof. Dr. Matei Bals Molecular Diagnostics Laboratory | Leontina Banica et al |
| EPI_ISL_1116447 | National Institute of Infectious Diseases-Prof. Dr. Matei Bals Molecular Diagnostics Laboratory | National Institute of Infectious Diseases-Prof. Dr. Matei Bals Molecular Diagnostics Laboratory | Leontina Banica et al |
| EPI_ISL_862836 | National Institute of Infectious Diseases-Prof. Dr. Matei Bals Molecular Diagnostics Laboratory | National Institute of Infectious Diseases-Prof. Dr. Matei Bals Molecular Diagnostics Laboratory | Leontina Banica et al |
| EPI_ISL_468137 | [Romania, Bucharest] National Institute for Infectious Diseases “Prof. Dr. Matei Balș” | [Romania, Bucharest] National Institute for Infectious Diseases “Prof. Dr. Matei Balș” | Leontina Banica et al |

| | | | |
|---|---|---|---|
| EPI_ISL_468139 | [Romania, Bucharest] National Institute for Infectious Diseases “Prof. Dr. Matei Balș” | [Romania, Bucharest] National Institute for Infectious Diseases “Prof. Dr. Matei Balș” | Leontina Banica et al |
| EPI_ISL_468141 | [Romania, Bucharest] National Institute for Infectious Diseases “Prof. Dr. Matei Balș” | [Romania, Bucharest] National Institute for Infectious Diseases “Prof. Dr. Matei Balș” | Leontina Banica et al |
| EPI_ISL_468143 | [Romania, Bucharest] National Institute for Infectious Diseases “Prof. Dr. Matei Balș” | [Romania, Bucharest] National Institute for Infectious Diseases “Prof. Dr. Matei Balș” | Leontina Banica et al |
| EPI_ISL_682298 | Respiratory virus Laboratory, Chinese Academy of Medical Science | Respiratory virus Laboratory, Chinese Academy of Medical Science | Li et al |
| EPI_ISL_417420 | Jiangxi province Center for Disease Control and Prevention | Jiangxi province Center for Disease Control and Prevention | Li jian Xiong et al |
| EPI_ISL_498226 | LIC | LIC | LIC et al |
| EPI_ISL_1098895 | Laboratory of Communicable Diseasea | Laboratory of Communicable Diseasea | Liidia Dotsenko et al |
| EPI_ISL_1138533 | Laboratory of Communicable Diseases | 1. Laboratory of Communicable Diseases (Estonia); 2. Eurofins Genomics Europe Sequencing GmbH | Liidia Dotsenko et al |
| EPI_ISL_1138531 | Laboratory of Communicable Diseases | 1. Laboratory of Communicable Diseases (Estonia); 2. Eurofins Genomics Europe Sequencing GmbH | Liidia Dotsenko et al |
| EPI_ISL_1138512 | SA Pärnu Hospital Laboratory | 1. Laboratory of Communicable Diseases (Estonia); 2. Eurofins Genomics Europe Sequencing GmbH | Liidia Dotsenko et al |
| EPI_ISL_1138513 | SA Pärnu Hospital Laboratory | 1. Laboratory of Communicable Diseases (Estonia); 2. Eurofins Genomics Europe Sequencing GmbH | Liidia Dotsenko et al |
| EPI_ISL_1138514 | SA Pärnu Hospital Laboratory | 1. Laboratory of Communicable Diseases (Estonia); 2. Eurofins Genomics Europe Sequencing GmbH | Liidia Dotsenko et al |
| EPI_ISL_1138515 | SA Pärnu Hospital Laboratory | 1. Laboratory of Communicable Diseases (Estonia); 2. Eurofins Genomics Europe Sequencing GmbH | Liidia Dotsenko et al |
| EPI_ISL_1259062 | Synlab Eesti OÜ | 1. Laboratory of Communicable Diseases (Estonia); 2. Eurofins Genomics Europe Sequencing GmbH | Liidia Dotsenko et al. |
| EPI_ISL_1470225 | Synlab Eesti OÜ | 1. Laboratory of Communicable Diseases (Estonia); 2. Eurofins Genomics Europe Sequencing GmbH | Liidia Dotsenko et al. |
| EPI_ISL_1470151 | Synlab Eesti OÜ | 1. Laboratory of Communicable Diseases (Estonia); 2. Eurofins Genomics Europe Sequencing GmbH | Liidia Dotsenko et al. |
| EPI_ISL_1319299 | Synlab Eesti OÜ | 1. Laboratory of Communicable Diseases (Estonia); 2. Eurofins Genomics Europe Sequencing GmbH | Liidia Dotsenko et al. |
| EPI_ISL_1259086 | Synlab Eesti OÜ | 1. Laboratory of Communicable Diseases (Estonia); 2. Eurofins Genomics Europe Sequencing GmbH | Liidia Dotsenko et al. |
| EPI_ISL_1470026 | Synlab Eesti OÜ | 1. Laboratory of Communicable Diseases (Estonia); 2. Eurofins Genomics Europe Sequencing GmbH | Liidia Dotsenko et al. |
| EPI_ISL_1319321 | Synlab Eesti OÜ | 1. Laboratory of Communicable Diseases (Estonia); 2. Eurofins Genomics Europe Sequencing GmbH | Liidia Dotsenko et al. |
| EPI_ISL_1470138 | Synlab Eesti OÜ | 1. Laboratory of Communicable Diseases (Estonia); 2. Eurofins Genomics Europe Sequencing GmbH | Liidia Dotsenko et al. |
| EPI_ISL_1470134 | North Estonia Medical Centre laboratory | 1. Laboratory of Communicable Diseases (Estonia); 2. Eurofins Genomics Europe Sequencing GmbH | Liidia Dotsenko et al. |
| EPI_ISL_1470176 | Synlab Eesti OÜ | 1. Laboratory of Communicable Diseases (Estonia); 2. Eurofins Genomics Europe Sequencing GmbH | Liidia Dotsenko et al. |

| | | | |
|---|---|---|---|
| EPI_ISL_1470177 | Synlab Eesti OÜ | 1. Laboratory of Communicable Diseases (Estonia); 2. Eurofins Genomics Europe Sequencing GmbH | Liidia Dotsenko et al. |
| EPI_ISL_1470189 | Synlab Eesti OÜ | 1. Laboratory of Communicable Diseases (Estonia); 2. Eurofins Genomics Europe Sequencing GmbH | Liidia Dotsenko et al. |
| EPI_ISL_1470197 | Synlab Eesti OÜ | 1. Laboratory of Communicable Diseases (Estonia); 2. Eurofins Genomics Europe Sequencing GmbH | Liidia Dotsenko et al. |
| EPI_ISL_1470287 | Synlab Eesti OÜ | 1. Laboratory of Communicable Diseases (Estonia); 2. Eurofins Genomics Europe Sequencing GmbH | Liidia Dotsenko et al. |
| EPI_ISL_1259026 | North Estonia Medical Centre laboratory | 1. Laboratory of Communicable Diseases (Estonia); 2. Eurofins Genomics Europe Sequencing GmbH | Liidia Dotsenko et al. |
| EPI_ISL_431180 | Fujian Center for Disease Control and Prevention | Fujian Center for Disease Control and Prevention | Lin Qi et al |
| EPI_ISL_431240 | Fujian Center for Disease Control and Prevention | Fujian Center for Disease Control and Prevention | Lin Qi et al |
| EPI_ISL_452236 | VC Sorgenfrimottagningen | The Public Health Agency of Sweden | Lisa Esbjornsson Klemendz et al |
| EPI_ISL_450816 | orestadsklinikens VC | The Public Health Agency of Sweden | Lisa Kjellberg / Laura Plavitu et al |
| EPI_ISL_445241 | Ã–restadsklinikens VC | The Public Health Agency of Sweden | Lisa Kjellberg / Laura Plavitu et al |
| EPI_ISL_678397 | Suceava County Emergency Hospital | Stefan cel Mare University Metagenomics Lab | Lobiuc Andrei et al |
| EPI_ISL_486834 | Suceava County Energency Hospital Sf. Ioan cel Nou | SMU Metagenomics lab | Lobiuc Andrei et al |
| EPI_ISL_491036 | Suceava County Emergency Hospital | Stefan cel Mare University Metagenomics Lab | Lobiuc Andrei et al |
| EPI_ISL_491038 | Suceava County Emergency Hospital | Stefan cel Mare University Metagenomics Lab | Lobiuc Andrei et al |
| EPI_ISL_491059 | Suceava County Emergency Hospital | Stefan cel Mare University Metagenomics Lab | Lobiuc Andrei et al |
| EPI_ISL_896374 | University of Medicine and Pharmacy of Craiova | Stefan cel Mare University Metagenomics Lab | Lobiuc Andrei et al |
| EPI_ISL_896375 | University of Medicine and Pharmacy of Craiova | Stefan cel Mare University Metagenomics Lab | Lobiuc Andrei et al |
| EPI_ISL_891227 | The Oncology Institute Prof. Dr. Ion Chiricuta Cluj Napoca | Stefan cel Mare University Metagenomics Lab | Lobiuc Andrei et al |
| EPI_ISL_891230 | The Oncology Institute Prof. Dr. Ion Chiricuta Cluj Napoca | Stefan cel Mare University Metagenomics Lab | Lobiuc Andrei et al |
| EPI_ISL_891224 | The Oncology Institute Prof. Dr. Ion Chiricuta Cluj Napoca | Stefan cel Mare University Metagenomics Lab | Lobiuc Andrei et al |
| EPI_ISL_891226 | The Oncology Institute Prof. Dr. Ion Chiricuta Cluj Napoca | Stefan cel Mare University Metagenomics Lab | Lobiuc Andrei et al |
| EPI_ISL_891225 | The Oncology Institute Prof. Dr. Ion Chiricuta Cluj Napoca | Stefan cel Mare University Metagenomics Lab | Lobiuc Andrei et al |
| EPI_ISL_486855 | Emergency county Hospital Suceava | Stefan cel Mare University Metagenomics Lab | Lobiuc Andrei et al. |
| EPI_ISL_491039 | Suceava County Emergency Hospital | Stefan cel Mare University Metagenomics Lab | Lobiuc Andrei et al. |
| EPI_ISL_491048 | Suceava County Emergency Hospital | Stefan cel Mare University Metagenomics Lab | Lobiuc Andrei et al. |
| EPI_ISL_491051 | Suceava County Emergency Hospital | Stefan cel Mare University Metagenomics Lab | Lobiuc Andrei et al. |
| EPI_ISL_491052 | Suceava County Emergency Hospital | Stefan cel Mare University Metagenomics Lab | Lobiuc Andrei et al. |
| EPI_ISL_491055 | Suceava County Emergency Hospital | Stefan cel Mare University Metagenomics Lab | Lobiuc Andrei et al. |

| | | | |
|---|---|---|---|
| EPI_ISL_491074 | Suceava County Emergency Hospital | Stefan cel Mare University Metagenomics Lab | Lobiuc Andrei et al. |
| EPI_ISL_491089 | Suceava County Emergency Hospital | Stefan cel Mare University Metagenomics Lab | Lobiuc Andrei et al. |
| EPI_ISL_444998 | Naval Health Research Center | Naval Medical Research Center Biological Defense Research Directorate | Logan Voegtly et al |
| EPI_ISL_444999 | Naval Health Research Center | Naval Medical Research Center Biological Defense Research Directorate | Logan Voegtly et al |
| EPI_ISL_445000 | Naval Health Research Center | Naval Medical Research Center Biological Defense Research Directorate | Logan Voegtly et al |
| EPI_ISL_849656 | Servizio Igiene Epidemiologia e Sanità Pubblica (SIESP)-L'Aquila | Istitituto Zooprofilattico Sperimentale dell'Abruzzo e Molise G.Caporale | Lorusso A et al |
| EPI_ISL_849657 | Servizio Igiene Epidemiologia e Sanità Pubblica (SIESP)-L'Aquila | Istitituto Zooprofilattico Sperimentale dell'Abruzzo e Molise G.Caporale | Lorusso A et al |
| EPI_ISL_849658 | Servizio Igiene Epidemiologia e Sanità Pubblica (SIESP)-L'Aquila | Istitituto Zooprofilattico Sperimentale dell'Abruzzo e Molise G.Caporale | Lorusso A et al |
| EPI_ISL_849659 | Servizio Igiene Epidemiologia e Sanità Pubblica (SIESP)-L'Aquila | Istitituto Zooprofilattico Sperimentale dell'Abruzzo e Molise G.Caporale | Lorusso A et al |
| EPI_ISL_849660 | Servizio Igiene Epidemiologia e Sanità Pubblica (SIESP)-L'Aquila | Istitituto Zooprofilattico Sperimentale dell'Abruzzo e Molise G.Caporale | Lorusso A et al |
| EPI_ISL_875566 | SIESP L'AQUILA | Istituto Zooprofilattico Sperimentale dell’Abruzzo e Molise “G. Caporale” | Lorusso A et al |
| EPI_ISL_875567 | SIESP L'AQUILA | Istituto Zooprofilattico Sperimentale dell’Abruzzo e Molise “G. Caporale” | Lorusso A et al |
| EPI_ISL_875568 | SIESP L'AQUILA | Istituto Zooprofilattico Sperimentale dell’Abruzzo e Molise “G. Caporale” | Lorusso A et al |
| EPI_ISL_528934 | Agenzia di Tutela della Salute di Bergamo | Istitituto Zooprofilattico Sperimentale dell'Abruzzo e Molise G.Caporale | Lorusso A et al |
| EPI_ISL_528942 | Agenzia di Tutela della Salute di Bergamo | Istitituto Zooprofilattico Sperimentale dell'Abruzzo e Molise G.Caporale | Lorusso A et al |
| EPI_ISL_528945 | Agenzia di Tutela della Salute di Bergamo | Istitituto Zooprofilattico Sperimentale dell'Abruzzo e Molise G.Caporale | Lorusso A et al |
| EPI_ISL_528946 | Agenzia di Tutela della Salute di Bergamo | Istitituto Zooprofilattico Sperimentale dell'Abruzzo e Molise G.Caporale | Lorusso A et al |
| EPI_ISL_514131 | Rondônia Central Public Health Laboratory (LACEN/RO), vinculated to State Health Secretariat of Rondônia (SESAU/RO) | Molecular Virology Laboratory of Oswaldo Cruz Foundation of Rondônia | Luan Felipo Botelho-Souza et al |
| EPI_ISL_514132 | Rondônia Central Public Health Laboratory (LACEN/RO), vinculated to State Health Secretariat of Rondônia (SESAU/RO) | Molecular Virology Laboratory of Oswaldo Cruz Foundation of Rondônia | Luan Felipo Botelho-Souza et al |
| EPI_ISL_514133 | Rondônia Central Public Health Laboratory (LACEN/RO), vinculated to State Health Secretariat of Rondônia (SESAU/RO) | Molecular Virology Laboratory of Oswaldo Cruz Foundation of Rondônia | Luan Felipo Botelho-Souza et al |
| EPI_ISL_514134 | Rondônia Central Public Health Laboratory (LACEN/RO), vinculated to State Health Secretariat of Rondônia (SESAU/RO) | Molecular Virology Laboratory of Oswaldo Cruz Foundation of Rondônia | Luan Felipo Botelho-Souza et al |
| EPI_ISL_514135 | Rondônia Central Public Health Laboratory (LACEN/RO), vinculated to State Health Secretariat of Rondônia (SESAU/RO) | Molecular Virology Laboratory of Oswaldo Cruz Foundation of Rondônia | Luan Felipo Botelho-Souza et al |
| EPI_ISL_514136 | Rondônia Central Public Health Laboratory (LACEN/RO), vinculated to State Health Secretariat of Rondônia (SESAU/RO) | Molecular Virology Laboratory of Oswaldo Cruz Foundation of Rondônia | Luan Felipo Botelho-Souza et al |
| EPI_ISL_514137 | Rondônia Central Public Health Laboratory (LACEN/RO), vinculated to State Health Secretariat of Rondônia (SESAU/RO) | Molecular Virology Laboratory of Oswaldo Cruz Foundation of Rondônia | Luan Felipo Botelho-Souza et al |
| EPI_ISL_514138 | Rondônia Central Public Health Laboratory (LACEN/RO), vinculated to State Health Secretariat of Rondônia (SESAU/RO) | Molecular Virology Laboratory of Oswaldo Cruz Foundation of Rondônia | Luan Felipo Botelho-Souza et al |
| EPI_ISL_1626618 | Laboratorio de Salud Pública Bogota | Gencore - Universidad de los Andes | Luisa Sacristan et al |
| EPI_ISL_1626621 | Laboratorio de Salud Pública Bogota | Gencore - Universidad de los Andes | Luisa Sacristan et al |

| | | | |
|---|---|---|---|
| EPI_ISL_1626619 | Laboratorio de Salud Pública Bogota | Gencore - Universidad de los Andes | Luisa Sacristan et al |
| EPI_ISL_1626615 | Gencore - Universidad de los Andes | Gencore - Universidad de los Andes | Luisa Sacristan et al |
| EPI_ISL_1626613 | Gencore - Universidad de los Andes | Gencore - Universidad de los Andes | Luisa Sacristan et al |
| EPI_ISL_1626616 | Gencore - Universidad de los Andes | Gencore - Universidad de los Andes | Luisa Sacristan et al |
| EPI_ISL_1626611 | Gencore - Universidad de los Andes | Gencore - Universidad de los Andes | Luisa Sacristan et al |
| EPI_ISL_1664115 | Laboratorio Central Noel Nutels | Bioinformatics Laboratory / LNCC | Luiz G P de Almeida et al |
| EPI_ISL_1664116 | Laboratorio Central Noel Nutels | Bioinformatics Laboratory / LNCC | Luiz G P de Almeida et al |
| EPI_ISL_1664121 | Laboratorio Central Noel Nutels | Bioinformatics Laboratory / LNCC | Luiz G P de Almeida et al |
| EPI_ISL_1664123 | Laboratorio Central Noel Nutels | Bioinformatics Laboratory / LNCC | Luiz G P de Almeida et al |
| EPI_ISL_1664156 | Laboratorio Central Noel Nutels | Bioinformatics Laboratory / LNCC | Luiz G P de Almeida et al |
| EPI_ISL_1664157 | Laboratorio Central Noel Nutels | Bioinformatics Laboratory / LNCC | Luiz G P de Almeida et al |
| EPI_ISL_1664160 | Laboratorio Central Noel Nutels | Bioinformatics Laboratory / LNCC | Luiz G P de Almeida et al |
| EPI_ISL_1664164 | Laboratorio Central Noel Nutels | Bioinformatics Laboratory / LNCC | Luiz G P de Almeida et al |
| EPI_ISL_1664165 | Laboratorio Central Noel Nutels | Bioinformatics Laboratory / LNCC | Luiz G P de Almeida et al |
| EPI_ISL_1664166 | Laboratorio Central Noel Nutels | Bioinformatics Laboratory / LNCC | Luiz G P de Almeida et al |
| EPI_ISL_1664167 | Laboratorio Central Noel Nutels | Bioinformatics Laboratory / LNCC | Luiz G P de Almeida et al |
| EPI_ISL_1664168 | Laboratorio Central Noel Nutels | Bioinformatics Laboratory / LNCC | Luiz G P de Almeida et al |
| EPI_ISL_1664169 | Laboratorio Central Noel Nutels | Bioinformatics Laboratory / LNCC | Luiz G P de Almeida et al |
| EPI_ISL_1664170 | Laboratorio Central Noel Nutels | Bioinformatics Laboratory / LNCC | Luiz G P de Almeida et al |
| EPI_ISL_1664171 | Laboratorio Central Noel Nutels | Bioinformatics Laboratory / LNCC | Luiz G P de Almeida et al |
| EPI_ISL_1664172 | Laboratorio Central Noel Nutels | Bioinformatics Laboratory / LNCC | Luiz G P de Almeida et al |
| EPI_ISL_1664173 | Laboratorio Central Noel Nutels | Bioinformatics Laboratory / LNCC | Luiz G P de Almeida et al |
| EPI_ISL_1664180 | Laboratorio Central Noel Nutels | Bioinformatics Laboratory / LNCC | Luiz G P de Almeida et al |
| EPI_ISL_1664191 | Laboratorio Central Noel Nutels | Bioinformatics Laboratory / LNCC | Luiz G P de Almeida et al |
| EPI_ISL_1664199 | Laboratorio Central Noel Nutels | Bioinformatics Laboratory / LNCC | Luiz G P de Almeida et al |
| EPI_ISL_910339 | Laboratory for Respiratory Viruses, Cantacuzino National Military-Medical Institute for Research and Development | Cantacuzino Institute Virology | Luiza Ustea et al |
| EPI_ISL_853819 | Department of Pathology, Landeskrankenhaus Graz II, Medical University Graz | Bergthaler laboratory, CeMM Research Center for Molecular Medicine of the Austrian Academy of Sciences | Lukas Endler et al |
| EPI_ISL_853886 | Center for Virology, Medical University of Vienna | Bergthaler laboratory, CeMM Research Center for Molecular Medicine of the Austrian Academy of Sciences | Lukas Endler et al |
| EPI_ISL_853836 | Center for Virology, Medical University of Vienna | Bergthaler laboratory, CeMM Research Center for Molecular Medicine of the Austrian Academy of Sciences | Lukas Endler et al |

| | | | |
|---|---|---|---|
| EPI_ISL_853837 | Center for Virology, Medical University of Vienna | Bergthaler laboratory, CeMM Research Center for Molecular Medicine of the Austrian Academy of Sciences | Lukas Endler et al |
| EPI_ISL_853840 | Center for Virology, Medical University of Vienna | Bergthaler laboratory, CeMM Research Center for Molecular Medicine of the Austrian Academy of Sciences | Lukas Endler et al |
| EPI_ISL_853751 | Center for Virology, Medical University of Vienna | Bergthaler laboratory, CeMM Research Center for Molecular Medicine of the Austrian Academy of Sciences | Lukas Endler et al |
| EPI_ISL_853892 | Center for Virology, Medical University of Vienna | Bergthaler laboratory, CeMM Research Center for Molecular Medicine of the Austrian Academy of Sciences | Lukas Endler et al |
| EPI_ISL_853893 | Center for Virology, Medical University of Vienna | Bergthaler laboratory, CeMM Research Center for Molecular Medicine of the Austrian Academy of Sciences | Lukas Endler et al |
| EPI_ISL_853897 | Center for Virology, Medical University of Vienna | Bergthaler laboratory, CeMM Research Center for Molecular Medicine of the Austrian Academy of Sciences | Lukas Endler et al |
| EPI_ISL_853899 | Center for Virology, Medical University of Vienna | Bergthaler laboratory, CeMM Research Center for Molecular Medicine of the Austrian Academy of Sciences | Lukas Endler et al |
| EPI_ISL_853849 | Center for Virology, Medical University of Vienna | Bergthaler laboratory, CeMM Research Center for Molecular Medicine of the Austrian Academy of Sciences | Lukas Endler et al |
| EPI_ISL_853911 | Center for Virology, Medical University of Vienna | Bergthaler laboratory, CeMM Research Center for Molecular Medicine of the Austrian Academy of Sciences | Lukas Endler et al |
| EPI_ISL_853979 | Austrian Agency for Health and Food Safety (AGES) | Bergthaler laboratory, CeMM Research Center for Molecular Medicine of the Austrian Academy of Sciences | Lukas Endler et al |
| EPI_ISL_854277 | Austrian Agency for Health and Food Safety (AGES) | Bergthaler laboratory, CeMM Research Center for Molecular Medicine of the Austrian Academy of Sciences | Lukas Endler et al |
| EPI_ISL_853977 | Austrian Agency for Health and Food Safety (AGES) | Bergthaler laboratory, CeMM Research Center for Molecular Medicine of the Austrian Academy of Sciences | Lukas Endler et al |
| EPI_ISL_1495157 | Department of Laboratory Medicine, Division of Clinical Virology, University of Medicine, Vienna | Bergthaler laboratory, CeMM Research Center for Molecular Medicine of the Austrian Academy of Sciences | Lukas Endler et al |
| EPI_ISL_1495159 | Department of Laboratory Medicine, Division of Clinical Virology, University of Medicine, Vienna | Bergthaler laboratory, CeMM Research Center for Molecular Medicine of the Austrian Academy of Sciences | Lukas Endler et al |
| EPI_ISL_1495160 | Department of Laboratory Medicine, Division of Clinical Virology, University of Medicine, Vienna | Bergthaler laboratory, CeMM Research Center for Molecular Medicine of the Austrian Academy of Sciences | Lukas Endler et al |
| EPI_ISL_1495161 | Department of Laboratory Medicine, Division of Clinical Virology, University of Medicine, Vienna | Bergthaler laboratory, CeMM Research Center for Molecular Medicine of the Austrian Academy of Sciences | Lukas Endler et al |
| EPI_ISL_1495162 | Department of Laboratory Medicine, Division of Clinical Virology, University of Medicine, Vienna | Bergthaler laboratory, CeMM Research Center for Molecular Medicine of the Austrian Academy of Sciences | Lukas Endler et al |
| EPI_ISL_1495164 | Department of Laboratory Medicine, Division of Clinical Virology, University of Medicine, Vienna | Bergthaler laboratory, CeMM Research Center for Molecular Medicine of the Austrian Academy of Sciences | Lukas Endler et al |
| EPI_ISL_1008372 | Department of Laboratory Medicine, Division of Clinical Virology, University of Medicine, Vienna | Bergthaler laboratory, CeMM Research Center for Molecular Medicine of the Austrian Academy of Sciences | Lukas Endler et al |
| EPI_ISL_1008373 | Department of Laboratory Medicine, Division of Clinical Virology, University of Medicine, Vienna | Bergthaler laboratory, CeMM Research Center for Molecular Medicine of the Austrian Academy of Sciences | Lukas Endler et al |
| EPI_ISL_934434 | Austrian Agency for Health and Food Safety (AGES) | Bergthaler laboratory, CeMM Research Center for Molecular Medicine of the Austrian Academy of Sciences | Lukas Endler et al |
| EPI_ISL_1008182 | Austrian Agency for Health and Food Safety (AGES) | Bergthaler laboratory, CeMM Research Center for Molecular Medicine of the Austrian Academy of Sciences | Lukas Endler et al |

| | | | |
|---|---|---|---|
| EPI_ISL_1008185 | Austrian Agency for Health and Food Safety (AGES) | Bergthaler laboratory, CeMM Research Center for Molecular Medicine of the Austrian Academy of Sciences | Lukas Endler et al |
| EPI_ISL_934667 | Department of Laboratory Medicine, Division of Clinical Virology, University of Medicine, Vienna | Bergthaler laboratory, CeMM Research Center for Molecular Medicine of the Austrian Academy of Sciences | Lukas Endler et al |
| EPI_ISL_934669 | Department of Laboratory Medicine, Division of Clinical Virology, University of Medicine, Vienna | Bergthaler laboratory, CeMM Research Center for Molecular Medicine of the Austrian Academy of Sciences | Lukas Endler et al |
| EPI_ISL_1008235 | Department of Laboratory Medicine, Division of Clinical Virology, University of Medicine, Vienna | Bergthaler laboratory, CeMM Research Center for Molecular Medicine of the Austrian Academy of Sciences | Lukas Endler et al |
| EPI_ISL_1008258 | Department of Laboratory Medicine, Division of Clinical Virology, University of Medicine, Vienna | Bergthaler laboratory, CeMM Research Center for Molecular Medicine of the Austrian Academy of Sciences | Lukas Endler et al |
| EPI_ISL_1008343 | Department of Laboratory Medicine, Division of Clinical Virology, University of Medicine, Vienna | Bergthaler laboratory, CeMM Research Center for Molecular Medicine of the Austrian Academy of Sciences | Lukas Endler et al |
| EPI_ISL_1180828 | Austrian Agency for Health and Food Safety (AGES) | Bergthaler laboratory, CeMM Research Center for Molecular Medicine of the Austrian Academy of Sciences | Lukas Endler et al |
| EPI_ISL_1118072 | Elling group, Institute of Molecular Biotechnology (IMBA) | Bergthaler laboratory, CeMM Research Center for Molecular Medicine of the Austrian Academy of Sciences | Lukas Endler et al |
| EPI_ISL_1117865 | Austrian Agency for Health and Food Safety (AGES) | Bergthaler laboratory, CeMM Research Center for Molecular Medicine of the Austrian Academy of Sciences | Lukas Endler et al |
| EPI_ISL_1180860 | Department of Laboratory Medicine, Division of Clinical Virology, University of Medicine, Vienna | Bergthaler laboratory, CeMM Research Center for Molecular Medicine of the Austrian Academy of Sciences | Lukas Endler et al |
| EPI_ISL_1180878 | Department of Laboratory Medicine, Division of Clinical Virology, University of Medicine, Vienna | Bergthaler laboratory, CeMM Research Center for Molecular Medicine of the Austrian Academy of Sciences | Lukas Endler et al |
| EPI_ISL_1495283 | Institute for Laboratory Diagnostics and Microbiology, Klinikum Klagenfurt am Wörthersee | Bergthaler laboratory, CeMM Research Center for Molecular Medicine of the Austrian Academy of Sciences | Lukas Endler et al |
| EPI_ISL_1495300 | Department of Laboratory Medicine, Division of Clinical Virology, University of Medicine, Vienna | Bergthaler laboratory, CeMM Research Center for Molecular Medicine of the Austrian Academy of Sciences | Lukas Endler et al |
| EPI_ISL_1495576 | Vienna COVID-19 Detection Initiative (VCDI) | Bergthaler laboratory, CeMM Research Center for Molecular Medicine of the Austrian Academy of Sciences | Lukas Endler et al |
| EPI_ISL_1495706 | Austrian Agency for Health and Food Safety (AGES) | Bergthaler laboratory, CeMM Research Center for Molecular Medicine of the Austrian Academy of Sciences | Lukas Endler et al |
| EPI_ISL_1583371 | Austrian Agency for Health and Food Safety (AGES) | Bergthaler laboratory, CeMM Research Center for Molecular Medicine of the Austrian Academy of Sciences | Lukas Endler et al |
| EPI_ISL_1495815 | Department of Laboratory Medicine, Division of Clinical Virology, University of Medicine, Vienna | Bergthaler laboratory, CeMM Research Center for Molecular Medicine of the Austrian Academy of Sciences | Lukas Endler et al |
| EPI_ISL_1583534 | Austrian Agency for Health and Food Safety (AGES) | Bergthaler laboratory, CeMM Research Center for Molecular Medicine of the Austrian Academy of Sciences | Lukas Endler et al |
| EPI_ISL_1671064 | Austrian Agency for Health and Food Safety (AGES) | Bergthaler laboratory, CeMM Research Center for Molecular Medicine of the Austrian Academy of Sciences | Lukas Endler et al |
| EPI_ISL_1671014 | Austrian Agency for Health and Food Safety (AGES) | Bergthaler laboratory, CeMM Research Center for Molecular Medicine of the Austrian Academy of Sciences | Lukas Endler et al |
| EPI_ISL_1670958 | Austrian Agency for Health and Food Safety (AGES) | Bergthaler laboratory, CeMM Research Center for Molecular Medicine of the Austrian Academy of Sciences | Lukas Endler et al |
| EPI_ISL_1671037 | Austrian Agency for Health and Food Safety (AGES) | Bergthaler laboratory, CeMM Research Center for Molecular Medicine of the Austrian Academy of Sciences | Lukas Endler et al |

| | | | |
|---|---|---|---|
| EPI_ISL_1671010 | Austrian Agency for Health and Food Safety (AGES) | Bergthaler laboratory, CeMM Research Center for Molecular Medicine of the Austrian Academy of Sciences | Lukas Endler et al |
| EPI_ISL_1671056 | Austrian Agency for Health and Food Safety (AGES) | Bergthaler laboratory, CeMM Research Center for Molecular Medicine of the Austrian Academy of Sciences | Lukas Endler et al |
| EPI_ISL_1670969 | Austrian Agency for Health and Food Safety (AGES) | Bergthaler laboratory, CeMM Research Center for Molecular Medicine of the Austrian Academy of Sciences | Lukas Endler et al |
| EPI_ISL_1671115 | Austrian Agency for Health and Food Safety (AGES) | Bergthaler laboratory, CeMM Research Center for Molecular Medicine of the Austrian Academy of Sciences | Lukas Endler et al |
| EPI_ISL_1671060 | Austrian Agency for Health and Food Safety (AGES) | Bergthaler laboratory, CeMM Research Center for Molecular Medicine of the Austrian Academy of Sciences | Lukas Endler et al |
| EPI_ISL_1671017 | Austrian Agency for Health and Food Safety (AGES) | Bergthaler laboratory, CeMM Research Center for Molecular Medicine of the Austrian Academy of Sciences | Lukas Endler et al |
| EPI_ISL_1180680 | Public Health Authority of the Slovak Republic | Bergthaler laboratory, CeMM Research Center for Molecular Medicine of the Austrian Academy of Sciences | Lukas Endler et al |
| EPI_ISL_1180691 | Public Health Authority of the Slovak Republic | Bergthaler laboratory, CeMM Research Center for Molecular Medicine of the Austrian Academy of Sciences | Lukas Endler et al |
| EPI_ISL_1180692 | Public Health Authority of the Slovak Republic | Bergthaler laboratory, CeMM Research Center for Molecular Medicine of the Austrian Academy of Sciences | Lukas Endler et al |
| EPI_ISL_1209258 | Public Health Authority of the Slovak Republic | Bergthaler laboratory, CeMM Research Center for Molecular Medicine of the Austrian Academy of Sciences | Lukas Endler et al |
| EPI_ISL_1209260 | Public Health Authority of the Slovak Republic | Bergthaler laboratory, CeMM Research Center for Molecular Medicine of the Austrian Academy of Sciences | Lukas Endler et al |
| EPI_ISL_603118 | Lithuanian University of Health Sciences Hospital, Department of Laboratory Medicine | Lithuanian University of Health Sciences, Molecular cardiology lab. | Lukas Zemaitis et al |
| EPI_ISL_603088 | Lithuanian University of Health Sciences Hospital, Department of Laboratory Medicine | Lithuanian University of Health Sciences, Molecular cardiology lab. | Lukas Zemaitis et al |
| EPI_ISL_1657243 | NVSPL | National Public Health Surveillance Laboratory | Lukas Zemaitis et al |
| EPI_ISL_1657152 | NMVRVI | National Public Health Surveillance Laboratory | Lukas Zemaitis et al |
| EPI_ISL_1655018 | NMVRVI | National Public Health Surveillance Laboratory | Lukas Zemaitis et al |
| EPI_ISL_1655991 | NMVRVI | National Public Health Surveillance Laboratory | Lukas Zemaitis et al |
| EPI_ISL_1580062 | NMVRVI | National Public Health Surveillance Laboratory | Lukas Zemaitis et al |
| EPI_ISL_1580272 | NMVRVI | National Public Health Surveillance Laboratory | Lukas Zemaitis et al |
| EPI_ISL_1656121 | NMVRVI | National Public Health Surveillance Laboratory | Lukas Zemaitis et al |
| EPI_ISL_1580281 | NMVRVI | National Public Health Surveillance Laboratory | Lukas Zemaitis et al |
| EPI_ISL_1580331 | NMVRVI | National Public Health Surveillance Laboratory | Lukas Zemaitis et al |
| EPI_ISL_1579767 | Rezus LT | National Public Health Surveillance Laboratory | Lukas Zemaitis et al |
| EPI_ISL_636604 | Lithuanian University of Health Sciences Hospital, Department of Laboratory Medicine | Lithuanian University of Health Sciences, Molecular cardiology lab. | Lukas Zemaitis et al |
| EPI_ISL_636839 | Lithuanian University of Health Sciences Hospital, Department of Laboratory Medicine | Lithuanian University of Health Sciences, Molecular cardiology lab. | Lukas Zemaitis et al |

| | | | |
|---|---|---|---|
| EPI_ISL_636840 | Lithuanian University of Health Sciences Hospital, Department of Laboratory Medicine | Lithuanian University of Health Sciences, Molecular cardiology lab. | Lukas Zemaitis et al |
| EPI_ISL_636841 | Lithuanian University of Health Sciences Hospital, Department of Laboratory Medicine | Lithuanian University of Health Sciences, Molecular cardiology lab. | Lukas Zemaitis et al |
| EPI_ISL_636842 | Lithuanian University of Health Sciences Hospital, Department of Laboratory Medicine | Lithuanian University of Health Sciences, Molecular cardiology lab. | Lukas Zemaitis et al |
| EPI_ISL_636843 | Lithuanian University of Health Sciences Hospital, Department of Laboratory Medicine | Lithuanian University of Health Sciences, Molecular cardiology lab. | Lukas Zemaitis et al |
| EPI_ISL_636844 | Lithuanian University of Health Sciences Hospital, Department of Laboratory Medicine | Lithuanian University of Health Sciences, Molecular cardiology lab. | Lukas Zemaitis et al |
| EPI_ISL_603119 | Lithuanian University of Health Sciences Hospital, Department of Laboratory Medicine | Lithuanian University of Health Sciences, Molecular cardiology lab. | Lukas Zemaitis et al |
| EPI_ISL_636903 | Lithuanian University of Health Sciences Hospital, Department of Laboratory Medicine | Lithuanian University of Health Sciences, Molecular cardiology lab. | Lukas Zemaitis et al |
| EPI_ISL_636906 | Lithuanian University of Health Sciences Hospital, Department of Laboratory Medicine | Lithuanian University of Health Sciences, Molecular cardiology lab. | Lukas Zemaitis et al |
| EPI_ISL_770499 | Lithuanian University of Health Sciences Hospital, Department of Laboratory Medicine | Lithuanian University of Health Sciences, Molecular cardiology lab. | Lukas Zemaitis et al |
| EPI_ISL_541873 | Lithuanian University of Health Sciences Hospital, Department of Laboratory Medicine | Lithuanian University of Health Sciences, Laboratory of Molecular Cardiology | Lukas Zemaitis et al |
| EPI_ISL_1084608 | Hospital for Infectious Diseases, Molecular Diagnostics Laboratory, Warsaw, Poland | 43. Laboratory of Recombinant Vaccines, Intercollegiate Faculty of Biotechnology University of Gdansk and Medical University of Gdansk, 2. ViroGenetics - BSL3 Laboratory of Virology, Małopolska Centre of Biotechnology, Jagiellonian University | Lukasz Rabalski et al |
| EPI_ISL_906711 | Hematology Laboratory, Section of Molecular Diagnostics, University Clinical Centre, Medical University of Gdansk | Laboratory of Recombinant Vaccines | Lukasz Rabalski et al |
| EPI_ISL_906732 | Hematology Laboratory, Section of Molecular Diagnostics, University Clinical Centre, Medical University of Gdansk | Laboratory of Recombinant Vaccines | Lukasz Rabalski et al |
| EPI_ISL_1229508 | Hematology Laboratory, Section of Molecular Diagnostics, University Clinical Centre, Medical University of Gdansk | 1. Laboratory of Recombinant Vaccines 2. Department of Biology and Medical Genetics 3. Laboratory of Clinical Genetics | Lukasz Rabalski et al |
| EPI_ISL_1229410 | Hematology Laboratory, Section of Molecular Diagnostics, University Clinical Centre, Medical University of Gdansk | Laboratory of Recombinant Vaccines | Lukasz Rabalski et al |
| EPI_ISL_906752 | Hematology Laboratory, Section of Molecular Diagnostics, University Clinical Centre, Medical University of Gdansk | Laboratory of Recombinant Vaccines | Lukasz Rabalski et al |
| EPI_ISL_440499 | Department of Pathology, University of Cambridge | Wellcome Sanger Institute for the COVID-19 Genomics UK (COG-UK) consortium | Luke W Meredith et al |
| EPI_ISL_941944 | Centro de Investigaciones en Microbiología y Biotecnología-UR (CIMBIUR), Facultad de Ciencias Naturales, Universidad del Rosario, Bogotá, Colombia | Centro de Investigaciones en Microbiología y Biotecnología-UR (CIMBIUR), Facultad de Ciencias Naturales, Universidad del Rosario, Bogotá, Colombia Instituto Nacional de Salud, Bogotá, Colombia Icahn School of Medicine at Mount Sinai, New York, USA | Luz Helena Patiño et al |
| EPI_ISL_942004 | Centro de Investigaciones en Microbiología y Biotecnología-UR (CIMBIUR), Facultad de Ciencias Naturales, Universidad del Rosario, Bogotá, Colombia | Centro de Investigaciones en Microbiología y Biotecnología-UR (CIMBIUR), Facultad de Ciencias Naturales, Universidad del Rosario, Bogotá, Colombia Instituto Nacional de Salud, Bogotá, Colombia Icahn School of Medicine at Mount Sinai, New York, USA | Luz Helena Patiño et al |

| | | | |
|---|---|---|---|
| EPI_ISL_941997 | Centro de Investigaciones en Microbiología y Biotecnología-UR (CIMBIUR), Facultad de Ciencias Naturales, Universidad del Rosario, Bogotá, Colombia | Centro de Investigaciones en Microbiología y Biotecnología-UR (CIMBIUR), Facultad de Ciencias Naturales, Universidad del Rosario, Bogotá, Colombia Instituto Nacional de Salud, Bogotá, Colombia Icahn School of Medicine at Mount Sinai, New York, USA | Luz Helena Patiño et al |
| EPI_ISL_941998 | Centro de Investigaciones en Microbiología y Biotecnología-UR (CIMBIUR), Facultad de Ciencias Naturales, Universidad del Rosario, Bogotá, Colombia | Centro de Investigaciones en Microbiología y Biotecnología-UR (CIMBIUR), Facultad de Ciencias Naturales, Universidad del Rosario, Bogotá, Colombia Instituto Nacional de Salud, Bogotá, Colombia Icahn School of Medicine at Mount Sinai, New York, USA | Luz Helena Patiño et al |
| EPI_ISL_941947 | Centro de Investigaciones en Microbiología y Biotecnología-UR (CIMBIUR), Facultad de Ciencias Naturales, Universidad del Rosario, Bogotá, Colombia | Centro de Investigaciones en Microbiología y Biotecnología-UR (CIMBIUR), Facultad de Ciencias Naturales, Universidad del Rosario, Bogotá, Colombia Instituto Nacional de Salud, Bogotá, Colombia Icahn School of Medicine at Mount Sinai, New York, USA | Luz Helena Patiño et al |
| EPI_ISL_941942 | Instituto Nacional de Salud, Bogotá, Colombia | Centro de Investigaciones en Microbiología y Biotecnología-UR (CIMBIUR), Facultad de Ciencias Naturales, Universidad del Rosario, Bogotá, Colombia Instituto Nacional de Salud, Bogotá, Colombia Icahn School of Medicine at Mount Sinai, New York, USA | Luz Helena Patiño et al |
| EPI_ISL_941943 | Instituto Nacional de Salud, Bogotá, Colombia | Centro de Investigaciones en Microbiología y Biotecnología-UR (CIMBIUR), Facultad de Ciencias Naturales, Universidad del Rosario, Bogotá, Colombia Instituto Nacional de Salud, Bogotá, Colombia Icahn School of Medicine at Mount Sinai, New York, USA | Luz Helena Patiño et al |
| EPI_ISL_941994 | Instituto Nacional de Salud, Bogotá, Colombia | Centro de Investigaciones en Microbiología y Biotecnología-UR (CIMBIUR), Facultad de Ciencias Naturales, Universidad del Rosario, Bogotá, Colombia Instituto Nacional de Salud, Bogotá, Colombia Icahn School of Medicine at Mount Sinai, New York, USA | Luz Helena Patiño et al |
| EPI_ISL_941955 | Instituto Nacional de Salud, Bogotá, Colombia | Centro de Investigaciones en Microbiología y Biotecnología-UR (CIMBIUR), Facultad de Ciencias Naturales, Universidad del Rosario, Bogotá, Colombia Instituto Nacional de Salud, Bogotá, Colombia Icahn School of Medicine at Mount Sinai, New York, USA | Luz Helena Patiño et al |
| EPI_ISL_964903 | Laboratorio Central Mg. Luis Alfredo Pianciola on behalf of 'Proyecto Argentino Interinstitucional de genomica de SARS-CoV-2' (PAIS Consortium) | Laboratorio Central Mg. Luis Alfredo Pianciola on behalf of 'Proyecto Argentino Interinstitucional de genomica de SARS-CoV-2' (PAIS Consortium) | M Mazzeo et al |
| EPI_ISL_528835 | CSIR-Centre for Cellular and Molecular Biology | CSIR-Centre for Cellular and Molecular Biology | M Soujanya Reddy et al |
| EPI_ISL_1000972 | Hospital Universitario de Gran Canaria Dr. Negrín | SeqCOVID-SPAIN consortium/IBV(CSIC) | M. Carmen Pérez González et al |
| EPI_ISL_1000974 | Hospital Universitario de Gran Canaria Dr. Negrín | SeqCOVID-SPAIN consortium/IBV(CSIC) | M. Carmen Pérez González et al |
| EPI_ISL_962942 | Hospital Universitario de Gran Canaria Dr. Negrín | SeqCOVID-SPAIN consortium/IBV(CSIC) | M. Carmen Pérez González et al |
| EPI_ISL_962973 | Hospital Universitario de Gran Canaria Dr. Negrín | SeqCOVID-SPAIN consortium/IBV(CSIC) | M. Carmen Pérez González et al |
| EPI_ISL_455472 | Laboratory for Respiratory Viruses, Cantacuzino National Military-Medical Institute for Research and Development | Cantacuzino Institute | M.Lazar et al |
| EPI_ISL_491474 | Research Institute for Tropical Medicine | Research Institute for Tropical Medicine | Ma. Angelica Tujan et al |
| EPI_ISL_512301 | Hematology Laboratory, Section of Molecular Diagnostics, University Clinical Centre, Medical University of Gdansk | Department of Virology, Faculty of Medicine, University of Helsinki, Helsinki, Finland | Maciej Grzybek et al |
| EPI_ISL_512303 | Hematology Laboratory, Section of Molecular Diagnostics, University Clinical Centre, Medical University of Gdansk | Department of Virology, Faculty of Medicine, University of Helsinki, Helsinki, Finland | Maciej Grzybek et al |
| EPI_ISL_1008733 | genXone SA, Molecular Diagnostics Laboratory / NZOZ | genXone SA, Research & Development Laboratory | Maciej Sykulski et al |

| | | | |
|---|---|---|---|
| EPI_ISL_876001 | Laboratory of Molecular Biology, Diagnostyka sp. z o.o. | genXone SA, Research & Development Laboratory | Maciej Sykulski et al |
| EPI_ISL_876010 | Laboratory of Molecular Biology, Diagnostyka sp. z o.o. | genXone SA, Research & Development Laboratory | Maciej Sykulski et al |
| EPI_ISL_925041 | Laboratory of Molecular Biology, Diagnostyka sp. z o.o. | genXone SA, Research & Development Laboratory | Maciej Sykulski et al |
| EPI_ISL_876015 | Laboratory of Molecular Biology, Diagnostyka sp. z o.o. | genXone SA, Research & Development Laboratory | Maciej Sykulski et al |
| EPI_ISL_1295181 | Laboratory Diagnostics and Clinical Immunology of Developmental Age, Medical University of Warsaw | genXone SA, Research & Development Laboratory; The Faculty of Mathematics, Informatics and Mechanics of the University of Warsaw | Maciej Sykulski et al |
| EPI_ISL_754904 | Laboratory Diagnostics and Clinical Immunology of Developmental Age, Medical University of Warsaw | genXone SA, Research & Development Laboratory; The Faculty of Mathematics, Informatics and Mechanics of the University of Warsaw | Maciej Sykulski et al |
| EPI_ISL_754906 | Laboratory Diagnostics and Clinical Immunology of Developmental Age, Medical University of Warsaw | genXone SA, Research & Development Laboratory; The Faculty of Mathematics, Informatics and Mechanics of the University of Warsaw | Maciej Sykulski et al |
| EPI_ISL_754907 | Laboratory Diagnostics and Clinical Immunology of Developmental Age, Medical University of Warsaw | genXone SA, Research & Development Laboratory; The Faculty of Mathematics, Informatics and Mechanics of the University of Warsaw | Maciej Sykulski et al |
| EPI_ISL_754913 | Laboratory Diagnostics and Clinical Immunology of Developmental Age, Medical University of Warsaw | genXone SA, Research & Development Laboratory; The Faculty of Mathematics, Informatics and Mechanics of the University of Warsaw | Maciej Sykulski et al |
| EPI_ISL_754909 | Laboratory Diagnostics and Clinical Immunology of Developmental Age, Medical University of Warsaw | genXone SA, Research & Development Laboratory; The Faculty of Mathematics, Informatics and Mechanics of the University of Warsaw | Maciej Sykulski et al |
| EPI_ISL_754910 | Laboratory Diagnostics and Clinical Immunology of Developmental Age, Medical University of Warsaw | genXone SA, Research & Development Laboratory; The Faculty of Mathematics, Informatics and Mechanics of the University of Warsaw | Maciej Sykulski et al |
| EPI_ISL_754902 | Laboratory Diagnostics and Clinical Immunology of Developmental Age, Medical University of Warsaw | genXone SA, Research & Development Laboratory; The Faculty of Mathematics, Informatics and Mechanics of the University of Warsaw | Maciej Sykulski et al |
| EPI_ISL_754908 | Laboratory Diagnostics and Clinical Immunology of Developmental Age, Medical University of Warsaw | genXone SA, Research & Development Laboratory; The Faculty of Mathematics, Informatics and Mechanics of the University of Warsaw | Maciej Sykulski et al |
| EPI_ISL_527918 | University Hospital Basel, Clinical Virology | University Hospital Basel, Clinical Bacteriology | Madlen Stange et al |
| EPI_ISL_527919 | University Hospital Basel, Clinical Virology | University Hospital Basel, Clinical Bacteriology | Madlen Stange et al |
| EPI_ISL_528291 | University Hospital Basel, Clinical Virology | University Hospital Basel, Clinical Bacteriology | Madlen Stange et al |
| EPI_ISL_463749 | Pasteur Institute of Iran | Rapid Response Team | Mahboobeh Rafigh et al |
| EPI_ISL_636765 | National Centre for Disease control (NCDC) | NCDC/CSIR-IGIB | Mahesh S. Dhar1* et al |
| EPI_ISL_949180 | Laboratory of Virology and Molecular Diagnostics | Institute of Public Health of Republic of North Macedonia Laboratory of Virology and Molecular Diagnostics | Maja Kuzmanovska et al |
| EPI_ISL_418993 | National Public Health Laboratory, National Centre for Infectious Diseases | National Public Health Laboratory, National Centre for Infectious Diseases | Mak TM et al |
| EPI_ISL_462318 | National Public Health Laboratory, National Centre for Infectious Diseases | National Public Health Laboratory, National Centre for Infectious Diseases | Mak TM et al |
| EPI_ISL_462342 | National Public Health Laboratory, National Centre for Infectious Diseases | National Public Health Laboratory, National Centre for Infectious Diseases | Mak TM et al |
| EPI_ISL_462346 | National Public Health Laboratory, National Centre for Infectious Diseases | National Public Health Laboratory, National Centre for Infectious Diseases | Mak TM et al |
| EPI_ISL_469122 | National Public Health Laboratory, National Centre for Infectious Diseases | National Public Health Laboratory, National Centre for Infectious Diseases | Mak TM et al |
| EPI_ISL_475938 | National Public Health Laboratory, National Centre for Infectious Diseases | National Public Health Laboratory, National Centre for Infectious Diseases | Mak TM et al |
| EPI_ISL_475940 | National Public Health Laboratory, National Centre for Infectious Diseases | National Public Health Laboratory, National Centre for Infectious Diseases | Mak TM et al |
| EPI_ISL_475948 | National Public Health Laboratory, National Centre for Infectious Diseases | National Public Health Laboratory, National Centre for Infectious Diseases | Mak TM et al |

| | | | |
|---|---|---|---|
| EPI_ISL_475952 | National Public Health Laboratory, National Centre for Infectious Diseases | National Public Health Laboratory, National Centre for Infectious Diseases | Mak TM et al |
| EPI_ISL_475997 | National Public Health Laboratory, National Centre for Infectious Diseases | National Public Health Laboratory, National Centre for Infectious Diseases | Mak TM et al |
| EPI_ISL_475999 | National Public Health Laboratory, National Centre for Infectious Diseases | National Public Health Laboratory, National Centre for Infectious Diseases | Mak TM et al |
| EPI_ISL_479591 | National Public Health Laboratory, National Centre for Infectious Diseases | National Public Health Laboratory, National Centre for Infectious Diseases | Mak TM et al |
| EPI_ISL_490067 | National Public Health Laboratory, National Centre for Infectious Diseases | National Public Health Laboratory, National Centre for Infectious Diseases | Mak TM et al |
| EPI_ISL_493390 | National Public Health Laboratory, National Centre for Infectious Diseases | National Public Health Laboratory, National Centre for Infectious Diseases | Mak TM et al |
| EPI_ISL_498599 | National Public Health Laboratory, National Centre for Infectious Diseases | National Public Health Laboratory, National Centre for Infectious Diseases | Mak TM et al |
| EPI_ISL_516815 | National Public Health Laboratory, National Centre for Infectious Diseases | National Public Health Laboratory, National Centre for Infectious Diseases | Mak TM et al |
| EPI_ISL_516819 | National Public Health Laboratory, National Centre for Infectious Diseases | National Public Health Laboratory, National Centre for Infectious Diseases | Mak TM et al |
| EPI_ISL_516824 | National Public Health Laboratory, National Centre for Infectious Diseases | National Public Health Laboratory, National Centre for Infectious Diseases | Mak TM et al |
| EPI_ISL_536431 | National Public Health Laboratory, National Centre for Infectious Diseases | National Public Health Laboratory, National Centre for Infectious Diseases | Mak TM et al |
| EPI_ISL_536435 | National Public Health Laboratory, National Centre for Infectious Diseases | National Public Health Laboratory, National Centre for Infectious Diseases | Mak TM et al |
| EPI_ISL_536445 | National Public Health Laboratory, National Centre for Infectious Diseases | National Public Health Laboratory, National Centre for Infectious Diseases | Mak TM et al |
| EPI_ISL_536447 | National Public Health Laboratory, National Centre for Infectious Diseases | National Public Health Laboratory, National Centre for Infectious Diseases | Mak TM et al |
| EPI_ISL_536451 | National Public Health Laboratory, National Centre for Infectious Diseases | National Public Health Laboratory, National Centre for Infectious Diseases | Mak TM et al |
| EPI_ISL_443187 | National Virology Reference Laboratory | National Public Health Laboratory, National Centre for Infectious Diseases | Mak Tze Minn et al |
| EPI_ISL_435674 | National Virology Reference Laboratory | National Public Health Laboratory, National Centre for Infectious Diseases | Mak Tze Minn et al |
| EPI_ISL_435677 | National Virology Reference Laboratory | National Public Health Laboratory, National Centre for Infectious Diseases | Mak Tze Minn et al |
| EPI_ISL_435686 | National Public Health Laboratory, National Centre for Infectious Diseases | National Public Health Laboratory, National Centre for Infectious Diseases | Mak Tze Minn et al |
| EPI_ISL_1196017 | Manzini RRT | National Institute for Communicable Diseases of the National Health Laboratory Service | Maphalala GP et al |
| EPI_ISL_1196002 | Women correctional | National Institute for Communicable Diseases of the National Health Laboratory Service | Maphalala GP et al |
| EPI_ISL_1196003 | Homecare | National Institute for Communicable Diseases of the National Health Laboratory Service | Maphalala GP et al |
| EPI_ISL_1196004 | Mbabane PHU | National Institute for Communicable Diseases of the National Health Laboratory Service | Maphalala GP et al |
| EPI_ISL_1196005 | RSP Clinic | National Institute for Communicable Diseases of the National Health Laboratory Service | Maphalala GP et al |
| EPI_ISL_1196006 | TLC Clinic | National Institute for Communicable Diseases of the National Health Laboratory Service | Maphalala GP et al |
| EPI_ISL_1196008 | Nhlangano Health Centre | National Institute for Communicable Diseases of the National Health Laboratory Service | Maphalala GP et al |
| EPI_ISL_1196009 | Nhlangano Health Centre | National Institute for Communicable Diseases of the National Health Laboratory Service | Maphalala GP et al |

| | | | |
|---|---|---|---|
| EPI_ISL_1196010 | Nhlangano Health Centre | National Institute for Communicable Diseases of the National Health Laboratory Service | Maphalala GP et al |
| EPI_ISL_1196011 | Homecare | National Institute for Communicable Diseases of the National Health Laboratory Service | Maphalala GP et al |
| EPI_ISL_1196012 | Homecare | National Institute for Communicable Diseases of the National Health Laboratory Service | Maphalala GP et al |
| EPI_ISL_1196013 | Hlathikhulu Government Hospital | National Institute for Communicable Diseases of the National Health Laboratory Service | Maphalala GP et al |
| EPI_ISL_1196014 | Homecare | National Institute for Communicable Diseases of the National Health Laboratory Service | Maphalala GP et al |
| EPI_ISL_1595918 | NSTU COVID-19 Diagnostic Center | NSU Genome Research Institute (NGRI) | Maqsud Hossain et al |
| EPI_ISL_1626485 | NSTU COVID-19 Diagnostic Center | NSU Genome Research Institute (NGRI) | Maqsud Hossain et al |
| EPI_ISL_1626488 | NSTU COVID-19 Diagnostic Center | NSU Genome Research Institute (NGRI) | Maqsud Hossain et al |
| EPI_ISL_1626489 | NSTU COVID-19 Diagnostic Center | NSU Genome Research Institute (NGRI) | Maqsud Hossain et al |
| EPI_ISL_1626490 | NSTU COVID-19 Diagnostic Center | NSU Genome Research Institute (NGRI) | Maqsud Hossain et al |
| EPI_ISL_1626495 | NSTU COVID-19 Diagnostic Center | NSU Genome Research Institute (NGRI) | Maqsud Hossain et al |
| EPI_ISL_1626502 | NSTU COVID-19 Diagnostic Center | NSU Genome Research Institute (NGRI) | Maqsud Hossain et al |
| EPI_ISL_1626509 | NSTU COVID-19 Diagnostic Center | NSU Genome Research Institute (NGRI) | Maqsud Hossain et al |
| EPI_ISL_1626514 | NSTU COVID-19 Diagnostic Center | NSU Genome Research Institute (NGRI) | Maqsud Hossain et al |
| EPI_ISL_1626517 | NSTU COVID-19 Diagnostic Center | NSU Genome Research Institute (NGRI) | Maqsud Hossain et al |
| EPI_ISL_1626519 | NSTU COVID-19 Diagnostic Center | NSU Genome Research Institute (NGRI) | Maqsud Hossain et al |
| EPI_ISL_1626524 | NSTU COVID-19 Diagnostic Center | NSU Genome Research Institute (NGRI) | Maqsud Hossain et al |
| EPI_ISL_1208770 | Microbiology Department, Laboratori Clínic Metropolitana Nord. Hospital Universitari Germans Trias i Pujol. | Can Ruti SARS-CoV-2 Sequencing Hub (HUGTiP/IrsiCaixa/IGTP) | Marc Noguera-Julian et al |
| EPI_ISL_1583556 | Microbiology Department, Laboratori Clínic Metropolitana Nord. Hospital Universitari Germans Trias i Pujol. | Can Ruti SARS-CoV-2 Sequencing Hub (HUGTiP/IrsiCaixa/IGTP) | Marc Noguera-Julian et al |
| EPI_ISL_1583601 | Microbiology Department, Laboratori Clínic Metropolitana Nord. Hospital Universitari Germans Trias i Pujol. | Can Ruti SARS-CoV-2 Sequencing Hub (HUGTiP/IrsiCaixa/IGTP) | Marc Noguera-Julian et al |
| EPI_ISL_1583612 | Microbiology Department, Laboratori Clínic Metropolitana Nord. Hospital Universitari Germans Trias i Pujol. | Can Ruti SARS-CoV-2 Sequencing Hub (HUGTiP/IrsiCaixa/IGTP) | Marc Noguera-Julian et al |
| EPI_ISL_1661933 | Microbiology Department, Laboratori Clínic Metropolitana Nord. Hospital Universitari Germans Trias i Pujol. | Can Ruti SARS-CoV-2 Sequencing Hub (HUGTiP/IrsiCaixa/IGTP) | Marc Noguera-Julian et al |
| EPI_ISL_1661943 | Microbiology Department, Laboratori Clínic Metropolitana Nord. Hospital Universitari Germans Trias i Pujol. | Can Ruti SARS-CoV-2 Sequencing Hub (HUGTiP/IrsiCaixa/IGTP) | Marc Noguera-Julian et al |
| EPI_ISL_1661956 | Microbiology Department, Laboratori Clínic Metropolitana Nord. Hospital Universitari Germans Trias i Pujol. | Can Ruti SARS-CoV-2 Sequencing Hub (HUGTiP/IrsiCaixa/IGTP) | Marc Noguera-Julian et al |
| EPI_ISL_1547769 | Microbiology Department, Laboratori Clínic Metropolitana Nord. Hospital Universitari Germans Trias i Pujol. | Can Ruti SARS-CoV-2 Sequencing Hub (HUGTiP/IrsiCaixa/IGTP) | Marc Noguera-Julian et al |

| | | | |
|---|---|---|---|
| EPI_ISL_1636455 | Microbiology Department, Laboratori Clínic Metropolitana Nord. Hospital Universitari Germans Trias i Pujol. | Can Ruti SARS-CoV-2 Sequencing Hub (HUGTiP/IrsiCaixa/IGTP) | Marc Noguera-Julian et al |
| EPI_ISL_832103 | Hospital Universitari Germans Trias i Pujol(HUGTiP)/Fundació Lluita contra la SIDA (FLSida) | IrsiCaixa AIDS Research Lab | Marc Noguera-Julian et al |
| EPI_ISL_1239012 | Laboratório Central de Saúde Pública do Estado de Pernambuco (LACEN-PE) | WallauLab, Aggeu Magalhaes Institute | Marcelo Henrique dos Santos Paiva et al |
| EPI_ISL_1239013 | Laboratório Central de Saúde Pública do Estado de Pernambuco (LACEN-PE) | WallauLab, Aggeu Magalhaes Institute | Marcelo Henrique dos Santos Paiva et al |
| EPI_ISL_1239015 | Laboratório Central de Saúde Pública do Estado de Pernambuco (LACEN-PE) | WallauLab, Aggeu Magalhaes Institute | Marcelo Henrique dos Santos Paiva et al |
| EPI_ISL_572373 | Laboratório Central de Saúde Pública do Estado de Pernambuco (LACEN-PE) | WallauLab, Aggeu Magalhaes Institute | Marcelo Henrique Santos Paiva et al |
| EPI_ISL_572374 | Laboratório Central de Saúde Pública do Estado de Pernambuco (LACEN-PE) | WallauLab, Aggeu Magalhaes Institute | Marcelo Henrique Santos Paiva et al |
| EPI_ISL_572375 | Laboratório Central de Saúde Pública do Estado de Pernambuco (LACEN-PE) | WallauLab, Aggeu Magalhaes Institute | Marcelo Henrique Santos Paiva et al |
| EPI_ISL_572379 | Laboratório Central de Saúde Pública do Estado de Pernambuco (LACEN-PE) | WallauLab, Aggeu Magalhaes Institute | Marcelo Henrique Santos Paiva et al |
| EPI_ISL_572380 | Laboratório Central de Saúde Pública do Estado de Pernambuco (LACEN-PE) | WallauLab, Aggeu Magalhaes Institute | Marcelo Henrique Santos Paiva et al |
| EPI_ISL_572381 | Laboratório Central de Saúde Pública do Estado de Pernambuco (LACEN-PE) | WallauLab, Aggeu Magalhaes Institute | Marcelo Henrique Santos Paiva et al |
| EPI_ISL_572382 | Laboratório Central de Saúde Pública do Estado de Pernambuco (LACEN-PE) | WallauLab, Aggeu Magalhaes Institute | Marcelo Henrique Santos Paiva et al |
| EPI_ISL_572384 | Laboratório Central de Saúde Pública do Estado de Pernambuco (LACEN-PE) | WallauLab, Aggeu Magalhaes Institute | Marcelo Henrique Santos Paiva et al |
| EPI_ISL_572385 | Laboratório Central de Saúde Pública do Estado de Pernambuco (LACEN-PE) | WallauLab, Aggeu Magalhaes Institute | Marcelo Henrique Santos Paiva et al |
| EPI_ISL_572386 | Laboratório Central de Saúde Pública do Estado de Pernambuco (LACEN-PE) | WallauLab, Aggeu Magalhaes Institute | Marcelo Henrique Santos Paiva et al |
| EPI_ISL_572388 | Laboratório Central de Saúde Pública do Estado de Pernambuco (LACEN-PE) | WallauLab, Aggeu Magalhaes Institute | Marcelo Henrique Santos Paiva et al |
| EPI_ISL_500463 | Laboratório Central de Saúde Pública do Estado de Pernambuco (LACEN-PE) | WallauLab, Aggeu Magalhaes Institute | Marcelo Henrique Santos Paiva et al |
| EPI_ISL_572394 | Laboratório Central de Saúde Pública do Estado de Pernambuco (LACEN-PE) | WallauLab, Aggeu Magalhaes Institute | Marcelo Henrique Santos Paiva et al |
| EPI_ISL_572396 | Laboratório Central de Saúde Pública do Estado de Pernambuco (LACEN-PE) | WallauLab, Aggeu Magalhaes Institute | Marcelo Henrique Santos Paiva et al |
| EPI_ISL_500465 | Laboratório Central de Saúde Pública do Estado de Pernambuco (LACEN-PE) | WallauLab, Aggeu Magalhaes Institute | Marcelo Henrique Santos Paiva et al |
| EPI_ISL_500867 | Laboratório Central de Saúde Pública do Estado de Pernambuco (LACEN-PE) | WallauLab, Aggeu Magalhaes Institute | Marcelo Henrique Santos Paiva et al |
| EPI_ISL_500467 | Laboratório Central de Saúde Pública do Estado de Pernambuco (LACEN-PE) | WallauLab, Aggeu Magalhaes Institute | Marcelo Henrique Santos Paiva et al |
| EPI_ISL_500873 | Laboratório Central de Saúde Pública do Estado de Pernambuco (LACEN-PE) | WallauLab, Aggeu Magalhaes Institute | Marcelo Henrique Santos Paiva et al |
| EPI_ISL_500479 | Laboratório Central de Saúde Pública do Estado de Pernambuco (LACEN-PE) | WallauLab, Aggeu Magalhaes Institute | Marcelo Henrique Santos Paiva et al |
| EPI_ISL_500875 | Laboratório Central de Saúde Pública do Estado de Pernambuco (LACEN-PE) | WallauLab, Aggeu Magalhaes Institute | Marcelo Henrique Santos Paiva et al |
| EPI_ISL_572336 | Laboratório Central de Saúde Pública do Estado de Pernambuco (LACEN-PE) | WallauLab, Aggeu Magalhaes Institute | Marcelo Henrique Santos Paiva et al |
| EPI_ISL_500481 | Laboratório Central de Saúde Pública do Estado de Pernambuco (LACEN-PE) | WallauLab, Aggeu Magalhaes Institute | Marcelo Henrique Santos Paiva et al |
| EPI_ISL_572337 | Laboratório Central de Saúde Pública do Estado de Pernambuco (LACEN-PE) | WallauLab, Aggeu Magalhaes Institute | Marcelo Henrique Santos Paiva et al |
| EPI_ISL_572338 | Laboratório Central de Saúde Pública do Estado de Pernambuco (LACEN-PE) | WallauLab, Aggeu Magalhaes Institute | Marcelo Henrique Santos Paiva et al |
| EPI_ISL_572349 | Laboratório Central de Saúde Pública do Estado de Pernambuco (LACEN-PE) | WallauLab, Aggeu Magalhaes Institute | Marcelo Henrique Santos Paiva et al |

| | | | |
|---|---|---|---|
| EPI_ISL_500483 | Laboratório Central de Saúde Pública do Estado de Pernambuco (LACEN-PE) | WallauLab, Aggeu Magalhaes Institute | Marcelo Henrique Santos Paiva et al |
| EPI_ISL_572351 | Laboratório Central de Saúde Pública do Estado de Pernambuco (LACEN-PE) | WallauLab, Aggeu Magalhaes Institute | Marcelo Henrique Santos Paiva et al |
| EPI_ISL_572358 | Laboratório Central de Saúde Pública do Estado de Pernambuco (LACEN-PE) | WallauLab, Aggeu Magalhaes Institute | Marcelo Henrique Santos Paiva et al |
| EPI_ISL_572359 | Laboratório Central de Saúde Pública do Estado de Pernambuco (LACEN-PE) | WallauLab, Aggeu Magalhaes Institute | Marcelo Henrique Santos Paiva et al |
| EPI_ISL_572360 | Laboratório Central de Saúde Pública do Estado de Pernambuco (LACEN-PE) | WallauLab, Aggeu Magalhaes Institute | Marcelo Henrique Santos Paiva et al |
| EPI_ISL_572361 | Laboratório Central de Saúde Pública do Estado de Pernambuco (LACEN-PE) | WallauLab, Aggeu Magalhaes Institute | Marcelo Henrique Santos Paiva et al |
| EPI_ISL_572362 | Laboratório Central de Saúde Pública do Estado de Pernambuco (LACEN-PE) | WallauLab, Aggeu Magalhaes Institute | Marcelo Henrique Santos Paiva et al |
| EPI_ISL_572363 | Laboratório Central de Saúde Pública do Estado de Pernambuco (LACEN-PE) | WallauLab, Aggeu Magalhaes Institute | Marcelo Henrique Santos Paiva et al |
| EPI_ISL_572366 | Laboratório Central de Saúde Pública do Estado de Pernambuco (LACEN-PE) | WallauLab, Aggeu Magalhaes Institute | Marcelo Henrique Santos Paiva et al |
| EPI_ISL_572367 | Laboratório Central de Saúde Pública do Estado de Pernambuco (LACEN-PE) | WallauLab, Aggeu Magalhaes Institute | Marcelo Henrique Santos Paiva et al |
| EPI_ISL_572371 | Laboratório Central de Saúde Pública do Estado de Pernambuco (LACEN-PE) | WallauLab, Aggeu Magalhaes Institute | Marcelo Henrique Santos Paiva et al |
| EPI_ISL_572372 | Laboratório Central de Saúde Pública do Estado de Pernambuco (LACEN-PE) | WallauLab, Aggeu Magalhaes Institute | Marcelo Henrique Santos Paiva et al |
| EPI_ISL_451971 | 1. ViroGenetics - BSL3 Laboratory of Virology, Małopolska Centre of Biotechnology, Jagiellonian University; 2. II Department of Internal Medicine, Faculty of Medicine, Jagiellonian University Medical College; 3. DIAGNOSTYKA Ltd. | 1. ViroGenetics - BSL3 Laboratory of Virology, Małopolska Centre of Biotechnology, Jagiellonian University; 2. II Department of Internal Medicine, Faculty of Medicine, Jagiellonian University Medical College. | Marek Sanak et al |
| EPI_ISL_451973 | 1. ViroGenetics - BSL3 Laboratory of Virology, Małopolska Centre of Biotechnology, Jagiellonian University; 2. II Department of Internal Medicine, Faculty of Medicine, Jagiellonian University Medical College; 3. DIAGNOSTYKA Ltd. | 1. ViroGenetics - BSL3 Laboratory of Virology, Małopolska Centre of Biotechnology, Jagiellonian University; 2. II Department of Internal Medicine, Faculty of Medicine, Jagiellonian University Medical College. | Marek Sanak et al |
| EPI_ISL_451975 | 1. ViroGenetics - BSL3 Laboratory of Virology, Małopolska Centre of Biotechnology, Jagiellonian University; 2. II Department of Internal Medicine, Faculty of Medicine, Jagiellonian University Medical College; 3. DIAGNOSTYKA Ltd. | 1. ViroGenetics - BSL3 Laboratory of Virology, Małopolska Centre of Biotechnology, Jagiellonian University; 2. II Department of Internal Medicine, Faculty of Medicine, Jagiellonian University Medical College. | Marek Sanak et al |
| EPI_ISL_451980 | 1. ViroGenetics - BSL3 Laboratory of Virology, Małopolska Centre of Biotechnology, Jagiellonian University; 2. II Department of Internal Medicine, Faculty of Medicine, Jagiellonian University Medical College; 3. DIAGNOSTYKA Ltd. | 1. ViroGenetics - BSL3 Laboratory of Virology, Małopolska Centre of Biotechnology, Jagiellonian University; 2. II Department of Internal Medicine, Faculty of Medicine, Jagiellonian University Medical College. | Marek Sanak et al |
| EPI_ISL_451981 | 1. ViroGenetics - BSL3 Laboratory of Virology, Małopolska Centre of Biotechnology, Jagiellonian University; 2. II Department of Internal Medicine, Faculty of Medicine, Jagiellonian University Medical College; 3. DIAGNOSTYKA Ltd. | 1. ViroGenetics - BSL3 Laboratory of Virology, Małopolska Centre of Biotechnology, Jagiellonian University; 2. II Department of Internal Medicine, Faculty of Medicine, Jagiellonian University Medical College. | Marek Sanak et al |
| EPI_ISL_451982 | 1. ViroGenetics - BSL3 Laboratory of Virology, Małopolska Centre of Biotechnology, Jagiellonian University; 2. II Department of Internal Medicine, Faculty of Medicine, Jagiellonian University Medical College; 3. DIAGNOSTYKA Ltd. | 1. ViroGenetics - BSL3 Laboratory of Virology, Małopolska Centre of Biotechnology, Jagiellonian University; 2. II Department of Internal Medicine, Faculty of Medicine, Jagiellonian University Medical College. | Marek Sanak et al |
| EPI_ISL_451984 | 1. ViroGenetics - BSL3 Laboratory of Virology, Małopolska Centre of Biotechnology, Jagiellonian University; 2. II Department of Internal Medicine, Faculty of Medicine, Jagiellonian University Medical College; 3. DIAGNOSTYKA Ltd. | 1. ViroGenetics - BSL3 Laboratory of Virology, Małopolska Centre of Biotechnology, Jagiellonian University; 2. II Department of Internal Medicine, Faculty of Medicine, Jagiellonian University Medical College. | Marek Sanak et al |

| | | | |
|---|---|---|---|
| EPI_ISL_451985 | 1. ViroGenetics - BSL3 Laboratory of Virology, Małopolska Centre of Biotechnology, Jagiellonian University; 2. II Department of Internal Medicine, Faculty of Medicine, Jagiellonian University Medical College; 3. DIAGNOSTYKA Ltd. | 1. ViroGenetics - BSL3 Laboratory of Virology, Małopolska Centre of Biotechnology, Jagiellonian University; 2. II Department of Internal Medicine, Faculty of Medicine, Jagiellonian University Medical College. | Marek Sanak et al |
| EPI_ISL_451986 | 1. ViroGenetics - BSL3 Laboratory of Virology, Małopolska Centre of Biotechnology, Jagiellonian University; 2. II Department of Internal Medicine, Faculty of Medicine, Jagiellonian University Medical College; 3. DIAGNOSTYKA Ltd. | 1. ViroGenetics - BSL3 Laboratory of Virology, Małopolska Centre of Biotechnology, Jagiellonian University; 2. II Department of Internal Medicine, Faculty of Medicine, Jagiellonian University Medical College. | Marek Sanak et al |
| EPI_ISL_430339 | NYU Langone Health | Departments of Pathology and Medicine, New York University School of Medicine | Maria Aguero-Rosenfeld et al |
| EPI_ISL_476838 | National Influenza Centre for Northern Greece | National Influenza Centre for Northern Greece | Maria Christoforidi et al |
| EPI_ISL_1550388 | Instituto Oswaldo Cruz | Laboratorio de Virologia, Faculdade de Medicina, Universidade Federal de Mato Grosso, campus Cuiabá | María de Fátima Ferreira et al |
| EPI_ISL_1395830 | Inmunología del Hospital Perrando e Instituto de Medicina Regional de la UNNE | Grupo de Genómica y Bioinformática del Instituto de Investigación de la Cadena Láctea CONICET-INTA on behalf of 'Proyecto Argentino Interinstitucional de genomica de SARS-CoV-2' (PAIS Consortium) | María Delia Foussal et al |
| EPI_ISL_1335887 | North Sumatera Health Office | Faculty of Medicine, Universitas Sumatera Utara; Institute of Tropical Disease, Universitas Airlangga | Maria I Lusida et al |
| EPI_ISL_614301 | Faroese National Reference Laboratory for Fish and Animal Diseases | Faroese National Reference Laboratory for Fish and Animal Diseases | Maria Marjunardóttir Dahl et al |
| EPI_ISL_614302 | Faroese National Reference Laboratory for Fish and Animal Diseases | Faroese National Reference Laboratory for Fish and Animal Diseases | Maria Marjunardóttir Dahl et al |
| EPI_ISL_614305 | Faroese National Reference Laboratory for Fish and Animal Diseases | Faroese National Reference Laboratory for Fish and Animal Diseases | Maria Marjunardóttir Dahl et al |
| EPI_ISL_614306 | Faroese National Reference Laboratory for Fish and Animal Diseases | Faroese National Reference Laboratory for Fish and Animal Diseases | Maria Marjunardóttir Dahl et al |
| EPI_ISL_854594 | Faroese National Reference Laboratory for Fish and Animal Diseases | Faroese National Reference Laboratory for Fish and Animal Diseases | Maria Marjunardóttir Dahl et al |
| EPI_ISL_455107 | Narhalsan Mellerud | The Public Health Agency of Sweden | Maria Nykvist et al |
| EPI_ISL_410546 | INMI Lazzaro Spallanzani IRCCS | Laboratory of Virology, INMI Lazzaro Spallanzani IRCCS | Maria R. Capobianchi et al |
| EPI_ISL_410545 | INMI Lazzaro Spallanzani IRCCS | Laboratory of Virology, INMI Lazzaro Spallanzani IRCCS | Maria R. Capobianchi et al |
| EPI_ISL_983221 | University Health Network/Mount Sinai Hospital Department of Microbiology | Ontario Institute for Cancer Research | Marie-Ming Aynaud et al |
| EPI_ISL_1511399 | Laboratorio de Patologia Clinica - UNICAMP | Laboratorio de Estudos de Virus Emergentes - UNICAMP | Mariene R. Amorim et al |
| EPI_ISL_1404193 | Lab voor klinische biologie | Lab voor klinische biologie | Marija Janevska et al |
| EPI_ISL_420142 | Department for Virology, Molecular Biology and Genome Research, R. G. Lugar Center for Public Health Research, National Center for Disease Control and Public Health (NCDC) of Georgia. | Department for Virology, Molecular Biology and Genome Research, R. G. Lugar Center for Public Health Research, National Center for Disease Control and Public Health (NCDC) of Georgia. | Marine Murtskhvaladze et al |
| EPI_ISL_416477 | R. G. Lugar Center for Public Health Research, National Center for Disease Control and Public Health (NCDC) of Georgia. | R. G. Lugar Center for Public Health Research, National Center for Disease Control and Public Health (NCDC) of Georgia. | Marine Murtskhvaladze et al |
| EPI_ISL_1583049 | Hospital | National Reference Center for Viruses of Respiratory Infections, Institut Pasteur, Paris | Marion Barbet et al |
| EPI_ISL_832144 | Hospital | National Reference Center for Viruses of Respiratory Infections, Institut Pasteur, Paris | Marion Barbet et al |
| EPI_ISL_1594450 | Labo Analyses Med | National Reference Center for Viruses of Respiratory Infections, Institut Pasteur, Paris | Marion Barbet et al |

| | | | |
|---|---|---|---|
| EPI_ISL_890353 | Labo Analyses Med | National Reference Center for Viruses of Respiratory Infections, Institut Pasteur, Paris | Marion Barbet et al |
| EPI_ISL_1060587 | Outre mer | National Reference Center for Viruses of Respiratory Infections, Institut Pasteur, Paris | Marion Barbet et al |
| EPI_ISL_1060591 | Outre mer | National Reference Center for Viruses of Respiratory Infections, Institut Pasteur, Paris | Marion Barbet et al |
| EPI_ISL_1096262 | Outre mer | National Reference Center for Viruses of Respiratory Infections, Institut Pasteur, Paris | Marion Barbet et al |
| EPI_ISL_1201532 | Outre Mer | National Reference Center for Viruses of Respiratory Infections, Institut Pasteur, Paris | Marion Barbet et al |
| EPI_ISL_1381215 | Hospital | National Reference Center for Viruses of Respiratory Infections, Institut Pasteur, Paris | Marion Barbet et al |
| EPI_ISL_613420 | Institut Pasteur de la Guadeloupe | Institut Pasteur de la Guadeloupe | Marion Barbet et al |
| EPI_ISL_613422 | Institut Pasteur de la Guadeloupe | Institut Pasteur de la Guadeloupe | Marion Barbet et al |
| EPI_ISL_613425 | Institut Pasteur de la Guadeloupe | Institut Pasteur de la Guadeloupe | Marion Barbet et al |
| EPI_ISL_613436 | Institut Pasteur de la Guadeloupe | Institut Pasteur de la Guadeloupe | Marion Barbet et al |
| EPI_ISL_613441 | Institut Pasteur de la Guadeloupe | Institut Pasteur de la Guadeloupe | Marion Barbet et al |
| EPI_ISL_613442 | Institut Pasteur de la Guadeloupe | Institut Pasteur de la Guadeloupe | Marion Barbet et al |
| EPI_ISL_613445 | Institut Pasteur de la Guadeloupe | Institut Pasteur de la Guadeloupe | Marion Barbet et al |
| EPI_ISL_1013418 | Institut Pasteur de Guadeloupe | National Reference Center for Viruses of Respiratory Infections, Institut Pasteur, Paris | Marion Barbet et al |
| EPI_ISL_1013425 | Institut Pasteur de Guadeloupe | National Reference Center for Viruses of Respiratory Infections, Institut Pasteur, Paris | Marion Barbet et al |
| EPI_ISL_1013426 | Institut Pasteur de Guadeloupe | National Reference Center for Viruses of Respiratory Infections, Institut Pasteur, Paris | Marion Barbet et al |
| EPI_ISL_1443892 | Outre mer | National Reference Center for Viruses of Respiratory Infections, Institut Pasteur, Paris | Marion Barbet et al |
| EPI_ISL_1416969 | Outre mer | National Reference Center for Viruses of Respiratory Infections, Institut Pasteur, Paris | Marion Barbet et al |
| EPI_ISL_1416970 | Outre mer | National Reference Center for Viruses of Respiratory Infections, Institut Pasteur, Paris | Marion Barbet et al |
| EPI_ISL_1416971 | Outre mer | National Reference Center for Viruses of Respiratory Infections, Institut Pasteur, Paris | Marion Barbet et al |
| EPI_ISL_1443894 | Outre mer | National Reference Center for Viruses of Respiratory Infections, Institut Pasteur, Paris | Marion Barbet et al |
| EPI_ISL_1416978 | Outre mer | National Reference Center for Viruses of Respiratory Infections, Institut Pasteur, Paris | Marion Barbet et al |
| EPI_ISL_1416979 | Outre mer | National Reference Center for Viruses of Respiratory Infections, Institut Pasteur, Paris | Marion Barbet et al |
| EPI_ISL_1416983 | Outre mer | National Reference Center for Viruses of Respiratory Infections, Institut Pasteur, Paris | Marion Barbet et al |

| | | | |
|---|---|---|---|
| EPI_ISL_1416985 | Outre mer | National Reference Center for Viruses of Respiratory Infections, Institut Pasteur, Paris | Marion Barbet et al |
| EPI_ISL_1416988 | Outre mer | National Reference Center for Viruses of Respiratory Infections, Institut Pasteur, Paris | Marion Barbet et al |
| EPI_ISL_1416990 | Outre mer | National Reference Center for Viruses of Respiratory Infections, Institut Pasteur, Paris | Marion Barbet et al |
| EPI_ISL_1660296 | Virology Unit, Institut Pasteur de Madagascar | Virology Unit, Institut Pasteur de Madagascar | Marion Barbet et al |
| EPI_ISL_1660305 | Virology Unit, Institut Pasteur de Madagascar | Virology Unit, Institut Pasteur de Madagascar | Marion Barbet et al |
| EPI_ISL_1660310 | Virology Unit, Institut Pasteur de Madagascar | Virology Unit, Institut Pasteur de Madagascar | Marion Barbet et al |
| EPI_ISL_1660312 | Virology Unit, Institut Pasteur de Madagascar | Virology Unit, Institut Pasteur de Madagascar | Marion Barbet et al |
| EPI_ISL_1660313 | Virology Unit, Institut Pasteur de Madagascar | Virology Unit, Institut Pasteur de Madagascar | Marion Barbet et al |
| EPI_ISL_1660315 | Virology Unit, Institut Pasteur de Madagascar | Virology Unit, Institut Pasteur de Madagascar | Marion Barbet et al |
| EPI_ISL_1660317 | Virology Unit, Institut Pasteur de Madagascar | Virology Unit, Institut Pasteur de Madagascar | Marion Barbet et al |
| EPI_ISL_1660318 | Virology Unit, Institut Pasteur de Madagascar | Virology Unit, Institut Pasteur de Madagascar | Marion Barbet et al |
| EPI_ISL_1660322 | Virology Unit, Institut Pasteur de Madagascar | Virology Unit, Institut Pasteur de Madagascar | Marion Barbet et al |
| EPI_ISL_1660323 | Virology Unit, Institut Pasteur de Madagascar | Virology Unit, Institut Pasteur de Madagascar | Marion Barbet et al |
| EPI_ISL_1660329 | Virology Unit, Institut Pasteur de Madagascar | Virology Unit, Institut Pasteur de Madagascar | Marion Barbet et al |
| EPI_ISL_1660331 | Virology Unit, Institut Pasteur de Madagascar | Virology Unit, Institut Pasteur de Madagascar | Marion Barbet et al |
| EPI_ISL_1201507 | Hopital | National Reference Center for Viruses of Respiratory Infections, Institut Pasteur, Paris | Marion Barbet et al |
| EPI_ISL_1219962 | Labo Analyses Med | National Reference Center for Viruses of Respiratory Infections, Institut Pasteur, Paris | Marion Barbet et al |
| EPI_ISL_1219949 | Labo Analyses Med | National Reference Center for Viruses of Respiratory Infections, Institut Pasteur, Paris | Marion Barbet et al |
| EPI_ISL_1219963 | Labo Analyses Med | National Reference Center for Viruses of Respiratory Infections, Institut Pasteur, Paris | Marion Barbet et al |
| EPI_ISL_459970 | Institut Pasteur du Maroc | Institut Pasteur du Maroc | Marion Barbet et al |
| EPI_ISL_459971 | Institut Pasteur du Maroc | Institut Pasteur du Maroc | Marion Barbet et al |
| EPI_ISL_459972 | Institut Pasteur du Maroc | Institut Pasteur du Maroc | Marion Barbet et al |
| EPI_ISL_459974 | Institut Pasteur du Maroc | Institut Pasteur du Maroc | Marion Barbet et al |
| EPI_ISL_459976 | Institut Pasteur du Maroc | Institut Pasteur du Maroc | Marion Barbet et al |
| EPI_ISL_459981 | Institut Pasteur du Maroc | Institut Pasteur du Maroc | Marion Barbet et al |
| EPI_ISL_459982 | Institut Pasteur du Maroc | Institut Pasteur du Maroc | Marion Barbet et al |
| EPI_ISL_613429 | Institut Pasteur de la Guadeloupe | Institut Pasteur de la Guadeloupe | Marion Barbet et al |
| EPI_ISL_613446 | Institut Pasteur de la Guadeloupe | Institut Pasteur de la Guadeloupe | Marion Barbet et al |

| | | | |
|---|---|---|---|
| EPI_ISL_613447 | Institut Pasteur de la Guadeloupe | Institut Pasteur de la Guadeloupe | Marion Barbet et al |
| EPI_ISL_613448 | Institut Pasteur de la Guadeloupe | Institut Pasteur de la Guadeloupe | Marion Barbet et al |
| EPI_ISL_613450 | Institut Pasteur de la Guadeloupe | Institut Pasteur de la Guadeloupe | Marion Barbet et al |
| EPI_ISL_613451 | Institut Pasteur de la Guadeloupe | Institut Pasteur de la Guadeloupe | Marion Barbet et al |
| EPI_ISL_613452 | Institut Pasteur de la Guadeloupe | Institut Pasteur de la Guadeloupe | Marion Barbet et al |
| EPI_ISL_613453 | Institut Pasteur de la Guadeloupe | Institut Pasteur de la Guadeloupe | Marion Barbet et al |
| EPI_ISL_613454 | Institut Pasteur de la Guadeloupe | Institut Pasteur de la Guadeloupe | Marion Barbet et al |
| EPI_ISL_613455 | Institut Pasteur de la Guadeloupe | Institut Pasteur de la Guadeloupe | Marion Barbet et al |
| EPI_ISL_613456 | Institut Pasteur de la Guadeloupe | Institut Pasteur de la Guadeloupe | Marion Barbet et al |
| EPI_ISL_1034760 | Bundeswehr Institute of Microbiology | Bundeswehr Institute of Microbiology | Markus Antwerpen et al |
| EPI_ISL_1000998 | Bundeswehr Institute of Microbiology | Bundeswehr Institute of Microbiology | Markus Antwerpen et al |
| EPI_ISL_1000999 | Bundeswehr Institute of Microbiology | Bundeswehr Institute of Microbiology | Markus Antwerpen et al |
| EPI_ISL_1001000 | Bundeswehr Institute of Microbiology | Bundeswehr Institute of Microbiology | Markus Antwerpen et al |
| EPI_ISL_1001001 | Bundeswehr Institute of Microbiology | Bundeswehr Institute of Microbiology | Markus Antwerpen et al |
| EPI_ISL_1001002 | Bundeswehr Institute of Microbiology | Bundeswehr Institute of Microbiology | Markus Antwerpen et al |
| EPI_ISL_1001003 | Bundeswehr Institute of Microbiology | Bundeswehr Institute of Microbiology | Markus Antwerpen et al |
| EPI_ISL_1001004 | Bundeswehr Institute of Microbiology | Bundeswehr Institute of Microbiology | Markus Antwerpen et al |
| EPI_ISL_1034755 | Bundeswehr Institute of Microbiology | Bundeswehr Institute of Microbiology | Markus Antwerpen et al |
| EPI_ISL_1034757 | Bundeswehr Institute of Microbiology | Bundeswehr Institute of Microbiology | Markus Antwerpen et al |
| EPI_ISL_732530 | Bundeswehr Institute of Microbiology | Bundeswehr Institute of Microbiology | Markus Antwerpen et al |
| EPI_ISL_732526 | Bundeswehr Institute of Microbiology | Bundeswehr Institute of Microbiology | Markus Antwerpen et al |
| EPI_ISL_754239 | Santosa Hospital Bandung Central | School of Life Sciences and Technology & School of Pharmacy-Institut Teknologi Bandung; Molecular Genetics Laboratory-Faculty of Medicine-Universitas Padjadjaran; Laboratorium Kesehatan Provinsi Jawa Barat | Marselina Irasonia Tan et al |
| EPI_ISL_1492637 | Laboratorio Central de Salud Publica de Paraguay | Laboratorio Central de Salud Publica de Paraguay | Marta Giovanetti et al |
| EPI_ISL_1492646 | Laboratorio Central de Salud Publica de Paraguay | Laboratorio Central de Salud Publica de Paraguay | Marta Giovanetti et al |
| EPI_ISL_1492647 | Laboratorio Central de Salud Publica de Paraguay | Laboratorio Central de Salud Publica de Paraguay | Marta Giovanetti et al |
| EPI_ISL_1492648 | Laboratorio Central de Salud Publica de Paraguay | Laboratorio Central de Salud Publica de Paraguay | Marta Giovanetti et al |
| EPI_ISL_1492649 | Laboratorio Central de Salud Publica de Paraguay | Laboratorio Central de Salud Publica de Paraguay | Marta Giovanetti et al |
| EPI_ISL_1492650 | Laboratorio Central de Salud Publica de Paraguay | Laboratorio Central de Salud Publica de Paraguay | Marta Giovanetti et al |
| EPI_ISL_1492651 | Laboratorio Central de Salud Publica de Paraguay | Laboratorio Central de Salud Publica de Paraguay | Marta Giovanetti et al |

| | | | |
|---|---|---|---|
| EPI_ISL_1492652 | Laboratorio Central de Salud Publica de Paraguay | Laboratorio Central de Salud Publica de Paraguay | Marta Giovanetti et al |
| EPI_ISL_1492654 | Laboratorio Central de Salud Publica de Paraguay | Laboratorio Central de Salud Publica de Paraguay | Marta Giovanetti et al |
| EPI_ISL_1492657 | Laboratorio Central de Salud Publica de Paraguay | Laboratorio Central de Salud Publica de Paraguay | Marta Giovanetti et al |
| EPI_ISL_1492658 | Laboratorio Central de Salud Publica de Paraguay | Laboratorio Central de Salud Publica de Paraguay | Marta Giovanetti et al |
| EPI_ISL_1492659 | Laboratorio Central de Salud Publica de Paraguay | Laboratorio Central de Salud Publica de Paraguay | Marta Giovanetti et al |
| EPI_ISL_1492660 | Laboratorio Central de Salud Publica de Paraguay | Laboratorio Central de Salud Publica de Paraguay | Marta Giovanetti et al |
| EPI_ISL_1492661 | Laboratorio Central de Salud Publica de Paraguay | Laboratorio Central de Salud Publica de Paraguay | Marta Giovanetti et al |
| EPI_ISL_1492662 | Laboratorio Central de Salud Publica de Paraguay | Laboratorio Central de Salud Publica de Paraguay | Marta Giovanetti et al |
| EPI_ISL_1492639 | Laboratorio Central de Salud Publica de Paraguay | Laboratorio Central de Salud Publica de Paraguay | Marta Giovanetti et al |
| EPI_ISL_1492670 | Laboratorio Central de Salud Publica de Paraguay | Laboratorio Central de Salud Publica de Paraguay | Marta Giovanetti et al |
| EPI_ISL_1492640 | Laboratorio Central de Salud Publica de Paraguay | Laboratorio Central de Salud Publica de Paraguay | Marta Giovanetti et al |
| EPI_ISL_1492676 | Laboratorio Central de Salud Publica de Paraguay | Laboratorio Central de Salud Publica de Paraguay | Marta Giovanetti et al |
| EPI_ISL_1492677 | Laboratorio Central de Salud Publica de Paraguay | Laboratorio Central de Salud Publica de Paraguay | Marta Giovanetti et al |
| EPI_ISL_1492641 | Laboratorio Central de Salud Publica de Paraguay | Laboratorio Central de Salud Publica de Paraguay | Marta Giovanetti et al |
| EPI_ISL_1492642 | Laboratorio Central de Salud Publica de Paraguay | Laboratorio Central de Salud Publica de Paraguay | Marta Giovanetti et al |
| EPI_ISL_1492643 | Laboratorio Central de Salud Publica de Paraguay | Laboratorio Central de Salud Publica de Paraguay | Marta Giovanetti et al |
| EPI_ISL_1492644 | Laboratorio Central de Salud Publica de Paraguay | Laboratorio Central de Salud Publica de Paraguay | Marta Giovanetti et al |
| EPI_ISL_1492645 | Laboratorio Central de Salud Publica de Paraguay | Laboratorio Central de Salud Publica de Paraguay | Marta Giovanetti et al |
| EPI_ISL_436322 | Servicio de Microbiologia. Hospital Clinico Universitario de Valencia | Sequencing and Bioinformatics Service and Molecular Epidemiology Research Group. FISABIO-Public Health | Marta Pla Diaz et al |
| EPI_ISL_1585338 | Laboratorio di Microbiologia | Laboratorio di Microbiologia | Martinetti Lucchini Gladys et al |
| EPI_ISL_1163532 | LABCOVID_HCPA | LABRESIS_HCPA | Martins AF et al |
| EPI_ISL_1163530 | LABCOVID_HCPA | LABRESIS_HCPA | Martins AF et al |
| EPI_ISL_1163736 | LABCOVID_HCPA | LABCOVID_HCPA | Martins AF et al |
| EPI_ISL_1163737 | LABCOVID_HCPA | LABRESIS_HCPA | Martins AF et al |
| EPI_ISL_1133128 | LABCOVID_HCPA | LABRESIS_HCPA | Martins AF et al |
| EPI_ISL_1163694 | LABCOVID_HCPA | LABRESIS_HCPA | Martins AF et al |
| EPI_ISL_1133129 | LABCOVID_HCPA | LABRESIS_HCPA | Martins AF et al |
| EPI_ISL_1163695 | LABCOVID_HCPA | LABRESIS_HCPA | Martins AF et al |
| EPI_ISL_1133130 | LABCOVID_HCPA | LABRESIS_HCPA | Martins AF et al |
| EPI_ISL_1163696 | LABCOVID_HCPA | LABRESIS_HCPA | Martins AF et al |

| | | | |
|---|---|---|---|
| EPI_ISL_1133131 | LABCOVID_HCPA | LABRESIS_HCPA | Martins AF et al |
| EPI_ISL_1133132 | LABCOVID_HCPA | LABRESIS_HCPA | Martins AF et al |
| EPI_ISL_1163697 | LABCOVID_HCPA | LABRESIS_HCPA | Martins AF et al |
| EPI_ISL_1161401 | LABCOVID_HCPA | LABRESIS_HCPA | Martins AF et al |
| EPI_ISL_1163698 | LABCOVID_HCPA | LABRESIS_HCPA | Martins AF et al |
| EPI_ISL_1163699 | LABCOVID_HCPA | LABRESIS_HCPA | Martins AF et al |
| EPI_ISL_1133133 | LABCOVID_HCPA | LABRESIS_HCPA | Martins AF et al |
| EPI_ISL_1161402 | LABCOVID_HCPA | LABRESIS_HCPA | Martins AF et al |
| EPI_ISL_1163700 | LABCOVID_HCPA | LABRESIS_HCPA | Martins AF et al |
| EPI_ISL_1161403 | LABCOVID_HCPA | LABRESIS_HCPA | Martins AF et al |
| EPI_ISL_1163701 | LABCOVID_HCPA | LABRESIS_HCPA | Martins AF et al |
| EPI_ISL_1133120 | LABCOVID_HCPA | LABRESIS_HCPA | Martins AF et al |
| EPI_ISL_1163702 | LABCOVID_HCPA | LABRESIS_HCPA | Martins AF et al |
| EPI_ISL_1163703 | LABCOVID_HCPA | LABRESIS_HCPA | Martins AF et al |
| EPI_ISL_1163704 | LABCOVID_HCPA | LABRESIS_HCPA | Martins AF et al |
| EPI_ISL_1163705 | LABCOVID_HCPA | LABRESIS_HCPA | Martins AF et al |
| EPI_ISL_1163706 | LABCOVID_HCPA | LABRESIS_HCPA | Martins AF et al |
| EPI_ISL_1163707 | LABCOVID_HCPA | LABRESIS_HCPA | Martins AF et al |
| EPI_ISL_1133134 | LABCOVID_HCPA | LABRESIS_HCPA | Martins AF et al |
| EPI_ISL_1163708 | LABCOVID_HCPA | LABRESIS_HCPA | Martins AF et al |
| EPI_ISL_1163709 | LABCOVID_HCPA | LABRESIS_HCPA | Martins AF et al |
| EPI_ISL_1163710 | LABCOVID_HCPA | LABRESIS_HCPA | Martins AF et al |
| EPI_ISL_1163711 | LABCOVID_HCPA | LABRESIS_HCPA | Martins AF et al |
| EPI_ISL_1133135 | LABCOVID_HCPA | LABRESIS_HCPA | Martins AF et al |
| EPI_ISL_1161413 | LABCOVID_HCPA | LABRESIS_HCPA | Martins AF et al |
| EPI_ISL_1163712 | LABCOVID_HCPA | LABRESIS_HCPA | Martins AF et al |
| EPI_ISL_1161414 | LABCOVID_HCPA | LABRESIS_HCPA | Martins AF et al |
| EPI_ISL_1163713 | LABCOVID_HCPA | LABRESIS_HCPA | Martins AF et al |
| EPI_ISL_1133136 | LABCOVID_HCPA | LABRESIS_HCPA | Martins AF et al |
| EPI_ISL_1133137 | LABCOVID_HCPA | LABRESIS_HCPA | Martins AF et al |

| | | | |
|---|---|---|---|
| EPI_ISL_1133138 | LABCOVID_HCPA | LABRESIS_HCPA | Martins AF et al |
| EPI_ISL_1163714 | LABCOVID_HCPA | LABRESIS_HCPA | Martins AF et al |
| EPI_ISL_1133139 | LABCOVID_HCPA | LABRESIS_HCPA | Martins AF et al |
| EPI_ISL_1163715 | LABCOVID_HCPA | LABRESIS_HCPA | Martins AF et al |
| EPI_ISL_1163716 | LABCOVID_HCPA | LABRESIS_HCPA | Martins AF et al |
| EPI_ISL_1133140 | LABCOVID_HCPA | LABRESIS_HCPA | Martins AF et al |
| EPI_ISL_1133121 | LABCOVID_HCPA | LABRESIS_HCPA | Martins AF et al |
| EPI_ISL_1161415 | LABCOVID_HCPA | LABRESIS_HCPA | Martins AF et al |
| EPI_ISL_1133141 | LABCOVID_HCPA | LABRESIS_HCPA | Martins AF et al |
| EPI_ISL_1133142 | LABCOVID_HCPA | LABRESIS_HCPA | Martins AF et al |
| EPI_ISL_1133143 | LABCOVID_HCPA | LABRESIS_HCPA | Martins AF et al |
| EPI_ISL_1133144 | LABCOVID_HCPA | LABRESIS_HCPA | Martins AF et al |
| EPI_ISL_1133122 | LABCOVID_HCPA | LABRESIS_HCPA | Martins AF et al |
| EPI_ISL_1133123 | LABCOVID_HCPA | LABRESIS_HCPA | Martins AF et al |
| EPI_ISL_1133124 | LABCOVID_HCPA | LABRESIS_HCPA | Martins AF et al |
| EPI_ISL_1133125 | LABCOVID_HCPA | LABRESIS_HCPA | Martins AF et al |
| EPI_ISL_1133126 | LABCOVID_HCPA | LABRESIS_HCPA | Martins AF et al |
| EPI_ISL_1163693 | LABCOVID_HCPA | LABRESIS_HCPA | Martins AF et al |
| EPI_ISL_1133127 | LABCOVID_HCPA | LABRESIS_HCPA | Martins AF et al |
| EPI_ISL_596672 | St.Vincent's University Hospital | St.Vincent's University Hospital | Mary Lucey et al |
| EPI_ISL_757285 | Department of Virology, Public Health Laboratories Division | Department of Virology, Public Health Laboratories Division | Massab Umair et al |
| EPI_ISL_757286 | Department of Virology, Public Health Laboratories Division | Department of Virology, Public Health Laboratories Division | Massab Umair et al |
| EPI_ISL_468159 | unknown | Department of Virology, Public Health Laboratories Division, National Institute of Health | Massab Umair et al |
| EPI_ISL_1385784 | Department of Virology | Department of Virology | Massab Umair et al |
| EPI_ISL_1385785 | Department of Virology | Department of Virology | Massab Umair et al |
| EPI_ISL_1385786 | Department of Virology | Department of Virology | Massab Umair et al |
| EPI_ISL_1385789 | Department of Virology | Department of Virology | Massab Umair et al |
| EPI_ISL_1397958 | Department of Virology | Department of Virology | Massab Umair et al |
| EPI_ISL_1398039 | Department of Virology | Department of Virology | Massab Umair et al |
| EPI_ISL_1406391 | Department of Virology | Department of Virology | Massab Umair et al |

| | | | |
|---|---|---|---|
| EPI_ISL_1406393 | Department of Virology | Department of Virology | Massab Umair et al |
| EPI_ISL_1406394 | Department of Virology | Department of Virology | Massab Umair et al |
| EPI_ISL_1385791 | Department of Virology | Department of Virology | Massab Umair et al |
| EPI_ISL_1385794 | Department of Virology | Department of Virology | Massab Umair et al |
| EPI_ISL_1406399 | Department of Virology | Department of Virology | Massab Umair et al |
| EPI_ISL_1385800 | Department of Virology | Department of Virology | Massab Umair et al |
| EPI_ISL_1385810 | Department of Virology | Department of Virology | Massab Umair et al |
| EPI_ISL_1385812 | Department of Virology | Department of Virology | Massab Umair et al |
| EPI_ISL_468163 | Department of Virology, Public Health Laboratories Division, National Institute of Health | Department of Virology, Public Health Laboratories Division, National Institute of Health | Massab Umair et al |
| EPI_ISL_1372711 | Maine Health and Environmental Testing Laboratory | Tewhey Lab, The Jackson Laboratory | Matluk et al |
| EPI_ISL_1372713 | Maine Health and Environmental Testing Laboratory | Tewhey Lab, The Jackson Laboratory | Matluk et al |
| EPI_ISL_1372731 | Maine Health and Environmental Testing Laboratory | Tewhey Lab, The Jackson Laboratory | Matluk et al |
| EPI_ISL_1372863 | Maine Health and Environmental Testing Laboratory | Tewhey Lab, The Jackson Laboratory | Matluk et al |
| EPI_ISL_417479 | Minnesota Department of Health, Public Health Laboratory | Minnesota Department of Health, Public Health Laboratory | Matt Plumb et al |
| EPI_ISL_417502 | Minnesota Department of Health, Public Health Laboratory | Minnesota Department of Health, Public Health Laboratory | Matt Plumb et al |
| EPI_ISL_413490 | Auckland Hospital | Institute of Environmental Science and Research (ESR) | Matt Storey et al |
| EPI_ISL_456288 | Southern Community Labs Dunedin | Institute of Environmental Science and Research (ESR) | Matt Storey et al |
| EPI_ISL_456291 | Southern Community Labs Dunedin | Institute of Environmental Science and Research (ESR) | Matt Storey et al |
| EPI_ISL_456293 | Southern Community Labs Dunedin | Institute of Environmental Science and Research (ESR) | Matt Storey et al |
| EPI_ISL_456303 | Southern Community Labs Dunedin | Institute of Environmental Science and Research (ESR) | Matt Storey et al |
| EPI_ISL_456313 | Southern Community Labs Dunedin | Institute of Environmental Science and Research (ESR) | Matt Storey et al |
| EPI_ISL_456315 | Southern Community Labs Dunedin | Institute of Environmental Science and Research (ESR) | Matt Storey et al |
| EPI_ISL_456341 | Wellington SCL | Institute of Environmental Science and Research (ESR) | Matt Storey et al |
| EPI_ISL_456354 | Canterbury Health Laboratories | Institute of Environmental Science and Research (ESR) | Matt Storey et al |
| EPI_ISL_871801 | AIID | Irish Coronavirus Sequencing Consortium-Teagasc Grange | Matthew McCabe et al |
| EPI_ISL_871802 | AIID | Irish Coronavirus Sequencing Consortium-Teagasc Grange | Matthew McCabe et al |
| EPI_ISL_602539 | Center of Medical Microbiology, Virology, and Hospital Hygiene, University of Duesseldorf | Center of Medical Microbiology, Virology, and Hospital Hygiene, University of Duesseldorf | Maximilian Damagnez et al |
| EPI_ISL_602515 | Center of Medical Microbiology, Virology, and Hospital Hygiene, University of Duesseldorf | Center of Medical Microbiology, Virology, and Hospital Hygiene, University of Duesseldorf | Maximilian Damagnez et al |
| EPI_ISL_1346939 | Center of Medical Microbiology, Virology, and Hospital Hygiene, University of Duesseldorf | Center of Medical Microbiology, Virology, and Hospital Hygiene, University of Duesseldorf | Maximilian Damagnez et al |

| | | | |
|---|---|---|---|
| EPI_ISL_1659701 | Swedish national genomic surveillance program of SARS-CoV-2 | The Public Health Agency of Sweden | Maximilian Riess et al |
| EPI_ISL_1659714 | Swedish national genomic surveillance program of SARS-CoV-2 | The Public Health Agency of Sweden | Maximilian Riess et al |
| EPI_ISL_1659866 | Swedish national genomic surveillance program of SARS-CoV-2 | The Public Health Agency of Sweden | Maximilian Riess et al |
| EPI_ISL_1660117 | Swedish national genomic surveillance program of SARS-CoV-2 | The Public Health Agency of Sweden | Maximilian Riess et al |
| EPI_ISL_1499123 | 1. Glowny Inspektorat Sanitarny. 2. Diagnostyka. Laboratoria Medyczne. | 1. ViroGenetics - BSL3 Laboratory of Virology, Małopolska Centre of Biotechnology, Jagiellonian University; 2. genXone SA, Research & Development Laboratory | Mazur-Panasiuk et al |
| EPI_ISL_1402488 | 1. Glowny Inspektorat Sanitarny. 2. Diagnostyka. Laboratoria Medyczne. | 1. ViroGenetics - BSL3 Laboratory of Virology, Małopolska Centre of Biotechnology, Jagiellonian University; 2. genXone SA, Research & Development Laboratory | Mazur-Panasiuk et al |
| EPI_ISL_1402519 | 1. Glowny Inspektorat Sanitarny. 2. Diagnostyka. Laboratoria Medyczne. | 1. ViroGenetics - BSL3 Laboratory of Virology, Małopolska Centre of Biotechnology, Jagiellonian University; 2. genXone SA, Research & Development Laboratory | Mazur-Panasiuk et al |
| EPI_ISL_1402524 | 1. Glowny Inspektorat Sanitarny. 2. Diagnostyka. Laboratoria Medyczne. | 1. ViroGenetics - BSL3 Laboratory of Virology, Małopolska Centre of Biotechnology, Jagiellonian University; 2. genXone SA, Research & Development Laboratory | Mazur-Panasiuk et al |
| EPI_ISL_1402537 | 1. Glowny Inspektorat Sanitarny. 2. Diagnostyka. Laboratoria Medyczne. | 1. ViroGenetics - BSL3 Laboratory of Virology, Małopolska Centre of Biotechnology, Jagiellonian University; 2. genXone SA, Research & Development Laboratory | Mazur-Panasiuk et al |
| EPI_ISL_1406337 | 1. Glowny Inspektorat Sanitarny. 2. Diagnostyka. Laboratoria Medyczne. | 1. ViroGenetics - BSL3 Laboratory of Virology, Małopolska Centre of Biotechnology, Jagiellonian University; 2. genXone SA, Research & Development Laboratory | Mazur-Panasiuk et al |
| EPI_ISL_573249 | Virology Department, Royal Infirmary of Edinburgh, NHS Lothian / School of Biological Sciences, University of Edinburgh / Institute of Genetics and Molecular Medicine, University of Edinburgh | COVID-19 Genomics UK (COG-UK) Consortium | McHugh M et al |
| EPI_ISL_1104214 | Virology Department, Royal Infirmary of Edinburgh, NHS Lothian / School of Biological Sciences, University of Edinburgh | COVID-19 Genomics UK (COG-UK) Consortium | McHugh M et al |
| EPI_ISL_1104216 | Virology Department, Royal Infirmary of Edinburgh, NHS Lothian / School of Biological Sciences, University of Edinburgh | COVID-19 Genomics UK (COG-UK) Consortium | McHugh M et al |
| EPI_ISL_1636703 | Virology Department, Royal Infirmary of Edinburgh, NHS Lothian / School of Biological Sciences, University of Edinburgh | COVID-19 Genomics UK (COG-UK) Consortium | McHugh M et al |
| EPI_ISL_478249 | Virology Department, Royal Infirmary of Edinburgh, NHS Lothian / School of Biological Sciences, University of Edinburgh / Institute of Genetics and Molecular Medicine, University of Edinburgh | COVID-19 Genomics UK (COG-UK) Consortium | McHugh M et al |
| EPI_ISL_1494726 | Centro de Investigación en Ciencias de la Salud y Biomedicina, U.A.S.LP. | Centro de Investigación en Ciencias de la Salud y Biomedicina, U.A.S.LP. | MD PhD Sofía Bernal Silva et al |
| EPI_ISL_943566 | National Institute of Laboratory Medicine and Referral Center | Genomic Research Lab, BCSIR | Md. Ahashan Habib et al |
| EPI_ISL_747283 | DNA SOLUTION LTD. | DNA SOLUTION LTD. | Md. Imran Khan et al |
| EPI_ISL_774897 | Designated Reference Institute for Chemical Measurements (DRICM) | DNA SOLUTION LTD. | Md. Imran Khan et al |
| EPI_ISL_774894 | Designated Reference Institute for Chemical Measurements (DRICM) | DNA SOLUTION LTD. | Md. Imran Khan et al |
| EPI_ISL_774898 | Designated Reference Institute for Chemical Measurements (DRICM) | DNA SOLUTION LTD. | Md. Imran Khan et al |
| EPI_ISL_775000 | Designated Reference Institute for Chemical Measurements (DRICM) | DNA SOLUTION LTD. | Md. Imran Khan et al |

| | | | |
|---|---|---|---|
| EPI_ISL_774979 | Designated Reference Institute for Chemical Measurements (DRICM) | DNA SOLUTION LTD. | Md. Imran Khan et al |
| EPI_ISL_735496 | Chattogram Veterinary and Animal Sciences University | Central Biological Research Laboratory and Department of Biochemistry and Molecular Biology | Md. Imranul Hoq et al |
| EPI_ISL_959309 | National Institute of Laboratory Medicine and Referral Center | Genomic Research Lab, BCSIR | Md. Maruf Ahmed Molla et al |
| EPI_ISL_447904 | National Institute of Biotechnology | National Institute of Biotechnology | Md. Moniruzzaman et al |
| EPI_ISL_1585269 | Armed Forces Institute of Pathology (AFIP), Dhaka Cantonment | Genomic Research Lab, BCSIR | Md. Murshed Hasan Sarkar et al |
| EPI_ISL_1636521 | DNA Solution Ltd. | Genomic Research Lab, BCSIR | Md. Murshed Hasan Sarkar et al |
| EPI_ISL_1636522 | DNA Solution Ltd. | Genomic Research Lab, BCSIR | Md. Murshed Hasan Sarkar et al |
| EPI_ISL_1493059 | National Institute of Laboratory Medicine and Referral Center | Bangladesh Council of Scientific and Industrial Research | Md. Murshed Hasan Sarkar et al |
| EPI_ISL_1033158 | National Institute of Laboratory Medicine and Referral Center | Genomic Research Lab, BCSIR | Md. Murshed Hasan Sarkar et al |
| EPI_ISL_1508950 | National Institute of Laboratory Medicine and Referral Center | Genomic Research Lab, BCSIR | Md. Murshed Hasan Sarkar et al |
| EPI_ISL_1542073 | Department of Genetic Engineering and Biotechnology, Shahjalal University of Science and Technology | Genomic Research Lab, BCSIR | Md. Murshed Hasan Sarkar et al |
| EPI_ISL_1563631 | Department of Genetic Engineering and Biotechnology, Shahjalal University of Science and Technology | Genomic Research Lab, BCSIR | Md. Shamsul Haque Prodhan et al |
| EPI_ISL_430456 | Rizal Medical Center | Research Institute for Tropical Medicine | Medado et al |
| EPI_ISL_430845 | Pasig City General Hospital | Research Institute for Tropical Medicine | Medado et al |
| EPI_ISL_610245 | Virology, National Institute for Biological Standards and Control | Virology, National Institute for Biological Standards and Control | Mee et al |
| EPI_ISL_455719 | T.C. Sağlık Bakanlığı Adıyaman İl Sağlık Müdürlüğü Adıyaman Eğitim Ve Araştırma Hastanesi | VETAL Animal Health Products Company, BSL3+ Production Laboratuary /Turkey | Mehmet Turgut et al |
| EPI_ISL_418241 | NIC Viral Respiratory Unit - Institut Pasteur of Algeria | National Reference Center for Viruses of Respiratory Infections, Institut Pasteur, Paris | Mélanie Albert et al |
| EPI_ISL_418242 | NIC Viral Respiratory Unit - Institut Pasteur of Algeria | National Reference Center for Viruses of Respiratory Infections, Institut Pasteur, Paris | Mélanie Albert et al |
| EPI_ISL_420037 | NIC Viral Respiratory Unit - Institut Pasteur of Algeria | National Reference Center for Viruses of Respiratory Infections, Institut Pasteur, Paris | Mélanie Albert et al |
| EPI_ISL_766864 | NIC Viral Respiratory Unit - Institut Pasteur of Algeria | National Reference Center for Viruses of Respiratory Infections, Institut Pasteur, Paris | Mélanie Albert et al |
| EPI_ISL_766867 | NIC Viral Respiratory Unit - Institut Pasteur of Algeria | National Reference Center for Viruses of Respiratory Infections, Institut Pasteur, Paris | Mélanie Albert et al |
| EPI_ISL_766874 | NIC Viral Respiratory Unit - Institut Pasteur of Algeria | National Reference Center for Viruses of Respiratory Infections, Institut Pasteur, Paris | Mélanie Albert et al |
| EPI_ISL_406597 | Department of Infectious and Tropical Diseases, Bichat Claude Bernard Hospital, Paris | National Reference Center for Viruses of Respiratory Infections, Institut Pasteur, Paris | Mélanie Albert et al |
| EPI_ISL_410984 | Department of Infectious and Tropical Diseases, Bichat Claude Bernard Hospital, Paris | National Reference Center for Viruses of Respiratory Infections, Institut Pasteur, Paris | Mélanie Albert et al |
| EPI_ISL_408430 | Department of Infectious and Tropical Diseases, Bichat Claude Bernard Hospital, Paris | National Reference Center for Viruses of Respiratory Infections, Institut Pasteur, Paris | Mélanie Albert et al |

| | | | |
|---|---|---|---|
| EPI_ISL_414623 | Laboratoire de Virologie Institut de Virologie - INSERM U 1109 Hôpitaux Universitaires de Strasbourg | National Reference Center for Viruses of Respiratory Infections, Institut Pasteur, Paris | Mélnie Albert et al |
| EPI_ISL_414600 | Laboratoire de Virologie Institut de Virologie - INSERM U 1109 Hôpitaux Universitaires de Strasbourg | National Reference Center for Viruses of Respiratory Infections, Institut Pasteur, Paris | Mélnie Albert et al |
| EPI_ISL_1296252 | Università degli Studi di Perugia | Istituto Zooprofilattico Sperimentale dell'Abruzzo e Molise "G. Caporale" | Mencacci A et al |
| EPI_ISL_1670647 | Università degli Studi di Perugia | Istituto Zooprofilattico Sperimentale dell'Abruzzo e Molise "G. Caporale" | Mencacci A et al |
| EPI_ISL_500168 | Hospital Universitario Virgen de las Nieves de Granada-SAS | SeqCOVID-SPAIN consortium/IBV(CSIC) | Mercedes Pérez Ruiz et al |
| EPI_ISL_471529 | Department for Virology, Molecular Biology and Genome Research, R. G. Lugar Center for Public Health Research, National Center for Disease Control and Public Health (NCDC) of Georgia. | Department for Virology, Molecular Biology and Genome Research, R. G. Lugar Center for Public Health Research, National Center for Disease Control and Public Health (NCDC) of Georgia. | Meri Pantsulaia et al |
| EPI_ISL_754181 | Department for Virology, Molecular Biology and Genome Research, R. G. Lugar Center for Public Health Research, National Center for Disease Control and Public Health (NCDC) of Georgia. | Department for Virology, Molecular Biology and Genome Research, R. G. Lugar Center for Public Health Research, National Center for Disease Control and Public Health (NCDC) of Georgia. | Meri Pantsulaia et al |
| EPI_ISL_469051 | LNR National Reference Laboratory, Mohammed VI University of Health Sciences | Medical Biotechnology Laboratory, Rabat Medical and Pharmacy School, Mohammed The Vth University in Rabat | Meriem LAAMARTI et al |
| EPI_ISL_482739 | LNR National Reference Laboratory, Mohammed VI University of Health Sciences | Medical Biotechnology Laboratory, Rabat Medical and Pharmacy School, Mohammed The Vth University in Rabat | Meriem LAAMARTI et al |
| EPI_ISL_482740 | LNR National Reference Laboratory, Mohammed VI University of Health Sciences | Medical Biotechnology Laboratory, Rabat Medical and Pharmacy School, Mohammed The Vth University in Rabat | Meriem LAAMARTI et al |
| EPI_ISL_1061413 | Istanbul University-Cerrahpasa, Cerrahpasa School of Medicine, COVID-19 Laboratory | Istanbul University-Cerrahpasa, Cerrahpasa School of Medicine, COVID-19 Laboratory | Mert Ahmet Kuskucu et al |
| EPI_ISL_1662234 | IU-Cerrahpasa, Cerrahpasa School of Medicine, COVID-19 Lab | IU-Cerrahpasa, Cerrahpasa School of Medicine, COVID-19 Lab | Mert Kuskucu et al |
| EPI_ISL_1662593 | IU-Cerrahpasa, Cerrahpasa School of Medicine, COVID-19 Lab | IU-Cerrahpasa, Cerrahpasa School of Medicine, COVID-19 Lab | Mert Kuskucu et al |
| EPI_ISL_1663062 | IU-Cerrahpasa, Cerrahpasa School of Medicine, COVID-19 Lab | Illumina Miseq | Mert Kuskucu et al |
| EPI_ISL_1663581 | IU-Cerrahpasa, Cerrahpasa School of Medicine, COVID-19 Lab | IU-Cerrahpasa, Cerrahpasa School of Medicine, COVID-19 Lab | Mert Kuskucu et al |
| EPI_ISL_959706 | National Virus Reference Laboratory | National Virus Reference Laboratory | Michael Carr et al |
| EPI_ISL_644338 | CEPHR / Vincent's Hospital | Irish Coronavirus Sequencing Consortium - National Virus Reference Laboratory | Michael Carr et al |
| EPI_ISL_528470 | National Virus Reference Laboratory | National Virus Reference Laboratory | Michael Carr et al |
| EPI_ISL_732464 | National Virus Reference Laboratory | National Virus Reference Laboratory | Michael Carr et al |
| EPI_ISL_501261 | National Virus Reference Laboratory | National Virus Reference Laboratory | Michael Carr et al |
| EPI_ISL_500592 | National Virus Reference Laboratory | National Virus Reference Laboratory | Michael Carr et al |
| EPI_ISL_528465 | National Virus Reference Laboratory | National Virus Reference Laboratory | Michael Carr et al |
| EPI_ISL_837362 | National Virus Reference Laboratory | National Virus Reference Laboratory | Michael Carr et al |
| EPI_ISL_875351 | National Virus Reference Laboratory | National Virus Reference Laboratory | Michael Carr et al |
| EPI_ISL_644214 | CEPHR / Vincent's Hospital | Irish Coronavirus Sequencing Consortium - National Virus Reference Laboratory | Michael Carr et al |

| | | | |
|---|---|---|---|
| EPI_ISL_671903 | National Virus Reference Laboratory | National Virus Reference Laboratory | Michael Carr et al |
| EPI_ISL_512106 | National Virus Reference Laboratory | National Virus Reference Laboratory | Michael Carr et al |
| EPI_ISL_525363 | National Virus Reference Laboratory | National Virus Reference Laboratory | Michael Carr et al |
| EPI_ISL_644213 | CEPHR / Vincent's Hospital | Irish Coronavirus Sequencing Consortium - National Virus Reference Laboratory | Michael Carr et al |
| EPI_ISL_501260 | National Virus Reference Laboratory | National Virus Reference Laboratory | Michael Carr et al |
| EPI_ISL_671921 | National Virus Reference Laboratory | National Virus Reference Laboratory | Michael Carr et al |
| EPI_ISL_639900 | National Virus Reference Laboratory | National Virus Reference Laboratory | Michael Carr et al |
| EPI_ISL_639899 | National Virus Reference Laboratory | National Virus Reference Laboratory | Michael Carr et al |
| EPI_ISL_639898 | National Virus Reference Laboratory | National Virus Reference Laboratory | Michael Carr et al |
| EPI_ISL_639906 | National Virus Reference Laboratory | National Virus Reference Laboratory | Michael Carr et al |
| EPI_ISL_639892 | National Virus Reference Laboratory | National Virus Reference Laboratory | Michael Carr et al |
| EPI_ISL_639903 | National Virus Reference Laboratory | National Virus Reference Laboratory | Michael Carr et al |
| EPI_ISL_639831 | National Virus Reference Laboratory | National Virus Reference Laboratory | Michael Carr et al |
| EPI_ISL_525354 | National Virus Reference Laboratory | National Virus Reference Laboratory | Michael Carr et al |
| EPI_ISL_528453 | National Virus Reference Laboratory | National Virus Reference Laboratory | Michael Carr et al |
| EPI_ISL_578235 | National Virus Reference Laboratory | National Virus Reference Laboratory | Michael Carr et al |
| EPI_ISL_752592 | National Virus Reference Laboratory | National Virus Reference Laboratory | Michael Carr et al |
| EPI_ISL_644224 | CEPHR / Vincent's Hospital | Irish Coronavirus Sequencing Consortium - National Virus Reference Laboratory | Michael Carr et al |
| EPI_ISL_1301441 | Department of Infectious and Tropical Diseases, Bichat Claude Bernard Hospital, Paris | NBACC | Michael Schuit et al |
| EPI_ISL_708512 | National Virus Reference Laboratory | Irish Coronavirus Sequencing Consortium - Teagasc Oakpark | Michele Della Bartola et al |
| EPI_ISL_708519 | National Virus Reference Laboratory | Irish Coronavirus Sequencing Consortium - Teagasc Oakpark | Michele Della Bartola et al |
| EPI_ISL_861712 | Laboratory for Respiratory Viruses, Cantacuzino National Military-Medical Institute for Research and Development | Cantacuzino Institute Virology | Mihaela Lazar et al |
| EPI_ISL_861713 | Laboratory for Respiratory Viruses, Cantacuzino National Military-Medical Institute for Research and Development | Cantacuzino Institute Virology | Mihaela Lazar et al |
| EPI_ISL_861711 | Laboratory for Respiratory Viruses, Cantacuzino National Military-Medical Institute for Research and Development | Cantacuzino Institute Virology | Mihaela Lazar et al |
| EPI_ISL_1663578 | Fimlab Laboratories | Fimlab Laboratories | Minna Paloniemi et al |
| EPI_ISL_1663568 | Fimlab Laboratories | Fimlab Laboratories | Minna Paloniemi et al |
| EPI_ISL_1663575 | Fimlab Laboratories | Fimlab Laboratories | Minna Paloniemi et al |
| EPI_ISL_1072985 | University of Balamand | Microbial Genomics Lab, Lebanese American University, Byblos | Mira El Chaar et al |

| | | | |
|---|---|---|---|
| EPI_ISL_978545 | Centre de Virologie des Maladies infectueuses Tropicales | Functional Genomic Platform UATRS-biology, CNRST | Mly Abdelaziz ELALAOUI et al |
| EPI_ISL_430819 | Center of Scientific Excellence for Influenza Viruses,National Research Centre (NRC), Egypt. | Center of Scientific Excellence for Influenza Viruses,National Research Centre (NRC), Egypt. | Mohamed Ahmed Ali et al |
| EPI_ISL_442044 | Kawsar Human Genetic Research Center | Kawsar Human Genetic Research Center | Mohammad Ali Khosravi et al |
| EPI_ISL_904015 | Molecular Biology and Virology lab, Faculty of Veterinary Medicine, Jordan University of Science and Technology | Molecular Biology and Virology lab, Faculty of Veterinary Medicine, Jordan University of Science and Technology | Mohammad Hussien Alboom et al |
| EPI_ISL_904016 | Molecular Biology and Virology lab, Faculty of Veterinary Medicine, Jordan University of Science and Technology | Molecular Biology and Virology lab, Faculty of Veterinary Medicine, Jordan University of Science and Technology | Mohammad Hussien Alboom et al |
| EPI_ISL_1593857 | Armed Forces Institute of Pathology (AFIP), Dhaka Cantonment | Genomic Research Lab, BCSIR | Mohammad Mizanur Rahman et al |
| EPI_ISL_1133253 | University of Chittagong | Central Biological Research Laboratory and Department of Biochemistry and Molecular Biology | Mohammad Omar Faruque et al |
| EPI_ISL_977585 | Department of Medical Microbiology, Hospital Pengajar Universiti Putra Malaysia | Malaysia Genome Institute | Mohd Noor Mat Isa et al |
| EPI_ISL_861725 | Department of Medical Microbiology, Hospital Pengajar Universiti Putra Malaysia | Malaysia Genome Institute | Mohd Noor Mat Isa et al |
| EPI_ISL_861726 | Department of Medical Microbiology, Hospital Pengajar Universiti Putra Malaysia | Malaysia Genome Institute | Mohd Noor Mat Isa et al |
| EPI_ISL_455793 | Institute for Medical Research, Infectious Disease Research Centre, National Institutes of Health, Ministry of Health Malaysia | Malaysia Genome Institute | Mohd Noor Mat Isa et al |
| EPI_ISL_455790 | Institute for Medical Research, Infectious Disease Research Centre, National Institutes of Health, Ministry of Health Malaysia | Malaysia Genome Institute | Mohd Noor Mat Isa et al |
| EPI_ISL_455791 | Institute for Medical Research, Infectious Disease Research Centre, National Institutes of Health, Ministry of Health Malaysia | Malaysia Genome Institute | Mohd Noor Mat Isa et al |
| EPI_ISL_455792 | Institute for Medical Research, Infectious Disease Research Centre, National Institutes of Health, Ministry of Health Malaysia | Malaysia Genome Institute | Mohd Noor Mat Isa et al |
| EPI_ISL_582124 | Malaysia Genome Institute | Malaysia Genome Institute | Mohd Noor Mat Isa et al |
| EPI_ISL_528743 | Malaysia Genome Institute | Malaysia Genome Institute | Mohd Noor Mat Isa et al |
| EPI_ISL_528744 | Malaysia Genome Institute | Malaysia Genome Institute | Mohd Noor Mat Isa et al |
| EPI_ISL_416829 | National Public Health Laboratory | Malaysia Genome Institute | Mohd Noor Mat Isa et al |
| EPI_ISL_416866 | National Public Health Laboratory | Malaysia Genome Institute | Mohd Noor Mat Isa et al |
| EPI_ISL_416885 | National Public Health Laboratory | Malaysia Genome Institute | Mohd Noor Mat Isa et al |
| EPI_ISL_416886 | National Public Health Laboratory | Malaysia Genome Institute | Mohd Noor Mat Isa et al |
| EPI_ISL_416907 | National Public Health Laboratory | Malaysia Genome Institute | Mohd Noor Mat Isa et al |
| EPI_ISL_807153 | Makmal Kesihatan Awam Kebangsaan, Kementerian Kesihatan Malaysia | Integrative Pharmacogenomics Institute (iPROMISE) | Mohd Zaki Salleh et al |
| EPI_ISL_807152 | Makmal Kesihatan Awam Kebangsaan, Kementerian Kesihatan Malaysia | Integrative Pharmacogenomics Institute (iPROMISE) | Mohd Zaki Salleh et al |
| EPI_ISL_407071 | Respiratory Virus Unit, Microbiology Services Colindale, Public Health England | Respiratory Virus Unit, Microbiology Services Colindale, Public Health England | Monica Galiano et al |

| | | | |
|---|---|---|---|
| EPI_ISL_407073 | Respiratory Virus Unit, Microbiology Services Colindale, Public Health England | Respiratory Virus Unit, Microbiology Services Colindale, Public Health England | Monica Galiano et al |
| EPI_ISL_414522 | Respiratory Virus Unit, Microbiology Services Colindale, Public Health England | Respiratory Virus Unit, Microbiology Services Colindale, Public Health England | Monica Galiano et al |
| EPI_ISL_424154 | Respiratory Virus Unit, Microbiology Services Colindale, Public Health England | Respiratory Virus Unit, Microbiology Services Colindale, Public Health England | Monica Galiano et al |
| EPI_ISL_1589576 | Hospital Universitario Marqués de Valdecilla - IDIVAL (Santander, Cantabria) | SeqCOVID-SPAIN consortium/IBV(CSIC) | Mónica Gozalo Margüello et al |
| EPI_ISL_1133186 | Molecular Virology Unit, Microbiology and Virology Depertment, Fondazione IRCCS Policlinico San Matteo, Pavia | Molecular Virology Unit, Microbiology and Virology Depertment, Fondazione IRCCS Policlinico San Matteo, Pavia | Monica Tallarita et al |
| EPI_ISL_1133197 | Molecular Virology Unit, Microbiology and Virology Depertment, Fondazione IRCCS Policlinico San Matteo, Pavia | Molecular Virology Unit, Microbiology and Virology Depertment, Fondazione IRCCS Policlinico San Matteo, Pavia | Monica Tallarita et al |
| EPI_ISL_1133201 | Molecular Virology Unit, Microbiology and Virology Depertment, Fondazione IRCCS Policlinico San Matteo, Pavia | Molecular Virology Unit, Microbiology and Virology Depertment, Fondazione IRCCS Policlinico San Matteo, Pavia | Monica Tallarita et al |
| EPI_ISL_1133202 | Molecular Virology Unit, Microbiology and Virology Depertment, Fondazione IRCCS Policlinico San Matteo, Pavia | Molecular Virology Unit, Microbiology and Virology Depertment, Fondazione IRCCS Policlinico San Matteo, Pavia | Monica Tallarita et al |
| EPI_ISL_417877 | Institute of Virology, Biomedical Research Center of the Slovak Academy of Sciences, Bratislava; Public Health Authority of the Slovak Republic, Bratislava | Institute of Virology, Biomedical Research Center of the Slovak Academy of Sciences, Bratislava; Comenius University Science Park, Bratislava | Monika Slávikova et al |
| EPI_ISL_417878 | Institute of Virology, Biomedical Research Center of the Slovak Academy of Sciences, Bratislava; Public Health Authority of the Slovak Republic, Bratislava | Institute of Virology, Biomedical Research Center of the Slovak Academy of Sciences, Bratislava; Comenius University Science Park, Bratislava | Monika Slávikova et al |
| EPI_ISL_417879 | Institute of Virology, Biomedical Research Center of the Slovak Academy of Sciences, Bratislava; Public Health Authority of the Slovak Republic, Bratislava | Institute of Virology, Biomedical Research Center of the Slovak Academy of Sciences, Bratislava; Comenius University Science Park, Bratislava | Monika Slávikova et al |
| EPI_ISL_417880 | Institute of Virology, Biomedical Research Center of the Slovak Academy of Sciences, Bratislava; Public Health Authority of the Slovak Republic, Bratislava | Institute of Virology, Biomedical Research Center of the Slovak Academy of Sciences, Bratislava; Comenius University Science Park, Bratislava | Monika Slávikova et al |
| EPI_ISL_1347884 | Instituto Nacional de Investigação em Saúde | KRISP, KZN Research Innovation and Sequencing Platform | Morais J et al |
| EPI_ISL_1347887 | Instituto Nacional de Investigação em Saúde | KRISP, KZN Research Innovation and Sequencing Platform | Morais J et al |
| EPI_ISL_1347896 | Instituto Nacional de Investigação em Saúde | KRISP, KZN Research Innovation and Sequencing Platform | Morais J et al |
| EPI_ISL_1347897 | Instituto Nacional de Investigação em Saúde | KRISP, KZN Research Innovation and Sequencing Platform | Morais J et al |
| EPI_ISL_1347898 | Instituto Nacional de Investigação em Saúde | KRISP, KZN Research Innovation and Sequencing Platform | Morais J et al |
| EPI_ISL_1347910 | Instituto Nacional de Investigação em Saúde | KRISP, KZN Research Innovation and Sequencing Platform | Morais J et al |
| EPI_ISL_1347935 | Instituto Nacional de Investigação em Saúde | KRISP, KZN Research Innovation and Sequencing Platform | Morais J et al |
| EPI_ISL_1347937 | Instituto Nacional de Investigação em Saúde | KRISP, KZN Research Innovation and Sequencing Platform | Morais J et al |
| EPI_ISL_1347940 | Instituto Nacional de Investigação em Saúde | KRISP, KZN Research Innovation and Sequencing Platform | Morais J et al |
| EPI_ISL_1347941 | Instituto Nacional de Investigação em Saúde | KRISP, KZN Research Innovation and Sequencing Platform | Morais J et al |
| EPI_ISL_1347942 | Instituto Nacional de Investigação em Saúde | KRISP, KZN Research Innovation and Sequencing Platform | Morais J et al |
| EPI_ISL_1545290 | Instituto Nacional de Investigação em Saúde | KRISP, KZN Research Innovation and Sequencing Platform | Morais J et al |
| EPI_ISL_1545304 | Instituto Nacional de Investigação em Saúde | KRISP, KZN Research Innovation and Sequencing Platform | Morais J et al |
| EPI_ISL_1545307 | Instituto Nacional de Investigação em Saúde | KRISP, KZN Research Innovation and Sequencing Platform | Morais J et al |

| | | | |
|---|---|---|---|
| EPI_ISL_1545308 | Instituto Nacional de Investigação em Saúde | KRISP, KZN Research Innovation and Sequencing Platform | Morais J et al |
| EPI_ISL_1545309 | Instituto Nacional de Investigação em Saúde | KRISP, KZN Research Innovation and Sequencing Platform | Morais J et al |
| EPI_ISL_1545313 | Instituto Nacional de Investigação em Saúde | KRISP, KZN Research Innovation and Sequencing Platform | Morais J et al |
| EPI_ISL_1545321 | Instituto Nacional de Investigação em Saúde | KRISP, KZN Research Innovation and Sequencing Platform | Morais J et al |
| EPI_ISL_1545323 | Instituto Nacional de Investigação em Saúde | KRISP, KZN Research Innovation and Sequencing Platform | Morais J et al |
| EPI_ISL_1545325 | Instituto Nacional de Investigação em Saúde | KRISP, KZN Research Innovation and Sequencing Platform | Morais J et al |
| EPI_ISL_1545326 | Instituto Nacional de Investigação em Saúde | KRISP, KZN Research Innovation and Sequencing Platform | Morais J et al |
| EPI_ISL_1545353 | Instituto Nacional de Investigação em Saúde | KRISP, KZN Research Innovation and Sequencing Platform | Morais J et al |
| EPI_ISL_1545370 | Instituto Nacional de Investigação em Saúde | KRISP, KZN Research Innovation and Sequencing Platform | Morais J et al |
| EPI_ISL_1545372 | Instituto Nacional de Investigação em Saúde | KRISP, KZN Research Innovation and Sequencing Platform | Morais J et al |
| EPI_ISL_944663 | Department of Biochemistry, Cell and Molecular Biology, West African Centre for Cell Biology of Infectious Pathogens (WACCBIP), University of Ghana | Department of Biochemistry, Cell and Molecular Biology, West African Centre for Cell Biology of Infectious Pathogens (WACCBIP), University of Ghana | Morang'a et al |
| EPI_ISL_944662 | Department of Biochemistry, Cell and Molecular Biology, West African Centre for Cell Biology of Infectious Pathogens (WACCBIP), University of Ghana | Department of Biochemistry, Cell and Molecular Biology, West African Centre for Cell Biology of Infectious Pathogens (WACCBIP), University of Ghana | Morang'a et al |
| EPI_ISL_944735 | Department of Biochemistry, Cell and Molecular Biology, West African Centre for Cell Biology of Infectious Pathogens (WACCBIP), University of Ghana | Department of Biochemistry, Cell and Molecular Biology, West African Centre for Cell Biology of Infectious Pathogens (WACCBIP), University of Ghana | Morang'a et al |
| EPI_ISL_416142 | Department of Virus and Microbiological Special diagnostics, Statens Serum Institut, Copenhagen, Denmark. | Statens Serum Institute | Morten Rasmussen et al |
| EPI_ISL_416143 | Department of Virus and Microbiological Special diagnostics, Statens Serum Institut, Copenhagen, Denmark. | ViFU | Morten Rasmussen et al |
| EPI_ISL_769863 | Laboratoire Biolife | Laboratoire de Biotechnologie | Mouna Ouadghiri et al |
| EPI_ISL_775221 | Laboratoire Biolife | Laboratoire de Biotechnologie | Mouna Ouadghiri et al |
| EPI_ISL_775256 | Laboratoire Biolife | Laboratoire de Biotechnologie | Mouna Ouadghiri et al |
| EPI_ISL_775258 | Laboratoire Biolife | Laboratoire de Biotechnologie | Mouna Ouadghiri et al |
| EPI_ISL_775263 | Laboratoire Biolife | Laboratoire de Biotechnologie | Mouna Ouadghiri et al |
| EPI_ISL_1415293 | Institut National d'hygiène | Unité Mixte Internationale TransVIHMI (UMI 233 IRD – U1175 INSERM - Université de Montpellier) IRD (Institut de recherche pour le développement) | Mounerou SALOU et al |
| EPI_ISL_1440879 | Institut National d'Hygiène | Unité Mixte Internationale TransVIHMI (UMI 233 IRD – U1175 INSERM - Université de Montpellier) IRD (Institut de recherche pour le développement) | Mounerou SALOU et al |
| EPI_ISL_1442993 | Institut National d'hygiène | Unité Mixte Internationale TransVIHMI (UMI 233 IRD – U1175 INSERM - Université de Montpellier) IRD (Institut de recherche pour le développement) | Mounerou SALOU et al |

| | | | |
|---|---|---|---|
| EPI_ISL_1416192 | Institut National d'hygiène | Unité Mixte Internationale TransVIHMI (UMI 233 IRD – U1175 INSERM - Université de Montpellier) IRD (Institut de recherche pour le développement) | Mounerou SALOU et al |
| EPI_ISL_1416222 | Institut National d'hygiène | Unité Mixte Internationale TransVIHMI (UMI 233 IRD – U1175 INSERM - Université de Montpellier) IRD (Institut de recherche pour le développement) | Mounerou SALOU et al |
| EPI_ISL_1433132 | Institut National d'Hygiène | Unité Mixte Internationale TransVIHMI (UMI 233 IRD – U1175 INSERM - Université de Montpellier) IRD (Institut de recherche pour le développement) | Mounerou SALOU et al |
| EPI_ISL_1434443 | Institut National d'Hygiène | Unité Mixte Internationale TransVIHMI (UMI 233 IRD – U1175 INSERM - Université de Montpellier) IRD (Institut de recherche pour le développement) | Mounerou SALOU et al |
| EPI_ISL_1435763 | Institut National d'Hygiène | Unité Mixte Internationale TransVIHMI (UMI 233 IRD – U1175 INSERM - Université de Montpellier) IRD (Institut de recherche pour le développement) | Mounerou SALOU et al |
| EPI_ISL_1436817 | Institut National d'Hygiène | Unité Mixte Internationale TransVIHMI (UMI 233 IRD – U1175 INSERM - Université de Montpellier) IRD (Institut de recherche pour le développement) | Mounerou SALOU et al |
| EPI_ISL_1303031 | Institut National d'Hygiène (INH) | Unité Mixte Internationale TransVIHMI (UMI 233 IRD – U1175 INSERM - Université de Montpellier)IRD (Institut de recherche pour le développement) | Mounerou SALOU et al |
| EPI_ISL_1443001 | Institut National d'hygiène | Unité Mixte Internationale TransVIHMI (UMI 233 IRD – U1175 INSERM - Université de Montpellier) IRD (Institut de recherche pour le développement) | Mounerou SALOU et al |
| EPI_ISL_1437749 | Institut National d'Hygiène | Unité Mixte Internationale TransVIHMI (UMI 233 IRD – U1175 INSERM - Université de Montpellier) IRD (Institut de recherche pour le développement) | Mounerou SALOU et al |
| EPI_ISL_1439647 | Institut National d'Hygiène | Unité Mixte Internationale TransVIHMI (UMI 233 IRD – U1175 INSERM - Université de Montpellier) IRD (Institut de recherche pour le développement) | Mounerou SALOU et al |
| EPI_ISL_1443002 | Institut National d'hygiène | Unité Mixte Internationale TransVIHMI (UMI 233 IRD – U1175 INSERM - Université de Montpellier) IRD (Institut de recherche pour le développement) | Mounerou SALOU et al |
| EPI_ISL_1443003 | Institut National d'hygiène | Unité Mixte Internationale TransVIHMI (UMI 233 IRD – U1175 INSERM - Université de Montpellier) IRD (Institut de recherche pour le développement) | Mounerou SALOU et al |
| EPI_ISL_1445275 | Institut National d'Hygiène (INH) | Unité Mixte Internationale TransVIHMI (UMI 233 IRD – U1175 INSERM - Université de Montpellier)IRD (Institut de recherche pour le développement) | Mounerou SALOU et al |
| EPI_ISL_1438551 | Institut National d'Hygiène | Unité Mixte Internationale TransVIHMI (UMI 233 IRD – U1175 INSERM - Université de Montpellier) IRD (Institut de recherche pour le développement) | Mounerou SALOU et al |
| EPI_ISL_1321604 | Institut National d'Hygiène (INH) | Unité Mixte Internationale TransVIHMI (UMI 233 IRD – U1175 INSERM - Université de Montpellier)IRD (Institut de recherche pour le développement) | Mounerou SALOU et al |
| EPI_ISL_1404615 | Institut National d'Hygiène | Unité Mixte Internationale TransVIHMI (UMI 233 IRD – U1175 INSERM - Université de Montpellier) IRD (Institut de recherche pour le développement) | Mounerou SALOU et al |

| | | | |
|---|---|---|---|
| EPI_ISL_1406144 | Institut National d’Hygiène (INH) | Unité Mixte Internationale TransVIHMI (UMI 233 IRD – U1175 INSERM - Université de Montpellier)IRD (Institut de recherche pour le développement) | Mounerou SALOU et al |
| EPI_ISL_1293351 | Institut National d’Hygiène (INH) | Unité Mixte Internationale TransVIHMI (UMI 233 IRD – U1175 INSERM - Université de Montpellier)IRD (Institut de recherche pour le développement) | Mounerou SALOU et al |
| EPI_ISL_1587449 | Laboratoire Biolim/FSS/UL | Unité Mixte Internationale TransVIHMI (UMI 233 IRD – U1175 INSERM - Université de Montpellier) IRD (Institut de recherche pour le développement) | Mounerou SALOU et al |
| EPI_ISL_1654210 | Laboratoire Biolim/FSS/UL | Unité Mixte Internationale TransVIHMI (UMI 233 IRD – U1175 INSERM - Université de Montpellier) IRD (Institut de recherche pour le développement) | Mounerou SALOU et al |
| EPI_ISL_1575297 | Laboratoire Biolim/FSS/UL | Unité Mixte Internationale TransVIHMI (UMI 233 IRD – U1175 INSERM - Université de Montpellier) IRD (Institut de recherche pour le développement) | Mounerou SALOU et al |
| EPI_ISL_1575310 | Laboratoire Biolim/FSS/UL | Unité Mixte Internationale TransVIHMI (UMI 233 IRD – U1175 INSERM - Université de Montpellier) IRD (Institut de recherche pour le développement) | Mounerou SALOU et al |
| EPI_ISL_1575436 | Laboratoire Biolim/FSS/UL | Unité Mixte Internationale TransVIHMI (UMI 233 IRD – U1175 INSERM - Université de Montpellier) IRD (Institut de recherche pour le développement) | Mounerou SALOU et al |
| EPI_ISL_1575441 | Laboratoire Biolim/FSS/UL | Unité Mixte Internationale TransVIHMI (UMI 233 IRD – U1175 INSERM - Université de Montpellier) IRD (Institut de recherche pour le développement) | Mounerou SALOU et al |
| EPI_ISL_1508956 | Laboratoire Biolim/FSS/UL | Unité Mixte Internationale TransVIHMI (UMI 233 IRD – U1175 INSERM - Université de Montpellier) IRD (Institut de recherche pour le développement) | Mounerou SALOU et al |
| EPI_ISL_1500948 | Institut National d’Hygiène (INH) | Unité Mixte Internationale TransVIHMI (UMI 233 IRD – U1175 INSERM - Université de Montpellier)IRD (Institut de recherche pour le développement) | Mounerou SALOU et al |
| EPI_ISL_1508849 | Laboratoire Biolim/FSS/UL | Unité Mixte Internationale TransVIHMI (UMI 233 IRD – U1175 INSERM - Université de Montpellier) IRD (Institut de recherche pour le développement) | Mounerou SALOU et al |
| EPI_ISL_1508890 | Laboratoire Biolim/FSS/UL | Unité Mixte Internationale TransVIHMI (UMI 233 IRD – U1175 INSERM - Université de Montpellier) IRD (Institut de recherche pour le développement) | Mounerou SALOU et al |
| EPI_ISL_1585941 | Laboratoire Biolim/FSS/UL | Unité Mixte Internationale TransVIHMI (UMI 233 IRD – U1175 INSERM - Université de Montpellier) IRD (Institut de recherche pour le développement) | Mounerou SALOU et al |
| EPI_ISL_1508894 | Laboratoire Biolim/FSS/UL | Unité Mixte Internationale TransVIHMI (UMI 233 IRD – U1175 INSERM - Université de Montpellier) IRD (Institut de recherche pour le développement) | Mounerou SALOU et al |
| EPI_ISL_1508916 | Laboratoire Biolim/FSS/UL | Unité Mixte Internationale TransVIHMI (UMI 233 IRD – U1175 INSERM - Université de Montpellier) IRD (Institut de recherche pour le développement) | Mounerou SALOU et al |
| EPI_ISL_1508945 | Laboratoire Biolim/FSS/UL | Unité Mixte Internationale TransVIHMI (UMI 233 IRD – U1175 INSERM - Université de Montpellier) IRD (Institut de recherche pour le développement) | Mounerou SALOU et al |

| | | | |
|---|---|---|---|
| EPI_ISL_1508959 | Laboratoire Biolim/FSS/UL | Unité Mixte Internationale TransVIHMI (UMI 233 IRD – U1175 INSERM - Université de Montpellier) IRD (Institut de recherche pour le développement) | Mounerou SALOU et al |
| EPI_ISL_1509102 | Laboratoire Biolim/FSS/UL | Unité Mixte Internationale TransVIHMI (UMI 233 IRD – U1175 INSERM - Université de Montpellier) IRD (Institut de recherche pour le développement) | Mounerou SALOU et al |
| EPI_ISL_1524717 | Laboratoire Biolim/FSS/UL | Unité Mixte Internationale TransVIHMI (UMI 233 IRD – U1175 INSERM - Université de Montpellier) IRD (Institut de recherche pour le développement) | Mounerou SALOU et al |
| EPI_ISL_1508949 | Laboratoire Biolim/FSS/UL | Unité Mixte Internationale TransVIHMI (UMI 233 IRD – U1175 INSERM - Université de Montpellier) IRD (Institut de recherche pour le développement) | Mounerou SALOU et al |
| EPI_ISL_1508953 | Institut National d'hygiène | Unité Mixte Internationale TransVIHMI (UMI 233 IRD – U1175 INSERM - Université de Montpellier) IRD (Institut de recherche pour le développement) | Mounerou SALOU et al |
| EPI_ISL_1508958 | Institut National d'hygiène | Unité Mixte Internationale TransVIHMI (UMI 233 IRD – U1175 INSERM - Université de Montpellier) IRD (Institut de recherche pour le développement) | Mounerou SALOU et al |
| EPI_ISL_1622433 | RSJ Dr. Soeharto Heerdjan | Eijkman Institute for Molecular Biology, Ministry of Research and Technology/National Agency for Research and Innovation | Muhammad Rezki Rasyak et al |
| EPI_ISL_678211 | Pathogen Genomics Lab King Abdullah University of Science and Technology (KAUST) | Pathogen Genomics Lab King Abdullah University of Science and Technology(KAUST) | Muhammad Shuaib et al |
| EPI_ISL_678238 | Pathogen Genomics Lab King Abdullah University of Science and Technology (KAUST) | Pathogen Genomics Lab King Abdullah University of Science and Technology(KAUST) | Muhammad Shuaib et al |
| EPI_ISL_495423 | Kafkas University, Faculty of Medicine, Department of Medical Microbiology | Kafkas University, Faculty of Medicine, Department of Medical Microbiology | Murat Karamese et al |
| EPI_ISL_1080932 | National Health Laboratory Services | National Health Laboratory Services | Mushal Ali et al |
| EPI_ISL_1407113 | National HIV Reference Laboratory, Ministry of Health, Public Health Institute of Malawi | KRISP, KZN Research Innovation and Sequencing Platform | Mvula B et al |
| EPI_ISL_1407115 | National HIV Reference Laboratory, Ministry of Health, Public Health Institute of Malawi | KRISP, KZN Research Innovation and Sequencing Platform | Mvula B et al |
| EPI_ISL_1407116 | National HIV Reference Laboratory, Ministry of Health, Public Health Institute of Malawi | KRISP, KZN Research Innovation and Sequencing Platform | Mvula B et al |
| EPI_ISL_1407131 | National HIV Reference Laboratory, Ministry of Health, Public Health Institute of Malawi | KRISP, KZN Research Innovation and Sequencing Platform | Mvula B et al |
| EPI_ISL_1407148 | National HIV Reference Laboratory, Ministry of Health, Public Health Institute of Malawi | KRISP, KZN Research Innovation and Sequencing Platform | Mvula B et al |
| EPI_ISL_1407151 | National HIV Reference Laboratory, Ministry of Health, Public Health Institute of Malawi | KRISP, KZN Research Innovation and Sequencing Platform | Mvula B et al |
| EPI_ISL_1407158 | National HIV Reference Laboratory, Ministry of Health, Public Health Institute of Malawi | KRISP, KZN Research Innovation and Sequencing Platform | Mvula B et al |
| EPI_ISL_1407201 | National HIV Reference Laboratory, Ministry of Health, Public Health Institute of Malawi | KRISP, KZN Research Innovation and Sequencing Platform | Mvula B et al |
| EPI_ISL_1407212 | National HIV Reference Laboratory, Ministry of Health, Public Health Institute of Malawi | KRISP, KZN Research Innovation and Sequencing Platform | Mvula B et al |

| | | | |
|---|---|---|---|
| EPI_ISL_1407253 | National HIV Reference Laboratory, Ministry of Health, Public Health Institute of Malawi | KRISP, KZN Research Innovation and Sequencing Platform | Mvula B et al |
| EPI_ISL_512844 | Department of Medical Research | DMR_Myanmar | Myat Htut Nyunt et al |
| EPI_ISL_1103569 | Laboratoire central de Virologie, Centre hospitalo-universitaire Ibn Sina, Hopital des spécialités, Rabat | Laboratoire de Biotechnologie | Myriam Seffar et al |
| EPI_ISL_1103576 | Laboratoire central de Virologie | Laboratoire de Biotechnologie | Myriam Seffar et al |
| EPI_ISL_1103579 | Laboratoire central de Virologie, Centre hospitalo-universitaire Ibn Sina, Hôpital des spécialités | Laboratoire de Biotechnologie | Myriam Seffar et al |
| EPI_ISL_793013 | Laboratorio de Virología del Hospital de Niños Dr. Ricardo Gutierrez | Área de Secuenciación del Laboratorio de Virología del Hospital de Niños Dr. Ricardo Gutierrez on behalf of 'Proyecto Argentino Interinstitucional de genomica de SARS-CoV-2' (PAIS Consortium) | Nabaes Jodar et al |
| EPI_ISL_430815 | Laboratorio de Virología del Hospital de Niños Dr. Ricardo Gutierrez | Área de Secuenciación del Laboratorio de Virología del Hospital de Niños Dr. Ricardo Gutierrez on behalf of 'Proyecto Argentino Interinstitucional de genomica de SARS-CoV-2' (PAIS Consortium) | Nabaes Jodar et al |
| EPI_ISL_430817 | Laboratorio de Virología del Hospital de Niños Dr. Ricardo Gutierrez | Área de Secuenciación del Laboratorio de Virología del Hospital de Niños Dr. Ricardo Gutierrez on behalf of 'Proyecto Argentino Interinstitucional de genomica de SARS-CoV-2' (PAIS Consortium) | Nabaes Jodar et al |
| EPI_ISL_792223 | Laboratorio de Virología del Hospital de Niños Dr. Ricardo Gutierrez | Área de Secuenciación del Laboratorio de Virología del Hospital de Niños Dr. Ricardo Gutierrez on behalf of 'Proyecto Argentino Interinstitucional de genomica de SARS-CoV-2' (PAIS Consortium) | Nabaes Jodar et al |
| EPI_ISL_792235 | Laboratorio de Virología del Hospital de Niños Dr. Ricardo Gutierrez | Área de Secuenciación del Laboratorio de Virología del Hospital de Niños Dr. Ricardo Gutierrez on behalf of 'Proyecto Argentino Interinstitucional de genomica de SARS-CoV-2' (PAIS Consortium) | Nabaes Jodar et al |
| EPI_ISL_792302 | Unidad de Virología, Centro de Educación Médica en Investigaciones Clínicas CEMIC | Área de Secuenciación del Laboratorio de Virología del Hospital de Niños Dr. Ricardo Gutierrez on behalf of 'Proyecto Argentino Interinstitucional de genomica de SARS-CoV-2' (PAIS Consortium) | Nabaes Jodar et al |
| EPI_ISL_792303 | Unidad de Virología, Centro de Educación Médica en Investigaciones Clínicas CEMIC | Área de Secuenciación del Laboratorio de Virología del Hospital de Niños Dr. Ricardo Gutierrez on behalf of 'Proyecto Argentino Interinstitucional de genomica de SARS-CoV-2' (PAIS Consortium) | Nabaes Jodar et al |
| EPI_ISL_792317 | Laboratorio del Hospital El Cruce Dr. Néstor C. Kirchner | Área de Secuenciación del Laboratorio de Virología del Hospital de Niños Dr. Ricardo Gutierrez on behalf of 'Proyecto Argentino Interinstitucional de genomica de SARS-CoV-2' (PAIS Consortium) | Nabaes Jodar et al |
| EPI_ISL_792335 | Laboratorio del Hospital Interzonal General de Agudos Evita | Área de Secuenciación del Laboratorio de Virología del Hospital de Niños Dr. Ricardo Gutierrez on behalf of 'Proyecto Argentino Interinstitucional de genomica de SARS-CoV-2' (PAIS Consortium) | Nabaes Jodar et al |
| EPI_ISL_792336 | Laboratorio del Hospital Interzonal General de Agudos Evita | Área de Secuenciación del Laboratorio de Virología del Hospital de Niños Dr. Ricardo Gutierrez on behalf of 'Proyecto Argentino Interinstitucional de genomica de SARS-CoV-2' (PAIS Consortium) | Nabaes Jodar et al |
| EPI_ISL_792346 | Laboratorio del Hospital Interzonal General de Agudos Evita | Área de Secuenciación del Laboratorio de Virología del Hospital de Niños Dr. Ricardo Gutierrez on behalf of 'Proyecto Argentino Interinstitucional de genomica de SARS-CoV-2' (PAIS Consortium) | Nabaes Jodar et al |
| EPI_ISL_792376 | Laboratorio de salud pública, Facultad de Ciencias Exactas, Universidad Nacional de La Plata | Área de Secuenciación del Laboratorio de Virología del Hospital de Niños Dr. Ricardo Gutierrez on behalf of 'Proyecto Argentino Interinstitucional de genomica de SARS-CoV-2' (PAIS Consortium) | Nabaes Jodar et al |

| | | | |
|---|---|---|---|
| EPI_ISL_792377 | Laboratorio de salud pública, Facultad de Ciencias Exactas, Universidad Nacional de La Plata | Área de Secuenciación del Laboratorio de Virología del Hospital de Niños Dr. Ricardo Gutierrez on behalf of 'Proyecto Argentino Interinstitucional de genomica de SARS-CoV-2' (PAIS Consortium) | Nabaes Jodar et al |
| EPI_ISL_792380 | Laboratorio de salud pública, Facultad de Ciencias Exactas, Universidad Nacional de La Plata | Área de Secuenciación del Laboratorio de Virología del Hospital de Niños Dr. Ricardo Gutierrez on behalf of 'Proyecto Argentino Interinstitucional de genomica de SARS-CoV-2' (PAIS Consortium) | Nabaes Jodar et al |
| EPI_ISL_792383 | Laboratorio de salud pública, Facultad de Ciencias Exactas, Universidad Nacional de La Plata | Área de Secuenciación del Laboratorio de Virología del Hospital de Niños Dr. Ricardo Gutierrez on behalf of 'Proyecto Argentino Interinstitucional de genomica de SARS-CoV-2' (PAIS Consortium) | Nabaes Jodar et al |
| EPI_ISL_792385 | Laboratorio de salud pública, Facultad de Ciencias Exactas, Universidad Nacional de La Plata | Área de Secuenciación del Laboratorio de Virología del Hospital de Niños Dr. Ricardo Gutierrez on behalf of 'Proyecto Argentino Interinstitucional de genomica de SARS-CoV-2' (PAIS Consortium) | Nabaes Jodar et al |
| EPI_ISL_792395 | Plataforma de Servicios Biotecnólogicos: UTTIPP/PSB , Universidad Nacional de Quilmes. | Área de Secuenciación del Laboratorio de Virología del Hospital de Niños Dr. Ricardo Gutierrez on behalf of 'Proyecto Argentino Interinstitucional de genomica de SARS-CoV-2' (PAIS Consortium) | Nabaes Jodar et al |
| EPI_ISL_971451 | Cell culture Unit at CV-MIT belonging to HIMMV | Functional Genomic Platform UATRS-biology, CNRST | Nadia Touil et al |
| EPI_ISL_851051 | Gandhi Medical College and Hospital | CSIR-Centre for Cellular and Molecular Biology | Nagamani Kammili et al |
| EPI_ISL_850514 | Gandhi Medical College and Hospital | MDRU-DHR,Gandhi Medical College | Nagamani Kammili et al |
| EPI_ISL_660557 | The National Institute of Public Health | State Veterinary Institute Prague | Nagy et al |
| EPI_ISL_693664 | The National Institute of Public Health | State Veterinary Institute Prague | Nagy et al |
| EPI_ISL_693672 | The National Institute of Public Health | State Veterinary Institute Prague | Nagy et al |
| EPI_ISL_693663 | The National Institute of Public Health | State Veterinary Institute Prague | Nagy et al |
| EPI_ISL_1510357 | National Institute of Public Health | State Veterinary Institute Prague | Nagy et al |
| EPI_ISL_584080 | The National Institute of Public Health | State Veterinary Institute Prague | Nagy et al |
| EPI_ISL_584078 | The National Institute of Public Health | State Veterinary Institute Prague | Nagy et al |
| EPI_ISL_737014 | The National Institute of Public Health | State Veterinary Institute Prague | Nagy et al |
| EPI_ISL_737022 | The National Institute of Public Health | State Veterinary Institute Prague | Nagy et al |
| EPI_ISL_792690 | The National Institute of Public Health | State Veterinary Institute Prague | Nagy et al |
| EPI_ISL_792707 | The National Institute of Public Health | State Veterinary Institute Prague | Nagy et al |
| EPI_ISL_792697 | The National Institute of Public Health | State Veterinary Institute Prague | Nagy et al |
| EPI_ISL_960414 | The National Institute of Public Health | State Veterinary Institute Prague | Nagy et al |
| EPI_ISL_960423 | The National Institute of Public Health | State Veterinary Institute Prague | Nagy et al |
| EPI_ISL_960438 | The National Institute of Public Health | State Veterinary Institute Prague | Nagy et al |
| EPI_ISL_960424 | The National Institute of Public Health | State Veterinary Institute Prague | Nagy et al |
| EPI_ISL_882953 | The National Institute of Public Health | State Veterinary Institute Prague | Nagy et al |
| EPI_ISL_577626 | The National Institute of Public Health | State Veterinary Institute Prague | Nagy et al |

| | | | |
|---|---|---|---|
| EPI_ISL_577638 | The National Institute of Public Health | State Veterinary Institute Prague | Nagy et al |
| EPI_ISL_887455 | Instituto Nacional de Saude (INS), Mozambique | KRISP, KZN Research Innovation and Sequencing Platform | Nalia Ismael et al |
| EPI_ISL_887456 | Instituto Nacional de Saude (INS), Mozambique | KRISP, KZN Research Innovation and Sequencing Platform | Nalia Ismael et al |
| EPI_ISL_887458 | Instituto Nacional de Saude (INS), Mozambique | KRISP, KZN Research Innovation and Sequencing Platform | Nalia Ismael et al |
| EPI_ISL_887459 | Instituto Nacional de Saude (INS), Mozambique | KRISP, KZN Research Innovation and Sequencing Platform | Nalia Ismael et al |
| EPI_ISL_887460 | Instituto Nacional de Saude (INS), Mozambique | KRISP, KZN Research Innovation and Sequencing Platform | Nalia Ismael et al |
| EPI_ISL_887461 | Instituto Nacional de Saude (INS), Mozambique | KRISP, KZN Research Innovation and Sequencing Platform | Nalia Ismael et al |
| EPI_ISL_887464 | Instituto Nacional de Saude (INS), Mozambique | KRISP, KZN Research Innovation and Sequencing Platform | Nalia Ismael et al |
| EPI_ISL_887474 | Instituto Nacional de Saude (INS), Mozambique | KRISP, KZN Research Innovation and Sequencing Platform | Nalia Ismael et al |
| EPI_ISL_887428 | Instituto Nacional de Saude (INS), Mozambique | KRISP, KZN Research Innovation and Sequencing Platform | Nalia Ismael et al |
| EPI_ISL_887444 | Instituto Nacional de Saude (INS), Mozambique | KRISP, KZN Research Innovation and Sequencing Platform | Nalia Ismael et al |
| EPI_ISL_887445 | Instituto Nacional de Saude (INS), Mozambique | KRISP, KZN Research Innovation and Sequencing Platform | Nalia Ismael et al |
| EPI_ISL_887447 | Instituto Nacional de Saude (INS), Mozambique | KRISP, KZN Research Innovation and Sequencing Platform | Nalia Ismael et al |
| EPI_ISL_887420 | Instituto Nacional de Saude (INS), Mozambique | KRISP, KZN Research Innovation and Sequencing Platform | Nalia Ismael et al |
| EPI_ISL_887500 | Instituto Nacional de Saude (INS), Mozambique | KRISP, KZN Research Innovation and Sequencing Platform | Nalia Ismael et al |
| EPI_ISL_887501 | Instituto Nacional de Saude (INS), Mozambique | KRISP, KZN Research Innovation and Sequencing Platform | Nalia Ismael et al |
| EPI_ISL_887421 | Instituto Nacional de Saude (INS), Mozambique | KRISP, KZN Research Innovation and Sequencing Platform | Nalia Ismael et al |
| EPI_ISL_887434 | Instituto Nacional de Saude (INS), Mozambique | KRISP, KZN Research Innovation and Sequencing Platform | Nalia Ismael et al |
| EPI_ISL_887431 | Instituto Nacional de Saude (INS), Mozambique | KRISP, KZN Research Innovation and Sequencing Platform | Nalia Ismael et al |
| EPI_ISL_887426 | Instituto Nacional de Saude (INS), Mozambique | KRISP, KZN Research Innovation and Sequencing Platform | Nalia Ismael et al |
| EPI_ISL_887502 | Instituto Nacional de Saude (INS), Mozambique | KRISP, KZN Research Innovation and Sequencing Platform | Nalia Ismael et al |
| EPI_ISL_887503 | Instituto Nacional de Saude (INS), Mozambique | KRISP, KZN Research Innovation and Sequencing Platform | Nalia Ismael et al |
| EPI_ISL_887429 | Instituto Nacional de Saude (INS), Mozambique | KRISP, KZN Research Innovation and Sequencing Platform | Nalia Ismael et al |
| EPI_ISL_887433 | Instituto Nacional de Saude (INS), Mozambique | KRISP, KZN Research Innovation and Sequencing Platform | Nalia Ismael et al |
| EPI_ISL_887430 | Instituto Nacional de Saude (INS), Mozambique | KRISP, KZN Research Innovation and Sequencing Platform | Nalia Ismael et al |
| EPI_ISL_964923 | Instituto Nacional de Saude (INS), Mozambique | KRISP, KZN Research Innovation and Sequencing Platform | Nalia Ismael et al |
| EPI_ISL_1132824 | Instituto Nacional de Saude (INS), Mozambique | KRISP, KZN Research Innovation and Sequencing Platform | Nalia Ismael et al |
| EPI_ISL_1132836 | Instituto Nacional de Saude (INS), Mozambique | KRISP, KZN Research Innovation and Sequencing Platform | Nalia Ismael et al |
| EPI_ISL_1132843 | Instituto Nacional de Saude (INS), Mozambique | KRISP, KZN Research Innovation and Sequencing Platform | Nalia Ismael et al |
| EPI_ISL_626569 | National Centre for Communicable Disease (NCCD) National Influenza Center | National Centre for Communicable Disease (NCCD) National Influenza Center | Naranzul Ts et al |

| | | | |
|---|---|---|---|
| EPI_ISL_626566 | National Centre for Communication Disease (NCCD) National Influenza Center | National Centre for Communication Disease (NCCD) National Influenza Center | Naranzul Ts et al |
| EPI_ISL_626626 | National Centre for Communicable Disease (NCCD) National Influenza Center | National Centre for Communicable Disease (NCCD) National Influenza Center | Naranzul Ts et al |
| EPI_ISL_626627 | National Centre for Communicable Disease (NCCD) National Influenza Center | National Centre for Communicable Disease (NCCD) National Influenza Center | Naranzul Ts et al |
| EPI_ISL_467666 | Virology lab, NIC, NCCD, Ulaanbaatar, Mongolia | National Centre for Communicable Deseases (NCCD) | Naranzul Ts et al |
| EPI_ISL_1502568 | Biochemistry and Molecular Biology Department-Faculty of Medicine, Al-Quds University | Biochemistry and Molecular Biology Department-Faculty of Medicine, Al-Quds University | Nasereddin et al |
| EPI_ISL_1502570 | Biochemistry and Molecular Biology Department-Faculty of Medicine, Al-Quds University | Biochemistry and Molecular Biology Department-Faculty of Medicine, Al-Quds University | Nasereddin et al |
| EPI_ISL_1502574 | Biochemistry and Molecular Biology Department-Faculty of Medicine, Al-Quds University | Biochemistry and Molecular Biology Department-Faculty of Medicine, Al-Quds University | Nasereddin et al |
| EPI_ISL_1502578 | Biochemistry and Molecular Biology Department-Faculty of Medicine, Al-Quds University | Biochemistry and Molecular Biology Department-Faculty of Medicine, Al-Quds University | Nasereddin et al |
| EPI_ISL_1395939 | Laboratorio Central de Salud Pública | Grupo de Genómica y Bioinformática del Instituto de Investigación de la Cadena Láctea CONICET-INTA on behalf of 'Proyecto Argentino Interinstitucional de genomica de SARS-CoV-2' (PAIS Consortium) | Natalia Andrea Ayala y María Verónica Góm |
| EPI_ISL_941111 | Centro de Investigaciones en Microbiología y Biotecnología-UR (CIMBIUR), Facultad de Ciencias Naturales, Universidad del Rosario, Bogotá, Colombia | Centro de Investigaciones en Microbiología y Biotecnología-UR (CIMBIUR), Facultad de Ciencias Naturales, Universidad del Rosario, Bogotá, Colombia Icahn School of Medicine at Mount Sinai, New York, USA | Nathalia Ballesteros et al |
| EPI_ISL_941114 | Centro de Investigaciones en Microbiología y Biotecnología-UR (CIMBIUR), Facultad de Ciencias Naturales, Universidad del Rosario, Bogotá, Colombia | Centro de Investigaciones en Microbiología y Biotecnología-UR (CIMBIUR), Facultad de Ciencias Naturales, Universidad del Rosario, Bogotá, Colombia Icahn School of Medicine at Mount Sinai, New York, USA | Nathalia Ballesteros et al |
| EPI_ISL_941123 | Centro de Investigaciones en Microbiología y Biotecnología-UR (CIMBIUR), Facultad de Ciencias Naturales, Universidad del Rosario, Bogotá, Colombia | Centro de Investigaciones en Microbiología y Biotecnología-UR (CIMBIUR), Facultad de Ciencias Naturales, Universidad del Rosario, Bogotá, Colombia Icahn School of Medicine at Mount Sinai, New York, USA | Nathalia Ballesteros et al |
| EPI_ISL_941139 | Centro de Investigaciones en Microbiología y Biotecnología-UR (CIMBIUR), Facultad de Ciencias Naturales, Universidad del Rosario, Bogotá, Colombia | Centro de Investigaciones en Microbiología y Biotecnología-UR (CIMBIUR), Facultad de Ciencias Naturales, Universidad del Rosario, Bogotá, Colombia Icahn School of Medicine at Mount Sinai, New York, USA | Nathalia Ballesteros et al |
| EPI_ISL_941140 | Centro de Investigaciones en Microbiología y Biotecnología-UR (CIMBIUR), Facultad de Ciencias Naturales, Universidad del Rosario, Bogotá, Colombia | Centro de Investigaciones en Microbiología y Biotecnología-UR (CIMBIUR), Facultad de Ciencias Naturales, Universidad del Rosario, Bogotá, Colombia Icahn School of Medicine at Mount Sinai, New York, USA | Nathalia Ballesteros et al |
| EPI_ISL_941149 | Centro de Investigaciones en Microbiología y Biotecnología-UR (CIMBIUR), Facultad de Ciencias Naturales, Universidad del Rosario, Bogotá, Colombia | Centro de Investigaciones en Microbiología y Biotecnología-UR (CIMBIUR), Facultad de Ciencias Naturales, Universidad del Rosario, Bogotá, Colombia Icahn School of Medicine at Mount Sinai, New York, USA | Nathalia Ballesteros et al |
| EPI_ISL_941154 | Centro de Investigaciones en Microbiología y Biotecnología-UR (CIMBIUR), Facultad de Ciencias Naturales, Universidad del Rosario, Bogotá, Colombia | Centro de Investigaciones en Microbiología y Biotecnología-UR (CIMBIUR), Facultad de Ciencias Naturales, Universidad del Rosario, Bogotá, Colombia Icahn School of Medicine at Mount Sinai, New York, USA | Nathalia Ballesteros et al |
| EPI_ISL_415643 | R. G. Lugar Center for Public Health Research, National Center for Disease Control and Public Health (NCDC) of Georgia. | R. G. Lugar Center for Public Health Research, National Center for Disease Control and Public Health (NCDC) of Georgia. | Nato Kotaria et al |
| EPI_ISL_415641 | R. G. Lugar Center for Public Health Research, National Center for Disease Control and Public Health (NCDC) of Georgia. | R. G. Lugar Center for Public Health Research, National Center for Disease Control and Public Health (NCDC) of Georgia. | Nato Kotaria et al |
| EPI_ISL_415644 | R. G. Lugar Center for Public Health Research, National Center for Disease Control and Public Health (NCDC) of Georgia. | R. G. Lugar Center for Public Health Research, National Center for Disease Control and Public Health (NCDC) of Georgia. | Nato Kotaria et al |

ıez et al

| | | | |
|---|---|---|---|
| EPI_ISL_418206 | Institut Pasteur Dakar | Institut Pasteur de Dakar | Ndongo Dia et al |
| EPI_ISL_418216 | Institut Pasteur Dakar | Institut Pasteur de Dakar | Ndongo Dia et al |
| EPI_ISL_480789 | Institut Pasteur Dakar | Institut Pasteur de Dakar | Ndongo Dia et al |
| EPI_ISL_481220 | Institut Pasteur Dakar | Institut Pasteur de Dakar | Ndongo Dia et al |
| EPI_ISL_481234 | Institut Pasteur Dakar | Institut Pasteur de Dakar | Ndongo Dia et al |
| EPI_ISL_498238 | Institut Pasteur de Dakar | Institut Pasteur de Dakar | Ndongo Dia et al |
| EPI_ISL_498239 | Institut Pasteur de Dakar | Institut Pasteur de Dakar | Ndongo Dia et al |
| EPI_ISL_498241 | Institut Pasteur de Dakar | Institut Pasteur de Dakar | Ndongo Dia et al |
| EPI_ISL_420078 | Institut Pasteur Dakar | Institut Pasteur de Dakar | Ndongo Dia et al |
| EPI_ISL_498246 | Institut Pasteur de Dakar | Institut Pasteur de Dakar | Ndongo Dia et al |
| EPI_ISL_476560 | Institut Pasteur Dakar | Institut Pasteur de Dakar | Ndongo Dia et al |
| EPI_ISL_476570 | Institut Pasteur Dakar | Institut Pasteur de Dakar | Ndongo Dia et al |
| EPI_ISL_486869 | Institut Pasteur Dakar | Institut Pasteur de Dakar | Ndongo Dia et al |
| EPI_ISL_498235 | Institut Pasteur de Dakar | Institut Pasteur de Dakar | Ndongo Dia et al |
| EPI_ISL_498251 | Institut Pasteur de Dakar | Institut Pasteur de Dakar | Ndongo Dia et al |
| EPI_ISL_1209667 | Israel Central Virology laboratory | Israel National Consortium for SARS-CoV-2 sequencing | Neta Zuckerman et al |
| EPI_ISL_889051 | Israel Central Virology laboratory | Israel National Consortium for SARS-CoV-2 sequencing | Neta Zuckerman et al |
| EPI_ISL_1209489 | Israel Central Virology laboratory | Israel National Consortium for SARS-CoV-2 sequencing | Neta Zuckerman et al |
| EPI_ISL_1210310 | Israel Central Virology laboratory | Israel National Consortium for SARS-CoV-2 sequencing | Neta Zuckerman et al |
| EPI_ISL_1358495 | Israel Central Virology laboratory | Israel National Consortium for SARS-CoV-2 sequencing | Neta Zuckerman et al |
| EPI_ISL_1209816 | Israel Central Virology laboratory | Israel National Consortium for SARS-CoV-2 sequencing | Neta Zuckerman et al |
| EPI_ISL_804112 | Israel Central Virology laboratory | Israel National Consortium for SARS-CoV-2 sequencing | Neta Zuckerman et al |
| EPI_ISL_1240654 | Israel Central Virology laboratory | Israel National Consortium for SARS-CoV-2 sequencing | Neta Zuckerman et al |
| EPI_ISL_889025 | Israel Central Virology laboratory | Israel National Consortium for SARS-CoV-2 sequencing | Neta Zuckerman et al |
| EPI_ISL_889141 | Israel Central Virology laboratory | Israel National Consortium for SARS-CoV-2 sequencing | Neta Zuckerman et al |
| EPI_ISL_889144 | Israel Central Virology laboratory | Israel National Consortium for SARS-CoV-2 sequencing | Neta Zuckerman et al |
| EPI_ISL_1073156 | Israel Central Virology laboratory | Israel National Consortium for SARS-CoV-2 sequencing | Neta Zuckerman et al |
| EPI_ISL_889270 | Israel Central Virology laboratory | Israel National Consortium for SARS-CoV-2 sequencing | Neta Zuckerman et al |
| EPI_ISL_514308 | Israel Central Virology laboratory | Israel Central Virology laboratory | Neta Zuckerman et al |
| EPI_ISL_1240645 | Israel Central Virology Laboratory | Israel National Consortium for SARS-CoV-2 sequencing | Neta Zuckerman et al |

| | | | |
|---|---|---|---|
| EPI_ISL_1240647 | Israel Central Virology Laboratory | Israel National Consortium for SARS-CoV-2 sequencing | Neta Zuckerman et al |
| EPI_ISL_1240648 | Israel Central Virology Laboratory | Israel National Consortium for SARS-CoV-2 sequencing | Neta Zuckerman et al |
| EPI_ISL_1240649 | Israel Central Virology Laboratory | Israel National Consortium for SARS-CoV-2 sequencing | Neta Zuckerman et al |
| EPI_ISL_1240650 | Israel Central Virology Laboratory | Israel National Consortium for SARS-CoV-2 sequencing | Neta Zuckerman et al |
| EPI_ISL_1240651 | Israel Central Virology laboratory | Israel National Consortium for SARS-CoV-2 sequencing | Neta Zuckerman et al |
| EPI_ISL_516900 | Israel Central Virology laboratory | Israel Central Virology laboratory | Neta Zuckerman et al |
| EPI_ISL_514278 | Israel Central Virology laboratory | Israel Central Virology laboratory | Neta Zuckerman et al |
| EPI_ISL_514282 | Israel Central Virology laboratory | Israel Central Virology laboratory | Neta Zuckerman et al |
| EPI_ISL_514298 | Israel Central Virology laboratory | Israel Central Virology laboratory | Neta Zuckerman et al |
| EPI_ISL_649094 | Israel Central Virology laboratory | Israel Central Virology laboratory | Neta Zuckerman et al |
| EPI_ISL_575332 | Israel Central Virology laboratory | Israel Central Virology laboratory | Neta Zuckerman et al |
| EPI_ISL_575334 | Israel Central Virology laboratory | Israel Central Virology laboratory | Neta Zuckerman et al |
| EPI_ISL_516904 | Israel Central Virology laboratory | Israel Central Virology laboratory | Neta Zuckerman et al |
| EPI_ISL_516915 | Israel Central Virology laboratory | Israel Central Virology laboratory | Neta Zuckerman et al |
| EPI_ISL_483708 | Israel Central Virology laboratory | Israel Central Virology laboratory | Neta Zuckerman et al |
| EPI_ISL_483721 | Israel Central Virology laboratory | Israel Central Virology laboratory | Neta Zuckerman et al |
| EPI_ISL_515082 | Department of Biochemistry, Cell and Molecular Biology | WACCBIP, University of Ghana | Ngoi et al |
| EPI_ISL_515085 | Department of Biochemistry, Cell and Molecular Biology | WACCBIP, University of Ghana | Ngoi et al |
| EPI_ISL_515087 | Department of Biochemistry, Cell and Molecular Biology | WACCBIP, University of Ghana | Ngoi et al |
| EPI_ISL_515096 | Department of Biochemistry, Cell and Molecular Biology | WACCBIP, University of Ghana | Ngoi et al |
| EPI_ISL_515098 | Department of Biochemistry, Cell and Molecular Biology | WACCBIP, University of Ghana | Ngoi et al |
| EPI_ISL_515103 | Department of Biochemistry, Cell and Molecular Biology | WACCBIP, University of Ghana | Ngoi et al |
| EPI_ISL_515106 | Department of Biochemistry, Cell and Molecular Biology | WACCBIP, University of Ghana | Ngoi et al |
| EPI_ISL_884843 | Department of Biochemistry, Cell and Molecular Biology, West African Centre for Cell Biology of Infectious Pathogens (WACCBIP), University of Ghana | Department of Biochemistry, Cell and Molecular Biology, West African Centre for Cell Biology of Infectious Pathogens (WACCBIP), University of Ghana | Ngoi et al |
| EPI_ISL_884849 | Department of Biochemistry, Cell and Molecular Biology, West African Centre for Cell Biology of Infectious Pathogens (WACCBIP), University of Ghana | Department of Biochemistry, Cell and Molecular Biology, West African Centre for Cell Biology of Infectious Pathogens (WACCBIP), University of Ghana | Ngoi et al |
| EPI_ISL_884851 | Department of Biochemistry, Cell and Molecular Biology, West African Centre for Cell Biology of Infectious Pathogens (WACCBIP), University of Ghana | Department of Biochemistry, Cell and Molecular Biology, West African Centre for Cell Biology of Infectious Pathogens (WACCBIP), University of Ghana | Ngoi et al |
| EPI_ISL_481230 | Lab voor klinische biologie | Onderzoeksgroep Virologie | Nick Vereecke et al |
| EPI_ISL_717588 | Lab voor klinische biologie | Onderzoeksgroep Virologie | Nick Vereecke et al |

| | | | |
|---|---|---|---|
| EPI_ISL_1524330 | Biology Department, College of Science, Al Muthanna University and Public Health Laboratory, Al-Muthanna Health Directorate | Department of Virology, Faculty of Medicine, University of Helsinki, Helsinki, Finland generated and submitted to GISAID | Nihad Al-Rashedi et al |
| EPI_ISL_1524344 | Biology Department, College of Science, Al Muthanna University and Public Health Laboratory, Al-Muthanna Health Directorate | Department of Virology, Faculty of Medicine, University of Helsinki, Helsinki, Finland generated and submitted to GISAID | Nihad Al-Rashedi et al |
| EPI_ISL_1524347 | Biology Department, College of Science, Al Muthanna University and Public Health Laboratory, Al-Muthanna Health Directorate | Department of Virology, Faculty of Medicine, University of Helsinki, Helsinki, Finland generated and submitted to GISAID | Nihad Al-Rashedi et al |
| EPI_ISL_1524352 | Biology Department, College of Science, Al Muthanna University and Public Health Laboratory, Al-Muthanna Health Directorate | Department of Virology, Faculty of Medicine, University of Helsinki, Helsinki, Finland generated and submitted to GISAID | Nihad Al-Rashedi et al |
| EPI_ISL_1524356 | Biology Department, College of Science, Al Muthanna University and Public Health Laboratory, Al-Muthanna Health Directorate | Department of Virology, Faculty of Medicine, University of Helsinki, Helsinki, Finland generated and submitted to GISAID | Nihad Al-Rashedi et al |
| EPI_ISL_1524332 | Biology Department, College of Science, Al Muthanna University and Public Health Laboratory, Al-Muthanna Health Directorate | Department of Virology, Faculty of Medicine, University of Helsinki, Helsinki, Finland generated and submitted to GISAID | Nihad Al-Rashedi et al |
| EPI_ISL_1524378 | Biology Department, College of Science, Al Muthanna University and Public Health Laboratory, Al-Muthanna Health Directorate | Department of Virology, Faculty of Medicine, University of Helsinki, Helsinki, Finland generated and submitted to GISAID | Nihad Al-Rashedi et al |
| EPI_ISL_1524381 | Biology Department, College of Science, Al Muthanna University and Public Health Laboratory, Al-Muthanna Health Directorate | Department of Virology, Faculty of Medicine, University of Helsinki, Helsinki, Finland generated and submitted to GISAID | Nihad Al-Rashedi et al |
| EPI_ISL_1524337 | Biology Department, College of Science, Al Muthanna University and Public Health Laboratory, Al-Muthanna Health Directorate | Department of Virology, Faculty of Medicine, University of Helsinki, Helsinki, Finland generated and submitted to GISAID | Nihad Al-Rashedi et al |
| EPI_ISL_977655 | Caribbean Public Health Agency | Carrington Lab, Department of PreClinical Sciences, Building 36, First Floor Biochemistry Unit, Faculty of Medical Sciences, The University of the West Indies | Nikita S. D. Sahadeo et al |
| EPI_ISL_977656 | Caribbean Public Health Agency | Carrington Lab, Department of PreClinical Sciences, Building 36, First Floor Biochemistry Unit, Faculty of Medical Sciences, The University of the West Indies | Nikita S. D. Sahadeo et al |
| EPI_ISL_977658 | Caribbean Public Health Agency | Carrington Lab, Department of PreClinical Sciences, Building 36, First Floor Biochemistry Unit, Faculty of Medical Sciences, The University of the West Indies | Nikita S. D. Sahadeo et al |
| EPI_ISL_977659 | Caribbean Public Health Agency | Carrington Lab, Department of PreClinical Sciences, Building 36, First Floor Biochemistry Unit, Faculty of Medical Sciences, The University of the West Indies | Nikita S. D. Sahadeo et al |
| EPI_ISL_756367 | The Caribbean Public Health Agency | Carrington Lab, Department of PreClinical Sciences, Faculty of Medical Sciences, The University of the West Indies | Nikita S. D. Sahadeo et al |
| EPI_ISL_872193 | Caribbean Public Health Agency | Carrington Lab, Department ofBuilding 36, First Floor Biochemistry Unit, Faculty of Medical Sciences, The University of the West Indies | Nikita S. D. Sahadeo et al |
| EPI_ISL_872194 | Caribbean Public Health Agency | Carrington Lab, Department of PreClinical Sciences, Faculty of Medical Sciences, The University of the West Indies | Nikita S. D. Sahadeo et al |
| EPI_ISL_872195 | Caribbean Public Health Agency | Carrington Lab, Department of PreClinical Sciences, Faculty of Medical Sciences, The University of the West Indies | Nikita S. D. Sahadeo et al |
| EPI_ISL_977583 | Caribbean Public Health Agency | Carrington Lab, Department of PreClinical Sciences | Nikita S. D. Sahadeo et al |
| EPI_ISL_977538 | Caribbean Public Health Agency | Carrington Lab, Department of PreClinical Sciences | Nikita S. D. Sahadeo et al |
| EPI_ISL_1498262 | The Caribbean Public Health Agency | Carrington Lab, Department of PreClinical Sciences, Faculty of Medical Sciences, The University of the West Indies | Nikita S. D. Sahadeo et al |
| EPI_ISL_1498379 | The Caribbean Public Health Agency | Carrington Lab, Department of PreClinical Sciences, Faculty of Medical Sciences, The University of the West Indies | Nikita S. D. Sahadeo et al |

| | | | |
|---|---|---|---|
| EPI_ISL_1499115 | Trinidad Public Health Laboratory | Carrington Lab, Department of PreClinical Sciences, Faculty of Medical Sciences, The University of the West Indies | Nikita S. D. Sahadeo et al |
| EPI_ISL_1490252 | The Caribbean Public Health Agency | Carrington Lab, Department of PreClinical Sciences, Faculty of Medical Sciences, The University of the West Indies | Nikita S. D. Sahadeo et al |
| EPI_ISL_1490228 | The Caribbean Public Health Agency | Carrington Lab, Department of PreClinical Sciences, Faculty of Medical Sciences, The University of the West Indies | Nikita S. D. Sahadeo et al |
| EPI_ISL_1490226 | The Caribbean Public Health Agency | Carrington Lab, Department of PreClinical Sciences, Faculty of Medical Sciences, The University of the West Indies | Nikita S. D. Sahadeo et al |
| EPI_ISL_1591351 | Caribbean Public Health Agency | Carrington Lab, Department of PreClinical Sciences, Faculty of Medical Sciences, The University of the West Indies | Nikita S. D. Sahadeo et al |
| EPI_ISL_1591250 | Caribbean Public Health Agency | Carrington Lab, Department of PreClinical Sciences, Faculty of Medical Sciences, The University of the West Indies | Nikita S. D. Sahadeo et al |
| EPI_ISL_1591265 | Caribbean Public Health Agency | Carrington Lab, Department of PreClinical Sciences, Faculty of Medical Sciences, The University of the West Indies | Nikita S. D. Sahadeo et al |
| EPI_ISL_1490666 | The Caribbean Public Health Agency | Carrington Lab, Department of PreClinical Sciences, Faculty of Medical Sciences, The University of the West Indies | Nikita S. D. Sahadeo et al |
| EPI_ISL_1498095 | The Caribbean Public Health Agency | Carrington Lab, Department of PreClinical Sciences, Faculty of Medical Sciences, The University of the West Indies | Nikita S. D. Sahadeo et al |
| EPI_ISL_1588899 | Caribbean Public Health Agency | Carrington Lab, Department of PreClinical Sciences, Faculty of Medical Sciences, The University of the West Indies | Nikita S. D. Sahadeo et al |
| EPI_ISL_1588904 | Caribbean Public Health Agency | Carrington Lab, Department of PreClinical Sciences, Faculty of Medical Sciences, The University of the West Indies | Nikita S. D. Sahadeo et al |
| EPI_ISL_1589003 | Caribbean Public Health Agency | Carrington Lab, Department of PreClinical Sciences, Faculty of Medical Sciences, The University of the West Indies | Nikita S. D. Sahadeo et al |
| EPI_ISL_1589004 | Caribbean Public Health Agency | Carrington Lab, Department of PreClinical Sciences, Faculty of Medical Sciences, The University of the West Indies | Nikita S. D. Sahadeo et al |
| EPI_ISL_1589115 | Caribbean Public Health Agency | Carrington Lab, Department of PreClinical Sciences, Faculty of Medical Sciences, The University of the West Indies | Nikita S. D. Sahadeo et al |
| EPI_ISL_1591458 | Caribbean Public Health Agency | Carrington Lab, Department of PreClinical Sciences, Faculty of Medical Sciences, The University of the West Indies | Nikita S. D. Sahadeo et al |
| EPI_ISL_1591459 | Caribbean Public Health Agency | Carrington Lab, Department of PreClinical Sciences, Faculty of Medical Sciences, The University of the West Indies | Nikita S. D. Sahadeo et al |
| EPI_ISL_1591718 | Caribbean Public Health Agency | Carrington Lab, Department of PreClinical Sciences, Faculty of Medical Sciences, The University of the West Indies | Nikita S. D. Sahadeo et al |
| EPI_ISL_1591518 | Caribbean Public Health Agency | Carrington Lab, Department of PreClinical Sciences, Faculty of Medical Sciences, The University of the West Indies | Nikita S. D. Sahadeo et al |
| EPI_ISL_1591719 | Caribbean Public Health Agency | Carrington Lab, Department of PreClinical Sciences, Faculty of Medical Sciences, The University of the West Indies | Nikita S. D. Sahadeo et al |
| EPI_ISL_1591720 | Caribbean Public Health Laboratory | Carrington Lab, Department of PreClinical Sciences, Faculty of Medical Sciences, The University of the West Indies | Nikita S. D. Sahadeo et al |
| EPI_ISL_1591721 | Caribbean Public Health Agency | Carrington Lab, Department of PreClinical Sciences, Faculty of Medical Sciences, The University of the West Indies | Nikita S. D. Sahadeo et al |
| EPI_ISL_717694 | Trinidad Public Health Laboratory | Carrington Lab, Department of PreClinical Sciences, Faculty of Medical Sciences, The University of the West Indies | Nikita S. D. Sahadeo et al |

| | | | |
|---|---|---|---|
| EPI_ISL_756362 | Trinidad Public Health Laboratory | Carrington Lab, Department of PreClinical Sciences, Faculty of Medical Sciences, The University of the West Indies | Nikita S. D. Sahadeo et al |
| EPI_ISL_717695 | Trinidad Public Health Laboratory | Carrington Lab, Department of PreClinical Sciences, Faculty of Medical Sciences, The University of the West Indies | Nikita S. D. Sahadeo et al |
| EPI_ISL_717699 | Trinidad Public Health Laboratory | Carrington Lab, Department of PreClinical Sciences, Faculty of Medical Sciences, The University of the West Indies | Nikita S. D. Sahadeo et al |
| EPI_ISL_756311 | The Caribbean Public Health Agency | Carrington Lab, Department of PreClinical Sciences, Faculty of Medical Sciences, The University of the West Indies | Nikita S. D. Sahadeo et al |
| EPI_ISL_756357 | The Caribbean Public Health Agency | Carrington Lab, Department of PreClinical Sciences, Faculty of Medical Sciences, The University of the West Indies | Nikita S. D. Sahadeo et al |
| EPI_ISL_717697 | Trinidad Public Health Laboratory | Carrington Lab, Department of PreClinical Sciences, Faculty of Medical Sciences, The University of the West Indies | Nikita S. D. Sahadeo et al |
| EPI_ISL_756363 | Trinidad Public Health Laboratory | Carrington Lab, Department of PreClinical Sciences, Faculty of Medical Sciences, The University of the West Indies | Nikita S. D. Sahadeo et al |
| EPI_ISL_717692 | Trinidad Public Health Laboratory | Carrington Lab, Department of PreClinical Sciences, Faculty of Medical Sciences, The University of the West Indies | Nikita S. D. Sahadeo et al |
| EPI_ISL_717696 | Trinidad Public Health Laboratory | Carrington Lab, Department of PreClinical Sciences, Faculty of Medical Sciences, The University of the West Indies | Nikita S. D. Sahadeo et al |
| EPI_ISL_717700 | Trinidad Public Health Laboratory | Carrington Lab, Department of PreClinical Sciences, Faculty of Medical Sciences, The University of the West Indies | Nikita S. D. Sahadeo et al |
| EPI_ISL_717698 | Trinidad Public Health Laboratory | Carrington Lab, Department of PreClinical Sciences, Faculty of Medical Sciences, The University of the West Indies | Nikita S. D. Sahadeo et al |
| EPI_ISL_481483 | Department for Virology, Molecular Biology and Genome Research, R. G. Lugar Center for Public Health Research, National Center for Disease Control and Public Health (NCDC) of Georgia. | Department for Virology, Molecular Biology and Genome Research, R. G. Lugar Center for Public Health Research, National Center for Disease Control and Public Health (NCDC) of Georgia. | Nino Berishvili et al |
| EPI_ISL_596509 | Palestinian Ministry of Health | Molecular Genetics Lab | Nouar Qutob et al |
| EPI_ISL_596522 | Palestinian Ministry of Health | Molecular Genetics Lab | Nouar Qutob et al |
| EPI_ISL_596517 | Palestinian Ministry of Health | Molecular Genetics Lab | Nouar Qutob et al |
| EPI_ISL_596519 | Palestinian Ministry of Health | Molecular Genetics Lab | Nouar Qutob et al |
| EPI_ISL_596521 | Palestinian Ministry of Health | Molecular Genetics Lab | Nouar Qutob et al |
| EPI_ISL_596525 | Palestinian Ministry of Health | Molecular Genetics Lab | Nouar Qutob et al |
| EPI_ISL_596530 | Palestinian Ministry of Health | Molecular Genetics Lab | Nouar Qutob et al |
| EPI_ISL_596531 | Palestinian Ministry of Health | Molecular Genetics Lab | Nouar Qutob et al |
| EPI_ISL_596532 | Palestinian Ministry of Health | Molecular Genetics Lab | Nouar Qutob et al |
| EPI_ISL_596534 | Palestinian Ministry of Health | Molecular Genetics Lab | Nouar Qutob et al |
| EPI_ISL_596536 | Palestinian Ministry of Health | Molecular Genetics Lab | Nouar Qutob et al |
| EPI_ISL_596542 | Palestinian Ministry of Health | Molecular Genetics Lab | Nouar Qutob et al |
| EPI_ISL_596548 | Palestinian Ministry of Health | Molecular Genetics Lab | Nouar Qutob et al |

| | | | |
|---|---|---|---|
| EPI_ISL_596550 | Palestinian Ministry of Health | Molecular Genetics Lab | Nouar Qutob et al |
| EPI_ISL_596551 | Palestinian Ministry of Health | Molecular Genetics Lab | Nouar Qutob et al |
| EPI_ISL_596552 | Palestinian Ministry of Health | Molecular Genetics Lab | Nouar Qutob et al |
| EPI_ISL_596553 | Palestinian Ministry of Health | Molecular Genetics Lab | Nouar Qutob et al |
| EPI_ISL_596554 | Palestinian Ministry of Health | Molecular Genetics Lab | Nouar Qutob et al |
| EPI_ISL_596558 | Palestinian Ministry of Health | Molecular Genetics Lab | Nouar Qutob et al |
| EPI_ISL_596560 | Palestinian Ministry of Health | Molecular Genetics Lab | Nouar Qutob et al |
| EPI_ISL_596561 | Palestinian Ministry of Health | Molecular Genetics Lab | Nouar Qutob et al |
| EPI_ISL_596567 | Palestinian Ministry of Health | Molecular Genetics Lab | Nouar Qutob et al |
| EPI_ISL_804815 | DB Diagnosticos do Brasil | Laboratório de Parasitologia Médica - Instituto de Medicina Tropical - Universidade de São Paulo | Nuno Faria et al |
| EPI_ISL_804824 | DB Diagnosticos do Brasil | Laboratório de Parasitologia Médica - Instituto de Medicina Tropical - Universidade de São Paulo | Nuno Faria et al |
| EPI_ISL_804832 | DB Diagnosticos do Brasil | Laboratório de Parasitologia Médica - Instituto de Medicina Tropical - Universidade de São Paulo | Nuno Faria et al |
| EPI_ISL_804835 | DB Diagnosticos do Brasil | Laboratório de Parasitologia Médica - Instituto de Medicina Tropical - Universidade de São Paulo | Nuno Faria et al |
| EPI_ISL_804829 | DB Diagnosticos do Brasil | Laboratório de Parasitologia Médica - Instituto de Medicina Tropical - Universidade de São Paulo | Nuno Faria et al |
| EPI_ISL_804833 | DB Diagnosticos do Brasil | Laboratório de Parasitologia Médica - Instituto de Medicina Tropical - Universidade de São Paulo | Nuno Faria et al |
| EPI_ISL_583897 | Singapore General Hospital | Department of Microbiology | Nurdyana Abdul Rahman et al |
| EPI_ISL_583900 | Singapore General Hospital | Department of Microbiology | Nurdyana Abdul Rahman et al |
| EPI_ISL_482679 | Singapore General Hospital | Department of Microbiology | Nurdyana Abdul Rahman et al |
| EPI_ISL_500570 | Singapore General Hospital | Department of Microbiology | Nurdyana Abdul Rahman et al |
| EPI_ISL_518003 | Singapore General Hospital | Department of Microbiology | Nurdyana Abdul Rahman et al |
| EPI_ISL_1014682 | National Influenza Center, Virology Department | National Influenza Center | NZ Shafiei Jandaghi et al |
| EPI_ISL_862081 | National Influenza Center, Virology Department | National Influenza Center | NZ Shafiei Jandaghi et al |
| EPI_ISL_1014686 | National Influenza Center, Virology Department | National Influenza Center | NZ Shafiei Jandaghi et al |
| EPI_ISL_539259 | Hospital Universitario de La Ribera (Alzira, València) | SeqCOVID-SPAIN consortium/IBV(CSIC) | Olalla Martínez Macias et al |
| EPI_ISL_1093432 | Nigerian Centre for Disease Control (NCDC) | African Centre of Excellence for Genomics of Infectious Diseases (ACEGID), Redeemer’s University | Olawoye et al |
| EPI_ISL_1093450 | Nigerian Centre for Disease Control (NCDC) | African Centre of Excellence for Genomics of Infectious Diseases (ACEGID), Redeemer’s University | Olawoye et al |
| EPI_ISL_1173210 | Nigeria Centre for Disease Control (NCDC) | African Centre for Excellence for Genomics of Infectious Diseases (ACEGID), Redeemer’s University | Olawoye et al |

| | | | |
|---|---|---|---|
| EPI_ISL_1173243 | Nigeria Centre for Disease Control (NCDC) | African Centre for Excellence for Genomics of Infectious Diseases (ACEGID), Redeemer’s University | Olawoye et al |
| EPI_ISL_1235644 | Nigerian Centre for Disease Control (NCDC) | African Centre of Excellence for Genomics of Infectious Diseases (ACEGID), Redeemer’s University | Olawoye et al |
| EPI_ISL_1235666 | Nigerian Centre for Disease Control (NCDC) | African Centre of Excellence for Genomics of Infectious Diseases (ACEGID), Redeemer’s University | Olawoye et al |
| EPI_ISL_428916 | State Research Center of Virology and Biotechnology VECTOR, Department of Collection of Microorganisms | State Research Center of Virology and Biotechnology VECTOR, Department of Collection of Microorganisms | Oleg V. Pyankov et al |
| EPI_ISL_428893 | State Research Center of Virology and Biotechnology VECTOR, Department of Collection of Microorganisms | State Research Center of Virology and Biotechnology VECTOR, Department of Collection of Microorganisms | Oleg V. Pyankov et al |
| EPI_ISL_428883 | State Research Center of Virology and Biotechnology VECTOR, Department of Collection of Microorganisms | State Research Center of Virology and Biotechnology VECTOR, Department of Collection of Microorganisms | Oleg V. Pyankov et al |
| EPI_ISL_845548 | National Public Health Laboratory, Cameroon | African Centre of Excellence for Genomics of Infectious Diseases (ACEGID), Redeemer's University | Oluniyi P.E. et al |
| EPI_ISL_845546 | National Public Health Laboratory, Cameroon | African Centre of Excellence for Genomics of Infectious Diseases (ACEGID), Redeemer's University | Oluniyi P.E. et al |
| EPI_ISL_845549 | National Public Health Laboratory, Cameroon | African Centre of Excellence for Genomics of Infectious Diseases (ACEGID), Redeemer's University | Oluniyi P.E. et al |
| EPI_ISL_845551 | National Public Health Laboratory, Cameroon | African Centre of Excellence for Genomics of Infectious Diseases (ACEGID), Redeemer's University | Oluniyi P.E. et al |
| EPI_ISL_845550 | National Public Health Laboratory, Cameroon | African Centre of Excellence for Genomics of Infectious Diseases (ACEGID), Redeemer's University | Oluniyi P.E. et al |
| EPI_ISL_845553 | National Public Health Laboratory, Cameroon | African Centre of Excellence for Genomics of Infectious Diseases (ACEGID), Redeemer's University | Oluniyi P.E. et al |
| EPI_ISL_845552 | National Public Health Laboratory, Cameroon | African Centre of Excellence for Genomics of Infectious Diseases (ACEGID), Redeemer's University | Oluniyi P.E. et al |
| EPI_ISL_845554 | National Public Health Laboratory, Cameroon | African Centre of Excellence for Genomics of Infectious Diseases (ACEGID), Redeemer's University | Oluniyi P.E. et al |
| EPI_ISL_845564 | National Public Health Laboratory, Cameroon | African Centre of Excellence for Genomics of Infectious Diseases (ACEGID), Redeemer's University | Oluniyi P.E. et al |
| EPI_ISL_845563 | National Public Health Laboratory, Cameroon | African Centre of Excellence for Genomics of Infectious Diseases (ACEGID), Redeemer's University | Oluniyi P.E. et al |
| EPI_ISL_845565 | National Public Health Laboratory, Cameroon | African Centre of Excellence for Genomics of Infectious Diseases (ACEGID), Redeemer's University | Oluniyi P.E. et al |
| EPI_ISL_845562 | National Public Health Laboratory, Cameroon | African Centre of Excellence for Genomics of Infectious Diseases (ACEGID), Redeemer's University | Oluniyi P.E. et al |
| EPI_ISL_845560 | National Public Health Laboratory, Cameroon | African Centre of Excellence for Genomics of Infectious Diseases (ACEGID), Redeemer's University | Oluniyi P.E. et al |
| EPI_ISL_845561 | National Public Health Laboratory, Cameroon | African Centre of Excellence for Genomics of Infectious Diseases (ACEGID), Redeemer's University | Oluniyi P.E. et al |
| EPI_ISL_845558 | National Public Health Laboratory, Cameroon | African Centre of Excellence for Genomics of Infectious Diseases (ACEGID), Redeemer's University | Oluniyi P.E. et al |
| EPI_ISL_729993 | Nigeria Centre for Disease Control (NCDC) | African Centre of Excellence for Genomics of Infectious Diseases (ACEGID), Redeemer's University, Ede, Osun State, Nigeria | Oluniyi P.E. et al |

| | | | |
|---|---|---|---|
| EPI_ISL_729959 | Nigeria Centre for Disease Control (NCDC) | African Centre of Excellence for Genomics of Infectious Diseases (ACEGID), Redeemer's University, Ede, Osun State, Nigeria | Oluniyi P.E. et al |
| EPI_ISL_729995 | Nigeria Centre for Disease Control (NCDC) | African Centre of Excellence for Genomics of Infectious Diseases (ACEGID), Redeemer's University, Ede, Osun State, Nigeria | Oluniyi P.E. et al |
| EPI_ISL_729997 | Nigeria Centre for Disease Control (NCDC) | African Centre of Excellence for Genomics of Infectious Diseases (ACEGID), Redeemer's University, Ede, Osun State, Nigeria | Oluniyi P.E. et al |
| EPI_ISL_729999 | Nigeria Centre for Disease Control (NCDC) | African Centre of Excellence for Genomics of Infectious Diseases (ACEGID), Redeemer's University, Ede, Osun State, Nigeria | Oluniyi P.E. et al |
| EPI_ISL_730000 | Nigeria Centre for Disease Control (NCDC) | African Centre of Excellence for Genomics of Infectious Diseases (ACEGID), Redeemer's University, Ede, Osun State, Nigeria | Oluniyi P.E. et al |
| EPI_ISL_730001 | Nigeria Centre for Disease Control (NCDC) | African Centre of Excellence for Genomics of Infectious Diseases (ACEGID), Redeemer's University, Ede, Osun State, Nigeria | Oluniyi P.E. et al |
| EPI_ISL_730003 | Nigeria Centre for Disease Control (NCDC) | African Centre of Excellence for Genomics of Infectious Diseases (ACEGID), Redeemer's University, Ede, Osun State, Nigeria | Oluniyi P.E. et al |
| EPI_ISL_730004 | Nigeria Centre for Disease Control (NCDC) | African Centre of Excellence for Genomics of Infectious Diseases (ACEGID), Redeemer's University, Ede, Osun State, Nigeria | Oluniyi P.E. et al |
| EPI_ISL_730010 | Nigeria Centre for Disease Control (NCDC) | African Centre of Excellence for Genomics of Infectious Diseases (ACEGID), Redeemer's University, Ede, Osun State, Nigeria | Oluniyi P.E. et al |
| EPI_ISL_730013 | Nigeria Centre for Disease Control (NCDC) | African Centre of Excellence for Genomics of Infectious Diseases (ACEGID), Redeemer's University, Ede, Osun State, Nigeria | Oluniyi P.E. et al |
| EPI_ISL_730014 | Nigeria Centre for Disease Control (NCDC) | African Centre of Excellence for Genomics of Infectious Diseases (ACEGID), Redeemer's University, Ede, Osun State, Nigeria | Oluniyi P.E. et al |
| EPI_ISL_906298 | Nigeria Centre for Disease Control (NCDC) | African Centre of Excellence for Genomics of Infectious Diseases (ACEGID), Redeemer's University | Oluniyi P.E. et al |
| EPI_ISL_941277 | Nigeria Centre for Disease Control (NCDC) | African Centre of Excellence for Genomics of Infectious Diseases (ACEGID), Redeemer's University | Oluniyi P.E. et al |
| EPI_ISL_941280 | Nigeria Centre for Disease Control (NCDC) | African Centre of Excellence for Genomics of Infectious Diseases (ACEGID), Redeemer's University | Oluniyi P.E. et al |
| EPI_ISL_941281 | Nigeria Centre for Disease Control (NCDC) | African Centre of Excellence for Genomics of Infectious Diseases (ACEGID), Redeemer's University | Oluniyi P.E. et al |
| EPI_ISL_941287 | Nigeria Centre for Disease Control (NCDC) | African Centre of Excellence for Genomics of Infectious Diseases (ACEGID), Redeemer's University | Oluniyi P.E. et al |
| EPI_ISL_1242014 | Nigeria Centre for Disease Control (NCDC) | African Centre of Excellence for Genomics of Infectious Diseases (ACEGID), Redeemer's University | Oluniyi P.E. et al |
| EPI_ISL_527875 | Nigeria Centre for Disease Control (NCDC) | African Centre of Excellence for Genomics of Infectious Diseases (ACEGID), Redeemer's University, Ede, Osun State, Nigeria | Oluniyi P.E. et al |
| EPI_ISL_729921 | Nigeria Centre for Disease Control (NCDC) | African Centre of Excellence for Genomics of Infectious Diseases (ACEGID), Redeemer's University, Ede, Osun State, Nigeria | Oluniyi P.E. et al |
| EPI_ISL_729924 | Nigeria Centre for Disease Control (NCDC) | African Centre of Excellence for Genomics of Infectious Diseases (ACEGID), Redeemer's University, Ede, Osun State, Nigeria | Oluniyi P.E. et al |
| EPI_ISL_729931 | Nigeria Centre for Disease Control (NCDC) | African Centre of Excellence for Genomics of Infectious Diseases (ACEGID), Redeemer's University, Ede, Osun State, Nigeria | Oluniyi P.E. et al |
| EPI_ISL_729933 | Nigeria Centre for Disease Control (NCDC) | African Centre of Excellence for Genomics of Infectious Diseases (ACEGID), Redeemer's University, Ede, Osun State, Nigeria | Oluniyi P.E. et al |

| | | | |
|---|---|---|---|
| EPI_ISL_872603 | Nigeria Centre for Disease Control (NCDC) | African Centre of Excellence for Genomics of Infectious Diseases (ACEGID), Redeemer's University | Oluniyi P.E. et al |
| EPI_ISL_872604 | Nigeria Centre for Disease Control (NCDC) | African Centre of Excellence for Genomics of Infectious Diseases (ACEGID), Redeemer's University | Oluniyi P.E. et al |
| EPI_ISL_872606 | Nigeria Centre for Disease Control (NCDC) | African Centre of Excellence for Genomics of Infectious Diseases (ACEGID), Redeemer's University | Oluniyi P.E. et al |
| EPI_ISL_872609 | Nigeria Centre for Disease Control (NCDC) | African Centre of Excellence for Genomics of Infectious Diseases (ACEGID), Redeemer's University | Oluniyi P.E. et al |
| EPI_ISL_872611 | Nigeria Centre for Disease Control (NCDC) | African Centre of Excellence for Genomics of Infectious Diseases (ACEGID), Redeemer's University | Oluniyi P.E. et al |
| EPI_ISL_872622 | Nigeria Centre for Disease Control (NCDC) | African Centre of Excellence for Genomics of Infectious Diseases (ACEGID), Redeemer's University | Oluniyi P.E. et al |
| EPI_ISL_872624 | Nigeria Centre for Disease Control (NCDC) | African Centre of Excellence for Genomics of Infectious Diseases (ACEGID), Redeemer's University | Oluniyi P.E. et al |
| EPI_ISL_730023 | Nigeria Centre for Disease Control (NCDC) | African Centre of Excellence for Genomics of Infectious Diseases (ACEGID), Redeemer's University, Ede, Osun State, Nigeria | Oluniyi P.E. et al |
| EPI_ISL_730047 | Nigeria Centre for Disease Control (NCDC) | African Centre of Excellence for Genomics of Infectious Diseases (ACEGID), Redeemer's University, Ede, Osun State, Nigeria | Oluniyi P.E. et al |
| EPI_ISL_730048 | Nigeria Centre for Disease Control (NCDC) | African Centre of Excellence for Genomics of Infectious Diseases (ACEGID), Redeemer's University, Ede, Osun State, Nigeria | Oluniyi P.E. et al |
| EPI_ISL_729946 | Nigeria Centre for Disease Control (NCDC) | African Centre of Excellence for Genomics of Infectious Diseases (ACEGID), Redeemer's University, Ede, Osun State, Nigeria | Oluniyi P.E. et al |
| EPI_ISL_729947 | Nigeria Centre for Disease Control (NCDC) | African Centre of Excellence for Genomics of Infectious Diseases (ACEGID), Redeemer's University, Ede, Osun State, Nigeria | Oluniyi P.E. et al |
| EPI_ISL_730042 | Nigeria Centre for Disease Control (NCDC) | African Centre of Excellence for Genomics of Infectious Diseases (ACEGID), Redeemer's University, Ede, Osun State, Nigeria | Oluniyi P.E. et al |
| EPI_ISL_413550 | Centre for Human and Zoonotic Virology (CHAZVY), College of Medicine University of Lagos/Lagos University Teaching Hospital (LUTH), part of the Laboratory Network of the Nigeria Centre for Disease Control (NCDC) | African Centre of Excellence for Genomics of Infectious Diseases (ACEGID), Redeemer's University, Ede, Osun State, Nigeria | Oluniyi P.E. et al |
| EPI_ISL_729967 | Nigeria Centre for Disease Control (NCDC) | African Centre of Excellence for Genomics of Infectious Diseases (ACEGID), Redeemer's University, Ede, Osun State, Nigeria | Oluniyi P.E. et al |
| EPI_ISL_729978 | Nigeria Centre for Disease Control (NCDC) | African Centre of Excellence for Genomics of Infectious Diseases (ACEGID), Redeemer's University, Ede, Osun State, Nigeria | Oluniyi P.E. et al |
| EPI_ISL_729979 | Nigeria Centre for Disease Control (NCDC) | African Centre of Excellence for Genomics of Infectious Diseases (ACEGID), Redeemer's University, Ede, Osun State, Nigeria | Oluniyi P.E. et al |
| EPI_ISL_729980 | Nigeria Centre for Disease Control (NCDC) | African Centre of Excellence for Genomics of Infectious Diseases (ACEGID), Redeemer's University, Ede, Osun State, Nigeria | Oluniyi P.E. et al |
| EPI_ISL_729981 | Nigeria Centre for Disease Control (NCDC) | African Centre of Excellence for Genomics of Infectious Diseases (ACEGID), Redeemer's University, Ede, Osun State, Nigeria | Oluniyi P.E. et al |
| EPI_ISL_729982 | Nigeria Centre for Disease Control (NCDC) | African Centre of Excellence for Genomics of Infectious Diseases (ACEGID), Redeemer's University, Ede, Osun State, Nigeria | Oluniyi P.E. et al |
| EPI_ISL_729984 | Nigeria Centre for Disease Control (NCDC) | African Centre of Excellence for Genomics of Infectious Diseases (ACEGID), Redeemer's University, Ede, Osun State, Nigeria | Oluniyi P.E. et al |

| | | | |
|---|---|---|---|
| EPI_ISL_729985 | Nigeria Centre for Disease Control (NCDC) | African Centre of Excellence for Genomics of Infectious Diseases (ACEGID), Redeemer's University, Ede, Osun State, Nigeria | Oluniyi P.E. et al |
| EPI_ISL_455415 | Nigeria Centre for Disease Control (NCDC) | African Centre of Excellence for Genomics of Infectious Diseases (ACEGID), Redeemer's University, Ede, Osun State, Nigeria | Oluniyi P.E. et al |
| EPI_ISL_455418 | Nigeria Centre for Disease Control (NCDC) | African Centre of Excellence for Genomics of Infectious Diseases (ACEGID), Redeemer's University, Ede, Osun State, Nigeria | Oluniyi P.E. et al |
| EPI_ISL_455422 | Nigeria Centre for Disease Control | African Centre of Excellence for Genomics of Infectious Diseases (ACEGID), Redeemer's University, Ede, Osun State, Nigeria | Oluniyi P.E. et al |
| EPI_ISL_455423 | Nigeria Centre for Disease Control (NCDC) | African Centre of Excellence for Genomics of Infectious Diseases (ACEGID), Redeemer's University, Ede, Osun State, Nigeria | Oluniyi P.E. et al |
| EPI_ISL_455427 | Nigeria Centre for Disease Control (NCDC) | African Centre of Excellence for Genomics of Infectious Diseases (ACEGID), Redeemer's University, Ede, Osun State, Nigeria | Oluniyi P.E. et al |
| EPI_ISL_527895 | Nigeria Centre for Disease Control (NCDC) | African Centre of Excellence for Genomics of Infectious Diseases (ACEGID), Redeemer's University, Ede, Osun State, Nigeria | Oluniyi P.E. et al |
| EPI_ISL_527898 | Nigeria Centre for Disease Control (NCDC) | African Centre of Excellence for Genomics of Infectious Diseases (ACEGID), Redeemer's University, Ede, Osun State, Nigeria | Oluniyi P.E. et al |
| EPI_ISL_527899 | Nigeria Centre for Disease Control (NCDC) | African Centre of Excellence for Genomics of Infectious Diseases (ACEGID), Redeemer's University, Ede, Osun State, Nigeria | Oluniyi P.E. et al |
| EPI_ISL_527902 | Nigeria Centre for Disease Control (NCDC) | African Centre of Excellence for Genomics of Infectious Diseases (ACEGID), Redeemer's University, Ede, Osun State, Nigeria | Oluniyi P.E. et al |
| EPI_ISL_872605 | Nigeria Centre for Disease Control (NCDC) | African Centre of Excellence for Genomics of Infectious Diseases (ACEGID), Redeemer's University | Oluniyi P.E. et al |
| EPI_ISL_626228 | Institute for Virology, University Hospital Essen | Center of Medical Microbiology, Virology, and Hospital Hygiene, University of Duesseldorf | Olympia E. Anastasiou et al |
| EPI_ISL_1385804 | Alfa Diagnostica, Republic of Moldova | ONCOGENE LLC | ONCOGENE LLC et al |
| EPI_ISL_1385807 | Alfa Diagnostica LLC | ONCOGENE LLC | ONCOGENE LLC et al |
| EPI_ISL_1385811 | Alfa Diagnostica LLC | ONCOGENE LLC | ONCOGENE LLC et al |
| EPI_ISL_707938 | Pamukkale University Hospital | Pamukkale University Department of Medical Genetics | Onur TOKGUN et al. |
| EPI_ISL_707939 | Pamukkale University Hospital | Pamukkale University Department of Medical Genetics | Onur TOKGUN et al. |
| EPI_ISL_708185 | Pamukkale University Hospital | Pamukkale University Department of Medical Genetics | Onur TOKGUN et al. |
| EPI_ISL_707937 | Pamukkale University Hospital | Pamukkale University Department of Medical Genetics | Onur TOKGUN et al. |
| EPI_ISL_708190 | Pamukkale University Hospital | Pamukkale University Department of Medical Genetics | Onur TOKGUN et al. |
| EPI_ISL_708191 | Pamukkale University Hospital | Pamukkale University Department of Medical Genetics | Onur TOKGUN et al. |
| EPI_ISL_708193 | Pamukkale University Hospital | Pamukkale University Department of Medical Genetics | Onur TOKGUN et al. |
| EPI_ISL_849738 | Special Operations Medical Research Division, Defence Services Medical Research Centre | Special Operations Medical Research Division, Defence Services Medical Research Centre | Oo et al |
| EPI_ISL_849739 | Special Operations Medical Research Division, Defence Services Medical Research Centre | Special Operations Medical Research Division, Defence Services Medical Research Centre | Oo et al |
| EPI_ISL_849740 | Special Operations Medical Research Division, Defence Services Medical Research Centre | Special Operations Medical Research Division, Defence Services Medical Research Centre | Oo et al |

| | | | |
|---|---|---|---|
| EPI_ISL_849741 | Special Operations Medical Research Division, Defence Services Medical Research Centre | Special Operations Medical Research Division, Defence Services Medical Research Centre | Oo et al |
| EPI_ISL_849742 | Special Operations Medical Research Division, Defence Services Medical Research Centre | Special Operations Medical Research Division, Defence Services Medical Research Centre | Oo et al |
| EPI_ISL_849743 | Special Operations Medical Research Division, Defence Services Medical Research Centre | Special Operations Medical Research Division, Defence Services Medical Research Centre | Oo et al |
| EPI_ISL_413488 | Center of Medical Microbiology, Virology, and Hospital Hygiene, University of Duesseldorf | Center of Medical Microbiology, Virology, and Hospital Hygiene, University of Duesseldorf | Ortwin Adams et al |
| EPI_ISL_414506 | Center of Medical Microbiology, Virology, and Hospital Hygiene, University of Duesseldorf | Center of Medical Microbiology, Virology, and Hospital Hygiene, University of Duesseldorf | Ortwin Adams et al |
| EPI_ISL_414508 | Center of Medical Microbiology, Virology, and Hospital Hygiene, University of Duesseldorf | Center of Medical Microbiology, Virology, and Hospital Hygiene, University of Duesseldorf | Ortwin Adams et al |
| EPI_ISL_414574 | Center of Medical Microbiology, Virology, and Hospital Hygiene, University of Duesseldorf | Center of Medical Microbiology, Virology, and Hospital Hygiene, University of Duesseldorf | Ortwin Adams et al |
| EPI_ISL_1250842 | Clin & Gen Lab | Molecular Genetics Laboratory, Instituto de Investigaciones Químicas, Universidad Mayor de San Andrés | Oscar M. Rollano-Peñaloza et al |
| EPI_ISL_1278275 | Clin & Gen Lab | Molecular Genetics Laboratory, Instituto de Investigaciones Químicas, Universidad Mayor de San Andrés | Oscar M. Rollano-Peñaloza et al |
| EPI_ISL_1278276 | Clin & Gen Lab | Molecular Genetics Laboratory, Instituto de Investigaciones Químicas, Universidad Mayor de San Andrés | Oscar M. Rollano-Peñaloza et al |
| EPI_ISL_1363787 | Clin & Gen Lab | Microbiologia Molecular, Instituto SELADIS, Universidad Mayor de San Andrés | Oscar M. Rollano-Peñaloza et al |
| EPI_ISL_1278281 | Laboratorio de Biologia Molecular, SEDES-Potosi | Molecular Genetics Laboratory, Instituto de Investigaciones Químicas, Universidad Mayor de San Andrés | Oscar M. Rollano-Peñaloza et al |
| EPI_ISL_1278282 | Laboratorio de Biologia Molecular, SEDES-Potosi | Molecular Genetics Laboratory, Instituto de Investigaciones Químicas, Universidad Mayor de San Andrés | Oscar M. Rollano-Peñaloza et al |
| EPI_ISL_1278283 | Laboratorio de Biologia Molecular, SEDES-Potosi | Molecular Genetics Laboratory, Instituto de Investigaciones Químicas, Universidad Mayor de San Andrés | Oscar M. Rollano-Peñaloza et al |
| EPI_ISL_1278284 | Laboratorio de Biologia Molecular, SEDES-Potosi | Molecular Genetics Laboratory, Instituto de Investigaciones Químicas, Universidad Mayor de San Andrés | Oscar M. Rollano-Peñaloza et al |
| EPI_ISL_1278277 | Laboratorio de Biologia Molecular, Hospital San Pedro Claver | Molecular Genetics Laboratory, Instituto de Investigaciones Químicas, Universidad Mayor de San Andrés | Oscar M. Rollano-Peñaloza et al |
| EPI_ISL_1278278 | Laboratorio de Biologia Molecular, Hospital San Pedro Claver | Molecular Genetics Laboratory, Instituto de Investigaciones Químicas, Universidad Mayor de San Andrés | Oscar M. Rollano-Peñaloza et al |
| EPI_ISL_476139 | Folkhalsomyndigheten | The Public Health Agency of Sweden | Oskar Karlsson Lindsjo et al |
| EPI_ISL_475544 | Karolinska Universitetslaboratoriet | The Public Health Agency of Sweden | Oskar Karlsson Lindsjo et al |
| EPI_ISL_475545 | Karolinska Universitetslaboratoriet | The Public Health Agency of Sweden | Oskar Karlsson Lindsjo et al |
| EPI_ISL_475546 | Karolinska Universitetslaboratoriet | The Public Health Agency of Sweden | Oskar Karlsson Lindsjo et al |
| EPI_ISL_475547 | Karolinska Universitetslaboratoriet | The Public Health Agency of Sweden | Oskar Karlsson Lindsjo et al |
| EPI_ISL_475557 | Halmstad klinisk mikrobiologi | The Public Health Agency of Sweden | Oskar Karlsson Lindsjo et al |
| EPI_ISL_475511 | Orestadsklinikens VC | The Public Health Agency of Sweden | Oskar Karlsson Lindsjo et al |

| | | | |
|---|---|---|---|
| EPI_ISL_469060 | Narhalsan Sjobo vardcentral | The Public Health Agency of Sweden | Oskar Karlsson Lindsjo et al |
| EPI_ISL_469061 | Narhalsan Sjobo vardcentral | The Public Health Agency of Sweden | Oskar Karlsson Lindsjo et al |
| EPI_ISL_475559 | Karolinska Universitetslaboratoriet | The Public Health Agency of Sweden | Oskar Karlsson Lindsjo et al |
| EPI_ISL_510831 | Halmstad klinisk mikrobiologi | The Public Health Agency of Sweden | Oskar Karlsson Lindsjo et al |
| EPI_ISL_510842 | Klinisk mikrobiologi NAL Trollhattan | The Public Health Agency of Sweden | Oskar Karlsson Lindsjo et al |
| EPI_ISL_510850 | Skovde/Unilabs | The Public Health Agency of Sweden | Oskar Karlsson Lindsjo et al |
| EPI_ISL_509413 | Acibadem Labcell Cellular Therapy Laboratory | Acibadem Mehmet Ali Aydinlar University School of Medicine, Medical Genetics Department | Ozden Hatirnaz Ng et al |
| EPI_ISL_949630 | NHLS Universitas Academic | UFS Virology | PA Bester et al |
| EPI_ISL_912467 | NHLS Universitas Academic | UFS Virology | PA Bester et al |
| EPI_ISL_729906 | Instituto de Medicina Tropical, Universidad Nacional Toribio Rodríguez de Mendoza de Amazonas | Laboratorio de Genómica Microbiana, Universidad Peruana Cayetano Heredia | Pablo Tsukayama et al |
| EPI_ISL_729907 | Instituto de Medicina Tropical, Universidad Nacional Toribio Rodríguez de Mendoza de Amazonas | Laboratorio de Genómica Microbiana, Universidad Peruana Cayetano Heredia | Pablo Tsukayama et al |
| EPI_ISL_729908 | Instituto de Medicina Tropical, Universidad Nacional Toribio Rodríguez de Mendoza de Amazonas | Laboratorio de Genómica Microbiana, Universidad Peruana Cayetano Heredia | Pablo Tsukayama et al |
| EPI_ISL_729909 | Instituto de Medicina Tropical, Universidad Nacional Toribio Rodríguez de Mendoza de Amazonas | Laboratorio de Genómica Microbiana, Universidad Peruana Cayetano Heredia | Pablo Tsukayama et al |
| EPI_ISL_729912 | Instituto de Medicina Tropical, Universidad Nacional Toribio Rodríguez de Mendoza de Amazonas | Laboratorio de Genómica Microbiana, Universidad Peruana Cayetano Heredia | Pablo Tsukayama et al |
| EPI_ISL_729915 | Instituto de Medicina Tropical, Universidad Nacional Toribio Rodríguez de Mendoza de Amazonas | Laboratorio de Genómica Microbiana, Universidad Peruana Cayetano Heredia | Pablo Tsukayama et al |
| EPI_ISL_729916 | Instituto de Medicina Tropical, Universidad Nacional Toribio Rodríguez de Mendoza de Amazonas | Laboratorio de Genómica Microbiana, Universidad Peruana Cayetano Heredia | Pablo Tsukayama et al |
| EPI_ISL_729871 | Instituto de Medicina Tropical, Universidad Nacional Toribio Rodríguez de Mendoza de Amazonas | Laboratorio de Genómica Microbiana, Universidad Peruana Cayetano Heredia | Pablo Tsukayama et al |
| EPI_ISL_529067 | Laboratorio de Referencia Nacional de Virus Respiratorios, Instituto Nacional de Salud Peru | Laboratorio de Genómica Microbiana, Universidad Peruana Cayetano Heredia | Pablo Tsukayama et al |
| EPI_ISL_540926 | Laboratorio de Referencia Nacional de Virus Respiratorios, Instituto Nacional de Salud Peru | Laboratorio de Genómica Microbiana, Universidad Peruana Cayetano Heredia | Pablo Tsukayama et al |
| EPI_ISL_540934 | Laboratorio de Referencia Nacional de Virus Respiratorios, Instituto Nacional de Salud Peru | Laboratorio de Genómica Microbiana, Universidad Peruana Cayetano Heredia | Pablo Tsukayama et al |
| EPI_ISL_529075 | Laboratorio de Referencia Nacional de Virus Respiratorios, Instituto Nacional de Salud Peru | Laboratorio de Genómica Microbiana, Universidad Peruana Cayetano Heredia | Pablo Tsukayama et al |
| EPI_ISL_540945 | Laboratorio de Referencia Nacional de Virus Respiratorios, Instituto Nacional de Salud Peru | Laboratorio de Genómica Microbiana, Universidad Peruana Cayetano Heredia | Pablo Tsukayama et al |
| EPI_ISL_540952 | Laboratorio de Referencia Nacional de Virus Respiratorios, Instituto Nacional de Salud Peru | Laboratorio de Genómica Microbiana, Universidad Peruana Cayetano Heredia | Pablo Tsukayama et al |
| EPI_ISL_540959 | Laboratorio de Referencia Nacional de Virus Respiratorios, Instituto Nacional de Salud Peru | Laboratorio de Genómica Microbiana, Universidad Peruana Cayetano Heredia | Pablo Tsukayama et al |

| | | | |
|---|---|---|---|
| EPI_ISL_540992 | Laboratorio de Referencia Nacional de Virus Respiratorios, Instituto Nacional de Salud Peru | Laboratorio de Genómica Microbiana, Universidad Peruana Cayetano Heredia | Pablo Tsukayama et al |
| EPI_ISL_568517 | Laboratorio de Referencia Nacional de Virus Respiratorios, Instituto Nacional de Salud Peru | Laboratorio de Genómica Microbiana, Universidad Peruana Cayetano Heredia | Pablo Tsukayama et al |
| EPI_ISL_568527 | Laboratorio de Referencia Nacional de Virus Respiratorios, Instituto Nacional de Salud Peru | Laboratorio de Genómica Microbiana, Universidad Peruana Cayetano Heredia | Pablo Tsukayama et al |
| EPI_ISL_568536 | Laboratorio de Referencia Nacional de Virus Respiratorios, Instituto Nacional de Salud Peru | Laboratorio de Genómica Microbiana, Universidad Peruana Cayetano Heredia | Pablo Tsukayama et al |
| EPI_ISL_729872 | Laboratorio de Referencia Nacional de Virus Respiratorios, Instituto Nacional de Salud Peru | Laboratorio de Genómica Microbiana, Universidad Peruana Cayetano Heredia | Pablo Tsukayama et al |
| EPI_ISL_729875 | Laboratorio de Referencia Nacional de Virus Respiratorios, Instituto Nacional de Salud Peru | Laboratorio de Genómica Microbiana, Universidad Peruana Cayetano Heredia | Pablo Tsukayama et al |
| EPI_ISL_729876 | Laboratorio de Referencia Nacional de Virus Respiratorios, Instituto Nacional de Salud Peru | Laboratorio de Genómica Microbiana, Universidad Peruana Cayetano Heredia | Pablo Tsukayama et al |
| EPI_ISL_729896 | Laboratorio de Referencia Nacional de Virus Respiratorios, Instituto Nacional de Salud Peru | Laboratorio de Genómica Microbiana, Universidad Peruana Cayetano Heredia | Pablo Tsukayama et al |
| EPI_ISL_729897 | Laboratorio de Referencia Nacional de Virus Respiratorios, Instituto Nacional de Salud Peru | Laboratorio de Genómica Microbiana, Universidad Peruana Cayetano Heredia | Pablo Tsukayama et al |
| EPI_ISL_862476 | Kurnool Medical College (KMC) | CSIR Institute of Genomics and Integrative Biology | Pallavali Roja Rani et al |
| EPI_ISL_862479 | Kurnool Medical College (KMC) | CSIR Institute of Genomics and Integrative Biology | Pallavali Roja Rani et al |
| EPI_ISL_1424519 | Queensland Medical Laboratories | Victorian Infectious Diseases Reference Laboratory (VIDRL) and the Melbourne Diagnostic Unit Public Health Laboratory (MDU-PHL) | Palou et al |
| EPI_ISL_1424521 | Queensland Medical Laboratories | Victorian Infectious Diseases Reference Laboratory (VIDRL) and the Melbourne Diagnostic Unit Public Health Laboratory (MDU-PHL) | Palou et al |
| EPI_ISL_1424523 | Queensland Medical Laboratories | Victorian Infectious Diseases Reference Laboratory (VIDRL) and the Melbourne Diagnostic Unit Public Health Laboratory (MDU-PHL) | Palou et al |
| EPI_ISL_1424526 | Queensland Medical Laboratories | Victorian Infectious Diseases Reference Laboratory (VIDRL) and the Melbourne Diagnostic Unit Public Health Laboratory (MDU-PHL) | Palou et al |
| EPI_ISL_1424540 | Queensland Medical Laboratories | Victorian Infectious Diseases Reference Laboratory (VIDRL) and the Melbourne Diagnostic Unit Public Health Laboratory (MDU-PHL) | Palou et al |
| EPI_ISL_1424544 | Queensland Medical Laboratories | Victorian Infectious Diseases Reference Laboratory (VIDRL) and the Melbourne Diagnostic Unit Public Health Laboratory (MDU-PHL) | Palou et al |
| EPI_ISL_1424584 | Queensland Medical Laboratories | Victorian Infectious Diseases Reference Laboratory (VIDRL) and the Melbourne Diagnostic Unit Public Health Laboratory (MDU-PHL) | Palou et al |
| EPI_ISL_1424591 | Queensland Medical Laboratories | Victorian Infectious Diseases Reference Laboratory (VIDRL) and the Melbourne Diagnostic Unit Public Health Laboratory (MDU-PHL) | Palou et al |
| EPI_ISL_1424592 | Queensland Medical Laboratories | Victorian Infectious Diseases Reference Laboratory (VIDRL) and the Melbourne Diagnostic Unit Public Health Laboratory (MDU-PHL) | Palou et al |
| EPI_ISL_1424615 | Queensland Medical Laboratories | Victorian Infectious Diseases Reference Laboratory (VIDRL) and the Melbourne Diagnostic Unit Public Health Laboratory (MDU-PHL) | Palou et al |
| EPI_ISL_1424616 | Queensland Medical Laboratories | Victorian Infectious Diseases Reference Laboratory (VIDRL) and the Melbourne Diagnostic Unit Public Health Laboratory (MDU-PHL) | Palou et al |

| | | | |
|---|---|---|---|
| EPI_ISL_1424618 | Queensland Medical Laboratories | Victorian Infectious Diseases Reference Laboratory (VIDRL) and the Melbourne Diagnostic Unit Public Health Laboratory (MDU-PHL) | Palou et al |
| EPI_ISL_1424625 | Queensland Medical Laboratories | Victorian Infectious Diseases Reference Laboratory (VIDRL) and the Melbourne Diagnostic Unit Public Health Laboratory (MDU-PHL) | Palou et al |
| EPI_ISL_1424635 | Queensland Medical Laboratories | Victorian Infectious Diseases Reference Laboratory (VIDRL) and the Melbourne Diagnostic Unit Public Health Laboratory (MDU-PHL) | Palou et al |
| EPI_ISL_1424680 | Queensland Medical Laboratories | Melbourne Diagnostic Unit Public Health Laboratory (MDU-PHL) | Palou et al |
| EPI_ISL_1296263 | Azienda Ospedaliera Terni | Istituto Zooprofilattico Sperimentale dell'Abruzzo e Molise "G. Caporale" | Palumbo M et al |
| EPI_ISL_1299525 | Azienda Ospedaliera Terni | Istituto Zooprofilattico Sperimentale dell'Abruzzo e Molise "G. Caporale" | Palumbo m et al |
| EPI_ISL_1415419 | Reference Laboratory of the Ministry of Health | Laboratory of Respiratory Viruses and Measles, Oswaldo Cruz Institute, FIOCRUZ | Paola Resende et al |
| EPI_ISL_1415418 | Reference Laboratory of the Ministry of Health | Laboratory of Respiratory Viruses and Measles, Oswaldo Cruz Institute, FIOCRUZ | Paola Resende et al |
| EPI_ISL_1415417 | Reference Laboratory of the Ministry of Health | Laboratory of Respiratory Viruses and Measles, Oswaldo Cruz Institute, FIOCRUZ | Paola Resende et al |
| EPI_ISL_1415416 | Reference Laboratory of the Ministry of Health | Laboratory of Respiratory Viruses and Measles, Oswaldo Cruz Institute, FIOCRUZ | Paola Resende et al |
| EPI_ISL_1415415 | Reference Laboratory of the Ministry of Health | Laboratory of Respiratory Viruses and Measles, Oswaldo Cruz Institute, FIOCRUZ | Paola Resende et al |
| EPI_ISL_1415414 | Reference Laboratory of the Ministry of Health | Laboratory of Respiratory Viruses and Measles, Oswaldo Cruz Institute, FIOCRUZ | Paola Resende et al |
| EPI_ISL_1415413 | Reference Laboratory of the Ministry of Health | Laboratory of Respiratory Viruses and Measles, Oswaldo Cruz Institute, FIOCRUZ | Paola Resende et al |
| EPI_ISL_1415412 | Reference Laboratory of the Ministry of Health | Laboratory of Respiratory Viruses and Measles, Oswaldo Cruz Institute, FIOCRUZ | Paola Resende et al |
| EPI_ISL_1415411 | Reference Laboratory of the Ministry of Health | Laboratory of Respiratory Viruses and Measles, Oswaldo Cruz Institute, FIOCRUZ | Paola Resende et al |
| EPI_ISL_1415410 | Reference Laboratory of the Ministry of Health | Laboratory of Respiratory Viruses and Measles, Oswaldo Cruz Institute, FIOCRUZ | Paola Resende et al |
| EPI_ISL_1415409 | Reference Laboratory of the Ministry of Health | Laboratory of Respiratory Viruses and Measles, Oswaldo Cruz Institute, FIOCRUZ | Paola Resende et al |
| EPI_ISL_1415408 | Reference Laboratory of the Ministry of Health | Laboratory of Respiratory Viruses and Measles, Oswaldo Cruz Institute, FIOCRUZ | Paola Resende et al |
| EPI_ISL_1415407 | Reference Laboratory of the Ministry of Health | Laboratory of Respiratory Viruses and Measles, Oswaldo Cruz Institute, FIOCRUZ | Paola Resende et al |
| EPI_ISL_1415406 | Reference Laboratory of the Ministry of Health | Laboratory of Respiratory Viruses and Measles, Oswaldo Cruz Institute, FIOCRUZ | Paola Resende et al |
| EPI_ISL_1415404 | Reference Laboratory of the Ministry of Health | Laboratory of Respiratory Viruses and Measles, Oswaldo Cruz Institute, FIOCRUZ | Paola Resende et al |
| EPI_ISL_1415403 | Reference Laboratory of the Ministry of Health | Laboratory of Respiratory Viruses and Measles, Oswaldo Cruz Institute, FIOCRUZ | Paola Resende et al |
| EPI_ISL_1415399 | Reference Laboratory of the Ministry of Health | Laboratory of Respiratory Viruses and Measles, Oswaldo Cruz Institute, FIOCRUZ | Paola Resende et al |

| | | | |
|---|---|---|---|
| EPI_ISL_1415392 | Reference Laboratory of the Ministry of Health | Laboratory of Respiratory Viruses and Measles, Oswaldo Cruz Institute, FIOCRUZ | Paola Resende et al |
| EPI_ISL_837555 | Centro Nacional de Enfermedades Tropicales (CENETROP) | Laboratory of Respiratory Viruses and Measles, Oswaldo Cruz Institute, FIOCRUZ | Paola Resende et al |
| EPI_ISL_837556 | Centro Nacional de Enfermedades Tropicales (CENETROP) | Laboratory of Respiratory Viruses and Measles, Oswaldo Cruz Institute, FIOCRUZ | Paola Resende et al |
| EPI_ISL_837557 | Centro Nacional de Enfermedades Tropicales (CENETROP) | Laboratory of Respiratory Viruses and Measles, Oswaldo Cruz Institute, FIOCRUZ | Paola Resende et al |
| EPI_ISL_837558 | Centro Nacional de Enfermedades Tropicales (CENETROP) | Laboratory of Respiratory Viruses and Measles, Oswaldo Cruz Institute, FIOCRUZ | Paola Resende et al |
| EPI_ISL_837554 | Centro Nacional de Enfermedades Tropicales (CENETROP) | Laboratory of Respiratory Viruses and Measles, Oswaldo Cruz Institute, FIOCRUZ | Paola Resende et al |
| EPI_ISL_837559 | Centro Nacional de Enfermedades Tropicales (CENETROP) | Laboratory of Respiratory Viruses and Measles, Oswaldo Cruz Institute, FIOCRUZ | Paola Resende et al |
| EPI_ISL_837560 | Centro Nacional de Enfermedades Tropicales (CENETROP) | Laboratory of Respiratory Viruses and Measles, Oswaldo Cruz Institute, FIOCRUZ | Paola Resende et al |
| EPI_ISL_837561 | Centro Nacional de Enfermedades Tropicales (CENETROP) | Laboratory of Respiratory Viruses and Measles, Oswaldo Cruz Institute, FIOCRUZ | Paola Resende et al |
| EPI_ISL_837562 | Centro Nacional de Enfermedades Tropicales (CENETROP) | Laboratory of Respiratory Viruses and Measles, Oswaldo Cruz Institute, FIOCRUZ | Paola Resende et al |
| EPI_ISL_837563 | Centro Nacional de Enfermedades Tropicales (CENETROP) | Laboratory of Respiratory Viruses and Measles, Oswaldo Cruz Institute, FIOCRUZ | Paola Resende et al |
| EPI_ISL_837564 | Centro Nacional de Enfermedades Tropicales (CENETROP) | Laboratory of Respiratory Viruses and Measles, Oswaldo Cruz Institute, FIOCRUZ | Paola Resende et al |
| EPI_ISL_837565 | Centro Nacional de Enfermedades Tropicales (CENETROP) | Laboratory of Respiratory Viruses and Measles, Oswaldo Cruz Institute, FIOCRUZ | Paola Resende et al |
| EPI_ISL_837553 | Centro Nacional de Enfermedades Tropicales (CENETROP) | Laboratory of Respiratory Viruses and Measles, Oswaldo Cruz Institute, FIOCRUZ | Paola Resende et al |
| EPI_ISL_837568 | Centro Nacional de Enfermedades Tropicales (CENETROP) | Laboratory of Respiratory Viruses and Measles, Oswaldo Cruz Institute, FIOCRUZ | Paola Resende et al |
| EPI_ISL_837569 | Centro Nacional de Enfermedades Tropicales (CENETROP) | Laboratory of Respiratory Viruses and Measles, Oswaldo Cruz Institute, FIOCRUZ | Paola Resende et al |
| EPI_ISL_837570 | Centro Nacional de Enfermedades Tropicales (CENETROP) | Laboratory of Respiratory Viruses and Measles, Oswaldo Cruz Institute, FIOCRUZ | Paola Resende et al |
| EPI_ISL_837571 | Centro Nacional de Enfermedades Tropicales (CENETROP) | Laboratory of Respiratory Viruses and Measles, Oswaldo Cruz Institute, FIOCRUZ | Paola Resende et al |
| EPI_ISL_837550 | Centro Nacional de Enfermedades Tropicales (CENETROP) | Laboratory of Respiratory Viruses and Measles, Oswaldo Cruz Institute, FIOCRUZ | Paola Resende et al |
| EPI_ISL_837572 | Centro Nacional de Enfermedades Tropicales (CENETROP) | Laboratory of Respiratory Viruses and Measles, Oswaldo Cruz Institute, FIOCRUZ | Paola Resende et al |
| EPI_ISL_837573 | Centro Nacional de Enfermedades Tropicales (CENETROP) | Laboratory of Respiratory Viruses and Measles, Oswaldo Cruz Institute, FIOCRUZ | Paola Resende et al |
| EPI_ISL_837575 | Centro Nacional de Enfermedades Tropicales (CENETROP) | Laboratory of Respiratory Viruses and Measles, Oswaldo Cruz Institute, FIOCRUZ | Paola Resende et al |

| | | | |
|---|---|---|---|
| EPI_ISL_837576 | Centro Nacional de Enfermedades Tropicales (CENETROP) | Laboratory of Respiratory Viruses and Measles, Oswaldo Cruz Institute, FIOCRUZ | Paola Resende et al |
| EPI_ISL_837577 | Centro Nacional de Enfermedades Tropicales (CENETROP) | Laboratory of Respiratory Viruses and Measles, Oswaldo Cruz Institute, FIOCRUZ | Paola Resende et al |
| EPI_ISL_837578 | Centro Nacional de Enfermedades Tropicales (CENETROP) | Laboratory of Respiratory Viruses and Measles, Oswaldo Cruz Institute, FIOCRUZ | Paola Resende et al |
| EPI_ISL_792639 | Laboratório Central de Saúde Pública do Estado de Alagoas (LACEN-AL) | Laboratory of Respiratory Viruses and Measles, Oswaldo Cruz Institute, FIOCRUZ | Paola Resende et al |
| EPI_ISL_792640 | Laboratório Central de Saúde Pública do Estado de Alagoas (LACEN-AL) | Laboratory of Respiratory Viruses and Measles, Oswaldo Cruz Institute, FIOCRUZ | Paola Resende et al |
| EPI_ISL_792641 | Laboratório Central de Saúde Pública do Estado de Alagoas (LACEN-AL) | Laboratory of Respiratory Viruses and Measles, Oswaldo Cruz Institute, FIOCRUZ | Paola Resende et al |
| EPI_ISL_792642 | Laboratório Central de Saúde Pública do Estado de Alagoas (LACEN-AL) | Laboratory of Respiratory Viruses and Measles, Oswaldo Cruz Institute, FIOCRUZ | Paola Resende et al |
| EPI_ISL_792643 | Laboratório Central de Saúde Pública do Estado de Alagoas (LACEN-AL) | Laboratory of Respiratory Viruses and Measles, Oswaldo Cruz Institute, FIOCRUZ | Paola Resende et al |
| EPI_ISL_792644 | Laboratório Central de Saúde Pública do Estado de Alagoas (LACEN-AL) | Laboratory of Respiratory Viruses and Measles, Oswaldo Cruz Institute, FIOCRUZ | Paola Resende et al |
| EPI_ISL_1181592 | Laboratorio Central de Saude Publica do Estado do Alagoas (LACEN-AL) | Laboratory of Respiratory Viruses and Measles, Oswaldo Cruz Institute, FIOCRUZ | Paola Resende et al |
| EPI_ISL_1181396 | Laboratorio Central de Saude Publica do Estado do Alagoas (LACEN-AL) | Laboratory of Respiratory Viruses and Measles, Oswaldo Cruz Institute, FIOCRUZ | Paola Resende et al |
| EPI_ISL_1181365 | Laboratorio Central de Saude Publica do Estado do Alagoas (LACEN-AL) | Laboratory of Respiratory Viruses and Measles, Oswaldo Cruz Institute, FIOCRUZ | Paola Resende et al |
| EPI_ISL_1219135 | Laboratorio Central de Saude Publica do Estado do Alagoas (LACEN-AL) | Laboratory of Respiratory Viruses and Measles, Oswaldo Cruz Institute, FIOCRUZ | Paola Resende et al |
| EPI_ISL_1219134 | Laboratorio Central de Saude Publica do Estado do Alagoas (LACEN-AL) | Laboratory of Respiratory Viruses and Measles, Oswaldo Cruz Institute, FIOCRUZ | Paola Resende et al |
| EPI_ISL_427292 | Laboratório Central de Saúde Pública do Estado de Alagoas (LACEN-AL) | Laboratory of Respiratory Viruses and Measles, Oswaldo Cruz Institute, FIOCRUZ | Paola Resende et al |
| EPI_ISL_1181354 | Gonçalo Moniz Institute, FIOCRUZ, Bahia | Laboratory of Respiratory Viruses and Measles, Oswaldo Cruz Institute, FIOCRUZ | Paola Resende et al |
| EPI_ISL_1181484 | Gonçalo Moniz Institute, FIOCRUZ, Bahia | Laboratory of Respiratory Viruses and Measles, Oswaldo Cruz Institute, FIOCRUZ | Paola Resende et al |
| EPI_ISL_1181485 | Gonçalo Moniz Institute, FIOCRUZ, Bahia | Laboratory of Respiratory Viruses and Measles, Oswaldo Cruz Institute, FIOCRUZ | Paola Resende et al |
| EPI_ISL_1181435 | Federal University of Southern Bahia (UFSB - Universidade Federal do Sul da Bahia) | Laboratory of Respiratory Viruses and Measles, Oswaldo Cruz Institute, FIOCRUZ | Paola Resende et al |
| EPI_ISL_1181436 | Federal University of Southern Bahia (UFSB - Universidade Federal do Sul da Bahia) | Laboratory of Respiratory Viruses and Measles, Oswaldo Cruz Institute, FIOCRUZ | Paola Resende et al |
| EPI_ISL_1181364 | Federal University of Southern Bahia (UFSB - Universidade Federal do Sul da Bahia) | Laboratory of Respiratory Viruses and Measles, Oswaldo Cruz Institute, FIOCRUZ | Paola Resende et al |
| EPI_ISL_1181443 | Federal University of Southern Bahia (UFSB - Universidade Federal do Sul da Bahia) | Laboratory of Respiratory Viruses and Measles, Oswaldo Cruz Institute, FIOCRUZ | Paola Resende et al |

| | | | |
|---|---|---|---|
| EPI_ISL_1181450 | Federal University of Southern Bahia (UFSB - Universidade Federal do Sul da Bahia) | Laboratory of Respiratory Viruses and Measles, Oswaldo Cruz Institute, FIOCRUZ | Paola Resende et al |
| EPI_ISL_1181449 | Gonçalo Moniz Institute, FIOCRUZ, Bahia | Laboratory of Respiratory Viruses and Measles, Oswaldo Cruz Institute, FIOCRUZ | Paola Resende et al |
| EPI_ISL_1181444 | Gonçalo Moniz Institute, FIOCRUZ, Bahia | Laboratory of Respiratory Viruses and Measles, Oswaldo Cruz Institute, FIOCRUZ | Paola Resende et al |
| EPI_ISL_1181445 | Gonçalo Moniz Institute, FIOCRUZ, Bahia | Laboratory of Respiratory Viruses and Measles, Oswaldo Cruz Institute, FIOCRUZ | Paola Resende et al |
| EPI_ISL_1181441 | Gonçalo Moniz Institute, FIOCRUZ, Bahia | Laboratory of Respiratory Viruses and Measles, Oswaldo Cruz Institute, FIOCRUZ | Paola Resende et al |
| EPI_ISL_1181440 | Gonçalo Moniz Institute, FIOCRUZ, Bahia | Laboratory of Respiratory Viruses and Measles, Oswaldo Cruz Institute, FIOCRUZ | Paola Resende et al |
| EPI_ISL_1181355 | Gonçalo Moniz Institute, FIOCRUZ, Bahia | Laboratory of Respiratory Viruses and Measles, Oswaldo Cruz Institute, FIOCRUZ | Paola Resende et al |
| EPI_ISL_1181429 | Gonçalo Moniz Institute, FIOCRUZ, Bahia | Laboratory of Respiratory Viruses and Measles, Oswaldo Cruz Institute, FIOCRUZ | Paola Resende et al |
| EPI_ISL_1181379 | Gonçalo Moniz Institute, FIOCRUZ, Bahia | Laboratory of Respiratory Viruses and Measles, Oswaldo Cruz Institute, FIOCRUZ | Paola Resende et al |
| EPI_ISL_1181424 | Gonçalo Moniz Institute, FIOCRUZ, Bahia | Laboratory of Respiratory Viruses and Measles, Oswaldo Cruz Institute, FIOCRUZ | Paola Resende et al |
| EPI_ISL_1181419 | Gonçalo Moniz Institute, FIOCRUZ, Bahia | Laboratory of Respiratory Viruses and Measles, Oswaldo Cruz Institute, FIOCRUZ | Paola Resende et al |
| EPI_ISL_1181408 | Gonçalo Moniz Institute, FIOCRUZ, Bahia | Laboratory of Respiratory Viruses and Measles, Oswaldo Cruz Institute, FIOCRUZ | Paola Resende et al |
| EPI_ISL_1219136 | Gonçalo Moniz Institute, FIOCRUZ, Bahia | Laboratory of Respiratory Viruses and Measles, Oswaldo Cruz Institute, FIOCRUZ | Paola Resende et al |
| EPI_ISL_1181356 | Oswaldo Cruz Foundation, FIOCRUZ - Ceara (Fiocruz-CE) | Laboratory of Respiratory Viruses and Measles, Oswaldo Cruz Institute, FIOCRUZ | Paola Resende et al |
| EPI_ISL_1181520 | Oswaldo Cruz Foundation, FIOCRUZ - Ceara (Fiocruz-CE) | Laboratory of Respiratory Viruses and Measles, Oswaldo Cruz Institute, FIOCRUZ | Paola Resende et al |
| EPI_ISL_1181522 | Oswaldo Cruz Foundation, FIOCRUZ - Ceara (Fiocruz-CE) | Laboratory of Respiratory Viruses and Measles, Oswaldo Cruz Institute, FIOCRUZ | Paola Resende et al |
| EPI_ISL_1181575 | Oswaldo Cruz Foundation, FIOCRUZ - Ceara (Fiocruz-CE) | Laboratory of Respiratory Viruses and Measles, Oswaldo Cruz Institute, FIOCRUZ | Paola Resende et al |
| EPI_ISL_1181577 | Oswaldo Cruz Foundation, FIOCRUZ - Ceara (Fiocruz-CE) | Laboratory of Respiratory Viruses and Measles, Oswaldo Cruz Institute, FIOCRUZ | Paola Resende et al |
| EPI_ISL_1181586 | Oswaldo Cruz Foundation, FIOCRUZ - Ceara (Fiocruz-CE) | Laboratory of Respiratory Viruses and Measles, Oswaldo Cruz Institute, FIOCRUZ | Paola Resende et al |
| EPI_ISL_1181588 | Oswaldo Cruz Foundation, FIOCRUZ - Ceara (Fiocruz-CE) | Laboratory of Respiratory Viruses and Measles, Oswaldo Cruz Institute, FIOCRUZ | Paola Resende et al |
| EPI_ISL_1181591 | Oswaldo Cruz Foundation, FIOCRUZ - Ceara (Fiocruz-CE) | Laboratory of Respiratory Viruses and Measles, Oswaldo Cruz Institute, FIOCRUZ | Paola Resende et al |
| EPI_ISL_1181593 | Oswaldo Cruz Foundation, FIOCRUZ - Ceara (Fiocruz-CE) | Laboratory of Respiratory Viruses and Measles, Oswaldo Cruz Institute, FIOCRUZ | Paola Resende et al |

| | | | |
|---|---|---|---|
| EPI_ISL_1181596 | Oswaldo Cruz Foundation, FIOCRUZ - Ceara (Fiocruz-CE) | Laboratory of Respiratory Viruses and Measles, Oswaldo Cruz Institute, FIOCRUZ | Paola Resende et al |
| EPI_ISL_1181598 | Oswaldo Cruz Foundation, FIOCRUZ - Ceara (Fiocruz-CE) | Laboratory of Respiratory Viruses and Measles, Oswaldo Cruz Institute, FIOCRUZ | Paola Resende et al |
| EPI_ISL_1181604 | Oswaldo Cruz Foundation, FIOCRUZ - Ceara (Fiocruz-CE) | Laboratory of Respiratory Viruses and Measles, Oswaldo Cruz Institute, FIOCRUZ | Paola Resende et al |
| EPI_ISL_1181621 | Oswaldo Cruz Foundation, FIOCRUZ - Ceara (Fiocruz-CE) | Laboratory of Respiratory Viruses and Measles, Oswaldo Cruz Institute, FIOCRUZ | Paola Resende et al |
| EPI_ISL_1181622 | Oswaldo Cruz Foundation, FIOCRUZ - Ceara (Fiocruz-CE) | Laboratory of Respiratory Viruses and Measles, Oswaldo Cruz Institute, FIOCRUZ | Paola Resende et al |
| EPI_ISL_427294 | Laboratory of Respiratory Viruses and Measles, Oswaldo Cruz Institute, FIOCRUZ | Laboratory of Respiratory Viruses and Measles, Oswaldo Cruz Institute, FIOCRUZ | Paola Resende et al |
| EPI_ISL_427295 | Laboratory of Respiratory Viruses and Measles, Oswaldo Cruz Institute, FIOCRUZ | Laboratory of Respiratory Viruses and Measles, Oswaldo Cruz Institute, FIOCRUZ | Paola Resende et al |
| EPI_ISL_427296 | Laboratory of Respiratory Viruses and Measles, Oswaldo Cruz Institute, FIOCRUZ | Laboratory of Respiratory Viruses and Measles, Oswaldo Cruz Institute, FIOCRUZ | Paola Resende et al |
| EPI_ISL_427297 | Laboratory of Respiratory Viruses and Measles, Oswaldo Cruz Institute, FIOCRUZ | Laboratory of Respiratory Viruses and Measles, Oswaldo Cruz Institute, FIOCRUZ | Paola Resende et al |
| EPI_ISL_427298 | Laboratory of Respiratory Viruses and Measles, Oswaldo Cruz Institute, FIOCRUZ | Laboratory of Respiratory Viruses and Measles, Oswaldo Cruz Institute, FIOCRUZ | Paola Resende et al |
| EPI_ISL_415128 | Laboratório Central de Saúde Pública do Estado do Espírito Santo (LACEN-ES) | Laboratory of Respiratory Viruses and Measles, Oswaldo Cruz Institute, FIOCRUZ | Paola Resende et al |
| EPI_ISL_1181432 | Laboratorio Central de Saude Publica do Estado do Espitito Santo (LACEN-ES) | Laboratory of Respiratory Viruses and Measles, Oswaldo Cruz Institute, FIOCRUZ | Paola Resende et al |
| EPI_ISL_1181418 | Laboratorio Central de Saude Publica do Estado do Espitito Santo (LACEN-ES) | Laboratory of Respiratory Viruses and Measles, Oswaldo Cruz Institute, FIOCRUZ | Paola Resende et al |
| EPI_ISL_1181420 | Laboratorio Central de Saude Publica do Estado do Espitito Santo (LACEN-ES) | Laboratory of Respiratory Viruses and Measles, Oswaldo Cruz Institute, FIOCRUZ | Paola Resende et al |
| EPI_ISL_1181421 | Laboratorio Central de Saude Publica do Estado do Espitito Santo (LACEN-ES) | Laboratory of Respiratory Viruses and Measles, Oswaldo Cruz Institute, FIOCRUZ | Paola Resende et al |
| EPI_ISL_1181423 | Laboratorio Central de Saude Publica do Estado do Espitito Santo (LACEN-ES) | Laboratory of Respiratory Viruses and Measles, Oswaldo Cruz Institute, FIOCRUZ | Paola Resende et al |
| EPI_ISL_1181397 | Laboratorio Central de Saude Publica do Estado do Espitito Santo (LACEN-ES) | Laboratory of Respiratory Viruses and Measles, Oswaldo Cruz Institute, FIOCRUZ | Paola Resende et al |
| EPI_ISL_1181395 | Laboratorio Central de Saude Publica do Estado do Espitito Santo (LACEN-ES) | Laboratory of Respiratory Viruses and Measles, Oswaldo Cruz Institute, FIOCRUZ | Paola Resende et al |
| EPI_ISL_1181583 | Laboratorio Central de Saude Publica do Estado do Espitito Santo (LACEN-ES) | Laboratory of Respiratory Viruses and Measles, Oswaldo Cruz Institute, FIOCRUZ | Paola Resende et al |
| EPI_ISL_1181585 | Laboratorio Central de Saude Publica do Estado do Espitito Santo (LACEN-ES) | Laboratory of Respiratory Viruses and Measles, Oswaldo Cruz Institute, FIOCRUZ | Paola Resende et al |
| EPI_ISL_1181582 | Laboratorio Central de Saude Publica do Estado do Espitito Santo (LACEN-ES) | Laboratory of Respiratory Viruses and Measles, Oswaldo Cruz Institute, FIOCRUZ | Paola Resende et al |
| EPI_ISL_1181480 | Laboratorio de Imunologia de Transplantes de Goias LTDA (HLAGYN) | Laboratory of Respiratory Viruses and Measles, Oswaldo Cruz Institute, FIOCRUZ | Paola Resende et al |

| | | | |
|---|---|---|---|
| EPI_ISL_1181478 | Laboratorio de Imunologia de Transplantes de Goias LTDA (HLAGYN) | Laboratory of Respiratory Viruses and Measles, Oswaldo Cruz Institute, FIOCRUZ | Paola Resende et al |
| EPI_ISL_1181479 | Laboratorio de Imunologia de Transplantes de Goias LTDA (HLAGYN) | Laboratory of Respiratory Viruses and Measles, Oswaldo Cruz Institute, FIOCRUZ | Paola Resende et al |
| EPI_ISL_1181546 | Laboratorio de Imunologia de Transplantes de Goias LTDA (HLAGYN) | Laboratory of Respiratory Viruses and Measles, Oswaldo Cruz Institute, FIOCRUZ | Paola Resende et al |
| EPI_ISL_1181547 | Laboratorio de Imunologia de Transplantes de Goias LTDA (HLAGYN) | Laboratory of Respiratory Viruses and Measles, Oswaldo Cruz Institute, FIOCRUZ | Paola Resende et al |
| EPI_ISL_1181548 | Laboratorio de Imunologia de Transplantes de Goias LTDA (HLAGYN) | Laboratory of Respiratory Viruses and Measles, Oswaldo Cruz Institute, FIOCRUZ | Paola Resende et al |
| EPI_ISL_1181549 | Laboratorio de Imunologia de Transplantes de Goias LTDA (HLAGYN) | Laboratory of Respiratory Viruses and Measles, Oswaldo Cruz Institute, FIOCRUZ | Paola Resende et al |
| EPI_ISL_1181550 | Laboratorio de Imunologia de Transplantes de Goias LTDA (HLAGYN) | Laboratory of Respiratory Viruses and Measles, Oswaldo Cruz Institute, FIOCRUZ | Paola Resende et al |
| EPI_ISL_1181471 | Laboratorio de Imunologia de Transplantes de Goias LTDA (HLAGYN) | Laboratory of Respiratory Viruses and Measles, Oswaldo Cruz Institute, FIOCRUZ | Paola Resende et al |
| EPI_ISL_1181472 | Laboratorio de Imunologia de Transplantes de Goias LTDA (HLAGYN) | Laboratory of Respiratory Viruses and Measles, Oswaldo Cruz Institute, FIOCRUZ | Paola Resende et al |
| EPI_ISL_1181473 | Laboratorio de Imunologia de Transplantes de Goias LTDA (HLAGYN) | Laboratory of Respiratory Viruses and Measles, Oswaldo Cruz Institute, FIOCRUZ | Paola Resende et al |
| EPI_ISL_1181487 | Laboratorio de Imunologia de Transplantes de Goias LTDA (HLAGYN) | Laboratory of Respiratory Viruses and Measles, Oswaldo Cruz Institute, FIOCRUZ | Paola Resende et al |
| EPI_ISL_1181492 | Laboratorio de Imunologia de Transplantes de Goias LTDA (HLAGYN) | Laboratory of Respiratory Viruses and Measles, Oswaldo Cruz Institute, FIOCRUZ | Paola Resende et al |
| EPI_ISL_1181525 | Laboratorio de Imunologia de Transplantes de Goias LTDA (HLAGYN) | Laboratory of Respiratory Viruses and Measles, Oswaldo Cruz Institute, FIOCRUZ | Paola Resende et al |
| EPI_ISL_1181535 | Laboratorio de Imunologia de Transplantes de Goias LTDA (HLAGYN) | Laboratory of Respiratory Viruses and Measles, Oswaldo Cruz Institute, FIOCRUZ | Paola Resende et al |
| EPI_ISL_1181532 | Laboratorio de Imunologia de Transplantes de Goias LTDA (HLAGYN) | Laboratory of Respiratory Viruses and Measles, Oswaldo Cruz Institute, FIOCRUZ | Paola Resende et al |
| EPI_ISL_1181516 | Laboratorio de Imunologia de Transplantes de Goias LTDA (HLAGYN) | Laboratory of Respiratory Viruses and Measles, Oswaldo Cruz Institute, FIOCRUZ | Paola Resende et al |
| EPI_ISL_1181569 | Laboratorio de Imunologia de Transplantes de Goias LTDA (HLAGYN) | Laboratory of Respiratory Viruses and Measles, Oswaldo Cruz Institute, FIOCRUZ | Paola Resende et al |
| EPI_ISL_1181452 | Laboratorio de Imunologia de Transplantes de Goias LTDA (HLAGYN) | Laboratory of Respiratory Viruses and Measles, Oswaldo Cruz Institute, FIOCRUZ | Paola Resende et al |
| EPI_ISL_1181378 | Laboratorio de Imunologia de Transplantes de Goias LTDA (HLAGYN) | Laboratory of Respiratory Viruses and Measles, Oswaldo Cruz Institute, FIOCRUZ | Paola Resende et al |
| EPI_ISL_1465194 | Laboratorio Central de Saude Publica do Estado do Maranhao (LACEN-MA) | Laboratory of Respiratory Viruses and Measles, Oswaldo Cruz Institute, FIOCRUZ | Paola Resende et al |
| EPI_ISL_1465217 | Laboratorio Central de Saude Publica do Estado do Maranhao (LACEN-MA) | Laboratory of Respiratory Viruses and Measles, Oswaldo Cruz Institute, FIOCRUZ | Paola Resende et al |
| EPI_ISL_1465215 | Laboratorio Central de Saude Publica do Estado do Maranhao (LACEN-MA) | Laboratory of Respiratory Viruses and Measles, Oswaldo Cruz Institute, FIOCRUZ | Paola Resende et al |

| | | | |
|---|---|---|---|
| EPI_ISL_1465203 | Laboratorio Central de Saude Publica do Estado do Maranhao (LACEN-MA) | Laboratory of Respiratory Viruses and Measles, Oswaldo Cruz Institute, FIOCRUZ | Paola Resende et al |
| EPI_ISL_1465206 | Laboratorio Central de Saude Publica do Estado do Maranhao (LACEN-MA) | Laboratory of Respiratory Viruses and Measles, Oswaldo Cruz Institute, FIOCRUZ | Paola Resende et al |
| EPI_ISL_1465213 | Laboratorio Central de Saude Publica do Estado do Maranhao (LACEN-MA) | Laboratory of Respiratory Viruses and Measles, Oswaldo Cruz Institute, FIOCRUZ | Paola Resende et al |
| EPI_ISL_1465212 | Laboratorio Central de Saude Publica do Estado do Maranhao (LACEN-MA) | Laboratory of Respiratory Viruses and Measles, Oswaldo Cruz Institute, FIOCRUZ | Paola Resende et al |
| EPI_ISL_1465208 | Laboratorio Central de Saude Publica do Estado do Maranhao (LACEN-MA) | Laboratory of Respiratory Viruses and Measles, Oswaldo Cruz Institute, FIOCRUZ | Paola Resende et al |
| EPI_ISL_1465209 | Laboratorio Central de Saude Publica do Estado do Maranhao (LACEN-MA) | Laboratory of Respiratory Viruses and Measles, Oswaldo Cruz Institute, FIOCRUZ | Paola Resende et al |
| EPI_ISL_1465210 | Laboratorio Central de Saude Publica do Estado do Maranhao (LACEN-MA) | Laboratory of Respiratory Viruses and Measles, Oswaldo Cruz Institute, FIOCRUZ | Paola Resende et al |
| EPI_ISL_1465199 | Laboratorio Central de Saude Publica do Estado do Maranhao (LACEN-MA) | Laboratory of Respiratory Viruses and Measles, Oswaldo Cruz Institute, FIOCRUZ | Paola Resende et al |
| EPI_ISL_1465201 | Laboratorio Central de Saude Publica do Estado do Maranhao (LACEN-MA) | Laboratory of Respiratory Viruses and Measles, Oswaldo Cruz Institute, FIOCRUZ | Paola Resende et al |
| EPI_ISL_1465202 | Laboratorio Central de Saude Publica do Estado do Maranhao (LACEN-MA) | Laboratory of Respiratory Viruses and Measles, Oswaldo Cruz Institute, FIOCRUZ | Paola Resende et al |
| EPI_ISL_1465192 | Laboratorio Central de Saude Publica do Estado do Maranhao (LACEN-MA) | Laboratory of Respiratory Viruses and Measles, Oswaldo Cruz Institute, FIOCRUZ | Paola Resende et al |
| EPI_ISL_1465189 | Laboratorio Central de Saude Publica do Estado do Maranhao (LACEN-MA) | Laboratory of Respiratory Viruses and Measles, Oswaldo Cruz Institute, FIOCRUZ | Paola Resende et al |
| EPI_ISL_1465225 | Laboratorio Central de Saude Publica do Estado do Maranhao (LACEN-MA) | Laboratory of Respiratory Viruses and Measles, Oswaldo Cruz Institute, FIOCRUZ | Paola Resende et al |
| EPI_ISL_1465195 | Laboratorio Central de Saude Publica do Estado do Maranhao (LACEN-MA) | Laboratory of Respiratory Viruses and Measles, Oswaldo Cruz Institute, FIOCRUZ | Paola Resende et al |
| EPI_ISL_1465205 | Laboratorio Central de Saude Publica do Estado do Maranhao (LACEN-MA) | Laboratory of Respiratory Viruses and Measles, Oswaldo Cruz Institute, FIOCRUZ | Paola Resende et al |
| EPI_ISL_1465196 | Laboratorio Central de Saude Publica do Estado do Maranhao (LACEN-MA) | Laboratory of Respiratory Viruses and Measles, Oswaldo Cruz Institute, FIOCRUZ | Paola Resende et al |
| EPI_ISL_1465191 | Laboratorio Central de Saude Publica do Estado do Maranhao (LACEN-MA) | Laboratory of Respiratory Viruses and Measles, Oswaldo Cruz Institute, FIOCRUZ | Paola Resende et al |
| EPI_ISL_1465216 | Laboratorio Central de Saude Publica do Estado do Maranhao (LACEN-MA) | Laboratory of Respiratory Viruses and Measles, Oswaldo Cruz Institute, FIOCRUZ | Paola Resende et al |
| EPI_ISL_1465221 | Laboratorio Central de Saude Publica do Estado do Maranhao (LACEN-MA) | Laboratory of Respiratory Viruses and Measles, Oswaldo Cruz Institute, FIOCRUZ | Paola Resende et al |
| EPI_ISL_1465219 | Laboratorio Central de Saude Publica do Estado do Maranhao (LACEN-MA) | Laboratory of Respiratory Viruses and Measles, Oswaldo Cruz Institute, FIOCRUZ | Paola Resende et al |
| EPI_ISL_1465220 | Laboratorio Central de Saude Publica do Estado do Maranhao (LACEN-MA) | Laboratory of Respiratory Viruses and Measles, Oswaldo Cruz Institute, FIOCRUZ | Paola Resende et al |
| EPI_ISL_1465261 | Laboratorio Central de Saude Publica do Estado do Maranhao (LACEN-MA) | Laboratory of Respiratory Viruses and Measles, Oswaldo Cruz Institute, FIOCRUZ | Paola Resende et al |

| | | | |
|---|---|---|---|
| EPI_ISL_1465246 | Laboratorio Central de Saude Publica do Estado do Maranhao (LACEN-MA) | Laboratory of Respiratory Viruses and Measles, Oswaldo Cruz Institute, FIOCRUZ | Paola Resende et al |
| EPI_ISL_1465275 | Laboratorio Central de Saude Publica do Estado do Maranhao (LACEN-MA) | Laboratory of Respiratory Viruses and Measles, Oswaldo Cruz Institute, FIOCRUZ | Paola Resende et al |
| EPI_ISL_1465277 | Laboratorio Central de Saude Publica do Estado do Maranhao (LACEN-MA) | Laboratory of Respiratory Viruses and Measles, Oswaldo Cruz Institute, FIOCRUZ | Paola Resende et al |
| EPI_ISL_1465271 | Laboratorio Central de Saude Publica do Estado do Maranhao (LACEN-MA) | Laboratory of Respiratory Viruses and Measles, Oswaldo Cruz Institute, FIOCRUZ | Paola Resende et al |
| EPI_ISL_1465222 | Laboratorio Central de Saude Publica do Estado do Maranhao (LACEN-MA) | Laboratory of Respiratory Viruses and Measles, Oswaldo Cruz Institute, FIOCRUZ | Paola Resende et al |
| EPI_ISL_1465268 | Laboratorio Central de Saude Publica do Estado do Maranhao (LACEN-MA) | Laboratory of Respiratory Viruses and Measles, Oswaldo Cruz Institute, FIOCRUZ | Paola Resende et al |
| EPI_ISL_1465262 | Laboratorio Central de Saude Publica do Estado do Maranhao (LACEN-MA) | Laboratory of Respiratory Viruses and Measles, Oswaldo Cruz Institute, FIOCRUZ | Paola Resende et al |
| EPI_ISL_1465267 | Laboratorio Central de Saude Publica do Estado do Maranhao (LACEN-MA) | Laboratory of Respiratory Viruses and Measles, Oswaldo Cruz Institute, FIOCRUZ | Paola Resende et al |
| EPI_ISL_1465270 | Laboratorio Central de Saude Publica do Estado do Maranhao (LACEN-MA) | Laboratory of Respiratory Viruses and Measles, Oswaldo Cruz Institute, FIOCRUZ | Paola Resende et al |
| EPI_ISL_1465278 | Laboratorio Central de Saude Publica do Estado do Maranhao (LACEN-MA) | Laboratory of Respiratory Viruses and Measles, Oswaldo Cruz Institute, FIOCRUZ | Paola Resende et al |
| EPI_ISL_1181414 | Laboratorio Central de Saude Publica do Estado Maranhao (LACEN-MA) | Laboratory of Respiratory Viruses and Measles, Oswaldo Cruz Institute, FIOCRUZ | Paola Resende et al |
| EPI_ISL_1181400 | Laboratorio Central de Saude Publica do Estado Maranhao (LACEN-MA) | Laboratory of Respiratory Viruses and Measles, Oswaldo Cruz Institute, FIOCRUZ | Paola Resende et al |
| EPI_ISL_1181426 | Laboratorio Central de Saude Publica do Estado de Minas Gerais (LACEN-MG) | Laboratory of Respiratory Viruses and Measles, Oswaldo Cruz Institute, FIOCRUZ | Paola Resende et al |
| EPI_ISL_1181427 | Laboratorio Central de Saude Publica do Estado de Minas Gerais (LACEN-MG) | Laboratory of Respiratory Viruses and Measles, Oswaldo Cruz Institute, FIOCRUZ | Paola Resende et al |
| EPI_ISL_1181428 | Laboratorio Central de Saude Publica do Estado de Minas Gerais (LACEN-MG) | Laboratory of Respiratory Viruses and Measles, Oswaldo Cruz Institute, FIOCRUZ | Paola Resende et al |
| EPI_ISL_1181399 | Laboratorio Central de Saude Publica do Estado de Minas Gerais (LACEN-MG) | Laboratory of Respiratory Viruses and Measles, Oswaldo Cruz Institute, FIOCRUZ | Paola Resende et al |
| EPI_ISL_1219137 | Laboratorio Central de Saude Publica do Estado de Minas Gerais (LACEN-MG) | Laboratory of Respiratory Viruses and Measles, Oswaldo Cruz Institute, FIOCRUZ | Paola Resende et al |
| EPI_ISL_1181469 | Federal University of Mato Grosso (UFMT) | Laboratory of Respiratory Viruses and Measles, Oswaldo Cruz Institute, FIOCRUZ | Paola Resende et al |
| EPI_ISL_1219138 | Federal University of Mato Grosso (UFMT) | Laboratory of Respiratory Viruses and Measles, Oswaldo Cruz Institute, FIOCRUZ | Paola Resende et al |
| EPI_ISL_792563 | Laboratório Central de Saúde Pública do Estado da Paraíba (LACEN-PB) | Laboratory of Respiratory Viruses and Measles, Oswaldo Cruz Institute, FIOCRUZ | Paola Resende et al |
| EPI_ISL_792564 | Laboratório Central de Saúde Pública do Estado da Paraíba (LACEN-PB) | Laboratory of Respiratory Viruses and Measles, Oswaldo Cruz Institute, FIOCRUZ | Paola Resende et al |
| EPI_ISL_792565 | Laboratório Central de Saúde Pública do Estado da Paraíba (LACEN-PB) | Laboratory of Respiratory Viruses and Measles, Oswaldo Cruz Institute, FIOCRUZ | Paola Resende et al |

| | | | |
|---|---|---|---|
| EPI_ISL_792566 | Laboratório Central de Saúde Pública do Estado da Paraíba (LACEN-PB) | Laboratory of Respiratory Viruses and Measles, Oswaldo Cruz Institute, FIOCRUZ | Paola Resende et al |
| EPI_ISL_792568 | Laboratório Central de Saúde Pública do Estado da Paraíba (LACEN-PB) | Laboratory of Respiratory Viruses and Measles, Oswaldo Cruz Institute, FIOCRUZ | Paola Resende et al |
| EPI_ISL_792569 | Laboratório Central de Saúde Pública do Estado da Paraíba (LACEN-PB) | Laboratory of Respiratory Viruses and Measles, Oswaldo Cruz Institute, FIOCRUZ | Paola Resende et al |
| EPI_ISL_792570 | Laboratório Central de Saúde Pública do Estado da Paraíba (LACEN-PB) | Laboratory of Respiratory Viruses and Measles, Oswaldo Cruz Institute, FIOCRUZ | Paola Resende et al |
| EPI_ISL_792571 | Laboratório Central de Saúde Pública do Estado da Paraíba (LACEN-PB) | Laboratory of Respiratory Viruses and Measles, Oswaldo Cruz Institute, FIOCRUZ | Paola Resende et al |
| EPI_ISL_792572 | Laboratório Central de Saúde Pública do Estado da Paraíba (LACEN-PB) | Laboratory of Respiratory Viruses and Measles, Oswaldo Cruz Institute, FIOCRUZ | Paola Resende et al |
| EPI_ISL_792573 | Laboratório Central de Saúde Pública do Estado da Paraíba (LACEN-PB) | Laboratory of Respiratory Viruses and Measles, Oswaldo Cruz Institute, FIOCRUZ | Paola Resende et al |
| EPI_ISL_792574 | Laboratório Central de Saúde Pública do Estado da Paraíba (LACEN-PB) | Laboratory of Respiratory Viruses and Measles, Oswaldo Cruz Institute, FIOCRUZ | Paola Resende et al |
| EPI_ISL_792575 | Laboratório Central de Saúde Pública do Estado da Paraíba (LACEN-PB) | Laboratory of Respiratory Viruses and Measles, Oswaldo Cruz Institute, FIOCRUZ | Paola Resende et al |
| EPI_ISL_792576 | Laboratório Central de Saúde Pública do Estado da Paraíba (LACEN-PB) | Laboratory of Respiratory Viruses and Measles, Oswaldo Cruz Institute, FIOCRUZ | Paola Resende et al |
| EPI_ISL_792577 | Laboratório Central de Saúde Pública do Estado da Paraíba (LACEN-PB) | Laboratory of Respiratory Viruses and Measles, Oswaldo Cruz Institute, FIOCRUZ | Paola Resende et al |
| EPI_ISL_792578 | Laboratório Central de Saúde Pública do Estado da Paraíba (LACEN-PB) | Laboratory of Respiratory Viruses and Measles, Oswaldo Cruz Institute, FIOCRUZ | Paola Resende et al |
| EPI_ISL_792579 | Laboratório Central de Saúde Pública do Estado da Paraíba (LACEN-PB) | Laboratory of Respiratory Viruses and Measles, Oswaldo Cruz Institute, FIOCRUZ | Paola Resende et al |
| EPI_ISL_792583 | Laboratório Central de Saúde Pública do Estado da Paraíba (LACEN-PB) | Laboratory of Respiratory Viruses and Measles, Oswaldo Cruz Institute, FIOCRUZ | Paola Resende et al |
| EPI_ISL_792585 | Laboratório Central de Saúde Pública do Estado da Paraíba (LACEN-PB) | Laboratory of Respiratory Viruses and Measles, Oswaldo Cruz Institute, FIOCRUZ | Paola Resende et al |
| EPI_ISL_792587 | Laboratório Central de Saúde Pública do Estado da Paraíba (LACEN-PB) | Laboratory of Respiratory Viruses and Measles, Oswaldo Cruz Institute, FIOCRUZ | Paola Resende et al |
| EPI_ISL_792588 | Laboratório Central de Saúde Pública do Estado da Paraíba (LACEN-PB) | Laboratory of Respiratory Viruses and Measles, Oswaldo Cruz Institute, FIOCRUZ | Paola Resende et al |
| EPI_ISL_792589 | Laboratório Central de Saúde Pública do Estado da Paraíba (LACEN-PB) | Laboratory of Respiratory Viruses and Measles, Oswaldo Cruz Institute, FIOCRUZ | Paola Resende et al |
| EPI_ISL_792590 | Laboratório Central de Saúde Pública do Estado da Paraíba (LACEN-PB) | Laboratory of Respiratory Viruses and Measles, Oswaldo Cruz Institute, FIOCRUZ | Paola Resende et al |
| EPI_ISL_792593 | Laboratório Central de Saúde Pública do Estado da Paraíba (LACEN-PB) | Laboratory of Respiratory Viruses and Measles, Oswaldo Cruz Institute, FIOCRUZ | Paola Resende et al |
| EPI_ISL_792594 | Laboratório Central de Saúde Pública do Estado da Paraíba (LACEN-PB) | Laboratory of Respiratory Viruses and Measles, Oswaldo Cruz Institute, FIOCRUZ | Paola Resende et al |
| EPI_ISL_792595 | Laboratório Central de Saúde Pública do Estado da Paraíba (LACEN-PB) | Laboratory of Respiratory Viruses and Measles, Oswaldo Cruz Institute, FIOCRUZ | Paola Resende et al |

| | | | |
|---|---|---|---|
| EPI_ISL_792596 | Laboratório Central de Saúde Pública do Estado da Paraíba (LACEN-PB) | Laboratory of Respiratory Viruses and Measles, Oswaldo Cruz Institute, FIOCRUZ | Paola Resende et al |
| EPI_ISL_792597 | Laboratório Central de Saúde Pública do Estado da Paraíba (LACEN-PB) | Laboratory of Respiratory Viruses and Measles, Oswaldo Cruz Institute, FIOCRUZ | Paola Resende et al |
| EPI_ISL_792598 | Laboratório Central de Saúde Pública do Estado da Paraíba (LACEN-PB) | Laboratory of Respiratory Viruses and Measles, Oswaldo Cruz Institute, FIOCRUZ | Paola Resende et al |
| EPI_ISL_792599 | Laboratório Central de Saúde Pública do Estado da Paraíba (LACEN-PB) | Laboratory of Respiratory Viruses and Measles, Oswaldo Cruz Institute, FIOCRUZ | Paola Resende et al |
| EPI_ISL_792561 | Laboratório Central de Saúde Pública do Estado da Paraíba (LACEN-PB) | Laboratory of Respiratory Viruses and Measles, Oswaldo Cruz Institute, FIOCRUZ | Paola Resende et al |
| EPI_ISL_792562 | Laboratório Central de Saúde Pública do Estado da Paraíba (LACEN-PB) | Laboratory of Respiratory Viruses and Measles, Oswaldo Cruz Institute, FIOCRUZ | Paola Resende et al |
| EPI_ISL_792600 | Laboratório Central de Saúde Pública do Estado da Paraíba (LACEN-PB) | Laboratory of Respiratory Viruses and Measles, Oswaldo Cruz Institute, FIOCRUZ | Paola Resende et al |
| EPI_ISL_792601 | Laboratório Central de Saúde Pública do Estado da Paraíba (LACEN-PB) | Laboratory of Respiratory Viruses and Measles, Oswaldo Cruz Institute, FIOCRUZ | Paola Resende et al |
| EPI_ISL_792602 | Laboratório Central de Saúde Pública do Estado da Paraíba (LACEN-PB) | Laboratory of Respiratory Viruses and Measles, Oswaldo Cruz Institute, FIOCRUZ | Paola Resende et al |
| EPI_ISL_792603 | Laboratório Central de Saúde Pública do Estado da Paraíba (LACEN-PB) | Laboratory of Respiratory Viruses and Measles, Oswaldo Cruz Institute, FIOCRUZ | Paola Resende et al |
| EPI_ISL_792604 | Laboratório Central de Saúde Pública do Estado da Paraíba (LACEN-PB) | Laboratory of Respiratory Viruses and Measles, Oswaldo Cruz Institute, FIOCRUZ | Paola Resende et al |
| EPI_ISL_792605 | Laboratório Central de Saúde Pública do Estado da Paraíba (LACEN-PB) | Laboratory of Respiratory Viruses and Measles, Oswaldo Cruz Institute, FIOCRUZ | Paola Resende et al |
| EPI_ISL_792606 | Laboratório Central de Saúde Pública do Estado da Paraíba (LACEN-PB) | Laboratory of Respiratory Viruses and Measles, Oswaldo Cruz Institute, FIOCRUZ | Paola Resende et al |
| EPI_ISL_792607 | Laboratório Central de Saúde Pública do Estado da Paraíba (LACEN-PB) | Laboratory of Respiratory Viruses and Measles, Oswaldo Cruz Institute, FIOCRUZ | Paola Resende et al |
| EPI_ISL_792608 | Laboratório Central de Saúde Pública do Estado da Paraíba (LACEN-PB) | Laboratory of Respiratory Viruses and Measles, Oswaldo Cruz Institute, FIOCRUZ | Paola Resende et al |
| EPI_ISL_792609 | Laboratório Central de Saúde Pública do Estado da Paraíba (LACEN-PB) | Laboratory of Respiratory Viruses and Measles, Oswaldo Cruz Institute, FIOCRUZ | Paola Resende et al |
| EPI_ISL_792610 | Laboratório Central de Saúde Pública do Estado da Paraíba (LACEN-PB) | Laboratory of Respiratory Viruses and Measles, Oswaldo Cruz Institute, FIOCRUZ | Paola Resende et al |
| EPI_ISL_792611 | Laboratório Central de Saúde Pública do Estado da Paraíba (LACEN-PB) | Laboratory of Respiratory Viruses and Measles, Oswaldo Cruz Institute, FIOCRUZ | Paola Resende et al |
| EPI_ISL_792612 | Laboratório Central de Saúde Pública do Estado da Paraíba (LACEN-PB) | Laboratory of Respiratory Viruses and Measles, Oswaldo Cruz Institute, FIOCRUZ | Paola Resende et al |
| EPI_ISL_792613 | Laboratório Central de Saúde Pública do Estado da Paraíba (LACEN-PB) | Laboratory of Respiratory Viruses and Measles, Oswaldo Cruz Institute, FIOCRUZ | Paola Resende et al |
| EPI_ISL_792614 | Laboratório Central de Saúde Pública do Estado da Paraíba (LACEN-PB) | Laboratory of Respiratory Viruses and Measles, Oswaldo Cruz Institute, FIOCRUZ | Paola Resende et al |
| EPI_ISL_792615 | Laboratório Central de Saúde Pública do Estado da Paraíba (LACEN-PB) | Laboratory of Respiratory Viruses and Measles, Oswaldo Cruz Institute, FIOCRUZ | Paola Resende et al |

| | | | |
|---|---|---|---|
| EPI_ISL_792616 | Laboratório Central de Saúde Pública do Estado da Paraíba (LACEN-PB) | Laboratory of Respiratory Viruses and Measles, Oswaldo Cruz Institute, FIOCRUZ | Paola Resende et al |
| EPI_ISL_792617 | Laboratório Central de Saúde Pública do Estado da Paraíba (LACEN-PB) | Laboratory of Respiratory Viruses and Measles, Oswaldo Cruz Institute, FIOCRUZ | Paola Resende et al |
| EPI_ISL_792618 | Laboratório Central de Saúde Pública do Estado da Paraíba (LACEN-PB) | Laboratory of Respiratory Viruses and Measles, Oswaldo Cruz Institute, FIOCRUZ | Paola Resende et al |
| EPI_ISL_792619 | Laboratório Central de Saúde Pública do Estado da Paraíba (LACEN-PB) | Laboratory of Respiratory Viruses and Measles, Oswaldo Cruz Institute, FIOCRUZ | Paola Resende et al |
| EPI_ISL_792620 | Laboratório Central de Saúde Pública do Estado da Paraíba (LACEN-PB) | Laboratory of Respiratory Viruses and Measles, Oswaldo Cruz Institute, FIOCRUZ | Paola Resende et al |
| EPI_ISL_792621 | Laboratório Central de Saúde Pública do Estado da Paraíba (LACEN-PB) | Laboratory of Respiratory Viruses and Measles, Oswaldo Cruz Institute, FIOCRUZ | Paola Resende et al |
| EPI_ISL_792622 | Laboratório Central de Saúde Pública do Estado da Paraíba (LACEN-PB) | Laboratory of Respiratory Viruses and Measles, Oswaldo Cruz Institute, FIOCRUZ | Paola Resende et al |
| EPI_ISL_792623 | Laboratório Central de Saúde Pública do Estado da Paraíba (LACEN-PB) | Laboratory of Respiratory Viruses and Measles, Oswaldo Cruz Institute, FIOCRUZ | Paola Resende et al |
| EPI_ISL_792624 | Laboratório Central de Saúde Pública do Estado da Paraíba (LACEN-PB) | Laboratory of Respiratory Viruses and Measles, Oswaldo Cruz Institute, FIOCRUZ | Paola Resende et al |
| EPI_ISL_792625 | Laboratório Central de Saúde Pública do Estado da Paraíba (LACEN-PB) | Laboratory of Respiratory Viruses and Measles, Oswaldo Cruz Institute, FIOCRUZ | Paola Resende et al |
| EPI_ISL_792626 | Laboratório Central de Saúde Pública do Estado da Paraíba (LACEN-PB) | Laboratory of Respiratory Viruses and Measles, Oswaldo Cruz Institute, FIOCRUZ | Paola Resende et al |
| EPI_ISL_792627 | Laboratório Central de Saúde Pública do Estado da Paraíba (LACEN-PB) | Laboratory of Respiratory Viruses and Measles, Oswaldo Cruz Institute, FIOCRUZ | Paola Resende et al |
| EPI_ISL_792628 | Laboratório Central de Saúde Pública do Estado da Paraíba (LACEN-PB) | Laboratory of Respiratory Viruses and Measles, Oswaldo Cruz Institute, FIOCRUZ | Paola Resende et al |
| EPI_ISL_792629 | Laboratório Central de Saúde Pública do Estado da Paraíba (LACEN-PB) | Laboratory of Respiratory Viruses and Measles, Oswaldo Cruz Institute, FIOCRUZ | Paola Resende et al |
| EPI_ISL_792630 | Laboratório Central de Saúde Pública do Estado da Paraíba (LACEN-PB) | Laboratory of Respiratory Viruses and Measles, Oswaldo Cruz Institute, FIOCRUZ | Paola Resende et al |
| EPI_ISL_792631 | Laboratório Central de Saúde Pública do Estado da Paraíba (LACEN-PB) | Laboratory of Respiratory Viruses and Measles, Oswaldo Cruz Institute, FIOCRUZ | Paola Resende et al |
| EPI_ISL_792632 | Laboratório Central de Saúde Pública do Estado da Paraíba (LACEN-PB) | Laboratory of Respiratory Viruses and Measles, Oswaldo Cruz Institute, FIOCRUZ | Paola Resende et al |
| EPI_ISL_792633 | Laboratório Central de Saúde Pública do Estado da Paraíba (LACEN-PB) | Laboratory of Respiratory Viruses and Measles, Oswaldo Cruz Institute, FIOCRUZ | Paola Resende et al |
| EPI_ISL_792634 | Laboratório Central de Saúde Pública do Estado da Paraíba (LACEN-PB) | Laboratory of Respiratory Viruses and Measles, Oswaldo Cruz Institute, FIOCRUZ | Paola Resende et al |
| EPI_ISL_792635 | Laboratório Central de Saúde Pública do Estado da Paraíba (LACEN-PB) | Laboratory of Respiratory Viruses and Measles, Oswaldo Cruz Institute, FIOCRUZ | Paola Resende et al |
| EPI_ISL_792636 | Laboratório Central de Saúde Pública do Estado da Paraíba (LACEN-PB) | Laboratory of Respiratory Viruses and Measles, Oswaldo Cruz Institute, FIOCRUZ | Paola Resende et al |
| EPI_ISL_792637 | Laboratório Central de Saúde Pública do Estado da Paraíba (LACEN-PB) | Laboratory of Respiratory Viruses and Measles, Oswaldo Cruz Institute, FIOCRUZ | Paola Resende et al |

| | | | |
|---|---|---|---|
| EPI_ISL_792638 | Laboratório Central de Saúde Pública do Estado da Paraíba (LACEN-PB) | Laboratory of Respiratory Viruses and Measles, Oswaldo Cruz Institute, FIOCRUZ | Paola Resende et al |
| EPI_ISL_1402429 | Laboratory of Respiratory Viruses and Measles, Oswaldo Cruz Institute, FIOCRUZ | Laboratory of Respiratory Viruses and Measles, Oswaldo Cruz Institute, FIOCRUZ | Paola Resende et al |
| EPI_ISL_1181416 | Laboratorio Central de Saude Publica do Estado da Paraiba (LACEN-PB) | Laboratory of Respiratory Viruses and Measles, Oswaldo Cruz Institute, FIOCRUZ | Paola Resende et al |
| EPI_ISL_1181422 | Laboratorio Central de Saude Publica do Estado da Paraiba (LACEN-PB) | Laboratory of Respiratory Viruses and Measles, Oswaldo Cruz Institute, FIOCRUZ | Paola Resende et al |
| EPI_ISL_1181362 | Instituto Aggeu Magalhães - Oswaldo Cruz Foundation, FIOCRUZ (FIOCRUZ-PE) | Laboratory of Respiratory Viruses and Measles, Oswaldo Cruz Institute, FIOCRUZ | Paola Resende et al |
| EPI_ISL_1181526 | Laboratorio Central de Saude Publica do Estado do Parana (LACEN-PR) | Laboratory of Respiratory Viruses and Measles, Oswaldo Cruz Institute, FIOCRUZ | Paola Resende et al |
| EPI_ISL_1181527 | Laboratorio Central de Saude Publica do Estado do Parana (LACEN-PR) | Laboratory of Respiratory Viruses and Measles, Oswaldo Cruz Institute, FIOCRUZ | Paola Resende et al |
| EPI_ISL_1181528 | Laboratorio Central de Saude Publica do Estado do Parana (LACEN-PR) | Laboratory of Respiratory Viruses and Measles, Oswaldo Cruz Institute, FIOCRUZ | Paola Resende et al |
| EPI_ISL_1181563 | Laboratorio Central de Saude Publica do Estado do Parana (LACEN-PR) | Laboratory of Respiratory Viruses and Measles, Oswaldo Cruz Institute, FIOCRUZ | Paola Resende et al |
| EPI_ISL_1181551 | Laboratorio Central de Saude Publica do Estado do Parana (LACEN-PR) | Laboratory of Respiratory Viruses and Measles, Oswaldo Cruz Institute, FIOCRUZ | Paola Resende et al |
| EPI_ISL_1181552 | Laboratorio Central de Saude Publica do Estado do Parana (LACEN-PR) | Laboratory of Respiratory Viruses and Measles, Oswaldo Cruz Institute, FIOCRUZ | Paola Resende et al |
| EPI_ISL_1181553 | Laboratorio Central de Saude Publica do Estado do Parana (LACEN-PR) | Laboratory of Respiratory Viruses and Measles, Oswaldo Cruz Institute, FIOCRUZ | Paola Resende et al |
| EPI_ISL_1181543 | Laboratorio Central de Saude Publica do Estado do Parana (LACEN-PR) | Laboratory of Respiratory Viruses and Measles, Oswaldo Cruz Institute, FIOCRUZ | Paola Resende et al |
| EPI_ISL_1181537 | Laboratorio Central de Saude Publica do Estado do Parana (LACEN-PR) | Laboratory of Respiratory Viruses and Measles, Oswaldo Cruz Institute, FIOCRUZ | Paola Resende et al |
| EPI_ISL_1181533 | Laboratorio Central de Saude Publica do Estado do Parana (LACEN-PR) | Laboratory of Respiratory Viruses and Measles, Oswaldo Cruz Institute, FIOCRUZ | Paola Resende et al |
| EPI_ISL_1181534 | Laboratorio Central de Saude Publica do Estado do Parana (LACEN-PR) | Laboratory of Respiratory Viruses and Measles, Oswaldo Cruz Institute, FIOCRUZ | Paola Resende et al |
| EPI_ISL_1181538 | Laboratorio Central de Saude Publica do Estado do Parana (LACEN-PR) | Laboratory of Respiratory Viruses and Measles, Oswaldo Cruz Institute, FIOCRUZ | Paola Resende et al |
| EPI_ISL_1181539 | Laboratorio Central de Saude Publica do Estado do Parana (LACEN-PR) | Laboratory of Respiratory Viruses and Measles, Oswaldo Cruz Institute, FIOCRUZ | Paola Resende et al |
| EPI_ISL_1181392 | Laboratorio Central de Saude Publica do Estado do Parana (LACEN-PR) | Laboratory of Respiratory Viruses and Measles, Oswaldo Cruz Institute, FIOCRUZ | Paola Resende et al |
| EPI_ISL_1181545 | Laboratorio Central de Saude Publica do Estado do Parana (LACEN-PR) | Laboratory of Respiratory Viruses and Measles, Oswaldo Cruz Institute, FIOCRUZ | Paola Resende et al |
| EPI_ISL_1181540 | Laboratorio Central de Saude Publica do Estado do Parana (LACEN-PR) | Laboratory of Respiratory Viruses and Measles, Oswaldo Cruz Institute, FIOCRUZ | Paola Resende et al |
| EPI_ISL_1181544 | Laboratorio Central de Saude Publica do Estado do Parana (LACEN-PR) | Laboratory of Respiratory Viruses and Measles, Oswaldo Cruz Institute, FIOCRUZ | Paola Resende et al |

| | | | |
|---|---|---|---|
| EPI_ISL_1181541 | Laboratorio Central de Saude Publica do Estado do Parana (LACEN-PR) | Laboratory of Respiratory Viruses and Measles, Oswaldo Cruz Institute, FIOCRUZ | Paola Resende et al |
| EPI_ISL_1181542 | Laboratorio Central de Saude Publica do Estado do Parana (LACEN-PR) | Laboratory of Respiratory Viruses and Measles, Oswaldo Cruz Institute, FIOCRUZ | Paola Resende et al |
| EPI_ISL_1181605 | Laboratorio Central de Saude Publica do Estado do Parana (LACEN-PR) | Laboratory of Respiratory Viruses and Measles, Oswaldo Cruz Institute, FIOCRUZ | Paola Resende et al |
| EPI_ISL_1181584 | Laboratorio Central de Saude Publica do Estado do Parana (LACEN-PR) | Laboratory of Respiratory Viruses and Measles, Oswaldo Cruz Institute, FIOCRUZ | Paola Resende et al |
| EPI_ISL_1181581 | Laboratorio Central de Saude Publica do Estado do Parana (LACEN-PR) | Laboratory of Respiratory Viruses and Measles, Oswaldo Cruz Institute, FIOCRUZ | Paola Resende et al |
| EPI_ISL_1181580 | Laboratorio Central de Saude Publica do Estado do Parana (LACEN-PR) | Laboratory of Respiratory Viruses and Measles, Oswaldo Cruz Institute, FIOCRUZ | Paola Resende et al |
| EPI_ISL_1181576 | Laboratorio Central de Saude Publica do Estado do Parana (LACEN-PR) | Laboratory of Respiratory Viruses and Measles, Oswaldo Cruz Institute, FIOCRUZ | Paola Resende et al |
| EPI_ISL_1181536 | Laboratorio Central de Saude Publica do Estado do Parana (LACEN-PR) | Laboratory of Respiratory Viruses and Measles, Oswaldo Cruz Institute, FIOCRUZ | Paola Resende et al |
| EPI_ISL_1181530 | Laboratorio Central de Saude Publica do Estado do Parana (LACEN-PR) | Laboratory of Respiratory Viruses and Measles, Oswaldo Cruz Institute, FIOCRUZ | Paola Resende et al |
| EPI_ISL_1181517 | Laboratorio Central de Saude Publica do Estado do Parana (LACEN-PR) | Laboratory of Respiratory Viruses and Measles, Oswaldo Cruz Institute, FIOCRUZ | Paola Resende et al |
| EPI_ISL_1181498 | Laboratorio Central de Saude Publica do Estado do Parana (LACEN-PR) | Laboratory of Respiratory Viruses and Measles, Oswaldo Cruz Institute, FIOCRUZ | Paola Resende et al |
| EPI_ISL_1534000 | Laboratorio Central de Saude Publica do Estado do Parana (LACEN-PR) | Laboratory of Respiratory Viruses and Measles, Oswaldo Cruz Institute, FIOCRUZ | Paola Resende et al |
| EPI_ISL_1534001 | Laboratorio Central de Saude Publica do Estado do Parana (LACEN-PR) | Laboratory of Respiratory Viruses and Measles, Oswaldo Cruz Institute, FIOCRUZ | Paola Resende et al |
| EPI_ISL_1533999 | Laboratorio Central de Saude Publica do Estado do Parana (LACEN-PR) | Laboratory of Respiratory Viruses and Measles, Oswaldo Cruz Institute, FIOCRUZ | Paola Resende et al |
| EPI_ISL_1533978 | Laboratorio Central de Saude Publica do Estado do Parana (LACEN-PR) | Laboratory of Respiratory Viruses and Measles, Oswaldo Cruz Institute, FIOCRUZ | Paola Resende et al |
| EPI_ISL_1181474 | Laboratorio Central de Saude Publica do Estado do Parana (LACEN-PR) | Laboratory of Respiratory Viruses and Measles, Oswaldo Cruz Institute, FIOCRUZ | Paola Resende et al |
| EPI_ISL_792645 | Laboratório Central de Saúde Pública do Estado do Paraná (LACEN-PR) | Laboratory of Respiratory Viruses and Measles, Oswaldo Cruz Institute, FIOCRUZ | Paola Resende et al |
| EPI_ISL_792646 | Laboratório Central de Saúde Pública do Estado do Paraná (LACEN-PR) | Laboratory of Respiratory Viruses and Measles, Oswaldo Cruz Institute, FIOCRUZ | Paola Resende et al |
| EPI_ISL_792647 | Laboratório Central de Saúde Pública do Estado do Paraná (LACEN-PR) | Laboratory of Respiratory Viruses and Measles, Oswaldo Cruz Institute, FIOCRUZ | Paola Resende et al |
| EPI_ISL_792648 | Laboratório Central de Saúde Pública do Estado do Paraná (LACEN-PR) | Laboratory of Respiratory Viruses and Measles, Oswaldo Cruz Institute, FIOCRUZ | Paola Resende et al |
| EPI_ISL_792649 | Laboratório Central de Saúde Pública do Estado do Paraná (LACEN-PR) | Laboratory of Respiratory Viruses and Measles, Oswaldo Cruz Institute, FIOCRUZ | Paola Resende et al |
| EPI_ISL_792650 | Laboratório Central de Saúde Pública do Estado do Paraná (LACEN-PR) | Laboratory of Respiratory Viruses and Measles, Oswaldo Cruz Institute, FIOCRUZ | Paola Resende et al |

| | | | |
|---|---|---|---|
| EPI_ISL_792651 | Laboratório Central de Saúde Pública do Estado do Paraná (LACEN-PR) | Laboratory of Respiratory Viruses and Measles, Oswaldo Cruz Institute, FIOCRUZ | Paola Resende et al |
| EPI_ISL_792652 | Laboratório Central de Saúde Pública do Estado do Paraná (LACEN-PR) | Laboratory of Respiratory Viruses and Measles, Oswaldo Cruz Institute, FIOCRUZ | Paola Resende et al |
| EPI_ISL_792653 | Laboratório Central de Saúde Pública do Estado do Paraná (LACEN-PR) | Laboratory of Respiratory Viruses and Measles, Oswaldo Cruz Institute, FIOCRUZ | Paola Resende et al |
| EPI_ISL_792654 | Laboratório Central de Saúde Pública do Estado do Paraná (LACEN-PR) | Laboratory of Respiratory Viruses and Measles, Oswaldo Cruz Institute, FIOCRUZ | Paola Resende et al |
| EPI_ISL_1181617 | Laboratorio Central de Saude Publica do Estado do Parana (LACEN-PR) | Laboratory of Respiratory Viruses and Measles, Oswaldo Cruz Institute, FIOCRUZ | Paola Resende et al |
| EPI_ISL_1181618 | Laboratorio Central de Saude Publica do Estado do Parana (LACEN-PR) | Laboratory of Respiratory Viruses and Measles, Oswaldo Cruz Institute, FIOCRUZ | Paola Resende et al |
| EPI_ISL_1181619 | Laboratorio Central de Saude Publica do Estado do Parana (LACEN-PR) | Laboratory of Respiratory Viruses and Measles, Oswaldo Cruz Institute, FIOCRUZ | Paola Resende et al |
| EPI_ISL_1181614 | Laboratorio Central de Saude Publica do Estado do Parana (LACEN-PR) | Laboratory of Respiratory Viruses and Measles, Oswaldo Cruz Institute, FIOCRUZ | Paola Resende et al |
| EPI_ISL_1181610 | Laboratorio Central de Saude Publica do Estado do Parana (LACEN-PR) | Laboratory of Respiratory Viruses and Measles, Oswaldo Cruz Institute, FIOCRUZ | Paola Resende et al |
| EPI_ISL_1181615 | Laboratorio Central de Saude Publica do Estado do Parana (LACEN-PR) | Laboratory of Respiratory Viruses and Measles, Oswaldo Cruz Institute, FIOCRUZ | Paola Resende et al |
| EPI_ISL_1181609 | Laboratorio Central de Saude Publica do Estado do Parana (LACEN-PR) | Laboratory of Respiratory Viruses and Measles, Oswaldo Cruz Institute, FIOCRUZ | Paola Resende et al |
| EPI_ISL_1181616 | Laboratorio Central de Saude Publica do Estado do Parana (LACEN-PR) | Laboratory of Respiratory Viruses and Measles, Oswaldo Cruz Institute, FIOCRUZ | Paola Resende et al |
| EPI_ISL_1219132 | Laboratorio Central de Saude Publica do Estado do Parana (LACEN-PR) | Laboratory of Respiratory Viruses and Measles, Oswaldo Cruz Institute, FIOCRUZ | Paola Resende et al |
| EPI_ISL_1181394 | Laboratorio Central de Saude Publica do Estado do Parana (LACEN-PR) | Laboratory of Respiratory Viruses and Measles, Oswaldo Cruz Institute, FIOCRUZ | Paola Resende et al |
| EPI_ISL_1219133 | Laboratorio Central de Saude Publica do Estado do Parana (LACEN-PR) | Laboratory of Respiratory Viruses and Measles, Oswaldo Cruz Institute, FIOCRUZ | Paola Resende et al |
| EPI_ISL_541340 | Laboratório Central de Saúde Pública do Estado do Paraná (LACEN-PR) | Laboratory of Respiratory Viruses and Measles, Oswaldo Cruz Institute, FIOCRUZ | Paola Resende et al |
| EPI_ISL_541341 | Laboratório Central de Saúde Pública do Estado do Paraná (LACEN-PR) | Laboratory of Respiratory Viruses and Measles, Oswaldo Cruz Institute, FIOCRUZ | Paola Resende et al |
| EPI_ISL_541342 | Laboratório Central de Saúde Pública do Estado do Paraná (LACEN-PR) | Laboratory of Respiratory Viruses and Measles, Oswaldo Cruz Institute, FIOCRUZ | Paola Resende et al |
| EPI_ISL_541343 | Laboratório Central de Saúde Pública do Estado do Paraná (LACEN-PR) | Laboratory of Respiratory Viruses and Measles, Oswaldo Cruz Institute, FIOCRUZ | Paola Resende et al |
| EPI_ISL_541344 | Laboratório Central de Saúde Pública do Estado do Paraná (LACEN-PR) | Laboratory of Respiratory Viruses and Measles, Oswaldo Cruz Institute, FIOCRUZ | Paola Resende et al |
| EPI_ISL_541345 | Laboratório Central de Saúde Pública do Estado do Paraná (LACEN-PR) | Laboratory of Respiratory Viruses and Measles, Oswaldo Cruz Institute, FIOCRUZ | Paola Resende et al |
| EPI_ISL_541346 | Laboratório Central de Saúde Pública do Estado do Paraná (LACEN-PR) | Laboratory of Respiratory Viruses and Measles, Oswaldo Cruz Institute, FIOCRUZ | Paola Resende et al |

| | | | |
|---|---|---|---|
| EPI_ISL_1181566 | Laboratorio Central de Saude Publica do Estado do Parana (LACEN-PR) | Laboratory of Respiratory Viruses and Measles, Oswaldo Cruz Institute, FIOCRUZ | Paola Resende et al |
| EPI_ISL_1181568 | Laboratorio Central de Saude Publica do Estado do Parana (LACEN-PR) | Laboratory of Respiratory Viruses and Measles, Oswaldo Cruz Institute, FIOCRUZ | Paola Resende et al |
| EPI_ISL_1181570 | Laboratorio Central de Saude Publica do Estado do Parana (LACEN-PR) | Laboratory of Respiratory Viruses and Measles, Oswaldo Cruz Institute, FIOCRUZ | Paola Resende et al |
| EPI_ISL_1181564 | Laboratorio Central de Saude Publica do Estado do Parana (LACEN-PR) | Laboratory of Respiratory Viruses and Measles, Oswaldo Cruz Institute, FIOCRUZ | Paola Resende et al |
| EPI_ISL_1181555 | Laboratorio Central de Saude Publica do Estado do Parana (LACEN-PR) | Laboratory of Respiratory Viruses and Measles, Oswaldo Cruz Institute, FIOCRUZ | Paola Resende et al |
| EPI_ISL_1181496 | Laboratory of Respiratory Viruses and Measles, Oswaldo Cruz Institute, FIOCRUZ | Laboratory of Respiratory Viruses and Measles, Oswaldo Cruz Institute, FIOCRUZ | Paola Resende et al |
| EPI_ISL_1181495 | Laboratory of Respiratory Viruses and Measles, Oswaldo Cruz Institute, FIOCRUZ | Laboratory of Respiratory Viruses and Measles, Oswaldo Cruz Institute, FIOCRUZ | Paola Resende et al |
| EPI_ISL_456076 | Laboratório Central de Saúde Pública Noel Nutels (LACEN-RJ) | Laboratory of Respiratory Viruses and Measles, Oswaldo Cruz Institute, FIOCRUZ | Paola Resende et al |
| EPI_ISL_1181489 | Laboratory of Respiratory Viruses and Measles, Oswaldo Cruz Institute, FIOCRUZ | Laboratory of Respiratory Viruses and Measles, Oswaldo Cruz Institute, FIOCRUZ | Paola Resende et al |
| EPI_ISL_1181361 | Laboratory of Respiratory Viruses and Measles, Oswaldo Cruz Institute, FIOCRUZ | Laboratory of Respiratory Viruses and Measles, Oswaldo Cruz Institute, FIOCRUZ | Paola Resende et al |
| EPI_ISL_1181488 | Laboratory of Respiratory Viruses and Measles, Oswaldo Cruz Institute, FIOCRUZ | Laboratory of Respiratory Viruses and Measles, Oswaldo Cruz Institute, FIOCRUZ | Paola Resende et al |
| EPI_ISL_1181486 | Laboratory of Respiratory Viruses and Measles, Oswaldo Cruz Institute, FIOCRUZ | Laboratory of Respiratory Viruses and Measles, Oswaldo Cruz Institute, FIOCRUZ | Paola Resende et al |
| EPI_ISL_1181483 | Laboratory of Respiratory Viruses and Measles, Oswaldo Cruz Institute, FIOCRUZ | Laboratory of Respiratory Viruses and Measles, Oswaldo Cruz Institute, FIOCRUZ | Paola Resende et al |
| EPI_ISL_1181388 | Laboratory of Respiratory Viruses and Measles, Oswaldo Cruz Institute, FIOCRUZ | Laboratory of Respiratory Viruses and Measles, Oswaldo Cruz Institute, FIOCRUZ | Paola Resende et al |
| EPI_ISL_1181613 | Laboratorio Central de Saude Publica do Estado do Rio de Janeiro (LACEN-RJ) | Laboratory of Respiratory Viruses and Measles, Oswaldo Cruz Institute, FIOCRUZ | Paola Resende et al |
| EPI_ISL_1181477 | Laboratory of Respiratory Viruses and Measles, Oswaldo Cruz Institute, FIOCRUZ | Laboratory of Respiratory Viruses and Measles, Oswaldo Cruz Institute, FIOCRUZ | Paola Resende et al |
| EPI_ISL_1181476 | Laboratory of Respiratory Viruses and Measles, Oswaldo Cruz Institute, FIOCRUZ | Laboratory of Respiratory Viruses and Measles, Oswaldo Cruz Institute, FIOCRUZ | Paola Resende et al |
| EPI_ISL_1181475 | Laboratory of Respiratory Viruses and Measles, Oswaldo Cruz Institute, FIOCRUZ | Laboratory of Respiratory Viruses and Measles, Oswaldo Cruz Institute, FIOCRUZ | Paola Resende et al |
| EPI_ISL_1181470 | Laboratory of Respiratory Viruses and Measles, Oswaldo Cruz Institute, FIOCRUZ | Laboratory of Respiratory Viruses and Measles, Oswaldo Cruz Institute, FIOCRUZ | Paola Resende et al |
| EPI_ISL_1534016 | Laboratory of Respiratory Viruses and Measles, Oswaldo Cruz Institute, FIOCRUZ | Laboratory of Respiratory Viruses and Measles, Oswaldo Cruz Institute, FIOCRUZ | Paola Resende et al |
| EPI_ISL_1181466 | Laboratory of Respiratory Viruses and Measles, Oswaldo Cruz Institute, FIOCRUZ | Laboratory of Respiratory Viruses and Measles, Oswaldo Cruz Institute, FIOCRUZ | Paola Resende et al |
| EPI_ISL_1181467 | Laboratory of Respiratory Viruses and Measles, Oswaldo Cruz Institute, FIOCRUZ | Laboratory of Respiratory Viruses and Measles, Oswaldo Cruz Institute, FIOCRUZ | Paola Resende et al |

| | | | |
|---|---|---|---|
| EPI_ISL_1181465 | Laboratory of Respiratory Viruses and Measles, Oswaldo Cruz Institute, FIOCRUZ | Laboratory of Respiratory Viruses and Measles, Oswaldo Cruz Institute, FIOCRUZ | Paola Resende et al |
| EPI_ISL_1533991 | Laboratorio de Virologia Molecular / UFRJ | Laboratory of Respiratory Viruses and Measles, Oswaldo Cruz Institute, FIOCRUZ | Paola Resende et al |
| EPI_ISL_1533995 | Laboratorio de Virologia Molecular / UFRJ | Laboratory of Respiratory Viruses and Measles, Oswaldo Cruz Institute, FIOCRUZ | Paola Resende et al |
| EPI_ISL_1533998 | Laboratorio de Virologia Molecular / UFRJ | Laboratory of Respiratory Viruses and Measles, Oswaldo Cruz Institute, FIOCRUZ | Paola Resende et al |
| EPI_ISL_1181366 | Laboratory of Respiratory Viruses and Measles, Oswaldo Cruz Institute, FIOCRUZ | Laboratory of Respiratory Viruses and Measles, Oswaldo Cruz Institute, FIOCRUZ | Paola Resende et al |
| EPI_ISL_1181464 | Laboratory of Respiratory Viruses and Measles, Oswaldo Cruz Institute, FIOCRUZ | Laboratory of Respiratory Viruses and Measles, Oswaldo Cruz Institute, FIOCRUZ | Paola Resende et al |
| EPI_ISL_1181463 | Laboratory of Respiratory Viruses and Measles, Oswaldo Cruz Institute, FIOCRUZ | Laboratory of Respiratory Viruses and Measles, Oswaldo Cruz Institute, FIOCRUZ | Paola Resende et al |
| EPI_ISL_1181374 | Laboratory of Respiratory Viruses and Measles, Oswaldo Cruz Institute, FIOCRUZ | Laboratory of Respiratory Viruses and Measles, Oswaldo Cruz Institute, FIOCRUZ | Paola Resende et al |
| EPI_ISL_1181461 | Laboratory of Respiratory Viruses and Measles, Oswaldo Cruz Institute, FIOCRUZ | Laboratory of Respiratory Viruses and Measles, Oswaldo Cruz Institute, FIOCRUZ | Paola Resende et al |
| EPI_ISL_1181457 | Laboratory of Respiratory Viruses and Measles, Oswaldo Cruz Institute, FIOCRUZ | Laboratory of Respiratory Viruses and Measles, Oswaldo Cruz Institute, FIOCRUZ | Paola Resende et al |
| EPI_ISL_1181458 | Laboratory of Respiratory Viruses and Measles, Oswaldo Cruz Institute, FIOCRUZ | Laboratory of Respiratory Viruses and Measles, Oswaldo Cruz Institute, FIOCRUZ | Paola Resende et al |
| EPI_ISL_1181462 | Laboratory of Respiratory Viruses and Measles, Oswaldo Cruz Institute, FIOCRUZ | Laboratory of Respiratory Viruses and Measles, Oswaldo Cruz Institute, FIOCRUZ | Paola Resende et al |
| EPI_ISL_1181367 | Laboratory of Respiratory Viruses and Measles, Oswaldo Cruz Institute, FIOCRUZ | Laboratory of Respiratory Viruses and Measles, Oswaldo Cruz Institute, FIOCRUZ | Paola Resende et al |
| EPI_ISL_1181377 | Laboratory of Respiratory Viruses and Measles, Oswaldo Cruz Institute, FIOCRUZ | Laboratory of Respiratory Viruses and Measles, Oswaldo Cruz Institute, FIOCRUZ | Paola Resende et al |
| EPI_ISL_1181455 | Laboratory of Respiratory Viruses and Measles, Oswaldo Cruz Institute, FIOCRUZ | Laboratory of Respiratory Viruses and Measles, Oswaldo Cruz Institute, FIOCRUZ | Paola Resende et al |
| EPI_ISL_1181360 | Laboratory of Respiratory Viruses and Measles, Oswaldo Cruz Institute, FIOCRUZ | Laboratory of Respiratory Viruses and Measles, Oswaldo Cruz Institute, FIOCRUZ | Paola Resende et al |
| EPI_ISL_1181456 | Laboratory of Respiratory Viruses and Measles, Oswaldo Cruz Institute, FIOCRUZ | Laboratory of Respiratory Viruses and Measles, Oswaldo Cruz Institute, FIOCRUZ | Paola Resende et al |
| EPI_ISL_1181454 | Laboratory of Respiratory Viruses and Measles, Oswaldo Cruz Institute, FIOCRUZ | Laboratory of Respiratory Viruses and Measles, Oswaldo Cruz Institute, FIOCRUZ | Paola Resende et al |
| EPI_ISL_1181382 | Laboratory of Respiratory Viruses and Measles, Oswaldo Cruz Institute, FIOCRUZ | Laboratory of Respiratory Viruses and Measles, Oswaldo Cruz Institute, FIOCRUZ | Paola Resende et al |
| EPI_ISL_1181448 | Laboratory of Respiratory Viruses and Measles, Oswaldo Cruz Institute, FIOCRUZ | Laboratory of Respiratory Viruses and Measles, Oswaldo Cruz Institute, FIOCRUZ | Paola Resende et al |
| EPI_ISL_1181446 | Laboratory of Respiratory Viruses and Measles, Oswaldo Cruz Institute, FIOCRUZ | Laboratory of Respiratory Viruses and Measles, Oswaldo Cruz Institute, FIOCRUZ | Paola Resende et al |
| EPI_ISL_1181447 | Laboratory of Respiratory Viruses and Measles, Oswaldo Cruz Institute, FIOCRUZ | Laboratory of Respiratory Viruses and Measles, Oswaldo Cruz Institute, FIOCRUZ | Paola Resende et al |

| | | | |
|---|---|---|---|
| EPI_ISL_1181381 | Laboratory of Respiratory Viruses and Measles, Oswaldo Cruz Institute, FIOCRUZ | Laboratory of Respiratory Viruses and Measles, Oswaldo Cruz Institute, FIOCRUZ | Paola Resende et al |
| EPI_ISL_1181383 | Laboratory of Respiratory Viruses and Measles, Oswaldo Cruz Institute, FIOCRUZ | Laboratory of Respiratory Viruses and Measles, Oswaldo Cruz Institute, FIOCRUZ | Paola Resende et al |
| EPI_ISL_1181442 | Laboratory of Respiratory Viruses and Measles, Oswaldo Cruz Institute, FIOCRUZ | Laboratory of Respiratory Viruses and Measles, Oswaldo Cruz Institute, FIOCRUZ | Paola Resende et al |
| EPI_ISL_456088 | Laboratório Central de Saúde Pública Noel Nutels (LACEN-RJ) | Laboratory of Respiratory Viruses and Measles, Oswaldo Cruz Institute, FIOCRUZ | Paola Resende et al |
| EPI_ISL_1181437 | Laboratory of Respiratory Viruses and Measles, Oswaldo Cruz Institute, FIOCRUZ | Laboratory of Respiratory Viruses and Measles, Oswaldo Cruz Institute, FIOCRUZ | Paola Resende et al |
| EPI_ISL_1181438 | Laboratory of Respiratory Viruses and Measles, Oswaldo Cruz Institute, FIOCRUZ | Laboratory of Respiratory Viruses and Measles, Oswaldo Cruz Institute, FIOCRUZ | Paola Resende et al |
| EPI_ISL_1181439 | Laboratory of Respiratory Viruses and Measles, Oswaldo Cruz Institute, FIOCRUZ | Laboratory of Respiratory Viruses and Measles, Oswaldo Cruz Institute, FIOCRUZ | Paola Resende et al |
| EPI_ISL_456102 | Laboratory of Respiratory Viruses and Measles, Oswaldo Cruz Institute, FIOCRUZ | Laboratory of Respiratory Viruses and Measles, Oswaldo Cruz Institute, FIOCRUZ | Paola Resende et al |
| EPI_ISL_541354 | Laboratory of Respiratory Viruses and Measles, Oswaldo Cruz Institute, FIOCRUZ | Laboratory of Respiratory Viruses and Measles, Oswaldo Cruz Institute, FIOCRUZ | Paola Resende et al |
| EPI_ISL_541355 | Laboratory of Respiratory Viruses and Measles, Oswaldo Cruz Institute, FIOCRUZ | Laboratory of Respiratory Viruses and Measles, Oswaldo Cruz Institute, FIOCRUZ | Paola Resende et al |
| EPI_ISL_1181353 | Laboratorio Central de Saude Publica do Estado do Rio de Janeiro (LACEN-RJ) | Laboratory of Respiratory Viruses and Measles, Oswaldo Cruz Institute, FIOCRUZ | Paola Resende et al |
| EPI_ISL_1181372 | Laboratorio Central de Saude Publica do Estado do Rio de Janeiro (LACEN-RJ) | Laboratory of Respiratory Viruses and Measles, Oswaldo Cruz Institute, FIOCRUZ | Paola Resende et al |
| EPI_ISL_1181431 | Laboratorio Central de Saude Publica do Estado do Rio de Janeiro (LACEN-RJ) | Laboratory of Respiratory Viruses and Measles, Oswaldo Cruz Institute, FIOCRUZ | Paola Resende et al |
| EPI_ISL_541359 | Laboratory of Respiratory Viruses and Measles, Oswaldo Cruz Institute, FIOCRUZ | Laboratory of Respiratory Viruses and Measles, Oswaldo Cruz Institute, FIOCRUZ | Paola Resende et al |
| EPI_ISL_1181398 | Laboratory of Respiratory Viruses and Measles, Oswaldo Cruz Institute, FIOCRUZ | Laboratory of Respiratory Viruses and Measles, Oswaldo Cruz Institute, FIOCRUZ | Paola Resende et al |
| EPI_ISL_467356 | Laboratory of Respiratory Viruses and Measles, Oswaldo Cruz Institute, FIOCRUZ | Laboratory of Respiratory Viruses and Measles, Oswaldo Cruz Institute, FIOCRUZ | Paola Resende et al |
| EPI_ISL_467359 | Laboratory of Respiratory Viruses and Measles, Oswaldo Cruz Institute, FIOCRUZ | Laboratory of Respiratory Viruses and Measles, Oswaldo Cruz Institute, FIOCRUZ | Paola Resende et al |
| EPI_ISL_467366 | Laboratory of Respiratory Viruses and Measles, Oswaldo Cruz Institute, FIOCRUZ | Laboratory of Respiratory Viruses and Measles, Oswaldo Cruz Institute, FIOCRUZ | Paola Resende et al |
| EPI_ISL_1181425 | Laboratorio Central de Saude Publica do Estado do Rio de Janeiro (LACEN-RJ) | Laboratory of Respiratory Viruses and Measles, Oswaldo Cruz Institute, FIOCRUZ | Paola Resende et al |
| EPI_ISL_1181410 | Laboratory of Respiratory Viruses and Measles, Oswaldo Cruz Institute, FIOCRUZ | Laboratory of Respiratory Viruses and Measles, Oswaldo Cruz Institute, FIOCRUZ | Paola Resende et al |
| EPI_ISL_1181415 | Laboratory of Respiratory Viruses and Measles, Oswaldo Cruz Institute, FIOCRUZ | Laboratory of Respiratory Viruses and Measles, Oswaldo Cruz Institute, FIOCRUZ | Paola Resende et al |
| EPI_ISL_1181578 | Laboratory of Respiratory Viruses and Measles, Oswaldo Cruz Institute, FIOCRUZ | Laboratory of Respiratory Viruses and Measles, Oswaldo Cruz Institute, FIOCRUZ | Paola Resende et al |

| | | | |
|---|---|---|---|
| EPI_ISL_1181384 | Laboratory of Respiratory Viruses and Measles, Oswaldo Cruz Institute, FIOCRUZ | Laboratory of Respiratory Viruses and Measles, Oswaldo Cruz Institute, FIOCRUZ | Paola Resende et al |
| EPI_ISL_1181574 | Laboratory of Respiratory Viruses and Measles, Oswaldo Cruz Institute, FIOCRUZ | Laboratory of Respiratory Viruses and Measles, Oswaldo Cruz Institute, FIOCRUZ | Paola Resende et al |
| EPI_ISL_1181359 | Laboratory of Respiratory Viruses and Measles, Oswaldo Cruz Institute, FIOCRUZ | Laboratory of Respiratory Viruses and Measles, Oswaldo Cruz Institute, FIOCRUZ | Paola Resende et al |
| EPI_ISL_1181531 | Laboratory of Respiratory Viruses and Measles, Oswaldo Cruz Institute, FIOCRUZ | Laboratory of Respiratory Viruses and Measles, Oswaldo Cruz Institute, FIOCRUZ | Paola Resende et al |
| EPI_ISL_1181529 | Laboratory of Respiratory Viruses and Measles, Oswaldo Cruz Institute, FIOCRUZ | Laboratory of Respiratory Viruses and Measles, Oswaldo Cruz Institute, FIOCRUZ | Paola Resende et al |
| EPI_ISL_1181523 | Laboratory of Respiratory Viruses and Measles, Oswaldo Cruz Institute, FIOCRUZ | Laboratory of Respiratory Viruses and Measles, Oswaldo Cruz Institute, FIOCRUZ | Paola Resende et al |
| EPI_ISL_1181369 | Laboratory of Respiratory Viruses and Measles, Oswaldo Cruz Institute, FIOCRUZ | Laboratory of Respiratory Viruses and Measles, Oswaldo Cruz Institute, FIOCRUZ | Paola Resende et al |
| EPI_ISL_1181521 | Laboratory of Respiratory Viruses and Measles, Oswaldo Cruz Institute, FIOCRUZ | Laboratory of Respiratory Viruses and Measles, Oswaldo Cruz Institute, FIOCRUZ | Paola Resende et al |
| EPI_ISL_1181519 | Laboratory of Respiratory Viruses and Measles, Oswaldo Cruz Institute, FIOCRUZ | Laboratory of Respiratory Viruses and Measles, Oswaldo Cruz Institute, FIOCRUZ | Paola Resende et al |
| EPI_ISL_1181357 | Laboratory of Respiratory Viruses and Measles, Oswaldo Cruz Institute, FIOCRUZ | Laboratory of Respiratory Viruses and Measles, Oswaldo Cruz Institute, FIOCRUZ | Paola Resende et al |
| EPI_ISL_1181514 | Laboratory of Respiratory Viruses and Measles, Oswaldo Cruz Institute, FIOCRUZ | Laboratory of Respiratory Viruses and Measles, Oswaldo Cruz Institute, FIOCRUZ | Paola Resende et al |
| EPI_ISL_1181515 | Laboratory of Respiratory Viruses and Measles, Oswaldo Cruz Institute, FIOCRUZ | Laboratory of Respiratory Viruses and Measles, Oswaldo Cruz Institute, FIOCRUZ | Paola Resende et al |
| EPI_ISL_1181511 | Laboratory of Respiratory Viruses and Measles, Oswaldo Cruz Institute, FIOCRUZ | Laboratory of Respiratory Viruses and Measles, Oswaldo Cruz Institute, FIOCRUZ | Paola Resende et al |
| EPI_ISL_427302 | Laboratory of Respiratory Viruses and Measles, Oswaldo Cruz Institute, FIOCRUZ | Laboratory of Respiratory Viruses and Measles, Oswaldo Cruz Institute, FIOCRUZ | Paola Resende et al |
| EPI_ISL_1534005 | Laboratory of Respiratory Viruses and Measles, Oswaldo Cruz Institute, FIOCRUZ | Laboratory of Respiratory Viruses and Measles, Oswaldo Cruz Institute, FIOCRUZ | Paola Resende et al |
| EPI_ISL_427303 | Laboratory of Respiratory Viruses and Measles, Oswaldo Cruz Institute, FIOCRUZ | Laboratory of Respiratory Viruses and Measles, Oswaldo Cruz Institute, FIOCRUZ | Paola Resende et al |
| EPI_ISL_427304 | Laboratory of Respiratory Viruses and Measles, Oswaldo Cruz Institute, FIOCRUZ | Laboratory of Respiratory Viruses and Measles, Oswaldo Cruz Institute, FIOCRUZ | Paola Resende et al |
| EPI_ISL_456071 | Laboratory of Respiratory Viruses and Measles, Oswaldo Cruz Institute, FIOCRUZ | Laboratory of Respiratory Viruses and Measles, Oswaldo Cruz Institute, FIOCRUZ | Paola Resende et al |
| EPI_ISL_1181608 | Laboratory of Respiratory Viruses and Measles, Oswaldo Cruz Institute, FIOCRUZ | Laboratory of Respiratory Viruses and Measles, Oswaldo Cruz Institute, FIOCRUZ | Paola Resende et al |
| EPI_ISL_1181499 | Laboratory of Respiratory Viruses and Measles, Oswaldo Cruz Institute, FIOCRUZ | Laboratory of Respiratory Viruses and Measles, Oswaldo Cruz Institute, FIOCRUZ | Paola Resende et al |
| EPI_ISL_1534013 | Laboratorio Central de Saude Publica do Estado do Rio Grande do Sul (LACEN-RS) | Laboratory of Respiratory Viruses and Measles, Oswaldo Cruz Institute, FIOCRUZ | Paola Resende et al |
| EPI_ISL_729801 | Laboratório Central de Saúde Pública do Estado do Rio Grande do Sul (LACEN-RS) | Laboratory of Respiratory Viruses and Measles, Oswaldo Cruz Institute, FIOCRUZ | Paola Resende et al |

| | | | |
|---|---|---|---|
| EPI_ISL_729844 | Laboratório Central de Saúde Pública do Estado do Rio Grande do Sul (LACEN-RS) | Laboratory of Respiratory Viruses and Measles, Oswaldo Cruz Institute, FIOCRUZ | Paola Resende et al |
| EPI_ISL_729850 | Laboratório Central de Saúde Pública do Estado do Rio Grande do Sul (LACEN-RS) | Laboratory of Respiratory Viruses and Measles, Oswaldo Cruz Institute, FIOCRUZ | Paola Resende et al |
| EPI_ISL_729856 | Laboratório Central de Saúde Pública do Estado do Rio Grande do Sul (LACEN-RS) | Laboratory of Respiratory Viruses and Measles, Oswaldo Cruz Institute, FIOCRUZ | Paola Resende et al |
| EPI_ISL_729857 | Laboratório Central de Saúde Pública do Estado do Rio Grande do Sul (LACEN-RS) | Laboratory of Respiratory Viruses and Measles, Oswaldo Cruz Institute, FIOCRUZ | Paola Resende et al |
| EPI_ISL_729848 | Laboratório Central de Saúde Pública do Estado do Rio Grande do Sul (LACEN-RS) | Laboratory of Respiratory Viruses and Measles, Oswaldo Cruz Institute, FIOCRUZ | Paola Resende et al |
| EPI_ISL_729845 | Laboratório Central de Saúde Pública do Estado do Rio Grande do Sul (LACEN-RS) | Laboratory of Respiratory Viruses and Measles, Oswaldo Cruz Institute, FIOCRUZ | Paola Resende et al |
| EPI_ISL_729851 | Laboratório Central de Saúde Pública do Estado do Rio Grande do Sul (LACEN-RS) | Laboratory of Respiratory Viruses and Measles, Oswaldo Cruz Institute, FIOCRUZ | Paola Resende et al |
| EPI_ISL_729852 | Laboratório Central de Saúde Pública do Estado do Rio Grande do Sul (LACEN-RS) | Laboratory of Respiratory Viruses and Measles, Oswaldo Cruz Institute, FIOCRUZ | Paola Resende et al |
| EPI_ISL_729858 | Laboratório Central de Saúde Pública do Estado do Rio Grande do Sul (LACEN-RS) | Laboratory of Respiratory Viruses and Measles, Oswaldo Cruz Institute, FIOCRUZ | Paola Resende et al |
| EPI_ISL_729802 | Laboratório Central de Saúde Pública do Estado do Rio Grande do Sul (LACEN-RS) | Laboratory of Respiratory Viruses and Measles, Oswaldo Cruz Institute, FIOCRUZ | Paola Resende et al |
| EPI_ISL_729853 | Laboratório Central de Saúde Pública do Estado do Rio Grande do Sul (LACEN-RS) | Laboratory of Respiratory Viruses and Measles, Oswaldo Cruz Institute, FIOCRUZ | Paola Resende et al |
| EPI_ISL_729859 | Laboratório Central de Saúde Pública do Estado do Rio Grande do Sul (LACEN-RS) | Laboratory of Respiratory Viruses and Measles, Oswaldo Cruz Institute, FIOCRUZ | Paola Resende et al |
| EPI_ISL_729803 | Laboratório Central de Saúde Pública do Estado do Rio Grande do Sul (LACEN-RS) | Laboratory of Respiratory Viruses and Measles, Oswaldo Cruz Institute, FIOCRUZ | Paola Resende et al |
| EPI_ISL_729860 | Laboratório Central de Saúde Pública do Estado do Rio Grande do Sul (LACEN-RS) | Laboratory of Respiratory Viruses and Measles, Oswaldo Cruz Institute, FIOCRUZ | Paola Resende et al |
| EPI_ISL_729849 | Laboratório Central de Saúde Pública do Estado do Rio Grande do Sul (LACEN-RS) | Laboratory of Respiratory Viruses and Measles, Oswaldo Cruz Institute, FIOCRUZ | Paola Resende et al |
| EPI_ISL_729854 | Laboratório Central de Saúde Pública do Estado do Rio Grande do Sul (LACEN-RS) | Laboratory of Respiratory Viruses and Measles, Oswaldo Cruz Institute, FIOCRUZ | Paola Resende et al |
| EPI_ISL_729799 | Laboratório Central de Saúde Pública do Estado do Rio Grande do Sul (LACEN-RS) | Laboratory of Respiratory Viruses and Measles, Oswaldo Cruz Institute, FIOCRUZ | Paola Resende et al |
| EPI_ISL_729855 | Laboratório Central de Saúde Pública do Estado do Rio Grande do Sul (LACEN-RS) | Laboratory of Respiratory Viruses and Measles, Oswaldo Cruz Institute, FIOCRUZ | Paola Resende et al |
| EPI_ISL_729861 | Laboratório Central de Saúde Pública do Estado do Rio Grande do Sul (LACEN-RS) | Laboratory of Respiratory Viruses and Measles, Oswaldo Cruz Institute, FIOCRUZ | Paola Resende et al |
| EPI_ISL_729804 | Laboratório Central de Saúde Pública do Estado do Rio Grande do Sul (LACEN-RS) | Laboratory of Respiratory Viruses and Measles, Oswaldo Cruz Institute, FIOCRUZ | Paola Resende et al |
| EPI_ISL_729805 | Laboratório Central de Saúde Pública do Estado do Rio Grande do Sul (LACEN-RS) | Laboratory of Respiratory Viruses and Measles, Oswaldo Cruz Institute, FIOCRUZ | Paola Resende et al |
| EPI_ISL_729806 | Laboratório Central de Saúde Pública do Estado do Rio Grande do Sul (LACEN-RS) | Laboratory of Respiratory Viruses and Measles, Oswaldo Cruz Institute, FIOCRUZ | Paola Resende et al |

| | | | |
|---|---|---|---|
| EPI_ISL_729807 | Laboratório Central de Saúde Pública do Estado do Rio Grande do Sul (LACEN-RS) | Laboratory of Respiratory Viruses and Measles, Oswaldo Cruz Institute, FIOCRUZ | Paola Resende et al |
| EPI_ISL_729808 | Laboratório Central de Saúde Pública do Estado do Rio Grande do Sul (LACEN-RS) | Laboratory of Respiratory Viruses and Measles, Oswaldo Cruz Institute, FIOCRUZ | Paola Resende et al |
| EPI_ISL_729809 | Laboratório Central de Saúde Pública do Estado do Rio Grande do Sul (LACEN-RS) | Laboratory of Respiratory Viruses and Measles, Oswaldo Cruz Institute, FIOCRUZ | Paola Resende et al |
| EPI_ISL_1181468 | Laboratorio Central de Saude Publica do Estado do Rio Grande do Sul (LACEN-RS) | Laboratory of Respiratory Viruses and Measles, Oswaldo Cruz Institute, FIOCRUZ | Paola Resende et al |
| EPI_ISL_729810 | Laboratório Central de Saúde Pública do Estado do Rio Grande do Sul (LACEN-RS) | Laboratory of Respiratory Viruses and Measles, Oswaldo Cruz Institute, FIOCRUZ | Paola Resende et al |
| EPI_ISL_1181451 | Laboratorio Central de Saude Publica do Estado do Rio Grande do Sul (LACEN-RS) | Laboratory of Respiratory Viruses and Measles, Oswaldo Cruz Institute, FIOCRUZ | Paola Resende et al |
| EPI_ISL_1181434 | Laboratorio Central de Saude Publica do Estado do Rio Grande do Sul (LACEN-RS) | Laboratory of Respiratory Viruses and Measles, Oswaldo Cruz Institute, FIOCRUZ | Paola Resende et al |
| EPI_ISL_1181433 | Laboratorio Central de Saude Publica do Estado do Rio Grande do Sul (LACEN-RS) | Laboratory of Respiratory Viruses and Measles, Oswaldo Cruz Institute, FIOCRUZ | Paola Resende et al |
| EPI_ISL_1181620 | Laboratorio Central de Saude Publica do Estado do Rio Grande do Sul (LACEN-RS) | Laboratory of Respiratory Viruses and Measles, Oswaldo Cruz Institute, FIOCRUZ | Paola Resende et al |
| EPI_ISL_729812 | Laboratório Central de Saúde Pública do Estado do Rio Grande do Sul (LACEN-RS) | Laboratory of Respiratory Viruses and Measles, Oswaldo Cruz Institute, FIOCRUZ | Paola Resende et al |
| EPI_ISL_729813 | Laboratório Central de Saúde Pública do Estado do Rio Grande do Sul (LACEN-RS) | Laboratory of Respiratory Viruses and Measles, Oswaldo Cruz Institute, FIOCRUZ | Paola Resende et al |
| EPI_ISL_729814 | Laboratório Central de Saúde Pública do Estado do Rio Grande do Sul (LACEN-RS) | Laboratory of Respiratory Viruses and Measles, Oswaldo Cruz Institute, FIOCRUZ | Paola Resende et al |
| EPI_ISL_729815 | Laboratório Central de Saúde Pública do Estado do Rio Grande do Sul (LACEN-RS) | Laboratory of Respiratory Viruses and Measles, Oswaldo Cruz Institute, FIOCRUZ | Paola Resende et al |
| EPI_ISL_729816 | Laboratório Central de Saúde Pública do Estado do Rio Grande do Sul (LACEN-RS) | Laboratory of Respiratory Viruses and Measles, Oswaldo Cruz Institute, FIOCRUZ | Paola Resende et al |
| EPI_ISL_729817 | Laboratório Central de Saúde Pública do Estado do Rio Grande do Sul (LACEN-RS) | Laboratory of Respiratory Viruses and Measles, Oswaldo Cruz Institute, FIOCRUZ | Paola Resende et al |
| EPI_ISL_729818 | Laboratório Central de Saúde Pública do Estado do Rio Grande do Sul (LACEN-RS) | Laboratory of Respiratory Viruses and Measles, Oswaldo Cruz Institute, FIOCRUZ | Paola Resende et al |
| EPI_ISL_1533979 | Laboratorio Central de Saude Publica do Estado do Rio Grande do Sul (LACEN-RS) | Laboratory of Respiratory Viruses and Measles, Oswaldo Cruz Institute, FIOCRUZ | Paola Resende et al |
| EPI_ISL_1181524 | Laboratorio Central de Saude Publica do Estado do Rio Grande do Sul (LACEN-RS) | Laboratory of Respiratory Viruses and Measles, Oswaldo Cruz Institute, FIOCRUZ | Paola Resende et al |
| EPI_ISL_1181368 | Laboratorio Central de Saude Publica do Estado do Rio Grande do Sul (LACEN-RS) | Laboratory of Respiratory Viruses and Measles, Oswaldo Cruz Institute, FIOCRUZ | Paola Resende et al |
| EPI_ISL_1181411 | Laboratorio Central de Saude Publica do Estado do Rio Grande do Sul (LACEN-RS) | Laboratory of Respiratory Viruses and Measles, Oswaldo Cruz Institute, FIOCRUZ | Paola Resende et al |
| EPI_ISL_729819 | Laboratório Central de Saúde Pública do Estado do Rio Grande do Sul (LACEN-RS) | Laboratory of Respiratory Viruses and Measles, Oswaldo Cruz Institute, FIOCRUZ | Paola Resende et al |
| EPI_ISL_729820 | Laboratório Central de Saúde Pública do Estado do Rio Grande do Sul (LACEN-RS) | Laboratory of Respiratory Viruses and Measles, Oswaldo Cruz Institute, FIOCRUZ | Paola Resende et al |

| | | | |
|---|---|---|---|
| EPI_ISL_729821 | Laboratório Central de Saúde Pública do Estado do Rio Grande do Sul (LACEN-RS) | Laboratory of Respiratory Viruses and Measles, Oswaldo Cruz Institute, FIOCRUZ | Paola Resende et al |
| EPI_ISL_729797 | Laboratório Central de Saúde Pública do Estado do Rio Grande do Sul (LACEN-RS) | Laboratory of Respiratory Viruses and Measles, Oswaldo Cruz Institute, FIOCRUZ | Paola Resende et al |
| EPI_ISL_729822 | Laboratório Central de Saúde Pública do Estado do Rio Grande do Sul (LACEN-RS) | Laboratory of Respiratory Viruses and Measles, Oswaldo Cruz Institute, FIOCRUZ | Paola Resende et al |
| EPI_ISL_729823 | Laboratório Central de Saúde Pública do Estado do Rio Grande do Sul (LACEN-RS) | Laboratory of Respiratory Viruses and Measles, Oswaldo Cruz Institute, FIOCRUZ | Paola Resende et al |
| EPI_ISL_729798 | Laboratório Central de Saúde Pública do Estado do Rio Grande do Sul (LACEN-RS) | Laboratory of Respiratory Viruses and Measles, Oswaldo Cruz Institute, FIOCRUZ | Paola Resende et al |
| EPI_ISL_729824 | Laboratório Central de Saúde Pública do Estado do Rio Grande do Sul (LACEN-RS) | Laboratory of Respiratory Viruses and Measles, Oswaldo Cruz Institute, FIOCRUZ | Paola Resende et al |
| EPI_ISL_729825 | Laboratório Central de Saúde Pública do Estado do Rio Grande do Sul (LACEN-RS) | Laboratory of Respiratory Viruses and Measles, Oswaldo Cruz Institute, FIOCRUZ | Paola Resende et al |
| EPI_ISL_729826 | Laboratório Central de Saúde Pública do Estado do Rio Grande do Sul (LACEN-RS) | Laboratory of Respiratory Viruses and Measles, Oswaldo Cruz Institute, FIOCRUZ | Paola Resende et al |
| EPI_ISL_729827 | Laboratório Central de Saúde Pública do Estado do Rio Grande do Sul (LACEN-RS) | Laboratory of Respiratory Viruses and Measles, Oswaldo Cruz Institute, FIOCRUZ | Paola Resende et al |
| EPI_ISL_729794 | Laboratório Central de Saúde Pública do Estado do Rio Grande do Sul (LACEN-RS) | Laboratory of Respiratory Viruses and Measles, Oswaldo Cruz Institute, FIOCRUZ | Paola Resende et al |
| EPI_ISL_729828 | Laboratório Central de Saúde Pública do Estado do Rio Grande do Sul (LACEN-RS) | Laboratory of Respiratory Viruses and Measles, Oswaldo Cruz Institute, FIOCRUZ | Paola Resende et al |
| EPI_ISL_729829 | Laboratório Central de Saúde Pública do Estado do Rio Grande do Sul (LACEN-RS) | Laboratory of Respiratory Viruses and Measles, Oswaldo Cruz Institute, FIOCRUZ | Paola Resende et al |
| EPI_ISL_729830 | Laboratório Central de Saúde Pública do Estado do Rio Grande do Sul (LACEN-RS) | Laboratory of Respiratory Viruses and Measles, Oswaldo Cruz Institute, FIOCRUZ | Paola Resende et al |
| EPI_ISL_729831 | Laboratório Central de Saúde Pública do Estado do Rio Grande do Sul (LACEN-RS) | Laboratory of Respiratory Viruses and Measles, Oswaldo Cruz Institute, FIOCRUZ | Paola Resende et al |
| EPI_ISL_729832 | Laboratório Central de Saúde Pública do Estado do Rio Grande do Sul (LACEN-RS) | Laboratory of Respiratory Viruses and Measles, Oswaldo Cruz Institute, FIOCRUZ | Paola Resende et al |
| EPI_ISL_729833 | Laboratório Central de Saúde Pública do Estado do Rio Grande do Sul (LACEN-RS) | Laboratory of Respiratory Viruses and Measles, Oswaldo Cruz Institute, FIOCRUZ | Paola Resende et al |
| EPI_ISL_729834 | Laboratório Central de Saúde Pública do Estado do Rio Grande do Sul (LACEN-RS) | Laboratory of Respiratory Viruses and Measles, Oswaldo Cruz Institute, FIOCRUZ | Paola Resende et al |
| EPI_ISL_729835 | Laboratório Central de Saúde Pública do Estado do Rio Grande do Sul (LACEN-RS) | Laboratory of Respiratory Viruses and Measles, Oswaldo Cruz Institute, FIOCRUZ | Paola Resende et al |
| EPI_ISL_729836 | Laboratório Central de Saúde Pública do Estado do Rio Grande do Sul (LACEN-RS) | Laboratory of Respiratory Viruses and Measles, Oswaldo Cruz Institute, FIOCRUZ | Paola Resende et al |
| EPI_ISL_729795 | Laboratório Central de Saúde Pública do Estado do Rio Grande do Sul (LACEN-RS) | Laboratory of Respiratory Viruses and Measles, Oswaldo Cruz Institute, FIOCRUZ | Paola Resende et al |
| EPI_ISL_729796 | Laboratório Central de Saúde Pública do Estado do Rio Grande do Sul (LACEN-RS) | Laboratory of Respiratory Viruses and Measles, Oswaldo Cruz Institute, FIOCRUZ | Paola Resende et al |
| EPI_ISL_729837 | Laboratório Central de Saúde Pública do Estado do Rio Grande do Sul (LACEN-RS) | Laboratory of Respiratory Viruses and Measles, Oswaldo Cruz Institute, FIOCRUZ | Paola Resende et al |

| | | | |
|---|---|---|---|
| EPI_ISL_729846 | Laboratório Central de Saúde Pública do Estado do Rio Grande do Sul (LACEN-RS) | Laboratory of Respiratory Viruses and Measles, Oswaldo Cruz Institute, FIOCRUZ | Paola Resende et al |
| EPI_ISL_729838 | Laboratório Central de Saúde Pública do Estado do Rio Grande do Sul (LACEN-RS) | Laboratory of Respiratory Viruses and Measles, Oswaldo Cruz Institute, FIOCRUZ | Paola Resende et al |
| EPI_ISL_729800 | Laboratório Central de Saúde Pública do Estado do Rio Grande do Sul (LACEN-RS) | Laboratory of Respiratory Viruses and Measles, Oswaldo Cruz Institute, FIOCRUZ | Paola Resende et al |
| EPI_ISL_729839 | Laboratório Central de Saúde Pública do Estado do Rio Grande do Sul (LACEN-RS) | Laboratory of Respiratory Viruses and Measles, Oswaldo Cruz Institute, FIOCRUZ | Paola Resende et al |
| EPI_ISL_729840 | Laboratório Central de Saúde Pública do Estado do Rio Grande do Sul (LACEN-RS) | Laboratory of Respiratory Viruses and Measles, Oswaldo Cruz Institute, FIOCRUZ | Paola Resende et al |
| EPI_ISL_1181567 | Laboratorio Central de Saude Publica do Estado do Rio Grande do Sul (LACEN-RS) | Laboratory of Respiratory Viruses and Measles, Oswaldo Cruz Institute, FIOCRUZ | Paola Resende et al |
| EPI_ISL_729841 | Laboratório Central de Saúde Pública do Estado do Rio Grande do Sul (LACEN-RS) | Laboratory of Respiratory Viruses and Measles, Oswaldo Cruz Institute, FIOCRUZ | Paola Resende et al |
| EPI_ISL_729842 | Laboratório Central de Saúde Pública do Estado do Rio Grande do Sul (LACEN-RS) | Laboratory of Respiratory Viruses and Measles, Oswaldo Cruz Institute, FIOCRUZ | Paola Resende et al |
| EPI_ISL_729843 | Laboratório Central de Saúde Pública do Estado do Rio Grande do Sul (LACEN-RS) | Laboratory of Respiratory Viruses and Measles, Oswaldo Cruz Institute, FIOCRUZ | Paola Resende et al |
| EPI_ISL_729847 | Laboratório Central de Saúde Pública do Estado do Rio Grande do Sul (LACEN-RS) | Laboratory of Respiratory Viruses and Measles, Oswaldo Cruz Institute, FIOCRUZ | Paola Resende et al |
| EPI_ISL_1181403 | Laboratorio Central de Saude Publica do Estado de Santa Catarina (LACEN-SC) | Laboratory of Respiratory Viruses and Measles, Oswaldo Cruz Institute, FIOCRUZ | Paola Resende et al |
| EPI_ISL_1534007 | Laboratorio Central de Saude Publica do Estado de Santa Catarina (LACEN-SC) | Laboratory of Respiratory Viruses and Measles, Oswaldo Cruz Institute, FIOCRUZ | Paola Resende et al |
| EPI_ISL_1534003 | Laboratorio Central de Saude Publica do Estado de Santa Catarina (LACEN-SC) | Laboratory of Respiratory Viruses and Measles, Oswaldo Cruz Institute, FIOCRUZ | Paola Resende et al |
| EPI_ISL_1534006 | Laboratorio Central de Saude Publica do Estado de Santa Catarina (LACEN-SC) | Laboratory of Respiratory Viruses and Measles, Oswaldo Cruz Institute, FIOCRUZ | Paola Resende et al |
| EPI_ISL_1534010 | Laboratorio Central de Saude Publica do Estado de Santa Catarina (LACEN-SC) | Laboratory of Respiratory Viruses and Measles, Oswaldo Cruz Institute, FIOCRUZ | Paola Resende et al |
| EPI_ISL_1534008 | Laboratorio Central de Saude Publica do Estado de Santa Catarina (LACEN-SC) | Laboratory of Respiratory Viruses and Measles, Oswaldo Cruz Institute, FIOCRUZ | Paola Resende et al |
| EPI_ISL_1534009 | Laboratorio Central de Saude Publica do Estado de Santa Catarina (LACEN-SC) | Laboratory of Respiratory Viruses and Measles, Oswaldo Cruz Institute, FIOCRUZ | Paola Resende et al |
| EPI_ISL_1534002 | Laboratorio Central de Saude Publica do Estado de Santa Catarina (LACEN-SC) | Laboratory of Respiratory Viruses and Measles, Oswaldo Cruz Institute, FIOCRUZ | Paola Resende et al |
| EPI_ISL_1533994 | Laboratorio Central de Saude Publica do Estado de Santa Catarina (LACEN-SC) | Laboratory of Respiratory Viruses and Measles, Oswaldo Cruz Institute, FIOCRUZ | Paola Resende et al |
| EPI_ISL_1533997 | Laboratorio Central de Saude Publica do Estado de Santa Catarina (LACEN-SC) | Laboratory of Respiratory Viruses and Measles, Oswaldo Cruz Institute, FIOCRUZ | Paola Resende et al |
| EPI_ISL_1533992 | Laboratorio Central de Saude Publica do Estado de Santa Catarina (LACEN-SC) | Laboratory of Respiratory Viruses and Measles, Oswaldo Cruz Institute, FIOCRUZ | Paola Resende et al |
| EPI_ISL_1533990 | Laboratorio Central de Saude Publica do Estado de Santa Catarina (LACEN-SC) | Laboratory of Respiratory Viruses and Measles, Oswaldo Cruz Institute, FIOCRUZ | Paola Resende et al |

| | | | |
|---|---|---|---|
| EPI_ISL_1533996 | Laboratorio Central de Saude Publica do Estado de Santa Catarina (LACEN-SC) | Laboratory of Respiratory Viruses and Measles, Oswaldo Cruz Institute, FIOCRUZ | Paola Resende et al |
| EPI_ISL_1533993 | Laboratorio Central de Saude Publica do Estado de Santa Catarina (LACEN-SC) | Laboratory of Respiratory Viruses and Measles, Oswaldo Cruz Institute, FIOCRUZ | Paola Resende et al |
| EPI_ISL_1181402 | Laboratory of Respiratory Viruses and Measles, Oswaldo Cruz Institute, FIOCRUZ | Laboratory of Respiratory Viruses and Measles, Oswaldo Cruz Institute, FIOCRUZ | Paola Resende et al |
| EPI_ISL_1181412 | Laboratorio Central de Saude Publica do Estado de Santa Catarina (LACEN-SC) | Laboratory of Respiratory Viruses and Measles, Oswaldo Cruz Institute, FIOCRUZ | Paola Resende et al |
| EPI_ISL_1181404 | Laboratorio Central de Saude Publica do Estado de Santa Catarina (LACEN-SC) | Laboratory of Respiratory Viruses and Measles, Oswaldo Cruz Institute, FIOCRUZ | Paola Resende et al |
| EPI_ISL_1181453 | Laboratorio Central de Saude Publica do Estado de Santa Catarina (LACEN-SC) | Laboratory of Respiratory Viruses and Measles, Oswaldo Cruz Institute, FIOCRUZ | Paola Resende et al |
| EPI_ISL_427305 | Laboratório Central de Saúde Pública do Estado de Santa Catarina (LACEN-SC) | Laboratory of Respiratory Viruses and Measles, Oswaldo Cruz Institute, FIOCRUZ | Paola Resende et al |
| EPI_ISL_427306 | Laboratório Central de Saúde Pública do Estado de Santa Catarina (LACEN-SC) | Laboratory of Respiratory Viruses and Measles, Oswaldo Cruz Institute, FIOCRUZ | Paola Resende et al |
| EPI_ISL_541370 | Laboratório Central de Saúde Pública do Estado de Santa Catarina (LACEN-SC) | Laboratory of Respiratory Viruses and Measles, Oswaldo Cruz Institute, FIOCRUZ | Paola Resende et al |
| EPI_ISL_541371 | Laboratório Central de Saúde Pública do Estado de Santa Catarina (LACEN-SC) | Laboratory of Respiratory Viruses and Measles, Oswaldo Cruz Institute, FIOCRUZ | Paola Resende et al |
| EPI_ISL_1534004 | Laboratorio Central de Saude Publica do Estado de Sergipe (LACEN-SE) | Laboratory of Respiratory Viruses and Measles, Oswaldo Cruz Institute, FIOCRUZ | Paola Resende et al |
| EPI_ISL_1181405 | Laboratorio Central de Saude Publica do Estado de Sergipe (LACEN-SE) | Laboratory of Respiratory Viruses and Measles, Oswaldo Cruz Institute, FIOCRUZ | Paola Resende et al |
| EPI_ISL_1181409 | Laboratorio Central de Saude Publica do Estado de Sergipe (LACEN-SE) | Laboratory of Respiratory Viruses and Measles, Oswaldo Cruz Institute, FIOCRUZ | Paola Resende et al |
| EPI_ISL_1181406 | Laboratorio Central de Saude Publica do Estado de Sergipe (LACEN-SE) | Laboratory of Respiratory Viruses and Measles, Oswaldo Cruz Institute, FIOCRUZ | Paola Resende et al |
| EPI_ISL_1181407 | Laboratorio Central de Saude Publica do Estado de Sergipe (LACEN-SE) | Laboratory of Respiratory Viruses and Measles, Oswaldo Cruz Institute, FIOCRUZ | Paola Resende et al |
| EPI_ISL_541372 | Laboratório Central de Saúde Pública do Estado de Sergipe (LACEN-SE) | Laboratory of Respiratory Viruses and Measles, Oswaldo Cruz Institute, FIOCRUZ | Paola Resende et al |
| EPI_ISL_541373 | Laboratório Central de Saúde Pública do Estado de Sergipe (LACEN-SE) | Laboratory of Respiratory Viruses and Measles, Oswaldo Cruz Institute, FIOCRUZ | Paola Resende et al |
| EPI_ISL_541374 | Laboratório Central de Saúde Pública do Estado de Sergipe (LACEN-SE) | Laboratory of Respiratory Viruses and Measles, Oswaldo Cruz Institute, FIOCRUZ | Paola Resende et al |
| EPI_ISL_541375 | Laboratório Central de Saúde Pública do Estado de Sergipe (LACEN-SE) | Laboratory of Respiratory Viruses and Measles, Oswaldo Cruz Institute, FIOCRUZ | Paola Resende et al |
| EPI_ISL_541376 | Laboratório Central de Saúde Pública do Estado de Sergipe (LACEN-SE) | Laboratory of Respiratory Viruses and Measles, Oswaldo Cruz Institute, FIOCRUZ | Paola Resende et al |
| EPI_ISL_541377 | Laboratório Central de Saúde Pública do Estado de Sergipe (LACEN-SE) | Laboratory of Respiratory Viruses and Measles, Oswaldo Cruz Institute, FIOCRUZ | Paola Resende et al |
| EPI_ISL_541378 | Laboratório Central de Saúde Pública do Estado de Sergipe (LACEN-SE) | Laboratory of Respiratory Viruses and Measles, Oswaldo Cruz Institute, FIOCRUZ | Paola Resende et al |

| | | | |
|---|---|---|---|
| EPI_ISL_541379 | Laboratório Central de Saúde Pública do Estado de Sergipe (LACEN-SE) | Laboratory of Respiratory Viruses and Measles, Oswaldo Cruz Institute, FIOCRUZ | Paola Resende et al |
| EPI_ISL_541380 | Laboratório Central de Saúde Pública do Estado de Sergipe (LACEN-SE) | Laboratory of Respiratory Viruses and Measles, Oswaldo Cruz Institute, FIOCRUZ | Paola Resende et al |
| EPI_ISL_1181590 | Laboratorio Central de Saude Publica do Estado de Sergipe (LACEN-SE) | Laboratory of Respiratory Viruses and Measles, Oswaldo Cruz Institute, FIOCRUZ | Paola Resende et al |
| EPI_ISL_541381 | Laboratório Central de Saúde Pública do Estado de Sergipe (LACEN-SE) | Laboratory of Respiratory Viruses and Measles, Oswaldo Cruz Institute, FIOCRUZ | Paola Resende et al |
| EPI_ISL_541382 | Laboratório Central de Saúde Pública do Estado de Sergipe (LACEN-SE) | Laboratory of Respiratory Viruses and Measles, Oswaldo Cruz Institute, FIOCRUZ | Paola Resende et al |
| EPI_ISL_541383 | Laboratório Central de Saúde Pública do Estado de Sergipe (LACEN-SE) | Laboratory of Respiratory Viruses and Measles, Oswaldo Cruz Institute, FIOCRUZ | Paola Resende et al |
| EPI_ISL_541384 | Laboratório Central de Saúde Pública do Estado de Sergipe (LACEN-SE) | Laboratory of Respiratory Viruses and Measles, Oswaldo Cruz Institute, FIOCRUZ | Paola Resende et al |
| EPI_ISL_541385 | Laboratório Central de Saúde Pública do Estado de Sergipe (LACEN-SE) | Laboratory of Respiratory Viruses and Measles, Oswaldo Cruz Institute, FIOCRUZ | Paola Resende et al |
| EPI_ISL_541386 | Laboratório Central de Saúde Pública do Estado de Sergipe (LACEN-SE) | Laboratory of Respiratory Viruses and Measles, Oswaldo Cruz Institute, FIOCRUZ | Paola Resende et al |
| EPI_ISL_541387 | Laboratório Central de Saúde Pública do Estado de Sergipe (LACEN-SE) | Laboratory of Respiratory Viruses and Measles, Oswaldo Cruz Institute, FIOCRUZ | Paola Resende et al |
| EPI_ISL_541388 | Laboratório Central de Saúde Pública do Estado de Sergipe (LACEN-SE) | Laboratory of Respiratory Viruses and Measles, Oswaldo Cruz Institute, FIOCRUZ | Paola Resende et al |
| EPI_ISL_541389 | Laboratório Central de Saúde Pública do Estado de Sergipe (LACEN-SE) | Laboratory of Respiratory Viruses and Measles, Oswaldo Cruz Institute, FIOCRUZ | Paola Resende et al |
| EPI_ISL_541390 | Laboratório Central de Saúde Pública do Estado de Sergipe (LACEN-SE) | Laboratory of Respiratory Viruses and Measles, Oswaldo Cruz Institute, FIOCRUZ | Paola Resende et al |
| EPI_ISL_541391 | Laboratório Central de Saúde Pública do Estado de Sergipe (LACEN-SE) | Laboratory of Respiratory Viruses and Measles, Oswaldo Cruz Institute, FIOCRUZ | Paola Resende et al |
| EPI_ISL_541392 | Laboratório Central de Saúde Pública do Estado de Sergipe (LACEN-SE) | Laboratory of Respiratory Viruses and Measles, Oswaldo Cruz Institute, FIOCRUZ | Paola Resende et al |
| EPI_ISL_1181595 | Laboratorio Central de Saude Publica do Estado de Sergipe (LACEN-SE) | Laboratory of Respiratory Viruses and Measles, Oswaldo Cruz Institute, FIOCRUZ | Paola Resende et al |
| EPI_ISL_541393 | Laboratório Central de Saúde Pública do Estado de Sergipe (LACEN-SE) | Laboratory of Respiratory Viruses and Measles, Oswaldo Cruz Institute, FIOCRUZ | Paola Resende et al |
| EPI_ISL_541394 | Laboratório Central de Saúde Pública do Estado de Sergipe (LACEN-SE) | Laboratory of Respiratory Viruses and Measles, Oswaldo Cruz Institute, FIOCRUZ | Paola Resende et al |
| EPI_ISL_541395 | Laboratório Central de Saúde Pública do Estado de Sergipe (LACEN-SE) | Laboratory of Respiratory Viruses and Measles, Oswaldo Cruz Institute, FIOCRUZ | Paola Resende et al |
| EPI_ISL_541396 | Laboratório Central de Saúde Pública do Estado de Sergipe (LACEN-SE) | Laboratory of Respiratory Viruses and Measles, Oswaldo Cruz Institute, FIOCRUZ | Paola Resende et al |
| EPI_ISL_1533988 | Laboratorio Nacional de Salud | Laboratory of Respiratory Viruses and Measles, Oswaldo Cruz Institute, FIOCRUZ | Paola Resende et al |
| EPI_ISL_1533989 | Laboratorio Nacional de Salud | Laboratory of Respiratory Viruses and Measles, Oswaldo Cruz Institute, FIOCRUZ | Paola Resende et al |

| | | | |
|---|---|---|---|
| EPI_ISL_837579 | Laboratorio Nacional de Salud | Laboratory of Respiratory Viruses and Measles, Oswaldo Cruz Institute, FIOCRUZ | Paola Resende et al |
| EPI_ISL_837580 | Laboratorio Nacional de Salud | Laboratory of Respiratory Viruses and Measles, Oswaldo Cruz Institute, FIOCRUZ | Paola Resende et al |
| EPI_ISL_837581 | Laboratorio Nacional de Salud | Laboratory of Respiratory Viruses and Measles, Oswaldo Cruz Institute, FIOCRUZ | Paola Resende et al |
| EPI_ISL_837582 | Laboratorio Nacional de Salud | Laboratory of Respiratory Viruses and Measles, Oswaldo Cruz Institute, FIOCRUZ | Paola Resende et al |
| EPI_ISL_837583 | Laboratorio Nacional de Salud | Laboratory of Respiratory Viruses and Measles, Oswaldo Cruz Institute, FIOCRUZ | Paola Resende et al |
| EPI_ISL_837551 | Laboratorio Nacional de Salud | Laboratory of Respiratory Viruses and Measles, Oswaldo Cruz Institute, FIOCRUZ | Paola Resende et al |
| EPI_ISL_837584 | Laboratorio Nacional de Salud | Laboratory of Respiratory Viruses and Measles, Oswaldo Cruz Institute, FIOCRUZ | Paola Resende et al |
| EPI_ISL_837586 | Laboratorio Nacional de Salud | Laboratory of Respiratory Viruses and Measles, Oswaldo Cruz Institute, FIOCRUZ | Paola Resende et al |
| EPI_ISL_837587 | Laboratorio Nacional de Salud | Laboratory of Respiratory Viruses and Measles, Oswaldo Cruz Institute, FIOCRUZ | Paola Resende et al |
| EPI_ISL_837588 | Laboratorio Nacional de Salud | Laboratory of Respiratory Viruses and Measles, Oswaldo Cruz Institute, FIOCRUZ | Paola Resende et al |
| EPI_ISL_837589 | Laboratorio Nacional de Salud | Laboratory of Respiratory Viruses and Measles, Oswaldo Cruz Institute, FIOCRUZ | Paola Resende et al |
| EPI_ISL_837592 | Laboratorio Nacional de Salud | Laboratory of Respiratory Viruses and Measles, Oswaldo Cruz Institute, FIOCRUZ | Paola Resende et al |
| EPI_ISL_837593 | Laboratorio Nacional de Salud | Laboratory of Respiratory Viruses and Measles, Oswaldo Cruz Institute, FIOCRUZ | Paola Resende et al |
| EPI_ISL_837595 | Laboratorio Nacional de Salud | Laboratory of Respiratory Viruses and Measles, Oswaldo Cruz Institute, FIOCRUZ | Paola Resende et al |
| EPI_ISL_837596 | Laboratorio Nacional de Salud | Laboratory of Respiratory Viruses and Measles, Oswaldo Cruz Institute, FIOCRUZ | Paola Resende et al |
| EPI_ISL_837552 | Laboratorio Nacional de Salud | Laboratory of Respiratory Viruses and Measles, Oswaldo Cruz Institute, FIOCRUZ | Paola Resende et al |
| EPI_ISL_1533981 | Laboratorio Nacional de Salud | Laboratory of Respiratory Viruses and Measles, Oswaldo Cruz Institute, FIOCRUZ | Paola Resende et al |
| EPI_ISL_1533982 | Laboratorio Nacional de Salud | Laboratory of Respiratory Viruses and Measles, Oswaldo Cruz Institute, FIOCRUZ | Paola Resende et al |
| EPI_ISL_1533984 | Laboratorio Nacional de Salud | Laboratory of Respiratory Viruses and Measles, Oswaldo Cruz Institute, FIOCRUZ | Paola Resende et al |
| EPI_ISL_1533985 | Laboratorio Nacional de Salud | Laboratory of Respiratory Viruses and Measles, Oswaldo Cruz Institute, FIOCRUZ | Paola Resende et al |
| EPI_ISL_1533987 | Laboratorio Nacional de Salud | Laboratory of Respiratory Viruses and Measles, Oswaldo Cruz Institute, FIOCRUZ | Paola Resende et al |
| EPI_ISL_1340751 | Departamento de Virologia, Laboratorio Central de Salud Pública, Avenida Venezuela y Teniente Escurra, Asunción, Paraguay | Laboratory of Respiratory Viruses and Measles, Oswaldo Cruz Institute, FIOCRUZ | Paola Resende et al |

| | | | |
|---|---|---|---|
| EPI_ISL_1340750 | Departamento de Virologia, Laboratorio Central de Salud Pública, Avenida Venezuela y Teniente Escurra, Asunción, Paraguay | Laboratory of Respiratory Viruses and Measles, Oswaldo Cruz Institute, FIOCRUZ | Paola Resende et al |
| EPI_ISL_1340764 | Departamento de Virologia, Laboratorio Central de Salud Pública, Avenida Venezuela y Teniente Escurra, Asunción, Paraguay | Laboratory of Respiratory Viruses and Measles, Oswaldo Cruz Institute, FIOCRUZ | Paola Resende et al |
| EPI_ISL_1340762 | Departamento de Virologia, Laboratorio Central de Salud Pública, Avenida Venezuela y Teniente Escurra, Asunción, Paraguay | Laboratory of Respiratory Viruses and Measles, Oswaldo Cruz Institute, FIOCRUZ | Paola Resende et al |
| EPI_ISL_1340763 | Departamento de Virologia, Laboratorio Central de Salud Pública, Avenida Venezuela y Teniente Escurra, Asunción, Paraguay | Laboratory of Respiratory Viruses and Measles, Oswaldo Cruz Institute, FIOCRUZ | Paola Resende et al |
| EPI_ISL_1340761 | Departamento de Virologia, Laboratorio Central de Salud Pública, Avenida Venezuela y Teniente Escurra, Asunción, Paraguay | Laboratory of Respiratory Viruses and Measles, Oswaldo Cruz Institute, FIOCRUZ | Paola Resende et al |
| EPI_ISL_1340760 | Departamento de Virologia, Laboratorio Central de Salud Pública, Avenida Venezuela y Teniente Escurra, Asunción, Paraguay | Laboratory of Respiratory Viruses and Measles, Oswaldo Cruz Institute, FIOCRUZ | Paola Resende et al |
| EPI_ISL_1340759 | Departamento de Virologia, Laboratorio Central de Salud Pública, Avenida Venezuela y Teniente Escurra, Asunción, Paraguay | Laboratory of Respiratory Viruses and Measles, Oswaldo Cruz Institute, FIOCRUZ | Paola Resende et al |
| EPI_ISL_1340758 | Departamento de Virologia, Laboratorio Central de Salud Pública, Avenida Venezuela y Teniente Escurra, Asunción, Paraguay | Laboratory of Respiratory Viruses and Measles, Oswaldo Cruz Institute, FIOCRUZ | Paola Resende et al |
| EPI_ISL_1340756 | Departamento de Virologia, Laboratorio Central de Salud Pública, Avenida Venezuela y Teniente Escurra, Asunción, Paraguay | Laboratory of Respiratory Viruses and Measles, Oswaldo Cruz Institute, FIOCRUZ | Paola Resende et al |
| EPI_ISL_1340753 | Departamento de Virologia, Laboratorio Central de Salud Pública, Avenida Venezuela y Teniente Escurra, Asunción, Paraguay | Laboratory of Respiratory Viruses and Measles, Oswaldo Cruz Institute, FIOCRUZ | Paola Resende et al |
| EPI_ISL_1340754 | Departamento de Virologia, Laboratorio Central de Salud Pública, Avenida Venezuela y Teniente Escurra, Asunción, Paraguay | Laboratory of Respiratory Viruses and Measles, Oswaldo Cruz Institute, FIOCRUZ | Paola Resende et al |
| EPI_ISL_1340755 | Departamento de Virologia, Laboratorio Central de Salud Pública, Avenida Venezuela y Teniente Escurra, Asunción, Paraguay | Laboratory of Respiratory Viruses and Measles, Oswaldo Cruz Institute, FIOCRUZ | Paola Resende et al |
| EPI_ISL_1340752 | Departamento de Virologia, Laboratorio Central de Salud Pública, Avenida Venezuela y Teniente Escurra, Asunción, Paraguay | Laboratory of Respiratory Viruses and Measles, Oswaldo Cruz Institute, FIOCRUZ | Paola Resende et al |
| EPI_ISL_875687 | Department of Infectious Diseases, Istituto Superiore di Sanità, Rome, Italy; UCO Igiene e Sanità Pubblica, ASUGI, Trieste, Italy; Laboratorio di Genomica ed Epigenomica sistema Argo, Area SciencePark, Trieste, Italy; Laboratorio di Virologia Molecolare, ICGEB, Trieste, Italy | Istituto Superiore di Sanità (ISS) | Paola Stefanelli et al |
| EPI_ISL_412974 | Department of Infectious Diseases, Istituto Superiore di Sanità, Rome, Italy | Virology Laboratory, Scientific Department, Army Medical Center | Paola Stefanelli et al |
| EPI_ISL_1443188 | CHWAPI - SITE NOTRE DAME | Institut de Pathologie et Genetique (IPG) | Pascale Hilbert et al |
| EPI_ISL_1451467 | CHWAPI - SITE NOTRE DAME | Institut de Pathologie et Genetique (IPG) | Pascale Hilbert et al |
| EPI_ISL_1451473 | CHWAPI - SITE NOTRE DAME | Institut de Pathologie et Genetique (IPG) | Pascale Hilbert et al |
| EPI_ISL_1299241 | CHWAPI - SITE NOTRE DAME | Institut de Pathologie et Genetique (IPG) | Pascale Hilbert et al |
| EPI_ISL_596709 | PathWest Laboratory Medicine WA | PathWest Laboratory Medicine WA Microbial Surveillance Unit | PathWest Laboratory Medicine WA Microbia |
| EPI_ISL_596764 | PathWest Laboratory Medicine WA | PathWest Laboratory Medicine WA Microbial Surveillance Unit | PathWest Laboratory Medicine WA Microbia |
| EPI_ISL_596766 | PathWest Laboratory Medicine WA | PathWest Laboratory Medicine WA Microbial Surveillance Unit | PathWest Laboratory Medicine WA Microbia |
| EPI_ISL_596777 | PathWest Laboratory Medicine WA | PathWest Laboratory Medicine WA Microbial Surveillance Unit | PathWest Laboratory Medicine WA Microbia |
| EPI_ISL_605879 | PathWest Laboratory Medicine WA | PathWest Laboratory Medicine WA Microbial Surveillance Unit | PathWest Laboratory Medicine WA Microbia |

al Surveillance Unit et al
al Surveillance Unit et al
al Surveillance Unit et al
al Surveillance Unit et al
al Surveillance Unit et al

| | | | |
|---|---|---|---|
| EPI_ISL_672653 | PathWest Laboratory Medicine WA | PathWest Laboratory Medicine WA Microbial Surveillance Unit | PathWest Laboratory Medicine WA Microbia |
| EPI_ISL_708745 | PathWest Laboratory Medicine WA | PathWest Laboratory Medicine WA Microbial Surveillance Unit | PathWest Laboratory Medicine WA Microbia |
| EPI_ISL_794715 | PathWest Laboratory Medicine WA | PathWest Laboratory Medicine WA Microbial Surveillance Unit | PathWest Laboratory Medicine WA Microbia |
| EPI_ISL_794726 | PathWest Laboratory Medicine WA | PathWest Laboratory Medicine WA Microbial Surveillance Unit | PathWest Laboratory Medicine WA Microbia |
| EPI_ISL_794672 | PathWest Laboratory Medicine WA | PathWest Laboratory Medicine WA Microbial Surveillance Unit | PathWest Laboratory Medicine WA Microbia |
| EPI_ISL_933795 | PathWest Laboratory Medicine WA | PathWest Laboratory Medicine WA Microbial Surveillance Unit | PathWest Laboratory Medicine WA Microbia |
| EPI_ISL_1184507 | PathWest Laboratory Medicine WA | PathWest Laboratory Medicine WA Microbial Surveillance Unit | PathWest Laboratory Medicine WA Microbia |
| EPI_ISL_1416322 | PathWest Laboratory Medicine WA | PathWest Laboratory Medicine WA Microbial Surveillance Unit | PathWest Laboratory Medicine WA Microbia |
| EPI_ISL_1508996 | PathWest Laboratory Medicine WA | PathWest Laboratory Medicine WA Microbial Surveillance Unit | PathWest Laboratory Medicine WA Microbia |
| EPI_ISL_1620802 | PathWest Laboratory Medicine WA | PathWest Laboratory Medicine WA Microbial Surveillance Unit | PathWest Laboratory Medicine WA Microbia |
| EPI_ISL_1633680 | PathWest Laboratory Medicine WA | PathWest Laboratory Medicine WA Microbial Surveillance Unit | PathWest Laboratory Medicine WA Microbia |
| EPI_ISL_1669122 | PathWest Laboratory Medicine WA | PathWest Laboratory Medicine WA Microbial Surveillance Unit | PathWest Laboratory Medicine WA Microbia |
| EPI_ISL_1580504 | Laboratório de Biologia Molecular do Hospital das Clínicas da Faculdade de Medicina de Botucatu/SP | Laboratórios de Genômica Funcional (FCA/UNESP) e Biologia Molecular (FMB-HC/UNESP) - Rede de Vigilância Genômica (Vigenômica)/UNESP | Patrícia Akemi Assato et al |
| EPI_ISL_1580502 | Laboratório de Biologia Molecular do Hospital das Clínicas da Faculdade de Medicina de Botucatu/SP | Laboratórios de Genômica Funcional (FCA/UNESP) e Biologia Molecular (FMB-HC/UNESP) - Rede de Vigilância Genômica (Vigenômica)/UNESP | Patrícia Akemi Assato et al |
| EPI_ISL_681887 | National Virus Reference Laboratory | Irish Coronavirus Sequencing Consortium - Teagasc Moorepark | Paul Cotter et al |
| EPI_ISL_1324139 | UW Virology Lab | UW Virology Lab | Pavitra Roychoudhury et al |
| EPI_ISL_1324149 | UW Virology Lab | UW Virology Lab | Pavitra Roychoudhury et al |
| EPI_ISL_1324142 | UW Virology Lab | UW Virology Lab | Pavitra Roychoudhury et al |
| EPI_ISL_1324145 | UW Virology Lab | UW Virology Lab | Pavitra Roychoudhury et al |
| EPI_ISL_1324137 | UW Virology Lab | UW Virology Lab | Pavitra Roychoudhury et al |
| EPI_ISL_1324140 | UW Virology Lab | UW Virology Lab | Pavitra Roychoudhury et al |
| EPI_ISL_1324138 | UW Virology Lab | UW Virology Lab | Pavitra Roychoudhury et al |
| EPI_ISL_1324135 | UW Virology Lab | UW Virology Lab | Pavitra Roychoudhury et al |
| EPI_ISL_1324144 | UW Virology Lab | UW Virology Lab | Pavitra Roychoudhury et al |
| EPI_ISL_538512 | Balai Penelitian dan Pengembangan Biomedis Papua | National Institute of Health Research and Development | Pawestri et al |
| EPI_ISL_910075 | CSIR-Centre for Cellular and Molecular Biology | CSIR-Centre for Cellular and Molecular Biology | Payel Mukherjee et al |
| EPI_ISL_910082 | CSIR-Centre for Cellular and Molecular Biology | CSIR-Centre for Cellular and Molecular Biology | Payel Mukherjee et al |
| EPI_ISL_910184 | CSIR-Centre for Cellular and Molecular Biology | CSIR-Centre for Cellular and Molecular Biology | Payel Mukherjee et al |
| EPI_ISL_910186 | CSIR-Centre for Cellular and Molecular Biology | CSIR-Centre for Cellular and Molecular Biology | Payel Mukherjee et al |
| EPI_ISL_910273 | CSIR-Centre for Cellular and Molecular Biology | CSIR-Centre for Cellular and Molecular Biology | Payel Mukherjee et al |

al Surveillance Unit et al

al Surveillance Unit et al

al Surveillance Unit et al

al Surveillance Unit et al

al Surveillance Unit et al

al Surveillance Unit et al

al Surveillance Unit et al

al Surveillance Unit et al

al Surveillance Unit et al

al Surveillance Unit et al

al Surveillance Unit et al

al Surveillance Unit et al

| | | | |
|---|---|---|---|
| EPI_ISL_910276 | CSIR-Centre for Cellular and Molecular Biology | CSIR-Centre for Cellular and Molecular Biology | Payel Mukherjee et al |
| EPI_ISL_1380090 | CSIR-Centre for Cellular and Molecular Biology | CSIR-Centre for Cellular and Molecular Biology | Payel Mukherjee et al |
| EPI_ISL_1380102 | CSIR-Centre for Cellular and Molecular Biology | CSIR-Centre for Cellular and Molecular Biology | Payel Mukherjee et al |
| EPI_ISL_1380150 | CSIR-Centre for Cellular and Molecular Biology | CSIR-Centre for Cellular and Molecular Biology | Payel Mukherjee et al |
| EPI_ISL_1380167 | CSIR-Centre for Cellular and Molecular Biology | CSIR-Centre for Cellular and Molecular Biology | Payel Mukherjee et al |
| EPI_ISL_1380196 | CSIR-Centre for Cellular and Molecular Biology | CSIR-Centre for Cellular and Molecular Biology | Payel Mukherjee et al |
| EPI_ISL_1380299 | CSIR-Centre for Cellular and Molecular Biology | CSIR-Centre for Cellular and Molecular Biology | Payel Mukherjee et al |
| EPI_ISL_1380470 | CSIR-Centre for Cellular and Molecular Biology | CSIR-Centre for Cellular and Molecular Biology | Payel Mukherjee et al |
| EPI_ISL_539720 | CSIR-Centre for Cellular and Molecular Biology | CSIR-Centre for Cellular and Molecular Biology | Payel Mukherjee et al |
| EPI_ISL_1260835 | BSL-3 Lab, National Institute for Viral Disease Control and Prevention, Chinese Center for Disease Control and Prevention | BSL-3 Lab, National Institute for Viral Disease Control and Prevention, Chinese Center for Disease Control and Prevention | Peipei Liu et al |
| EPI_ISL_430737 | Chinese PLA Institute for Disease Control and Prevention | Chinese PLA Institute for Disease Control and Prevention | Peng Li et al |
| EPI_ISL_815257 | Centogene | Centogene | Peter Bauer et al |
| EPI_ISL_815398 | Centogene | Centogene | Peter Bauer et al |
| EPI_ISL_815256 | Centogene | Centogene | Peter Bauer et al |
| EPI_ISL_815255 | Centogene | Centogene | Peter Bauer et al |
| EPI_ISL_815390 | Centogene | Centogene | Peter Bauer et al |
| EPI_ISL_435410 | Virological Research Group, Szentágothai Research Centre | Bioinformatics Research Group, Szentágothai Research Centre | Péter Urbán et al |
| EPI_ISL_435416 | Virological Research Group, Szentágothai Research Centre | Bioinformatics Research Group, Szentágothai Research Centre | Péter Urbán et al |
| EPI_ISL_435412 | Virological Research Group, Szentágothai Research Centre | Bioinformatics Research Group, Szentágothai Research Centre | Péter Urbán et al |
| EPI_ISL_435415 | Virological Research Group, Szentágothai Research Centre | Bioinformatics Research Group, Szentágothai Research Centre | Péter Urbán et al |
| EPI_ISL_1027843 | Laboratory Corporation of America | Respiratory Viruses Branch, Division of Viral Diseases, Centers for Disease Control and Prevention | Peter W. Cook et al |
| EPI_ISL_1252981 | Quest Diagnostics Incorporated | Centers for Disease Control and Prevention Division of Viral Diseases, Pathogen Discovery | Peter W. Cook et al |
| EPI_ISL_1268990 | Helix/Illumina | Centers for Disease Control and Prevention Division of Viral Diseases, Pathogen Discovery | Peter W. Cook et al |
| EPI_ISL_1267799 | Helix/Illumina | Centers for Disease Control and Prevention Division of Viral Diseases, Pathogen Discovery | Peter W. Cook et al |
| EPI_ISL_1088103 | Quest Diagnostics Incorporated | Respiratory Viruses Branch, Division of Viral Diseases, Centers for Disease Control and Prevention | Peter W. Cook et al |
| EPI_ISL_889336 | The University Hospital Brno | Institute of Applied Biotechnologies a.s. | Petr Klempt et al |
| EPI_ISL_889346 | The University Hospital Brno | Institute of Applied Biotechnologies a.s. | Petr Klempt et al |
| EPI_ISL_889349 | The University Hospital Brno | Institute of Applied Biotechnologies a.s. | Petr Klempt et al |

| | | | |
|---|---|---|---|
| EPI_ISL_889350 | The University Hospital Brno | Institute of Applied Biotechnologies a.s. | Petr Klempt et al |
| EPI_ISL_889351 | The University Hospital Brno | Institute of Applied Biotechnologies a.s. | Petr Klempt et al |
| EPI_ISL_889371 | Olomouc University Hospital | Institute of Applied Biotechnologies a.s. | Petr Klempt et al |
| EPI_ISL_889359 | Motol University Hospital | Institute of Applied Biotechnologies a.s. | Petr Klempt et al |
| EPI_ISL_889362 | Motol University Hospital | Institute of Applied Biotechnologies a.s. | Petr Klempt et al |
| EPI_ISL_889428 | Olomouc University Hospital | Institute of Applied Biotechnologies a.s. | Petr Klempt et al |
| EPI_ISL_889347 | The University Hospital Brno | Institute of Applied Biotechnologies a.s. | Petr Klempt et al |
| EPI_ISL_889360 | Motol University Hospital | Institute of Applied Biotechnologies a.s. | Petr Klempt et al |
| EPI_ISL_1588739 | Bioptická laboratoř s.r.o. | Bioptická laboratoř s.r.o. | Petr Šteiner et al |
| EPI_ISL_1588734 | Bioptická laboratoř s.r.o. | Bioptická laboratoř s.r.o. | Petr Šteiner et al |
| EPI_ISL_1588736 | Bioptická laboratoř s.r.o. | Bioptická laboratoř s.r.o. | Petr Šteiner et al |
| EPI_ISL_1588735 | Bioptická laboratoř s.r.o. | Bioptická laboratoř s.r.o. | Petr Šteiner et al |
| EPI_ISL_1299856 | Unit of lab surveillance of viral emerging diseases, National Lab of Influenza | Respiratory Virus Unit, National Infection Service, Public Health England | PHE Covid Sequencing Team et al |
| EPI_ISL_1299858 | Unit of lab surveillance of viral emerging diseases, National Lab of Influenza | Respiratory Virus Unit, National Infection Service, Public Health England | PHE Covid Sequencing Team et al |
| EPI_ISL_1299861 | Unit of lab surveillance of viral emerging diseases, National Lab of Influenza | Respiratory Virus Unit, National Infection Service, Public Health England | PHE Covid Sequencing Team et al |
| EPI_ISL_1299862 | Unit of lab surveillance of viral emerging diseases, National Lab of Influenza | Respiratory Virus Unit, National Infection Service, Public Health England | PHE Covid Sequencing Team et al |
| EPI_ISL_1299863 | Unit of lab surveillance of viral emerging diseases, National Lab of Influenza | Respiratory Virus Unit, National Infection Service, Public Health England | PHE Covid Sequencing Team et al |
| EPI_ISL_1299864 | Unit of lab surveillance of viral emerging diseases, National Lab of Influenza | Respiratory Virus Unit, National Infection Service, Public Health England | PHE Covid Sequencing Team et al |
| EPI_ISL_1299867 | Unit of lab surveillance of viral emerging diseases, National Lab of Influenza | Respiratory Virus Unit, National Infection Service, Public Health England | PHE Covid Sequencing Team et al |
| EPI_ISL_1299871 | Unit of lab surveillance of viral emerging diseases, National Lab of Influenza | Respiratory Virus Unit, National Infection Service, Public Health England | PHE Covid Sequencing Team et al |
| EPI_ISL_1299873 | Unit of lab surveillance of viral emerging diseases, National Lab of Influenza | Respiratory Virus Unit, National Infection Service, Public Health England | PHE Covid Sequencing Team et al |
| EPI_ISL_1299878 | Unit of lab surveillance of viral emerging diseases, National Lab of Influenza | Respiratory Virus Unit, National Infection Service, Public Health England | PHE Covid Sequencing Team et al |
| EPI_ISL_1299882 | Unit of lab surveillance of viral emerging diseases, National Lab of Influenza | Respiratory Virus Unit, National Infection Service, Public Health England | PHE Covid Sequencing Team et al |
| EPI_ISL_1299884 | Unit of lab surveillance of viral emerging diseases, National Lab of Influenza | Respiratory Virus Unit, National Infection Service, Public Health England | PHE Covid Sequencing Team et al |
| EPI_ISL_780385 | Bermuda Government Molecular Diagnostics Laboratory (MDL) | Respiratory Virus Unit, National Infection Service, Public Health England | PHE Covid Sequencing Team et al |
| EPI_ISL_780386 | Bermuda Government Molecular Diagnostics Laboratory (MDL) | Respiratory Virus Unit, National Infection Service, Public Health England | PHE Covid Sequencing Team et al |
| EPI_ISL_780392 | Bermuda Government Molecular Diagnostics Laboratory (MDL) | Respiratory Virus Unit, National Infection Service, Public Health England | PHE Covid Sequencing Team et al |
| EPI_ISL_780393 | Bermuda Government Molecular Diagnostics Laboratory (MDL) | Respiratory Virus Unit, National Infection Service, Public Health England | PHE Covid Sequencing Team et al |
| EPI_ISL_780384 | Bermuda Government Molecular Diagnostics Laboratory (MDL) | Respiratory Virus Unit, National Infection Service, Public Health England | PHE Covid Sequencing Team et al |
| EPI_ISL_780397 | Bermuda Government Molecular Diagnostics Laboratory (MDL) | Respiratory Virus Unit, National Infection Service, Public Health England | PHE Covid Sequencing Team et al |

| | | | |
|---|---|---|---|
| EPI_ISL_780398 | Bermuda Government Molecular Diagnostics Laboratory (MDL) | Respiratory Virus Unit, National Infection Service, Public Health England | PHE Covid Sequencing Team et al |
| EPI_ISL_780402 | Bermuda Government Molecular Diagnostics Laboratory (MDL) | Respiratory Virus Unit, National Infection Service, Public Health England | PHE Covid Sequencing Team et al |
| EPI_ISL_780403 | Bermuda Government Molecular Diagnostics Laboratory (MDL) | Respiratory Virus Unit, National Infection Service, Public Health England | PHE Covid Sequencing Team et al |
| EPI_ISL_780383 | Bermuda Government Molecular Diagnostics Laboratory (MDL) | Respiratory Virus Unit, National Infection Service, Public Health England | PHE Covid Sequencing Team et al |
| EPI_ISL_780404 | Bermuda Government Molecular Diagnostics Laboratory (MDL) | Respiratory Virus Unit, National Infection Service, Public Health England | PHE Covid Sequencing Team et al |
| EPI_ISL_780405 | Bermuda Government Molecular Diagnostics Laboratory (MDL) | Respiratory Virus Unit, National Infection Service, Public Health England | PHE Covid Sequencing Team et al |
| EPI_ISL_780406 | Bermuda Government Molecular Diagnostics Laboratory (MDL) | Respiratory Virus Unit, National Infection Service, Public Health England | PHE Covid Sequencing Team et al |
| EPI_ISL_780407 | Bermuda Government Molecular Diagnostics Laboratory (MDL) | Respiratory Virus Unit, National Infection Service, Public Health England | PHE Covid Sequencing Team et al |
| EPI_ISL_780408 | Bermuda Government Molecular Diagnostics Laboratory (MDL) | Respiratory Virus Unit, National Infection Service, Public Health England | PHE Covid Sequencing Team et al |
| EPI_ISL_780409 | Bermuda Government Molecular Diagnostics Laboratory (MDL) | Respiratory Virus Unit, National Infection Service, Public Health England | PHE Covid Sequencing Team et al |
| EPI_ISL_780410 | Bermuda Government Molecular Diagnostics Laboratory (MDL) | Respiratory Virus Unit, National Infection Service, Public Health England | PHE Covid Sequencing Team et al |
| EPI_ISL_464189 | Respiratory Virus Unit, Microbiology Services Colindale, Public Health England | Respiratory Virus Unit, Microbiology Services Colindale, Public Health England | PHE Covid Sequencing Team et al |
| EPI_ISL_464212 | Respiratory Virus Unit, Microbiology Services Colindale, Public Health England | Respiratory Virus Unit, Microbiology Services Colindale, Public Health England | PHE Covid Sequencing Team et al |
| EPI_ISL_464247 | Respiratory Virus Unit, Microbiology Services Colindale, Public Health England | Respiratory Virus Unit, Microbiology Services Colindale, Public Health England | PHE Covid Sequencing Team et al |
| EPI_ISL_466061 | Respiratory Virus Unit, Microbiology Services Colindale, Public Health England | Respiratory Virus Unit, Microbiology Services Colindale, Public Health England | PHE Covid Sequencing Team et al |
| EPI_ISL_465195 | Respiratory Virus Unit, Microbiology Services Colindale, Public Health England | Respiratory Virus Unit, Microbiology Services Colindale, Public Health England | PHE Covid Sequencing Team et al |
| EPI_ISL_547433 | Microbiology, Department of Pathology, St. Bernard's Hospital, Gibraltar Health Authority | Respiratory Virus Unit, Microbiology Services Colindale, Public Health England | PHE Covid Sequencing Team et al |
| EPI_ISL_547434 | Microbiology, Department of Pathology, St. Bernard's Hospital, Gibraltar Health Authority | Respiratory Virus Unit, Microbiology Services Colindale, Public Health England | PHE Covid Sequencing Team et al |
| EPI_ISL_547435 | Microbiology, Department of Pathology, St. Bernard's Hospital, Gibraltar Health Authority | Respiratory Virus Unit, Microbiology Services Colindale, Public Health England | PHE Covid Sequencing Team et al |
| EPI_ISL_547437 | Microbiology, Department of Pathology, St. Bernard's Hospital, Gibraltar Health Authority | Respiratory Virus Unit, Microbiology Services Colindale, Public Health England | PHE Covid Sequencing Team et al |
| EPI_ISL_487433 | Queen Astrid Military Hospital | Institute of Tropical Medicine | Philippe Selhorst et al |
| EPI_ISL_487434 | Queen Astrid Military Hospital | Institute of Tropical Medicine | Philippe Selhorst et al |
| EPI_ISL_487435 | Queen Astrid Military Hospital | Institute of Tropical Medicine | Philippe Selhorst et al |
| EPI_ISL_487436 | Queen Astrid Military Hospital | Institute of Tropical Medicine | Philippe Selhorst et al |
| EPI_ISL_877765 | UCO Igiene e Sanità Pubblica, ASUGI, Trieste, Italy; Laboratorio di Virologia Molecolare, ICGEB, Trieste, Italy | Laboratorio di Genomica ed Epigenomica sistema Argo, Area SciencePark, Trieste, Italy; | Pierlanfranco D'Agaro et al |
| EPI_ISL_450196 | bumrungrad international hospital | National Institute of Health. Department of medical Sciences, Ministry of Public Health, Thailand | Pilailuk et al |

| | | | |
|---|---|---|---|
| EPI_ISL_447920 | n/a | National Institute of Health. Department of medical Sciences, Ministry of Public Health, Thailand | Pilailuk et al |
| EPI_ISL_447913 | n/a | National Institute of Health. Department of medical Sciences, Ministry of Public Health, Thailand | Pilailuk et al |
| EPI_ISL_447918 | n/a | National Institute of Health. Department of medical Sciences, Ministry of Public Health, Thailand | Pilailuk et al |
| EPI_ISL_434695 | unknown | National Institute of Health. Department of medical Sciences, Ministry of Public Health, Thailand | Pilailuk et al |
| EPI_ISL_434700 | Ramkhamhaeng Hospital | National Institute of Health. Department of medical Sciences, Ministry of Public Health, Thailand | Pilailuk et al |
| EPI_ISL_434702 | Praram 9 Hospital | National Institute of Health. Department of medical Sciences, Ministry of Public Health, Thailand | Pilailuk et al |
| EPI_ISL_434707 | Thammasat University Hospital | National Institute of Health. Department of medical Sciences, Ministry of Public Health, Thailand | Pilailuk et al |
| EPI_ISL_447919 | n/a | National Institute of Health. Department of medical Sciences, Ministry of Public Health, Thailand | Pilailuk et al |
| EPI_ISL_708815 | Urban Institute for Disease Prevention and Control | National Institute of Health, Department of Medical Sciences, Ministry of Public Health, Thailand | Pilailuk Okada et al |
| EPI_ISL_708817 | Urban Institute for Disease Prevention and Control | National Institute of Health, Department of Medical Sciences, Ministry of Public Health, Thailand | Pilailuk Okada et al |
| EPI_ISL_708813 | World Medical Hospital | National Institute of Health, Department of Medical Sciences, Ministry of Public Health, Thailand | Pilailuk Okada et al |
| EPI_ISL_708812 | World Medical Hospital | National Institute of Health, Department of Medical Sciences, Ministry of Public Health, Thailand | Pilailuk Okada et al |
| EPI_ISL_768614 | Regional Medical Sciences Center 1/1 Chiang Rai | National Institute of Health, Department of Medical Sciences, Ministry of Public Health, Thailand | Pilailuk Okada et al |
| EPI_ISL_708737 | Regional medical sciences center 6 chonburi | National Institute of Health, Department of Medical Sciences, Ministry of Public Health, Thailand | Pilailuk Okada et al |
| EPI_ISL_708804 | Regional medical sciences center 6 chonburi | National Institute of Health, Department of Medical Sciences, Ministry of Public Health, Thailand | Pilailuk Okada et al |
| EPI_ISL_708802 | Regional medical sciences center 6 chonburi | National Institute of Health, Department of Medical Sciences, Ministry of Public Health, Thailand | Pilailuk Okada et al |
| EPI_ISL_708797 | Regional medical sciences center 6 chonburi | National Institute of Health, Department of Medical Sciences, Ministry of Public Health, Thailand | Pilailuk Okada et al |
| EPI_ISL_708800 | Regional medical sciences center 6 chonburi | National Institute of Health, Department of Medical Sciences, Ministry of Public Health, Thailand | Pilailuk Okada et al |
| EPI_ISL_708807 | Regional medical sciences center 6 chonburi | National Institute of Health, Department of Medical Sciences, Ministry of Public Health, Thailand | Pilailuk Okada et al |
| EPI_ISL_708808 | Regional medical sciences center 6 chonburi | National Institute of Health, Department of Medical Sciences, Ministry of Public Health, Thailand | Pilailuk Okada et al |
| EPI_ISL_708801 | Regional medical sciences center 6 chonburi | National Institute of Health, Department of Medical Sciences, Ministry of Public Health, Thailand | Pilailuk Okada et al |
| EPI_ISL_768610 | Regional medical sciences center 6 Chonburi | National Institute of Health, Department of Medical Sciences, Ministry of Public Health, Thailand | Pilailuk Okada et al |

| | | | |
|---|---|---|---|
| EPI_ISL_768611 | Regional medical sciences center 6 Chonburi | National Institute of Health, Department of Medical Sciences, Ministry of Public Health, Thailand | Pilailuk Okada et al |
| EPI_ISL_1497597 | The office of Disease Prevention and Control 6, Chonburi | National Institute of Health, Department of Medical Sciences, Ministry of Public Health, Thailand | Pilailuk Okada et al |
| EPI_ISL_755588 | Regional medical sciences center 6 Chonburi | National Institute of Health, Department of Medical Sciences, Ministry of Public Health, Thailand | Pilailuk Okada et al |
| EPI_ISL_768617 | Regional Medical Sciences Center 5 Samut Songkhram | National Institute of Health, Department of Medical Sciences, Ministry of Public Health, Thailand | Pilailuk Okada et al |
| EPI_ISL_768542 | Regional Medical Sciences Center 5 Samut Songkhram | National Institute of Health, Department of Medical Sciences, Ministry of Public Health, Thailand | Pilailuk Okada et al |
| EPI_ISL_708811 | Regional medical sciences center 2 Phitsanulok | National Institute of Health, Department of Medical Sciences, Ministry of Public Health, Thailand | Pilailuk Okada et al |
| EPI_ISL_471412 | Viral Respiratory Lab, National Institute for Biomedical Research (INRB) | Pathogen Sequencing Lab, National Institute for Biomedical Research (INRB) | Placide Mbala-Kingebeni et al |
| EPI_ISL_471413 | Viral Respiratory Lab, National Institute for Biomedical Research (INRB) | Pathogen Sequencing Lab, National Institute for Biomedical Research (INRB) | Placide Mbala-Kingebeni et al |
| EPI_ISL_527539 | Viral Respiratory Lab, National Institute for Biomedical Research (INRB) | Pathogen Sequencing Lab, National Institute for Biomedical Research (INRB) | Placide Mbala-Kingebeni et al |
| EPI_ISL_447596 | Viral Respiratory Lab, National Institute for Biomedical Research (INRB) | Pathogen Sequencing Lab, National Institute for Biomedical Research (INRB) | Placide Mbala-Kingebeni et al |
| EPI_ISL_965122 | Viral Respiratory Lab, National Institute for Biomedical Research (INRB) | Pathogen Sequencing Lab, National Institute for Biomedical Research (INRB) | Placide Mbala-Kingebeni et al |
| EPI_ISL_417950 | Viral Respiratory Lab, National Institute for Biomedical Research (INRB) | Pathogen Sequencing Lab, National Institute for Biomedical Research (INRB) | Placide Mbala-Kingebeni et al |
| EPI_ISL_591083 | Viral Respiratory Lab, National Institute for Biomedical Research (INRB) | Pathogen Sequencing Lab, National Institute for Biomedical Research (INRB) | Placide Mbala-Kingebeni et al |
| EPI_ISL_591084 | Viral Respiratory Lab, National Institute for Biomedical Research (INRB) | Pathogen Sequencing Lab, National Institute for Biomedical Research (INRB) | Placide Mbala-Kingebeni et al |
| EPI_ISL_591085 | Viral Respiratory Lab, National Institute for Biomedical Research (INRB) | Pathogen Sequencing Lab, National Institute for Biomedical Research (INRB) | Placide Mbala-Kingebeni et al |
| EPI_ISL_591086 | Viral Respiratory Lab, National Institute for Biomedical Research (INRB) | Pathogen Sequencing Lab, National Institute for Biomedical Research (INRB) | Placide Mbala-Kingebeni et al |
| EPI_ISL_420840 | Viral Respiratory Lab, National Institute for Biomedical Research (INRB) | Pathogen Sequencing Lab, National Institute for Biomedical Research (INRB) | Placide Mbala-Kingebeni et al |
| EPI_ISL_420841 | Viral Respiratory Lab, National Institute for Biomedical Research (INRB) | Pathogen Sequencing Lab, National Institute for Biomedical Research (INRB) | Placide Mbala-Kingebeni et al |
| EPI_ISL_447247 | Viral Respiratory Lab, National Institute for Biomedical Research (INRB) | Pathogen Sequencing Lab, National Institute for Biomedical Research (INRB) | Placide Mbala-Kingebeni et al |
| EPI_ISL_420853 | Viral Respiratory Lab, National Institute for Biomedical Research (INRB) | Pathogen Sequencing Lab, National Institute for Biomedical Research (INRB) | Placide Mbala-Kingebeni et al |
| EPI_ISL_513597 | Viral Respiratory Lab, National Institute for Biomedical Research (INRB) | Pathogen Sequencing Lab, National Institute for Biomedical Research (INRB) | Placide Mbala-Kingebeni et al |
| EPI_ISL_496498 | Viral Respiratory Lab, National Institute for Biomedical Research (INRB) | Pathogen Sequencing Lab, National Institute for Biomedical Research (INRB) | Placide Mbala-Kingebeni et al |

| | | | |
|---|---|---|---|
| EPI_ISL_961010 | Viral Respiratory Lab, National Institute for Biomedical Research (INRB) | Pathogen Sequencing Lab, National Institute for Biomedical Research (INRB) | Placide Mbala-Kingebeni et al |
| EPI_ISL_1623756 | Viral Respiratory Lab, National Institute for Biomedical Research (INRB) | Pathogen Sequencing Lab, National Institute for Biomedical Research (INRB) | Placide Mbala-Kingebeni et al |
| EPI_ISL_1623757 | Viral Respiratory Lab, National Institute for Biomedical Research (INRB) | Pathogen Sequencing Lab, National Institute for Biomedical Research (INRB) | Placide Mbala-Kingebeni et al |
| EPI_ISL_417944 | Viral Respiratory Lab, National Institute for Biomedical Research (INRB) | Pathogen Sequencing Lab, National Institute for Biomedical Research (INRB) | Placide Mbala-Kingebeni et al |
| EPI_ISL_965123 | Viral Respiratory Lab, National Institute for Biomedical Research (INRB) | Pathogen Sequencing Lab, National Institute for Biomedical Research (INRB) | Placide Mbala-Kingebeni et al |
| EPI_ISL_965124 | Viral Respiratory Lab, National Institute for Biomedical Research (INRB) | Pathogen Sequencing Lab, National Institute for Biomedical Research (INRB) | Placide Mbala-Kingebeni et al |
| EPI_ISL_1623758 | Viral Respiratory Lab, National Institute for Biomedical Research (INRB) | Pathogen Sequencing Lab, National Institute for Biomedical Research (INRB) | Placide Mbala-Kingebeni et al |
| EPI_ISL_417434 | Viral Respiratory Lab, National Institute for Biomedical Research (INRB) | Pathogen Sequencing Lab, National Institute for Biomedical Research (INRB) | Placide Mbala-Kingebeni et al |
| EPI_ISL_413522 | Indian Council of Medical Research - National Institute of Virology | National Influenza Center, Indian Council of Medical Research - National Institute of Virology | Potdar V et al |
| EPI_ISL_413523 | Indian Council of Medical Research-National Institute of Virology | National Influenza Center, Indian Council of Medical Research-National Institute of Virology | Potdar V et al |
| EPI_ISL_454528 | NIV Influenza | NIV Influenza | Potdar V et al |
| EPI_ISL_454534 | NIV Influenza | NIV Influenza | Potdar V et al |
| EPI_ISL_454537 | NIV Influenza | NIV Influenza | Potdar V et al |
| EPI_ISL_479572 | NIV Influenza | NIV Influenza | Potdar V et al |
| EPI_ISL_1034165 | NIV Influenza | NIV Influenza | Potdar V et al |
| EPI_ISL_1034166 | NIV Influenza | NIV Influenza | Potdar V et al |
| EPI_ISL_754068 | Nepal Korea Friendship Municipality Hospital | Nepal Health Research Council | Pradip Gyanwali et al |
| EPI_ISL_754069 | Nepal Korea Friendship Municipality Hospital | Nepal Health Research Council | Pradip Gyanwali et al |
| EPI_ISL_754062 | Kathmandu University | Nepal Health Research Council | Pradip Gyanwali et al |
| EPI_ISL_754063 | Kathmandu University | Nepal Health Research Council | Pradip Gyanwali et al |
| EPI_ISL_754064 | Kathmandu University | Nepal Health Research Council | Pradip Gyanwali et al |
| EPI_ISL_754065 | Kathmandu University | Nepal Health Research Council | Pradip Gyanwali et al |
| EPI_ISL_754067 | Kathmandu University | Nepal Health Research Council | Pradip Gyanwali et al |
| EPI_ISL_754073 | National Public Health Laboratory | Nepal Health Research Council | Pradip Gyanwali et al |
| EPI_ISL_754071 | Nepal Korea Friendship Municipality Hospital | Nepal Health Research Council | Pradip Gyanwali et al |
| EPI_ISL_516650 | Institute of Microbiology, Universidad San Francisco de Quito | Institute of Microbiology, Universidad San Francisco de Quito | Prado-Vivar et al |
| EPI_ISL_516651 | Institute of Microbiology, Universidad San Francisco de Quito | Institute of Microbiology, Universidad San Francisco de Quito | Prado-Vivar et al |

| | | | |
|---|---|---|---|
| EPI_ISL_516652 | Institute of Microbiology, Universidad San Francisco de Quito | Institute of Microbiology, Universidad San Francisco de Quito | Prado-Vivar et al |
| EPI_ISL_421670 | National Influenza Center, Indian Council of Medical Research - National Institute of Virology | Indian Council of Medical Research-National Institute of Virology, Microbial Containment Complex | Pragya D. Yadav et al |
| EPI_ISL_1061417 | Indian Council of Medical Research-National Institute of Virology, Microbial Containment Complex | Indian Council of Medical Research-National Institute of Virology, Microbial Containment Complex | Pragya D. Yadav et al |
| EPI_ISL_637099 | Indian Council of Medical Research-National Institute of Virology, Microbial Containment Complex | Indian Council of Medical Research-National Institute of Virology, Microbial Containment Complex | Pragya D. Yadav et al |
| EPI_ISL_637102 | Indian Council of Medical Research-National Institute of Virology, Microbial Containment Complex | Indian Council of Medical Research-National Institute of Virology, Microbial Containment Complex | Pragya D. Yadav et al |
| EPI_ISL_538348 | Kingston Health Sciences Centre / Queen's University | Ontario Institute for Cancer Research | Prameet M. Sheth et al |
| EPI_ISL_538349 | Kingston Health Sciences Centre / Queen's University | Ontario Institute for Cancer Research | Prameet M. Sheth et al |
| EPI_ISL_435089 | National Centre for Disease control (NCDC), CSIR-Institute of Genomics and Integrative Biology (CSIR-IGIB) | NCDC/CSIR-IGIB | Pramod Kumar et al |
| EPI_ISL_435073 | National Centre for Disease control (NCDC), CSIR-Institute of Genomics and Integrative Biology (CSIR-IGIB) | NCDC/CSIR-IGIB | Pramod Kumar et al |
| EPI_ISL_482628 | National Centre for Disease control (NCDC) | NCDC/CSIR-IGIB | Pramod Kumar# et al |
| EPI_ISL_482629 | National Centre for Disease control (NCDC) | NCDC/CSIR-IGIB | Pramod Kumar# et al |
| EPI_ISL_482630 | National Centre for Disease control (NCDC) | NCDC/CSIR-IGIB | Pramod Kumar# et al |
| EPI_ISL_482632 | National Centre for Disease control (NCDC) | NCDC/CSIR-IGIB | Pramod Kumar# et al |
| EPI_ISL_482641 | National Centre for Disease control (NCDC) | NCDC/CSIR-IGIB | Pramod Kumar# et al |
| EPI_ISL_436453 | National Centre for Disease control (NCDC) | NCDC/CSIR-IGIB | Pramod Kumar# et al |
| EPI_ISL_436463 | National Centre for Disease control (NCDC) | NCDC/CSIR-IGIB | Pramod Kumar# et al |
| EPI_ISL_1499201 | Associação Fundo de Incentivo à Pesquisa (AFIP) | Associação Fundo de Incentivo à Pesquisa (AFIP) | Priscila Farias Tempaku et al |
| EPI_ISL_1499202 | Associação Fundo de Incentivo à Pesquisa (AFIP) | Associação Fundo de Incentivo à Pesquisa (AFIP) | Priscila Farias Tempaku et al |
| EPI_ISL_1499297 | Associação Fundo de Incentivo à Pesquisa (AFIP) | Associação Fundo de Incentivo à Pesquisa (AFIP) | Priscila Farias Tempaku et al |
| EPI_ISL_1499205 | Associação Fundo de Incentivo à Pesquisa (AFIP) | Associação Fundo de Incentivo à Pesquisa (AFIP) | Priscila Farias Tempaku et al |
| EPI_ISL_1499298 | Associação Fundo de Incentivo à Pesquisa (AFIP) | Associação Fundo de Incentivo à Pesquisa (AFIP) | Priscila Farias Tempaku et al |
| EPI_ISL_1499299 | Associação Fundo de Incentivo à Pesquisa (AFIP) | Associação Fundo de Incentivo à Pesquisa (AFIP) | Priscila Farias Tempaku et al |
| EPI_ISL_1499114 | Associação Fundo de Incentivo à Pesquisa (AFIP) | Associação Fundo de Incentivo à Pesquisa (AFIP) | Priscila Farias Tempaku et al |
| EPI_ISL_1499300 | Associação Fundo de Incentivo à Pesquisa (AFIP) | Associação Fundo de Incentivo à Pesquisa (AFIP) | Priscila Farias Tempaku et al |
| EPI_ISL_1654211 | Fondation Congolaise pour la recherche medicale (FCRM), Francine Ntoumi | Institute of Tropical Medicine | Prof. Francine Ntoumi et al |
| EPI_ISL_1654212 | Fondation Congolaise pour la recherche medicale (FCRM), Francine Ntoumi | Institute of Tropical Medicine | Prof. Francine Ntoumi et al |
| EPI_ISL_1654213 | Fondation Congolaise pour la recherche medicale (FCRM), Francine Ntoumi | Institute of Tropical Medicine | Prof. Francine Ntoumi et al |
| EPI_ISL_1654214 | Fondation Congolaise pour la recherche medicale (FCRM), Francine Ntoumi | Institute of Tropical Medicine | Prof. Francine Ntoumi et al |

| | | | |
|---|---|---|---|
| EPI_ISL_1654217 | Fondation Congolaise pour la recherche medicale (FCRM), Francine Ntoumi | Institute of Tropical Medicine | Prof. Francine Ntoumi et al |
| EPI_ISL_1654218 | Fondation Congolaise pour la recherche medicale (FCRM), Francine Ntoumi | Institute of Tropical Medicine | Prof. Francine Ntoumi et al |
| EPI_ISL_968754 | BCCDC Public Health Laboratory | BCCDC Public Health Laboratory | Prystajecky Natalie et al |
| EPI_ISL_969102 | BCCDC Public Health Laboratory | BCCDC Public Health Laboratory | Prystajecky Natalie et al |
| EPI_ISL_969908 | BCCDC Public Health Laboratory | BCCDC Public Health Laboratory | Prystajecky Natalie et al |
| EPI_ISL_968466 | BCCDC Public Health Laboratory | BCCDC Public Health Laboratory | Prystajecky Natalie et al |
| EPI_ISL_968521 | BCCDC Public Health Laboratory | BCCDC Public Health Laboratory | Prystajecky Natalie et al |
| EPI_ISL_969755 | BCCDC Public Health Laboratory | BCCDC Public Health Laboratory | Prystajecky Natalie et al |
| EPI_ISL_976424 | BCCDC Public Health Laboratory | BCCDC Public Health Laboratory | Prystajecky Natalie et al |
| EPI_ISL_976425 | BCCDC Public Health Laboratory | BCCDC Public Health Laboratory | Prystajecky Natalie et al |
| EPI_ISL_974609 | BCCDC Public Health Laboratory | BCCDC Public Health Laboratory | Prystajecky Natalie et al |
| EPI_ISL_974610 | BCCDC Public Health Laboratory | BCCDC Public Health Laboratory | Prystajecky Natalie et al |
| EPI_ISL_1250695 | Wellington SCL (WN) | Institute of Environmental Science and Research (ESR) | Rachel Boyle et al |
| EPI_ISL_1082263 | LabPLUS | Institute of Environmental Science and Research (ESR) | Rachel Boyle et al |
| EPI_ISL_1172032 | LabPLUS | Institute of Environmental Science and Research (ESR) | Rachel Boyle et al |
| EPI_ISL_1172030 | LabPLUS | Institute of Environmental Science and Research (ESR) | Rachel Boyle et al |
| EPI_ISL_1172029 | LabPLUS | Institute of Environmental Science and Research (ESR) | Rachel Boyle et al |
| EPI_ISL_1250697 | Middlemore Hospital | Institute of Environmental Science and Research (ESR) | Rachel Boyle et al |
| EPI_ISL_1250700 | LabPLUS | Institute of Environmental Science and Research (ESR) | Rachel Boyle et al |
| EPI_ISL_1250699 | LabPLUS | Institute of Environmental Science and Research (ESR) | Rachel Boyle et al |
| EPI_ISL_1315318 | Middlemore Hospital | Institute of Environmental Science and Research (ESR) | Rachel Boyle et al |
| EPI_ISL_1469106 | LabPLUS | Institute of Environmental Science and Research (ESR) | Rachel Boyle et al |
| EPI_ISL_1621303 | LabPLUS | Institute of Environmental Science and Research (ESR) | Rachel Boyle et al |
| EPI_ISL_1621294 | LabPLUS | Institute of Environmental Science and Research (ESR) | Rachel Boyle et al |
| EPI_ISL_1621311 | Middlemore Hospital | Institute of Environmental Science and Research (ESR) | Rachel Boyle et al |
| EPI_ISL_1621325 | LabPLUS | Institute of Environmental Science and Research (ESR) | Rachel Boyle et al |
| EPI_ISL_1621305 | LabPLUS | Institute of Environmental Science and Research (ESR) | Rachel Boyle et al |
| EPI_ISL_1621304 | LabPLUS | Institute of Environmental Science and Research (ESR) | Rachel Boyle et al |
| EPI_ISL_1651917 | WSSE w Gdańsku | 1. Academic Center for Pathomorphological and Genetic-Molecular Diagnostics ltd, Bialystok, Poland 2. National Institute of Public Health - National Institute of Hygiene, Warsaw, Poland | Radosław Charkiewicz et al |

| | | | |
|---|---|---|---|
| EPI_ISL_1651931 | WSSE w Gdańsku | 1. Academic Center for Pathomorphological and Genetic-Molecular Diagnostics ltd, Bialystok, Poland 2. National Institute of Public Health - National Institute of Hygiene, Warsaw, Poland | Radosław Charkiewicz et al |
| EPI_ISL_1659587 | National Institute of Public Health - National Institute of Hygiene | 1. Academic Center for Pathomorphological and Genetic-Molecular Diagnostics ltd, Bialystok, Poland 2. National Institute of Public Health - National Institute of Hygiene, Warsaw, Poland | Radosław Charkiewicz et al |
| EPI_ISL_1659598 | National Institute of Public Health - National Institute of Hygiene | 1. Academic Center for Pathomorphological and Genetic-Molecular Diagnostics ltd, Bialystok, Poland 2. National Institute of Public Health - National Institute of Hygiene, Warsaw, Poland | Radosław Charkiewicz et al |
| EPI_ISL_1659607 | National Institute of Public Health - National Institute of Hygiene | 1. Academic Center for Pathomorphological and Genetic-Molecular Diagnostics ltd, Bialystok, Poland 2. National Institute of Public Health - National Institute of Hygiene, Warsaw, Poland | Radosław Charkiewicz et al |
| EPI_ISL_512880 | Pathogen Genomics Lab King Abdullah University of Science and Technology (KAUST) | Pathogen Genomics Lab King Abdullah University of Science and Technology(KAUST) | Raeece Naeem et al |
| EPI_ISL_512889 | Pathogen Genomics Lab King Abdullah University of Science and Technology (KAUST) | Pathogen Genomics Lab King Abdullah University of Science and Technology(KAUST) | Raeece Naeem et al |
| EPI_ISL_512891 | Pathogen Genomics Lab King Abdullah University of Science and Technology (KAUST) | Pathogen Genomics Lab King Abdullah University of Science and Technology(KAUST) | Raeece Naeem et al |
| EPI_ISL_512893 | Pathogen Genomics Lab King Abdullah University of Science and Technology (KAUST) | Pathogen Genomics Lab King Abdullah University of Science and Technology(KAUST) | Raeece Naeem et al |
| EPI_ISL_512894 | Pathogen Genomics Lab King Abdullah University of Science and Technology (KAUST) | Pathogen Genomics Lab King Abdullah University of Science and Technology(KAUST) | Raeece Naeem et al |
| EPI_ISL_512897 | Pathogen Genomics Lab King Abdullah University of Science and Technology (KAUST) | Pathogen Genomics Lab King Abdullah University of Science and Technology(KAUST) | Raeece Naeem et al |
| EPI_ISL_678191 | Pathogen Genomics Lab King Abdullah University of Science and Technology (KAUST) | Pathogen Genomics Lab King Abdullah University of Science and Technology(KAUST) | Raeece Naeem et al |
| EPI_ISL_513036 | Pathogen Genomics Lab King Abdullah University of Science and Technology (KAUST) | Pathogen Genomics Lab King Abdullah University of Science and Technology(KAUST) | Rahul P Salunke et al |
| EPI_ISL_513049 | Pathogen Genomics Lab King Abdullah University of Science and Technology (KAUST) | Pathogen Genomics Lab King Abdullah University of Science and Technology(KAUST) | Rahul P Salunke et al |
| EPI_ISL_513054 | Pathogen Genomics Lab King Abdullah University of Science and Technology (KAUST) | Pathogen Genomics Lab King Abdullah University of Science and Technology(KAUST) | Rahul P Salunke et al |
| EPI_ISL_513055 | Pathogen Genomics Lab King Abdullah University of Science and Technology (KAUST) | Pathogen Genomics Lab King Abdullah University of Science and Technology(KAUST) | Rahul P Salunke et al |
| EPI_ISL_513198 | Pathogen Genomics Lab King Abdullah University of Science and Technology (KAUST) | Pathogen Genomics Lab King Abdullah University of Science and Technology(KAUST) | Rahul P Salunke et al |
| EPI_ISL_528422 | TNMC & BYL NAIR CH. HOSPITAL | Institute of Genomics and Integrative Biology - Council of Scientific and Industrial Research | Rajesh Pandey et al |
| EPI_ISL_459913 | Devki Devi Foundation, a unit of Max Healthcare | CSIR-IGIB/Max | Rajesh Pandey# et al |
| EPI_ISL_459914 | Devki Devi Foundation, a unit of Max Healthcare | CSIR-IGIB/Max | Rajesh Pandey# et al |
| EPI_ISL_459916 | Devki Devi Foundation, a unit of Max Healthcare | CSIR-IGIB/Max | Rajesh Pandey# et al |
| EPI_ISL_459924 | Devki Devi Foundation, a unit of Max Healthcare | CSIR-IGIB/Max | Rajesh Pandey# et al |
| EPI_ISL_412972 | Instituto Nacional de Enfermedades Respiratorias | Instituto de Diagnostico y Referencia Epidemiologicos (INDRE) | Ramirez-Gonzalez Ernesto et al |

| | | | |
|---|---|---|---|
| EPI_ISL_452139 | Instituto de Diagnostico y Referencia Epidemiologicos (INDRE) | Instituto de diagnóstico y Referencia Epidemiologicos (INDRE) | Ramirez-Gonzalez Ernesto et al |
| EPI_ISL_985237 | Biorepository and Clinical Virology Laboratory | Ozer Lab | Ramon Lorenzo-Redondo et al |
| EPI_ISL_985238 | Biorepository and Clinical Virology Laboratory | Ozer Lab | Ramon Lorenzo-Redondo et al |
| EPI_ISL_1191597 | Virology Department, Victoria Hospital, Plaine-Wilhems, Mauritius | National Institute for Communicable Diseases of the National Health Laboratory Service | Ramuth M et al |
| EPI_ISL_1191600 | Virology Department, Victoria Hospital, Plaine-Wilhems, Mauritius | National Institute for Communicable Diseases of the National Health Laboratory Service | Ramuth M et al |
| EPI_ISL_1191602 | Virology Department, Victoria Hospital, Plaine-Wilhems, Mauritius | National Institute for Communicable Diseases of the National Health Laboratory Service | Ramuth M et al |
| EPI_ISL_1191604 | Virology Department, Victoria Hospital, Plaine-Wilhems, Mauritius | National Institute for Communicable Diseases of the National Health Laboratory Service | Ramuth M et al |
| EPI_ISL_1191605 | Virology Department, Victoria Hospital, Plaine-Wilhems, Mauritius | National Institute for Communicable Diseases of the National Health Laboratory Service | Ramuth M et al |
| EPI_ISL_569988 | Unity Health Toronto | Ontario Institute for Cancer Research | Ramzi Fattouh et al |
| EPI_ISL_526267 | Unity Health Toronto | Ontario Institute for Cancer Research | Ramzi Fattouh et al |
| EPI_ISL_1248874 | Unity Health Toronto | Ontario Institute for Cancer Research | Ramzi Fattouh et al |
| EPI_ISL_1248878 | Unity Health Toronto | Ontario Institute for Cancer Research | Ramzi Fattouh et al |
| EPI_ISL_1248880 | Unity Health Toronto | Ontario Institute for Cancer Research | Ramzi Fattouh et al |
| EPI_ISL_1248884 | Unity Health Toronto | Ontario Institute for Cancer Research | Ramzi Fattouh et al |
| EPI_ISL_1248890 | Unity Health Toronto | Ontario Institute for Cancer Research | Ramzi Fattouh et al |
| EPI_ISL_1248891 | Unity Health Toronto | Ontario Institute for Cancer Research | Ramzi Fattouh et al |
| EPI_ISL_1248887 | Unity Health Toronto | Ontario Institute for Cancer Research | Ramzi Fattouh et al |
| EPI_ISL_1045717 | Randox Laboratories | Wellcome Sanger Institute for the COVID-19 Genomics UK (COG-UK) Consortium | Randox Laboratories et al |
| EPI_ISL_1631841 | Randox Laboratories | Wellcome Sanger Institute for the COVID-19 Genomics UK (COG-UK) Consortium | Randox Laboratories et al |
| EPI_ISL_1631888 | Randox Laboratories | Wellcome Sanger Institute for the COVID-19 Genomics UK (COG-UK) Consortium | Randox Laboratories et al |
| EPI_ISL_1632206 | Randox Laboratories | Wellcome Sanger Institute for the COVID-19 Genomics UK (COG-UK) Consortium | Randox Laboratories et al |
| EPI_ISL_410301 | National Influenza Centre, National Public Health Laboratory, Kathmandu, Nepal | The University of Hong Kong | Ranjit Sah et al |
| EPI_ISL_444851 | Department of Virus and Microbiological Special Diagnostics, Statens Serum Institut, Copenhagen, Denmark, Artillerivej 5, 2300 Copenahgen S | Albertsen lab, Department of Chemistry and Bioscience, Aalborg University, Denmark | Rasmus Kirkegaard et al |
| EPI_ISL_444919 | Department of Virus and Microbiological Special Diagnostics, Statens Serum Institut, Copenhagen, Denmark, Artillerivej 5, 2300 Copenahgen S | Albertsen lab, Department of Chemistry and Bioscience, Aalborg University, Denmark | Rasmus Kirkegaard et al |
| EPI_ISL_429498 | Department of Virus and Microbiological Special Diagnostics, Statens Serum Institut, Copenhagen, Denmark, Artillerivej 5, 2300 Copenahgen S | Albertsen lab, Department of Chemistry and Bioscience, Aalborg University, Denmark | Rasmus Kirkegaard et al |

| | | | |
|---|---|---|---|
| EPI_ISL_527019 | Area of Virology, Serology and Virology Division (SAViD), New South Wales Health Pathology Randwick | Area of Virology, Serology and Virology Division (SAViD), New South Wales Health Pathology Randwick | Rawlinson et al |
| EPI_ISL_527032 | Area of Virology, Serology and Virology Division (SAViD), New South Wales Health Pathology Randwick | Area of Virology, Serology and Virology Division (SAViD), New South Wales Health Pathology Randwick | Rawlinson et al |
| EPI_ISL_877727 | Clinical Molecular Microbiology Laboratory, UNC Hospital | Dirk Dittmer | Razia Moorad et al |
| EPI_ISL_678253 | Institute for Lung Diseases in Children - Skopje | Research Center for Genetic Engineering and Biotechnology Georgi D. Efremov" , Macedonian Academy of Sciences and Arts | RCGEB - MASA et al |
| EPI_ISL_678256 | General Hospital - Ohrid | Research Center for Genetic Engineering and Biotechnology Georgi D. Efremov" , Macedonian Academy of Sciences and Arts | RCGEB - MASA et al |
| EPI_ISL_678257 | PHI Institute of Nephrology - Struga | Research Center for Genetic Engineering and Biotechnology Georgi D. Efremov" , Macedonian Academy of Sciences and Arts | RCGEB - MASA et al |
| EPI_ISL_678259 | Clinical Hospital - Shtip | Research Center for Genetic Engineering and Biotechnology Georgi D. Efremov" , Macedonian Academy of Sciences and Arts | RCGEB - MASA et al |
| EPI_ISL_918492 | General Hospital - Kumanovo | Research Center for Genetic Engineering and Biotechnology Georgi D. Efremov" , Macedonian Academy of Sciences and Arts | RCGEB - MASA et al |
| EPI_ISL_918488 | General Hospital - Strumica | Research Center for Genetic Engineering and Biotechnology Georgi D. Efremov" , Macedonian Academy of Sciences and Arts | RCGEB - MASA et al |
| EPI_ISL_933579 | General Hospital - Strumica | Research Center for Genetic Engineering and Biotechnology Georgi D. Efremov" , Macedonian Academy of Sciences and Arts | RCGEB - MASA et al |
| EPI_ISL_933717 | General Hospital - Strumica | Research Center for Genetic Engineering and Biotechnology Georgi D. Efremov" , Macedonian Academy of Sciences and Arts | RCGEB - MASA et al |
| EPI_ISL_933772 | General Hospital - Bitola | Research Center for Genetic Engineering and Biotechnology Georgi D. Efremov" , Macedonian Academy of Sciences and Arts | RCGEB - MASA et al |
| EPI_ISL_933786 | General Hospital - Bitola | Research Center for Genetic Engineering and Biotechnology Georgi D. Efremov" , Macedonian Academy of Sciences and Arts | RCGEB - MASA et al |
| EPI_ISL_934314 | General Hospital - Bitola | Research Center for Genetic Engineering and Biotechnology Georgi D. Efremov" , Macedonian Academy of Sciences and Arts | RCGEB - MASA et al |
| EPI_ISL_934413 | General Hospital - Shtip | Research Center for Genetic Engineering and Biotechnology Georgi D. Efremov" , Macedonian Academy of Sciences and Arts | RCGEB - MASA et al |
| EPI_ISL_677708 | Center for public health - Skopje | Research Center for Genetic Engineering and Biotechnology Georgi D. Efremov" , Macedonian Academy of Sciences and Arts | RCGEB - MASA et al |
| EPI_ISL_516428 | Clinical Hospital - Shtip | Research Center for Genetic Engineering and Biotechnology Georgi D. Efremov" , Macedonian Academy of Sciences and Arts | RCGEB - MASA et al |
| EPI_ISL_516429 | Clinical Hospital - Shtip | Research Center for Genetic Engineering and Biotechnology Georgi D. Efremov" , Macedonian Academy of Sciences and Arts | RCGEB - MASA et al |
| EPI_ISL_677716 | Clinical Hospital - Shtip | Research Center for Genetic Engineering and Biotechnology Georgi D. Efremov" , Macedonian Academy of Sciences and Arts | RCGEB - MASA et al |
| EPI_ISL_677674 | General Hospital - Prilep | Research Center for Genetic Engineering and Biotechnology Georgi D. Efremov" , Macedonian Academy of Sciences and Arts | RCGEB - MASA et al |
| EPI_ISL_677724 | Clinical Hospital - Shtip | Research Center for Genetic Engineering and Biotechnology Georgi D. Efremov" , Macedonian Academy of Sciences and Arts | RCGEB - MASA et al |
| EPI_ISL_678248 | Center for public health - Skopje | Research Center for Genetic Engineering and Biotechnology Georgi D. Efremov" , Macedonian Academy of Sciences and Arts | RCGEB - MASA et al |

| | | | |
|---|---|---|---|
| EPI_ISL_678249 | Center for public health - Skopje | Research Center for Genetic Engineering and Biotechnology Georgi D. Efremov" , Macedonian Academy of Sciences and Arts | RCGEB - MASA et al |
| EPI_ISL_678250 | Clinical Hospital - Shtip | Research Center for Genetic Engineering and Biotechnology Georgi D. Efremov" , Macedonian Academy of Sciences and Arts | RCGEB - MASA et al |
| EPI_ISL_933566 | General Hospital - Bitola | Research Center for Genetic Engineering and Biotechnology Georgi D. Efremov" , Macedonian Academy of Sciences and Arts | RCGEB - MASA et al |
| EPI_ISL_1315064 | Armed Forces Medical Research Laboratories and Blood Bank, Egypt | Department of Microbial Biotechnology, Genetic Engineering Division, National Research Centre, | Reham Dawood et al |
| EPI_ISL_1585247 | GHC Genetics, s.r.o. | GHC Genetics, s.r.o. | Renata Michalovská et al |
| EPI_ISL_467300 | General Hospital Abdulah Nakas | Alea Genetic Center | Rijad Konjhodzic et al |
| EPI_ISL_1653935 | Alea Genetic Center | Alea Genetic Center | Rijad Konjhodzic et al |
| EPI_ISL_910329 | ALEA dr Kandic | Alea Genetic Centre | Rijad Konjhodzic et al |
| EPI_ISL_922076 | General Hospital #Abdulah Nakas Sarajevo | Alea Genetic Centre | Rijad Konjhodzic et al |
| EPI_ISL_910334 | ALEA dr Kandic | Alea Genetic Centre | Rijad Konjhodzic et al |
| EPI_ISL_560991 | RSUD Dr. Soetomo | Institute of Tropical Disease, Universitas Airlangga | Rima R Prasetya et al |
| EPI_ISL_1165070 | Siti Khodijah Hospital | Institute of Tropical Disease, Universitas Airlangga | Rima R Prasetya et al |
| EPI_ISL_1324435 | National Food and Veterinary Risk Assessment Institute | Vilnius University Hospital Santaros Klinikos, Center of Laboratory Medicine | Rimvydas Norvilas et al |
| EPI_ISL_1041030 | Dr. Cipto Mangunkusumo (RSCM) Hospital | Faculty of Medicine, Universitas Indonesia | Rina La Distia Nora et al |
| EPI_ISL_450509 | Rafik Hariri University Hospital | Rafik Hariri University Hospital | Rita Feghali et al |
| EPI_ISL_450512 | Rafik Hariri University Hospital | Rafik Hariri University Hospital | Rita Feghali et al |
| EPI_ISL_450514 | Rafik Hariri University Hospital | Rafik Hariri University Hospital | Rita Feghali et al |
| EPI_ISL_450515 | Rafik Hariri University Hospital | Rafik Hariri University Hospital | Rita Feghali et al |
| EPI_ISL_671974 | Laboratorio de Virología y Microbiología Molecular, Depto. de Microbiología, Facultad de Medicina, Universidad de El Salvador/INS-laboratorio de Ref. Ministerio de Salud | Laboratorio de Virología y Microbiología Molecular, Depto. de Microbiología, Facultad de Medicina, Universidad de El Salvador/INS-laboratorio de Ref. Ministerio de Salud | Rivera NR et al |
| EPI_ISL_1187189 | Lighthouse Lab in Cambridge | Wellcome Sanger Institute for the COVID-19 Genomics UK (COG-UK) Consortium | Rob Howes et al |
| EPI_ISL_1327496 | Lighthouse Lab in Cambridge | Wellcome Sanger Institute for the COVID-19 Genomics UK (COG-UK) Consortium | Rob Howes et al |
| EPI_ISL_1327150 | Lighthouse Lab in Cambridge | Wellcome Sanger Institute for the COVID-19 Genomics UK (COG-UK) Consortium | Rob Howes et al |
| EPI_ISL_1455639 | Lighthouse Lab in Cambridge | Wellcome Sanger Institute for the COVID-19 Genomics UK (COG-UK) Consortium | Rob Howes et al |
| EPI_ISL_1653315 | Lighthouse Lab in Cambridge | Wellcome Sanger Institute for the COVID-19 Genomics UK (COG-UK) Consortium | Rob Howes et al |
| EPI_ISL_1653472 | Lighthouse Lab in Cambridge | Wellcome Sanger Institute for the COVID-19 Genomics UK (COG-UK) Consortium | Rob Howes et al |

| | | | |
|---|---|---|---|
| EPI_ISL_673528 | Lighthouse Lab in Cambridge | Wellcome Sanger Institute for the COVID-19 Genomics UK (COG-UK) Consortium | Rob Howes et al |
| EPI_ISL_710574 | University Hospital Dubrava | Ruđer Boškovic Institute; Forensic Science Centre Ivan Vučetić; University of Zagreb Faculty of Science | Robert Belužić et al |
| EPI_ISL_710568 | University Hospital Dubrava | Ruđer Boškovic Institute; Forensic Science Centre Ivan Vučetić; University of Zagreb Faculty of Science | Robert Belužić et al |
| EPI_ISL_710549 | University Hospital Dubrava | Ruđer Boškovic Institute; Forensic Science Centre Ivan Vučetić; University of Zagreb Faculty of Science | Robert Belužić et al |
| EPI_ISL_710555 | University Hospital Dubrava | Ruđer Boškovic Institute; Forensic Science Centre Ivan Vučetić; University of Zagreb Faculty of Science | Robert Belužić et al |
| EPI_ISL_710548 | University Hospital Dubrava | Ruđer Boškovic Institute; Forensic Science Centre Ivan Vučetić; University of Zagreb Faculty of Science | Robert Belužić et al |
| EPI_ISL_710567 | University Hospital Dubrava | Ruđer Boškovic Institute; Forensic Science Centre Ivan Vučetić; University of Zagreb Faculty of Science | Robert Belužić et al |
| EPI_ISL_710566 | University Hospital Dubrava | Ruđer Boškovic Institute; Forensic Science Centre Ivan Vučetić; University of Zagreb Faculty of Science | Robert Belužić et al |
| EPI_ISL_710570 | University Hospital Dubrava | Ruđer Boškovic Institute; Forensic Science Centre Ivan Vučetić; University of Zagreb Faculty of Science | Robert Belužić et al |
| EPI_ISL_710565 | University Hospital Dubrava | Ruđer Boškovic Institute; Forensic Science Centre Ivan Vučetić; University of Zagreb Faculty of Science | Robert Belužić et al |
| EPI_ISL_710573 | University Hospital Dubrava | Ruđer Boškovic Institute; Forensic Science Centre Ivan Vučetić; University of Zagreb Faculty of Science | Robert Belužić et al |
| EPI_ISL_710547 | Sestre Milosrdnice University Hospital Center | Ruđer Boškovic Institute; Forensic Science Centre Ivan Vučetić; University of Zagreb Faculty of Science | Robert Belužić et al |
| EPI_ISL_710554 | Sestre Milosrdnice University Hospital Center | Ruđer Boškovic Institute; Forensic Science Centre Ivan Vučetić; University of Zagreb Faculty of Science | Robert Belužić et al |
| EPI_ISL_710556 | University Hospital Dubrava | Ruđer Boškovic Institute; Forensic Science Centre Ivan Vučetić; University of Zagreb Faculty of Science | Robert Belužić et al |
| EPI_ISL_710571 | University Hospital Dubrava | Ruđer Boškovic Institute; Forensic Science Centre Ivan Vučetić; University of Zagreb Faculty of Science | Robert Belužić et al |
| EPI_ISL_710560 | University Hospital Dubrava | Ruđer Boškovic Institute; Forensic Science Centre Ivan Vučetić; University of Zagreb Faculty of Science | Robert Belužić et al |
| EPI_ISL_710557 | University Hospital Dubrava | Ruđer Boškovic Institute; Forensic Science Centre Ivan Vučetić; University of Zagreb Faculty of Science | Robert Belužić et al |
| EPI_ISL_710572 | University Hospital Dubrava | Ruđer Boškovic Institute; Forensic Science Centre Ivan Vučetić; University of Zagreb Faculty of Science | Robert Belužić et al |
| EPI_ISL_710558 | University Hospital Dubrava | Ruđer Boškovic Institute; Forensic Science Centre Ivan Vučetić; University of Zagreb Faculty of Science | Robert Belužić et al |
| EPI_ISL_525467 | Universidad Iberoamericana | International Centre for Genetic Engineering and Biotechnology (ICGEB) and ARGO Open Lab Platform | Robert Paulino-Ramirez et al |
| EPI_ISL_525468 | Universidad Iberoamericana | International Centre for Genetic Engineering and Biotechnology (ICGEB) and ARGO Open Lab Platform | Robert Paulino-Ramirez et al |
| EPI_ISL_525469 | Universidad Iberoamericana | International Centre for Genetic Engineering and Biotechnology (ICGEB) and ARGO Open Lab Platform | Robert Paulino-Ramirez et al |

| | | | |
|---|---|---|---|
| EPI_ISL_523811 | Universidad Iberoamericana, Instituto de Medicina Tropical & Salud Global | International Centre for Genetic Engineering and Biotechnology (ICGEB) and ARGO Open Lab Platform | Robert Paulino-Ramirez et al |
| EPI_ISL_523812 | Universidad Iberoamericana, Instituto de Medicina Tropical & Salud Global | International Centre for Genetic Engineering and Biotechnology (ICGEB) and ARGO Open Lab Platform | Robert Paulino-Ramirez et al |
| EPI_ISL_1123293 | Abdul Malek Ukil Medical College, Noakhali | Central Biological Research Laboratory and Department of Biochemistry and Molecular Biology | Robiul Hasan Bhuiyan et al |
| EPI_ISL_413595 | Centre for Infectious Diseases and Microbiology Laboratory Services | NSW Health Pathology - Institute of Clinical Pathology and Medical Research; Westmead Hospital; University of Sydney | Rockett R et al |
| EPI_ISL_437620 | unknown | Faculty of Medicine | Rodpan et al |
| EPI_ISL_437618 | unknown | Faculty of Medicine | Rodpan et al |
| EPI_ISL_437615 | unknown | Faculty of Medicine | Rodpan et al |
| EPI_ISL_437614 | unknown | Faculty of Medicine | Rodpan et al |
| EPI_ISL_437613 | unknown | Faculty of Medicine | Rodpan et al |
| EPI_ISL_437612 | unknown | Faculty of Medicine | Rodpan et al |
| EPI_ISL_877766 | Thai Red Cross Emerging Infectious Diseases Health Science Centre, Chulalongkorn Hospital, Faculty of Medicine, Chulalongkorn University | Thai Red Cross Emerging Infectious Diseases Center and Faculty of Medicine, Chulalongkorn University | Rome Buathong et al |
| EPI_ISL_981004 | Thai Red Cross Emerging Infectious Diseases Health Science Centre, Chulalongkorn Hospital, Faculty of Medicine, Chulalongkorn University | Thai Red Cross Emerging Infectious Diseases Center and Faculty of Medicine, Chulalongkorn University | Rome Buathong et al |
| EPI_ISL_1007658 | Thai Red Cross Emerging Infectious Diseases Health Science Centre, Chulalongkorn Hospital, Faculty of Medicine, Chulalongkorn University | Thai Red Cross Emerging Infectious Diseases Center and Faculty of Medicine, Chulalongkorn University | Rome Buathong et al |
| EPI_ISL_1111083 | Laboratorio de Referencia Nacional de Virus Respiratorio. Instituto Nacional de Salud Perú | Laboratorio de Referencia Nacional de Enteropatógenos. Instituto Nacional de Salud del Perú | Ronnie Gavilan Chavez et al |
| EPI_ISL_1111112 | Laboratorio de Referencia Nacional de Virus Respiratorio. Instituto Nacional de Salud Perú | Laboratorio de Referencia Nacional de Enteropatógenos. Instituto Nacional de Salud del Perú | Ronnie Gavilan Chavez et al |
| EPI_ISL_1111137 | Laboratorio de Referencia Nacional de Virus Respiratorio. Instituto Nacional de Salud Perú | Laboratorio de Referencia Nacional de Enteropatógenos. Instituto Nacional de Salud del Perú | Ronnie Gavilan Chavez et al |
| EPI_ISL_1111142 | Laboratorio de Referencia Nacional de Virus Respiratorio. Instituto Nacional de Salud Perú | Laboratorio de Referencia Nacional de Enteropatógenos. Instituto Nacional de Salud del Perú | Ronnie Gavilan Chavez et al |
| EPI_ISL_1111143 | Laboratorio de Referencia Nacional de Virus Respiratorio. Instituto Nacional de Salud Perú | Laboratorio de Referencia Nacional de Enteropatógenos. Instituto Nacional de Salud del Perú | Ronnie Gavilan Chavez et al |
| EPI_ISL_1111151 | Laboratorio de Referencia Nacional de Virus Respiratorio. Instituto Nacional de Salud Perú | Laboratorio de Referencia Nacional de Enteropatógenos. Instituto Nacional de Salud del Perú | Ronnie Gavilan Chavez et al |
| EPI_ISL_1111152 | Laboratorio de Referencia Nacional de Virus Respiratorio. Instituto Nacional de Salud Perú | Laboratorio de Referencia Nacional de Enteropatógenos. Instituto Nacional de Salud del Perú | Ronnie Gavilan Chavez et al |
| EPI_ISL_1111169 | Laboratorio de Referencia Nacional de Virus Respiratorio. Instituto Nacional de Salud Perú | Laboratorio de Referencia Nacional de Enteropatógenos. Instituto Nacional de Salud del Perú | Ronnie Gavilan Chavez et al |
| EPI_ISL_1111214 | Laboratorio de Referencia Nacional de Virus Respiratorio. Instituto Nacional de Salud Perú | Laboratorio de Referencia Nacional de Enteropatógenos. Instituto Nacional de Salud del Perú | Ronnie Gavilan Chavez et al |
| EPI_ISL_1111218 | Laboratorio de Referencia Nacional de Virus Respiratorio. Instituto Nacional de Salud Perú | Laboratorio de Referencia Nacional de Enteropatógenos. Instituto Nacional de Salud del Perú | Ronnie Gavilan Chavez et al |
| EPI_ISL_1111220 | Laboratorio de Referencia Nacional de Virus Respiratorio. Instituto Nacional de Salud Perú | Laboratorio de Referencia Nacional de Enteropatógenos. Instituto Nacional de Salud del Perú | Ronnie Gavilan Chavez et al |

| | | | |
|---|---|---|---|
| EPI_ISL_1111223 | Laboratorio de Referencia Nacional de Virus Respiratorio. Instituto Nacional de Salud Perú | Laboratorio de Referencia Nacional de Enteropatógenos. Instituto Nacional de Salud del Perú | Ronnie Gavilan Chavez et al |
| EPI_ISL_1111242 | Laboratorio de Referencia Nacional de Virus Respiratorio. Instituto Nacional de Salud Perú | Laboratorio de Referencia Nacional de Enteropatógenos. Instituto Nacional de Salud del Perú | Ronnie Gavilan Chavez et al |
| EPI_ISL_1111249 | Laboratorio de Referencia Nacional de Virus Respiratorio. Instituto Nacional de Salud Perú | Laboratorio de Referencia Nacional de Enteropatógenos. Instituto Nacional de Salud del Perú | Ronnie Gavilan Chavez et al |
| EPI_ISL_1111252 | Laboratorio de Referencia Nacional de Virus Respiratorio. Instituto Nacional de Salud Perú | Laboratorio de Referencia Nacional de Enteropatógenos. Instituto Nacional de Salud del Perú | Ronnie Gavilan Chavez et al |
| EPI_ISL_1111266 | Laboratorio de Referencia Nacional de Virus Respiratorio. Instituto Nacional de Salud Perú | Laboratorio de Referencia Nacional de Enteropatógenos. Instituto Nacional de Salud del Perú | Ronnie Gavilan Chavez et al |
| EPI_ISL_1111267 | Laboratorio de Referencia Nacional de Virus Respiratorio. Instituto Nacional de Salud Perú | Laboratorio de Referencia Nacional de Enteropatógenos. Instituto Nacional de Salud del Perú | Ronnie Gavilan Chavez et al |
| EPI_ISL_1111271 | Laboratorio de Referencia Nacional de Virus Respiratorio. Instituto Nacional de Salud Perú | Laboratorio de Referencia Nacional de Enteropatógenos. Instituto Nacional de Salud del Perú | Ronnie Gavilan Chavez et al |
| EPI_ISL_1111275 | Laboratorio de Referencia Nacional de Virus Respiratorio. Instituto Nacional de Salud Perú | Laboratorio de Referencia Nacional de Enteropatógenos. Instituto Nacional de Salud del Perú | Ronnie Gavilan Chavez et al |
| EPI_ISL_1111312 | Laboratorio de Referencia Nacional de Virus Respiratorio. Instituto Nacional de Salud Perú | Laboratorio de Referencia Nacional de Enteropatógenos. Instituto Nacional de Salud del Perú | Ronnie Gavilan Chavez et al |
| EPI_ISL_1111374 | Laboratorio de Referencia Nacional de Virus Respiratorio. Instituto Nacional de Salud Perú | Laboratorio de Referencia Nacional de Enteropatógenos. Instituto Nacional de Salud del Perú | Ronnie Gavilan Chavez et al |
| EPI_ISL_1111384 | Laboratorio de Referencia Nacional de Virus Respiratorio. Instituto Nacional de Salud Perú | Laboratorio de Referencia Nacional de Enteropatógenos. Instituto Nacional de Salud del Perú | Ronnie Gavilan Chavez et al |
| EPI_ISL_1111397 | Laboratorio de Referencia Nacional de Enteropatógenos. Instituto Nacional de Salud del Perú | Laboratorio de Referencia Nacional de Enteropatógenos. Instituto Nacional de Salud del Perú | Ronnie Gavilan Chavez et al |
| EPI_ISL_1111414 | Laboratorio de Referencia Nacional de Enteropatógenos. Instituto Nacional de Salud del Perú | Laboratorio de Referencia Nacional de Enteropatógenos. Instituto Nacional de Salud del Perú | Ronnie Gavilan Chavez et al |
| EPI_ISL_1111415 | Laboratorio de Referencia Nacional de Enteropatógenos. Instituto Nacional de Salud del Perú | Laboratorio de Referencia Nacional de Enteropatógenos. Instituto Nacional de Salud del Perú | Ronnie Gavilan Chavez et al |
| EPI_ISL_1111450 | Laboratorio de Referencia Nacional de Virus Respiratorio. Instituto Nacional de Salud Perú | Laboratorio de Referencia Nacional de Enteropatógenos. Instituto Nacional de Salud del Perú | Ronnie Gavilan Chavez et al |
| EPI_ISL_1111451 | Laboratorio de Referencia Nacional de Virus Respiratorio. Instituto Nacional de Salud Perú | Laboratorio de Referencia Nacional de Enteropatógenos. Instituto Nacional de Salud del Perú | Ronnie Gavilan Chavez et al |
| EPI_ISL_1111461 | Laboratorio de Referencia Nacional de Virus Respiratorio. Instituto Nacional de Salud Perú | Laboratorio de Referencia Nacional de Enteropatógenos. Instituto Nacional de Salud del Perú | Ronnie Gavilan Chavez et al |
| EPI_ISL_1111467 | Laboratorio de Referencia Nacional de Virus Respiratorio. Instituto Nacional de Salud Perú | Laboratorio de Referencia Nacional de Enteropatógenos. Instituto Nacional de Salud del Perú | Ronnie Gavilan Chavez et al |
| EPI_ISL_1111472 | Laboratorio de Referencia Nacional de Virus Respiratorio. Instituto Nacional de Salud Perú | Laboratorio de Referencia Nacional de Enteropatógenos. Instituto Nacional de Salud del Perú | Ronnie Gavilan Chavez et al |
| EPI_ISL_1111474 | Laboratorio de Referencia Nacional de Virus Respiratorio. Instituto Nacional de Salud Perú | Laboratorio de Referencia Nacional de Enteropatógenos. Instituto Nacional de Salud del Perú | Ronnie Gavilan Chavez et al |
| EPI_ISL_1111475 | Laboratorio de Referencia Nacional de Virus Respiratorio. Instituto Nacional de Salud Perú | Laboratorio de Referencia Nacional de Enteropatógenos. Instituto Nacional de Salud del Perú | Ronnie Gavilan Chavez et al |
| EPI_ISL_1111483 | Laboratorio de Referencia Nacional de Virus Respiratorio. Instituto Nacional de Salud Perú | Laboratorio de Referencia Nacional de Enteropatógenos. Instituto Nacional de Salud del Perú | Ronnie Gavilan Chavez et al |

| | | | |
|---|---|---|---|
| EPI_ISL_1111484 | Laboratorio de Referencia Nacional de Virus Respiratorio. Instituto Nacional de Salud Perú | Laboratorio de Referencia Nacional de Enteropatógenos. Instituto Nacional de Salud del Perú | Ronnie Gavilan Chavez et al |
| EPI_ISL_1111486 | Laboratorio de Referencia Nacional de Virus Respiratorio. Instituto Nacional de Salud Perú | Laboratorio de Referencia Nacional de Enteropatógenos. Instituto Nacional de Salud del Perú | Ronnie Gavilan Chavez et al |
| EPI_ISL_1111490 | Laboratorio de Referencia Nacional de Virus Respiratorio. Instituto Nacional de Salud Perú | Laboratorio de Referencia Nacional de Enteropatógenos. Instituto Nacional de Salud del Perú | Ronnie Gavilan Chavez et al |
| EPI_ISL_1111496 | Laboratorio de Referencia Nacional de Virus Respiratorio. Instituto Nacional de Salud Perú | Laboratorio de Referencia Nacional de Enteropatógenos. Instituto Nacional de Salud del Perú | Ronnie Gavilan Chavez et al |
| EPI_ISL_1395790 | Laboratorio de salud pública, Facultad de Ciencias Exactas, UNLP | Área de Secuenciación del Laboratorio de Virología del Hospital de Niños Dr. Ricardo Gutierrez on behalf of 'Proyecto Argentino Interinstitucional de genomica de SARS-CoV-2' (PAIS Consortium) | Rosana Isabel Toro et al |
| EPI_ISL_537988 | Servicio de Microbiología. Hospital General Universitario de Castellón | SeqCOVID-SPAIN consortium/IBV(CSIC) | Rosario Moreno et al |
| EPI_ISL_794599 | Central Laboratories, Egyptian Ministry of Health and Population | Central Laboratories, Egyptian Ministry of Health and Population | Roshdy et al |
| EPI_ISL_794595 | Central Laboratories, Egyptian Ministry of Health and Population | Central Laboratories, Egyptian Ministry of Health and Population | Roshdy et al |
| EPI_ISL_794594 | Central Laboratories, Egyptian Ministry of Health and Population | Central Laboratories, Egyptian Ministry of Health and Population | Roshdy et al |
| EPI_ISL_794593 | Central Laboratories, Egyptian Ministry of Health and Population | Central Laboratories, Egyptian Ministry of Health and Population | Roshdy et al |
| EPI_ISL_981226 | National Virus Reference Laboratory | Irish Coronavirus Sequencing Consortium - Helixworks | Sachin Chalapati et al |
| EPI_ISL_981259 | National Virus Reference Laboratory | Irish Coronavirus Sequencing Consortium - Helixworks | Sachin Chalapati et al |
| EPI_ISL_981262 | National Virus Reference Laboratory | Irish Coronavirus Sequencing Consortium - Helixworks | Sachin Chalapati et al |
| EPI_ISL_981269 | National Virus Reference Laboratory | Irish Coronavirus Sequencing Consortium - Helixworks | Sachin Chalapati et al |
| EPI_ISL_981231 | National Virus Reference Laboratory | Irish Coronavirus Sequencing Consortium - Helixworks | Sachin Chalapati et al |
| EPI_ISL_935039 | Biolab Diagnostic Laboratories | Princess Haya Biotechnology Center, Jordan University of Science and Technology | Saied Jaradat et al |
| EPI_ISL_935041 | Biolab Diagnostic Laboratories | Princess Haya Biotechnology Center, Jordan University of Science and Technology | Saied Jaradat et al |
| EPI_ISL_935043 | Biolab Diagnostic Laboratories | Princess Haya Biotechnology Center/ Jordan University of Science & Technology | Saied Jaradat et al |
| EPI_ISL_525423 | Oman-National Influenza Center | Biotechnology & OMICs Laboratory | Sajjad Asaf et al |
| EPI_ISL_525426 | Oman-National Influenza Center | Biotechnology & OMICs Laboratory | Sajjad Asaf et al |
| EPI_ISL_525428 | Oman-National Influenza Center | Biotechnology & OMICs Laboratory | Sajjad Asaf et al |
| EPI_ISL_639739 | Centre of Nanotechnologies, INCD IMT-Bucuresti (National Institute for Research and Development in Microtechnologies - Bucharest) | Centre of Nanotechnologies, INCD IMT-Bucuresti (National Institute for Research and Development in Microtechnologies - Bucharest) | Salceanu et al |
| EPI_ISL_722201 | Vitalis Mostar | Alea Genetic Center | Salihefendic L. et al |
| EPI_ISL_722209 | Dom Zdravlja Sarajevo | Alea Genetic Center | Salihefendic L. et al |
| EPI_ISL_903383 | MOH - Jaber Al-Ahmad Hospital (Innovation Research Laboratory) | MOH - Jaber Al-Ahmad Hospital (Innovation Research Laboratory) | Salman Al-Sabah et al |
| EPI_ISL_903376 | MOH - Jaber Al-Ahmad Hospital (Innovation Research Laboratory) | MOH - Jaber Al-Ahmad Hospital (Innovation Research Laboratory) | Salman Al-Sabah et al |

| | | | |
|---|---|---|---|
| EPI_ISL_903384 | MOH - Jaber Al-Ahmad Hospital (Innovation Research Laboratory) | MOH - Jaber Al-Ahmad Hospital (Innovation Research Laboratory) | Salman Al-Sabah et al |
| EPI_ISL_754180 | Department for Virology, Molecular Biology and Genome Research, R. G. Lugar Center for Public Health Research, National Center for Disease Control and Public Health (NCDC) of Georgia. | Department for Virology, Molecular Biology and Genome Research, R. G. Lugar Center for Public Health Research, National Center for Disease Control and Public Health (NCDC) of Georgia. | Salome Javashvili et al |
| EPI_ISL_453359 | Liverpool Clinical Laboratories | COVID-19 Genomics UK (COG-UK) Consortium | Sam Haldenby et al |
| EPI_ISL_1208399 | Laboratoire de virologie clinique - Institut Pasteur de Tunis | 1-Laboratory of Microbiology, National Reference Lab, Charles Nicolle Hospital; 2-University of Tunis ElManar, Faculty of Medicine of Tunis, LR99ES09, Tunis, Tunisia | Sameh Trabelsi et al |
| EPI_ISL_1197037 | Laboratoire de virologie clinique - Institut Pasteur de Tunis | 1-Laboratory of Microbiology, National Reference Lab, Charles Nicolle Hospital; 2-University of Tunis ElManar, Faculty of Medicine of Tunis, LR99ES09, Tunis, Tunisia | Sameh Trabelsi et al |
| EPI_ISL_766569 | Oman-National Influenza Center | Oman-National Influenza Center | Samiha Al-Kharusi et al |
| EPI_ISL_709542 | National Institute of Blood Diseases (NIBD), Molecular Biology Lab | Genomics Lab NIBD | Samina Naz Mukry et al |
| EPI_ISL_525424 | Oman-National Influenza Center | Biotechnology & OMICs Laboratory | Samira Al-Mahruqi et al |
| EPI_ISL_525425 | Oman-National Influenza Center | Biotechnology & OMICs Laboratory | Samira Al-Mahruqi et al |
| EPI_ISL_525429 | Oman-National Influenza Center | Biotechnology & OMICs Laboratory | Samira Al-Mahruqi et al |
| EPI_ISL_457706 | Oman-NIC | Oman-NIC | Samira Al-Maruqi et al |
| EPI_ISL_457937 | Oman-NIC | Oman-NIC | Samira Al-Maruqi et al |
| EPI_ISL_457991 | Oman-NIC | Oman-NIC | Samira Al-Maruqi et al |
| EPI_ISL_457701 | Oman-NIC | Oman-NIC | Samira Al-Maruqi et al |
| EPI_ISL_457704 | Oman-NIC | Oman-NIC | Samira Al-Maruqi et al |
| EPI_ISL_877428 | Institute of Microbiology and Immunology, Faculty of Medicine, University of Ljubljana | Institute of Microbiology and Immunology, Faculty of Medicine, University of Ljubljana | Samo Zakotnik et al |
| EPI_ISL_877439 | Institute of Microbiology and Immunology, Faculty of Medicine, University of Ljubljana | Institute of Microbiology and Immunology, Faculty of Medicine, University of Ljubljana | Samo Zakotnik et al |
| EPI_ISL_877443 | Institute of Microbiology and Immunology, Faculty of Medicine, University of Ljubljana | Institute of Microbiology and Immunology, Faculty of Medicine, University of Ljubljana | Samo Zakotnik et al |
| EPI_ISL_979246 | Institute of Microbiology and Immunology, Faculty of Medicine, University of Ljubljana | Institute of Microbiology and Immunology, Faculty of Medicine, University of Ljubljana | Samo Zakotnik et al |
| EPI_ISL_635201 | Institute of Microbiology and Immunology, Faculty of Medicine, University of Ljubljana | Institute of Microbiology and Immunology, Faculty of Medicine, University of Ljubljana | Samo Zakotnik et al |
| EPI_ISL_877432 | National laboratory of health, environment and food Celje | Institute of Microbiology and Immunology, Faculty of Medicine, University of Ljubljana | Samo Zakotnik et al |
| EPI_ISL_984640 | Laboratory of Virology of Federal Budget Health Care Institution Center of Hygiene and Epidemiology in Arkhangelsk region | Group of Genomics and Postgenomic Technologies of Central Research Institute of Epidemiology | Samoilov AE et al |
| EPI_ISL_1082311 | Laboratory of Virology of Federal Budget Health Care Institution Center of Hygiene and Epidemiology in Arkhangelsk region | Group of Genomics and Postgenomic Technologies of Central Research Institute of Epidemiology | Samoilov AE et al |
| EPI_ISL_1527385 | Laboratory of Virology of Federal Budget Health Care Institution Center of Hygiene and Epidemiology in Arkhangelsk region | Group of Genomics and Postgenomic Technologies of Central Research Institute of Epidemilology | Samoilov AE et al |

| | | | |
|---|---|---|---|
| EPI_ISL_1527368 | Molecular diagnostic laboratory of Federal Budget Institution of Science Central Research Institute of Epidemiology of The Federal Service on Customers' Rights Protection and Human Well-being Surveillance | Group of Genomics and Postgenomic Technologies of Central Research Institute of Epidemilology | Samoilov AE et al |
| EPI_ISL_476259 | Hospital da Clínicas da Faculdade de Medicina da Universidade de São Paulo | Instituto de Medicina Tropical da Univesidade de São Paulo | Samples: Ingra Morales Claro et al |
| EPI_ISL_476204 | Hospital da Clínicas da Faculdade de Medicina da Universidade de São Paulo | Instituto de Medicina Tropical da Univesidade de São Paulo | Samples: Ingra Morales Claro et al |
| EPI_ISL_476266 | Hospital da Clínicas da Faculdade de Medicina da Universidade de São Paulo | Instituto de Medicina Tropical da Univesidade de São Paulo | Samples: Ingra Morales Claro et al |
| EPI_ISL_476444 | Hospital da Clínicas da Faculdade de Medicina da Universidade de São Paulo | Instituto de Medicina Tropical da Univesidade de São Paulo | Samples: Ingra Morales Claro et al |
| EPI_ISL_476446 | Hospital da Clínicas da Faculdade de Medicina da Universidade de São Paulo | Instituto de Medicina Tropical da Univesidade de São Paulo | Samples: Ingra Morales Claro et al |
| EPI_ISL_476450 | Hospital da Clínicas da Faculdade de Medicina da Universidade de São Paulo | Instituto de Medicina Tropical da Univesidade de São Paulo | Samples: Ingra Morales Claro et al |
| EPI_ISL_476475 | Hospital da Clínicas da Faculdade de Medicina da Universidade de São Paulo | Instituto de Medicina Tropical da Univesidade de São Paulo | Samples: Ingra Morales Claro et al |
| EPI_ISL_476484 | Hospital da Clínicas da Faculdade de Medicina da Universidade de São Paulo | Instituto de Medicina Tropical da Univesidade de São Paulo | Samples: Ingra Morales Claro et al |
| EPI_ISL_476486 | Hospital da Clínicas da Faculdade de Medicina da Universidade de São Paulo | Instituto de Medicina Tropical da Univesidade de São Paulo | Samples: Ingra Morales Claro et al |
| EPI_ISL_476224 | DB Diagnósticos do Brasil | Instituto de Medicina Tropical da Univesidade de São Paulo | Samples: Nelson Gaburo Jr et al |
| EPI_ISL_476304 | DB Diagnósticos do Brasil | Instituto de Medicina Tropical da Univesidade de São Paulo | Samples: Nelson Gaburo Jr et al |
| EPI_ISL_476305 | DB Diagnósticos do Brasil | Instituto de Medicina Tropical da Univesidade de São Paulo | Samples: Nelson Gaburo Jr et al |
| EPI_ISL_476182 | DB Diagnósticos do Brasil | Instituto de Medicina Tropical da Univesidade de São Paulo | Samples: Nelson Gaburo Jr et al |
| EPI_ISL_476184 | DB Diagnósticos do Brasil | Instituto de Medicina Tropical da Univesidade de São Paulo | Samples: Nelson Gaburo Jr et al |
| EPI_ISL_476200 | DB Diagnósticos do Brasil | Instituto de Medicina Tropical da Univesidade de São Paulo | Samples: Nelson Gaburo Jr et al |
| EPI_ISL_476218 | DB Diagnósticos do Brasil | Instituto de Medicina Tropical da Univesidade de São Paulo | Samples: Nelson Gaburo Jr et al |
| EPI_ISL_476281 | DB Diagnósticos do Brasil | Instituto de Medicina Tropical da Univesidade de São Paulo | Samples: Nelson Gaburo Jr et al |
| EPI_ISL_476283 | DB Diagnósticos do Brasil | Instituto de Medicina Tropical da Univesidade de São Paulo | Samples: Nelson Gaburo Jr et al |
| EPI_ISL_476288 | DB Diagnósticos do Brasil | Instituto de Medicina Tropical da Univesidade de São Paulo | Samples: Nelson Gaburo Jr et al |
| EPI_ISL_476289 | DB Diagnósticos do Brasil | Instituto de Medicina Tropical da Univesidade de São Paulo | Samples: Nelson Gaburo Jr et al |
| EPI_ISL_476307 | DB Diagnósticos do Brasil | Instituto de Medicina Tropical da Univesidade de São Paulo | Samples: Nelson Gaburo Jr et al |
| EPI_ISL_476331 | DB Diagnósticos do Brasil | Instituto de Medicina Tropical da Univesidade de São Paulo | Samples: Nelson Gaburo Jr et al |
| EPI_ISL_476369 | DB Diagnósticos do Brasil | Instituto de Medicina Tropical da Univesidade de São Paulo | Samples: Nelson Gaburo Jr et al |
| EPI_ISL_476333 | DB Diagnósticos do Brasil | Instituto de Medicina Tropical da Univesidade de São Paulo | Samples: Nelson Gaburo Jr et al |
| EPI_ISL_476336 | DB Diagnósticos do Brasil | Instituto de Medicina Tropical da Univesidade de São Paulo | Samples: Nelson Gaburo Jr et al |
| EPI_ISL_476309 | DB Diagnósticos do Brasil | Instituto de Medicina Tropical da Univesidade de São Paulo | Samples: Nelson Gaburo Jr et al |
| EPI_ISL_476327 | DB Diagnósticos do Brasil | Instituto de Medicina Tropical da Univesidade de São Paulo | Samples: Nelson Gaburo Jr et al |
| EPI_ISL_476328 | DB Diagnósticos do Brasil | Instituto de Medicina Tropical da Univesidade de São Paulo | Samples: Nelson Gaburo Jr et al |

| | | | |
|---|---|---|---|
| EPI_ISL_476330 | DB Diagnósticos do Brasil | Instituto de Medicina Tropical da Univesidade de São Paulo | Samples: Nelson Gaburo Jr et al |
| EPI_ISL_476194 | DB Diagnósticos do Brasil | Instituto de Medicina Tropical da Univesidade de São Paulo | Samples: Nelson Gaburo Jr et al |
| EPI_ISL_476290 | DB Diagnósticos do Brasil | Instituto de Medicina Tropical da Univesidade de São Paulo | Samples: Nelson Gaburo Jr et al |
| EPI_ISL_476191 | DB Diagnósticos do Brasil | Instituto de Medicina Tropical da Univesidade de São Paulo | Samples: Nelson Gaburo Jr et al |
| EPI_ISL_476282 | DB Diagnósticos do Brasil | Instituto de Medicina Tropical da Univesidade de São Paulo | Samples: Nelson Gaburo Jr et al |
| EPI_ISL_476311 | DB Diagnósticos do Brasil | Instituto de Medicina Tropical da Univesidade de São Paulo | Samples: Nelson Gaburo Jr et al |
| EPI_ISL_476312 | DB Diagnósticos do Brasil | Instituto de Medicina Tropical da Univesidade de São Paulo | Samples: Nelson Gaburo Jr et al |
| EPI_ISL_476313 | DB Diagnósticos do Brasil | Instituto de Medicina Tropical da Univesidade de São Paulo | Samples: Nelson Gaburo Jr et al |
| EPI_ISL_476314 | DB Diagnósticos do Brasil | Instituto de Medicina Tropical da Univesidade de São Paulo | Samples: Nelson Gaburo Jr et al |
| EPI_ISL_476318 | DB Diagnósticos do Brasil | Instituto de Medicina Tropical da Univesidade de São Paulo | Samples: Nelson Gaburo Jr et al |
| EPI_ISL_476370 | DB Diagnósticos do Brasil | Instituto de Medicina Tropical da Univesidade de São Paulo | Samples: Nelson Gaburo Jr et al |
| EPI_ISL_476178 | DB Diagnósticos do Brasil | Instituto de Medicina Tropical da Univesidade de São Paulo | Samples: Nelson Gaburo Jr et al |
| EPI_ISL_476180 | DB Diagnósticos do Brasil | Instituto de Medicina Tropical da Univesidade de São Paulo | Samples: Nelson Gaburo Jr et al |
| EPI_ISL_476210 | DB Diagnósticos do Brasil | Instituto de Medicina Tropical da Univesidade de São Paulo | Samples: Nelson Gaburo Jr et al |
| EPI_ISL_476326 | DB Diagnósticos do Brasil | Instituto de Medicina Tropical da Univesidade de São Paulo | Samples: Nelson Gaburo Jr et al |
| EPI_ISL_476188 | DB Diagnósticos do Brasil | Instituto de Medicina Tropical da Univesidade de São Paulo | Samples: Nelson Gaburo Jr et al |
| EPI_ISL_476192 | DB Diagnósticos do Brasil | Instituto de Medicina Tropical da Univesidade de São Paulo | Samples: Nelson Gaburo Jr et al |
| EPI_ISL_476316 | DB Diagnósticos do Brasil | Instituto de Medicina Tropical da Univesidade de São Paulo | Samples: Nelson Gaburo Jr et al |
| EPI_ISL_476319 | DB Diagnósticos do Brasil | Instituto de Medicina Tropical da Univesidade de São Paulo | Samples: Nelson Gaburo Jr et al |
| EPI_ISL_476335 | DB Diagnósticos do Brasil | Instituto de Medicina Tropical da Univesidade de São Paulo | Samples: Nelson Gaburo Jr et al |
| EPI_ISL_476278 | DB Diagnósticos do Brasil | Instituto de Medicina Tropical da Univesidade de São Paulo | Samples: Nelson Gaburo Jr et al |
| EPI_ISL_476292 | DB Diagnósticos do Brasil | Instituto de Medicina Tropical da Univesidade de São Paulo | Samples: Nelson Gaburo Jr et al |
| EPI_ISL_476297 | DB Diagnósticos do Brasil | Instituto de Medicina Tropical da Univesidade de São Paulo | Samples: Nelson Gaburo Jr et al |
| EPI_ISL_476317 | DB Diagnósticos do Brasil | Instituto de Medicina Tropical da Univesidade de São Paulo | Samples: Nelson Gaburo Jr et al |
| EPI_ISL_476324 | DB Diagnósticos do Brasil | Instituto de Medicina Tropical da Univesidade de São Paulo | Samples: Nelson Gaburo Jr et al |
| EPI_ISL_476201 | DB Diagnósticos do Brasil | Instituto de Medicina Tropical da Univesidade de São Paulo | Samples: Nelson Gaburo Jr et al |
| EPI_ISL_476211 | DB Diagnósticos do Brasil | Instituto de Medicina Tropical da Univesidade de São Paulo | Samples: Nelson Gaburo Jr et al |
| EPI_ISL_476366 | DB Diagnósticos do Brasil | Instituto de Medicina Tropical da Univesidade de São Paulo | Samples: Nelson Gaburo Jr et al |
| EPI_ISL_897845 | University Hospitals of Geneva, Laboratory of Virology | HUG, Laboratory of Virology and the Health2030 Genome Center | Samuel Cordey et al |
| EPI_ISL_981706 | University Hospitals of Geneva, Laboratory of Virology | HUG, Laboratory of Virology and the Health2030 Genome Center | Samuel Cordey et al |

| | | | |
|---|---|---|---|
| EPI_ISL_981707 | University Hospitals of Geneva, Laboratory of Virology | HUG, Laboratory of Virology and the Health2030 Genome Center | Samuel Cordey et al |
| EPI_ISL_981708 | University Hospitals of Geneva, Laboratory of Virology | HUG, Laboratory of Virology and the Health2030 Genome Center | Samuel Cordey et al |
| EPI_ISL_981709 | University Hospitals of Geneva, Laboratory of Virology | HUG, Laboratory of Virology and the Health2030 Genome Center | Samuel Cordey et al |
| EPI_ISL_981710 | University Hospitals of Geneva, Laboratory of Virology | HUG, Laboratory of Virology and the Health2030 Genome Center | Samuel Cordey et al |
| EPI_ISL_981711 | University Hospitals of Geneva, Laboratory of Virology | HUG, Laboratory of Virology and the Health2030 Genome Center | Samuel Cordey et al |
| EPI_ISL_981712 | University Hospitals of Geneva, Laboratory of Virology | HUG, Laboratory of Virology and the Health2030 Genome Center | Samuel Cordey et al |
| EPI_ISL_1040363 | University Hospitals of Geneva, Laboratory of Virology | HUG, Laboratory of Virology and the Health2030 Genome Center | Samuel Cordey et al |
| EPI_ISL_1084930 | University Hospitals of Geneva, Laboratory of Virology | HUG, Laboratory of Virology and the Health2030 Genome Center | Samuel Cordey et al |
| EPI_ISL_1369531 | University Hospitals of Geneva, Laboratory of Virology | HUG, Laboratory of Virology and the Health2030 Genome Center | Samuel Cordey et al |
| EPI_ISL_1533404 | University Hospitals of Geneva, Laboratory of Virology | HUG, Laboratory of Virology and the Health2030 Genome Center | Samuel Cordey et al |
| EPI_ISL_1533406 | University Hospitals of Geneva, Laboratory of Virology | HUG, Laboratory of Virology and the Health2030 Genome Center | Samuel Cordey et al |
| EPI_ISL_1228877 | University Hospitals of Geneva, Laboratory of Virology | HUG, Laboratory of Virology and the Health2030 Genome Center | Samuel Cordey et al |
| EPI_ISL_981713 | University Hospitals of Geneva, Laboratory of Virology | HUG, Laboratory of Virology and the Health2030 Genome Center | Samuel Cordey et al |
| EPI_ISL_981714 | University Hospitals of Geneva, Laboratory of Virology | HUG, Laboratory of Virology and the Health2030 Genome Center | Samuel Cordey et al |
| EPI_ISL_1369541 | University Hospitals of Geneva, Laboratory of Virology | HUG, Laboratory of Virology and the Health2030 Genome Center | Samuel Cordey et al |
| EPI_ISL_1448613 | University Hospitals of Geneva, Laboratory of Virology | HUG, Laboratory of Virology and the Health2030 Genome Center | Samuel Cordey et al |
| EPI_ISL_1665100 | National Laboratory for Health, Environment and Food, OMM, Maribor | NLZOH (National Laboratory for Health, Environment and Food) / CISLD (Clinical Institute of Special Laboratory Diagnostics), University Children's Hospital, University Medical Center Ljubljana | Sandra Janezic et al |
| EPI_ISL_1664822 | National Laboratory for Health, Environment and Food, OMM, Celje | NLZOH (National Laboratory for Health, Environment and Food) / CISLD (Clinical Institute of Special Laboratory Diagnostics), University Children's Hospital, University Medical Center Ljubljana | Sandra Janezic et al |
| EPI_ISL_1664811 | National Laboratory for Health, Environment and Food, OMM, Celje | NLZOH (National Laboratory for Health, Environment and Food) / CISLD (Clinical Institute of Special Laboratory Diagnostics), University Children's Hospital, University Medical Center Ljubljana | Sandra Janezic et al |
| EPI_ISL_1664862 | National Laboratory for Health, Environment and Food, OMM, Celje | NLZOH (National Laboratory for Health, Environment and Food) / CISLD (Clinical Institute of Special Laboratory Diagnostics), University Children's Hospital, University Medical Center Ljubljana | Sandra Janezic et al |
| EPI_ISL_1523487 | Laboratoire de santé publique du Québec | Laboratoire de santé publique du Québec | Sandrine Moreira et al |
| EPI_ISL_1404870 | Laboratoire de santé publique du Québec | Laboratoire de santé publique du Québec | Sandrine Moreira et al |
| EPI_ISL_1241439 | Laboratoire de santé publique du Québec | Laboratoire de santé publique du Québec | Sandrine Moreira et al |
| EPI_ISL_1241665 | Laboratoire de santé publique du Québec | Laboratoire de santé publique du Québec | Sandrine Moreira et al |
| EPI_ISL_1534181 | Laboratoire de santé publique du Québec | Laboratoire de santé publique du Québec | Sandrine Moreira et al |
| EPI_ISL_1167303 | Laboratoire de santé publique du Québec | Laboratoire de santé publique du Québec | Sandrine Moreira et al |
| EPI_ISL_826002 | Laboratoire de santé publique du Québec | Laboratoire de santé publique du Québec | Sandrine Moreira et al |

| | | | |
|---|---|---|---|
| EPI_ISL_826072 | Laboratoire de santé publique du Québec | Laboratoire de santé publique du Québec | Sandrine Moreira et al |
| EPI_ISL_826243 | Laboratoire de santé publique du Québec | Laboratoire de santé publique du Québec | Sandrine Moreira et al |
| EPI_ISL_1403122 | Laboratoire de santé publique du Québec | Laboratoire de santé publique du Québec | Sandrine Moreira et al |
| EPI_ISL_1403125 | Laboratoire de santé publique du Québec | Laboratoire de santé publique du Québec | Sandrine Moreira et al |
| EPI_ISL_1365947 | Laboratoire de santé publique du Québec | Laboratoire de santé publique du Québec | Sandrine Moreira et al |
| EPI_ISL_535716 | Hôpital de Verdun | Laboratoire de santé publique du Québec | Sandrine Moreira et al |
| EPI_ISL_450303 | Centre hospitalier Anna-Laberge | Laboratoire de santé publique du Québec | Sandrine Moreira et al |
| EPI_ISL_465693 | Hôpital du Suroît | Laboratoire de santé publique du Québec | Sandrine Moreira et al |
| EPI_ISL_465696 | Hôpital Pierre-Boucher | Laboratoire de santé publique du Québec | Sandrine Moreira et al |
| EPI_ISL_848599 | Evandro Chagas Institute | Evandro Chagas Institute | Santos et al |
| EPI_ISL_848602 | Evandro Chagas Institute | Evandro Chagas Institute | Santos et al |
| EPI_ISL_458139 | Evandro Chagas Institute | Evandro Chagas Institute | Santos et al |
| EPI_ISL_848600 | Evandro Chagas Institute | Evandro Chagas Institute | Santos et al |
| EPI_ISL_848601 | Evandro Chagas Institute | Evandro Chagas Institute | Santos et al |
| EPI_ISL_848627 | Evandro Chagas Institute | Evandro Chagas Institute | Santos et al |
| EPI_ISL_848573 | Evandro Chagas Institute | Evandro Chagas Institute | Santos et al |
| EPI_ISL_848574 | Evandro Chagas Institute | Evandro Chagas Institute | Santos et al |
| EPI_ISL_848612 | Evandro Chagas Institute | Evandro Chagas Institute | Santos et al |
| EPI_ISL_848613 | Evandro Chagas Institute | Evandro Chagas Institute | Santos et al |
| EPI_ISL_925846 | LACEN - Laboratório Central de Saúde Pública do Amazonas | Evandro Chagas Institute Virology | Santos et al |
| EPI_ISL_918532 | LACEN - Laboratório Central de Saúde Pública do Amazonas | Evandro Chagas Institute | Santos et al |
| EPI_ISL_918533 | LACEN - Laboratório Central de Saúde Pública do Amazonas | Evandro Chagas Institute | Santos et al |
| EPI_ISL_1261694 | LACEN - Laboratório Central de Saúde Pública do Amazonas | Evandro Chagas Institute | Santos et al |
| EPI_ISL_918551 | LACEN - Laboratório Central de Saúde Pública do Amapa | Evandro Chagas Institute | Santos et al |
| EPI_ISL_918553 | LACEN - Laboratório Central de Saúde Pública do Amapa | Evandro Chagas Institute | Santos et al |
| EPI_ISL_918554 | LACEN - Laboratório Central de Saúde Pública do Amapa | Evandro Chagas Institute | Santos et al |
| EPI_ISL_918555 | LACEN - Laboratório Central de Saúde Pública do Amapa | Evandro Chagas Institute | Santos et al |
| EPI_ISL_918556 | LACEN - Laboratório Central de Saúde Pública do Amapa | Evandro Chagas Institute | Santos et al |
| EPI_ISL_918557 | LACEN - Laboratório Central de Saúde Pública do Amapa | Evandro Chagas Institute | Santos et al |
| EPI_ISL_918558 | LACEN - Laboratório Central de Saúde Pública do Amapa | Evandro Chagas Institute | Santos et al |

| | | | |
|---|---|---|---|
| EPI_ISL_918559 | LACEN - Laboratório Central de Saúde Pública do Amapa | Evandro Chagas Institute | Santos et al |
| EPI_ISL_918560 | LACEN - Laboratório Central de Saúde Pública do Amapa | Evandro Chagas Institute | Santos et al |
| EPI_ISL_918561 | LACEN - Laboratório Central de Saúde Pública do Amapa | Evandro Chagas Institute | Santos et al |
| EPI_ISL_1164985 | LACEN - Laboratório Central de Saúde Pública do Amapá | Evandro Chagas Institute | Santos et al |
| EPI_ISL_1164984 | LACEN - Laboratório Central de Saúde Pública do Amapá | Evandro Chagas Institute | Santos et al |
| EPI_ISL_1164981 | LACEN - Laboratório Central de Saúde Pública do Amapá | Evandro Chagas Institute | Santos et al |
| EPI_ISL_1164982 | LACEN - Laboratório Central de Saúde Pública do Amapá | Evandro Chagas Institute | Santos et al |
| EPI_ISL_1164976 | LACEN - Laboratório Central de Saúde Pública do Amapá | Evandro Chagas Institute | Santos et al |
| EPI_ISL_1261686 | LACEN - Laboratório Central de Saúde Pública do Amapá | Evandro Chagas Institute | Santos et al |
| EPI_ISL_1261696 | LACEN - Laboratório Central de Saúde Pública do Amapá | Evandro Chagas Institute | Santos et al |
| EPI_ISL_1261692 | LACEN - Laboratório Central de Saúde Pública do Amapá | Evandro Chagas Institute | Santos et al |
| EPI_ISL_1261695 | LACEN - Laboratório Central de Saúde Pública do Amapá | Evandro Chagas Institute | Santos et al |
| EPI_ISL_1261688 | LACEN - Laboratório Central de Saúde Pública do Amapá | Evandro Chagas Institute | Santos et al |
| EPI_ISL_1261689 | LACEN - Laboratório Central de Saúde Pública do Amapá | Evandro Chagas Institute | Santos et al |
| EPI_ISL_450873 | Evandro Chagas Institute | Evandro Chagas Institute | Santos et al |
| EPI_ISL_524792 | Evandro Chagas Institute | Evandro Chagas Institute | Santos et al |
| EPI_ISL_458138 | Evandro Chagas Institute | Evandro Chagas Institute | Santos et al |
| EPI_ISL_458142 | Evandro Chagas Institute | Evandro Chagas Institute | Santos et al |
| EPI_ISL_524795 | Evandro Chagas Institute | Evandro Chagas Institute | Santos et al |
| EPI_ISL_458144 | Evandro Chagas Institute | Evandro Chagas Institute | Santos et al |
| EPI_ISL_458143 | Evandro Chagas Institute | Evandro Chagas Institute | Santos et al |
| EPI_ISL_458145 | Evandro Chagas Institute | Evandro Chagas Institute | Santos et al |
| EPI_ISL_524796 | Evandro Chagas Institute | Evandro Chagas Institute | Santos et al |
| EPI_ISL_524784 | Evandro Chagas Institute | Evandro Chagas Institute | Santos et al |
| EPI_ISL_524793 | Evandro Chagas Institute | Evandro Chagas Institute | Santos et al |
| EPI_ISL_848584 | Evandro Chagas Institute | Evandro Chagas Institute | Santos et al |
| EPI_ISL_848578 | Evandro Chagas Institute | Evandro Chagas Institute | Santos et al |
| EPI_ISL_848579 | Evandro Chagas Institute | Evandro Chagas Institute | Santos et al |
| EPI_ISL_848577 | Evandro Chagas Institute | Evandro Chagas Institute | Santos et al |
| EPI_ISL_848575 | Evandro Chagas Institute | Evandro Chagas Institute | Santos et al |

| | | | |
|---|---|---|---|
| EPI_ISL_848576 | Evandro Chagas Institute | Evandro Chagas Institute | Santos et al |
| EPI_ISL_1261697 | LACEN - Laboratório Central de Saúde Pública do Ceará | Evandro Chagas Institute | Santos et al |
| EPI_ISL_918537 | LACEN - Laboratório Central de Saúde Pública do Ceara | Evandro Chagas Institute | Santos et al |
| EPI_ISL_918538 | LACEN - Laboratório Central de Saúde Pública do Ceara | Evandro Chagas Institute | Santos et al |
| EPI_ISL_918539 | LACEN - Laboratório Central de Saúde Pública do Ceara | Evandro Chagas Institute | Santos et al |
| EPI_ISL_918540 | LACEN - Laboratório Central de Saúde Pública do Ceara | Evandro Chagas Institute | Santos et al |
| EPI_ISL_918541 | LACEN - Laboratório Central de Saúde Pública do Ceara | Evandro Chagas Institute | Santos et al |
| EPI_ISL_918542 | LACEN - Laboratório Central de Saúde Pública do Ceara | Evandro Chagas Institute | Santos et al |
| EPI_ISL_918543 | LACEN - Laboratório Central de Saúde Pública do Ceara | Evandro Chagas Institute | Santos et al |
| EPI_ISL_918544 | LACEN - Laboratório Central de Saúde Pública do Ceara | Evandro Chagas Institute | Santos et al |
| EPI_ISL_1164980 | LACEN - Laboratório Central de Saúde Pública do Ceará | Evandro Chagas Institute | Santos et al |
| EPI_ISL_1164986 | LACEN - Laboratório Central de Saúde Pública do Ceará | Evandro Chagas Institute | Santos et al |
| EPI_ISL_1164995 | LACEN - Laboratório Central de Saúde Pública do Ceará | Evandro Chagas Institute | Santos et al |
| EPI_ISL_1164993 | LACEN - Laboratório Central de Saúde Pública do Ceará | Evandro Chagas Institute | Santos et al |
| EPI_ISL_1164973 | LACEN - Laboratório Central de Saúde Pública do Ceará | Evandro Chagas Institute | Santos et al |
| EPI_ISL_1164970 | LACEN - Laboratório Central de Saúde Pública do Ceará | Evandro Chagas Institute | Santos et al |
| EPI_ISL_1164971 | LACEN - Laboratório Central de Saúde Pública do Ceará | Evandro Chagas Institute | Santos et al |
| EPI_ISL_1261693 | LACEN - Laboratório Central de Saúde Pública do Ceará | Evandro Chagas Institute | Santos et al |
| EPI_ISL_1261684 | LACEN - Laboratório Central de Saúde Pública do Ceará | Evandro Chagas Institute | Santos et al |
| EPI_ISL_1164996 | LACEN - Laboratório Central de Saúde Pública do Maranhao | Evandro Chagas Institute | Santos et al |
| EPI_ISL_1164998 | LACEN - Laboratório Central de Saúde Pública do Maranhao | Evandro Chagas Institute | Santos et al |
| EPI_ISL_524797 | Evandro Chagas Institute | Evandro Chagas Institute | Santos et al |
| EPI_ISL_848626 | Evandro Chagas Institute | Evandro Chagas Institute | Santos et al |
| EPI_ISL_524791 | Evandro Chagas Institute | Evandro Chagas Institute | Santos et al |
| EPI_ISL_458149 | Evandro Chagas Institute | Evandro Chagas Institute | Santos et al |
| EPI_ISL_524788 | Evandro Chagas Institute | Evandro Chagas Institute | Santos et al |
| EPI_ISL_524790 | Evandro Chagas Institute | Evandro Chagas Institute | Santos et al |
| EPI_ISL_524794 | Evandro Chagas Institute | Evandro Chagas Institute | Santos et al |
| EPI_ISL_524789 | Evandro Chagas Institute | Evandro Chagas Institute | Santos et al |
| EPI_ISL_524799 | Evandro Chagas Institute | Evandro Chagas Institute | Santos et al |

| | | | |
|---|---|---|---|
| EPI_ISL_848580 | Evandro Chagas Institute | Evandro Chagas Institute | Santos et al |
| EPI_ISL_848581 | Evandro Chagas Institute | Evandro Chagas Institute | Santos et al |
| EPI_ISL_1086373 | LACEN - Laboratório Central de Saúde Pública do Maranhao | Evandro Chagas Institute | Santos et al |
| EPI_ISL_918514 | Evandro Chagas Institute | Evandro Chagas Institute | Santos et al |
| EPI_ISL_918515 | LACEN - Laboratório Central de Saúde Pública do Para | Evandro Chagas Institute | Santos et al |
| EPI_ISL_918516 | LACEN - Laboratório Central de Saúde Pública do Para | Evandro Chagas Institute | Santos et al |
| EPI_ISL_918517 | LACEN - Laboratório Central de Saúde Pública do Para | Evandro Chagas Institute | Santos et al |
| EPI_ISL_918518 | Evandro Chagas Institute | Evandro Chagas Institute | Santos et al |
| EPI_ISL_918519 | Evandro Chagas Institute | Evandro Chagas Institute | Santos et al |
| EPI_ISL_918520 | Evandro Chagas Institute | Evandro Chagas Institute | Santos et al |
| EPI_ISL_918521 | Evandro Chagas Institute | Evandro Chagas Institute | Santos et al |
| EPI_ISL_918523 | LACEN - Laboratório Central de Saúde Pública do Para | Evandro Chagas Institute | Santos et al |
| EPI_ISL_918524 | LACEN - Laboratório Central de Saúde Pública do Para | Evandro Chagas Institute | Santos et al |
| EPI_ISL_918525 | LACEN - Laboratório Central de Saúde Pública do Para | Evandro Chagas Institute | Santos et al |
| EPI_ISL_918527 | LACEN - Laboratório Central de Saúde Pública do Para | Evandro Chagas Institute | Santos et al |
| EPI_ISL_918528 | LACEN - Laboratório Central de Saúde Pública do Para | Evandro Chagas Institute | Santos et al |
| EPI_ISL_918529 | LACEN - Laboratório Central de Saúde Pública do Para | Evandro Chagas Institute | Santos et al |
| EPI_ISL_918545 | LACEN - Laboratório Central de Saúde Pública do Para | Evandro Chagas Institute | Santos et al |
| EPI_ISL_918546 | LACEN - Laboratório Central de Saúde Pública do Para | Evandro Chagas Institute | Santos et al |
| EPI_ISL_918547 | LACEN - Laboratório Central de Saúde Pública do Para | Evandro Chagas Institute | Santos et al |
| EPI_ISL_918548 | LACEN - Laboratório Central de Saúde Pública do Para | Evandro Chagas Institute | Santos et al |
| EPI_ISL_918549 | LACEN - Laboratório Central de Saúde Pública do Para | Evandro Chagas Institute | Santos et al |
| EPI_ISL_904120 | LACEN - Laboratório Central de Saúde Pública do Pará | Evandro Chagas Institute | Santos et al |
| EPI_ISL_904121 | LACEN - Laboratório Central de Saúde Pública do Pará | Evandro Chagas Institute | Santos et al |
| EPI_ISL_918550 | LACEN - Laboratório Central de Saúde Pública do Para | Evandro Chagas Institute | Santos et al |
| EPI_ISL_918552 | LACEN - Laboratório Central de Saúde Pública do Para | Evandro Chagas Institute | Santos et al |
| EPI_ISL_1164972 | LACEN - Laboratório Central de Saúde Pública do Pará | Evandro Chagas Institute | Santos et al |
| EPI_ISL_1164978 | LACEN - Laboratório Central de Saúde Pública do Pará | Evandro Chagas Institute | Santos et al |
| EPI_ISL_1164983 | LACEN - Laboratório Central de Saúde Pública do Pará | Evandro Chagas Institute | Santos et al |
| EPI_ISL_1164974 | LACEN - Laboratório Central de Saúde Pública do Pará | Evandro Chagas Institute | Santos et al |

| | | | |
|---|---|---|---|
| EPI_ISL_1164975 | LACEN - Laboratório Central de Saúde Pública do Pará | Evandro Chagas Institute | Santos et al |
| EPI_ISL_1261691 | Laboratório Paulo C. Azevedo | Evandro Chagas Institute | Santos et al |
| EPI_ISL_450874 | Evandro Chagas Institute | Evandro Chagas Institute | Santos et al |
| EPI_ISL_458140 | Evandro Chagas Institute | Evandro Chagas Institute | Santos et al |
| EPI_ISL_848624 | Evandro Chagas Institute | Evandro Chagas Institute | Santos et al |
| EPI_ISL_524787 | Evandro Chagas Institute | Evandro Chagas Institute | Santos et al |
| EPI_ISL_458146 | Evandro Chagas Institute | Evandro Chagas Institute | Santos et al |
| EPI_ISL_458147 | Evandro Chagas Institute | Evandro Chagas Institute | Santos et al |
| EPI_ISL_458141 | Evandro Chagas Institute | Evandro Chagas Institute | Santos et al |
| EPI_ISL_524783 | Evandro Chagas Institute | Evandro Chagas Institute | Santos et al |
| EPI_ISL_524786 | Evandro Chagas Institute | Evandro Chagas Institute | Santos et al |
| EPI_ISL_524785 | Evandro Chagas Institute | Evandro Chagas Institute | Santos et al |
| EPI_ISL_524800 | Evandro Chagas Institute | Evandro Chagas Institute | Santos et al |
| EPI_ISL_1063789 | Evandro Chagas Institute | Evandro Chagas Institute Virology | Santos et al |
| EPI_ISL_848622 | Evandro Chagas Institute | Evandro Chagas Institute | Santos et al |
| EPI_ISL_848628 | Evandro Chagas Institute | Evandro Chagas Institute | Santos et al |
| EPI_ISL_848629 | Evandro Chagas Institute | Evandro Chagas Institute | Santos et al |
| EPI_ISL_848597 | Evandro Chagas Institute | Evandro Chagas Institute | Santos et al |
| EPI_ISL_848596 | Evandro Chagas Institute | Evandro Chagas Institute | Santos et al |
| EPI_ISL_848625 | Evandro Chagas Institute | Evandro Chagas Institute | Santos et al |
| EPI_ISL_848598 | Evandro Chagas Institute | Evandro Chagas Institute | Santos et al |
| EPI_ISL_848617 | Evandro Chagas Institute | Evandro Chagas Institute | Santos et al |
| EPI_ISL_848618 | Evandro Chagas Institute | Evandro Chagas Institute | Santos et al |
| EPI_ISL_848620 | Evandro Chagas Institute | Evandro Chagas Institute | Santos et al |
| EPI_ISL_848593 | Evandro Chagas Institute | Evandro Chagas Institute | Santos et al |
| EPI_ISL_848591 | Evandro Chagas Institute | Evandro Chagas Institute | Santos et al |
| EPI_ISL_848588 | Evandro Chagas Institute | Evandro Chagas Institute | Santos et al |
| EPI_ISL_848590 | Evandro Chagas Institute | Evandro Chagas Institute | Santos et al |
| EPI_ISL_848571 | Evandro Chagas Institute | Evandro Chagas Institute | Santos et al |
| EPI_ISL_848583 | Evandro Chagas Institute | Evandro Chagas Institute | Santos et al |

| | | | |
|---|---|---|---|
| EPI_ISL_848615 | Evandro Chagas Institute | Evandro Chagas Institute | Santos et al |
| EPI_ISL_848582 | Evandro Chagas Institute | Evandro Chagas Institute | Santos et al |
| EPI_ISL_848611 | Evandro Chagas Institute | Evandro Chagas Institute | Santos et al |
| EPI_ISL_848568 | Evandro Chagas Institute | Evandro Chagas Institute | Santos et al |
| EPI_ISL_848567 | Evandro Chagas Institute | Evandro Chagas Institute | Santos et al |
| EPI_ISL_848572 | Evandro Chagas Institute | Evandro Chagas Institute | Santos et al |
| EPI_ISL_848566 | Evandro Chagas Institute | Evandro Chagas Institute | Santos et al |
| EPI_ISL_848614 | Evandro Chagas Institute | Evandro Chagas Institute | Santos et al |
| EPI_ISL_848570 | Evandro Chagas Institute | Evandro Chagas Institute | Santos et al |
| EPI_ISL_848569 | Evandro Chagas Institute | Evandro Chagas Institute | Santos et al |
| EPI_ISL_848564 | Evandro Chagas Institute | Evandro Chagas Institute | Santos et al |
| EPI_ISL_848609 | Evandro Chagas Institute | Evandro Chagas Institute | Santos et al |
| EPI_ISL_848561 | Evandro Chagas Institute | Evandro Chagas Institute | Santos et al |
| EPI_ISL_848565 | Evandro Chagas Institute | Evandro Chagas Institute | Santos et al |
| EPI_ISL_848562 | Evandro Chagas Institute | Evandro Chagas Institute | Santos et al |
| EPI_ISL_848563 | Evandro Chagas Institute | Evandro Chagas Institute | Santos et al |
| EPI_ISL_848608 | Evandro Chagas Institute | Evandro Chagas Institute | Santos et al |
| EPI_ISL_848555 | Evandro Chagas Institute | Evandro Chagas Institute | Santos et al |
| EPI_ISL_848607 | Evandro Chagas Institute | Evandro Chagas Institute | Santos et al |
| EPI_ISL_848559 | Evandro Chagas Institute | Evandro Chagas Institute | Santos et al |
| EPI_ISL_848560 | Evandro Chagas Institute | Evandro Chagas Institute | Santos et al |
| EPI_ISL_848558 | Evandro Chagas Institute | Evandro Chagas Institute | Santos et al |
| EPI_ISL_848606 | Evandro Chagas Institute | Evandro Chagas Institute | Santos et al |
| EPI_ISL_848557 | Evandro Chagas Institute | Evandro Chagas Institute | Santos et al |
| EPI_ISL_1164994 | LACEN - Laboratório Central de Saúde Pública do Paraiba | Evandro Chagas Institute | Santos et al |
| EPI_ISL_1164992 | LACEN - Laboratório Central de Saúde Pública do Paraiba | Evandro Chagas Institute | Santos et al |
| EPI_ISL_1164989 | LACEN - Laboratório Central de Saúde Pública do Paraiba | Evandro Chagas Institute | Santos et al |
| EPI_ISL_1164991 | LACEN - Laboratório Central de Saúde Pública do Paraiba | Evandro Chagas Institute | Santos et al |
| EPI_ISL_524801 | Evandro Chagas Institute | Evandro Chagas Institute | Santos et al |
| EPI_ISL_1086377 | LACEN - Laboratório Central de Saúde Pública do Paraiba | Evandro Chagas Institute | Santos et al |

| | | | |
|---|---|---|---|
| EPI_ISL_1261699 | LACEN - Laboratório Central de Saúde Pública de Pernambuco | Evandro Chagas Institute | Santos et al |
| EPI_ISL_1261698 | LACEN - Laboratório Central de Saúde Pública de Pernambuco | Evandro Chagas Institute | Santos et al |
| EPI_ISL_524798 | Evandro Chagas Institute | Evandro Chagas Institute | Santos et al |
| EPI_ISL_1164977 | LACEN - Laboratório Central de Saúde Pública do Rio Grande do Norte | Evandro Chagas Institute | Santos et al |
| EPI_ISL_1164997 | LACEN - Laboratório Central de Saúde Pública do Rio Grande do Norte | Evandro Chagas Institute | Santos et al |
| EPI_ISL_1086376 | LACEN - Laboratório Central de Saúde Pública do Rio Grande do Norte | Evandro Chagas Institute | Santos et al |
| EPI_ISL_1086375 | LACEN - Laboratório Central de Saúde Pública do Rio Grande do Norte | Evandro Chagas Institute | Santos et al |
| EPI_ISL_918513 | LACEN - Laboratório Central de Saúde Pública do Roraima | Evandro Chagas Institute | Santos et al |
| EPI_ISL_1261687 | LACEN - Laboratório Central de Saúde Pública de Roraima | Evandro Chagas Institute | Santos et al |
| EPI_ISL_848586 | Evandro Chagas Institute | Evandro Chagas Institute | Santos et al |
| EPI_ISL_848610 | Evandro Chagas Institute | Evandro Chagas Institute | Santos et al |
| EPI_ISL_751235 | Pathogen Genomics Lab King Abdullah University of Science and Technology (KAUST) | Pathogen Genomics Lab King Abdullah University of Science and Technology(KAUST) | Sara Mfarrej et al |
| EPI_ISL_751230 | Pathogen Genomics Lab King Abdullah University of Science and Technology (KAUST) | Pathogen Genomics Lab King Abdullah University of Science and Technology(KAUST) | Sara Mfarrej et al |
| EPI_ISL_751217 | Pathogen Genomics Lab King Abdullah University of Science and Technology (KAUST) | Pathogen Genomics Lab King Abdullah University of Science and Technology(KAUST) | Sara Mfarrej et al |
| EPI_ISL_678215 | Pathogen Genomics Lab King Abdullah University of Science and Technology (KAUST) | Pathogen Genomics Lab King Abdullah University of Science and Technology(KAUST) | Sara Mfarrej et al |
| EPI_ISL_678182 | Pathogen Genomics Lab King Abdullah University of Science and Technology (KAUST) | Pathogen Genomics Lab King Abdullah University of Science and Technology(KAUST) | Sara Mfarrej et al |
| EPI_ISL_678220 | Pathogen Genomics Lab King Abdullah University of Science and Technology (KAUST) | Pathogen Genomics Lab King Abdullah University of Science and Technology(KAUST) | Sara Mfarrej et al |
| EPI_ISL_508822 | Florida Bureau of Public Health Laboratories | Florida Bureau of Public Health Laboratories | Sarah Schmedes et al |
| EPI_ISL_514175 | Florida Bureau of Public Health Laboratories | Florida Bureau of Public Health Laboratories | Sarah Schmedes et al |
| EPI_ISL_514209 | Florida Bureau of Public Health Laboratories | Florida Bureau of Public Health Laboratories | Sarah Schmedes et al |
| EPI_ISL_594371 | Florida Bureau of Public Health Laboratories | Florida Bureau of Public Health Laboratories | Sarah Schmedes et al |
| EPI_ISL_594383 | Florida Bureau of Public Health Laboratories | Florida Bureau of Public Health Laboratories | Sarah Schmedes et al |
| EPI_ISL_848773 | Florida Bureau of Public Health Laboratories | Florida Bureau of Public Health Laboratories | Sarah Schmedes et al |
| EPI_ISL_1532786 | Translational Health Science and Technology Institute -ESIC medical college and hospital, Faridabad | THSTI Bioassay laboratory | Saurabh Kumar et al |
| EPI_ISL_476832 | Medical Biology Department, Kocaeli University | Medical Genetics Department, Kocaeli University | Savli H et al |
| EPI_ISL_907077 | Biology, MCL | Biology, MCL | Seadawy et al |
| EPI_ISL_907085 | Biology, MCL | Biology, MCL | Seadawy et al |
| EPI_ISL_1531880 | Centro de Diagnostico COVID-19 UABC Tijuana | Andersen lab at Scripps Research | SEARCH Alliance San Diego with Idanya R |

| | | | |
|---|---|---|---|
| EPI_ISL_1531899 | Centro de Diagnostico COVID-19 UABC Tijuana | Andersen lab at Scripps Research | SEARCH Alliance San Diego with Idanya R |
| EPI_ISL_1531917 | Centro de Diagnostico COVID-19 UABC Tijuana | Andersen lab at Scripps Research | SEARCH Alliance San Diego with Idanya R |
| EPI_ISL_635488 | Centro de Diagn√≥stico COVID-19 UABC Tijuana | Andersen lab at Scripps Research | SEARCH Alliance San Diego with Idanya R |
| EPI_ISL_635549 | Centro de Diagn√≥stico COVID-19 UABC Tijuana | Andersen lab at Scripps Research | SEARCH Alliance San Diego with Idanya R |
| EPI_ISL_730223 | Genomica Lab Molecular, M√©xico | Andersen lab at Scripps Research | SEARCH Alliance San Diego with Jonathan |
| EPI_ISL_563576 | Microbiological Diagnostic Unit - Public Health Laboratory (MDU-PHL) | MDU-PHL | Seemann et al |
| EPI_ISL_564763 | Microbiological Diagnostic Unit - Public Health Laboratory (MDU-PHL) | MDU-PHL | Seemann et al |
| EPI_ISL_565504 | Microbiological Diagnostic Unit - Public Health Laboratory (MDU-PHL) | MDU-PHL | Seemann et al |
| EPI_ISL_419731 | Microbiological Diagnostic Unit Public Health Laboratory | Microbiological Diagnostic Unit Public Health Laboratory | Seemann T. et al |
| EPI_ISL_592747 | Microbiological Diagnostic Unit - Public Health Laboratory (MDU-PHL) | MDU-PHL | Seemann T. et al |
| EPI_ISL_641220 | Microbiological Diagnostic Unit - Public Health Laboratory (MDU-PHL) | MDU-PHL | Seemann T. et al |
| EPI_ISL_521641 | Microbiological Diagnostic Unit - Public Health Laboratory (MDU-PHL) | MDU-PHL | Seemann T. et al |
| EPI_ISL_520615 | Microbiological Diagnostic Unit - Public Health Laboratory (MDU-PHL) | MDU-PHL | Seemann T. et al |
| EPI_ISL_430644 | Microbiological Diagnostic Unit Public Health Laboratory | Microbiological Diagnostic Unit Public Health Laboratory | Seemann T. et al |
| EPI_ISL_468071 | Child Health Research Foundation | Child Health Research Foundation | Senjuti Saha et al |
| EPI_ISL_468072 | Child Health Research Foundation | Child Health Research Foundation | Senjuti Saha et al |
| EPI_ISL_492028 | Child Health Research Foundation | Child Health Research Foundation | Senjuti Saha et al |
| EPI_ISL_625468 | Child Health Research Foundation | Child Health Research Foundation | Senjuti Saha et al |
| EPI_ISL_700339 | Child Health Research Foundation | Child Health Research Foundation | Senjuti Saha et al |
| EPI_ISL_700332 | Child Health Research Foundation | Child Health Research Foundation | Senjuti Saha et al |
| EPI_ISL_700344 | Child Health Research Foundation | Child Health Research Foundation | Senjuti Saha et al |
| EPI_ISL_700345 | Child Health Research Foundation | Child Health Research Foundation | Senjuti Saha et al |
| EPI_ISL_700347 | Child Health Research Foundation | Child Health Research Foundation | Senjuti Saha et al |
| EPI_ISL_906091 | Shimantik Pathology and Diagnostic Center | Child Health Research Foundation | Senjuti Saha et al |
| EPI_ISL_591070 | College of Veterinary Medicine, Chungnam National University | College of Veterinary Medicine, Chungnam National University | Seo et al |
| EPI_ISL_428901 | State Research Center of Virology and Biotechnology VECTOR, Department of Collection of Microorganisms | State Research Center of Virology and Biotechnology VECTOR, Department of Collection of Microorganisms | Sergey A. Bodnev et al |
| EPI_ISL_428898 | State Research Center of Virology and Biotechnology VECTOR, Department of Collection of Microorganisms | State Research Center of Virology and Biotechnology VECTOR, Department of Collection of Microorganisms | Sergey A. Bodnev et al |
| EPI_ISL_428897 | State Research Center of Virology and Biotechnology VECTOR, Department of Collection of Microorganisms | State Research Center of Virology and Biotechnology VECTOR, Department of Collection of Microorganisms | Sergey A. Bodnev et al |
| EPI_ISL_985408 | Hospital General Universitario Gregorio Marañón | Hospital General Universitario Gregorio Marañón | Sergio Buenestado Serrano et al |

ubi Serafin Higuera et al

ubi Serafin Higuera et al

ubí Serafín Higuera et al

ubí Serafín Higuera et al

Gonzalez Garcia et al

| | | | |
|---|---|---|---|
| EPI_ISL_985409 | Hospital General Universitario Gregorio Marañón | Hospital General Universitario Gregorio Marañón | Sergio Buenestado Serrano et al |
| EPI_ISL_471158 | MRCG at LSHTM Genomics lab | MRCG at LSHTM Genomics lab | Sesay et al |
| EPI_ISL_471159 | MRCG at LSHTM Genomics lab | MRCG at LSHTM Genomics lab | Sesay et al |
| EPI_ISL_471160 | MRCG at LSHTM Genomics lab | MRCG at LSHTM Genomics lab | Sesay et al |
| EPI_ISL_471165 | MRCG at LSHTM Genomics lab | MRCG at LSHTM Genomics lab | Sesay et al |
| EPI_ISL_428855 | MRCG at LSHTM Geomics lab | MRCG at LSHTM Genomics lab | Sesay et al |
| EPI_ISL_428856 | MRCG at LSHTM Genomics Lab | MRCG at LSHTM Genomics lab | Sesay et al |
| EPI_ISL_954776 | National Institute of Laboratory Medicine and Referral Center | Genomic Research Lab, BCSIR | Shahina Akter et al |
| EPI_ISL_1233953 | Jamil-ur-Rahman Center for Genome Research, Dr. Panjwani Center for Molecular Medicine and Drug Research | Jamil-ur-Rahman Center for Genome Research, Dr. Panjwani Center for Molecular Medicine and Drug Research | Shakeel et al |
| EPI_ISL_779264 | Jamil-ur-Rahman Center for Genome Research, Dr. Panjwani Center for Molecular Medicine and Drug Research | Jamil-ur-Rahman Center for Genome Research, Dr. Panjwani Center for Molecular Medicine and Drug Research | Shakeel et al |
| EPI_ISL_779255 | Jamil-ur-Rahman Center for Genome Research, Dr. Panjwani Center for Molecular Medicine and Drug Research | Jamil-ur-Rahman Center for Genome Research, Dr. Panjwani Center for Molecular Medicine and Drug Research | Shakeel et al |
| EPI_ISL_779256 | Jamil-ur-Rahman Center for Genome Research, Dr. Panjwani Center for Molecular Medicine and Drug Research | Jamil-ur-Rahman Center for Genome Research, Dr. Panjwani Center for Molecular Medicine and Drug Research | Shakeel et al |
| EPI_ISL_779258 | Jamil-ur-Rahman Center for Genome Research, Dr. Panjwani Center for Molecular Medicine and Drug Research | Jamil-ur-Rahman Center for Genome Research, Dr. Panjwani Center for Molecular Medicine and Drug Research | Shakeel et al |
| EPI_ISL_779290 | Jamil-ur-Rahman Center for Genome Research, Dr. Panjwani Center for Molecular Medicine and Drug Research | Jamil-ur-Rahman Center for Genome Research, Dr. Panjwani Center for Molecular Medicine and Drug Research | Shakeel et al |
| EPI_ISL_779272 | Jamil-ur-Rahman Center for Genome Research, Dr. Panjwani Center for Molecular Medicine and Drug Research | Jamil-ur-Rahman Center for Genome Research, Dr. Panjwani Center for Molecular Medicine and Drug Research | Shakeel et al |
| EPI_ISL_779276 | Jamil-ur-Rahman Center for Genome Research, Dr. Panjwani Center for Molecular Medicine and Drug Research | Jamil-ur-Rahman Center for Genome Research, Dr. Panjwani Center for Molecular Medicine and Drug Research | Shakeel et al |
| EPI_ISL_779257 | Jamil-ur-Rahman Center for Genome Research, Dr. Panjwani Center for Molecular Medicine and Drug Research | Jamil-ur-Rahman Center for Genome Research, Dr. Panjwani Center for Molecular Medicine and Drug Research | Shakeel et al |
| EPI_ISL_779289 | Jamil-ur-Rahman Center for Genome Research, Dr. Panjwani Center for Molecular Medicine and Drug Research | Jamil-ur-Rahman Center for Genome Research, Dr. Panjwani Center for Molecular Medicine and Drug Research | Shakeel et al |
| EPI_ISL_779282 | Jamil-ur-Rahman Center for Genome Research, Dr. Panjwani Center for Molecular Medicine and Drug Research | Jamil-ur-Rahman Center for Genome Research, Dr. Panjwani Center for Molecular Medicine and Drug Research | Shakeel et al |
| EPI_ISL_779263 | Jamil-ur-Rahman Center for Genome Research, Dr. Panjwani Center for Molecular Medicine and Drug Research | Jamil-ur-Rahman Center for Genome Research, Dr. Panjwani Center for Molecular Medicine and Drug Research | Shakeel et al |
| EPI_ISL_451958 | Jamil-ur-Rahman Center for Genome Research, Dr. Panjwani Center for Molecular Medicine and Drug Research, International Center for Chemical and Biological Sciences, University of Karachi | Jamil-ur-Rahman Center for Genome Research, Dr. Panjwani Center for Molecular Medicine and Drug Research, International Center for Chemical and Biological Sciences, University of Karachi | Shakeel et al |
| EPI_ISL_413015 | Public Health Ontario Laboratory | National Microbiology Laboratory | Shari Tyson et al |
| EPI_ISL_678079 | Pathogen Genomics Lab King Abdullah University of Science and Technology (KAUST) | Pathogen Genomics Lab King Abdullah University of Science and Technology(KAUST) | Sharif Hala et al |
| EPI_ISL_510531 | Communicable Disease Laboratory, Public Health Directorate | Communicable Disease Laboratory, Public Health Directorate | Shehab et al |

| | | | |
|---|---|---|---|
| EPI_ISL_416344 | Shanghai Public Health Clinical Center, Shanghai Medical College, Fudan University | National Research Center for Translational Medicine (Shanghai), Ruijin Hospital affiliated to Shanghai Jiao Tong University School of Medicine & Shanghai Public Health Clinical Center | Shengyue Wang et al |
| EPI_ISL_416346 | Shanghai Public Health Clinical Center, Shanghai Medical College, Fudan University | National Research Center for Translational Medicine (Shanghai), Ruijin Hospital affiliated to Shanghai Jiao Tong University School of Medicine & Shanghai Public Health Clinical Center | Shengyue Wang et al |
| EPI_ISL_416356 | Shanghai Public Health Clinical Center, Shanghai Medical College, Fudan University | National Research Center for Translational Medicine (Shanghai), Ruijin Hospital affiliated to Shanghai Jiao Tong University School of Medicine & Shanghai Public Health Clinical Center | Shengyue Wang et al |
| EPI_ISL_416371 | Shanghai Public Health Clinical Center, Shanghai Medical College, Fudan University | National Research Center for Translational Medicine (Shanghai), Ruijin Hospital affiliated to Shanghai Jiao Tong University School of Medicine & Shanghai Public Health Clinical Center | Shengyue Wang et al |
| EPI_ISL_416385 | Shanghai Public Health Clinical Center, Shanghai Medical College, Fudan University | National Research Center for Translational Medicine (Shanghai), Ruijin Hospital affiliated to Shanghai Jiao Tong University School of Medicine & Shanghai Public Health Clinical Center | Shengyue Wang et al |
| EPI_ISL_1365364 | Laboratory of virology, National center of expertise | RSE National Center for Biotechnology and RSE National Center of Expertise | Shevtsov Alexandr et al |
| EPI_ISL_1334570 | Laboratory of virology, National center of expertise | RSE National Center for Biotechnology and RSE National Center of Expertise | Shevtsov Alexandr et al |
| EPI_ISL_1365365 | Laboratory of virology, National center of expertise | RSE National Center for Biotechnology and RSE National Center of Expertise | Shevtsov Alexandr et al |
| EPI_ISL_1364617 | Laboratory of virology, National center of expertise | RSE National Center for Biotechnology and RSE National Center of Expertise | Shevtsov Alexandr et al |
| EPI_ISL_1364865 | Laboratory of virology, National center of expertise | RSE National Center for Biotechnology and RSE National Center of Expertise | Shevtsov Alexandr et al |
| EPI_ISL_1364864 | Laboratory of virology, National center of expertise | RSE National Center for Biotechnology and RSE National Center of Expertise | Shevtsov Alexandr et al |
| EPI_ISL_1364879 | Laboratory of virology, National center of expertise | RSE National Center for Biotechnology and RSE National Center of Expertise | Shevtsov Alexandr et al |
| EPI_ISL_1334572 | Laboratory of virology, National center of expertise | RSE National Center for Biotechnology and RSE National Center of Expertise | Shevtsov Alexandr et al |
| EPI_ISL_1334580 | Laboratory of virology, National center of expertise | RSE National Center for Biotechnology and RSE National Center of Expertise | Shevtsov Alexandr et al |
| EPI_ISL_1364880 | Laboratory of virology, National center of expertise | RSE National Center for Biotechnology and RSE National Center of Expertise | Shevtsov Alexandr et al |
| EPI_ISL_1448018 | Laboratory of virology, National center of expertise | RSE National Center for Biotechnology and RSE National Center of Expertise | Shevtsov Alexandr et al |
| EPI_ISL_1334569 | Laboratory of virology, National center of expertise | RSE National Center for Biotechnology and RSE National Center of Expertise | Shevtsov Alexandr et al |
| EPI_ISL_1334574 | Laboratory of virology, National center of expertise | RSE National Center for Biotechnology and RSE National Center of Expertise | Shevtsov Alexandr et al |
| EPI_ISL_1448020 | Laboratory of virology, National center of expertise | RSE National Center for Biotechnology and RSE National Center of Expertise | Shevtsov Alexandr et al |
| EPI_ISL_408489 | Department of Laboratory Medicine, National Taiwan University Hospital | Microbial Genomics Core Lab, National Taiwan University Centers of Genomic and Precision Medicine | Shiou-Hwei Yeh et al |

| | | | |
|---|---|---|---|
| EPI_ISL_447622 | Department of Laboratory Medicine, National Taiwan University Hospital | Microbial Genomics Core Lab, National Taiwan University Centers of Genomic and Precision Medicine | Shiou-Hwei Yeh et al |
| EPI_ISL_534336 | Department of Laboratory Medicine, National Taiwan University Hospital | Microbial Genomics Core Lab, National Taiwan University Centers of Genomic and Precision Medicine | Shiou-Hwei Yeh et al |
| EPI_ISL_693303 | Department of Laboratory Medicine, National Taiwan University Hospital | Microbial Genomics Core Lab, National Taiwan University Centers of Genomic and Precision Medicine | Shiou-Hwei Yeh et al |
| EPI_ISL_693305 | Department of Laboratory Medicine, National Taiwan University Hospital | Microbial Genomics Core Lab, National Taiwan University Centers of Genomic and Precision Medicine | Shiou-Hwei Yeh et al |
| EPI_ISL_693306 | Department of Laboratory Medicine, National Taiwan University Hospital | Microbial Genomics Core Lab, National Taiwan University Centers of Genomic and Precision Medicine | Shiou-Hwei Yeh et al |
| EPI_ISL_738064 | Department of Laboratory Medicine, National Taiwan University Hospital | Microbial Genomics Core Lab, National Taiwan University Centers of Genomic and Precision Medicine | Shiou-Hwei Yeh et al |
| EPI_ISL_740547 | Department of Laboratory Medicine, National Taiwan University Hospital | Microbial Genomics Core Lab, National Taiwan University Centers of Genomic and Precision Medicine | Shiou-Hwei Yeh et al |
| EPI_ISL_872591 | Department of Laboratory Medicine, National Taiwan University Hospital | Microbial Genomics Core Lab, National Taiwan University Centers of Genomic and Precision Medicine | Shiou-Hwei Yeh et al |
| EPI_ISL_872592 | Department of Laboratory Medicine, National Taiwan University Hospital | Microbial Genomics Core Lab, National Taiwan University Centers of Genomic and Precision Medicine | Shiou-Hwei Yeh et al |
| EPI_ISL_872594 | Department of Laboratory Medicine, National Taiwan University Hospital | Microbial Genomics Core Lab, National Taiwan University Centers of Genomic and Precision Medicine | Shiou-Hwei Yeh et al |
| EPI_ISL_872597 | Department of Laboratory Medicine, National Taiwan University Hospital | Microbial Genomics Core Lab, National Taiwan University Centers of Genomic and Precision Medicine | Shiou-Hwei Yeh et al |
| EPI_ISL_872598 | Department of Laboratory Medicine, National Taiwan University Hospital | Microbial Genomics Core Lab, National Taiwan University Centers of Genomic and Precision Medicine | Shiou-Hwei Yeh et al |
| EPI_ISL_872599 | Department of Laboratory Medicine, National Taiwan University Hospital | Microbial Genomics Core Lab, National Taiwan University Centers of Genomic and Precision Medicine | Shiou-Hwei Yeh et al |
| EPI_ISL_1667469 | Department of Laboratory Medicine, National Taiwan University Hospital | Microbial Genomics Core Lab, National Taiwan University Centers of Genomic and Precision Medicine | Shiou-Hwei Yeh et al |
| EPI_ISL_1667471 | Department of Laboratory Medicine, National Taiwan University Hospital | Microbial Genomics Core Lab, National Taiwan University Centers of Genomic and Precision Medicine | Shiou-Hwei Yeh et al |
| EPI_ISL_1667472 | Department of Laboratory Medicine, National Taiwan University Hospital | Microbial Genomics Core Lab, National Taiwan University Centers of Genomic and Precision Medicine | Shiou-Hwei Yeh et al |
| EPI_ISL_1667474 | Department of Laboratory Medicine, National Taiwan University Hospital | Microbial Genomics Core Lab, National Taiwan University Centers of Genomic and Precision Medicine | Shiou-Hwei Yeh et al |
| EPI_ISL_1667475 | Department of Laboratory Medicine, National Taiwan University Hospital | Microbial Genomics Core Lab, National Taiwan University Centers of Genomic and Precision Medicine | Shiou-Hwei Yeh et al |
| EPI_ISL_796018 | Hebei Provincial Center for Disease Control and Prevention, Shijiazhuang, Hebei Province; National Institute for Viral Disease Control and Prevention, China CDC | Hebei Provincial Center for Disease Control and Prevention, Shijiazhuang, Hebei Province; National Institute for Viral Disease Control and Prevention, China CDC | Shunxiang Qi et al |
| EPI_ISL_796022 | Hebei Provincial Center for Disease Control and Prevention, Shijiazhuang, Hebei Province; National Institute for Viral Disease Control and Prevention, China CDC | Hebei Provincial Center for Disease Control and Prevention, Shijiazhuang, Hebei Province; National Institute for Viral Disease Control and Prevention, China CDC | Shunxiang Qi et al |
| EPI_ISL_796023 | Hebei Provincial Center for Disease Control and Prevention, Shijiazhuang, Hebei Province; National Institute for Viral Disease Control and Prevention, China CDC | Hebei Provincial Center for Disease Control and Prevention, Shijiazhuang, Hebei Province; National Institute for Viral Disease Control and Prevention, China CDC | Shunxiang Qi et al |

| | | | |
|---|---|---|---|
| EPI_ISL_1516870 | Botswana Harvard HIV Reference Laboratory | Botswana Harvard HIV Reference Laboratory | Sikhulile Dorcas Maruapula et al |
| EPI_ISL_1516871 | Botswana Harvard HIV Reference Laboratory | Botswana Harvard HIV Reference Laboratory | Sikhulile Dorcas Maruapula et al |
| EPI_ISL_1516874 | Botswana Harvard HIV Reference Laboratory | Botswana Harvard HIV Reference Laboratory | Sikhulile Dorcas Maruapula et al |
| EPI_ISL_1516877 | Botswana Harvard HIV Reference Laboratory | Botswana Harvard HIV Reference Laboratory | Sikhulile Dorcas Maruapula et al |
| EPI_ISL_949068 | Botswana Harvard HIV Reference Laboratory | Botswana Harvard HIV Reference Laboratory | Sikhulile Moyo et al |
| EPI_ISL_965264 | Botswana Harvard HIV Reference Laboratory | Botswana Harvard HIV Reference Laboratory | Sikhulile Moyo et al |
| EPI_ISL_968234 | Botswana Harvard HIV Reference Laboratory | Botswana Harvard HIV Reference Laboratory | Sikhulile Moyo et al |
| EPI_ISL_969034 | Botswana Harvard HIV Reference Laboratory | Botswana Harvard HIV Reference Laboratory | Sikhulile Moyo et al |
| EPI_ISL_1307686 | Botswana Harvard HIV Reference Laboratory | Botswana Harvard HIV Reference Laboratory | Sikhulile Moyo et al |
| EPI_ISL_1516834 | Botswana Harvard HIV Reference Laboratory | Botswana Harvard HIV Reference Laboratory | Sikhulile Wonderful T. Choga et al |
| EPI_ISL_1516835 | Botswana Harvard HIV Reference Laboratory | Botswana Harvard HIV Reference Laboratory | Sikhulile Wonderful T. Choga et al |
| EPI_ISL_1516836 | Botswana Harvard HIV Reference Laboratory | Botswana Harvard HIV Reference Laboratory | Sikhulile Wonderful T. Choga et al |
| EPI_ISL_1516837 | Botswana Harvard HIV Reference Laboratory | Botswana Harvard HIV Reference Laboratory | Sikhulile Wonderful T. Choga et al |
| EPI_ISL_1516838 | Botswana Harvard HIV Reference Laboratory | Botswana Harvard HIV Reference Laboratory | Sikhulile Wonderful T. Choga et al |
| EPI_ISL_1516853 | Botswana Harvard HIV Reference Laboratory | Botswana Harvard HIV Reference Laboratory | Sikhulile Wonderful T. Choga et al |
| EPI_ISL_1516829 | Botswana Harvard HIV Reference Laboratory | Botswana Harvard HIV Reference Laboratory | Sikhulile Wonderful T. Choga et al |
| EPI_ISL_1629722 | SOMER | Universidad Nacional de Colombia - Laboratorio Genómico One Health | Simón Villegas Velásquez et al |
| EPI_ISL_610210 | Department of Health Technology and Informatics, The Hong Kong Polytechnic University | Department of Health Technology and Informatics, The Hong Kong Polytechnic University | Siu et al |
| EPI_ISL_610216 | Department of Health Technology and Informatics, The Hong Kong Polytechnic University | Department of Health Technology and Informatics, The Hong Kong Polytechnic University | Siu et al |
| EPI_ISL_610225 | Department of Health Technology and Informatics, The Hong Kong Polytechnic University | Department of Health Technology and Informatics, The Hong Kong Polytechnic University | Siu et al |
| EPI_ISL_452334 | Laboratory of Infectious Diseases Center of Beijing Ditan Hospital | Laboratory of Infectious Diseases Center of Beijing Ditan Hospital | Siyuan Yang et al |
| EPI_ISL_452336 | Laboratory of Infectious Diseases Center of Beijing Ditan Hospital | Laboratory of Infectious Diseases Center of Beijing Ditan Hospital | Siyuan Yang et al |
| EPI_ISL_456598 | National Health Laboratory, Timor-Leste | Microbiological Diagnostic Unit Public Health Laboratory, The Peter Doherty Institute for Infection and Immunity | Soares da Silva et al |
| EPI_ISL_456601 | National Health Laboratory, Timor-Leste | Microbiological Diagnostic Unit Public Health Laboratory, The Peter Doherty Institute for Infection and Immunity | Soares da Silva et al |
| EPI_ISL_456602 | National Health Laboratory, Timor-Leste | Microbiological Diagnostic Unit Public Health Laboratory, The Peter Doherty Institute for Infection and Immunity | Soares da Silva et al |
| EPI_ISL_456604 | National Health Laboratory, Timor-Leste | Microbiological Diagnostic Unit Public Health Laboratory, The Peter Doherty Institute for Infection and Immunity | Soares da Silva et al |
| EPI_ISL_456605 | National Health Laboratory, Timor-Leste | Microbiological Diagnostic Unit Public Health Laboratory, The Peter Doherty Institute for Infection and Immunity | Soares da Silva et al |

| | | | |
|---|---|---|---|
| EPI_ISL_456610 | National Health Laboratory, Timor-Leste | Microbiological Diagnostic Unit Public Health Laboratory, The Peter Doherty Institute for Infection and Immunity | Soares da Silva et al |
| EPI_ISL_456599 | National Health Laboratory, Timor-Leste | Microbiological Diagnostic Unit Public Health Laboratory, The Peter Doherty Institute for Infection and Immunity | Soares da Silva et al |
| EPI_ISL_456612 | National Health Laboratory, Timor-Leste | Microbiological Diagnostic Unit Public Health Laboratory, The Peter Doherty Institute for Infection and Immunity | Soares da Silva et al |
| EPI_ISL_456600 | National Health Laboratory, Timor-Leste | Microbiological Diagnostic Unit Public Health Laboratory, The Peter Doherty Institute for Infection and Immunity | Soares da Silva et al |
| EPI_ISL_918368 | Virology Unit, Institut Pasteur du Cambodge | Virology Unit, Institut Pasteur du Cambodge | Sokhoun Yann et al |
| EPI_ISL_918366 | Virology Unit, Institut Pasteur du Cambodge | Virology Unit, Institut Pasteur du Cambodge | Sokhoun Yann et al |
| EPI_ISL_918359 | Virology Unit, Institut Pasteur du Cambodge | Virology Unit, Institut Pasteur du Cambodge | Sokhoun Yann et al |
| EPI_ISL_918361 | Virology Unit, Institut Pasteur du Cambodge | Virology Unit, Institut Pasteur du Cambodge | Sokhoun Yann et al |
| EPI_ISL_1532806 | Virology Unit, Institut Pasteur du Cambodge | Virology Unit, Institut Pasteur du Cambodge | Sokhoun Yann et al |
| EPI_ISL_1532811 | Virology Unit, Institut Pasteur du Cambodge | Virology Unit, Institut Pasteur du Cambodge | Sokhoun Yann et al |
| EPI_ISL_1534542 | Virology Unit, Institut Pasteur du Cambodge | Virology Unit, Institut Pasteur du Cambodge | Sokhoun Yann et al |
| EPI_ISL_1534540 | Virology Unit, Institut Pasteur du Cambodge | Virology Unit, Institut Pasteur du Cambodge | Sokhoun Yann et al |
| EPI_ISL_1534536 | Virology Unit, Institut Pasteur du Cambodge | Virology Unit, Institut Pasteur du Cambodge | Sokhoun Yann et al |
| EPI_ISL_1534543 | Virology Unit, Institut Pasteur du Cambodge | Virology Unit, Institut Pasteur du Cambodge | Sokhoun Yann et al |
| EPI_ISL_1532803 | Cambodian National Public Health Laboratory, National Institute of Public Health | Virology Unit, Institut Pasteur du Cambodge | Sokhoun Yann et al |
| EPI_ISL_1096139 | Cambodian National Public Health Laboratory, National Institute of Public Health | Virology Unit, Institut Pasteur du Cambodge | Sokhoun Yann et al |
| EPI_ISL_1096140 | Cambodian National Public Health Laboratory, National Institute of Public Health | Virology Unit, Institut Pasteur du Cambodge | Sokhoun Yann et al |
| EPI_ISL_576143 | Public Health Virology Laboratory, Forensic and Scientific Services (PHV-FSS) | Public Health Virology Laboratory, Forensic and Scientific Services (PHV-FSS) | Son Nguyen et al |
| EPI_ISL_693257 | Public Health Virology Laboratory, Forensic and Scientific Services (PHV-FSS) | Public Health Virology Laboratory, Forensic and Scientific Services (PHV-FSS) | Son Nguyen et al |
| EPI_ISL_693278 | Public Health Virology Laboratory, Forensic and Scientific Services (PHV-FSS) | Public Health Virology Laboratory, Forensic and Scientific Services (PHV-FSS) | Son Nguyen et al |
| EPI_ISL_849680 | Public Health Virology Laboratory, Forensic and Scientific Services (PHV-FSS) | Public Health Virology Laboratory, Forensic and Scientific Services (PHV-FSS) | Son Nguyen et al |
| EPI_ISL_849730 | Public Health Virology Laboratory, Forensic and Scientific Services (PHV-FSS) | Public Health Virology Laboratory, Forensic and Scientific Services (PHV-FSS) | Son Nguyen et al |
| EPI_ISL_849733 | Public Health Virology Laboratory, Forensic and Scientific Services (PHV-FSS) | Public Health Virology Laboratory, Forensic and Scientific Services (PHV-FSS) | Son Nguyen et al |
| EPI_ISL_1300529 | Public Health Virology-Forensic and Scientific Services (PHV-FSS) | Public Health Virology-Forensic and Scientific Services (PHV-FSS) | Son Nguyen et al |
| EPI_ISL_906056 | Tilia Laboratories s.r.o. | Tilia Laboratories s.r.o. | Sona Pekova et al |

| | | | |
|---|---|---|---|
| EPI_ISL_906063 | Tilia Laboratories s.r.o. | Tilia Laboratories s.r.o. | Sona Pekova et al |
| EPI_ISL_906064 | Tilia Laboratories s.r.o. | Tilia Laboratories s.r.o. | Sona Pekova et al |
| EPI_ISL_906054 | Tilia Laboratories s.r.o. | Tilia Laboratories s.r.o. | Sona Pekova et al |
| EPI_ISL_906057 | Tilia Laboratories s.r.o. | Tilia Laboratories s.r.o. | Sona Pekova et al |
| EPI_ISL_906062 | Tilia Laboratories s.r.o. | Tilia Laboratories s.r.o. | Sona Pekova et al |
| EPI_ISL_906053 | Tilia Laboratories s.r.o. | Tilia Laboratories s.r.o. | Sona Pekova et al |
| EPI_ISL_906055 | Tilia Laboratories s.r.o. | Tilia Laboratories s.r.o. | Sona Pekova MD et al |
| EPI_ISL_728237 | LNR National Reference Laboratory, Mohammed VI University of Health Sciences | Medical Biotechnology Laboratory, Rabat Medical and Pharmacy School, Mohammed The Vth University in Rabat | Souad KARTTI et al |
| EPI_ISL_1116467 | Laboratoire de Microbiologie- CHU Habib Bourguiba – Sfax | Laboratoire des Procédés de Criblage Moléculaire et Cellulaire-Centre de Biotechnologie de Sfax | Souissi et al |
| EPI_ISL_1118884 | Laboratoire de Microbiologie- CHU Habib Bourguiba – Sfax | Laboratoire des Procédés de Criblage Moléculaire et Cellulaire-Centre de Biotechnologie de Sfax | Souissi et al |
| EPI_ISL_1116464 | Laboratoire de Microbiologie- CHU Habib Bourguiba – Sfax | Laboratoire des Procédés de Criblage Moléculaire et Cellulaire-Centre de Biotechnologie de Sfax | Souissi et al |
| EPI_ISL_1116469 | Laboratoire de Microbiologie- CHU Habib Bourguiba – Sfax | Laboratoire des Procédés de Criblage Moléculaire et Cellulaire-Centre de Biotechnologie de Sfax | Souissi et al |
| EPI_ISL_1118923 | Laboratoire de Microbiologie- CHU Habib Bourguiba – Sfax | Laboratoire des Procédés de Criblage Moléculaire et Cellulaire-Centre de Biotechnologie de Sfax | Souissi et al |
| EPI_ISL_1116468 | Laboratoire de Microbiologie- CHU Habib Bourguiba – Sfax | Laboratoire des Procédés de Criblage Moléculaire et Cellulaire-Centre de Biotechnologie de Sfax | Souissi et al |
| EPI_ISL_710575 | Hôpital Fattouma-Bourguiba de Monastir | Laboratoire des Procédés de Criblage Moléculaire et Cellulaire-Centre de Biotechnologie de Sfax | Souissi et al |
| EPI_ISL_710537 | Hôpital Fattouma-Bourguiba de Monastir | Laboratoire des Procédés de Criblage Moléculaire et Cellulaire-Centre de Biotechnologie de Sfax | Souissi et al |
| EPI_ISL_711057 | Hôpital Fattouma-Bourguiba de Monastir | Laboratoire des Procédés de Criblage Moléculaire et Cellulaire-Centre de Biotechnologie de Sfax | Souissi et al |
| EPI_ISL_712069 | Laboratoire de Microbiologie- CHU Habib Bourguiba – Sfax adresse | Laboratoire des Procédés de Criblage Moléculaire et Cellulaire-Centre de Biotechnologie de Sfax | Souissi et al |
| EPI_ISL_712065 | Laboratoire de Microbiologie- CHU Habib Bourguiba – Sfax adresse | Laboratoire des Procédés de Criblage Moléculaire et Cellulaire-Centre de Biotechnologie de Sfax | Souissi et al |
| EPI_ISL_712066 | Laboratoire de Microbiologie- CHU Habib Bourguiba – Sfax adresse | Laboratoire des Procédés de Criblage Moléculaire et Cellulaire-Centre de Biotechnologie de Sfax | Souissi et al |
| EPI_ISL_1137609 | Département de Maladies Infectieuses, CHU Farhat Hached Sousse, Tunisie | Laboratoire des Procédés de Criblage Moléculaire et Cellulaire-Centre de Biotechnologie de Sfax | Souissi et al |
| EPI_ISL_1167128 | Iressef Genomics lab | L'institut de Recherche en Santé, de Surveillance Épidémiologique et de Formation (IRESSEF) | Souleymane MBOUP et al |
| EPI_ISL_1633465 | Institute for Health Research, Epidemiological Surveillance and Training (IRESSEF) | Abbott | Souleymane Mboup et al |
| EPI_ISL_1633466 | Institute for Health Research, Epidemiological Surveillance and Training (IRESSEF) | Abbott | Souleymane Mboup et al |

| | | | |
|---|---|---|---|
| EPI_ISL_1633467 | Institute for Health Research, Epidemiological Surveillance and Training (IRESSEF) | Abbott | Souleymane Mboup et al |
| EPI_ISL_1630272 | IRESSEF | Abbott | Souleymane Mboup et al |
| EPI_ISL_1630258 | IRESSEF | Abbott | Souleymane Mboup et al |
| EPI_ISL_1167181 | Iressef Genomics lab | L'institut de Recherche en Santé, de Surveillance Épidémiologique et de Formation (IRESSEF) | Souleymane MBOUP et al |
| EPI_ISL_1167131 | Iressef Genomics lab | L'institut de Recherche en Santé, de Surveillance Épidémiologique et de Formation (IRESSEF) | Souleymane MBOUP et al |
| EPI_ISL_1167125 | Iressef Genomics lab | L'institut de Recherche en Santé, de Surveillance Épidémiologique et de Formation (IRESSEF) | Souleymane MBOUP et al |
| EPI_ISL_1167162 | Iressef Genomics lab | L'institut de Recherche en Santé, de Surveillance Épidémiologique et de Formation (IRESSEF) | Souleymane MBOUP et al |
| EPI_ISL_1167163 | Iressef Genomics lab | L'institut de Recherche en Santé, de Surveillance Épidémiologique et de Formation (IRESSEF) | Souleymane MBOUP et al |
| EPI_ISL_1167134 | Iressef Genomics lab | L'institut de Recherche en Santé, de Surveillance Épidémiologique et de Formation (IRESSEF) | Souleymane MBOUP et al |
| EPI_ISL_1167127 | Iressef Genomics lab | L'institut de Recherche en Santé, de Surveillance Épidémiologique et de Formation (IRESSEF) | Souleymane MBOUP et al |
| EPI_ISL_1167171 | Iressef Genomics lab | L'institut de Recherche en Santé, de Surveillance Épidémiologique et de Formation (IRESSEF) | Souleymane MBOUP et al |
| EPI_ISL_1167126 | Iressef Genomics lab | L'institut de Recherche en Santé, de Surveillance Épidémiologique et de Formation (IRESSEF) | Souleymane MBOUP et al |
| EPI_ISL_1630257 | IRESSEF | Abbott | Souleymane Mboup et al |
| EPI_ISL_1630267 | IRESSEF | Abbott | Souleymane Mboup et al |
| EPI_ISL_1630268 | IRESSEF | Abbott | Souleymane Mboup et al |
| EPI_ISL_1261763 | Iressef Genomics lab | IRESSEF | Souleymane MBOUP et al |
| EPI_ISL_1630264 | IRESSEF | Abbott | Souleymane Mboup et al |
| EPI_ISL_636838 | Laboratório de Imunofarmacologia - Instituto Oswaldo Cruz | Laboratório de Imunofarmacologia - Instituto Oswaldo Cruz | Souza et al |
| EPI_ISL_1020200 | RS Trimitra, Bogor, West java | Biosafety Level-3 Laboratory, Indonesian Institute of Sciences (LIPI) | Sri Swasthikawati et al |
| EPI_ISL_413019 | Department of Internal Medicine, Triemli Hospital | Institute of Medical Virology, University of Zurich | Stefan Schmutz et al |
| EPI_ISL_413021 | Klinik Hirslanden Zurich | Institute of Medical Virology, University of Zurich | Stefan Schmutz et al |
| EPI_ISL_413023 | Division of Infectious Diseases, University Hospital Zurich | Institute of Medical Virology, University of Zurich | Stefan Schmutz et al |
| EPI_ISL_1068315 | Central Public Health Laboratory - LACEN -Bahia, Salvador, Brazil | Central Public Health Laboratory - LACEN -Bahia, Salvador, Brazil | Stephane Tosta et al |
| EPI_ISL_1068316 | Central Public Health Laboratory - LACEN -Bahia, Salvador, Brazil | Central Public Health Laboratory - LACEN -Bahia, Salvador, Brazil | Stephane Tosta et al |
| EPI_ISL_1068317 | Central Public Health Laboratory - LACEN -Bahia, Salvador, Brazil | Central Public Health Laboratory - LACEN -Bahia, Salvador, Brazil | Stephane Tosta et al |
| EPI_ISL_1068319 | Central Public Health Laboratory - LACEN -Bahia, Salvador, Brazil | Central Public Health Laboratory - LACEN -Bahia, Salvador, Brazil | Stephane Tosta et al |
| EPI_ISL_1068320 | Central Public Health Laboratory - LACEN -Bahia, Salvador, Brazil | Central Public Health Laboratory - LACEN -Bahia, Salvador, Brazil | Stephane Tosta et al |

| | | | |
|---|---|---|---|
| EPI_ISL_1068323 | Central Public Health Laboratory - LACEN -Bahia, Salvador, Brazil | Central Public Health Laboratory - LACEN -Bahia, Salvador, Brazil | Stephane Tosta et al |
| EPI_ISL_978495 | Central Public Health Laboratory - LACEN -Bahia, Salvador, Brazil | Central Public Health Laboratory - LACEN -Bahia, Salvador, Brazil | Stephane Tosta et al |
| EPI_ISL_1068385 | Central Public Health Laboratory - LACEN -Bahia, Salvador, Brazil | Central Public Health Laboratory - LACEN -Bahia, Salvador, Brazil | Stephane Tosta et al |
| EPI_ISL_1068387 | Central Public Health Laboratory - LACEN -Bahia, Salvador, Brazil | Central Public Health Laboratory - LACEN -Bahia, Salvador, Brazil | Stephane Tosta et al |
| EPI_ISL_1068388 | Central Public Health Laboratory - LACEN -Bahia, Salvador, Brazil | Central Public Health Laboratory - LACEN -Bahia, Salvador, Brazil | Stephane Tosta et al |
| EPI_ISL_1068389 | Central Public Health Laboratory - LACEN -Bahia, Salvador, Brazil | Central Public Health Laboratory - LACEN -Bahia, Salvador, Brazil | Stephane Tosta et al |
| EPI_ISL_1068390 | Central Public Health Laboratory - LACEN -Bahia, Salvador, Brazil | Central Public Health Laboratory - LACEN -Bahia, Salvador, Brazil | Stephane Tosta et al |
| EPI_ISL_978496 | Central Public Health Laboratory - LACEN -Bahia, Salvador, Brazil | Central Public Health Laboratory - LACEN -Bahia, Salvador, Brazil | Stephane Tosta et al |
| EPI_ISL_1068393 | Central Public Health Laboratory - LACEN -Bahia, Salvador, Brazil | Central Public Health Laboratory - LACEN -Bahia, Salvador, Brazil | Stephane Tosta et al |
| EPI_ISL_1068394 | Central Public Health Laboratory - LACEN -Bahia, Salvador, Brazil | Central Public Health Laboratory - LACEN -Bahia, Salvador, Brazil | Stephane Tosta et al |
| EPI_ISL_978497 | Central Public Health Laboratory - LACEN -Bahia, Salvador, Brazil | Central Public Health Laboratory - LACEN -Bahia, Salvador, Brazil | Stephane Tosta et al |
| EPI_ISL_978498 | Central Public Health Laboratory - LACEN -Bahia, Salvador, Brazil | Central Public Health Laboratory - LACEN -Bahia, Salvador, Brazil | Stephane Tosta et al |
| EPI_ISL_978499 | Central Public Health Laboratory - LACEN -Bahia, Salvador, Brazil | Central Public Health Laboratory - LACEN -Bahia, Salvador, Brazil | Stephane Tosta et al |
| EPI_ISL_978500 | Central Public Health Laboratory - LACEN -Bahia, Salvador, Brazil | Central Public Health Laboratory - LACEN -Bahia, Salvador, Brazil | Stephane Tosta et al |
| EPI_ISL_978502 | Central Public Health Laboratory - LACEN -Bahia, Salvador, Brazil | Central Public Health Laboratory - LACEN -Bahia, Salvador, Brazil | Stephane Tosta et al |
| EPI_ISL_978503 | Central Public Health Laboratory - LACEN -Bahia, Salvador, Brazil | Central Public Health Laboratory - LACEN -Bahia, Salvador, Brazil | Stephane Tosta et al |
| EPI_ISL_978504 | Central Public Health Laboratory - LACEN -Bahia, Salvador, Brazil | Central Public Health Laboratory - LACEN -Bahia, Salvador, Brazil | Stephane Tosta et al |
| EPI_ISL_978505 | Central Public Health Laboratory - LACEN -Bahia, Salvador, Brazil | Central Public Health Laboratory - LACEN -Bahia, Salvador, Brazil | Stephane Tosta et al |
| EPI_ISL_978506 | Central Public Health Laboratory - LACEN -Bahia, Salvador, Brazil | Central Public Health Laboratory - LACEN -Bahia, Salvador, Brazil | Stephane Tosta et al |
| EPI_ISL_978507 | Central Public Health Laboratory - LACEN -Bahia, Salvador, Brazil | Central Public Health Laboratory - LACEN -Bahia, Salvador, Brazil | Stephane Tosta et al |
| EPI_ISL_978508 | Central Public Health Laboratory - LACEN -Bahia, Salvador, Brazil | Central Public Health Laboratory - LACEN -Bahia, Salvador, Brazil | Stephane Tosta et al |
| EPI_ISL_978510 | Central Public Health Laboratory - LACEN -Bahia, Salvador, Brazil | Central Public Health Laboratory - LACEN -Bahia, Salvador, Brazil | Stephane Tosta et al |
| EPI_ISL_978512 | Central Public Health Laboratory - LACEN -Bahia, Salvador, Brazil | Central Public Health Laboratory - LACEN -Bahia, Salvador, Brazil | Stephane Tosta et al |
| EPI_ISL_978513 | Central Public Health Laboratory - LACEN -Bahia, Salvador, Brazil | Central Public Health Laboratory - LACEN -Bahia, Salvador, Brazil | Stephane Tosta et al |
| EPI_ISL_978514 | Central Public Health Laboratory - LACEN -Bahia, Salvador, Brazil | Central Public Health Laboratory - LACEN -Bahia, Salvador, Brazil | Stephane Tosta et al |
| EPI_ISL_978515 | Central Public Health Laboratory - LACEN -Bahia, Salvador, Brazil | Central Public Health Laboratory - LACEN -Bahia, Salvador, Brazil | Stephane Tosta et al |
| EPI_ISL_978516 | Central Public Health Laboratory - LACEN -Bahia, Salvador, Brazil | Central Public Health Laboratory - LACEN -Bahia, Salvador, Brazil | Stephane Tosta et al |
| EPI_ISL_978517 | Central Public Health Laboratory - LACEN -Bahia, Salvador, Brazil | Central Public Health Laboratory - LACEN -Bahia, Salvador, Brazil | Stephane Tosta et al |
| EPI_ISL_978518 | Central Public Health Laboratory - LACEN -Bahia, Salvador, Brazil | Central Public Health Laboratory - LACEN -Bahia, Salvador, Brazil | Stephane Tosta et al |
| EPI_ISL_978520 | Central Public Health Laboratory - LACEN -Bahia, Salvador, Brazil | Central Public Health Laboratory - LACEN -Bahia, Salvador, Brazil | Stephane Tosta et al |

| | | | |
|---|---|---|---|
| EPI_ISL_978522 | Central Public Health Laboratory - LACEN -Bahia, Salvador, Brazil | Central Public Health Laboratory - LACEN -Bahia, Salvador, Brazil | Stephane Tosta et al |
| EPI_ISL_978525 | Central Public Health Laboratory - LACEN -Bahia, Salvador, Brazil | Central Public Health Laboratory - LACEN -Bahia, Salvador, Brazil | Stephane Tosta et al |
| EPI_ISL_978528 | Central Public Health Laboratory - LACEN -Bahia, Salvador, Brazil | Central Public Health Laboratory - LACEN -Bahia, Salvador, Brazil | Stephane Tosta et al |
| EPI_ISL_978529 | Central Public Health Laboratory - LACEN -Bahia, Salvador, Brazil | Central Public Health Laboratory - LACEN -Bahia, Salvador, Brazil | Stephane Tosta et al |
| EPI_ISL_978530 | Central Public Health Laboratory - LACEN -Bahia, Salvador, Brazil | Central Public Health Laboratory - LACEN -Bahia, Salvador, Brazil | Stephane Tosta et al |
| EPI_ISL_978531 | Central Public Health Laboratory - LACEN -Bahia, Salvador, Brazil | Central Public Health Laboratory - LACEN -Bahia, Salvador, Brazil | Stephane Tosta et al |
| EPI_ISL_978532 | Central Public Health Laboratory - LACEN -Bahia, Salvador, Brazil | Central Public Health Laboratory - LACEN -Bahia, Salvador, Brazil | Stephane Tosta et al |
| EPI_ISL_1067737 | Central Public Health Laboratory - LACEN -Bahia, Salvador, Brazil | Central Public Health Laboratory - LACEN -Bahia, Salvador, Brazil | Stephane Tosta et al |
| EPI_ISL_1067730 | Central Public Health Laboratory - LACEN -Bahia, Salvador, Brazil | Central Public Health Laboratory - LACEN -Bahia, Salvador, Brazil | Stephane Tosta et al |
| EPI_ISL_1067738 | Central Public Health Laboratory - LACEN -Bahia, Salvador, Brazil | Central Public Health Laboratory - LACEN -Bahia, Salvador, Brazil | Stephane Tosta et al |
| EPI_ISL_1067731 | Central Public Health Laboratory - LACEN -Bahia, Salvador, Brazil | Central Public Health Laboratory - LACEN -Bahia, Salvador, Brazil | Stephane Tosta et al |
| EPI_ISL_1067733 | Central Public Health Laboratory - LACEN -Bahia, Salvador, Brazil | Central Public Health Laboratory - LACEN -Bahia, Salvador, Brazil | Stephane Tosta et al |
| EPI_ISL_1067729 | Central Public Health Laboratory - LACEN -Bahia, Salvador, Brazil | Central Public Health Laboratory - LACEN -Bahia, Salvador, Brazil | Stephane Tosta et al |
| EPI_ISL_1067734 | Central Public Health Laboratory - LACEN -Bahia, Salvador, Brazil | Central Public Health Laboratory - LACEN -Bahia, Salvador, Brazil | Stephane Tosta et al |
| EPI_ISL_1067735 | Central Public Health Laboratory - LACEN -Bahia, Salvador, Brazil | Central Public Health Laboratory - LACEN -Bahia, Salvador, Brazil | Stephane Tosta et al |
| EPI_ISL_1067736 | Center for Biotechnology and Cell Therapy, São Rafael Hospital, Salvador, Brazil | Central Public Health Laboratory - LACEN -Bahia, Salvador, Brazil | Stephane Tosta et al |
| EPI_ISL_1067728 | Center for Biotechnology and Cell Therapy, São Rafael Hospital, Salvador, Brazil | Central Public Health Laboratory - LACEN -Bahia, Salvador, Brazil | Stephane Tosta et al |
| EPI_ISL_1067732 | Center for Biotechnology and Cell Therapy, São Rafael Hospital, Salvador, Brazil | Central Public Health Laboratory - LACEN -Bahia, Salvador, Brazil | Stephane Tosta et al |
| EPI_ISL_1068363 | Central Public Health Laboratory - LACEN -Bahia, Salvador, Brazil | Central Public Health Laboratory - LACEN -Bahia, Salvador, Brazil | Stephane Tosta et al |
| EPI_ISL_1068364 | Central Public Health Laboratory - LACEN -Bahia, Salvador, Brazil | Central Public Health Laboratory - LACEN -Bahia, Salvador, Brazil | Stephane Tosta et al |
| EPI_ISL_1068366 | Central Public Health Laboratory - LACEN -Bahia, Salvador, Brazil | Central Public Health Laboratory - LACEN -Bahia, Salvador, Brazil | Stephane Tosta et al |
| EPI_ISL_1068367 | Central Public Health Laboratory - LACEN -Bahia, Salvador, Brazil | Central Public Health Laboratory - LACEN -Bahia, Salvador, Brazil | Stephane Tosta et al |
| EPI_ISL_1068368 | Central Public Health Laboratory - LACEN -Bahia, Salvador, Brazil | Central Public Health Laboratory - LACEN -Bahia, Salvador, Brazil | Stephane Tosta et al |
| EPI_ISL_1068369 | Central Public Health Laboratory - LACEN -Bahia, Salvador, Brazil | Central Public Health Laboratory - LACEN -Bahia, Salvador, Brazil | Stephane Tosta et al |
| EPI_ISL_1068370 | Central Public Health Laboratory - LACEN -Bahia, Salvador, Brazil | Central Public Health Laboratory - LACEN -Bahia, Salvador, Brazil | Stephane Tosta et al |
| EPI_ISL_1068371 | Central Public Health Laboratory - LACEN -Bahia, Salvador, Brazil | Central Public Health Laboratory - LACEN -Bahia, Salvador, Brazil | Stephane Tosta et al |
| EPI_ISL_1068372 | Central Public Health Laboratory - LACEN -Bahia, Salvador, Brazil | Central Public Health Laboratory - LACEN -Bahia, Salvador, Brazil | Stephane Tosta et al |
| EPI_ISL_1068373 | Central Public Health Laboratory - LACEN -Bahia, Salvador, Brazil | Central Public Health Laboratory - LACEN -Bahia, Salvador, Brazil | Stephane Tosta et al |
| EPI_ISL_1068374 | Central Public Health Laboratory - LACEN -Bahia, Salvador, Brazil | Central Public Health Laboratory - LACEN -Bahia, Salvador, Brazil | Stephane Tosta et al |

| | | | |
|---|---|---|---|
| EPI_ISL_1068375 | Central Public Health Laboratory - LACEN -Bahia, Salvador, Brazil | Central Public Health Laboratory - LACEN -Bahia, Salvador, Brazil | Stephane Tosta et al |
| EPI_ISL_1068376 | Central Public Health Laboratory - LACEN -Bahia, Salvador, Brazil | Central Public Health Laboratory - LACEN -Bahia, Salvador, Brazil | Stephane Tosta et al |
| EPI_ISL_1068377 | Central Public Health Laboratory - LACEN -Bahia, Salvador, Brazil | Central Public Health Laboratory - LACEN -Bahia, Salvador, Brazil | Stephane Tosta et al |
| EPI_ISL_1068378 | Central Public Health Laboratory - LACEN -Bahia, Salvador, Brazil | Central Public Health Laboratory - LACEN -Bahia, Salvador, Brazil | Stephane Tosta et al |
| EPI_ISL_1068379 | Central Public Health Laboratory - LACEN -Bahia, Salvador, Brazil | Central Public Health Laboratory - LACEN -Bahia, Salvador, Brazil | Stephane Tosta et al |
| EPI_ISL_1068380 | Central Public Health Laboratory - LACEN -Bahia, Salvador, Brazil | Central Public Health Laboratory - LACEN -Bahia, Salvador, Brazil | Stephane Tosta et al |
| EPI_ISL_1068382 | Central Public Health Laboratory - LACEN -Bahia, Salvador, Brazil | Central Public Health Laboratory - LACEN -Bahia, Salvador, Brazil | Stephane Tosta et al |
| EPI_ISL_1583642 | Central Public Health Laboratory - LACEN -Bahia, Salvador, Brazil | Central Public Health Laboratory - LACEN -Bahia, Salvador, Brazil | Stephane Tosta et al |
| EPI_ISL_1583643 | Central Public Health Laboratory - LACEN -Bahia, Salvador, Brazil | Central Public Health Laboratory - LACEN -Bahia, Salvador, Brazil | Stephane Tosta et al |
| EPI_ISL_1583644 | Central Public Health Laboratory - LACEN -Bahia, Salvador, Brazil | Central Public Health Laboratory - LACEN -Bahia, Salvador, Brazil | Stephane Tosta et al |
| EPI_ISL_1583645 | Central Public Health Laboratory - LACEN -Bahia, Salvador, Brazil | Central Public Health Laboratory - LACEN -Bahia, Salvador, Brazil | Stephane Tosta et al |
| EPI_ISL_1583646 | Central Public Health Laboratory - LACEN -Bahia, Salvador, Brazil | Central Public Health Laboratory - LACEN -Bahia, Salvador, Brazil | Stephane Tosta et al |
| EPI_ISL_1583647 | Central Public Health Laboratory - LACEN -Bahia, Salvador, Brazil | Central Public Health Laboratory - LACEN -Bahia, Salvador, Brazil | Stephane Tosta et al |
| EPI_ISL_1583648 | Central Public Health Laboratory - LACEN -Bahia, Salvador, Brazil | Central Public Health Laboratory - LACEN -Bahia, Salvador, Brazil | Stephane Tosta et al |
| EPI_ISL_1583649 | Central Public Health Laboratory - LACEN -Bahia, Salvador, Brazil | Central Public Health Laboratory - LACEN -Bahia, Salvador, Brazil | Stephane Tosta et al |
| EPI_ISL_1583651 | Central Public Health Laboratory - LACEN -Bahia, Salvador, Brazil | Central Public Health Laboratory - LACEN -Bahia, Salvador, Brazil | Stephane Tosta et al |
| EPI_ISL_1583652 | Central Public Health Laboratory - LACEN -Bahia, Salvador, Brazil | Central Public Health Laboratory - LACEN -Bahia, Salvador, Brazil | Stephane Tosta et al |
| EPI_ISL_1583654 | Central Public Health Laboratory - LACEN -Bahia, Salvador, Brazil | Central Public Health Laboratory - LACEN -Bahia, Salvador, Brazil | Stephane Tosta et al |
| EPI_ISL_1583656 | Central Public Health Laboratory - LACEN -Bahia, Salvador, Brazil | Central Public Health Laboratory - LACEN -Bahia, Salvador, Brazil | Stephane Tosta et al |
| EPI_ISL_1583659 | Central Public Health Laboratory - LACEN -Bahia, Salvador, Brazil | Central Public Health Laboratory - LACEN -Bahia, Salvador, Brazil | Stephane Tosta et al |
| EPI_ISL_1583660 | Central Public Health Laboratory - LACEN -Bahia, Salvador, Brazil | Central Public Health Laboratory - LACEN -Bahia, Salvador, Brazil | Stephane Tosta et al |
| EPI_ISL_1583661 | Central Public Health Laboratory - LACEN -Bahia, Salvador, Brazil | Central Public Health Laboratory - LACEN -Bahia, Salvador, Brazil | Stephane Tosta et al |
| EPI_ISL_1583663 | Central Public Health Laboratory - LACEN -Bahia, Salvador, Brazil | Central Public Health Laboratory - LACEN -Bahia, Salvador, Brazil | Stephane Tosta et al |
| EPI_ISL_1583664 | Central Public Health Laboratory - LACEN -Bahia, Salvador, Brazil | Central Public Health Laboratory - LACEN -Bahia, Salvador, Brazil | Stephane Tosta et al |
| EPI_ISL_1583665 | Central Public Health Laboratory - LACEN -Bahia, Salvador, Brazil | Central Public Health Laboratory - LACEN -Bahia, Salvador, Brazil | Stephane Tosta et al |
| EPI_ISL_1583668 | Central Public Health Laboratory - LACEN -Bahia, Salvador, Brazil | Central Public Health Laboratory - LACEN -Bahia, Salvador, Brazil | Stephane Tosta et al |
| EPI_ISL_1583669 | Central Public Health Laboratory - LACEN -Bahia, Salvador, Brazil | Central Public Health Laboratory - LACEN -Bahia, Salvador, Brazil | Stephane Tosta et al |
| EPI_ISL_1583670 | Central Public Health Laboratory - LACEN -Bahia, Salvador, Brazil | Central Public Health Laboratory - LACEN -Bahia, Salvador, Brazil | Stephane Tosta et al |
| EPI_ISL_1583672 | Central Public Health Laboratory - LACEN -Bahia, Salvador, Brazil | Central Public Health Laboratory - LACEN -Bahia, Salvador, Brazil | Stephane Tosta et al |
| EPI_ISL_1583673 | Central Public Health Laboratory - LACEN -Bahia, Salvador, Brazil | Central Public Health Laboratory - LACEN -Bahia, Salvador, Brazil | Stephane Tosta et al |

| | | | |
|---|---|---|---|
| EPI_ISL_1583675 | DNA Laboratory | Central Public Health Laboratory - LACEN -Bahia, Salvador, Brazil | Stephane Tosta et al |
| EPI_ISL_1583676 | Central Public Health Laboratory - LACEN -Bahia, Salvador, Brazil | Central Public Health Laboratory - LACEN -Bahia, Salvador, Brazil | Stephane Tosta et al |
| EPI_ISL_1583679 | Central Public Health Laboratory - LACEN -Bahia, Salvador, Brazil | Central Public Health Laboratory - LACEN -Bahia, Salvador, Brazil | Stephane Tosta et al |
| EPI_ISL_1583680 | Central Public Health Laboratory - LACEN -Bahia, Salvador, Brazil | Central Public Health Laboratory - LACEN -Bahia, Salvador, Brazil | Stephane Tosta et al |
| EPI_ISL_1583681 | Central Public Health Laboratory - LACEN -Bahia, Salvador, Brazil | Central Public Health Laboratory - LACEN -Bahia, Salvador, Brazil | Stephane Tosta et al |
| EPI_ISL_1583686 | Central Public Health Laboratory - LACEN -Bahia, Salvador, Brazil | Central Public Health Laboratory - LACEN -Bahia, Salvador, Brazil | Stephane Tosta et al |
| EPI_ISL_1583689 | Central Public Health Laboratory - LACEN -Bahia, Salvador, Brazil | Central Public Health Laboratory - LACEN -Bahia, Salvador, Brazil | Stephane Tosta et al |
| EPI_ISL_1583694 | Central Public Health Laboratory - LACEN -Bahia, Salvador, Brazil | Central Public Health Laboratory - LACEN -Bahia, Salvador, Brazil | Stephane Tosta et al |
| EPI_ISL_1583697 | Central Public Health Laboratory - LACEN -Bahia, Salvador, Brazil | Central Public Health Laboratory - LACEN -Bahia, Salvador, Brazil | Stephane Tosta et al |
| EPI_ISL_1583700 | Central Public Health Laboratory - LACEN -Bahia, Salvador, Brazil | Central Public Health Laboratory - LACEN -Bahia, Salvador, Brazil | Stephane Tosta et al |
| EPI_ISL_1583703 | Central Public Health Laboratory - LACEN -Bahia, Salvador, Brazil | Central Public Health Laboratory - LACEN -Bahia, Salvador, Brazil | Stephane Tosta et al |
| EPI_ISL_1583705 | Central Public Health Laboratory - LACEN -Bahia, Salvador, Brazil | Central Public Health Laboratory - LACEN -Bahia, Salvador, Brazil | Stephane Tosta et al |
| EPI_ISL_1583714 | Central Public Health Laboratory - LACEN -Bahia, Salvador, Brazil | Central Public Health Laboratory - LACEN -Bahia, Salvador, Brazil | Stephane Tosta et al |
| EPI_ISL_1583716 | Central Public Health Laboratory - LACEN -Bahia, Salvador, Brazil | Central Public Health Laboratory - LACEN -Bahia, Salvador, Brazil | Stephane Tosta et al |
| EPI_ISL_1583725 | Central Public Health Laboratory - LACEN -Bahia, Salvador, Brazil | Central Public Health Laboratory - LACEN -Bahia, Salvador, Brazil | Stephane Tosta et al |
| EPI_ISL_1583727 | Central Public Health Laboratory - LACEN -Bahia, Salvador, Brazil | Central Public Health Laboratory - LACEN -Bahia, Salvador, Brazil | Stephane Tosta et al |
| EPI_ISL_1583730 | Central Public Health Laboratory - LACEN -Bahia, Salvador, Brazil | Central Public Health Laboratory - LACEN -Bahia, Salvador, Brazil | Stephane Tosta et al |
| EPI_ISL_444008 | PHE South West Regional Laboratory, National Infection Service | Wellcome Sanger Institute for the COVID-19 Genomics UK (COG-UK) consortium | Stephanie Hutchings et al |
| EPI_ISL_447469 | Clinical Microbiology Laboratory, Sheba Medical Center | Stern Lab | Stern Lab et al |
| EPI_ISL_447464 | Clinical Microbiology Laboratory, Sheba Medical Center | Stern Lab | Stern Lab et al |
| EPI_ISL_447462 | Clinical Microbiology Laboratory, Sheba Medical Center | Stern Lab | Stern Lab et al |
| EPI_ISL_1425044 | Clalit Health Services Laboratories, Israel | Stern Lab | Stern Lab et al |
| EPI_ISL_1278926 | Clalit Health Services Laboratories, Israel | Stern Lab | Stern Lab et al |
| EPI_ISL_1278865 | Clalit Health Services Laboratories, Israel | Stern Lab | Stern Lab et al |
| EPI_ISL_1424934 | Clalit Health Services Laboratories, Israel | Stern Lab | Stern Lab et al |
| EPI_ISL_1424942 | Clalit Health Services Laboratories, Israel | Stern Lab | Stern Lab et al |
| EPI_ISL_1424974 | Clalit Health Services Laboratories, Israel | Stern Lab | Stern Lab et al |
| EPI_ISL_447398 | Clinical Microbiology Laboratory, The Baruch Padeh Medical Center, Poriya | Stern Lab | Stern Lab et al |
| EPI_ISL_1278872 | Clalit Health Services Laboratories, Israel | Stern Lab | Stern Lab et al |
| EPI_ISL_1278853 | Clalit Health Services Laboratories, Israel | Stern Lab | Stern Lab et al |

| | | | |
|---|---|---|---|
| EPI_ISL_1278775 | Clalit Health Services Laboratories, Israel | Stern Lab | Stern Lab et al |
| EPI_ISL_1424887 | Clalit Health Services Laboratories, Israel | Stern Lab | Stern Lab et al |
| EPI_ISL_447304 | Microbiology Division, Barzilai University Medical Center | Stern Lab | Stern Lab et al |
| EPI_ISL_447298 | Microbiology Division, Barzilai University Medical Center | Stern Lab | Stern Lab et al |
| EPI_ISL_447270 | Microbiology laboratory, Assuta Ashdod University-Affiliated Hospital | Stern Lab | Stern Lab et al |
| EPI_ISL_1222766 | Central Scientific Research Department, Gomel State Medical University, | Laboratory of Genomics and Bioinformatics of the Forest Research Institute of the NAS of Belarus | Stoma et al |
| EPI_ISL_1169052 | National Institute of Health Research and Development | National Institute of Health Research and Development | Subangkit et al |
| EPI_ISL_1534706 | National Institute Health Research and Development | National Institute of Health Research and Development | Subangkit et al |
| EPI_ISL_1469264 | National Institute Health Research and Development | National Institute of Health Research and Development | Subangkit et al |
| EPI_ISL_775599 | Balai Besar Veteriner Maros | National Institute of Health Research and Development | Subangkit et al |
| EPI_ISL_944762 | CSIR-Indian Institute of Chemical Biology, MEDICA Supercpecialty Hospital Kolkata | CSIR-Indian Institute of Chemical Biology, MEDICA Supercpecialty Hospital Kolkata | Sujay Krishna Maity et al |
| EPI_ISL_1265456 | BTKLPP Kelas I Makassar | Eijkman Institute for Molecular Biology, Ministry of Research and Technology/National Agency for Research and Innovation; National Institute of Health Research and Development | Sukma Oktavianthi et al |
| EPI_ISL_417482 | Institute of Microbiology, Universidad San Francisco de Quito | Institute of Microbiology, Universidad San Francisco de Quito | Sully Márquez et al |
| EPI_ISL_471267 | Hospital IESS Babahoyo | Institute of Microbiology, Universidad San Francisco de Quito | Sully Márquez et al |
| EPI_ISL_477015 | Institute of Microbiology, Universidad San Francisco de Quito | Institute of Microbiology, Universidad San Francisco de Quito | Sully Márquez et al |
| EPI_ISL_660531 | Institute of Microbiology, Universidad San Francisco de Quito | Institute of Microbiology, Universidad San Francisco de Quito | Sully Márquez et al |
| EPI_ISL_660532 | Institute of Microbiology, Universidad San Francisco de Quito | Institute of Microbiology, Universidad San Francisco de Quito | Sully Márquez et al |
| EPI_ISL_660533 | Institute of Microbiology, Universidad San Francisco de Quito | Institute of Microbiology, Universidad San Francisco de Quito | Sully Márquez et al |
| EPI_ISL_660534 | Institute of Microbiology, Universidad San Francisco de Quito | Institute of Microbiology, Universidad San Francisco de Quito | Sully Márquez et al |
| EPI_ISL_660535 | Institute of Microbiology, Universidad San Francisco de Quito | Institute of Microbiology, Universidad San Francisco de Quito | Sully Márquez et al |
| EPI_ISL_660536 | Institute of Microbiology, Universidad San Francisco de Quito | Institute of Microbiology, Universidad San Francisco de Quito | Sully Márquez et al |
| EPI_ISL_660538 | Institute of Microbiology, Universidad San Francisco de Quito | Institute of Microbiology, Universidad San Francisco de Quito | Sully Márquez et al |
| EPI_ISL_728202 | Institute of Microbiology, Universidad San Francisco de Quito | Institute of Microbiology, Universidad San Francisco de Quito | Sully Márquez et al |
| EPI_ISL_728204 | Institute of Microbiology, Universidad San Francisco de Quito | Institute of Microbiology, Universidad San Francisco de Quito | Sully Márquez et al |
| EPI_ISL_803098 | Institute of Microbiology, Universidad San Francisco de Quito | Institute of Microbiology, Universidad San Francisco de Quito | Sully Márquez et al |
| EPI_ISL_1372080 | Microbiology Department , Late Shri Lakhiram Agrawal Memory Hospital & College, Raigarh, Chhattisgarh | Institute of Life Sciences - INSACOG | Sunil K. Raghav et al |
| EPI_ISL_1372081 | Microbiology Department , Late Shri Lakhiram Agrawal Memory Hospital & College, Raigarh, Chhattisgarh | Institute of Life Sciences - INSACOG | Sunil K. Raghav et al |
| EPI_ISL_1372086 | Immunogenomics lab, Institute of Life Sciences, Bhubaneswar | Institute of Life Sciences - INSACOG | Sunil K. Raghav et al |

| | | | |
|---|---|---|---|
| EPI_ISL_1372087 | Immunogenomics lab, Institute of Life Sciences, Bhubaneswar | Institute of Life Sciences - INSACOG | Sunil K. Raghav et al |
| EPI_ISL_1663490 | Capital hospital, Bhubaneswar | Institute of Life Sciences - INSACOG | Sunil K. Raghav et al |
| EPI_ISL_1663491 | Capital hospital, Bhubaneswar | Institute of Life Sciences - INSACOG | Sunil K. Raghav et al |
| EPI_ISL_1663492 | Capital hospital, Bhubaneswar | Institute of Life Sciences - INSACOG | Sunil K. Raghav et al |
| EPI_ISL_1663538 | MGM Medical College, Jamshedpur | Institute of Life Sciences - INSACOG | Sunil K. Raghav et al |
| EPI_ISL_1063848 | REGIONAL VRDL, ICMR-RMRC BBSR | Immunogenomics lab, Institute of Life Sciences, Bhubaneswar | Sunil K. Raghav et al |
| EPI_ISL_481161 | Immunogenomics lab, Institute of Life Sciences, Bhubaneswar | Immunogenomics lab, Institute of Life Sciences, Bhubaneswar | Sunil Raghav et al |
| EPI_ISL_718291 | Institute for Medical Research, Infectious Disease Research Centre, National Institutes of Health, Ministry of Health Malaysia | Institute for Medical Research, Infectious Disease Research Centre, National Institutes of Health, Ministry of Health Malaysia | Suppiah J et al |
| EPI_ISL_718300 | Institute for Medical Research, Infectious Disease Research Centre, National Institutes of Health, Ministry of Health Malaysia | Institute for Medical Research, Infectious Disease Research Centre, National Institutes of Health, Ministry of Health Malaysia | Suppiah J et al |
| EPI_ISL_877329 | University College Sedaya International (UCSI University) | Institute for Medical Research, Infectious Disease Research Centre, National Institutes of Health, Ministry of Health Malaysia | Suppiah J et al |
| EPI_ISL_718284 | Institute for Medical Research, Infectious Disease Research Centre, National Institutes of Health, Ministry of Health Malaysia | Institute for Medical Research, Infectious Disease Research Centre, National Institutes of Health, Ministry of Health Malaysia | Suppiah J et al |
| EPI_ISL_728157 | Institute for Medical Research, Infectious Disease Research Centre, National Institutes of Health, Ministry of Health Malaysia | Institute for Medical Research, Infectious Disease Research Centre, National Institutes of Health, Ministry of Health Malaysia | Suppiah J et al |
| EPI_ISL_728161 | Institute for Medical Research, Infectious Disease Research Centre, National Institutes of Health, Ministry of Health Malaysia | Institute for Medical Research, Infectious Disease Research Centre, National Institutes of Health, Ministry of Health Malaysia | Suppiah J et al |
| EPI_ISL_728165 | Institute for Medical Research, Infectious Disease Research Centre, National Institutes of Health, Ministry of Health Malaysia | Institute for Medical Research, Infectious Disease Research Centre, National Institutes of Health, Ministry of Health Malaysia | Suppiah J et al |
| EPI_ISL_728241 | Institute for Medical Research, Infectious Disease Research Centre, National Institutes of Health, Ministry of Health Malaysia | Institute for Medical Research, Infectious Disease Research Centre, National Institutes of Health, Ministry of Health Malaysia | Suppiah J et al |
| EPI_ISL_728242 | Institute for Medical Research, Infectious Disease Research Centre, National Institutes of Health, Ministry of Health Malaysia | Institute for Medical Research, Infectious Disease Research Centre, National Institutes of Health, Ministry of Health Malaysia | Suppiah J et al |
| EPI_ISL_728248 | Institute for Medical Research, Infectious Disease Research Centre, National Institutes of Health, Ministry of Health Malaysia | Institute for Medical Research, Infectious Disease Research Centre, National Institutes of Health, Ministry of Health Malaysia | Suppiah J et al |
| EPI_ISL_489994 | Institute for Medical Research, Infectious Disease Research Centre, National Institutes of Health, Ministry of Health Malaysia | Institute for Medical Research, Infectious Disease Research Centre, National Institutes of Health, Ministry of Health Malaysia | Suppiah J et al |
| EPI_ISL_718270 | Institute for Medical Research, Infectious Disease Research Centre, National Institutes of Health, Ministry of Health Malaysia | Institute for Medical Research, Infectious Disease Research Centre, National Institutes of Health, Ministry of Health Malaysia | Suppiah J et al |
| EPI_ISL_718274 | Institute for Medical Research, Infectious Disease Research Centre, National Institutes of Health, Ministry of Health Malaysia | Institute for Medical Research, Infectious Disease Research Centre, National Institutes of Health, Ministry of Health Malaysia | Suppiah J et al |
| EPI_ISL_718272 | Institute for Medical Research, Infectious Disease Research Centre, National Institutes of Health, Ministry of Health Malaysia | Institute for Medical Research, Infectious Disease Research Centre, National Institutes of Health, Ministry of Health Malaysia | Suppiah J et al |
| EPI_ISL_718275 | Institute for Medical Research, Infectious Disease Research Centre, National Institutes of Health, Ministry of Health Malaysia | Institute for Medical Research, Infectious Disease Research Centre, National Institutes of Health, Ministry of Health Malaysia | Suppiah J et al |
| EPI_ISL_718276 | Institute for Medical Research, Infectious Disease Research Centre, National Institutes of Health, Ministry of Health Malaysia | Institute for Medical Research, Infectious Disease Research Centre, National Institutes of Health, Ministry of Health Malaysia | Suppiah J et al |
| EPI_ISL_718278 | Institute for Medical Research, Infectious Disease Research Centre, National Institutes of Health, Ministry of Health Malaysia | Institute for Medical Research, Infectious Disease Research Centre, National Institutes of Health, Ministry of Health Malaysia | Suppiah J et al |

| | | | |
|---|---|---|---|
| EPI_ISL_718279 | Institute for Medical Research, Infectious Disease Research Centre, National Institutes of Health, Ministry of Health Malaysia | Institute for Medical Research, Infectious Disease Research Centre, National Institutes of Health, Ministry of Health Malaysia | Suppiah J et al |
| EPI_ISL_718311 | Institute for Medical Research, Infectious Disease Research Centre, National Institutes of Health, Ministry of Health Malaysia | Institute for Medical Research, Infectious Disease Research Centre, National Institutes of Health, Ministry of Health Malaysia | Suppiah J et al |
| EPI_ISL_1186028 | Institute for Medical Research, Infectious Disease Research Centre, National Institutes of Health, Ministry of Health Malaysia | Institute for Medical Research, Infectious Disease Research Centre, National Institutes of Health, Ministry of Health Malaysia | Suppiah J et al |
| EPI_ISL_1585270 | Labuan Hospital | Institute for Medical Research, Infectious Disease Research Centre, National Institutes of Health, Ministry of Health Malaysia | Suppiah J et al |
| EPI_ISL_644672 | Institute for Medical Research, Infectious Disease Research Centre, National Institutes of Health, Ministry of Health Malaysia | Institute for Medical Research, Infectious Disease Research Centre, National Institutes of Health, Ministry of Health Malaysia | Suppiah J et al |
| EPI_ISL_1424130 | Institute for Medical Research, Infectious Disease Research Centre, National Institutes of Health, Ministry of Health Malaysia | Institute for Medical Research, Infectious Disease Research Centre, National Institutes of Health, Ministry of Health Malaysia | Suppiah J et al |
| EPI_ISL_1263458 | Institute for Medical Research, Infectious Disease Research Centre, National Institutes of Health, Ministry of Health Malaysia | Institute for Medical Research, Infectious Disease Research Centre, National Institutes of Health, Ministry of Health Malaysia | Suppiah J et al |
| EPI_ISL_1424143 | Institute for Medical Research, Infectious Disease Research Centre, National Institutes of Health, Ministry of Health Malaysia | Institute for Medical Research, Infectious Disease Research Centre, National Institutes of Health, Ministry of Health Malaysia | Suppiah J et al |
| EPI_ISL_1424466 | Institute for Medical Research, Infectious Disease Research Centre, National Institutes of Health, Ministry of Health Malaysia | Institute for Medical Research, Infectious Disease Research Centre, National Institutes of Health, Ministry of Health Malaysia | Suppiah J et al |
| EPI_ISL_1264762 | Institute for Medical Research, Infectious Disease Research Centre, National Institutes of Health, Ministry of Health Malaysia | Institute for Medical Research, Infectious Disease Research Centre, National Institutes of Health, Ministry of Health Malaysia | Suppiah J et al |
| EPI_ISL_1424471 | Institute for Medical Research, Infectious Disease Research Centre, National Institutes of Health, Ministry of Health Malaysia | Institute for Medical Research, Infectious Disease Research Centre, National Institutes of Health, Ministry of Health Malaysia | Suppiah J et al |
| EPI_ISL_1424470 | Institute for Medical Research, Infectious Disease Research Centre, National Institutes of Health, Ministry of Health Malaysia | Institute for Medical Research, Infectious Disease Research Centre, National Institutes of Health, Ministry of Health Malaysia | Suppiah J et al |
| EPI_ISL_1424468 | Institute for Medical Research, Infectious Disease Research Centre, National Institutes of Health, Ministry of Health Malaysia | Institute for Medical Research, Infectious Disease Research Centre, National Institutes of Health, Ministry of Health Malaysia | Suppiah J et al |
| EPI_ISL_1424467 | Institute for Medical Research, Infectious Disease Research Centre, National Institutes of Health, Ministry of Health Malaysia | Institute for Medical Research, Infectious Disease Research Centre, National Institutes of Health, Ministry of Health Malaysia | Suppiah J et al |
| EPI_ISL_1424513 | Institute for Medical Research, Infectious Disease Research Centre, National Institutes of Health, Ministry of Health Malaysia | Institute for Medical Research, Infectious Disease Research Centre, National Institutes of Health, Ministry of Health Malaysia | Suppiah J et al |
| EPI_ISL_1583006 | Institute for Medical Research, Infectious Disease Research Centre, National Institutes of Health, Ministry of Health Malaysia | Institute for Medical Research, Infectious Disease Research Centre, National Institutes of Health, Ministry of Health Malaysia | Suppiah J et al |
| EPI_ISL_1583007 | Institute for Medical Research, Infectious Disease Research Centre, National Institutes of Health, Ministry of Health Malaysia | Institute for Medical Research, Infectious Disease Research Centre, National Institutes of Health, Ministry of Health Malaysia | Suppiah J et al |
| EPI_ISL_912397 | Institute for Medical Research, Infectious Disease Research Centre, National Institutes of Health, Ministry of Health Malaysia | Institute for Medical Research, Infectious Disease Research Centre, National Institutes of Health, Ministry of Health Malaysia | Suppiah J et al |
| EPI_ISL_944096 | Institute for Medical Research, Infectious Disease Research Centre, National Institutes of Health, Ministry of Health Malaysia | Institute for Medical Research, Infectious Disease Research Centre, National Institutes of Health, Ministry of Health Malaysia | Suppiah J et al |
| EPI_ISL_944103 | Institute for Medical Research, Infectious Disease Research Centre, National Institutes of Health, Ministry of Health Malaysia | Institute for Medical Research, Infectious Disease Research Centre, National Institutes of Health, Ministry of Health Malaysia | Suppiah J et al |
| EPI_ISL_944102 | Institute for Medical Research, Infectious Disease Research Centre, National Institutes of Health, Ministry of Health Malaysia | Institute for Medical Research, Infectious Disease Research Centre, National Institutes of Health, Ministry of Health Malaysia | Suppiah J et al |
| EPI_ISL_944104 | Institute for Medical Research, Infectious Disease Research Centre, National Institutes of Health, Ministry of Health Malaysia | Institute for Medical Research, Infectious Disease Research Centre, National Institutes of Health, Ministry of Health Malaysia | Suppiah J et al |

| | | | |
|---|---|---|---|
| EPI_ISL_944099 | Institute for Medical Research, Infectious Disease Research Centre, National Institutes of Health, Ministry of Health Malaysia | Institute for Medical Research, Infectious Disease Research Centre, National Institutes of Health, Ministry of Health Malaysia | Suppiah J et al |
| EPI_ISL_1138750 | Institute for Medical Research, Infectious Disease Research Centre, National Institutes of Health, Ministry of Health Malaysia | Institute for Medical Research, Infectious Disease Research Centre, National Institutes of Health, Ministry of Health Malaysia | Suppiah J et al |
| EPI_ISL_1138598 | Institute for Medical Research, Infectious Disease Research Centre, National Institutes of Health, Ministry of Health Malaysia | Institute for Medical Research, Infectious Disease Research Centre, National Institutes of Health, Ministry of Health Malaysia | Suppiah J et al |
| EPI_ISL_1263540 | Institute for Medical Research, Infectious Disease Research Centre, National Institutes of Health, Ministry of Health Malaysia | Institute for Medical Research, Infectious Disease Research Centre, National Institutes of Health, Ministry of Health Malaysia | Suppiah J et al |
| EPI_ISL_1265161 | Institute for Medical Research, Infectious Disease Research Centre, National Institutes of Health, Ministry of Health Malaysia | Institute for Medical Research, Infectious Disease Research Centre, National Institutes of Health, Ministry of Health Malaysia | Suppiah J et al |
| EPI_ISL_490094 | Institute for Medical Research, Infectious Disease Research Centre, National Institutes of Health, Ministry of Health Malaysia | Institute for Medical Research, Infectious Disease Research Centre, National Institutes of Health, Ministry of Health Malaysia | Suppiah J et al |
| EPI_ISL_490096 | Institute for Medical Research, Infectious Disease Research Centre, National Institutes of Health, Ministry of Health Malaysia | Institute for Medical Research, Infectious Disease Research Centre, National Institutes of Health, Ministry of Health Malaysia | Suppiah J et al |
| EPI_ISL_490099 | Institute for Medical Research, Infectious Disease Research Centre, National Institutes of Health, Ministry of Health Malaysia | Institute for Medical Research, Infectious Disease Research Centre, National Institutes of Health, Ministry of Health Malaysia | Suppiah J et al |
| EPI_ISL_718269 | Institute for Medical Research, Infectious Disease Research Centre, National Institutes of Health, Ministry of Health Malaysia | Institute for Medical Research, Infectious Disease Research Centre, National Institutes of Health, Ministry of Health Malaysia | Suppiah J et al |
| EPI_ISL_936494 | Hospital Enche Besar Hajjah Khalsom Kluang | Institute for Medical Research, Infectious Disease Research Centre, National Institutes of Health, Ministry of Health Malaysia | Suppiah J et al |
| EPI_ISL_936496 | Institute for Medical Research, Infectious Disease Research Centre, National Institutes of Health, Ministry of Health Malaysia | Institute for Medical Research, Infectious Disease Research Centre, National Institutes of Health, Ministry of Health Malaysia | Suppiah J et al |
| EPI_ISL_430443 | Institute for Medical Research, Infectious Disease Research Centre, National Institutes of Health, Ministry of Health Malaysia | Institute for Medical Research, Infectious Disease Research Centre, National Institutes of Health, Ministry of Health Malaysia | Suppiah.J et al |
| EPI_ISL_430441 | Institute for Medical Research, Infectious Disease Research Centre, National Institutes of Health, Ministry of Health Malaysia | Institute for Medical Research, Infectious Disease Research Centre, National Institutes of Health, Ministry of Health Malaysia | Suppiah.J et al |
| EPI_ISL_430444 | Institute for Medical Research, Infectious Disease Research Centre, National Institutes of Health, Ministry of Health Malaysia | Institute for Medical Research, Infectious Disease Research Centre, National Institutes of Health, Ministry of Health Malaysia | Suppiah.J et al |
| EPI_ISL_745162 | Elsies River CHC wc ERP | National Health Laboratory Service (NHLS), Tygerberg | Susan Engelbrecht et al |
| EPI_ISL_745185 | Tygerberg Hospital wc TBH | National Health Laboratory Service (NHLS), Tygerberg | Susan Engelbrecht et al |
| EPI_ISL_912401 | PathCare | National Health Laboratory Service (NHLS), Tygerberg | Susan Engelbrecht et al |
| EPI_ISL_1591408 | Division of Medical Virology, National Health Laboratory Service (NHLS), Tygerberg Hospital / Stellenbosch University | Division of Medical Virology, Stellenbosch University and NHLS Tygerberg Hospital | Susan Engelbrecht et al |
| EPI_ISL_1502148 | Division of Medical Virology, National Health Laboratory Service (NHLS), Tygerberg Hospital / Stellenbosch University | Division of Medical Virology, Stellenbosch University and NHLS Tygerberg Hospital | Susan Engelbrecht et al |
| EPI_ISL_1502167 | Division of Medical Virology, National Health Laboratory Service (NHLS), Tygerberg Hospital / Stellenbosch University | Division of Medical Virology, Stellenbosch University and NHLS Tygerberg Hospital | Susan Engelbrecht et al |
| EPI_ISL_491296 | Instituto Gulbenkian de Ciência | Instituto Gulbenkian de Ciência | Susana Ladeiro et al |
| EPI_ISL_855569 | Department of Virology, Principal Military Hospital of Instruction of Tunis | Bundeswehr Institute of Microbiology | Susann Handrick et al |
| EPI_ISL_855560 | Department of Virology, Principal Military Hospital of Instruction of Tunis | Bundeswehr Institute of Microbiology | Susann Handrick et al |
| EPI_ISL_855557 | Department of Virology, Principal Military Hospital of Instruction of Tunis | Bundeswehr Institute of Microbiology | Susann Handrick et al |

| | | | |
|---|---|---|---|
| EPI_ISL_1661454 | Microvida | Microvida | Suzan D. Pas et al |
| EPI_ISL_510535 | Molecular Virology, Instituto Carlos Chagas / Fiocruz Paraná | Universidade Federal do Parana (UFPR) | Suzukawa et al |
| EPI_ISL_1617677 | Swedish national genomic surveillance program of SARS-CoV-2 | The Public Health Agency of Sweden | Swedish national genomic surveillance prog |
| EPI_ISL_1606241 | Swedish national genomic surveillance program of SARS-CoV-2 | The Public Health Agency of Sweden | Swedish national genomic surveillance prog |
| EPI_ISL_1364175 | Swedish national genomic surveillance program of SARS-CoV-2 | The Public Health Agency of Sweden | Swedish national genomic surveillance prog |
| EPI_ISL_1618017 | Swedish national genomic surveillance program of SARS-CoV-2 | The Public Health Agency of Sweden | Swedish national genomic surveillance prog |
| EPI_ISL_1601927 | Swedish national genomic surveillance program of SARS-CoV-2 | The Public Health Agency of Sweden | Swedish national genomic surveillance prog |
| EPI_ISL_1605767 | Swedish national genomic surveillance program of SARS-CoV-2 | The Public Health Agency of Sweden | Swedish national genomic surveillance prog |
| EPI_ISL_1606420 | Swedish national genomic surveillance program of SARS-CoV-2 | The Public Health Agency of Sweden | Swedish national genomic surveillance prog |
| EPI_ISL_1604430 | Swedish national genomic surveillance program of SARS-CoV-2 | The Public Health Agency of Sweden | Swedish national genomic surveillance prog |
| EPI_ISL_1290087 | Swedish national genomic surveillance program of SARS-CoV-2 | The Public Health Agency of Sweden | Swedish national genomic surveillance prog |
| EPI_ISL_1290603 | Swedish national genomic surveillance program of SARS-CoV-2 | The Public Health Agency of Sweden | Swedish national genomic surveillance prog |
| EPI_ISL_1290605 | Swedish national genomic surveillance program of SARS-CoV-2 | The Public Health Agency of Sweden | Swedish national genomic surveillance prog |
| EPI_ISL_1200012 | Swedish national genomic surveillance program of SARS-CoV-2 | The Public Health Agency of Sweden | Swedish national genomic surveillance prog |
| EPI_ISL_1200011 | Swedish national genomic surveillance program of SARS-CoV-2 | The Public Health Agency of Sweden | Swedish national genomic surveillance prog |
| EPI_ISL_833497 | Klinik Apotek Dein, Jakarta, Indonesia | Biosafety Level-3 Laboratory, Indonesian Institute of Sciences (LIPI) | Syam Budi Iryanto et al |
| EPI_ISL_560587 | Hopital | National Reference Center for Viruses of Respiratory Infections, Institut Pasteur, Paris | Sylvie Behillil et al |
| EPI_ISL_560590 | Hopital | National Reference Center for Viruses of Respiratory Infections, Institut Pasteur, Paris | Sylvie Behillil et al |
| EPI_ISL_560636 | Hospital | National Reference Center for Viruses of Respiratory Infections, Institut Pasteur, Paris | Sylvie Behillil et al |
| EPI_ISL_560637 | Labo Analyses Med | National Reference Center for Viruses of Respiratory Infections, Institut Pasteur, Paris | Sylvie Behillil et al |
| EPI_ISL_593911 | Hospital, Le Chesnay Cédex | National Reference Center for Viruses of Respiratory Infections, Institut Pasteur, Paris | Sylvie Behillil et al |
| EPI_ISL_593929 | Labo Analyses Med, Puteaux | National Reference Center for Viruses of Respiratory Infections, Institut Pasteur, Paris | Sylvie Behillil et al |
| EPI_ISL_1627241 | Wojewodzka Stacja Sanitarno-Epidemiologiczna w Olsztynie, Laboratorium Badan Epidemiologiczno-Klinicznych | Wojewodzka Stacja Sanitarno-Epidemiologiczna w Olsztynie, Laboratorium Badan Epidemiologiczno-Klinicznych | Sylwia Krzetowska et al |
| EPI_ISL_1627090 | Pro-Vitam Diagnostics and Research Laboratory | Pro-Vitam Diagnostics and Research Laboratory | Szilard N. Fejer et al |
| EPI_ISL_582032 | Institute of Human Genetics, Polish Academy of Sciences | Institute of Human Genetics, Polish Academy of Sciences | Szymon Hryhorowicz et al |
| EPI_ISL_485399 | Institute of Human Genetics, Polish Academy of Sciences | Institute of Human Genetics, Polish Academy of Sciences | Szymon Hryhorowicz et al |
| EPI_ISL_455432 | Instituto de Diagnostico y Referencia Epidemiologicos (INDRE) | Instituto de Diagnostico y Referencia Epidemiologicos (INDRE) | Taboada Ramírez Blanca. Ramirez-Gonzale |
| EPI_ISL_847827 | COVID-19 National Reference Laboratory | COVID-19 National Reference Laboratory | Tahmineh Jalali et al |

gram of SARS-CoV-2 et al
gram of SARS-CoV-2 et al
gram of SARS-CoV-2 et al
gram of SARS-CoV-2 et al
gram of SARS-CoV-2 et al
gram of SARS-CoV-2 et al
gram of SARS-CoV-2 et al
gram of SARS-CoV-2 et al
gram of SARS-CoV-2 et al
gram of SARS-CoV-2 et al
gram of SARS-CoV-2 et al
gram of SARS-CoV-2 et al
gram of SARS-CoV-2 et al

ez Ernesto et al

| | | | |
|---|---|---|---|
| EPI_ISL_429676 | Central Public Health Laboratory/Octávio Magalhães Institute (IOM) from the Ezequiel Dias Foundation (FUNED) | Instituto Octávio Magalhães / Fundação Ezequiel Dias (IOM/Funed) | Talita Adelino et al |
| EPI_ISL_429679 | Central Public Health Laboratory/Octávio Magalhães Institute (IOM) from the Ezequiel Dias Foundation (FUNED) | Instituto Octávio Magalhães / Fundação Ezequiel Dias (IOM/Funed) | Talita Adelino et al |
| EPI_ISL_429665 | Central Public Health Laboratory/Octávio Magalhães Institute (IOM) from the Ezequiel Dias Foundation (FUNED) | Instituto Octávio Magalhães / Fundação Ezequiel Dias (IOM/Funed) | Talita Adelino et al |
| EPI_ISL_429684 | Central Public Health Laboratory/Octávio Magalhães Institute (IOM) from the Ezequiel Dias Foundation (FUNED) | Instituto Octávio Magalhães / Fundação Ezequiel Dias (IOM/Funed) | Talita Adelino et al |
| EPI_ISL_429688 | Central Public Health Laboratory/Octávio Magalhães Institute (IOM) from the Ezequiel Dias Foundation (FUNED) | Instituto Octávio Magalhães / Fundação Ezequiel Dias (IOM/Funed) | Talita Adelino et al |
| EPI_ISL_429689 | Central Public Health Laboratory/Octávio Magalhães Institute (IOM) from the Ezequiel Dias Foundation (FUNED) | Instituto Octávio Magalhães / Fundação Ezequiel Dias (IOM/Funed) | Talita Adelino et al |
| EPI_ISL_429701 | Central Public Health Laboratory/Octávio Magalhães Institute (IOM) from the Ezequiel Dias Foundation (FUNED) | Instituto Octávio Magalhães / Fundação Ezequiel Dias (IOM/Funed) | Talita Adelino et al |
| EPI_ISL_429671 | Central Public Health Laboratory/Octávio Magalhães Institute (IOM) from the Ezequiel Dias Foundation (FUNED) | Instituto Octávio Magalhães / Fundação Ezequiel Dias (IOM/Funed) | Talita Adelino et al |
| EPI_ISL_429672 | Central Public Health Laboratory/Octávio Magalhães Institute (IOM) from the Ezequiel Dias Foundation (FUNED) | Instituto Octávio Magalhães / Fundação Ezequiel Dias (IOM/Funed) | Talita Adelino et al |
| EPI_ISL_943561 | National Institute of Laboratory Medicine and Referral Center | Genomic Research Lab, BCSIR | Tanjina Akhtar Banu et al |
| EPI_ISL_1544143 | SSG Medical College, Baroda | Gujarat Biotechnology Research Centre | Tanuja B. Javadekar et al |
| EPI_ISL_651545 | Oxford Viromics, NDM, University of Oxford; Oxford University Hospitals; Basingstoke and North Hampshire Hospital | COVID-19 Genomics UK (COG-UK) Consortium | Tanya Golubchik et al |
| EPI_ISL_906851 | Respiratory Viruses Branch, Centers for Disease Control and Prevention | Respiratory Viruses Branch, Centers for Disease Control and Prevention | Tao et al |
| EPI_ISL_477169 | Department for Virology, Molecular Biology and Genome Research, R. G. Lugar Center for Public Health Research, National Center for Disease Control and Public Health (NCDC) of Georgia. | Department for Virology, Molecular Biology and Genome Research, R. G. Lugar Center for Public Health Research, National Center for Disease Control and Public Health (NCDC) of Georgia. | Tata Imnadze et al |
| EPI_ISL_1280095 | Department of Genetics, Medirex | Laboratory of Genomics and Bioinformatics, Comenius University Science Park | Tatiana Sedláčková et al |
| EPI_ISL_1280161 | Department of Genetics, Medirex | Laboratory of Genomics and Bioinformatics, Comenius University Science Park | Tatiana Sedláčková et al |
| EPI_ISL_1280115 | Department of Genetics, Medirex | Laboratory of Genomics and Bioinformatics, Comenius University Science Park | Tatiana Sedláčková et al |
| EPI_ISL_1280124 | Department of Genetics, Medirex | Laboratory of Genomics and Bioinformatics, Comenius University Science Park | Tatiana Sedláčková et al |
| EPI_ISL_1280140 | Department of Genetics, Medirex | Laboratory of Genomics and Bioinformatics, Comenius University Science Park | Tatiana Sedláčková et al |
| EPI_ISL_1280146 | Department of Genetics, Medirex | Laboratory of Genomics and Bioinformatics, Comenius University Science Park | Tatiana Sedláčková et al |
| EPI_ISL_1280151 | Department of Genetics, Medirex | Laboratory of Genomics and Bioinformatics, Comenius University Science Park | Tatiana Sedláčková et al |
| EPI_ISL_1280152 | Department of Genetics, Medirex | Laboratory of Genomics and Bioinformatics, Comenius University Science Park | Tatiana Sedláčková et al |

| | | | |
|---|---|---|---|
| EPI_ISL_1280121 | Department of Genetics, Medirex | Laboratory of Genomics and Bioinformatics, Comenius University Science Park | Tatiana Sedláčková et al |
| EPI_ISL_1599311 | Public Health Authority of the Slovak Republic | Laboratory of Genomics and Bioinformatics, Comenius University Science Park | Tatiana Sedláčková et al |
| EPI_ISL_1599337 | Public Health Authority of the Slovak Republic | Laboratory of Genomics and Bioinformatics, Comenius University Science Park | Tatiana Sedláčková et al |
| EPI_ISL_1653929 | Public Health Authority of the Slovak Republic | Laboratory of Genomics and Bioinformatics, Comenius University Science Park | Tatiana Sedláčková et al |
| EPI_ISL_1647652 | Public Health Authority of the Slovak Republic | Laboratory of Genomics and Bioinformatics, Comenius University Science Park | Tatiana Sedláčková et al |
| EPI_ISL_1647755 | Public Health Authority of the Slovak Republic | Laboratory of Genomics and Bioinformatics, Comenius University Science Park | Tatiana Sedláčková et al |
| EPI_ISL_1647718 | Public Health Authority of the Slovak Republic | Laboratory of Genomics and Bioinformatics, Comenius University Science Park | Tatiana Sedláčková et al |
| EPI_ISL_1647687 | Public Health Authority of the Slovak Republic | Laboratory of Genomics and Bioinformatics, Comenius University Science Park | Tatiana Sedláčková et al |
| EPI_ISL_1401572 | Public Health Authority of the Slovak Republic | Laboratory of Genomics and Bioinformatics, Comenius University Science Park | Tatiana Sedláčková et al |
| EPI_ISL_1599275 | Public Health Authority of the Slovak Republic | Laboratory of Genomics and Bioinformatics, Comenius University Science Park | Tatiana Sedláčková et al |
| EPI_ISL_1524015 | Public Health Authority of the Slovak Republic | Laboratory of Genomics and Bioinformatics, Comenius University Science Park | Tatiana Sedláčková et al |
| EPI_ISL_1524032 | Public Health Authority of the Slovak Republic | Laboratory of Genomics and Bioinformatics, Comenius University Science Park | Tatiana Sedláčková et al |
| EPI_ISL_407079 | Lapland Central Hospital | Department of Virology, University of Helsinki and Helsinki University Hospital, Helsinki, Finland | Teemu Smura et al |
| EPI_ISL_1496712 | Department of Virology and Immunology, University of Helsinki and Helsinki University Hospital, Huslab Finland | Department of Virology, Faculty of Medicine, University of Helsinki, Helsinki, Finland | Teemu Smura et al |
| EPI_ISL_481533 | Department of Virology and Immunology, University of Helsinki and Helsinki University Hospital, Huslab Finland | Department of Virology, Faculty of Medicine, University of Helsinki, Helsinki, Finland | Teemu Smura et al |
| EPI_ISL_1218646 | Department of Virology and Immunology, University of Helsinki and Helsinki University Hospital, Huslab Finland | Department of Virology, Faculty of Medicine, University of Helsinki, Helsinki, Finland | Teemu Smura et al |
| EPI_ISL_1218648 | Department of Virology and Immunology, University of Helsinki and Helsinki University Hospital, Huslab Finland | Department of Virology, Faculty of Medicine, University of Helsinki, Helsinki, Finland | Teemu Smura et al |
| EPI_ISL_732569 | Department of Virology and Immunology, University of Helsinki and Helsinki University Hospital, Huslab Finland | Department of Virology, Faculty of Medicine, University of Helsinki, Helsinki, Finland | Teemu Smura et al |
| EPI_ISL_1595762 | Department of Virology and Immunology, University of Helsinki and Helsinki University Hospital, Huslab Finland | Department of Virology, Faculty of Medicine, University of Helsinki, Helsinki, Finland | Teemu Smura et al |
| EPI_ISL_1595773 | Department of Virology and Immunology, University of Helsinki and Helsinki University Hospital, Huslab Finland | Department of Virology, Faculty of Medicine, University of Helsinki, Helsinki, Finland | Teemu Smura et al |
| EPI_ISL_1595775 | Department of Virology and Immunology, University of Helsinki and Helsinki University Hospital, Huslab Finland | Department of Virology, Faculty of Medicine, University of Helsinki, Helsinki, Finland | Teemu Smura et al |
| EPI_ISL_1595791 | Department of Virology and Immunology, University of Helsinki and Helsinki University Hospital, Huslab Finland | Department of Virology, Faculty of Medicine, University of Helsinki, Helsinki, Finland | Teemu Smura et al |

| | | | |
|---|---|---|---|
| EPI_ISL_756247 | Department of Virology and Immunology, University of Helsinki and Helsinki University Hospital, Huslab Finland | Department of Virology, Faculty of Medicine, University of Helsinki, Helsinki, Finland | Teemu Smura et al |
| EPI_ISL_737210 | Department of Virology and Immunology, University of Helsinki and Helsinki University Hospital, Huslab Finland | Department of Virology, Faculty of Medicine, University of Helsinki, Helsinki, Finland | Teemu Smura et al |
| EPI_ISL_737228 | Department of Virology and Immunology, University of Helsinki and Helsinki University Hospital, Huslab Finland | Department of Virology, Faculty of Medicine, University of Helsinki, Helsinki, Finland | Teemu Smura et al |
| EPI_ISL_1217198 | Department of Virology and Immunology, University of Helsinki and Helsinki University Hospital, Huslab Finland | Department of Virology, Faculty of Medicine, University of Helsinki, Helsinki, Finland | Teemu Smura et al |
| EPI_ISL_737234 | Department of Virology and Immunology, University of Helsinki and Helsinki University Hospital, Huslab Finland | Department of Virology, Faculty of Medicine, University of Helsinki, Helsinki, Finland | Teemu Smura et al |
| EPI_ISL_1497002 | Department of Virology and Immunology, University of Helsinki and Helsinki University Hospital, Huslab Finland | Department of Virology, Faculty of Medicine, University of Helsinki, Helsinki, Finland | Teemu Smura et al |
| EPI_ISL_1219684 | Department of Virology and Immunology, University of Helsinki and Helsinki University Hospital, Huslab Finland | Department of Virology, Faculty of Medicine, University of Helsinki, Helsinki, Finland | Teemu Smura et al |
| EPI_ISL_1219696 | Department of Virology and Immunology, University of Helsinki and Helsinki University Hospital, Huslab Finland | Department of Virology, Faculty of Medicine, University of Helsinki, Helsinki, Finland | Teemu Smura et al |
| EPI_ISL_1219721 | Department of Virology and Immunology, University of Helsinki and Helsinki University Hospital, Huslab Finland | Department of Virology, Faculty of Medicine, University of Helsinki, Helsinki, Finland | Teemu Smura et al |
| EPI_ISL_757302 | Department of Virology and Immunology, University of Helsinki and Helsinki University Hospital, Huslab Finland | Department of Virology, Faculty of Medicine, University of Helsinki, Helsinki, Finland | Teemu Smura et al |
| EPI_ISL_756284 | Department of Virology and Immunology, University of Helsinki and Helsinki University Hospital, Huslab Finland | Department of Virology, Faculty of Medicine, University of Helsinki, Helsinki, Finland | Teemu Smura et al |
| EPI_ISL_757336 | Department of Virology and Immunology, University of Helsinki and Helsinki University Hospital, Huslab Finland | Department of Virology, Faculty of Medicine, University of Helsinki, Helsinki, Finland | Teemu Smura et al |
| EPI_ISL_757377 | Department of Virology and Immunology, University of Helsinki and Helsinki University Hospital, Huslab Finland | Department of Virology, Faculty of Medicine, University of Helsinki, Helsinki, Finland | Teemu Smura et al |
| EPI_ISL_757379 | Department of Virology and Immunology, University of Helsinki and Helsinki University Hospital, Huslab Finland | Department of Virology, Faculty of Medicine, University of Helsinki, Helsinki, Finland | Teemu Smura et al |
| EPI_ISL_1233202 | Department of Virology and Immunology, University of Helsinki and Helsinki University Hospital, Huslab Finland | Department of Virology, Faculty of Medicine, University of Helsinki, Helsinki, Finland | Teemu Smura et al |
| EPI_ISL_1233188 | Department of Virology and Immunology, University of Helsinki and Helsinki University Hospital, Huslab Finland | Department of Virology, Faculty of Medicine, University of Helsinki, Helsinki, Finland | Teemu Smura et al |
| EPI_ISL_759839 | Department of Virology and Immunology, University of Helsinki and Helsinki University Hospital, Huslab Finland | Department of Virology, Faculty of Medicine, University of Helsinki, Helsinki, Finland | Teemu Smura et al |
| EPI_ISL_759940 | Department of Virology and Immunology, University of Helsinki and Helsinki University Hospital, Huslab Finland | Department of Virology, Faculty of Medicine, University of Helsinki, Helsinki, Finland | Teemu Smura et al |
| EPI_ISL_902786 | Department of Virology and Immunology, University of Helsinki and Helsinki University Hospital, Huslab Finland | Department of Virology, Faculty of Medicine, University of Helsinki, Helsinki, Finland | Teemu Smura et al |
| EPI_ISL_1240078 | Department of Virology and Immunology, University of Helsinki and Helsinki University Hospital, Huslab Finland | Department of Virology, Faculty of Medicine, University of Helsinki, Helsinki, Finland | Teemu Smura et al |
| EPI_ISL_1240150 | Department of Virology and Immunology, University of Helsinki and Helsinki University Hospital, Huslab Finland | Department of Virology, Faculty of Medicine, University of Helsinki, Helsinki, Finland | Teemu Smura et al |
| EPI_ISL_1240168 | Department of Virology and Immunology, University of Helsinki and Helsinki University Hospital, Huslab Finland | Department of Virology, Faculty of Medicine, University of Helsinki, Helsinki, Finland | Teemu Smura et al |

| | | | |
|---|---|---|---|
| EPI_ISL_1240166 | Department of Virology and Immunology, University of Helsinki and Helsinki University Hospital, Huslab Finland | Department of Virology, Faculty of Medicine, University of Helsinki, Helsinki, Finland | Teemu Smura et al |
| EPI_ISL_862052 | Department of Virology and Immunology, University of Helsinki and Helsinki University Hospital, Huslab Finland | Department of Virology, Faculty of Medicine, University of Helsinki, Helsinki, Finland | Teemu Smura et al |
| EPI_ISL_1240950 | Department of Virology and Immunology, University of Helsinki and Helsinki University Hospital, Huslab Finland | Department of Virology, Faculty of Medicine, University of Helsinki, Helsinki, Finland | Teemu Smura et al |
| EPI_ISL_1240964 | Department of Virology and Immunology, University of Helsinki and Helsinki University Hospital, Huslab Finland | Department of Virology, Faculty of Medicine, University of Helsinki, Helsinki, Finland | Teemu Smura et al |
| EPI_ISL_481705 | Department of Virology and Immunology, University of Helsinki and Helsinki University Hospital, Huslab Finland | Department of Virology, Faculty of Medicine, University of Helsinki, Helsinki, Finland | Teemu Smura et al |
| EPI_ISL_1240972 | Department of Virology and Immunology, University of Helsinki and Helsinki University Hospital, Huslab Finland | Department of Virology, Faculty of Medicine, University of Helsinki, Helsinki, Finland | Teemu Smura et al |
| EPI_ISL_1240984 | Department of Virology and Immunology, University of Helsinki and Helsinki University Hospital, Huslab Finland | Department of Virology, Faculty of Medicine, University of Helsinki, Helsinki, Finland | Teemu Smura et al |
| EPI_ISL_481718 | Department of Virology and Immunology, University of Helsinki and Helsinki University Hospital, Huslab Finland | Department of Virology, Faculty of Medicine, University of Helsinki, Helsinki, Finland | Teemu Smura et al |
| EPI_ISL_755956 | Department of Virology and Immunology, University of Helsinki and Helsinki University Hospital, Huslab Finland | Department of Virology, Faculty of Medicine, University of Helsinki, Helsinki, Finland | Teemu Smura et al |
| EPI_ISL_412971 | HUS Diagnostiikkakeskus, Hallinto | Department of Virology Faculty of Medicine, Medicum University of Helsinki | Teemu Smura et al |
| EPI_ISL_1279111 | Department of Virology and Immunology, University of Helsinki and Helsinki University Hospital, Huslab Finland | Department of Virology, Faculty of Medicine, University of Helsinki, Helsinki, Finland | Teemu Smura et al |
| EPI_ISL_1279107 | Department of Virology and Immunology, University of Helsinki and Helsinki University Hospital, Huslab Finland | Department of Virology, Faculty of Medicine, University of Helsinki, Helsinki, Finland | Teemu Smura et al |
| EPI_ISL_1278956 | Department of Virology and Immunology, University of Helsinki and Helsinki University Hospital, Huslab Finland | Department of Virology, Faculty of Medicine, University of Helsinki, Helsinki, Finland | Teemu Smura et al |
| EPI_ISL_1278946 | Department of Virology and Immunology, University of Helsinki and Helsinki University Hospital, Huslab Finland | Department of Virology, Faculty of Medicine, University of Helsinki, Helsinki, Finland | Teemu Smura et al |
| EPI_ISL_1278953 | Department of Virology and Immunology, University of Helsinki and Helsinki University Hospital, Huslab Finland | Department of Virology, Faculty of Medicine, University of Helsinki, Helsinki, Finland | Teemu Smura et al |
| EPI_ISL_732630 | Department of Virology and Immunology, University of Helsinki and Helsinki University Hospital, Huslab Finland | Department of Virology, Faculty of Medicine, University of Helsinki, Helsinki, Finland | Teemu Smura et al |
| EPI_ISL_737062 | Department of Virology and Immunology, University of Helsinki and Helsinki University Hospital, Huslab Finland | Department of Virology, Faculty of Medicine, University of Helsinki, Helsinki, Finland | Teemu Smura et al |
| EPI_ISL_555586 | Lighthouse Lab in Alderley Park | Wellcome Sanger Institute for the COVID-19 Genomics UK (COG-UK) consortium | The Lighthouse Lab in Alderley Park et al |
| EPI_ISL_558200 | Lighthouse Lab in Alderley Park | Wellcome Sanger Institute for the COVID-19 Genomics UK (COG-UK) consortium | The Lighthouse Lab in Alderley Park et al |
| EPI_ISL_551467 | Lighthouse Lab in Alderley Park | Wellcome Sanger Institute for the COVID-19 Genomics UK (COG-UK) consortium | The Lighthouse Lab in Alderley Park et al |
| EPI_ISL_552985 | Lighthouse Lab in Alderley Park | Wellcome Sanger Institute for the COVID-19 Genomics UK (COG-UK) consortium | The Lighthouse Lab in Alderley Park et al |
| EPI_ISL_552955 | Lighthouse Lab in Alderley Park | Wellcome Sanger Institute for the COVID-19 Genomics UK (COG-UK) consortium | The Lighthouse Lab in Alderley Park et al |

| | | | |
|---|---|---|---|
| EPI_ISL_555725 | Lighthouse Lab in Alderley Park | Wellcome Sanger Institute for the COVID-19 Genomics UK (COG-UK) consortium | The Lighthouse Lab in Alderley Park et al |
| EPI_ISL_554084 | Lighthouse Lab in Alderley Park | Wellcome Sanger Institute for the COVID-19 Genomics UK (COG-UK) consortium | The Lighthouse Lab in Alderley Park et al |
| EPI_ISL_553594 | Lighthouse Lab in Alderley Park | Wellcome Sanger Institute for the COVID-19 Genomics UK (COG-UK) consortium | The Lighthouse Lab in Alderley Park et al |
| EPI_ISL_1084155 | Lighthouse Lab in Milton Keynes | Wellcome Sanger Institute for the COVID-19 Genomics UK (COG-UK) Consortium | The Lighthouse Lab in Milton Keynes et al |
| EPI_ISL_1084042 | Lighthouse Lab in Milton Keynes | Wellcome Sanger Institute for the COVID-19 Genomics UK (COG-UK) Consortium | The Lighthouse Lab in Milton Keynes et al |
| EPI_ISL_550429 | Lighthouse Lab in Milton Keynes | Wellcome Sanger Institute for the COVID-19 Genomics UK (COG-UK) consortium | The Lighthouse Lab in Milton Keynes et al |
| EPI_ISL_552436 | Lighthouse Lab in Milton Keynes | Wellcome Sanger Institute for the COVID-19 Genomics UK (COG-UK) consortium | The Lighthouse Lab in Milton Keynes et al |
| EPI_ISL_549833 | Lighthouse Lab in Milton Keynes | Wellcome Sanger Institute for the COVID-19 Genomics UK (COG-UK) consortium | The Lighthouse Lab in Milton Keynes et al |
| EPI_ISL_631036 | Lighthouse Lab in Milton Keynes | Wellcome Sanger Institute for the COVID-19 Genomics UK (COG-UK) consortium | The Lighthouse Lab in Milton Keynes et al |
| EPI_ISL_630998 | Lighthouse Lab in Milton Keynes | Wellcome Sanger Institute for the COVID-19 Genomics UK (COG-UK) consortium | The Lighthouse Lab in Milton Keynes et al |
| EPI_ISL_760963 | Lighthouse Lab in Milton Keynes | Wellcome Sanger Institute for the COVID-19 Genomics UK (COG-UK) Consortium | The Lighthouse Lab in Milton Keynes et al |
| EPI_ISL_762436 | Lighthouse Lab in Milton Keynes | Wellcome Sanger Institute for the COVID-19 Genomics UK (COG-UK) Consortium | The Lighthouse Lab in Milton Keynes et al |
| EPI_ISL_762502 | Lighthouse Lab in Milton Keynes | Wellcome Sanger Institute for the COVID-19 Genomics UK (COG-UK) Consortium | The Lighthouse Lab in Milton Keynes et al |
| EPI_ISL_924202 | Virology Department, Sheffield Teaching Hospitals NHS Foundation Trust/Department of Infection, Immunity and Cardiovascular Disease, The Medical School, University of Sheffield | COVID-19 Genomics UK (COG-UK) Consortium | Thushan de Silva et al |
| EPI_ISL_420268 | Virology Department, Sheffield Teaching Hospitals NHS Foundation Trust | Department of Infection, Immunity and Cardiovascular Disease, The Florey Institute, The Medical School, University of Sheffield | Thushan de Silva et al |
| EPI_ISL_1233628 | Labormedizinisches Zentrum Dr Risch | University Hospital Basel, Clinical Bacteriology | Tim Roloff et al |
| EPI_ISL_1233629 | Labormedizinisches Zentrum Dr Risch | University Hospital Basel, Clinical Bacteriology | Tim Roloff et al |
| EPI_ISL_1233630 | Labormedizinisches Zentrum Dr Risch | University Hospital Basel, Clinical Bacteriology | Tim Roloff et al |
| EPI_ISL_1233663 | Labormedizinisches Zentrum Dr Risch | University Hospital Basel, Clinical Bacteriology | Tim Roloff et al |
| EPI_ISL_1233664 | Labormedizinisches Zentrum Dr Risch | University Hospital Basel, Clinical Bacteriology | Tim Roloff et al |
| EPI_ISL_1233691 | Labormedizinisches Zentrum Dr Risch | University Hospital Basel, Clinical Bacteriology | Tim Roloff et al |
| EPI_ISL_1233692 | Labormedizinisches Zentrum Dr Risch | University Hospital Basel, Clinical Bacteriology | Tim Roloff et al |
| EPI_ISL_1299921 | Labormedizinisches Zentrum Dr Risch | University Hospital Basel, Clinical Bacteriology | Tim Roloff et al |
| EPI_ISL_1299915 | Labormedizinisches Zentrum Dr Risch | University Hospital Basel, Clinical Bacteriology | Tim Roloff et al |

| | | | |
|---|---|---|---|
| EPI_ISL_1036127 | Labormedizinisches Zentrum Dr Risch | University Hospital Basel, Clinical Bacteriology | Tim Roloff et al |
| EPI_ISL_1036070 | Labormedizinisches Zentrum Dr Risch | University Hospital Basel, Clinical Bacteriology | Tim Roloff et al |
| EPI_ISL_830810 | University Hospital Basel, Clinical Virology | University Hospital Basel, Clinical Bacteriology | Tim Roloff et al |
| EPI_ISL_1014691 | University Hospital Basel, Clinical Virology | University Hospital Basel, Clinical Bacteriology | Tim Roloff et al |
| EPI_ISL_450408 | unknown | Microbiology | To et al |
| EPI_ISL_1260768 | National Laboratory for Health, Environment and Food | Department for Public Health Microbiology Ljubljana, National Laboratory for Health, Environment and Food | Tom Koritnik et al |
| EPI_ISL_1402180 | Department of Public Health Microbiology Ljubljana, National Laboratory for Health, Environment and Food | Department of Public Health Microbiology Ljubljana, National Laboratory for Health, Environment and Food | Tom Koritnik et al |
| EPI_ISL_1654321 | Department of Public Health Microbiology Ljubljana, National Laboratory for Health, Environment and Food | Department for Public Health Microbiology Ljubljana, National Laboratory for Health, Environment and Food | Tom Koritnik et al |
| EPI_ISL_636460 | Institute of Microbiology and Immunology, Faculty of Medicine, University of Ljubljana | Institute of Microbiology and Immunology, Faculty of Medicine, University of Ljubljana | Tomaž Mark Zorec et al |
| EPI_ISL_635272 | Institute of Microbiology and Immunology, Faculty of Medicine, University of Ljubljana | Institute of Microbiology and Immunology, Faculty of Medicine, University of Ljubljana | Tomaž Mark Zorec et al |
| EPI_ISL_635279 | Institute of Microbiology and Immunology, Faculty of Medicine, University of Ljubljana | Institute of Microbiology and Immunology, Faculty of Medicine, University of Ljubljana | Tomaž Mark Zorec et al |
| EPI_ISL_635283 | Institute of Microbiology and Immunology, Faculty of Medicine, University of Ljubljana | Institute of Microbiology and Immunology, Faculty of Medicine, University of Ljubljana | Tomaž Mark Zorec et al |
| EPI_ISL_635297 | Institute of Microbiology and Immunology, Faculty of Medicine, University of Ljubljana | Institute of Microbiology and Immunology, Faculty of Medicine, University of Ljubljana | Tomaž Mark Zorec et al |
| EPI_ISL_458235 | KU Leuven, Rega Institute, Clinical and Epidemiological Virology | KU Leuven, Rega Institute, Clinical and Epidemiological Virology | Tony Wawina-Bokalanga et al |
| EPI_ISL_464075 | KU Leuven, Rega Institute, Clinical and Epidemiological Virology | KU Leuven, Rega Institute, Clinical and Epidemiological Virology | Tony Wawina-Bokalanga et al |
| EPI_ISL_734637 | UZ Leuven, National Reference Laboratory for Coronaviruses, Laboratory Medicine, Leuven, Belgium | KU Leuven, Rega Institute, Clinical and Epidemiological Virology | Tony Wawina-Bokalanga et al |
| EPI_ISL_734645 | UZ Leuven, National Reference Laboratory for Coronaviruses, Laboratory Medicine, Leuven, Belgium | KU Leuven, Rega Institute, Clinical and Epidemiological Virology | Tony Wawina-Bokalanga et al |
| EPI_ISL_734687 | UZ Leuven, National Reference Laboratory for Coronaviruses, Laboratory Medicine, Leuven, Belgium | KU Leuven, Rega Institute, Clinical and Epidemiological Virology | Tony Wawina-Bokalanga et al |
| EPI_ISL_734720 | UZ Leuven, National Reference Laboratory for Coronaviruses, Laboratory Medicine, Leuven, Belgium | KU Leuven, Rega Institute, Clinical and Epidemiological Virology | Tony Wawina-Bokalanga et al |
| EPI_ISL_477007 | KU Leuven, Rega Institute, Clinical and Epidemiological Virology | KU Leuven, Rega Institute, Clinical and Epidemiological Virology | Tony Wawina-Bokalanga et al |
| EPI_ISL_734730 | UZ Leuven, National Reference Laboratory for Coronaviruses, Laboratory Medicine, Leuven, Belgium | KU Leuven, Rega Institute, Clinical and Epidemiological Virology | Tony Wawina-Bokalanga et al |
| EPI_ISL_734732 | UZ Leuven, National Reference Laboratory for Coronaviruses, Laboratory Medicine, Leuven, Belgium | KU Leuven, Rega Institute, Clinical and Epidemiological Virology | Tony Wawina-Bokalanga et al |
| EPI_ISL_734733 | UZ Leuven, National Reference Laboratory for Coronaviruses, Laboratory Medicine, Leuven, Belgium | KU Leuven, Rega Institute, Clinical and Epidemiological Virology | Tony Wawina-Bokalanga et al |
| EPI_ISL_734763 | UZ Leuven, National Reference Laboratory for Coronaviruses, Laboratory Medicine, Leuven, Belgium | KU Leuven, Rega Institute, Clinical and Epidemiological Virology | Tony Wawina-Bokalanga et al |

| | | | |
|---|---|---|---|
| EPI_ISL_734764 | UZ Leuven, National Reference Laboratory for Coronaviruses, Laboratory Medicine, Leuven, Belgium | KU Leuven, Rega Institute, Clinical and Epidemiological Virology | Tony Wawina-Bokalanga et al |
| EPI_ISL_734768 | UZ Leuven, National Reference Laboratory for Coronaviruses, Laboratory Medicine, Leuven, Belgium | KU Leuven, Rega Institute, Clinical and Epidemiological Virology | Tony Wawina-Bokalanga et al |
| EPI_ISL_734782 | UZ Leuven, National Reference Laboratory for Coronaviruses, Laboratory Medicine, Leuven, Belgium | KU Leuven, Rega Institute, Clinical and Epidemiological Virology | Tony Wawina-Bokalanga et al |
| EPI_ISL_522350 | KU Leuven, Rega Institute, Clinical and Epidemiological Virology | KU Leuven, Rega Institute, Clinical and Epidemiological Virology | Tony Wawina-Bokalanga et al |
| EPI_ISL_734825 | UZ Leuven, National Reference Laboratory for Coronaviruses, Laboratory Medicine, Leuven, Belgium | KU Leuven, Rega Institute, Clinical and Epidemiological Virology | Tony Wawina-Bokalanga et al |
| EPI_ISL_734865 | UZ Leuven, National Reference Laboratory for Coronaviruses, Laboratory Medicine, Leuven, Belgium | KU Leuven, Rega Institute, Clinical and Epidemiological Virology | Tony Wawina-Bokalanga et al |
| EPI_ISL_735016 | UZ Leuven, National Reference Laboratory for Coronaviruses, Laboratory Medicine, Leuven, Belgium | KU Leuven, Rega Institute, Clinical and Epidemiological Virology | Tony Wawina-Bokalanga et al |
| EPI_ISL_735085 | UZ Leuven, National Reference Laboratory for Coronaviruses, Laboratory Medicine, Leuven, Belgium | KU Leuven, Rega Institute, Clinical and Epidemiological Virology | Tony Wawina-Bokalanga et al |
| EPI_ISL_735087 | UZ Leuven, National Reference Laboratory for Coronaviruses, Laboratory Medicine, Leuven, Belgium | KU Leuven, Rega Institute, Clinical and Epidemiological Virology | Tony Wawina-Bokalanga et al |
| EPI_ISL_1322686 | KU Leuven, Rega Institute, Clinical and Epidemiological Virology | KU Leuven, Rega Institute, Clinical and Epidemiological Virology | Tony Wawina-Bokalanga et al |
| EPI_ISL_1322689 | KU Leuven, Rega Institute, Clinical and Epidemiological Virology | KU Leuven, Rega Institute, Clinical and Epidemiological Virology | Tony Wawina-Bokalanga et al |
| EPI_ISL_1380845 | KU Leuven, Rega Institute, Clinical and Epidemiological Virology | KU Leuven, Rega Institute, Clinical and Epidemiological Virology | Tony Wawina-Bokalanga et al |
| EPI_ISL_1382423 | KU Leuven, Rega Institute, Clinical and Epidemiological Virology | KU Leuven, Rega Institute, Clinical and Epidemiological Virology | Tony Wawina-Bokalanga et al |
| EPI_ISL_1382789 | KU Leuven, Rega Institute, Clinical and Epidemiological Virology | KU Leuven, Rega Institute, Clinical and Epidemiological Virology | Tony Wawina-Bokalanga et al |
| EPI_ISL_966940 | Technical Support Units for Scientific Research (UATRS), National Centre for Scientific and Technical Research (CNRST) | Technical Support Units for Scientific Research (UATRS), National Centre for Scientific and Technical Research (CNRST) | Touil et al |
| EPI_ISL_1205370 | Arizona State Public Health Laboratory | Arizona State Public Health Laboratory | Trung Huynh et al |
| EPI_ISL_1184565 | Arizona State Public Health Laboratory | Arizona State Public Health Laboratory | Trung Huynh et al |
| EPI_ISL_407084 | Department of Virology III, National Institute of Infectious Diseases | Pathogen Genomics Center, National Institute of Infectious Diseases | Tsuyoshi Sekizuka et al |
| EPI_ISL_416568 | Japanese Quarantine Stations | Pathogen Genomics Center, National Institute of Infectious Diseases | Tsuyoshi Sekizuka et al |
| EPI_ISL_416613 | Japanese Quarantine Stations | Pathogen Genomics Center, National Institute of Infectious Diseases | Tsuyoshi Sekizuka et al |
| EPI_ISL_416615 | Japanese Quarantine Stations | Pathogen Genomics Center, National Institute of Infectious Diseases | Tsuyoshi Sekizuka et al |
| EPI_ISL_591352 | Pathogen Genomics Center, National Institute of Infectious Diseases | Pathogen Genomics Center, National Institute of Infectious Diseases | Tsuyoshi Sekizuka et al |
| EPI_ISL_591372 | Pathogen Genomics Center, National Institute of Infectious Diseases | Pathogen Genomics Center, National Institute of Infectious Diseases | Tsuyoshi Sekizuka et al |
| EPI_ISL_591390 | Pathogen Genomics Center, National Institute of Infectious Diseases | Pathogen Genomics Center, National Institute of Infectious Diseases | Tsuyoshi Sekizuka et al |
| EPI_ISL_591402 | Pathogen Genomics Center, National Institute of Infectious Diseases | Pathogen Genomics Center, National Institute of Infectious Diseases | Tsuyoshi Sekizuka et al |
| EPI_ISL_591411 | Pathogen Genomics Center, National Institute of Infectious Diseases | Pathogen Genomics Center, National Institute of Infectious Diseases | Tsuyoshi Sekizuka et al |
| EPI_ISL_591449 | Pathogen Genomics Center, National Institute of Infectious Diseases | Pathogen Genomics Center, National Institute of Infectious Diseases | Tsuyoshi Sekizuka et al |

| | | | |
|---|---|---|---|
| EPI_ISL_667560 | Pathogen Genomics Center, National Institute of Infectious Diseases | Pathogen Genomics Center, National Institute of Infectious Diseases | Tsuyoshi Sekizuka et al |
| EPI_ISL_667626 | Pathogen Genomics Center, National Institute of Infectious Diseases | Pathogen Genomics Center, National Institute of Infectious Diseases | Tsuyoshi Sekizuka et al |
| EPI_ISL_667639 | Pathogen Genomics Center, National Institute of Infectious Diseases | Pathogen Genomics Center, National Institute of Infectious Diseases | Tsuyoshi Sekizuka et al |
| EPI_ISL_667649 | Pathogen Genomics Center, National Institute of Infectious Diseases | Pathogen Genomics Center, National Institute of Infectious Diseases | Tsuyoshi Sekizuka et al |
| EPI_ISL_667650 | Pathogen Genomics Center, National Institute of Infectious Diseases | Pathogen Genomics Center, National Institute of Infectious Diseases | Tsuyoshi Sekizuka et al |
| EPI_ISL_667663 | Pathogen Genomics Center, National Institute of Infectious Diseases | Pathogen Genomics Center, National Institute of Infectious Diseases | Tsuyoshi Sekizuka et al |
| EPI_ISL_667738 | Pathogen Genomics Center, National Institute of Infectious Diseases | Pathogen Genomics Center, National Institute of Infectious Diseases | Tsuyoshi Sekizuka et al |
| EPI_ISL_721601 | Pathogen Genomics Center, National Institute of Infectious Diseases | Pathogen Genomics Center, National Institute of Infectious Diseases | Tsuyoshi Sekizuka et al |
| EPI_ISL_768654 | Pathogen Genomics Center, National Institute of Infectious Diseases | Pathogen Genomics Center, National Institute of Infectious Diseases | Tsuyoshi Sekizuka et al |
| EPI_ISL_768709 | Pathogen Genomics Center, National Institute of Infectious Diseases | Pathogen Genomics Center, National Institute of Infectious Diseases | Tsuyoshi Sekizuka et al |
| EPI_ISL_779207 | Pathogen Genomics Center, National Institute of Infectious Diseases | Pathogen Genomics Center, National Institute of Infectious Diseases | Tsuyoshi Sekizuka et al |
| EPI_ISL_792680 | Pathogen Genomics Center, National Institute of Infectious Diseases | Pathogen Genomics Center, National Institute of Infectious Diseases | Tsuyoshi Sekizuka et al |
| EPI_ISL_792682 | Pathogen Genomics Center, National Institute of Infectious Diseases | Pathogen Genomics Center, National Institute of Infectious Diseases | Tsuyoshi Sekizuka et al |
| EPI_ISL_792683 | Pathogen Genomics Center, National Institute of Infectious Diseases | Pathogen Genomics Center, National Institute of Infectious Diseases | Tsuyoshi Sekizuka et al |
| EPI_ISL_779245 | Pathogen Genomics Center, National Institute of Infectious Diseases | Pathogen Genomics Center, National Institute of Infectious Diseases | Tsuyoshi Sekizuka et al |
| EPI_ISL_779246 | Pathogen Genomics Center, National Institute of Infectious Diseases | Pathogen Genomics Center, National Institute of Infectious Diseases | Tsuyoshi Sekizuka et al |
| EPI_ISL_779676 | Pathogen Genomics Center, National Institute of Infectious Diseases | Pathogen Genomics Center, National Institute of Infectious Diseases | Tsuyoshi Sekizuka et al |
| EPI_ISL_1425749 | SARS-CoV-2 testing team, National Institute of Infectious Diseases | Pathogen Genomics Center, National Institute of Infectious Diseases | Tsuyoshi Sekizuka et al |
| EPI_ISL_1425720 | SARS-CoV-2 testing team, National Institute of Infectious Diseases | Pathogen Genomics Center, National Institute of Infectious Diseases | Tsuyoshi Sekizuka et al |
| EPI_ISL_1425714 | SARS-CoV-2 testing team, National Institute of Infectious Diseases | Pathogen Genomics Center, National Institute of Infectious Diseases | Tsuyoshi Sekizuka et al |
| EPI_ISL_408669 | Dept. of Virology III, National Institute of Infectious Diseases | Pathogen Genomics Center, National Institute of Infectious Diseases | Tsuyoshi Sekizuka et al |
| EPI_ISL_410531 | Dept. of Pathology, National Institute of Infectious Diseases | Pathogen Genomics Center, National Institute of Infectious Diseases | Tsuyoshi Sekizuka et al |
| EPI_ISL_410532 | Dept. of Pathology, National Institute of Infectious Diseases | Pathogen Genomics Center, National Institute of Infectious Diseases | Tsuyoshi Sekizuka et al |
| EPI_ISL_479799 | Sapporo City Institute of Public Health | Pathogen Genomics Center, National Institute of Infectious Diseases | Tsuyoshi Sekizuka et al |
| EPI_ISL_479800 | Sapporo City Institute of Public Health | Pathogen Genomics Center, National Institute of Infectious Diseases | Tsuyoshi Sekizuka et al |
| EPI_ISL_684112 | Pathogen Genomics Center, National Institute of Infectious Diseases | Pathogen Genomics Center, National Institute of Infectious Diseases | Tsuyoshi Sekizuka et al |
| EPI_ISL_684127 | Gunma Prefectural Institute of Public Health and Environmental Sciences | Pathogen Genomics Center, National Institute of Infectious Diseases | Tsuyoshi Sekizuka et al |
| EPI_ISL_684147 | Sakai City Institute of Public Health | Pathogen Genomics Center, National Institute of Infectious Diseases | Tsuyoshi Sekizuka et al |
| EPI_ISL_480044 | Nagoya City Public Health Research Institute | Pathogen Genomics Center, National Institute of Infectious Diseases | Tsuyoshi Sekizuka et al |
| EPI_ISL_900909 | Pathogen Genomics Center, National Institute of Infectious Diseases | Pathogen Genomics Center, National Institute of Infectious Diseases | Tsuyoshi Sekizuka et al |

| | | | |
|---|---|---|---|
| EPI_ISL_901462 | Gunma Prefectural Institute of Public Health and Environmental Sciences | Pathogen Genomics Center, National Institute of Infectious Diseases | Tsuyoshi Sekizuka et al |
| EPI_ISL_901731 | Pathogen Genomics Center, National Institute of Infectious Diseases | Pathogen Genomics Center, National Institute of Infectious Diseases | Tsuyoshi Sekizuka et al |
| EPI_ISL_898049 | Sakai City Institute of Public Health | Pathogen Genomics Center, National Institute of Infectious Diseases | Tsuyoshi Sekizuka et al |
| EPI_ISL_896614 | Pathogen Genomics Center, National Institute of Infectious Diseases | Pathogen Genomics Center, National Institute of Infectious Diseases | Tsuyoshi Sekizuka et al |
| EPI_ISL_892373 | Pathogen Genomics Center, National Institute of Infectious Diseases | Pathogen Genomics Center, National Institute of Infectious Diseases | Tsuyoshi Sekizuka et al |
| EPI_ISL_892381 | Pathogen Genomics Center, National Institute of Infectious Diseases | Pathogen Genomics Center, National Institute of Infectious Diseases | Tsuyoshi Sekizuka et al |
| EPI_ISL_892382 | Pathogen Genomics Center, National Institute of Infectious Diseases | Pathogen Genomics Center, National Institute of Infectious Diseases | Tsuyoshi Sekizuka et al |
| EPI_ISL_892388 | Pathogen Genomics Center, National Institute of Infectious Diseases | Pathogen Genomics Center, National Institute of Infectious Diseases | Tsuyoshi Sekizuka et al |
| EPI_ISL_892533 | Osaka Institute of Public Health, Morinomiya Center | Pathogen Genomics Center, National Institute of Infectious Diseases | Tsuyoshi Sekizuka et al |
| EPI_ISL_892811 | Pathogen Genomics Center, National Institute of Infectious Diseases | Pathogen Genomics Center, National Institute of Infectious Diseases | Tsuyoshi Sekizuka et al |
| EPI_ISL_685238 | Pathogen Genomics Center, National Institute of Infectious Diseases | Pathogen Genomics Center, National Institute of Infectious Diseases | Tsuyoshi Sekizuka et al |
| EPI_ISL_1131143 | Osaka Institute of Public Health, Morinomiya Center | Pathogen Genomics Center, National Institute of Infectious Diseases | Tsuyoshi Sekizuka et al |
| EPI_ISL_1127231 | Pathogen Genomics Center, National Institute of Infectious Diseases | Pathogen Genomics Center, National Institute of Infectious Diseases | Tsuyoshi Sekizuka et al |
| EPI_ISL_1127124 | Pathogen Genomics Center, National Institute of Infectious Diseases | Pathogen Genomics Center, National Institute of Infectious Diseases | Tsuyoshi Sekizuka et al |
| EPI_ISL_1426592 | Sakai City Institute of Public Health | Pathogen Genomics Center, National Institute of Infectious Diseases | Tsuyoshi Sekizuka et al |
| EPI_ISL_1429772 | Pathogen Genomics Center, National Institute of Infectious Diseases | Pathogen Genomics Center, National Institute of Infectious Diseases | Tsuyoshi Sekizuka et al |
| EPI_ISL_1429779 | Pathogen Genomics Center, National Institute of Infectious Diseases | Pathogen Genomics Center, National Institute of Infectious Diseases | Tsuyoshi Sekizuka et al |
| EPI_ISL_1429787 | Pathogen Genomics Center, National Institute of Infectious Diseases | Pathogen Genomics Center, National Institute of Infectious Diseases | Tsuyoshi Sekizuka et al |
| EPI_ISL_1429788 | Pathogen Genomics Center, National Institute of Infectious Diseases | Pathogen Genomics Center, National Institute of Infectious Diseases | Tsuyoshi Sekizuka et al |
| EPI_ISL_1429790 | Pathogen Genomics Center, National Institute of Infectious Diseases | Pathogen Genomics Center, National Institute of Infectious Diseases | Tsuyoshi Sekizuka et al |
| EPI_ISL_1430687 | Saitama Prefectural Institute of Public Health | Pathogen Genomics Center, National Institute of Infectious Diseases | Tsuyoshi Sekizuka et al |
| EPI_ISL_1430688 | Saitama Prefectural Institute of Public Health | Pathogen Genomics Center, National Institute of Infectious Diseases | Tsuyoshi Sekizuka et al |
| EPI_ISL_1430690 | Saitama Prefectural Institute of Public Health | Pathogen Genomics Center, National Institute of Infectious Diseases | Tsuyoshi Sekizuka et al |
| EPI_ISL_1430697 | SARS-CoV-2 testing team, National Institute of Infectious Diseases | Pathogen Genomics Center, National Institute of Infectious Diseases | Tsuyoshi Sekizuka et al |
| EPI_ISL_690818 | Kanagawa Prefectural Institute of Public Health | Pathogen Genomics Center, National Institute of Infectious Diseases | Tsuyoshi Sekizuka et al |
| EPI_ISL_690824 | Kanagawa Prefectural Institute of Public Health | Pathogen Genomics Center, National Institute of Infectious Diseases | Tsuyoshi Sekizuka et al |
| EPI_ISL_1430698 | Saitama Prefectural Institute of Public Health | Pathogen Genomics Center, National Institute of Infectious Diseases | Tsuyoshi Sekizuka et al |
| EPI_ISL_690635 | Pathogen Genomics Center, National Institute of Infectious Diseases | Pathogen Genomics Center, National Institute of Infectious Diseases | Tsuyoshi Sekizuka et al |
| EPI_ISL_690267 | Pathogen Genomics Center, National Institute of Infectious Diseases | Pathogen Genomics Center, National Institute of Infectious Diseases | Tsuyoshi Sekizuka et al |
| EPI_ISL_690519 | Fukuoka Institute of Health and Environmental Sciences | Pathogen Genomics Center, National Institute of Infectious Diseases | Tsuyoshi Sekizuka et al |

| | | | |
|---|---|---|---|
| EPI_ISL_688923 | Fukuoka Institute of Health and Environmental Sciences | Pathogen Genomics Center, National Institute of Infectious Diseases | Tsuyoshi Sekizuka et al |
| EPI_ISL_689271 | Pathogen Genomics Center, National Institute of Infectious Diseases | Pathogen Genomics Center, National Institute of Infectious Diseases | Tsuyoshi Sekizuka et al |
| EPI_ISL_689294 | Pathogen Genomics Center, National Institute of Infectious Diseases | Pathogen Genomics Center, National Institute of Infectious Diseases | Tsuyoshi Sekizuka et al |
| EPI_ISL_686339 | Pathogen Genomics Center, National Institute of Infectious Diseases | Pathogen Genomics Center, National Institute of Infectious Diseases | Tsuyoshi Sekizuka et al |
| EPI_ISL_686346 | Pathogen Genomics Center, National Institute of Infectious Diseases | Pathogen Genomics Center, National Institute of Infectious Diseases | Tsuyoshi Sekizuka et al |
| EPI_ISL_686354 | Pathogen Genomics Center, National Institute of Infectious Diseases | Pathogen Genomics Center, National Institute of Infectious Diseases | Tsuyoshi Sekizuka et al |
| EPI_ISL_902344 | Toyama Institute of Health | Pathogen Genomics Center, National Institute of Infectious Diseases | Tsuyoshi Sekizuka et al |
| EPI_ISL_408665 | Dept. of Virology III, National Institute of Infectious Diseases | Pathogen Genomics Center, National Institute of Infectious Diseases | Tsuyoshi Sekizuka et al |
| EPI_ISL_408666 | Dept. of Virology III, National Institute of Infectious Diseases | Pathogen Genomics Center, National Institute of Infectious Diseases | Tsuyoshi Sekizuka et al |
| EPI_ISL_408667 | Dept. of Virology III, National Institute of Infectious Diseases | Pathogen Genomics Center, National Institute of Infectious Diseases | Tsuyoshi Sekizuka et al |
| EPI_ISL_491298 | Rural Health Unit - Calauan, Laguna | Research Institute for Tropical Medicine | Tujan et al |
| EPI_ISL_1122419 | National Public Health Laboratory, National Centre for Infectious Diseases | National Virology Reference Laboratory | Tze Minn Mak et al |
| EPI_ISL_1122420 | National Public Health Laboratory, National Centre for Infectious Diseases | National Virology Reference Laboratory | Tze Minn Mak et al |
| EPI_ISL_1122421 | National Public Health Laboratory, National Centre for Infectious Diseases | National Virology Reference Laboratory | Tze Minn Mak et al |
| EPI_ISL_693471 | Central Public Health Laboratory | National Public Health Laboratory, National Centre for Infectious Diseases | Tze Minn Mak et al |
| EPI_ISL_693480 | Central Public Health Laboratory | National Public Health Laboratory, National Centre for Infectious Diseases | Tze Minn Mak et al |
| EPI_ISL_693481 | Central Public Health Laboratory | National Public Health Laboratory, National Centre for Infectious Diseases | Tze Minn Mak et al |
| EPI_ISL_693482 | Central Public Health Laboratory | National Public Health Laboratory, National Centre for Infectious Diseases | Tze Minn Mak et al |
| EPI_ISL_1322329 | Central Public Health Laboratory | National Public Health Laboratory, National Centre for Infectious Diseases | Tze Minn Mak et al |
| EPI_ISL_693483 | Central Public Health Laboratory | National Public Health Laboratory, National Centre for Infectious Diseases | Tze Minn Mak et al |
| EPI_ISL_1322330 | Central Public Health Laboratory | National Public Health Laboratory, National Centre for Infectious Diseases | Tze Minn Mak et al |
| EPI_ISL_693472 | Central Public Health Laboratory | National Public Health Laboratory, National Centre for Infectious Diseases | Tze Minn Mak et al |
| EPI_ISL_693473 | Central Public Health Laboratory | National Public Health Laboratory, National Centre for Infectious Diseases | Tze Minn Mak et al |
| EPI_ISL_1307659 | Central Public Health Laboratory | National Public Health Laboratory, National Centre for Infectious Diseases | Tze Minn Mak et al |
| EPI_ISL_1307660 | Central Public Health Laboratory | National Public Health Laboratory, National Centre for Infectious Diseases | Tze Minn Mak et al |
| EPI_ISL_693474 | Central Public Health Laboratory | National Public Health Laboratory, National Centre for Infectious Diseases | Tze Minn Mak et al |
| EPI_ISL_693475 | Central Public Health Laboratory | National Public Health Laboratory, National Centre for Infectious Diseases | Tze Minn Mak et al |
| EPI_ISL_693476 | Central Public Health Laboratory | National Public Health Laboratory, National Centre for Infectious Diseases | Tze Minn Mak et al |
| EPI_ISL_693477 | Central Public Health Laboratory | National Public Health Laboratory, National Centre for Infectious Diseases | Tze Minn Mak et al |
| EPI_ISL_1307677 | Central Public Health Laboratory | National Public Health Laboratory, National Centre for Infectious Diseases | Tze Minn Mak et al |

| | | | |
|---|---|---|---|
| EPI_ISL_693478 | Central Public Health Laboratory | National Public Health Laboratory, National Centre for Infectious Diseases | Tze Minn Mak et al |
| EPI_ISL_693479 | Central Public Health Laboratory | National Public Health Laboratory, National Centre for Infectious Diseases | Tze Minn Mak et al |
| EPI_ISL_574531 | National Public Health Laboratory, National Centre for Infectious Diseases | National Public Health Laboratory, National Centre for Infectious Diseases | Tze Minn Mak et al |
| EPI_ISL_891244 | National Public Health Laboratory, National Centre for Infectious Diseases | National Public Health Laboratory, National Centre for Infectious Diseases | Tze Minn Mak et al |
| EPI_ISL_574541 | National Public Health Laboratory, National Centre for Infectious Diseases | National Public Health Laboratory, National Centre for Infectious Diseases | Tze Minn Mak et al |
| EPI_ISL_596478 | National Public Health Laboratory, National Centre for Infectious Diseases | National Public Health Laboratory, National Centre for Infectious Diseases | Tze Minn Mak et al |
| EPI_ISL_596483 | National Public Health Laboratory, National Centre for Infectious Diseases | National Public Health Laboratory, National Centre for Infectious Diseases | Tze Minn Mak et al |
| EPI_ISL_596488 | National Public Health Laboratory, National Centre for Infectious Diseases | National Public Health Laboratory, National Centre for Infectious Diseases | Tze Minn Mak et al |
| EPI_ISL_626646 | National Public Health Laboratory, National Centre for Infectious Diseases | National Public Health Laboratory, National Centre for Infectious Diseases | Tze Minn Mak et al |
| EPI_ISL_906840 | National Public Health Laboratory, National Centre for Infectious Diseases | National Public Health Laboratory, National Centre for Infectious Diseases | Tze Minn Mak et al |
| EPI_ISL_937515 | National Public Health Laboratory, National Centre for Infectious Diseases | National Public Health Laboratory, National Centre for Infectious Diseases | Tze Minn Mak et al |
| EPI_ISL_937518 | National Public Health Laboratory, National Centre for Infectious Diseases | National Public Health Laboratory, National Centre for Infectious Diseases | Tze Minn Mak et al |
| EPI_ISL_937528 | National Public Health Laboratory, National Centre for Infectious Diseases | National Public Health Laboratory, National Centre for Infectious Diseases | Tze Minn Mak et al |
| EPI_ISL_645115 | National Public Health Laboratory, National Centre for Infectious Diseases | National Public Health Laboratory, National Centre for Infectious Diseases | Tze Minn Mak et al |
| EPI_ISL_645117 | National Public Health Laboratory, National Centre for Infectious Diseases | National Public Health Laboratory, National Centre for Infectious Diseases | Tze Minn Mak et al |
| EPI_ISL_654813 | National Public Health Laboratory, National Centre for Infectious Diseases | National Public Health Laboratory, National Centre for Infectious Diseases | Tze Minn Mak et al |
| EPI_ISL_693308 | National Public Health Laboratory, National Centre for Infectious Diseases | National Public Health Laboratory, National Centre for Infectious Diseases | Tze Minn Mak et al |
| EPI_ISL_693314 | National Public Health Laboratory, National Centre for Infectious Diseases | National Public Health Laboratory, National Centre for Infectious Diseases | Tze Minn Mak et al |
| EPI_ISL_693318 | National Public Health Laboratory, National Centre for Infectious Diseases | National Public Health Laboratory, National Centre for Infectious Diseases | Tze Minn Mak et al |
| EPI_ISL_953388 | National Public Health Laboratory, National Centre for Infectious Diseases | National Public Health Laboratory, National Centre for Infectious Diseases | Tze Minn Mak et al |
| EPI_ISL_728187 | National Public Health Laboratory, National Centre for Infectious Diseases | National Public Health Laboratory, National Centre for Infectious Diseases | Tze Minn Mak et al |
| EPI_ISL_754079 | National Public Health Laboratory, National Centre for Infectious Diseases | National Public Health Laboratory, National Centre for Infectious Diseases | Tze Minn Mak et al |
| EPI_ISL_981027 | National Public Health Laboratory, National Centre for Infectious Diseases | National Public Health Laboratory, National Centre for Infectious Diseases | Tze Minn Mak et al |
| EPI_ISL_1034270 | National Public Health Laboratory, National Centre for Infectious Diseases | National Public Health Laboratory, National Centre for Infectious Diseases | Tze Minn Mak et al |
| EPI_ISL_1164354 | National Public Health Laboratory, National Centre for Infectious Diseases | National Public Health Laboratory, National Centre for Infectious Diseases | Tze Minn Mak et al |
| EPI_ISL_1367558 | National Public Health Laboratory, National Centre for Infectious Diseases | National Public Health Laboratory, National Centre for Infectious Diseases | Tze Minn Mak et al |
| EPI_ISL_1524783 | National Public Health Laboratory, National Centre for Infectious Diseases | National Public Health Laboratory, National Centre for Infectious Diseases | Tze Minn Mak et al |
| EPI_ISL_1524796 | National Public Health Laboratory, National Centre for Infectious Diseases | National Public Health Laboratory, National Centre for Infectious Diseases | Tze Minn Mak et al |
| EPI_ISL_1519355 | National Public Health Laboratory, National Centre for Infectious Diseases | National Public Health Laboratory, National Centre for Infectious Diseases | Tze Minn Mak et al |
| EPI_ISL_1543939 | National Public Health Laboratory, National Centre for Infectious Diseases | National Public Health Laboratory, National Centre for Infectious Diseases | Tze Minn Mak et al |

| | | | |
|---|---|---|---|
| EPI_ISL_833376 | National Public Health Laboratory, National Centre for Infectious Diseases | National Public Health Laboratory, National Centre for Infectious Diseases | Tze Minn Mak et al |
| EPI_ISL_1620144 | National Public Health Laboratory, National Centre for Infectious Diseases | National Public Health Laboratory, National Centre for Infectious Diseases | Tze Minn Mak et al |
| EPI_ISL_1620131 | National Public Health Laboratory, National Centre for Infectious Diseases | National Public Health Laboratory, National Centre for Infectious Diseases | Tze Minn Mak et al |
| EPI_ISL_1652091 | National Public Health Laboratory, National Centre for Infectious Diseases | National Public Health Laboratory, National Centre for Infectious Diseases | Tze Minn Mak et al |
| EPI_ISL_833384 | National Public Health Laboratory, National Centre for Infectious Diseases | National Public Health Laboratory, National Centre for Infectious Diseases | Tze Minn Mak et al |
| EPI_ISL_1652117 | National Public Health Laboratory, National Centre for Infectious Diseases | National Public Health Laboratory, National Centre for Infectious Diseases | Tze Minn Mak et al |
| EPI_ISL_1652120 | National Public Health Laboratory, National Centre for Infectious Diseases | National Public Health Laboratory, National Centre for Infectious Diseases | Tze Minn Mak et al |
| EPI_ISL_1652123 | National Public Health Laboratory, National Centre for Infectious Diseases | National Public Health Laboratory, National Centre for Infectious Diseases | Tze Minn Mak et al |
| EPI_ISL_891236 | National Public Health Laboratory, National Centre for Infectious Diseases | National Public Health Laboratory, National Centre for Infectious Diseases | Tze Minn Mak et al |
| EPI_ISL_1271944 | Universidade Federal do Norte do Tocantins (UFNT) | Laboratório de Bioinformática e Biotecnologia (Labinftec/UFT) | Ueric José Borges de Souza et al |
| EPI_ISL_1272074 | Universidade Federal do Norte do Tocantins (UFNT) | Laboratório de Bioinformática e Biotecnologia (Labinftec/UFT) | Ueric José Borges de Souza et al |
| EPI_ISL_1272076 | Universidade Federal do Norte do Tocantins (UFNT) | Laboratório de Bioinformática e Biotecnologia (Labinftec/UFT) | Ueric José Borges de Souza et al |
| EPI_ISL_1272188 | Universidade Federal do Norte do Tocantins (UFNT) | Laboratório de Bioinformática e Biotecnologia (Labinftec/UFT) | Ueric José Borges de Souza et al |
| EPI_ISL_1272236 | Universidade Federal do Norte do Tocantins (UFNT) | Laboratório de Bioinformática e Biotecnologia (Labinftec/UFT) | Ueric José Borges de Souza et al |
| EPI_ISL_516934 | Nicolae Testemitanu State University of Medicine and Pharmacy | International Centre for Genetic Engineering and Biotechnology (ICGEB) and ARGO Open Lab Platform for Genome Sequencing | Ulinici M et al |
| EPI_ISL_516938 | Nicolae Testemitanu State University of Medicine and Pharmacy | International Centre for Genetic Engineering and Biotechnology (ICGEB) and ARGO Open Lab Platform for Genome Sequencing | Ulinici M et al |
| EPI_ISL_516936 | Nicolae Testemitanu State University of Medicine and Pharmacy | International Centre for Genetic Engineering and Biotechnology (ICGEB) and ARGO Open Lab Platform for Genome Sequencing | Ulinici M et al |
| EPI_ISL_1373189 | Protzer Lab | Protzer Lab, Institut für Medizinische Mikrobiologie und Hygiene, Gagneur Lab | Ulrike Protzer et al |
| EPI_ISL_1373184 | Protzer Lab | Protzer Lab, Institut für Medizinische Mikrobiologie und Hygiene, Gagneur Lab | Ulrike Protzer et al |
| EPI_ISL_1373167 | Protzer Lab | Protzer Lab, Institut für Medizinische Mikrobiologie und Hygiene, Gagneur Lab | Ulrike Protzer et al |
| EPI_ISL_1373258 | Protzer Lab | Protzer Lab, Institut für Medizinische Mikrobiologie und Hygiene, Gagneur Lab | Ulrike Protzer et al |
| EPI_ISL_1373269 | Protzer Lab | Protzer Lab, Institut für Medizinische Mikrobiologie und Hygiene, Gagneur Lab | Ulrike Protzer et al |
| EPI_ISL_1373163 | Protzer Lab | Protzer Lab, Institut für Medizinische Mikrobiologie und Hygiene, Gagneur Lab | Ulrike Protzer et al |
| EPI_ISL_1373186 | Protzer Lab | Protzer Lab, Institut für Medizinische Mikrobiologie und Hygiene, Gagneur Lab | Ulrike Protzer et al |
| EPI_ISL_1373276 | Protzer Lab | Protzer Lab, Institut für Medizinische Mikrobiologie und Hygiene, Gagneur Lab | Ulrike Protzer et al |
| EPI_ISL_1373281 | Protzer Lab | Protzer Lab, Institut für Medizinische Mikrobiologie und Hygiene, Gagneur Lab | Ulrike Protzer et al |

| | | | |
|---|---|---|---|
| EPI_ISL_1373222 | Protzer Lab | Protzer Lab, Institut für Medizinische Mikrobiologie und Hygiene, Gagneur Lab | Ulrike Protzer et al |
| EPI_ISL_1373214 | Protzer Lab | Protzer Lab, Institut für Medizinische Mikrobiologie und Hygiene, Gagneur Lab | Ulrike Protzer et al |
| EPI_ISL_1373294 | Protzer Lab | Protzer Lab, Institut für Medizinische Mikrobiologie und Hygiene, Gagneur Lab | Ulrike Protzer et al |
| EPI_ISL_1373303 | Protzer Lab | Protzer Lab, Institut für Medizinische Mikrobiologie und Hygiene, Gagneur Lab | Ulrike Protzer et al |
| EPI_ISL_1373229 | Protzer Lab | Protzer Lab, Institut für Medizinische Mikrobiologie und Hygiene, Gagneur Lab | Ulrike Protzer et al |
| EPI_ISL_1373188 | Protzer Lab | Protzer Lab, Institut für Medizinische Mikrobiologie und Hygiene, Gagneur Lab | Ulrike Protzer et al |
| EPI_ISL_1373177 | Protzer Lab | Protzer Lab, Institut für Medizinische Mikrobiologie und Hygiene, Gagneur Lab | Ulrike Protzer et al |
| EPI_ISL_1373180 | Protzer Lab | Protzer Lab, Institut für Medizinische Mikrobiologie und Hygiene, Gagneur Lab | Ulrike Protzer et al |
| EPI_ISL_466680 | Nebraska Public Health Laboratory | UNMC COVID-19 Response Team | UNMC COVID-19 Response Team et al |
| EPI_ISL_959275 | National Influenza Center, Virology Department | National Influenza Center | V Salimi et al |
| EPI_ISL_1014676 | National Influenza Center, Virology Department | National Influenza Center | V Salimi et al |
| EPI_ISL_862075 | National Influenza Center, Virology Department | National Influenza Center | V Salimi et al |
| EPI_ISL_1239127 | Laboratório Central de Saúde Pública do Espírito Santo | Coordenação Geral de Laboratórios de Saúde Pública (CGLAB) | Vagner Fonseca et al |
| EPI_ISL_1239136 | Laboratório Central de Saúde Pública do Espírito Santo | Coordenação Geral de Laboratórios de Saúde Pública (CGLAB) | Vagner Fonseca et al |
| EPI_ISL_1239117 | Laboratório Central de Saúde Pública do Espírito Santo | Coordenação Geral de Laboratórios de Saúde Pública (CGLAB) | Vagner Fonseca et al |
| EPI_ISL_1239125 | Laboratório Central de Saúde Pública do Espírito Santo | Coordenação Geral de Laboratórios de Saúde Pública (CGLAB) | Vagner Fonseca et al |
| EPI_ISL_1239135 | Laboratório Central de Saúde Pública do Espírito Santo | Coordenação Geral de Laboratórios de Saúde Pública (CGLAB) | Vagner Fonseca et al |
| EPI_ISL_1239131 | Laboratório Central de Saúde Pública do Espírito Santo | Coordenação Geral de Laboratórios de Saúde Pública (CGLAB) | Vagner Fonseca et al |
| EPI_ISL_1239126 | Laboratório Central de Saúde Pública do Espírito Santo | Coordenação Geral de Laboratórios de Saúde Pública (CGLAB) | Vagner Fonseca et al |
| EPI_ISL_1239115 | Laboratório Central de Saúde Pública do Espírito Santo | Coordenação Geral de Laboratórios de Saúde Pública (CGLAB) | Vagner Fonseca et al |
| EPI_ISL_1239133 | Laboratório Central de Saúde Pública do Espírito Santo | Coordenação Geral de Laboratórios de Saúde Pública (CGLAB) | Vagner Fonseca et al |
| EPI_ISL_1239118 | Laboratório Central de Saúde Pública do Espírito Santo | Coordenação Geral de Laboratórios de Saúde Pública (CGLAB) | Vagner Fonseca et al |
| EPI_ISL_1239128 | Laboratório Central de Saúde Pública do Espírito Santo | Coordenação Geral de Laboratórios de Saúde Pública (CGLAB) | Vagner Fonseca et al |
| EPI_ISL_1239134 | Laboratório Central de Saúde Pública do Espírito Santo | Coordenação Geral de Laboratórios de Saúde Pública (CGLAB) | Vagner Fonseca et al |
| EPI_ISL_1239120 | Laboratório Central de Saúde Pública do Espírito Santo | Coordenação Geral de Laboratórios de Saúde Pública (CGLAB) | Vagner Fonseca et al |
| EPI_ISL_1239123 | Laboratório Central de Saúde Pública do Espírito Santo | Coordenação Geral de Laboratórios de Saúde Pública (CGLAB) | Vagner Fonseca et al |
| EPI_ISL_1239132 | Laboratório Central de Saúde Pública do Espírito Santo | Coordenação Geral de Laboratórios de Saúde Pública (CGLAB) | Vagner Fonseca et al |

| | | | |
|---|---|---|---|
| EPI_ISL_1239114 | Laboratório Central de Saúde Pública do Espírito Santo | Coordenação Geral de Laboratórios de Saúde Pública (CGLAB) | Vagner Fonseca et al |
| EPI_ISL_1239122 | Laboratório Central de Saúde Pública do Espírito Santo | Coordenação Geral de Laboratórios de Saúde Pública (CGLAB) | Vagner Fonseca et al |
| EPI_ISL_1239116 | Laboratório Central de Saúde Pública do Espírito Santo | Coordenação Geral de Laboratórios de Saúde Pública (CGLAB) | Vagner Fonseca et al |
| EPI_ISL_1239121 | Laboratório Central de Saúde Pública do Espírito Santo | Coordenação Geral de Laboratórios de Saúde Pública (CGLAB) | Vagner Fonseca et al |
| EPI_ISL_1239129 | Laboratório Central de Saúde Pública do Espírito Santo | Coordenação Geral de Laboratórios de Saúde Pública (CGLAB) | Vagner Fonseca et al |
| EPI_ISL_1239130 | Laboratório Central de Saúde Pública do Espírito Santo | Coordenação Geral de Laboratórios de Saúde Pública (CGLAB) | Vagner Fonseca et al |
| EPI_ISL_1182574 | Fundação Ezequiel Dias (FUNED) | Coordenação Geral de Laboratórios de Saúde Pública (CGLAB/DAEVS/SVS/MS) | Vagner Fonseca et al |
| EPI_ISL_1182573 | Fundação Ezequiel Dias (FUNED) | Coordenação Geral de Laboratórios de Saúde Pública (CGLAB/DAEVS/SVS/MS) | Vagner Fonseca et al |
| EPI_ISL_1182555 | Fundação Ezequiel Dias (FUNED) | Coordenação Geral de Laboratórios de Saúde Pública (CGLAB/DAEVS/SVS/MS) | Vagner Fonseca et al |
| EPI_ISL_1182548 | Fundação Ezequiel Dias (FUNED) | Coordenação Geral de Laboratórios de Saúde Pública (CGLAB/DAEVS/SVS/MS) | Vagner Fonseca et al |
| EPI_ISL_1182544 | Fundação Ezequiel Dias (FUNED) | Coordenação Geral de Laboratórios de Saúde Pública (CGLAB/DAEVS/SVS/MS) | Vagner Fonseca et al |
| EPI_ISL_1182550 | Fundação Ezequiel Dias (FUNED) | Coordenação Geral de Laboratórios de Saúde Pública (CGLAB/DAEVS/SVS/MS) | Vagner Fonseca et al |
| EPI_ISL_1182579 | Fundação Ezequiel Dias (FUNED) | Coordenação Geral de Laboratórios de Saúde Pública (CGLAB/DAEVS/SVS/MS) | Vagner Fonseca et al |
| EPI_ISL_1182569 | Fundação Ezequiel Dias (FUNED) | Coordenação Geral de Laboratórios de Saúde Pública (CGLAB/DAEVS/SVS/MS) | Vagner Fonseca et al |
| EPI_ISL_1182561 | Fundação Ezequiel Dias (FUNED) | Coordenação Geral de Laboratórios de Saúde Pública (CGLAB/DAEVS/SVS/MS) | Vagner Fonseca et al |
| EPI_ISL_1182560 | Fundação Ezequiel Dias (FUNED) | Coordenação Geral de Laboratórios de Saúde Pública (CGLAB/DAEVS/SVS/MS) | Vagner Fonseca et al |
| EPI_ISL_1182566 | Fundação Ezequiel Dias (FUNED) | Coordenação Geral de Laboratórios de Saúde Pública (CGLAB/DAEVS/SVS/MS) | Vagner Fonseca et al |
| EPI_ISL_1182564 | Fundação Ezequiel Dias (FUNED) | Coordenação Geral de Laboratórios de Saúde Pública (CGLAB/DAEVS/SVS/MS) | Vagner Fonseca et al |
| EPI_ISL_1182551 | Fundação Ezequiel Dias (FUNED) | Coordenação Geral de Laboratórios de Saúde Pública (CGLAB/DAEVS/SVS/MS) | Vagner Fonseca et al |
| EPI_ISL_1182559 | Fundação Ezequiel Dias (FUNED) | Coordenação Geral de Laboratórios de Saúde Pública (CGLAB/DAEVS/SVS/MS) | Vagner Fonseca et al |
| EPI_ISL_1182541 | Fundação Ezequiel Dias (FUNED) | Coordenação Geral de Laboratórios de Saúde Pública (CGLAB/DAEVS/SVS/MS) | Vagner Fonseca et al |
| EPI_ISL_1239124 | Fundação Ezequiel Dias | Coordenação Geral de Laboratórios de Saúde Pública (CGLAB) | Vagner Fonseca et al |
| EPI_ISL_1239139 | Fundação Ezequiel Dias | Coordenação Geral de Laboratórios de Saúde Pública (CGLAB) | Vagner Fonseca et al |
| EPI_ISL_1182625 | Fundação Ezequiel Dias (FUNED) | Coordenação Geral de Laboratórios de Saúde Pública (CGLAB/DAEVS/SVS/MS) | Vagner Fonseca et al |

| | | | |
|---|---|---|---|
| EPI_ISL_1182618 | Fundação Ezequiel Dias (FUNED) | Coordenação Geral de Laboratórios de Saúde Pública (CGLAB/DAEVS/SVS/MS) | Vagner Fonseca et al |
| EPI_ISL_1182615 | Fundação Ezequiel Dias (FUNED) | Coordenação Geral de Laboratórios de Saúde Pública (CGLAB/DAEVS/SVS/MS) | Vagner Fonseca et al |
| EPI_ISL_1182612 | Fundação Ezequiel Dias (FUNED) | Coordenação Geral de Laboratórios de Saúde Pública (CGLAB/DAEVS/SVS/MS) | Vagner Fonseca et al |
| EPI_ISL_1182611 | Fundação Ezequiel Dias (FUNED) | Coordenação Geral de Laboratórios de Saúde Pública (CGLAB/DAEVS/SVS/MS) | Vagner Fonseca et al |
| EPI_ISL_1182609 | Fundação Ezequiel Dias (FUNED) | Coordenação Geral de Laboratórios de Saúde Pública (CGLAB/DAEVS/SVS/MS) | Vagner Fonseca et al |
| EPI_ISL_1182607 | Fundação Ezequiel Dias (FUNED) | Coordenação Geral de Laboratórios de Saúde Pública (CGLAB/DAEVS/SVS/MS) | Vagner Fonseca et al |
| EPI_ISL_1182598 | Fundação Ezequiel Dias (FUNED) | Coordenação Geral de Laboratórios de Saúde Pública (CGLAB/DAEVS/SVS/MS) | Vagner Fonseca et al |
| EPI_ISL_1182600 | Fundação Ezequiel Dias (FUNED) | Coordenação Geral de Laboratórios de Saúde Pública (CGLAB/DAEVS/SVS/MS) | Vagner Fonseca et al |
| EPI_ISL_1182601 | Fundação Ezequiel Dias (FUNED) | Coordenação Geral de Laboratórios de Saúde Pública (CGLAB/DAEVS/SVS/MS) | Vagner Fonseca et al |
| EPI_ISL_1182593 | Fundação Ezequiel Dias (FUNED) | Coordenação Geral de Laboratórios de Saúde Pública (CGLAB/DAEVS/SVS/MS) | Vagner Fonseca et al |
| EPI_ISL_1182591 | Fundação Ezequiel Dias (FUNED) | Coordenação Geral de Laboratórios de Saúde Pública (CGLAB/DAEVS/SVS/MS) | Vagner Fonseca et al |
| EPI_ISL_1182570 | Fundação Ezequiel Dias (FUNED) | Coordenação Geral de Laboratórios de Saúde Pública (CGLAB/DAEVS/SVS/MS) | Vagner Fonseca et al |
| EPI_ISL_1182587 | Fundação Ezequiel Dias (FUNED) | Coordenação Geral de Laboratórios de Saúde Pública (CGLAB/DAEVS/SVS/MS) | Vagner Fonseca et al |
| EPI_ISL_1182578 | Fundação Ezequiel Dias (FUNED) | Coordenação Geral de Laboratórios de Saúde Pública (CGLAB/DAEVS/SVS/MS) | Vagner Fonseca et al |
| EPI_ISL_1182586 | Fundação Ezequiel Dias (FUNED) | Coordenação Geral de Laboratórios de Saúde Pública (CGLAB/DAEVS/SVS/MS) | Vagner Fonseca et al |
| EPI_ISL_1182577 | Fundação Ezequiel Dias (FUNED) | Coordenação Geral de Laboratórios de Saúde Pública (CGLAB/DAEVS/SVS/MS) | Vagner Fonseca et al |
| EPI_ISL_1182571 | Laboratório Central do Estado do Paraná | Coordenação Geral de Laboratórios de Saúde Pública (CGLAB/DAEVS/SVS/MS) | Vagner Fonseca et al |
| EPI_ISL_1182575 | Laboratório Central do Estado do Paraná | Coordenação Geral de Laboratórios de Saúde Pública (CGLAB/DAEVS/SVS/MS) | Vagner Fonseca et al |
| EPI_ISL_1182568 | Laboratório Central do Estado do Paraná | Coordenação Geral de Laboratórios de Saúde Pública (CGLAB/DAEVS/SVS/MS) | Vagner Fonseca et al |
| EPI_ISL_1182567 | Laboratório Central do Estado do Paraná | Coordenação Geral de Laboratórios de Saúde Pública (CGLAB/DAEVS/SVS/MS) | Vagner Fonseca et al |
| EPI_ISL_1182565 | Laboratório Central do Estado do Paraná | Coordenação Geral de Laboratórios de Saúde Pública (CGLAB/DAEVS/SVS/MS) | Vagner Fonseca et al |
| EPI_ISL_1182546 | Laboratório Central do Estado do Paraná | Coordenação Geral de Laboratórios de Saúde Pública (CGLAB/DAEVS/SVS/MS) | Vagner Fonseca et al |

| | | | |
|---|---|---|---|
| EPI_ISL_1182562 | Laboratório Central do Estado do Paraná | Coordenação Geral de Laboratórios de Saúde Pública (CGLAB/DAEVS/SVS/MS) | Vagner Fonseca et al |
| EPI_ISL_1182563 | Laboratório Central do Estado do Paraná | Coordenação Geral de Laboratórios de Saúde Pública (CGLAB/DAEVS/SVS/MS) | Vagner Fonseca et al |
| EPI_ISL_1182542 | Laboratório Central do Estado do Paraná | Coordenação Geral de Laboratórios de Saúde Pública (CGLAB/DAEVS/SVS/MS) | Vagner Fonseca et al |
| EPI_ISL_1182624 | Laboratório Central do Estado do Paraná | Coordenação Geral de Laboratórios de Saúde Pública (CGLAB/DAEVS/SVS/MS) | Vagner Fonseca et al |
| EPI_ISL_1182622 | Laboratório Central do Estado do Paraná | Coordenação Geral de Laboratórios de Saúde Pública (CGLAB/DAEVS/SVS/MS) | Vagner Fonseca et al |
| EPI_ISL_1182617 | Laboratório Central do Estado do Paraná | Coordenação Geral de Laboratórios de Saúde Pública (CGLAB/DAEVS/SVS/MS) | Vagner Fonseca et al |
| EPI_ISL_1182613 | Laboratório Central do Estado do Paraná | Coordenação Geral de Laboratórios de Saúde Pública (CGLAB/DAEVS/SVS/MS) | Vagner Fonseca et al |
| EPI_ISL_1182614 | Laboratório Central do Estado do Paraná | Coordenação Geral de Laboratórios de Saúde Pública (CGLAB/DAEVS/SVS/MS) | Vagner Fonseca et al |
| EPI_ISL_1182603 | Laboratório Central do Estado do Paraná | Coordenação Geral de Laboratórios de Saúde Pública (CGLAB/DAEVS/SVS/MS) | Vagner Fonseca et al |
| EPI_ISL_1182604 | Laboratório Central do Estado do Paraná | Coordenação Geral de Laboratórios de Saúde Pública (CGLAB/DAEVS/SVS/MS) | Vagner Fonseca et al |
| EPI_ISL_1182596 | Laboratório Central do Estado do Paraná | Coordenação Geral de Laboratórios de Saúde Pública (CGLAB/DAEVS/SVS/MS) | Vagner Fonseca et al |
| EPI_ISL_1182594 | Laboratório Central do Estado do Paraná | Coordenação Geral de Laboratórios de Saúde Pública (CGLAB/DAEVS/SVS/MS) | Vagner Fonseca et al |
| EPI_ISL_1182595 | Laboratório Central do Estado do Paraná | Coordenação Geral de Laboratórios de Saúde Pública (CGLAB/DAEVS/SVS/MS) | Vagner Fonseca et al |
| EPI_ISL_1182592 | Laboratório Central do Estado do Paraná | Coordenação Geral de Laboratórios de Saúde Pública (CGLAB/DAEVS/SVS/MS) | Vagner Fonseca et al |
| EPI_ISL_1182589 | Laboratório Central do Estado do Paraná | Coordenação Geral de Laboratórios de Saúde Pública (CGLAB/DAEVS/SVS/MS) | Vagner Fonseca et al |
| EPI_ISL_1182588 | Laboratório Central do Estado do Paraná | Coordenação Geral de Laboratórios de Saúde Pública (CGLAB/DAEVS/SVS/MS) | Vagner Fonseca et al |
| EPI_ISL_1182584 | Laboratório Central do Estado do Paraná | Coordenação Geral de Laboratórios de Saúde Pública (CGLAB/DAEVS/SVS/MS) | Vagner Fonseca et al |
| EPI_ISL_1182583 | Laboratório Central do Estado do Paraná | Coordenação Geral de Laboratórios de Saúde Pública (CGLAB/DAEVS/SVS/MS) | Vagner Fonseca et al |
| EPI_ISL_1182582 | Laboratório Central do Estado do Paraná | Coordenação Geral de Laboratórios de Saúde Pública (CGLAB/DAEVS/SVS/MS) | Vagner Fonseca et al |
| EPI_ISL_1182580 | Laboratório Central do Estado do Paraná | Coordenação Geral de Laboratórios de Saúde Pública (CGLAB/DAEVS/SVS/MS) | Vagner Fonseca et al |
| EPI_ISL_1182581 | Laboratório Central do Estado do Paraná | Coordenação Geral de Laboratórios de Saúde Pública (CGLAB/DAEVS/SVS/MS) | Vagner Fonseca et al |
| EPI_ISL_1182576 | Laboratório Central do Estado do Paraná | Coordenação Geral de Laboratórios de Saúde Pública (CGLAB/DAEVS/SVS/MS) | Vagner Fonseca et al |

| | | | |
|---|---|---|---|
| EPI_ISL_1182572 | Laboratório Central do Estado do Paraná | Coordenação Geral de Laboratórios de Saúde Pública (CGLAB/DAEVS/SVS/MS) | Vagner Fonseca et al |
| EPI_ISL_1182557 | Laboratório Central do Estado do Rio de Janeiro | Coordenação Geral de Laboratórios de Saúde Pública (CGLAB/DAEVS/SVS/MS) | Vagner Fonseca et al |
| EPI_ISL_1182556 | Laboratório Central do Estado do Rio de Janeiro | Coordenação Geral de Laboratórios de Saúde Pública (CGLAB/DAEVS/SVS/MS) | Vagner Fonseca et al |
| EPI_ISL_1182552 | Laboratório Central do Estado do Rio de Janeiro | Coordenação Geral de Laboratórios de Saúde Pública (CGLAB/DAEVS/SVS/MS) | Vagner Fonseca et al |
| EPI_ISL_1182554 | Laboratório Central do Estado do Rio de Janeiro | Coordenação Geral de Laboratórios de Saúde Pública (CGLAB/DAEVS/SVS/MS) | Vagner Fonseca et al |
| EPI_ISL_1182553 | Laboratório Central do Estado do Rio de Janeiro | Coordenação Geral de Laboratórios de Saúde Pública (CGLAB/DAEVS/SVS/MS) | Vagner Fonseca et al |
| EPI_ISL_1239112 | Laboratório Central de Saúde Pública Noel Nutels | Coordenação Geral de Laboratórios de Saúde Pública (CGLAB) | Vagner Fonseca et al |
| EPI_ISL_1239113 | Laboratório Central de Saúde Pública Noel Nutels | Coordenação Geral de Laboratórios de Saúde Pública (CGLAB) | Vagner Fonseca et al |
| EPI_ISL_1239111 | Laboratório Central de Saúde Pública Noel Nutels | Coordenação Geral de Laboratórios de Saúde Pública (CGLAB) | Vagner Fonseca et al |
| EPI_ISL_1182621 | Laboratório Central de Saúde Pública do Rio Grande do Sul | Coordenação Geral de Laboratórios de Saúde Pública (CGLAB/DAEVS/SVS/MS) | Vagner Fonseca et al |
| EPI_ISL_1182547 | Laboratório Central de Saúde Pública do Rio Grande do Sul | Coordenação Geral de Laboratórios de Saúde Pública (CGLAB/DAEVS/SVS/MS) | Vagner Fonseca et al |
| EPI_ISL_1182623 | Laboratório Central de Saúde Pública do Rio Grande do Sul | Coordenação Geral de Laboratórios de Saúde Pública (CGLAB/DAEVS/SVS/MS) | Vagner Fonseca et al |
| EPI_ISL_1182619 | Laboratório Central de Saúde Pública do Rio Grande do Sul | Coordenação Geral de Laboratórios de Saúde Pública (CGLAB/DAEVS/SVS/MS) | Vagner Fonseca et al |
| EPI_ISL_1182620 | Laboratório Central de Saúde Pública do Rio Grande do Sul | Coordenação Geral de Laboratórios de Saúde Pública (CGLAB/DAEVS/SVS/MS) | Vagner Fonseca et al |
| EPI_ISL_1182616 | Laboratório Central de Saúde Pública do Rio Grande do Sul | Coordenação Geral de Laboratórios de Saúde Pública (CGLAB/DAEVS/SVS/MS) | Vagner Fonseca et al |
| EPI_ISL_1182606 | Laboratório Central de Saúde Pública do Rio Grande do Sul | Coordenação Geral de Laboratórios de Saúde Pública (CGLAB/DAEVS/SVS/MS) | Vagner Fonseca et al |
| EPI_ISL_1182608 | Laboratório Central de Saúde Pública do Rio Grande do Sul | Coordenação Geral de Laboratórios de Saúde Pública (CGLAB/DAEVS/SVS/MS) | Vagner Fonseca et al |
| EPI_ISL_1182610 | Laboratório Central de Saúde Pública do Rio Grande do Sul | Coordenação Geral de Laboratórios de Saúde Pública (CGLAB/DAEVS/SVS/MS) | Vagner Fonseca et al |
| EPI_ISL_1182605 | Laboratório Central de Saúde Pública do Rio Grande do Sul | Coordenação Geral de Laboratórios de Saúde Pública (CGLAB/DAEVS/SVS/MS) | Vagner Fonseca et al |
| EPI_ISL_1182599 | Laboratório Central de Saúde Pública do Rio Grande do Sul | Coordenação Geral de Laboratórios de Saúde Pública (CGLAB/DAEVS/SVS/MS) | Vagner Fonseca et al |
| EPI_ISL_1182597 | Laboratório Central de Saúde Pública do Rio Grande do Sul | Coordenação Geral de Laboratórios de Saúde Pública (CGLAB/DAEVS/SVS/MS) | Vagner Fonseca et al |
| EPI_ISL_1240640 | Laboratório Central de Saúde Pública do Espírito Santo | Coordenação Geral de Laboratórios de Saúde Pública (CGLAB) | Vagner Fonseca et al. |
| EPI_ISL_1240641 | Laboratório Central de Saúde Pública do Espírito Santo | Coordenação Geral de Laboratórios de Saúde Pública (CGLAB) | Vagner Fonseca et al. |
| EPI_ISL_1240639 | Laboratório Central de Saúde Pública do Espírito Santo | Coordenação Geral de Laboratórios de Saúde Pública (CGLAB) | Vagner Fonseca et al. |

| | | | |
|---|---|---|---|
| EPI_ISL_1240642 | Fundação Ezequiel Dias | Coordenação Geral de Laboratórios de Saúde Pública (CGLAB) | Vagner Fonseca et al. |
| EPI_ISL_417034 | Laboratorio de Ecologia de Doencas Transmissiveis na Amazonia, Instituto Leonidas e Maria Deane - Fiocruz Amazonia | Laboratorio de Ecologia de Doencas Transmissiveis na Amazonia, Instituto Leonidas e Maria Deane - Fiocruz Amazonia | Valdinete Nascimento et al |
| EPI_ISL_1068078 | Laboratorio de Ecologia de Doencas Transmissiveis na Amazonia, Instituto Leonidas e Maria Deane - Fiocruz Amazonia | Laboratorio de Ecologia de Doencas Transmissiveis na Amazonia, Instituto Leonidas e Maria Deane - Fiocruz Amazonia | Valdinete Nascimento et al |
| EPI_ISL_811148 | Laboratorio de Ecologia de Doencas Transmissiveis na Amazonia, Instituto Leonidas e Maria Deane - Fiocruz Amazonia | Laboratorio de Ecologia de Doencas Transmissiveis na Amazonia, Instituto Leonidas e Maria Deane - Fiocruz Amazonia | Valdinete Nascimento et al |
| EPI_ISL_1068079 | Laboratorio de Ecologia de Doencas Transmissiveis na Amazonia, Instituto Leonidas e Maria Deane - Fiocruz Amazonia | Laboratorio de Ecologia de Doencas Transmissiveis na Amazonia, Instituto Leonidas e Maria Deane - Fiocruz Amazonia | Valdinete Nascimento et al |
| EPI_ISL_1068082 | Laboratorio de Ecologia de Doencas Transmissiveis na Amazonia, Instituto Leonidas e Maria Deane - Fiocruz Amazonia | Laboratorio de Ecologia de Doencas Transmissiveis na Amazonia, Instituto Leonidas e Maria Deane - Fiocruz Amazonia | Valdinete Nascimento et al |
| EPI_ISL_1068083 | Laboratorio de Ecologia de Doencas Transmissiveis na Amazonia, Instituto Leonidas e Maria Deane - Fiocruz Amazonia | Laboratorio de Ecologia de Doencas Transmissiveis na Amazonia, Instituto Leonidas e Maria Deane - Fiocruz Amazonia | Valdinete Nascimento et al |
| EPI_ISL_1068084 | Laboratorio de Ecologia de Doencas Transmissiveis na Amazonia, Instituto Leonidas e Maria Deane - Fiocruz Amazonia | Laboratorio de Ecologia de Doencas Transmissiveis na Amazonia, Instituto Leonidas e Maria Deane - Fiocruz Amazonia | Valdinete Nascimento et al |
| EPI_ISL_1068086 | Laboratorio de Ecologia de Doencas Transmissiveis na Amazonia, Instituto Leonidas e Maria Deane - Fiocruz Amazonia | Laboratorio de Ecologia de Doencas Transmissiveis na Amazonia, Instituto Leonidas e Maria Deane - Fiocruz Amazonia | Valdinete Nascimento et al |
| EPI_ISL_1068089 | Laboratorio de Ecologia de Doencas Transmissiveis na Amazonia, Instituto Leonidas e Maria Deane - Fiocruz Amazonia | Laboratorio de Ecologia de Doencas Transmissiveis na Amazonia, Instituto Leonidas e Maria Deane - Fiocruz Amazonia | Valdinete Nascimento et al |
| EPI_ISL_1068090 | Laboratorio de Ecologia de Doencas Transmissiveis na Amazonia, Instituto Leonidas e Maria Deane - Fiocruz Amazonia | Laboratorio de Ecologia de Doencas Transmissiveis na Amazonia, Instituto Leonidas e Maria Deane - Fiocruz Amazonia | Valdinete Nascimento et al |
| EPI_ISL_1068091 | Laboratorio de Ecologia de Doencas Transmissiveis na Amazonia, Instituto Leonidas e Maria Deane - Fiocruz Amazonia | Laboratorio de Ecologia de Doencas Transmissiveis na Amazonia, Instituto Leonidas e Maria Deane - Fiocruz Amazonia | Valdinete Nascimento et al |
| EPI_ISL_1068092 | Laboratorio de Ecologia de Doencas Transmissiveis na Amazonia, Instituto Leonidas e Maria Deane - Fiocruz Amazonia | Laboratorio de Ecologia de Doencas Transmissiveis na Amazonia, Instituto Leonidas e Maria Deane - Fiocruz Amazonia | Valdinete Nascimento et al |
| EPI_ISL_1068095 | Laboratorio de Ecologia de Doencas Transmissiveis na Amazonia, Instituto Leonidas e Maria Deane - Fiocruz Amazonia | Laboratorio de Ecologia de Doencas Transmissiveis na Amazonia, Instituto Leonidas e Maria Deane - Fiocruz Amazonia | Valdinete Nascimento et al |
| EPI_ISL_1068096 | Laboratorio de Ecologia de Doencas Transmissiveis na Amazonia, Instituto Leonidas e Maria Deane - Fiocruz Amazonia | Laboratorio de Ecologia de Doencas Transmissiveis na Amazonia, Instituto Leonidas e Maria Deane - Fiocruz Amazonia | Valdinete Nascimento et al |
| EPI_ISL_1068097 | Laboratorio de Ecologia de Doencas Transmissiveis na Amazonia, Instituto Leonidas e Maria Deane - Fiocruz Amazonia | Laboratorio de Ecologia de Doencas Transmissiveis na Amazonia, Instituto Leonidas e Maria Deane - Fiocruz Amazonia | Valdinete Nascimento et al |
| EPI_ISL_1068098 | Laboratorio de Ecologia de Doencas Transmissiveis na Amazonia, Instituto Leonidas e Maria Deane - Fiocruz Amazonia | Laboratorio de Ecologia de Doencas Transmissiveis na Amazonia, Instituto Leonidas e Maria Deane - Fiocruz Amazonia | Valdinete Nascimento et al |
| EPI_ISL_801386 | Laboratorio de Ecologia de Doencas Transmissiveis na Amazonia, Instituto Leonidas e Maria Deane - Fiocruz Amazonia | Laboratorio de Ecologia de Doencas Transmissiveis na Amazonia, Instituto Leonidas e Maria Deane - Fiocruz Amazonia | Valdinete Nascimento et al |
| EPI_ISL_801387 | Laboratorio de Ecologia de Doencas Transmissiveis na Amazonia, Instituto Leonidas e Maria Deane - Fiocruz Amazonia | Laboratorio de Ecologia de Doencas Transmissiveis na Amazonia, Instituto Leonidas e Maria Deane - Fiocruz Amazonia | Valdinete Nascimento et al |
| EPI_ISL_801388 | Laboratorio de Ecologia de Doencas Transmissiveis na Amazonia, Instituto Leonidas e Maria Deane - Fiocruz Amazonia | Laboratorio de Ecologia de Doencas Transmissiveis na Amazonia, Instituto Leonidas e Maria Deane - Fiocruz Amazonia | Valdinete Nascimento et al |
| EPI_ISL_1068099 | Laboratorio de Ecologia de Doencas Transmissiveis na Amazonia, Instituto Leonidas e Maria Deane - Fiocruz Amazonia | Laboratorio de Ecologia de Doencas Transmissiveis na Amazonia, Instituto Leonidas e Maria Deane - Fiocruz Amazonia | Valdinete Nascimento et al |
| EPI_ISL_801389 | Laboratorio de Ecologia de Doencas Transmissiveis na Amazonia, Instituto Leonidas e Maria Deane - Fiocruz Amazonia | Laboratorio de Ecologia de Doencas Transmissiveis na Amazonia, Instituto Leonidas e Maria Deane - Fiocruz Amazonia | Valdinete Nascimento et al |

| | | | |
|---|---|---|---|
| EPI_ISL_1068101 | Laboratorio de Ecologia de Doencas Transmissiveis na Amazonia, Instituto Leonidas e Maria Deane - Fiocruz Amazonia | Laboratorio de Ecologia de Doencas Transmissiveis na Amazonia, Instituto Leonidas e Maria Deane - Fiocruz Amazonia | Valdinete Nascimento et al |
| EPI_ISL_1068102 | Laboratorio de Ecologia de Doencas Transmissiveis na Amazonia, Instituto Leonidas e Maria Deane - Fiocruz Amazonia | Laboratorio de Ecologia de Doencas Transmissiveis na Amazonia, Instituto Leonidas e Maria Deane - Fiocruz Amazonia | Valdinete Nascimento et al |
| EPI_ISL_1068103 | Laboratorio de Ecologia de Doencas Transmissiveis na Amazonia, Instituto Leonidas e Maria Deane - Fiocruz Amazonia | Laboratorio de Ecologia de Doencas Transmissiveis na Amazonia, Instituto Leonidas e Maria Deane - Fiocruz Amazonia | Valdinete Nascimento et al |
| EPI_ISL_1068104 | Laboratorio de Ecologia de Doencas Transmissiveis na Amazonia, Instituto Leonidas e Maria Deane - Fiocruz Amazonia | Laboratorio de Ecologia de Doencas Transmissiveis na Amazonia, Instituto Leonidas e Maria Deane - Fiocruz Amazonia | Valdinete Nascimento et al |
| EPI_ISL_801390 | Laboratorio de Ecologia de Doencas Transmissiveis na Amazonia, Instituto Leonidas e Maria Deane - Fiocruz Amazonia | Laboratorio de Ecologia de Doencas Transmissiveis na Amazonia, Instituto Leonidas e Maria Deane - Fiocruz Amazonia | Valdinete Nascimento et al |
| EPI_ISL_1068107 | Laboratorio de Ecologia de Doencas Transmissiveis na Amazonia, Instituto Leonidas e Maria Deane - Fiocruz Amazonia | Laboratorio de Ecologia de Doencas Transmissiveis na Amazonia, Instituto Leonidas e Maria Deane - Fiocruz Amazonia | Valdinete Nascimento et al |
| EPI_ISL_801391 | Laboratorio de Ecologia de Doencas Transmissiveis na Amazonia, Instituto Leonidas e Maria Deane - Fiocruz Amazonia | Laboratorio de Ecologia de Doencas Transmissiveis na Amazonia, Instituto Leonidas e Maria Deane - Fiocruz Amazonia | Valdinete Nascimento et al |
| EPI_ISL_801392 | Laboratorio de Ecologia de Doencas Transmissiveis na Amazonia, Instituto Leonidas e Maria Deane - Fiocruz Amazonia | Laboratorio de Ecologia de Doencas Transmissiveis na Amazonia, Instituto Leonidas e Maria Deane - Fiocruz Amazonia | Valdinete Nascimento et al |
| EPI_ISL_833131 | Laboratorio de Ecologia de Doencas Transmissiveis na Amazonia, Instituto Leonidas e Maria Deane - Fiocruz Amazonia | Laboratorio de Ecologia de Doencas Transmissiveis na Amazonia, Instituto Leonidas e Maria Deane - Fiocruz Amazonia | Valdinete Nascimento et al |
| EPI_ISL_1068116 | Laboratorio de Ecologia de Doencas Transmissiveis na Amazonia, Instituto Leonidas e Maria Deane - Fiocruz Amazonia | Laboratorio de Ecologia de Doencas Transmissiveis na Amazonia, Instituto Leonidas e Maria Deane - Fiocruz Amazonia | Valdinete Nascimento et al |
| EPI_ISL_1068120 | Laboratorio de Ecologia de Doencas Transmissiveis na Amazonia, Instituto Leonidas e Maria Deane - Fiocruz Amazonia | Laboratorio de Ecologia de Doencas Transmissiveis na Amazonia, Instituto Leonidas e Maria Deane - Fiocruz Amazonia | Valdinete Nascimento et al |
| EPI_ISL_1068122 | Laboratorio de Ecologia de Doencas Transmissiveis na Amazonia, Instituto Leonidas e Maria Deane - Fiocruz Amazonia | Laboratorio de Ecologia de Doencas Transmissiveis na Amazonia, Instituto Leonidas e Maria Deane - Fiocruz Amazonia | Valdinete Nascimento et al |
| EPI_ISL_1068123 | Laboratorio de Ecologia de Doencas Transmissiveis na Amazonia, Instituto Leonidas e Maria Deane - Fiocruz Amazonia | Laboratorio de Ecologia de Doencas Transmissiveis na Amazonia, Instituto Leonidas e Maria Deane - Fiocruz Amazonia | Valdinete Nascimento et al |
| EPI_ISL_1068124 | Laboratorio de Ecologia de Doencas Transmissiveis na Amazonia, Instituto Leonidas e Maria Deane - Fiocruz Amazonia | Laboratorio de Ecologia de Doencas Transmissiveis na Amazonia, Instituto Leonidas e Maria Deane - Fiocruz Amazonia | Valdinete Nascimento et al |
| EPI_ISL_801393 | Laboratorio de Ecologia de Doencas Transmissiveis na Amazonia, Instituto Leonidas e Maria Deane - Fiocruz Amazonia | Laboratorio de Ecologia de Doencas Transmissiveis na Amazonia, Instituto Leonidas e Maria Deane - Fiocruz Amazonia | Valdinete Nascimento et al |
| EPI_ISL_801394 | Laboratorio de Ecologia de Doencas Transmissiveis na Amazonia, Instituto Leonidas e Maria Deane - Fiocruz Amazonia | Laboratorio de Ecologia de Doencas Transmissiveis na Amazonia, Instituto Leonidas e Maria Deane - Fiocruz Amazonia | Valdinete Nascimento et al |
| EPI_ISL_1068125 | Laboratorio de Ecologia de Doencas Transmissiveis na Amazonia, Instituto Leonidas e Maria Deane - Fiocruz Amazonia | Laboratorio de Ecologia de Doencas Transmissiveis na Amazonia, Instituto Leonidas e Maria Deane - Fiocruz Amazonia | Valdinete Nascimento et al |
| EPI_ISL_801395 | Laboratorio de Ecologia de Doencas Transmissiveis na Amazonia, Instituto Leonidas e Maria Deane - Fiocruz Amazonia | Laboratorio de Ecologia de Doencas Transmissiveis na Amazonia, Instituto Leonidas e Maria Deane - Fiocruz Amazonia | Valdinete Nascimento et al |
| EPI_ISL_1661251 | Laboratorio de Ecologia de Doencas Transmissiveis na Amazonia, Instituto Leonidas e Maria Deane - Fiocruz Amazonia | Laboratorio de Ecologia de Doencas Transmissiveis na Amazonia, Instituto Leonidas e Maria Deane - Fiocruz Amazonia | Valdinete Nascimento et al |
| EPI_ISL_1068129 | Laboratorio de Ecologia de Doencas Transmissiveis na Amazonia, Instituto Leonidas e Maria Deane - Fiocruz Amazonia | Laboratorio de Ecologia de Doencas Transmissiveis na Amazonia, Instituto Leonidas e Maria Deane - Fiocruz Amazonia | Valdinete Nascimento et al |
| EPI_ISL_1068130 | Laboratorio de Ecologia de Doencas Transmissiveis na Amazonia, Instituto Leonidas e Maria Deane - Fiocruz Amazonia | Laboratorio de Ecologia de Doencas Transmissiveis na Amazonia, Instituto Leonidas e Maria Deane - Fiocruz Amazonia | Valdinete Nascimento et al |
| EPI_ISL_801396 | Laboratorio de Ecologia de Doencas Transmissiveis na Amazonia, Instituto Leonidas e Maria Deane - Fiocruz Amazonia | Laboratorio de Ecologia de Doencas Transmissiveis na Amazonia, Instituto Leonidas e Maria Deane - Fiocruz Amazonia | Valdinete Nascimento et al |

| | | | |
|---|---|---|---|
| EPI_ISL_1068134 | Laboratorio de Ecologia de Doencas Transmissiveis na Amazonia, Instituto Leonidas e Maria Deane - Fiocruz Amazonia | Laboratorio de Ecologia de Doencas Transmissiveis na Amazonia, Instituto Leonidas e Maria Deane - Fiocruz Amazonia | Valdinete Nascimento et al |
| EPI_ISL_1068137 | Laboratorio de Ecologia de Doencas Transmissiveis na Amazonia, Instituto Leonidas e Maria Deane - Fiocruz Amazonia | Laboratorio de Ecologia de Doencas Transmissiveis na Amazonia, Instituto Leonidas e Maria Deane - Fiocruz Amazonia | Valdinete Nascimento et al |
| EPI_ISL_1068139 | Laboratorio de Ecologia de Doencas Transmissiveis na Amazonia, Instituto Leonidas e Maria Deane - Fiocruz Amazonia | Laboratorio de Ecologia de Doencas Transmissiveis na Amazonia, Instituto Leonidas e Maria Deane - Fiocruz Amazonia | Valdinete Nascimento et al |
| EPI_ISL_1068142 | Laboratorio de Ecologia de Doencas Transmissiveis na Amazonia, Instituto Leonidas e Maria Deane - Fiocruz Amazonia | Laboratorio de Ecologia de Doencas Transmissiveis na Amazonia, Instituto Leonidas e Maria Deane - Fiocruz Amazonia | Valdinete Nascimento et al |
| EPI_ISL_801397 | Laboratório Central de Saúde Pública do Estado do Amazonas (LACEN-AM) | Laboratorio de Ecologia de Doencas Transmissiveis na Amazonia, Instituto Leonidas e Maria Deane - Fiocruz Amazonia | Valdinete Nascimento et al |
| EPI_ISL_801398 | Laboratório Central de Saúde Pública do Estado do Amazonas (LACEN-AM) | Laboratorio de Ecologia de Doencas Transmissiveis na Amazonia, Instituto Leonidas e Maria Deane - Fiocruz Amazonia | Valdinete Nascimento et al |
| EPI_ISL_1068163 | Laboratorio de Ecologia de Doencas Transmissiveis na Amazonia, Instituto Leonidas e Maria Deane - Fiocruz Amazonia | Laboratorio de Ecologia de Doencas Transmissiveis na Amazonia, Instituto Leonidas e Maria Deane - Fiocruz Amazonia | Valdinete Nascimento et al |
| EPI_ISL_801399 | Laboratório Central de Saúde Pública do Estado do Amazonas (LACEN-AM) | Laboratorio de Ecologia de Doencas Transmissiveis na Amazonia, Instituto Leonidas e Maria Deane - Fiocruz Amazonia | Valdinete Nascimento et al |
| EPI_ISL_1068166 | Laboratorio de Ecologia de Doencas Transmissiveis na Amazonia, Instituto Leonidas e Maria Deane - Fiocruz Amazonia | Laboratorio de Ecologia de Doencas Transmissiveis na Amazonia, Instituto Leonidas e Maria Deane - Fiocruz Amazonia | Valdinete Nascimento et al |
| EPI_ISL_1068167 | Laboratorio de Ecologia de Doencas Transmissiveis na Amazonia, Instituto Leonidas e Maria Deane - Fiocruz Amazonia | Laboratorio de Ecologia de Doencas Transmissiveis na Amazonia, Instituto Leonidas e Maria Deane - Fiocruz Amazonia | Valdinete Nascimento et al |
| EPI_ISL_1068168 | Laboratorio de Ecologia de Doencas Transmissiveis na Amazonia, Instituto Leonidas e Maria Deane - Fiocruz Amazonia | Laboratorio de Ecologia de Doencas Transmissiveis na Amazonia, Instituto Leonidas e Maria Deane - Fiocruz Amazonia | Valdinete Nascimento et al |
| EPI_ISL_1068171 | Laboratorio de Ecologia de Doencas Transmissiveis na Amazonia, Instituto Leonidas e Maria Deane - Fiocruz Amazonia | Laboratorio de Ecologia de Doencas Transmissiveis na Amazonia, Instituto Leonidas e Maria Deane - Fiocruz Amazonia | Valdinete Nascimento et al |
| EPI_ISL_1068172 | Laboratorio de Ecologia de Doencas Transmissiveis na Amazonia, Instituto Leonidas e Maria Deane - Fiocruz Amazonia | Laboratorio de Ecologia de Doencas Transmissiveis na Amazonia, Instituto Leonidas e Maria Deane - Fiocruz Amazonia | Valdinete Nascimento et al |
| EPI_ISL_1068173 | Laboratorio de Ecologia de Doencas Transmissiveis na Amazonia, Instituto Leonidas e Maria Deane - Fiocruz Amazonia | Laboratorio de Ecologia de Doencas Transmissiveis na Amazonia, Instituto Leonidas e Maria Deane - Fiocruz Amazonia | Valdinete Nascimento et al |
| EPI_ISL_1068175 | Laboratorio de Ecologia de Doencas Transmissiveis na Amazonia, Instituto Leonidas e Maria Deane - Fiocruz Amazonia | Laboratorio de Ecologia de Doencas Transmissiveis na Amazonia, Instituto Leonidas e Maria Deane - Fiocruz Amazonia | Valdinete Nascimento et al |
| EPI_ISL_801400 | Laboratório Central de Saúde Pública do Estado do Amazonas (LACEN-AM) | Laboratorio de Ecologia de Doencas Transmissiveis na Amazonia, Instituto Leonidas e Maria Deane - Fiocruz Amazonia | Valdinete Nascimento et al |
| EPI_ISL_801401 | Laboratório Central de Saúde Pública do Estado do Amazonas (LACEN-AM) | Laboratorio de Ecologia de Doencas Transmissiveis na Amazonia, Instituto Leonidas e Maria Deane - Fiocruz Amazonia | Valdinete Nascimento et al |
| EPI_ISL_1068180 | Laboratorio de Ecologia de Doencas Transmissiveis na Amazonia, Instituto Leonidas e Maria Deane - Fiocruz Amazonia | Laboratorio de Ecologia de Doencas Transmissiveis na Amazonia, Instituto Leonidas e Maria Deane - Fiocruz Amazonia | Valdinete Nascimento et al |
| EPI_ISL_1068182 | Laboratorio de Ecologia de Doencas Transmissiveis na Amazonia, Instituto Leonidas e Maria Deane - Fiocruz Amazonia | Laboratorio de Ecologia de Doencas Transmissiveis na Amazonia, Instituto Leonidas e Maria Deane - Fiocruz Amazonia | Valdinete Nascimento et al |
| EPI_ISL_1068186 | Laboratorio de Ecologia de Doencas Transmissiveis na Amazonia, Instituto Leonidas e Maria Deane - Fiocruz Amazonia | Laboratorio de Ecologia de Doencas Transmissiveis na Amazonia, Instituto Leonidas e Maria Deane - Fiocruz Amazonia | Valdinete Nascimento et al |
| EPI_ISL_801402 | Laboratório Central de Saúde Pública do Estado do Amazonas (LACEN-AM) | Laboratorio de Ecologia de Doencas Transmissiveis na Amazonia, Instituto Leonidas e Maria Deane - Fiocruz Amazonia | Valdinete Nascimento et al |
| EPI_ISL_1068189 | Laboratorio de Ecologia de Doencas Transmissiveis na Amazonia, Instituto Leonidas e Maria Deane - Fiocruz Amazonia | Laboratorio de Ecologia de Doencas Transmissiveis na Amazonia, Instituto Leonidas e Maria Deane - Fiocruz Amazonia | Valdinete Nascimento et al |

| | | | |
|---|---|---|---|
| EPI_ISL_1068190 | Laboratorio de Ecologia de Doencas Transmissiveis na Amazonia, Instituto Leonidas e Maria Deane - Fiocruz Amazonia | Laboratorio de Ecologia de Doencas Transmissiveis na Amazonia, Instituto Leonidas e Maria Deane - Fiocruz Amazonia | Valdinete Nascimento et al |
| EPI_ISL_1068192 | Laboratorio de Ecologia de Doencas Transmissiveis na Amazonia, Instituto Leonidas e Maria Deane - Fiocruz Amazonia | Laboratorio de Ecologia de Doencas Transmissiveis na Amazonia, Instituto Leonidas e Maria Deane - Fiocruz Amazonia | Valdinete Nascimento et al |
| EPI_ISL_1068195 | Laboratorio de Ecologia de Doencas Transmissiveis na Amazonia, Instituto Leonidas e Maria Deane - Fiocruz Amazonia | Laboratorio de Ecologia de Doencas Transmissiveis na Amazonia, Instituto Leonidas e Maria Deane - Fiocruz Amazonia | Valdinete Nascimento et al |
| EPI_ISL_1068197 | Laboratorio de Ecologia de Doencas Transmissiveis na Amazonia, Instituto Leonidas e Maria Deane - Fiocruz Amazonia | Laboratorio de Ecologia de Doencas Transmissiveis na Amazonia, Instituto Leonidas e Maria Deane - Fiocruz Amazonia | Valdinete Nascimento et al |
| EPI_ISL_1068200 | Laboratorio de Ecologia de Doencas Transmissiveis na Amazonia, Instituto Leonidas e Maria Deane - Fiocruz Amazonia | Laboratorio de Ecologia de Doencas Transmissiveis na Amazonia, Instituto Leonidas e Maria Deane - Fiocruz Amazonia | Valdinete Nascimento et al |
| EPI_ISL_1068201 | Laboratorio de Ecologia de Doencas Transmissiveis na Amazonia, Instituto Leonidas e Maria Deane - Fiocruz Amazonia | Laboratorio de Ecologia de Doencas Transmissiveis na Amazonia, Instituto Leonidas e Maria Deane - Fiocruz Amazonia | Valdinete Nascimento et al |
| EPI_ISL_1068202 | Laboratorio de Ecologia de Doencas Transmissiveis na Amazonia, Instituto Leonidas e Maria Deane - Fiocruz Amazonia | Laboratorio de Ecologia de Doencas Transmissiveis na Amazonia, Instituto Leonidas e Maria Deane - Fiocruz Amazonia | Valdinete Nascimento et al |
| EPI_ISL_1068204 | Laboratorio de Ecologia de Doencas Transmissiveis na Amazonia, Instituto Leonidas e Maria Deane - Fiocruz Amazonia | Laboratorio de Ecologia de Doencas Transmissiveis na Amazonia, Instituto Leonidas e Maria Deane - Fiocruz Amazonia | Valdinete Nascimento et al |
| EPI_ISL_1068205 | Laboratorio de Ecologia de Doencas Transmissiveis na Amazonia, Instituto Leonidas e Maria Deane - Fiocruz Amazonia | Laboratorio de Ecologia de Doencas Transmissiveis na Amazonia, Instituto Leonidas e Maria Deane - Fiocruz Amazonia | Valdinete Nascimento et al |
| EPI_ISL_1068206 | Laboratorio de Ecologia de Doencas Transmissiveis na Amazonia, Instituto Leonidas e Maria Deane - Fiocruz Amazonia | Laboratorio de Ecologia de Doencas Transmissiveis na Amazonia, Instituto Leonidas e Maria Deane - Fiocruz Amazonia | Valdinete Nascimento et al |
| EPI_ISL_1068207 | Laboratorio de Ecologia de Doencas Transmissiveis na Amazonia, Instituto Leonidas e Maria Deane - Fiocruz Amazonia | Laboratorio de Ecologia de Doencas Transmissiveis na Amazonia, Instituto Leonidas e Maria Deane - Fiocruz Amazonia | Valdinete Nascimento et al |
| EPI_ISL_1068208 | Laboratorio de Ecologia de Doencas Transmissiveis na Amazonia, Instituto Leonidas e Maria Deane - Fiocruz Amazonia | Laboratorio de Ecologia de Doencas Transmissiveis na Amazonia, Instituto Leonidas e Maria Deane - Fiocruz Amazonia | Valdinete Nascimento et al |
| EPI_ISL_1068209 | Laboratorio de Ecologia de Doencas Transmissiveis na Amazonia, Instituto Leonidas e Maria Deane - Fiocruz Amazonia | Laboratorio de Ecologia de Doencas Transmissiveis na Amazonia, Instituto Leonidas e Maria Deane - Fiocruz Amazonia | Valdinete Nascimento et al |
| EPI_ISL_1068210 | Laboratorio de Ecologia de Doencas Transmissiveis na Amazonia, Instituto Leonidas e Maria Deane - Fiocruz Amazonia | Laboratorio de Ecologia de Doencas Transmissiveis na Amazonia, Instituto Leonidas e Maria Deane - Fiocruz Amazonia | Valdinete Nascimento et al |
| EPI_ISL_1068211 | Laboratorio de Ecologia de Doencas Transmissiveis na Amazonia, Instituto Leonidas e Maria Deane - Fiocruz Amazonia | Laboratorio de Ecologia de Doencas Transmissiveis na Amazonia, Instituto Leonidas e Maria Deane - Fiocruz Amazonia | Valdinete Nascimento et al |
| EPI_ISL_1068213 | Laboratorio de Ecologia de Doencas Transmissiveis na Amazonia, Instituto Leonidas e Maria Deane - Fiocruz Amazonia | Laboratorio de Ecologia de Doencas Transmissiveis na Amazonia, Instituto Leonidas e Maria Deane - Fiocruz Amazonia | Valdinete Nascimento et al |
| EPI_ISL_1068214 | Laboratorio de Ecologia de Doencas Transmissiveis na Amazonia, Instituto Leonidas e Maria Deane - Fiocruz Amazonia | Laboratorio de Ecologia de Doencas Transmissiveis na Amazonia, Instituto Leonidas e Maria Deane - Fiocruz Amazonia | Valdinete Nascimento et al |
| EPI_ISL_1068216 | Laboratorio de Ecologia de Doencas Transmissiveis na Amazonia, Instituto Leonidas e Maria Deane - Fiocruz Amazonia | Laboratorio de Ecologia de Doencas Transmissiveis na Amazonia, Instituto Leonidas e Maria Deane - Fiocruz Amazonia | Valdinete Nascimento et al |
| EPI_ISL_1068217 | Laboratorio de Ecologia de Doencas Transmissiveis na Amazonia, Instituto Leonidas e Maria Deane - Fiocruz Amazonia | Laboratorio de Ecologia de Doencas Transmissiveis na Amazonia, Instituto Leonidas e Maria Deane - Fiocruz Amazonia | Valdinete Nascimento et al |
| EPI_ISL_1068218 | Laboratorio de Ecologia de Doencas Transmissiveis na Amazonia, Instituto Leonidas e Maria Deane - Fiocruz Amazonia | Laboratorio de Ecologia de Doencas Transmissiveis na Amazonia, Instituto Leonidas e Maria Deane - Fiocruz Amazonia | Valdinete Nascimento et al |
| EPI_ISL_1068220 | Laboratorio de Ecologia de Doencas Transmissiveis na Amazonia, Instituto Leonidas e Maria Deane - Fiocruz Amazonia | Laboratorio de Ecologia de Doencas Transmissiveis na Amazonia, Instituto Leonidas e Maria Deane - Fiocruz Amazonia | Valdinete Nascimento et al |
| EPI_ISL_1068224 | Laboratorio de Ecologia de Doencas Transmissiveis na Amazonia, Instituto Leonidas e Maria Deane - Fiocruz Amazonia | Laboratorio de Ecologia de Doencas Transmissiveis na Amazonia, Instituto Leonidas e Maria Deane - Fiocruz Amazonia | Valdinete Nascimento et al |

| | | | |
|---|---|---|---|
| EPI_ISL_1068227 | Laboratorio de Ecologia de Doencas Transmissiveis na Amazonia, Instituto Leonidas e Maria Deane - Fiocruz Amazonia | Laboratorio de Ecologia de Doencas Transmissiveis na Amazonia, Instituto Leonidas e Maria Deane - Fiocruz Amazonia | Valdinete Nascimento et al |
| EPI_ISL_1068228 | Laboratorio de Ecologia de Doencas Transmissiveis na Amazonia, Instituto Leonidas e Maria Deane - Fiocruz Amazonia | Laboratorio de Ecologia de Doencas Transmissiveis na Amazonia, Instituto Leonidas e Maria Deane - Fiocruz Amazonia | Valdinete Nascimento et al |
| EPI_ISL_1068229 | Laboratorio de Ecologia de Doencas Transmissiveis na Amazonia, Instituto Leonidas e Maria Deane - Fiocruz Amazonia | Laboratorio de Ecologia de Doencas Transmissiveis na Amazonia, Instituto Leonidas e Maria Deane - Fiocruz Amazonia | Valdinete Nascimento et al |
| EPI_ISL_1068230 | Laboratorio de Ecologia de Doencas Transmissiveis na Amazonia, Instituto Leonidas e Maria Deane - Fiocruz Amazonia | Laboratorio de Ecologia de Doencas Transmissiveis na Amazonia, Instituto Leonidas e Maria Deane - Fiocruz Amazonia | Valdinete Nascimento et al |
| EPI_ISL_1068231 | Laboratorio de Ecologia de Doencas Transmissiveis na Amazonia, Instituto Leonidas e Maria Deane - Fiocruz Amazonia | Laboratorio de Ecologia de Doencas Transmissiveis na Amazonia, Instituto Leonidas e Maria Deane - Fiocruz Amazonia | Valdinete Nascimento et al |
| EPI_ISL_1068232 | Laboratorio de Ecologia de Doencas Transmissiveis na Amazonia, Instituto Leonidas e Maria Deane - Fiocruz Amazonia | Laboratorio de Ecologia de Doencas Transmissiveis na Amazonia, Instituto Leonidas e Maria Deane - Fiocruz Amazonia | Valdinete Nascimento et al |
| EPI_ISL_1068233 | Laboratorio de Ecologia de Doencas Transmissiveis na Amazonia, Instituto Leonidas e Maria Deane - Fiocruz Amazonia | Laboratorio de Ecologia de Doencas Transmissiveis na Amazonia, Instituto Leonidas e Maria Deane - Fiocruz Amazonia | Valdinete Nascimento et al |
| EPI_ISL_1068234 | Laboratorio de Ecologia de Doencas Transmissiveis na Amazonia, Instituto Leonidas e Maria Deane - Fiocruz Amazonia | Laboratorio de Ecologia de Doencas Transmissiveis na Amazonia, Instituto Leonidas e Maria Deane - Fiocruz Amazonia | Valdinete Nascimento et al |
| EPI_ISL_801403 | Laboratório Central de Saúde Pública do Estado do Amazonas (LACEN-AM) | Laboratorio de Ecologia de Doencas Transmissiveis na Amazonia, Instituto Leonidas e Maria Deane - Fiocruz Amazonia | Valdinete Nascimento et al |
| EPI_ISL_1068236 | Laboratorio de Ecologia de Doencas Transmissiveis na Amazonia, Instituto Leonidas e Maria Deane - Fiocruz Amazonia | Laboratorio de Ecologia de Doencas Transmissiveis na Amazonia, Instituto Leonidas e Maria Deane - Fiocruz Amazonia | Valdinete Nascimento et al |
| EPI_ISL_1068240 | Laboratorio de Ecologia de Doencas Transmissiveis na Amazonia, Instituto Leonidas e Maria Deane - Fiocruz Amazonia | Laboratorio de Ecologia de Doencas Transmissiveis na Amazonia, Instituto Leonidas e Maria Deane - Fiocruz Amazonia | Valdinete Nascimento et al |
| EPI_ISL_1068241 | Laboratorio de Ecologia de Doencas Transmissiveis na Amazonia, Instituto Leonidas e Maria Deane - Fiocruz Amazonia | Laboratorio de Ecologia de Doencas Transmissiveis na Amazonia, Instituto Leonidas e Maria Deane - Fiocruz Amazonia | Valdinete Nascimento et al |
| EPI_ISL_1068242 | Laboratorio de Ecologia de Doencas Transmissiveis na Amazonia, Instituto Leonidas e Maria Deane - Fiocruz Amazonia | Laboratorio de Ecologia de Doencas Transmissiveis na Amazonia, Instituto Leonidas e Maria Deane - Fiocruz Amazonia | Valdinete Nascimento et al |
| EPI_ISL_1068244 | Laboratorio de Ecologia de Doencas Transmissiveis na Amazonia, Instituto Leonidas e Maria Deane - Fiocruz Amazonia | Laboratorio de Ecologia de Doencas Transmissiveis na Amazonia, Instituto Leonidas e Maria Deane - Fiocruz Amazonia | Valdinete Nascimento et al |
| EPI_ISL_1068245 | Laboratorio de Ecologia de Doencas Transmissiveis na Amazonia, Instituto Leonidas e Maria Deane - Fiocruz Amazonia | Laboratorio de Ecologia de Doencas Transmissiveis na Amazonia, Instituto Leonidas e Maria Deane - Fiocruz Amazonia | Valdinete Nascimento et al |
| EPI_ISL_1068246 | Laboratorio de Ecologia de Doencas Transmissiveis na Amazonia, Instituto Leonidas e Maria Deane - Fiocruz Amazonia | Laboratorio de Ecologia de Doencas Transmissiveis na Amazonia, Instituto Leonidas e Maria Deane - Fiocruz Amazonia | Valdinete Nascimento et al |
| EPI_ISL_1068247 | Laboratorio de Ecologia de Doencas Transmissiveis na Amazonia, Instituto Leonidas e Maria Deane - Fiocruz Amazonia | Laboratorio de Ecologia de Doencas Transmissiveis na Amazonia, Instituto Leonidas e Maria Deane - Fiocruz Amazonia | Valdinete Nascimento et al |
| EPI_ISL_1068250 | Laboratorio de Ecologia de Doencas Transmissiveis na Amazonia, Instituto Leonidas e Maria Deane - Fiocruz Amazonia | Laboratorio de Ecologia de Doencas Transmissiveis na Amazonia, Instituto Leonidas e Maria Deane - Fiocruz Amazonia | Valdinete Nascimento et al |
| EPI_ISL_1068252 | Laboratorio de Ecologia de Doencas Transmissiveis na Amazonia, Instituto Leonidas e Maria Deane - Fiocruz Amazonia | Laboratorio de Ecologia de Doencas Transmissiveis na Amazonia, Instituto Leonidas e Maria Deane - Fiocruz Amazonia | Valdinete Nascimento et al |
| EPI_ISL_1068253 | Laboratorio de Ecologia de Doencas Transmissiveis na Amazonia, Instituto Leonidas e Maria Deane - Fiocruz Amazonia | Laboratorio de Ecologia de Doencas Transmissiveis na Amazonia, Instituto Leonidas e Maria Deane - Fiocruz Amazonia | Valdinete Nascimento et al |
| EPI_ISL_1068254 | Laboratorio de Ecologia de Doencas Transmissiveis na Amazonia, Instituto Leonidas e Maria Deane - Fiocruz Amazonia | Laboratorio de Ecologia de Doencas Transmissiveis na Amazonia, Instituto Leonidas e Maria Deane - Fiocruz Amazonia | Valdinete Nascimento et al |
| EPI_ISL_1533609 | Laboratorio de Ecologia de Doencas Transmissiveis na Amazonia, Instituto Leonidas e Maria Deane - Fiocruz Amazonia | Laboratorio de Ecologia de Doencas Transmissiveis na Amazonia, Instituto Leonidas e Maria Deane - Fiocruz Amazonia | Valdinete Nascimento et al |

| | | | |
|---|---|---|---|
| EPI_ISL_1068258 | Laboratorio de Ecologia de Doencas Transmissiveis na Amazonia, Instituto Leonidas e Maria Deane - Fiocruz Amazonia | Laboratorio de Ecologia de Doencas Transmissiveis na Amazonia, Instituto Leonidas e Maria Deane - Fiocruz Amazonia | Valdinete Nascimento et al |
| EPI_ISL_1015159 | Department of Virology, Pitié-Salpêtrière hospital | Department of Virology, Pitié-Salpêtrière hospital | Valentin Leducq et al |
| EPI_ISL_1395782 | Instituto de Investigaciones Biomédicas en Retrovirus y SIDA (INBIRS) | Área de Secuenciación del Laboratorio de Virología del Hospital de Niños Dr. Ricardo Gutierrez on behalf of 'Proyecto Argentino Interinstitucional de genomica de SARS-CoV-2' (PAIS Consortium) | Vanesa Seery et al |
| EPI_ISL_1395783 | Instituto de Investigaciones Biomédicas en Retrovirus y SIDA (INBIRS) | Área de Secuenciación del Laboratorio de Virología del Hospital de Niños Dr. Ricardo Gutierrez on behalf of 'Proyecto Argentino Interinstitucional de genomica de SARS-CoV-2' (PAIS Consortium) | Vanesa Seery et al |
| EPI_ISL_1395784 | Instituto de Investigaciones Biomédicas en Retrovirus y SIDA (INBIRS) | Área de Secuenciación del Laboratorio de Virología del Hospital de Niños Dr. Ricardo Gutierrez on behalf of 'Proyecto Argentino Interinstitucional de genomica de SARS-CoV-2' (PAIS Consortium) | Vanesa Seery et al |
| EPI_ISL_1395802 | Instituto de Investigaciones Biomédicas en Retrovirus y SIDA (INBIRS) | Área de Secuenciación del Laboratorio de Virología del Hospital de Niños Dr. Ricardo Gutierrez on behalf of 'Proyecto Argentino Interinstitucional de genomica de SARS-CoV-2' (PAIS Consortium) | Vanesa Seery et al |
| EPI_ISL_1251212 | Raimundo Reginaldo de Almeida | MOA Lab | Vanessa C. Nicolete et al |
| EPI_ISL_1251214 | Raimundo Reginaldo de Almeida | MOA Lab | Vanessa C. Nicolete et al |
| EPI_ISL_1251217 | Raimundo Reginaldo de Almeida | MOA Lab | Vanessa C. Nicolete et al |
| EPI_ISL_1251224 | Raimundo Reginaldo de Almeida | MOA Lab | Vanessa C. Nicolete et al |
| EPI_ISL_1251222 | Raimundo Reginaldo de Almeida | MOA Lab | Vanessa C. Nicolete et al |
| EPI_ISL_1251225 | Raimundo Reginaldo de Almeida | MOA Lab | Vanessa C. Nicolete et al |
| EPI_ISL_1251213 | Raimundo Reginaldo de Almeida | MOA Lab | Vanessa C. Nicolete et al |
| EPI_ISL_1251216 | Raimundo Reginaldo de Almeida | MOA Lab | Vanessa C. Nicolete et al |
| EPI_ISL_1251223 | Raimundo Reginaldo de Almeida | MOA Lab | Vanessa C. Nicolete et al |
| EPI_ISL_430469 | Hellenic Pasteur Institute, Public Health Laboratories | Hellenic Pasteur Institute, Public Health Laboratories, Unit of Bioinformatics and Applied Genomics | Vasiliki Pogka et al |
| EPI_ISL_501248 | Hellenic Pasteur Institute, National Influenza Reference laboratory of Southern Greece & Unit of Bioinformatics and Applied Genomics | Hellenic Pasteur Institute, National Influenza Reference laboratory of Southern Greece & Unit of Bioinformatics and Applied Genomics | Vasiliki Pogka et al |
| EPI_ISL_501249 | Hellenic Pasteur Institute, National Influenza Reference laboratory of Southern Greece & Unit of Bioinformatics and Applied Genomics | Hellenic Pasteur Institute, National Influenza Reference laboratory of Southern Greece & Unit of Bioinformatics and Applied Genomics | Vasiliki Pogka et al |
| EPI_ISL_501230 | Hellenic Pasteur Institute, Public Health Laboratories | Hellenic Pasteur Institute, National Influenza Reference laboratory of Southern Greece & Unit of Bioinformatics and Applied Genomics | Vasiliki Pogka et al |
| EPI_ISL_501233 | Hellenic Pasteur Institute, National Influenza Reference laboratory of Southern Greece & Unit of Bioinformatics and Applied Genomics | Hellenic Pasteur Institute, National Influenza Reference laboratory of Southern Greece & Unit of Bioinformatics and Applied Genomics | Vasiliki Pogka et al |
| EPI_ISL_501250 | Hellenic Pasteur Institute, National Influenza Reference laboratory of Southern Greece & Unit of Bioinformatics and Applied Genomics | Hellenic Pasteur Institute, National Influenza Reference laboratory of Southern Greece & Unit of Bioinformatics and Applied Genomics | Vasiliki Pogka et al |
| EPI_ISL_501242 | Hellenic Pasteur Institute, National Influenza Reference laboratory of Southern Greece & Unit of Bioinformatics and Applied Genomics | Hellenic Pasteur Institute, National Influenza Reference laboratory of Southern Greece & Unit of Bioinformatics and Applied Genomics | Vasiliki Pogka et al |
| EPI_ISL_501251 | Hellenic Pasteur Institute, National Influenza Reference laboratory of Southern Greece & Unit of Bioinformatics and Applied Genomics | Hellenic Pasteur Institute, National Influenza Reference laboratory of Southern Greece & Unit of Bioinformatics and Applied Genomics | Vasiliki Pogka et al |

| | | | |
|---|---|---|---|
| EPI_ISL_451311 | Hellenic Pasteur Institute, National Influenza Reference laboratory of Southern Greece & Unit of Bioinformatics and Applied Genomics | Hellenic Pasteur Institute, National Influenza Reference laboratory of Southern Greece & Unit of Bioinformatics and Applied Genomics | Vasiliki Pogka et al |
| EPI_ISL_451310 | Hellenic Pasteur Institute, National Influenza Reference laboratory of Southern Greece & Unit of Bioinformatics and Applied Genomics | Hellenic Pasteur Institute, National Influenza Reference laboratory of Southern Greece & Unit of Bioinformatics and Applied Genomics | Vasiliki Pogka et al |
| EPI_ISL_1360411 | Laboratorio de Enfermedades Emergentes y Reemergentes | Laboratorio de Enfermedades Emergentes y Reemergentes | Vega-Magaña Natali et al |
| EPI_ISL_1278371 | Thai Red Cross Emerging Infectious Diseases Health Science Centre, Chulalongkorn Hospital, Faculty of Medicine, Chulalongkorn University | Thai Red Cross Emerging Infectious Diseases Center and Faculty of Medicine, Chulalongkorn University | Vichan Pawun et al |
| EPI_ISL_861457 | WHO/Minsk | Charité Universitätsmedizin Berlin, Institut für Virologie | Victor M Corman et al |
| EPI_ISL_860805 | WHO/Minsk | Charité Universitätsmedizin Berlin, Institut für Virologie | Victor M Corman et al |
| EPI_ISL_860811 | WHO/Minsk | Charité Universitätsmedizin Berlin, Institut für Virologie | Victor M Corman et al |
| EPI_ISL_462450 | Clinical Center, University of Sarajevo | Charite Universitatsmedizin Berlin, Institute of Virology | Victor M Corman et al |
| EPI_ISL_462462 | Clinical Center, University of Sarajevo | Charite Universitatsmedizin Berlin, Institute of Virology | Victor M Corman et al |
| EPI_ISL_462463 | Clinical Center, University of Sarajevo | Charite Universitatsmedizin Berlin, Institute of Virology | Victor M Corman et al |
| EPI_ISL_462467 | Clinical Center, University of Sarajevo | Charite Universitatsmedizin Berlin, Institute of Virology | Victor M Corman et al |
| EPI_ISL_462476 | Clinical Center, University of Sarajevo | Charite Universitatsmedizin Berlin, Institute of Virology | Victor M Corman et al |
| EPI_ISL_462472 | Clinical Center, University of Sarajevo | Charite Universitatsmedizin Berlin, Institute of Virology | Victor M Corman et al |
| EPI_ISL_462461 | Clinical Center, University of Sarajevo | Charite Universitatsmedizin Berlin, Institute of Virology | Victor M Corman et al |
| EPI_ISL_462466 | Clinical Center, University of Sarajevo | Charite Universitatsmedizin Berlin, Institute of Virology | Victor M Corman et al |
| EPI_ISL_462470 | Clinical Center, University of Sarajevo | Charite Universitatsmedizin Berlin, Institute of Virology | Victor M Corman et al |
| EPI_ISL_416742 | NRL for Influenza, Centrum Epidemiology and Microbiology of National Institute of Public Health, Czech Republic | Charite Universitaetsmedizin Berlin, Institute of Virology | Victor M Corman et al |
| EPI_ISL_753746 | Charité Universitätsmedizin Berlin, Institut für Virologie/Labor Berlin | Charité Universitätsmedizin Berlin, Institut für Virologie | Victor M Corman et al |
| EPI_ISL_753706 | Charité Universitätsmedizin Berlin, Institut für Virologie/Labor Berlin | Charité Universitätsmedizin Berlin, Institut für Virologie | Victor M Corman et al |
| EPI_ISL_753726 | Charité Universitätsmedizin Berlin, Institut für Virologie/Labor Berlin | Charité Universitätsmedizin Berlin, Institut für Virologie | Victor M Corman et al |
| EPI_ISL_753982 | Charité Universitätsmedizin Berlin, Institut für Virologie/Labor Berlin | Charité Universitätsmedizin Berlin, Institut für Virologie | Victor M Corman et al |
| EPI_ISL_753986 | Charité Universitätsmedizin Berlin, Institut für Virologie/Labor Berlin | Charité Universitätsmedizin Berlin, Institut für Virologie | Victor M Corman et al |
| EPI_ISL_753708 | Charité Universitätsmedizin Berlin, Institut für Virologie/Labor Berlin | Charité Universitätsmedizin Berlin, Institut für Virologie | Victor M Corman et al |
| EPI_ISL_729473 | Charité Universitätsmedizin Berlin, Institut für Virologie/Labor Berlin | Charité Universitätsmedizin Berlin, Institut für Virologie | Victor M Corman et al |
| EPI_ISL_406862 | Charité Universitätsmedizin Berlin, Institute of Virology; Institut für Mikrobiologie der Bundeswehr, Munich | Charité Universitätsmedizin Berlin, Institute of Virology | Victor M Corman et al |
| EPI_ISL_729573 | A. Krumbholz, Labor Dr. Krause und Kollegen MVZ GmbH, Kiel | Charité Universitätsmedizin Berlin, Institut für Virologie | Victor M Corman et al |
| EPI_ISL_729589 | A. Krumbholz, Labor Dr. Krause und Kollegen MVZ GmbH, Kiel | Charité Universitätsmedizin Berlin, Institut für Virologie | Victor M Corman et al |
| EPI_ISL_729660 | A. Krumbholz, Labor Dr. Krause und Kollegen MVZ GmbH, Kiel | Charité Universitätsmedizin Berlin, Institut für Virologie | Victor M Corman et al |

| | | | |
|---|---|---|---|
| EPI_ISL_1027653 | Department of Microbiology, National Institute for Public Health of Kosova | Charité Universitätsmedizin Berlin, Institut für Virologie | Victor M Corman et al |
| EPI_ISL_1027654 | Department of Microbiology, National Institute for Public Health of Kosova | Charité Universitätsmedizin Berlin, Institut für Virologie | Victor M Corman et al |
| EPI_ISL_1027652 | Department of Microbiology, National Institute for Public Health of Kosova | Charité Universitätsmedizin Berlin, Institut für Virologie | Victor M Corman et al |
| EPI_ISL_1027638 | Department of Microbiology, National Institute for Public Health of Kosova | Charité Universitätsmedizin Berlin, Institut für Virologie | Victor M Corman et al |
| EPI_ISL_1027640 | Department of Microbiology, National Institute for Public Health of Kosova | Charité Universitätsmedizin Berlin, Institut für Virologie | Victor M Corman et al |
| EPI_ISL_1027645 | Department of Microbiology, National Institute for Public Health of Kosova | Charité Universitätsmedizin Berlin, Institut für Virologie | Victor M Corman et al |
| EPI_ISL_1027646 | Department of Microbiology, National Institute for Public Health of Kosova | Charité Universitätsmedizin Berlin, Institut für Virologie | Victor M Corman et al |
| EPI_ISL_1027648 | Department of Microbiology, National Institute for Public Health of Kosova | Charité Universitätsmedizin Berlin, Institut für Virologie | Victor M Corman et al |
| EPI_ISL_1027650 | Department of Microbiology, National Institute for Public Health of Kosova | Charité Universitätsmedizin Berlin, Institut für Virologie | Victor M Corman et al |
| EPI_ISL_1027651 | Department of Microbiology, National Institute for Public Health of Kosova | Charité Universitätsmedizin Berlin, Institut für Virologie | Victor M Corman et al |
| EPI_ISL_523960 | National Agency for Public Health, Republic of Moldova | Charite Universitatsmedizin Berlin, Institute of Virology | Victor M Corman et al |
| EPI_ISL_523962 | National Agency for Public Health, Republic of Moldova | Charite Universitatsmedizin Berlin, Institute of Virology | Victor M Corman et al |
| EPI_ISL_523964 | National Agency for Public Health, Republic of Moldova | Charite Universitatsmedizin Berlin, Institute of Virology | Victor M Corman et al |
| EPI_ISL_523968 | National Agency for Public Health, Republic of Moldova | Charite Universitatsmedizin Berlin, Institute of Virology | Victor M Corman et al |
| EPI_ISL_523966 | National Agency for Public Health, Republic of Moldova | Charite Universitatsmedizin Berlin, Institute of Virology | Victor M Corman et al |
| EPI_ISL_516922 | Department for Molecular Diagnostics, Centre for Medical Microbiology, Institute of Public Health of Montenegro | Charité Universitätsmedizin Berlin, Institut für Virologie | Victor M Corman et al |
| EPI_ISL_516927 | Department for Molecular Diagnostics, Centre for Medical Microbiology, Institute of Public Health of Montenegro | Charité Universitätsmedizin Berlin, Institut für Virologie | Victor M Corman et al |
| EPI_ISL_516928 | Department for Molecular Diagnostics, Centre for Medical Microbiology, Institute of Public Health of Montenegro | Charité Universitätsmedizin Berlin, Institut für Virologie | Victor M Corman et al |
| EPI_ISL_516929 | Department for Molecular Diagnostics, Centre for Medical Microbiology, Institute of Public Health of Montenegro | Charité Universitätsmedizin Berlin, Institut für Virologie | Victor M Corman et al |
| EPI_ISL_516930 | Department for Molecular Diagnostics, Centre for Medical Microbiology, Institute of Public Health of Montenegro | Charité Universitätsmedizin Berlin, Institut für Virologie | Victor M Corman et al |
| EPI_ISL_516931 | Department for Molecular Diagnostics, Centre for Medical Microbiology, Institute of Public Health of Montenegro | Charité Universitätsmedizin Berlin, Institut für Virologie | Victor M Corman et al |
| EPI_ISL_516932 | Department for Molecular Diagnostics, Centre for Medical Microbiology, Institute of Public Health of Montenegro | Charité Universitätsmedizin Berlin, Institut für Virologie | Victor M Corman et al |
| EPI_ISL_1013598 | Department for Molecular Diagnostics, Centre for Medical Microbiology, Institute of Public Health, Montenegro | Charité Universitätsmedizin Berlin, Institut für Virologie | Victor M Corman et al |
| EPI_ISL_1013601 | Department for Molecular Diagnostics, Centre for Medical Microbiology, Institute of Public Health, Montenegro | Charité Universitätsmedizin Berlin, Institut für Virologie | Victor M Corman et al |
| EPI_ISL_1013614 | Department for Molecular Diagnostics, Centre for Medical Microbiology, Institute of Public Health, Montenegro | Charité Universitätsmedizin Berlin, Institut für Virologie | Victor M Corman et al |
| EPI_ISL_492076 | Institute for Public Health of the Republic of North Macedonia | Charite Universitatsmedizin Berlin, Institute of Virology | Victor M Corman et al |
| EPI_ISL_541658 | Laboratory Diagnostic, Veterinary Specialized Institute Kraljevo | Laboratory Diagnostic, Veterinary Specialized Institute Kraljevo | Vidanovic et al |

| | | | |
|---|---|---|---|
| EPI_ISL_582530 | Laboratory Diagnostic, Veterinary Specialized Institute Kraljevo | Laboratory Diagnostic, Veterinary Specialized Institute Kraljevo | Vidanovic et al |
| EPI_ISL_644567 | Laboratory Diagnostic, Veterinary Specialized Institute Kraljevo | Laboratory Diagnostic, Veterinary Specialized Institute Kraljevo | Vidanovic et al |
| EPI_ISL_541655 | Laboratory Diagnostic, Veterinary Specialized Institute Kraljevo | Laboratory Diagnostic, Veterinary Specialized Institute Kraljevo | Vidanovic et al |
| EPI_ISL_541657 | Laboratory Diagnostic, Veterinary Specialized Institute Kraljevo | Laboratory Diagnostic, Veterinary Specialized Institute Kraljevo | Vidanovic et al |
| EPI_ISL_904005 | Scientific Veterinary Institute Novi Sad | Veterinary Specialized Institute Kraljevo, Serbia | Vidanovic et al |
| EPI_ISL_582525 | Veterinary Specialized Institute Kraljevo, Serbia | Veterinary Specialized Institute Kraljevo, Serbia | Vidanovic et al |
| EPI_ISL_904007 | Veterinary Specialized Institute Kraljevo | Veterinary Specialized Institute Kraljevo, Serbia | Vidanovic et al |
| EPI_ISL_462435 | unknown | Laboratory Diagnostic | Vidanovic et al |
| EPI_ISL_514426 | Laboratory Diagnostic, Veterinary Specialized Institute Kraljevo | Laboratory Diagnostic, Veterinary Specialized Institute Kraljevo | Vidanovic et al |
| EPI_ISL_514428 | Laboratory Diagnostic, Veterinary Specialized Institute Kraljevo | Laboratory Diagnostic, Veterinary Specialized Institute Kraljevo | Vidanovic et al |
| EPI_ISL_462438 | unknown | Laboratory Diagnostic | Vidanovic et al |
| EPI_ISL_661189 | Scientific Veterinary Institute Novi Sad | Veterinary Specialized Institute Kraljevo, Serbia | Vidanovic et al |
| EPI_ISL_644572 | Veterinary Specialized Institute Kraljevo, Serbia | Veterinary Specialized Institute Kraljevo, Serbia | Vidanovic et al |
| EPI_ISL_582532 | Veterinary Specialized Institute Kraljevo, Serbia | Veterinary Specialized Institute Kraljevo, Serbia | Vidanovic et al |
| EPI_ISL_904010 | Veterinary Specialized Institute Kraljevo | Veterinary Specialized Institute Kraljevo, Serbia | Vidanovic et al |
| EPI_ISL_541652 | Laboratory Diagnostic, Veterinary Specialized Institute Kraljevo | Laboratory Diagnostic, Veterinary Specialized Institute Kraljevo | Vidanovic et al |
| EPI_ISL_541654 | Laboratory Diagnostic, Veterinary Specialized Institute Kraljevo | Laboratory Diagnostic, Veterinary Specialized Institute Kraljevo | Vidanovic et al |
| EPI_ISL_541661 | Laboratory Diagnostic, Veterinary Specialized Institute Kraljevo | Laboratory Diagnostic, Veterinary Specialized Institute Kraljevo | Vidanovic et al |
| EPI_ISL_833517 | Veterinary Specialized Instute Nis | Veterinary Specialized Institute Kraljevo, Serbia | Vidanovic et al |
| EPI_ISL_661185 | Scientific Veterinary Institute Novi Sad | Veterinary Specialized Institute Kraljevo, Serbia | Vidanovic et al |
| EPI_ISL_676583 | Scientific Veterinary Institute Novi Sad | Veterinary Specialized Institute Kraljevo, Serbia | Vidanovic et al |
| EPI_ISL_661192 | Scientific Veterinary Institute Novi Sad | Veterinary Specialized Institute Kraljevo, Serbia | Vidanovic et al |
| EPI_ISL_904009 | Scientific Veterinary Institute Novi Sad | Veterinary Specialized Institute Kraljevo, Serbia | Vidanovic et al |
| EPI_ISL_676576 | Scientific Veterinary Institute Novi Sad | Veterinary Specialized Institute Kraljevo, Serbia | Vidanovic et al |
| EPI_ISL_676594 | Scientific Veterinary Institute Novi Sad | Veterinary Specialized Institute Kraljevo, Serbia | Vidanovic et al |
| EPI_ISL_661188 | Scientific Veterinary Institute Novi Sad | Veterinary Specialized Institute Kraljevo, Serbia | Vidanovic et al |
| EPI_ISL_833574 | Veterinary Specialized Instute Nis | Veterinary Specialized Institute Sabac, Serbia | Vidanovic et al |
| EPI_ISL_678488 | Veterinary Specialized Institute Sabac, Serbia | Veterinary Specialized Institute Kraljevo, Serbia | Vidanovic et al |
| EPI_ISL_904011 | Veterinary Specialized Institute Kraljevo | Veterinary Specialized Institute Kraljevo, Serbia | Vidanovic et al |
| EPI_ISL_676585 | Scientific Veterinary Institute Novi Sad | Veterinary Specialized Institute Kraljevo, Serbia | Vidanovic et al |

| | | | |
|---|---|---|---|
| EPI_ISL_676589 | Scientific Veterinary Institute Novi Sad | Veterinary Specialized Institute Kraljevo, Serbia | Vidanovic et al |
| EPI_ISL_676595 | Scientific Veterinary Institute Novi Sad | Veterinary Specialized Institute Kraljevo, Serbia | Vidanovic et al |
| EPI_ISL_1626760 | Institute of Virology, Biomedical Research Center of the Slovak Academy of Sciences, Bratislava | Faculty of Natural Sciences, Comenius University, Bratislava | Viktoria Cabanova et al |
| EPI_ISL_959638 | Institute of Virology, Biomedical Research Center of the Slovak Academy of Sciences, Bratislava | Faculty of Natural Sciences, Comenius University, Bratislava | Viktória Čabanová et al |
| EPI_ISL_1239416 | Institute of Virology, Biomedical Research Center of the Slovak Academy of Sciences, Bratislava | Faculty of Natural Sciences, Comenius University, Bratislava | Viktória Čabanová et al |
| EPI_ISL_875538 | Institute of Virology, Biomedical Research Center of the Slovak Academy of Sciences, Bratislava | Faculty of Natural Sciences, Comenius University, Bratislava | Viktória Čabanová et al |
| EPI_ISL_577734 | Institute of Virology, Biomedical Research Center of the Slovak Academy of Sciences, Bratislava | Faculty of Natural Sciences, Comenius University, Bratislava | Viktória Hodorová et al |
| EPI_ISL_577736 | Institute of Virology, Biomedical Research Center of the Slovak Academy of Sciences, Bratislava | Faculty of Natural Sciences, Comenius University, Bratislava | Viktória Hodorová et al |
| EPI_ISL_577738 | Institute of Virology, Biomedical Research Center of the Slovak Academy of Sciences, Bratislava | Faculty of Natural Sciences, Comenius University, Bratislava | Viktória Hodorová et al |
| EPI_ISL_583481 | Institute of Virology, Biomedical Research Center of the Slovak Academy of Sciences, Bratislava | Faculty of Natural Sciences, Comenius University, Bratislava | Viktória Hodorová et al |
| EPI_ISL_1608137 | LBM ALPHABIO, Marseille | LBM ALPHABIO, Marseille | Vincent GARCIA et al |
| EPI_ISL_1608138 | LBM ALPHABIO, Marseille | LBM ALPHABIO, Marseille | Vincent GARCIA et al |
| EPI_ISL_831474 | Laboratório de Microbiologia Molecular - Universidade FEEVALE | Universidade Federal de Ciências da Saúde de Porto Alegre | Vinícius Bonetti Franceschi et al |
| EPI_ISL_831645 | Laboratório de Microbiologia Molecular - Universidade FEEVALE | Universidade Federal de Ciências da Saúde de Porto Alegre | Vinícius Bonetti Franceschi et al |
| EPI_ISL_831646 | Laboratório de Microbiologia Molecular - Universidade FEEVALE | Universidade Federal de Ciências da Saúde de Porto Alegre | Vinícius Bonetti Franceschi et al |
| EPI_ISL_831660 | Laboratório de Microbiologia Molecular - Universidade FEEVALE | Universidade Federal de Ciências da Saúde de Porto Alegre | Vinícius Bonetti Franceschi et al |
| EPI_ISL_831681 | Laboratório de Microbiologia Molecular - Universidade FEEVALE | Universidade Federal de Ciências da Saúde de Porto Alegre | Vinícius Bonetti Franceschi et al |
| EPI_ISL_831683 | Laboratório de Microbiologia Molecular - Universidade FEEVALE | Universidade Federal de Ciências da Saúde de Porto Alegre | Vinícius Bonetti Franceschi et al |
| EPI_ISL_831685 | Laboratório de Microbiologia Molecular - Universidade FEEVALE | Universidade Federal de Ciências da Saúde de Porto Alegre | Vinícius Bonetti Franceschi et al |
| EPI_ISL_831688 | Laboratório de Microbiologia Molecular - Universidade FEEVALE | Universidade Federal de Ciências da Saúde de Porto Alegre | Vinícius Bonetti Franceschi et al |
| EPI_ISL_831689 | Laboratório de Microbiologia Molecular - Universidade FEEVALE | Universidade Federal de Ciências da Saúde de Porto Alegre | Vinícius Bonetti Franceschi et al |
| EPI_ISL_831892 | Laboratório de Microbiologia Molecular - Universidade FEEVALE | Universidade Federal de Ciências da Saúde de Porto Alegre | Vinícius Bonetti Franceschi et al |
| EPI_ISL_831898 | Laboratório de Microbiologia Molecular - Universidade FEEVALE | Universidade Federal de Ciências da Saúde de Porto Alegre | Vinícius Bonetti Franceschi et al |
| EPI_ISL_831913 | Laboratório de Microbiologia Molecular - Universidade FEEVALE | Universidade Federal de Ciências da Saúde de Porto Alegre | Vinícius Bonetti Franceschi et al |
| EPI_ISL_831938 | Laboratório de Microbiologia Molecular - Universidade FEEVALE | Universidade Federal de Ciências da Saúde de Porto Alegre | Vinícius Bonetti Franceschi et al |
| EPI_ISL_831939 | Laboratório de Microbiologia Molecular - Universidade FEEVALE | Universidade Federal de Ciências da Saúde de Porto Alegre | Vinícius Bonetti Franceschi et al |
| EPI_ISL_831940 | Laboratório de Microbiologia Molecular - Universidade FEEVALE | Universidade Federal de Ciências da Saúde de Porto Alegre | Vinícius Bonetti Franceschi et al |

| | | | |
|---|---|---|---|
| EPI_ISL_832009 | Laboratório de Microbiologia Molecular - Universidade FEEVALE | Universidade Federal de Ciências da Saúde de Porto Alegre | Vinícius Bonetti Franceschi et al |
| EPI_ISL_832010 | Laboratório de Microbiologia Molecular - Universidade FEEVALE | Universidade Federal de Ciências da Saúde de Porto Alegre | Vinícius Bonetti Franceschi et al |
| EPI_ISL_832011 | Laboratório de Microbiologia Molecular - Universidade FEEVALE | Universidade Federal de Ciências da Saúde de Porto Alegre | Vinícius Bonetti Franceschi et al |
| EPI_ISL_832012 | Laboratório de Microbiologia Molecular - Universidade FEEVALE | Universidade Federal de Ciências da Saúde de Porto Alegre | Vinícius Bonetti Franceschi et al |
| EPI_ISL_832013 | Laboratório de Microbiologia Molecular - Universidade FEEVALE | Universidade Federal de Ciências da Saúde de Porto Alegre | Vinícius Bonetti Franceschi et al |
| EPI_ISL_831678 | Laboratório de Microbiologia Molecular - Universidade FEEVALE | Universidade Federal de Ciências da Saúde de Porto Alegre | Vinícius Bonetti Franceschi et al |
| EPI_ISL_956391 | Isolation - Virology Unit, Institut Pasteur du Cambodge; Sequencing - US National Institute of Allergy and Infectious Diseases Cambodia, US Naval Medical Research Unit -2, Cambodia National Institute for Public Health | Virology Unit, Institut Pasteur du Cambodge | Vireak Heang et al |
| EPI_ISL_956392 | Isolation - Virology Unit, Institut Pasteur du Cambodge; Sequencing - US National Institute of Allergy and Infectious Diseases Cambodia, US Naval Medical Research Unit -2, Cambodia National Institute for Public Health | Virology Unit, Institut Pasteur du Cambodge | Vireak Heang et al |
| EPI_ISL_956393 | Isolation - Virology Unit, Institut Pasteur du Cambodge; Sequencing - US National Institute of Allergy and Infectious Diseases Cambodia, US Naval Medical Research Unit -2, Cambodia National Institute for Public Health | Virology Unit, Institut Pasteur du Cambodge | Vireak Heang et al |
| EPI_ISL_1469286 | RS Khusus Paru Karawang | National Institute of Health Research and Development | Vivi Setiawaty et al |
| EPI_ISL_1469254 | RS Lira Medika Karawang | National Institute of Health Research and Development | Vivi Setiawaty et al |
| EPI_ISL_1257832 | BTKL-PP Ambon | National Institute of Health Research and Development | Vivi Setiawaty et al |
| EPI_ISL_444610 | U.S. Naval Medical Research Center Biological Defense Research Directorate | U.S. Naval Medical Research Center Biological Defense Research Directorate | Voegtly et al |
| EPI_ISL_406798 | General Hospital of Central Theater Command of People's Liberation Army of China | BGI & Institute of Microbiology, Chinese Academy of Sciences & Shandong First Medical University & Shandong Academy of Medical Sciences & General Hospital of Central Theater Command of People's Liberation Army of China | Weijun Chen et al |
| EPI_ISL_1170957 | HIV Molecular laboratory, Ethiopian Public Health Institute | HIV Molecular Laboratory | Weldemariam et al |
| EPI_ISL_1170958 | HIV Molecular Laboratory, Ethiopian Public Health Institute, Ethiopia | HIV molecular lab, Ethiopian Public Health Institute, Ethiopian | Weldemariam et al |
| EPI_ISL_1169919 | HIV Molecular Laboratory, Ethiopian Public Health Institute | HIV Molecular Laboratory, Ethiopian Public Health Institute | Weldemariam et al |
| EPI_ISL_1170956 | HIV Molecular Lab, Ethiopian Public Health Institute | HIV Molecular Lab | Weldemariam et al |
| EPI_ISL_469254 | National Institute for Viral Disease Control and Prevention, China CDC | Institute of Viral Disease Control and Prevention, China CDC | Wenjie Tan et al |
| EPI_ISL_824656 | Cedars-Sinai Medical Center, Department of Pathology & Laboratory Medicine, Molecular Pathology Laboratory | Cedars-Sinai Medical Center, Molecular Pathology Laboratory of Department of Pathology & Laboratory Medicine and Genomic Core | Wenjuan Zhang et al |
| EPI_ISL_1117388 | Nucleo de Pesquisa em Inovacao Terapeutica - UFPE | LABBE, Federal University of Pernambuco | Wilson Jose da Silva Junior et al |
| EPI_ISL_1117390 | Nucleo de Pesquisa em Inovacao Terapeutica - UFPE | LABBE, Federal University of Pernambuco | Wilson Jose da Silva Junior et al |
| EPI_ISL_1117440 | Nucleo de Pesquisa em Inovacao Terapeutica - UFPE | LABBE, Federal University of Pernambuco | Wilson Jose da Silva Junior et al |
| EPI_ISL_1117399 | Nucleo de Pesquisa em Inovacao Terapeutica - UFPE | LABBE, Federal University of Pernambuco | Wilson Jose da Silva Junior et al |
| EPI_ISL_1117393 | Nucleo de Pesquisa em Inovacao Terapeutica - UFPE | LABBE, Federal University of Pernambuco | Wilson Jose da Silva Junior et al |

| | | | |
|---|---|---|---|
| EPI_ISL_1117395 | Nucleo de Pesquisa em Inovacao Terapeutica - UFPE | LABBE, Federal University of Pernambuco | Wilson Jose da Silva Junior et al |
| EPI_ISL_1117443 | Nucleo de Pesquisa em Inovacao Terapeutica - UFPE | LABBE, Federal University of Pernambuco | Wilson Jose da Silva Junior et al |
| EPI_ISL_1117435 | Nucleo de Pesquisa em Inovacao Terapeutica - UFPE | LABBE, Federal University of Pernambuco | Wilson Jose da Silva Junior et al |
| EPI_ISL_1117428 | Nucleo de Pesquisa em Inovacao Terapeutica - UFPE | LABBE, Federal University of Pernambuco | Wilson Jose da Silva Junior et al |
| EPI_ISL_1117405 | Nucleo de Pesquisa em Inovacao Terapeutica - UFPE | LABBE, Federal University of Pernambuco | Wilson Jose da Silva Junior et al |
| EPI_ISL_1117384 | Nucleo de Pesquisa em Inovacao Terapeutica - UFPE | LABBE, Federal University of Pernambuco | Wilson Jose da Silva Junior et al |
| EPI_ISL_1117386 | Nucleo de Pesquisa em Inovacao Terapeutica - UFPE | LABBE, Federal University of Pernambuco | Wilson Jose da Silva Junior et al |
| EPI_ISL_1117432 | Nucleo de Pesquisa em Inovacao Terapeutica - UFPE | LABBE, Federal University of Pernambuco | Wilson Jose da Silva Junior et al |
| EPI_ISL_1117433 | Nucleo de Pesquisa em Inovacao Terapeutica - UFPE | LABBE, Federal University of Pernambuco | Wilson Jose da Silva Junior et al |
| EPI_ISL_1117434 | Nucleo de Pesquisa em Inovacao Terapeutica - UFPE | LABBE, Federal University of Pernambuco | Wilson Jose da Silva Junior et al |
| EPI_ISL_1117436 | Nucleo de Pesquisa em Inovacao Terapeutica - UFPE | LABBE, Federal University of Pernambuco | Wilson Jose da Silva Junior et al |
| EPI_ISL_1117442 | Nucleo de Pesquisa em Inovacao Terapeutica - UFPE | LABBE, Federal University of Pernambuco | Wilson Jose da Silva Junior et al |
| EPI_ISL_1117404 | Nucleo de Pesquisa em Inovacao Terapeutica - UFPE | LABBE, Federal University of Pernambuco | Wilson Jose da Silva Junior et al |
| EPI_ISL_1117401 | Nucleo de Pesquisa em Inovacao Terapeutica - UFPE | LABBE, Federal University of Pernambuco | Wilson Jose da Silva Junior et al |
| EPI_ISL_1117402 | Nucleo de Pesquisa em Inovacao Terapeutica - UFPE | LABBE, Federal University of Pernambuco | Wilson Jose da Silva Junior et al |
| EPI_ISL_1117379 | Nucleo de Pesquisa em Inovacao Terapeutica - UFPE | LABBE, Federal University of Pernambuco | Wilson Jose da Silva Junior et al |
| EPI_ISL_1117380 | Nucleo de Pesquisa em Inovacao Terapeutica - UFPE | LABBE, Federal University of Pernambuco | Wilson Jose da Silva Junior et al |
| EPI_ISL_1117381 | Nucleo de Pesquisa em Inovacao Terapeutica - UFPE | LABBE, Federal University of Pernambuco | Wilson Jose da Silva Junior et al |
| EPI_ISL_1117382 | Nucleo de Pesquisa em Inovacao Terapeutica - UFPE | LABBE, Federal University of Pernambuco | Wilson Jose da Silva Junior et al |
| EPI_ISL_1117449 | Nucleo de Pesquisa em Inovacao Terapeutica - UFPE | LABBE, Federal University of Pernambuco | Wilson Jose da Silva Junior et al |
| EPI_ISL_1117385 | Nucleo de Pesquisa em Inovacao Terapeutica - UFPE | LABBE, Federal University of Pernambuco | Wilson Jose da Silva Junior et al |
| EPI_ISL_1117429 | Nucleo de Pesquisa em Inovacao Terapeutica - UFPE | LABBE, Federal University of Pernambuco | Wilson Jose da Silva Junior et al |
| EPI_ISL_1117408 | Nucleo de Pesquisa em Inovacao Terapeutica - UFPE | LABBE, Federal University of Pernambuco | Wilson Jose da Silva Junior et al |
| EPI_ISL_1117417 | Nucleo de Pesquisa em Inovacao Terapeutica - UFPE | LABBE, Federal University of Pernambuco | Wilson Jose da Silva Junior et al |
| EPI_ISL_882957 | Specjalistyczny Szpital im. Dra A. Sokołowskiego | National Institute of Public Health - National Institute of Hygiene | Wołkowicz Tomasz et al |
| EPI_ISL_882960 | Laboratorium Diagnostyi Mikrobiologicznej z Pracownią Prątka Gruźlicy SPSzW im. Jana Bożego w Lublinie | National Institute of Public Health - National Institute of Hygiene | Wołkowicz Tomasz et al |
| EPI_ISL_882961 | Laboratorium Diagnostyi Mikrobiologicznej z Pracownią Prątka Gruźlicy SPSzW im. Jana Bożego w Lublinie | National Institute of Public Health - National Institute of Hygiene | Wołkowicz Tomasz et al |
| EPI_ISL_1239619 | Laboratorum Epidemiologii WSSE w Szczecinie | 1. National Institute of Public Health - National Institute of Hygiene; 2. Eurofins Genomics Europe Sequencing GmbH | Wołkowicz Tomasz et al |
| EPI_ISL_1393660 | Dział Diagnostyki Laboratoryjnej SPZOZ | 1. National Institute of Public Health - National Institute of Hygiene; 2. Eurofins Genomics Europe Sequencing GmbH | Wołkowicz Tomasz et al |

| | | | |
|---|---|---|---|
| EPI_ISL_1393683 | Labolatorium Biologi Molekularnej USK | 1. National Institute of Public Health - National Institute of Hygiene; 2. Eurofins Genomics Europe Sequencing GmbH | Wołkowicz Tomasz et al |
| EPI_ISL_1393827 | Laboratorium Medyczne LAB-MED | 1. National Institute of Public Health - National Institute of Hygiene; 2. Eurofins Genomics Europe Sequencing GmbH | Wołkowicz Tomasz et al |
| EPI_ISL_413557 | California Department of Public Health | Chiu Laboratory, University of California, San Francisco | Xianding Deng et al |
| EPI_ISL_435585 | Santa Clara County Public Health Department | Chiu Laboratory, University of California, San Francisco | Xianding Deng et al |
| EPI_ISL_850948 | National Institute for Viral Disease Control and Prevention, China CDC | National Institute for Viral Disease Control and Prevention, China CDC | Xiang Zhao et al |
| EPI_ISL_469255 | National Institute for Viral Disease Control and Prevention, China CDC | Institute of Viral Disease Control and Prevention, China CDC | Xiang Zhao et al |
| EPI_ISL_850946 | National Institute for Viral Disease Control and Prevention, China CDC | National Institute for Viral Disease Control and Prevention, China CDC | Xiang Zhao et al |
| EPI_ISL_850950 | National Institute for Viral Disease Control and Prevention, China CDC | National Institute for Viral Disease Control and Prevention, China CDC | Xiang Zhao et al |
| EPI_ISL_498692 | National Institute for Viral Disease Control and Prevention, China CDC | National Institute for Viral Disease Control and Prevention, China CDC | Xiang Zhao et al |
| EPI_ISL_498694 | National Institute for Viral Disease Control and Prevention, China CDC | National Institute for Viral Disease Control and Prevention, China CDC | Xiang Zhao et al |
| EPI_ISL_498693 | National Institute for Viral Disease Control and Prevention, China CDC | National Institute for Viral Disease Control and Prevention, China CDC | Xiang Zhao et al |
| EPI_ISL_498691 | National Institute for Viral Disease Control and Prevention, China CDC | National Institute for Viral Disease Control and Prevention, China CDC | Xiang Zhao et al |
| EPI_ISL_850949 | National Institute for Viral Disease Control and Prevention, China CDC | National Institute for Viral Disease Control and Prevention, China CDC | Xiang Zhao et al |
| EPI_ISL_962578 | Infectious Disease Control and Prevention Institute | Infectious Disease Control and Prevention Institute | Xiaolin Jiang et al |
| EPI_ISL_962568 | Infectious Disease Control and Prevention Institute | Infectious Disease Control and Prevention Institute | Xiaolin Jiang et al |
| EPI_ISL_962872 | Infectious Disease Control and Prevention Institute | Infectious Disease Control and Prevention Institute | Xiaolin Jiang et al |
| EPI_ISL_962544 | Infectious Disease Control and Prevention Institute | Infectious Disease Control and Prevention Institute | Xiaolin Jiang et al |
| EPI_ISL_962871 | Infectious Disease Control and Prevention Institute | Infectious Disease Control and Prevention Institute | Xiaolin Jiang et al |
| EPI_ISL_962868 | Infectious Disease Control and Prevention Institute | Infectious Disease Control and Prevention Institute | Xiaolin Jiang et al |
| EPI_ISL_548104 | LabTests | Institute of Environmental Science and Research (ESR) | Xiaoyun Ren et al |
| EPI_ISL_548105 | LabTests | Institute of Environmental Science and Research (ESR) | Xiaoyun Ren et al |
| EPI_ISL_579059 | LabPLUS | Institute of Environmental Science and Research (ESR) | Xiaoyun Ren et al |
| EPI_ISL_548110 | LabPLUS | Institute of Environmental Science and Research (ESR) | Xiaoyun Ren et al |
| EPI_ISL_548114 | Middlemore Hospital | Institute of Environmental Science and Research (ESR) | Xiaoyun Ren et al |
| EPI_ISL_548115 | Middlemore Hospital | Institute of Environmental Science and Research (ESR) | Xiaoyun Ren et al |
| EPI_ISL_622788 | Canterbury Health Laboratories | Institute of Environmental Science and Research (ESR) | Xiaoyun Ren et al |
| EPI_ISL_579100 | Middlemore Hospital | Institute of Environmental Science and Research (ESR) | Xiaoyun Ren et al |
| EPI_ISL_579114 | LabPLUS | Institute of Environmental Science and Research (ESR) | Xiaoyun Ren et al |
| EPI_ISL_622774 | LabPLUS | Institute of Environmental Science and Research (ESR) | Xiaoyun Ren et al |
| EPI_ISL_622772 | Middlemore Hospital | Institute of Environmental Science and Research (ESR) | Xiaoyun Ren et al |

| | | | |
|---|---|---|---|
| EPI_ISL_622818 | Middlemore Hospital | Institute of Environmental Science and Research (ESR) | Xiaoyun Ren et al |
| EPI_ISL_661260 | Canterbury Health Laboratories | Institute of Environmental Science and Research (ESR) | Xiaoyun Ren et al |
| EPI_ISL_682291 | LabPLUS | Institute of Environmental Science and Research (ESR) | Xiaoyun Ren et al |
| EPI_ISL_707794 | Waikato Hospital | Institute of Environmental Science and Research (ESR) | Xiaoyun Ren et al |
| EPI_ISL_707800 | LabPLUS | Institute of Environmental Science and Research (ESR) | Xiaoyun Ren et al |
| EPI_ISL_732962 | LabPLUS | Institute of Environmental Science and Research (ESR) | Xiaoyun Ren et al |
| EPI_ISL_755623 | Canterbury Health Laboratories | Institute of Environmental Science and Research (ESR) | Xiaoyun Ren et al |
| EPI_ISL_755626 | PathLab Bay of Plenty | Institute of Environmental Science and Research (ESR) | Xiaoyun Ren et al |
| EPI_ISL_755633 | LabPLUS | Institute of Environmental Science and Research (ESR) | Xiaoyun Ren et al |
| EPI_ISL_755634 | Middlemore Hospital | Institute of Environmental Science and Research (ESR) | Xiaoyun Ren et al |
| EPI_ISL_755635 | Middlemore Hospital | Institute of Environmental Science and Research (ESR) | Xiaoyun Ren et al |
| EPI_ISL_755638 | Canterbury Health Laboratories | Institute of Environmental Science and Research (ESR) | Xiaoyun Ren et al |
| EPI_ISL_579220 | Canterbury Health Laboratories | Institute of Environmental Science and Research (ESR) | Xiaoyun Ren et al |
| EPI_ISL_579403 | LabPLUS | Institute of Environmental Science and Research (ESR) | Xiaoyun Ren et al |
| EPI_ISL_579406 | North Shore Hospital | Institute of Environmental Science and Research (ESR) | Xiaoyun Ren et al |
| EPI_ISL_579407 | North Shore Hospital | Institute of Environmental Science and Research (ESR) | Xiaoyun Ren et al |
| EPI_ISL_579422 | LabTests | Institute of Environmental Science and Research (ESR) | Xiaoyun Ren et al |
| EPI_ISL_579425 | LabPLUS | Institute of Environmental Science and Research (ESR) | Xiaoyun Ren et al |
| EPI_ISL_579426 | Canterbury Health Laboratories | Institute of Environmental Science and Research (ESR) | Xiaoyun Ren et al |
| EPI_ISL_548130 | Middlemore Hospital | Institute of Environmental Science and Research (ESR) | Xiaoyun Ren et al |
| EPI_ISL_548133 | Middlemore Hospital | Institute of Environmental Science and Research (ESR) | Xiaoyun Ren et al |
| EPI_ISL_548138 | Canterbury Health Laboratories | Institute of Environmental Science and Research (ESR) | Xiaoyun Ren et al |
| EPI_ISL_548139 | Canterbury Health Laboratories | Institute of Environmental Science and Research (ESR) | Xiaoyun Ren et al |
| EPI_ISL_548140 | Canterbury Health Laboratories | Institute of Environmental Science and Research (ESR) | Xiaoyun Ren et al |
| EPI_ISL_548145 | Middlemore Hospital | Institute of Environmental Science and Research (ESR) | Xiaoyun Ren et al |
| EPI_ISL_548095 | Middlemore Hospital | Institute of Environmental Science and Research (ESR) | Xiaoyun Ren et al |
| EPI_ISL_843193 | LabPLUS | Institute of Environmental Science and Research (ESR) | Xiaoyun Ren et al |
| EPI_ISL_877209 | LabPLUS | Institute of Environmental Science and Research (ESR) | Xiaoyun Ren et al |
| EPI_ISL_877212 | LabPLUS | Institute of Environmental Science and Research (ESR) | Xiaoyun Ren et al |
| EPI_ISL_877216 | Middlemore Hospital | Institute of Environmental Science and Research (ESR) | Xiaoyun Ren et al |

| | | | |
|---|---|---|---|
| EPI_ISL_877217 | Middlemore Hospital | Institute of Environmental Science and Research (ESR) | Xiaoyun Ren et al |
| EPI_ISL_877221 | LabPLUS | Institute of Environmental Science and Research (ESR) | Xiaoyun Ren et al |
| EPI_ISL_877226 | LabPLUS | Institute of Environmental Science and Research (ESR) | Xiaoyun Ren et al |
| EPI_ISL_1016868 | Middlemore Hospital | Institute of Environmental Science and Research (ESR) | Xiaoyun Ren et al |
| EPI_ISL_1016874 | Middlemore Hospital | Institute of Environmental Science and Research (ESR) | Xiaoyun Ren et al |
| EPI_ISL_1016847 | LabPLUS | Institute of Environmental Science and Research (ESR) | Xiaoyun Ren et al |
| EPI_ISL_1016846 | LabPLUS | Institute of Environmental Science and Research (ESR) | Xiaoyun Ren et al |
| EPI_ISL_1016869 | LabPLUS | Institute of Environmental Science and Research (ESR) | Xiaoyun Ren et al |
| EPI_ISL_1016865 | Middlemore Hospital | Institute of Environmental Science and Research (ESR) | Xiaoyun Ren et al |
| EPI_ISL_1016873 | LabPLUS | Institute of Environmental Science and Research (ESR) | Xiaoyun Ren et al |
| EPI_ISL_1016872 | Middlemore Hospital | Institute of Environmental Science and Research (ESR) | Xiaoyun Ren et al |
| EPI_ISL_1016844 | Middlemore Hospital | Institute of Environmental Science and Research (ESR) | Xiaoyun Ren et al |
| EPI_ISL_1016871 | LabPLUS | Institute of Environmental Science and Research (ESR) | Xiaoyun Ren et al |
| EPI_ISL_1016854 | LabPLUS | Institute of Environmental Science and Research (ESR) | Xiaoyun Ren et al |
| EPI_ISL_476827 | Laboratoire des Fièvres Hémorragiques Virales du Benin | Charité-Universitätsmedizin Berlin | Yadouleton et al |
| EPI_ISL_476830 | Laboratoire des Fièvres Hémorragiques Virales du Benin | Charité-Universitätsmedizin Berlin | Yadouleton et al |
| EPI_ISL_476831 | Laboratoire des Fièvres Hémorragiques Virales du Benin | Charité-Universitätsmedizin Berlin | Yadouleton et al |
| EPI_ISL_476823 | Laboratoire des Fièvres Hémorragiques Virales du Benin | Charité-Universitätsmedizin Berlin | Yadouleton et al |
| EPI_ISL_476834 | Laboratoire des Fièvres Hémorragiques Virales du Benin | Charité-Universitätsmedizin Berlin | Yadouleton et al |
| EPI_ISL_476833 | Laboratoire des Fièvres Hémorragiques Virales du Benin | Charité-Universitätsmedizin Berlin | Yadouleton et al |
| EPI_ISL_476829 | Laboratoire des Fièvres Hémorragiques Virales du Benin | Charité-Universitätsmedizin Berlin | Yadouleton et al |
| EPI_ISL_1225337 | Gorgas Memorial Laboratory of Health Studies | Gorgas Memorial Laboratory of Health Studies | Yamilka Diaz et al |
| EPI_ISL_1225364 | Gorgas Memorial Laboratory of Health Studies | Gorgas Memorial Laboratory of Health Studies | Yamilka Diaz et al |
| EPI_ISL_1225373 | Gorgas Memorial Laboratory of Health Studies | Gorgas Memorial Laboratory of Health Studies | Yamilka Diaz et al |
| EPI_ISL_1225382 | Gorgas Memorial Laboratory of Health Studies | Gorgas Memorial Laboratory of Health Studies | Yamilka Diaz et al |
| EPI_ISL_1225391 | Gorgas Memorial Laboratory of Health Studies | Gorgas Memorial Laboratory of Health Studies | Yamilka Diaz et al |
| EPI_ISL_1225424 | Gorgas Memorial Laboratory of Health Studies | Gorgas Memorial Laboratory of Health Studies | Yamilka Diaz et al |
| EPI_ISL_1225429 | Gorgas Memorial Laboratory of Health Studies | Gorgas Memorial Laboratory of Health Studies | Yamilka Diaz et al |
| EPI_ISL_1225463 | Gorgas Memorial Laboratory of Health Studies | Gorgas Memorial Laboratory of Health Studies | Yamilka Diaz et al |
| EPI_ISL_1225481 | Gorgas Memorial Laboratory of Health Studies | Gorgas Memorial Laboratory of Health Studies | Yamilka Diaz et al |

| | | | |
|---|---|---|---|
| EPI_ISL_1225490 | Gorgas Memorial Laboratory of Health Studies | Gorgas Memorial Laboratory of Health Studies | Yamilka Diaz et al |
| EPI_ISL_1225491 | Gorgas Memorial Laboratory of Health Studies | Gorgas Memorial Laboratory of Health Studies | Yamilka Diaz et al |
| EPI_ISL_1225534 | Gorgas Memorial Laboratory of Health Studies | Gorgas Memorial Laboratory of Health Studies | Yamilka Diaz et al |
| EPI_ISL_1225537 | Gorgas Memorial Laboratory of Health Studies | Gorgas Memorial Laboratory of Health Studies | Yamilka Diaz et al |
| EPI_ISL_1225555 | Gorgas Memorial Laboratory of Health Studies | Gorgas Memorial Laboratory of Health Studies | Yamilka Diaz et al |
| EPI_ISL_1225556 | Gorgas Memorial Laboratory of Health Studies | Gorgas Memorial Laboratory of Health Studies | Yamilka Diaz et al |
| EPI_ISL_1225560 | Gorgas Memorial Laboratory of Health Studies | Gorgas Memorial Laboratory of Health Studies | Yamilka Diaz et al |
| EPI_ISL_1225572 | Gorgas Memorial Laboratory of Health Studies | Gorgas Memorial Laboratory of Health Studies | Yamilka Diaz et al |
| EPI_ISL_1225574 | Gorgas Memorial Laboratory of Health Studies | Gorgas Memorial Laboratory of Health Studies | Yamilka Diaz et al |
| EPI_ISL_1225577 | Gorgas Memorial Laboratory of Health Studies | Gorgas Memorial Laboratory of Health Studies | Yamilka Diaz et al |
| EPI_ISL_450793 | Jamaica Ministry of Health and Wellness | Pathogen Discovery, Respiratory Viruses Branch, Division of Viral Diseases, Centers for Disease Control and Prevention | Yan Li et al |
| EPI_ISL_450794 | Jamaica Ministry of Health and Wellness | Pathogen Discovery, Respiratory Viruses Branch, Division of Viral Diseases, Centers for Disease Control and Prevention | Yan Li et al |
| EPI_ISL_450795 | Jamaica Ministry of Health and Wellness | Pathogen Discovery, Respiratory Viruses Branch, Division of Viral Diseases, Centers for Disease Control and Prevention | Yan Li et al |
| EPI_ISL_450796 | Jamaica Ministry of Health and Wellness | Pathogen Discovery, Respiratory Viruses Branch, Division of Viral Diseases, Centers for Disease Control and Prevention | Yan Li et al |
| EPI_ISL_450797 | Jamaica Ministry of Health and Wellness | Pathogen Discovery, Respiratory Viruses Branch, Division of Viral Diseases, Centers for Disease Control and Prevention | Yan Li et al |
| EPI_ISL_450799 | Jamaica Ministry of Health and Wellness | Pathogen Discovery, Respiratory Viruses Branch, Division of Viral Diseases, Centers for Disease Control and Prevention | Yan Li et al |
| EPI_ISL_1591094 | NAMRU-6 | Pathogen Discovery, Respiratory Viruses Branch, Division of Viral Diseases, Centers for Disease Control and Prevention | Yan Li et al |
| EPI_ISL_1591098 | NAMRU-6 | Pathogen Discovery, Respiratory Viruses Branch, Division of Viral Diseases, Centers for Disease Control and Prevention | Yan Li et al |
| EPI_ISL_1591097 | NAMRU-6 | Pathogen Discovery, Respiratory Viruses Branch, Division of Viral Diseases, Centers for Disease Control and Prevention | Yan Li et al |
| EPI_ISL_1591096 | NAMRU-6 | Pathogen Discovery, Respiratory Viruses Branch, Division of Viral Diseases, Centers for Disease Control and Prevention | Yan Li et al |
| EPI_ISL_1591095 | NAMRU-6 | Pathogen Discovery, Respiratory Viruses Branch, Division of Viral Diseases, Centers for Disease Control and Prevention | Yan Li et al |
| EPI_ISL_410045 | IL Department of Public Health Chicago Laboratory | Pathogen Discovery, Respiratory Viruses Branch, Division of Viral Diseases, Centers for Disease Control and Prevention | Yan Li et al |
| EPI_ISL_1117378 | Beijing Center for Disease Prevention and Control | Beijing Center for Disease Prevention and Control | Yang Pan et al |
| EPI_ISL_1117377 | Beijing Center for Disease Prevention and Control | Beijing Center for Disease Prevention and Control | Yang Pan et al |
| EPI_ISL_1117376 | Beijing Center for Disease Prevention and Control | Beijing Center for Disease Prevention and Control | Yang Pan et al |
| EPI_ISL_1121993 | Emergency department of COVID-19, Beijing Ditan Hospital | National Institute for Viral Disease Control and Prevention | Yang Song2* et al |

| | | | |
|---|---|---|---|
| EPI_ISL_1122015 | Emergency department of COVID-19, Beijing Ditan Hospital | National Institute for Viral Disease Control and Prevention | Yang Song2* et al |
| EPI_ISL_1122016 | Emergency department of COVID-19, Beijing Ditan Hospital | National Institute for Viral Disease Control and Prevention | Yang Song2* et al |
| EPI_ISL_1122017 | Emergency department of COVID-19, Beijing Ditan Hospital, Capital Medical University, Beijing, China. | National Institute for Viral Disease Control and Prevention | Yang Song2* et al |
| EPI_ISL_548945 | Institute of Microbiology, University of Veterinary and Animal sciences | Institute of Microbiology, University of Veterinary and Animal sciences | Yaqub et al |
| EPI_ISL_509695 | Guatemala Ministry of Public Health | Pathogen Discovery, Respiratory Viruses Branch, Division of Viral Diseases, Centers for Disease Control and Prevention | Ying Tao et al |
| EPI_ISL_509696 | Guatemala Ministry of Public Health | Pathogen Discovery, Respiratory Viruses Branch, Division of Viral Diseases, Centers for Disease Control and Prevention | Ying Tao et al |
| EPI_ISL_509700 | Guatemala Ministry of Public Health | Pathogen Discovery, Respiratory Viruses Branch, Division of Viral Diseases, Centers for Disease Control and Prevention | Ying Tao et al |
| EPI_ISL_509701 | Guatemala Ministry of Public Health | Pathogen Discovery, Respiratory Viruses Branch, Division of Viral Diseases, Centers for Disease Control and Prevention | Ying Tao et al |
| EPI_ISL_509702 | Guatemala Ministry of Public Health | Pathogen Discovery, Respiratory Viruses Branch, Division of Viral Diseases, Centers for Disease Control and Prevention | Ying Tao et al |
| EPI_ISL_509703 | Guatemala Ministry of Public Health | Pathogen Discovery, Respiratory Viruses Branch, Division of Viral Diseases, Centers for Disease Control and Prevention | Ying Tao et al |
| EPI_ISL_406223 | Arizona Department of Health Services | Pathogen Discovery, Respiratory Viruses Branch, Division of Viral Diseases, Centers for Disease Control and Prevention | Ying Tao et al |
| EPI_ISL_408010 | California Department of Health | Pathogen Discovery, Respiratory Viruses Branch, Division of Viral Diseases, Centers for Disease Control and Prevention | Ying Tao et al |
| EPI_ISL_404253 | IL Department of Public Health Chicago Laboratory | Pathogen Discovery, Respiratory Viruses Branch, Division of Viral Diseases, Centers for Disease Control and Prevention | Ying Tao et al |
| EPI_ISL_501197 | Department of Medical Microbiology, University Malaya Medical Centre | Department of Medical Microbiology, Faculty of Medicine, University of Malaya | Yoong Min CHONG et al |
| EPI_ISL_506998 | Department of Medical Microbiology, University Malaya Medical Centre | Department of Medical Microbiology, Faculty of Medicine, University of Malaya | Yoong Min CHONG et al |
| EPI_ISL_501202 | Department of Medical Microbiology, University Malaya Medical Centre | Department of Medical Microbiology, Faculty of Medicine, University of Malaya | Yoong Min CHONG et al |
| EPI_ISL_501208 | Department of Medical Microbiology, University Malaya Medical Centre | Department of Medical Microbiology, Faculty of Medicine, University of Malaya | Yoong Min CHONG et al |
| EPI_ISL_501211 | Department of Medical Microbiology, University Malaya Medical Centre | Department of Medical Microbiology, Faculty of Medicine, University of Malaya | Yoong Min CHONG et al |
| EPI_ISL_506999 | Department of Medical Microbiology, University Malaya Medical Centre | Department of Medical Microbiology, Faculty of Medicine, University of Malaya | Yoong Min CHONG et al |
| EPI_ISL_501213 | Department of Medical Microbiology, University Malaya Medical Centre | Department of Medical Microbiology, Faculty of Medicine, University of Malaya | Yoong Min CHONG et al |
| EPI_ISL_501216 | Department of Medical Microbiology, University Malaya Medical Centre | Department of Medical Microbiology, Faculty of Medicine, University of Malaya | Yoong Min CHONG et al |
| EPI_ISL_501217 | Department of Medical Microbiology, University Malaya Medical Centre | Department of Medical Microbiology, Faculty of Medicine, University of Malaya | Yoong Min CHONG et al |
| EPI_ISL_501220 | Department of Medical Microbiology, University Malaya Medical Centre | Department of Medical Microbiology, Faculty of Medicine, University of Malaya | Yoong Min CHONG et al |

| | | | |
|---|---|---|---|
| EPI_ISL_507000 | Department of Medical Microbiology, University Malaya Medical Centre | Department of Medical Microbiology, Faculty of Medicine, University of Malaya | Yoong Min CHONG et al |
| EPI_ISL_1595847 | Genome Analysis Center, Yamanashi Central Hospital | Genome Analysis Center, Yamanashi Central Hospital | Yosuke Hirotsu et al |
| EPI_ISL_1595849 | Genome Analysis Center, Yamanashi Central Hospital | Genome Analysis Center, Yamanashi Central Hospital | Yosuke Hirotsu et al |
| EPI_ISL_1595851 | Genome Analysis Center, Yamanashi Central Hospital | Genome Analysis Center, Yamanashi Central Hospital | Yosuke Hirotsu et al |
| EPI_ISL_1669765 | Genome Analysis Center, Yamanashi Central Hospital | Genome Analysis Center, Yamanashi Central Hospital | Yosuke Hirotsu et al |
| EPI_ISL_925850 | Nucleic Acid Testing, National Reference Laboratory | GIGA Medical Genomics | Yvan Butera et al |
| EPI_ISL_925851 | Nucleic Acid Testing, National Reference Laboratory | GIGA Medical Genomics | Yvan Butera et al |
| EPI_ISL_925852 | Nucleic Acid Testing, National Reference Laboratory | GIGA Medical Genomics | Yvan Butera et al |
| EPI_ISL_925883 | Nucleic Acid Testing, National Reference Laboratory | GIGA Medical Genomics | Yvan Butera et al |
| EPI_ISL_925898 | Nucleic Acid Testing, National Reference Laboratory | GIGA Medical Genomics | Yvan Butera et al |
| EPI_ISL_925901 | Nucleic Acid Testing, National Reference Laboratory | GIGA Medical Genomics | Yvan Butera et al |
| EPI_ISL_925902 | Nucleic Acid Testing, National Reference Laboratory | GIGA Medical Genomics | Yvan Butera et al |
| EPI_ISL_925904 | Nucleic Acid Testing, National Reference Laboratory | GIGA Medical Genomics | Yvan Butera et al |
| EPI_ISL_1020288 | Nucleic Acid Testing, National Reference Laboratory | GIGA Medical Genomics | Yvan Butera et al |
| EPI_ISL_960228 | Nucleic Acid Testing, National Reference Laboratory | GIGA Medical Genomics | Yvan Butera et al |
| EPI_ISL_960278 | Nucleic Acid Testing, National Reference Laboratory | GIGA Medical Genomics | Yvan Butera et al |
| EPI_ISL_960302 | Nucleic Acid Testing, National Reference Laboratory | GIGA Medical Genomics | Yvan Butera et al |
| EPI_ISL_960301 | Nucleic Acid Testing, National Reference Laboratory | GIGA Medical Genomics | Yvan Butera et al |
| EPI_ISL_960300 | Nucleic Acid Testing, National Reference Laboratory | GIGA Medical Genomics | Yvan Butera et al |
| EPI_ISL_960288 | Nucleic Acid Testing, National Reference Laboratory | GIGA Medical Genomics | Yvan Butera et al |
| EPI_ISL_1301779 | Nucleic Acid Testing, National Reference Laboratory | GIGA Medical Genomics | Yvan Butera et al |
| EPI_ISL_1301784 | Nucleic Acid Testing, National Reference Laboratory | GIGA Medical Genomics | Yvan Butera et al |
| EPI_ISL_1301790 | Nucleic Acid Testing, National Reference Laboratory | GIGA Medical Genomics | Yvan Butera et al |
| EPI_ISL_1301741 | Nucleic Acid Testing, National Reference Laboratory | GIGA Medical Genomics | Yvan Butera et al |
| EPI_ISL_1301793 | Nucleic Acid Testing, National Reference Laboratory | GIGA Medical Genomics | Yvan Butera et al |
| EPI_ISL_1301742 | Nucleic Acid Testing, National Reference Laboratory | GIGA Medical Genomics | Yvan Butera et al |
| EPI_ISL_1301794 | Nucleic Acid Testing, National Reference Laboratory | GIGA Medical Genomics | Yvan Butera et al |
| EPI_ISL_1301736 | Nucleic Acid Testing, National Reference Laboratory | GIGA Medical Genomics | Yvan Butera et al |
| EPI_ISL_625683 | National Reference Laboratory for COVID-19, Pasteur Institute of Iran | National Reference Laboratory for COVID-19, Pasteur Institute of Iran | Zahra Ahmadi et al |
| EPI_ISL_833578 | National Reference Laboratory for COVID-19, Pasteur Institute of Iran | National Reference Laboratory for COVID-19, Pasteur Institute of Iran | Zahra Ahmadi et al |

| | | | |
|---|---|---|---|
| EPI_ISL_1139147 | National Center of Disease Control and Prevention of the Republic of Armenia | Institute of Molecular Biology NAS RA, Republic of Armenia, Department of Bioengineering, BioinformaticsInstitute and Molecular Biology IBMPh RAU, Republic of Armenia | Zaven Karalyan et al |
| EPI_ISL_1299979 | OKMI FN Brno | CEITEC MU | Zdrazilova Dubska L. et al |
| EPI_ISL_1300005 | OKMI FN Brno | CEITEC MU | Zdrazilova Dubska L. et al |
| EPI_ISL_468062 | Egyptian National Cancer Institute (ENCI) | Egyptian National Cancer Institute (ENCI) | Zekri et al |
| EPI_ISL_907096 | Cancer Biology Department, National Cancer Institute | Cancer Biology Department, National Cancer Institute | Zekri et al |
| EPI_ISL_907097 | Cancer Biology Department, National Cancer Institute | Cancer Biology Department, National Cancer Institute | Zekri et al |
| EPI_ISL_907099 | Cancer Biology Department, National Cancer Institute | Cancer Biology Department, National Cancer Institute | Zekri et al |
| EPI_ISL_475724 | Egyptian National Cancer Institute (ENCI) | Egyptian National Cancer Institute (ENCI) | Zekri et al |
| EPI_ISL_857332 | Cancer Biology Department, National Cancer Institute | Cancer Biology Department, National Cancer Institute | Zekri et al |
| EPI_ISL_857330 | Cancer Biology Department, National Cancer Institute | Cancer Biology Department, National Cancer Institute | Zekri et al |
| EPI_ISL_857336 | Cancer Biology Department, National Cancer Institute | Cancer Biology Department, National Cancer Institute | Zekri et al |
| EPI_ISL_857347 | Cancer Biology Department, National Cancer Institute | Cancer Biology Department, National Cancer Institute | Zekri et al |
| EPI_ISL_402125 | National Institute for Communicable Disease Control and Prevention (ICDC) Chinese Center for Disease Control and Prevention (China CDC) | National Institute for Communicable Disease Control and Prevention (ICDC) Chinese Center for Disease Control and Prevention (China CDC) | Zhang et al |
| EPI_ISL_430847 | HS mikrobiologi virus | The Public Health Agency of Sweden | Zhibing Yun et al |
| EPI_ISL_1055106 | National Virus Reference Laboratory | National Virus Reference Laboratory | Zoe Yandle et al |
| EPI_ISL_1180385 | National Virus Reference Laboratory | National Virus Reference Laboratory | Zoe Yandle et al |
| EPI_ISL_1180388 | National Virus Reference Laboratory | National Virus Reference Laboratory | Zoe Yandle et al |
| EPI_ISL_1623011 | National Virus Reference Laboratory | National Virus Reference Laboratory | Zoe Yandle et al |
| EPI_ISL_1623005 | National Virus Reference Laboratory | National Virus Reference Laboratory | Zoe Yandle et al |
| EPI_ISL_1623006 | National Virus Reference Laboratory | National Virus Reference Laboratory | Zoe Yandle et al |
| EPI_ISL_1552560 | National Virus Reference Laboratory | National Virus Reference Laboratory | Zoe Yandle et al |
| EPI_ISL_1117702 | National Virus Reference Laboratory | National Virus Reference Laboratory | Zoe Yandle et al |
| EPI_ISL_1657504 | National Virus Reference Laboratory | National Virus Reference Laboratory | Zoe Yandle et al |
| EPI_ISL_1657422 | National Virus Reference Laboratory | National Virus Reference Laboratory | Zoe Yandle et al |
| EPI_ISL_1164141 | National Virus Reference Laboratory | National Virus Reference Laboratory | Zoe Yandle et al |
| EPI_ISL_1191448 | National Virus Reference Laboratory | National Virus Reference Laboratory | Zoe Yandle et al |
| EPI_ISL_1577467 | National Virus Reference Laboratory | National Virus Reference Laboratory | Zoe Yandle et al |
| EPI_ISL_1577559 | National Virus Reference Laboratory | National Virus Reference Laboratory | Zoe Yandle et al |
| EPI_ISL_594188 | Department of Pathology, School of Medicine, Imam Khomeini Hospital, Tehran University of Medical Sciences | Genetics Research Center, University of Social Welfare and Rehabilitation Sciences | Zohreh Fattahi et al |

| | | | |
|---|---|---|---|
| EPI_ISL_596451 | Department of Pathology, School of Medicine, Imam Khomeini Hospital, Tehran University of Medical Sciences | Genetics Research Center, University of Social Welfare and Rehabilitation Sciences | Zohreh Fattahi et al |
| EPI_ISL_596454 | Infectious Disease and Tropical Medicine Research Center, Resistant Tuberculosis Institute, Zahedan University of Medical Sciences, Zahedan, Iran. | Genetics Research Center, University of Social Welfare and Rehabilitation Sciences | Zohreh Fattahi et al |
| EPI_ISL_596452 | Department of Pathology, School of Medicine, Imam Khomeini Hospital, Tehran University of Medical Sciences | Genetics Research Center, University of Social Welfare and Rehabilitation Sciences | Zohreh Fattahi et al |
| EPI_ISL_596455 | Department of Pathology, School of Medicine, Imam Khomeini Hospital, Tehran University of Medical Sciences | Genetics Research Center, University of Social Welfare and Rehabilitation Sciences | Zohreh Fattahi et al |
